# Genomic Epidemiology of SARS-CoV-2 in Seychelles, 2020-2021

**DOI:** 10.1101/2022.03.18.22272503

**Authors:** John Mwita Morobe, Brigitte Pool, Lina Marie, Dwayne Didon, Arnold W. Lambisia, Timothy Makori, Khadija Said Mohammed, Zaydah R. de Laurent, Leonard Ndwiga, Maureen W. Mburu, Edidah Moraa, Nickson Murunga, Jennifer Musyoki, Jedida Mwacharo, Lydia Nyamako, Debra Riako, Pariken Ephnatus, Faith Gambo, Josephine Naimani, Joyce Namulondo, Susan Zimba Tembo, Edwin Ogendi, Thierno Balde, Fred Athanasius Dratibi, Yahaya Ali Ahmed, Nicksy Gumede, Rachel A. Achilla, Peter K. Borus, Dorcas Wanjohi, Sofonias K. Tessema, Joseph Mwangangi, Philip Bejon, D. James Nokes, Lynette Isabella Ochola-Oyier, George Githinji, Leon Biscornet, Charles N. Agoti

## Abstract

Seychelles, an archipelago of 155 islands in the Indian Ocean, had confirmed 24,788 cases of severe acute respiratory syndrome coronavirus 2 (SARS-CoV-2) by the 31^st^ December 2021. The first SARS-CoV-2 cases in Seychelles were reported on the 14^th^ of March 2020, but cases remained low until January 2021, when a surge of SARS-CoV-2 cases was observed on the islands. Here, we investigated the potential drivers of the surge by genomic analysis 1,056 SARS-CoV-2 positive samples collected in Seychelles between 14^th^ March 2020 and 31^st^ December 2021. The Seychelles genomes were classified into 32 Pango lineages, 1,042 of which fell within four variants of concern i.e., Alpha, Beta, Delta and Omicron. Sporadic cases of SARS-CoV-2 detected in Seychelles in 2020 were mainly of lineage B.1 (European origin) but this lineage was rapidly replaced by Beta variant starting January 2021, and which was also subsequently replaced by the Delta variant in May 2021 that dominated till November 2021 when Omicron cases were identified. Using ancestral state reconstruction approach, we estimated at least 78 independent SARS-CoV-2 introduction events into Seychelles during the study period. Majority of viral introductions into Seychelles occurred in 2021, despite substantial COVID-19 restrictions in place during this period. We conclude that the surge of SARS-CoV-2 cases in Seychelles in January 2021 was primarily due to the introduction of more transmissible SARS-CoV-2 variants into the islands.

## BACKGROUND

Seychelles, an archipelago of 155 islands in the Indian Ocean with a population size of approximately 99,202^1^, confirmed its first cases of severe acute respiratory syndrome coronavirus 2 (SARS-CoV-2), the aetiological agent of coronavirus disease 2019 (COVID-19), on the 14^th^ March 2020. This was shortly after the World Health Organisation (WHO) declared COVID-19 a global pandemic on the 11^th^ March 2020. However, the number of COVID-19 cases in Seychelles remained low (average of 1 case/day) until January 2021 when a surge was observed in the country (average of 72 cases/day). As of 31^st^ December 2021, Seychelles had reported 24,788 laboratory-confirmed COVID-19 cases, >98% of which were recorded in 2021^2^. The surge of the number of COVID-19 cases in Seychelles in 2021 could be due to 2 major factors: (a) the relaxation of government COVID-19 stringency measures and (b) the arrival of more transmissible SARS-CoV-2 variants on the islands. Our analysis looked at these factors in an attempt to improve understanding of the COVID-19 transmission dynamics in Seychelles.

The first round of COVID-19 countermeasures by the Seychelles government to curb introductions and spread of the virus were announced in March 2020. These measures included: a 14-day quarantine for people returning from countries with significant COVID-19 community transmission on the 16^th^ March 2020, closure of day care centres and learning institutions, and ban on international arrivals and foreign travel by Seychellois citizens, except for medical emergencies beginning 23^rd^ March 2020, a 21-day nationwide lockdown, tracing, isolation and monitoring of all persons who had close contact with COVID-19 patients for 14 days beginning 6^th^ April 2020, closure of all shops except those that sell food items, groceries, pharmaceutical products beginning 6^th^ April 2020, workplace closures and restriction of outdoor movement except for essential services on 9^th^ April. On the 4^th^ of May 2020, the Seychelles government eased some of the COVID-19 restrictions, including opening of all day care and learning institutions, opening of all shops, and lifting of the ban on restrictions of movement of people. In June 2020, the Seychelles government lifted the ban on international travel and allowed visitors (international tourists) from countries categorised as low-risk countries, however with a requirement to show a COVID-19 negative certificate (RT-PCR test). Despite this removal of many of the Government restrictions, the number of SARS-CoV-2 cases in Seychelles throughout 2020 remained low.

Towards the end of 2020 and in 2021, in widely different geographical locations, five SARS-CoV-2 variants of concern (VOC) - Alpha, Beta, Gamma, Delta and Omicron, five variants of variants of interest (VOI) – Eta, Kappa, Iota, Epsilon and Theta and over 10 variants under monitoring (VUM) - emerged that appeared to be considerably more transmissible and with potential to escape pre-existing immunity^3,4^ Notably, the surge of COVID-19 cases in Seychelles in early 2021 coincided with their emergence. Further, in the last quarter of 2021, the Omicron SARS-CoV-2 variants of concern (VOCs) was detected globally^4^. The emergence and spread of these variants of concern globally, has exacerbated the effect of the COVID-19 pandemic^5^. The objective of this study was to describe the genomic epidemiology of SARS-CoV-2 in Seychelles and in particular lineages coinciding with the surge of cases that began in January 2021, with the aim of improving the understanding on the introduction and transmission of SARS-CoV-2 in Seychelles.

## MATERIALS AND METHODS

### Ethical statement

The SARS-CoV-2 positive samples were sequenced at the Kenya Medical Research Institute (KEMRI) - Wellcome Trust Research Programme (KWTRP) as part of a regional collaborative COVID-19 public health rapid response initiative overseen by WHO-AFRO and Africa-CDC. KWTRP Kilifi is one of the 12 designated WHO-AFRO/Africa-CDC regional reference laboratories for SARS-CoV-2 genomic surveillance in Africa. The whole genome sequencing study protocol was reviewed and approved by the Scientific and Ethics Review Committee (SERU) at KEMRI, (SERU #4035). Individual patient consent requirement was waivered by the committee as the sequenced samples were part of the public health emergency response.

### Study site and samples

A total of 1,298 SARS-CoV-2 real-time polymerase chain reaction (qRT-PCR)-confirmed nasopharyngeal and oropharyngeal (NP/OP) positive swab samples collected between 14^th^ March 2020 and 31^st^ December 2021 were targeted for whole genome sequencing. The samples received for sequencing at KWTRP for whole genome sequencing were selected considering cycle threshold (Ct) value cut off <30. The monthly temporal distribution of samples selected for whole genome sequencing is shown in **Supplementary Figure 1**.

### Laboratory procedures

#### RNA extraction, cDNA synthesis and amplification

The NP/OP swab samples on arrival at KWTRP laboratories were re-extracted using the QIAamp Viral RNA Mini Kit (Qiagen, Manchester, UK) following the manufacturer’s instructions, starting volume 140 µl, and elution volume of 60 μl. The RNA was then re-assayed to confirm SARS-CoV-2 genetic material using one of three commercial kits namely, Da An Gene Co. Ltd.’s Detection Kit (targeting N gene or ORF1ab), SD Biosensor’s Standard M Real Time Detection Kit (targeting E gene and ORF1ab) or KH Medical’s RADI COVID-19 Detection Kit (targeting RdRp and S genes) while following manufacturer’s instructions. Samples with Ct values <33 were selected for cDNA synthesis.

Extracted RNA was reverse transcribed using the LunaScript® RT SuperMix Kit (New England Biolabs, US). For each of the selected sample, 2 μl of the LunaScript® RT SuperMix was added to 8 μl of RNA template, incubated at 25°C for 2 minutes, 55°C for 10 minutes, held at 95°C for 1 minute and placed on ice for 1 minute. The generated viral cDNA was amplified using the Q5® Hot Start High-Fidelity 2X Master Mix (NEB, US) along with ARTIC nCoV-2019 version 3 primers (primer pools A and B), as documented previously^6^. The thermocycling conditions involved a touchdown PCR with the following conditions: heat activation at 98°C for 30 seconds, followed by 40 amplification cycles (i.e., 25 cycles of 98°C for 15 seconds and 65°C for 5 minutes, and 15 cycles of 62.5°C for 5 minutes and 98°C for 15 seconds and one cycle at 62.5°C for 5 minutes), final extension at 62.5°C for 5 minutes, followed by a final hold at 4°C. To overcome amplicon dropouts in regions 3, 9, 17, 26, 64, 66, 67, 68, 74 88, 91 and 92 of the genomes^6^, primer pairs for the aforementioned regions were constituted in an additional pool, named herein pool C. After the multiplex PCR, an agarose gel electrophoresis step was included to exclude samples with no visible bands from further processing.

#### Oxford Nanopore library preparation and sequencing

For each sample, the PCR products of primer pools A, B and C were combined to make a total of 23μl (all of pool A (10μl), pool B (10μl) and pool C(3µl)) and cleaned using 1x AMPure XP beads (Beckman Coulter, US), followed by two ethanol (80%) washes. The pellet was resuspended in 20 μl nuclease-free water and 1 μl of the eluted sample was quantified using the Qubit dsDNA HS Assay Kit (ThermoFisher Scientific, US). End-prep reaction was performed according to the ARTIC nCoV-2019 sequencing protocol v3 (LoCost) with 200 fmol (50ng) of amplicons and the NEBNext Ultra II End repair/dA-tailing Kit (NEB, US) and incubated at 20°C for 5 minutes and 65°C for 5 minutes. From this, 1μl of DNA was used for barcode ligation using Native Barcoding Expansion 96 (Oxford Nanopore Technology, UK) and NEBNext Ultra II Ligation Module reagents (NEB, US). Incubation was performed at 20°C for 20 minutes and at 65°C for 10 minutes. This step was eventually modified to employ NEBNext Blunt/TA Ligase Master Mix (NEB, US) using the same barcodes and incubation conditions.

The barcoded samples were pooled together. The pooled and barcoded DNA samples were cleaned using 0.4X AMPure XP beads followed by two ethanol (80%) washes and eluted in a 1/14 of the original volume of nuclease-free water. Adapter ligation was performed using 50-100 ng of the barcoded amplicon pool, NEBNext Quick Ligation Module reagents (NEB, US) and Adaptor Mix II (ONT, UK), and incubated at room temperature for 20 minutes. Final clean-up was performed using 1X AMPure XP beads and 125 μl of Short Fragment Buffer (ONT, UK). The library was eluted in 15 μl Elution Buffer (ONT, UK). The final library was normalised to 15-70 ng, loaded on a SpotON R9 flow cell and sequenced on a MinION Mk1B or GridION device^6^.

#### SARS-CoV-2 genome consensus assembly

The data generated via the MinION and GridION devices were processed using the ARTIC bioinformatic protocol (https://artic.network/ncov-2019/ncov2019-bioinformatics-sop.html). In brief, raw FAST5 files were base called and demultiplexed using ONT’s Guppy (version 4.0.5) in high accuracy mode using a minimum Q score of 7. FASTQ reads between 300 bps and 750 bps were filtered using the guppyplex module. The consensus sequences were generated by aligning base called reads against the SARS-CoV-2 reference genome (GenBank accession MN908947.3) using MiniMap2^7^. All positions with a genome coverage of less than 20 reads were masked with Ns. The consensus sequences were then polished using Nanopolish toolkit (version 0.13.3) using the raw signals.

#### Lineage and VOC assignment

Additional quality control, clade assignment and mutation profiles were obtained using the NextClade tool v1.13.2^8^ using a SARS-CoV-2 reference genome (accession NC_045512). All consensus sequences with a genome coverage >70% were classified using the PANGO lineage assignment tool (Pangolin version 3.1.20 and PangoLearn v.02/02/2022)^9^.

#### Global comparison sequences

Seychelles sequences were analysed against a backdrop of globally representative SARS-CoV-2 lineages. To ensure global representation of sequences, we downloaded all the sequences (n= 8,916,634) from the Global Initiative on Sharing All Influenza Data (GISAID) database collected before 31^st^ December and used an in-house R script to randomly select a sub-sample of 5,179 genomes while considering Pango lineage (lineages detected in Seychelles only), continent and date of collection. These global random genomes were collected from 150 countries and territories between 2^nd^ May 2020 and 31^st^ December 2021.

#### Phylogenetic reconstruction

The retrieved global sequence dataset, the sequences from Seychelles were aligned using Nextalign version 1.4.1. against the reference SARS-CoV-2 genome (accession NC_045512). A maximum likelihood (ML) phylogeny was inferred using IQTREE version 2.1.3 (http://www.iqtree.org/). The software initiates tree reconstruction after assessment and selection of the best model of nucleotide substitution for the alignment. TreeTime version 0.8.1 was used to transform the ML tree topology into a time calibrated phylogenetic tree. The resulting trees were visualized using the Bioconductor ggTree version 2.2.4 package^10^ in R^11^.

#### Estimation of virus importations and exportations into Seychelles

The global ML tree topology was used to estimate the number of viral transmission events between Seychelles and the rest of the world as described previously^12^. TreeTime was used to transform the ML tree topology into a dated phylogenetic tree. Outlier sequences (n=208) were identified by TreeTime and excluded during this process. A mugration model was fitted using the time-scaled phylogenetic tree, mapping the location status of the genomes from Seychelles at both the tips and internal nodes. Using the date and location annotated tree topology, we counted the number of transitions between Seychelles and the rest of the world and plotted this using ggplot2 version 3.3.3^13^.

#### Statistical analysis

All statistical analyses were performed using R version 4.1.0^11^.

## RESULTS

### Sequenced COVID-19 cases in Seychelles

The rise of COVID-19 cases in 2021 was preceded by a period of relaxed countermeasures to curb the spread of the virus **(Figure1 A and B)**. Of 1,298 SARS-CoV-2 positive samples received at KWTRP for genome sequencing, near complete genomes (>70% genome coverage) were recovered from 1,056 samples, and these were used in the subsequent lineage and phylogenetic analysis **(Supplementary Figure 2)**. A summary of the demographic details for the samples successfully sequenced and those that failed are provided in **Table 1**.

**Table 1.**
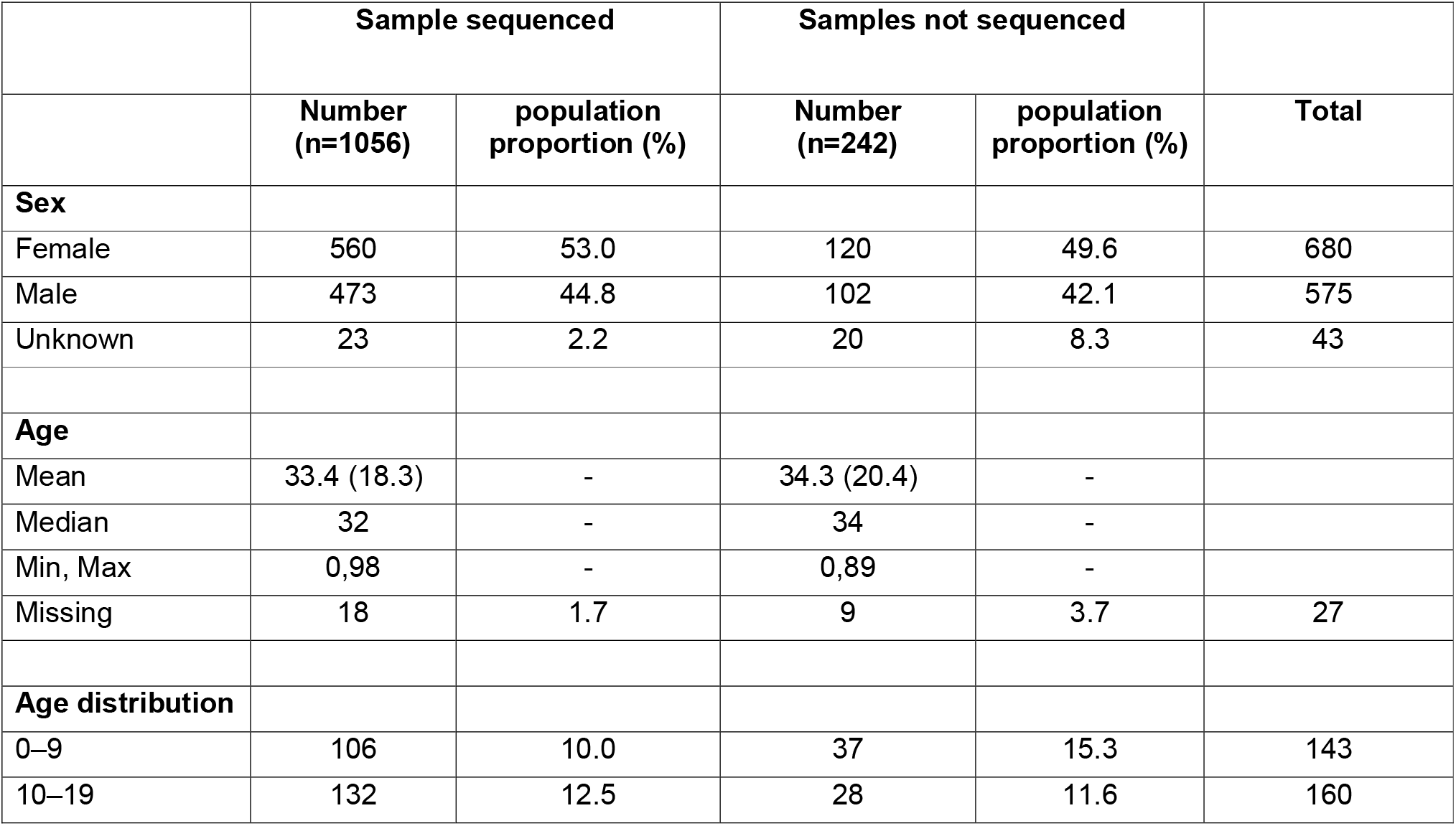
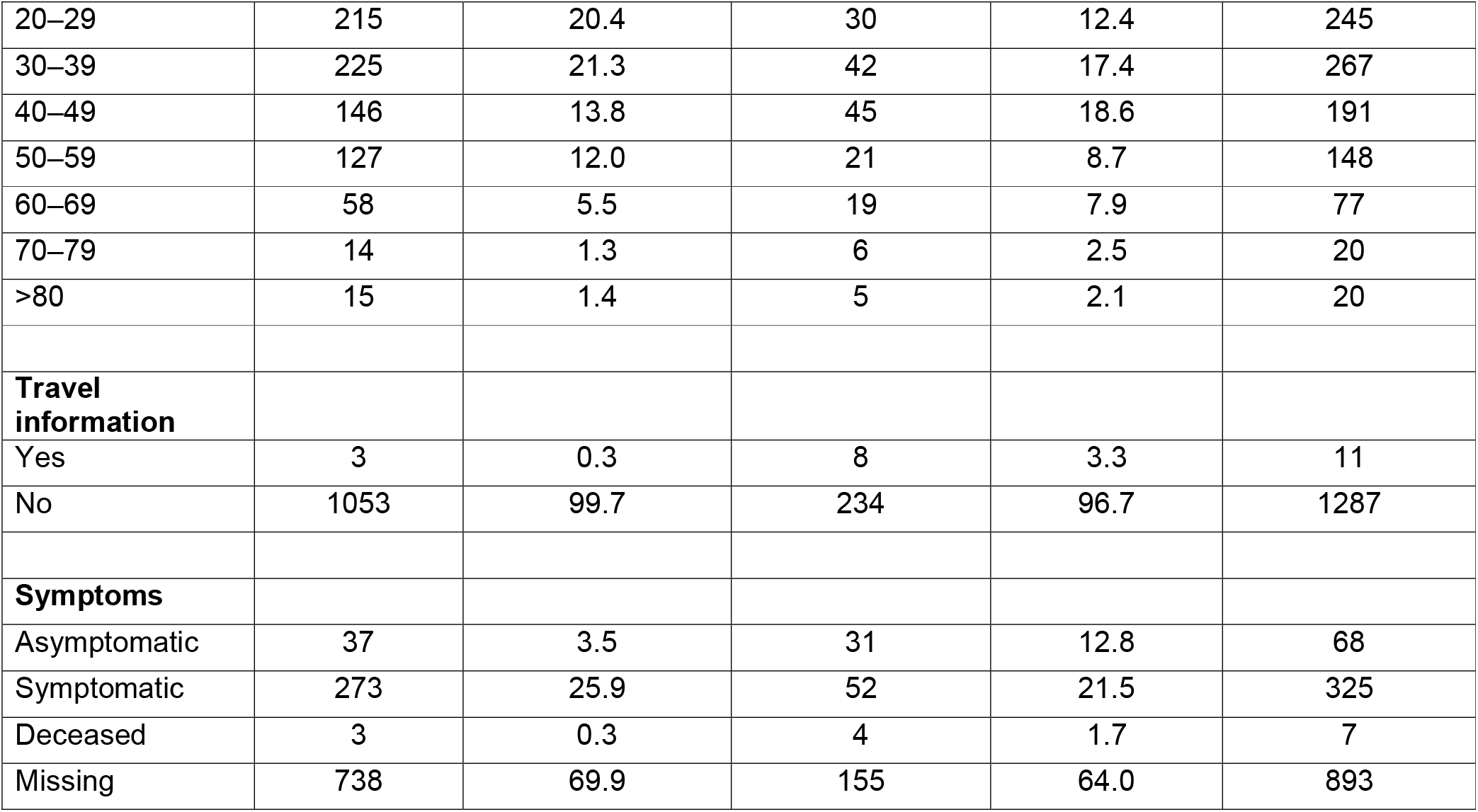
Demographic characteristics of SARS-CoV-2 positive samples received for sequencing at KWTRP. Sample were collected between 14^th^ March 2020 and 31^st^ December 2021 (n = 1,298).

|  | Sample sequenced |  | Samples not sequenced |  |  |
| --- | --- | --- | --- | --- | --- |
|  | Number<br>(n=1056) | population<br>proportion (%) | Number<br>(n=242) | population<br>proportion (%) | Total |
| **Sex** |  |  |  |  |  |
| Female | 560 | 53.0 | 120 | 49.6 | 680 |
| Male | 473 | 44.8 | 102 | 42.1 | 575 |
| Unknown | 23 | 2.2 | 20 | 8.3 | 43 |
| **Age** |  |  |  |  |  |
| Mean | 33.4 (18.3) | - | 34.3 (20.4) | - |  |
| Median | 32 | - | 34 | - |  |
| Min, Max | 0,98 | - | 0,89 | - |  |
| Missing | 18 | 1.7 | 9 | 3.7 | 27 |
| **Age distribution** |  |  |  |  |  |
| 0–9 | 106 | 10.0 | 37 | 15.3 | 143 |
| 10–19 | 132 | 12.5 | 28 | 11.6 | 160 |
| 20–29 | 215 | 20.4 | 30 | 12.4 | 245 |
| 30–39 | 225 | 21.3 | 42 | 17.4 | 267 |
| 40–49 | 146 | 13.8 | 45 | 18.6 | 191 |
| 50–59 | 127 | 12.0 | 21 | 8.7 | 148 |
| 60–69 | 58 | 5.5 | 19 | 7.9 | 77 |
| 70–79 | 14 | 1.3 | 6 | 2.5 | 20 |
| >80 | 15 | 1.4 | 5 | 2.1 | 20 |
| **Travel information** |  |  |  |  |  |
| Yes | 3 | 0.3 | 8 | 3.3 | 11 |
| No | 1053 | 99.7 | 234 | 96.7 | 1287 |
| **Symptoms** |  |  |  |  |  |
| Asymptomatic | 37 | 3.5 | 31 | 12.8 | 68 |
| Symptomatic | 273 | 25.9 | 52 | 21.5 | 325 |
| Deceased | 3 | 0.3 | 4 | 1.7 | 7 |
| Missing | 738 | 69.9 | 155 | 64.0 | 893 |

### SARS-CoV-2 lineages circulating in Seychelles

The recovered 1,056 genomes were classified into 32 distinct Pango lineages, 28 of which occurred within VOC, VOI or VUM: Alpha VOC (n=1), Beta VOC (n=1), Delta VOC (n=21) and Omicron VOC (n=3) and Kappa VOI (n=1) and B.1.640.2 VUM (n=1) **(Table 2)**. A total of four non-VOC/non-VOI/non-VUM lineages were detected among the sequenced infections in Seychelles: B.1 (n=9), B.1.1 (n=1), B.1.1.50 (n=1) and lineage B.1.1.277 (n=1). Lineage B.1 (predominantly detected in Europe) was the first lineage to be detected in Seychelles, in a sequenced sample from June 2020, followed by lineage B.1.1.277 (predominantly detected in Europe) in October 2020, B.1.1.50 (Predominantly detected in Israel and Palestine) in January 2021 followed by B.1.1 in May 2021. The non-VOC/VOI lineages were replaced by the Beta VOC (B.1.351) in February 2021, and which was later subsequently replaced by the Delta VOC in May 2021 that dominated until November 2021 when Omicron cases were first identified **(Figure 1C and Figure 2)**.

**Table 2.**
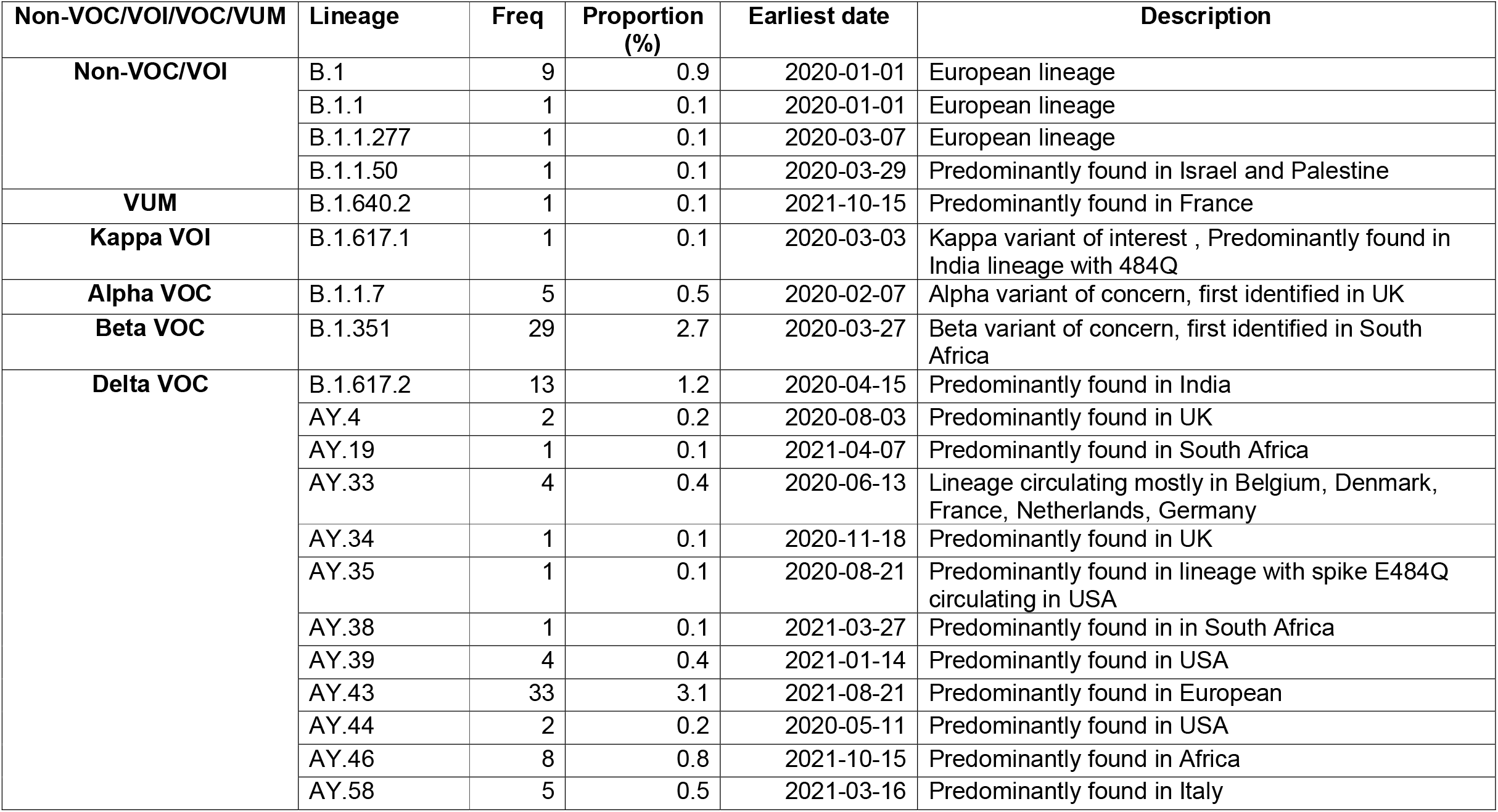
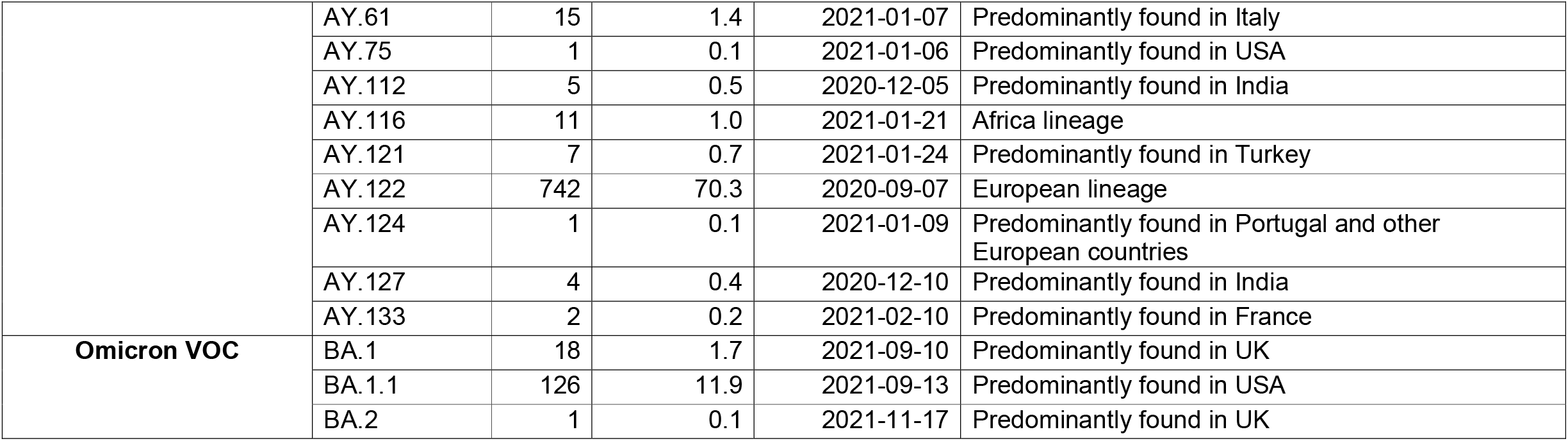
Description of SARS-CoV-2 lineages observed in Seychelles, their global history and VOC/VOI status

| Non-VOC/VOI/VOC/VUM | Lineage | Freq | Proportion (%) | Earliest date | Description |
| --- | --- | --- | --- | --- | --- |
| **Non-VOC/VOI** | B.1 | 9 | 0.9 | 2020-01-01 | European lineage |
|  | B.1.1 | 1 | 0.1 | 2020-01-01 | European lineage |
|  | B.1.1.277 | 1 | 0.1 | 2020-03-07 | European lineage |
|  | B.1.1.50 | 1 | 0.1 | 2020-03-29 | Predominantly found in Israel and Palestine |
| **VUM** | B.1.640.2 | 1 | 0.1 | 2021-10-15 | Predominantly found in France |
| **Kappa VOI** | B.1.617.1 | 1 | 0.1 | 2020-03-03 | Kappa variant of interest , Predominantly found in India lineage with 484Q |
| **Alpha VOC** | B.1.1.7 | 5 | 0.5 | 2020-02-07 | Alpha variant of concern, first identified in UK |
| **Beta VOC** | B.1.351 | 29 | 2.7 | 2020-03-27 | Beta variant of concern, first identified in South Africa |
| **Delta VOC** | B.1.617.2 | 13 | 1.2 | 2020-04-15 | Predominantly found in India |
|  | AY.4 | 2 | 0.2 | 2020-08-03 | Predominantly found in UK |
|  | AY.19 | 1 | 0.1 | 2021-04-07 | Predominantly found in South Africa |
|  | AY.33 | 4 | 0.4 | 2020-06-13 | Lineage circulating mostly in Belgium, Denmark, France, Netherlands, Germany |
|  | AY.34 | 1 | 0.1 | 2020-11-18 | Predominantly found in UK |
|  | AY.35 | 1 | 0.1 | 2020-08-21 | Predominantly found in lineage with spike E484Q circulating in USA |
|  | AY.38 | 1 | 0.1 | 2021-03-27 | Predominantly found in in South Africa |
|  | AY.39 | 4 | 0.4 | 2021-01-14 | Predominantly found in USA |
|  | AY.43 | 33 | 3.1 | 2021-08-21 | Predominantly found in European |
|  | AY.44 | 2 | 0.2 | 2020-05-11 | Predominantly found in USA |
|  | AY.46 | 8 | 0.8 | 2021-10-15 | Predominantly found in Africa |
|  | AY.58 | 5 | 0.5 | 2021-03-16 | Predominantly found in Italy |
|  | AY.61 | 15 | 1.4 | 2021-01-07 | Predominantly found in Italy |
|  | AY.75 | 1 | 0.1 | 2021-01-06 | Predominantly found in USA |
|  | AY.112 | 5 | 0.5 | 2020-12-05 | Predominantly found in India |
|  | AY.116 | 11 | 1.0 | 2021-01-21 | Africa lineage |
|  | AY.121 | 7 | 0.7 | 2021-01-24 | Predominantly found in Turkey |
|  | AY.122 | 742 | 70.3 | 2020-09-07 | European lineage |
|  | AY.124 | 1 | 0.1 | 2021-01-09 | Predominantly found in Portugal and other European countries |
|  | AY.127 | 4 | 0.4 | 2020-12-10 | Predominantly found in India |
|  | AY.133 | 2 | 0.2 | 2021-02-10 | Predominantly found in France |
| **Omicron VOC** | BA.1 | 18 | 1.7 | 2021-09-10 | Predominantly found in UK |
|  | BA.1.1 | 126 | 11.9 | 2021-09-13 | Predominantly found in USA |
|  | BA.2 | 1 | 0.1 | 2021-11-17 | Predominantly found in UK |

**Figure 1.**
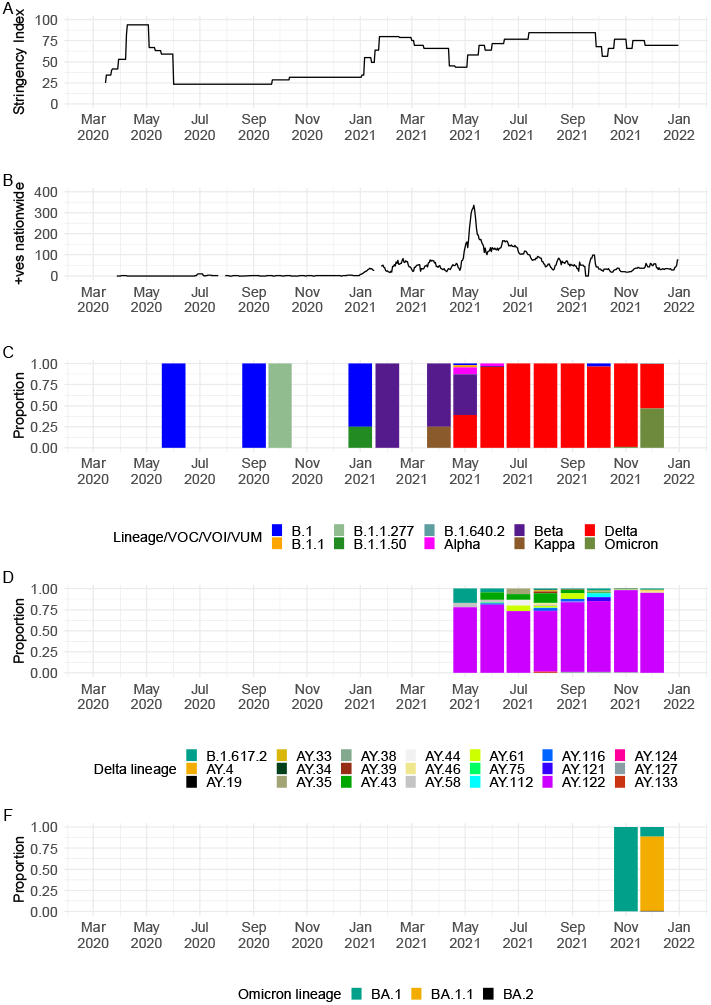
**A** Seychelles government intervention levels as measured by the Oxford stringency index ^29^. **B** An epidemic curve for Seychelles derived from the daily positive case numbers obtained from https://ourworldindata.org/coronavirus/country/seychelles. **C** Monthly temporal pattern of SARS-CoV-2 lineages and variants in Seychelles among the 1,056 samples sequenced from COVID-19 positive cases from the Seychelles (25^th^ June 2020, to 31^st^ December 2021). **D** Monthly temporal distribution of Delta VOC lineages among samples sequenced from COVID-19 positive cases from the Seychelles (25^th^ June 2020, to 31^st^ December 2021). **E** Monthly temporal distribution of Omicron VOC lineages among samples sequenced from COVID-19 positive cases from the Seychelles (25^th^ June 2020, to 31^st^ December 2021).

**Figure 2.**
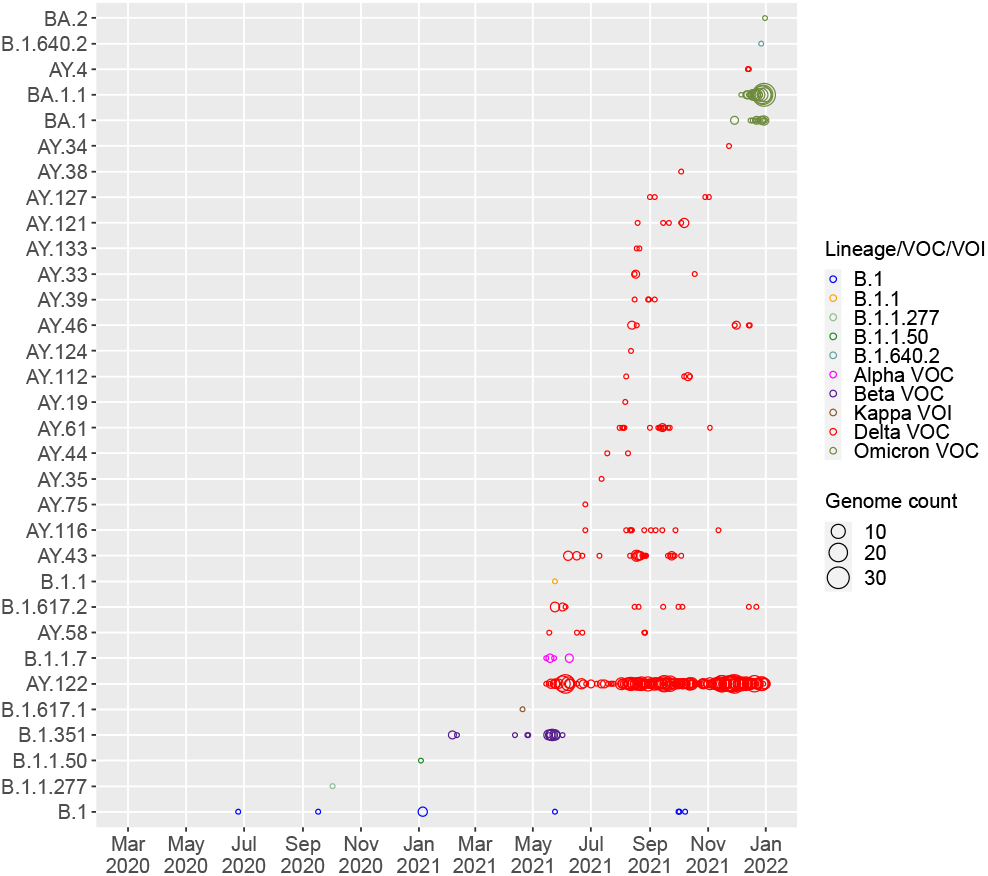
SARS-CoV-2 Pango lineages in the sequenced 1,056 Seychelles samples and timing of detections (circle size scaled by number of daily detections).

Detection of Beta VOC in February 2021 coincided with the start of a surge of COVID-19 cases, with a further sharp surge observed in May 2021 coinciding with detection of Delta VOC in May 2021 **(Figure 1B and C)**. Since the emergence of Delta VOC in Seychelles in May 2021, a total of 21 Delta VOC lineages co-circulated with varying frequency. Lineage AY.122 (n= 742) was the most prevalent, with detections until the end of the surveillance period covered in this analysis **(Figure 1D and Figure 2)**. Other common Delta lineages were AY.43 (n=33) and B.1.617.2 (n=13). We observed the start of another surge in November 2021 due to Omicron (lineages BA.1 (n=126), BA.1.1 (n=18) and BA.2 (n=1) **(Figure 1B and Supplementary Figure 1A)** which peaked in mid-January 2022 (not shown).

### SARS-CoV-2 diversity in Seychelles

Genetic distance-resolved phylogeny of the Seychelles genomes, including global reference sequences (n=5,179) revealed that most of the Seychelles sequences were interspersed as clusters (>2 sequences) or singletons across the phylogenetic trees suggesting multiple viral introductions into Seychelles **(Figure 3)**. In the VOC/non-VOC-specific phylogenies, Delta VOC (n=863) grouped into 14 clusters (>2 sequences) clusters and 12 singletons on the global phylogenetic tree pointing to separate introductions of the Delta VOC into the country whereas Omicron VOC (n=145) clustered into 10 clusters and 11 singletons (**Figures 3D and E**). Seychelles’ Beta (n=29) and Alpha (n=5) viruses clustered closely amongst themselves suggesting few introductions that led to onward transmission in Seychelles **(Figures 3B and C)**. Seychelles B.1 viruses sampled in 2020 were dispersed on the global phylogeny as singletons most likely pointing to multiple introductions into Seychelles during the initial phase of the pandemic **(Figure 3A)**. Sequences from different locations (i.e., districts) in Seychelles clustered closely or together on the phylogenetic tree, a feature suggesting rapid spread of the virus within the country over a short period of time (not shown).

**Figure 3.**
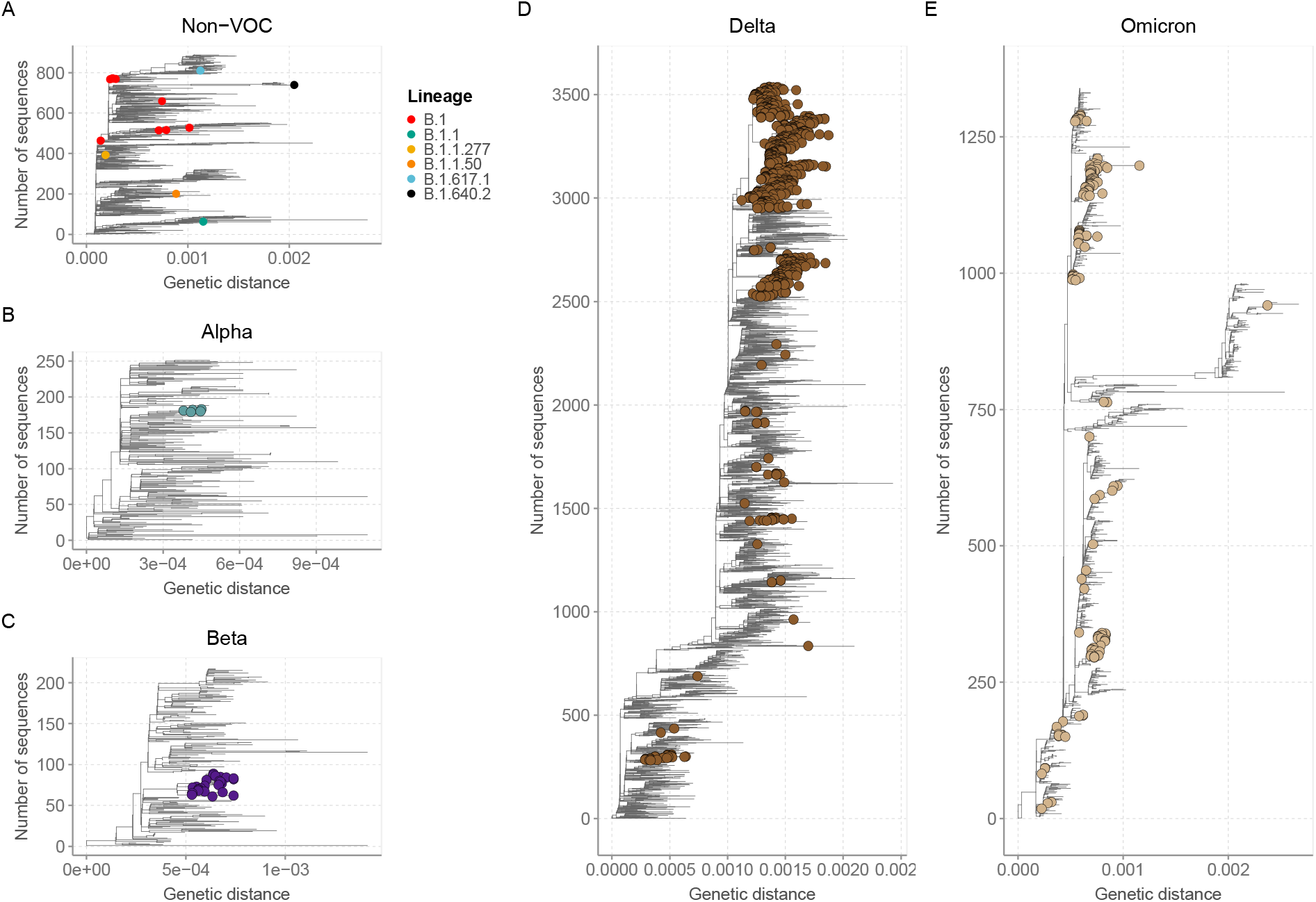
Genetic distance-resolved lineage-specific phylogenetic trees for Omicron, Alpha, Beta, Delta VOC, and Non-VOC. Seychelles genomes are indicated with colored tip labels. **A** Phylogeny of Delta VOC that combined 863 Seychelles sequences and 2,676 global sequences. **B** Phylogeny of Omicron VOC that combined 145 Seychelles sequences and 1,195 global sequences **C** Phylogeny of Alpha VOC that combined 5 Seychelles sequences and 246 global sequences. **D** Phylogeny of Beta VOC that combined 29 Seychelles sequences and 187 global sequences. **E** Phylogeny of Non-VOC that combined 14 Seychelles sequences and 875 global sequences.

SARS-CoV-2 variant analysis comparing sequences from Seychelles’ sequences to the Wuhan reference sequence (NC_045512.2) detected a total of 703 amino acid mutations across different gene regions; however, focusing on the frequent occurring mutations only, we identified a total of 27 amino acid mutations and two deletions that had a prevalence of >50% in the sequenced cases **(Supplementary Figure 3A and B)**. The most prevalent amino acid mutation was D614G (A23403G) (98.9%) occurring in the spike glycoprotein followed by P314L (C14408T) (93.3%) in the open reading frame 1b (ORF1b) **(Supplementary Figure 3A)**.

### Export and import of SARS-CoV-2 lineages in Seychelles

Ancestral location state reconstruction of the dated global phylogeny **(Figure 4A)** was used to infer the number of viral importations and exportations. In total, between the 25^th^ June 2020 and the 31^st^ December 2021, we inferred at least 78 importations into Seychelles with 28 (35%) of the introductions coming from Europe, 21 separate introductions from Africa, 15 separate introductions from Asia, 6 introductions from North America, 5 separate introductions from Oceania and 3 introductions from South America. **(Figure 4B)**. Of the 78 detected viral imports into Seychelles, 66 occurred between January and December 2021 after the rise in COVID-19 cases was experienced in the Seychelles (**Figure 4C)**. From the analysis, we also inferred 32 export events from Seychelles to the rest of the world, mainly Asia (n=10) Europe (n=8) and Africa (n=6).

**Figure 4.**
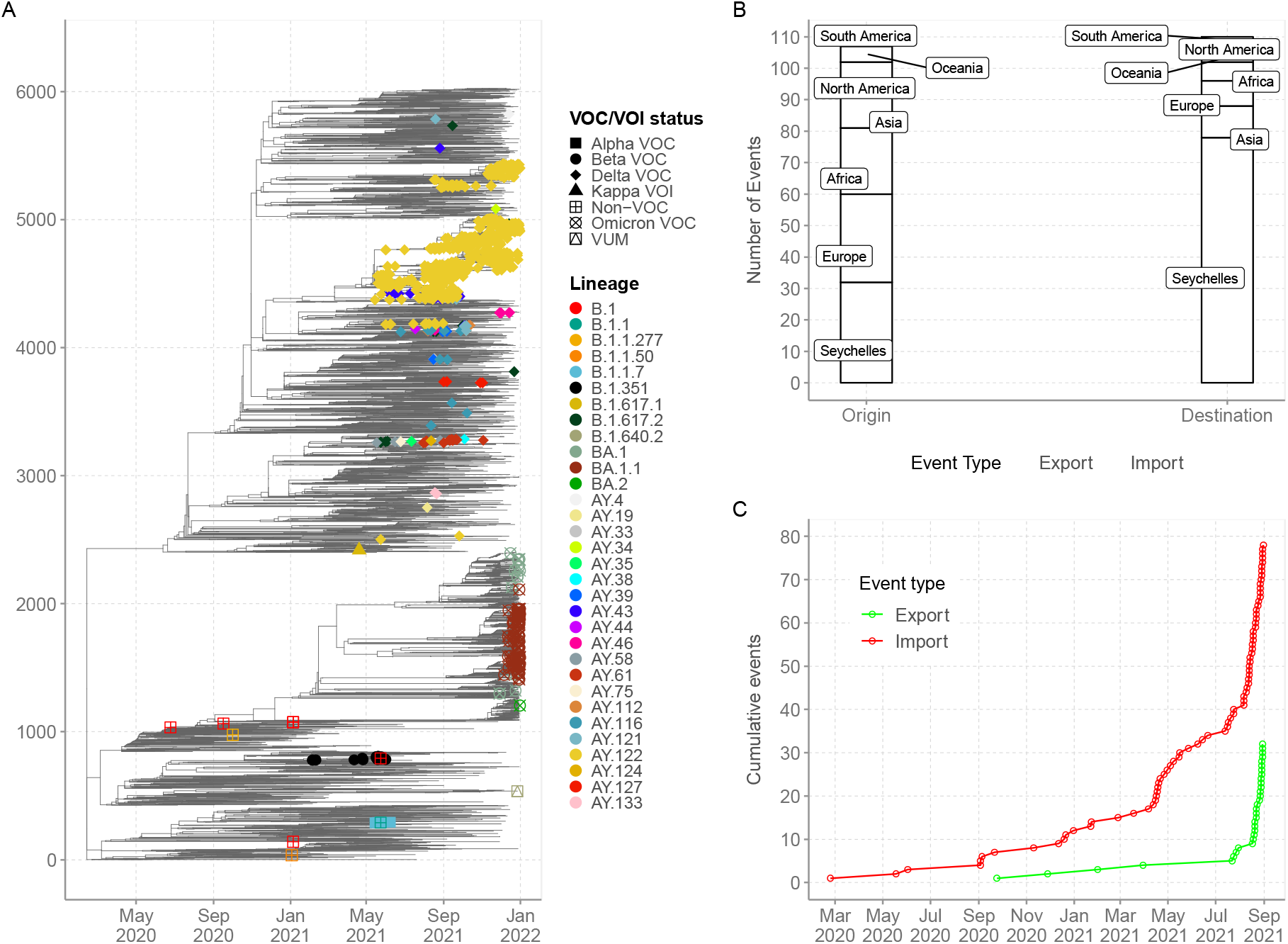
**A**. Time-resolved global phylogeny that combined 1,056 Seychelles sequences (coloured tip labels) and 5,179 global reference sequences. **B** The number of viral imports and exports into and out of Seychelles. **C** Cumulative number of viral imports and export over time into Seychelles.

## DISCUSSION

We estimated at least 78 independent introductions of SARS-CoV-2 into Seychelles between 25^th^ June 2020 to 31^st^ December 2021, with the importations likely originating from all the continents in the world. Notably, the surge of COVID-19 cases from January to December 2021 was characterised by detections of VOCs in the country. The Beta variant of concern was the dominant strain in circulation from February to April 2021 and probably responsible for the surge in cases in January 2021 (highest daily number of infections at 231, on the 4^th^ March 2021), but the introduction and rapid spread of the Delta variant seen from May to December 2021 caused a further surge (highest daily number of infections at 484, on the 6^th^ May 2021).

The low number of cases detected for non-VOCs (B.1, B.1.1, B.1.1.50 and B.1.1.277) lineages in 2020, could be explained by the fact that this insular population experienced very low importations due to the COVID-19 restriction countermeasures that were in place in 2020 but also that the non-VOCs were not highly transmissible. We ascertain that the introduction of the VOCs with high transmissibility was probably the cause of the surge of cases in Seychelles in 2021. However, these assertions should be taken with caution since extensive genomic surveillance in the country began in early 2021. To note, the detection of VOCs in the country preceded period of relaxed COVID-19 restriction countermeasures, including allowing tourists into the country and the resumption of in person school classes. This may have led to increased viral importations, as detected in our dataset, and to spread in the country.

The detection of Delta VOCs in May 2021 coincided with a period of relaxed COVID-19 restriction countermeasures (Oxford stringency index < 50) and the surge of cases during this time may have been due to multiple introductions and rapid spread of the virus in the country. By December, a total of 21 different Delta lineages were detected in Seychelles. The first Omicron detection in Seychelles was on the 29^th^ of November 2021. By 31st December 2021, three Omicron lineages had been detected in Seychelles including BA.2 which has been noted to be rapidly growing across the world^14–16^

Unsurprisingly, the majority of the Seychelles sequences harboured important spike mutations e.g., D614G mutation occurring in 98.2% of all the sequenced cases. The D614G amino acid change has been associated with stronger interaction between the virus and the angiotensin-converting enzyme 2 (ACE2) affinity, leading to higher infectivity and transmissibility^17,18^, Other important mutations included : i. Beta VOC; K417N, E484K and N501Y occurring in the spike receptor binding domain (RBD), which have been associated with reduced sensitivity to convalescent and post-vaccination sera^19^; ii. Delta VOC, L452R, P681R and D950N mutations, which have been linked to reduced sensitivity to neutralizing antibodies and higher transmissibility^20^; iii. Alpha VOC; N501Y and P681H mutations and iv. Omicron VOC: Q498R and N501Y occurring in the RBD have been linked to ACE2 binding affinity^21^. These RBD mutations coupled with four amino acid substitutions (i.e., A67V, T95I, and L212) and three deletion (67-70, 142-144 and 211) and an insertion (EPE at position 214) in the N-terminal domain (NTD) are linked to reduced sensitivity of Omicron to reduced sensitivity to convalescent and post-vaccination sera^22,23^. A cluster of three mutations occurring at S1-S2 furin cleavage site (H655Y, N679K and P681H) have been associated with increased transmissibility^24^.

Our phylogenetic analysis showed that Seychelles sequences virus diversity was nested within the global virus diversity (i.e., Seychelles sequences clustered with sequences sampled from different countries, suggestive of global spread of SARS-CoV-2 lineages). The close association between the viruses and those from other countries reflects global transmission of the virus as a result of global migration, increased connectivity, and social mixing. Further, focusing on the VOCs, we observed patterns of SARS-CoV-2 viral diversity inside Seychelles; phylogenetic clusters consisted of viruses which were derived from different geographic locations and formed a deep hierarchical structure, indicating an extensive and persistent nation-wide transmission of the virus.

Our findings are consistent with findings from Island countries such as Comoros, Reunion and New Zealand, where these countries were able to contain the first pandemic wave starting in March 2020, due COVID-19 strict countermeasures such as ban on international arrivals, which may have led to limited viral introductions into the islands, or perhaps viral introductions during the early phase of the pandemic did not results to community transmission due to government countermeasure on COVID-19 such as countrywide lockdown or self-isolation of the entire population^25,26^. Surge of COVID-19 cases in Islandic populations appears to be majorly driven by introductions of VOCs. For example, Comoros seems to have experienced its first surge of COVID-19 cases after introduction of Beta VOCs into the population in January 2021^27^, similar to the period when Seychelles saw its first surge also due Beta VOC.

This study had some limitations. First, our import/export inferences can be influenced by sampling biases and the low rate of sequencing in Seychelles. Therefore, the true number of international introductions are likely significantly higher than what is reported here. Second, incomplete metadata for some samples limited the scope of our analysis, for example, lack of location of samples collected disallowed us to investigate the transmission pathway of viruses within the country. Third, SARS-CoV-2 sequences from Seychelles are only available from a very small fraction of the number of confirmed cases into the country.

These data reinforce the importance of genomic surveillance in Seychelles as a tool for monitoring and providing real-time information on spread of emerging SARS-CoV-2 variants in the population with important implications for public health and immunization strategies. Surge of COVID-19 cases due to VOCs during a period of heightened COVID-19 countermeasures raises questions on the optimal timing of the introduction of public health interventions. When the interventions are introduced after a surge has started, it is often too late, and the control strategies should focus on local transmission to understand characteristics and origins of locally circulating SARS-CoV-2 diversity to prevent further spread^28^. Moreover, studies on genomic surveillance would also be useful in investigating vaccine effectiveness against circulating variants which appear to have a high turnover.

## Data Availability

All data generated and analysis script for this manuscript will be available from the Virus Epidemiology and Control, Kenya Medical Research Institute (KEMRI) Wellcome Trust Research Programme data server, https://doi.org/10.7910/DVN/AYT2UA.

## Acknowledgements

We thank the Seychelles Ministry of Health, Public Health Laboratory, Africa-CDC, WHO-Afro, WHO-Kenya Office, WHO-Seychelles Office for facilitating sharing of SARS-CoV-2 samples that were sequenced. Further, we thanks all the laboratories that have shared SARS-CoV-2 sequence data in GISAID that we included as comparison data in our analysis (see list in appendix 1).

## Grant Information

This works was supported by the National Institute for Health Research (NIHR) (project references 17/63/82 and 16/136/33) using UK aid from the UK Government to support global health research, The UK Foreign, Commonwealth and Development Office and Wellcome Trust (grant# 220985/Z/20/Z) The views expressed in this publication are those of the author (s) and not necessarily those of NIHR or the Department of Health and Social Care, Foreign Commonwealth and Development Office. This work was also supported by the Seychelles Public Health Laboratory (Public Health Authority), Africa-CDC, WHO-Afro, WHO-Seychelles, ASLM, and WHO-Kenya offices.

## Data availability

All data generated and analysis script for this manuscript will be available from the Virus Epidemiology and Control, Kenya Medical Research Institute (KEMRI)– Wellcome Trust Research Programme data server, https://doi.org/10.7910/DVN/AYT2UA. Whole genome sequences are available from GISAID database accession number EPI_ISL_4880527 - EPI_ISL_4880796, EPI_ISL_5942854 - EPI_ISL_6705093, EPI_ISL_8424404 - EPI_ISL_8424612 and EPI_ISL_11060102 - EPI_ISL_11060407.

## Conflicts of Interest

The authors have declared no competing interest.

## SUPPLEMENTARY FIGURES

**Supplementary Figure 1.**
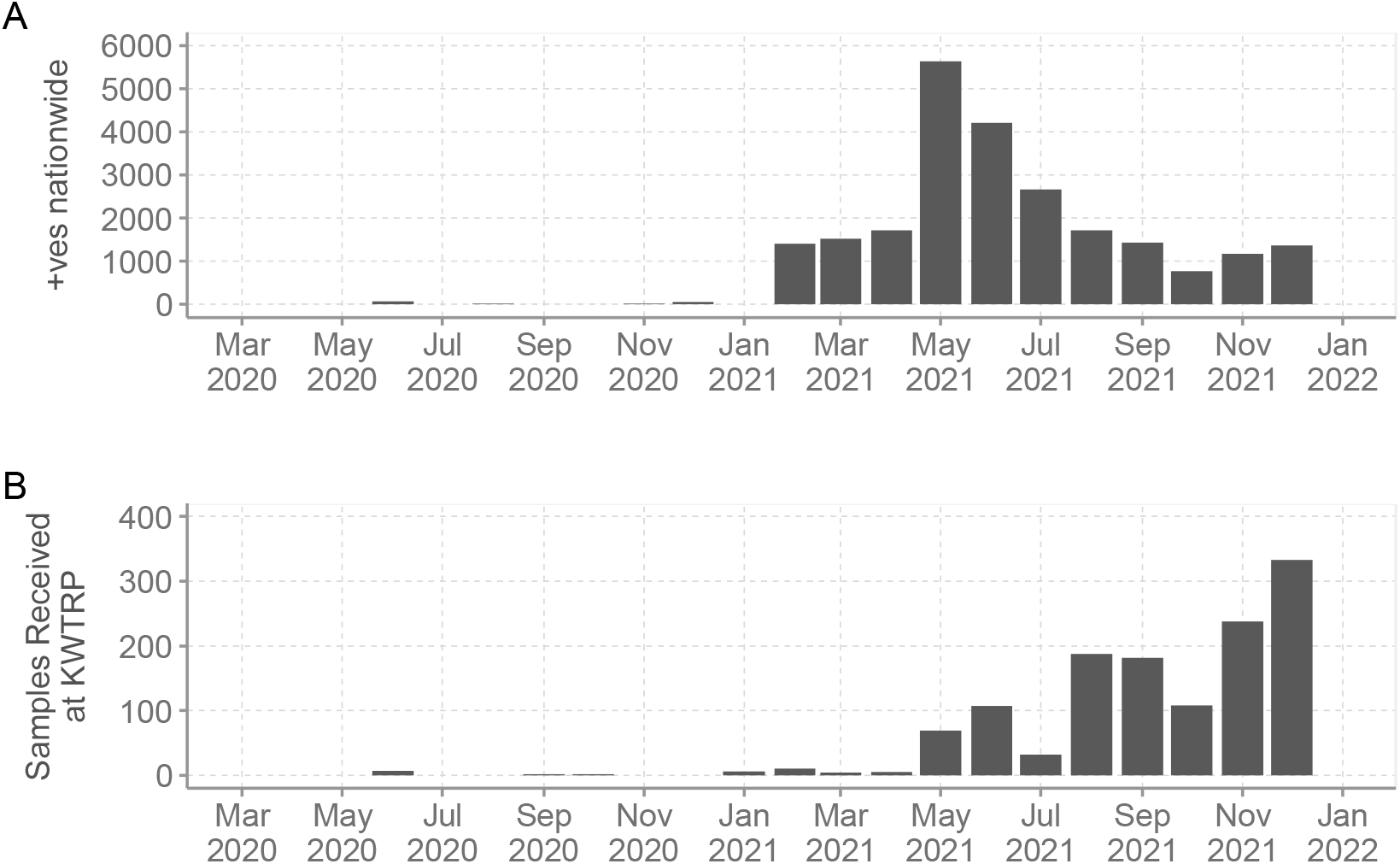
**A** The monthly temporal distribution of SARS-CoV-2 cases between March 2020 and 31^st^ December 2021 in Seychelles vs SARS-CoV-2 positives samples received at KEMRI-WT for whole genome sequencing.

**Supplementary Figure 2.**
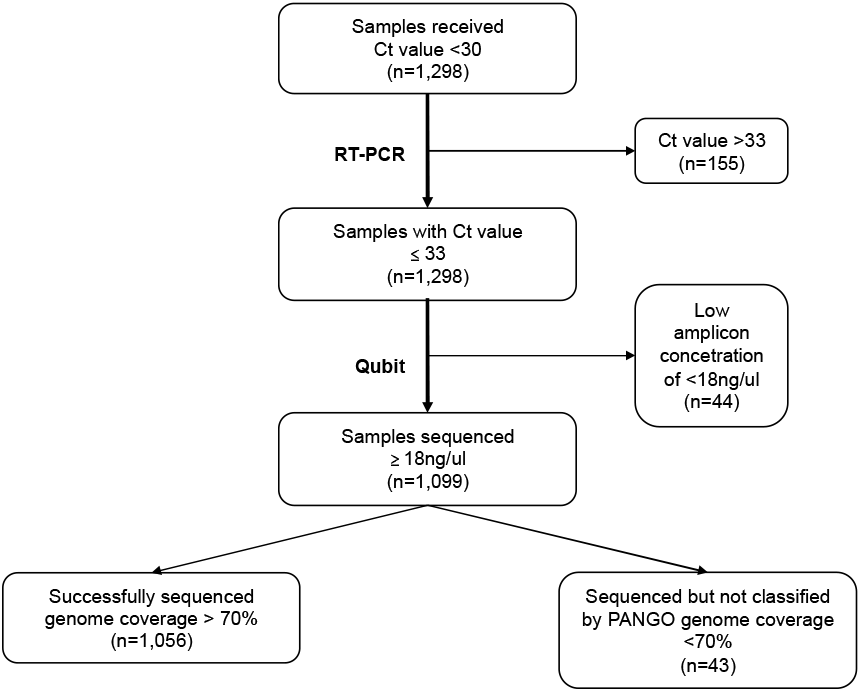
Flow chart describing processing of SARS-CoV-2 samples received at KEMRI-WT for whole genome sequencing.

**Supplementary Figure 3.**
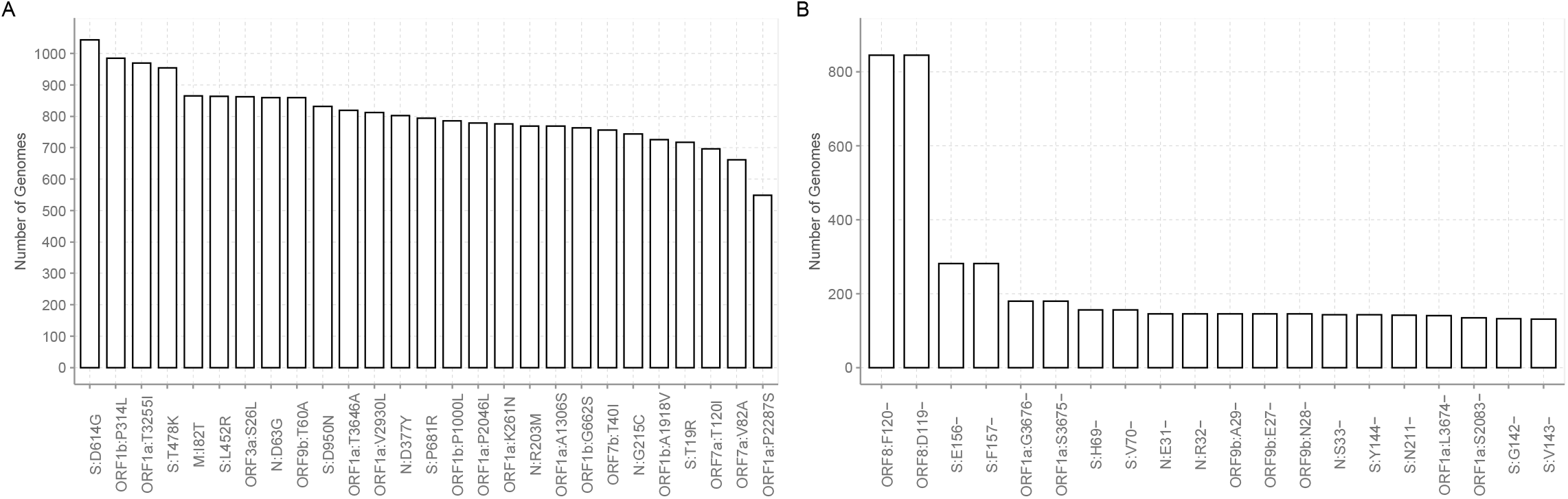
**A** Prevalent amino acid mutation among the 1,056 genomes from Seychelles. **B** Prevalent amino acid deletions among the 1,056 genomes from Seychelles.

## Notes

### Author Declarations

The whole genome sequencing study protocol was reviewed and approved by the Scientific and Ethics Review Committee (SERU) at KEMRI, (SERU #4035). Individual patient consent requirement was waivered by the committee as the sequenced samples were part of the public health emergency response.

### Summary of Updates

The list of authors and affiliations have been updated. The first paragraph in the abstract has been rephrased.

