## Supplementary figures and images for "Genomic Epidemiology of SARS-CoV-2 in Seychelles, 2020-2021"

### SupplementaryFigure1

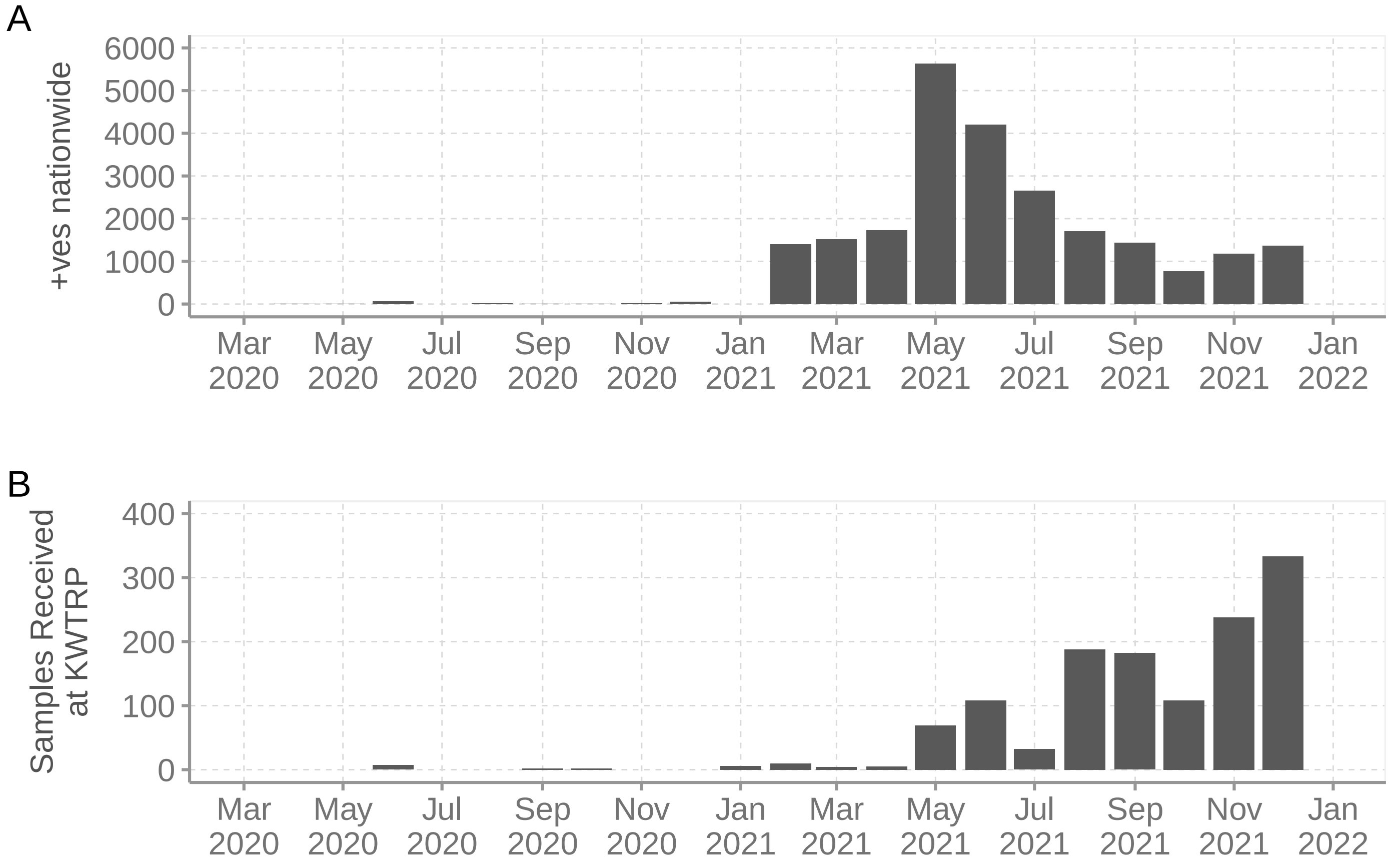

### SupplementaryFigure2

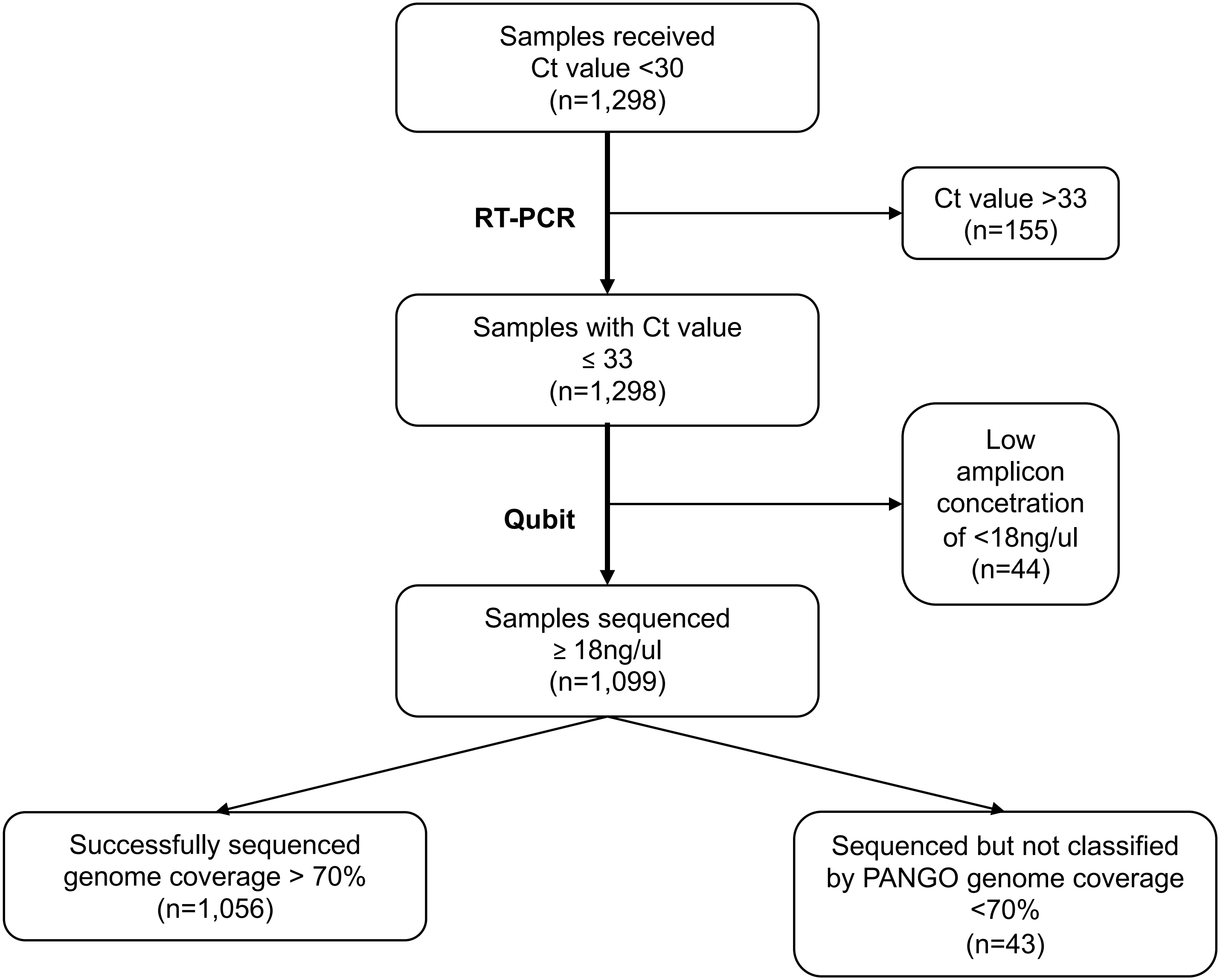

### SupplementaryFigure3

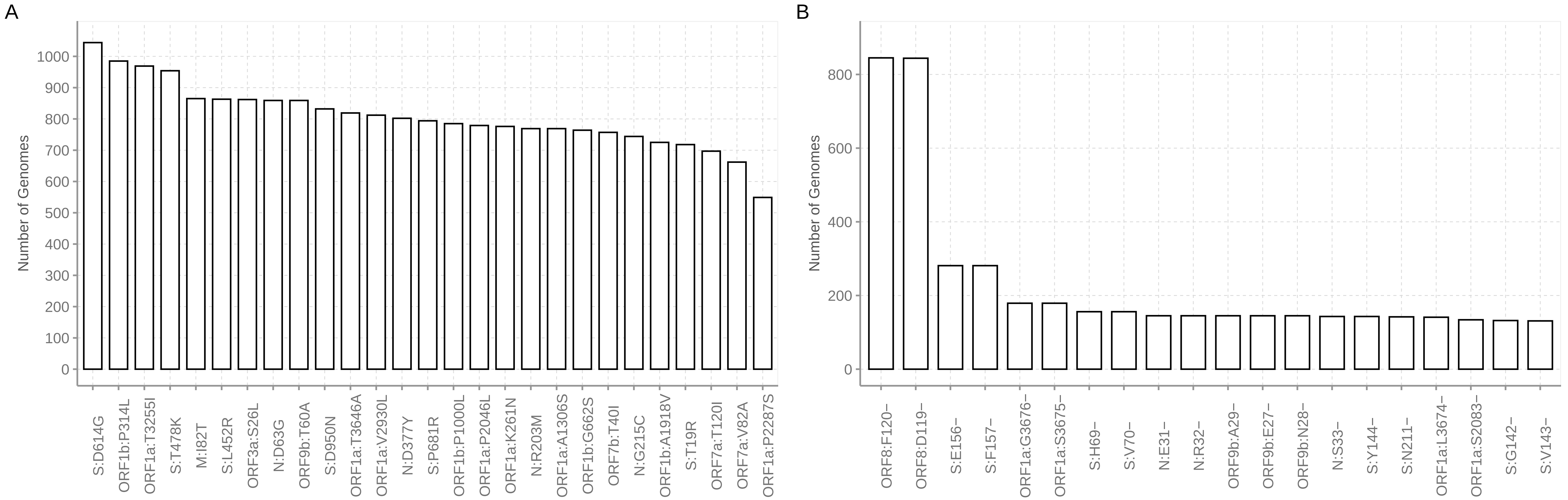
