## Appendix1 for "Genomic Epidemiology of SARS-CoV-2 in Seychelles, 2020-2021"

We gratefully acknowledge the following Authors from the Originating laboratories responsible for obtaining the specimens and the Submitting laboratories where genetic sequence data were generated and shared via the GISAID Initiative, on which this research is based

| Virus name | Accession | Collected | Originating laboratory | Submitting laboratory | Authors |
|---|---|---|---|---|---|
| hCoV-19/USA/AK-PHL9114/2021 | EPI_ISL_2421690 | 2021-05-20 | Alaska State Virology Laboratory | Alaska State Virology Laboratory | Stephanie DeRonde, Elva House, Jacob Zidek, Lisa Smith, Ph.D., Jack Chen, Ph.D. |
| hCoV-19/Ghana/WACCBIP-GS593/2021 | EPI_ISL_2422569 | 2021-03-30 | West African Centre for Cell Biology of Infectious Pathogen, University of Ghana, Legon | WACCBIP, University of Ghana, Volta Road, Legon, Accra | Collins M. Morang'a, Peter K Quashie, Joyce M. Ngoi, Vincent Appiah, Dominic S.Y. Amuzu, Evelyn B. Quansah, Philip M Soglo, Violet MCormack, Samirah Saiid, Ivy A Asante, Joseph HK Bonney, Evelyn Y Bonney, John K. Odoom, Nicaise T. Ndam, Frederick Tei-Maya |
| hCoV-19/Uruguay/UY-NYULH2060/2021 | EPI_ISL_2427638 | 2021-02-24 | Laboratorio de Biologv≠a Molecular Mv©dica Uruguaya | Departments of Pathology and Medicine, New York University School of Medicine | Maria Victoria Elizondo, Maria Noel Zubillaga, Gonzalo Manrique, Cecilia Sorhouet, Maria Cristina Mogdasy, Paul Zappile, Dacia Dimartino, Christian Marier, Adriana Heguy |
| hCoV-19/England/PLYM-165A52D/2021 | EPI_ISL_2570575 | 2021-06-08 | Lighthouse Laboratory Plymouth | Wellcome Sanger Institute for the COVID-19 Genomics UK (COG-UK) Consortium | Lighthouse Laboratory Plymouth and Alex Alderton, Roberto Amato, Jeffrey Barrett, Sonia Goncalves, Ewan Harrison, David K. Jackson, Ian Johnston, Dominic Kwiatkowski, Cordelia Langford, John Sillitoe on behalf of the Wellcome Sanger Institute COVID-19 Sur |
| hCoV-19/SouthAfrica/NICD-N00281/2020 | EPI_ISL_2433786 | 2020-09-10 | LANCET, LABORATORIES | Diseases of the National Health Laboratory Service | Amoako DG, Scheepers C, Mohale T, Ntuli N, Mahlangu B, Ismail A, Bhiman JN |
| hCoV-19/SouthAfrica/NICD-N00312/2020 | EPI_ISL_2433816 | 2020-09-09 | National Institute for Communicable Diseases of the National Health Laboratory Service | Diseases of the National Health Laboratory Service | Amoako DG, Scheepers C, Mohale T, Ntuli N, Mahlangu B, Ismail A, Bhiman JN |
| hCoV-19/SouthAfrica/VIDA-KRISP-K011335/2020 | EPI_ISL_2360689 | 2020-12-28 | Vaccines and Infectious Diseases Analytics Research Unit (VIDA) | KRISP, KZn Research Innovation and Sequencing Platform | Baillie Vicky, du Plessis Jeanine, Giandhari Jennifer, Pillay Sureshnee, Naidoo Yeshnee, Tegally Houriiyah, San James, Wilkinson Eduan, de Oliveira Tulio, Madhi Shabir |
| hCoV-19/England/ALDP-15F3633/2021 | EPI_ISL_2435009 | 2021-05-29 | Lighthouse Lab in Alderley Park | Wellcome Sanger Institute for the COVID-19 Genomics UK (COG-UK) Consortium | Jacquelyn Wynn, Mairead Hyland, The Lighthouse Lab in Alderley Park and Alex Alderton, Roberto Amato, Jeffrey Barrett, Sonia Goncalves, Ewan Harrison, David K. Jackson, Ian Johnston, Dominic Kwiatkowski, Cordelia Langford, John Sillitoe on behalf of the W |
| hCoV-19/SouthAfrica/N6479/2021 | EPI_ISL_2582924 | 2021-03-17 | KIMBERLEY LABORATORY | Diseases of the National Health Laboratory Service | Amoako DG, Everatt J, Scheepers C, Mohale T, Ntuli N, Mahlangu B, Mnguni A, Ismail A, Bhiman JN |
| hCoV-19/USA/GA-CDC-STM-000073558/2021 | EPI_ISL_2440306 | 2021-05-11 | Helix/Illumina | Centers for Disease Control and Prevention Division of Viral Diseases, Pathogen Discovery | Dakota Howard, Dhwani Batra, Peter W. Cook, Kara Moser, Adrian Paskey, Jason Caravas, Benjamin Rambo-Martin, Shatavia Morrison, Christopher Gulvick, Scott Sammons, Yvette Unoarumhi, Darlene Wagner, Matthew Schmerer, Eileen de Feo, Jan Antico, Christine Tr |

| | | | | | |
|---|---|---|---|---|---|
| hCoV-19/Canada/SK-NML-62504/2021 | EPI_ISL_2585976 | 2021-04-27 | SK-Roy Romanow Provincial Laboratory | National Microbiology Laboratory (NML) | Anna Majer, Shari Tyson, Grace Seo, Philip Mabon, Elsie Grudeski, Rhiannon Huzarewich, Russell Mandes, Anneliese Landgraff, Jennifer Tanner, Natalie Knox, Morag Graham, Gary Van Domselaar, Ryan McDonald, Amanda Lang, Rachel DePaulo, Jessica Minion, Nathal |
| hCoV-19/India/TG-CCMB-CIA2893/2021 | EPI_ISL_2441636 | 2021-05-13 | CSIR-Centre for Cellular and Molecular Biology | CSIR-Centre for Cellular and Molecular Biology-INSACOG | Lamuk Zaveri,Tulasi Nagabandi,Payel Mukherjee,Onkar Kulkarni, Sofia Banu, Ara Sreenivas,Shreekant Verma, Amareshwar Vodapalli ,B Himasri,Valli Nagalakshmi Undamatla, Sumedha Avadhanula, Priya Nurkurthy, Vidhyadhari Methuku, Archana Bharadwaj Siva, Karthik |
| hCoV-19/Reunion/CHU_7210334277/2021 | EPI_ISL_2441786 | 2021-03-31 | CHU de la R√©union - Nord | Laboratoire de virologie, CNR arbovirus Associ√©, Chu de la R√©union | Etienne Frumence, Marie-Christine Jaffar Bandjee, Nicolas Traversier, Sabrina Petit Genet |
| hCoV-19/Brazil/PR-FIOCRUZ-22792/2021 | EPI_ISL_2443545 | 2021-04-26 | Lboratorio Central de Saude Publica do Estado do Parana (LACEN/PR) | Laboratory of Respiratory Viruses and Measles, Oswaldo Cruz Institute, FIOCRUZ | Paola Resende, Luciana Appolinario, Fernando Motta, Anna Carolina Paixao, Ana Carolina Mendonca, Alice Sampaio Rocha, Taina Venas, Elisa Cavalcante Pereira, Renata Serrano Lopes, Irina Riediger, Marilda Siqueira on behalf of the Fiocruz COVID-19 Genomic S |
| hCoV-19/Portugal/PT8384/2021 | EPI_ISL_2445958 | 2021-05-11 | Affidea | Instituto Nacional de Saude (INSA) | Borges et al |
| hCoV-19/India/MP-NCDC-1209278/2021 | EPI_ISL_2448239 | 2021-02-19 | National Centre for Disease Control (NCDC) Biotechnology Division, Delhi | NCDC Delhi, Biotechnology Division INSACOG | Mahesh S Dhar, Kalaiarasan Ponnusamy, Meena Datta, Priyanka Singh, Uma Sharma, Manoj K Singh, Radhakrishnan V. S, Robin Marwal, Hemlata Lall, Hema Gogia, Preeti Madan, Sandhya Kabra, Sujeet K Singh, Partha Rakshit |
| hCoV-19/Germany/BE-RKI-I-148356/2021 | EPI_ISL_2260146 | 2021-04-29 | Labor 28 MVZ GmbH | Robert Koch Institute | unknown |
| hCoV-19/Belgium/IPG-225/2021 | EPI_ISL_1655960 | 2021-04-08 | CHWAPI - SITE NOTRE DAME | (IPG) | Pascale Hilbert, J√©r√©mie Gras |
| hCoV-19/Russia/SPE-RII-MH18155S/2021 | EPI_ISL_2450559 | 2021-05-17 | City Hospital No 40 | WHO National Influenza Centre Russian Federation | Andrey Komissarov, Artem Fadeev, Kseniya Komissarova, Oula Mansour, Kirill Varchenko, Mikhail Bakaev, Tamila Musaeva, Maria Timofeeva, Veronika Eder, Maria Pisareva, Nikita Yolshin, Daria Danilenko, Olga Shneider, Sergey Scherbak, Ksenia Safina, Elena Nab |
| hCoV-19/USA/CO-CDC-MMB08823378/2021 | EPI_ISL_2451072 | 2021-05-16 | Mako Medical | Centers for Disease Control and Prevention Division of Viral Diseases, Pathogen Discovery | Dakota Howard, Dhwani Batra, Peter W. Cook, Kara Moser, Adrian Paskey, Jason Caravas, Benjamin Rambo-Martin, Shatavia Morrison, Christopher Gulvick, Scott Sammons, Yvette Unoarumhi, Darlene Wagner, Matthew Schmerer,Matthew Tugwell, Lauren Moon, Clinton R. |
| hCoV-19/SouthAfrica/NICD-N6028/2021 | EPI_ISL_2454692 | 2021-04-04 | ROB FERREIRA LABORATORY | Diseases of the National Health Laboratory Service | Amoako DG, Everatt J, Scheepers C, Mohale T, Ntuli N, Mahlangu B, Ismail A, Bhiman JN |
| hCoV-19/Germany/BY-RKI-I-104055/2021 | EPI_ISL_1848610 | 2021-04-16 | LabKom - Labor Augsburg MVZ GmbH | Robert Koch Institute | unknown |
| hCoV-19/USA/AZ-ASPHL-580/2020 | EPI_ISL_2597734 | 2020-05-05 | Arizona State Public Health Laboratory | Arizona State Public Health Laboratory | Trung Huynh, Jessica Escobar, Katherine Fullerton, Nobuko Fukushima, Matthew Contursi, Stacy White, Linda Getsinger, Victor Waddell |

| | | | | | |
|---|---|---|---|---|---|
| hCoV-19/India/DL-NCDC-1009145/2020 | EPI_ISL_2459905 | 2020-12-22 | National Centre for Disease Control (NCDC) Biotechnology Division, Delhi | NCDC Delhi, Biotechnology Division INSACOG | Mahesh S Dhar, Kalaiarasan Ponnusamy, Meena Datta, Priyanka Singh,¬†Uma Sharma, Manoj K Singh,¬†Radhakrishnan V. S, Robin Marwal, Hemlata Lall, Hema Gogia, Preeti Madan, Sandhya Kabra, Sujeet K Singh, Partha Rakshit |
| hCoV-19/Madagascar/IPM-00513/2021 | EPI_ISL_1660235 | 2021-01-06 | Virology Unit, Institut Pasteur de Madagascar | Virology Unit, Institut Pasteur de Madagascar | Christian Ranaivoson, Cara E. Brook, Vida Ahyong, Soa Fy Andriamandimby, Vololoniaina Raharinosy, Tsiry Randriambolamanantsoa, Helisoa Razafimanjato, Norosoa Razanajatovo, Michelle Tan, Cristina M. Tato, Joseph L. DeRisi, Jean-Michel Heraud, Philippe Duss |
| hCoV-19/Madagascar/IPM-00566/2021 | EPI_ISL_1660297 | 2021-01-07 | Virology Unit, Institut Pasteur de Madagascar | Virology Unit, Institut Pasteur de Madagascar | Marion Barbet, Sylvie Behillil, Fr√©d√©ric Lemoine, Corinne Maufrais, Christophe Malabat, Emmanuelle Permal, M√©line Bizard, Angela Brisebarre, Camille Capel, Louise Lefran√ßois, Etienne Simon-Lori√®re, Vincent Enouf, Maud Vanpeene, Christian Ranaivoson, |
| hCoV-19/India/DL-NCDC-1709214/2021 | EPI_ISL_2460606 | 2021-03-19 | National Centre for Disease Control (NCDC) Biotechnology Division, Delhi | NCDC Delhi, Biotechnology Division INSACOG | Kalaiarasan Ponnusamy, Meena Datta, Priyanka Singh,¬†Uma Sharma, Hemlata Lall, Manoj K Singh,¬†Radhakrishnan V. S, Robin Marwal, Mahesh S Dhar, Hema Gogia, Preeti Madan, Sandhya Kabra, Sujeet K Singh, Partha Rakshit |
| hCoV-19/India/RJ-NCDC-1809302/2020 | EPI_ISL_2460672 | 2020-10-13 | National Centre for Disease Control (NCDC) Biotechnology Division, Delhi | NCDC Delhi, Biotechnology Division INSACOG | Kalaiarasan Ponnusamy, Meena Datta, Priyanka Singh,¬†Uma Sharma, Hemlata Lall, Manoj K Singh,¬†Radhakrishnan V. S, Robin Marwal, Mahesh S Dhar, Hema Gogia, Preeti Madan, Sandhya Kabra, Sujeet K Singh, Partha Rakshit |
| hCoV-19/India/DL-NCDC-2809334/2021 | EPI_ISL_2461101 | 2021-05-04 | National Centre for Disease Control (NCDC) Biotechnology Division, Delhi | NCDC Delhi, Biotechnology Division INSACOG | Priyanka Singh, Uma Sharma, Hemlata Lall, Manoj K Singh, Radhakrishnan V. S, Robin Marwal, Mahesh S Dhar, Kalaiarasan Ponnusamy, Meena Datta, Hema Gogia, Preeti Madan, Sandhya Kabra, Sujeet K Singh, Partha Rakshit |
| hCoV-19/India/DL-NCDC-2309377/2021 | EPI_ISL_2461162 | 2021-04-12 | National Centre for Disease Control (NCDC) Biotechnology Division, Delhi | NCDC Delhi, Biotechnology Division INSACOG | Hemlata Lall, Manoj K Singh,¬†Radhakrishnan V. S, Robin Marwal, Mahesh S Dhar, Kalaiarasan Ponnusamy, Meena Datta, Priyanka Singh,¬†Uma Sharma, Hema Gogia, Preeti Madan, Sandhya Kabra, Sujeet K Singh, Partha Rakshit |
| hCoV-19/India/MP-NCDC-2509230/2020 | EPI_ISL_2461258 | 2020-09-07 | National Centre for Disease Control (NCDC) Biotechnology Division, Delhi | NCDC Delhi, Biotechnology Division INSACOG | Radhakrishnan V. S, Robin Marwal, Mahesh S Dhar, Kalaiarasan Ponnusamy, Meena Datta, Priyanka Singh,¬†Uma Sharma, Hemlata Lall, Manoj K Singh,¬†Hema Gogia, Preeti Madan, Sandhya Kabra, Sujeet K Singh, Partha Rakshit |
| hCoV-19/India/DL-NCDC-250919/2021 | EPI_ISL_2461383 | 2021-04-13 | National Centre for Disease Control (NCDC) Biotechnology Division, Delhi | NCDC Delhi, Biotechnology Division INSACOG | Manoj K Singh,¬†Radhakrishnan V. S, Robin Marwal, Mahesh S Dhar, Kalaiarasan Ponnusamy, Meena Datta, Priyanka Singh,¬†Uma Sharma, Hemlata Lall, Hema Gogia, Preeti Madan, Sandhya Kabra, Sujeet K Singh, Partha Rakshit |

| | | | | | |
|---|---|---|---|---|---|
| hCoV-19/India/JK-NCDC-260984/2020 | EPI_ISL_2461515 | 2020-12-05 | National Centre for Disease Control (NCDC) Biotechnology Division, Delhi | NCDC Delhi, Biotechnology Division INSACOG | Robin Marwal, Mahesh S Dhar, Kalaiarasan Ponnusamy, Meena Datta, Priyanka Singh,¬†Uma Sharma, Hemlata Lall, Manoj K Singh,¬†Radhakrishnan V. S, Hema Gogia, Preeti Madan, Sandhya Kabra, Sujeet K Singh, Partha Rakshit |
| hCoV-19/India/PB-NCDC-2709371/2021 | EPI_ISL_2461832 | 2021-04-23 | National Centre for Disease Control (NCDC) Biotechnology Division, Delhi | NCDC Delhi, Biotechnology Division INSACOG | Meena Datta, Priyanka Singh,¬†Uma Sharma, Hemlata Lall, Manoj K Singh,¬†Radhakrishnan V. S, Robin Marwal, Mahesh S Dhar, Kalaiarasan Ponnusamy, Hema Gogia, Preeti Madan, Sandhya Kabra, Sujeet K Singh, Partha Rakshit |
| hCoV-19/India/JK-NCDC-27092/2020 | EPI_ISL_2461949 | 2020-11-22 | National Centre for Disease Control (NCDC) Biotechnology Division, Delhi | NCDC Delhi, Biotechnology Division INSACOG | Mahesh S Dhar, Kalaiarasan Ponnusamy, Meena Datta, Priyanka Singh,¬†Uma Sharma, Hemlata Lall, Manoj K Singh,¬†Radhakrishnan V. S, Robin Marwal, Hema Gogia, Preeti Madan, Sandhya Kabra, Sujeet K Singh, Partha Rakshit |
| hCoV-19/India/JK-NCDC-270925/2021 | EPI_ISL_2461954 | 2021-01-03 | National Centre for Disease Control (NCDC) Biotechnology Division, Delhi | NCDC Delhi, Biotechnology Division INSACOG | Mahesh S Dhar, Kalaiarasan Ponnusamy, Meena Datta, Priyanka Singh,¬†Uma Sharma, Hemlata Lall, Manoj K Singh,¬†Radhakrishnan V. S, Robin Marwal, Hema Gogia, Preeti Madan, Sandhya Kabra, Sujeet K Singh, Partha Rakshit |
| hCoV-19/India/JK-NCDC-27095/2020 | EPI_ISL_2461957 | 2020-12-01 | National Centre for Disease Control (NCDC) Biotechnology Division, Delhi | NCDC Delhi, Biotechnology Division INSACOG | Mahesh S Dhar, Kalaiarasan Ponnusamy, Meena Datta, Priyanka Singh,¬†Uma Sharma, Hemlata Lall, Manoj K Singh,¬†Radhakrishnan V. S, Robin Marwal, Hema Gogia, Preeti Madan, Sandhya Kabra, Sujeet K Singh, Partha Rakshit |
| hCoV-19/India/PB-NCDC-2709304/2021 | EPI_ISL_2462018 | 2021-04-23 | National Centre for Disease Control (NCDC) Biotechnology Division, Delhi | NCDC Delhi, Biotechnology Division INSACOG | Meena Datta, Priyanka Singh,¬†Uma Sharma, Hemlata Lall, Manoj K Singh,¬†Radhakrishnan V. S, Robin Marwal, Mahesh S Dhar, Kalaiarasan Ponnusamy, Hema Gogia, Preeti Madan, Sandhya Kabra, Sujeet K Singh, Partha Rakshit |
| hCoV-19/Australia/SA0704/2021 | EPI_ISL_2462347 | 2021-04-25 | SA Pathology | SA Pathology | Caitlin Selway, Lex Leong, Chuan Kok Lim, Mark Turra, Ivan Bastian, Geoff Higgins |
| hCoV-19/Australia/SA0727/2021 | EPI_ISL_2462363 | 2021-05-23 | SA Pathology | SA Pathology | Caitlin Selway, Lex Leong, Chuan Kok Lim, Mark Turra, Ivan Bastian, Geoff Higgins |
| hCoV-19/Belgium/UMONS-J70-32319-01/2021 | EPI_ISL_2756196 | 2021-06-11 | National Platform bis UMONS/Jolimont | National Platform bis UMONS/Jolimont | Fran√ßois Dufrasne, Guillaume Bayon-Vicente, Florian Juszczak, Eric Tarantino, Gautier Detry, Ruddy Wattiez |
| hCoV-19/India/OR-ILSGS00655/2021 | EPI_ISL_1662437 | 2021-03-25 | Capital hospital, Bhubaneswar | Institute of Life Sciences - INSACOG | Sunil K. Raghav, Safal Walia, Arup Ghosh, Atimukta Jha, Amol M. Kanampalliwar, Shifu Aggarwal, Rupesh Dash, Rajeeb Swain, Punit Prasad, INSACOG Consortium, Ajay Parida |
| hCoV-19/India/OR-ILSGS00682/2021 | EPI_ISL_1662451 | 2021-03-18 | Immunogenomics lab, Institute of Life Sciences, Bhubaneswar | Institute of Life Sciences - INSACOG | Sunil K. Raghav, Safal Walia, Arup Ghosh, Atimukta Jha, Amol M. Kanampalliwar, Shifu Aggarwal, Rupesh Dash, Rajeeb Swain, Punit Prasad, INSACOG Consortium, Ajay Parida |

| | | | | | |
|---|---|---|---|---|---|
| hCoV-19/USA/NJ-CDC-ASC210039868/2021 | EPI_ISL_2600023 | 2021-03-31 | Aegis Sciences Corporation | Centers for Disease Control and Prevention Division of Viral Diseases, Pathogen Discovery | Dakota Howard, Dhwani Batra, Peter W. Cook, Kara Moser, Adrian Paskey, Jason Caravas, Benjamin Rambo-Martin, Shatavia Morrison, Christopher Gulvick, Scott Sammons, Yvette Unoarumhi, Darlene Wagner, Matthew Schmerer, Cyndi Clark, Patrick Campbell, Rob Case |
| hCoV-19/Italy/UMB-IZSGC-282514.1.12/2021 | EPI_ISL_2600539 | 2021-05-25 | Universit√† degli Studi di Perugia | Istituto Zooprofilattico Sperimentale dell‚ÄôAbruzzo e Molise ‚ÄúG. Caporale‚Äù | Mencacci A, Camilloni B, Biagetti M, Giammarioli M, Lorusso A, Marcacci M, Di Domenico M, Ancora M, Curini V, Mangone I, Rinaldi A, Delli Compagni E, Scialabba S, Di Pasquale A, Camm√† C, Puglia I, Calistri P, Savini G |
| hCoV-19/Kenya/C92031/2021 | EPI_ISL_2602562 | 2021-04-12 | KEMRI-Wellcome Trust Research Programme,Kilifi | KEMRI-Wellcome Trust Research Programme,Kilifi | Githinji G.,Mohamed K.S.,de Laurent Z.,Mburu M.W.,Matoke D.,Onyango C., |
| hCoV-19/Kenya/C93178/2021 | EPI_ISL_2602612 | 2021-04-23 | KEMRI-Wellcome Trust Research Programme,Kilifi | KEMRI-Wellcome Trust Research Programme,Kilifi | Githinji G.,Mohamed K.S.,de Laurent Z.,Mburu M.W.,Matoke D.,Onyango C., |
| hCoV-19/Kenya/C98175/2021 | EPI_ISL_2602742 | 2021-05-26 | KEMRI-Wellcome Trust Research Programme,Kilifi | KEMRI-Wellcome Trust Research Programme,Kilifi | Githinji G.,Mohamed K.S.,de Laurent Z.,Mburu M.W.,Matoke D.,Onyango C., |
| hCoV-19/Kenya/SS311/2021 | EPI_ISL_2603039 | 2021-04-29 | KEMRI-Wellcome Trust Research Programme,Kilifi | KEMRI-Wellcome Trust Research Programme,Kilifi | Githinji G.,Mohamed K.S.,de Laurent Z.,Mburu M.W.,Matoke D.,Onyango C., |
| hCoV-19/Kenya/SS318/2021 | EPI_ISL_2603045 | 2021-04-29 | KEMRI-Wellcome Trust Research Programme,Kilifi | KEMRI-Wellcome Trust Research Programme,Kilifi | Githinji G.,Mohamed K.S.,de Laurent Z.,Mburu M.W.,Matoke D.,Onyango C., |
| hCoV-19/Kenya/SS297/2021 | EPI_ISL_2603048 | 2021-05-03 | KEMRI-Wellcome Trust Research Programme,Kilifi | KEMRI-Wellcome Trust Research Programme,Kilifi | Githinji G.,Mohamed K.S.,de Laurent Z.,Mburu M.W.,Matoke D.,Onyango C., |
| hCoV-19/Kenya/SS366/2021 | EPI_ISL_2603058 | 2021-04-07 | KEMRI-Wellcome Trust Research Programme,Kilifi | KEMRI-Wellcome Trust Research Programme,Kilifi | Githinji G.,Mohamed K.S.,de Laurent Z.,Mburu M.W.,Matoke D.,Onyango C., |
| hCoV-19/Ecuador/UEES-HLV-45/2021 | EPI_ISL_2604888 | 2021-06-10 | Omics Sciences Laboratory | Omics Sciences Laboratory | Derly Andrade Molina, Gabriel Morey Le√≥n, Darlyn Amaya, Rub√©n Armas Gonz√°lez, Juan Carlos Fern√°ndez Cadena |
| hCoV-19/Brazil/RJ-FIOCRUZ-25683/2021 | EPI_ISL_2466268 | 2021-05-24 | Laboratorio Central de Saude Publica do Estado do Rio de Janeiro (LACEN-RJ) | Laboratory of Respiratory Viruses and Measles, Oswaldo Cruz Institute, FIOCRUZ | Paola Resende, Luciana Appolinario, Fernando Motta, Anna Carolina Paixao, Ana Carolina Mendonca, Alice Sampaio Rocha, Taina Venas, Elisa Cavalcante Pereira, Renata Serrano Lopes, Andrea Cony Cavalcanti, Marilda Siqueira on behalf of the Fiocruz COVID-19 G |
| hCoV-19/Angola/CERI-KRISPK013397/2021 | EPI_ISL_2609530 | 2021-03-30 | Instituto Nacional de Investiga√ß√£o em Sa√∫de | KRISP, KZN Research Innovation and Sequencing Platform | Morais J, Neto Z, Afonso P, Miranda J, David K, Ingl√™s L, Pereira A, Paulo A Carralero RR Paix√£o JP, Freitas RH, Mufinda M, Lutucuta S, Giandhari J, Pillay S, Naidoo Y, Emmanuel SJ, Tegally H, Wilkinson E, de Oliveira T |
| hCoV-19/Angola/CERI-KRISPK013566/2021 | EPI_ISL_2609548 | 2021-04-01 | Instituto Nacional de Investiga√ß√£o em Sa√∫de | KRISP, KZN Research Innovation and Sequencing Platform | Morais J, Neto Z, Afonso P, Miranda J, David K, Ingl√™s L, Pereira A, Paulo A Carralero RR Paix√£o JP, Freitas RH, Mufinda M, Lutucuta S, Giandhari J, Pillay S, Naidoo Y, Emmanuel SJ, Tegally H, Wilkinson E, de Oliveira T |
| hCoV-19/Netherlands/NH-RIVM-38972/2021 | EPI_ISL_2610433 | 2021-05-31 | Dutch COVID-19 response team | National Institute for Public Health and the Environment (RIVM) | Adam Meijer, Harry Vennema, Dirk Eggink, Jeroen Cremer, Sharon van den Brink, Bas van der Veer, AnneMarie van den Brandt, Lisa Wijsman, Kim Freriks, Ryanne Jaarsma, Eun√≠ce Then, Lynn Aarts, Sanne Bos, Melissa van Tuil, Linda van de Nes, Florian Zwagemake |

| | | | | | |
|---|---|---|---|---|---|
| hCoV-19/India/ILSGS00931/2021 | EPI_ISL_1663507 | 2021-03-16 | AIIMS, Patna | Institute of Life Sciences - INSACOG | Sunil K. Raghav, Safal Walia, Arup Ghosh, Atimukta Jha, Amol M. Kanampalliwar, Shifu Aggarwal, Rupesh Dash, Rajeeb Swain, Punit Prasad, INSACOG Consortium, Ajay Parida |
| hCoV-19/India/ILSGS00941/2020 | EPI_ISL_1663516 | 2020-12-12 | AIIMS, Patna | Institute of Life Sciences - INSACOG | Sunil K. Raghav, Safal Walia, Arup Ghosh, Atimukta Jha, Amol M. Kanampalliwar, Shifu Aggarwal, Rupesh Dash, Rajeeb Swain, Punit Prasad, INSACOG Consortium, Ajay Parida |
| hCoV-19/India/ILSGS00977/2021 | EPI_ISL_1663548 | 2021-02-19 | MGM Medical College, Jamshedpur | Institute of Life Sciences - INSACOG | Sunil K. Raghav, Safal Walia, Arup Ghosh, Atimukta Jha, Amol M. Kanampalliwar, Shifu Aggarwal, Rupesh Dash, Rajeeb Swain, Punit Prasad, INSACOG Consortium, Ajay Parida |
| hCoV-19/India/ILSGS00978/2021 | EPI_ISL_1663549 | 2021-02-19 | MGM Medical College, Jamshedpur | Institute of Life Sciences - INSACOG | Sunil K. Raghav, Safal Walia, Arup Ghosh, Atimukta Jha, Amol M. Kanampalliwar, Shifu Aggarwal, Rupesh Dash, Rajeeb Swain, Punit Prasad, INSACOG Consortium, Ajay Parida |
| hCoV-19/Mexico/TAM-INMEGEN-06-01-222/2021 | EPI_ISL_2616931 | 2021-06-02 | Salud Digna | Instituto Nacional de Medicina Genomica | Hidalgo-Miranda A, Cedro-Tanda A, Mendoza-Vargas A, Reyes-Grajeda JP, Abraham Campos-Romero, Moreno-Camacho Jos√© Luis, Rodriguez-Gallegos Jorge, Luna-Ruiz Marco, Gonzalez-Barrera D, Rangel-DeLeon D, Munguia-Garza P, Ramirez-Vega O, Escobar-Arrazola, M, H |
| hCoV-19/Chile/RM-0263-04929326/2021 | EPI_ISL_2467138 | 2021-04-24 | Clinica INDISA | Facultad de Ciencias de la Vida, UNAB | Claudio Meneses, Eduardo Castro, Waldo Diaz, Macarena Bastias, Ricardo Yusta, Susana Saez, Dayan Sanhueza, Sebastian Wolter, Daniel Leon, Gonzalo Campa√±a, Paola Pidal, V√≠ctor Monreal, Claudio Olmos, Ariel Orellana |
| hCoV-19/Angola/CERI-KRISP-K015459/2021 | EPI_ISL_2617133 | 2021-05-05 | Instituto Nacional de Investiga√ß√£o em Sa√∫de | and Innvoation, Stellenbosch University and KRISP, KZN Research Innovation and Sequencing Platform, UKZN. | Morais J, Neto Z, Afonso P, Miranda J, David K, Ingl√™s L, Pereira A, Paulo A Carralero RR Paix√£o JP, Freitas RH, Mufinda M, Lutucuta S, Giandhari J, Pillay S, Naidoo Y, Emmanuel SJ, Tegally H, Wilkinson E, de Oliveira T |
| hCoV-19/Chile/RM-0263-04973530/2021 | EPI_ISL_2467251 | 2021-05-05 | Clinica INDISA | Facultad de Ciencias de la Vida, UNAB | Claudio Meneses, Eduardo Castro, Waldo Diaz, Macarena Bastias, Ricardo Yusta, Susana Saez, Dayan Sanhueza, Sebastian Wolter, Daniel Leon, Gonzalo Campa√±a, Paola Pidal, V√≠ctor Monreal, Claudio Olmos, Ariel Orellana |
| hCoV-19/Chile/RM-0705-05012073/2021 | EPI_ISL_2467360 | 2021-05-13 | Clinica INDISA | Facultad de Ciencias de la Vida, UNAB | Claudio Meneses, Eduardo Castro, Waldo Diaz, Macarena Bastias, Ricardo Yusta, Susana Saez, Dayan Sanhueza, Sebastian Wolter, Daniel Leon, Gonzalo Campa√±a, Paola Pidal, V√≠ctor Monreal, Claudio Olmos, Ariel Orellana |
| hCoV-19/USA/WA-UW-21051308994/2021 | EPI_ISL_2618698 | 2021-05-13 | UW Virology Lab | UW Virology Lab | Pavitra Roychoudhury, Hong Xie, Lasata Shrestha, Shah Mohamed Bakhash, Tien V. Nguyen, Noah R. Baker, Sean Ellis, Meei-Li Huang, Keith R Jerome, Alexander Greninger |
| hCoV-19/Sweden/20352014P/2021 | EPI_ISL_2619612 | 2021-05-31 | Swedish national genomic surveillance program of SARS-CoV-2 | The Public Health Agency of Sweden | Maximilian Riess, Maria Lind Karlberg, Alma Brolund, Swedish national genomic surveillance program of SARS-CoV-2 |

| | | | | | |
|---|---|---|---|---|---|
| hCoV-19/SouthAfrica/NHLS-UCT-GS-3543/2020 | EPI_ISL_2621093 | 2020-10-10 | Conville CDC wc CVC | NHLS/UCT | Arash Iranzadeh, Deelan Doolabh, Lynn Tyers, Bruna Galvao, Innocent Mudau, Marvin Hsiao, Gert Marais, Diana Hardie, Stephen Korsman, Carolyn Williamson |
| hCoV-19/Japan/IC-1338/2021 | EPI_ISL_2621749 | 2021-06-05 | SARS-CoV-2 testing team, National Institute of Infectious Diseases | Pathogen Genomics Center, National Institute of Infectious Diseases | Tsuyoshi Sekizuka, Kentaro Itokawa, Rina Tanaka, Masanori Hashino, Nozomu Hanaoka, Masumichi Saito, Naomi Nojiri, Hazuka Y Furihata, Sana Uchikoba, Hiromizu Takahashi, Tsuguto Fujimoto, Makoto Kuroda |
| hCoV-19/Netherlands/DR-EMC-9/2020 | EPI_ISL_523127 | 2020-06-03 | Dutch COVID-19 response team | Erasmus Medical Center | Bas Oude Munnink, David Nieuwenhuijse, Reina Sikkema, Claudia Schapendonk, Irina Chestakova, Anne van der Linden, Theo Bestebroer, Stefan van Nieuwkoop, Mark Pronk, Pascal Lexmond, Corien Swaan, Manon Haverkate, Madelief Mollers, Mart Stein, Sandra Kengne |
| hCoV-19/Netherlands/GE-EMC-299/2020 | EPI_ISL_523278 | 2020-06-05 | Dutch COVID-19 response team | Erasmus Medical Center | Bas Oude Munnink, David Nieuwenhuijse, Reina Sikkema, Claudia Schapendonk, Irina Chestakova, Anne van der Linden, Theo Bestebroer, Stefan van Nieuwkoop, Mark Pronk, Pascal Lexmond, Corien Swaan, Manon Haverkate, Madelief Mollers, Mart Stein, Sandra Kengne |
| hCoV-19/NewZealand/20VR3911/2020 | EPI_ISL_548103 | 2020-08-13 | Middlemore Hospital | Institute of Environmental Science and Research (ESR) | Xiaoyun Ren, Matt Storey, Nikki Freed, Muhammad Faisal, Jing Wang, Hermes Perez, Anja Werno, Antje van der Linden, Arlo Upton, Chris Mansell, David Hammer, Dragana Drinkovic, Gary McAuliffe, Hana Sofia Andersson, James Ussher, Jill Sherwood, Josh Freeman, |
| hCoV-19/Australia/VIC6995/2020 | EPI_ISL_562463 | 2020-07-23 | Victorian Infectious Diseases Reference Laboratory (VIDRL) | VIDRL and MDU-PHL | Caly, L., Seemann, T., Sait, M., Schultz, M. B., Druce J., Sherry, N. |
| hCoV-19/Kenya/C6934/2020 | EPI_ISL_568818 | 2020-05-15 | KEMRI-Wellcome Trust Research Programme/KEMRI-CGMR-C Kilifi | KEMRI-Wellcome Trust Research Programme/KEMRI-CGMR-C Kilifi | Githinji et al 2020 |
| hCoV-19/England/NORW-13DF9AF/2021 | EPI_ISL_2624214 | 2021-05-21 | Quadram Institute Bioscience | COVID-19 Genomics UK (COG-UK) Consortium | Dave J. Baker, Gemma L. Kay, Alp Aydin, Thanh Le-Viet, Steven Rudder, Ana P. Tedim, Anastasia Kolyva, Maria Diaz, Leonardo de Oliveira Martins, Nabil-Fareed Alikhan, Lizzie Meadows, Rachael Stanley, Ngozi Elumogo, Muhammed Yasir, Nicholas M. Thomson, Alex |
| hCoV-19/NewZealand/20VR3060/2020 | EPI_ISL_579403 | 2020-05-02 | LabPLUS | Institute of Environmental Science and Research (ESR) | Xiaoyun Ren, Matt Storey, Nikki Freed, Muhammad Faisal, Jing Wang, Hermes Perez, Anja Werno, Antje van der Linden, Arlo Upton, Chris Mansell, David Hammer, Dragana Drinkovic, Gary McAuliffe, Hana Sofia Andersson, James Ussher, Jill Sherwood, Josh Freeman, |
| hCoV-19/England/SHEF-10E356B/2021 | EPI_ISL_2625431 | 2021-06-12 | Virology Department, Sheffield Teaching Hospitals NHS Foundation Trust/Department of Infection, Immunity and Cardiovascular Disease, The Medical School, University of Sheffield | COVID-19 Genomics UK (COG-UK) Consortium | Thushan de Silva, Matthew Parker, Nikki Smith, Adri Angyal, Rebecca Brown, Luke Green, Rachel Tucker, Paul Parsons, Danielle Groves, Katie Johnson, Laura Carrilero, Alex Keeley, Dave Partridge, Matthew Wyles, Benjamin Lindsey, Mehmet Yavuz, Mohammad Raza, |
| hCoV-19/Portugal/PT8722/2021 | EPI_ISL_2628943 | 2021-06-06 | UNILABS | Instituto Nacional de Saude (INSA) | Borges et al |
| hCoV-19/UnitedArabEmirates/skmc-3122376/2020 | EPI_ISL_582649 | 2020-08-29 | Sheikh Khalifa Medical City | Molecular/Surveillance lab Sheikh Khalifa Medical City | Amirtharaj Francis, Sajeed Abdul, Hala Imambaccus, Sahar Almarzooqi, Hiba Saud, Stefan Weber |

| | | | | | |
|---|---|---|---|---|---|
| hCoV-19/Australia/WA680/2021 | EPI_ISL_1669119 | 2021-04-16 | PathWest Laboratory Medicine WA | PathWest Laboratory Medicine WA Microbial Surveillance Unit | PathWest Laboratory Medicine WA Microbial Surveillance Unit |
| hCoV-19/Australia/WA683/2021 | EPI_ISL_1669122 | 2021-04-17 | PathWest Laboratory Medicine WA | PathWest Laboratory Medicine WA Microbial Surveillance Unit | PathWest Laboratory Medicine WA Microbial Surveillance Unit |
| hCoV-19/England/ALDP-9E92FF/2020 | EPI_ISL_584201 | 2020-09-24 | Centre for Enzyme Innovation, University of Portsmouth / Translational Research Laboratory, Portsmouth Hospitals NHS Trust | COVID-19 Genomics UK (COG-UK) Consortium | Angela Beckett,Yann Bourgeois,Garry Scarlett,Sharon Glaysher,Scott Elliott,Kelly Bicknell,Robert Impey,Allyson Lloyd,Sarah Wyllie,Ethan Butcher,Anoop Chauhan,Samuel Robson |
| hCoV-19/USA/CA-ALSR-102007/2020 | EPI_ISL_2628489 | 2020-07-08 | San Diego County Public Health Laboratory | Andersen lab at Scripps Research | SEARCH Alliance San Diego with Ashleigh Murphy, Jovan Shephard, Brett Austin |
| hCoV-19/USA/CA-ALSR-102014/2021 | EPI_ISL_2628494 | 2021-05-18 | San Diego County Public Health Laboratory | Andersen lab at Scripps Research | SEARCH Alliance San Diego with Ashleigh Murphy, Jovan Shephard, Brett Austin |
| hCoV-19/Portugal/PT8623/2021 | EPI_ISL_2628806 | 2021-06-04 | UNILABS | Instituto Nacional de Saude (INSA) | Borges et al |
| hCoV-19/USA/GA-GPHL-0495/2021 | EPI_ISL_2629587 | 2021-04-29 | GA Department of Public Health | GA Department of Public Health | Stacy Reeves, Jonathan Edwards, Cynthia Dixey, Tonia Parrott, Aliyah Fields, Taylor Smith |
| hCoV-19/Aruba/AW-RIVM-36938/2021 | EPI_ISL_2475379 | 2021-05-25 | Dutch COVID-19 response team | National Institute for Public Health and the Environment (RIVM) | Adam Meijer, Harry Vennema, Dirk Eggink, Jeroen Cremer, Sharon van den Brink, Bas van der Veer, AnneMarie van den Brandt, Lisa Wijsman, Kim Freriks, Ryanne Jaarsma, Eun√≠ce Then, Lynn Aarts, Sanne Bos, Melissa van Tuil, Linda van de Nes, Florian Zwagemake |
| hCoV-19/Netherlands/GE-RIVM-37308/2021 | EPI_ISL_2475713 | 2021-05-28 | Dutch COVID-19 response team | National Institute for Public Health and the Environment (RIVM) | Adam Meijer, Harry Vennema, Dirk Eggink, Jeroen Cremer, Sharon van den Brink, Bas van der Veer, AnneMarie van den Brandt, Lisa Wijsman, Kim Freriks, Ryanne Jaarsma, Eun√≠ce Then, Lynn Aarts, Sanne Bos, Melissa van Tuil, Linda van de Nes, Florian Zwagemake |
| hCoV-19/Israel/CVL-9072/2021 | EPI_ISL_2084111 | 2021-03-01 | Israel Central Virology laboratory | Israel National Consortium for SARS-CoV-2 sequencing | Neta Zuckerman, Efrat Dahan Bucris, Michal Mandelboim, Dana Bar-Ilan, Oran Erster, Tzvia Mann, Omer Murik, David A. Zeevi, Assaf Rokney, Joseph Jaffe, Eva Nachum, Maya Davidovich Cohen, Ephraim Fass, Gal Zizelski Valenci, Mor Rubinstein, Efrat Rorman, Isr |
| hCoV-19/Israel/CVL-9199/2021 | EPI_ISL_2084225 | 2021-03-01 | Israel Central Virology laboratory | Israel National Consortium for SARS-CoV-2 sequencing | Neta Zuckerman, Efrat Dahan Bucris, Michal Mandelboim, Dana Bar-Ilan, Oran Erster, Tzvia Mann, Omer Murik, David A. Zeevi, Assaf Rokney, Joseph Jaffe, Eva Nachum, Maya Davidovich Cohen, Ephraim Fass, Gal Zizelski Valenci, Mor Rubinstein, Efrat Rorman, Isr |
| hCoV-19/USA/NY-PRL-2021_0611_52N01/2020 | EPI_ISL_2633778 | 2020-12-21 | Pandemic Response Lab - NYC | Pandemic Response Lab, R&D | Henry Lee, Michael Hammerling, Melissa Hopkins, Cybill del Castillo, Shinyoung Clair Kang, William Ward, Pradeep Bugga, Sol Rey, Dylan Law, Katharine Nelson, Haiping Hao, Jon Laurent |
| hCoV-19/USA/FL-CDC-LC0065550/2021 | EPI_ISL_2480454 | 2021-05-29 | Laboratory Corporation of America | Centers for Disease Control and Prevention Division of Viral Diseases, Pathogen Discovery | Dakota Howard, Dhwani Batra, Peter W. Cook, Kara Moser, Adrian Paskey, Jason Caravas, Benjamin Rambo-Martin, Shatavia Morrison, Christopher Gulvick, Scott Sammons, Yvette Unoarumhi, Darlene Wagner, Matthew Schmerer, Minoo Agarwal, Eyad Almasri, Debbie Bol |

| | | | | | |
|---|---|---|---|---|---|
| hCoV-19/Israel/CVL-4682/2021 | EPI_ISL_1674134 | 2021-01-28 | Israel Central Virology laboratory | Israel National Consortium for SARS-CoV-2 sequencing | Neta Zuckerman, Efrat Dahan Bucris, Michal Mandelboim, Dana Bar-Ilan, Oran Erster, Tzvia Mann, Omer Murik, David A. Zeevi, Assaf Rokney, Joseph Jaffe, Eva Nachum, Maya Davidovich Cohen, Ephraim Fass, Gal Zizelski Valenci, Mor Rubinstein, Efrat Rorman, Is |
| hCoV-19/Australia/VIC18653/2021 | EPI_ISL_2482444 | 2021-06-02 | Microbiological Diagnostic Unit - Public Health Laboratory (MDU-PHL) | MDU-PHL | Seemann T., Sait, M.L., Sherry, N.L. |
| hCoV-19/Belgium/ULG-16287/2021 | EPI_ISL_2483898 | 2021-05-30 | SYNLAB | GIGA Medical Genomics | Keith Durkin, Maria Artesi, S√©bastien Bontems, Rapha√´l Boreux, Bouchra Boujemla, Nathalie Renotte, C√©cile Meex, Pierrette Melin, Marie-Pierre Hayette, Vincent Bours |
| hCoV-19/Colombia/ANT-CWOHC-VG-SEC00092N/2021 | EPI_ISL_1675304 | 2021-03-04 | UDEA | Universidad Nacional de Colombia - Laboratorio Gen√≥mico One Health | Sim√≥n Villegas Vel√°squez, Andres F. Cardona-Rios, Maria Stella L√≥pez, Daniel O. Maldonado-Perez, Lina M. Hurtado, Laura Silvana Perez, Karl A Ciuoderis, Maria Ang√©lica Maya, Rita Almanza Payares, Idabely Betancur Ortiz, Sandra Ines Cano, Diego A. √Ålv |
| hCoV-19/Switzerland/GL-UHB-727017500/2021 | EPI_ISL_2484361 | 2021-01-30 | Labormedizinisches Zentrum Dr Risch | Clinical Bacteriology | Tim Roloff, Fanny Wegner, Helena MB Seth-Smith, Alfredo Mari, Karoline Leuzinger, Julia Bielicki, Nadia Wohlwend,Martin Risch, Lorenz Risch, Manuel Battegay, Hans Hirsch, Adrian Egli |
| hCoV-19/India/WB-1931300315544/2021 | EPI_ISL_2548856 | 2021-05-06 | INSACOG-WB | National Institute of Biomedical Genomics ‚Äì INSACOG | Arindam Maitra, Bhaswati Bandyopadhyay, Nidhan Kumar Biswas, Tamal Ghosh, Sreedhar Chinnaswamy, Ajay Chakraborti, Saumitra Das |
| hCoV-19/Switzerland/GR-UHB-4182029201/2021 | EPI_ISL_2484480 | 2021-01-30 | Labormedizinisches Zentrum Dr Risch | Clinical Bacteriology | Tim Roloff, Fanny Wegner, Helena MB Seth-Smith, Alfredo Mari, Karoline Leuzinger, Julia Bielicki, Nadia Wohlwend,Martin Risch, Lorenz Risch, Manuel Battegay, Hans Hirsch, Adrian Egli |
| hCoV-19/Italy/CAM-TIGEM-IZSM-COLLI-16088/2021 | EPI_ISL_2641871 | 2021-06-09 | Istituto Zooprofilattico Sperimentale del Mezzogiorno | Telethon Institute of Genetics and Medicine (TIGEM) | Antonio Grimaldi Patrizia Annunziata Francesco Panariello Biancamaria Pierri Claudia Tiberio Teresa Giuliano Valentina Bouche Chiara Colantuono Maria Concetta Cuomo Denise Di Concilio Lucio Di Filippo Anna Manfredi Marcello Salvi Antonio Limone Luigi Atri |
| hCoV-19/USA/CO-CDC-MMB08704173/2021 | EPI_ISL_2646411 | 2021-05-07 | Mako Medical | Centers for Disease Control and Prevention Division of Viral Diseases, Pathogen Discovery | Dakota Howard, Dhwani Batra, Peter W. Cook, Kara Moser, Adrian Paskey, Jason Caravas, Benjamin Rambo-Martin, Shatavia Morrison, Christopher Gulvick, Scott Sammons, Yvette Unoarumhi, Darlene Wagner, Matthew Schmerer,Matthew Tugwell, Lauren Moon, Clinton R. |
| hCoV-19/Spain/CL-COV02253/2021 | EPI_ISL_2649056 | 2021-04-15 | SARS-CoV-2 Sequencing Castilla y Leon-Spain Consortium | SARS-CoV-2 Sequencing Castilla y Leon-Spain Consortium | Antonio Ordu√±a-Domingo, Marta Hernandez, David Abad, Marta Dominguez-Gil, Silvia Rojo, Laura S√°nchez de Prada, Sonsoles Garcinu√±o P√©rez, Carmen Aldea-Mansilla, M¬™ Fe Brezmes-Valdivieso, Gregoria Meg√≠as Lob√≥n, Mar√≠a Antonia Garc√≠a Castro, Carmen G |
| hCoV-19/Australia/NSW1648/2021 | EPI_ISL_2650469 | 2021-06-15 | Sydney South West Pathology Service (SSWPS) - Royal Prince Alfred Hospital - NSW Health Pathology | Clinical Pathology and Medical Research; Westmead Hospital; University of Sydney | CIDM-PH et al. |

| | | | | | |
|---|---|---|---|---|---|
| hCoV-19/Italy/PIE-15840830/2021 | EPI_ISL_2650652 | 2021-04-20 | A.S.L. CITTA' DI TORINO - OSPEDALE AMEDEO DI SAVOIA - SC MICROBIOLOGIA E VIROLOGIA - | Fondazione del Piemonte per l'Oncologia IRCCS | Antonino Sottile, Silvia Brossa, Paola Marino, Giorgio Giardina |
| hCoV-19/Italy/PIE-15826615/2021 | EPI_ISL_2650852 | 2021-03-18 | AZ. OSPEDAL. S. CROCE E CARLE - S.C. LABORATORIO ANALISI CHIMICO CLINICHE E MICROBIOLOGIA | Fondazione del Piemonte per l'Oncologia IRCCS | Antonino Sottile, Silvia Brossa, Paola Marino, Giorgio Giardina |
| hCoV-19/Scotland/QEUH-1628BCC/2021 | EPI_ISL_2486443 | 2021-06-03 | Lighthouse Lab in Glasgow | Wellcome Sanger Institute for the COVID-19 Genomics UK (COG-UK) Consortium | Harper VanSteenhouse, Yumi Kasai, David Gray, Carol Clugston, Anna Dominiczak and Alex Alderton, Roberto Amato, Jeffrey Barrett, Sonia Goncalves, Ewan Harrison, David K. Jackson, Ian Johnston, Dominic Kwiatkowski, Cordelia Langford, John Sillitoe on behal |
| hCoV-19/USA/CA-ALSR-2843/2020 | EPI_ISL_635709 | 2020-08-06 | San Diego County Public Health Laboratory | Andersen lab at Scripps Research | SEARCH Alliance San Diego with Tracy Basler, Jovan Shephard, Brett Austin |
| hCoV-19/Ecuador/USFQ-1478/2021 | EPI_ISL_2488762 | 2021-05-31 | Institute of Microbiology, Universidad San Francisco de Quito | Institute of Microbiology, Universidad San Francisco de Quito | Rommel Guevara, Juan Jos√© Guadalupe, Sully M√°rquez, Bel√©n Prado-Vivar, Fernanda Zurita, Monica Becerra-Wong, Bernardo Guti√©rrez, Manuel Jibaja, Ver√≥nica Barrag√°n, Patricio Rojas-Silva, Gabriel Trueba, Michelle Grunauer, Pa√∫l C√°rdenas |
| hCoV-19/Australia/WA495/2020 | EPI_ISL_708789 | 2020-11-24 | PathWest Laboratory Medicine WA | PathWest Laboratory Medicine WA Microbial Surveillance Unit | PathWest Laboratory Medicine WA Microbial Surveillance Unit |
| hCoV-19/USA/IL-CDC-ASC210105888/2021 | EPI_ISL_2489711 | 2021-05-27 | Aegis Sciences Corporation | Centers for Disease Control and Prevention Division of Viral Diseases, Pathogen Discovery | Dakota Howard, Dhwani Batra, Peter W. Cook, Kara Moser, Adrian Paskey, Jason Caravas, Benjamin Rambo-Martin, Shatavia Morrison, Christopher Gulvick, Scott Sammons, Yvette Unoarumhi, Darlene Wagner, Matthew Schmerer, Cyndi Clark, Patrick Campbell, Rob Case |
| hCoV-19/Belgium/ULG-15040/2021 | EPI_ISL_1970770 | 2021-04-21 | SYNLAB | GIGA Medical Genomics | Keith Durkin, Maria Artesi, S√©bastien Bontems, Rapha√´l Boreux, Bouchra Boujemla, Nathalie Renotte, C√©cile Meex, Pierrette Melin, Marie-Pierre Hayette, Vincent Bours |
| hCoV-19/Zimbabwe/CERI-KRISP-K011643/2020 | EPI_ISL_2492577 | 2020-07-09 | National Microbiology Reference Laboratory, Ministry of Health, Harare, Zimbabwe | CERI, Centre for Epidemic Response and Innvoation, Stellenbosch University and KRISP, KZN Research Innovation and Sequencing Platform, UKZN. | Air Comodor Dr J. Chimedza,¬†Dr Raiva Simbi,¬†Agnes Juru,¬†Tapfumanei Mashe,¬†Kenneth Maeka,¬†Hlanai Gumbo,¬†Tatenda Takawira,¬†Charles Nyagupe, Giandhari J, Pillay S, Naidoo Y, Emmanuel SJ, Tegally H, Wilkinson E, de Oliveira T |
| hCoV-19/Angola/CERI-KRISP-K013423/2020 | EPI_ISL_2492777 | 2020-08-25 | Instituto Nacional de Investiga√ß√£o em Sa√∫de | and Innvoation, Stellenbosch University and KRISP, KZN Research Innovation and Sequencing Platform, UKZN. | Morais J, Neto Z, Afonso P, Miranda J, David K, Ingl√™s L, Pereira A, Paulo A Carralero RR Paix√£o JP, Freitas RH, Mufinda M, Lutucuta S, Giandhari J, Pillay S, Naidoo Y, Emmanuel SJ, Tegally H, Wilkinson E, de Oliveira T |
| hCoV-19/Angola/CERI-KRISP-K013436/2021 | EPI_ISL_2492870 | 2021-04-12 | Instituto Nacional de Investiga√ß√£o em Sa√∫de | and Innvoation, Stellenbosch University and KRISP, KZN Research Innovation and Sequencing Platform, UKZN. | Morais J, Neto Z, Afonso P, Miranda J, David K, Ingl√™s L, Pereira A, Paulo A Carralero RR Paix√£o JP, Freitas RH, Mufinda M, Lutucuta S, Giandhari J, Pillay S, Naidoo Y, Emmanuel SJ, Tegally H, Wilkinson E, de Oliveira T |

| | | | | | |
|---|---|---|---|---|---|
| hCoV-19/Malawi/CERI-KRISP-K015274/2021 | EPI_ISL_2493069 | 2021-04-28 | National HIV Reference Laboratory, Ministry of Health, Public Health Institute of Malawi | CERI, Centre for Epidemic Response and Innvoation, Stellenbosch University and KRISP, KZN Research Innovation and Sequencing Platform, UKZN. | Mvula B, Chilima B, Chiwaula M, Mwangomba W, Panja L, Kasambara W, Auld A, Kim L, Kampira E, Kaba M, Wadonda N, Maida A, Giandhari J, Pillay S, Naidoo Y, Lessells R, Emmanuel SJ, Tegally H, Wilkinson E, de Oliveira T |
| hCoV-19/Turkey/HSGM-B10476/2021 | EPI_ISL_2363606 | 2021-04-26 | Ministry of Health Turkey | Ministry of Health Turkey | Fatma Bayrakdar, Yasemin Cosgun, Suleyman Yalcin, Gulay Korukluoglu |
| hCoV-19/SouthAfrica/VIDA-KRISP-K012837/2020 | EPI_ISL_2494919 | 2020-09-18 | Vaccines and Infectious Diseases Analytics Research Unit (VIDA) | KRISP, KZN Research Innovation and Sequencing Platform | Baillie Vicky, du Plessis Jeanine, Giandhari Jennifer, Pillay Sureshnee, Naidoo Yeshnee, Tegally Houriiyah, San James, Wilkinson Eduan, de Oliveira Tulio, Madhi Shabir |
| hCoV-19/Malawi/CERI-KRISP-K015281/2021 | EPI_ISL_2494966 | 2021-04-30 | National HIV Reference Laboratory, Ministry of Health, Public Health Institute of Malawi | KRISP, KZN Research Innovation and Sequencing Platform | Mvula B, Chilima B, Chiwaula M, Mwangomba W, Panja L, Kasambara W, Auld A, Kim L, Kampira E, Kaba M, Wadonda N, Maida A, Giandhari J, Pillay S, Naidoo Y, Lessells R, Emmanuel SJ, Tegally H, Wilkinson E, de Oliveira T |
| hCoV-19/SouthAfrica/VIDA-KRISP-K012925/2020 | EPI_ISL_2494968 | 2020-09-18 | Vaccines and Infectious Diseases Analytics Research Unit (VIDA) | KRISP, KZN Research Innovation and Sequencing Platform | Baillie Vicky, du Plessis Jeanine, Giandhari Jennifer, Pillay Sureshnee, Naidoo Yeshnee, Tegally Houriiyah, San James, Wilkinson Eduan, de Oliveira Tulio, Madhi Shabir |
| hCoV-19/Malawi/CERI-KRISP-K015315/2021 | EPI_ISL_2495039 | 2021-04-28 | National HIV Reference Laboratory, Ministry of Health, Public Health Institute of Malawi | KRISP, KZN Research Innovation and Sequencing Platform | Mvula B, Chilima B, Chiwaula M, Mwangomba W, Panja L, Kasambara W, Auld A, Kim L, Kampira E, Kaba M, Wadonda N, Maida A, Giandhari J, Pillay S, Naidoo Y, Lessells R, Emmanuel SJ, Tegally H, Wilkinson E, de Oliveira T |
| hCoV-19/France/PAC-IHU-12089_Nova1/2021 | EPI_ISL_2659743 | 2021-06-11 | MEPHI, Aix Marseille University | MEPHI, Aix Marseille University | Anthony LEVASSEUR |
| hCoV-19/France/PAC-IHU-12250_Nova1/2020 | EPI_ISL_2659891 | 2020-05-02 | MEPHI, Aix Marseille University | MEPHI, Aix Marseille University | Anthony LEVASSEUR |
| hCoV-19/India/AP-CCMB-CIA3120/2021 | EPI_ISL_2660770 | 2021-05-15 | AP SSO | CSIR-Centre for Cellular and Molecular Biology-INSACOG | Ara Sreenivas,Shreekant Verma, Amareshwar Vodapalli ,B Himasri,Valli Nagalakshmi Undamatla,Payel Mukherjee, Lamuk Zaveri, Onkar Kulkarni,Tulasi Nagabandi, Sofia Banu, ,Priya Nurkurthy, Vidhyadhari Methuku, Sumedha Avadhanula, Archana Bharadwaj Siva, Karth |
| hCoV-19/India/ML-CCMB-CIA3198/2021 | EPI_ISL_2660871 | 2021-05-07 | PASTEUR MOLECULAR LABORATORY - INSACOG MEGHALAYA | CSIR-Centre for Cellular and Molecular Biology-INSACOG | R. Lyngdoh, E Shadap , E Marak,Tulasi Nagabandi,Lamuk Zaveri,Payel Mukherjee,Onkar Kulkarni, Sofia Banu, Ara Sreenivas,Shreekant Verma, Amareshwar Vodapalli ,B Himasri,Valli Nagalakshmi Undamatla, Priya Nurkurthy,Sumedha Avadhanula, Vidhyadhari Methuku, A |
| hCoV-19/Australia/QLD1938/2021 | EPI_ISL_2661514 | 2021-06-16 | Queensland Health Forensic and Scientific Services | Queensland Health Forensic and Scientific Services | Son Nguyen |
| hCoV-19/Australia/QLD1952/2021 | EPI_ISL_2661522 | 2021-06-20 | Queensland Health Forensic and Scientific Services | Queensland Health Forensic and Scientific Services | Son Nguyen |

| | | | | | |
|---|---|---|---|---|---|
| hCoV-19/Brazil/CE-FIOCRUZ-6574/2021 | EPI_ISL_2661909 | 2021-01-03 | Oswaldo Cruz Institute, FIOCRUZ/CE | Analytical Competence Molecular Epidemiology Lab/ACME, Oswaldo Cruz Foundation, Ceara (FIOCRUZ CE) | Paola Resende, Fabio Miyajima, Luciana Appolinario, Fernando Motta, Anna Carolina Paixao, Ana Carolina Mendonca, Alice Sampaio Rocha, Taina Venas, Elisa Cavalcante Pereira, Renata Serrano Lopes, Marilda Siqueira on behalf of the Fiocruz COVID-19 Genomic S |
| hCoV-19/Canada/BC-BCCDC-37076/2020 | EPI_ISL_2499346 | 2020-12-24 | BCCDC Public Health Laboratory | BCCDC Public Health Laboratory | Prystajecky Natalie, Linda Hoang, Dan Fornika, John Tyson, Shannon Russell, Kim Macdonald, Kimia Kamelian, Ana Pacagnella, Corrinne Ng, Loretta Janz, Robert Azana, Mel Krajden |
| hCoV-19/Canada/BC-BCCDC-37087/2020 | EPI_ISL_2499355 | 2020-12-21 | BCCDC Public Health Laboratory | BCCDC Public Health Laboratory | Prystajecky Natalie, Linda Hoang, Dan Fornika, John Tyson, Shannon Russell, Kim Macdonald, Kimia Kamelian, Ana Pacagnella, Corrinne Ng, Loretta Janz, Robert Azana, Mel Krajden |
| hCoV-19/Canada/BC-BCCDC-37156/2020 | EPI_ISL_2499412 | 2020-12-19 | BCCDC Public Health Laboratory | BCCDC Public Health Laboratory | Prystajecky Natalie, Linda Hoang, Dan Fornika, John Tyson, Shannon Russell, Kim Macdonald, Kimia Kamelian, Ana Pacagnella, Corrinne Ng, Loretta Janz, Robert Azana, Mel Krajden |
| hCoV-19/Indonesia/JK-GSILab-490269/2021 | EPI_ISL_2499963 | 2021-05-28 | Genomik Solidaritas Indonesia Laboratorium | Genomik Solidaritas Indonesia Laboratorium | Vania Gavrila Wikasa, Carissa Sintca Wijaya, Alfin Mohammad Abdillah, Dhahlia Agustina Cahyono, Normastuti Adhini Tantyo, Annisa Muthiah Sukirman, Gracia Felias Enos Korompis, Pratiwi Sudarmono, Budi Wiweko, Meutia Ayuputeri Kumaheri, Anindya Pradipta Sus |
| hCoV-19/Japan/Donner288/2020 | EPI_ISL_1973147 | 2020-10-06 | Keio University School of Medicine | Keio University School of Medicine | Kenjiro Kosaki, Yuka Iwasaki, Hirotsugu Ishizu, Haruhiko Siomi, Kodai Abe |
| hCoV-19/India/MP-NCDC-210947/2021 | EPI_ISL_2502120 | 2021-03-25 | National Centre for Disease Control (NCDC) Biotechnology Division, Delhi | NCDC Delhi, Biotechnology Division INSACOG | Priyanka Singh,¬†Uma Sharma, Hemlata Lall, Manoj K Singh,¬†Radhakrishnan V. S, Robin Marwal, Mahesh S Dhar, Kalaiarasan Ponnusamy, Meena Datta, Hema Gogia, Preeti Madan, Sandhya Kabra, Sujeet K Singh, Partha Rakshit |
| hCoV-19/Germany/BW-RKI-I-011520/2021 | EPI_ISL_1141650 | 2021-01-24 | Institut f√ºr Virologie des Universit√§tsklinikums T√ºbingen | Robert Koch Institute | unknown |
| hCoV-19/Germany/SN-RKI-I-021632/2021 | EPI_ISL_1151797 | 2021-02-15 | St√§dtisches Klinikum Dresden | Robert Koch Institute | unknown |
| hCoV-19/Gambia/NPHL_7981/2020 | EPI_ISL_1216112 | 2020-08-24 | MRCG at LSHTM Genomics lab | MRCG at LSHTM Genomics lab | Abdul Karim sesay, Abdoulie Kanteh, Jarra Manneh, Mariama Kujabi, Bakary Sanyang |
| hCoV-19/Germany/NW-RKI-I-022222/2021 | EPI_ISL_1152388 | 2021-02-05 | Universit√§tsklinikum Leipzig - Institut f√ºr Medizinische Mikrobiologie und Virologie virologisches Labor | Robert Koch Institute | unknown |
| hCoV-19/USA/WA-CDC-LC0043887/2021 | EPI_ISL_1684712 | 2021-04-07 | Laboratory Corporation of America | Centers for Disease Control and Prevention Division of Viral Diseases, Pathogen Discovery | Dakota Howard, Dhwani Batra, Peter W. Cook, Kara Moser, Adrian Paskey, Jason Caravas, Benjamin Rambo-Martin, Shatavia Morrison, Christopher Gulvick, Scott Sammons, Yvette Unoarumhi, Darlene Wagner, Matthew Schmerer, Minoo Agarwal, Eyad Almasri, Debbie Bol |
| hCoV-19/Germany/NW-RKI-I-028631/2021 | EPI_ISL_1211715 | 2021-02-22 | Labor Dr. Wisplinghoff - K√∂ln | Robert Koch Institute | unknown |

| | | | | | |
|---|---|---|---|---|---|
| hCoV-19/USA/WA-UW-21042667869/2021 | EPI_ISL_2502898 | 2021-04-26 | UW Virology Lab | UW Virology Lab | Pavitra Roychoudhury, Hong Xie, Lasata Shrestha, Shah Mohamed Bakhash, Tien V. Nguyen, Noah R. Baker, Sean Ellis, Meei-Li Huang, Keith R Jerome, Alexander Greninger |
| hCoV-19/India/WB-1931300260606/2021 | EPI_ISL_2503770 | 2021-04-07 | INSACOG-WB | National Institute of Biomedical Genomics ,Äì INSACOG | Arindam Maitra, Bhaswati Bandyopadhyay, Nidhan Kumar Biswas, Tamal Ghosh, Sreedhar Chinnaswamy, Ajay Chakraborti, Saumitra Das |
| hCoV-19/Germany/un-RKI-I-060621/2021 | EPI_ISL_1566126 | 2021-03-15 | SYNLAB MVZ Heidelberg | Robert Koch Institute | unknown |
| hCoV-19/Australia/QLD1913/2021 | EPI_ISL_2507086 | 2021-06-07 | Queensland Health Forensic and Scientific Services | Queensland Health Forensic and Scientific Services | Son Nguyen |
| hCoV-19/Australia/QLD1922/2021 | EPI_ISL_2507090 | 2021-06-10 | Queensland Health Forensic and Scientific Services | Queensland Health Forensic and Scientific Services | Son Nguyen |
| hCoV-19/Australia/QLD1928/2021 | EPI_ISL_2507094 | 2021-06-11 | Queensland Health Forensic and Scientific Services | Queensland Health Forensic and Scientific Services | Son Nguyen |
| hCoV-19/USA/SC-DHEC-1826/2020 | EPI_ISL_2508437 | 2020-05-07 | Microbiology Division, SC DHEC | Microbiology Division, SC DHEC | Freeman,J., Flores,H. |
| hCoV-19/Singapore/1457/2021 | EPI_ISL_2509110 | 2021-05-31 | National Public Health Laboratory, National Centre for Infectious Diseases | National Centre for Infectious Diseases | Tze Minn Mak, Zhenyang Zhou, Royce Ang, Lin Cui, Raymond Tzer Pin Lin |
| hCoV-19/England/HSLL-1642230/2021 | EPI_ISL_2512134 | 2021-06-05 | Health Services Laboratories | Wellcome Sanger Institute for the COVID-19 Genomics UK (COG-UK) Consortium | Health Services Laboratories and Alex Alderton, Roberto Amato, Jeffrey Barrett, Sonia Goncalves, Ewan Harrison, David K. Jackson, Ian Johnston, Dominic Kwiatkowski, Cordelia Langford, John Sillitoe on behalf of the Wellcome Sanger Institute COVID-19 Surve |
| hCoV-19/England/MILK-1630B85/2021 | EPI_ISL_2512401 | 2021-06-04 | Lighthouse Lab in Milton Keynes | Wellcome Sanger Institute for the COVID-19 Genomics UK (COG-UK) Consortium | The Lighthouse Lab in Milton Keynes and Alex Alderton, Roberto Amato, Jeffrey Barrett, Sonia Goncalves, Ewan Harrison, David K. Jackson, Ian Johnston, Dominic Kwiatkowski, Cordelia Langford, John Sillitoe on behalf of the Wellcome Sanger Institute COVID-1 |
| hCoV-19/Germany/NW-RKI-I-075415/2021 | EPI_ISL_1641213 | 2021-04-07 | MVZ Labor Krone GbR | Robert Koch Institute | unknown |
| hCoV-19/Germany/BY-RKI-I-083430/2021 | EPI_ISL_1720919 | 2021-04-01 | MVZ Labor Passau GbR | Robert Koch Institute | unknown |
| hCoV-19/Germany/un-RKI-I-128704/2021 | EPI_ISL_2112265 | 2021-04-27 | SYNLAB MVZ Leinfelden-Echterdingen | Robert Koch Institute | unknown |
| hCoV-19/Germany/BE-RKI-I-129573/2021 | EPI_ISL_2112773 | 2021-05-03 | MDI Limbach Berlin GmbH; MVZ Labor Berlin | Robert Koch Institute | unknown |
| hCoV-19/Germany/BE-RKI-I-137079/2021 | EPI_ISL_2120490 | 2021-05-04 | MDI Limbach Berlin GmbH; MVZ Labor Berlin | Robert Koch Institute | unknown |
| hCoV-19/Germany/SN-RKI-I-141289/2021 | EPI_ISL_2125024 | 2021-04-29 | Fraunhofer-Institut fur Zelltherapie und Immunologie IZI AG Next-Generation Diagnostics | Robert Koch Institute | unknown |
| hCoV-19/Germany/NW-RKI-I-156361/2021 | EPI_ISL_2266815 | 2021-05-18 | SYNLAB MVZ Leverkusen | Robert Koch Institute | unknown |
| hCoV-19/Germany/BE-RKI-I-158147/2021 | EPI_ISL_2313343 | 2021-05-21 | Robert Koch-Institut ZBS1 | Robert Koch Institute | Annika Brinkmann |

| | | | | | |
|---|---|---|---|---|---|
| hCoV-19/Germany/NW-RKI-I-162518/2021 | EPI_ISL_2386460 | 2021-05-19 | SYNLAB MVZ Leverkusen | Robert Koch Institute | unknown |
| hCoV-19/England/LOND-1350EC5/2021 | EPI_ISL_2517351 | 2021-06-06 | University College London, Great Ormond Street Hospital for Children NHS Foundation Trust, Imperial College Healthcare NHS Trust | COVID-19 Genomics UK (COG-UK) Consortium | Sergi Castellano, Rachel Williams, Mark Kristiansen, Paola Resende Silva, Sunando Roy, Tony Brooks, Helena Tutill, Paola Niola, Patricia Dyal, Charlotte Williams, Leysa Forrest, Yasmin Panchbhaya, Jacqueline Findlay, Samuel Weeks, Julianne Brown, Kathryn |
| hCoV-19/Germany/NW-RKI-I-165734/2021 | EPI_ISL_2389112 | 2021-05-18 | Medizinische Laboratorien Dvºsseldorf | Robert Koch Institute | unknown |
| hCoV-19/Germany/NW-RKI-I-165739/2021 | EPI_ISL_2389117 | 2021-05-18 | Medizinische Laboratorien Dvºsseldorf | Robert Koch Institute | unknown |
| hCoV-19/England/NORT-1BCDECB/2021 | EPI_ISL_2518872 | 2021-05-30 | Northumbria University / South Tees Hospitals NHS Foundation Trust / North Cumbria Integrated Care NHS Foundation Trust / North Tees and Hartlepool NHS Foundation Trust / Newcastle Hospitals NHS Foundation Trust | COVID-19 Genomics UK (COG-UK) Consortium | Darren L Smith,Andrew Nelson,Matthew Bashton,Greg R Young,Joshua Loh,John Allan,Mohammad A Tariq,Giles S Holt,Gary Black,Wen C Yew,Lynn Dover,Paul Baker,Steve Liggett,Sarah Essex,Jane Greenaway,Debra Padgett,Clive Graham,Garren Scott,Edward Barton,Emma Sw |
| hCoV-19/Germany/HH-RKI-I-171229/2021 | EPI_ISL_2471055 | 2021-05-26 | Labor Dr. Heidrich & Kollegen MVZ GmbH Hamburg | Robert Koch Institute | unknown |
| hCoV-19/Australia/SA0646/2021 | EPI_ISL_1701832 | 2021-03-14 | SA Pathology | SA Pathology | Lex Leong, Julien Soubrier, Chuan Kok Lim, Song Gao, Mark Turra, Karin Kassahn, Ivan Bastian, Geoff Higgins |
| hCoV-19/Germany/HE-RKI-I-177040/2021 | EPI_ISL_2633569 | 2021-06-05 | CENTOGENE Frankfurt Laboratory: Niederlassung Industriepark Hv∂chst | Robert Koch Institute | unknown |
| hCoV-19/Canada/BC-BCCDC-38684/2021 | EPI_ISL_2524412 | 2021-01-19 | BCCDC Public Health Laboratory | BCCDC Public Health Laboratory | Prystajecky Natalie, Linda Hoang, Dan Fornika, John Tyson, Shannon Russell, Kim Macdonald, Kimia Kamelian, Ana Pacagnella, Corrinne Ng, Loretta Janz, Robert Azana, Mel Krajden |
| hCoV-19/USA/CT-CDC-QDX23678350/2021 | EPI_ISL_1702662 | 2021-04-05 | Quest Diagnostics Incorporated | Centers for Disease Control and Prevention Division of Viral Diseases, Pathogen Discovery | Dakota Howard, Dhwani Batra, Peter W. Cook, Kara Moser, Adrian Paskey, Jason Caravas, Benjamin Rambo-Martin, Shatavia Morrison, Christopher Gulvick, Scott Sammons, Yvette Unoarumhi, Darlene Wagner, Matthew Schmerer, S. H. Rosenthal, A. Gerasimova, R. M. K |
| hCoV-19/Germany/SN-RKI-I-179830/2021 | EPI_ISL_2636270 | 2021-05-15 | Diagnosticum - Labor Neukirchen | Robert Koch Institute | unknown |
| hCoV-19/India/GJ-ICMR-NIV-INSACOG-GSEQ-1167/2020 | EPI_ISL_1703896 | 2020-11-27 | ICMR-National Institute of Virology - INSACOG | NIV Influenza | Potdar, Varsha on behalf of National Influenza Centre, NIV, Pune |
| hCoV-19/Germany/HE-RKI-I-183131/2021 | EPI_ISL_2761815 | 2021-06-16 | CENTOGENE Frankfurt Laboratory: Niederlassung Industriepark Hv∂chst | Robert Koch Institute | unknown |
| hCoV-19/Germany/NW-RKI-I-183678/2021 | EPI_ISL_2762119 | 2021-06-17 | MVZ Labor Krone GbR | Robert Koch Institute | unknown |
| hCoV-19/Australia/SA0637/2021 | EPI_ISL_1704674 | 2021-03-11 | SA Pathology | SA Pathology | Lex Leong, Julien Soubrier, Chuan Kok Lim, Song Gao, Mark Turra, Karin Kassahn, Ivan Bastian, Geoff Higgins |
| hCoV-19/Australia/SA0638/2021 | EPI_ISL_1704675 | 2021-03-11 | SA Pathology | SA Pathology | Lex Leong, Julien Soubrier, Chuan Kok Lim, Song Gao, Mark Turra, Karin Kassahn, Ivan Bastian, Geoff Higgins |
| hCoV-19/Australia/SA0640/2021 | EPI_ISL_1704786 | 2021-03-13 | SA Pathology | SA Pathology | Lex Leong, Julien Soubrier, Chuan Kok Lim, Song Gao, Mark Turra, Karin Kassahn, Ivan Bastian, Geoff Higgins |

| | | | | | |
|---|---|---|---|---|---|
| hCoV-19/USA/MI-CDC-ASC210106488/2021 | EPI_ISL_2528622 | 2021-06-01 | Aegis Sciences Corporation | Centers for Disease Control and Prevention Division of Viral Diseases, Pathogen Discovery | Dakota Howard, Dhwani Batra, Peter W. Cook, Kara Moser, Adrian Paskey, Jason Caravas, Benjamin Rambo-Martin, Shatavia Morrison, Christopher Gulvick, Scott Sammons, Yvette Unoarumhi, Darlene Wagner, Matthew Schmerer, Cyndi Clark, Patrick Campbell, Rob Case |
| hCoV-19/NewZealand/21CV0012/2021 | EPI_ISL_794617 | 2021-01-01 | LabPLUS | Institute of Environmental Science and Research (ESR) | Xiaoyun Ren, Matt Storey, Nikki Freed, Muhammad Faisal, Jing Wang, Hermes Perez, Anja Werno, Antje van der Linden, Arlo Upton, Chris Mansell, David Hammer, Dragana Drinkovic, Gary McAuliffe, Hana Sofia Andersson, James Ussher, Jill Sherwood, Josh Freeman, |
| hCoV-19/Australia/SA0684/2021 | EPI_ISL_1706366 | 2021-04-20 | SA Pathology | SA Pathology | Lex Leong, Chuan Kok Lim, Mark Turra, Ivan Bastian, Geoff Higgins |
| hCoV-19/USA/OR-OHSU-5897/2020 | EPI_ISL_832803 | 2020-11-25 | OHSU Lab Services Molecular Microbiology Lab | Oregon SARS-CoV-2 Genome Sequencing Center | Brendan L. O'Connell, Ruth V. Nichols, Sally Grindstaff, Alec J. Hirsch, Donna Hansel, Guang Fan, Daniel N. Streblow, William B. Messer, Andrew C. Adey, Benjamin N. Bimber, Brian J. O‚ÄôRoak |
| hCoV-19/Canada/BC-BCCDC-55917/2021 | EPI_ISL_2530933 | 2021-04-06 | BCCDC Public Health Laboratory | BCCDC Public Health Laboratory | Prystajecky Natalie, Linda Hoang, Dan Fornika, John Tyson, Shannon Russell, Kim Macdonald, Kimia Kamelian, Ana Pacagnella, Corrinne Ng, Loretta Janz, Robert Azana, Mel Krajden |
| hCoV-19/Canada/BC-BCCDC-55987/2021 | EPI_ISL_2531060 | 2021-04-12 | BCCDC Public Health Laboratory | BCCDC Public Health Laboratory | Prystajecky Natalie, Linda Hoang, Dan Fornika, John Tyson, Shannon Russell, Kim Macdonald, Kimia Kamelian, Ana Pacagnella, Corrinne Ng, Loretta Janz, Robert Azana, Mel Krajden |
| hCoV-19/Norway/14203/2021 | EPI_ISL_2673737 | 2021-06-10 | Furst Medical Laboratory | Norwegian Institute of Public Health, Department of Virology | Kathrine Stene-Johansen, Kamilla Heddeland Instefjord, Hilde Elshaug, Garcia Llorente Ignacio, Jon Br√•te, Engebretsen Serina Beate,Pedersen Benedikte Nevjen, Line Victoria Moen, Debech Nadia, Atiya R Ali,Marie Paulsen Madsen, Rasmus Riis Kopperud, Hilde |
| hCoV-19/Aruba/AW-RIVM-16039/2021 | EPI_ISL_1232714 | 2021-02-25 | Dutch COVID-19 response team | National Institute for Public Health and the Environment (RIVM) | Adam Meijer, Harry Vennema, Dirk Eggink, Jeroen Cremer, Sharon van den Brink, Bas van der Veer, AnneMarie van den Brandt, Florian Zwagemaker, Dennis Schmitz, Chantal Reusken, on behalf of the national COVID-19 response team |
| hCoV-19/Aruba/AW-RIVM-16063/2021 | EPI_ISL_1232738 | 2021-02-26 | Dutch COVID-19 response team | National Institute for Public Health and the Environment (RIVM) | Adam Meijer, Harry Vennema, Dirk Eggink, Jeroen Cremer, Sharon van den Brink, Bas van der Veer, AnneMarie van den Brandt, Florian Zwagemaker, Dennis Schmitz, Chantal Reusken, on behalf of the national COVID-19 response team |
| hCoV-19/India/KA-040424_2_S42_R1_001/2020 | EPI_ISL_1708354 | 2020-09-21 | Thindlu PHC | inStem NCBS - INSACOG | Uma Ramakrishnan Dasaradhi Palakodeti Aswin SaiNarain |
| hCoV-19/Ireland/D-NVRL-e22IRL89411/2021 | EPI_ISL_2533476 | 2021-05-10 | National Virus Reference Laboratory | National Virus Reference Laboratory | Zoe Yandle, Charlene Bennett, Gabriel Gonzalez, Michael Carr, Jonathan Dean, Cillian F De Gascun |

| | | | | | |
|---|---|---|---|---|---|
| hCoV-19/Qatar/QA-QU_12-11-A6/2020 | EPI_ISL_1713243 | 2020-11-26 | Ministry of Public Health / Hamad Medical Corporation | Biomedical Research Center (BRC), Qatar University / Qatar Genome Project (QGP) | BRC: Fatiha M. Benslimane, Heba A. Al-Khatib, Oal Al-Jamal, Dana Al-Batesh, Hadi M. Yassine, Asmaa A. Al-Thani. MOPH and HMC: Abdullatif Al-Khal, Muna A. S. Al-Maslamani, Mashael A. Al-Bader, Hamda Alromaihi, Roberto Bertollini, Peter V. Coyle, Einas A. |
| hCoV-19/India/WB-NIBMG-1931700254822/2021 | EPI_ISL_2676011 | 2021-04-30 | INSACOG-WB | National Institute of Biomedical Genomics ‚Äì INSACOG | Arindam Maitra, Bhaswati Bandyopadhyay, Nidhan Kumar Biswas, Tamal Ghosh, Sreedhar Chinnaswamy, Ajay Chakraborti, Saumitra Das |
| hCoV-19/Qatar/QA-QU_18-5-3-B7/2021 | EPI_ISL_1713719 | 2021-02-27 | Ministry of Public Health / Hamad Medical Corporation | Biomedical Research Center (BRC), Qatar University / Qatar Genome Project (QGP) | BRC: Fatiha M. Benslimane, Heba A. Al-Khatib, Oal Al-Jamal, Dana Al-Batesh, Hadi M. Yassine, Asmaa A. Al-Thani. MOPH and HMC: Abdullatif Al-Khal, Muna A. S. Al-Maslamani, Mashael A. Al-Bader, Hamda Alromaihi, Roberto Bertollini, Peter V. Coyle, Einas A. |
| hCoV-19/India/WB-NIBMG-1931300379204/2021 | EPI_ISL_2676243 | 2021-06-09 | INSACOG-WB | National Institute of Biomedical Genomics ‚Äì INSACOG | Arindam Maitra, Bhaswati Bandyopadhyay, Nidhan Kumar Biswas, Tamal Ghosh, Sreedhar Chinnaswamy, Ajay Chakraborti, Saumitra Das |
| hCoV-19/Qatar/QA-WCMQ_FD18163103/2020 | EPI_ISL_1714373 | 2020-08-20 | Ministry of Public Health / Hamad Medical Corporation | Weill Cornell Medical College - Qatar (WCM-Q), Genomics Core Laboratory / Qatar Genome Project (QGP) | WCMQ: Ayeda A. Ahmed, Meryem Bensaad, Shameem Younuskunju, Yasmin Mohamoud, Laith Abu-Raddad, Joel A Malek. QGP: Fatima H. Al-Kuwari, Chadi Saad MOPH and HMC: Abdullatif Al-Khal, Muna A. S. Al-Maslamani, Mashael A. Al-Bader, Hamda Alromaihi, Roberto Ber |
| hCoV-19/Qatar/QA-WCMQ_FD18163135/2020 | EPI_ISL_1714404 | 2020-08-14 | Ministry of Public Health / Hamad Medical Corporation | Weill Cornell Medical College - Qatar (WCM-Q), Genomics Core Laboratory / Qatar Genome Project (QGP) | WCMQ: Ayeda A. Ahmed, Meryem Bensaad, Shameem Younuskunju, Yasmin Mohamoud, Laith Abu-Raddad, Joel A Malek. QGP: Fatima H. Al-Kuwari, Chadi Saad MOPH and HMC: Abdullatif Al-Khal, Muna A. S. Al-Maslamani, Mashael A. Al-Bader, Hamda Alromaihi, Roberto Ber |
| hCoV-19/Portugal/PT8354/2021 | EPI_ISL_2445928 | 2021-05-20 | H Santarem | Instituto Nacional de Saude (INSA) | Borges et al |
| hCoV-19/Nigeria/CV1066/2020 | EPI_ISL_1715414 | 2020-12-29 | Nigerian Centre for Disease Control (NCDC) | Genomics of Infectious Diseases, Redeemer‚Äôs University | Olawoye, I.B. et al |
| hCoV-19/Chile/RM-98045/2021 | EPI_ISL_2543479 | 2021-05-22 | Genetica Molecular and Subdepartamento de Virologia ISP Chile | Instituto de Salud Publica de Chile | Karen Orostica, Constanza Campano, Barbara Parra, Loredana Arata, Gisselle Barra, Patricia Bustos, Rodrigo Fasce, Javier Tognarelli, Andres Castillo, Soledad Ulloa, Jorge Fernandez |
| hCoV-19/Poland/PZH-LUM-1362/2021 | EPI_ISL_2543732 | 2021-05-29 | UM im. Karola Marcinkowskiego w Poznaniu Laboratorium UCA_Covid-19 | 1. National Institute of Public Health National Institute of Hygiene, Warsaw, Poland 2. Biobank Lab, University of Lodz 3. Laboratory of Respiratory Viruses, Teaching and Clinical Center of the Medical University of Lodz | Dominik Strapagiel, Marta Sobalska-Kwapis, Klaudyna Kr√≥likowska, Jakub Lach, Marcin S≈Çomka, Tomasz P≈Çoszaj, Magdalena Traczyk-Borszy≈Ñska, Izabela Dr√≥≈ºd≈º, Maciej Borowiec, Katarzyna Zacharczuk, Magdalena Nowakowska, Ma≈Çgorzata Sadkowska-Todys, Toma |
| hCoV-19/Barbados/71164/2021 | EPI_ISL_2678164 | 2021-05-24 | The Caribbean Public Health Agency | Carrington Lab, Department of Preclinical Sciences, Faculty of Medical Sciences, The University of the West Indies, St Augustine Campus | Nikita S. D. Sahadeo, Arianne Brown-Jordan, Sarah Hill, Vernie Ramkissoon, Anushka Ramjag, Kenneth George, Avery Hinds, Risha Singh, SueMin Nathaniel, Nuno Faria, Oliver Pybus, Gabriel Escobar, Christine V. F. Carrington |
| hCoV-19/Germany/BY-RKI-I-181236/2021 | EPI_ISL_2678385 | 2021-05-10 | Virologisches Institut des Universit√§tsklinikums Erlangen | Robert Koch Institute | unknown |

| | | | | | |
|---|---|---|---|---|---|
| hCoV-19/USA/IN-INSPHL-02055/2021 | EPI_ISL_2544594 | 2021-05-29 | IN State Department of Health Laboratory Services | IN State Department of Health Laboratory Services | Cassandra Campion, Jamie Yeadon, Brian Pope, Lixia Liu, Kyle Brownlee, Melissa Hindenlang, Mark Glazier |
| hCoV-19/Germany/BE-RKI-I-181554/2021 | EPI_ISL_2678662 | 2021-06-16 | MDI Limbach Berlin GmbH; MVZ Labor Berlin | Robert Koch Institute | unknown |
| hCoV-19/Canada/AB-ABPHL-04590/2021 | EPI_ISL_2164355 | 2021-01-15 | Alberta Precision Labs (APL) | Public Health Agency of Canada (PHAC) National Microbiology Laboratory | Buss, E, Croxen M, Deo A, Dieu P, Gill K, Ferrato C, Khan F, Koleva P, Li V, Lloyd C, Lynch T, Ma R, Murphy S, Pabbaraju K, Shokoples S, Tipples G, Thayer J, Whitehouse M, Wong A, Yu C, Zelyas N |
| hCoV-19/England/MILK-1512D84/2021 | EPI_ISL_1719097 | 2021-04-17 | Lighthouse Lab in Milton Keynes | Wellcome Sanger Institute for the COVID-19 Genomics UK (COG-UK) Consortium | The Lighthouse Lab in Milton Keynes and Alex Alderton, Roberto Amato, Jeffrey Barrett, Sonia Goncalves, Ewan Harrison, David K. Jackson, Ian Johnston, Dominic Kwiatkowski, Cordelia Langford, John Sillitoe on behalf of the Wellcome Sanger Institute COVID-1 |
| hCoV-19/India/MH-IGIB-NIV-INSACOG-GSEQ-1455/2021 | EPI_ISL_2546078 | 2021-03-19 | ICMR-National Institute of Virology - INSACOG | NIV Influenza | Potdar, Varsha on behalf of National Influenza Centre, NIV, Pune |
| hCoV-19/India/MH-IGIB-NIV-INSACOG-GSEQ-1857/2021 | EPI_ISL_2546308 | 2021-03-27 | ICMR-National Institute of Virology - INSACOG | NIV Influenza | Potdar, Varsha on behalf of National Influenza Centre, NIV, Pune |
| hCoV-19/India/MH-IGIB-NIV-INSACOG-GSEQ-1982/2021 | EPI_ISL_2546364 | 2021-03-30 | ICMR-National Institute of Virology - INSACOG | NIV Influenza | Potdar, Varsha on behalf of National Influenza Centre, NIV, Pune |
| hCoV-19/India/MH-IGIB-NIV-INSACOG-GSEQ-2137/2021 | EPI_ISL_2546422 | 2021-03-22 | ICMR-National Institute of Virology - INSACOG | NIV Influenza | Potdar, Varsha on behalf of National Influenza Centre, NIV, Pune |
| hCoV-19/India/GJ-ICMR-NIV-INSACOG-GSEQ-3616/2021 | EPI_ISL_2546755 | 2021-05-12 | ICMR-National Institute of Virology - INSACOG | NIV Influenza | Potdar, Varsha on behalf of National Influenza Centre, NIV, Pune |
| hCoV-19/India/ML-1727400000034/2021 | EPI_ISL_2547441 | 2021-04-06 | INSACOG-MEGHALAYA | National Institute of Biomedical Genomics ‚Äì INSACOG | Arindam Maitra, R. Lyngdoh, Nidhan Kumar Biswas, E Shadap, Sreedhar Chinnaswamy,¬†E Marak, Saumitra Das |
| hCoV-19/Australia/VIC1946/2020 | EPI_ISL_480671 | 2020-06-09 | Victorian Infectious Diseases Reference Laboratory (VIDRL) | VIDRL and MDU-PHL | Caly L., Seemann T., Sait, M., Schultz M., Druce J., Sherry, N. |
| hCoV-19/India/WB-1930300609940/2021 | EPI_ISL_2548646 | 2021-03-27 | INSACOG-WB | National Institute of Biomedical Genomics ‚Äì INSACOG | Arindam Maitra, Bhaswati Bandyopadhyay, Nidhan Kumar Biswas, Tamal Ghosh, Sreedhar Chinnaswamy, Ajay Chakraborti, Saumitra Das |
| hCoV-19/Argentina/PAIS-H0069/2020 | EPI_ISL_1396484 | 2020-10-16 | Centro de Tecnolog√≠a en Salud P√∫blica de la Universidad Nacional de Rosario | Laboratorio Mixto de Biotecnolog√≠a Acu√°tica (LMBA) on behalf of 'Proyecto Argentino Interinstitucional de genomica de SARS-CoV-2' (PAIS Consortium) | Joaqu√≠n Ezpeleta, Ignacio Garc√≠a Labari, Victoria Posner, Vanina Villanova, Pablo Casal, Pilar Bulacio, Sof√≠a Lavista Llanos, Federico Remes Lenicov, Ana Paletta, Leandro Ciappina, Flavio Spetale, Agustina Cerri, Silvana Spinelli, Elisa Bolatti, Diego |
| hCoV-19/India/WB-1932000254981/2021 | EPI_ISL_2549034 | 2021-04-19 | INSACOG-WB | National Institute of Biomedical Genomics ‚Äì INSACOG | Arindam Maitra, Bhaswati Bandyopadhyay, Nidhan Kumar Biswas, Tamal Ghosh, Sreedhar Chinnaswamy, Ajay Chakraborti, Saumitra Das |
| hCoV-19/England/LOND-1337170/2021 | EPI_ISL_1730346 | 2021-04-18 | University College London, Great Ormond Street Hospital for Children NHS Foundation Trust, Imperial College Healthcare NHS Trust | COVID-19 Genomics UK (COG-UK) Consortium | Sergi Castellano, Rachel Williams, Mark Kristiansen, Paola Resende Silva, Sunando Roy, Tony Brooks, Helena Tutill, Paola Niola, Patricia Dyal, Charlotte Williams, Leysa Forrest, Yasmin Panchbhaya, Jacqueline Findlay, Samuel Weeks, Julianne Brown, Kathryn |

| | | | | | |
|---|---|---|---|---|---|
| hCoV-19/USA/PA-MGEL-00300/2020 | EPI_ISL_1732507 | 2020-05-02 | UPMC Clinical Microbiology Laboratory | Center; Microbial Genomic Epidemiology Laboratory, University of Pittsburgh | Lee H. Harrison, Jane W. Marsh, Marissa P. Griffith, Stephanie L. Mitchell, Vatsala R. Srinivasa, Kady D. Waggle, Daniel J. Snyder, Vaughn S. Cooper |
| hCoV-19/Mauritius/104672/2021 | EPI_ISL_2549199 | 2021-05-08 | Virology Department, Central Health Laboratory | The Francis Crick Institute | Ramuth M, Manraj SS, Sonoo J, Bahadoor BS, Crawford M, Patel H, Harvey R, Goldstone R, Daniels RS, Nicod J |
| hCoV-19/England/MILK-1517790/2021 | EPI_ISL_1741398 | 2021-04-19 | Lighthouse Lab in Milton Keynes | Wellcome Sanger Institute for the COVID-19 Genomics UK (COG-UK) Consortium | The Lighthouse Lab in Milton Keynes and Alex Alderton, Roberto Amato, Jeffrey Barrett, Sonia Goncalves, Ewan Harrison, David K. Jackson, Ian Johnston, Dominic Kwiatkowski, Cordelia Langford, John Sillitoe on behalf of the Wellcome Sanger Institute COVID-1 |
| hCoV-19/Switzerland/VD-ETHZ-580364/2021 | EPI_ISL_1750600 | 2021-04-12 | Viollier AG | Department of Biosystems Science and Engineering, ETH Zvºrich | Christian Beisel, Sarah Nadeau, Chaoran Chen, Ivan Topolsky, Philipp Jablonski, Lara Fuhrmann, David Dreifuss, Katharina Jahn, Rebecca Denes, Mirjam Feldkamp, Ina Nissen, Natascha Santacroce, Elodie Burcklen, Christiane Beckmann, Maurice Redondo, Olivier |
| hCoV-19/Germany/BY-MVP-000002683/2020 | EPI_ISL_1751401 | 2020-12-24 | Max von Pettenkofer Institute, Virology, National Reference Center for Retroviruses, LMU Munich | Laboratory for Functional Genome Analysis; Dept. Genomics; Gene Center of the LMU Munich | Max Muenchhoff; Stefan Krebs; Alexander Graf; Oliver Keppler; Helmut Blum |
| hCoV-19/Germany/BY-MVP-000002576/2021 | EPI_ISL_1752261 | 2021-01-26 | Max von Pettenkofer Institute, Virology, National Reference Center for Retroviruses, LMU Munich | Laboratory for Functional Genome Analysis; Dept. Genomics; Gene Center of the LMU Munich | Max Muenchhoff; Stefan Krebs; Alexander Graf; Oliver Keppler; Helmut Blum |
| hCoV-19/Australia/NSW-R0222/2021 | EPI_ISL_1756022 | 2021-04-23 | New South Wales Health Pathology Royal Prince Alfred Hospital | Microbiology RPAH | Foster, C.; Au, J.; Ruiz Silva, M.; Deveson, I.; Bull, R.; Van Hal, S.; Rawlinson, W. |
| hCoV-19/Israel/CVL-4282/2021 | EPI_ISL_1762147 | 2021-01-27 | Israel Central Virology laboratory | Israel National Consortium for SARS-CoV-2 sequencing | Neta Zuckerman, Efrat Dahan Bucris, Michal Mandelboim, Dana Bar-Ilan, Oran Erster, Tzvia Mann, Omer Murik, David A. Zeevi, Assaf Rokney, Joseph Jaffe, Eva Nachum, Maya Davidovich Cohen, Ephraim Fass, Gal Zizelski Valenci, Mor Rubinstein, Efrat Rorman, Isr |
| hCoV-19/Israel/CVL-6303/2021 | EPI_ISL_1762343 | 2021-01-07 | Israel Central Virology laboratory | Israel National Consortium for SARS-CoV-2 sequencing | Neta Zuckerman, Efrat Dahan Bucris, Michal Mandelboim, Dana Bar-Ilan, Oran Erster, Tzvia Mann, Omer Murik, David A. Zeevi, Assaf Rokney, Joseph Jaffe, Eva Nachum, Maya Davidovich Cohen, Ephraim Fass, Gal Zizelski Valenci, Mor Rubinstein, Efrat Rorman, Isr |
| hCoV-19/Israel/CVL-6374/2021 | EPI_ISL_1762374 | 2021-01-11 | Israel Central Virology laboratory | Israel National Consortium for SARS-CoV-2 sequencing | Neta Zuckerman, Efrat Dahan Bucris, Michal Mandelboim, Dana Bar-Ilan, Oran Erster, Tzvia Mann, Omer Murik, David A. Zeevi, Assaf Rokney, Joseph Jaffe, Eva Nachum, Maya Davidovich Cohen, Ephraim Fass, Gal Zizelski Valenci, Mor Rubinstein, Efrat Rorman, Isr |
| hCoV-19/Israel/CVL-2862/2021 | EPI_ISL_1762558 | 2021-01-11 | Israel Central Virology laboratory | Israel National Consortium for SARS-CoV-2 sequencing | Neta Zuckerman, Efrat Dahan Bucris, Michal Mandelboim, Dana Bar-Ilan, Oran Erster, Tzvia Mann, Omer Murik, David A. Zeevi, Assaf Rokney, Joseph Jaffe, Eva Nachum, Maya Davidovich Cohen, Ephraim Fass, Gal Zizelski Valenci, Mor Rubinstein, Efrat Rorman, Isr |

| | | | | | |
|---|---|---|---|---|---|
| hCoV-19/Israel/CVL-6590/2021 | EPI_ISL_1763296 | 2021-02-28 | Israel Central Virology laboratory | Israel National Consortium for SARS-CoV-2 sequencing | Neta Zuckerman, Efrat Dahan Bucris, Michal Mandelboim, Dana Bar-Ilan, Oran Erster, Tzvia Mann, Omer Murik, David A. Zeevi, Assaf Rokney, Joseph Jaffe, Eva Nachum, Maya Davidovich Cohen, Ephraim Fass, Gal Zizelski Valenci, Mor Rubinstein, Efrat Rorman, Isr |
| hCoV-19/Israel/CVL-7070/2021 | EPI_ISL_1763575 | 2021-02-22 | Israel Central Virology laboratory | Israel National Consortium for SARS-CoV-2 sequencing | Neta Zuckerman, Efrat Dahan Bucris, Michal Mandelboim, Dana Bar-Ilan, Oran Erster, Tzvia Mann, Omer Murik, David A. Zeevi, Assaf Rokney, Joseph Jaffe, Eva Nachum, Maya Davidovich Cohen, Ephraim Fass, Gal Zizelski Valenci, Mor Rubinstein, Efrat Rorman, Isr |
| hCoV-19/Israel/CVL-7079/2021 | EPI_ISL_1763583 | 2021-02-22 | Israel Central Virology laboratory | Israel National Consortium for SARS-CoV-2 sequencing | Neta Zuckerman, Efrat Dahan Bucris, Michal Mandelboim, Dana Bar-Ilan, Oran Erster, Tzvia Mann, Omer Murik, David A. Zeevi, Assaf Rokney, Joseph Jaffe, Eva Nachum, Maya Davidovich Cohen, Ephraim Fass, Gal Zizelski Valenci, Mor Rubinstein, Efrat Rorman, Isr |
| hCoV-19/Israel/CVL-7370/2021 | EPI_ISL_1763775 | 2021-02-23 | Israel Central Virology laboratory | Israel National Consortium for SARS-CoV-2 sequencing | Neta Zuckerman, Efrat Dahan Bucris, Michal Mandelboim, Dana Bar-Ilan, Oran Erster, Tzvia Mann, Omer Murik, David A. Zeevi, Assaf Rokney, Joseph Jaffe, Eva Nachum, Maya Davidovich Cohen, Ephraim Fass, Gal Zizelski Valenci, Mor Rubinstein, Efrat Rorman, Isr |
| hCoV-19/England/MILK-1523236/2021 | EPI_ISL_1790678 | 2021-04-20 | Lighthouse Lab in Milton Keynes | Wellcome Sanger Institute for the COVID-19 Genomics UK (COG-UK) Consortium | The Lighthouse Lab in Milton Keynes and Alex Alderton, Roberto Amato, Jeffrey Barrett, Sonia Goncalves, Ewan Harrison, David K. Jackson, Ian Johnston, Dominic Kwiatkowski, Cordelia Langford, John Sillitoe on behalf of the Wellcome Sanger Institute COVID-1 |
| hCoV-19/England/MILK-152460C/2021 | EPI_ISL_1790954 | 2021-04-20 | Lighthouse Lab in Milton Keynes | Wellcome Sanger Institute for the COVID-19 Genomics UK (COG-UK) Consortium | The Lighthouse Lab in Milton Keynes and Alex Alderton, Roberto Amato, Jeffrey Barrett, Sonia Goncalves, Ewan Harrison, David K. Jackson, Ian Johnston, Dominic Kwiatkowski, Cordelia Langford, John Sillitoe on behalf of the Wellcome Sanger Institute COVID-1 |
| hCoV-19/Netherlands/NB-RIVM-27390/2021 | EPI_ISL_1792907 | 2021-04-15 | Dutch COVID-19 response team | National Institute for Public Health and the Environment (RIVM) | Adam Meijer, Harry Vennema, Dirk Eggink, Jeroen Cremer, Sharon van den Brink, Bas van der Veer, AnneMarie van den Brandt, Lisa Wijsman, Kim Freriks, Ryanne Jaarsma, Eun√≠ce Then, Lynn Aarts, Sanne Bos, Melissa van Tuil, Robert Kohl, Linda van de Nes, Sjoe |
| hCoV-19/Brazil/SP-844329/2021 | EPI_ISL_1795072 | 2021-03-21 | LABORATORIO DR PAULO EMILIO DALESSANDRO PINDAMONHANGABA | Instituto Butantan / ESALQ-Piracicaba | Instituto Butantan: Alexander Roberto Precioso, Dimas Tadeu Covas, Sandra Coccuzzo Sampaio, Maria Carolina Elias, Jos√© Salvatore Leister Patan√©, Vincent Louis Viala, Antonio Jorge Martins, Ricardo Haddad, Claudia Renata dos Santos Barros, Elaine Cristin |
| hCoV-19/USA/MI-MDHHS-SC25921/2021 | EPI_ISL_1527956 | 2021-03-23 | Michigan Department of Health and Human Services, Bureau of Laboratories | Human Services, Bureau of Laboratories | Blankenship HM, Riner D, Soehnlen MK |

| | | | | | |
|---|---|---|---|---|---|
| hCoV-19/England/MILK-1528538/2021 | EPI_ISL_1806433 | 2021-04-21 | Lighthouse Lab in Milton Keynes | Wellcome Sanger Institute for the COVID-19 Genomics UK (COG-UK) Consortium | The Lighthouse Lab in Milton Keynes and Alex Alderton, Roberto Amato, Jeffrey Barrett, Sonia Goncalves, Ewan Harrison, David K. Jackson, Ian Johnston, Dominic Kwiatkowski, Cordelia Langford, John Sillitoe on behalf of the Wellcome Sanger Institute COVID-1 |
| hCoV-19/England/ALDP-152CF27/2021 | EPI_ISL_1806561 | 2021-04-22 | Lighthouse Lab in Alderley Park | Wellcome Sanger Institute for the COVID-19 Genomics UK (COG-UK) Consortium | Jacquelyn Wynn, Mairead Hyland, The Lighthouse Lab in Alderley Park and Alex Alderton, Roberto Amato, Jeffrey Barrett, Sonia Goncalves, Ewan Harrison, David K. Jackson, Ian Johnston, Dominic Kwiatkowski, Cordelia Langford, John Sillitoe on behalf of the W |
| hCoV-19/USA/CA-SEARCH-11906/2021 | EPI_ISL_1814748 | 2021-02-12 | EXCITE Lab | Andersen lab at Scripps Research | Nicole L Washington, Simon White, Geraint Levan, Kelly Schiabor Barrett, Elizabeth Cirulli, Alexandre Bolze, Charlotte Rivera-Garcia, Ryan Cho, Jason Nguyen, Sherry Wang, Jimmy Ramirez, Celena Andrade, Alice Summerfield, Tyler Cassens, Efren Sandoval, Fra |
| hCoV-19/Singapore/709/2021 | EPI_ISL_1816953 | 2021-04-25 | National Public Health Laboratory, National Centre for Infectious Diseases | National Centre for Infectious Diseases | Tze Minn Mak, Zhenyang Zhou, Grace Jie Yin Ngan, Royce Ang, Lin Cui, Raymond Tzer Pin Lin |
| hCoV-19/USA/NE-CUMC_21109060/2021 | EPI_ISL_1822557 | 2021-04-15 | Creighton University Medical Center | Creighton COVID Consortium | Holly Stessman, Michael Belshan, Richard Goering, Cynthia Watson, Jake Siedlik |
| hCoV-19/USA/CA-COLOR-UCSF-CC144/2021 | EPI_ISL_1824863 | 2021-02-12 | Color Genomics | Chiu Laboratory, University of California, San Francisco | Charles Chiu, Venice Servellita, Candace Wang, Alicia Sotomayor-Gonzalez, Yueyuan Zhang, Daniel Wong, Scott Topper, Madyson Wanner, Erika Torres, Alicia Zhou |
| hCoV-19/Mauritius/N4638/2021 | EPI_ISL_1827657 | 2021-03-12 | Virology Department, Victoria Hospital, Plaine-Wilhems, Mauritius | National Institute for Communicable Diseases of the National Health Laboratory Service | Ramuth M, Manraj SS, Sonoo J, Baboo SB, Amoako DG, Mohale T, Ntuli N, Mahlangu B, Allam M, Ismail A, Bhiman JN |
| hCoV-19/USA/NY-PRL-2021_0426_00A15/2021 | EPI_ISL_1828260 | 2021-04-23 | Pandemic Response Lab - NYC | Pandemic Response Lab, R&D | Henry Lee, Michael Hammerling, Melissa Hopkins, Cybill del Castillo, Shinyoung Clair Kang, William Ward, Pradeep Bugga, Sol Rey, Dylan Law, Katharine Nelson, Haiping Hao, Jon Laurent |
| hCoV-19/England/MILK-152447B/2021 | EPI_ISL_1831277 | 2021-04-19 | Lighthouse Lab in Milton Keynes | Wellcome Sanger Institute for the COVID-19 Genomics UK (COG-UK) Consortium | The Lighthouse Lab in Milton Keynes and Alex Alderton, Roberto Amato, Jeffrey Barrett, Sonia Goncalves, Ewan Harrison, David K. Jackson, Ian Johnston, Dominic Kwiatkowski, Cordelia Langford, John Sillitoe on behalf of the Wellcome Sanger Institute COVID-1 |
| hCoV-19/England/CAMC-133CE0B/2021 | EPI_ISL_1831538 | 2021-02-26 | Lighthouse Lab in Cambridge | Wellcome Sanger Institute for the COVID-19 Genomics UK (COG-UK) Consortium | Rob Howes, The Lighthouse Lab in Cambridge and Alex Alderton, Roberto Amato, Jeffrey Barrett, Sonia Goncalves, Ewan Harrison, David K. Jackson, Ian Johnston, Dominic Kwiatkowski, Cordelia Langford, John Sillitoe on behalf of the Wellcome Sanger Institute |
| hCoV-19/India/MH-NS1239/2021 | EPI_ISL_1838272 | 2021-04-01 | The National Centre for Cell Science | CSIR-Centre for Cellular and Molecular Biology-INSACOG | Dhiraj Paul, Mohak P Gujare, Shivang P. Bhanushali, Mitali Inamdar, Sonal Manik Chavan, Manoj Kumar Bhat, Ajay Pillai, INSACOG Consortium team, Yogesh Shouche,Payel Mukherjee,Lamuk Zaveri,Tulasi Nagabandi, Ara Sreenivas,Valli Nagalakshmi Undamatla,Shreeka |

| | | | | | |
|---|---|---|---|---|---|
| hCoV-19/India/AP-CCMB-BO889/2021 | EPI_ISL_1838697 | 2021-04-12 | CSIR-Centre for Cellular and Molecular Biology | CSIR-Centre for Cellular and Molecular Biology-INSACOG | Payel Mukherjee,Lamuk Zaveri,Tulasi Nagabandi, Ara Sreenivas,Shreekant Verma, Amareshwar Vodapalli ,Blessy B John,Viswagithe S L,B Himasri,Valli Nagalakshmi Undamatla,Onkar Kulkarni,Sofia Banu,Archana Bharadwaj Siva,Sharath Chandra Thota,Karthik Bharadwaj |
| hCoV-19/Denmark/DCGC-70035/2020 | EPI_ISL_1878606 | 2020-11-23 | Department of Virus and Microbiological Special Diagnostics, Statens Serum Institut, Copenhagen, Denmark | Aalborg University | Danish Covid-19 Genome Consortium |
| hCoV-19/India/GJ-NCDC-NIV-INSACOG-240938/2021 | EPI_ISL_2555672 | 2021-03-31 | ICMR-National Institute of Virology | NCDC Delhi, Biotechnology Division | V.Vipat,Meena Datta,S.Jadhav,Priyanka Singh,U.Saha ,M.Das,Uma Sharma,G.Divekar,T.Sanjeevi,Manoj K Singh,S Jadhav,Radhakrishnan V. S,A Walimbe ,Robin Marwal,V Malik,Mahesh S Dhar,K.Iyengar,Kalaiarasan Ponnusamy,K P.Shinde,Hemlata Lall,K.Korabu,Z. Sayyed,He |
| hCoV-19/India/GJ-NCDC-NIV-INSACOG-240991/2021 | EPI_ISL_2555683 | 2021-03-26 | ICMR-National Institute of Virology | NCDC Delhi, Biotechnology Division | V.Vipat,Meena Datta,S.Jadhav,Priyanka Singh,U.Saha ,M.Das,Uma Sharma,G.Divekar,T.Sanjeevi,Manoj K Singh,S Jadhav,Radhakrishnan V. S,A Walimbe ,Robin Marwal,V Malik,Mahesh S Dhar,K.Iyengar,Kalaiarasan Ponnusamy,K P.Shinde,Hemlata Lall,K.Korabu,Z. Sayyed,He |
| hCoV-19/India/DL-NCDC-2509373/2021 | EPI_ISL_2555807 | 2021-04-23 | National Centre for Disease Control (NCDC) Biotechnology Division, Delhi | NCDC Delhi, Biotechnology Division INSACOG | Radhakrishnan V. S, Robin Marwal, Mahesh S Dhar, Kalaiarasan Ponnusamy, Meena Datta, Priyanka Singh, Uma Sharma, Hemlata Lall, Manoj K Singh, Hema Gogia, Preeti Madan, Sandhya Kabra, Sujeet K Singh, Partha Rakshit |
| hCoV-19/India/UK-NCDC-2609307/2021 | EPI_ISL_2555885 | 2021-04-20 | National Centre for Disease Control (NCDC) Biotechnology Division, Delhi | NCDC Delhi, Biotechnology Division INSACOG | Mahesh S Dhar, Kalaiarasan Ponnusamy, Meena Datta, Priyanka Singh, Uma Sharma, Hemlata Lall, Manoj K Singh, Radhakrishnan V. S, Robin Marwal, Hema Gogia, Preeti Madan, Sandhya Kabra, Sujeet K Singh, Partha Rakshit |
| hCoV-19/India/JK-NCDC-270918/2021 | EPI_ISL_2555909 | 2021-01-29 | National Centre for Disease Control (NCDC) Biotechnology Division, Delhi | NCDC Delhi, Biotechnology Division INSACOG | Mahesh S Dhar, Kalaiarasan Ponnusamy, Meena Datta, Priyanka Singh, Uma Sharma, Hemlata Lall, Manoj K Singh, Radhakrishnan V. S, Robin Marwal, Hema Gogia, Preeti Madan, Sandhya Kabra, Sujeet K Singh, Partha Rakshit |
| hCoV-19/India/JK-NCDC-270976/2021 | EPI_ISL_2555931 | 2021-03-09 | National Centre for Disease Control (NCDC) Biotechnology Division, Delhi | NCDC Delhi, Biotechnology Division INSACOG | Mahesh S Dhar, Kalaiarasan Ponnusamy, Meena Datta, Priyanka Singh, Uma Sharma, Hemlata Lall, Manoj K Singh, Radhakrishnan V. S, Robin Marwal, Hema Gogia, Preeti Madan, Sandhya Kabra, Sujeet K Singh, Partha Rakshit |
| hCoV-19/India/RJ-NCDC-1809178/2020 | EPI_ISL_2556700 | 2020-09-08 | National Centre for Disease Control (NCDC) Biotechnology Division, Delhi | NCDC Delhi, Biotechnology Division INSACOG | Kalaiarasan Ponnusamy, Meena Datta, Priyanka Singh, Uma Sharma, Hemlata Lall, Manoj K Singh, Radhakrishnan V. S, Robin Marwal, Mahesh S Dhar, Hema Gogia, Preeti Madan, Sandhya Kabra, Sujeet K Singh, Partha Rakshit |

| | | | | | |
|---|---|---|---|---|---|
| hCoV-19/India/DL-NCDC-1809194/2021 | EPI_ISL_2556711 | 2021-03-13 | National Centre for Disease Control (NCDC) Biotechnology Division, Delhi | NCDC Delhi, Biotechnology Division INSACOG | Kalaiarasan Ponnusamy, Meena Datta, Priyanka Singh, Uma Sharma, Hemlata Lall, Manoj K Singh, Radhakrishnan V. S, Robin Marwal, Mahesh S Dhar, Hema Gogia, Preeti Madan, Sandhya Kabra, Sujeet K Singh, Partha Rakshit |
| hCoV-19/Finland/THL-202118454/2021 | EPI_ISL_2557202 | 2021-05-19 | Fimlab Laboratoriot Oy Tampere | Expert Microbiology, National Institute for Health and Welfare | Soile Blomqvist, Jani Halkilahti, Kirsi Liitsola, Haider al-Hello, P√§ivi Laurila, Erika Lindh, Teemu Smura, Ravi Kant, Phuoc Truong, Olli Vapalahti, Sari Hannula, Pekka Ellonen, Niina Ikonen, Carita Savolainen-Kopra |
| hCoV-19/Spain/MD-HRYC-11878254/2021 | EPI_ISL_2558018 | 2021-05-26 | Servicio de Microbiologia Hospital Ramon y Cajal | Servicio de Microbiologia Hospital Ramon y Cajal | Ponce M, Galan JC, Martinez L. Abreu M, y Gonzalez-Alba JM |
| hCoV-19/USA/WI-WSLH-215380/2021 | EPI_ISL_2558570 | 2021-05-19 | Wisconsin State Laboratory of Hygiene Communicable Disease Division | Wisconsin State Laboratory of Hygiene Communicable Disease Division | Abigail C. Shockey, Alicia J. Mooney, Erika M. Hanson, Tonya Danz, Richard Griesser, Sara Wagner, Kelsey R. Florek |
| hCoV-19/USA/WI-WSLH-215457/2021 | EPI_ISL_2558644 | 2021-05-17 | Wisconsin State Laboratory of Hygiene Communicable Disease Division | Wisconsin State Laboratory of Hygiene Communicable Disease Division | Abigail C. Shockey, Alicia J. Mooney, Erika M. Hanson, Tonya Danz, Richard Griesser, Sara Wagner, Kelsey R. Florek |
| hCoV-19/USA/WY-WYPHL-20131357/2020 | EPI_ISL_2560439 | 2020-11-11 | Wyoming Public Health Laboratory | Wyoming Public Health Laboratory | Jim Mildenberger, Taylor Fearing, Channing Weber, Ashley Norberg, Chayse Rowley, Marley Goetz, Brian Dominguez, Elliot Thomasson, Sam Britz, Cari Sloma, Robert Petit, and Rob Christensen |
| hCoV-19/USA/NE-CUMC_21130087/2021 | EPI_ISL_2562542 | 2021-04-20 | Creighton University Medical Center | Creighton COVID Consortium | Holly Stessman, Michael Belshan, Richard Goering, Cynthia Watson, Jake Siedlik |
| hCoV-19/USA/ND-NDDH-01991/2021 | EPI_ISL_2565914 | 2021-05-28 | North Dakota Department of Health, Public Health Laboratory | North Dakota Department of Health, Public Health Laboratory | Lisa Wingerter |
| hCoV-19/Israel/SMC-7003221/2021 | EPI_ISL_2567013 | 2021-05-16 | Shamir Medical Center (Asaf Harofe) | Shamir Medical Center (Asaf Harofe) | Nir Rainy,Chen Weiner,Reut Sorek Abramovich,Yevgeni Yegorov,Anna Vishnevsky,Patricia Benveniste-Lekovitz,Abu Hamad Ramzia,Adina Bar Chaim |
| hCoV-19/Scotland/QEUH-170F544/2021 | EPI_ISL_2683368 | 2021-06-16 | Lighthouse Lab in Glasgow | Wellcome Sanger Institute for the COVID-19 Genomics UK (COG-UK) Consortium | Harper VanSteenhouse, Yumi Kasai, David Gray, Carol Clugston, Anna Dominiczak and Alex Alderton, Roberto Amato, Jeffrey Barrett, Sonia Goncalves, Ewan Harrison, David K. Jackson, Ian Johnston, Dominic Kwiatkowski, Cordelia Langford, John Sillitoe on behal |
| hCoV-19/England/AUV2_CP52100184012326/2021 | EPI_ISL_2686123 | 2021-05-30 | Amazon Dx, AUV2 Laboratory | Amazon Dx, AUV2 Laboratory | Luke Meredith |
| hCoV-19/BurkinaFaso/LNRFHV_1344/2020 | EPI_ISL_2686246 | 2020-12-22 | Laboratoire National de R√©f√©rence pour les Fi√®vres H√©morragiques Virales, Centre Muraz | Centre Muraz | Ars√®ne Zongo, Ange Badjo, Sa√Ødou Ouedraogo, Jasmin Schlotterbeck, Essia Belarbi, Grit Schubert, Fabian Leendertz, Soumeya Ouangraoua, Th√©r√®se Kagone |
| hCoV-19/BurkinaFaso/M_LBC_3870/2021 | EPI_ISL_2686356 | 2021-01-20 | Laboratoire de Biologie Clinique, Centre Muraz | Centre Muraz | Ars√®ne Zongo, Ange Badjo, Jasmin Schlotterbeck, Essia Belarbi, Grit Schubert, Fabian Leendertz, Soumeya Ouangraoua, Amariane M. M. Kon√©, Th√©r√®se Kagone |

| | | | | | |
|---|---|---|---|---|---|
| hCoV-19/USA/TX-CDC-FG-037687/2021 | EPI_ISL_2686714 | 2021-06-07 | Fulgent Genetics | Centers for Disease Control and Prevention Division of Viral Diseases, Pathogen Discovery | Dakota Howard, Dhwani Batra, Peter W. Cook, Kara Moser, Adrian Paskey, Jason Caravas, Benjamin Rambo-Martin, Shatavia Morrison, Christopher Gulvick, Scott Sammons, Yvette Unoarumhi, Darlene Wagner, Matthew Schmerer, Harry Gao, Mickey Li, John Gao, Joseph |
| hCoV-19/Belgium/UMONS-P02-04406-01/2021 | EPI_ISL_2687961 | 2021-06-11 | National Platform bis UMONS/Jolimont | National Platform bis UMONS/Jolimont | Fran√ßois Dufrasne, Guillaume Bayon-Vicente, Florian Juszczak, Gautier Detry, Ruddy Wattiez |
| hCoV-19/Uganda/UG514/2021 | EPI_ISL_2690490 | 2021-06-08 | MRC/UVRI & LSHTM Uganda¬†Research Unit, Central Public Health Laboratories, Rakai Health Sciences Program | MRC/UVRI & LSHTM Uganda¬†Research Unit, Central Public Health Laboratories | Matthew Cotten, Dan Lule Bugembe, My V.T. Phan, Pontiano Kaleebu,Isaac Sseeewanyana, Patrick Semanda, Susan Nabadda, Hellen Nansumba |
| hCoV-19/SouthAfrica/KRISP-K016684/2021 | EPI_ISL_2693826 | 2021-05-24 | NHLS_VIRO Eastern Cape | KRISP, KZN Research Innovation and Sequencing Platform | Giandhari Jennifer, Pillay Sureshnee, Yajna Ramphal, Naidoo Yeshnee, Tshabuila Derek,Tegally Houriiyah, San James, Wilkinson Eduan, de Oliveira Tulio |
| hCoV-19/SouthAfrica/NICD-N8704/2021 | EPI_ISL_2694613 | 2021-06-03 | TAMBO MEMORIAL | Diseases of the National Health Laboratory Service | Amoako DG, Everatt J, Scheepers C, Mohale T, Ntuli N, Mahlangu B, Mnguni A, Ismail A, Bhiman JN |
| hCoV-19/SouthAfrica/NICD-N8881/2021 | EPI_ISL_2695518 | 2021-06-03 | TAMBO MEMORIAL | Diseases of the National Health Laboratory Service | Amoako DG, Everatt J, Scheepers C, Mohale T, Ntuli N, Mahlangu B, Mnguni A, Ismail A, Bhiman JN |
| hCoV-19/SouthAfrica/NICD-N8180/2021 | EPI_ISL_2695663 | 2021-03-12 | KIMBERLEY LABORATORY | Diseases of the National Health Laboratory Service | Amoako DG, Everatt J, Scheepers C, Mohale T, Ntuli N, Mahlangu B, Mnguni A, Ismail A, Bhiman JN |
| hCoV-19/SouthAfrica/NICD-N8411/2021 | EPI_ISL_2695756 | 2021-05-03 | ROB FERREIRA LABORATORY | Diseases of the National Health Laboratory Service | Amoako DG, Everatt J, Scheepers C, Mohale T, Ntuli N, Mahlangu B, Mnguni A, Ismail A, Bhiman JN |
| hCoV-19/SouthAfrica/NICD-N8415/2021 | EPI_ISL_2695758 | 2021-05-18 | ROB FERREIRA LABORATORY | Diseases of the National Health Laboratory Service | Amoako DG, Everatt J, Scheepers C, Mohale T, Ntuli N, Mahlangu B, Mnguni A, Ismail A, Bhiman JN |
| hCoV-19/Wales/PHWC-PYG1YE/2021 | EPI_ISL_2707654 | 2021-06-15 | Originating lab: Wales Specialist Virology Centre Sequencing lab: Pathogen Genomics Unit | Public Health Wales Microbiology Cardiff Wales Specialist Virology Centre | Catherine Moore, Johnathan Evans, Laura Gifford, Malorie Perry, Simon Cottrell, Angela Marchbank, Alec Birchley, Alexander Adams, Amy Gaskin, Bree Gatica-Wilcox, Jason Coombes, Joel Southgate, Lauren Gilbert, Lee Graham, Nicole Pacchiarini, Sara Kumziene- |
| hCoV-19/England/SHEF-10DB8D6/2021 | EPI_ISL_2708493 | 2021-06-18 | Virology Department, Sheffield Teaching Hospitals NHS Foundation Trust/Department of Infection, Immunity and Cardiovascular Disease, The Medical School, University of Sheffield | COVID-19 Genomics UK (COG-UK) Consortium | Thushan de Silva, Matthew Parker, Nikki Smith, Adri Angyal, Rebecca Brown, Luke Green, Rachel Tucker, Paul Parsons, Danielle Groves, Katie Johnson, Laura Carrilero, Alex Keeley, Dave Partridge, Matthew Wyles, Benjamin Lindsey, Mehmet Yavuz, Mohammad Raza, |
| hCoV-19/SouthAfrica/NICD-N9070/2021 | EPI_ISL_2709860 | 2021-06-07 | POLOKWANE MANKWENG HOSPITAL | Diseases of the National Health Laboratory Service | Amoako DG, Everatt J, Scheepers C, Mohale T, Ntuli N, Mahlangu B, Mnguni A, Ismail A, Bhiman JN |
| hCoV-19/SouthAfrica/NICD-N8546/2021 | EPI_ISL_2709954 | 2021-05-31 | TAMBO MEMORIAL LABORATORY | Diseases of the National Health Laboratory Service | Amoako DG, Everatt J, Scheepers C, Mohale T, Ntuli N, Mahlangu B, Mnguni A, Ismail A, Bhiman JN |
| hCoV-19/SouthAfrica/NICD-N8551/2021 | EPI_ISL_2709958 | 2021-05-29 | TAMBO MEMORIAL LABORATORY | Diseases of the National Health Laboratory Service | Amoako DG, Everatt J, Scheepers C, Mohale T, Ntuli N, Mahlangu B, Mnguni A, Ismail A, Bhiman JN |
| hCoV-19/SouthAfrica/NICD-N8573/2021 | EPI_ISL_2709975 | 2021-05-28 | TAMBO MEMORIAL LABORATORY | Diseases of the National Health Laboratory Service | Amoako DG, Everatt J, Scheepers C, Mohale T, Ntuli N, Mahlangu B, Mnguni A, Ismail A, Bhiman JN |
| hCoV-19/Singapore/1624/2021 | EPI_ISL_2710085 | 2021-06-20 | National Public Health Laboratory, National Centre for Infectious Diseases | National Centre for Infectious Diseases | Tze Minn Mak, Katherine Ching, Zhenyang Zhou, Royce Ang, Lin Cui, Raymond Tzer Pin Lin |
| hCoV-19/SouthAfrica/KRISP-K018713/2021 | EPI_ISL_2710315 | 2021-06-02 | BARC / Lancet | KRISP, KZN Research Innovation and Sequencing Platform | Sisonke, Giandhari Jennifer, Pillay Sureshnee, Yajna Ramphal, Naidoo Yeshnee, Tshabuila Derek,Tegally Houriiyah, San James, Wilkinson Eduan, de Oliveira Tulio |

| | | | | | |
|---|---|---|---|---|---|
| hCoV-19/Canada/BC-BCCDC-77810/2021 | EPI_ISL_2719897 | 2021-04-28 | BCCDC Public Health Laboratory | BCCDC Public Health Laboratory | Prystajecky Natalie, Linda Hoang, Dan Fornika, John Tyson, Shannon Russell, Kim Macdonald, Kimia Kamelian, Ana Pacagnella, Corrinne Ng, Loretta Janz, Robert Azana, Mel Krajden |
| hCoV-19/Canada/BC-BCCDC-78099/2021 | EPI_ISL_2720128 | 2021-04-28 | BCCDC Public Health Laboratory | BCCDC Public Health Laboratory | Prystajecky Natalie, Linda Hoang, Dan Fornika, John Tyson, Shannon Russell, Kim Macdonald, Kimia Kamelian, Ana Pacagnella, Corrinne Ng, Loretta Janz, Robert Azana, Mel Krajden |
| hCoV-19/India/TN-CL-333_S144_R1_001/2021 | EPI_ISL_2723866 | 2021-04-27 | Dr S raju, Director of public heath and preventive medicine | inStem NCBS ‚Äì INSACOG | Uma Ramakrishnan Dasaradhi Palakodeti Aswin SaiNarain |
| hCoV-19/India/TN-CL-415_S226_R1_001/2021 | EPI_ISL_2723917 | 2021-02-05 | Dr S raju, Director of public heath and preventive medicine | inStem NCBS ‚Äì INSACOG | Uma Ramakrishnan Dasaradhi Palakodeti Aswin SaiNarain |
| hCoV-19/India/TN-CL_811_S75_R1_001/2021 | EPI_ISL_2724083 | 2021-04-30 | Dr S raju, Director of public heath and preventive medicine | inStem NCBS ‚Äì INSACOG | Uma Ramakrishnan Dasaradhi Palakodeti Aswin SaiNarain |
| hCoV-19/India/KA-CBP323_S329_R1_001/2021 | EPI_ISL_2724314 | 2021-05-05 | Bangalore Baptist Hospital | inStem NCBS ‚Äì INSACOG | Uma Ramakrishnan Dasaradhi Palakodeti Aswin SaiNarain |
| hCoV-19/Belgium/rega-9051/2021 | EPI_ISL_2726508 | 2021-05-05 | KU Leuven, Rega Institute, Clinical and Epidemiological Virology | KU Leuven, Rega Institute, Clinical and Epidemiological Virology | Tony Wawina-Bokalanga, Bert Vanmechelen, Joan Marti-Carerras, Piet Maes |
| hCoV-19/SouthAfrica/KRISP-K018544/2021 | EPI_ISL_2727329 | 2021-05-28 | ZARV/NHLS,¬†Department Medical Virology,¬†University of Pretoria | KRISP, KZn Research Innovation and Sequencing Platform | Amy Strydom, Adriano Mendes, Micheala Davids, Sim Mayaphi and Marietjie Venter, Giandhari J, Pillay S, Naidoo Y, Ramphal U, Lessells R, Emmanuel SJ, Tegally H, Wilkinson E, de Oliveira T |
| hCoV-19/SouthAfrica/KRISP-K018565/2021 | EPI_ISL_2727347 | 2021-05-12 | ZARV/NHLS,¬†Department Medical Virology,¬†University of Pretoria | KRISP, KZn Research Innovation and Sequencing Platform | Amy Strydom, Adriano Mendes, Micheala Davids, Sim Mayaphi and Marietjie Venter, Giandhari J, Pillay S, Naidoo Y, Ramphal U, Lessells R, Emmanuel SJ, Tegally H, Wilkinson E, de Oliveira T |
| hCoV-19/SouthAfrica/KRISP-K018572/2021 | EPI_ISL_2727354 | 2021-06-18 | ZARV/NHLS,¬†Department Medical Virology,¬†University of Pretoria | KRISP, KZn Research Innovation and Sequencing Platform | Amy Strydom, Adriano Mendes, Micheala Davids, Sim Mayaphi and Marietjie Venter, Giandhari J, Pillay S, Naidoo Y, Ramphal U, Lessells R, Emmanuel SJ, Tegally H, Wilkinson E, de Oliveira T |
| hCoV-19/SouthAfrica/KRISP-K018584/2021 | EPI_ISL_2727366 | 2021-06-16 | ZARV/NHLS,¬†Department Medical Virology,¬†University of Pretoria | KRISP, KZn Research Innovation and Sequencing Platform | Amy Strydom, Adriano Mendes, Micheala Davids, Sim Mayaphi and Marietjie Venter, Giandhari J, Pillay S, Naidoo Y, Ramphal U, Lessells R, Emmanuel SJ, Tegally H, Wilkinson E, de Oliveira T |
| hCoV-19/SouthAfrica/KRISP-K018606/2021 | EPI_ISL_2727385 | 2021-06-16 | NHLS_VIRO | KRISP, KZn Research Innovation and Sequencing Platform | Giandhari Jennifer, Pillay Sureshnee, Yajna Ramphal, Naidoo Yeshnee, Tshabuila Derek,Tegally Houriiyah, San James, Wilkinson Eduan, de Oliveira Tulio |
| hCoV-19/SouthAfrica/KRISP-K018608/2021 | EPI_ISL_2727387 | 2021-06-16 | NHLS_VIRO | KRISP, KZn Research Innovation and Sequencing Platform | Giandhari Jennifer, Pillay Sureshnee, Yajna Ramphal, Naidoo Yeshnee, Tshabuila Derek,Tegally Houriiyah, San James, Wilkinson Eduan, de Oliveira Tulio |
| hCoV-19/USA/NJ-PHEL-V21000284/2021 | EPI_ISL_2727575 | 2021-06-11 | NJDOH, Public Health and Environmental Laboratories | NJ_PHEL | Lindsey Bodnar, Shiv K. Verma, Jacquelyn Deverell, Ryan Pachucki, Dana Woell, Allison Roder, Byeong Jeong |
| hCoV-19/Australia/QLD1971/2021 | EPI_ISL_2727643 | 2021-06-28 | Queensland Health Forensic and Scientific Services | Queensland Health Forensic and Scientific Services | Son Nguyen |

| | | | | | |
|---|---|---|---|---|---|
| hCoV-19/USA/WI-WSLH-215608/2021 | EPI_ISL_2727785 | 2021-06-02 | Wisconsin State Laboratory of Hygiene Communicable Disease Division | Wisconsin State Laboratory of Hygiene Communicable Disease Division | Abigail C. Shockey, Alicia J. Mooney, Erika M. Hanson, Tonya Danz, Richard Griesser, Sara Wagner, Kelsey R. Florek |
| hCoV-19/USA/WY-WYPHL-20126874/2020 | EPI_ISL_2728094 | 2020-11-09 | Wyoming Public Health Laboratory | Wyoming Public Health Laboratory | Jim Mildenberger, Taylor Fearing, Channing Weber, Ashley Norberg, Chayse Rowley, Marley Goetz, Brian Dominguez, Elliot Thomasson, Sam Britz, Cari Sloma, Robert Petit, and Rob Christensen |
| hCoV-19/India/KA-NIMH-SEQ-1201/2021 | EPI_ISL_2728269 | 2021-06-05 | BBMP Urban | INSACOG-KA, NIMHANS | Chitra Pattabiraman, Anson Kunjumon George,Darshan Sreenivas,Ellango Ramasamy, Sony Sharma, Mahesh Kumar.C.S, Ananthapadmanabha Kotambail, Chetan G K, Gautham Arunachal Udupi, Anita S Desai, V Ravi |
| hCoV-19/India/KA-NIMH-SEQ-1327/2021 | EPI_ISL_2728366 | 2021-04-29 | DSU VIJAYAPURA | INSACOG-KA, NIMHANS | Chitra Pattabiraman, Anson Kunjumon George,Darshan Sreenivas,Ellango Ramasamy, Sony Sharma, Mahesh Kumar.C.S, Ananthapadmanabha Kotambail, Chetan G K, Gautham Arunachal Udupi, Anita S Desai, V Ravi |
| hCoV-19/India/KA-NIMH-SEQ-1493/2021 | EPI_ISL_2728503 | 2021-05-05 | RAILWAY HOSPITAL | INSACOG-KA, NIMHANS | Chitra Pattabiraman, Anson Kunjumon George,Darshan Sreenivas,Ellango Ramasamy, Sony Sharma, Mahesh Kumar.C.S, Ananthapadmanabha Kotambail, Chetan G K, Gautham Arunachal Udupi, Anita S Desai, V Ravi |
| hCoV-19/India/KA-NIMH-SEQ-1522/2021 | EPI_ISL_2728525 | 2021-04-21 | RAILWAY HOSPITAL | INSACOG-KA, NIMHANS | Chitra Pattabiraman, Anson Kunjumon George,Darshan Sreenivas,Ellango Ramasamy, Sony Sharma, Mahesh Kumar.C.S, Ananthapadmanabha Kotambail, Chetan G K, Gautham Arunachal Udupi, Anita S Desai, V Ravi |
| hCoV-19/Brazil/SP-FIOCRUZ-35260/2021 | EPI_ISL_2731624 | 2021-05-17 | Laboratv≥rio de Biotecnologia Aplicada (LBA) - Laboratv≥rio de Biologia Molecular - Hospital das Clv≠nicas, Faculdade de Medicina de Botucatu, Departamento de Bioprocessos e Biotecnologia - Faculdade de Civ™ncias Agronv¥micas, UNESP ‚Äì Botucatu/SP | Laboratory of Respiratory Viruses and Measles, Oswaldo Cruz Institute, FIOCRUZ | Paola Resende, Jayme Augusto de Souza Neto, Rejane Maria Tommasini, Leonardo Nazario de Moraes, Felipe Allan da Silva Costa, Patricia Akemi Assato, Luciana Appolinario, Fernando Motta, Anna Carolina Paixao, Ana Carolina Mendonca, Alice Sampaio Rocha, Tain |
| hCoV-19/England/BRIS-134A8C/2020 | EPI_ISL_2733224 | 2020-10-08 | Respiratory Virus Unit, Microbiology Services Colindale, Public Health England | COVID-19 Genomics UK (COG-UK) Consortium | PHE Covid Sequencing Team |
| hCoV-19/England/PHEC-311124/2021 | EPI_ISL_2742983 | 2021-05-21 | Respiratory Virus Unit, Microbiology Services Colindale, Public Health England | COVID-19 Genomics UK (COG-UK) Consortium | PHE Covid Sequencing Team |
| hCoV-19/Spain/CL-IBV-98029694/2020 | EPI_ISL_2754269 | 2020-05-19 | Universidad de Lev≥n | SeqCOVID-SPAIN consortium/IBV(CSIC) | Ana Carvajal, Vicente Martv≠n, Hv©ctor Argvºello, Juan M. Fregeneda, Tania Fernv°ndez-Villa, Antonio J. Molina and SeqCOVID-SPAIN consortium |
| hCoV-19/Spain/CL-IBV-98029695/2020 | EPI_ISL_2754270 | 2020-05-19 | Universidad de Lev≥n | SeqCOVID-SPAIN consortium/IBV(CSIC) | Ana Carvajal, Vicente Martv≠n, Hv©ctor Argvºello, Juan M. Fregeneda, Tania Fernv°ndez-Villa, Antonio J. Molina and SeqCOVID-SPAIN consortium |
| hCoV-19/Chile/ML-111500/2021 | EPI_ISL_2756159 | 2021-06-13 | Genetica Molecular and Subdepartamento de Virologia ISP Chile | Instituto de Salud Publica de Chile | Karen Orostica, Constanza Campano, Barbara Parra, Loredana Arata, Gisselle Barra, Patricia Bustos, Rodrigo Fasce, Javier Tognarelli, Andres Castillo, Soledad Ulloa, Jorge Fernandez |
| hCoV-19/Austria/KILM655/2021 | EPI_ISL_2757918 | 2021-06-18 | Lifebrain Covid Labor GmbH | Dept. of Laboratory Medicine | Harald Esterbauer, Anna Gschaider, Petra Jurkowitsch, Fabian Konig, Sabina Plumer, Robert Strassl, Claudia Weber, Oswald Wagner |

| | | | | | |
|---|---|---|---|---|---|
| hCoV-19/Brazil/PR-FIOCRUZ-9952-TG-177/2020 | EPI_ISL_2759071 | 2020-06-04 | Laboratorio Central de Saude Publica do Estado do Parana (Instituto de Biologia Molecular do Paranv° (LAC)EN-PR) | Instituto Carlos Chagas - Fiocruz | Mauro de Medeiros Oliveira, Michelle Orane Schemberger, Andreia Akemi Suzukawa, Irina Nastassja Riediger, Maria do Carmo Debur, Guilherme Becker, Paola Cristina Resende, Tiago Grv§f, Eduardo Balsanelli, Valter Antv¥nio de Baura, Emanuel Maltempi de Souza, |
| hCoV-19/Germany/NI-RKI-I-183652/2021 | EPI_ISL_2762095 | 2021-06-19 | Sonic - MVZ Medizinisches Labor Oldenburg | Robert Koch Institute | unknown |
| hCoV-19/Germany/SN-RKI-I-184100/2021 | EPI_ISL_2762703 | 2021-06-21 | Bioscientia Labor Wermsdorf | Robert Koch Institute | unknown |
| hCoV-19/SouthAfrica/KRISP-K018778/2021 | EPI_ISL_2770468 | 2021-05-17 | NHLS Charlotte Maxeke Johannesburg Academic Hospital and the University of the Witwatersrand | KRISP, KZN Research Innovation and Sequencing Platform | Florette Treurnicht, Bulelani Manene, Kathleen Subramoney, Giandhari Jennifer, Pillay Sureshnee, Yajna Ramphal, Naidoo Yeshnee, Tshabuila Derek,Tegally Houriiyah, San James, Wilkinson Eduan, de Oliveira Tulio |
| hCoV-19/Japan/PG-57857/2021 | EPI_ISL_2772093 | 2021-05-21 | SARS-CoV-2 testing team, National Institute of Infectious Diseases | Pathogen Genomics Center, National Institute of Infectious Diseases | Tsuyoshi Sekizuka, Kentaro Itokawa, Rina Tanaka, Masanori Hashino, Nozomu Hanaoka, Masumichi Saito, Naomi Nojiri, Hazuka Y Furihata, Hiromizu Takahashi, Tsuguto Fujimoto, Makoto Kuroda |
| hCoV-19/Japan/PG-48705/2021 | EPI_ISL_2772767 | 2021-05-13 | Fukuoka Institute of Health and Environmental Sciences | Fukuoka Institute of Health and Environmental Sciences | Saori Ueda, Yoshiki Etoh, Hiroaki Shigemura, Akira Ohishi, Takayuki Kobayashi, Carl Yuki, Chiharu Katamune, Mitsuhiro Hamasaki, Yuki Ashizuka, Tsuyoshi Sekizuka, Kentaro Itokawa, Rina Tanaka, Masanori Hashino, Makoto Kuroda |
| hCoV-19/India/TG-CCMB-CIA4804/2021 | EPI_ISL_2774924 | 2021-05-31 | CSIR-Centre for Cellular and Molecular Biology | CSIR-Centre for Cellular and Molecular Biology-INSACOG | Ara Sreenivas,Shreekant Verma, Amareshwar Vodapalli ,B Himasri,Valli Nagalakshmi Undamatla,Payel Mukherjee, Lamuk Zaveri, Onkar Kulkarni,Jandhyala Sai Krishna,Tulasi Nagabandi, Sofia Banu, ,Priya Nurkurthy, Vidhyadhari Methuku, Sumedha Avadhanula, Archana |
| hCoV-19/Kenya/BUS-453-20/2020 | EPI_ISL_2779423 | 2020-09-11 | GK Prison Busia (Matayo)Matayos Prison | USAMRD-A, Basic Science Laboratory | Gathii Kimita, Josphat Nyataya, Esther Omuseni, Faith Sigei, Alan Lemtudo, Eric Muthanje, Brian Andika, Rehema Liyai, Rachel Githii, Clement Masakwe, Stephen Ochola, George Awinda, Carol Kifude, Beth Mutai, John Waitumbi |
| hCoV-19/Kenya/MOH-K-BTK-DS-368/2021 | EPI_ISL_2779510 | 2021-02-15 | BATUK | USAMRD-A, Basic Science Laboratory | Gathii Kimita, Josphat Nyataya, Esther Omuseni, Faith Sigei, Alan Lemtudo, Eric Muthanje, Brian Andika, Rehema Liyai, Rachel Githii, Clement Masakwe, Stephen Ochola, George Awinda, Carol Kifude, Beth Mutai, John Waitumbi |
| hCoV-19/Kenya/SOMP-IW-052/2020 | EPI_ISL_2779549 | 2020-07-15 | MOH-K | USAMRD-A, Basic Science Laboratory | Gathii Kimita, Josphat Nyataya, Esther Omuseni, Faith Sigei, Alan Lemtudo, Eric Muthanje, Brian Andika, Rehema Liyai, Rachel Githii, Clement Masakwe, Stephen Ochola, George Awinda, Carol Kifude, Beth Mutai, John Waitumbi |

| | | | | | |
|---|---|---|---|---|---|
| hCoV-19/USA/TX-HMH-MCoV-28136/2021 | EPI_ISL_1237918 | 2021-02-09 | Houston Methodist Hospital | Houston Methodist Hospital | S. Wesley Long, Randall J. Olsen, Paul A. Christensen, Sishir Subedi, Robert Olson, James J. Davis, Matthew Ojeda Saavedra, Prasanti Yerramilli, Layne Pruitt, Kristina Reppond, Madison N. Shyer, Jessica Cambric, Ilya J. Finkelstein, Jimmy Gollihar, and |
| hCoV-19/USA/NY-CDC-LC0033581/2021 | EPI_ISL_1514693 | 2021-03-18 | Laboratory Corporation of America | Centers for Disease Control and Prevention Division of Viral Diseases, Pathogen Discovery | Dakota Howard, Dhwani Batra, Peter W. Cook, Kara Moser, Adrian Paskey, Jason Caravas, Benjamin Rambo-Martin, Shatavia Morrison, Christopher Gulvick, Scott Sammons, Yvette Unoarumhi, Darlene Wagner, Matthew Schmerer, Minoo Agarwal, Eyad Almasri, Debbie Bol |
| hCoV-19/USA/HI-H211426/2021 | EPI_ISL_1515827 | 2021-03-09 | State Laboratories Division, Hawaii State Department of Health | State Laboratories Division, Hawaii State Department of Health | Pamela O'Brien, Drew Kuwazaki, Ayana Garnet, Razvan Sultana, Edward Desmond |
| hCoV-19/USA/CA-CDPH-2000018730/2021 | EPI_ISL_2783510 | 2021-01-09 | CDPH VBL | California Department of Public Health | CDPH-COVIDNet,UCI Genome Sciences Center/GHTF |
| hCoV-19/Australia/VIC1894/2020 | EPI_ISL_480650 | 2020-05-31 | Victorian Infectious Diseases Reference Laboratory (VIDRL) | VIDRL and MDU-PHL | Caly L., Seemann T., Sait, M., Schultz M., Druce J., Sherry, N. |
| hCoV-19/Australia/VIC1958/2020 | EPI_ISL_480682 | 2020-06-11 | Victorian Infectious Diseases Reference Laboratory (VIDRL) | VIDRL and MDU-PHL | Caly L., Seemann T., Sait, M., Schultz M., Druce J., Sherry, N. |
| hCoV-19/Scotland/QEUH-14BCD7E/2021 | EPI_ISL_1584953 | 2021-04-02 | Lighthouse Lab in Glasgow | Wellcome Sanger Institute for the COVID-19 Genomics UK (COG-UK) Consortium | Harper VanSteenhouse, Yumi Kasai, David Gray, Carol Clugston, Anna Dominiczak and Alex Alderton, Roberto Amato, Jeffrey Barrett, Sonia Goncalves, Ewan Harrison, David K. Jackson, Ian Johnston, Dominic Kwiatkowski, Cordelia Langford, John Sillitoe on behal |
| hCoV-19/Canada/NL-NML-17515/2021 | EPI_ISL_1587837 | 2021-02-20 | NL-Dr. Leonard A. Miller Centre for Health Services | National Microbiology Laboratory (NML) | Anna Majer, Shari Tyson, Grace Seo, Philip Mabon, Elsie Grudeski, Rhiannon Huzarewich, Russell Mandes, Anneliese Landgraff, Jennifer Tanner, Natalie Knox, Morag Graham, Gary Van Domselaar, Robert Needle, Yang Yu, Adel Malek, Laura Gilbert, George Zahariad |
| hCoV-19/India/JH-1931500983945/2021 | EPI_ISL_1589860 | 2021-03-23 | INSACOG-WB | National Institute of Biomedical Genomics ‚Äì INSACOG | Arindam Maitra, Bhaswati Bandyopadhyay, Nidhan Kumar Biswas, Tamal Ghosh, Sreedhar Chinnaswamy, Ajay Chakraborti, Saumitra Das |
| hCoV-19/Netherlands/LI-RIVM-23438/2021 | EPI_ISL_1596319 | 2021-03-22 | Dutch COVID-19 response team | National Institute for Public Health and the Environment (RIVM) | Adam Meijer, Harry Vennema, Dirk Eggink, Jeroen Cremer, Sharon van den Brink, Bas van der Veer, AnneMarie van den Brandt, Lisa Wijsman, Kim Freriks, Ryanne Jaarsma, Eun√≠ce Then, Jolienke Hardeman, Lynn Aarts, Sanne Bos, Melissa van Tuil, Robert Kohl, Lin |
| hCoV-19/Netherlands/ZH-RIVM-24157/2021 | EPI_ISL_1597037 | 2021-03-06 | Dutch COVID-19 response team | National Institute for Public Health and the Environment (RIVM) | Adam Meijer, Harry Vennema, Dirk Eggink, Jeroen Cremer, Sharon van den Brink, Bas van der Veer, AnneMarie van den Brandt, Lisa Wijsman, Kim Freriks, Ryanne Jaarsma, Eun√≠ce Then, Jolienke Hardeman, Lynn Aarts, Sanne Bos, Melissa van Tuil, Robert Kohl, Lin |
| hCoV-19/USA/GA-CDC-LC0037682/2021 | EPI_ISL_1610781 | 2021-03-31 | Laboratory Corporation of America | Centers for Disease Control and Prevention Division of Viral Diseases, Pathogen Discovery | Dakota Howard, Dhwani Batra, Peter W. Cook, Kara Moser, Adrian Paskey, Jason Caravas, Benjamin Rambo-Martin, Shatavia Morrison, Christopher Gulvick, Scott Sammons, Yvette Unoarumhi, Darlene Wagner, Matthew Schmerer, Minoo Agarwal, Eyad Almasri, Debbie Bol |

| | | | | | |
|---|---|---|---|---|---|
| hCoV-19/USA/PA-CDC-LC0038275/2021 | EPI_ISL_1611372 | 2021-03-31 | Laboratory Corporation of America | Centers for Disease Control and Prevention Division of Viral Diseases, Pathogen Discovery | Dakota Howard, Dhwani Batra, Peter W. Cook, Kara Moser, Adrian Paskey, Jason Caravas, Benjamin Rambo-Martin, Shatavia Morrison, Christopher Gulvick, Scott Sammons, Yvette Unoarumhi, Darlene Wagner, Matthew Schmerer, Minoo Agarwal, Eyad Almasri, Debbie Bol |
| hCoV-19/NewZealand/21MV0338/2021 | EPI_ISL_1621305 | 2021-04-08 | LabPLUS | Institute of Environmental Science and Research (ESR) | Rachel Boyle, SallyAnn Harbison, Olivia Stroeven, Xiaoyun Ren, Matt Storey, Nikki Freed, Muhammad Faisal, Jing Wang, Hermes Perez, Anja Werno, Antje van der Linden, Arlo Upton, Chris Mansell, David Hammer, Dragana Drinkovic, Gary McAuliffe, Hana Sofia And |
| hCoV-19/NewZealand/21MV0256/2021 | EPI_ISL_1621312 | 2021-03-30 | LabPLUS | Institute of Environmental Science and Research (ESR) | Rachel Boyle, SallyAnn Harbison, Olivia Stroeven, Xiaoyun Ren, Matt Storey, Nikki Freed, Muhammad Faisal, Jing Wang, Hermes Perez, Anja Werno, Antje van der Linden, Arlo Upton, Chris Mansell, David Hammer, Dragana Drinkovic, Gary McAuliffe, Hana Sofia And |
| hCoV-19/USA/WV-WVU-WV091014/2021 | EPI_ISL_1624970 | 2021-03-31 | Berkeley Medical Center | WVU and Marshall University Combined Genomics Core Facilities | James Denvir, Peter Stoilov, Peter Perrotta, Wesley Kimble, Ryan Percifield |
| hCoV-19/FrenchGuiana/IPG2021124/2021 | EPI_ISL_1627110 | 2021-04-07 | Laboratoire Carage | Institut Pasteur de la Guyane | Anne Lavergne, Dominique Rousset |
| hCoV-19/Reunion/PIMIT_00992/2021 | EPI_ISL_1629001 | 2021-02-17 | CHU | UMR PIMIT | Dr David A Wilkinson, Dr Patrick Mavingui, Dr Camille Lebarbenchon, Magali Turpin |
| hCoV-19/Peru/ARE-UPCH-0373/2021 | EPI_ISL_1629765 | 2021-01-01 | Hospital Carlos Alberto Seguv≠n Escobedo - EsSalud | Laboratorio de Genv≥mica Microbiana, Universidad Peruana Cayetano Heredia | Lenin Maturrano, Pablo Tsukayama, Alejandra Dv°vila-Barclay, Guillermo Salvatierra, Luis Gonzv°lez, Pedro E. Romero, Janet Huancachoque, Pool Marcos, Fernando Sv°nchez Fragoso, Lin Zevallos Cuarite |
| hCoV-19/England/LIVE-E0E16B/2021 | EPI_ISL_1636862 | 2021-03-31 | Liverpool Clinical Laboratories | COVID-19 Genomics UK (COG-UK) Consortium | Sam Haldenby, Alistair Darby, Steve Paterson, Anita Lucaci, Julian Hiscox, M Almsaud, A Alrezaihi, Muhannad Alruwaili, Stuart D Armstrong, Jones Benjamin, Eleanor G Bentley, Anu Chawla, Jordan J Clark, Angela Cowell, Richard Eccles, Isabel Garcv≠a-Doriva |
| hCoV-19/Australia/NSW1555/2021 | EPI_ISL_1904462 | 2021-04-25 | South Eastern Area Laboratory Services (SEALS) | Clinical Pathology and Medical Research; Westmead Hospital; University of Sydney | CIDM-PH et al. |
| hCoV-19/Australia/NSW1557/2021 | EPI_ISL_1904463 | 2021-04-26 | South Eastern Area Laboratory Services (SEALS) | Clinical Pathology and Medical Research; Westmead Hospital; University of Sydney | CIDM-PH et al. |
| hCoV-19/Australia/NSW1564/2021 | EPI_ISL_1904468 | 2021-04-27 | South Eastern Area Laboratory Services (SEALS) | Clinical Pathology and Medical Research; Westmead Hospital; University of Sydney | CIDM-PH et al. |
| hCoV-19/India/HR-NCDC-INSACOG-1309157/2021 | EPI_ISL_1908756 | 2021-03-08 | Biotechnology Division, NCDC Delhi | NCDC Delhi, Biotechnology Division | Radhakrishnan V. S, Robin Marwal, Mahesh S Dhar, Kalaiarasan Ponnusamy, Meena Datta, Priyanka Singh, Uma Sharma, Manoj K Singh, Hemlata Lall, Hema Gogia, Preeti Madan, Sandhya Kabra, Sujeet K Singh, Partha Rakshit |

| | | | | | |
|---|---|---|---|---|---|
| hCoV-19/Australia/NSW4594/2021 | EPI_ISL_1911185 | 2021-04-25 | Area of Virology, Serology and Virology Division (SAViD), New South Wales Health Pathology Randwick | Virology, Serology and Virology Division (SAViD), New South Wales Health Pathology Randwick | Foster, C.; Au, J.; Ruiz Silva, M.; Deveson, I.; Bull, R.; Van Hal, S.; Rawlinson, W. |
| hCoV-19/Mexico/NLE-LESPNL-20210407040/2021 | EPI_ISL_1911264 | 2021-03-27 | LESPNL | LESPNL | (in alphabetical order) Consuelo Treviv±o-Garza, Eduardo Isaac de la Rosa-Moreno, Else del Carmen Garcia-Garcia, Gloria Alejandra Jasso-de la Pev±a, Manuel Enrique de la O-Cavazos, Olv≠n Medina-Chv°vez, Yulianna Mayre Cordero-Cruz |
| hCoV-19/India/CT-AIIMS-Raipur-L-1040/2021 | EPI_ISL_1914596 | 2021-03-30 | State Virus Research and Diagnostic Laboratory (VRDL), AIIMS Raipur | State Virus Research and Diagnostic Laboratory (VRDL), AIIMS Raipur | Pushpendra Singh, Kuldeep Sharma, Priyanka Singh, Somya Sharma, Sanjay Singh Negi, Anudita Bhargava |
| hCoV-19/Australia/WA715/2021 | EPI_ISL_1914668 | 2021-05-01 | PathWest Laboratory Medicine WA | PathWest Laboratory Medicine WA Microbial Surveillance Unit | PathWest Laboratory Medicine WA Microbial Surveillance Unit |
| hCoV-19/SouthAfrica/NICD-N3324/2021 | EPI_ISL_1914810 | 2021-01-05 | National Institute for Communicable Diseases,National Health Laboratory Services, Gauteng, South Africa | National Institute for Communicable Diseases of the National Health Laboratory Service | Amoako DG, Scheepers C, Mohale T, Ntuli N, Mahlangu B, Ismail A, Bhiman JN |
| hCoV-19/France/NOR-IPP10456/2021 | EPI_ISL_1915155 | 2021-04-20 | Labo Analyses Med | National Reference Center for Viruses of Respiratory Infections, Institut Pasteur, Paris | Marion Barbet, Sylvie Behillil, Mv©line Bizard, Angela Brisebarre, Camille Capel, Vincent Enouf, Louise Lefranv§ois, Frv©dv©ric Lemoine, Christophe Malabat, Corinne Maufrais, Emmanuelle Permal, Etienne Simon-Loriv®re, Maud Vanpeene, Sylvie Van der Werf ,F |
| hCoV-19/India/MH-NEERI-NGP-36422/2021 | EPI_ISL_1916460 | 2021-02-20 | CSIR-National Environmental Engineering Research Institute | CSIR-Centre for Cellular and Molecular Biology - INSACOG | Ara Sreenivas, Onkar Kulkarni, Lamuk Zaveri, Sofia Banu, Shreekant Verma, Amareshwar Vodapalli , Viswagithe S L, B Himasri, Sharath Chandra Thota, Karthik Bharadwaj Tallapaka, Krishna Khairnar, Rakesh K Mishra,Divya Tej Sowpati |
| hCoV-19/India/MH-NEERI-NGP-41285/2021 | EPI_ISL_1916474 | 2021-03-13 | CSIR-National Environmental Engineering Research Institute | CSIR-Centre for Cellular and Molecular Biology - INSACOG | Ara Sreenivas, Onkar Kulkarni, Lamuk Zaveri, Sofia Banu, Shreekant Verma, Amareshwar Vodapalli , Viswagithe S L, B Himasri, Sharath Chandra Thota, Karthik Bharadwaj Tallapaka, Krishna Khairnar, Rakesh K Mishra,Divya Tej Sowpati |
| hCoV-19/USA/MD-CDC-LC0049707/2021 | EPI_ISL_1925936 | 2021-04-15 | Laboratory Corporation of America | Centers for Disease Control and Prevention Division of Viral Diseases, Pathogen Discovery | Dakota Howard, Dhwani Batra, Peter W. Cook, Kara Moser, Adrian Paskey, Jason Caravas, Benjamin Rambo-Martin, Shatavia Morrison, Christopher Gulvick, Scott Sammons, Yvette Unoarumhi, Darlene Wagner, Matthew Schmerer, Minoo Agarwal, Eyad Almasri, Debbie Bol |
| hCoV-19/USA/PA-CDC-LC0050945/2021 | EPI_ISL_1928166 | 2021-04-22 | Laboratory Corporation of America | Centers for Disease Control and Prevention Division of Viral Diseases, Pathogen Discovery | Dakota Howard, Dhwani Batra, Peter W. Cook, Kara Moser, Adrian Paskey, Jason Caravas, Benjamin Rambo-Martin, Shatavia Morrison, Christopher Gulvick, Scott Sammons, Yvette Unoarumhi, Darlene Wagner, Matthew Schmerer, Minoo Agarwal, Eyad Almasri, Debbie Bol |
| hCoV-19/India/MH-ICMR-NIV-INSACOG-GSEQ-2910/2021 | EPI_ISL_1928522 | 2021-04-17 | ICMR-National Institute of Virology - INSACOG | NIV Influenza | Potdar, Varsha on behalf of National Influenza Centre, NIV, Pune |
| hCoV-19/India/MH-ICMR-NIV-INSACOG-GSEQ-3182/2021 | EPI_ISL_1928595 | 2021-04-20 | ICMR-National Institute of Virology - INSACOG | NIV Influenza | Potdar, Varsha on behalf of National Influenza Centre, NIV, Pune |

| | | | | | |
|---|---|---|---|---|---|
| hCoV-19/USA/KY-CDC-LC0051408/2021 | EPI_ISL_1929003 | 2021-04-24 | Laboratory Corporation of America | Centers for Disease Control and Prevention Division of Viral Diseases, Pathogen Discovery | Dakota Howard, Dhwani Batra, Peter W. Cook, Kara Moser, Adrian Paskey, Jason Caravas, Benjamin Rambo-Martin, Shatavia Morrison, Christopher Gulvick, Scott Sammons, Yvette Unoarumhi, Darlene Wagner, Matthew Schmerer, Minoo Agarwal, Eyad Almasri, Debbie Bol |
| hCoV-19/USA/TX-CDC-LC0052589/2021 | EPI_ISL_1930735 | 2021-04-19 | Laboratory Corporation of America | Centers for Disease Control and Prevention Division of Viral Diseases, Pathogen Discovery | Dakota Howard, Dhwani Batra, Peter W. Cook, Kara Moser, Adrian Paskey, Jason Caravas, Benjamin Rambo-Martin, Shatavia Morrison, Christopher Gulvick, Scott Sammons, Yvette Unoarumhi, Darlene Wagner, Matthew Schmerer, Minoo Agarwal, Eyad Almasri, Debbie Bol |
| hCoV-19/Japan/PG-31288/2021 | EPI_ISL_1932178 | 2021-02-16 | Pathogen Genomics Center, National Institute of Infectious Diseases | Pathogen Genomics Center, National Institute of Infectious Diseases | Tsuyoshi Sekizuka, Kentaro Itokawa, Rina Tanaka, Masanori Hashino, Makoto Kuroda |
| hCoV-19/Japan/PG-35430/2021 | EPI_ISL_1934015 | 2021-04-01 | Pathogen Genomics Center, National Institute of Infectious Diseases | Pathogen Genomics Center, National Institute of Infectious Diseases | Tsuyoshi Sekizuka, Kentaro Itokawa, Rina Tanaka, Masanori Hashino, Makoto Kuroda |
| hCoV-19/Brazil/SP-IB_100200/2021 | EPI_ISL_1966235 | 2021-03-15 | CENTRO DE SAUDE III SALES OLIVEIRA | Instituto Butantan / Mendelics | Instituto Butantan: Dimas Tadeu Covas, Sandra Coccuzzo Sampaio, Maria Carolina Elias, Jos√© Salvatore Leister Patan√©, Vincent Louis Viala, Antonio Jorge Martins, Ricardo Haddad, Claudia Renata dos Santos Barros, Elaine Cristina Marqueze, Raul Machado Net |
| hCoV-19/India/KA-SEQ_444_S47_R1_001/2021 | EPI_ISL_1940014 | 2021-03-04 | HOSAHALLI | inStem NCBS - INSACOG | Uma Ramakrishnan Dasaradhi Palakodeti Aswin SaiNarain |
| hCoV-19/India/KA-SEQ_546_S155_R1_001/2021 | EPI_ISL_1940110 | 2021-04-06 | GOVINDRAJNAGAR | inStem NCBS - INSACOG | Uma Ramakrishnan Dasaradhi Palakodeti Aswin SaiNarain |
| hCoV-19/Brazil/SP-IB_100620/2021 | EPI_ISL_1966629 | 2021-04-05 | SECAO CENTRO DE DIAGNOSTICO SECEDI | Instituto Butantan / Mendelics | Instituto Butantan: Dimas Tadeu Covas, Sandra Coccuzzo Sampaio, Maria Carolina Elias, Jos√© Salvatore Leister Patan√©, Vincent Louis Viala, Antonio Jorge Martins, Ricardo Haddad, Claudia Renata dos Santos Barros, Elaine Cristina Marqueze, Raul Machado Net |
| hCoV-19/Aruba/AW-RIVM-29821/2021 | EPI_ISL_1962180 | 2021-04-22 | Dutch COVID-19 response team | National Institute for Public Health and the Environment (RIVM) | Adam Meijer, Harry Vennema, Dirk Eggink, Jeroen Cremer, Sharon van den Brink, Bas van der Veer, AnneMarie van den Brandt, Lisa Wijsman, Kim Freriks, Ryanne Jaarsma, Eun√≠ce Then, Lynn Aarts, Sanne Bos, Melissa van Tuil, Linda van de Nes, Sjoerd Kuiling, J |
| hCoV-19/Netherlands/ZH-RIVM-28879/2021 | EPI_ISL_1962267 | 2021-04-19 | Dutch COVID-19 response team | National Institute for Public Health and the Environment (RIVM) | Adam Meijer, Harry Vennema, Dirk Eggink, Jeroen Cremer, Sharon van den Brink, Bas van der Veer, AnneMarie van den Brandt, Lisa Wijsman, Kim Freriks, Ryanne Jaarsma, Eun√≠ce Then, Lynn Aarts, Sanne Bos, Melissa van Tuil, Linda van de Nes, Sjoerd Kuiling, J |
| hCoV-19/Aruba/AW-RIVM-29740/2021 | EPI_ISL_1962940 | 2021-04-22 | Dutch COVID-19 response team | National Institute for Public Health and the Environment (RIVM) | Adam Meijer, Harry Vennema, Dirk Eggink, Jeroen Cremer, Sharon van den Brink, Bas van der Veer, AnneMarie van den Brandt, Lisa Wijsman, Kim Freriks, Ryanne Jaarsma, Eun√≠ce Then, Lynn Aarts, Sanne Bos, Melissa van Tuil, Linda van de Nes, Sjoerd Kuiling, J |

| | | | | | |
|---|---|---|---|---|---|
| hCoV-19/Aruba/AW-RIVM-28065/2021 | EPI_ISL_1962944 | 2021-04-19 | Dutch COVID-19 response team | National Institute for Public Health and the Environment (RIVM) | Adam Meijer, Harry Vennema, Dirk Eggink, Jeroen Cremer, Sharon van den Brink, Bas van der Veer, AnneMarie van den Brandt, Lisa Wijsman, Kim Freriks, Ryanne Jaarsma, Eun√≠ce Then, Lynn Aarts, Sanne Bos, Melissa van Tuil, Linda van de Nes, Sjoerd Kuiling, J |
| hCoV-19/USA/NY-CUIMC-NP-6257/2021 | EPI_ISL_1965786 | 2021-03-24 | Clinical Microbiology Laboratory, NewYork Presbyterian Hospital/Columbia University Irving Medical Center | Uhlemann Laboratory, Columbia University Irving Medical Center | Medini K. Annavajhala, Anne Kelley, Maya Tagliavia, Anne-Catrin Uhlemann |
| hCoV-19/Indonesia/KT-NIHRD-WGS02181/2021 | EPI_ISL_1969246 | 2021-03-19 | Rumah Sakit Umum Daerah Palangkaraya | National Institute of Health Research and Development | Subangkit, Hana Apsari Pawestri, Kartika Dewi Puspa, Arie Ardiansyah Nugraha, Hartanti Dian Ikawati, Krisna Pangesti, Triyani Soekarso, Nelly Puspandari, Vivi Setiawaty |
| hCoV-19/India/DEL-NCDC-INSACOG-2209110/2021 | EPI_ISL_1970153 | 2021-04-01 | Biotechnology Division, NCDC Delhi | NCDC Delhi, Biotechnology Division | Kalaiarasan Ponnusamy, Meena Datta,¬†Priyanka Singh,¬†Uma Sharma,¬†Manoj K Singh,¬†Radhakrishnan V. S, Robin Marwal, Mahesh S Dhar, Hemlata Lall, Hema Gogia, Preeti Madan, Sandhya Kabra, Sujeet K Singh, Partha Rakshit |
| hCoV-19/England/ALDP-154E3E7/2021 | EPI_ISL_1987156 | 2021-04-30 | Lighthouse Lab in Alderley Park | Wellcome Sanger Institute for the COVID-19 Genomics UK (COG-UK) Consortium | Jacquelyn Wynn, Mairead Hyland, The Lighthouse Lab in Alderley Park and Alex Alderton, Roberto Amato, Jeffrey Barrett, Sonia Goncalves, Ewan Harrison, David K. Jackson, Ian Johnston, Dominic Kwiatkowski, Cordelia Langford, John Sillitoe on behalf of the W |
| hCoV-19/India/MH-ICMR-NIV-INSAGOG-GSEQ-3240/2021 | EPI_ISL_1970535 | 2021-04-26 | ICMR-National Institute of Virology - INSACOG | NIV Influenza | Potdar, Varsha on behalf of National Influenza Centre, NIV, Pune |
| hCoV-19/USA/MA-CDCBI-CRSP_BOQ4QIXLVPOYIOPW/2021 | EPI_ISL_1971653 | 2021-04-30 | Broad Institute Clinical Research Sequencing Platform | Infectious Disease Program, Broad Institute of Harvard and MIT | Siddle,K.J., Adams,G., Pearlman,L., Gladden-Young,A., Vicente,G., Blumenstiel,B., DeFelice,M., Lee,M., McGovern,S., Lagerborg,K., Rudy,M., DeRuff,K., Carter,A., Normandin,E., Bauer,M., Reilly,S., Tomkins-Tinch,C., Loreth,C., Chaluvadi,S., Meldrim,J., Gran |
| hCoV-19/England/NORT-1BA4422/2021 | EPI_ISL_1978628 | 2021-04-24 | Northumbria University / South Tees Hospitals NHS Foundation Trust / North Cumbria Integrated Care NHS Foundation Trust / North Tees and Hartlepool NHS Foundation Trust / Newcastle Hospitals NHS Foundation Trust | COVID-19 Genomics UK (COG-UK) Consortium | Darren L Smith,Andrew Nelson,Matthew Bashton,Greg R Young,Joshua Loh,John Allan,Mohammad A Tariq,Giles S Holt,Gary Black,Wen C Yew,Lynn Dover,Paul Baker,Steve Liggett,Sarah Essex,Jane Greenaway,Debra Padgett,Clive Graham,Garren Scott,Edward Barton,Emma Sw |
| hCoV-19/Wales/PHWC-PYRO8Y/2021 | EPI_ISL_1979377 | 2021-04-26 | Originating lab: Wales Specialist Virology Centre Sequencing lab: Pathogen Genomics Unit | Public Health Wales Microbiology Cardiff Wales Specialist Virology Centre | Catherine Moore, Johnathan Evans, Laura Gifford, Malorie Perry, Simon Cottrell, Angela Marchbank, Alec Birchley, Alexander Adams, Amy Gaskin, Bree Gatica-Wilcox, Jason Coombes, Joel Southgate, Lauren Gilbert, Lee Graham, Nicole Pacchiarini, Sara Kumziene- |
| hCoV-19/England/MILK-155318B/2021 | EPI_ISL_1986293 | 2021-04-30 | Lighthouse Lab in Milton Keynes | Wellcome Sanger Institute for the COVID-19 Genomics UK (COG-UK) Consortium | The Lighthouse Lab in Milton Keynes and Alex Alderton, Roberto Amato, Jeffrey Barrett, Sonia Goncalves, Ewan Harrison, David K. Jackson, Ian Johnston, Dominic Kwiatkowski, Cordelia Langford, John Sillitoe on behalf of the Wellcome Sanger Institute COVID-1 |

| | | | | | |
|---|---|---|---|---|---|
| hCoV-19/Germany/BE-ChVir-LB-210305-4404/2021 | EPI_ISL_1989297 | 2021-03-02 | Labor Berlin CharitéÃÅ Vivantes GmbH / Institut fuÃàr Virologie | CharitéÃÅ UniversitaÃàtsmedizin Berlin, Institut fuÃàr Virologie/Labor Berlin | Peter Menzel, Christine Stephan, Rolf Schwarzer, Victor M Corman, Barbara MuÃàhlemann, Terry Jones, Christian Drosten |
| hCoV-19/EquatorialGuinea/94388/2021 | EPI_ISL_1989339 | 2021-02-13 | Laboratorio de Investigaciones de Baney | Swiss Tropical and Public Health Institute | Salome Hosch, Carlos Cortes, Claudia Daubenberger, Guillermo Garcia, Bonifacio Manguire Nlavo, Maximilian Mpina, Elizabeth Nyakarungu, Diosdado Odjama Nseng Ada, Mitoha Ondo O Ayekaba, Tobias Schindler, Philipp Wagner, Philip Wonder Phiri |
| hCoV-19/USA/CA-CDC-FG-028497/2021 | EPI_ISL_1992133 | 2021-04-29 | Fulgent Genetics | Centers for Disease Control and Prevention Division of Viral Diseases, Pathogen Discovery | Dakota Howard, Dhwani Batra, Peter W. Cook, Kara Moser, Adrian Paskey, Jason Caravas, Benjamin Rambo-Martin, Shatavia Morrison, Christopher Gulvick, Scott Sammons, Yvette Unoarumhi, Darlene Wagner, Matthew Schmerer, Harry Gao, Mickey Li, John Gao, Joseph |
| hCoV-19/USA/CO-CDC-FG-028311/2021 | EPI_ISL_1992395 | 2021-04-22 | Fulgent Genetics | Centers for Disease Control and Prevention Division of Viral Diseases, Pathogen Discovery | Dakota Howard, Dhwani Batra, Peter W. Cook, Kara Moser, Adrian Paskey, Jason Caravas, Benjamin Rambo-Martin, Shatavia Morrison, Christopher Gulvick, Scott Sammons, Yvette Unoarumhi, Darlene Wagner, Matthew Schmerer, Harry Gao, Mickey Li, John Gao, Joseph |
| hCoV-19/Ghana/NMIMR-NTRA-21-37138/2021 | EPI_ISL_2001076 | 2021-03-16 | Immunology, Noguchi Memorial Institute for Medical Research | Immunology, Noguchi Memorial Institute for Medical Research | Adu,B., Egyir,B., Kumordjie,S., Agbodzi,B., Yeboah,C., Mohktar,Q., Oteng,F., Owusu-Nyantakyi,C., Asare,K.M., Appiah-Kubi,J., Bonney,J.K., Odoom,J.K. |
| hCoV-19/Ecuador/USFQ-1239/2021 | EPI_ISL_2004114 | 2021-04-12 | Institute of Microbiology, Universidad San Francisco de Quito | Institute of Microbiology, Universidad San Francisco de Quito | Sully M√°rquez, Juan Jos√© Guadalupe, Bel√©n Prado-Vivar, Monica Becerra-Wong, Bernardo Guti√©rrez, Victor Vinicio Rom√°n, Ver√≥nica Barrag√°n, Patricio Rojas-Silva, Gabriel Trueba, Michelle Grunauer, Pa√∫l C√°rdenas |
| hCoV-19/England/ALDP-15578BF/2021 | EPI_ISL_2004426 | 2021-05-01 | Lighthouse Lab in Alderley Park | Wellcome Sanger Institute for the COVID-19 Genomics UK (COG-UK) Consortium | Jacquelyn Wynn, Mairead Hyland, The Lighthouse Lab in Alderley Park and Alex Alderton, Roberto Amato, Jeffrey Barrett, Sonia Goncalves, Ewan Harrison, David K. Jackson, Ian Johnston, Dominic Kwiatkowski, Cordelia Langford, John Sillitoe on behalf of the W |
| hCoV-19/England/MILK-1541FB3/2021 | EPI_ISL_2005525 | 2021-04-26 | Lighthouse Lab in Milton Keynes | Wellcome Sanger Institute for the COVID-19 Genomics UK (COG-UK) Consortium | The Lighthouse Lab in Milton Keynes and Alex Alderton, Roberto Amato, Jeffrey Barrett, Sonia Goncalves, Ewan Harrison, David K. Jackson, Ian Johnston, Dominic Kwiatkowski, Cordelia Langford, John Sillitoe on behalf of the Wellcome Sanger Institute COVID-1 |
| hCoV-19/England/RAND-15540F6/2021 | EPI_ISL_2006007 | 2021-04-29 | Randox Laboratories | Wellcome Sanger Institute for the COVID-19 Genomics UK (COG-UK) Consortium | Randox Laboratories and Alex Alderton, Roberto Amato, Jeffrey Barrett, Sonia Goncalves, Ewan Harrison, David K. Jackson, Ian Johnston, Dominic Kwiatkowski, Cordelia Langford, John Sillitoe on behalf of the Wellcome Sanger Institute COVID-19 Surveillance T |
| hCoV-19/England/HSLL-153F2CE/2021 | EPI_ISL_2006543 | 2021-04-25 | Health Services Laboratories | Wellcome Sanger Institute for the COVID-19 Genomics UK (COG-UK) Consortium | Health Services Laboratories and Alex Alderton, Roberto Amato, Jeffrey Barrett, Sonia Goncalves, Ewan Harrison, David K. Jackson, Ian Johnston, Dominic Kwiatkowski, Cordelia Langford, John Sillitoe on behalf of the Wellcome Sanger Institute COVID-19 Surve |

| | | | | | |
|---|---|---|---|---|---|
| hCoV-19/England/HSLL-153F2A0/2021 | EPI_ISL_2006613 | 2021-04-25 | Health Services Laboratories | Wellcome Sanger Institute for the COVID-19 Genomics UK (COG-UK) Consortium | Health Services Laboratories and Alex Alderton, Roberto Amato, Jeffrey Barrett, Sonia Goncalves, Ewan Harrison, David K. Jackson, Ian Johnston, Dominic Kwiatkowski, Cordelia Langford, John Sillitoe on behalf of the Wellcome Sanger Institute COVID-19 Surve |
| hCoV-19/Germany/BY-MVP-000005645/2021 | EPI_ISL_2006677 | 2021-03-18 | Max von Pettenkofer Institute, Virology, National Reference Center for Retroviruses, LMU Munich | Laboratory for Functional Genome Analysis; Dept. Genomics; Gene Center of the LMU Munich | Max Muenchhoff; Stefan Krebs; Alexander Graf; Oliver Keppler; Helmut Blum |
| hCoV-19/USA/MA-MASPHL-03521/2021 | EPI_ISL_2008162 | 2021-04-25 | Massachusetts State Public Health Laboratory | Massachusetts State Public Health Laboratory | Andrew Lang, Timelia Fink, Glen Gallagher, Sandra Smole |
| hCoV-19/Chile/RM-75715/2021 | EPI_ISL_2009293 | 2021-04-10 | Genetica Molecular and Subdepartamento de Virologia ISP Chile | Instituto de Salud Publica de Chile | Javier Tognarelli, Karen Orostica, Barbara Parra, Loredana Arata, Gisselle Barra, Patricia Bustos, Rodrigo Fasce, Andres Castillo, Soledad Ulloa, Jorge Fernandez |
| hCoV-19/India/GJ-GBRC645a/2021 | EPI_ISL_2017734 | 2021-03-29 | VRDL, Goverment Medical College (GMC), Surat | Gujarat Biotechnology Research Centre | Janvi Raval,Zarna Patel,Nitin Savaliya,Dinesh Kumar,Twinkle Soni,Sonal Sharma ,Zuber Saiyed,Ramesh Pandit,Neeta Khandelwal,Umang Mishra,Nitesh Shah,Chaitanya Joshi,Madhvi Joshi |
| hCoV-19/England/MILK-155E4A5/2021 | EPI_ISL_2021270 | 2021-04-26 | Lighthouse Lab in Milton Keynes | Wellcome Sanger Institute for the COVID-19 Genomics UK (COG-UK) Consortium | The Lighthouse Lab in Milton Keynes and Alex Alderton, Roberto Amato, Jeffrey Barrett, Sonia Goncalves, Ewan Harrison, David K. Jackson, Ian Johnston, Dominic Kwiatkowski, Cordelia Langford, John Sillitoe on behalf of the Wellcome Sanger Institute COVID-1 |
| hCoV-19/England/MILK-155D9F2/2021 | EPI_ISL_2021661 | 2021-05-04 | Lighthouse Lab in Milton Keynes | Wellcome Sanger Institute for the COVID-19 Genomics UK (COG-UK) Consortium | The Lighthouse Lab in Milton Keynes and Alex Alderton, Roberto Amato, Jeffrey Barrett, Sonia Goncalves, Ewan Harrison, David K. Jackson, Ian Johnston, Dominic Kwiatkowski, Cordelia Langford, John Sillitoe on behalf of the Wellcome Sanger Institute COVID-1 |
| hCoV-19/England/HSLL-15524ED/2021 | EPI_ISL_2022558 | 2021-04-30 | Health Services Laboratories | Wellcome Sanger Institute for the COVID-19 Genomics UK (COG-UK) Consortium | Health Services Laboratories and Alex Alderton, Roberto Amato, Jeffrey Barrett, Sonia Goncalves, Ewan Harrison, David K. Jackson, Ian Johnston, Dominic Kwiatkowski, Cordelia Langford, John Sillitoe on behalf of the Wellcome Sanger Institute COVID-19 Surve |
| hCoV-19/USA/WA-UW-21042239411/2021 | EPI_ISL_2031503 | 2021-04-22 | UW Virology Lab | UW Virology Lab | Pavitra Roychoudhury, Hong Xie, Lasata Shrestha, Tien V. Nguyen, Shah Mohamed Bakhash, Michelle Lin, Noah R. Baker, Sean Ellis, Meei-Li Huang, Keith R Jerome, Alexander Greninger |
| hCoV-19/Belgium/AZDelta-15561-2117R/2021 | EPI_ISL_2036113 | 2021-05-01 | AZDelta | AZ Delta Medical Laboratories in Roeselare, Belgium | Geert Martens, Dieter De Smet, Merijn Vanhee, on behalf of AZ Delta COVID-19 Genomics core (member of Genomic surveillance of SARS-CoV-2 in Belgium network) |
| hCoV-19/Spain/CT-LabRefCat-798217/2021 | EPI_ISL_2036267 | 2021-04-30 | Laboratori de Referencia de Catalunya | Laboratori de Referencia de Catalunya | Ramirez A., Bellosillo B., Padilla E., Vilas A., Hernandez JJ., Canal M. |
| hCoV-19/France/PAC-IHU-5013-N1/2021 | EPI_ISL_2036317 | 2021-04-19 | MEPHI, Aix Marseille University | MEPHI, Aix Marseille University | Anthony LEVASSEUR |

| | | | | | |
|---|---|---|---|---|---|
| hCoV-19/USA/NY-PRL-2021_0506_00I06/2021 | EPI_ISL_2037360 | 2021-05-03 | Pandemic Response Lab - NYC | Pandemic Response Lab, R&D | Henry Lee, Michael Hammerling, Melissa Hopkins, Cybill del Castillo, Shinyoung Clair Kang, William Ward, Pradeep Bugga, Sol Rey, Dylan Law, Katharine Nelson, Haiping Hao, Jon Laurent |
| hCoV-19/USA/NC-CDC-ASC210074323/2021 | EPI_ISL_2040976 | 2021-04-18 | Aegis Sciences Corporation | Centers for Disease Control and Prevention Division of Viral Diseases, Pathogen Discovery | Dakota Howard, Dhwani Batra, Peter W. Cook, Kara Moser, Adrian Paskey, Jason Caravas, Benjamin Rambo-Martin, Shatavia Morrison, Christopher Gulvick, Scott Sammons, Yvette Unoarumhi, Darlene Wagner, Matthew Schmerer, Cyndi Clark, Patrick Campbell, Rob Case |
| hCoV-19/USA/CA-UCSC-531/2021 | EPI_ISL_2081239 | 2021-04-16 | UCSC Genomics Institute | UCSC Genomics Institute | Mark Akeson, Ikenna Anigbogu, Eric Beraut, Isabel Bjork, Molly Cassatt-Johnstone, Terren Chang, Russell Corbett-Detig, Namrita Dhillon, Maximilian Haeussler, David Haussler, Angie Hinrichs, Miten Jain, Joshua Kapp, A. Marm Kilpatrick, Jakob McBroome, Hugh |
| hCoV-19/Israel/CVL-8287/2021 | EPI_ISL_2083542 | 2021-02-26 | Israel Central Virology laboratory | Israel National Consortium for SARS-CoV-2 sequencing | Neta Zuckerman, Efrat Dahan Bucris, Michal Mandelboim, Dana Bar-Ilan, Oran Erster, Tzvia Mann, Omer Murik, David A. Zeevi, Assaf Rokney, Joseph Jaffe, Eva Nachum, Maya Davidovich Cohen, Ephraim Fass, Gal Zizelski Valenci, Mor Rubinstein, Efrat Rorman, Isr |
| hCoV-19/Israel/CVL-10533/2021 | EPI_ISL_2085164 | 2021-03-01 | Israel Central Virology laboratory | Israel National Consortium for SARS-CoV-2 sequencing | Neta Zuckerman, Efrat Dahan Bucris, Michal Mandelboim, Dana Bar-Ilan, Miranda Geva, Netanel Abu, Oran Erster, Efrat Glick-Saar, Omri Nayshool, Gideon Rechavi, Ella Mendelson, Orna Mor |
| hCoV-19/SouthAfrica/KRISP-K014748/2021 | EPI_ISL_2086227 | 2021-05-05 | ShipMed Inc | KRISP, KZN Research Innovation and Sequencing Platform | Giandhari J, Pillay S, Naidoo Y, Ramphal U, Lessells R, Emmanuel SJ, Tegally H, Wilkinson E, de Oliveira T |
| hCoV-19/Ecuador/USFQ-1167/2021 | EPI_ISL_2086715 | 2021-04-21 | Institute of Microbiology, Universidad San Francisco de Quito | Institute of Microbiology, Universidad San Francisco de Quito | Sully M√°rquez, Bel√©n Prado-Vivar, Juan Jos√© Guadalupe, Monica Becerra-Wong, Bernardo Guti√©rrez, Maria Nataly Velastegui Peralta, Ver√≥nica Barrag√°n, Patricio Rojas-Silva, Gabriel Trueba, Michelle Grunauer, Pa√∫l C√°rdenas |
| hCoV-19/SouthAfrica/NICD-N5070/2021 | EPI_ISL_2087000 | 2021-02-02 | AMPATH LABORATORIES | Diseases of the National Health Laboratory Service | Amoako DG, Scheepers C, Mohale T, Ntuli N, Mahlangu B, Ismail A, Bhiman JN |
| hCoV-19/USA/ID-IBL-726959/2021 | EPI_ISL_2090646 | 2021-05-04 | Idaho Bureau of Laboratories | Idaho Bureau of Laboratories | R. Beukelman, Matthew Charles Burns, Aimee Ceniseros, Robert L. Voermans, Christopher Ball |
| hCoV-19/Curacao/CW-RIVM-30433/2021 | EPI_ISL_2094396 | 2021-04-23 | Dutch COVID-19 response team | National Institute for Public Health and the Environment (RIVM) | Adam Meijer, Harry Vennema, Dirk Eggink, Jeroen Cremer, Sharon van den Brink, Bas van der Veer, AnneMarie van den Brandt, Lisa Wijsman, Kim Freriks, Ryanne Jaarsma, Eun√≠ce Then, Lynn Aarts, Sanne Bos, Melissa van Tuil, Linda van de Nes, Sjoerd Kuiling, J |
| hCoV-19/Germany/BY-MVP-000004245/2021 | EPI_ISL_2095286 | 2021-03-21 | Max von Pettenkofer Institute, Virology, National Reference Center for Retroviruses, LMU Munich | Laboratory for Functional Genome Analysis; Dept. Genomics; Gene Center of the LMU Munich | Max Muenchhoff; Stefan Krebs; Alexander Graf; Oliver Keppler; Helmut Blum |

| | | | | | |
|---|---|---|---|---|---|
| hCoV-19/Russia/MOW-RCOV_069/2021 | EPI_ISL_2096201 | 2021-01-10 | FEDERAL STATE BUDGETARY INSTITUTION OF HEALTHCARE HOSPITAL OF THE RUSSIAN ACADEMY OF SCIENCES (TROITSK), Academic Consortium | FEDERAL RESEARCH AND CLINICAL CENTER OF PHYSICAL-CHEMICAL MEDICINE OF FEDERAL MEDICAL BIOLOGICAL AGENCY, Academic Consortium | Vladislav Babenko, Ramiz Bakhtyev, Vladimir Baklaushev, Larisa Balykova, Pavel Bashkirov, Anna Blagonravova, Daria Boldyreva, Dmitry Fedorov, Ilshat Gafurov, Raushaniya Gaifullina, Yulia Galeeva, Elena Galova, Vadim Govorun, Elena Ilina, Konstantin Ivanov |
| hCoV-19/Israel/HMO-VL-2429879/2020 | EPI_ISL_2096775 | 2020-09-16 | Hadassah Medical Center Clinical Virology Laboratory, Hadassah Ein Kerem | Hadassah Hebrew University Viral Sequencing Group, Hadassah Hebrew University Medical Center | Hadar Golan Berman, Esther Oiknine-Djian, Mila Rivkin, Sheera Adar, Dana G. Wolf |
| hCoV-19/Israel/HMO-VL-2497871/2020 | EPI_ISL_2096776 | 2020-10-13 | Hadassah Medical Center Clinical Virology Laboratory, Hadassah Ein Kerem | Hadassah Hebrew University Viral Sequencing Group, Hadassah Hebrew University Medical Center | Hadar Golan Berman, Esther Oiknine-Djian, Mila Rivkin, Sheera Adar, Dana G. Wolf |
| hCoV-19/Israel/HMO-VL-2681855/2021 | EPI_ISL_2096948 | 2021-04-19 | Hadassah Medical Center Clinical Virology Laboratory, Hadassah Ein Kerem | Hadassah Hebrew University Viral Sequencing Group, Hadassah Hebrew University Medical Center | Hadar Golan Berman, Esther Oiknine-Djian, Mila Rivkin, Sheera Adar, Dana G. Wolf |
| hCoV-19/Brazil/SP-LNN00646/2021 | EPI_ISL_2101402 | 2021-04-21 | Laboratorio Central Noel Nutels | Bioinformatics Laboratory / LNCC | Luiz G P de Almeida, Alessandra P Lamarca, Ronaldo da Silva F Jr, Liliane Cavalcante, Alexandra L Gerber, Ana Paula de C Guimaraes, Douglas Terra Machado, Cassia Alves, Diana Mariani, Cintia Policarpo, Gleidson da Silva de Oliveira, Mario Sergio Ribeiro, |
| hCoV-19/NewZealand/21MV0414/2021 | EPI_ISL_2103201 | 2021-05-06 | LabPLUS | Institute of Environmental Science and Research (ESR) | Rachel Boyle, SallyAnn Harbison, Olivia Stroeven, Xiaoyun Ren, Matt Storey, Nikki Freed, Muhammad Faisal, Jing Wang, Hermes Perez, Anja Werno, Antje van der Linden, Arlo Upton, Chris Mansell, David Hammer, Dragana Drinkovic, Gary McAuliffe, Hana Sofia And |
| hCoV-19/India/MH-NCCS-ND16364/2021 | EPI_ISL_2107017 | 2021-03-21 | National Centre For Cell Science ,Äì INSACOG | National Centre For Cell Science | Dhiraj Paul, Shivang P. Bhanushali, Mohak P Gujare, Mitali Inamdar, Manoj Kumar Bhat, Ajay Pillai, INSACOG Consortium team, Yogesh Shouche |
| hCoV-19/India/MH-NCCS-ND16322/2021 | EPI_ISL_2107022 | 2021-03-21 | National Centre For Cell Science ,Äì INSACOG | National Centre For Cell Science | Dhiraj Paul, Shivang P. Bhanushali, Mohak P Gujare, Mitali Inamdar, Manoj Kumar Bhat, Ajay Pillai, INSACOG Consortium team, Yogesh Shouche |
| hCoV-19/Brazil/SP-SJRP129949/2021 | EPI_ISL_2107297 | 2021-04-27 | Laboratorio de Pesquisa em Virologia, FAMERP, SJRP | Laboratorio de Pesquisa em Virologia, FAMERP, SJRP | Cecv≠lia Artico Banho; Lv≠via Sacchetto; Fv°bio Sossai Possebon; Leila Sabrina Ullmann; Cv≠ntia Bittar; Guilherme Campos; Helena Lage Ferreira; Jorge A. Petroli Marchesi; Maisa C. Pereira Parra; Marv≠lia Moraes; Paula Rahal; Paulo Inacio da Costa; Jov£o |
| hCoV-19/Australia/NSW-R0239/2021 | EPI_ISL_2107500 | 2021-04-30 | New South Wales Health Pathology Royal Prince Alfred Hospital | Microbiology RPAH | Foster, C.; Au, J.; Ruiz Silva, M.; Deveson, I.; Bull, R.; Van Hal, S.; Rawlinson, W. |
| hCoV-19/Australia/NSW-R0266/2021 | EPI_ISL_2107531 | 2021-05-10 | New South Wales Health Pathology Royal Prince Alfred Hospital | Microbiology RPAH | Foster, C.; Au, J.; Ruiz Silva, M.; Deveson, I.; Bull, R.; Van Hal, S.; Rawlinson, W. |
| hCoV-19/Australia/NSW-R0270/2021 | EPI_ISL_2107535 | 2021-05-12 | New South Wales Health Pathology Royal Prince Alfred Hospital | Microbiology RPAH | Foster, C.; Au, J.; Ruiz Silva, M.; Deveson, I.; Bull, R.; Van Hal, S.; Rawlinson, W. |
| hCoV-19/USA/CA-CDC-FG-038608/2021 | EPI_ISL_2785951 | 2021-06-24 | Fulgent Genetics | Centers for Disease Control and Prevention Division of Viral Diseases, Pathogen Discovery | Dakota Howard, Dhwani Batra, Peter W. Cook, Kara Moser, Adrian Paskey, Jason Caravas, Benjamin Rambo-Martin, Shatavia Morrison, Christopher Gulvick, Scott Sammons, Yvette Unoarumhi, Darlene Wagner, Matthew Schmerer, Harry Gao, Mickey Li, John Gao, Joseph |

| | | | | | |
|---|---|---|---|---|---|
| hCoV-19/England/MILK-156362F/2021 | EPI_ISL_2121856 | 2021-05-08 | Lighthouse Lab in Milton Keynes | Wellcome Sanger Institute for the COVID-19 Genomics UK (COG-UK) Consortium | The Lighthouse Lab in Milton Keynes and Alex Alderton, Roberto Amato, Jeffrey Barrett, Sonia Goncalves, Ewan Harrison, David K. Jackson, Ian Johnston, Dominic Kwiatkowski, Cordelia Langford, John Sillitoe on behalf of the Wellcome Sanger Institute COVID-1 |
| hCoV-19/USA/NC-CDC-MMB09188222/2021 | EPI_ISL_2786546 | 2021-06-17 | Mako Medical | Centers for Disease Control and Prevention Division of Viral Diseases, Pathogen Discovery | Dakota Howard, Dhwani Batra, Peter W. Cook, Kara Moser, Adrian Paskey, Jason Caravas, Benjamin Rambo-Martin, Shatavia Morrison, Christopher Gulvick, Scott Sammons, Yvette Unoarumhi, Darlene Wagner, Matthew Schmerer,Matthew Tugwell, Lauren Moon, Clinton R. |
| hCoV-19/Ireland/MH-NVRL-22IRL57999/2021 | EPI_ISL_2132109 | 2021-04-30 | National Virus Reference Laboratory | National Virus Reference Laboratory | Zoe Yandle, Charlene Bennett, Gabriel Gonzalez, Michael Carr, Jonathan Dean, Cillian F De Gascun |
| hCoV-19/England/MILK-157AF48/2021 | EPI_ISL_2138009 | 2021-05-10 | Lighthouse Lab in Milton Keynes | Wellcome Sanger Institute for the COVID-19 Genomics UK (COG-UK) Consortium | The Lighthouse Lab in Milton Keynes and Alex Alderton, Roberto Amato, Jeffrey Barrett, Sonia Goncalves, Ewan Harrison, David K. Jackson, Ian Johnston, Dominic Kwiatkowski, Cordelia Langford, John Sillitoe on behalf of the Wellcome Sanger Institute COVID-1 |
| hCoV-19/England/ALDP-15786DC/2021 | EPI_ISL_2138129 | 2021-05-09 | Lighthouse Lab in Alderley Park | Wellcome Sanger Institute for the COVID-19 Genomics UK (COG-UK) Consortium | Jacquelyn Wynn, Mairead Hyland, The Lighthouse Lab in Alderley Park and Alex Alderton, Roberto Amato, Jeffrey Barrett, Sonia Goncalves, Ewan Harrison, David K. Jackson, Ian Johnston, Dominic Kwiatkowski, Cordelia Langford, John Sillitoe on behalf of the W |
| hCoV-19/England/MILK-1579204/2021 | EPI_ISL_2138278 | 2021-05-11 | Lighthouse Lab in Milton Keynes | Wellcome Sanger Institute for the COVID-19 Genomics UK (COG-UK) Consortium | The Lighthouse Lab in Milton Keynes and Alex Alderton, Roberto Amato, Jeffrey Barrett, Sonia Goncalves, Ewan Harrison, David K. Jackson, Ian Johnston, Dominic Kwiatkowski, Cordelia Langford, John Sillitoe on behalf of the Wellcome Sanger Institute COVID-1 |
| hCoV-19/USA/IN-CDC-STM-000045992/2021 | EPI_ISL_1615349 | 2021-03-29 | Helix/Illumina | Centers for Disease Control and Prevention Division of Viral Diseases, Pathogen Discovery | Dakota Howard, Dhwani Batra, Peter W. Cook, Kara Moser, Adrian Paskey, Jason Caravas, Benjamin Rambo-Martin, Shatavia Morrison, Christopher Gulvick, Scott Sammons, Yvette Unoarumhi, Darlene Wagner, Matthew Schmerer, Eileen de Feo, Jan Antico, Christine Tr |
| hCoV-19/USA/IN-CDC-STM-000064674/2021 | EPI_ISL_2144594 | 2021-04-27 | Helix/Illumina | Centers for Disease Control and Prevention Division of Viral Diseases, Pathogen Discovery | Dakota Howard, Dhwani Batra, Peter W. Cook, Kara Moser, Adrian Paskey, Jason Caravas, Benjamin Rambo-Martin, Shatavia Morrison, Christopher Gulvick, Scott Sammons, Yvette Unoarumhi, Darlene Wagner, Matthew Schmerer, Eileen de Feo, Jan Antico, Christine Tr |
| hCoV-19/USA/IL-CDC-ASC210031444/2021 | EPI_ISL_2149545 | 2021-04-23 | Aegis Sciences Corporation | Centers for Disease Control and Prevention Division of Viral Diseases, Pathogen Discovery | Dakota Howard, Dhwani Batra, Peter W. Cook, Kara Moser, Adrian Paskey, Jason Caravas, Benjamin Rambo-Martin, Shatavia Morrison, Christopher Gulvick, Scott Sammons, Yvette Unoarumhi, Darlene Wagner, Matthew Schmerer, Cyndi Clark, Patrick Campbell, Rob Case |

| | | | | | |
|---|---|---|---|---|---|
| hCoV-19/USA/IL-CDC-ASC210031445/2021 | EPI_ISL_2149546 | 2021-04-23 | Aegis Sciences Corporation | Centers for Disease Control and Prevention Division of Viral Diseases, Pathogen Discovery | Dakota Howard, Dhwani Batra, Peter W. Cook, Kara Moser, Adrian Paskey, Jason Caravas, Benjamin Rambo-Martin, Shatavia Morrison, Christopher Gulvick, Scott Sammons, Yvette Unoarumhi, Darlene Wagner, Matthew Schmerer, Cyndi Clark, Patrick Campbell, Rob Case |
| hCoV-19/India/MH-1122500047002/2021 | EPI_ISL_2151097 | 2021-03-27 | INSACOG-Sikkim | National Institute of Biomedical Genomics ,Äì INSACOG | Arindam Maitra, Tara D Sharma, Nidhan Kumar Biswas, Kaden Zangmu Bhutia, Sreedhar Chinnaswamy Tshering Ongmu Bhutia, Saumitra Das |
| hCoV-19/Philippines/PH-PGC-00128/2020 | EPI_ISL_2153916 | 2020-12-09 | Philippine Genome Center - Biobank (NCR) | Philippine Genome Center | Francis A. Tablizo, Kenneth M. Kim, Carlo M. Lapid, Marc Jerrone R. Castro, Maria Sofia L. Yangzon, Benedict A. Maralit, Marc Edsel C. Ayes, Eva Maria Cutiongco-de la Paz, Alethea R. de Guzman, Jan Michael C. Yap, Jo-Hannah S. Llames, Sheila Mae M. Araiza |
| hCoV-19/Brazil/SP-SJRP326/2021 | EPI_ISL_2154956 | 2021-03-25 | Laboratorio de Pesquisa em Virologia, FAMERP, SJRP | Laboratorio de Pesquisa em Virologia, FAMERP, SJRP | Cecv≠lia Artico Banho; Lv≠via Sacchetto; Fv°bio Sossai Possebon; Leila Sabrina Ullmann; Cv≠ntia Bittar; Guilherme Campos; Helena Lage Ferreira; Jorge A. Petroli Marchesi; Maisa C. Pereira Parra; Marv≠lia Moraes; Paula Rahal; Paulo Inacio da Costa; Jov£o |
| hCoV-19/Philippines/PH-PGC-03130/2020 | EPI_ISL_2155074 | 2020-11-09 | Research Institute for Tropical Medicine, Inc. (RITM) | Philippine Genome Center | Francis A. Tablizo, Kenneth M. Kim, Carlo M. Lapid, Marc Jerrone R. Castro, Maria Sofia L. Yangzon, Benedict A. Maralit, Marc Edsel C. Ayes, Eva Maria Cutiongco-de la Paz, Alethea R. de Guzman, Jan Michael C. Yap, Jo-Hannah S. Llames, Sheila Mae M. Araiza |
| hCoV-19/Philippines/PH-PGC-04672/2021 | EPI_ISL_2155090 | 2021-01-27 | Amosup Seamen's Hospital | Philippine Genome Center | Francis A. Tablizo, Kenneth M. Kim, Carlo M. Lapid, Marc Jerrone R. Castro, Maria Sofia L. Yangzon, Benedict A. Maralit, Marc Edsel C. Ayes, Eva Maria Cutiongco-de la Paz, Alethea R. de Guzman, Jan Michael C. Yap, Jo-Hannah S. Llames, Sheila Mae M. Araiza |
| hCoV-19/Philippines/PH-PGC-07832/2021 | EPI_ISL_2156258 | 2021-02-24 | THE LORD'S GRACE MEDICAL AND INDUSTRIAL CLINIC | Philippine Genome Center | Francis A. Tablizo, Kenneth M. Kim, Carlo M. Lapid, Marc Jerrone R. Castro, Maria Sofia L. Yangzon, Elcid Aaron R. Pangilinan, Benedict A. Maralit, Marc Edsel C. Ayes, Eva Maria Cutiongco-de la Paz, Alethea R. de Guzman, Jan Michael C. Yap, Jo-Hannah S. L |
| hCoV-19/India/TG-CCMB-CIA1532/2021 | EPI_ISL_2157154 | 2021-04-23 | CSIR-Centre for Cellular and Molecular Biology | CSIR-Centre for Cellular and Molecular Biology-INSACOG | Payel Mukherjee,Lamuk Zaveri,Tulasi Nagabandi, Ara Sreenivas,Shreekant Verma, Amareshwar Vodapalli ,Blessy B John,Viswagithe S L,B Himasri,Valli Nagalakshmi Undamatla,Sofia Banu,Archana Bharadwaj Siva,Sharath Chandra Thota,Karthik Bharadwaj Tallapaka,Rake |
| hCoV-19/Haiti/34050/2020 | EPI_ISL_2157361 | 2020-12-24 | Laboratoire National de Santv© Publique ,Äì LNSP (HAITI - LNSP) | Laboratory of Respiratory Viruses and Measles, Oswaldo Cruz Institute, FIOCRUZ | Paola Resende, Patrick Delly, Jaques Boncy, Ito Journel, Luciana Appolinario, Fernando Motta, Anna Carolina Paixao, Ana Carolina Mendonca, Alice Sampaio Rocha, Taina Venas, Elisa Cavalcante Pereira, Renata Serrano Lopes, Marilda Siqueira on behalf of the |
| hCoV-19/Turkey/HSGM-B11266/2021 | EPI_ISL_2158039 | 2021-04-28 | Ministry of Health Turkey | Ministry of Health Turkey | Fatma Bayrakdar, Yasemin Cosgun, Suleyman Yalcin, Gulay Korukluoglu |

| | | | | | |
|---|---|---|---|---|---|
| hCoV-19/Canada/NS-NML-48435/2021 | EPI_ISL_2162120 | 2021-04-19 | NS-QEII Health Sciences Centre | National Microbiology Laboratory (NML) | Anna Majer, Shari Tyson, Grace Seo, Philip Mabon, Elsie Grudeski, Rhiannon Huzarewich, Russell Mandes, Anneliese Landgraff, Jennifer Tanner, Natalie Knox, Morag Graham, Gary Van Domselaar, Todd Hatchette, Jason LeBlanc, Janice Pettipas, Dan Gaston, Nathal |
| hCoV-19/Canada/ABPHL-09430/2021 | EPI_ISL_2167539 | 2021-03-18 | Alberta Precision Labs (APL) | Public Health Agency of Canada (PHAC) National Microbiology Laboratory | Buss, E, Croxen M, Deo A, Dieu P, Gill K, Ferrato C, Khan F, Koleva P, Li V, Lloyd C, Lynch T, Ma R, Murphy S, Pabbaraju K, Shokoples S, Tipples G, Thayer J, Whitehouse M, Wong A, Yu C, Zelyas N |
| hCoV-19/England/ALDP-153526E/2021 | EPI_ISL_2172223 | 2021-04-23 | Lighthouse Lab in Alderley Park | Wellcome Sanger Institute for the COVID-19 Genomics UK (COG-UK) Consortium | Jacquelyn Wynn, Mairead Hyland, The Lighthouse Lab in Alderley Park and Alex Alderton, Roberto Amato, Jeffrey Barrett, Sonia Goncalves, Ewan Harrison, David K. Jackson, Ian Johnston, Dominic Kwiatkowski, Cordelia Langford, John Sillitoe on behalf of the W |
| hCoV-19/England/ALDP-1537FFC/2021 | EPI_ISL_2172328 | 2021-04-24 | Lighthouse Lab in Alderley Park | Wellcome Sanger Institute for the COVID-19 Genomics UK (COG-UK) Consortium | Jacquelyn Wynn, Mairead Hyland, The Lighthouse Lab in Alderley Park and Alex Alderton, Roberto Amato, Jeffrey Barrett, Sonia Goncalves, Ewan Harrison, David K. Jackson, Ian Johnston, Dominic Kwiatkowski, Cordelia Langford, John Sillitoe on behalf of the W |
| hCoV-19/Israel/CVL-13488/2021 | EPI_ISL_2183706 | 2021-05-06 | Israel Central Virology laboratory | Israel National Consortium for SARS-CoV-2 sequencing | Neta Zuckerman, Efrat Dahan Bucris, Michal Mandelboim, Dana Bar-Ilan, Miranda Geva, Netanel Abu, Oran Erster, Efrat Glick-Saar, Omri Nayshool, Gideon Rechavi, Ella Mendelson, Orna Mor |
| hCoV-19/USA/LA-CDC-LC0056679/2021 | EPI_ISL_2181668 | 2021-05-02 | Laboratory Corporation of America | Centers for Disease Control and Prevention Division of Viral Diseases, Pathogen Discovery | Dakota Howard, Dhwani Batra, Peter W. Cook, Kara Moser, Adrian Paskey, Jason Caravas, Benjamin Rambo-Martin, Shatavia Morrison, Christopher Gulvick, Scott Sammons, Yvette Unoarumhi, Darlene Wagner, Matthew Schmerer, Minoo Agarwal, Eyad Almasri, Debbie Bol |
| hCoV-19/Israel/CVL-12262/2021 | EPI_ISL_2182335 | 2021-04-08 | Israel Central Virology laboratory | Israel National Consortium for SARS-CoV-2 sequencing | Neta Zuckerman, Efrat Dahan Bucris, Michal Mandelboim, Dana Bar-Ilan, Miranda Geva, Netanel Abu, Oran Erster, Efrat Glick-Saar, Omri Nayshool, Gideon Rechavi, Ella Mendelson, Orna Mor |
| hCoV-19/Israel/CVL-12553/2021 | EPI_ISL_2182689 | 2021-04-12 | Israel Central Virology laboratory | Israel National Consortium for SARS-CoV-2 sequencing | Neta Zuckerman, Efrat Dahan Bucris, Michal Mandelboim, Dana Bar-Ilan, Miranda Geva, Netanel Abu, Oran Erster, Efrat Glick-Saar, Omri Nayshool, Gideon Rechavi, Ella Mendelson, Orna Mor |
| hCoV-19/Israel/CVL-13470/2021 | EPI_ISL_2183679 | 2021-05-06 | Israel Central Virology laboratory | Israel National Consortium for SARS-CoV-2 sequencing | Neta Zuckerman, Efrat Dahan Bucris, Michal Mandelboim, Dana Bar-Ilan, Miranda Geva, Netanel Abu, Oran Erster, Efrat Glick-Saar, Omri Nayshool, Gideon Rechavi, Ella Mendelson, Orna Mor |
| hCoV-19/USA/NV-CDC-LC0058808/2021 | EPI_ISL_2185019 | 2021-05-05 | Laboratory Corporation of America | Centers for Disease Control and Prevention Division of Viral Diseases, Pathogen Discovery | Dakota Howard, Dhwani Batra, Peter W. Cook, Kara Moser, Adrian Paskey, Jason Caravas, Benjamin Rambo-Martin, Shatavia Morrison, Christopher Gulvick, Scott Sammons, Yvette Unoarumhi, Darlene Wagner, Matthew Schmerer, Minoo Agarwal, Eyad Almasri, Debbie Bol |

| | | | | | |
|---|---|---|---|---|---|
| hCoV-19/Philippines/PH-PGC-16021/2021 | EPI_ISL_2189512 | 2021-03-20 | Pasig City Children's Hospital - Child's Hope | Philippine Genome Center | Francis A. Tablizo, Kenneth M. Kim, Maria Sofia L. Yangzon, Elcid Aaron R. Pangilinan, Renato Jacinto Q. Mantaring, Benedict A. Maralit, Marc Edsel C. Ayes, Eva Maria Cutiongco-de la Paz, Alethea R. de Guzman, Jan Michael C. Yap, Jo-Hannah S. Llames, Shei |
| hCoV-19/Belgium/UGent-6078/2021 | EPI_ISL_2192265 | 2021-04-27 | Lab voor klinische biologie | Lab voor klinische biologie | Marija Janevska, Hannelore Hamerlinck, Bruno Verhasselt |
| hCoV-19/Belgium/UGent-6242/2021 | EPI_ISL_2192404 | 2021-04-29 | Lab voor klinische biologie | Lab voor klinische biologie | Marija Janevska, Hannelore Hamerlinck, Bruno Verhasselt |
| hCoV-19/SouthAfrica/NICD-R01891/2021 | EPI_ISL_2193358 | 2021-05-06 | Ampath Laboratories | Diseases of the National Health Laboratory Service | Amoako DG, Scheepers C, Mohale T, Ntuli N, Mahlangu B, Viana R, Glass A, Marshall T, Gottberg A, Bhiman JN |
| hCoV-19/USA/NJ-NYGC-NJ-BioR-557-Ampliseq/2020 | EPI_ISL_2193497 | 2020-09-23 | Hackensack Medical Center | New York Genome Center | Michael Zody, Andre Corvelo, Dayna M. Oschwald, Samantha Fennessey, Tom Maniatis, Liang Chen, Jose Mediavilla, Marcus Cunningham, Kaelea Composto, Kar Chow, David Perlin, Barry Kreiswirth |
| hCoV-19/Scotland/QEUH-1583D42/2021 | EPI_ISL_2199010 | 2021-05-12 | Lighthouse Lab in Glasgow | Wellcome Sanger Institute for the COVID-19 Genomics UK (COG-UK) Consortium | Harper VanSteenhouse, Yumi Kasai, David Gray, Carol Clugston, Anna Dominiczak and Alex Alderton, Roberto Amato, Jeffrey Barrett, Sonia Goncalves, Ewan Harrison, David K. Jackson, Ian Johnston, Dominic Kwiatkowski, Cordelia Langford, John Sillitoe on behal |
| hCoV-19/USA/TX-HMH-MCoV-35567/2021 | EPI_ISL_2200411 | 2021-03-19 | Houston Methodist Hospital | Houston Methodist Hospital | Randall J. Olsen, Paul A. Christensen, S. Wesley Long, Sishir Subedi, Robert Olson, Marcus Nguyen, James J. Davis, Matthew Ojeda Saavedra, Prasanti Yerramilli, Layne Pruitt, Kristina Reppond, Madison N. Shyer, Jessica Cambric, Ryan Gadd, Ilya J. Finkels |
| hCoV-19/Sweden/20038566P/2021 | EPI_ISL_2209223 | 2021-04-24 | Swedish national genomic surveillance program of SARS-CoV-2 | The Public Health Agency of Sweden | Maximilian Riess, Maria Lind Karlberg, Alma Brolund, Swedish national genomic surveillance program of SARS-CoV-2 |
| hCoV-19/Netherlands/ZE-RIVM-32425/2021 | EPI_ISL_2218905 | 2021-04-25 | Dutch COVID-19 response team | National Institute for Public Health and the Environment (RIVM) | Adam Meijer, Harry Vennema, Dirk Eggink, Jeroen Cremer, Sharon van den Brink, Bas van der Veer, AnneMarie van den Brandt, Lisa Wijsman, Kim Freriks, Ryanne Jaarsma, Eun√≠ce Then, Lynn Aarts, Sanne Bos, Melissa van Tuil, Linda van de Nes, Sjoerd Kuiling, J |
| hCoV-19/USA/TX-HMH-MCoV-35192/2020 | EPI_ISL_2220194 | 2020-08-11 | Houston Methodist Hospital | Houston Methodist Hospital | Randall J. Olsen, Paul A. Christensen, S. Wesley Long, Sishir Subedi, Robert Olson, Marcus Nguyen, James J. Davis, Matthew Ojeda Saavedra, Prasanti Yerramilli, Layne Pruitt, Kristina Reppond, Madison N. Shyer, Jessica Cambric, Ryan Gadd, Ilya J. Finkels |
| hCoV-19/USA/TX-HMH-MCoV-42224/2021 | EPI_ISL_2221359 | 2021-04-13 | Houston Methodist Hospital | Houston Methodist Hospital | Randall J. Olsen, Paul A. Christensen, S. Wesley Long, Sishir Subedi, Robert Olson, Marcus Nguyen, James J. Davis, Matthew Ojeda Saavedra, Prasanti Yerramilli, Layne Pruitt, Kristina Reppond, Madison N. Shyer, Jessica Cambric, Ryan Gadd, Ilya J. Finkels |

| | | | | | |
|---|---|---|---|---|---|
| hCoV-19/USA/TX-HMH-MCoV-40515/2020 | EPI_ISL_2223698 | 2020-08-02 | Houston Methodist Hospital | Houston Methodist Hospital | Randall J. Olsen, Paul A. Christensen, S. Wesley Long, Sishir Subedi, Robert Olson, Marcus Nguyen, James J. Davis, Matthew Ojeda Saavedra, Prasanti Yerramilli, Layne Pruitt, Kristina Reppond, Madison N. Shyer, Jessica Cambric, Ryan Gadd, Ilya J. Finkels |
| hCoV-19/USA/GU-CDC-2-4356537/2021 | EPI_ISL_2229094 | 2021-04-22 | Guam Public Health Laboratory | Centers for Disease Control and Prevention Division of Viral Diseases, Pathogen Discovery | Mili Sheth, Sarah Nobles, Jasmine Padilla, Mark Burroughs, Shoshona Le, Katie Dillon, Peter Cook, Clinton R. Paden, Dhwani Batra, Krista Queen, Kristen Knipe, Dakota Howard, Yvette Unoarumhi, Darlene Wagner, Matthew Schmerer, Ben L. Rambo-Martin, Kristine |
| hCoV-19/USA/GU-CDC-2-4356532/2021 | EPI_ISL_2229105 | 2021-04-26 | Guam Public Health Laboratory | Centers for Disease Control and Prevention Division of Viral Diseases, Pathogen Discovery | Mili Sheth, Sarah Nobles, Jasmine Padilla, Mark Burroughs, Shoshona Le, Katie Dillon, Peter Cook, Clinton R. Paden, Dhwani Batra, Krista Queen, Kristen Knipe, Dakota Howard, Yvette Unoarumhi, Darlene Wagner, Matthew Schmerer, Ben L. Rambo-Martin, Kristine |
| hCoV-19/USA/MA-MASPHL-03690/2021 | EPI_ISL_2229531 | 2021-04-30 | Massachusetts State Public Health Laboratory | Massachusetts State Public Health Laboratory | Andrew Lang, Timelia Fink, Glen Gallagher, Sandra Smole |
| hCoV-19/India/PB-90/2021 | EPI_ISL_2233544 | 2021-03-27 | National Centre for Disease Control | CDFD -INSACOG | Murali Bashyam, Asmita Gupta, Pratyusha Bala, Vinay Donipadi, Divya Vashisht, Ashwin Dalal |
| hCoV-19/England/QEUH-157313A/2021 | EPI_ISL_2238153 | 2021-05-08 | Lighthouse Lab in Glasgow | Wellcome Sanger Institute for the COVID-19 Genomics UK (COG-UK) Consortium | Harper VanSteenhouse, Yumi Kasai, David Gray, Carol Clugston, Anna Dominiczak and Alex Alderton, Roberto Amato, Jeffrey Barrett, Sonia Goncalves, Ewan Harrison, David K. Jackson, Ian Johnston, Dominic Kwiatkowski, Cordelia Langford, John Sillitoe on behal |
| hCoV-19/England/MILK-1565B5B/2021 | EPI_ISL_2238695 | 2021-04-28 | Lighthouse Lab in Milton Keynes | Wellcome Sanger Institute for the COVID-19 Genomics UK (COG-UK) Consortium | The Lighthouse Lab in Milton Keynes and Alex Alderton, Roberto Amato, Jeffrey Barrett, Sonia Goncalves, Ewan Harrison, David K. Jackson, Ian Johnston, Dominic Kwiatkowski, Cordelia Langford, John Sillitoe on behalf of the Wellcome Sanger Institute COVID-1 |
| hCoV-19/England/LIVE-E12498/2021 | EPI_ISL_2238872 | 2021-05-09 | Liverpool Clinical Laboratories | COVID-19 Genomics UK (COG-UK) Consortium | Sam Haldenby, Alistair Darby, Steve Paterson, Anita Lucaci, Julian Hiscox, M Almsaud, A Alrezaihi, Muhannad Alruwaili, Stuart D Armstrong, Jones Benjamin, Eleanor G Bentley, Anu Chawla, Jordan J Clark, Angela Cowell, Richard Eccles, Isabel Garc√≠a-Doriva |
| hCoV-19/England/LOND-12FDB88/2021 | EPI_ISL_2238961 | 2021-05-15 | University College London, Great Ormond Street Hospital for Children NHS Foundation Trust, Imperial College Healthcare NHS Trust | COVID-19 Genomics UK (COG-UK) Consortium | Sergi Castellano, Rachel Williams, Mark Kristiansen, Paola Resende Silva, Sunando Roy, Tony Brooks, Helena Tutill, Paola Niola, Patricia Dyal, Charlotte Williams, Leysa Forrest, Yasmin Panchbhaya, Jacqueline Findlay, Samuel Weeks, Julianne Brown, Kathryn |
| hCoV-19/USA/OH-ODH-SC1056549/2021 | EPI_ISL_2240731 | 2021-05-03 | Ohio Department of Health Laboratory | Ohio Department of Health Laboratory | Holmes, Jennifer; Eric Brandt, Keoni Omura, Glen McGillivary, Caitlin McDonnell, Jade Mowery, Stephanie Mccracken, Tyler Payne, Kirtana Ramadugu, Erica Leasure, Brent Lee, Kelsey Florek, Heather Blankenship, Allison Black, Quanta Brown, and Tammy Banne |
| hCoV-19/Brazil/PB-BA1200-250026294/2020 | EPI_ISL_2241527 | 2020-11-16 | Laborat√≥rio Central de Sa√∫de P√∫blica da Paraiba | Laborat√≥rios de Sa√∫de P√∫blica (CGLAB/DAEVS/SVS/MS) | Vagner Fonseca, et al. |

| | | | | | |
|---|---|---|---|---|---|
| hCoV-19/Brazil/PB-BA1200-250034845/2020 | EPI_ISL_2241570 | 2020-11-26 | Laboratv≥rio Central de Sav∫de Pv∫blica da Paraiba | Laboratv≥rios de Sav∫de Pv∫blica (CGLAB/DAEVS/SVS/MS) | Vagner Fonseca, et al. |
| hCoV-19/Russia/SPE-RII-29769S/2021 | EPI_ISL_2245643 | 2021-03-19 | WHO National Influenza Centre Russian Federation | WHO National Influenza Centre Russian Federation | Andrey Komissarov, Artem Fadeev, Kseniya Komissarova, Oula Masour, Kirill Varchenko, Mikhail Bakaev, Tamila Musaeva, Maria Timofeeva, Veronika Eder, Maria Pisareva, Nikita Yolshin, Daria Danilenko, Ksenia Safina, Elena Nabieva, Georgii Bazykin, Dmitry Lio |
| hCoV-19/Guinea/Conakry_IPG19296/2021 | EPI_ISL_2245913 | 2021-03-16 | Institut Pasteur de Guinv©e | Institut Pateur de Dakar | Grayo Solene, Diagne Moussa Mov∅se, Dia Ndongo, Diallo Amadou, Mbengue Safietou Sankhe, Ndiaye Ndack, Diop Mamadou, Maimouna Mbanne, Mohamed Kane, Loucoubar Cheikh, Tordo Noel, Faye Ousmane, Sall Amadou Alpha |
| hCoV-19/USA/WY-WYPHL20148013/2020 | EPI_ISL_2246812 | 2020-11-23 | Wyoming Public Health Laboratory | Wyoming Public Health Laboratory | Jim Mildenberger, Taylor Fearing, Channing Weber, Ashley Norberg, Chayse Rowley, Marley Goetz, Brian Dominguez, Elliot Thomasson, Brittany Oher, Sam Britz, Cari Sloma, and Rob Christensen |
| hCoV-19/Mexico/SIN-InDRE_FB15453_S2030/2021 | EPI_ISL_2246834 | 2021-05-13 | LESP Sinaloa | Instituto de Diagnostico y Referencia Epidemiologicos (INDRE) | Claudia Wong-Arambula, Abril Rodriguez-Maldonado, Vanessa Rivero-Arredondo, Ariadna Medina-Benitez, Joaquin Quiroz-Mercado, Sergio Rangel-Guerrero, Natividad Cruz-Ortiz, Tatiana Nunez-Garcia, Gisela Barrera-Badillo, Lucia Hernandez-Rivas, Irma Lopez-Marti |
| hCoV-19/England/MILK-1582745/2021 | EPI_ISL_2253799 | 2021-05-12 | Lighthouse Lab in Milton Keynes | Wellcome Sanger Institute for the COVID-19 Genomics UK (COG-UK) Consortium | The Lighthouse Lab in Milton Keynes and Alex Alderton, Roberto Amato, Jeffrey Barrett, Sonia Goncalves, Ewan Harrison, David K. Jackson, Ian Johnston, Dominic Kwiatkowski, Cordelia Langford, John Sillitoe on behalf of the Wellcome Sanger Institute COVID-1 |
| hCoV-19/Singapore/1716/2021 | EPI_ISL_2790362 | 2021-06-26 | National Public Health Laboratory, National Centre for Infectious Diseases | National Centre for Infectious Diseases | Tze Minn Mak, Katherine Ching, Zhenyang Zhou, Royce Ang, Lin Cui, Raymond Tzer Pin Lin |
| hCoV-19/USA/FL-CDC-QDX24713358/2021 | EPI_ISL_2268940 | 2021-05-07 | Quest Diagnostics Incorporated | Centers for Disease Control and Prevention Division of Viral Diseases, Pathogen Discovery | Dakota Howard, Dhwani Batra, Peter W. Cook, Kara Moser, Adrian Paskey, Jason Caravas, Benjamin Rambo-Martin, Shatavia Morrison, Christopher Gulvick, Scott Sammons, Yvette Unoarumhi, Darlene Wagner, Matthew Schmerer, S. H. Rosenthal, A. Gerasimova, R. M. K |
| hCoV-19/USA/AK-PHL9696/2021 | EPI_ISL_2790790 | 2021-06-16 | Alaska State Virology Laboratory | Alaska State Virology Laboratory | Stephanie DeRonde, Elva House, Jacob Zidek, Lisa Smith, Ph.D., Jack Chen, Ph.D. |
| hCoV-19/USA/KS-KHEL-1913/2020 | EPI_ISL_2271712 | 2020-05-20 | Kansas Health and Environmental Lab | Kansas Health and Environmental Lab | Mike Grose, Katherine Wiggins, Jonathan Barnell, Ben Olsen, and Phil Adam |
| hCoV-19/India/PY-Seq_978_S63_R1_001/2021 | EPI_ISL_2272917 | 2021-04-21 | JIPMER Puducherry | inStem NCBS - INSACOG | Uma Ramakrishnan Dasaradhi Palakodeti Aswin SaiNarain |
| hCoV-19/Australia/QLD1890/2021 | EPI_ISL_2274246 | 2021-05-20 | Queensland Health Forensic and Scientific Services | Queensland Health Forensic and Scientific Services | Son Nguyen |
| hCoV-19/Qatar/QA.QU_18.11.D8/2021 | EPI_ISL_2274345 | 2021-04-08 | Ministry of Public Health / Hamad Medical Corporation | Biomedical Research Center (BRC), Qatar University / Qatar Genome Project (QGP) | BRC: Fatiha M. Benslimane, Heba A. Al-Khatib, Oal Al-Jamal, Dana Al-Batesh, Hadi M. Yassine, Asmaa A. Al-Thani. MOPH and HMC: Abdullatif Al-Khal, Muna A. S. Al-Maslamani, Mashael A. Al-Bader, Hamda Alromaihi, Roberto Bertollini, Peter V. Coyle, Einas A. E |

| | | | | | |
|---|---|---|---|---|---|
| hCoV-19/Brazil/SP-2898/2021 | EPI_ISL_2274979 | 2021-05-22 | Aeroporto Internacional de Guarulhos | Instituto Adolfo Lutz, Interdiciplinary Procedures Center, Strategic Laboratory | Claudio Tavares Sacchi, Claudia Regina Gonvßalves, Erica Valessa Ramos Gomes, Karoline Rodrigues Campos, Caio Vinicius Dias Lopes, Leonardo Jose Tadeu de Araujo |
| hCoV-19/Scotland/QEUH-1596869/2021 | EPI_ISL_2275689 | 2021-05-17 | Lighthouse Lab in Glasgow | Wellcome Sanger Institute for the COVID-19 Genomics UK (COG-UK) Consortium | Harper VanSteenhouse, Yumi Kasai, David Gray, Carol Clugston, Anna Dominiczak and Alex Alderton, Roberto Amato, Jeffrey Barrett, Sonia Goncalves, Ewan Harrison, David K. Jackson, Ian Johnston, Dominic Kwiatkowski, Cordelia Langford, John Sillitoe on behal |
| hCoV-19/USA/IL-CDC-ASC210064165/2021 | EPI_ISL_2280386 | 2021-05-16 | Aegis Sciences Corporation | Centers for Disease Control and Prevention Division of Viral Diseases, Pathogen Discovery | Dakota Howard, Dhwani Batra, Peter W. Cook, Kara Moser, Adrian Paskey, Jason Caravas, Benjamin Rambo-Martin, Shatavia Morrison, Christopher Gulvick, Scott Sammons, Yvette Unoarumhi, Darlene Wagner, Matthew Schmerer, Cyndi Clark, Patrick Campbell, Rob Case |
| hCoV-19/USA/TX-CDC-ASC210063979/2021 | EPI_ISL_2280656 | 2021-05-15 | Aegis Sciences Corporation | Centers for Disease Control and Prevention Division of Viral Diseases, Pathogen Discovery | Dakota Howard, Dhwani Batra, Peter W. Cook, Kara Moser, Adrian Paskey, Jason Caravas, Benjamin Rambo-Martin, Shatavia Morrison, Christopher Gulvick, Scott Sammons, Yvette Unoarumhi, Darlene Wagner, Matthew Schmerer, Cyndi Clark, Patrick Campbell, Rob Case |
| hCoV-19/USA/MN-CDC-IBX459671412588/2021 | EPI_ISL_2282904 | 2021-05-07 | Infinity Biologix | Centers for Disease Control and Prevention Division of Viral Diseases, Pathogen Discovery | Dakota Howard, Dhwani Batra, Peter W. Cook, Kara Moser, Adrian Paskey, Jason Caravas, Benjamin Rambo-Martin, Shatavia Morrison, Christopher Gulvick, Scott Sammons, Yvette Unoarumhi, Darlene Wagner, Matthew Schmerer, Christian Bixby, Yihe Wang, Jonathan Sc |
| hCoV-19/Ireland/D-NVRL-t90IRL06995/2021 | EPI_ISL_2284347 | 2021-04-28 | National Virus Reference Laboratory | National Virus Reference Laboratory | Fiona Crispie, Calum Walsh, Matthew McCabe, Zoe Yandle, Charlene Bennet, Gabriel Gonzalez, Michael Carr, Jonathan Dean, Paul Cotter, Cillian F De Gascun |
| hCoV-19/Ireland/C-NVRL-t90IRL09938/2021 | EPI_ISL_2284375 | 2021-04-28 | National Virus Reference Laboratory | National Virus Reference Laboratory | Fiona Crispie, Calum Walsh, Matthew McCabe, Zoe Yandle, Charlene Bennet, Gabriel Gonzalez, Michael Carr, Jonathan Dean, Paul Cotter, Cillian F De Gascun |
| hCoV-19/Ghana/WACCBIP-TRA720/2021 | EPI_ISL_2285860 | 2021-04-20 | National Influenza Centre | National Influenza Centre | William K. Ampofo, Michael Marks, Ivy A. Asante, Sharon Hsu, Benjamiin B. Lindsey, Benjamin H. Foulkes, Mildred Adusei-Poku, Linda Boatemaa, Lorreta Kwasah, Joseph Oliver-Commey, Ernest Asiedu, Franklin Asiedu-Bekoe, Gordon Awandare, Joyce Ngoi, Dennis La |
| hCoV-19/France/PAC-IHU-3962_Nano1/2020 | EPI_ISL_2286796 | 2020-10-27 | MEPHI, Aix Marseille University | MEPHI, Aix Marseille University | Anthony LEVASSEUR |
| hCoV-19/France/PAC-IHU-3957_Nano1/2020 | EPI_ISL_2286836 | 2020-09-03 | MEPHI, Aix Marseille University | MEPHI, Aix Marseille University | Anthony LEVASSEUR |
| hCoV-19/USA/UT-UPHL-2105811114/2021 | EPI_ISL_2292612 | 2021-04-24 | Utah Public Health Laboratory | Utah Public Health Laboratory | Erin L. Young, Kelly F. Oakeson, Tara Gallagher |
| hCoV-19/FrenchGuiana/IPP12143/2021 | EPI_ISL_2293372 | 2021-04-24 | Outre mer | National Reference Center for Viruses of Respiratory Infections, Institut Pasteur, Paris | Marion Barbet, Sylvie Behillil, M√©line Bizard, Angela Brisebarre, Camille Capel, Vincent Enouf, Louise Lefran√ßois, Fr√©d√©ric Lemoine, Christophe Malabat, Corinne Maufrais, Etienne Simon-Lori√®re, Maud Vanpeene, Sylvie Van der Werf ,Dominique Rousset |

| | | | | | |
|---|---|---|---|---|---|
| hCoV-19/Canada/Qc-L00323965001/2021 | EPI_ISL_2293948 | 2021-02-11 | Laboratoire de sant√© publique du Qu√©bec | Laboratoire de sant√© publique du Qu√©bec | Sandrine Moreira, Ioannis Ragoussis, Guillaume Bourque, Jesse Shapiro, Mark Lathrop and Michel Roger on behalf of the CoVSeQ research group (http://covseq.ca/researchgroup) |
| hCoV-19/USA/IN-PU-LH000187551/2021 | EPI_ISL_2304444 | 2021-04-22 | Indiana Animal Disease Diagnostic Laboratory | Carpi Laboratory - Purdue University | Jack Dorman, Ilinca I Ciubotariu, Nicole M Perry, Jobin J Kattoor, Lev Gorenstein, Abebe A Fola, G Kenitra Hendrix, Rebecca P Wilkes, Giovanna Carpi |
| hCoV-19/USA/IN-CDC-LC0059876/2021 | EPI_ISL_2304847 | 2021-05-10 | Laboratory Corporation of America | Centers for Disease Control and Prevention Division of Viral Diseases, Pathogen Discovery | Dakota Howard, Dhwani Batra, Peter W. Cook, Kara Moser, Adrian Paskey, Jason Caravas, Benjamin Rambo-Martin, Shatavia Morrison, Christopher Gulvick, Scott Sammons, Yvette Unoarumhi, Darlene Wagner, Matthew Schmerer, Minoo Agarwal, Eyad Almasri, Debbie Bol |
| hCoV-19/USA/WA-CDC-LC0060245/2021 | EPI_ISL_2305588 | 2021-05-18 | Laboratory Corporation of America | Centers for Disease Control and Prevention Division of Viral Diseases, Pathogen Discovery | Dakota Howard, Dhwani Batra, Peter W. Cook, Kara Moser, Adrian Paskey, Jason Caravas, Benjamin Rambo-Martin, Shatavia Morrison, Christopher Gulvick, Scott Sammons, Yvette Unoarumhi, Darlene Wagner, Matthew Schmerer, Minoo Agarwal, Eyad Almasri, Debbie Bol |
| hCoV-19/Switzerland/BE-ETHZ-620326/2021 | EPI_ISL_2307575 | 2021-05-10 | Viollier AG | Department of Biosystems Science and Engineering, ETH Z√ºrich | Christian Beisel, Sarah Nadeau, Chaoran Chen, Ivan Topolsky, Philipp Jablonski, Lara Fuhrmann, David Dreifuss, Katharina Jahn, Rebecca Denes, Mirjam Feldkamp, Ina Nissen, Natascha Santacroce, Elodie Burcklen, Christiane Beckmann, Maurice Redondo, Olivier |
| hCoV-19/Germany/BE-RKI-I-158387/2021 | EPI_ISL_2313650 | 2021-05-14 | Labor 28 MVZ GmbH | Robert Koch Institute | unknown |
| hCoV-19/England/ALDP-15A11D9/2021 | EPI_ISL_2317576 | 2021-05-17 | Lighthouse Lab in Alderley Park | Wellcome Sanger Institute for the COVID-19 Genomics UK (COG-UK) Consortium | Jacquelyn Wynn, Mairead Hyland, The Lighthouse Lab in Alderley Park and Alex Alderton, Roberto Amato, Jeffrey Barrett, Sonia Goncalves, Ewan Harrison, David K. Jackson, Ian Johnston, Dominic Kwiatkowski, Cordelia Langford, John Sillitoe on behalf of the W |
| hCoV-19/Georgia/Tb-SNGS132/2021 | EPI_ISL_2319170 | 2021-05-20 | Department for Virology, Molecular Biology and Genome Research, R. G. Lugar Center for Public Health Research, National Center for Disease Control and Public Health (NCDC) of Georgia. | Biology and Genome Research, R. G. Lugar Center for Public Health Research, National Center for Disease Control and Public Health (NCDC) of Georgia. | Giorgi Tomashvili, Meri Pantsulaia, Gvantsa Brachveli, Giorgi Gogoladze, Nino Berishvili, Tata Imnadze, Ana Papkiauri, Gvantsa Chanturia, Ann Machablishvili, Nato Kotaria, Marine Murtskhvaladze, Lela Sabadze, Mari Gavashelidze, Tamar Jashiashvili, Tea Tev |
| hCoV-19/Japan/PG-38966/2021 | EPI_ISL_2327939 | 2021-03-24 | Pathogen Genomics Center, National Institute of Infectious Diseases | Pathogen Genomics Center, National Institute of Infectious Diseases | Tsuyoshi Sekizuka, Kentaro Itokawa, Rina Tanaka, Masanori Hashino, Makoto Kuroda |
| hCoV-19/Japan/PG-39130/2021 | EPI_ISL_2328103 | 2021-03-31 | Pathogen Genomics Center, National Institute of Infectious Diseases | Pathogen Genomics Center, National Institute of Infectious Diseases | Tsuyoshi Sekizuka, Kentaro Itokawa, Rina Tanaka, Masanori Hashino, Makoto Kuroda |
| hCoV-19/Japan/PG-40112/2021 | EPI_ISL_2329098 | 2021-04-19 | SARS-CoV-2 testing team, National Institute of Infectious Diseases | Pathogen Genomics Center, National Institute of Infectious Diseases | Tsuyoshi Sekizuka, Kentaro Itokawa, Rina Tanaka, Masanori Hashino, Nozomu Hanaoka, Masumichi Saito, Naomi Nojiri, Hazuka Y Furihata, Sana Uchikoba, Tsuguto Fujimoto, Makoto Kuroda |
| hCoV-19/Colombia/DC-C21/2020 | EPI_ISL_2339882 | 2020-10-10 | Centro de Estudio de Enfermedades Autoinmunes (CREA), Universidad del Rosario, Bogota, Colombia | Microbiolog√≠a y Biotecnolog√≠a-UR (CIMBIUR), Facultad de Ciencias Naturales, Universidad del Rosario, Bogot√°, Colombia | Juan-Manuel Anaya, Gustavo Salguero, Juan Esteban Gallo, Carolina Ram√≠rez-Santana, Sergio Castañeda, Nathalia Ballesteros, Marina Mu√±oz, Luz H. Pati√±o, Juan David Ram√≠rez |

| | | | | | |
|---|---|---|---|---|---|
| hCoV-19/Portugal/PT7634/2021 | EPI_ISL_2340336 | 2021-05-11 | Germano de Sousa | and BioSystems & Integrative Sciences Institute (BioISI) Genomics Unit, FCUL | Borges et al |
| hCoV-19/India/OR-ILSGS01219/2021 | EPI_ISL_2341666 | 2021-04-03 | Veer Surendra Sai Institute of Medical Sciences and Research, Burla, Sambalpur | Institute of Life Sciences - INSACOG | Sunil K. Raghav, Safal Walia, Arup Ghosh, Atimukta Jha, Amol M. Kanampalliwar, Omprakash Shiriwas, Sana Fatma, Shifu Aggarwal, Rupesh Dash, Rajeeb Swain, Punit Prasad, INSACOG Consortium, Ajay Parida |
| hCoV-19/India/CT-ILSGS01062/2021 | EPI_ISL_2341885 | 2021-03-17 | Pt. Jawahar Lal Nehru Memorial Medical College, Raipur | Institute of Life Sciences - INSACOG | Sunil K. Raghav, Safal Walia, Arup Ghosh, Atimukta Jha, Amol M. Kanampalliwar, Omprakash Shiriwas, Sana Fatma, Shifu Aggarwal, Rupesh Dash, Rajeeb Swain, Punit Prasad, INSACOG Consortium, Ajay Parida |
| hCoV-19/India/JH-ILSGS01744/2021 | EPI_ISL_2342653 | 2021-04-15 | MGM Medical College & Hospital, Jamshedpur | Institute of Life Sciences - INSACOG | Sunil K. Raghav, Safal Walia, Arup Ghosh, Atimukta Jha, Amol M. Kanampalliwar, Omprakash Shiriwas, Sana Fatma, Shifu Aggarwal, Rupesh Dash, Rajeeb Swain, Punit Prasad, INSACOG Consortium, Ajay Parida |
| hCoV-19/India/CT-ILSGS01442/2021 | EPI_ISL_2342880 | 2021-04-05 | BRLSABVM Government Medical College, Rajnandgaon, Chhattisgarh | Institute of Life Sciences - INSACOG | Sunil K. Raghav, Safal Walia, Arup Ghosh, Atimukta Jha, Amol M. Kanampalliwar, Omprakash Shiriwas, Sana Fatma, Shifu Aggarwal, Rupesh Dash, Rajeeb Swain, Punit Prasad, INSACOG Consortium, Ajay Parida |
| hCoV-19/USA/VA-DCLS-5125/2021 | EPI_ISL_2344473 | 2021-05-03 | Virginia Division of Consolidated Laboratory Services | Virginia Division of Consolidated Laboratory Services | Virginia DCLS |
| hCoV-19/England/MILK-15A9D64/2021 | EPI_ISL_2346736 | 2021-05-21 | Lighthouse Lab in Milton Keynes | Wellcome Sanger Institute for the COVID-19 Genomics UK (COG-UK) Consortium | The Lighthouse Lab in Milton Keynes and Alex Alderton, Roberto Amato, Jeffrey Barrett, Sonia Goncalves, Ewan Harrison, David K. Jackson, Ian Johnston, Dominic Kwiatkowski, Cordelia Langford, John Sillitoe on behalf of the Wellcome Sanger Institute COVID-1 |
| hCoV-19/Sweden/Orebro-48527430/2021 | EPI_ISL_2350915 | 2021-05-12 | Orebro University Hospital | Orebro University Hospital | Sundqvist M et al |
| hCoV-19/England/MILK-15B9806/2021 | EPI_ISL_2353445 | 2021-05-24 | Lighthouse Lab in Milton Keynes | Wellcome Sanger Institute for the COVID-19 Genomics UK (COG-UK) Consortium | The Lighthouse Lab in Milton Keynes and Alex Alderton, Roberto Amato, Jeffrey Barrett, Sonia Goncalves, Ewan Harrison, David K. Jackson, Ian Johnston, Dominic Kwiatkowski, Cordelia Langford, John Sillitoe on behalf of the Wellcome Sanger Institute COVID-1 |
| hCoV-19/England/GSTT-210201TCBC38/2021 | EPI_ISL_2355225 | 2021-01-29 | Centre for Clinical Infection and Diagnostics Research and Genomics Innovation Unit, Guy's and St. Thomas' NHS Trust | COVID-19 Genomics UK (COG-UK) Consortium | Chloe Fisher, Luke Snell, Penny Cliff, Rahul Batra, Jonathan Edgeworth, Ali Raza Awan |
| hCoV-19/SouthAfrica/VIDA-KRISP-K012794/2020 | EPI_ISL_2360855 | 2020-10-30 | Vaccines and Infectious Diseases Analytics Research Unit (VIDA) | KRISP, KZn Research Innovation and Sequencing Platform | Baillie Vicky, du Plessis Jeanine, Giandhari Jennifer, Pillay Sureshnee, Naidoo Yeshnee, Tegally Houriiyah, San James, Wilkinson Eduan, de Oliveira Tulio, Madhi Shabir |
| hCoV-19/Ecuador/USFQ-1335/2021 | EPI_ISL_2361459 | 2021-05-05 | Institute of Microbiology, Universidad San Francisco de Quito | Institute of Microbiology, Universidad San Francisco de Quito | Bel√©n Prado-Vivar, Sully M√°rquez, Juan Jos√© Guadalupe, Fernada Zurita, Monica Becerra-Wong, Bernardo Guti√©rrez, Miriam Ruiz, Hugo Hern√°ndez, Ver√≥nica Barrag√°n, Patricio Rojas-Silva, Gabriel Trueba, Michelle Grunauer, Pa√∫l C√°rdenas |
| hCoV-19/Ireland/C-NVRL-30IRL20722/2021 | EPI_ISL_2363198 | 2021-05-17 | National Virus Reference Laboratory | National Virus Reference Laboratory | Zoe Yandle, Charlene Bennett, Gabriel Gonzalez, Michael Carr, Jonathan Dean, Cillian F De Gascun |

| | | | | | |
|---|---|---|---|---|---|
| hCoV-19/Sweden/KUH-660/2020 | EPI_ISL_2365961 | 2020-11-08 | Karolinska University Hospital | Karolinska University Hospital | Jan Albert ,Tobias Allander ,Annelie Bjerkner ,Sandra Broddesson ,Robert Dyrdak ,Martin Ekman ,Lynda Eneh ,Lina Guerra Blomqvist ,Karolina Ininbergs ,Tanja Normark ,Isak Sylvin ,Zhibing Yun ,Martina Wahlund ,Valtteri Wirta |
| hCoV-19/Sweden/KUH-663/2020 | EPI_ISL_2365964 | 2020-11-12 | Karolinska University Hospital | Karolinska University Hospital | Jan Albert ,Tobias Allander ,Annelie Bjerkner ,Sandra Broddesson ,Robert Dyrdak ,Martin Ekman ,Lynda Eneh ,Lina Guerra Blomqvist ,Karolina Ininbergs ,Tanja Normark ,Isak Sylvin ,Zhibing Yun ,Martina Wahlund ,Valtteri Wirta |
| hCoV-19/USA/FL-CDC-ASC210065831/2021 | EPI_ISL_2370135 | 2021-05-21 | Aegis Sciences Corporation | Centers for Disease Control and Prevention Division of Viral Diseases, Pathogen Discovery | Dakota Howard, Dhwani Batra, Peter W. Cook, Kara Moser, Adrian Paskey, Jason Caravas, Benjamin Rambo-Martin, Shatavia Morrison, Christopher Gulvick, Scott Sammons, Yvette Unoarumhi, Darlene Wagner, Matthew Schmerer, Cyndi Clark, Patrick Campbell, Rob Case |
| hCoV-19/USA/WA-CDC-QDX24940788/2021 | EPI_ISL_2372241 | 2021-05-12 | Quest Diagnostics Incorporated | Centers for Disease Control and Prevention Division of Viral Diseases, Pathogen Discovery | Dakota Howard, Dhwani Batra, Peter W. Cook, Kara Moser, Adrian Paskey, Jason Caravas, Benjamin Rambo-Martin, Shatavia Morrison, Christopher Gulvick, Scott Sammons, Yvette Unoarumhi, Darlene Wagner, Matthew Schmerer, S. H. Rosenthal, A. Gerasimova, R. M. K |
| hCoV-19/Spain/CT-HUGTiPM017SA8G11/2021 | EPI_ISL_2372761 | 2021-05-18 | Microbiologia¬†CATLAB | Can Ruti SARS-CoV-2 Sequencing Hub (HUGTiP/IrsiCaixa/IGTP) | Marc Noguera-Julian, Pilar Armengol, Ignacio Blanco, Antoni E Bordoy, Francesc Catala-Moll, Pere-Joan Cardona, Maria Casadell√†, Cristina Casa√±, Gemma Clara, Bonaventura Clotet, Cristina Esteban, Montserrat Gim√©nez, Mercedes Guerrero, Anna Not, Roger Pa |
| hCoV-19/India/AP-CCMB-CIA2035/2021 | EPI_ISL_2373233 | 2021-05-06 | AP SSO | CSIR-Centre for Cellular and Molecular Biology-INSACOG | Ara Sreenivas,Shreekant Verma,Tulasi Nagabandi, Lamuk Zaveri, Amareshwar Vodapalli ,Blessy B John,Viswagithe S L,B Himasri,Valli Nagalakshmi Undamatla,Payel Mukherjee,Sofia Banu,Archana Bharadwaj Siva,Sharath Chandra Thota,Karthik Bharadwaj Tallapaka,Rake |
| hCoV-19/India/UP-CCMB-CIA1733/2021 | EPI_ISL_2373503 | 2021-04-25 | Banaras Hindu University | CSIR-Centre for Cellular and Molecular Biology-INSACOG | Royana Singh, Saurabh Singh, Jaya Chakraborty, Priyoneel Basu, Chetan Sahni, Deepa Devadas, Saumya Singh, Manpreet Kaur, Nitish Kumar Singh, Abhay Kumar Yadav, Ajay Kumar Yadav, Ashish, Shani Vishwakarma,Payel Mukherjee,Lamuk Zaveri,Tulasi Nagabandi, ,Ble |
| hCoV-19/Norway/12882/2021 | EPI_ISL_2374511 | 2021-05-19 | Furst Medical Laboratory | Norwegian Institute of Public Health, Department of Virology | Kathrine Stene-Johansen, Kamilla Heddeland Instefjord, Hilde Elshaug, Garcia Llorente Ignacio, Jon Br√•te, Line Victoria Moen, Engebretsen Serina Beate,Pedersen Benedikte Nevjen, Debech Nadia, Atiya R Ali,Marie Paulsen Madsen, Rasmus Riis Kopperud, Hilde |
| hCoV-19/Australia/VIC18582/2021 | EPI_ISL_2379277 | 2021-05-26 | Microbiological Diagnostic Unit - Public Health Laboratory (MDU-PHL) | MDU-PHL | Seemann T., Sait, M.L., Sherry, N.L. |
| hCoV-19/India/KA-NIMH-SEQ-919/2021 | EPI_ISL_2379379 | 2021-05-09 | PANTHRPALYA | INSACOG-KA, NIMHANS | Chitra Pattabiraman, Anson Kunjumon George,Darshan Sreenivas,Pramada Prasad, Ananthapadmanabha Kotambail, Chetan G K, Gautham Arunachal Udupi, Anita S Desai, V Ravi |

| | | | | | |
|---|---|---|---|---|---|
| hCoV-19/India/TN-C_134_S185_R1_001/2021 | EPI_ISL_2379496 | 2021-01-02 | Dr S Raju, Director of Public Heath and Preventive Medicine | inStem NCBS - INSACOG | Uma Ramakrishnan Dasaradhi Palakodeti Aswin SaiNarain |
| hCoV-19/Egypt/NRC-6746/2020 | EPI_ISL_2380070 | 2020-09-18 | Center of Scientific Excellence for Influenza Viruses (CSEIV), National Research Centre | Center of Scientific Excellence for Influenza Viruses (CSEIV), National Research Centre | Rabeh El-Shesheny, Ahmed E Kayed, Ahmed El-Taweel, Mokhtar Gomaa, Sara Mahmoud, Yassmin Moatasim, Omnia Kutkat, Mina Kamel, Noura M Abo Shama, Mohamed El Sayes, Mahmoud Shehata, Ahmed Mostafa, Ahmed Kandeil, Richard Webby, Ghazi Kayali, Mohamed Ahmed Ali |
| hCoV-19/SouthAfrica/NICD-N6625/2021 | EPI_ISL_2382251 | 2021-05-05 | CHRIS HANI BARAGWANATH LABORATORY | Diseases of the National Health Laboratory Service | Amoako DG, Scheepers C, Mohale T, Ntuli N, Mahlangu B, Ismail A, Bhiman JN |
| hCoV-19/India/GA-NCCS-NS2570/2021 | EPI_ISL_2382951 | 2021-05-14 | National Centre For Cell Science | National Centre For Cell Science-INSACOG | Dhiraj Paul, Shivang P. Bhanushali, Mohak P Gujare, Mitali Inamdar, Manoj Kumar Bhat, Ajay Pillai, INSACOG Consortium team, Yogesh Shouche |
| hCoV-19/Russia/SPE-RII-34902S/2021 | EPI_ISL_2385185 | 2021-05-19 | WHO National Influenza Centre Russian Federation | WHO National Influenza Centre Russian Federation | Andrey Komissarov, Artem Fadeev, Kseniya Komissarova, Oula Masour, Kirill Varchenko, Mikhail Bakaev, Tamila Musaeva, Maria Timofeeva, Veronika Eder, Maria Pisareva, Nikita Yolshin, Daria Danilenko, Maria Baturova, Alexey Masharsky, Ksenia Safina, Elena Na |
| hCoV-19/Sweden/20257199P/2021 | EPI_ISL_2793995 | 2021-05-17 | Swedish national genomic surveillance program of SARS-CoV-2 | The Public Health Agency of Sweden | Maximilian Riess, Maria Lind Karlberg, Alma Brolund, Swedish national genomic surveillance program of SARS-CoV-2 |
| hCoV-19/England/MILK-15DD6A8/2021 | EPI_ISL_2392616 | 2021-05-29 | Lighthouse Lab in Milton Keynes | Wellcome Sanger Institute for the COVID-19 Genomics UK (COG-UK) Consortium | The Lighthouse Lab in Milton Keynes and Alex Alderton, Roberto Amato, Jeffrey Barrett, Sonia Goncalves, Ewan Harrison, David K. Jackson, Ian Johnston, Dominic Kwiatkowski, Cordelia Langford, John Sillitoe on behalf of the Wellcome Sanger Institute COVID-1 |
| hCoV-19/Mozambique/INS-K011778/2020 | EPI_ISL_2396892 | 2020-08-25 | Instituto Nacional de Saude (INS), Mozambique | KRISP, KZN Research Innovation and Sequencing Platform | Nalia Ismael, Nadia Sitoe, Paulo Arnaldo, Nedio Mabunda, Giandhari J, Pillay S, Emmanuel S, Tegally H, Wilkinson E, de Oliveira T |
| hCoV-19/USA/NC-CDC-LC0063580/2021 | EPI_ISL_2398629 | 2021-05-13 | Laboratory Corporation of America | Centers for Disease Control and Prevention Division of Viral Diseases, Pathogen Discovery | Dakota Howard, Dhwani Batra, Peter W. Cook, Kara Moser, Adrian Paskey, Jason Caravas, Benjamin Rambo-Martin, Shatavia Morrison, Christopher Gulvick, Scott Sammons, Yvette Unoarumhi, Darlene Wagner, Matthew Schmerer, Minoo Agarwal, Eyad Almasri, Debbie Bol |
| hCoV-19/Turkey/HSGM-B2457/2021 | EPI_ISL_2403511 | 2021-03-30 | Ministry of Health Turkey | Ministry of Health Turkey | Fatma Bayrakdar, Yasemin Cosgun, Suleyman Yalcin, Gulay Korukluoglu |
| hCoV-19/Australia/NSW1575/2021 | EPI_ISL_2405111 | 2021-04-27 | Sydney South West Pathology Service (SSWPS) - Royal Prince Alfred Hospital - NSW Health Pathology | Clinical Pathology and Medical Research; Westmead Hospital; University of Sydney | CIDM-PH et al. |
| hCoV-19/Australia/VIC18613/2021 | EPI_ISL_2405357 | 2021-05-30 | Microbiological Diagnostic Unit - Public Health Laboratory (MDU-PHL) | MDU-PHL | Seemann T., Sait, M.L., Sherry, N.L. |
| hCoV-19/NewZealand/21MV0452/2021 | EPI_ISL_2406492 | 2021-05-20 | Middlemore Hospital | Institute of Environmental Science and Research (ESR) | Rachel Boyle, SallyAnn Harbison, Olivia Stroeven, Xiaoyun Ren, Matt Storey, Nikki Freed, Muhammad Faisal, Jing Wang, Hermes Perez, Anja Werno, Antje van der Linden, Arlo Upton, Chris Mansell, David Hammer, Dragana Drinkovic, Gary McAuliffe, Hana Sofia And |

| | | | | | |
|---|---|---|---|---|---|
| hCoV-19/Qatar/QA.QU_18.14.G2/2021 | EPI_ISL_2408378 | 2021-05-06 | Ministry of Public Health / Hamad Medical Corporation | Biomedical Research Center (BRC), Qatar University / Qatar Genome Project (QGP) | BRC: Fatiha M. Benslimane, Heba A. Al-Khatib, Oal Al-Jamal, Dana Al-Batesh, Hadi M. Yassine, Asmaa A. Al-Thani. MOPH and HMC: Abdullatif Al-Khal, Muna A. S. Al-Maslamani, Mashael A. Al-Bader, Hamda Alromaihi, Roberto Bertollini, Peter V. Coyle, Einas A. E |
| hCoV-19/Colombia/ANT-CWOHC-VG-SEC00042P/2021 | EPI_ISL_1629737 | 2021-01-20 | SURA | Universidad Nacional de Colombia - Laboratorio Gen√≥mico One Health | Karl A Ciuoderis, Andres F. Cardona-Rios, Daniel O. Maldonado-Perez, Maria Stella L√≥pez, Sim√≥n Villegas Vel√°squez, Laura Silvana Perez, Idabely Betancur Ortiz, Sandra Ines Cano, Eliana Patricia Calvo Tapiero, Diego A. √Ålvarez-D√≠az, Carlos Franco-Mu√± |
| hCoV-19/Sweden/37002052XXP/2021 | EPI_ISL_2795212 | 2021-06-09 | Swedish national genomic surveillance program of SARS-CoV-2 | The Public Health Agency of Sweden | Maximilian Riess, Maria Lind Karlberg, Alma Brolund, Swedish national genomic surveillance program of SARS-CoV-2 |
| hCoV-19/USA/NV-NSPHL-LV363309/2021 | EPI_ISL_2801494 | 2021-06-22 | Nevada State Public Health Laboratory | Nevada State Public Health Laboratory | Andrew Gorzalski, Mark Pandori |
| hCoV-19/SouthAfrica/NHLS-UCT-GP-6549/2021 | EPI_ISL_2802126 | 2021-05-24 | Villiersdorp Clinic wc VDP | NHLS/UCT | Arash Iranzadeh, Deelan Doolabh, Lynn Tyers, Bruna Galvao, Innocent Mudau, Marvin Hsiao, Gert Marais, Diana Hardie, Stephen Korsman, Carolyn Williamson |
| hCoV-19/SriLanka/CDR199/2021 | EPI_ISL_2803196 | 2021-06-11 | Centre for Dengue Research and AICBU, Department of Immunology and Molecular Medicine | Centre for Dengue Research and AICBU, Department of Immunology and Molecular Medicine | Chandima Jeewandara, Deshni Jayathilaka, Deshan Madhusanka, Diyanath Ranasinghe, Gathsaurie Neelika Malavige |
| hCoV-19/SriLanka/CDR193/2021 | EPI_ISL_2803271 | 2021-06-22 | Centre for Dengue Research and AICBU, Department of Immunology and Molecular Medicine | Centre for Dengue Research and AICBU, Department of Immunology and Molecular Medicine | Chandima Jeewandara, Deshni Jayathilaka, Deshan Madhusanka, Diyanath Ranasinghe, Gathsaurie Neelika Malavige |
| hCoV-19/SriLanka/CMC119426/2021 | EPI_ISL_2803280 | 2021-06-22 | Centre for Dengue Research and AICBU, Department of Immunology and Molecular Medicine | Centre for Dengue Research and AICBU, Department of Immunology and Molecular Medicine | Chandima Jeewandara, Deshni Jayathilaka, Deshan Madhusanka, Diyanath Ranasinghe, Gathsaurie Neelika Malavige |
| hCoV-19/Zambia/ZMB-CH14212/2021 | EPI_ISL_2803450 | 2021-05-02 | University of Zambia, School of Veterinary Medicine | UNZAVET and PATH | Mulenga Mwenda-Chimfwembe, Ngonda Saasa, Daniel Bridges, ZNPHI and ZGSC |
| hCoV-19/Zambia/ZMB-97824/2021 | EPI_ISL_2803664 | 2021-05-24 | University of Zambia, School of Veterinary Medicine | UNZAVET and PATH | Mulenga Mwenda-Chimfwembe, Ngonda Saasa, Daniel Bridges, ZNPHI and ZGSC |
| hCoV-19/Zambia/ZMB-98174/2021 | EPI_ISL_2803710 | 2021-05-27 | University of Zambia, School of Veterinary Medicine | UNZAVET and PATH | Mulenga Mwenda-Chimfwembe, Ngonda Saasa, Daniel Bridges, ZNPHI and ZGSC |
| hCoV-19/Nigeria/NCDC-FCT-1X/2021 | EPI_ISL_2803747 | 2021-03-19 | Everight Diagnostic Laboratories | National Reference Laboratory, Nigeria Centre for Disease Control | Dr Ndodo Nnaemeka, Olusola Anuoluwapo Akanbi, Chimaobi Chukwu, Dr Omoare Adesuyi, Grace Esebanmen, Anthony Ahumibe, Catherine Okoi, Nwando Mba, Dr Chikwe Ihekweazu |
| hCoV-19/Israel/CVL_S10-2/2020 | EPI_ISL_2811914 | 2020-05-03 | IDF medical corps | Central Virology Laboratory, Ministry of Health | Neta S. Zuckerman, Efrat Bucris, Oran Erster, Michal Mandelboim, Amos Adler, Saar Burstein, Noam Protter, Moran Shwartswort-Cohen, Ella Mendelson, Orna Mor |
| hCoV-19/NewZealand/21CH0033/2021 | EPI_ISL_2811951 | 2021-06-02 | Canterbury Health Laboratories | Institute of Environmental Science and Research (ESR) | Paula scholes, Susan Lin, Xiaoyun Ren, Matt Storey, Nikki Freed, Muhammad Faisal, Jing Wang, Hermes Perez, Anja Werno, Antje van der Linden, Arlo Upton, Chris Mansell, David Hammer, Dragana Drinkovic, Gary McAuliffe, Hana Sofia Andersson, James Ussher, Ji |

| | | | | | |
|---|---|---|---|---|---|
| hCoV-19/NewZealand/21MV0522/2021 | EPI_ISL_2811954 | 2021-06-19 | Middlemore Hospital | Institute of Environmental Science and Research (ESR) | Rachel Boyle, SallyAnn Harbison, Olivia Stroeven, Xiaoyun Ren, Matt Storey, Nikki Freed, Muhammad Faisal, Jing Wang, Hermes Perez, Anja Werno, Antje van der Linden, Arlo Upton, Chris Mansell, David Hammer, Dragana Drinkovic, Gary McAuliffe, Hana Sofia And |
| hCoV-19/Scotland/CVR7897/2021 | EPI_ISL_2815921 | 2021-06-17 | West of Scotland Specialist Virology Centre, NHSGGC / MRC-University of Glasgow Centre for Virus Research | COVID-19 Genomics UK (COG-UK) Consortium | Ana da Silva Filipe, Natasha Johnson, Kathy Smollett, Daniel Mair, Stephen Carmichael, Alice Broos, Lily Tong, Jenna Nichols, Kyriaki Nomikou; Sarah McDonald; Ioulia Tsatsani, Richard Orton, Joseph Hughes, Sreenu Vattipally, David L Robertson; Alasdair Ma |
| hCoV-19/England/PHEC-L308L5BB/2021 | EPI_ISL_2818085 | 2021-06-20 | Respiratory Virus Unit, Microbiology Services Colindale, Public Health England | COVID-19 Genomics UK (COG-UK) Consortium | PHE Covid Sequencing Team |
| hCoV-19/England/PORT-1E8121F/2021 | EPI_ISL_2819882 | 2021-06-23 | Centre for Enzyme Innovation, University of Portsmouth / Translational Research Laboratory, Portsmouth Hospitals NHS Trust / Hampshire Hospitals NHS Foundation Trust / Bournemouth University / University Hospitals Dorset NHS Foundation Trust / Brighton an | COVID-19 Genomics UK (COG-UK) Consortium | Samuel Robson, Angela Beckett, Salman Goudarzi, Christopher Fearn, Kate Cook, Katie Loveson, Sharon Glaysher, Scott Elliott, Kelly Bicknell, Sarah Wyllie, Allyson Lloyd, Robert Impey, Anoop Chauhan, Stephen Kidd, Nathan Moore, Nick Cortes, Claire Thomas, |
| hCoV-19/Botswana/R20B31_BHP_AAB70078/2021 | EPI_ISL_2820407 | 2021-06-15 | Kasane Primary Hospital Laboratory | Botswana Harvard HIV Reference Laboratory | Sikhulile Moyo, Wonderful T. Choga, Dorcas Maruapula, Agnes Karutwaeng, Rose Munyere, Keoratile Ntshambiwa, Thongbotho Mphoyakgosi, Boitumelo Zuze, Botshelo Radibe, Legodile Kooepile, Ontlametse T. Bareng, Roger Shapiro, Shahin Lockman, Joseph Makhema, Ma |
| hCoV-19/Botswana/R20B33_BHP_AAB70080/2021 | EPI_ISL_2820409 | 2021-06-17 | Kasane Primary Hospital Laboratory | Botswana Harvard HIV Reference Laboratory | Sikhulile Moyo, Wonderful T. Choga, Dorcas Maruapula, Agnes Karutwaeng, Rose Munyere, Keoratile Ntshambiwa, Thongbotho Mphoyakgosi, Boitumelo Zuze, Botshelo Radibe, Legodile Kooepile, Ontlametse T. Bareng, Roger Shapiro, Shahin Lockman, Joseph Makhema, Ma |
| hCoV-19/Gambia/36175/2021 | EPI_ISL_2820697 | 2021-05-17 | MRCG at LSHTM Genomics lab | MRCG at LSHTM Genomics lab | Abdul Karim sesay, Abdoulie Kanteh, Jarra Manneh, Mariama Kujabi, Bakary Sanyang,Dabiri Damilari |
| hCoV-19/Italy/APU-IZSPB-1525PT/2021 | EPI_ISL_2820733 | 2021-06-28 | Presidio di Brindisi Di Summa - Perrino | Istituto Zooprofilattico Sperimentale della Puglia e della Basilicata | Parisi A., Bianco A., Capozzi L., Del Sambro L., Simone D., Difato L., Santoro A., Ridolfi D., Giannico A., Manzari C., Pesole G. |
| hCoV-19/Djibouti/CNSS00206-IHU1031243458/2020 | EPI_ISL_2820757 | 2020-05-31 | Laboratoire de Diagnostic, Centre de Soins 1, Caisse Nationale de S√©curit√© Sociale (CNSS) de Djibouti. | IHU M√©diterran√©e Infection. | Ikram Omar Osman; Anthony Levasseur; Ludivine Brechar; Iman Abdillahi Hassan; Idil Salah abdillahi; Zeinab Ali Waberi; Jeremy Delerce; Linda Houhamdi; Pierre-Edouard Fournier; Philippe Colson; Mohamed Houmed Aboubaker; Didier Raoult; Christian A. Devaux. |
| hCoV-19/Ireland/LK-NVRL-tQ23IRL92963/2021 | EPI_ISL_2821224 | 2021-06-17 | National Virus Reference Laboratory | National Virus Reference Laboratory | Fiona Crispie, Calum Walsh, Matthew McCabe, Zoe Yandle, Charlene Bennet, Gabriel Gonzalez, Michael Carr, Jonathan Dean, Paul Cotter, Cillian F De Gascun |

| | | | | | |
|---|---|---|---|---|---|
| hCoV-19/Canada/QC-1nMCY-S9101798/2021 | EPI_ISL_2822125 | 2021-01-10 | Laboratoire de santV© publique du QuV©bec | Laboratoire de santV© publique du QuV©bec | Sandrine Moreira, Ioannis Ragoussis, Guillaume Bourque, Jesse Shapiro, Mark Lathrop and Michel Roger on behalf of the CoVSeQ research group (http://covseq.ca/researchgroup) |
| hCoV-19/USA/CA-CDPH-2000044784/2021 | EPI_ISL_2825796 | 2021-03-30 | CDPH VBL | California Department of Public Health | CDPH-COVIDNet,UCLA Technology Center for Genomics & Bioinformatics |
| hCoV-19/USA/CA-CDPH-2000045602/2021 | EPI_ISL_2825904 | 2021-04-20 | CDPH VBL | California Department of Public Health | CDPH-COVIDNet,UCLA Technology Center for Genomics & Bioinformatics |
| hCoV-19/Australia/WA707/2021 | EPI_ISL_1817559 | 2021-04-28 | PathWest Laboratory Medicine WA | PathWest Laboratory Medicine WA Microbial Surveillance Unit | PathWest Laboratory Medicine WA Microbial Surveillance Unit |
| hCoV-19/Australia/WA708/2021 | EPI_ISL_1817560 | 2021-04-18 | PathWest Laboratory Medicine WA | PathWest Laboratory Medicine WA Microbial Surveillance Unit | PathWest Laboratory Medicine WA Microbial Surveillance Unit |
| hCoV-19/India/TG-CCMB-CIA5136/2021 | EPI_ISL_2774705 | 2021-06-10 | Mapmygenome | CSIR-Centre for Cellular and Molecular Biology-INSACOG | Shreekant Verma, Amareshwar Vodapalli ,Ara Sreenivas,B Himasri,Valli Nagalakshmi Undamatla,Payel Mukherjee, Lamuk Zaveri, Onkar Kulkarni,Tulasi Nagabandi, Sofia Banu, Jandhyala Sai Krishna,Sumedha Avadhanula, Priya Nurkurthy, Vidhyadhari Methuku, Archana |
| hCoV-19/India/TG-CCMB-CIA4901/2021 | EPI_ISL_2774991 | 2021-06-04 | CSIR-Centre for Cellular and Molecular Biology | CSIR-Centre for Cellular and Molecular Biology-INSACOG | B Himasri,Amareshwar Vodapalli,Ara Sreenivas,Shreekant Verma,Valli Nagalakshmi Undamatla,Payel Mukherjee, Lamuk Zaveri, Onkar Kulkarni,Tulasi Nagabandi, Sofia Banu, Vidhyadhari Methuku,Priya Nurkurthy, Jandhyala Sai Krishna,Sumedha Avadhanula, Archana Bha |
| hCoV-19/England/MILK-14F618B/2021 | EPI_ISL_1634517 | 2021-04-11 | Lighthouse Lab in Milton Keynes | Wellcome Sanger Institute for the COVID-19 Genomics UK (COG-UK) Consortium | The Lighthouse Lab in Milton Keynes and Alex Alderton, Roberto Amato, Jeffrey Barrett, Sonia Goncalves, Ewan Harrison, David K. Jackson, Ian Johnston, Dominic Kwiatkowski, Cordelia Langford, John Sillitoe on behalf of the Wellcome Sanger Institute COVID-1 |
| hCoV-19/England/LIVE-A0873/2020 | EPI_ISL_1636748 | 2020-06-17 | Liverpool Clinical Laboratories | COVID-19 Genomics UK (COG-UK) Consortium | Sam Haldenby, Alistair Darby, Steve Paterson, Anita Lucaci, Julian Hiscox, M Almsaud, A Alrezaihi, Muhannad Alruwaili, Stuart D Armstrong, Jones Benjamin, Eleanor G Bentley, Anu Chawla, Jordan J Clark, Angela Cowell, Richard Eccles, Isabel GarcV≠a-Doriva |
| hCoV-19/USA/DE-Curative-024494/2021 | EPI_ISL_2784203 | 2021-06-07 | Curative Labs | Curative Labs | Mikhail Hanewich-Hollatz, J.Cesar Ignacio-Espinoza, Elias L. Salfati, Nabjot Sandhu, George Way, Zhiyi Xie, Eugenia Khorosheva, Sophia Quasem, Janet Chen, Vladimir Slepnev |
| hCoV-19/Australia/WA736/2021 | EPI_ISL_2784344 | 2021-06-29 | PathWest Laboratory Medicine WA | PathWest Laboratory Medicine WA Microbial Surveillance Unit | PathWest Laboratory Medicine WA Microbial Surveillance Unit |
| hCoV-19/Turkey/HSGM-B11741/2021 | EPI_ISL_2403326 | 2021-05-04 | Ministry of Health Turkey | Ministry of Health Turkey | Fatma Bayrakdar, Yasemin Cosgun, Suleyman Yalcin, Gulay Korukluoglu |
| hCoV-19/Ireland/D-NVRL-S21IRL09122/2021 | EPI_ISL_2789716 | 2021-06-23 | National Virus Reference Laboratory | National Virus Reference Laboratory | Zoe Yandle, Charlene Bennett, Gabriel Gonzalez, Michael Carr, Jonathan Dean, Cillian F De Gascun |
| hCoV-19/Kenya/C95262/2021 | EPI_ISL_2602646 | 2021-05-06 | KEMRI-Wellcome Trust Research Programme,Kilifi | KEMRI-Wellcome Trust Research Programme,Kilifi | Githinji G.,Mohamed K.S.,de Laurent Z.,Mburu M.W.,Matoke D.,Onyango C., |

| | | | | | |
|---|---|---|---|---|---|
| hCoV-19/Kenya/SS298/2021 | EPI_ISL_2603054 | 2021-05-03 | KEMRI-Wellcome Trust Research Programme,Kilifi | KEMRI-Wellcome Trust Research Programme,Kilifi | Githinji G.,Mohamed K.S.,de Laurent Z.,Mburu M.W.,Matoke D.,Onyango C., |
| hCoV-19/USA/LA-EVTL659/2020 | EPI_ISL_653351 | 2020-07-11 | LSUHS Emerging Viral Threat Laboratory | Microbial Genome Sequencing Center | Maarten Van Diest, Jeremy P. Kamil, Rona S. Scott, Malgorzata Bienkowska-Haba, Katarzyna Zwolinska, Andrew D. Yurochko, Christopher G. Kevil, Martin J. Sapp, Daniel J. Snyder, Vaughn S. Cooper, John A. Vanchiere |
| hCoV-19/India/TN-CL-287_S98_R1_001/2021 | EPI_ISL_2723831 | 2021-03-05 | Dr S raju, Director of public heath and preventive medicine | inStem NCBS ‚Äì INSACOG | Uma Ramakrishnan Dasaradhi Palakodeti Aswin SaiNarain |
| hCoV-19/USA/LA-EVTL823/2020 | EPI_ISL_653514 | 2020-07-28 | LSUHS Emerging Viral Threat Laboratory | Microbial Genome Sequencing Center | Jeremy P. Kamil, Rona S. Scott, Maarten Van Diest, Malgorzata Bienkowska-Haba, Katarzyna Zwolinska, Andrew D. Yurochko, Christopher G. Kevil, Martin J. Sapp, Daniel J. Snyder, Vaughn S. Cooper, John A. Vanchiere |
| hCoV-19/USA/WA-UW-21042249403/2021 | EPI_ISL_2037537 | 2021-04-22 | UW Virology Lab | UW Virology Lab | Pavitra Roychoudhury, Hong Xie, Lasata Shrestha, Tien V. Nguyen, Shah Mohamed Bakhash, Michelle Lin, Noah R. Baker, Sean Ellis, Meei-Li Huang, Keith R Jerome, Alexander Greninger |
| hCoV-19/USA/WA-UW-21041694101/2021 | EPI_ISL_1849851 | 2021-04-16 | UW Virology Lab | UW Virology Lab | Pavitra Roychoudhury, Hong Xie, Lasata Shrestha, Tien V. Nguyen, Shah Mohamed Bakhash, Michelle Lin, Noah R. Baker, Sean Ellis, Meei-Li Huang, Keith R Jerome, Alexander Greninger |
| hCoV-19/Russia/MOW-CRIE-L188P0160/2021 | EPI_ISL_2626269 | 2021-04-19 | Molecular diagnostic laboratory of Federal Budget Institution of Science "Central Research Institute of Epidemiology" of The Federal Service on Customers' Rights Protection and Human Well-being Surveillance | Group of Genomics and Postgenomic Technologies of Central Research Institute of Epidemiology | Samoilov A.E., Kaptelova V.V., Korneenko E.V., Sinitsyn S.O., Saenko S.S., Nadtoka M.I., Speranskaya A.S., Tivanova E.V., Kondrasheva L.Y., Smirnova Y.S., Akimkin V.G. |
| hCoV-19/USA/NY-Wadsworth-21063427-01/2021 | EPI_ISL_2627178 | 2021-05-25 | NORTHWELL HEALTH LABORATORIES | Wadsworth Center, New York State Department of Health | Kirsten St. George, Daryl M. Lamson, Alexis Russell, Matthew Shudt, Melissa A Leisner, Jonathan Plitnick, Catharine Prussing, Navjot Singh, John Kelly, Erasmus Schneider, Erica Lasek-Nesselquist |
| hCoV-19/Belgium/ULG-15608/2021 | EPI_ISL_2245441 | 2021-05-10 | SYNLAB | GIGA Medical Genomics | Keith Durkin, Maria Artesi, S√©bastien Bontems, Rapha√´l Boreux, Bouchra Boujemla, Nathalie Renotte, C√©cile Meex, Pierrette Melin, Marie-Pierre Hayette, Vincent Bours |
| hCoV-19/Australia/VIC18354/2021 | EPI_ISL_1249996 | 2021-02-24 | Victorian Infectious Diseases Reference Laboratory (VIDRL) | VIDRL and MDU-PHL | Caly L., Seemann T., Sait, M.L., Druce J., Sherry, N.L. |
| hCoV-19/NewZealand/21MV0156/2021 | EPI_ISL_1250705 | 2021-03-04 | LabPLUS | Institute of Environmental Science and Research (ESR) | Rachel Boyle, SallyAnn Harbison, Olivia Stroeven, Xiaoyun Ren, Matt Storey, Nikki Freed, Muhammad Faisal, Jing Wang, Hermes Perez, Anja Werno, Antje van der Linden, Arlo Upton, Chris Mansell, David Hammer, Dragana Drinkovic, Gary McAuliffe, Hana Sofia And |
| hCoV-19/USA/TX-DSHS-1637/2020 | EPI_ISL_710074 | 2020-06-24 | Texas Department of State Health Services | Texas Department of State Health Services | Anita Pokharel, Bonnie Oh, James Daniel Bonser, Rashmi Tuladhar, Mayela Pedrueza, Jenny Zhang, Maliha Rahman, Myong Koag, Chung Wang, Rachel Lee, Grace Kubin |

| | | | | | |
|---|---|---|---|---|---|
| hCoV-19/Ghana/WACCBIP-GS257/2020 | EPI_ISL_1255166 | 2020-12-09 | West African Centre for Cell Biology of Infectious Pathogens (WACCBIP), University of Ghana, Accra, Ghana | West African Centre for Cell Biology of Infectious Pathogens (WACCBIP), University of Ghana, Volta Road, Legon-Accra, Ghana | Collins M. Morang'a, Joyce M. Ngoi, Evelyn B. Quansah, Samirah Saiid, Dominic S.Y. Amuzu, Vincent Appiah, Philip M. Soglo, Vanessa Magnussen, Aisha Mohammed, Kesego Tapela, Nelson Kibinge, Abdoulaye B Diallo, Frederick Kumi-Ansah, Theophilus Odoom, Oliver |
| hCoV-19/Ghana/WACCBIP-GS297/2020 | EPI_ISL_1255192 | 2020-11-23 | West African Centre for Cell Biology of Infectious Pathogens (WACCBIP), University of Ghana, Accra, Ghana | West African Centre for Cell Biology of Infectious Pathogens (WACCBIP), University of Ghana, Volta Road, Legon-Accra, Ghana | Collins M. Morang'a, Joyce M. Ngoi, Evelyn B. Quansah, Samirah Saiid, Dominic S.Y. Amuzu, Vincent Appiah, Philip M. Soglo, Vanessa Magnussen, Aisha Mohammed, Kesego Tapela, Nelson Kibinge, Abdoulaye B Diallo, Frederick Kumi-Ansah, Theophilus Odoom, Oliver |
| hCoV-19/Ghana/WACCBIP-GS358/2020 | EPI_ISL_1255235 | 2020-11-27 | West African Centre for Cell Biology of Infectious Pathogens (WACCBIP), University of Ghana, Accra, Ghana | West African Centre for Cell Biology of Infectious Pathogens (WACCBIP), University of Ghana, Volta Road, Legon-Accra, Ghana | Collins M. Morang'a, Joyce M. Ngoi, Evelyn B. Quansah, Samirah Saiid, Dominic S.Y. Amuzu, Vincent Appiah, Philip M. Soglo, Vanessa Magnussen, Aisha Mohammed, Kesego Tapela, Nelson Kibinge, Abdoulaye B Diallo, Frederick Kumi-Ansah, Theophilus Odoom, Oliver |
| hCoV-19/USA/MN-MDH-3605/2020 | EPI_ISL_1267294 | 2020-11-19 | Vault Health | Minnesota Department of Health, Public Health Laboratory | Alexandra Lorentz, Jacob Garfin, Matt Plumb, and Xiong Wang |
| hCoV-19/Palestine/AAS17/2020 | EPI_ISL_1273085 | 2020-11-30 | Biochemistry and Molecular Biology Department-Faculty of Medicine, Al-Quds University | Biochemistry and Molecular Biology Department-Faculty of Medicine, Al-Quds University | Ereqat,S., Al-Jawabreh,A., Al-Jawabreh,H., Dumaidi,K., Nasereddin,A. |
| hCoV-19/Palestine/AAS29/2020 | EPI_ISL_1273092 | 2020-11-30 | Biochemistry and Molecular Biology Department-Faculty of Medicine, Al-Quds University | Biochemistry and Molecular Biology Department-Faculty of Medicine, Al-Quds University | Ereqat,S., Al-Jawabreh,A., Al-Jawabreh,H., Dumaidi,K., Nasereddin,A. |
| hCoV-19/Palestine/AAS11/2020 | EPI_ISL_1273098 | 2020-11-05 | Faculty of Medicine, Al-Quds University | University | Ereqat,S., Al-Jawabreh,A., Dumaidi,K., Nasereddin,A. |
| hCoV-19/England/QEUH-13A82D1/2021 | EPI_ISL_1276556 | 2021-03-07 | Lighthouse Lab in Glasgow | Wellcome Sanger Institute for the COVID-19 Genomics UK (COG-UK) Consortium | Harper VanSteenhouse, Yumi Kasai, David Gray, Carol Clugston, Anna Dominiczak and Alex Alderton, Roberto Amato, Jeffrey Barrett, Sonia Goncalves, Ewan Harrison, David K. Jackson, Ian Johnston, Dominic Kwiatkowski, Cordelia Langford, John Sillitoe on behal |
| hCoV-19/Germany/BW-RKI-I-049180/2021 | EPI_ISL_1356606 | 2021-03-03 | Eurofins LifeCodexx GmbH | Robert Koch Institute | unknown |
| hCoV-19/USA/NY-CDC-LC0067207/2021 | EPI_ISL_2481456 | 2021-05-29 | Laboratory Corporation of America | Centers for Disease Control and Prevention Division of Viral Diseases, Pathogen Discovery | Dakota Howard, Dhwani Batra, Peter W. Cook, Kara Moser, Adrian Paskey, Jason Caravas, Benjamin Rambo-Martin, Shatavia Morrison, Christopher Gulvick, Scott Sammons, Yvette Unoarumhi, Darlene Wagner, Matthew Schmerer, Minoo Agarwal, Eyad Almasri, Debbie Bol |
| hCoV-19/Indonesia/JK-EIJK236/2021 | EPI_ISL_1284204 | 2021-01-26 | Klinik Ciputra Medical Center | Eijkman Institute for Molecular Biology, Ministry of Research and Technology/National Agency for Research and Innovation | Frilasita A Yudhaputri, Hidayat Trimarsanto, Iskandar Adnan, Lydia V. Panggalo, Sukma Oktavianthi, Willy Agustine, Edison Johar, Safarina G Malik, Khin Saw Myint, Amin Soebandrio |
| hCoV-19/USA/FL-CDC-LC0055077/2021 | EPI_ISL_2045668 | 2021-04-28 | Laboratory Corporation of America | Centers for Disease Control and Prevention Division of Viral Diseases, Pathogen Discovery | Dakota Howard, Dhwani Batra, Peter W. Cook, Kara Moser, Adrian Paskey, Jason Caravas, Benjamin Rambo-Martin, Shatavia Morrison, Christopher Gulvick, Scott Sammons, Yvette Unoarumhi, Darlene Wagner, Matthew Schmerer, Minoo Agarwal, Eyad Almasri, Debbie Bol |

| | | | | | |
|---|---|---|---|---|---|
| hCoV-19/India/MN-333906939/2021 | EPI_ISL_2812618 | 2021-06-04 | INSACOG-MANIPUR | National Institute of Biomedical Genomics ‚Äì INSACOG | Arindam Maitra, Nanaocha Sharma, Nidhan Kumar Biswas, R K Manglem Singh, Sreedhar Chinnaswamy, Khumanthem Ranjana, Saumitra Das |
| hCoV-19/Portugal/PT7353/2021 | EPI_ISL_2248935 | 2021-05-11 | Joaquim Chaves Saude | Instituto Nacional de Saude (INSA) | Borges et al |
| hCoV-19/USA/AZ-CDC-LC0055682/2021 | EPI_ISL_2046043 | 2021-04-28 | Laboratory Corporation of America | Centers for Disease Control and Prevention Division of Viral Diseases, Pathogen Discovery | Dakota Howard, Dhwani Batra, Peter W. Cook, Kara Moser, Adrian Paskey, Jason Caravas, Benjamin Rambo-Martin, Shatavia Morrison, Christopher Gulvick, Scott Sammons, Yvette Unoarumhi, Darlene Wagner, Matthew Schmerer, Minoo Agarwal, Eyad Almasri, Debbie Bol |
| hCoV-19/USA/FL-CDC-FG-019893/2021 | EPI_ISL_1666997 | 2021-03-31 | Fulgent Genetics | Centers for Disease Control and Prevention Division of Viral Diseases, Pathogen Discovery | Dakota Howard, Dhwani Batra, Peter W. Cook, Kara Moser, Adrian Paskey, Jason Caravas, Benjamin Rambo-Martin, Shatavia Morrison, Christopher Gulvick, Scott Sammons, Yvette Unoarumhi, Darlene Wagner, Matthew Schmerer, Harry Gao, Mickey Li, John Gao, Joseph |
| hCoV-19/India/MH-NEERI-NGP-44241/2021 | EPI_ISL_1916414 | 2021-03-23 | CSIR-National Environmental Engineering Research Institute | CSIR-Centre for Cellular and Molecular Biology - INSACOG | Onkar Kulkarni, Lamuk Zaveri, Ara Sreenivas, Sofia Banu, Shreekant Verma, Amareshwar Vodapalli , Viswagithe S L, B Himasri, Sharath Chandra Thota, Karthik Bharadwaj Tallapaka, Krishna Khairnar, Rakesh K Mishra,Divya Tej Sowpati |
| hCoV-19/Australia/NT159/2021 | EPI_ISL_2250193 | 2021-04-15 | Royal Darwin Hospital Pathology | Microbiological Diagnostic Unit Public Health Laboratory (MDU-PHL) | Meumann, E., Caly L., Seemann T., Sait, M.L., Druce J., Sherry, N.L. |
| hCoV-19/Scotland/QEUH-13C2295/2021 | EPI_ISL_1293776 | 2021-03-11 | Lighthouse Lab in Glasgow | Wellcome Sanger Institute for the COVID-19 Genomics UK (COG-UK) Consortium | Harper VanSteenhouse, Yumi Kasai, David Gray, Carol Clugston, Anna Dominiczak and Alex Alderton, Roberto Amato, Jeffrey Barrett, Sonia Goncalves, Ewan Harrison, David K. Jackson, Ian Johnston, Dominic Kwiatkowski, Cordelia Langford, John Sillitoe on behal |
| hCoV-19/USA/TX-HHD-2105125610/2021 | EPI_ISL_2313096 | 2021-05-06 | Houston Health Department, Disease Prevention and Control | Houston Health Department, Disease Prevention and Control | Penn,R., Brown,P., Lara,A., Lai,Y. |
| hCoV-19/Colombia/ANT-CWOHC-VG-SEC00149S/2021 | EPI_ISL_2136279 | 2021-03-21 | SURA | Universidad Nacional de Colombia - Laboratorio Gen√≥mico One Health | Maria Stella L√≥pez, Andres F. Cardona-Rios, Daniel O. Maldonado-Perez, Sim√≥n Villegas Vel√°squez, Lina M. Hurtado, Laura Silvana Perez, Karl A Ciuoderis, Maria Ang√©lica Maya, Carolina Mu√±oz-Arango, Celeny Ortiz, Rita Almanza Payares, Idabely Betancur |
| hCoV-19/Angola/CERI-KRISP-K013430/2021 | EPI_ISL_2492899 | 2021-04-09 | Instituto Nacional de Investiga√ß√£o em Sa√∫de | and Innvoation, Stellenbosch University and KRISP, KZN Research Innovation and Sequencing Platform, UKZN. | Morais J, Neto Z, Afonso P, Miranda J, David K, Ingl√™s L, Pereira A, Paulo A Carralero RR Paix√£o JP, Freitas RH, Mufinda M, Lutucuta S, Giandhari J, Pillay S, Naidoo Y, Emmanuel SJ, Tegally H, Wilkinson E, de Oliveira T |
| hCoV-19/SouthAfrica/KRISP-K012232/2021 | EPI_ISL_2494445 | 2021-03-16 | NHLS_VIRO | KRISP, KZn Research Innovation and Sequencing Platform | Baillie Vicky, du Plessis Jeanine, Giandhari Jennifer, Pillay Sureshnee, Naidoo Yeshnee, Tshabuila Derek,Tegally Houriiyah, San James, Wilkinson Eduan, de Oliveira Tulio, Madhi Shabir |
| hCoV-19/SouthAfrica/VIDA-KRISP-K012902/2020 | EPI_ISL_2495012 | 2020-09-07 | Vaccines and Infectious Diseases Analytics Research Unit (VIDA) | KRISP, KZN Research Innovation and Sequencing Platform | Baillie Vicky, du Plessis Jeanine, Giandhari Jennifer, Pillay Sureshnee, Naidoo Yeshnee, Tegally Houriiyah, San James, Wilkinson Eduan, de Oliveira Tulio, Madhi Shabir |

| | | | | | |
|---|---|---|---|---|---|
| hCoV-19/Malawi/CERI-KRISP-K015319/2021 | EPI_ISL_2495042 | 2021-04-28 | National HIV Reference Laboratory, Ministry of Health, Public Health Institute of Malawi | KRISP, KZN Research Innovation and Sequencing Platform | Mvula B, Chilima B, Chiwaula M, Mwangomba W, Panja L, Kasambara W, Auld A, Kim L, Kampira E, Kaba M, Wadonda N, Maida A, Giandhari J, Pillay S, Naidoo Y, Lessells R, Emmanuel SJ, Tegally H, Wilkinson E, de Oliveira T |
| hCoV-19/England/MILK-15FAA30/2021 | EPI_ISL_2447083 | 2021-06-01 | Lighthouse Lab in Milton Keynes | Wellcome Sanger Institute for the COVID-19 Genomics UK (COG-UK) Consortium | The Lighthouse Lab in Milton Keynes and Alex Alderton, Roberto Amato, Jeffrey Barrett, Sonia Goncalves, Ewan Harrison, David K. Jackson, Ian Johnston, Dominic Kwiatkowski, Cordelia Langford, John Sillitoe on behalf of the Wellcome Sanger Institute COVID-1 |
| hCoV-19/NewZealand/21MV0459/2021 | EPI_ISL_2650020 | 2021-05-26 | LabPLUS | Institute of Environmental Science and Research (ESR) | Rachel Boyle, SallyAnn Harbison, Olivia Stroeven, Xiaoyun Ren, Matt Storey, Nikki Freed, Muhammad Faisal, Jing Wang, Hermes Perez, Anja Werno, Antje van der Linden, Arlo Upton, Chris Mansell, David Hammer, Dragana Drinkovic, Gary McAuliffe, Hana Sofia And |
| hCoV-19/Australia/NSW1642/2021 | EPI_ISL_2650466 | 2021-06-15 | South Eastern Area Laboratory Services (SEALS) | Clinical Pathology and Medical Research; Westmead Hospital; University of Sydney | CIDM-PH et al. |
| hCoV-19/India/KA-NIMH-SEQ-1048/2021 | EPI_ISL_2651350 | 2021-05-21 | NIMHANS | INSACOG-KA, NIMHANS | Chitra Pattabiraman, Anson Kunjumon George,Darshan Sreenivas,Pramada Prasad, Ellango Ramasamy, Sony Sharma, Mahesh Kumar.C.S, Ananthapadmanabha Kotambail, Chetan G K, Gautham Arunachal Udupi, Anita S Desai, V Ravi |
| hCoV-19/Poland/PZH-GUM-1689/2021 | EPI_ISL_2657177 | 2021-06-01 | UCK | [illegible] consortium: University of Gdansk, Medical University of Gdansk, Vaxican Ltd., Invicta Ltd. 2. National Institute of Public Health - National Institute of Hygiene, Warsaw, Poland | Maciej Kosinski, Celina Cybulska, Krystyna Bienkowska Szewczyk, Maciej Grzybek, Karolina Gackowska, Marcin Lubocki, Katarzyna Groth, Lukasz Rabalski, Katarzyna Zacharczuk, Magdalena Nowakowska, Ma≈Çgorzata Sadkowska-Todys, Tomasz Wo≈Çkowicz |
| hCoV-19/Chile/AR-192898/2020 | EPI_ISL_1300457 | 2020-09-21 | Genetica Molecular and Subdepartamento de Virologia ISP Chile | Instituto de Salud Publica de Chile | Javier Tognarelli, Karen Orostica, Barbara Parra, Loredana Arata, Jaime Lagos, Gisselle Barra, Patricia Bustos, Rodrigo Fasce, Andres Castillo, Jorge Fernandez |
| hCoV-19/Australia/NSW-R0377/2021 | EPI_ISL_2274250 | 2021-05-23 | New South Wales Health Pathology Royal Prince Alfred Hospital | Microbiology RPAH | Foster, C.; Au, J.; Ruiz Silva, M.; Deveson, I.; Bull, R.; Van Hal, S.; Rawlinson, W. |
| hCoV-19/Australia/NSW-R0249/2021 | EPI_ISL_2107448 | 2021-05-04 | New South Wales Health Pathology Royal Prince Alfred Hospital | Microbiology RPAH | Foster, C.; Au, J.; Ruiz Silva, M.; Deveson, I.; Bull, R.; Van Hal, S.; Rawlinson, W. |
| hCoV-19/Australia/NSW-R0245/2021 | EPI_ISL_2107446 | 2021-05-03 | New South Wales Health Pathology Royal Prince Alfred Hospital | Microbiology RPAH | Foster, C.; Au, J.; Ruiz Silva, M.; Deveson, I.; Bull, R.; Van Hal, S.; Rawlinson, W. |
| hCoV-19/Australia/NSW-R0262/2021 | EPI_ISL_2107506 | 2021-05-07 | New South Wales Health Pathology Royal Prince Alfred Hospital | Microbiology RPAH | Foster, C.; Au, J.; Ruiz Silva, M.; Deveson, I.; Bull, R.; Van Hal, S.; Rawlinson, W. |
| hCoV-19/Australia/NSW-R0238/2021 | EPI_ISL_2107499 | 2021-04-29 | New South Wales Health Pathology Royal Prince Alfred Hospital | Microbiology RPAH | Foster, C.; Au, J.; Ruiz Silva, M.; Deveson, I.; Bull, R.; Van Hal, S.; Rawlinson, W. |
| hCoV-19/England/CAMC-1322EBD/2021 | EPI_ISL_1333725 | 2021-02-22 | Lighthouse Lab in Cambridge | Wellcome Sanger Institute for the COVID-19 Genomics UK (COG-UK) Consortium | Rob Howes, The Lighthouse Lab in Cambridge and Alex Alderton, Roberto Amato, Jeffrey Barrett, Sonia Goncalves, Ewan Harrison, David K. Jackson, Ian Johnston, Dominic Kwiatkowski, Cordelia Langford, John Sillitoe on behalf of the Wellcome Sanger Institute |

| | | | | | |
|---|---|---|---|---|---|
| hCoV-19/India/MH-NCDC-1109337/2021 | EPI_ISL_2460194 | 2021-02-05 | National Centre for Disease Control (NCDC) Biotechnology Division, Delhi | NCDC Delhi, Biotechnology Division INSACOG | Uma Sharma, Manoj K Singh,¬†Radhakrishnan V. S, Robin Marwal, Mahesh S Dhar, Kalaiarasan Ponnusamy, Meena Datta, Priyanka Singh,¬†Hemlata Lall, Hema Gogia, Preeti Madan, Sandhya Kabra, Sujeet K Singh, Partha Rakshit |
| hCoV-19/NewZealand/21MV0171/2021 | EPI_ISL_1315322 | 2021-03-09 | LabPLUS | Institute of Environmental Science and Research (ESR) | Rachel Boyle, SallyAnn Harbison, Olivia Stroeven, Xiaoyun Ren, Matt Storey, Nikki Freed, Muhammad Faisal, Jing Wang, Hermes Perez, Anja Werno, Antje van der Linden, Arlo Upton, Chris Mansell, David Hammer, Dragana Drinkovic, Gary McAuliffe, Hana Sofia And |
| hCoV-19/India/MP-NCDC-2509239/2020 | EPI_ISL_2461256 | 2020-11-28 | National Centre for Disease Control (NCDC) Biotechnology Division, Delhi | NCDC Delhi, Biotechnology Division INSACOG | Radhakrishnan V. S, Robin Marwal, Mahesh S Dhar, Kalaiarasan Ponnusamy, Meena Datta, Priyanka Singh,¬†Uma Sharma, Hemlata Lall, Manoj K Singh,¬†Hema Gogia, Preeti Madan, Sandhya Kabra, Sujeet K Singh, Partha Rakshit |
| hCoV-19/Australia/SA0716/2021 | EPI_ISL_2462354 | 2021-05-03 | SA Pathology | SA Pathology | Caitlin Selway, Lex Leong, Chuan Kok Lim, Mark Turra, Ivan Bastian, Geoff Higgins |
| hCoV-19/USA/TX-DSHS-3928/2021 | EPI_ISL_1291335 | 2021-02-22 | TXDSHS | TXDSHS | Rashmi Tuladhar, Bonnie Oh, Jenny Zhang, Maliha Rahman, Mayela Pedrueza, Anita Pokharel, Myong Koag, Chun Wang, Rachel Lee, Grace Kubin |
| hCoV-19/Australia/QLD1951/2021 | EPI_ISL_2661523 | 2021-06-20 | Queensland Health Forensic and Scientific Services | Queensland Health Forensic and Scientific Services | Son Nguyen |
| hCoV-19/India/MH-ICMR-NIV-INSACOG-2019nCoV-279084/2020 | EPI_ISL_2131509 | 2020-12-10 | ICMR-National Institute of Virology - INSACOG | NIV Influenza | Potdar, Varsha on behalf of National Influenza Centre, NIV, Pune |
| hCoV-19/Israel/CVL-5325/2021 | EPI_ISL_1357806 | 2021-02-07 | Israel Central Virology laboratory | Israel National Consortium for SARS-CoV-2 sequencing | Neta Zuckerman, Efrat Dahan Bucris, Michal Mandelboim, Dana Bar-Ilan, Oran Erster, Tzvia Mann, Omer Murik, David A. Zeevi, Assaf Rokney, Joseph Jaffe, Eva Nachum, Maya Davidovich Cohen, Ephraim Fass, Gal Zizelski Valenci, Mor Rubinstein, Efrat Rorman, Isr |
| hCoV-19/Israel/CVL-2793/2021 | EPI_ISL_1358435 | 2021-01-18 | Israel Central Virology laboratory | Israel National Consortium for SARS-CoV-2 sequencing | Neta Zuckerman, Efrat Dahan Bucris, Michal Mandelboim, Dana Bar-Ilan, Oran Erster, Tzvia Mann, Omer Murik, David A. Zeevi, Assaf Rokney, Joseph Jaffe, Eva Nachum, Maya Davidovich Cohen, Ephraim Fass, Gal Zizelski Valenci, Mor Rubinstein, Efrat Rorman, Isr |
| hCoV-19/Israel/CVL-4542/2021 | EPI_ISL_1358645 | 2021-02-02 | Israel Central Virology laboratory | Israel National Consortium for SARS-CoV-2 sequencing | Neta Zuckerman, Efrat Dahan Bucris, Michal Mandelboim, Dana Bar-Ilan, Oran Erster, Tzvia Mann, Omer Murik, David A. Zeevi, Assaf Rokney, Joseph Jaffe, Eva Nachum, Maya Davidovich Cohen, Ephraim Fass, Gal Zizelski Valenci, Mor Rubinstein, Efrat Rorman, Isr |
| hCoV-19/Scotland/QEUH-152F30A/2021 | EPI_ISL_2022593 | 2021-04-23 | Lighthouse Lab in Glasgow | Wellcome Sanger Institute for the COVID-19 Genomics UK (COG-UK) Consortium | Harper VanSteenhouse, Yumi Kasai, David Gray, Carol Clugston, Anna Dominiczak and Alex Alderton, Roberto Amato, Jeffrey Barrett, Sonia Goncalves, Ewan Harrison, David K. Jackson, Ian Johnston, Dominic Kwiatkowski, Cordelia Langford, John Sillitoe on behal |

| | | | | | |
|---|---|---|---|---|---|
| hCoV-19/India/MH-NEERI-NGP-31713/2021 | EPI_ISL_1360342 | 2021-01-19 | CSIR-National Environmental Engineering Research Institute | CSIR-Centre for Cellular and Molecular Biology - INSACOG | Sofia Banu, Lamuk Zaveri, Ara Sreenivas, Shreekant Verma, Onkar Kulkarni, Amareshwar Vodapalli ,Viswagithe S L, B Himasri, Sharath Chandra Thota, Karthik Bharadwaj Tallapaka, Krishna Khairnar, Rakesh K Mishra,Divya Tej Sowpati |
| hCoV-19/Australia/NSW4663/2021 | EPI_ISL_2404988 | 2021-05-03 | Area of Virology, Serology and Virology Division (SAViD), New South Wales Health Pathology Randwick | Virology, Serology and Virology Division (SAViD), New South Wales Health Pathology Randwick | Foster, C.; Au, J.; Ruiz Silva, M.; Deveson, I.; Bull, R.; Van Hal, S.; Rawlinson, W. |
| hCoV-19/SouthAfrica/NCV31/2020 | EPI_ISL_1363148 | 2020-07-29 | National Health Laboratory Services | National Health Laboratory Services | Kathleen Subramoney, Florette Treurnicht, Arshad Ismail |
| hCoV-19/England/CAMC-133CCF2/2021 | EPI_ISL_1831443 | 2021-02-26 | Lighthouse Lab in Cambridge | Wellcome Sanger Institute for the COVID-19 Genomics UK (COG-UK) Consortium | Rob Howes, The Lighthouse Lab in Cambridge and Alex Alderton, Roberto Amato, Jeffrey Barrett, Sonia Goncalves, Ewan Harrison, David K. Jackson, Ian Johnston, Dominic Kwiatkowski, Cordelia Langford, John Sillitoe on behalf of the Wellcome Sanger Institute |
| hCoV-19/USA/VA-DCLS-3612/2021 | EPI_ISL_1364578 | 2021-02-05 | Virginia Division of Consolidated Laboratory Services | Virginia Division of Consolidated Laboratory Services | Virginia DCLS |
| hCoV-19/SouthAfrica/KRISP-K010439/2021 | EPI_ISL_1371933 | 2021-02-23 | National Health Laboratory Service, South Africa | KRISP, KZn Research Innovation and Sequencing Platform | Giandhari J, Pillay S, Maslo C, Sitharam L, Lessells R, Mdlalose K, York D, Khan S, Emmanuel SJ, Tegally H, Wilkinson E, de Oliveira T |
| hCoV-19/India/AP-CCMB-BI361/2021 | EPI_ISL_1372268 | 2021-03-02 | AP SSO | CSIR-Centre for Cellular and Molecular Biology - INSACOG | Lamuk Zaveri, Ara Sreenivas, Onkar Kulkarni, Sofia Banu, Shreekant Verma,Amareshwar Vodapalli, Viswagithe S L, B Himasri, Sharath Chandra Thota, Karthik Bharadwaj Tallapaka, Rakesh K Mishra,Divya Tej Sowpati |
| hCoV-19/Denmark/DCGC-122407/2021 | EPI_ISL_2784666 | 2021-06-25 | Department of Bacteria, Parasites and Fungi, Statens Serum Institut, Copenhagen, Denmark | Statens Serum Institut Bioinformatics and Microbial Genomics | Danish Covid-19 Genome Consortium |
| hCoV-19/Poland/0951WSN2021_wsseol/2021 | EPI_ISL_2776155 | 2021-06-17 | Wojewodzka Stacja Sanitarno-Epidemiologiczna w Olsztynie, Laboratorium Badan Epidemiologiczno-Klinicznych | Wojewodzka Stacja Sanitarno-Epidemiologiczna w Olsztynie, Laboratorium Badan Epidemiologiczno-Klinicznych | Sylwia Krzetowska, Monika Czerminska, Ewa Liszewska, Tomasz Jakubczak, Marta Lukian, Patryk Bielecki, Aleksandra Kobiatko, Emilia Tarabasz, Paulina Rozycka, Barbara Dolinska |
| hCoV-19/India/TG-CCMB-BO867/2021 | EPI_ISL_1838907 | 2021-04-12 | CSIR-Centre for Cellular and Molecular Biology | CSIR-Centre for Cellular and Molecular Biology-INSACOG | Payel Mukherjee,Lamuk Zaveri,Tulasi Nagabandi, Ara Sreenivas,Shreekant Verma, Amareshwar Vodapalli ,Blessy B John,Viswagithe S L,B Himasri,Valli Nagalakshmi Undamatla,Onkar Kulkarni,Sofia Banu,Archana Bharadwaj Siva,Sharath Chandra Thota,Karthik Bharadwaj |
| hCoV-19/Kenya/C90935/2021 | EPI_ISL_2602567 | 2021-04-06 | KEMRI-Wellcome Trust Research Programme,Kilifi | KEMRI-Wellcome Trust Research Programme,Kilifi | Githinji G.,Mohamed K.S.,de Laurent Z.,Mburu M.W.,Matoke D.,Onyango C., |
| hCoV-19/Argentina/PAIS-E0083/2020 | EPI_ISL_1395897 | 2020-12-11 | Laboratorio Central de la Ciudad de Santa Fe | [illegible] Bioinformv°tica del Instituto de Investigaciv≥n de la Cadena Lv°ctea CONICET-INTA on behalf of 'Proyecto Argentino Interinstitucional de genomica de SARS-CoV-2' (PAIS | Eberhardt, MF; Irazoqui, JM; Ojeda, G; Rompato, G; Mugna, V; Pastor, C; Amadio, AF |

| | | | | | |
|---|---|---|---|---|---|
| hCoV-19/Argentina/PAIS-E0148/2020 | EPI_ISL_1395932 | 2020-08-05 | Laboratorio Central De Redes y Programas | [illegible] Bioinform√°tica del Instituto de Investigaci√≥n de la Cadena L√°ctea CONICET-INTA on behalf of 'Proyecto Argentino Interinstitucional de genomica de SARS-CoV-2' (PAIS | Natalia Ruiz Diaz, Gerardo Andino, Antonieta Cayr√©, Laura Lescano, Eberhardt, MF, Irazoqui, Amadio, AF |
| hCoV-19/Argentina/PAIS-G0074/2020 | EPI_ISL_1396007 | 2020-06-21 | Laboratorio Central, Ministerio de Salud Cordoba | INTA) on behalf of 'Proyecto Argentino Interinstitucional de genomica de SARS-CoV-2' (PAIS Consortium) | Fern√°ndez, FD; Marquez, N.; Debat, HJ.; Re, V.; Pisano, M.B.; Castro, G.; Barbas, G. |
| hCoV-19/Argentina/PAIS-H0090/2020 | EPI_ISL_1396501 | 2020-09-30 | Centro de Tecnolog√≠a en Salud P√∫blica de la Universidad Nacional de Rosario | Laboratorio Mixto de Biotecnolog√≠a Acu√°tica (LMBA) on behalf of 'Proyecto Argentino Interinstitucional de genomica de SARS-CoV-2' (PAIS Consortium) | Joaqu√≠n Ezpeleta, Ignacio Garc√≠a Labari, Victoria Posner, Vanina Villanova, Pablo Casal, Pilar Bulacio, Sof√≠a Lavista Llanos, Federico Remes Lenicov, Ana Paletta, Leandro Ciappina, Flavio Spetale, Agustina Cerri, Silvana Spinelli, Elisa Bolatti, Diego |
| hCoV-19/India/WB-1931700218712/2021 | EPI_ISL_1589936 | 2021-03-19 | INSACOG-WB | National Institute of Biomedical Genomics ‚Äì INSACOG | Arindam Maitra, Bhaswati Bandyopadhyay, Nidhan Kumar Biswas, Tamal Ghosh, Sreedhar Chinnaswamy, Ajay Chakraborti, Saumitra Das |
| hCoV-19/USA/PA-VSP0937/2021 | EPI_ISL_1402980 | 2021-03-08 | Hospital of the University of Pennsylvania Molecular Pathology Lab | Bushman Lab - University of Pennsylvania | John Everett, Kyle Rodino, Shantan Reddy, Pascha Hokama, Aoife M. Roche, Young Hwang, Abigail Glascock, Scott Sherrill-Mix, Samantha A. Whiteside, Jevon Graham-Wooten, Layla A. Khatib, Ayannah S. Fitzgerald, Arupa Ganguly, Mike Feldman, Brendan Kelly, Ron |
| hCoV-19/Italy/PIE-15840603/2021 | EPI_ISL_2650717 | 2021-04-20 | A.S.L. VERCELLI | Fondazione del Piemonte per l'Oncologia IRCCS | Antonino Sottile, Silvia Brossa, Paola Marino, Giorgio Giardina |
| hCoV-19/Italy/LOM-Policlinico-25687009/2021 | EPI_ISL_2728614 | 2021-06-23 | Fondazione IRCCS Ca' Granda Ospedale Maggiore Policlinico | Fondazione IRCCS Ca' Granda Ospedale Maggiore Policlinico | Ferruccio Ceriotti, Sara Uceda Renteria |
| hCoV-19/TrinidadandTobago/57842/2021 | EPI_ISL_1591456 | 2021-03-05 | Caribbean Public Health Agency | Carrington Lab, Department of PreClinical Sciences, Faculty of Medical Sciences, The University of the West Indies | Nikita S. D. Sahadeo, Arianne Brown-Jordan, Sarah Hill, Vernie Ramkissoon, Roshan Parasram, Naresh Nandram, Avery Hinds, Jerome Foster, Stanley Giddings, Karla Georges, Marsha Ivey, Rahul Naidu, Risha Singh, SueMin Nathaniel, Rajini Haraksingh, Jaya Jayar |
| hCoV-19/England/NORW-13D30E2/2021 | EPI_ISL_2519049 | 2021-05-31 | Quadram Institute Bioscience | COVID-19 Genomics UK (COG-UK) Consortium | Dave J. Baker, Gemma L. Kay, Alp Aydin, Thanh Le-Viet, Steven Rudder, Ana P. Tedim, Anastasia Kolyva, Maria Diaz, Leonardo de Oliveira Martins, Nabil-Fareed Alikhan, Lizzie Meadows, Rachael Stanley, Ngozi Elumogo, Muhammed Yasir, Nicholas M. Thomson, Alex |
| hCoV-19/USA/NV-NSPHL-NV2041450/2020 | EPI_ISL_1404612 | 2020-11-09 | Nevada State Public Health Laboratory | Nevada State Public Health Laboratory | Andrew Gorzalski, Mark Pandori |
| hCoV-19/Spain/CL-IBV-98029697/2020 | EPI_ISL_2754272 | 2020-05-19 | Universidad de Le√≥n | SeqCOVID-SPAIN consortium/IBV(CSIC) | Ana Carvajal, Vicente Mart√≠n, H√©ctor Arg√ºello, Juan M. Fregeneda, Tania Fern√°ndez-Villa, Antonio J. Molina and SeqCOVID-SPAIN consortium |
| hCoV-19/Japan/PG-40689/2021 | EPI_ISL_2359503 | 2021-05-01 | Department of Infectious Diseases, Kobe Institute of Health | Department of Infectious Diseases, Kobe Institute of Health | Ryohei Nomoto, Noriko Nakanishi, Tomotada Iwamoto, Tsuyoshi Sekizuka, Kentaro Itokawa, Rina Tanaka, Masanori Hashino, Makoto Kuroda |

| | | | | | |
|---|---|---|---|---|---|
| hCoV-19/Malawi/KRISP-K010161/2021 | EPI_ISL_1407122 | 2021-01-22 | National HIV Reference Laboratory, Ministry of Health, Public Health Institute of Malawi | KRISP, KZN Research Innovation and Sequencing Platform | Mvula B, Chilima B, Chiwaula M, Mwangomba W, Panja L, Kasambara W, Auld A, Kim L, Kampira E, Kaba M, Wadonda N, Maida A, Giandhari J, Pillay S, Naidoo Y, Lessells R, Emmanuel SJ, Tegally H, Wilkinson E, de Oliveira T |
| hCoV-19/Australia/QLD1889/2021 | EPI_ISL_2274245 | 2021-05-18 | Queensland Health Forensic and Scientific Services | Queensland Health Forensic and Scientific Services | Son Nguyen |
| hCoV-19/Kenya/C87486/2021 | EPI_ISL_1440266 | 2021-03-16 | KEMRI-Wellcome Trust Research Programme,Kilifi | KEMRI-Wellcome Trust Research Programme,Kilifi | Githinji G.,Mohamed K.S.,deLaurent Z.,Mburu M.W. |
| hCoV-19/USA/NY-Wadsworth-21032996-01/2021 | EPI_ISL_1465717 | 2021-03-01 | NYC Pandemic Response Lab | Wadsworth Center, New York State Department of Health | Kirsten St. George, Daryl M. Lamson, Alexis Russell, Matthew Shudt, Melissa A Leisner, Jonathan Plitnick, Catharine Prussing, Navjot Singh, John Kelly, Erasmus Schneider, Erica Lasek-Nesselquist |
| hCoV-19/Brazil/RS-14039CVL/2020 | EPI_ISL_1469556 | 2020-08-11 | Secretaria Municipal de Sa√∫de de S√£o Leopoldo | Epiclin | Fernando Hayashi Sant,ÄôAnna, Ana Paula Muterle, Janira Prichula, Juliana Comerlato, Carolina Comerlato, Eliana M√°rcia Da Ros Wendland |
| hCoV-19/Brazil/RS-30291CVL/2020 | EPI_ISL_1469839 | 2020-11-15 | HOSPITAL DE CAMPO BOM DR LAURO REUS | Epiclin | Fernando Hayashi Sant,ÄôAnna, Ana Paula Muterle, Janira Prichula, Juliana Comerlato, Carolina Comerlato, Eliana M√°rcia Da Ros Wendland |
| hCoV-19/Chile/AP-53845/2021 | EPI_ISL_1470427 | 2021-03-15 | Genetica Molecular and Subdepartamento de Virologia ISP Chile | Instituto de Salud Publica de Chile | Javier Tognarelli, Karen Orostica, Barbara Parra, Loredana Arata, Jaime Lagos, Gisselle Barra, Patricia Bustos, Rodrigo Fasce, Andres Castillo, Jorge Fernandez |
| hCoV-19/Luxembourg/LNS5818306/2021 | EPI_ISL_1477110 | 2021-03-01 | Centre Hospitalier du Nord | Laboratoire national de sante, Microbiology, Microbial Genomics Platform | Anke Wienecke-Baldacchino, Catherine Ragimbeau,Jessica Tapp, Fatu Djabi, Lise Pignon, Raoul Salmon, Fatiha Boulmerka, Tamir Abdelrahman |
| hCoV-19/SouthKorea/KDCA2305/2021 | EPI_ISL_1489626 | 2021-03-03 | Division of Emerging Infectious Diseases, Bureau of Infectious Diseases Diagnosis Control, Korea Disease Control and Prevention Agency | Diseases, Bureau of Infectious Diseases Diagnosis Control, Korea Disease Control and Prevention Agency | Ae Kyung Park, Il-Hwan Kim, Heui Man Kim, Jeong-Min Kim, Jeong-Ah Kim, Chae Young Lee, Jin Sun No, Eun-Jin Kim |
| hCoV-19/Sweden/21-51785/2021 | EPI_ISL_1300308 | 2021-02-28 | Klinisk mikrobiologi | The Public Health Agency of Sweden | Samuel Ohman, Maria Lind Karlberg, Anna-Malin Linde, Carlo Berg, Oskar Karlsson Lindsjo, Sofia Stamouli, Reza Advani, Mattias Haukland, Petra Holmstrom, Noura Walai, Petra Edquist, Mia Brytting, Anna Risberg, Karin Tegmark-Wisell |
| hCoV-19/India/MH-NEERI-NGP-29989/2021 | EPI_ISL_1360329 | 2021-01-06 | CSIR-National Environmental Engineering Research Institute | CSIR-Centre for Cellular and Molecular Biology - INSACOG | Lamuk Zaveri, Ara Sreenivas, Sofia Banu, Onkar Kulkarni, Shreekant Verma, Amareshwar Vodapalli , Viswagithe S L, B Himasri, Sharath Chandra Thota, Karthik Bharadwaj Tallapaka, Krishna Khairnar, Rakesh K Mishra,Divya Tej Sowpati |
| hCoV-19/Australia/NSW-R0167/2021 | EPI_ISL_1315070 | 2021-03-18 | New South Wales Health Pathology Royal Prince Alfred Hospital | Microbiology RPAH | Foster, C.; Au, J.; Ruiz Silva, M.; Deveson, I.; Bull, R.; Van Hal, S.; Rawlinson, W. |
| hCoV-19/Chile/RM-52474/2021 | EPI_ISL_1470499 | 2021-03-15 | Genetica Molecular and Subdepartamento de Virologia ISP Chile | Instituto de Salud Publica de Chile | Javier Tognarelli, Karen Orostica, Barbara Parra, Loredana Arata, Jaime Lagos, Gisselle Barra, Patricia Bustos, Rodrigo Fasce, Andres Castillo, Jorge Fernandez |

| | | | | | |
|---|---|---|---|---|---|
| hCoV-19/England/QEUH-7A8C2D/2020 | EPI_ISL_533086 | 2020-07-14 | Lighthouse Lab in Glasgow | Wellcome Sanger Institute for the COVID-19 Genomics UK (COG-UK) Consortium | Harper VanSteenhouse, Yumi Kasai, David Gray, Carol Clugston, Anna Dominiczak and Alex Alderton, Roberto Amato, Sonia Goncalves, Ewan Harrison, David K. Jackson, Ian Johnston, Dominic Kwiatkowski, Cordelia Langford, John Sillitoe on behalf of the Wellcome |
| hCoV-19/Japan/IC-0126/2020 | EPI_ISL_591467 | 2020-08-19 | Pathogen Genomics Center, National Institute of Infectious Diseases | Pathogen Genomics Center, National Institute of Infectious Diseases | Tsuyoshi Sekizuka, Kentaro Itokawa, Rina Tanaka, Masanori Hashino, Makoto Kuroda |
| hCoV-19/USA/AZ-TG794421/2020 | EPI_ISL_1500648 | 2020-11-03 | Sonora Quest Laboratories | TGen North | Jolene Bowers, Heather Centner, Chris French, Hayley Yaglom, Ashlyn Pfeiffer, Darrin Lemmer, Dave Engelthaler, The Arizona COVID Genomics Union (ACGU) |
| hCoV-19/Slovenia/57237/2020 | EPI_ISL_1510959 | 2020-07-17 | Institute of Microbiology and Immunology, Faculty of Medicine, University of Ljubljana | Institute of Microbiology and Immunology, Faculty of Medicine, University of Ljubljana | Alen Suljiƒç, Samo Zakotnik, Toma≈æ Mark Zorec, Matic Brvar, Doroteja Vlaj, Andra≈æ Celar, Dominika ≈†turm, Patricija Pozvek, ≈†pela Pleh, Mi≈°a Korva, Mario Poljak, Tatjana Av≈°iƒç ,Äì ≈Ωupanc |
| hCoV-19/Botswana/R8B32_BHP_AAB10536/2020 | EPI_ISL_1516849 | 2020-12-11 | Botswana Harvard HIV Reference Laboratory | Botswana Harvard HIV Reference Laboratory | Sikhulile Wonderful T. Choga, Dorcas Maruapula, Thongbotho Mphoyakgosi, Boitumelo Zuze, Botshelo Radibe, Legodile Kooepile, David Lawrence, Roger Shapiro, Shahin Lockman, Mosepele Mosepele, Joseph Makhema, Simani Gaseitsiwe |
| hCoV-19/England/CAMC-14ABF6D/2021 | EPI_ISL_1519300 | 2021-03-31 | Lighthouse Lab in Cambridge | Wellcome Sanger Institute for the COVID-19 Genomics UK (COG-UK) Consortium | Rob Howes, The Lighthouse Lab in Cambridge and Alex Alderton, Roberto Amato, Jeffrey Barrett, Sonia Goncalves, Ewan Harrison, David K. Jackson, Ian Johnston, Dominic Kwiatkowski, Cordelia Langford, John Sillitoe on behalf of the Wellcome Sanger Institute |
| hCoV-19/Australia/SA0499/2020 | EPI_ISL_602580 | 2020-10-12 | SA Pathology | SA Pathology | Lex Leong, Julien Soubrier, Chuan Kok Lim, Song Gao, Mark Turra, Karin Kassahn, Ivan Bastian, Geoff Higgins |
| hCoV-19/Curacao/CW-RIVM-21646/2021 | EPI_ISL_1521078 | 2021-03-21 | Dutch COVID-19 response team | National Institute for Public Health and the Environment (RIVM) | Adam Meijer, Harry Vennema, Dirk Eggink, Jeroen Cremer, Sharon van den Brink, Bas van der Veer, AnneMarie van den Brandt, Lisa Wijsman, Kim Freriks, Ryanne Jaarsma, Eun√≠ce Then, Jolienke Hardeman, Lynn Aarts, Sanne Bos, Melissa van Tuil, Robert Kohl, Lin |
| hCoV-19/Australia/SA0551/2020 | EPI_ISL_654798 | 2020-11-14 | SA Pathology | SA Pathology | Lex Leong, Julien Soubrier, Chuan Kok Lim, Song Gao, Mark Turra, Karin Kassahn, Ivan Bastian, Geoff Higgins |
| hCoV-19/Japan/PG-1684/2020 | EPI_ISL_685290 | 2020-05-05 | Pathogen Genomics Center, National Institute of Infectious Diseases | Pathogen Genomics Center, National Institute of Infectious Diseases | Tsuyoshi Sekizuka, Kentaro Itokawa, Rina Tanaka, Masanori Hashino, Makoto Kuroda |
| hCoV-19/Canada/QC-DYN-0621935/2020 | EPI_ISL_1523443 | 2020-12-18 | Laboratoire de sant√© publique du Qu√©bec | Laboratoire de sant√© publique du Qu√©bec | Sandrine Moreira, Ioannis Ragoussis, Guillaume Bourque, Jesse Shapiro, Mark Lathrop and Michel Roger on behalf of the CoVSeQ research group (http://covseq.ca/researchgroup) |
| hCoV-19/Peru/AMA-INS-766/2020 | EPI_ISL_1532165 | 2020-05-30 | Laboratorio de Referencia Nacional de Virus Respiratorio. Instituto Nacional de Salud Per√∫ | Laboratorio de Referencia Nacional de Biotecnolog√≠a y Biolog√≠a Molecular. Instituto Nacional de Salud Per√∫ | Carlos Padilla Rojas, Karolyn Vega Chozo, Luis Barcena, Priscila Lope Pari, Omar Caceres Rey, Marco Galarza Perez, Maribel Huaringa Nu√±ez, Johanna Balbuena Torrez, Henri Bailon Calderon, Nancy Rojas Serrano |

| | | | | | |
|---|---|---|---|---|---|
| hCoV-19/Peru/CAL-INS-767/2020 | EPI_ISL_1532166 | 2020-07-10 | Laboratorio de Referencia Nacional de Virus Respiratorio. Instituto Nacional de Salud Per√∫ | Laboratorio de Referencia Nacional de Biotecnolog√≠a y Biolog√≠a Molecular. Instituto Nacional de Salud Per√∫ | Carlos Padilla Rojas, Karolyn Vega Chozo, Luis Barcena, Priscila Lope Pari, Omar Caceres Rey, Marco Galarza Perez, Maribel Huaringa Nu√±ez, Johanna Balbuena Torrez, Henri Bailon Calderon, Nancy Rojas Serrano |
| hCoV-19/Japan/PG-6750/2020 | EPI_ISL_687860 | 2020-08-11 | Pathogen Genomics Center, National Institute of Infectious Diseases | Pathogen Genomics Center, National Institute of Infectious Diseases | Tsuyoshi Sekizuka, Kentaro Itokawa, Rina Tanaka, Masanori Hashino, Makoto Kuroda |
| hCoV-19/India/MH-NCCS-CHN21026142/2021 | EPI_ISL_1533778 | 2021-02-22 | National Centre For Cell Science | National Centre For Cell Science ‚Äì INSACOG | Dhiraj Paul, Sonal Manik Chavan, Mohak P Gujare, Shivang P. Bhanushali, Mitali Inamdar, Manoj Kumar Bhat, Ajay Pillai, INSACOG Consortium team, Yogesh Shouche |
| hCoV-19/Japan/PG-3117/2020 | EPI_ISL_691239 | 2020-05-17 | Pathogen Genomics Center, National Institute of Infectious Diseases | Pathogen Genomics Center, National Institute of Infectious Diseases | Tsuyoshi Sekizuka, Kentaro Itokawa, Rina Tanaka, Masanori Hashino, Makoto Kuroda |
| hCoV-19/Switzerland/VS-ETHZ-370483/2020 | EPI_ISL_693969 | 2020-11-16 | Viollier AG | Department of Biosystems Science and Engineering, ETH Z√ºrich | Christian Beisel, Sarah Nadeau, Chaoran Chen, Ivan Topolsky, Pedro Ferreira, Philipp Jablonski, Susana Posada-C√©spedes, Tobias Sch√§r, Ina Nissen, Natascha Santacroce, Elodie Burcklen, Christiane Beckmann, Maurice Redondo, Olivier Kobel, Christoph Noppen |
| hCoV-19/India/MH-AFCO-15534/2020 | EPI_ISL_722200 | 2020-07-27 | Armed Forces Medical College | National Centre For Cell Science | Dhiraj Paul, Kunal Jani, Radha Chauhan, Janesh Kumar, Vasudevan Seshadri, Girdhari Lal, Rajesh Karyakarte, Suvarna Joshi, Murlidhar Tambe, Sourav Sen, Santosh Karade, Kavita Bala Anand, Shelinder Pal Singh Shergill, Rajiv Mohan Gupta, Manoj Kumar Bhat, Ar |
| hCoV-19/Australia/SA0593/2020 | EPI_ISL_732961 | 2020-12-19 | SA Pathology | SA Pathology | Lex Leong, Julien Soubrier, Chuan Kok Lim, Song Gao, Mark Turra, Karin Kassahn, Ivan Bastian, Geoff Higgins |
| hCoV-19/Belgium/rega-0722802/2020 | EPI_ISL_734815 | 2020-07-22 | UZ Leuven, National Reference Laboratory for Coronaviruses, Laboratory Medicine, Leuven, Belgium | KU Leuven, Rega Institute, Clinical and Epidemiological Virology | Tony Wawina-Bokalanga, Joan Marti-Carerras, Bert Vanmechelen, Piet Maes |
| hCoV-19/Brazil/SP-900B/2020 | EPI_ISL_763074 | 2020-12-21 | Diagnosticos da America - DASA | Procedures Center, Strategic Laboratory | Claudio Tavares Sacchi, Claudia Regina Gon√ßalves, Erica Valessa Ramos Gomes, Karoline Rodrigues Campos |
| hCoV-19/Japan/IC-0470/2020 | EPI_ISL_768679 | 2020-12-20 | Pathogen Genomics Center, National Institute of Infectious Diseases | Pathogen Genomics Center, National Institute of Infectious Diseases | Tsuyoshi Sekizuka, Kentaro Itokawa, Rina Tanaka, Masanori Hashino, Makoto Kuroda |
| hCoV-19/USA/CA-CDC-STM-021/2020 | EPI_ISL_802605 | 2020-12-28 | Helix/Illumina | Genomics and Discovery, Respiratory Viruses Branch, Division of Viral Diseases, Centers for Disease Control and Prevention | Peter W. Cook, Dhwani Batra, Ben L. Rambo-Martin Eileen de Feo, Jan Antico, Christine Tran, Matthew Tolentino, Shannon Wickline, Kim Gietzen, Brad Sickler, Jingtao Liu, Eric Allen, Phil Febbo, Summer Galloway, Nicole L. Washington, Simon White, Geraint Le |
| hCoV-19/USA/CA-CDC-STM-070/2020 | EPI_ISL_802615 | 2020-12-26 | Helix/Illumina | Genomics and Discovery, Respiratory Viruses Branch, Division of Viral Diseases, Centers for Disease Control and Prevention | Peter W. Cook, Dhwani Batra, Ben L. Rambo-Martin Eileen de Feo, Jan Antico, Christine Tran, Matthew Tolentino, Shannon Wickline, Kim Gietzen, Brad Sickler, Jingtao Liu, Eric Allen, Phil Febbo, Summer Galloway, Nicole L. Washington, Simon White, Geraint Le |
| hCoV-19/Bolivia/26953/2020 | EPI_ISL_837550 | 2020-05-03 | Centro Nacional de Enfermedades Tropicales (CENETROP) | Laboratory of Respiratory Viruses and Measles, Oswaldo Cruz Institute, FIOCRUZ | Paola Resende, Roxana Loayza, Cinthia Avila, Luciana Appolinario, Fernando Motta, Anna Carolina Paixao, Ana Carolina Mendonca, Marilda Siqueira on behalf of the Fiocruz COVID-19 Genomic Surveillance Network |

| | | | | | |
|---|---|---|---|---|---|
| hCoV-19/Switzerland/GE-32860303/2021 | EPI_ISL_860241 | 2021-01-03 | University Hospitals of Geneva, Laboratory of Virology | HUG, Laboratory of Virology and Universit√§tsspital Basel | Samuel Cordey, Ana Rita Goncalves, Laurent Kaiser, Tim Roloff, Madlen Stange, Helena MB Seth-Smith, Alfredo Mari, Karoline Leuzinger, Julia Bielicki, Manuel Battegay, Hans Hirsch, Adrian Egli |
| hCoV-19/Ireland/WD-NVRL-g21IRL49158/2021 | EPI_ISL_1552253 | 2021-02-26 | National Virus Reference Laboratory | National Virus Reference Laboratory | Guerrino Macori, Gabriel Gonzalez, Michael Carr, Zoe Yandle, Charlene Bennett, Jonathan Dean, Seamus Fanning, Cillian F De Gascun |
| hCoV-19/Japan/PG-15780/2020 | EPI_ISL_894589 | 2020-11-20 | Pathogen Genomics Center, National Institute of Infectious Diseases | Pathogen Genomics Center, National Institute of Infectious Diseases | Tsuyoshi Sekizuka, Kentaro Itokawa, Rina Tanaka, Masanori Hashino, Makoto Kuroda |
| hCoV-19/Australia/NSW4453/2021 | EPI_ISL_1563681 | 2021-04-10 | Area of Virology, Serology and Virology Division (SAViD), New South Wales Health Pathology Randwick | Virology, Serology and Virology Division (SAViD), New South Wales Health Pathology Randwick | Foster, C.; Au, J.; Ruiz Silva, M.; Deveson, I.; Bull, R.; Van Hal, S.; Rawlinson, W. |
| hCoV-19/Japan/PG-9840/2020 | EPI_ISL_902550 | 2020-10-13 | Department of Infectious Diseases, Kobe Institute of Health | Pathogen Genomics Center, National Institute of Infectious Diseases | Tsuyoshi Sekizuka, Kentaro Itokawa, Rina Tanaka, Masanori Hashino, Makoto Kuroda |
| hCoV-19/USA/MN-CDC-2-4044442/2020 | EPI_ISL_903713 | 2020-12-06 | MN PHL Division, Minnesota Department of Health | Genomics and Discovery, Respiratory Viruses Branch, Division of Viral Diseases, Centers for Disease Control and Prevention | Krista Queen, Yan Li, Ying Tao, Jing Zhang, Anna Uehara, Anna Montmayeur, Clinton R. Paden, Peter W. Cook, Rachel Marine, Mili Sheth, Jasmine Padilla, Sarah Nobles, Mark Burroughs, Lori Rowe, Haibin Wang, Ben L. Rambo-Martin, Dhwani Batra, Justin Lee, Sux |
| hCoV-19/Suriname/SR-RIVM-22492/2021 | EPI_ISL_1522126 | 2021-03-22 | Dutch COVID-19 response team | National Institute for Public Health and the Environment (RIVM) | Adam Meijer, Harry Vennema, Dirk Eggink, Jeroen Cremer, Sharon van den Brink, Bas van der Veer, AnneMarie van den Brandt, Lisa Wijsman, Kim Freriks, Ryanne Jaarsma, Eun√≠ce Then, Jolienke Hardeman, Lynn Aarts, Sanne Bos, Melissa van Tuil, Robert Kohl, Lin |
| hCoV-19/Australia/QLD1481/2020 | EPI_ISL_944736 | 2020-12-26 | Queensland Health Forensic and Scientific Services | Queensland Health Forensic and Scientific Services | Son Nguyen et al |
| hCoV-19/Australia/QLD1487/2020 | EPI_ISL_944737 | 2020-12-29 | Queensland Health Forensic and Scientific Services | Queensland Health Forensic and Scientific Services | Son Nguyen et al |
| hCoV-19/Australia/QLD1527/2021 | EPI_ISL_944739 | 2021-01-17 | Queensland Health Forensic and Scientific Services | Queensland Health Forensic and Scientific Services | Son Nguyen et al |
| hCoV-19/England/NORT-1B7B503/2020 | EPI_ISL_950235 | 2020-12-31 | Northumbria University / South Tees Hospitals NHS Foundation Trust / North Cumbria Integrated Care NHS Foundation Trust / North Tees and Hartlepool NHS Foundation Trust / Newcastle Hospitals NHS Foundation Trust | COVID-19 Genomics UK (COG-UK) Consortium | Darren L Smith,Andrew Nelson,Matthew Bashton,Greg R Young,Joshua Loh,John Allan,Mohammad A Tariq,Giles S Holt,Gary Black,Wen C Yew,Lynn Dover,Paul Baker,Steve Liggett,Sarah Essex,Jane Greenaway,Debra Padgett,Clive Graham,Garren Scott,Edward Barton,Emma Sw |
| hCoV-19/Australia/QLD1468/2020 | EPI_ISL_849744 | 2020-12-06 | Queensland Health Forensic and Scientific Services | Queensland Health Forensic and Scientific Services | Son Nguyen et al |
| hCoV-19/Germany/BW-ChVir21958/2020 | EPI_ISL_818342 | 2020-12-30 | Charit√© Universit√§tsmedizin Berlin, Institute of Virology | Charit√© Universit√§tsmedizin Berlin, Institute of Virology | Victor M Corman, Julia Schneider, J√∂rn Beheim-Schwarzbach, Barbara M√ºhlemann, Talitha Veith, Terry Jones, Christian Drosten |
| hCoV-19/Japan/PG-17992/2021 | EPI_ISL_1131144 | 2021-01-12 | Pathogen Genomics Center, National Institute of Infectious Diseases | Pathogen Genomics Center, National Institute of Infectious Diseases | Tsuyoshi Sekizuka, Kentaro Itokawa, Rina Tanaka, Masanori Hashino, Makoto Kuroda |
| hCoV-19/Germany/NW-RKI-I-017548/2021 | EPI_ISL_1147710 | 2021-02-04 | amedes MVZ DIAMEDIS Sennestadt | Robert Koch Institute | unknown |

| | | | | | |
|---|---|---|---|---|---|
| hCoV-19/USA/TX-HMH-MCoV-28703/2021 | EPI_ISL_1238487 | 2021-02-05 | Houston Methodist Hospital | Houston Methodist Hospital | S. Wesley Long, Randall J. Olsen, Paul A. Christensen, Sishir Subedi, Robert Olson, James J. Davis, Matthew Ojeda Saavedra, Prasanti Yerramilli, Layne Pruitt, Kristina Reppond, Madison N. Shyer, Jessica Cambric, Ilya J. Finkelstein, Jimmy Gollihar, and |
| hCoV-19/Aruba/AW-RIVM-14890/2021 | EPI_ISL_1165530 | 2021-02-15 | Dutch COVID-19 response team | National Institute for Public Health and the Environment (RIVM) | Adam Meijer, Harry Vennema, Dirk Eggink, Jeroen Cremer, Sharon van den Brink, Bas van der Veer, AnneMarie van den Brandt, Florian Zwagemaker, Dennis Schmitz, Chantal Reusken, on behalf of the national COVID-19 response team |
| hCoV-19/PuertoRico/PR-CDC-LC0013726/2021 | EPI_ISL_1037989 | 2021-02-02 | Laboratory Corporation of America | Respiratory Viruses Branch, Division of Viral Diseases, Centers for Disease Control and Prevention | Peter W. Cook, Dakota Howard, Dhwani Batra, Ben L. Rambo-Martin, Clinton R. Paden, Suxiang Tong, Duncan MacCannell |
| hCoV-19/USA/TX-HMH-MCoV-10119/2020 | EPI_ISL_1201907 | 2020-07-06 | Houston Methodist Hospital | Houston Methodist Hospital | S. Wesley Long, Randall J. Olsen, Paul A. Christensen, Sishir Subedi, Robert Olson, James J. Davis, Matthew Ojeda Saavedra, Prasanti Yerramilli, Layne Pruitt, Kristina Reppond, Madison N. Shyer, Jessica Cambric, Ilya J. Finkelstein, Jimmy Gollihar, and |
| hCoV-19/Belgium/CHUNamur1321085101/2021 | EPI_ISL_1220090 | 2021-03-07 | Plateforme de testing Namuroise | Plateforme de testing Namuroise | ; Otto Gaetan ; Denis Olivier ; Degosserie Jonathan ; Mullier Fran√ßois |
| hCoV-19/Bonaire/BQ-RIVM-15865/2021 | EPI_ISL_1232540 | 2021-02-19 | Dutch COVID-19 response team | National Institute for Public Health and the Environment (RIVM) | Adam Meijer, Harry Vennema, Dirk Eggink, Jeroen Cremer, Sharon van den Brink, Bas van der Veer, AnneMarie van den Brandt, Florian Zwagemaker, Dennis Schmitz, Chantal Reusken, on behalf of the national COVID-19 response team |
| hCoV-19/Switzerland/GR-ETHZ-510678/2021 | EPI_ISL_1259874 | 2021-02-23 | Viollier AG | Department of Biosystems Science and Engineering, ETH Z√ºrich | Christian Beisel, Sarah Nadeau, Chaoran Chen, Ivan Topolsky, Philipp Jablonski, Lara Fuhrmann, David Dreifuss, Katharina Jahn, Rebecca Denes, Mirjam Feldkamp, Ina Nissen, Natascha Santacroce, Elodie Burcklen, Christiane Beckmann, Maurice Redondo, Olivier |
| hCoV-19/Bonaire/BQ-RIVM-16951/2021 | EPI_ISL_1288882 | 2021-02-28 | Dutch COVID-19 response team | National Institute for Public Health and the Environment (RIVM) | Adam Meijer, Harry Vennema, Dirk Eggink, Jeroen Cremer, Sharon van den Brink, Bas van der Veer, AnneMarie van den Brandt, Florian Zwagemaker, Dennis Schmitz, Chantal Reusken, on behalf of the national COVID-19 response team |
| hCoV-19/Australia/QLD1739/2021 | EPI_ISL_1306138 | 2021-03-19 | Queensland Health Forensic and Scientific Services | Queensland Health Forensic and Scientific Services | Son Nguyen |
| hCoV-19/USA/GA-CDC-LC0017613/2021 | EPI_ISL_1160619 | 2021-02-16 | Laboratory Corporation of America | Respiratory Viruses Branch, Division of Viral Diseases, Centers for Disease Control and Prevention | Peter W. Cook, Dakota Howard, Dhwani Batra, Ben L. Rambo-Martin, Minoo Agarwal, Eyad Almasri Debbie Boles, Ayla Burns, Nuthawin Charoensri, Oren Cohen, Susan Countryman, Mary Ann Cristobal, Bobbi Croy, Suzanne Dale, Hrushikesh Deshmukh, Amanda Douglas, Vi |
| hCoV-19/Cameroon/Ebolowa-20v-13103/2020 | EPI_ISL_1001066 | 2020-05-12 | Centre Pasteur du Cameroun | Institut Pasteur de Dakar | Njouom Richard |

| | | | | | |
|---|---|---|---|---|---|
| hCoV-19/Aruba/AW-RIVM-11747/2021 | EPI_ISL_1014330 | 2021-02-01 | Dutch COVID-19 response team | National Institute for Public Health and the Environment (RIVM) | Adam Meijer, Harry Vennema, Dirk Eggink, Jeroen Cremer, Sharon van den Brink, Bas van der Veer, AnneMarie van den Brandt, Florian Zwagemaker, Dennis Schmitz, Chantal Reusken, on behalf of the national COVID-19 response team |
| hCoV-19/India/GJ-NIV-VOC-78/2020 | EPI_ISL_1034171 | 2020-12-24 | NIV Influenza | NIV Influenza | Potdar, Varsha on behalf of National Influenza Centre, NIV, Pune |
| hCoV-19/Belgium/ULG-12382/2021 | EPI_ISL_1076419 | 2021-02-14 | Department of Clinical Microbiology | GIGA Medical Genomics | Keith Durkin, Maria Artesi, S√©bastien Bontems, Rapha√´l Boreux, Bouchra Boujemla, Nathalie Renotte, C√©cile Meex, Pierrette Melin, Marie-Pierre Hayette, Vincent Bours |
| hCoV-19/USA/TX-HMH-MCoV-22928/2020 | EPI_ISL_1077458 | 2020-12-27 | Houston Methodist Hospital | Houston Methodist Hospital | S. Wesley Long, Randall J. Olsen, Paul A. Christensen, Sishir Subedi, Robert Olson, James J. Davis, Matthew Ojeda Saavedra, Prasanti Yerramilli, Layne Pruitt, Kristina Reppond, Madison N. Shyer, Jessica Cambric, Ilya J. Finkelstein, Jimmy Gollihar, and |
| hCoV-19/Belgium/rega-3424/2021 | EPI_ISL_1093292 | 2021-02-15 | KU Leuven, Rega Institute, Clinical and Epidemiological Virology | KU Leuven, Rega Institute, Clinical and Epidemiological Virology | Tony Wawina-Bokalanga, Bert Vanmechelen, Joan Marti-Carerras, Piet Maes |
| hCoV-19/USA/MD-CDC-LC0022432/2021 | EPI_ISL_1319921 | 2021-02-28 | Laboratory Corporation of America | Centers for Disease Control and Prevention Division of Viral Diseases, Pathogen Discovery | Peter W. Cook, Dakota Howard, Dhwani Batra, Ben L. Rambo-Martin, Minoo Agarwal, Eyad Almasri, Debbie Boles, Ayla Burns, Nuthawin Charoensri, Oren Cohen, Susan Countryman, Mary Ann Cristobal, Bobbi Croy, Suzanne Dale, Hrushikesh Deshmukh, Amanda Douglas, V |
| hCoV-19/Belgium/rega-4962/2021 | EPI_ISL_1382788 | 2021-02-25 | KU Leuven, Rega Institute, Clinical and Epidemiological Virology | KU Leuven, Rega Institute, Clinical and Epidemiological Virology | Tony Wawina-Bokalanga, Bert Vanmechelen, Joan Marti-Carerras, Piet Maes |
| hCoV-19/Argentina/PAIS-E0133/2020 | EPI_ISL_1395947 | 2020-07-14 | Laboratorio Central de Salud P√∫blica | [illegible] Bioinform√°tica del Instituto de Investigaci√≥n de la Cadena L√°ctea CONICET-INTA on behalf of 'Proyecto Argentino Interinstitucional de genomica de SARS-CoV-2' (PAIS | Natalia Andrea Ayala y Mar√≠a Ver√≥nica G√≥mez, Erica Struss, Esteban Paredes, Antonieta Cayr√©, Laura Lescano, Eberhardt, MF, Irazoqui, Amadio, AF |
| hCoV-19/Russia/KAM-RII-MH12214S/2020 | EPI_ISL_1400540 | 2020-12-15 | WHO National Influenza Centre Russian Federation | WHO National Influenza Centre Russian Federation | Andrey Komissarov, Artem Fadeev, Anna Ivanova, Kseniya Komissarova, Dmitry Bazhenov, Mikhail Bakaev, Daria Danilenko, Ksenia Safina, Elena Nabieva, Georgii Bazykin, Dmitry Lioznov |
| hCoV-19/Russia/SPE-RII-MH13423S/2020 | EPI_ISL_1400553 | 2020-12-24 | HELIX LLC | WHO National Influenza Centre Russian Federation | Andrey Komissarov, Artem Fadeev, Anna Ivanova, Kseniya Komissarova, Dmitry Bazhenov, Tamila Musaeva, Maria Timofeeva, Veronika Eder, Maria Pisareva, Daria Danilenko, Ksenia Safina, Elena Nabieva, Georgii Bazykin, Dmitry Lioznov |
| hCoV-19/Switzerland/VS-ETHZ-410100/2020 | EPI_ISL_1407057 | 2020-12-15 | Viollier AG | Department of Biosystems Science and Engineering, ETH Z√ºrich | Chaoran Chen, Sarah Nadeau, Catharine Aquino, Ivan Topolsky, Philipp Jablonski, Lara Fuhrmann, David Dreifuss, Katharina Jahn, Andreia Cabral de Gouvea, Maria Domenica Moccia, Simon Gr√ºter, Timothy Sykes, Lennart Opitz, Griffin White, Laura Neff, Doris P |

| | | | | | |
|---|---|---|---|---|---|
| hCoV-19/Malawi/KRISP-K010155/2020 | EPI_ISL_1407116 | 2020-11-25 | National HIV Reference Laboratory, Ministry of Health, Public Health Institute of Malawi | KRISP, KZN Research Innovation and Sequencing Platform | Mvula B, Chilima B, Chiwaula M, Mwangomba W, Panja L, Kasambara W, Auld A, Kim L, Kampira E, Kaba M, Wadonda N, Maida A, Giandhari J, Pillay S, Naidoo Y, Lessells R, Emmanuel SJ, Tegally H, Wilkinson E, de Oliveira T |
| hCoV-19/India/WB-1931300129338/2020 | EPI_ISL_1419278 | 2020-10-20 | INSACOG-WB | National Institute of Biomedical Genomics ,Äì INSACOG | Arindam Maitra, Bhaswati Bandyopadhyay, Nidhan Kumar Biswas, Tamal Ghosh, Sreedhar Chinnaswamy, Ajay Chakraborti, Saumitra Das |
| hCoV-19/India/WB-1931300076600/2020 | EPI_ISL_1419318 | 2020-09-03 | INSACOG-WB | National Institute of Biomedical Genomics ,Äì INSACOG | Arindam Maitra, Bhaswati Bandyopadhyay, Nidhan Kumar Biswas, Tamal Ghosh, Sreedhar Chinnaswamy, Ajay Chakraborti, Saumitra Das |
| hCoV-19/India/WB-1931300076580/2020 | EPI_ISL_1419334 | 2020-09-03 | INSACOG-WB | National Institute of Biomedical Genomics ,Äì INSACOG | Arindam Maitra, Bhaswati Bandyopadhyay, Nidhan Kumar Biswas, Tamal Ghosh, Sreedhar Chinnaswamy, Ajay Chakraborti, Saumitra Das |
| hCoV-19/USA/SC-CDC-STM-000026311/2021 | EPI_ISL_1420959 | 2021-02-28 | Helix/Illumina | Centers for Disease Control and Prevention Division of Viral Diseases, Pathogen Discovery | Peter W. Cook, Dakota Howard, Dhwani Batra, Ben L. Rambo-Martin, Eileen de Feo, Jan Antico, Christine Tran, Matthew Tolentino, Shannon Wickline, Kim Gietzen, Brad Sickler, Jingtao Liu, Eric Allen, Phil Febbo, Summer Galloway, Nicole L. Washington, Simon W |
| hCoV-19/India/MH-NCCS-DHW50406/2020 | EPI_ISL_1415298 | 2020-12-25 | National Centre For Cell Science | National Centre For Cell Science ,Äì INSACOG | Dhiraj Paul, Mitali Inamdar, Sonal Manik Chavan, Mohak P Gujare, Shivang P. Bhanushali, Manoj Kumar Bhat, Ajay Pillai, INSACOG Consortium team, Yogesh Shouche. |
| hCoV-19/Japan/PG-22654/2021 | EPI_ISL_1427333 | 2021-01-27 | Pathogen Genomics Center, National Institute of Infectious Diseases | Pathogen Genomics Center, National Institute of Infectious Diseases | Tsuyoshi Sekizuka, Kentaro Itokawa, Rina Tanaka, Masanori Hashino, Makoto Kuroda |
| hCoV-19/env/India/GJ-GBRC745a/2020 | EPI_ISL_2484895 | 2020-11-26 | Amdavad Municipal Corporation, Ahmedabad | Gujarat Biotechnology Research Centre | Manish Kumar,Twinkle Soni,Sonal Sharma ,Nitin Savaliya,Dinesh Kumar,Ramesh Pandit,Dalip Singh Rathore,Hiren Mandalia,Chaitanya Joshi,Madhvi Joshi |
| hCoV-19/Australia/QLD1795/2021 | EPI_ISL_1465881 | 2021-03-30 | Queensland Health Forensic and Scientific Services | Queensland Health Forensic and Scientific Services | Son Nguyen |
| hCoV-19/Australia/QLD1817/2021 | EPI_ISL_1483029 | 2021-04-03 | Queensland Health Forensic and Scientific Services | Queensland Health Forensic and Scientific Services | Son Nguyen |
| hCoV-19/USA/VA-CDC-LC0033407/2021 | EPI_ISL_1514467 | 2021-03-16 | Laboratory Corporation of America | Centers for Disease Control and Prevention Division of Viral Diseases, Pathogen Discovery | Dakota Howard, Dhwani Batra, Peter W. Cook, Kara Moser, Adrian Paskey, Jason Caravas, Benjamin Rambo-Martin, Shatavia Morrison, Christopher Gulvick, Scott Sammons, Yvette Unoarumhi, Darlene Wagner, Matthew Schmerer, Minoo Agarwal, Eyad Almasri, Debbie Bol |
| hCoV-19/Netherlands/GE-RIVM-21747/2021 | EPI_ISL_1521524 | 2021-03-17 | Dutch COVID-19 response team | National Institute for Public Health and the Environment (RIVM) | Adam Meijer, Harry Vennema, Dirk Eggink, Jeroen Cremer, Sharon van den Brink, Bas van der Veer, AnneMarie van den Brandt, Lisa Wijsman, Kim Freriks, Ryanne Jaarsma, Eun√≠ce Then, Jolienke Hardeman, Lynn Aarts, Sanne Bos, Melissa van Tuil, Robert Kohl, Lin |
| hCoV-19/Canada/Qc-L00313381001/2020 | EPI_ISL_1532723 | 2020-11-30 | Laboratoire de sant√© publique du Qu√©bec | Laboratoire de sant√© publique du Qu√©bec | Sandrine Moreira, Ioannis Ragoussis, Guillaume Bourque, Jesse Shapiro, Mark Lathrop and Michel Roger on behalf of the CoVSeQ research group |

| | | | | | |
|---|---|---|---|---|---|
| hCoV-19/India/MH-NCCS-P1162000182735/2021 | EPI_ISL_1544014 | 2021-02-27 | National Centre For Cell Science | National Centre For Cell Science ‚Äì INSACOG | Dhiraj Paul, Mohak P Gujare, Shivang P. Bhanushali, Mitali Inamdar, Sonal Manik Chavan, Manoj Kumar Bhat, Ajay Pillai, INSACOG Consortium team, Yogesh Shouche |
| hCoV-19/USA/FL-CDC-FG-011257/2021 | EPI_ISL_1555352 | 2021-03-06 | Fulgent Genetics | Centers for Disease Control and Prevention Division of Viral Diseases, Pathogen Discovery | Dakota Howard, Dhwani Batra, Peter W. Cook, Kara Moser, Adrian Paskey, Jason Caravas, Benjamin Rambo-Martin, Shatavia Morrison, Christopher Gulvick, Scott Sammons, Yvette Unoarumhi, Darlene Wagner, Matthew Schmerer, Harry Gao, Mickey Li, John Gao, Joseph |
| hCoV-19/Australia/VIC1665/2020 | EPI_ISL_456566 | 2020-05-09 | Victorian Infectious Diseases Reference Laboratory (VIDRL) | Health Laboratory and Victorian Infectious Diseases Reference Laboratory, Doherty Institute | Caly L., Seemann T., Sait, M., Schultz M., Druce J., Sherry, N. |
| hCoV-19/Australia/VIC2258/2020 | EPI_ISL_521987 | 2020-06-24 | Victorian Infectious Diseases Reference Laboratory (VIDRL) | VIDRL and MDU-PHL | Caly L., Seemann T., Sait, M., Schultz M., Druce J., Sherry, N. |
| hCoV-19/India/GJ-GBRC88b/2020 | EPI_ISL_447550 | 2020-05-03 | GMERS Medical College and Hospital, Gandhinagar | Gujarat Biotechnology Research Centre | Akanksha Verma, Nitin Savaliya, Raghawendra Kumar, Dinesh Kumar, Zuber Saiyed, Dipa Kinariwala, Disha Patel, Binita Aring, Neeta Khandelwal, Geeta Vaghela, Sonia Barve, Bhavesh Modi, Kairavi Joshi, Gaurishankar Shrimali, Nidhi Sood, Pranay Shah, R D Dixit |
| hCoV-19/Australia/NSW552/2020 | EPI_ISL_455091 | 2020-05-13 | South Eastern Area Laboratory Services | Clinical Pathology and Medical Research; Westmead Hospital; University of Sydney | CIDM-PH et al. |
| hCoV-19/Australia/NSW554/2020 | EPI_ISL_455093 | 2020-05-20 | South Eastern Area Laboratory Services | Clinical Pathology and Medical Research; Westmead Hospital; University of Sydney | CIDM-PH et al. |
| hCoV-19/England/NORW-EC6EA/2020 | EPI_ISL_457552 | 2020-05-11 | Quadram Institute Bioscience | COVID-19 Genomics UK (COG-UK) Consortium | Dave J. Baker, Gemma L. Kay, Alp Aydin, Thanh Le-Viet, Steven Rudder, Ana P. Tedim, Anastasia Kolyva, Maria Diaz, Leonardo de Oliveira Martins, Nabil-Fareed Alikhan, Lizzie Meadows, Rachael Stanley, Ngozi Elumogo, Muhammed Yasir, Nicholas M. Thomson, Alex |
| hCoV-19/India/OR-ILSCV14743/2020 | EPI_ISL_463032 | 2020-05-10 | Institute of Life Sciences, Bhubaneswar | Immunogenomics lab, Institute of Life Sciences, Bhubaneswar | Sunil Raghav, Arup Ghosh, Atimukta Jha, Viplov K. Biswas, Swati Madhulika, Manasi Priyadarshini, Shuchi Smita, O. P. Shriwas, Priyanka Mohapatra, Satya Ranjan Sahu, Aliva Minz, Debyashrita Barik, Rupesh Dash, Soma Chattopadhyay, Ghulam Hussain Syed, Shant |
| hCoV-19/England/LIVE-9FC5D/2020 | EPI_ISL_472051 | 2020-06-01 | Liverpool Clinical Laboratories | COVID-19 Genomics UK (COG-UK) Consortium | Sam Haldenby, Anita Lucaci, Steve Paterson, Julian Hiscox, Alistair Darby, M Almsaud, A Alrezaihi, Muhannad Alruwaili, Stuart D Armstrong, Jones Benjamin, Eleanor G Bentley, Anu Chawla, Jordan J Clark, Angela Cowell, Richard Eccles, Isabel Garc√≠a-Dorival |
| hCoV-19/Senegal/57065/2020 | EPI_ISL_476148 | 2020-06-09 | Institut Pasteur Dakar | Institut Pasteur de Dakar | Ndongo Dia, Moussa Moise Diagne, Mamadou Diop, Ousmane Faye, Amadou Alpha Sall |
| hCoV-19/Argentina/Heritas_HG019/2020 | EPI_ISL_476573 | 2020-05-22 | Hospital de Pediatr√≠a "Prof. Dr. Juan P Garrahan" | H√©ritas | Dalmacio Pereyra, Roberta Crespo, Mauricio Grisolia, Cristian Rohr, Andrea Mangano, Maria Florencia Fernandez, Fabian Fay, Martin Vazquez |

| | | | | | |
|---|---|---|---|---|---|
| hCoV-19/Bangladesh/CHRF-0031/2020 | EPI_ISL_477140 | 2020-06-16 | Child Health Research Foundation | Child Health Research Foundation | Senjuti Saha, Md Saiful Islam Sajib, Roly Malaker, Md Hafizur Rahman, Afroza Akter Tanni, Syed Muktadir Al Sium, Maksuda Islam, Samir K Saha |
| hCoV-19/Egypt/CUNCI-HGC3I05/2020 | EPI_ISL_479690 | 2020-06-02 | Egyptian National Cancer Institute (ENCI) | Egyptian National Cancer Institute (ENCI) | Zekri, Abdel Rahman N, Amer,K.E., Ahmed,O.S., Soliman,H.K., Hafez,M.M., Bahnassy,A.A., Abdelhamid,W., Gad,A., Ali,M., Hassan,W., Samir,M., Raouf,A., Hamdy,M.S., Soliman,M.S., Elsissy,M.H., Elkhateeb,S.M., Ezzelarab,M.H., Abouelhoda, Mohamed |
| hCoV-19/Egypt/CUNCI-HGC5I012/2020 | EPI_ISL_479713 | 2020-06-02 | Egyptian National Cancer Institute (ENCI) | Egyptian National Cancer Institute (ENCI) | Zekri, Abdel Rahman N, Amer,K.E., Ahmed,O.S., Soliman,H.K., Hafez,M.M., Bahnassy,A.A., Abdelhamid,W., Gad,A., Ali,M., Hassan,W., Samir,M., Raouf,A., Hamdy,M.S., Soliman,M.S., Elsissy,M.H., Elkhateeb,S.M., Ezzelarab,M.H., Abouelhoda, Mohamed |
| hCoV-19/England/HECH-2599C94/2020 | EPI_ISL_482092 | 2020-05-05 | Microbiology Department, Hereford County Hospital | Wellcome Sanger Institute for the COVID-19 Genomics UK (COG-UK) consortium | Alison Johnson, Venkat Sivaprakasam, Fenella Halstead, Jane Thomas, Wendy Hogsden, Samantha Lamb and Alex Alderton, Roberto Amato, Sonia Goncalves, Ewan Harrison, David K. Jackson, Ian Johnston, Dominic Kwiatkowski, Cordelia Langford, John Sillitoe on beh |
| hCoV-19/England/NORT-296410/2020 | EPI_ISL_488017 | 2020-05-13 | NU-OMICS DNA Sequencing research facility, Northumbria University | Wellcome Sanger Institute for the COVID-19 Genomics UK (COG-UK) consortium | Chris Duncan, Sheia Waugh, Shirelle Burton-Fanning, Gary Eltringham, Jennifer Collins, Brendan Payne, Yusri Taha, Emma Swindells, Jane Greenaway, Edward Barton, Garren Scott, Debra Padgett, Clive Graham, Sarah Essex, Steve Ligget, Paul Baker, Lynn Dover, |
| hCoV-19/USA/FL-BPHL-0421/2020 | EPI_ISL_489830 | 2020-05-13 | Florida Bureau of Public Health Laboratories | Florida Bureau of Public Health Laboratories | Sarah Schmedes, Jason Blanton |
| hCoV-19/Australia/NSW613/2020 | EPI_ISL_490022 | 2020-06-07 | South Eastern Area Laboratory Services (SEALS) | Clinical Pathology and Medical Research; Westmead Hospital; University of Sydney | CIDM-PH et al. |
| hCoV-19/England/LIVE-A3904/2020 | EPI_ISL_490389 | 2020-06-09 | Liverpool Clinical Laboratories | COVID-19 Genomics UK (COG-UK) Consortium | Sam Haldenby, Anita Lucaci, Steve Paterson, Julian Hiscox, Alistair Darby, M Almsaud, A Alrezaihi, Muhannad Alruwaili, Stuart D Armstrong, Jones Benjamin, Eleanor G Bentley, Anu Chawla, Jordan J Clark, Angela Cowell, Richard Eccles, Isabel Garc√≠a-Dorival |
| hCoV-19/Ecuador/USFQ-139/2020 | EPI_ISL_491936 | 2020-06-30 | Institute of Microbiology, Universidad San Francisco de Quito | Institute of Microbiology, Universidad San Francisco de Quito | Bel√©n Prado-Vivar, Sully M√°rquez, Juan Jos√© Guadalupe, Monica Becerra-Wong, Bernardo Guti√©rrez, Carlos Guerrero, Ver√≥nica Barrag√°n, Patricio Rojas-Silva, Gabriel Trueba, Michelle Grunauer, Pa√∫l C√°rdenas |
| hCoV-19/India/TG-CCMB-M81/2020 | EPI_ISL_495246 | 2020-06-05 | CSIR-Centre for Cellular and Molecular Biology | CSIR-Centre for Cellular and Molecular Biology | Lamuk Zaveri, Shagufta Khan, Namami Gaur, Sakshi Shambhavi, Nikhil Hajirnis, M Soujanya Reddy, Pratheusa Maccha, Tulasi Nagabandi, Purushotham Vodnala, Payel Mukherjee, Sofia Banu, Priya Singh, Onkar Kulkarni, Dhiviya Vedagiri, Divya Gupta, Vishal Sah, Sa |
| hCoV-19/USA/WA-UW-12727/2020 | EPI_ISL_501117 | 2020-06-29 | University of Washington Virology Lab | University of Washington Virology Lab | Pavitra Roychoudhury, Hong Xie, Lasata Shrestha, Amin Addetia, Truong Nguyen, Victoria M Rachleff, Meei-Li Huang, Keith R Jerome, Alexander Greninger |

| | | | | | |
|---|---|---|---|---|---|
| hCoV-19/USA/NM-UNM-00389/2020 | EPI_ISL_507988 | 2020-05-05 | New Mexico Department of Health Scientific Laboratory Division | Center for Global Health, University of New Mexico Health Sciences Center | Daryl Domman, Kurt Schwalm, Twila Kunde, Joseph Hicks, Michael Edwards, Darrell Dinwiddie |
| hCoV-19/SouthAfrica/KRISP-K001054/2020 | EPI_ISL_509312 | 2020-07-18 | NHLS-IALCH | KRISP, KZN Research Innovation and Sequencing Platform | Giandhari J, Pillay S, Lessells R, Mdlalose K, York D, Tegally H, Wilkinson E, de Oliveira T |
| hCoV-19/Australia/WA67/2020 | EPI_ISL_512720 | 2020-06-04 | PathWest Laboratory Medicine WA | PathWest Laboratory Medicine WA Microbial Surveillance Unit | PathWest Laboratory Medicine WA Microbial Surveillance Unit |
| hCoV-19/Australia/WA71/2020 | EPI_ISL_512724 | 2020-07-04 | PathWest Laboratory Medicine WA | PathWest Laboratory Medicine WA Microbial Surveillance Unit | PathWest Laboratory Medicine WA Microbial Surveillance Unit |
| hCoV-19/SaudiArabia/KAUST-JEDDAH1010/2020 | EPI_ISL_512881 | 2020-06-17 | Pathogen Genomics Lab King Abdullah University of Science and Technology(KAUST) | Pathogen Genomics Lab King Abdullah University of Science and Technology(KAUST) | Raeece Naeem, Rahul P Salunke, Sharif Hala, Sara Mfarrej, Amit Kumar Subudhi, Fadwa Alofi, Fathia Ben Rached, Afrah Alsomali, Asim Khogeer, Ahmad Bakur Mahmoud, Anwar Hashem, Naif Almontashiri, Arnab Pain |
| hCoV-19/Brazil/RJ-INCA-I44/2020 | EPI_ISL_513580 | 2020-05-04 | Programa de Oncovirologia, Instituto Nacional de C√¢ncer | Programa de Oncovirologia, Instituto Nacional de C√¢ncer | Juliana D. Siqueira, Livia R. Goes, Brunna M. Alves, Claudia Cicala,James Arthos, Jo√£o P.B. Viola, Andreia C. de Melo, Marcelo A. Soares |
| hCoV-19/Israel/CVL4659/2020 | EPI_ISL_516916 | 2020-06-16 | Israel Central Virology laboratory | Israel Central Virology laboratory | Neta Zuckerman, Efrat Dahan Bucris, Oran Erster, Ella Mendelson, Michal Mandelboim |
| hCoV-19/SouthKorea/KCDC2709/2020 | EPI_ISL_522488 | 2020-07-08 | Division of Viral Diseases, Center for Laboratory Control of Infectious Diseases, Korea Centers for Diseases Control and Prevention | Division of Viral Diseases, Center for Laboratory Control of Infectious Diseases, Korea Centers for Diseases Control and Prevention | Jeong-Min Kim, Yoon-Seok Chung, Namjoo Lee, Sang Hee Woo, Hye-Jun Jo, Heui Man Kim, Jun-Sub Kim, Myung Guk Han |
| hCoV-19/Netherlands/OV-EMC-60/2020 | EPI_ISL_523444 | 2020-05-24 | Dutch COVID-19 response team | Erasmus Medical Center | Bas Oude Munnink, David Nieuwenhuijse, Reina Sikkema, Claudia Schapendonk, Irina Chestakova, Anne van der Linden, Theo Bestebroer, Stefan van Nieuwkoop, Mark Pronk, Pascal Lexmond, Corien Swaan, Manon Haverkate, Madelief Mollers, Mart Stein, Sandra Kengne |
| hCoV-19/Netherlands/OV-EMC-80/2020 | EPI_ISL_523464 | 2020-07-10 | Dutch COVID-19 response team | Erasmus Medical Center | Bas Oude Munnink, David Nieuwenhuijse, Reina Sikkema, Claudia Schapendonk, Irina Chestakova, Anne van der Linden, Theo Bestebroer, Stefan van Nieuwkoop, Mark Pronk, Pascal Lexmond, Corien Swaan, Manon Haverkate, Madelief Mollers, Mart Stein, Sandra Kengne |
| hCoV-19/Netherlands/OV-EMC-81/2020 | EPI_ISL_523465 | 2020-07-10 | Dutch COVID-19 response team | Erasmus Medical Center | Bas Oude Munnink, David Nieuwenhuijse, Reina Sikkema, Claudia Schapendonk, Irina Chestakova, Anne van der Linden, Theo Bestebroer, Stefan van Nieuwkoop, Mark Pronk, Pascal Lexmond, Corien Swaan, Manon Haverkate, Madelief Mollers, Mart Stein, Sandra Kengne |
| hCoV-19/Netherlands/UT-EMC-57/2020 | EPI_ISL_523534 | 2020-07-11 | Dutch COVID-19 response team | Erasmus Medical Center | Bas Oude Munnink, David Nieuwenhuijse, Reina Sikkema, Claudia Schapendonk, Irina Chestakova, Anne van der Linden, Theo Bestebroer, Stefan van Nieuwkoop, Mark Pronk, Pascal Lexmond, Corien Swaan, Manon Haverkate, Madelief Mollers, Mart Stein, Sandra Kengne |

| | | | | | |
|---|---|---|---|---|---|
| hCoV-19/Netherlands/UT-EMC-62/2020 | EPI_ISL_523539 | 2020-08-02 | Dutch COVID-19 response team | Erasmus Medical Center | Bas Oude Munnink, David Nieuwenhuijse, Reina Sikkema, Claudia Schapendonk, Irina Chestakova, Anne van der Linden, Theo Bestebroer, Stefan van Nieuwkoop, Mark Pronk, Pascal Lexmond, Corien Swaan, Manon Haverkate, Madelief Mollers, Mart Stein, Sandra Kengne |
| hCoV-19/DRC/11676/2020 | EPI_ISL_527531 | 2020-05-26 | Viral Respiratory Lab, National Institute for Biomedical Research (INRB) | Pathogen Sequencing Lab, National Institute for Biomedical Research (INRB) | Placide Mbala-Kingebeni, Edith Nkwembe, Eddy Kinganda-Lusamaki, Amuri Aziza, Francisca Muyembe Mawete, Emmanuel Lokilo Lofiko, Catherine Pratt, Matthias Pauthner, Josh Quick, Allison Black, James Hadfield, Trevor Bedford, Ian Goodfellow, Andrew Rambaut, N |
| hCoV-19/SouthAfrica/KRISP-K002738/2020 | EPI_ISL_529736 | 2020-08-14 | NHLS-IALCH | KRISP, KZN Research Innovation and Sequencing Platform | Giandhari J, Pillay S, Lessells R, Mdlalose K, York D, Khan S, Tegally H, Wilkinson E, de Oliveira T |
| hCoV-19/SouthAfrica/KRISP-K002785/2020 | EPI_ISL_529779 | 2020-08-24 | NHLS-IALCH | KRISP, KZN Research Innovation and Sequencing Platform | Giandhari J, Pillay S, Lessells R, Mdlalose K, York D, Khan S, Tegally H, Wilkinson E, de Oliveira T |
| hCoV-19/USA/WA-S2782/2020 | EPI_ISL_530139 | 2020-08-10 | Seattle Flu Study | Seattle Flu Study | Deborah A. Nickerson, Chris D. Frazar, Jover Lee, Benjamin Pelle, Matthew Richardson, Amanda Adler, Elisabeth Brandstetter, Peter D. Han, Kairsten Fay, Misja Ilcisin, Kirsten Lacombe, Thomas R. Sibley, Melissa Truong, Caitlin R. Wolf, Karen Cowgill, Steph |
| hCoV-19/England/QEUH-99046A/2020 | EPI_ISL_530374 | 2020-08-26 | Lighthouse Lab in Glasgow | Wellcome Sanger Institute for the COVID-19 Genomics UK (COG-UK) consortium | Harper VanSteenhouse, Yumi Kasai, David Gray, Carol Clugston, Anna Dominiczak and Alex Alderton, Roberto Amato, Sonia Goncalves, Ewan Harrison, David K. Jackson, Ian Johnston, Dominic Kwiatkowski, Cordelia Langford, John Sillitoe |
| hCoV-19/England/QEUH-556C99/2020 | EPI_ISL_532206 | 2020-05-11 | Lighthouse Lab in Glasgow | Wellcome Sanger Institute for the COVID-19 Genomics UK (COG-UK) consortium | Harper VanSteenhouse, Yumi Kasai, David Gray, Carol Clugston, Anna Dominiczak and Alex Alderton, Roberto Amato, Sonia Goncalves, Ewan Harrison, David K. Jackson, Ian Johnston, Dominic Kwiatkowski, Cordelia Langford, John Sillitoe |
| hCoV-19/Sweden/20-52637/2020 | EPI_ISL_534257 | 2020-08-21 | Laboratoriemedicin Vasternorrland | The Public Health Agency of Sweden | Anna-Malin Linde, Maria Lind Karlberg, Mattias Haukland, Reza Advani, Olov Svartstrom, Oskar Karlsson Lindsjo, Sandra Broddesson, Petra Edquist, Mia Brytting, Anna Risberg, Karin Tegmark-Wisell |
| hCoV-19/Switzerland/UR-ETHZ-260095/2020 | EPI_ISL_539415 | 2020-08-24 | Viollier AG | Department of Biosystems Science and Engineering, ETH Z√ºrich | Christian Beisel, Sarah Nadeau, Ivan Topolsky, Pedro Ferreira, Philipp Jablonski, Susana Posada-C√©spedes, Tobias Sch√§r, Ina Nissen, Natascha Santacroce, Elodie Burcklen, Christiane Beckmann, Maurice Redondo, Olivier Kobel, Christoph Noppen, Sophie Seide |
| hCoV-19/USA/TX-HMH-3180/2020 | EPI_ISL_544054 | 2020-06-11 | Houston Methodist Hospital | Houston Methodist Hospital | S. Wesley Long, Randall J. Olsen, Paul A. Christensen, David W. Bernard, James J. Davis, Maulik Shukla, Marcus Nguyen, Matthew Ojeda Saavedra, Concepcion C. Cantu, Prasanti Yerramilli, Layne Pruitt, Sishir Subedi, Hung-Che Kuo, Heather Hendrickson, Ghazal |

| | | | | | |
|---|---|---|---|---|---|
| hCoV-19/NewZealand/20VR3879/2020 | EPI_ISL_548145 | 2020-07-23 | Middlemore Hospital | Institute of Environmental Science and Research (ESR) | Xiaoyun Ren, Matt Storey, Nikki Freed, Muhammad Faisal, Jing Wang, Hermes Perez, Anja Werno, Antje van der Linden, Arlo Upton, Chris Mansell, David Hammer, Dragana Drinkovic, Gary McAuliffe, Hana Sofia Andersson, James Ussher, Jill Sherwood, Josh Freeman, |
| hCoV-19/England/ALDP-951FFB/2020 | EPI_ISL_551328 | 2020-06-09 | Lighthouse Lab in Alderley Park | Wellcome Sanger Institute for the COVID-19 Genomics UK (COG-UK) consortium | The Lighthouse Lab in Alderley Park and Alex Alderton, Roberto Amato, Sonia Goncalves, Ewan Harrison, David K. Jackson, Ian Johnston, Dominic Kwiatkowski, Cordelia Langford, John Sillitoe on behalf of the Wellcome Sanger Institute COVID-19 Surveillance Te |
| hCoV-19/England/MILK-97157B/2020 | EPI_ISL_553054 | 2020-08-18 | Lighthouse Lab in Milton Keynes | Wellcome Sanger Institute for the COVID-19 Genomics UK (COG-UK) consortium | The Lighthouse Lab in Milton Keynes and Alex Alderton, Roberto Amato, Sonia Goncalves, Ewan Harrison, David K. Jackson, Ian Johnston, Dominic Kwiatkowski, Cordelia Langford, John Sillitoe on behalf of the Wellcome Sanger Institute COVID-19 Surveillance Te |
| hCoV-19/England/MILK-971502/2020 | EPI_ISL_553060 | 2020-08-18 | Lighthouse Lab in Milton Keynes | Wellcome Sanger Institute for the COVID-19 Genomics UK (COG-UK) consortium | The Lighthouse Lab in Milton Keynes and Alex Alderton, Roberto Amato, Sonia Goncalves, Ewan Harrison, David K. Jackson, Ian Johnston, Dominic Kwiatkowski, Cordelia Langford, John Sillitoe on behalf of the Wellcome Sanger Institute COVID-19 Surveillance Te |
| hCoV-19/England/CAMC-94AC94/2020 | EPI_ISL_556802 | 2020-07-14 | Lighthouse Lab in Cambridge | Wellcome Sanger Institute for the COVID-19 Genomics UK (COG-UK) consortium | Rob Howes, The Lighthouse Lab in Cambridge and Alex Alderton, Roberto Amato, Sonia Goncalves, Ewan Harrison, David K. Jackson, Ian Johnston, Dominic Kwiatkowski, Cordelia Langford, John Sillitoe on behalf of the Wellcome Sanger Institute COVID-19 Surveill |
| hCoV-19/England/CAMC-9424EF/2020 | EPI_ISL_556886 | 2020-07-20 | Lighthouse Lab in Cambridge | Wellcome Sanger Institute for the COVID-19 Genomics UK (COG-UK) consortium | Rob Howes, The Lighthouse Lab in Cambridge and Alex Alderton, Roberto Amato, Sonia Goncalves, Ewan Harrison, David K. Jackson, Ian Johnston, Dominic Kwiatkowski, Cordelia Langford, John Sillitoe on behalf of the Wellcome Sanger Institute COVID-19 Surveill |
| hCoV-19/England/MILK-60D8C9/2020 | EPI_ISL_557866 | 2020-07-06 | Lighthouse Lab in Milton Keynes | Wellcome Sanger Institute for the COVID-19 Genomics UK (COG-UK) consortium | The Lighthouse Lab in Milton Keynes and Alex Alderton, Roberto Amato, Sonia Goncalves, Ewan Harrison, David K. Jackson, Ian Johnston, Dominic Kwiatkowski, Cordelia Langford, John Sillitoe on behalf of the Wellcome Sanger Institute COVID-19 Surveillance Te |
| hCoV-19/England/ALDP-5B57B5/2020 | EPI_ISL_558771 | 2020-06-19 | Lighthouse Lab in Alderley Park | Wellcome Sanger Institute for the COVID-19 Genomics UK (COG-UK) consortium | The Lighthouse Lab in Alderley Park and Alex Alderton, Roberto Amato, Sonia Goncalves, Ewan Harrison, David K. Jackson, Ian Johnston, Dominic Kwiatkowski, Cordelia Langford, John Sillitoe on behalf of the Wellcome Sanger Institute COVID-19 Surveillance Te |
| hCoV-19/Gambia/NPHL-1446/2020 | EPI_ISL_561230 | 2020-07-04 | MRCG at LSHTM Genomics lab | MRCG at LSHTM Genomics lab | Abdul Karim sesay, Abdoulie Kanteh, Jarra Manneh, Mariama Kujabi, Bakary Sanyang |
| hCoV-19/Australia/VIC10901/2020 | EPI_ISL_562434 | 2020-09-02 | Microbiological Diagnostic Unit - Public Health Laboratory (MDU-PHL) | MDU-PHL | Seemann, T., Schultz M. B., Sait, M., Sherry, N. |
| hCoV-19/Australia/VIC10984/2020 | EPI_ISL_562507 | 2020-08-01 | Microbiological Diagnostic Unit - Public Health Laboratory (MDU-PHL) | MDU-PHL | Seemann, T., Schultz M. B., Sait, M., Sherry, N. |

| | | | | | |
|---|---|---|---|---|---|
| hCoV-19/Peru/LIM-UPCH-0128/2020 | EPI_ISL_568524 | 2020-08-28 | Laboratorio de Referencia Nacional de Virus Respiratorios, Instituto Nacional de Salud Peru | Laboratorio de Gen√≥mica Microbiana, Universidad Peruana Cayetano Heredia | Pablo Tsukayama, Alejandra D√°vila-Barclay, Luis Gonz√°lez, Pedro E. Romero, Brenda Ayzanoa, Janet Huancachoque, Pool Marcos, Maribel Huaringa, Camila Castillo-Vilcahuaman, Guillermo Salvatierra |
| hCoV-19/Brazil/PE-FIOCRUZ-IAM939/2020 | EPI_ISL_572361 | 2020-05-03 | Laborat√≥rio Central de Sa√∫de P√∫blica do Estado de Pernambuco (LACEN-PE) | WallauLab, Aggeu Magalhaes Institute | Marcelo Henrique Santos Paiva, Duschinka Ribeiro Duarte Guedes, C√°ssia Docena, Matheus Filgueira Bezerra, Filipe Zimmer Dezordi, La√≠s Ceschini Machado, Larissa Krokovsky, Elisama Helvecio, Alexandre Freitas da Silva, Luydson Richardson Silva Vasconcelos |
| hCoV-19/England/MILK-973245/2020 | EPI_ISL_575819 | 2020-08-20 | Lighthouse Lab in Milton Keynes | Wellcome Sanger Institute for the COVID-19 Genomics UK (COG-UK) consortium | The Lighthouse Lab in Milton Keynes and Alex Alderton, Roberto Amato, Sonia Goncalves, Ewan Harrison, David K. Jackson, Ian Johnston, Dominic Kwiatkowski, Cordelia Langford, John Sillitoe on behalf of the Wellcome Sanger Institute COVID-19 Surveillance Te |
| hCoV-19/Scotland/QEUH-9AE77F/2020 | EPI_ISL_577371 | 2020-09-02 | University of Exeter | COVID-19 Genomics UK (COG-UK) Consortium | Ben Temperton,Aaron Jeffries,Michelle Michelsen,Joanna Warwick-Dugdale,Audrey Farbos,Robyn Manley,Stephen Michell,Jane Masoli |
| hCoV-19/Australia/NSW1068/2020 | EPI_ISL_593708 | 2020-08-01 | South Eastern Area Laboratory Services (SEALS) | Clinical Pathology and Medical Research; Westmead Hospital; University of Sydney | CIDM-PH et al. |
| hCoV-19/Australia/NSW1110/2020 | EPI_ISL_593725 | 2020-08-29 | South Eastern Area Laboratory Services (SEALS) | Clinical Pathology and Medical Research; Westmead Hospital; University of Sydney | CIDM-PH et al. |
| hCoV-19/England/BIRM-6529A/2020 | EPI_ISL_594544 | 2020-09-30 | University of Birmingham | COVID-19 Genomics UK (COG-UK) Consortium | Institute of Microbiology, University of Birmingham: Claire McMurray, Joanne Stockton, Samuel Nicholls, Radoslaw Poplawski, Will Rowe, Josh Quick, Nicholas Loman. University of Birmingham Testing Laboratory: Celina M Whalley, Andrew Bosworth, Charlotte P |
| hCoV-19/Wales/PHWC-47EA83/2020 | EPI_ISL_595545 | 2020-10-09 | Wales Specialist Virology Centre Sequencing lab: Pathogen Genomics Unit | COVID-19 Genomics UK (COG-UK) Consortium | Catherine Moore, Johnathan Evans, Laura Gifford, Malorie Perry, Simon Cottrell, Angela Marchbank, Alec Birchley, Alexander Adams, Amy Gaskin, Bree Gatica-Wilcox, Jason Coombes, Joel Southgate, Lauren Gilbert, Lee Graham, Nicole Pacchiarini, Sara Kumziene- |
| hCoV-19/USA/WA-S2874/2020 | EPI_ISL_596405 | 2020-09-22 | Seattle Flu Study | Seattle Flu Study | Deborah A. Nickerson, Chris D. Frazar, Jover Lee, Benjamin Pelle, Matthew Richardson, Amanda Adler, Elisabeth Brandstetter, Peter D. Han, Kairsten Fay, Misja Ilcisin, Kirsten Lacombe, Thomas R. Sibley, Melissa Truong, Caitlin R. Wolf, Karen Cowgill, Steph |
| hCoV-19/Palestine/2/2020 | EPI_ISL_596514 | 2020-07-15 | Palestinian Ministry of Health | Molecular Genetics Lab | Nouar Qutob, Zaidoun Salah, Damien Richard, Hisham Darwish, Husam Sallam, Issa Shtayeh, Osama Najjar, Mahmoud Ruzayqat, Dana Najjar, Francois Balloux, Lucy van Dorp |
| hCoV-19/Palestine/39/2020 | EPI_ISL_596521 | 2020-07-27 | Palestinian Ministry of Health | Molecular Genetics Lab | Nouar Qutob, Zaidoun Salah, Damien Richard, Hisham Darwish, Husam Sallam, Issa Shtayeh, Osama Najjar, Mahmoud Ruzayqat, Dana Najjar, Francois Balloux, Lucy van Dorp |

| | | | | | |
|---|---|---|---|---|---|
| hCoV-19/Palestine/40/2020 | EPI_ISL_596523 | 2020-08-15 | Palestinian Ministry of Health | Molecular Genetics Lab | Nouar Qutob, Zaidoun Salah, Damien Richard, Hisham Darwish, Husam Sallam, Issa Shtayeh, Osama Najjar, Mahmoud Ruzayqat, Dana Najjar, Francois Balloux, Lucy van Dorp |
| hCoV-19/Palestine/42/2020 | EPI_ISL_596525 | 2020-08-17 | Palestinian Ministry of Health | Molecular Genetics Lab | Nouar Qutob, Zaidoun Salah, Damien Richard, Hisham Darwish, Husam Sallam, Issa Shtayeh, Osama Najjar, Mahmoud Ruzayqat, Dana Najjar, Francois Balloux, Lucy van Dorp |
| hCoV-19/Palestine/45/2020 | EPI_ISL_596528 | 2020-08-17 | Palestinian Ministry of Health | Molecular Genetics Lab | Nouar Qutob, Zaidoun Salah, Damien Richard, Hisham Darwish, Husam Sallam, Issa Shtayeh, Osama Najjar, Mahmoud Ruzayqat, Dana Najjar, Francois Balloux, Lucy van Dorp |
| hCoV-19/Palestine/66/2020 | EPI_ISL_596543 | 2020-06-23 | Palestinian Ministry of Health | Molecular Genetics Lab | Nouar Qutob, Zaidoun Salah, Damien Richard, Hisham Darwish, Husam Sallam, Issa Shtayeh, Osama Najjar, Mahmoud Ruzayqat, Dana Najjar, Francois Balloux, Lucy van Dorp |
| hCoV-19/Palestine/75/2020 | EPI_ISL_596551 | 2020-07-10 | Palestinian Ministry of Health | Molecular Genetics Lab | Nouar Qutob, Zaidoun Salah, Damien Richard, Hisham Darwish, Husam Sallam, Issa Shtayeh, Osama Najjar, Mahmoud Ruzayqat, Dana Najjar, Francois Balloux, Lucy van Dorp |
| hCoV-19/Palestine/78/2020 | EPI_ISL_596554 | 2020-06-27 | Palestinian Ministry of Health | Molecular Genetics Lab | Nouar Qutob, Zaidoun Salah, Damien Richard, Hisham Darwish, Husam Sallam, Issa Shtayeh, Osama Najjar, Mahmoud Ruzayqat, Dana Najjar, Francois Balloux, Lucy van Dorp |
| hCoV-19/Scotland/QEUH-9FB1FD/2020 | EPI_ISL_600560 | 2020-09-29 | Lighthouse Lab in Glasgow | Wellcome Sanger Institute for the COVID-19 Genomics UK (COG-UK) consortium | Harper VanSteenhouse, Yumi Kasai, David Gray, Carol Clugston, Anna Dominiczak and Alex Alderton, Roberto Amato, Sonia Goncalves, Ewan Harrison, David K. Jackson, Ian Johnston, Dominic Kwiatkowski, Cordelia Langford, John Sillitoe on behalf of the Wellcome |
| hCoV-19/England/MILK-9E05B3/2020 | EPI_ISL_601443 | 2020-09-20 | Lighthouse Lab in Milton Keynes | Wellcome Sanger Institute for the COVID-19 Genomics UK (COG-UK) consortium | The Lighthouse Lab in Milton Keynes and Alex Alderton, Roberto Amato, Sonia Goncalves, Ewan Harrison, David K. Jackson, Ian Johnston, Dominic Kwiatkowski, Cordelia Langford, John Sillitoe on behalf of the Wellcome Sanger Institute COVID-19 Surveillance Te |
| hCoV-19/SouthAfrica/KRISP-K003532/2020 | EPI_ISL_602850 | 2020-09-23 | NHLS-IALCH | KRISP, KZN Research Innovation and Sequencing Platform | Giandhari J, Pillay S, Lessells R, Mdlalose K, York D, Khan S, Tegally H, Wilkinson E, de Oliveira T |
| hCoV-19/Italy/LAZ-INMI-61/2020 | EPI_ISL_603168 | 2020-06-07 | INMI Lazzaro Spallanzani IRCCS | INMI Lazzaro Spallanzani IRCCS | Martina Rueca, Barbara Bartolini, Cesare E.M. Gruber, Simone Lanini, Emanuela Giombini, Francesco Messina, Fulvia Pimpinelli, Antonino Di Caro, Maria R. Capobianchi |
| hCoV-19/England/MILK-A7AA90/2020 | EPI_ISL_606414 | 2020-10-15 | Lighthouse Lab in Milton Keynes | Wellcome Sanger Institute for the COVID-19 Genomics UK (COG-UK) consortium | The Lighthouse Lab in Milton Keynes and Alex Alderton, Roberto Amato, Sonia Goncalves, Ewan Harrison, David K. Jackson, Ian Johnston, Dominic Kwiatkowski, Cordelia Langford, John Sillitoe on behalf of the Wellcome Sanger Institute COVID-19 Surveillance Te |

| | | | | | |
|---|---|---|---|---|---|
| hCoV-19/USA/NM-UNM-00830/2020 | EPI_ISL_610268 | 2020-05-04 | New Mexico Department of Health Scientific Laboratory Division | Center for Global Health, University of New Mexico Health Sciences Center | Daryl Domman, Kurt Schwalm, Twila Kunde, Joseph Hicks, Michael Edwards, Darrell Dinwiddie |
| hCoV-19/NewZealand/20CV0463/2020 | EPI_ISL_622776 | 2020-10-20 | Middlemore Hospital | Institute of Environmental Science and Research (ESR) | Xiaoyun Ren, Matt Storey, Nikki Freed, Muhammad Faisal, Jing Wang, Hermes Perez, Anja Werno, Antje van der Linden, Arlo Upton, Chris Mansell, David Hammer, Dragana Drinkovic, Gary McAuliffe, Hana Sofia Andersson, James Ussher, Jill Sherwood, Josh Freeman, |
| hCoV-19/NewZealand/20CV0518/2020 | EPI_ISL_622786 | 2020-11-02 | Canterbury Health Laboratories | Institute of Environmental Science and Research (ESR) | Xiaoyun Ren, Matt Storey, Nikki Freed, Muhammad Faisal, Jing Wang, Hermes Perez, Anja Werno, Antje van der Linden, Arlo Upton, Chris Mansell, David Hammer, Dragana Drinkovic, Gary McAuliffe, Hana Sofia Andersson, James Ussher, Jill Sherwood, Josh Freeman, |
| hCoV-19/NewZealand/20CV0499/2020 | EPI_ISL_622827 | 2020-10-25 | Canterbury Health Laboratories | Institute of Environmental Science and Research (ESR) | Xiaoyun Ren, Matt Storey, Nikki Freed, Muhammad Faisal, Jing Wang, Hermes Perez, Anja Werno, Antje van der Linden, Arlo Upton, Chris Mansell, David Hammer, Dragana Drinkovic, Gary McAuliffe, Hana Sofia Andersson, James Ussher, Jill Sherwood, Josh Freeman, |
| hCoV-19/Denmark/DCGC-8944/2020 | EPI_ISL_626168 | 2020-10-26 | Department of Virus and Microbiological Special Diagnostics, Statens Serum Institut, Denmark | Albertsen lab, Department of Chemistry and Bioscience, Aalborg University, Denmark | Danish Covid-19 Genome Consortia |
| hCoV-19/NewZealand/20CV0515/2020 | EPI_ISL_626353 | 2020-10-30 | LabPLUS | Institute of Environmental Science and Research (ESR) | Xiaoyun Ren, Matt Storey, Nikki Freed, Muhammad Faisal, Jing Wang, Hermes Perez, Anja Werno, Antje van der Linden, Arlo Upton, Chris Mansell, David Hammer, Dragana Drinkovic, Gary McAuliffe, Hana Sofia Andersson, James Ussher, Jill Sherwood, Josh Freeman, |
| hCoV-19/England/MILK-AC8E97/2020 | EPI_ISL_629533 | 2020-10-22 | Lighthouse Lab in Milton Keynes | Wellcome Sanger Institute for the COVID-19 Genomics UK (COG-UK) consortium | The Lighthouse Lab in Milton Keynes and Alex Alderton, Roberto Amato, Sonia Goncalves, Ewan Harrison, David K. Jackson, Ian Johnston, Dominic Kwiatkowski, Cordelia Langford, John Sillitoe on behalf of the Wellcome Sanger Institute COVID-19 Surveillance Te |
| hCoV-19/USA/NY-NYCPHL-000473/2020 | EPI_ISL_631942 | 2020-05-02 | St Barnabas Hospital | New York City Public Health Laboratory | Jade Wang, et al. |
| hCoV-19/Netherlands/UT-EMC-115/2020 | EPI_ISL_632638 | 2020-10-12 | Dutch COVID-19 response team | Erasmus Medical Center | Bas Oude Munnink, David Nieuwenhuijse, Reina Sikkema, Claudia Schapendonk, Irina Chestakova, Anne van der Linden, Theo Bestebroer, Stefan van Nieuwkoop, Mark Pronk, Pascal Lexmond, Corien Swaan, Manon Haverkate, Madelief Mollers, Mart Stein, Sandra Kengne |
| hCoV-19/SouthAfrica/KRISP-K004006/2020 | EPI_ISL_634991 | 2020-09-29 | National Health Laboratory Service - Inkosi Albert Luthuli Central Hospital (NHLS-IALCH) | KRISP, KZN Research Innovation and Sequencing Platform | Giandhari J, Pillay S, Lessells R, Mdlalose K, York D, Khan S, Tegally H, Wilkinson E, de Oliveira T |
| hCoV-19/England/ALDP-9E89CE/2020 | EPI_ISL_638065 | 2020-09-24 | Centre for Enzyme Innovation, University of Portsmouth / Translational Research Laboratory, Portsmouth Hospitals NHS Trust | COVID-19 Genomics UK (COG-UK) Consortium | Angela Beckett,Yann Bourgeois,Garry Scarlett,Sharon Glaysher,Scott Elliott,Kelly Bicknell,Robert Impey,Allyson Lloyd,Sarah Wyllie,Ethan Butcher,Anoop Chauhan,Samuel Robson |

| | | | | | |
|---|---|---|---|---|---|
| hCoV-19/England/QEUH-A79220/2020 | EPI_ISL_638790 | 2020-10-12 | Oxford Viromics, NDM, University of Oxford; Oxford University Hospitals; Basingstoke and North Hampshire Hospital | COVID-19 Genomics UK (COG-UK) Consortium | Tanya Golubchik, David Bonsall, George Macintyre, Amy Trebes, Mariateresa de Cesare, Catrin Moore, Alex Mobbs, Anita Justice, Robert Shaw, Monique Andersson, Timothy Peto, Emma Wise, Nathan Moore, Jessica Lynch, Nick Cortes, Matilde Mori, Stephen Kidd, Da |
| hCoV-19/England/CAMC-B07C19/2020 | EPI_ISL_643684 | 2020-10-29 | Lighthouse Lab in Cambridge | Wellcome Sanger Institute for the COVID-19 Genomics UK (COG-UK) Consortium | Rob Howes, The Lighthouse Lab in Cambridge and Alex Alderton, Roberto Amato, Sonia Goncalves, Ewan Harrison, David K. Jackson, Ian Johnston, Dominic Kwiatkowski, Cordelia Langford, John Sillitoe on behalf of the Wellcome Sanger Institute COVID-19 Surveill |
| hCoV-19/England/CAMC-B0738D/2020 | EPI_ISL_643784 | 2020-10-29 | Lighthouse Lab in Cambridge | Wellcome Sanger Institute for the COVID-19 Genomics UK (COG-UK) Consortium | Rob Howes, The Lighthouse Lab in Cambridge and Alex Alderton, Roberto Amato, Sonia Goncalves, Ewan Harrison, David K. Jackson, Ian Johnston, Dominic Kwiatkowski, Cordelia Langford, John Sillitoe on behalf of the Wellcome Sanger Institute COVID-19 Surveill |
| hCoV-19/NewZealand/20CV0554/2020 | EPI_ISL_649122 | 2020-11-09 | Middlemore Hospital | Institute of Environmental Science and Research (ESR) | Xiaoyun Ren, Matt Storey, Nikki Freed, Muhammad Faisal, Jing Wang, Hermes Perez, Anja Werno, Antje van der Linden, Arlo Upton, Chris Mansell, David Hammer, Dragana Drinkovic, Gary McAuliffe, Hana Sofia Andersson, James Ussher, Jill Sherwood, Josh Freeman, |
| hCoV-19/England/MILK-B3863F/2020 | EPI_ISL_658784 | 2020-11-01 | Lighthouse Lab in Milton Keynes | Wellcome Sanger Institute for the COVID-19 Genomics UK (COG-UK) Consortium | The Lighthouse Lab in Milton Keynes and Alex Alderton, Roberto Amato, Sonia Goncalves, Ewan Harrison, David K. Jackson, Ian Johnston, Dominic Kwiatkowski, Cordelia Langford, John Sillitoe on behalf of the Wellcome Sanger Institute COVID-19 Surveillance Te |
| hCoV-19/England/CAMC-B3584B/2020 | EPI_ISL_659669 | 2020-11-05 | Lighthouse Lab in Cambridge | Wellcome Sanger Institute for the COVID-19 Genomics UK (COG-UK) Consortium | Rob Howes, The Lighthouse Lab in Cambridge and Alex Alderton, Roberto Amato, Sonia Goncalves, Ewan Harrison, David K. Jackson, Ian Johnston, Dominic Kwiatkowski, Cordelia Langford, John Sillitoe on behalf of the Wellcome Sanger Institute COVID-19 Surveill |
| hCoV-19/SouthAfrica/KRISP-K004532-CS1/2020 | EPI_ISL_660230 | 2020-11-16 | KRISP, KZN Research Innovation and Sequencing Platform | KRISP, KZN Research Innovation and Sequencing Platform | Giandhari J, Pillay S, Lessells R, Mdlalose K, York D, Khan S, Tegally H, Wilkinson E, de Oliveira T |
| hCoV-19/Australia/NSW1217/2020 | EPI_ISL_672661 | 2020-11-13 | South Eastern Area Laboratory Services (SEALS) | Clinical Pathology and Medical Research; Westmead Hospital; University of Sydney | CIDM-PH et al. |
| hCoV-19/England/CAMC-B09C9F/2020 | EPI_ISL_672951 | 2020-10-29 | Lighthouse Lab in Cambridge | Wellcome Sanger Institute for the COVID-19 Genomics UK (COG-UK) Consortium | Rob Howes, The Lighthouse Lab in Cambridge and Alex Alderton, Roberto Amato, Sonia Goncalves, Ewan Harrison, David K. Jackson, Ian Johnston, Dominic Kwiatkowski, Cordelia Langford, John Sillitoe on behalf of the Wellcome Sanger Institute COVID-19 Surveill |
| hCoV-19/SaudiArabia/KAUST-RIYADH1446/2020 | EPI_ISL_678180 | 2020-06-27 | Pathogen Genomics Lab King Abdullah University of Science and Technology(KAUST) | Pathogen Genomics Lab King Abdullah University of Science and Technology(KAUST) | Olga Douvropoulou, Sara Mfarrej, Raushan Nugmanova, Sharif Hala, Raeece Naeem, Asim Khogeer, Fadwa Alofi, Afrah Alsomali, Jumana Taha, Abdulaziz Alahmadi, Kahled Algithami, Anwar Hashem, Naif Almontashiri, Arnab Pain |

| | | | | | |
|---|---|---|---|---|---|
| hCoV-19/Australia/NSW2760/2020 | EPI_ISL_678343 | 2020-09-15 | Area of Virology, Serology and Virology Division (SAViD), New South Wales Health Pathology Randwick | Virology, Serology and Virology Division (SAViD), New South Wales Health Pathology Randwick | Foster, C.; Au, J.; Ruiz Silva, M.; Deveson, I.; Bull, R.; Van Hal, S.; Rawlinson, W. |
| hCoV-19/Australia/NSW2776/2020 | EPI_ISL_678345 | 2020-09-08 | Area of Virology, Serology and Virology Division (SAViD), New South Wales Health Pathology Randwick | Virology, Serology and Virology Division (SAViD), New South Wales Health Pathology Randwick | Foster, C.; Au, J.; Ruiz Silva, M.; Deveson, I.; Bull, R.; Van Hal, S.; Rawlinson, W. |
| hCoV-19/Australia/NSW2830/2020 | EPI_ISL_678353 | 2020-10-04 | Area of Virology, Serology and Virology Division (SAViD), New South Wales Health Pathology Randwick | Virology, Serology and Virology Division (SAViD), New South Wales Health Pathology Randwick | Foster, C.; Au, J.; Ruiz Silva, M.; Deveson, I.; Bull, R.; Van Hal, S.; Rawlinson, W. |
| hCoV-19/Australia/NSW3456/2020 | EPI_ISL_678386 | 2020-11-30 | Area of Virology, Serology and Virology Division (SAViD), New South Wales Health Pathology Randwick | Virology, Serology and Virology Division (SAViD), New South Wales Health Pathology Randwick | Foster, C.; Au, J.; Ruiz Silva, M.; Deveson, I.; Bull, R.; Van Hal, S.; Rawlinson, W. |
| hCoV-19/Serbia/Sabac-000012108/2020 | EPI_ISL_678486 | 2020-08-20 | Veterinary Specialized Institute "Sabac", Serbia | Veterinary Specialized Institute "Kraljevo", Serbia | Vidanovic,D., Tesovic,B., Knezevic,A., Jovanovic,T., Jankovic,M., Sekler,M., Banovic Djeri,B., Petrovic,T., Mrkovacki, S., Volkening,J., Afonso,C. |
| hCoV-19/England/204742305/2020 | EPI_ISL_678727 | 2020-11-18 | Respiratory Virus Unit, Microbiology Services Colindale, Public Health England | COVID-19 Genomics UK (COG-UK) Consortium | PHE Covid Sequencing Team |
| hCoV-19/Japan/PG-2846/2020 | EPI_ISL_690973 | 2020-05-04 | Pathogen Genomics Center, National Institute of Infectious Diseases | Pathogen Genomics Center, National Institute of Infectious Diseases | Tsuyoshi Sekizuka, Kentaro Itokawa, Rina Tanaka, Masanori Hashino, Makoto Kuroda |
| hCoV-19/SouthAfrica/NHLS-UCT-GS-3553-KRISP/2020 | EPI_ISL_696481 | 2020-11-19 | George Hospital wc GRH & NHLS/UCT | KRISP, KZN Research Innovation and Sequencing Platform | Arash Iranzadeh, Deelan Doolabh, Lynn Tyers, Bruna Galvao, Innocent Mudau, Marvin Hsiao, Kruger Marais, Jennifer Giandhari, Sureshnee Pillay, Houriiyah Tegally, Emanuel James San, Tulio de Oliveira, Diana Hardie, Stephen Korsman, Carolyn Williamson |
| hCoV-19/UnitedArabEmirates/0165/2020 | EPI_ISL_698352 | 2020-05-18 | Group 42 (G42) Healthcare, Abu Dhabi, United Arab Emirates; Department of Health, The United Arab Emirates | G42 Healthcare | Rong Liu, Pei Wu, Sally Mahmoud, Ke Liang, Pauline Ogrodzki, Pengjuan Liu, Stephen S. Francis, Tao Ma, Hanif Khalak, Fang Chen, Denghui Liu, Junhua Li, Weibin Liu, Wenjun He, Xinyu Huang, Zhaorong Yuan, Long Lin, Nan Qiao, Xin Meng, Budoor Alqarni, Javier |
| hCoV-19/UnitedArabEmirates/0634/2020 | EPI_ISL_698790 | 2020-05-15 | Group 42 (G42) Healthcare, Abu Dhabi, United Arab Emirates; Department of Health, The United Arab Emirates | G42 Healthcare | Rong Liu, Pei Wu, Sally Mahmoud, Ke Liang, Pauline Ogrodzki, Pengjuan Liu, Stephen S. Francis, Tao Ma, Hanif Khalak, Fang Chen, Denghui Liu, Junhua Li, Weibin Liu, Wenjun He, Xinyu Huang, Zhaorong Yuan, Long Lin, Nan Qiao, Xin Meng, Budoor Alqarni, Javier |
| hCoV-19/Ecuador/USFQ-539/2020 | EPI_ISL_697800 | 2020-11-30 | Institute of Microbiology, Universidad San Francisco de Quito | Institute of Microbiology, Universidad San Francisco de Quito | Belv©n Prado-Vivar, Sully Mv°rquez, Juan Josv© Guadalupe, Monica Becerra-Wong, Maureen Mosquera, Bernardo Gutiv©rrez, Verv≥nica Barragv°n, Patricio Rojas-Silva, Gabriel Trueba, Michelle Grunauer, Pav∫l Cv°rdenas |
| hCoV-19/Australia/NSW1197/2020 | EPI_ISL_667797 | 2020-10-17 | South Eastern Area Laboratory Services (SEALS) | Clinical Pathology and Medical Research; Westmead Hospital; University of Sydney | CIDM-PH et al. |
| hCoV-19/USA/CO-CDPHE-2009053291/2020 | EPI_ISL_677284 | 2020-09-03 | Colorado Department of Public Health and Environment | Colorado Department of Puplic Health and Environment | Laura Bankers, Molly Hetherington-Rauth, Shannon Ely, Shannon R. Matzinger, Sarah Elizabeth Totten, Emily A. Travanty |

| | | | | | |
|---|---|---|---|---|---|
| hCoV-19/SouthAfrica/NHLS-UCT-GS-1407/2020 | EPI_ISL_700501 | 2020-08-09 | Knysna Hospital wc KNY | NHLS/UCT | Arash Iranzadeh, Deelan Doolabh, Lynn Tyers, Bruna Galvao, Innocent Mudau, Marvin Hsiao, Kruger Marais, Diana Hardie, Stephen Korsman, Carolyn Williamson |
| hCoV-19/SouthAfrica/NHLS-UCT-GS-0812/2020 | EPI_ISL_700551 | 2020-10-15 | Pacaltsdorp Clinic wc PAC | NHLS/UCT | Houriiyah Tegally, Arash Iranzadeh, Deelan Doolabh, Lynn Tyers, Bruna Galvao, Innocent Mudau, Marvin Hsiao, Kruger Marais, Diana Hardie, Stephen Korsman, Carolyn Williamson |
| hCoV-19/SouthAfrica/NHLS-UCT-GS-1486/2020 | EPI_ISL_700565 | 2020-08-11 | Herbertsdale Sat Clinic wc HBD | NHLS/UCT | Arash Iranzadeh, Deelan Doolabh, Lynn Tyers, Bruna Galvao, Innocent Mudau, Marvin Hsiao, Kruger Marais, Diana Hardie, Stephen Korsman, Carolyn Williamson |
| hCoV-19/England/CAMC-BD1489/2020 | EPI_ISL_702220 | 2020-11-29 | Lighthouse Lab in Cambridge | Wellcome Sanger Institute for the COVID-19 Genomics UK (COG-UK) Consortium | Rob Howes, The Lighthouse Lab in Cambridge and Alex Alderton, Roberto Amato, Sonia Goncalves, Ewan Harrison, David K. Jackson, Ian Johnston, Dominic Kwiatkowski, Cordelia Langford, John Sillitoe on behalf of the Wellcome Sanger Institute COVID-19 Surveill |
| hCoV-19/England/MILK-B3D5D3/2020 | EPI_ISL_704692 | 2020-11-02 | Oxford Viromics, NDM, University of Oxford; Oxford University Hospitals; Basingstoke and North Hampshire Hospital | COVID-19 Genomics UK (COG-UK) Consortium | Tanya Golubchik, David Bonsall, George Macintyre, Amy Trebes, Mariateresa de Cesare, Catrin Moore, Alex Mobbs, Anita Justice, Robert Shaw, Monique Andersson, Timothy Peto, Emma Wise, Nathan Moore, Jessica Lynch, Nick Cortes, Matilde Mori, Stephen Kidd, Da |
| hCoV-19/England/CAMC-BC1C84/2020 | EPI_ISL_705130 | 2020-11-22 | Lighthouse Lab in Cambridge | Wellcome Sanger Institute for the COVID-19 Genomics UK (COG-UK) Consortium | Rob Howes, The Lighthouse Lab in Cambridge and Alex Alderton, Roberto Amato, Sonia Goncalves, Ewan Harrison, David K. Jackson, Ian Johnston, Dominic Kwiatkowski, Cordelia Langford, John Sillitoe on behalf of the Wellcome Sanger Institute COVID-19 Surveill |
| hCoV-19/England/MILK-B3CF9A/2020 | EPI_ISL_706210 | 2020-11-02 | Oxford Viromics, NDM, University of Oxford; Oxford University Hospitals; Basingstoke and North Hampshire Hospital | COVID-19 Genomics UK (COG-UK) Consortium | Tanya Golubchik, David Bonsall, George Macintyre, Amy Trebes, Mariateresa de Cesare, Catrin Moore, Alex Mobbs, Anita Justice, Robert Shaw, Monique Andersson, Timothy Peto, Emma Wise, Nathan Moore, Jessica Lynch, Nick Cortes, Matilde Mori, Stephen Kidd, Da |
| hCoV-19/NewZealand/20CV0652/2020 | EPI_ISL_707806 | 2020-12-03 | Middlemore Hospital | Institute of Environmental Science and Research (ESR) | Xiaoyun Ren, Matt Storey, Nikki Freed, Muhammad Faisal, Jing Wang, Hermes Perez, Anja Werno, Antje van der Linden, Arlo Upton, Chris Mansell, David Hammer, Dragana Drinkovic, Gary McAuliffe, Hana Sofia Andersson, James Ussher, Jill Sherwood, Josh Freeman, |
| hCoV-19/Australia/NSW1225/2020 | EPI_ISL_710123 | 2020-11-19 | South Eastern Area Laboratory Services (SEALS) | CIDM-PH et al. | CIDM-PH et al. |
| hCoV-19/Brazil/RS-FIOCRUZ-15285/2020 | EPI_ISL_729859 | 2020-08-14 | Laboratv≥rio Central de Sav∫de Pv∫blica do Estado do Rio Grande do Sul (LACEN-RS) | Laboratory of Respiratory Viruses and Measles, Oswaldo Cruz Institute, FIOCRUZ | Paola Resende, Luciana Appolinario, Fernando Motta, Anna Carolina Paixv£o, Ana Carolina Mendonvßa, Tatiana Schaffer Gregianini, Marilda Tereza Mar da Rosa, Marilda Siqueira on behalf of the Fiocruz COVID-19 Genomic Surveillance Network |
| hCoV-19/USA/NM-DOH-2020316957/2020 | EPI_ISL_732724 | 2020-08-28 | New Mexico Department of Health Scientific Laboratory | New Mexico Department of Health Scientific Laboratory | Ellie Johnson, Anastacia Griego-Fisher, D'Eldra Malone |

| | | | | | |
|---|---|---|---|---|---|
| hCoV-19/mink/Poland/Pomorskie_49/2020 | EPI_ISL_732953 | 2020-11-17 | Department of Tropical Parasitology | Laboratory of Recombinant Vaccines | Lukasz Rabalski, Maciej Kosinski, Teemu Smura, Kirsi Aaltonen, Ravi Kant,¬† Tarja Sironen, Boguslaw Szewczyk,¬† Maciej Grzybek |
| hCoV-19/NewZealand/20CV0668/2020 | EPI_ISL_732968 | 2020-12-09 | Middlemore Hospital | Institute of Environmental Science and Research (ESR) | Xiaoyun Ren, Matt Storey, Nikki Freed, Muhammad Faisal, Jing Wang, Hermes Perez, Anja Werno, Antje van der Linden, Arlo Upton, Chris Mansell, David Hammer, Dragana Drinkovic, Gary McAuliffe, Hana Sofia Andersson, James Ussher, Jill Sherwood, Josh Freeman, |
| hCoV-19/USA/CA-CZB-13838/2020 | EPI_ISL_739073 | 2020-08-12 | Alameda County Public Health Lab | Chan-Zuckerberg Biohub | CZB Cliahub Consortium |
| hCoV-19/Colombia/SAN-INS-0597/2020 | EPI_ISL_739666 | 2020-10-31 | Instituto Nacional de Salud, Bogot√°, Colombia | Instituto Nacional de Salud, Bogot√°, Colombia | Katherine Laiton-Donato, Diego A. √Ålvarez-D√≠az, Carlos Franco-Mu√±oz, Mauricio Pacheco-Montealegre, Jonathan Reales, Diego Andr√©s Prada, Sheryl Corchuelo, Magdalena Weisner, Martha Lucia Ospina Martinez, Marcela Mercado-Reyes |
| hCoV-19/Australia/NSW1242/2020 | EPI_ISL_740873 | 2020-12-09 | South Eastern Area Laboratory Services (SEALS) | Clinical Pathology and Medical Research; Westmead Hospital; University of Sydney | CIDM-PH et al. |
| hCoV-19/Chile/RM-109586/2020 | EPI_ISL_746558 | 2020-06-24 | Genetica Molecular and Subdepartamento de Virologia ISP Chile | Instituto de Salud Publica de Chile | Javier Tognarelli, Barbara Parra, Loredana Arata, Jaime Lagos, Gisselle Barra, Patricia Bustos, Rodrigo Fasce, Andres Castillo, Jorge Fernandez |
| hCoV-19/Chile/RM-165835/2020 | EPI_ISL_746695 | 2020-08-31 | Genetica Molecular and Subdepartamento de Virologia ISP Chile | Instituto de Salud Publica de Chile | Javier Tognarelli, Barbara Parra, Loredana Arata, Jaime Lagos, Gisselle Barra, Patricia Bustos, Rodrigo Fasce, Andres Castillo, Jorge Fernandez |
| hCoV-19/Chile/RM-187968/2020 | EPI_ISL_746737 | 2020-09-23 | Genetica Molecular and Subdepartamento de Virologia ISP Chile | Instituto de Salud Publica de Chile | Javier Tognarelli, Barbara Parra, Loredana Arata, Jaime Lagos, Gisselle Barra, Patricia Bustos, Rodrigo Fasce, Andres Castillo, Jorge Fernandez |
| hCoV-19/Germany/BE-ChVir-W1636-6724/2020 | EPI_ISL_753703 | 2020-07-30 | Charit√© Universit√§tsmedizin Berlin, Institut f√ºr Virologie/Labor Berlin | Charit√© Universit√§tsmedizin Berlin, Institut f√ºr Virologie | Victor M Corman, J√∂rn Beheim-Schwarzbach, Barbara M√ºhlemann, Julia Schneider, Talitha Veith, Terry Jones, Christian Drosten |
| hCoV-19/Georgia/Tb-97937/2020 | EPI_ISL_754181 | 2020-09-23 | Department for Virology, Molecular Biology and Genome Research, R. G. Lugar Center for Public Health Research, National Center for Disease Control and Public Health (NCDC) of Georgia. | [illegible] Biology and Genome Research, R. G. Lugar Center for Public Health Research, National Center for Disease Control and Public Health (NCDC) of Georgia. | Meri Pantsulaia, Nino Berishvili, Tata Imnadze, Giorgi Tomashvili, Ana Papkiauri, Gvantsa Brachveli, Gvantsa Chanturia, Ann Machablishvili, Nato Kotaria, Marine Murtskhvaladze, Lela Sabadze, Mari Gavashelidze, Tamar Jashiashvili, Tea Tevdoradze, Ketevan S |
| hCoV-19/NewZealand/20CV0687/2020 | EPI_ISL_755626 | 2020-12-20 | PathLab Bay of Plenty | Institute of Environmental Science and Research (ESR) | Xiaoyun Ren, Matt Storey, Nikki Freed, Muhammad Faisal, Jing Wang, Hermes Perez, Anja Werno, Antje van der Linden, Arlo Upton, Chris Mansell, David Hammer, Dragana Drinkovic, Gary McAuliffe, Hana Sofia Andersson, James Ussher, Jill Sherwood, Josh Freeman, |
| hCoV-19/England/QEUH-CAF17D/2020 | EPI_ISL_760739 | 2020-12-15 | Lighthouse Lab in Glasgow | Wellcome Sanger Institute for the COVID-19 Genomics UK (COG-UK) Consortium | Harper VanSteenhouse, Yumi Kasai, David Gray, Carol Clugston, Anna Dominiczak and Alex Alderton, Roberto Amato, Sonia Goncalves, Ewan Harrison, David K. Jackson, Ian Johnston, Dominic Kwiatkowski, Cordelia Langford, John Sillitoe on behalf of the Wellcome |

| | | | | | |
|---|---|---|---|---|---|
| hCoV-19/England/QEUH-C92EFE/2020 | EPI_ISL_762092 | 2020-12-17 | Lighthouse Lab in Glasgow | Wellcome Sanger Institute for the COVID-19 Genomics UK (COG-UK) Consortium | Harper VanSteenhouse, Yumi Kasai, David Gray, Carol Clugston, Anna Dominiczak and Alex Alderton, Roberto Amato, Sonia Goncalves, Ewan Harrison, David K. Jackson, Ian Johnston, Dominic Kwiatkowski, Cordelia Langford, John Sillitoe on behalf of the Wellcome |
| hCoV-19/USA/TX-DSHS-2016/2020 | EPI_ISL_766691 | 2020-06-01 | Texas Department of State Health Services | Texas Department of State Health Services | Rashmi Tuladhar, Bonnie Oh, Jenny Zhang, Maliha Rahman, Anita Pokharel, Myong Koag, Chung Wang, Rachel Lee, Grace Kubin, Mayela Pedrueza, James Daniel Bonser |
| hCoV-19/Norway/6605/2020 | EPI_ISL_775403 | 2020-12-07 | Nordland Hospital - Bodo, Laboratory Department, Molecular Biology Unit | Norwegian Institute of Public Health, Department of Virology | Kathrine Stene-Johansen, Kamilla Heddeland Instefjord, Hilde Elshaug, Atiya R Ali,Marie Paulsen Madsen, Rasmus Riis Kopperud, Hilde Vollan, Karoline Bragstad, Olav Hungnes |
| hCoV-19/Israel/CVL-2-ngs/2020 | EPI_ISL_776593 | 2020-12-24 | Israel Central Virology laboratory | Israel Central Virology laboratory | Neta Zuckerman, Efrat Dahan Bucris, Oran Erster, Michal Mandelboim, Orna Mor, Ella Mendelson |
| hCoV-19/USA/TX-HMH-MCoV-9309/2020 | EPI_ISL_780504 | 2020-07-03 | Houston Methodist Hospital | Houston Methodist Hospital | S. Wesley Long, Randall J. Olsen, Paul A. Christensen, David W. Bernard, James J. Davis, Maulik Shukla, Marcus Nguyen, Matthew Ojeda Saavedra, Prasanti Yerramilli, Layne Pruitt, Sishir Subedi, Heather Hendrickson, and James M. Musser |
| hCoV-19/USA/TX-HMH-MCoV-14974/2020 | EPI_ISL_786458 | 2020-10-16 | Houston Methodist Hospital | Houston Methodist Hospital | S. Wesley Long, Randall J. Olsen, Paul A. Christensen, David W. Bernard, James J. Davis, Maulik Shukla, Marcus Nguyen, Matthew Ojeda Saavedra, Prasanti Yerramilli, Layne Pruitt, Sishir Subedi, Heather Hendrickson, and James M. Musser |
| hCoV-19/USA/TX-HMH-MCoV-14509/2020 | EPI_ISL_787794 | 2020-10-09 | Houston Methodist Hospital | Houston Methodist Hospital | S. Wesley Long, Randall J. Olsen, Paul A. Christensen, David W. Bernard, James J. Davis, Maulik Shukla, Marcus Nguyen, Matthew Ojeda Saavedra, Prasanti Yerramilli, Layne Pruitt, Sishir Subedi, Heather Hendrickson, and James M. Musser |
| hCoV-19/Brazil/PB-FIOCRUZ-16617/2020 | EPI_ISL_792597 | 2020-06-30 | Laboratv≥rio Central de Sav∫de Pv∫blica do Estado da Parav≠ba (LACEN-PB) | Laboratory of Respiratory Viruses and Measles, Oswaldo Cruz Institute, FIOCRUZ | Paola Resende, Luciana Appolinario, Fernando Motta, Anna Carolina Paixao, Ana Carolina Mendonca, Jov£o Felipe Bezerra, Romero Henrique Teixeira de Vasconcelos, Dalane Loudal Florentino Teixeira, Thiago Franco de Oliveira Carneiro, Marilda Siqueira on beha |
| hCoV-19/Australia/WA555/2020 | EPI_ISL_794727 | 2020-12-17 | PathWest Laboratory Medicine WA | PathWest Laboratory Medicine WA Microbial Surveillance Unit | PathWest Laboratory Medicine WA Microbial Surveillance Unit |
| hCoV-19/England/CAMC-DD9892/2020 | EPI_ISL_797639 | 2020-12-29 | Lighthouse Lab in Cambridge | Wellcome Sanger Institute for the COVID-19 Genomics UK (COG-UK) Consortium | Rob Howes, The Lighthouse Lab in Cambridge and Alex Alderton, Roberto Amato, Sonia Goncalves, Ewan Harrison, David K. Jackson, Ian Johnston, Dominic Kwiatkowski, Cordelia Langford, John Sillitoe on behalf of the Wellcome Sanger Institute COVID-19 Surveill |

| | | | | | |
|---|---|---|---|---|---|
| hCoV-19/USA/NY-MSHSPSP-PV17762/2020 | EPI_ISL_801862 | 2020-09-03 | MSHS Clinical Microbiology Laboratories | MSHS Pathogen Surveillance Program | Ana S. Gonzalez-Reiche, Hala Alshammary, Mitchell J. Sullivan, Brianne Ciferri, Ajay Obla, Angela Amoako, Mahmoud Awawda, Elena Hirsch, Ashley S. Salimbangon, Levy Sominsky, Katherine Beach, Kayla Russo, Charles Gleason, Shelcie Fabre, Giulio Kleiner, Zen |
| hCoV-19/USA/NY-MSHSPSP-PV19497/2020 | EPI_ISL_802324 | 2020-10-01 | MSHS Clinical Microbiology Laboratories | MSHS Pathogen Surveillance Program | Ana S. Gonzalez-Reiche, Hala Alshammary, Mitchell J. Sullivan, Brianne Ciferri, Ajay Obla, Angela Amoako, Mahmoud Awawda, Elena Hirsch, Ashley S. Salimbangon, Levy Sominsky, Katherine Beach, Kayla Russo, Charles Gleason, Shelcie Fabre, Giulio Kleiner, Zen |
| hCoV-19/Israel/CVL-503ngs/2020 | EPI_ISL_804073 | 2020-12-28 | Israel Central Virology laboratory | Israel National Consortium for SARS-CoV-2 sequencing | Neta Zuckerman, Efrat Dahan Bucris, Michal Mandelboim, Dana Bar-Ilan, Oran Erster, Tzvia Mann, Omer Murik, David A. Zeevi, Assaf Rokney, Joseph Jaffe, Eva Nachum, Maya Davidovich Cohen, Ephraim Fass, Gal Zizelski Valenci, Mor Rubinstein, Efrat Rorman, Is |
| hCoV-19/Australia/VIC18081/2021 | EPI_ISL_812441 | 2021-01-08 | Victorian Infectious Diseases Reference Laboratory (VIDRL) | VIDRL and MDU-PHL | Caly L., Seemann T., Sait, M.L., Druce J., Sherry, N.L. |
| hCoV-19/Norway/7234/2020 | EPI_ISL_813975 | 2020-12-27 | Akershus University Hospital, Department for Microbiology and Infectious Disease Control | Norwegian Institute of Public Health, Department of Virology | Kathrine Stene-Johansen, Kamilla Heddeland Instefjord, Hilde Elshaug, Atiya R Ali,Marie Paulsen Madsen, Rasmus Riis Kopperud, Hilde Vollan, Karoline Bragstad, Olav Hungnes |
| hCoV-19/Israel/CVL-256-ngs/2020 | EPI_ISL_814265 | 2020-12-27 | Israel Central Virology laboratory | Israel National Consortium for SARS-CoV-2 sequencing | Neta Zuckerman, Efrat Dahan Bucris, Michal Mandelboim, Dana Bar-Ilan, Oran Erster, Tzvia Mann, Omer Murik, David A. Zeevi, Assaf Rokney, Joseph Jaffe, Eva Nachum, Maya Davidovich Cohen, Ephraim Fass, Gal Zizelski Valenci, Mor Rubinstein, Efrat Rorman, Is |
| hCoV-19/Ecuador/USFQ-564/2020 | EPI_ISL_824291 | 2020-12-30 | Institute of Microbiology, Universidad San Francisco de Quito | Institute of Microbiology, Universidad San Francisco de Quito | Bel√©n Prado-Vivar, Sully M√°rquez, Juan Jos√© Guadalupe, Monica Becerra-Wong, Bernardo Guti√©rrez, Andrea Cunguan, Nabih Dahik, Ver√≥nica Barrag√°n, Patricio Rojas-Silva, Gabriel Trueba, Michelle Grunauer, Pa√∫l C√°rdenas |
| hCoV-19/Ecuador/USFQ-578/2021 | EPI_ISL_824293 | 2021-01-06 | Institute of Microbiology, Universidad San Francisco de Quito | Institute of Microbiology, Universidad San Francisco de Quito | Bel√©n Prado-Vivar, Sully M√°rquez, Juan Jos√© Guadalupe, Monica Becerra-Wong, Bernardo Guti√©rrez, Eulalia Pazmi√±o, Katalina Pacheco, Ver√≥nica Barrag√°n, Patricio Rojas-Silva, Gabriel Trueba, Michelle Grunauer, Pa√∫l C√°rdenas |
| hCoV-19/USA/NY-AECOM_133/2020 | EPI_ISL_826667 | 2020-10-13 | Montefiore Medical Center | Albert Einstein College of Medicine, Dept. of Microbiology & Immunology, Chandran lab | J. Maximilian Fels, Saad Khan, Ryan Forster, Karin A. Skalina, Surksha Sirichand, Amy S. Fox, Aviv Bergman, William B. Mitchell, Lucia R. Wolgast, Wendy Szymczak, Robert H. Bortz III, M. Eugenia Dieterle, Catalina Florez, Denise Haslwanter, Rohit K. Jangr |
| hCoV-19/Ecuador/EC-M-95299/2020 | EPI_ISL_826825 | 2020-07-28 | INSPI-CRN DE INFLUENZA Y OTROS VIRUS RESPIRATORIOS | Instituto de Salud Publica de Chile | Javier Tognarelli, Barbara Parra, Loredana Arata, Jaime Lagos, Gisselle Barra, Alfredo Bruno, Domenica de Mora, Solon Narvaez, Jimmy Garcez, Michelle Paez, Martiza Olmedo, Manuel Gonzalez, Patricia Bustos, Rodrigo Fasce, Andres Castillo, Jorge Fernandez |

| | | | | | |
|---|---|---|---|---|---|
| hCoV-19/USA/NM-DOH-2020045837/2020 | EPI_ISL_831904 | 2020-05-04 | New Mexico Department of Health Scientific Laboratory | New Mexico Department of Health Scientific Laboratory | Ellie Johnson, Anastacia Griego-Fisher, D'eldra Malone |
| hCoV-19/NewZealand/21CV0047/2021 | EPI_ISL_843195 | 2021-01-06 | Canterbury Health Laboratories | Institute of Environmental Science and Research (ESR) | Xiaoyun Ren, Matt Storey, Nikki Freed, Muhammad Faisal, Jing Wang, Hermes Perez, Anja Werno, Antje van der Linden, Arlo Upton, Chris Mansell, David Hammer, Dragana Drinkovic, Gary McAuliffe, Hana Sofia Andersson, James Ussher, Jill Sherwood, Josh Freeman, |
| hCoV-19/USA/MI-UM-10036933724/2020 | EPI_ISL_846662 | 2020-11-13 | University of Michigan Clinical Microbiology Laboratory | Department of Microbiology and Immunology | Valesano |
| hCoV-19/USA/IL-IDPH-I-000118/2020 | EPI_ISL_848247 | 2020-10-08 | Illinois Department of Public Health | Gagnon Lab, Southern Illinois University | Keith Gagnon |
| hCoV-19/USA/UT-UPHL-2101037509/2020 | EPI_ISL_849234 | 2020-12-15 | Utah Public Health Laboratory | Utah Public Health Laboratory | Erin L. Young, Kelly F. Oakeson, Tara Gallagher |
| hCoV-19/SouthKorea/KDCA0556/2020 | EPI_ISL_850247 | 2020-07-24 | Division of Emerging Infectious Diseases, Bureau of Infectious Diseases Diagnosis Control, Korea Disease Control and Prevention Agency | Diseases, Bureau of Infectious Diseases Diagnosis Control, Korea Disease Control and Prevention Agency | Ae Kyung Park, Il-Hwan Kim, Heui Man Kim, Jeong-Min Kim, Namjoo Lee, Chaeyoung Lee, Sang Hee Woo, Eun-Jin Kim |
| hCoV-19/SouthKorea/KDCA0683/2020 | EPI_ISL_850364 | 2020-08-17 | Division of Emerging Infectious Diseases, Bureau of Infectious Diseases Diagnosis Control, Korea Disease Control and Prevention Agency | Diseases, Bureau of Infectious Diseases Diagnosis Control, Korea Disease Control and Prevention Agency | Ae Kyung Park, Il-Hwan Kim, Heui Man Kim, Jeong-Min Kim, Namjoo Lee, Chaeyoung Lee, Sang Hee Woo, Eun-Jin Kim |
| hCoV-19/Australia/WA601/2021 | EPI_ISL_857468 | 2021-01-12 | PathWest Laboratory Medicine WA | PathWest Laboratory Medicine WA Microbial Surveillance Unit | PathWest Laboratory Medicine WA Microbial Surveillance Unit |
| hCoV-19/UnitedArabEmirates/3875/2020 | EPI_ISL_859885 | 2020-11-24 | BTC, Khalifa University | BTC, Khalifa University | Al Safar et al |
| hCoV-19/UnitedArabEmirates/4359/2020 | EPI_ISL_860088 | 2020-12-26 | BTC, Khalifa University | BTC, Khalifa University | Al Safar et al |
| hCoV-19/Ghana/UHAS-W247/2020 | EPI_ISL_861459 | 2020-12-03 | UHAS COVID-19 Lab | UHAS COVID-19 Lab | Kwabena O. Duedu, Jones Gyamfi, Reuben Ayivor-Djanie, John O. Gyapong and the UHAS COVID-19 Lab Team |
| hCoV-19/India/AP-CS0357/2020 | EPI_ISL_862282 | 2020-07-04 | Kurnool Medical College (KMC) | CSIR Institute of Genomics and Integrative Biology | Pallavali Roja Rani, Mohamed Imran, J. Vijaya Lakshmi, Bani Jolly, S. Afsar, Abhinav Jain, Mohit Kumar Divakar, Panyam Suresh, Disha Sharma, Nambi Rajesh, Rahul C Bhoyar, Dasari Ankaiah, Sanaga Shanthi Kumari, Gyan Ranjan, Valluri Anitha Lavanya, Mercy Ro |
| hCoV-19/NewZealand/21CV0063/2021 | EPI_ISL_877208 | 2021-01-06 | LabPLUS | Institute of Environmental Science and Research (ESR) | Xiaoyun Ren, Matt Storey, Nikki Freed, Muhammad Faisal, Jing Wang, Hermes Perez, Anja Werno, Antje van der Linden, Arlo Upton, Chris Mansell, David Hammer, Dragana Drinkovic, Gary McAuliffe, Hana Sofia Andersson, James Ussher, Jill Sherwood, Josh Freeman, |
| hCoV-19/Australia/VIC18163/2021 | EPI_ISL_877584 | 2021-01-22 | Victorian Infectious Diseases Reference Laboratory (VIDRL) | VIDRL and MDU-PHL | Caly L., Seemann T., Sait, M.L., Druce J., Sherry, N.L. |

| | | | | | |
|---|---|---|---|---|---|
| hCoV-19/USA/NJ-CDC-LC00003021/2020 | EPI_ISL_887648 | 2020-12-30 | Labcorp | Genomics and Discovery, Respiratory Viruses Branch, Division of Viral Diseases, Centers for Disease Control and Prevention | Peter W. Cook,Dhwani Batra,Ben L. Rambo-Martin,Summer Galloway,Brian Krueger,Minoo Agarwal,Eyad Almasri,Debbie Boles,Ayla Burns,Nuthawin Charoensri,Oren Cohen,Susan Countryman,Mary Ann Cristobal,Bobbi Croy,Suzanne Dale,Hrushikesh Deshmukh,Amanda Douglas,V |
| hCoV-19/Israel/CVL-148ngs/2020 | EPI_ISL_889040 | 2020-12-27 | Israel Central Virology laboratory | Israel National Consortium for SARS-CoV-2 sequencing | Neta Zuckerman, Efrat Dahan Bucris, Michal Mandelboim, Dana Bar-Ilan, Oran Erster, Tzvia Mann, Omer Murik, David A. Zeevi, Assaf Rokney, Joseph Jaffe, Eva Nachum, Maya Davidovich Cohen, Ephraim Fass, Gal Zizelski Valenci, Mor Rubinstein, Efrat Rorman, Is |
| hCoV-19/Switzerland/GE-33002169/2021 | EPI_ISL_897657 | 2021-01-18 | University Hospitals of Geneva, Laboratory of Virology | HUG, Laboratory of Virology and the Health2030 Genome Center | Samuel Cordey, Ana Rita Goncalves, Laurent Kaiser, Lorenzo Cerutti, Henri Pegeot, Melyssa Elies, Deborah Penet, Keith Harshman, Ioannis Xenarios, Emmanouil Dermitzakis |
| hCoV-19/USA/MD-MDH-0688/2021 | EPI_ISL_902930 | 2021-01-14 | Maryland Public Health Laboratory | Maryland Public Health Laboratory | Maryland Department of Health Laboratories Administration |
| hCoV-19/Brazil/SP-1044/2021 | EPI_ISL_906074 | 2021-01-19 | Searom Laboratorio Diagnostico | Procedures Center, Strategic Laboratory | Claudio Tavares Sacchi, Claudia Regina Gonv∫alves, Erica Valessa Ramos Gomes, Karoline Rodrigues Campos |
| hCoV-19/Nigeria/CV605/2020 | EPI_ISL_906298 | 2020-12-17 | Nigeria Centre for Disease Control (NCDC) | African Centre of Excellence for Genomics of Infectious Diseases (ACEGID), Redeemer's University | Oluniyi P.E. et al |
| hCoV-19/Colombia/LAG-INS-VG-320/2021 | EPI_ISL_906532 | 2021-01-12 | Hospital San Jose de Maicao | Instituto Nacional de Salud Direcci√≥n de Investigaci√≥n en Salud P√∫blica, Universidad de los Andes-Applied genomics research group, Vicerrectoria de Investigaci√≥n y Creaci√≥n, Universidad de los Andes-Systems and Computing Engineering | Katherine Laiton-Donato, Diego A. √Ålvarez-D√≠az, Carlos Franco-Mu√±oz, Mauricio Pacheco-Montealegre, H√©ctor Alejandro Ruiz-Moreno, Maria T. Herrera-Sep√∫lveda, Diego Andr√©s Prada, Jhonnatan Reales-Gonz√°lez, Sheryll Corchuelo, Julian Naizaque, Gerardo |
| hCoV-19/Colombia/QUI-INS-VG-336/2020 | EPI_ISL_906533 | 2020-12-02 | Laboratorio de Virologia-Instituto Nacional de Salud | Instituto Nacional de Salud Direcci√≥n de Investigaci√≥n en Salud P√∫blica, Universidad de los Andes-Applied genomics research group, Vicerrectoria de Investigaci√≥n y Creaci√≥n, Universidad de los Andes-Systems and Computing Engineering | Katherine Laiton-Donato, Diego A. √Ålvarez-D√≠az, Carlos Franco-Mu√±oz, Mauricio Pacheco-Montealegre, H√©ctor Alejandro Ruiz-Moreno, Maria T. Herrera-Sep√∫lveda, Diego Andr√©s Prada, Jhonnatan Reales-Gonz√°lez, Sheryll Corchuelo, Julian Naizaque, Gerardo |
| hCoV-19/England/CAMC-106EF4C/2021 | EPI_ISL_908107 | 2021-01-16 | Lighthouse Lab in Cambridge | Wellcome Sanger Institute for the COVID-19 Genomics UK (COG-UK) Consortium | Rob Howes, The Lighthouse Lab in Cambridge and Alex Alderton, Roberto Amato, Sonia Goncalves, Ewan Harrison, David K. Jackson, Ian Johnston, Dominic Kwiatkowski, Cordelia Langford, John Sillitoe on behalf of the Wellcome Sanger Institute COVID-19 Surveill |
| hCoV-19/India/TG-CCMB-AI310/2020 | EPI_ISL_910157 | 2020-09-09 | CSIR-Centre for Cellular and Molecular Biology | CSIR-Centre for Cellular and Molecular Biology | Payel Mukherjee,Pratheusa Maccha,Namami Gaur,Lamuk Zaveri,Tulasi Nagabandi,Purushotham Vodnala,Blessy B John,Viswagithe S L,B Himasri,Sofia Banu,Priya Singh,Archana Bharadwaj Siva,Karthik Bharadwaj Tallapaka,Rakesh K Mishra,Divya Tej Sowpati |

| | | | | | |
|---|---|---|---|---|---|
| hCoV-19/England/BIRM-687B6/2020 | EPI_ISL_918578 | 2020-12-01 | University of Birmingham | COVID-19 Genomics UK (COG-UK) Consortium | Institute of Microbiology, University of Birmingham: Claire McMurray, Joanne Stockton, Samuel Nicholls, Radoslaw Poplawski, Will Rowe, Josh Quick, Nicholas Loman. University of Birmingham Testing Laboratory: Celina M Whalley, Andrew Bosworth, Charlotte P |
| hCoV-19/Australia/WA608/2021 | EPI_ISL_933787 | 2021-01-21 | PathWest Laboratory Medicine WA | PathWest Laboratory Medicine WA Microbial Surveillance Unit | PathWest Laboratory Medicine WA Microbial Surveillance Unit |
| hCoV-19/Australia/WA612/2021 | EPI_ISL_933794 | 2021-01-25 | PathWest Laboratory Medicine WA | PathWest Laboratory Medicine WA Microbial Surveillance Unit | PathWest Laboratory Medicine WA Microbial Surveillance Unit |
| hCoV-19/Australia/WA617/2021 | EPI_ISL_933800 | 2021-01-28 | PathWest Laboratory Medicine WA | PathWest Laboratory Medicine WA Microbial Surveillance Unit | PathWest Laboratory Medicine WA Microbial Surveillance Unit |
| hCoV-19/SouthAfrica/VIDA-KRISP-K008053/2020 | EPI_ISL_940872 | 2020-11-02 | Vaccines¬†and¬†Infectious Diseases Analytics¬†Research Unit (VIDA) | KRISP, KZN Research Innovation and Sequencing Platform | Baillie Vicky, du Plessis Jeanine, Giandhari Jennifer, Pillay Sureshnee, Naidoo Yeshnee, Tegally Houriiyah, de Oliveira Tulio, Madhi Shabir |
| hCoV-19/Colombia/AMA-GUR-06828/2020 | EPI_ISL_941161 | 2020-05-05 | Centro de Investigaciones en Microbiolog√≠a y Biotecnolog√≠a-UR (CIMBIUR), Facultad de Ciencias Naturales, Universidad del Rosario, Bogot√°, Colombia | [illegible] Microbiolog√≠a y Biotecnolog√≠a-UR (CIMBIUR), Facultad de Ciencias Naturales, Universidad del Rosario, Bogot√°, Colombia Icahn School of Medicine at Mount Sinai, New York, | Nathalia Ballesteros, Marina Mu√±oz, Luz Helena Pati√±o, Carolina Hern√°ndez, Felipe Gonz√°lez-Casabianca, Iv√°n Carroll, Mauricio Santos-Vega, Jaime Cascante, Andr√©s Angel, Alejandro Feged-Rivadeneira, M√≥nica Palma-Cuero, Carolina Fl√≥rez, Sergio Gomez |
| hCoV-19/Israel/CVL-1901/2020 | EPI_ISL_944225 | 2020-12-31 | Israel Central Virology laboratory | Israel National Consortium for SARS-CoV-2 sequencing | Neta Zuckerman, Efrat Dahan Bucris, Michal Mandelboim, Dana Bar-Ilan, Oran Erster, Tzvia Mann, Omer Murik, David A. Zeevi, Assaf Rokney, Joseph Jaffe, Eva Nachum, Maya Davidovich Cohen, Ephraim Fass, Gal Zizelski Valenci, Mor Rubinstein, Efrat Rorman, Is |
| hCoV-19/Ghana/WACCBIP-GS141/2021 | EPI_ISL_944693 | 2021-01-08 | Department of Biochemistry, Cell and Molecular Biology, West African Centre for Cell Biology of Infectious Pathogens (WACCBIP), University of Ghana | Department of Biochemistry, Cell and Molecular Biology, West African Centre for Cell Biology of Infectious Pathogens (WACCBIP), University of Ghana | Morang'a,C.M., Ngoi,J.M., Quansah,E.B., Saiid,S., Amuzu,D.S., Asante,I., Bonney,J.H., Bonney,E., Odoom,J.K., Ndam,N.T., Tei-Maya,F., Adusei-Poku,M., Ofori-Boadu,L., Ampofo,W.K., Amenga-Etego,L.N., Quashie,P., Bediako,Y., Awandare,G.A. |
| hCoV-19/India/WB-IICB-053/2020 | EPI_ISL_944789 | 2020-10-26 | CSIR-Indian Institute of Chemical Biology, MEDICA Supercpecialty Hospital Kolkata | CSIR-Indian Institute of Chemical Biology, MEDICA Supercpecialty Hospital Kolkata | Sujay Krishna Maity, Priyanka Mallick, Debaleena Bhowmik, Abhishake Lahiri, Dr. Aviral Roy, Dr. Soumen Saha, Dr. Arpita Ghosh Mitra, Dr. Rajesh Pandey, Dr. Sandip Paul, Dr. Partha Chakrabarti, Dr. Saikat Chakrabarti |
| hCoV-19/Indonesia/JB-EIJK68/2020 | EPI_ISL_947260 | 2020-11-11 | RSU Bunda Mulia | Eijkman Institute for Molecular Biology, Ministry of Research and Technology/National Agency for Research and Innovation | Iskandar Adnan, Lydia V. Panggalo, Sukma Oktavianthi, Willy Agustine, Edison Johar, Hidayat Trimarsanto, Frilasita A Yudhaputri, Safarina G Malik, Khin Saw Myint, Amin Soebandrio |
| hCoV-19/France/PAC-IHU-0925_Nova2/2020 | EPI_ISL_2659879 | 2020-05-27 | MEPHI, Aix Marseille University | MEPHI, Aix Marseille University | Anthony LEVASSEUR |
| hCoV-19/USA/KS-KHEL-0131/2020 | EPI_ISL_1004232 | 2020-06-11 | Kansas Health and Environmental Lab | Kansas Health and Environmental Lab | Mike Grose, Carissa Robertson, Ben Olsen, and Phil Adam |

| | | | | | |
|---|---|---|---|---|---|
| hCoV-19/England/MILK-12065CA/2021 | EPI_ISL_1006967 | 2021-02-01 | Lighthouse Lab in Milton Keynes | Wellcome Sanger Institute for the COVID-19 Genomics UK (COG-UK) Consortium | The Lighthouse Lab in Milton Keynes and Alex Alderton, Roberto Amato, Sonia Goncalves, Ewan Harrison, David K. Jackson, Ian Johnston, Dominic Kwiatkowski, Cordelia Langford, John Sillitoe on behalf of the Wellcome Sanger Institute COVID-19 Surveillance Te |
| hCoV-19/USA/UT-UPHL-210603523403/2021 | EPI_ISL_2456737 | 2021-05-19 | Utah Public Health Laboratory | Utah Public Health Laboratory | Erin L. Young, Kelly F. Oakeson, Tara Gallagher |
| hCoV-19/England/CAMC-1216530/2021 | EPI_ISL_1007216 | 2021-02-05 | Lighthouse Lab in Cambridge | Wellcome Sanger Institute for the COVID-19 Genomics UK (COG-UK) Consortium | Rob Howes, The Lighthouse Lab in Cambridge and Alex Alderton, Roberto Amato, Sonia Goncalves, Ewan Harrison, David K. Jackson, Ian Johnston, Dominic Kwiatkowski, Cordelia Langford, John Sillitoe on behalf of the Wellcome Sanger Institute COVID-19 Surveill |
| hCoV-19/Brazil/SE-FIOCRUZ-33574/2020 | EPI_ISL_2660688 | 2020-07-04 | Laboratorio Central de Saude Publica do Estado de Sergipe (LACEN/SE) | Laboratory of Respiratory Viruses and Measles, Oswaldo Cruz Institute, FIOCRUZ | Paola Resende, Luciana Appolinario, Fernando Motta, Anna Carolina Paixao, Ana Carolina Mendonca, Alice Sampaio Rocha, Tain√° Moreira Martins Venas, Elisa Cavalcante Pereira, Renata Serrano Lopes, Cliomar Alves dos Santos, Marilda Siqueira on behalf of the |
| hCoV-19/France/IDF_PSL_324/2020 | EPI_ISL_1015127 | 2020-07-28 | Department of Virology, Piti√©-Salp√™tri√®re hospital | Department of Virology, Piti√©-Salp√™tri√®re hospital | Valentin Leducq, Aude Jary, Karen Zafilaza, St√©phane Marot, Vincent Calvez, Anne-Genevi√®ve Marcelin |
| hCoV-19/France/IDF_PSL_252/2020 | EPI_ISL_1015146 | 2020-08-25 | Department of Virology, Piti√©-Salp√™tri√®re hospital | Department of Virology, Piti√©-Salp√™tri√®re hospital | Valentin Leducq, Aude Jary, Karen Zafilaza, St√©phane Marot, Vincent Calvez, Anne-Genevi√®ve Marcelin |
| hCoV-19/NewZealand/21CV0121/2021 | EPI_ISL_1016845 | 2021-01-15 | Middlemore Hospital | Institute of Environmental Science and Research (ESR) | Xiaoyun Ren, Matt Storey, Nikki Freed, Muhammad Faisal, Jing Wang, Hermes Perez, Anja Werno, Antje van der Linden, Arlo Upton, Chris Mansell, David Hammer, Dragana Drinkovic, Gary McAuliffe, Hana Sofia Andersson, James Ussher, Jill Sherwood, Josh Freeman, |
| hCoV-19/NewZealand/21MV0006/2021 | EPI_ISL_1016866 | 2021-01-26 | North Shore Hospital | Institute of Environmental Science and Research (ESR) | Xiaoyun Ren, Matt Storey, Nikki Freed, Muhammad Faisal, Jing Wang, Hermes Perez, Anja Werno, Antje van der Linden, Arlo Upton, Chris Mansell, David Hammer, Dragana Drinkovic, Gary McAuliffe, Hana Sofia Andersson, James Ussher, Jill Sherwood, Josh Freeman, |
| hCoV-19/NewZealand/21MV0065/2021 | EPI_ISL_1016871 | 2021-02-08 | LabPLUS | Institute of Environmental Science and Research (ESR) | Xiaoyun Ren, Matt Storey, Nikki Freed, Muhammad Faisal, Jing Wang, Hermes Perez, Anja Werno, Antje van der Linden, Arlo Upton, Chris Mansell, David Hammer, Dragana Drinkovic, Gary McAuliffe, Hana Sofia Andersson, James Ussher, Jill Sherwood, Josh Freeman, |
| hCoV-19/NewZealand/21MV0056/2021 | EPI_ISL_1016873 | 2021-02-05 | LabPLUS | Institute of Environmental Science and Research (ESR) | Xiaoyun Ren, Matt Storey, Nikki Freed, Muhammad Faisal, Jing Wang, Hermes Perez, Anja Werno, Antje van der Linden, Arlo Upton, Chris Mansell, David Hammer, Dragana Drinkovic, Gary McAuliffe, Hana Sofia Andersson, James Ussher, Jill Sherwood, Josh Freeman, |
| hCoV-19/Serbia/Belgrade-2515952809/2020 | EPI_ISL_1017698 | 2020-09-28 | Institute of Virology, Vaccines and Sera ''Torlak'' | Immunology, Faculty of Medicine, University of Belgrade | Knezevic,A., Jankovic,M., Vidanovic,D., Milicevic,O., Tesovic,B., Sekler,M., Jovanovic,T. |

| | | | | | |
|---|---|---|---|---|---|
| hCoV-19/HongKong/HKPU-01319/2020 | EPI_ISL_1018133 | 2020-07-08 | Department of Health Technology and Informatics, The Hong Kong Polytechnic University | Department of Health Technology and Informatics, The Hong Kong Polytechnic University | Gilman Kit-Hang Siu, Lam-Kwong Lee, Kenneth Siu-Sing Leung, Jake Siu-Lun Leung, Timothy Ting-Leung Ng, Chloe Toi-Mei Chan, Kingsley King-Gee Tam, Hiu-Yin Lao, Denise Sze-Hang Wong, Alan Ka-Lun Wu, Miranda Chong-Yee Yau, Yvette Wai-Man Lai, Kitty Sau-Chun |
| hCoV-19/England/ALDP-1278E31/2021 | EPI_ISL_1018529 | 2021-02-09 | Lighthouse Lab in Alderley Park | Wellcome Sanger Institute for the COVID-19 Genomics UK (COG-UK) Consortium | Jacquelyn Wynn, Mairead Hyland, The Lighthouse Lab in Alderley Park and Alex Alderton, Roberto Amato, Jeffrey Barrett, Sonia Goncalves, Ewan Harrison, David K. Jackson, Ian Johnston, Dominic Kwiatkowski, Cordelia Langford, John Sillitoe on behalf of the W |
| hCoV-19/NorthMacedonia/33023/2021 | EPI_ISL_1020141 | 2021-01-30 | University Clinic of Nephrology- Skopje | Engineering and Biotechnology "Georgi D. Efremov,Äù , Macedonian Academy of Sciences and Arts | Aleksandar J. Dimovski, Dijana Plasheska-Karanfilska, Predrag Noveski, Gjorgji Bozinovski, Milena Jakimovska |
| hCoV-19/Australia/WA565/2020 | EPI_ISL_794673 | 2020-12-31 | PathWest Laboratory Medicine WA | PathWest Laboratory Medicine WA Microbial Surveillance Unit | PathWest Laboratory Medicine WA Microbial Surveillance Unit |
| hCoV-19/Colombia/DC-CIMBIUR-D18/2020 | EPI_ISL_1040923 | 2020-07-13 | Grupo de Investigaci√≥n en Enfermedades Tropicales del Ej√©rcito (GINETEJ), Laboratorio de Referencia e Investigaci√≥n, Direcci√≥n de Sanidad Ej√©rcito, Bogot√°, Colombia | Centro de Investigaciones en Microbiolog√≠a y Biotecnolog√≠a-UR (CIMBIUR), Facultad de Ciencias Naturales, Universidad del Rosario, Bogot√°, Colombia | Juan David Ram√≠rez, Marina Mu√±oz, Nathalia Ballesteros, Luz H. Pati√±o, Sergio Casta√±eda, Carlos A. Rinc√≥n, Claudia P. Mendez, Magda C. Oliveros, Julie J. Perez, Elizabeth K. Marquez, Frank de los Santos Ortiz, Camilo Correa-Cardenas, Maria C. Duque, |
| hCoV-19/USA/WA-S4841/2021 | EPI_ISL_1049689 | 2021-01-12 | Altius Institute for Biomedical Sciences | Seattle Flu Study | Deborah A. Nickerson, Chris D. Frazar, Jover Lee, Benjamin Pelle, Erica Ryke, Matthew Richardson, Amanda Adler, Elisabeth Brandstetter, Peter D. Han, Kairsten Fay, Misja Ilcisin, Kirsten Lacombe, Thomas R. Sibley, Melissa Truong, Caitlin R. Wolf, Ryan Ale |
| hCoV-19/Switzerland/ZH-ETHZ-471383/2021 | EPI_ISL_1059842 | 2021-01-28 | Viollier AG | Department of Biosystems Science and Engineering, ETH Z√ºrich | Chaoran Chen, Sarah Nadeau, Ivan Topolsky, Emmanouil Dermitzakis, Keith Harshman, Ioannis Xenarios, Henri Pegeot, Lorenzo Cerutti, Deborah Penet, Philipp Jablonski, Lara Fuhrmann, David Dreifuss, Katharina Jahn, Christiane Beckmann, Maurice Redondo, Olivi |
| hCoV-19/Australia/QLD1611/2021 | EPI_ISL_1061309 | 2021-02-20 | Public Health Virology-Forensic and Scientific Services | Public Health Virology-Forensic and Scientific Services | Son Nguyen |
| hCoV-19/SouthAfrica/NICD-N6939/2021 | EPI_ISL_2662589 | 2021-05-12 | TAMBO MEMORIAL LABORATORY | Diseases of the National Health Laboratory Service | Amoako DG, Everatt J, Scheepers C, Mohale T, Ntuli N, Mahlangu B, Mnguni A, Ismail A, Bhiman JN |
| hCoV-19/SouthKorea/KDCA1344/2020 | EPI_ISL_1063653 | 2020-07-05 | Division of Emerging Infectious Diseases, Bureau of Infectious Diseases Diagnosis Control, Korea Disease Control and Prevention Agency | Diseases, Bureau of Infectious Diseases Diagnosis Control, Korea Disease Control and Prevention Agency | Ae Kyung Park, Il-Hwan Kim, Heui Man Kim, Jeong-Min Kim, Namjoo Lee, Chaeyoung Lee, Sang Hee Woo, Eun-Jin Kim |
| hCoV-19/USA/NM-UNM-01857/2021 | EPI_ISL_1064216 | 2021-01-17 | New Mexico Department of Health Scientific Laboratory | Center for Global Health, University of New Mexico Health Sciences Center | Daryl Domman, Kurt Schwalm, Twila Kunde, Joseph Hicks, Anastacia Griego, Michael Edwards, Darrell Dinwiddie |
| hCoV-19/USA/DE-B1099371/2021 | EPI_ISL_2662752 | 2021-05-26 | DPHL | Delaware Public Health Lab | Rebecca Savage |
| hCoV-19/Australia/WA621/2021 | EPI_ISL_1069390 | 2021-02-04 | PathWest Laboratory Medicine WA | PathWest Laboratory Medicine WA Microbial Surveillance Unit | PathWest Laboratory Medicine WA Microbial Surveillance Unit |

| | | | | | |
|---|---|---|---|---|---|
| hCoV-19/Israel/CVL-2681/2021 | EPI_ISL_1073480 | 2021-01-04 | Israel Central Virology laboratory | Israel National Consortium for SARS-CoV-2 sequencing | Neta Zuckerman, Efrat Dahan Bucris, Michal Mandelboim, Dana Bar-Ilan, Oran Erster, Tzvia Mann, Omer Murik, David A. Zeevi, Assaf Rokney, Joseph Jaffe, Eva Nachum, Maya Davidovich Cohen, Ephraim Fass, Gal Zizelski Valenci, Mor Rubinstein, Efrat Rorman, Is |
| hCoV-19/Israel/CVL-870/2020 | EPI_ISL_1073591 | 2020-12-29 | Israel Central Virology laboratory | Israel National Consortium for SARS-CoV-2 sequencing | Neta Zuckerman, Efrat Dahan Bucris, Michal Mandelboim, Dana Bar-Ilan, Oran Erster, Tzvia Mann, Omer Murik, David A. Zeevi, Assaf Rokney, Joseph Jaffe, Eva Nachum, Maya Davidovich Cohen, Ephraim Fass, Gal Zizelski Valenci, Mor Rubinstein, Efrat Rorman, Is |
| hCoV-19/Turkey/HSGM-D290/2021 | EPI_ISL_1073728 | 2021-02-08 | Ministry of Health Turkey | Ministry of Health Turkey | Fatma Bayrakdar, Yasemin Co≈ügun, S√ºleyman Yalcin, Ay≈üe Ba≈üak Alta≈ü, G√ºlay Korukluoƒülu |
| hCoV-19/Sweden/SUS0000005/2021 | EPI_ISL_1081847 | 2021-02-01 | Clinical Microbiology, Infection Prevention and Control | Section for Molecular Diagnostics | Bj√∂rn Hallstr√∂m, Jonas Bj√∂rkman |
| hCoV-19/USA/CT-Yale-1369/2020 | EPI_ISL_1091839 | 2020-08-19 | Connecticut Veterans' Affairs Hospital | Grubaugh Lab - Yale School of Public Health | Mary Petrone, Joseph Fauver, Tara Alpert, Chantal Vogels, Isabel Ott, Shaili Gupta, Danielle Plank, Nathan Grubaugh |
| hCoV-19/Colombia/CES-INS-VG-248/2020 | EPI_ISL_1092005 | 2020-12-26 | Laboratorio de Salud Publica de Cesar | Instituto Nacional de Salud-Direcci√≥n de Investigaci√≥n en Salud P√∫blica | Katherine Laiton-Donato, Diego A. √Ålvarez-D√≠az, Carlos Franco-Mu√±oz, Hector Alejandro Ruiz-Moreno, Maria T. Herrera-Sep√∫lveda, Diego Andr√©s Prada, Jhonnatan Reales-Gonz√°lez, Sheryll Corchuelo, Julian Naizaque, Gerardo Santamar√≠a, Magdalena Wiesner, |
| hCoV-19/Peru/ICA-INS-267/2020 | EPI_ISL_1093158 | 2020-09-22 | Laboratorio de Referencia Nacional de Virus Respiratorio. Instituto Nacional de Salud Per√∫ | Laboratorio de Referencia Nacional de Biotecnolog√≠a y Biolog√≠a Molecular. Instituto Nacional de Salud Per√∫ | Carlos Padilla Rojas, Karolyn Vega Chozo, Luis Barcena, Priscila Lope Pari, Omar Caceres Rey, Marco Galarza Perez, Maribel Huaringa Nu√±ez, Johanna Balbuena Torrez, Henri Bailon Calderon, Nancy Rojas Serrano |
| hCoV-19/Portugal/PT8977/2021 | EPI_ISL_2695184 | 2021-06-07 | ULS Litoral Alentejano | Instituto Nacional de Saude (INSA) and Institute of Biomedicine (iBiMed), Universidade de Aveiro | Borges et al |
| hCoV-19/SouthAfrica/NICD-N9047/2021 | EPI_ISL_2695576 | 2021-06-07 | POLOKWANE MANKWENG HOSPITAL | Diseases of the National Health Laboratory Service | Amoako DG, Everatt J, Scheepers C, Mohale T, Ntuli N, Mahlangu B, Mnguni A, Ismail A, Bhiman JN |
| hCoV-19/SouthAfrica/NICD-N8123/2021 | EPI_ISL_2695627 | 2021-05-17 | ROB FERREIRA LABORATORY | Diseases of the National Health Laboratory Service | Amoako DG, Everatt J, Scheepers C, Mohale T, Ntuli N, Mahlangu B, Mnguni A, Ismail A, Bhiman JN |
| hCoV-19/SouthAfrica/NICD-N8195/2021 | EPI_ISL_2695675 | 2021-03-10 | KIMBERLEY LABORATORY | Diseases of the National Health Laboratory Service | Amoako DG, Everatt J, Scheepers C, Mohale T, Ntuli N, Mahlangu B, Mnguni A, Ismail A, Bhiman JN |
| hCoV-19/SouthAfrica/NICD-N8205/2021 | EPI_ISL_2695681 | 2021-03-15 | KIMBERLEY LABORATORY | Diseases of the National Health Laboratory Service | Amoako DG, Everatt J, Scheepers C, Mohale T, Ntuli N, Mahlangu B, Mnguni A, Ismail A, Bhiman JN |
| hCoV-19/SouthAfrica/NICD-N8383/2021 | EPI_ISL_2695735 | 2021-06-05 | HELEN JOSEPH | Diseases of the National Health Laboratory Service | Amoako DG, Everatt J, Scheepers C, Mohale T, Ntuli N, Mahlangu B, Mnguni A, Ismail A, Bhiman JN |
| hCoV-19/Australia/NSW4745/2021 | EPI_ISL_2695792 | 2021-06-09 | Area of Virology, Serology and Virology Division (SAViD), New South Wales Health Pathology Randwick | Virology, Serology and Virology Division (SAViD), New South Wales Health Pathology Randwick | Foster, C.; Au, J.; Ruiz Silva, M.; Deveson, I.; Bull, R.; Van Hal, S.; Rawlinson, W. |
| hCoV-19/Peru/LIM-INS-549/2020 | EPI_ISL_1111442 | 2020-08-24 | Laboratorio de Referencia Nacional de Enteropat√≥genos. Instituto Nacional de Salud del Per√∫ | Laboratorio de Referencia Nacional de Enteropat√≥genos. Instituto Nacional de Salud del Per√∫ | Ronnie Gavilan Chavez, Junior Caro Castro, Willi Quino Sifuentes, Veronica Hurtado Vela, Iris Silva Molina, Fiorella Orellana Peralta |

| | | | | | |
|---|---|---|---|---|---|
| hCoV-19/Australia/QLD1962/2021 | EPI_ISL_2695787 | 2021-06-25 | Queensland Health Forensic and Scientific Services | Queensland Health Forensic and Scientific Services | Son Nguyen |
| hCoV-19/Ukraine/ChVir23535_19/2021 | EPI_ISL_1112299 | 2021-01-10 | Public Health Center of Ukraine | ChariteÃÅ UniversitaÃàtsmedizin Berlin, Institute of Virology | Victor M Corman, Barbara M√ºhlemann, J√∂rn Beheim-Schwarzbach, Talitha Veith, Julia Schneider, Terry Jones, Roman Rodyna, Ihor Kuzin, Liudmyla Chernenko, Iryna Demchyshyna, Christian Drosten |
| hCoV-19/Ukraine/ChVir23535_20/2021 | EPI_ISL_1112300 | 2021-01-10 | Public Health Center of Ukraine | ChariteÃÅ UniversitaÃàtsmedizin Berlin, Institute of Virology | Victor M Corman, Barbara M√ºhlemann, J√∂rn Beheim-Schwarzbach, Talitha Veith, Julia Schneider, Terry Jones, Roman Rodyna, Ihor Kuzin, Liudmyla Chernenko, Iryna Demchyshyna, Christian Drosten |
| hCoV-19/USA/WA-S5208/2021 | EPI_ISL_1114365 | 2021-02-21 | Washington State Department of Health | Seattle Flu Study | Deborah A. Nickerson, Chris D. Frazar, Jover Lee, Benjamin Pelle, Erica Ryke, Matthew Richardson, Amanda Adler, Elisabeth Brandstetter, Peter D. Han, Kairsten Fay, Misja Ilcisin, Kirsten Lacombe, Thomas R. Sibley, Melissa Truong, Caitlin R. Wolf, Romesh G |
| hCoV-19/Brazil/PE-UFPE011/2020 | EPI_ISL_1117392 | 2020-06-25 | Nucleo de Pesquisa em Inovacao Terapeutica - UFPE | LABBE, Federal University of Pernambuco | Wilson Jose da Silva Junior, Marcos da Silveira Regueira Neto, Heidi Lacerda Alves da Cruz, Bruno Sampaio, Reginaldo Goncalves de Lima Neto, Maira Galdino da Rocha Pitta, Michelly Cristiny Pereira, Marco Katzenberger, Valdir de Queiroz Balbino |
| hCoV-19/Austria/CeMM4028/2021 | EPI_ISL_1117959 | 2021-01-20 | HG Pharma GmbH | Bergthaler laboratory, CeMM Research Center for Molecular Medicine of the Austrian Academy of Sciences | Lukas Endler, Anna Schedl, Thomas Penz, Benedikt Agerer, Maelle Le Moing, Michael Schuster, Bekir Erguner, Jan Laine, Martin Senekowitsch, Christoph Bock, Andreas Bergthaler |
| hCoV-19/Switzerland/SO-ETHZ-501543/2020 | EPI_ISL_1119303 | 2020-11-10 | Viollier AG | Department of Biosystems Science and Engineering, ETH Z√ºrich | Chaoran Chen, Sarah Nadeau, Catharine Aquino, Ivan Topolsky, Philipp Jablonski, Lara Fuhrmann, David Dreifuss, Katharina Jahn, Andreia Cabral de Gouvea, Maria Domenica Moccia, Simon Gr√ºter, Timothy Sykes, Lennart Opitz, Griffin White, Laura Neff, Doris P |
| hCoV-19/Australia/NSW-R0092/2021 | EPI_ISL_1121981 | 2021-02-24 | New South Wales Health Pathology Royal Prince Alfred Hospital | Microbiology RPAH | Foster, C.; Au, J.; Ruiz Silva, M.; Deveson, I.; Bull, R.; Van Hal, S.; Rawlinson, W. |
| hCoV-19/Japan/TY8-612-P1/2021 | EPI_ISL_1123289 | 2021-01-29 | Department of Virology I, National Institute of Infectious Diseases | National Institute of Infectious Diseases | Yudai Kuroda, Tsukasa Yamamoto, Keita Ishijima, Souichi Yamada, Shuetsu Fukushi, Ken Maeda |
| hCoV-19/Australia/NT190/2021 | EPI_ISL_2321161 | 2021-04-18 | Royal Darwin Hospital Pathology | MDU-PHL | Meumann, E., Caly L., Seemann T., Sait, M.L., Druce J., Sherry, N.L. |
| hCoV-19/Switzerland/LU-ETHZ-470164/2021 | EPI_ISL_1130134 | 2021-01-22 | Viollier AG | Department of Biosystems Science and Engineering, ETH Z√ºrich | Christian Beisel, Sarah Nadeau, Chaoran Chen, Ivan Topolsky, Philipp Jablonski, Lara Fuhrmann, David Dreifuss, Katharina Jahn, Rebecca Denes, Mirjam Feldkamp, Ina Nissen, Natascha Santacroce, Elodie Burcklen, Christiane Beckmann, Maurice Redondo, Olivier |
| hCoV-19/SouthAfrica/VIDA-KRISP-CP161203493/2020 | EPI_ISL_1132698 | 2020-12-17 | Vaccines¬†and¬†Infectious Diseases Analytics¬†Research Unit (VIDA) | KRISP, KZn Research Innovation and Sequencing Platform | Baillie Vicky, du Plessis Jeanine, Giandhari Jennifer, Pillay Sureshnee, Naidoo Yeshnee, Tegally Houriiyah, de Oliveira Tulio, Madhi Shabir |

| | | | | | |
|---|---|---|---|---|---|
| hCoV-19/Brazil/MT-LBI39/2021 | EPI_ISL_1133274 | 2021-01-14 | Laboratv≥rio Hermes Pardini | Laboratv≥rio de Biologia Integrativa, Instituto de Civ™ncias Biolv≥gicas, Universidade Federal de Minas Gerais | Filipe Romero Rebello Moreira, Diego Menezes Bonfim, Danielle Alves Gomes Zauli, Joice do Prado Silva, Aline Brito de Lima, Frederico Scott Varella Malta, Alessandro Clayton de Souza Ferreira, Victor Cavalcanti Pardini, Daniel Costa Queiroz, Rafael Marque |
| hCoV-19/Peru/PIU-INS-741/2020 | EPI_ISL_1138422 | 2020-12-29 | Laboratorio de Referencia Nacional de Virus Respiratorio. Instituto Nacional de Salud Perv∫ | Laboratorio de Referencia Nacional de Biotecnologv≠a y Biologv≠a Molecular. Instituto Nacional de Salud Perv∫ | Carlos Padilla Rojas, Karolyn Vega Chozo, Luis Barcena, Priscila Lope Pari, Omar Caceres Rey, Marco Galarza Perez, Maribel Huaringa Nuv±ez, Johanna Balbuena Torrez, Henri Bailon Calderon, Nancy Rojas Serrano |
| hCoV-19/Chile/RM-209059/2020 | EPI_ISL_1167678 | 2020-10-16 | Genetica Molecular and Subdepartamento de Virologia ISP Chile | Instituto de Salud Publica de Chile | Javier Tognarelli, Karen Orostica, Barbara Parra, Loredana Arata, Jaime Lagos, Gisselle Barra, Patricia Bustos, Rodrigo Fasce, Andres Castillo, Jorge Fernandez |
| hCoV-19/Chile/RM-215103/2020 | EPI_ISL_1167686 | 2020-10-25 | Genetica Molecular and Subdepartamento de Virologia ISP Chile | Instituto de Salud Publica de Chile | Javier Tognarelli, Karen Orostica, Barbara Parra, Loredana Arata, Jaime Lagos, Gisselle Barra, Patricia Bustos, Rodrigo Fasce, Andres Castillo, Jorge Fernandez |
| hCoV-19/Chile/RM-266418/2020 | EPI_ISL_1167921 | 2020-12-30 | Genetica Molecular and Subdepartamento de Virologia ISP Chile | Instituto de Salud Publica de Chile | Javier Tognarelli, Karen Orostica, Barbara Parra, Loredana Arata, Jaime Lagos, Gisselle Barra, Patricia Bustos, Rodrigo Fasce, Andres Castillo, Jorge Fernandez |
| hCoV-19/Chile/RM-266423/2020 | EPI_ISL_1167925 | 2020-12-30 | Genetica Molecular and Subdepartamento de Virologia ISP Chile | Instituto de Salud Publica de Chile | Javier Tognarelli, Karen Orostica, Barbara Parra, Loredana Arata, Jaime Lagos, Gisselle Barra, Patricia Bustos, Rodrigo Fasce, Andres Castillo, Jorge Fernandez |
| hCoV-19/Japan/PG-20110/2021 | EPI_ISL_1169029 | 2021-01-13 | Department of Infectious Diseases, Kobe Institute of Health | Department of Infectious Diseases, Kobe Institute of Health | Ryohei Nomoto, Noriko Nakanishi, Tomotada Iwamoto, Tsuyoshi Sekizuka, Kentaro Itokawa, Rina Tanaka, Masanori Hashino, Makoto Kuroda |
| hCoV-19/NorthernMarianaIslands/CDC-2-3831042/2020 | EPI_ISL_1169508 | 2020-07-14 | Commonwealth Healthcare Center | Genomics and Discovery, Respiratory Viruses Branch, Division of Viral Diseases, Centers for Disease Control and Prevention | Krista Queen, Yan Li, Ying Tao, Jing Zhang, Anna Uehara, Anna Montmayeur, Clinton R. Paden, Peter W. Cook, Rachel Marine, Mili Sheth, Jasmine Padilla, Sarah Nobles, Mark Burroughs, Lori Rowe, Haibin Wang, Ben L. Rambo-Martin, Dhwani Batra, Justin Lee, Sux |
| hCoV-19/NorthernMarianaIslands/CDC-2-3831108/2020 | EPI_ISL_1169519 | 2020-09-26 | Commonwealth Healthcare Center | Genomics and Discovery, Respiratory Viruses Branch, Division of Viral Diseases, Centers for Disease Control and Prevention | Krista Queen, Yan Li, Ying Tao, Jing Zhang, Anna Uehara, Anna Montmayeur, Clinton R. Paden, Peter W. Cook, Rachel Marine, Mili Sheth, Jasmine Padilla, Sarah Nobles, Mark Burroughs, Lori Rowe, Haibin Wang, Ben L. Rambo-Martin, Dhwani Batra, Justin Lee, Sux |
| hCoV-19/NorthernMarianaIslands/CDC-2-3831103/2020 | EPI_ISL_1169540 | 2020-12-05 | Commonwealth Healthcare Center | Genomics and Discovery, Respiratory Viruses Branch, Division of Viral Diseases, Centers for Disease Control and Prevention | Krista Queen, Yan Li, Ying Tao, Jing Zhang, Anna Uehara, Anna Montmayeur, Clinton R. Paden, Peter W. Cook, Rachel Marine, Mili Sheth, Jasmine Padilla, Sarah Nobles, Mark Burroughs, Lori Rowe, Haibin Wang, Ben L. Rambo-Martin, Dhwani Batra, Justin Lee, Sux |
| hCoV-19/Australia/ACT0095/2021 | EPI_ISL_1170946 | 2021-03-03 | ACT Pathology | Schwessinger Lab | Ashley Jones, Benjamin Schwessinger, Robert Lanfear, Megan McDonald, Ming-Dao Chia, Kevin Murray, Robyn N Hall, Craig Kennedy, Karina Kennedy |
| hCoV-19/Kuwait/JAH3101610/2021 | EPI_ISL_2556115 | 2021-05-31 | MOH - Jaber Al-Ahmad Hospital (Innovation Research Laboratory) | MOH - Jaber Al-Ahmad Hospital (Innovation Research Laboratory) | Salman Al-Sabah , Mohammad Alghounaim |
| hCoV-19/Turkey/HSGM-5121/2021 | EPI_ISL_1183620 | 2021-02-19 | Ministry of Health Turkey | Ministry of Health Turkey | Fatma Bayrakdar, Yasemin Cosgun, Suleyman Yalcin, Gulay Korukluoglu |

| | | | | | |
|---|---|---|---|---|---|
| hCoV-19/Turkey/HSGM-5211/2021 | EPI_ISL_1183647 | 2021-02-19 | Ministry of Health Turkey | Ministry of Health Turkey | Fatma Bayrakdar, Yasemin Cosgun, Suleyman Yalcin, Gulay Korukluoglu |
| hCoV-19/Australia/NSW-R0120/2021 | EPI_ISL_1184097 | 2021-03-03 | New South Wales Health Pathology Royal Prince Alfred Hospital | Microbiology RPAH | Foster, C.; Au, J.; Ruiz Silva, M.; Deveson, I.; Bull, R.; Van Hal, S.; Rawlinson, W. |
| hCoV-19/Belgium/AZSL_2102-30973/2021 | EPI_ISL_2322747 | 2021-02-24 | AZ Sint-Lucas Gent | AZ Sint-Lucas Gent | Jos Van Acker, Charlotte Verfaillie, Elke Vanlaere, Klara De Rauw |
| hCoV-19/Zimbabwe/ZW-34550/2020 | EPI_ISL_1191832 | 2020-10-19 | National Microbiology Reference Laboratory | Quadram Institute Bioscience | Tapfumanei Mashe, Faustinos T Takawira, Hlanai Gumbo, Kenneth K Maeka, Agnes Juru, Charles Nyagupe, Sekesai Zinyowera, Muchaneta Mugabe, Thanh Le Viet, Justin O‚ÄôGrady, Gemma Kay, David Baker, Gaetan Thilliez, Ana-Victoria Gutierrez, Robert Kingsley, Leo |
| hCoV-19/Sweden/SE00163874/2021 | EPI_ISL_1192763 | 2021-01-18 | Swedish national genomic surveillance program of SARS-CoV-2 | The Public Health Agency of Sweden | Swedish national genomic surveillance program of SARS-CoV-2 |
| hCoV-19/India/WB-1931300253474/2021 | EPI_ISL_2503194 | 2021-03-30 | INSACOG-WB | National Institute of Biomedical Genomics ‚Äì INSACOG | Arindam Maitra, Bhaswati Bandyopadhyay, Nidhan Kumar Biswas, Tamal Ghosh, Sreedhar Chinnaswamy, Ajay Chakraborti, Saumitra Das |
| hCoV-19/India/WB-1931501009882/2021 | EPI_ISL_2503401 | 2021-03-29 | INSACOG-WB | National Institute of Biomedical Genomics ‚Äì INSACOG | Arindam Maitra, Bhaswati Bandyopadhyay, Nidhan Kumar Biswas, Tamal Ghosh, Sreedhar Chinnaswamy, Ajay Chakraborti, Saumitra Das |
| hCoV-19/India/WB-1930400407225/2021 | EPI_ISL_2503421 | 2021-03-22 | INSACOG-WB | National Institute of Biomedical Genomics ‚Äì INSACOG | Arindam Maitra, Bhaswati Bandyopadhyay, Nidhan Kumar Biswas, Tamal Ghosh, Sreedhar Chinnaswamy, Ajay Chakraborti, Saumitra Das |
| hCoV-19/India/DL-NCDC-1209135/2021 | EPI_ISL_2504415 | 2021-02-27 | National Centre for Disease Control (NCDC) Biotechnology Division, Delhi | NCDC Delhi, Biotechnology Division INSACOG | Robin Marwal, Mahesh S Dhar, Kalaiarasan Ponnusamy, Meena Datta, Priyanka Singh, Uma Sharma, Manoj K Singh, Radhakrishnan V. S, Hemlata Lall, Hema Gogia, Preeti Madan, Sandhya Kabra, Sujeet K Singh, Partha Rakshit |
| hCoV-19/India/JK-NCDC-INSACOG-1309135/2021 | EPI_ISL_2504613 | 2021-03-07 | Biotechnology Division, NCDC Delhi | NCDC Delhi, Biotechnology Division | Radhakrishnan V. S, Robin Marwal, Mahesh S Dhar, Kalaiarasan Ponnusamy, Meena Datta, Priyanka Singh, Uma Sharma, Manoj K Singh, Hemlata Lall, Hema Gogia, Preeti Madan, Sandhya Kabra, Sujeet K Singh, Partha Rakshit |
| hCoV-19/Australia/VIC16724/2020 | EPI_ISL_663326 | 2020-08-07 | Microbiological Diagnostic Unit - Public Health Laboratory (MDU-PHL) | MDU-PHL | Seemann T., Schultz M.B., Sait, M.L., Sherry, N.L. |
| hCoV-19/England/PHEP-005422/2021 | EPI_ISL_3779442 | 2021-07-13 | Respiratory Virus Unit, Microbiology Services Colindale, Public Health England | COVID-19 Genomics UK (COG-UK) Consortium | PHE Covid Sequencing Team |
| hCoV-19/USA/NC-CDC-ASC210159843/2021 | EPI_ISL_3325195 | 2021-07-19 | Aegis Sciences Corporation | Centers for Disease Control and Prevention Division of Viral Diseases, Pathogen Discovery | Dakota Howard, Dhwani Batra, Peter W. Cook, Kara Moser, Adrian Paskey, Jason Caravas, Benjamin Rambo-Martin, Shatavia Morrison, Christopher Gulvick, Scott Sammons, Yvette Unoarumhi, Darlene Wagner, Matthew Schmerer, Cyndi Clark, Patrick Campbell, Rob Case |

| | | | | | |
|---|---|---|---|---|---|
| hCoV-19/USA/FL-CDC-ASC210277165/2021 | EPI_ISL_4056830 | 2021-08-05 | Aegis Sciences Corporation | Centers for Disease Control and Prevention Division of Viral Diseases, Pathogen Discovery | Dakota Howard,Dhwani Batra,Peter Cook,Kara Moser,Adrian Paskey,Jason Caravas,Benjamin Rambo-Martin,Shatavia Morrison,Christopher Gulvick,Scott Sammons,Yvette Unoarumhi,Darlene Wagner,Matthew Schmerer,Cyndi Clark,Patrick Campbell,Rob Case,Vikramsinha Ghorp |
| hCoV-19/India/UP-CDFD-MBU2355/2021 | EPI_ISL_4057119 | 2021-05-07 | Banaras Hindu University | CDFD | Murali Bashyam, Asmita Gupta, Vinay Donipadi, Reelina Basu, Gyaneshwer Chaubey, Divya Vashisht, Ashwin Dalal |
| hCoV-19/USA/NY-PRL-2021_0803_02F04/2021 | EPI_ISL_3332094 | 2021-07-26 | Pandemic Response Lab - NYC | Pandemic Response Lab, R&D | Henry Lee, Michael Hammerling, Melissa Hopkins, Cybill del Castillo, Shinyoung Clair Kang, William Ward, Pradeep Bugga, Sol Rey, Dylan Law, Alex Carpio, Isabel Fernandez Escapa, Haiping Hao, Jon Laurent |
| hCoV-19/Brazil/PR-FIOCRUZ-45422/2021 | EPI_ISL_4061438 | 2021-07-19 | Laboratorio Central de Saude Publica do Estado do Parana (LACEN/PR) | Laboratory of Respiratory Viruses and Measles, Oswaldo Cruz Institute, FIOCRUZ | Paola Resende, Luciana Appolinario, Fernando Motta, Anna Carolina Paixao, Ighor Leonardo Arantes, Agatha Soares, Ana Carolina Mendonca, Alice Sampaio Rocha, Taina Venas, Elisa Cavalcante Pereira, Renata Serrano Lopes, Irina Riediger, Marilda Siqueira on b |
| hCoV-19/Oman/rega-OM-90/2021 | EPI_ISL_4062648 | 2021-01-06 | Sultan Qaboos University Hospital, Department of Microbiology & Immunology, Molecular Biology Section | KU Leuven, Rega Institute, Clinical and Epidemiological Virology | Tony Wawina-Bokalanga, Zakaryia Almuharrmi, Khuloud Al Maamari, Fatma BaAlawi, Abdullah Balkhair, Ishraq Al Kindi, Azza Alqayoudhi, Faiza Alnamaani, Omar Balkhair, Piet Maes |
| hCoV-19/Oman/rega-OM-154/2021 | EPI_ISL_4062992 | 2021-05-07 | Sultan Qaboos University Hospital, Department of Microbiology & Immunology, Molecular Biology Section | KU Leuven, Rega Institute, Clinical and Epidemiological Virology | Tony Wawina-Bokalanga, Zakaryia Almuharrmi, Khuloud Al Maamari, Fatma BaAlawi, Abdullah Balkhair, Ishraq Al Kindi, Azza Alqayoudhi, Faiza Alnamaani, Omar Balkhair, Piet Maes |
| hCoV-19/NewZealand/21MV1453/2021 | EPI_ISL_4063317 | 2021-09-07 | LabPLUS | Institute of Environmental Science and Research (ESR) | Rachel Boyle, SallyAnn Harbison, Olivia Stroeven, Xiaoyun Ren, Matt Storey, Nikki Freed, Muhammad Faisal, Jing Wang, Hermes Perez, Anja Werno, Antje van der Linden, Arlo Upton, Chris Mansell, David Hammer, Dragana Drinkovic, Gary McAuliffe, Hana Sofia And |
| hCoV-19/Ethiopia/AFCPH_IDSWH_28972/2021 | EPI_ISL_4063323 | 2021-08-09 | Armed Force Comprehensive Specialized Hospital laboratory departmentÔºåEthiopian Defense Force Health Main Department | Promotion and Disease Prevention Department, Ethiopian Defense Force Health Main Department; Department of Infectious Diseases, Southwest Hospital, Army Medical University | Yacob Mohamed Yusuf, Pei Zhou, Hu Luo, Xiuhua Wang, Fengming Sun, Alemu Tike Debela, Guohong Deng |
| hCoV-19/Australia/NSW-RPAH-1460/2021 | EPI_ISL_4063324 | 2021-08-31 | New South Wales Health Pathology Royal Prince Alfred Hospital | Microbiology RPAH | Foster, C.; Au, J.; Ruiz Silva, M.; Deveson, I.; Bull, R.; Van Hal, S.; Rawlinson, W. |
| hCoV-19/TrinidadandTobago/63609/2021 | EPI_ISL_4063331 | 2021-08-06 | Trinidad Public Health Laboratory | Carrington Lab, Department of Preclinical Sciences, Faculty of Medical Sciences, The University of the West Indies, St Augustine Campus | Nikita S. D. Sahadeo, Arianne Brown-Jordan, Sarah Hill, Vernie Ramkissoon, Anushka Ramjag, Roshan Parasram, Avery Hinds, Risha Singh, SueMin Nathaniel, Nuno Faria, Oliver Pybus, Christopher Oura, Gabriel Escobar, Christine V. F. Carrington |

| | | | | | |
|---|---|---|---|---|---|
| hCoV-19/USA/IN-CDC-LC0038455/2021 | EPI_ISL_4381524 | 2021-03-30 | Laboratory Corporation of America | Centers for Disease Control and Prevention Division of Viral Diseases, Pathogen Discovery | Dakota Howard,Dhwani Batra,Peter Cook,Jason Caravas,Benjamin Rambo-Martin,Scott Sammons,Yvette Unoarumhi,Matthew Schmerer,Kristine Lacek,Tymeckia Kendall,Victoria Caban Figueroa,Shatavia Morrison,Christopher Gulvick,Minoo Agarwal,Eyad Almasri,Debbie Boles |
| hCoV-19/USA/WA-S10811/2021 | EPI_ISL_4071457 | 2021-06-30 | Seattle Flu Study | Seattle Flu Study | Deborah A. Nickerson, Chris D. Frazar, Jover Lee, Benjamin Pelle, Erica Ryke, Matthew Richardson, Amanda Adler, Elisabeth Brandstetter, Peter D. Han, Kairsten Fay, Misja Ilcisin, Kirsten Lacombe, Thomas R. Sibley, Melissa Truong, Caitlin R. Wolf, Michael |
| hCoV-19/USA/WA-S10645/2021 | EPI_ISL_4071465 | 2021-07-24 | Overlake Hospital | Seattle Flu Study | Deborah A. Nickerson, Chris D. Frazar, Jover Lee, Benjamin Pelle, Erica Ryke, Matthew Richardson, Amanda Adler, Elisabeth Brandstetter, Peter D. Han, Kairsten Fay, Misja Ilcisin, Kirsten Lacombe, Thomas R. Sibley, Melissa Truong, Caitlin R. Wolf, Romesh G |
| hCoV-19/SouthAfrica/NICD-N2814/2021 | EPI_ISL_3342331 | 2021-01-10 | Mmametlhake laboratory | Diseases of the National Health Laboratory Service | Amoako DG, Everatt J, Scheepers C, Mohale T, Ntuli N, Mahlangu B, Mnguni A, Ismail A, Bhiman JN |
| hCoV-19/Aruba/AW-RIVM-55234/2021 | EPI_ISL_4077242 | 2021-08-05 | Dutch COVID-19 response team | National Institute for Public Health and the Environment (RIVM) | Adam Meijer, Harry Vennema, Dirk Eggink, Jeroen Cremer, Sharon van den Brink, Bas van der Veer, AnneMarie van den Brandt, Lisa Wijsman, Kim Freriks, Ryanne Jaarsma, Eun√≠ce Then, Lynn Aarts, Sanne Bos, Stijn van Rossum, Florian Zwagemaker, Dennis Schmitz, |
| hCoV-19/Guam/Yigo-USAFSAM-S4904/2021 | EPI_ISL_4078922 | 2021-07-26 | US Air Force School of Aerospace Medicine | US Air Force School of Aerospace Medicine | Anthony Fries, Deanna Muehleman, Jennifer Meyer, William Gruner, Kelsey Lanter, Carol Garrett, Sarah Purves, Clarise Starr, Elizabeth Macias |
| hCoV-19/Norway/17391/2021 | EPI_ISL_4080004 | 2021-08-13 | Medical Microbiology Unit, Department for Laboratory Medicine, Drammen Hospital, Vestre Viken Health Trust, | Norwegian Institute of Public Health, Department of Virology | Kathrine Stene-Johansen, Kamilla Heddeland Instefjord, Hilde Elshaug, Garcia Llorente Ignacio, Jon Br√•te, Engebretsen Serina Beate,Pedersen Benedikte Nevjen, Line Victoria Moen, Debech Nadia, Atiya R Ali,Marie Paulsen Madsen, Rasmus Riis Kopperud, Hilde |
| hCoV-19/Peru/CAL-INS-4240/2021 | EPI_ISL_4080297 | 2021-08-04 | Laboratorio de Referencia Nacional de Virus Respiratorios. Centro Nacional de Salud Publica. Instituto Nacional de Salud Peru. | Laboratorio de Referencia Nacional de Virus Respiratorios. Centro Nacional de Salud Publica. Instituto Nacional de Salud Peru. | Carlos Padilla Rojas, Veronica Hurtado Vela, Orson Mestanza Millones, Iris Silva Molina, Luis Barcena Flores, Sandra Morales Ruiz, Steve Acedo Lazo, Wendy Lizarraga Olivares, Victor Jimenez Vasquez, Sara Gordillo Vilchez, Nieves Sevilla Casta√±eda, Prince |
| hCoV-19/Peru/CAL-INS-4241/2021 | EPI_ISL_4080298 | 2021-08-04 | Laboratorio de Referencia Nacional de Virus Respiratorios. Centro Nacional de Salud Publica. Instituto Nacional de Salud Peru. | Laboratorio de Referencia Nacional de Virus Respiratorios. Centro Nacional de Salud Publica. Instituto Nacional de Salud Peru. | Carlos Padilla Rojas, Veronica Hurtado Vela, Orson Mestanza Millones, Iris Silva Molina, Luis Barcena Flores, Sandra Morales Ruiz, Steve Acedo Lazo, Wendy Lizarraga Olivares, Victor Jimenez Vasquez, Sara Gordillo Vilchez, Nieves Sevilla Casta√±eda, Prince |
| hCoV-19/Norway/17499/2021 | EPI_ISL_4080366 | 2021-08-17 | Ostfold Hospital Trust - Kalnes, Centre for Laboratory Medicine, Section for gene technology and infection serology | Norwegian Institute of Public Health, Department of Virology | Kathrine Stene-Johansen, Kamilla Heddeland Instefjord, Hilde Elshaug, Garcia Llorente Ignacio, Jon Br√•te, Engebretsen Serina Beate,Pedersen Benedikte Nevjen, Line Victoria Moen, Debech Nadia, Atiya R Ali,Marie Paulsen Madsen, Rasmus Riis Kopperud, Hilde |

| | | | | | |
|---|---|---|---|---|---|
| hCoV-19/Argentina/INEI107495/2021 | EPI_ISL_4083179 | 2021-07-10 | Servicio Virosis Respiratorias-Departamento Virologв‰ a-INEI | Instituto Nacional Enfermedades Infecciosas C.G.Malbran | Baumeister E., Avaro M., Benedetti E., Russo M., Dattero ME, Pontoriero A., Cisterna D., Molina V., Perandones C., Tuduri E., De Belder D., Sanchez Loria J., Haim MS., Lorenzo F., Poklepovich T., Campos J. |
| hCoV-19/Argentina/INEI107730/2021 | EPI_ISL_4083327 | 2021-07-14 | Servicio Virosis Respiratorias-Departamento Virolog√≠a-INEI | Instituto Nacional Enfermedades Infecciosas C.G.Malbran | Baumeister E., Avaro M., Benedetti E., Russo M., Dattero ME, Pontoriero A., Cisterna D., Molina V., Perandones C., Tuduri E., De Belder D., Sanchez Loria J., Haim MS., Lorenzo F., Poklepovich T., Campos J. |
| hCoV-19/Argentina/INEI108197/2021 | EPI_ISL_4083493 | 2021-07-21 | Servicio Virosis Respiratorias-Departamento Virolog√≠a-INEI | Instituto Nacional Enfermedades Infecciosas C.G.Malbran | Baumeister E., Avaro M., Benedetti E., Russo M., Dattero ME, Pontoriero A., Cisterna D., Molina V., Perandones C., Tuduri E., De Belder D., Sanchez Loria J., Haim MS., Lorenzo F., Poklepovich T., Campos J. |
| hCoV-19/USA/CA-CDC-FG-056704/2021 | EPI_ISL_3348902 | 2021-07-30 | Fulgent Genetics | Centers for Disease Control and Prevention Division of Viral Diseases, Pathogen Discovery | Dakota Howard, Dhwani Batra, Peter W. Cook, Kara Moser, Adrian Paskey, Jason Caravas, Benjamin Rambo-Martin, Shatavia Morrison, Christopher Gulvick, Scott Sammons, Yvette Unoarumhi, Darlene Wagner, Matthew Schmerer, Harry Gao, Mickey Li, John Gao, Joseph |
| hCoV-19/USA/CA-CDPH-UCSF-CC108/2021 | EPI_ISL_4279954 | 2021-06-03 | California Department of Public Health | Chiu Laboratory, University of California, San Francisco | Charles Chiu, Venice Servellita, Baolin Wang, Mary Kate Morris, Carl Hanson, Debra Wadford |
| hCoV-19/USA/CA-CDPH-UCSF-CC100/2021 | EPI_ISL_4279956 | 2021-02-04 | California Department of Public Health | Chiu Laboratory, University of California, San Francisco | Charles Chiu, Venice Servellita, Baolin Wang, Mary Kate Morris, Carl Hanson, Debra Wadford |
| hCoV-19/Germany/NI-RKI-I-188163/2021 | EPI_ISL_3044397 | 2021-07-10 | Sonic - MVZ Medizinisches Labor Oldenburg | Robert Koch Institute | unknown |
| hCoV-19/USA/UT-UPHL-210911956336/2021 | EPI_ISL_4177334 | 2021-08-31 | Utah Public Health Laboratory | Utah Public Health Laboratory | Erin L. Young, Kelly F. Oakeson, Pooja Gupta, John Arnn, Olinto Linares-Perdomo |
| hCoV-19/USA/SC-MUSC-02580/2021 | EPI_ISL_4280865 | 2021-08-17 | MUSC Molecular Pathology Laboratory | MUSC Molecular Pathology Laboratory | Julie W. Hirschhorn, W. Bailey Glen Jr, Dariusz Pytel, Jaclyn Dunne, Kristen Maurer, Jim Madory, Scott Curry, Frederick S. Nolte |
| hCoV-19/Reunion/HCL0211521767/2021 | EPI_ISL_4281044 | 2021-08-10 | CNR Virus des Infections Respiratoires - France SUD | CNR Virus des Infections Respiratoires - France SUD | Antonin Bal, Gregory Destras, Gwendolyne Burfin, Hadrien Regue, Quentin Semanas, Martine Valette, Bruno Lina, Laurence Josset |
| hCoV-19/France/ARA-HCL0211532056/2021 | EPI_ISL_4282389 | 2021-08-16 | CNR Virus des Infections Respiratoires - France SUD | CNR Virus des Infections Respiratoires - France SUD | Antonin Bal, Gregory Destras, Gwendolyne Burfin, Hadrien Regue, Quentin Semanas, Martine Valette, Bruno Lina, Laurence Josset |
| hCoV-19/Finland/9470/2021 | EPI_ISL_4093168 | 2021-06-27 | Department of Virology and Immunology, University of Helsinki and Helsinki University Hospital, Huslab Finland | Department of Virology, Faculty of Medicine, University of Helsinki, Helsinki, Finland | Teemu Smura, Ravi Kant, Phuoc Truong, Hussein Alburkat, Hannimari Kallio-Kokko, Jenni Virtanen, Maija Suvanto, Essi Korhonen, Sari Hannula, Harri Kangas, Hanna Liimatainen, Satu Kurkela, Hanna Jarva, Maija Lappalainen, Pekka Ellonen, Olli Vapalahti |
| hCoV-19/India/TG-CDFD-Xi667/2021 | EPI_ISL_3356516 | 2021-04-24 | CDFD | CDFD | Murali Bashyam, Asmita Gupta, Reelina Basu, Vinay Donipadi, Divya Vashisht, Ashwin Dalal |
| hCoV-19/India/TG-CDFD-Xi677/2021 | EPI_ISL_3356526 | 2021-04-26 | CDFD | CDFD | Murali Bashyam, Asmita Gupta, Reelina Basu, Vinay Donipadi, Divya Vashisht, Ashwin Dalal |
| hCoV-19/India/TG-CDFD-Xi684/2021 | EPI_ISL_3356533 | 2021-04-28 | CDFD | CDFD | Murali Bashyam, Asmita Gupta, Reelina Basu, Vinay Donipadi, Divya Vashisht, Ashwin Dalal |

| | | | | | |
|---|---|---|---|---|---|
| hCoV-19/USA/AR-UMGC-6944/2021 | EPI_ISL_3358282 | 2021-07-29 | Arkansas Public Health Laboratory, Arkansas Department of Health | University of Minnesota Genomics Center | Daryl M. Gohl, John Garbe, Jaquelyn Kuriger-Laber, Corbin Dirkx |
| hCoV-19/Germany/SN-RKI-I-213551/2021 | EPI_ISL_3889058 | 2021-08-23 | Bioscientia Labor Wermsdorf | Robert Koch Institute | unknown |
| hCoV-19/Turkey/HSGM-E2961/2021 | EPI_ISL_4101643 | 2021-03-09 | Ministry of Health Turkey | Ministry of Health Turkey | Fatma Bayrakdar, Yasemin Cosgun, Suleyman Yalcin, Gulay Korukluoglu |
| hCoV-19/Turkey/HSGM-E3178/2021 | EPI_ISL_4101824 | 2021-03-09 | Ministry of Health Turkey | Ministry of Health Turkey | Fatma Bayrakdar, Yasemin Cosgun, Suleyman Yalcin, Gulay Korukluoglu |
| hCoV-19/India/OR-ILSGS06068/2021 | EPI_ISL_4103095 | 2021-06-19 | Maharaja Krushna Chandra Gajapati Medical College and Hospital, Brahmapur | Institute of Life Sciences-INSACOG | Sunil K. Raghav, Safal Walia, Arup Ghosh, Atimukta Jha, Amol M. Kanampalliwar, Sana Fatma, Shifu Aggarwal, Rupesh Dash, Rajeeb Swain, Punit Prasad, INSACOG Consortium, Ajay Parida |
| hCoV-19/India/CT-ILSGS05872/2021 | EPI_ISL_4103122 | 2021-05-19 | Late Shri Lakhi Ram Agrawal Memorial Government Medical College, Raipur | Institute of Life Sciences-INSACOG | Sunil K. Raghav, Safal Walia, Arup Ghosh, Atimukta Jha, Amol M. Kanampalliwar, Sana Fatma, Shifu Aggarwal, Rupesh Dash, Rajeeb Swain, Punit Prasad, INSACOG Consortium, Ajay Parida |
| hCoV-19/India/OR-ILSGS07720/2021 | EPI_ISL_4104006 | 2021-07-10 | Srirama Chandra Bhanja Medical College and Hospital, Cuttack | Institute of Life Sciences-INSACOG | Sunil K. Raghav, Safal Walia, Arup Ghosh, Atimukta Jha, Amol M. Kanampalliwar, Sana Fatma, Shifu Aggarwal, Rupesh Dash, Rajeeb Swain, Punit Prasad, INSACOG Consortium, Ajay Parida |
| hCoV-19/Chile/RM-131100/2021 | EPI_ISL_3363689 | 2021-07-26 | Genetica Molecular and Subdepartamento de Virologia ISP Chile | Instituto de Salud Publica de Chile | Karen Orostica, Constanza Campano, Barbara Parra, Loredana Arata, Gisselle Barra, Patricia Bustos, Rodrigo Fasce, Javier Tognarelli, Andres Castillo, Soledad Ulloa, Jorge Fernandez |
| hCoV-19/England/ALDP-1AF032B/2021 | EPI_ISL_3363971 | 2021-08-04 | Lighthouse Lab in Alderley Park | Wellcome Sanger Institute for the COVID-19 Genomics UK (COG-UK) Consortium | Jacquelyn Wynn, Mairead Hyland, The Lighthouse Lab in Alderley Park and Alex Alderton, Roberto Amato, Jeffrey Barrett, Sonia Goncalves, Ewan Harrison, David K. Jackson, Ian Johnston, Dominic Kwiatkowski, Cordelia Langford, John Sillitoe on behalf of the W |
| hCoV-19/England/MILK-1AD2E2E/2021 | EPI_ISL_3364748 | 2021-08-02 | Lighthouse Lab in Milton Keynes | Wellcome Sanger Institute for the COVID-19 Genomics UK (COG-UK) Consortium | The Lighthouse Lab in Milton Keynes and Alex Alderton, Roberto Amato, Jeffrey Barrett, Sonia Goncalves, Ewan Harrison, David K. Jackson, Ian Johnston, Dominic Kwiatkowski, Cordelia Langford, John Sillitoe on behalf of the Wellcome Sanger Institute COVID-1 |
| hCoV-19/USA/OR-OHSU-212140635/2021 | EPI_ISL_3825587 | 2021-07-06 | Kaiser Permanente NW Reginal Lab | OHSU MM Lab | Yun Wu |
| hCoV-19/SouthAfrica/KRISP-UFS-K022236/2021 | EPI_ISL_4108311 | 2021-06-09 | UFS Virology | KRISP, KZn Research Innovation and Sequencing Platform | PA Bester, MM Nyaga, P Nthiga, MT Mogotsi, Emmanuel Ogunbayo, D Goedhals, Giandhari J, Pillay S, Yajna Ramphal, Naidoo Yeshnee, Tshabuila Derek, Emmanuel S, Tegally H, Wilkinson E, de Oliveira T |
| hCoV-19/Turkey/HSGM-C7783/2021 | EPI_ISL_3370067 | 2021-07-27 | Ministry of Health Turkey | Ministry of Health Turkey | Fatma Bayrakdar, Yasemin Co≈ügun, S√ºleyman Yalcin, G√ºlay Korukluoƒülu |
| hCoV-19/USA/NV-NSPHL-LV386393/2021 | EPI_ISL_3372417 | 2021-07-21 | Nevada State Public Health Laboratory | Nevada State Public Health Laboratory | Andrew Gorzalski, Mark Pandori |

| | | | | | |
|---|---|---|---|---|---|
| hCoV-19/USA/MA-CDCBI-CRSP_IALOM2NCFEA5FHJD/2021 | EPI_ISL_2991693 | 2021-05-24 | Broad Institute Clinical Research Sequencing Platform | Infectious Disease Program, Broad Institute of Harvard and MIT | Lemieux,J.E., Siddle,K.J., Shaw,B., Adams,G., Pierce,V., Turbett,S., Anahtar,M., Branda,J., Slater,D., Harris,J., Lin,A.E., Gladden-Young,A., Lagerborg,K., Rudy,M., DeRuff,K., Carter,A., Normandin,E., Bauer,M., Reilly,S., Tomkins-Tinch,C., Loreth,C., Chal |
| hCoV-19/USA/MA-CDCBI-CRSP_VFRFXUZRQVCY3VLU/2021 | EPI_ISL_2991884 | 2021-05-26 | Broad Institute Clinical Research Sequencing Platform | Infectious Disease Program, Broad Institute of Harvard and MIT | Lemieux,J.E., Siddle,K.J., Shaw,B., Adams,G., Pierce,V., Turbett,S., Anahtar,M., Branda,J., Slater,D., Harris,J., Lin,A.E., Gladden-Young,A., Lagerborg,K., Rudy,M., DeRuff,K., Carter,A., Normandin,E., Bauer,M., Reilly,S., Tomkins-Tinch,C., Loreth,C., Chal |
| hCoV-19/Brazil/SP-IB_123906/2021 | EPI_ISL_3374338 | 2021-07-28 | AFIP | Instituto Butantan | Dimas Tadeu Covas, Antonio Jorge Martins, Claudia Renata dos Santos Barros, David Schlesinger, Debora Botequio Moretti, Elaine Cristina Marqueze, Elaine Vieira Santos, Evandra Strazza Rodrigues, Heidge Fukumasu, Jayme Augusto de Souza-Neto, Jos√© Salvator |
| hCoV-19/France/ARA-HCL021146926901/2021 | EPI_ISL_4292911 | 2021-08-12 | unknown | CNR Virus des Infections Respiratoires - France SUD | Antonin Bal, Gregory Destras, Gwendolyne Burfin, Hadrien Regue, Quentin Semanas, Martine Valette, Bruno Lina, Laurence Josset |
| hCoV-19/Reunion/HCL021133132501/2021 | EPI_ISL_3382611 | 2021-07-13 | REUNILAB | CNR Virus des Infections Respiratoires - France SUD | Antonin Bal, Gregory Destras, Gwendolyne Burfin, Hadrien Regue, Quentin Semanas, Martine Valette, Bruno Lina, Laurence Josset |
| hCoV-19/Denmark/DCGC-149018/2021 | EPI_ISL_3384347 | 2021-08-08 | Department of Bacteria, Parasites and Fungi, Statens Serum Institut, Copenhagen, Denmark | Statens Serum Institut Bioinformatics and Microbial Genomics | Danish Covid-19 Genome Consortium |
| hCoV-19/USA/AZ-ASU10204/2021 | EPI_ISL_3387525 | 2021-01-30 | Arizona State University | Arizona State University | Ajeet Bains, LaRinda A. Holland, Matthew F. Smith, Peter T. Skidmore, Rabia Maqsood, Nicholas J. Mellor, Nathaniel Johnson, Joshua LaBaer, Vel Murugan, Efrem S. Lim |
| hCoV-19/India/MH-IGIB-GP129/2021 | EPI_ISL_4125352 | 2021-04-10 | Genepath, Pune | IISER Pune | Krishanpal Karmodiya, Aurnab Ghose, Joy Merwin Monteiro, Suvarna Joshi, Rajesh Karyakarte |
| hCoV-19/Austria/KILM1291/2021 | EPI_ISL_3394978 | 2021-08-01 | Dept. of Laboratory Medicine | Dept. of Laboratory Medicine | Harald Esterbauer, Anna Gschaider, Petra Jurkowitsch, Fabian Konig, Sabina Plumer, Robert Strassl, Claudia Weber, Oswald Wagner |
| hCoV-19/FrenchGuiana/IPG2021881/2021 | EPI_ISL_3398106 | 2021-08-09 | CNR Institut Pasteur de la Guyane | Institut Pasteur de la Guyane | Anne Lavergne, Dominique Rousset, Antoine Enfissi, Arielle Salmier |
| hCoV-19/Turkey/HSGM-GF16144/2021 | EPI_ISL_3400095 | 2021-08-04 | Ministry of Health Turkey | Ministry of Health Turkey | Fatma Bayrakdar, Yasemin Cosgun, Suleyman Yalcin, Gulay Korukluoglu |
| hCoV-19/Turkey/HSGM-GF15155/2021 | EPI_ISL_3400607 | 2021-08-04 | Ministry of Health Turkey | Ministry of Health Turkey | Fatma Bayrakdar, Yasemin Cosgun, Suleyman Yalcin, Gulay Korukluoglu |
| hCoV-19/Sweden/SUS0003295/2021 | EPI_ISL_3402091 | 2021-08-03 | Clinical Microbiology, Infection Prevention and Control | Section for Molecular Diagnostics | Bj√∂rn Hallstr√∂m, Jonas Bj√∂rkman |
| hCoV-19/Turkey/HSGM-FS44/2021 | EPI_ISL_3402613 | 2021-08-05 | Ministry of Health Turkey | Ministry of Health Turkey | Fatma Bayrakdar, Yasemin Cosgun, Suleyman Yalcin, Gulay Korukluoglu |
| hCoV-19/Turkey/HSGM-FS87/2021 | EPI_ISL_3402614 | 2021-08-05 | Ministry of Health Turkey | Ministry of Health Turkey | Fatma Bayrakdar, Yasemin Cosgun, Suleyman Yalcin, Gulay Korukluoglu |

| | | | | | |
|---|---|---|---|---|---|
| hCoV-19/India/MH-IGIB-Sample102/2021 | EPI_ISL_4126749 | 2021-01-14 | BJMC, Pune | IISER Pune | Krishanpal Karmodiya, Aurnab Ghose, Joy Merwin Monteiro, Suvarna Joshi, Rajesh Karyakarte |
| hCoV-19/USA/MA-UMASSMED-P014H6/2021 | EPI_ISL_3410250 | 2021-04-07 | UMass Memorial Medical Center | Center for Microbiome Research | Richard T. Ellison III, Karl J. Simon, Doyle V. Ward |
| hCoV-19/India/UP-IGIB-LUCN0040164113/2021 | EPI_ISL_4127271 | 2021-07-06 | INSACOG Surveillance | INSACOG at CSIR Institute of Genomics and Integrative Biology | INSACOG |
| hCoV-19/SouthAfrica/NICD-N8310/2021 | EPI_ISL_3411797 | 2021-04-20 | KIMBERLEY LABORATORY | Diseases of the National Health Laboratory Service | Amoako DG, Everatt J, Scheepers C, Mohale T, Ntuli N, Mahlangu B, Mnguni A, Ismail A, Bhiman JN |
| hCoV-19/USA/PA-VSP5115/2021 | EPI_ISL_3411922 | 2021-06-14 | Hospital of the University of Pennsylvania Molecular Pathology Lab | Bushman | John K. Everett, Kyle Rodino, Andrew D. Marques, Shantan Reddy, Aoife M. Roche, Young Hwang, Scott Sherrill-Mix, Samantha A. Whiteside, Jevon Graham-Wooten, Layla A. Khatib, Ayannah S. Fitzgerald, Arupa Ganguly, Mike Feldman, Brendan Kelly, Ronald G. Coll |
| hCoV-19/Turkey/HSGM-FS3730/2021 | EPI_ISL_3413139 | 2021-08-07 | Ministry of Health Turkey | Ministry of Health Turkey | Fatma Bayrakdar, Yasemin Cosgun, Suleyman Yalcin, Gulay Korukluoglu |
| hCoV-19/England/PHEC-U30AU926/2021 | EPI_ISL_3777194 | 2021-06-28 | Respiratory Virus Unit, Microbiology Services Colindale, Public Health England | COVID-19 Genomics UK (COG-UK) Consortium | PHE Covid Sequencing Team |
| hCoV-19/Turkey/HSGM-FS7375/2021 | EPI_ISL_3417217 | 2021-08-08 | Ministry of Health Turkey | Ministry of Health Turkey | Fatma Bayrakdar, Yasemin Cosgun, Suleyman Yalcin, Gulay Korukluoglu |
| hCoV-19/Australia/NSW-RPAH-0970/2021 | EPI_ISL_3417447 | 2021-08-05 | New South Wales Health Pathology Royal Prince Alfred Hospital | Microbiology RPAH | Foster, C.; Au, J.; Ruiz Silva, M.; Deveson, I.; Bull, R.; Van Hal, S.; Rawlinson, W. |
| hCoV-19/USA/NC-CORVASEQ-CLT-000236/2021 | EPI_ISL_3419020 | 2021-07-10 | StarMed Healthcare | UNC Charlotte Environmental Monitoring Laboratory | Jessica Schlueter, Jannatul Ferdous, Erin Stiers, Adam France, Sam Kunkleman, Kevin Lambirth, Cynthia Gibas |
| hCoV-19/Italy/LAZ-IFO-07293269/2021 | EPI_ISL_3419883 | 2021-07-29 | IRCCS San Gallicano Dermatological Institute | IRCCS Regina Elena National Cancer Institute | Ludovica Ciuffreda, Sara Donzelli, Matteo Pallocca, Valentina Ricca, Fabrizio Ensoli, Fulvia Pimpinelli, Maurizio Fanciulli, Giovanni Blandino, Aldo Morrone, Gennaro Ciliberto |
| hCoV-19/Belize/CML-109/2021 | EPI_ISL_4296413 | 2021-06-30 | Central Medical Laboratory | Baylor College of Medicine | Alexander Kneubehl, Sarah Gunter, Shannon Ronca, Allison Lino, Adrianna Maliga, Sarah Strobel, Russell Manzanero, Gerhaldine Morazan, Kristy Murray |
| hCoV-19/Belize/CML-99/2021 | EPI_ISL_4296403 | 2021-03-07 | Central Medical Laboratory | Baylor College of Medicine | Alexander Kneubehl, Sarah Gunter, Shannon Ronca, Allison Lino, Adrianna Maliga, Sarah Strobel, Russell Manzanero, Gerhaldine Morazan, Kristy Murray |
| hCoV-19/Belize/CML-102/2021 | EPI_ISL_4296406 | 2021-01-07 | Central Medical Laboratory | Baylor College of Medicine | Alexander Kneubehl, Sarah Gunter, Shannon Ronca, Allison Lino, Adrianna Maliga, Sarah Strobel, Russell Manzanero, Gerhaldine Morazan, Kristy Murray |
| hCoV-19/India/MH-IGIB-Sample755/2020 | EPI_ISL_4129896 | 2020-12-19 | BJMC, Pune | IISER Pune | Krishanpal Karmodiya, Aurnab Ghose, Joy Merwin Monteiro, Suvarna Joshi, Rajesh Karyakarte |
| hCoV-19/England/PHEP-262264/2021 | EPI_ISL_4129943 | 2021-01-11 | Respiratory Virus Unit, Microbiology Services Colindale, Public Health England | COVID-19 Genomics UK (COG-UK) Consortium | PHE Covid Sequencing Team |
| hCoV-19/England/PHEP-263338/2021 | EPI_ISL_4130289 | 2021-08-10 | Respiratory Virus Unit, Microbiology Services Colindale, Public Health England | COVID-19 Genomics UK (COG-UK) Consortium | PHE Covid Sequencing Team |

| | | | | | |
|---|---|---|---|---|---|
| hCoV-19/USA/GA-CDC-MMB09364687/2021 | EPI_ISL_3427552 | 2021-07-13 | Mako Medical | Centers for Disease Control and Prevention Division of Viral Diseases, Pathogen Discovery | Dakota Howard, Dhwani Batra, Peter W. Cook, Kara Moser, Adrian Paskey, Jason Caravas, Benjamin Rambo-Martin, Shatavia Morrison, Christopher Gulvick, Scott Sammons, Yvette Unoarumhi, Darlene Wagner, Matthew Schmerer, Matthew Tugwell, Lauren Moon, Clinton R |
| hCoV-19/USA/AK-USAFSAM-S5293/2021 | EPI_ISL_4001654 | 2021-07-29 | US Air Force School of Aerospace Medicine | US Air Force School of Aerospace Medicine | Anthony Fries, Deanna Muehleman, Jennifer Meyer, William Gruner, Kelsey Lanter, Carol Garrett, Sarah Purves, Clarise Starr, Elizabeth Macias |
| hCoV-19/Australia/VIC18480/2021 | EPI_ISL_2920933 | 2021-04-27 | Victorian Infectious Diseases Reference Laboratory (VIDRL) | VIDRL and MDU-PHL | Caly L., Seemann T., Sait, M.L., Druce J., Sherry, N.L. |
| hCoV-19/Botswana/R31B27_BHP_000744852/2021 | EPI_ISL_4299826 | 2021-09-03 | Palapye Primary Hospital Laboratory | Botswana Harvard HIV Reference Laboratory | Sikhulile Moyo, Wonderful T. Choga, Dorcas Maruapula, Keoratile Ntshambiwa, Sefetogi Ramaologa, Thongbotho Mphoyakgosi, Boitumelo Zuze, Botshelo Radibe, Legodile Kooepile, Ontlametse T. Bareng, Patrick T. Mokgethi, Pamela Smith-Lawrence, Kgomotso Moruisi, |
| hCoV-19/USA/CA-COLOR-UCSF-CC622/2021 | EPI_ISL_4300524 | 2021-04-30 | Color Genomics | Chiu Laboratory, University of California, San Francisco | Charles Chiu, Venice Servellita, Alicia Sotomayor-Gonzalez, Baolin Wang, Scott Topper, Erika Torres, Noah Brazer, Daniel Wong, Yueyuan Zhang, Madyson Wanner, Alicia Zhou |
| hCoV-19/SouthKorea/KDCA7665/2021 | EPI_ISL_3452039 | 2021-07-23 | Division of Emerging Infectious Diseases, Bureau of Infectious Diseases Diagnosis Control, Korea Disease Control and Prevention Agency | Diseases, Bureau of Infectious Diseases Diagnosis Control, Korea Disease Control and Prevention Agency | Ae Kyung Park, Il-Hwan Kim, Heui Man Kim, Jeong-Ah Kim, Chae Young Lee, Eun-Jin Kim |
| hCoV-19/USA/VT-CDCBI-CRSP_GE5TMP2FCQ5MKTTC/2021 | EPI_ISL_3431657 | 2021-08-07 | Broad Institute Clinical Research Sequencing Platform | Infectious Disease Program, Broad Institute of Harvard and MIT | Siddle,K.J., Adams,G., Pearlman,L., Gladden-Young,A., Vicente,G., Blumenstiel,B., DeFelice,M., Lee,M., McGovern,S., Lagerborg,K., Rudy,M., DeRuff,K., Carter,A., Normandin,E., Bauer,M., Reilly,S., Tomkins-Tinch,C., Loreth,C., Chaluvadi,S., Meldrim,J., Gran |
| hCoV-19/Portugal/PT13803/2021 | EPI_ISL_3432087 | 2021-08-02 | H Divino Espirito Santo - Ponta Delgada | Instituto Nacional de Saude (INSA) | Borges et al |
| hCoV-19/France/PAC-IHU-19923_Nova1/2021 | EPI_ISL_3432504 | 2021-07-20 | MEPHI, Aix Marseille University | MEPHI, Aix Marseille University | Anthony LEVASSEUR |
| hCoV-19/Paraguay/209776/2021 | EPI_ISL_4303344 | 2021-07-30 | Laboratorio Central de Salud P√∫blica | Laboratorio Central de Salud P√∫blica | Sandra Gonzalez, Andrea G√≥mez de la Fuente, Cynthia Vazquez, Julio Barrios, Cesar Cantero, Tania Alfonzo, Shirley Villalba, Juan Torales, Maria Liz Gamarra |
| hCoV-19/France/PAC-IHU-20036_Nova1/2021 | EPI_ISL_3432994 | 2021-07-23 | MEPHI, Aix Marseille University | MEPHI, Aix Marseille University | Anthony LEVASSEUR |
| hCoV-19/Italy/FVG-UD-409757/2021 | EPI_ISL_4533767 | 2021-09-17 | Dipartimento di Medicina di Laboratorio, Azienda sanitaria universitaria Friuli Centrale (ASU FC) | Dipartimento di Medicina di Laboratorio, Azienda sanitaria universitaria Friuli Centrale (ASU FC) | Corrado Pipan, Catia Mio, Chiara Dal Secco, Federica Baldan, Stefania Marzinotto, Francesco Curcio |
| hCoV-19/Australia/QLD2069/2021 | EPI_ISL_3161825 | 2021-07-27 | Queensland Health Forensic and Scientific Services | Queensland Health Forensic and Scientific Services | Chenwei Wang on behalf of Q-PHIRE Genomics |
| hCoV-19/Turkey/HSGM-C8408/2021 | EPI_ISL_3533335 | 2021-07-28 | Ministry of Health Turkey | Ministry of Health Turkey | Fatma Bayrakdar, Yasemin Cosgun, Suleyman Yalcin, Gulay Korukluoglu |

| | | | | | |
|---|---|---|---|---|---|
| hCoV-19/England/MILK-1B5FAF7/2021 | EPI_ISL_3437821 | 2021-08-09 | Lighthouse Lab in Milton Keynes | Wellcome Sanger Institute for the COVID-19 Genomics UK (COG-UK) Consortium | The Lighthouse Lab in Milton Keynes and Alex Alderton, Roberto Amato, Jeffrey Barrett, Sonia Goncalves, Ewan Harrison, David K. Jackson, Ian Johnston, Dominic Kwiatkowski, Cordelia Langford, John Sillitoe on behalf of the Wellcome Sanger Institute COVID-1 |
| hCoV-19/USA/SC-CDC-LC0196068/2021 | EPI_ISL_4150156 | 2021-08-16 | Laboratory Corporation of America | Centers for Disease Control and Prevention Division of Viral Diseases, Pathogen Discovery | Dakota Howard,Dhwani Batra,Peter Cook,Kara Moser,Adrian Paskey,Jason Caravas,Benjamin Rambo-Martin,Shatavia Morrison,Christopher Gulvick,Scott Sammons,Yvette Unoarumhi,Darlene Wagner,Matthew Schmerer,Minoo Agarwal,Eyad Almasri,Debbie Boles,Ayla Burns,Nuth |
| hCoV-19/England/PHEC-YYBFOEI/2021 | EPI_ISL_4310436 | 2021-08-08 | Respiratory Virus Unit, Microbiology Services Colindale, Public Health England | COVID-19 Genomics UK (COG-UK) Consortium | PHE Covid Sequencing Team |
| hCoV-19/Peru/CAL-INS-4244/2021 | EPI_ISL_4080301 | 2021-08-04 | Laboratorio de Referencia Nacional de Virus Respiratorios. Centro Nacional de Salud Publica. Instituto Nacional de Salud Peru. | Laboratorio de Referencia Nacional de Virus Respiratorios. Centro Nacional de Salud Publica. Instituto Nacional de Salud Peru. | Carlos Padilla Rojas, Veronica Hurtado Vela, Orson Mestanza Millones, Iris Silva Molina, Luis Barcena Flores, Sandra Morales Ruiz, Steve Acedo Lazo, Wendy Lizarraga Olivares, Victor Jimenez Vasquez, Sara Gordillo Vilchez, Nieves Sevilla Casta√±eda, Prince |
| hCoV-19/India/WB-INSACOG-1930301157996/2021 | EPI_ISL_4312178 | 2021-06-30 | INSACOG-WB | National Institute of Biomedical Genomics ,Äì¬†INSACOG | Arindam Maitra, Bhaswati Bandyopadhyay, Nidhan Kumar Biswas, Tamal Ghosh, Sreedhar Chinnaswamy, Ajay Chakraborti, Saumitra Das |
| hCoV-19/India/SK-INSACOG-1122500081830/2021 | EPI_ISL_4313077 | 2021-07-20 | INSACOG-Sikkim | National Institute of Biomedical Genomics ,Äì¬†INSACOG | Arindam Maitra, Shrijana Gurung, Nidhan Kumar Biswas, Kaden Zangmu Bhutia, Sreedhar Chinnaswamy Tshering Ongmu Bhutia, Saumitra Das |
| hCoV-19/Italy/LAZ-INMI-1775/2021 | EPI_ISL_3446984 | 2021-08-05 | RM1/Osp. S.Filippo Neri | National Institute for Infectious Diseases (INMI) L. Spallanzani I.R.C.C.S | E Giombini, B Bartolini, M Rueca, G Bonfiglio, O Butera, CEM Gruber, F Santini, F Messina, A Di Caro, MR Capobianchi |
| hCoV-19/USA/IL-S21WGS3374/2021 | EPI_ISL_3447299 | 2021-08-07 | Illinois Department of Public Health - Springfield Lab | Illinois Department of Public Health - Springfield Lab | Bryan Sim, Marshall Smock |
| hCoV-19/SouthAfrica/KRISP-K020660/2021 | EPI_ISL_3447732 | 2021-07-08 | NHLS_IALCH | KRISP, KZN Research Innovation and Sequencing Platform | Giandhari Jennifer, Pillay Sureshnee, Yajna Ramphal, Naidoo Yeshnee, Tshiabuila Derek,Tegally Houriiyah, San James, Wilkinson Eduan, de Oliveira Tulio |
| hCoV-19/SouthAfrica/NICD-N12669/2021 | EPI_ISL_3451237 | 2021-07-15 | POLOKWANE LABORATORY | Diseases of the National Health Laboratory Service | Amoako DG, Everatt J, Scheepers C, Mohale T, Ntuli N, Mahlangu B, Mnguni A, Ismail A, Bhiman JN |
| hCoV-19/SouthAfrica/NICD-N12699/2021 | EPI_ISL_3451259 | 2021-07-06 | KIMBERLEY LABORATORY | Diseases of the National Health Laboratory Service | Amoako DG, Everatt J, Scheepers C, Mohale T, Ntuli N, Mahlangu B, Mnguni A, Ismail A, Bhiman JN |
| hCoV-19/SouthAfrica/NICD-N12708/2021 | EPI_ISL_3451268 | 2021-07-07 | KIMBERLEY LABORATORY | Diseases of the National Health Laboratory Service | Amoako DG, Everatt J, Scheepers C, Mohale T, Ntuli N, Mahlangu B, Mnguni A, Ismail A, Bhiman JN |
| hCoV-19/India/RJ-CCMB-CIA6166/2020 | EPI_ISL_3453376 | 2020-10-30 | RUHS College of Medical Sciences | CSIR-Centre for Cellular and Molecular Biology - INSACOG | Dr. Gaurav Dalela,Dr. Vaseem Naheed Baig,Dr. Sonali Sharma,Dr. Ramesh Sharma,Dr. Nilofer Khayyam,Dr. Jitendra Panda,Dr. Sudhanshu Kacker,Dr. Raja Babu Panwar,Dr. Rajeev Gupta, Lamuk Zaveri,Tulasi Nagabandi,Payel Mukherjee,Onkar Kulkarni,Jandhyala Sai Kris |

| | | | | | |
|---|---|---|---|---|---|
| hCoV-19/India/AP-CCMB-CIA6573/2021 | EPI_ISL_3453437 | 2021-07-08 | AP SSO | CSIR-Centre for Cellular and Molecular Biology-INSACOG | Payel Mukherjee, Lamuk Zaveri, Onkar Kulkarni,Jandhyala Sai Krishna,Tulasi Nagabandi, Sofia Banu, Ara Sreenivas,Shreekant Verma, Amareshwar Vodapalli ,B Himasri,Valli Nagalakshmi Undamatla, Sumedha Avadhanula, Vidhyadhari Methuku, Priya Nurkurthy, Archana |
| hCoV-19/India/RJ-CCMB-CIA6152/2020 | EPI_ISL_3453544 | 2020-10-23 | RUHS College of Medical Sciences | CSIR-Centre for Cellular and Molecular Biology - INSACOG | Dr. Gaurav Dalela,Dr. Vaseem Naheed Baig,Dr. Sonali Sharma,Dr. Ramesh Sharma,Dr. Nilofer Khayyam,Dr. Jitendra Panda,Dr. Sudhanshu Kacker,Dr. Raja Babu Panwar,Dr. Rajeev Gupta, Payel Mukherjee, Lamuk Zaveri, Jandhyala Sai Krishna,Onkar Kulkarni,Tulasi Naga |
| hCoV-19/India/RJ-CCMB-CIA6237/2021 | EPI_ISL_3453549 | 2021-01-18 | RUHS College of Medical Sciences | CSIR-Centre for Cellular and Molecular Biology - INSACOG | Dr. Gaurav Dalela,Dr. Vaseem Naheed Baig,Dr. Sonali Sharma,Dr. Ramesh Sharma,Dr. Nilofer Khayyam,Dr. Jitendra Panda,Dr. Sudhanshu Kacker,Dr. Raja Babu Panwar,Dr. Rajeev Gupta, Amareshwar Vodapalli,Ara Sreenivas,Shreekant Verma, B Himasri,Valli Nagalakshmi |
| hCoV-19/India/RJ-CCMB-CIA6229/2021 | EPI_ISL_3453559 | 2021-01-15 | RUHS College of Medical Sciences | CSIR-Centre for Cellular and Molecular Biology - INSACOG | Dr. Gaurav Dalela,Dr. Vaseem Naheed Baig,Dr. Sonali Sharma,Dr. Ramesh Sharma,Dr. Nilofer Khayyam,Dr. Jitendra Panda,Dr. Sudhanshu Kacker,Dr. Raja Babu Panwar,Dr. Rajeev Gupta, Shreekant Verma, Amareshwar Vodapalli ,Ara Sreenivas,B Himasri,Valli Nagalakshm |
| hCoV-19/India/RJ-CCMB-CIA6182/2020 | EPI_ISL_3453562 | 2020-11-05 | RUHS College of Medical Sciences | CSIR-Centre for Cellular and Molecular Biology - INSACOG | Dr. Gaurav Dalela,Dr. Vaseem Naheed Baig,Dr. Sonali Sharma,Dr. Ramesh Sharma,Dr. Nilofer Khayyam,Dr. Jitendra Panda,Dr. Sudhanshu Kacker,Dr. Raja Babu Panwar,Dr. Rajeev Gupta, Lamuk Zaveri,Tulasi Nagabandi,Payel Mukherjee,Onkar Kulkarni,Jandhyala Sai Kris |
| hCoV-19/India/RJ-CCMB-CIA6217/2020 | EPI_ISL_3453568 | 2020-12-31 | RUHS College of Medical Sciences | CSIR-Centre for Cellular and Molecular Biology - INSACOG | Dr. Gaurav Dalela,Dr. Vaseem Naheed Baig,Dr. Sonali Sharma,Dr. Ramesh Sharma,Dr. Nilofer Khayyam,Dr. Jitendra Panda,Dr. Sudhanshu Kacker,Dr. Raja Babu Panwar,Dr. Rajeev Gupta, Ara Sreenivas,Shreekant Verma, Amareshwar Vodapalli ,B Himasri,Jandhyala Sai Kr |
| hCoV-19/India/RJ-CCMB-CIA6150/2020 | EPI_ISL_3453581 | 2020-10-23 | RUHS College of Medical Sciences | CSIR-Centre for Cellular and Molecular Biology - INSACOG | Dr. Gaurav Dalela,Dr. Vaseem Naheed Baig,Dr. Sonali Sharma,Dr. Ramesh Sharma,Dr. Nilofer Khayyam,Dr. Jitendra Panda,Dr. Sudhanshu Kacker,Dr. Raja Babu Panwar,Dr. Rajeev Gupta, Payel Mukherjee, Lamuk Zaveri, Jandhyala Sai Krishna,Onkar Kulkarni,Tulasi Naga |
| hCoV-19/India/RJ-CCMB-CIA6235/2021 | EPI_ISL_3453588 | 2021-01-18 | RUHS College of Medical Sciences | CSIR-Centre for Cellular and Molecular Biology - INSACOG | Dr. Gaurav Dalela,Dr. Vaseem Naheed Baig,Dr. Sonali Sharma,Dr. Ramesh Sharma,Dr. Nilofer Khayyam,Dr. Jitendra Panda,Dr. Sudhanshu Kacker,Dr. Raja Babu Panwar,Dr. Rajeev Gupta, Amareshwar Vodapalli,Ara Sreenivas,Shreekant Verma, B Himasri,Valli Nagalakshmi |

| | | | | | |
|---|---|---|---|---|---|
| hCoV-19/India/RJ-CCMB-CIA6193/2020 | EPI_ISL_3453590 | 2020-11-06 | RUHS College of Medical Sciences | CSIR-Centre for Cellular and Molecular Biology - INSACOG | Dr. Gaurav Dalela,Dr. Vaseem Naheed Baig,Dr. Sonali Sharma,Dr. Ramesh Sharma,Dr. Nilofer Khayyam,Dr. Jitendra Panda,Dr. Sudhanshu Kacker,Dr. Raja Babu Panwar,Dr. Rajeev Gupta, Tulasi Nagabandi,Lamuk Zaveri,Jandhyala Sai Krishna,Payel Mukherjee,Onkar Kulka |
| hCoV-19/India/RJ-CCMB-CIA6164/2020 | EPI_ISL_3453628 | 2020-10-29 | RUHS College of Medical Sciences | CSIR-Centre for Cellular and Molecular Biology - INSACOG | Dr. Gaurav Dalela,Dr. Vaseem Naheed Baig,Dr. Sonali Sharma,Dr. Ramesh Sharma,Dr. Nilofer Khayyam,Dr. Jitendra Panda,Dr. Sudhanshu Kacker,Dr. Raja Babu Panwar,Dr. Rajeev Gupta, Lamuk Zaveri,Tulasi Nagabandi,Payel Mukherjee,Onkar Kulkarni,Jandhyala Sai Kris |
| hCoV-19/India/AP-CCMB-CIA6479/2021 | EPI_ISL_3453662 | 2021-07-05 | AP SSO | CSIR-Centre for Cellular and Molecular Biology-INSACOG | Payel Mukherjee, Lamuk Zaveri, Onkar Kulkarni,Tulasi Nagabandi, Sofia Banu, Ara Sreenivas,Shreekant Verma, Amareshwar Vodapalli ,B Himasri,Valli Nagalakshmi Undamatla, Sumedha Avadhanula, Vidhyadhari Methuku, Priya Nurkurthy, Archana Bharadwaj Siva, Karth |
| hCoV-19/USA/FL-CDC-QDX27272206/2021 | EPI_ISL_3456165 | 2021-07-25 | Quest Diagnostics Incorporated | Centers for Disease Control and Prevention Division of Viral Diseases, Pathogen Discovery | Dakota Howard, Dhwani Batra, Peter W. Cook, Kara Moser, Adrian Paskey, Jason Caravas, Benjamin Rambo-Martin, Shatavia Morrison, Christopher Gulvick, Scott Sammons, Yvette Unoarumhi, Darlene Wagner, Matthew Schmerer, S. H. Rosenthal, A. Gerasimova, R. M. K |
| hCoV-19/USA/NC-CDC-LC0124997/2021 | EPI_ISL_4157597 | 2021-07-23 | Laboratory Corporation of America | Centers for Disease Control and Prevention Division of Viral Diseases, Pathogen Discovery | Dakota Howard,Dhwani Batra,Peter Cook,Kara Moser,Adrian Paskey,Jason Caravas,Benjamin Rambo-Martin,Shatavia Morrison,Christopher Gulvick,Scott Sammons,Yvette Unoarumhi,Darlene Wagner,Matthew Schmerer,Minoo Agarwal,Eyad Almasri,Debbie Boles,Ayla Burns,Nuth |
| hCoV-19/USA/CA-CDC-LC0127073/2021 | EPI_ISL_4159262 | 2021-07-22 | Laboratory Corporation of America | Centers for Disease Control and Prevention Division of Viral Diseases, Pathogen Discovery | Dakota Howard,Dhwani Batra,Peter Cook,Kara Moser,Adrian Paskey,Jason Caravas,Benjamin Rambo-Martin,Shatavia Morrison,Christopher Gulvick,Scott Sammons,Yvette Unoarumhi,Darlene Wagner,Matthew Schmerer,Minoo Agarwal,Eyad Almasri,Debbie Boles,Ayla Burns,Nuth |
| hCoV-19/Canada/QC-L00323995001/2021 | EPI_ISL_3458095 | 2021-01-13 | Laboratoire de sant√© publique du Qu√©bec | Laboratoire de sant√© publique du Qu√©bec | Sandrine Moreira, Ioannis Ragoussis, Guillaume Bourque, Jesse Shapiro, Mark Lathrop and Michel Roger on behalf of the CoVSeQ research group |
| hCoV-19/Canada/QC-L00359815001/2021 | EPI_ISL_3458961 | 2021-06-11 | Laboratoire de sant√© publique du Qu√©bec | Laboratoire de sant√© publique du Qu√©bec | Sandrine Moreira, Ioannis Ragoussis, Guillaume Bourque, Jesse Shapiro, Mark Lathrop and Michel Roger on behalf of the CoVSeQ research group |
| hCoV-19/Canada/QC-L00364012001/2021 | EPI_ISL_3459079 | 2021-06-30 | Laboratoire de sant√© publique du Qu√©bec | Laboratoire de sant√© publique du Qu√©bec | Sandrine Moreira, Ioannis Ragoussis, Guillaume Bourque, Jesse Shapiro, Mark Lathrop and Michel Roger on behalf of the CoVSeQ research group |

| | | | | | |
|---|---|---|---|---|---|
| hCoV-19/USA/CA-CDC-FG-097723/2021 | EPI_ISL_4160977 | 2021-08-22 | Fulgent Genetics | Centers for Disease Control and Prevention Division of Viral Diseases, Pathogen Discovery | Dakota Howard,Dhwani Batra,Peter Cook,Kara Moser,Adrian Paskey,Jason Caravas,Benjamin Rambo-Martin,Shatavia Morrison,Christopher Gulvick,Scott Sammons,Yvette Unoarumhi,Darlene Wagner,Matthew Schmerer,Harry Gao,Mickey Li,John Gao,Joseph Fierro,Benafsh Sapr |
| hCoV-19/Suriname/SR-435/2021 | EPI_ISL_3462611 | 2021-06-30 | Central Laboratory, Bureau of Public Health (BOG) and Academic Hospital Paramaribo | Erasmus Medical Center | Bas B Oude Munnink, Cherise Beek, Consuella Partowidjojo, Dion Gajadin, Ed PF IJzerman, Emmanuelle Munger, Gary Gummels, Ingrid SK Krishnadath, Lycke Woittiez, Marion PG Koopmans, Mireille Van de Veer, Phyllis Pinas, Princes Wongsowidjojo, Radjesh Ori, Ra |
| hCoV-19/Switzerland/VD-ETHZ-33641277/2021 | EPI_ISL_3465751 | 2021-08-02 | Viollier AG | Department of Biosystems Science and Engineering, ETH Z√ºrich | Chaoran Chen, Sarah Nadeau, Louis du Plessis, Catharine Aquino, Ivan Topolsky, Kim Philipp Jablonski, Lara Fuhrmann, Daniel Ehrsam, Isabel St√ºrmer, Andreia Cabral de Gouvea, Maria Domenica Moccia, Simon Gr√ºter, Timothy Sykes, Lennart Opitz, Griffin Whit |
| hCoV-19/Germany/BY-RKI-I-185943/2021 | EPI_ISL_2956127 | 2021-06-29 | Synlab MVZ Augsburg | Robert Koch Institute | unknown |
| hCoV-19/Germany/NW-RKI-I-186075/2021 | EPI_ISL_2956254 | 2021-07-06 | SYNLAB MVZ Leverkusen | Robert Koch Institute | unknown |
| hCoV-19/Germany/BW-RKI-I-186870/2021 | EPI_ISL_2956835 | 2021-06-20 | Eurofins LifeCodexx GmbH | Robert Koch Institute | unknown |
| hCoV-19/Germany/BY-RKI-I-187119/2021 | EPI_ISL_2957145 | 2021-06-22 | Bayerisches Landesamt f√ºr Gesundheit und Lebensmittelsicherheit (LGL) | Robert Koch Institute | unknown |
| hCoV-19/Germany/BY-RKI-I-187152/2021 | EPI_ISL_2957181 | 2021-06-21 | Bayerisches Landesamt f√ºr Gesundheit und Lebensmittelsicherheit (LGL) | Robert Koch Institute | unknown |
| hCoV-19/Germany/NW-RKI-I-188795/2021 | EPI_ISL_3044976 | 2021-07-15 | Labor M√∂nchengladbach MVZ Dr. Stein + Kollegen GbR | Robert Koch Institute | unknown |
| hCoV-19/Ireland/D-NVRL-S21IRL00126935/2021 | EPI_ISL_3467561 | 2021-07-23 | National Virus Reference Laboratory | National Virus Reference Laboratory | Zoe Yandle, Charlene Bennett, Gabriel Gonzalez, Michael Carr, Jonathan Dean, Cillian F De Gascun |
| hCoV-19/Germany/BW-RKI-I-191194/2021 | EPI_ISL_3122208 | 2021-07-05 | Eurofins LifeCodexx GmbH | Robert Koch Institute | unknown |
| hCoV-19/Germany/BE-RKI-I-191619/2021 | EPI_ISL_3122600 | 2021-07-22 | MDI Limbach Berlin GmbH; MVZ Labor Berlin | Robert Koch Institute | unknown |
| hCoV-19/Germany/BY-RKI-I-192418/2021 | EPI_ISL_3237930 | 2021-07-16 | Uniklinikum Regensburg; Institut f√ºr klinische Mikrobiologie und Hygiene | Robert Koch Institute | unknown |
| hCoV-19/Germany/NW-RKI-I-195425/2021 | EPI_ISL_3494952 | 2021-07-28 | amedes MVZ DIAMEDIS Sennestadt | Robert Koch Institute | unknown |
| hCoV-19/Germany/BW-RKI-I-198130/2021 | EPI_ISL_3497157 | 2021-08-05 | Limbach - MVZ Labor Ludwigsburg | Robert Koch Institute | unknown |
| hCoV-19/France/ARA-HCL021137857101/2021 | EPI_ISL_3472315 | 2021-07-31 | CH GIVORS | CNR Virus des Infections Respiratoires - France SUD | Antonin Bal, Gregory Destras, Gwendolyne Burfin, Hadrien Regue, Quentin Semanas, Martine Valette, Bruno Lina, Laurence Josset |
| hCoV-19/Germany/NW-RKI-I-209474/2021 | EPI_ISL_3879776 | 2021-08-21 | SYNLAB MVZ Leverkusen | Robert Koch Institute | unknown |

| | | | | | |
|---|---|---|---|---|---|
| hCoV-19/Germany/NW-RKI-I-211042/2021 | EPI_ISL_3885383 | 2021-08-20 | amedes MVZ DIAMEDIS Sennestadt | Robert Koch Institute | unknown |
| hCoV-19/Germany/NW-RKI-I-212995/2021 | EPI_ISL_3888131 | 2021-08-24 | Labor M√∂nchengladbach MVZ Dr. Stein + Kollegen GbR | Robert Koch Institute | unknown |
| hCoV-19/Brazil/SP-IB_129626/2021 | EPI_ISL_4168209 | 2021-08-23 | INSIDE DIAGNOSTICOS | Instituto Butantan | Dimas Tadeu Covas, Antonio Jorge Martins, Claudia Renata dos Santos Barros, David Schlesinger, Debora Botequio Moretti, Elaine Cristina Marqueze, Elaine Vieira Santos, Evandra Strazza Rodrigues, Heidge Fukumasu, Jayme Augusto de Souza-Neto, Jos√© Salvator |
| hCoV-19/Switzerland/SO-ETHZ-33638969/2021 | EPI_ISL_3476263 | 2021-08-02 | Viollier AG | Department of Biosystems Science and Engineering, ETH Z√ºrich | Chaoran Chen, Sarah Nadeau, Louis du Plessis, Catharine Aquino, Ivan Topolsky, Kim Philipp Jablonski, Lara Fuhrmann, Daniel Ehrsam, Isabel St√ºrmer, Andreia Cabral de Gouvea, Maria Domenica Moccia, Simon Gr√ºter, Timothy Sykes, Lennart Opitz, Griffin Whit |
| hCoV-19/Germany/NW-RKI-I-224152/2021 | EPI_ISL_4043521 | 2021-08-31 | SYNLAB MVZ Leverkusen | Robert Koch Institute | unknown |
| hCoV-19/Germany/BY-RKI-I-224325/2021 | EPI_ISL_4043689 | 2021-08-30 | SYNLAB MVZ Weiden | Robert Koch Institute | unknown |
| hCoV-19/France/IDF-HMN-21082020803/2021 | EPI_ISL_3476439 | 2021-07-31 | GH A.CHENEVIER-H.MONDOR | Mondor University Hospital, Assistance Publique H√¥pitaux de Paris, Universit√© Paris-Est Cr√©teil, INSERM U955 | Christophe Rodriguez, Slim Fourati, Vanessa Demontant, Guillaume Gricourt, Melissa N'Debi, Alexandre Soulier, Elisabeth Trawinski, Jean-Michel Pawlotsky |
| hCoV-19/Germany/NW-RKI-I-226889/2021 | EPI_ISL_4222262 | 2021-09-03 | SYNLAB MVZ Leverkusen | Robert Koch Institute | unknown |
| hCoV-19/Germany/NW-RKI-I-227165/2021 | EPI_ISL_4222604 | 2021-09-03 | SYNLAB MVZ Leverkusen | Robert Koch Institute | unknown |
| hCoV-19/NewZealand/21MV0638/2021 | EPI_ISL_3477082 | 2021-07-29 | Middlemore Hospital | Institute of Environmental Science and Research (ESR) | Rachel Boyle, SallyAnn Harbison, Olivia Stroeven, Xiaoyun Ren, Matt Storey, Nikki Freed, Muhammad Faisal, Jing Wang, Hermes Perez, Anja Werno, Antje van der Linden, Arlo Upton, Chris Mansell, David Hammer, Dragana Drinkovic, Gary McAuliffe, Hana Sofia And |
| hCoV-19/France/BFC-HMN-21082040513/2021 | EPI_ISL_3477428 | 2021-07-27 | BIOLLAN Brognard | Mondor University Hospital, Assistance Publique H√¥pitaux de Paris, Universit√© Paris-Est Cr√©teil, INSERM U955 | Christophe Rodriguez, Slim Fourati, Vanessa Demontant, Guillaume Gricourt, Melissa N'Debi, Alexandre Soulier, Elisabeth Trawinski, Jean-Michel Pawlotsky |
| hCoV-19/Germany/HE-RKI-I-232581/2021 | EPI_ISL_4226918 | 2021-08-28 | CENTOGENE Frankfurt Laboratory: Niederlassung Industriepark H√∂chst | Robert Koch Institute | unknown |
| hCoV-19/Germany/BE-RKI-I-232843/2021 | EPI_ISL_4227208 | 2021-09-07 | MDI Limbach Berlin GmbH; MVZ Labor Berlin | Robert Koch Institute | unknown |
| hCoV-19/France/IDF-HMN-21072260892/2021 | EPI_ISL_3478481 | 2021-07-20 | Biogroup Bio Lam-LCD Saint-Denis | Mondor University Hospital, Assistance Publique H√¥pitaux de Paris, Universit√© Paris-Est Cr√©teil, INSERM U955 | Christophe Rodriguez, Slim Fourati, Vanessa Demontant, Guillaume Gricourt, Melissa N'Debi, Alexandre Soulier, Elisabeth Trawinski, Jean-Michel Pawlotsky |
| hCoV-19/Germany/BY-RKI-I-235935/2021 | EPI_ISL_4229457 | 2021-08-26 | Bayerisches Landesamt f√ºr Gesundheit und Lebensmittelsicherheit (LGL) | Robert Koch Institute | unknown |

| | | | | | |
|---|---|---|---|---|---|
| hCoV-19/Germany/HE-RKI-I-243222/2021 | EPI_ISL_4440393 | 2021-09-08 | CENTOGENE Frankfurt Laboratory: Niederlassung Industriepark H√∂chst | Robert Koch Institute | unknown |
| hCoV-19/Germany/BE-RKI-I-247603/2021 | EPI_ISL_4444175 | 2021-09-15 | MDI Limbach Berlin GmbH; MVZ Labor Berlin | Robert Koch Institute | unknown |
| hCoV-19/Germany/BY-RKI-I-247815/2021 | EPI_ISL_4444387 | 2021-09-06 | Bayerisches Landesamt f√ºr Gesundheit und Lebensmittelsicherheit (LGL) | Robert Koch Institute | unknown |
| hCoV-19/SouthAfrica/Tygerberg_1573/2021 | EPI_ISL_3482596 | 2021-06-22 | Division of Medical Virology, National Health Laboratory Service (NHLS), Tygerberg Hospital / Stellenbosch University | Health Laboratory Service (NHLS), Tygerberg Hospital / Stellenbosch University | Susan Engelbrecht, Tongai Maponga, Bronwyn Kleinhans, Shannon Wilson, Tania Stander, Kamela Mahlakwane, Gert van Zyl, Wolfgang Preiser |
| hCoV-19/Germany/SN-RKI-I-250339/2021 | EPI_ISL_4446890 | 2021-09-07 | Diagnosticum - Labor Neukirchen | Robert Koch Institute | unknown |
| hCoV-19/Brazil/SP-IB_128589/2021 | EPI_ISL_4168770 | 2021-08-24 | POSTO DE SAUDE DO CENTRO ATIBAIA | Instituto Butantan | Dimas Tadeu Covas, Antonio Jorge Martins, Claudia Renata dos Santos Barros, David Schlesinger, Debora Botequio Moretti, Elaine Cristina Marqueze, Elaine Vieira Santos, Evandra Strazza Rodrigues, Heidge Fukumasu, Jayme Augusto de Souza-Neto, Jos√© Salvator |
| hCoV-19/Germany/BW-RKI-I-256031/2021 | EPI_ISL_4612791 | 2021-09-17 | SYNLAB MVZ Leinfelden-Echterdingen | Robert Koch Institute | unknown |
| hCoV-19/Brazil/SP-IB_129322/2021 | EPI_ISL_4168877 | 2021-08-27 | USF II ALADIA ARAN RODRIGUES | Instituto Butantan | Dimas Tadeu Covas, Antonio Jorge Martins, Claudia Renata dos Santos Barros, David Schlesinger, Debora Botequio Moretti, Elaine Cristina Marqueze, Elaine Vieira Santos, Evandra Strazza Rodrigues, Heidge Fukumasu, Jayme Augusto de Souza-Neto, Jos√© Salvator |
| hCoV-19/Canada/BC-BCCDC-91011/2021 | EPI_ISL_3487304 | 2021-05-21 | British Columbia Centre For Disease Control | BCCDC Public Health Laboratory | Prystajecky Natalie, Linda Hoang, Dan Fornika, John Tyson, Shannon Russell, Kim Macdonald, Kimia Kamelian, Ana Pacagnella, Corrinne Ng, Loretta Janz, Robert Azana, Mel Krajden |
| hCoV-19/Canada/BC-BCCDC-103593/2021 | EPI_ISL_3488665 | 2021-05-27 | British Columbia Centre For Disease Control | BCCDC Public Health Laboratory | Prystajecky Natalie, Linda Hoang, Dan Fornika, John Tyson, Shannon Russell, Kim Macdonald, Kimia Kamelian, Ana Pacagnella, Corrinne Ng, Loretta Janz, Robert Azana, Mel Krajden |
| hCoV-19/Guam/GU-CDC-2-4718128/2021 | EPI_ISL_3491413 | 2021-07-12 | Guam Public Health Laboratory | Centers for Disease Control and Prevention Division of Viral Diseases, Pathogen Discovery | Mili Sheth,Sarah Nobles,Jasmine Padilla,Alex Burgin,Meghan Bentz,Mark Burroughs,Peter Cook,Clinton Paden,Dhwani Batra,Krista Queen,Kristen Knipe,Dakota Howard,Yvette Unoarumhi,Matthew Schmerer,Ben Rambo-Martin,Kristine Lacek,Sam Shepard,Dave Wentworth,Viv |
| hCoV-19/USA/MS-UMMC-M756A7-506625/2021 | EPI_ISL_3492856 | 2021-07-29 | University of Mississippi Medical Center, Department of Pathology | University of Mississippi Medical Center, Molecular and Genomics Core Facility | Ashley C. Johnson, Wenjie Wu, Ithiel J. Frame, Krishna K. Ayyalasomayajula, Michael R. Garrett, D. Ashley Robinson |
| hCoV-19/Brazil/PR-FIOCRUZ-43884/2021 | EPI_ISL_4170298 | 2021-07-29 | Laboratorio Central de Saude Publica do Estado do Parana (LACEN/PR) | Laboratory of Respiratory Viruses and Measles, Oswaldo Cruz Institute, FIOCRUZ | Paola Resende, Luciana Appolinario, Fernando Motta, Anna Carolina Paixao, Ighor Leonardo Arantes, Agatha Soares, Ana Carolina Mendonca, Alice Sampaio Rocha, Taina Venas, Elisa Cavalcante Pereira, Renata Serrano Lopes, Irina Riediger, Marilda Siqueira on b |

| | | | | | |
|---|---|---|---|---|---|
| hCoV-19/Germany/BW-RKI-I-201522/2021 | EPI_ISL_3501343 | 2021-07-26 | Landesgesundheitsamt Baden-W√ºrttemberg | Robert Koch Institute | unknown |
| hCoV-19/Germany/BW-RKI-I-201525/2021 | EPI_ISL_3501346 | 2021-07-26 | Landesgesundheitsamt Baden-W√ºrttemberg | Robert Koch Institute | unknown |
| hCoV-19/Italy/LAZ-UCBM-255/2021 | EPI_ISL_3501818 | 2021-08-12 | Microbiology and Virology IFO, Rome, Italy | University Campus Bio-Medico of Rome (UCBM) | Riva E., De Florio L., Fogolari M., Francesconi M., Lintas C., Veralli R., Angeletti S. |
| hCoV-19/Italy/LAZ-UCBM-270/2021 | EPI_ISL_3501846 | 2021-08-12 | Microbiology and Virology IFO, Rome, Italy | University Campus Bio-Medico of Rome (UCBM) | Riva E., De Florio L., Fogolari M., Francesconi M., Lintas C., Veralli R., Angeletti S. |
| hCoV-19/Ecuador/USFQ-1926/2021 | EPI_ISL_3505581 | 2021-08-05 | Center for Research on Health in Latin America | Institute of Microbiology, Universidad San Francisco de Quito | Sully M√°rquez, Juan Jos√© Guadalupe, Bel√©n Prado-Vivar, Monica Becerra-Wong, Erika Mu√±oz, Fernanda Zurita, Bernardo Guti√©rrez, Jaime Costales, Ver√≥nica Barrag√°n, Patricio Rojas-Silva, Gabriel Trueba, Michelle Grunauer, Pa√∫l C√°rdenas |
| hCoV-19/Ecuador/USFQ-1878/2021 | EPI_ISL_3505615 | 2021-08-02 | Institute of Microbiology, Universidad San Francisco de Quito | Institute of Microbiology, Universidad San Francisco de Quito | Sully M√°rquez, Juan Jos√© Guadalupe, Bel√©n Prado-Vivar, Erika B. Mu√±oz, Monica Becerra-Wong, Fernanda Zurita, Bernardo Guti√©rrez, Christian Zambrano, Juan Carlos Zu√±iga, Ver√≥nica Barrag√°n, Patricio Rojas-Silva, Gabriel Trueba, Michelle Grunauer, Pa |
| hCoV-19/Spain/CL-COV13881/2021 | EPI_ISL_4506394 | 2021-09-01 | SARS-CoV-2 Sequencing Castilla y Leon-Spain Consortium | SARS-CoV-2 Sequencing Castilla y Leon-Spain Consortium | Antonio Ordu√±a-Domingo, Marta Hernandez, David Abad, Marta Dominguez-Gil, Silvia Rojo, Gabriel March Rosello, Sonsoles Garcinu√±o P√©rez, Carmen Aldea-Mansilla, M¬™ Fe Brezmes-Valdivieso, Gregoria Meg√≠as Lob√≥n, Mar√≠a Antonia Garc√≠a Castro, Carmen Gim |
| hCoV-19/Belgium/CHUNamur13544973/2021 | EPI_ISL_3506177 | 2021-08-11 | Plateforme de testing Namuroise | Plateforme de testing Namuroise | Lesly Nyinkeu Kemamen; Nicolas Gilliard; Demars Aurore ; Maschietto C√©line ; Nobis Chlo√© ; Otto Gaetan ; Denis Olivier ; Degosserie Jonathan ; Mullier Fran√ßois |
| hCoV-19/USA/CA-CDPH-2000047529/2021 | EPI_ISL_3243768 | 2021-05-07 | CDPH VBL | California Department of Public Health | Emily Smith on behalf of CDPH-COVIDNet and UCSD EXCITE lab |
| hCoV-19/France/PDL-IPP19638/2021 | EPI_ISL_3506801 | 2021-07-27 | Labo Analyses Med | National Reference Center for Viruses of Respiratory Infections, Institut Pasteur, Paris | Marion Barbet, Sylvie Behillil, M√©line Bizard, Angela Brisebarre, Camille Capel, Vincent Enouf, Louise Lefran√ßois, Fr√©d√©ric Lemoine, Christophe Malabat, Corinne Maufrais, Hub de Bioinformatique et Biostatistique, Etienne Simon-Lori√®re, Maud Vanpeene, |
| hCoV-19/India/TG-CDFD-WGS26_GMC_SDPT/2021 | EPI_ISL_3507496 | 2021-07-23 | Government General Hospital | Centre for DNA Fingerprinting and Diagnostics | Murali Bashyam, Asmita Gupta, Reelina Basu, Vinay Donipadi, Nagamani Kammili, Divya Vashisht, Ashwin Dalal |
| hCoV-19/Canada/QC-1nEAQ-0667616/2021 | EPI_ISL_3508340 | 2021-01-26 | Laboratoire de sant√© publique du Qu√©bec | Laboratoire de sant√© publique du Qu√©bec | Sandrine Moreira, Ioannis Ragoussis, Guillaume Bourque, Jesse Shapiro, Mark Lathrop and Michel Roger on behalf of the CoVSeQ research group |
| hCoV-19/USA/NY-CDC-LC0228454/2021 | EPI_ISL_4173515 | 2021-08-20 | Laboratory Corporation of America | Centers for Disease Control and Prevention Division of Viral Diseases, Pathogen Discovery | Dakota Howard,Dhwani Batra,Peter Cook,Kara Moser,Adrian Paskey,Jason Caravas,Benjamin Rambo-Martin,Shatavia Morrison,Christopher Gulvick,Scott Sammons,Yvette Unoarumhi,Darlene Wagner,Matthew Schmerer,Minoo Agarwal,Eyad Almasri,Debbie Boles,Ayla Burns,Nuth |

| | | | | | |
|---|---|---|---|---|---|
| hCoV-19/USA/PA-CDC-LC0140657/2021 | EPI_ISL_3513562 | 2021-07-26 | Laboratory Corporation of America | Centers for Disease Control and Prevention Division of Viral Diseases, Pathogen Discovery | Dakota Howard,Dhwani Batra,Peter Cook,Kara Moser,Adrian Paskey,Jason Caravas,Benjamin Rambo-Martin,Shatavia Morrison,Christopher Gulvick,Scott Sammons,Yvette Unoarumhi,Darlene Wagner,Matthew Schmerer,Minoo Agarwal,Eyad Almasri,Debbie Boles,Ayla Burns,Nuth |
| hCoV-19/USA/NY-CDC-LC0141074/2021 | EPI_ISL_3514252 | 2021-07-27 | Laboratory Corporation of America | Centers for Disease Control and Prevention Division of Viral Diseases, Pathogen Discovery | Dakota Howard,Dhwani Batra,Peter Cook,Kara Moser,Adrian Paskey,Jason Caravas,Benjamin Rambo-Martin,Shatavia Morrison,Christopher Gulvick,Scott Sammons,Yvette Unoarumhi,Darlene Wagner,Matthew Schmerer,Minoo Agarwal,Eyad Almasri,Debbie Boles,Ayla Burns,Nuth |
| hCoV-19/Spain/CL-COV11373/2021 | EPI_ISL_3909024 | 2021-07-27 | SARS-CoV-2 Sequencing Castilla y Leon-Spain Consortium | SARS-CoV-2 Sequencing Castilla y Leon-Spain Consortium | Antonio Orduv±a-Domingo, Marta Hernandez, David Abad, Marta Dominguez-Gil, Silvia Rojo, Gabriel March Rosello, Sonsoles Garcinuv±o Pv©rez, Carmen Aldea-Mansilla, M¬™ Fe Brezmes-Valdivieso, Gregoria Megv≠as Lobv≥n, Marv≠a Antonia Garcv≠a Castro, Carmen Gim |
| hCoV-19/France/NAQ-HCL021157060001/2021 | EPI_ISL_4324117 | 2021-08-31 | BIOFFICE - LBM RAVEZIES | CNR Virus des Infections Respiratoires - France SUD | Antonin Bal, Gregory Destras, Gwendolyne Burfin, Hadrien Regue, Quentin Semanas, Martine Valette, Bruno Lina, Laurence Josset |
| hCoV-19/USA/FL-CDC-LC0154191/2021 | EPI_ISL_3517186 | 2021-07-29 | Laboratory Corporation of America | Centers for Disease Control and Prevention Division of Viral Diseases, Pathogen Discovery | Dakota Howard,Dhwani Batra,Peter Cook,Kara Moser,Adrian Paskey,Jason Caravas,Benjamin Rambo-Martin,Shatavia Morrison,Christopher Gulvick,Scott Sammons,Yvette Unoarumhi,Darlene Wagner,Matthew Schmerer,Minoo Agarwal,Eyad Almasri,Debbie Boles,Ayla Burns,Nuth |
| hCoV-19/Japan/PG-77718/2021 | EPI_ISL_3888717 | 2021-07-23 | Pathogen Genomics Center, National Institute of Infectious Diseases | Pathogen Genomics Center, National Institute of Infectious Diseases | Tsuyoshi Sekizuka, Kentaro Itokawa, Rina Tanaka, Masanori Hashino, Makoto Kuroda |
| hCoV-19/USA/MD-IGS-52122511301A/2021 | EPI_ISL_4178731 | 2021-08-13 | Maryland Genomics, Institute for Genome Sciences, University of Maryland School of Medicine | Maryland Genomics, Institute for Genome Sciences, University of Maryland School of Medicine | Tallon, Luke J; Sadzewicz, Lisa D; Humphrys, Mike; Ott, Sandra; Roussey, Holly; Santana-Cruz, Ivette; Vavikolanu, Kranthi; Lim, Jonathan; Hazen, Tracy; Fraser, Claire M; Ravel, Jacques |
| hCoV-19/USA/VA-CDC-LC0227629/2021 | EPI_ISL_4178865 | 2021-08-27 | Laboratory Corporation of America | Centers for Disease Control and Prevention Division of Viral Diseases, Pathogen Discovery | Dakota Howard,Dhwani Batra,Peter Cook,Kara Moser,Adrian Paskey,Jason Caravas,Benjamin Rambo-Martin,Shatavia Morrison,Christopher Gulvick,Scott Sammons,Yvette Unoarumhi,Darlene Wagner,Matthew Schmerer,Minoo Agarwal,Eyad Almasri,Debbie Boles,Ayla Burns,Nuth |
| hCoV-19/NewZealand/21CV0502/2021 | EPI_ISL_4433106 | 2021-09-16 | Middlemore Hospital | Institute of Environmental Science and Research (ESR) | Sarah Cockerton, Clare Gebbie, Dhanisha Patel, Rachel Boyle, SallyAnn Harbison, Olivia Stroeven, Xiaoyun Ren, Matt Storey, Nikki Freed, Muhammad Faisal, Jing Wang, Hermes Perez, Anja Werno, Antje van der Linden, Arlo Upton, Chris Mansell, David Hammer, Dr |

| | | | | | |
|---|---|---|---|---|---|
| hCoV-19/Canada/AB-ABPHL-26034/2021 | EPI_ISL_3524523 | 2021-07-19 | Edmonton Provincial Lab | Public Health Agency of Canada (PHAC) National Microbiology Laboratory | Buss, E, Croxen M, Deo A, Dieu P, Gill K, Ferrato C, Khan F, Koleva P, Li V, Lloyd C, Lynch T, Ma R, Murphy S, Pabbaraju K, Shokoples S, Tipples G, Thayer J, Whitehouse M, Wong A, Yu C, Zelyas N |
| hCoV-19/England/MILK-18DD450/2020 | EPI_ISL_2998424 | 2020-11-04 | Lighthouse Lab in Milton Keynes | Wellcome Sanger Institute for the COVID-19 Genomics UK (COG-UK) Consortium | The Lighthouse Lab in Milton Keynes and Alex Alderton, Roberto Amato, Jeffrey Barrett, Sonia Goncalves, Ewan Harrison, David K. Jackson, Ian Johnston, Dominic Kwiatkowski, Cordelia Langford, John Sillitoe on behalf of the Wellcome Sanger Institute COVID-1 |
| hCoV-19/USA/RI-CDCBI-CRSP_SKPU5FS5SJROWAJ7/2021 | EPI_ISL_3525889 | 2021-08-12 | Broad Institute Clinical Research Sequencing Platform | Infectious Disease Program, Broad Institute of Harvard and MIT | Siddle,K.J., Adams,G., Pearlman,L., Gladden-Young,A., Vicente,G., Blumenstiel,B., DeFelice,M., Lee,M., McGovern,S., Lagerborg,K., Rudy,M., DeRuff,K., Carter,A., Normandin,E., Bauer,M., Reilly,S., Tomkins-Tinch,C., Loreth,C., Chaluvadi,S., Meldrim,J., Gran |
| hCoV-19/Finland/TRE24866_7/2021 | EPI_ISL_3527593 | 2021-04-24 | Fimlab Laboratories | Fimlab Laboratories | Minna Paloniemi, Leena Huhti, Sara Lehtinen, Bruno Luukinen, Tapio Seiskari, Mauri Kein√§nen |
| hCoV-19/USA/PA-M21072707/2021 | EPI_ISL_3527730 | 2021-07-28 | Pennsylvania Department of Health Bureau of Laboratories | Pennsylvania Department of Health Bureau of Laboratories | Dongxiang Xia |
| hCoV-19/Denmark/DCGC-153706/2021 | EPI_ISL_3531008 | 2021-08-13 | Department of Bacteria, Parasites and Fungi, Statens Serum Institut, Copenhagen, Denmark | Statens Serum Institut Bioinformatics and Microbial Genomics | Danish Covid-19 Genome Consortium |
| hCoV-19/Chile/RM-116322/2021 | EPI_ISL_2892956 | 2021-06-23 | Genetica Molecular and Subdepartamento de Virologia ISP Chile | Instituto de Salud Publica de Chile | Karen Orostica, Constanza Campano, Barbara Parra, Loredana Arata, Gisselle Barra, Patricia Bustos, Rodrigo Fasce, Javier Tognarelli, Andres Castillo, Soledad Ulloa, Jorge Fernandez |
| hCoV-19/India/GJ-ICMR-NIV-INSACOG-GSEQ-5177/2021 | EPI_ISL_3533102 | 2021-07-29 | ICMR-National Institute of Virology - INSACOG | NIV Influenza | Potdar, Varsha on behalf of National Influenza Centre, NIV, Pune |
| hCoV-19/USA/CA-CDPH1281/2021 | EPI_ISL_3536968 | 2021-04-27 | California Department of Public Health | California Department of Public Health | CDPH IDLB COVIDNet et al |
| hCoV-19/Chile/RM-135436/2021 | EPI_ISL_3537360 | 2021-08-04 | Genetica Molecular and Subdepartamento de Virologia ISP Chile | Instituto de Salud Publica de Chile | Karen Orostica, Constanza Campano, Barbara Parra, Loredana Arata, Gisselle Barra, Patricia Bustos, Rodrigo Fasce, Javier Tognarelli, Andres Castillo, Soledad Ulloa, Jorge Fernandez |
| hCoV-19/USA/TX-CDC-FG-110640/2021 | EPI_ISL_4183338 | 2021-08-27 | Fulgent Genetics | Centers for Disease Control and Prevention Division of Viral Diseases, Pathogen Discovery | Dakota Howard,Dhwani Batra,Peter Cook,Jason Caravas,Benjamin Rambo-Martin,Scott Sammons,Yvette Unoarumhi,Matthew Schmerer,Kristine Lacek,Tymeckia Kendall,Victoria Caban Figueroa,Shatavia Morrison,Christopher Gulvick,Erisa Sula,Harry Gao,Mickey Li,John Gao |
| hCoV-19/Turkey/HSGM-MK005/2021 | EPI_ISL_3540627 | 2021-08-13 | Ministry of Health Turkey | Ministry of Health Turkey | Fatma Bayrakdar, Yasemin Co≈ügun, S√ºleyman Yalcin, G√ºlay Korukluoƒülu |
| hCoV-19/Armenia/IMB6-5/2021 | EPI_ISL_3543621 | 2021-07-30 | National Center of Disease Control and Prevention of the Republic of Armenia, Davidyants Laboratories, Yerevan, Armenia | [illegible] RA, Republic of Armenia, Department of Bioengineering, BioinformaticsInstitute and Molecular Biology IBMPh RAU, Republic of Armenia | Arsen Arakelyan, Diana Avetyan, Siras Hakobyan, Gisane Khachatyan, Maria Nikoghosyan, Tamara Sirunyan, Nelli Muradyan, Ani Melkonyan, Andranik Chavushyan, Hovsep Ghazaryan, Roksana Zakharyan, Lilit Ghukasyan, Shushan Sargrsyan, Gayane Melik-Pashayan |

| | | | | | |
|---|---|---|---|---|---|
| hCoV-19/FrenchGuiana/IPG2021988/2021 | EPI_ISL_3547316 | 2021-08-16 | CNR Institut Pasteur de la Guyane | Institut Pasteur de la Guyane | Anne Lavergne, Dominique Rousset, Antoine Enfissi, Arielle Salmier |
| hCoV-19/FrenchGuiana/IPG2021977/2021 | EPI_ISL_3547329 | 2021-08-16 | CNR Institut Pasteur de la Guyane | Institut Pasteur de la Guyane | Anne Lavergne, Dominique Rousset, Antoine Enfissi, Arielle Salmier |
| hCoV-19/Turkey/HSGM-D5363/2021 | EPI_ISL_3555462 | 2021-08-16 | Ministry of Health Turkey | Ministry of Health Turkey | Fatma Bayrakdar, Yasemin Cosgun, Suleyman Yalcin, Gulay Korukluoglu |
| hCoV-19/Turkey/HSGM-C156/2021 | EPI_ISL_3564722 | 2021-06-30 | Ministry of Health Turkey | Ministry of Health Turkey | Fatma Bayrakdar, Yasemin Cosgun, Suleyman Yalcin, Gulay Korukluoglu |
| hCoV-19/Australia/NSW-SAVID-4734/2021 | EPI_ISL_3568446 | 2021-06-06 | Area of Virology, Serology and Virology Division (SAViD), New South Wales Health Pathology Randwick | Virology, Serology and Virology Division (SAViD), New South Wales Health Pathology Randwick | Foster, C.; Au, J.; Ruiz Silva, M.; Deveson, I.; Bull, R.; Van Hal, S.; Rawlinson, W. |
| hCoV-19/Australia/NSW-SAVID-4833/2021 | EPI_ISL_3568475 | 2021-07-08 | Area of Virology, Serology and Virology Division (SAViD), New South Wales Health Pathology Randwick | Virology, Serology and Virology Division (SAViD), New South Wales Health Pathology Randwick | Foster, C.; Au, J.; Ruiz Silva, M.; Deveson, I.; Bull, R.; Van Hal, S.; Rawlinson, W. |
| hCoV-19/Australia/NSW-SAVID-4893/2021 | EPI_ISL_3568513 | 2021-07-20 | Area of Virology, Serology and Virology Division (SAViD), New South Wales Health Pathology Randwick | Virology, Serology and Virology Division (SAViD), New South Wales Health Pathology Randwick | Foster, C.; Au, J.; Ruiz Silva, M.; Deveson, I.; Bull, R.; Van Hal, S.; Rawlinson, W. |
| hCoV-19/Australia/NSW-SAVID-4916/2021 | EPI_ISL_3568532 | 2021-07-24 | Area of Virology, Serology and Virology Division (SAViD), New South Wales Health Pathology Randwick | Virology, Serology and Virology Division (SAViD), New South Wales Health Pathology Randwick | Foster, C.; Au, J.; Ruiz Silva, M.; Deveson, I.; Bull, R.; Van Hal, S.; Rawlinson, W. |
| hCoV-19/Australia/NSW-SAVID-4917/2021 | EPI_ISL_3568533 | 2021-07-24 | Area of Virology, Serology and Virology Division (SAViD), New South Wales Health Pathology Randwick | Virology, Serology and Virology Division (SAViD), New South Wales Health Pathology Randwick | Foster, C.; Au, J.; Ruiz Silva, M.; Deveson, I.; Bull, R.; Van Hal, S.; Rawlinson, W. |
| hCoV-19/Australia/NSW-SAVID-4921/2021 | EPI_ISL_3568537 | 2021-07-25 | Area of Virology, Serology and Virology Division (SAViD), New South Wales Health Pathology Randwick | Virology, Serology and Virology Division (SAViD), New South Wales Health Pathology Randwick | Foster, C.; Au, J.; Ruiz Silva, M.; Deveson, I.; Bull, R.; Van Hal, S.; Rawlinson, W. |
| hCoV-19/USA/TX-CDC-QDX26718549/2021 | EPI_ISL_4187878 | 2021-07-11 | Quest Diagnostics Incorporated | Centers for Disease Control and Prevention Division of Viral Diseases, Pathogen Discovery | Dakota Howard,Dhwani Batra,Peter Cook,Jason Caravas,Benjamin Rambo-Martin,Scott Sammons,Yvette Unoarumhi,Matthew Schmerer,Kristine Lacek,Tymeckia Kendall,Victoria Caban Figueroa,Shatavia Morrison,Christopher Gulvick,Erisa Sula,S. Rosenthal,A. Gerasimova,R |
| hCoV-19/England/NORT-1BF4BE2/2021 | EPI_ISL_3571201 | 2021-01-24 | Northumbria University / South Tees Hospitals NHS Foundation Trust / North Cumbria Integrated Care NHS Foundation Trust / North Tees and Hartlepool NHS Foundation Trust / Newcastle Hospitals NHS Foundation Trust | COVID-19 Genomics UK (COG-UK) Consortium | Darren L Smith,Andrew Nelson,Matthew Bashton,Greg R Young,Joshua Loh,John Allan,Mohammad A Tariq,Giles S Holt,Gary Black,Wen C Yew,Lynn Dover,Paul Baker,Steve Liggett,Sarah Essex,Jane Greenaway,Debra Padgett,Clive Graham,Garren Scott,Edward Barton,Emma Sw |
| hCoV-19/England/PHEP-009083/2021 | EPI_ISL_3574615 | 2021-07-24 | Respiratory Virus Unit, Microbiology Services Colindale, Public Health England | COVID-19 Genomics UK (COG-UK) Consortium | PHE Covid Sequencing Team |

| | | | | | |
|---|---|---|---|---|---|
| hCoV-19/Pakistan/UHS2-Pak-221/2021 | EPI_ISL_3483854 | 2021-06-11 | University of Health Sciences | Quadram Institute Bioscience | Shah Jahan, Muhammad Bilal Sarwar, Muhammad Roman, Muhammad Asif Naveed, Sidra-tul-muntaha, Almina Shafiq, Khushbakth kirn, Nadeem Afzal, Javed Akram, Hazir Rahman, Waseem Iqbal, Aamir Ashiq, Asghar javeed, Tahir Yaqoob, Muhammad Nawaz, Muhhmad Ilyas, Muh |
| hCoV-19/USA/CO-CDPHE-2101392085/2021 | EPI_ISL_3583293 | 2021-07-05 | Colorado Department of Public Health and Environment | Colorado Department of Public Health and Environment | Laura Bankers, Molly C. Hetherington-Rauth, Diana Ir, Alexandria Rossheim, Michael Martin, Mandy Waters, Shannon R. Matzinger, Sarah Elizabeth Totten, Emily A. Travanty |
| hCoV-19/USA/CO-CDPHE-2101376279/2021 | EPI_ISL_3583830 | 2021-06-30 | Colorado Department of Public Health and Environment | Colorado Department of Public Health and Environment | Laura Bankers, Molly C. Hetherington-Rauth, Diana Ir, Alexandria Rossheim, Michael Martin, Mandy Waters, Shannon R. Matzinger, Sarah Elizabeth Totten, Emily A. Travanty |
| hCoV-19/USA/CO-CDPHE-2101378944/2021 | EPI_ISL_3583900 | 2021-06-30 | Colorado Department of Public Health and Environment | Colorado Department of Public Health and Environment | Laura Bankers, Molly C. Hetherington-Rauth, Diana Ir, Alexandria Rossheim, Michael Martin, Mandy Waters, Shannon R. Matzinger, Sarah Elizabeth Totten, Emily A. Travanty |
| hCoV-19/Spain/CL-COV07717/2021 | EPI_ISL_3587064 | 2021-07-02 | SARS-CoV-2 Sequencing Castilla y Leon-Spain Consortium | SARS-CoV-2 Sequencing Castilla y Leon-Spain Consortium | Antonio Ordu√±a-Domingo, Marta Hernandez, David Abad, Marta Dominguez-Gil, Silvia Rojo, Gabriel March Rosello, Sonsoles Garcinu√±o P√©rez, Carmen Aldea-Mansilla, M¬™ Fe Brezmes-Valdivieso, Gregoria Meg√≠as Lob√≥n, Mar√≠a Antonia Garc√≠a Castro, Carmen Gim |
| hCoV-19/Canada/BC-BCCDC-134320/2021 | EPI_ISL_3600391 | 2021-05-06 | British Columbia Centre For Disease Control | BCCDC Public Health Laboratory | Prystajecky Natalie, Linda Hoang, Dan Fornika, John Tyson, Shannon Russell, Kim Macdonald, Kimia Kamelian, Ana Pacagnella, Corrinne Ng, Loretta Janz, Robert Azana, Mel Krajden |
| hCoV-19/India/MN-1425500118075/2021 | EPI_ISL_3614374 | 2021-07-15 | INSACOG-MANIPUR | National Institute of Biomedical Genomics ‚Äì INSACOG | Arindam Maitra,¬†R.K.Manojkumar Singh, Nidhan Kumar Biswas,¬†Kh. Ranjana Devi, Sreedhar Chinnaswamy,¬†L. Shivadutta Singh, Saumitra Das |
| hCoV-19/India/MN-1425500118003/2021 | EPI_ISL_3614453 | 2021-07-15 | INSACOG-MANIPUR | National Institute of Biomedical Genomics ‚Äì INSACOG | Arindam Maitra,¬†R.K.Manojkumar Singh, Nidhan Kumar Biswas,¬†Kh. Ranjana Devi, Sreedhar Chinnaswamy,¬†L. Shivadutta Singh, Saumitra Das |
| hCoV-19/Australia/QLD2235/2021 | EPI_ISL_3614826 | 2021-08-13 | Queensland Health Forensic and Scientific Services | Queensland Health Forensic and Scientific Services | Chenwei Wang on behalf of Q-PHIRE Genomics |
| hCoV-19/Australia/QLD2263/2021 | EPI_ISL_3614834 | 2021-08-20 | PHV-FSS | PHV-FSS | Chenwei Wang on behalf of Q-PHIRE Genomics |
| hCoV-19/Brazil/RJ-LNN04339/2021 | EPI_ISL_3615222 | 2021-07-21 | Laboratorio Central Noel Nutels | Bioinformatics Laboratory / LNCC | Luiz G P de Almeida, Alessandra P Lamarca, Ronaldo da Silva F Jr, Liliane Cavalcante, Alexandra L Gerber, Ana Paula de C Guimaraes, Douglas Terra Machado, Erica Ramos dos Santos Nascimento, Fernanda Leitao dos Santos, Ricardo Jose Barbosa Salviano, Cintia |

| | | | | | |
|---|---|---|---|---|---|
| hCoV-19/Brazil/RJ-LNN04417/2021 | EPI_ISL_3615233 | 2021-07-23 | Laboratorio Central Noel Nutels | Bioinformatics Laboratory / LNCC | Luiz G P de Almeida, Alessandra P Lamarca, Ronaldo da Silva F Jr, Liliane Cavalcante, Alexandra L Gerber, Ana Paula de C Guimaraes, Douglas Terra Machado, Erica Ramos dos Santos Nascimento, Fernanda Leitao dos Santos, Ricardo Jose Barbosa Salviano, Cintia |
| hCoV-19/India/AS-CVD-34936/2020 | EPI_ISL_4192891 | 2020-12-02 | INSACOG-Assam | National Institute of Biomedical Genomics ‚Äì INSACOG | Arindam Maitra, Ajanta Sharma, Nidhan Kumar Biswas, Lahari Saikia, Sreedhar Chinnaswamy,¬†Kailash Chamuah, Saumitra Das |
| hCoV-19/India/WB-1931300435952/2021 | EPI_ISL_4193028 | 2021-08-11 | INSACOG-WB | National Institute of Biomedical Genomics ‚Äì INSACOG | Arindam Maitra, Bhaswati Bandyopadhyay, Nidhan Kumar Biswas, Tamal Ghosh, Sreedhar Chinnaswamy, Ajay Chakraborti, Saumitra Das |
| hCoV-19/India/WB-1931300432281/2021 | EPI_ISL_4193096 | 2021-08-06 | INSACOG-WB | National Institute of Biomedical Genomics ‚Äì INSACOG | Arindam Maitra, Bhaswati Bandyopadhyay, Nidhan Kumar Biswas, Tamal Ghosh, Sreedhar Chinnaswamy, Ajay Chakraborti, Saumitra Das |
| hCoV-19/India/AS-1828600362104/2021 | EPI_ISL_4193109 | 2021-06-27 | INSACOG-Assam | National Institute of Biomedical Genomics ‚Äì INSACOG | Arindam Maitra, Ajanta Sharma, Nidhan Kumar Biswas, Lahari Saikia, Sreedhar Chinnaswamy,¬†Kailash Chamuah, Saumitra Das |
| hCoV-19/Reunion/HCL021142927401/2021 | EPI_ISL_3635605 | 2021-07-27 | LAM DE SAINT BENOIT | CNR Virus des Infections Respiratoires - France SUD | Antonin Bal, Gregory Destras, Gwendolyne Burfin, Hadrien Regue, Quentin Semanas, Martine Valette, Bruno Lina, Laurence Josset |
| hCoV-19/India/WB-1931501996076/2021 | EPI_ISL_4193335 | 2021-08-10 | INSACOG-WB | National Institute of Biomedical Genomics ‚Äì INSACOG | Arindam Maitra, Bhaswati Bandyopadhyay, Nidhan Kumar Biswas, Tamal Ghosh, Sreedhar Chinnaswamy, Ajay Chakraborti, Saumitra Das |
| hCoV-19/India/WB-1970200054629/2021 | EPI_ISL_4193337 | 2021-07-17 | INSACOG-WB | National Institute of Biomedical Genomics ‚Äì INSACOG | Arindam Maitra, Bhaswati Bandyopadhyay, Nidhan Kumar Biswas, Tamal Ghosh, Sreedhar Chinnaswamy, Ajay Chakraborti, Saumitra Das |
| hCoV-19/India/AS-1828300375855/2021 | EPI_ISL_4193358 | 2021-07-03 | INSACOG-Assam | National Institute of Biomedical Genomics ‚Äì INSACOG | Arindam Maitra, Ajanta Sharma, Nidhan Kumar Biswas, Lahari Saikia, Sreedhar Chinnaswamy,¬†Kailash Chamuah, Saumitra Das |
| hCoV-19/SouthAfrica/NICD-N9868/2021 | EPI_ISL_3644068 | 2021-06-08 | PORT ELIZABETH LABORATORY | Diseases of the National Health Laboratory Service | Amoako DG, Everatt J, Scheepers C, Mohale T, Ntuli N, Mahlangu B, Mnguni A, Ismail A, Bhiman JN |
| hCoV-19/Turkey/HSGM-D15680/2021 | EPI_ISL_3647342 | 2021-08-19 | Ministry of Health Turkey | Ministry of Health Turkey | Fatma Bayrakdar, Yasemin Co≈ügun, S√ºleyman Yalcin, G√ºlay Korukluoƒülu |
| hCoV-19/USA/NM-NMDOH-2021143918/2021 | EPI_ISL_3647937 | 2021-08-05 | New Mexico Department of Health Scientific Laboratory | New Mexico Department of Health Scientific Laboratory | Ellie Johnson, Anastacia Griego-Fisher, D'eldra Malone, Jennifer Benoit |
| hCoV-19/Turkey/HSGM-D21994/2021 | EPI_ISL_3653313 | 2021-08-20 | Ministry of Health Turkey | Ministry of Health Turkey | Fatma Bayrakdar, Yasemin Co≈ügun, S√ºleyman Yalcin, G√ºlay Korukluoƒülu |
| hCoV-19/Norway/18229/2021 | EPI_ISL_4195207 | 2021-08-22 | Department of Medical Microbiology, St. Olavs hospital | Norwegian Institute of Public Health, Department of Virology | Kathrine Stene-Johansen, Kamilla Heddeland Instefjord, Hilde Elshaug, Garcia Llorente Ignacio, Jon Br√•te, Engebretsen Serina Beate,Pedersen Benedikte Nevjen, Line Victoria Moen, Debech Nadia, Atiya R Ali,Marie Paulsen Madsen, Rasmus Riis Kopperud, Hilde |

| | | | | | |
|---|---|---|---|---|---|
| hCoV-19/USA/MN-CDC-IBX126418446065/2021 | EPI_ISL_4344456 | 2021-09-02 | Infinity Biologix | Centers for Disease Control and Prevention Division of Viral Diseases, Pathogen Discovery | Dakota Howard,Dhwani Batra,Peter Cook,Jason Caravas,Benjamin Rambo-Martin,Scott Sammons,Yvette Unoarumhi,Matthew Schmerer,Kristine Lacek,Tymeckia Kendall,Victoria Caban Figueroa,Shatavia Morrison,Christopher Gulvick,Erisa Sula,Christian Bixby,Yihe Wang,Jo |
| hCoV-19/Australia/WA806/2021 | EPI_ISL_4346654 | 2021-09-10 | PathWest Laboratory Medicine WA | PathWest Laboratory Medicine WA Microbial Surveillance Unit | PathWest Laboratory Medicine WA Microbial Surveillance Unit |
| hCoV-19/SouthAfrica/UFS-VIRO-NGS-MSQ090921BC01/2021 | EPI_ISL_4348729 | 2021-08-10 | NHLS Universitas Academic | UFS Virology | PA Bester, MM Nyaga, P Nthiga, MT Mogotsi, Emmanuel Ogunbayo, D Goedhals, T de Oliveira |
| hCoV-19/England/LSPA-1E8F9A3/2021 | EPI_ISL_4349563 | 2021-09-14 | Rosalind Franklin Laboratory | Wellcome Sanger Institute for the COVID-19 Genomics UK (COG-UK) Consortium | Donald Fraser, Suki Lee, Rob Howes, The Rosalind Franklin Laboratory and Alex Alderton, Roberto Amato, Jeffrey Barrett, Sonia Goncalves, Ewan Harrison, David K. Jackson, Ian Johnston, Dominic Kwiatkowski, Cordelia Langford, John Sillitoe on behalf of the |
| hCoV-19/Mozambique/CERI-KRISP-K021127/2021 | EPI_ISL_3663557 | 2021-07-01 | Instituto Nacional de Saude (INS), Mozambique | [illegible] and Innvoation, Stellenbosch University and KRISP, KZN Research Innovation and Sequencing Platform, UKZN. | Nalia Ismael, Nadia Sitoe, Paulo Arnaldo, Nedio Mabunda, Giandhari J, Pillay S, Yajna Ramphal, Naidoo Yeshnee, Tshabuila Derek, Emmanuel S, Tegally H, Wilkinson E, de Oliveira T |
| hCoV-19/Malawi/CERI-KRISP-K020854/2021 | EPI_ISL_3663612 | 2021-04-13 | National HIV Reference Laboratory, Ministry of Health, Public Health Institute of Malawi | KRISP, KZn Research Innovation and Sequencing Platform | Mvula B, Chilima B, Chiwaula M, Mwangomba W, Panja L, Kasambara W, Auld A, Kim L, Kampira E, Kaba M, Wadonda N, Maida A, Giandhari J, Pillay S, Naidoo Y, Lessells R, Emmanuel SJ, Tegally H,Tshiabuila Derek, Wilkinson E, de Oliveira T |
| hCoV-19/Malawi/CERI-KRISP-K020875/2021 | EPI_ISL_3663627 | 2021-06-18 | National HIV Reference Laboratory, Ministry of Health, Public Health Institute of Malawi | KRISP, KZn Research Innovation and Sequencing Platform | Mvula B, Chilima B, Chiwaula M, Mwangomba W, Panja L, Kasambara W, Auld A, Kim L, Kampira E, Kaba M, Wadonda N, Maida A, Giandhari J, Pillay S, Naidoo Y, Lessells R, Emmanuel SJ, Tegally H,Tshiabuila Derek, Wilkinson E, de Oliveira T |
| hCoV-19/Malawi/CERI-KRISP-K020968/2021 | EPI_ISL_3663663 | 2021-03-18 | National HIV Reference Laboratory, Ministry of Health, Public Health Institute of Malawi | KRISP, KZn Research Innovation and Sequencing Platform | Mvula B, Chilima B, Chiwaula M, Mwangomba W, Panja L, Kasambara W, Auld A, Kim L, Kampira E, Kaba M, Wadonda N, Maida A, Giandhari J, Pillay S, Naidoo Y, Lessells R, Emmanuel SJ, Tegally H,Tshiabuila Derek, Wilkinson E, de Oliveira T |
| hCoV-19/Malawi/CERI-KRISP-K020981/2021 | EPI_ISL_3663667 | 2021-03-17 | National HIV Reference Laboratory, Ministry of Health, Public Health Institute of Malawi | KRISP, KZn Research Innovation and Sequencing Platform | Mvula B, Chilima B, Chiwaula M, Mwangomba W, Panja L, Kasambara W, Auld A, Kim L, Kampira E, Kaba M, Wadonda N, Maida A, Giandhari J, Pillay S, Naidoo Y, Lessells R, Emmanuel SJ, Tegally H,Tshiabuila Derek, Wilkinson E, de Oliveira T |
| hCoV-19/Malawi/CERI-KRISP-K021019/2021 | EPI_ISL_3663673 | 2021-03-18 | National HIV Reference Laboratory, Ministry of Health, Public Health Institute of Malawi | KRISP, KZn Research Innovation and Sequencing Platform | Mvula B, Chilima B, Chiwaula M, Mwangomba W, Panja L, Kasambara W, Auld A, Kim L, Kampira E, Kaba M, Wadonda N, Maida A, Giandhari J, Pillay S, Naidoo Y, Lessells R, Emmanuel SJ, Tegally H,Tshiabuila Derek, Wilkinson E, de Oliveira T |

| | | | | | |
|---|---|---|---|---|---|
| hCoV-19/USA/CO-CDC-MMB09671148/2021 | EPI_ISL_4196457 | 2021-08-10 | Mako Medical | Centers for Disease Control and Prevention Division of Viral Diseases, Pathogen Discovery | Dakota Howard,Dhwani Batra,Peter Cook,Jason Caravas,Benjamin Rambo-Martin,Scott Sammons,Yvette Unoarumhi,Matthew Schmerer,Kristine Lacek,Tymeckia Kendall,Victoria Caban Figueroa,Shatavia Morrison,Christopher Gulvick,Erisa Sula,Matthew Tugwell,Lauren Moon, |
| hCoV-19/USA/MN-MDH-10452/2021 | EPI_ISL_3668301 | 2021-08-10 | Centracare Laboratory Services | Minnesota Department of Health, Public Health Laboratory | Alexandra Lorentz, Jacob Garfin, Matt Plumb, and Xiong Wang |
| hCoV-19/USA/CA-CDPH-2000049572/2021 | EPI_ISL_3669449 | 2021-06-09 | CDPH VBL | California Department of Public Health | Emily Smith on behalf of CDPH-COVIDNet and UCSD EXCITE lab |
| hCoV-19/Colombia/DC-INS-368587/2021 | EPI_ISL_4196897 | 2021-04-10 | LSP DISTRITAL BOGOTA | Gencore - Universidad de los Andes | Luisa Sacristan, Cristian Barrera, Gabriela Ariza, Marcela Guevara, Silvia Restrepo, Johana Hernandez, Gabriela Delgado, Alejandro Gomez |
| hCoV-19/USA/MN-MDH-10306/2021 | EPI_ISL_3672488 | 2021-07-23 | Minnesota Department of Health, Public Health Laboratory | Minnesota Department of Health, Public Health Laboratory | Alexandra Lorentz, Jacob Garfin, Matt Plumb, and Xiong Wang |
| hCoV-19/USA/IL-CDC-LC0170543/2021 | EPI_ISL_3679394 | 2021-07-30 | Laboratory Corporation of America | Centers for Disease Control and Prevention Division of Viral Diseases, Pathogen Discovery | Dakota Howard,Dhwani Batra,Peter Cook,Kara Moser,Adrian Paskey,Jason Caravas,Benjamin Rambo-Martin,Shatavia Morrison,Christopher Gulvick,Scott Sammons,Yvette Unoarumhi,Darlene Wagner,Matthew Schmerer,Minoo Agarwal,Eyad Almasri,Debbie Boles,Ayla Burns,Nuth |
| hCoV-19/USA/NJ-CDC-LC0172253/2021 | EPI_ISL_3685386 | 2021-08-07 | Laboratory Corporation of America | Centers for Disease Control and Prevention Division of Viral Diseases, Pathogen Discovery | Dakota Howard,Dhwani Batra,Peter Cook,Kara Moser,Adrian Paskey,Jason Caravas,Benjamin Rambo-Martin,Shatavia Morrison,Christopher Gulvick,Scott Sammons,Yvette Unoarumhi,Darlene Wagner,Matthew Schmerer,Minoo Agarwal,Eyad Almasri,Debbie Boles,Ayla Burns,Nuth |
| hCoV-19/USA/IN-INSPHL-07439/2021 | EPI_ISL_3688222 | 2021-08-04 | IN State Department of Health Laboratory Services | IN State Department of Health Laboratory Services | Cassandra Campion, Jamie Yeadon, Brian Pope, Lixia Liu, Kyle Brownlee, Melissa Hindenlang, Mark Glazier |
| hCoV-19/Turkey/HSGM-D25685/2021 | EPI_ISL_3688725 | 2021-08-21 | Ministry of Health Turkey | Ministry of Health Turkey | Fatma Bayrakdar, Yasemin Cosgun, Suleyman Yalcin, Gulay Korukluoglu |
| hCoV-19/India/AP-CCMB-CIA7109/2021 | EPI_ISL_3689735 | 2021-07-07 | AP SSO | CSIR-Centre for Cellular and Molecular Biology - INSACOG | Shreekant Verma, Amareshwar Vodapalli ,Ara Sreenivas,B Himasri,Valli Nagalakshmi Undamatla,Payel Mukherjee, Lamuk Zaveri, Onkar Kulkarni,Tulasi Nagabandi, Sofia Banu, Jandhyala Sai Krishna,Sumedha Avadhanula, Priya Nurkurthy, Vidhyadhari Methuku, Archana |
| hCoV-19/India/AP-CCMB-CIA6919/2021 | EPI_ISL_3689503 | 2021-06-30 | AP SSO | CSIR-Centre for Cellular and Molecular Biology - INSACOG | Payel Mukherjee, Lamuk Zaveri, Onkar Kulkarni,Tulasi Nagabandi, Sofia Banu, Ara Sreenivas,Shreekant Verma, Amareshwar Vodapalli ,B Himasri,Valli Nagalakshmi Undamatla, Sumedha Avadhanula, Vidhyadhari Methuku, Priya Nurkurthy, Archana Bharadwaj Siva, Karth |

| | | | | | |
|---|---|---|---|---|---|
| hCoV-19/Martinique/IPP20339/2021 | EPI_ISL_3690134 | 2021-07-26 | Hospital | National Reference Center for Viruses of Respiratory Infections, Institut Pasteur, Paris | Marion Barbet, Sylvie Behillil, M√©line Bizard, Angela Brisebarre, Camille Capel, Vincent Enouf, Louise Lefran√ßois, Fr√©d√©ric Lemoine, Christophe Malabat, Corinne Maufrais, Hub de Bioinformatique et Biostatistique, Etienne Simon-Lori√®re, Maud Vanpeene, |
| hCoV-19/Indonesia/JT-UGM-2123929/2021 | EPI_ISL_2964956 | 2021-06-04 | RSUD DR. MOEWARDI | Genetics Working Group (Pokja Genetik) Faculty of Medicine, Public Health and Nursing Universitas Gadjah Mada (FK-KMK UGM); Disease Investigation Center Wates Ministry of Agriculture Indonesia; Department of Microbiology FK-KMK UGM; Laboratorium Diagnosti | Gunadi, Hendra Wibawa, Marcellus, Mohamad S. Hakim, Edwin W. Daniwijaya, Ludhang P. Rizki, Endah Supriyati, Eggi Arguni, Titik Nuryastuti, Tri Wibawa, Dwi AA Nugrahaningsih, Afiahayati, Siswanto,Kristy Iskandar,Nungki Anggorowati, Pramesti G Dewi, Dwi Ind |
| hCoV-19/Sweden/6174459663/2021 | EPI_ISL_3690865 | 2021-08-10 | Sweden/Sodermanland/Unilabs | Unilabs/Eskilstuna/Sweden | Emma Arvidsson |
| hCoV-19/USA/CA-CDPH-500000089/2021 | EPI_ISL_3695256 | 2021-06-21 | TPMG Regional Laboratory | California Department of Public Health | Emily Smith on behalf of CDPH-COVIDNet and UCI Genome Sciences Center/GHTF |
| hCoV-19/Peru/LIM-UPCH-0946/2020 | EPI_ISL_3707361 | 2020-10-14 | Laboratorio de Gen√≥mica Microbiana, Universidad Peruana Cayetano Heredia | Laboratorio de Gen√≥mica Microbiana, Universidad Peruana Cayetano Heredia | Pablo Tsukayama, Alejandra D√°vila-Barclay, Guillermo Salvatierra, Luis Gonz√°lez, Pedro E. Romero, Diego Cuicapuza, Janet Huancachoque, Pool Marcos |
| hCoV-19/Chile/RM-USS-UNAB-BFT823/2021 | EPI_ISL_4199584 | 2021-08-28 | Universidad San Sebastian | Facultad de Ciencias de la Vida, UNAB | Claudio Meneses, Eduardo Castro, Waldo Diaz, Macarena Bastias, Ricardo Yusta, Susana Saez, Dayan Sanhueza, Sebastian Wolter, Daniel Leon, Gonzalo Campa√±a, Paola Pidal, V√≠ctor Monreal, Claudio Olmos, Ariel Orellana |
| hCoV-19/Chile/RM-HCUCH-UNAB-92228819/2021 | EPI_ISL_4199599 | 2021-08-23 | Lab. Hospital clinico Universidad de Chile | Facultad de Ciencias de la Vida, UNAB | Claudio Meneses, Eduardo Castro, Waldo Diaz, Macarena Bastias, Ricardo Yusta, Susana Saez, Dayan Sanhueza, Sebastian Wolter, Daniel Leon, Gonzalo Campa√±a, Paola Pidal, V√≠ctor Monreal, Claudio Olmos, Ariel Orellana |
| hCoV-19/Chile/RM-USS-UNAB-ADZ320/2021 | EPI_ISL_4199617 | 2021-08-25 | Universidad San Sebastian | Facultad de Ciencias de la Vida, UNAB | Claudio Meneses, Eduardo Castro, Waldo Diaz, Macarena Bastias, Ricardo Yusta, Susana Saez, Dayan Sanhueza, Sebastian Wolter, Daniel Leon, Gonzalo Campa√±a, Paola Pidal, V√≠ctor Monreal, Claudio Olmos, Ariel Orellana |
| hCoV-19/Denmark/DCGC-134407/2021 | EPI_ISL_3709241 | 2021-07-20 | Department of Bacteria, Parasites and Fungi, Statens Serum Institut, Copenhagen, Denmark | Statens Serum Institut Bioinformatics and Microbial Genomics | Danish Covid-19 Genome Consortium |
| hCoV-19/Denmark/DCGC-140969/2021 | EPI_ISL_3709509 | 2021-07-30 | Department of Bacteria, Parasites and Fungi, Statens Serum Institut, Copenhagen, Denmark | Statens Serum Institut Bioinformatics and Microbial Genomics | Danish Covid-19 Genome Consortium |
| hCoV-19/Australia/SA0937/2021 | EPI_ISL_3710513 | 2021-08-22 | SA Pathology | SA Pathology | Caitlin Selway, Lex Leong, Chuan Kok Lim, Mark Turra, Ivan Bastian |
| hCoV-19/France/OCC-CHU-TLS-2118112407/2021 | EPI_ISL_3710589 | 2021-06-30 | CHU Purpan - Laboratoire de Virologie - Institut F√©d√©ratif de Biologie | CHU Purpan - Laboratoire de Virologie - Institut F√©d√©ratif de Biologie | Latour J., Jeanne N., Ranger N., Carcenac R., Harter A., Boyer P., Ferrer V., Tremeaux R., Izopet J. |

| | | | | | |
|---|---|---|---|---|---|
| hCoV-19/Madagascar/IPM-01847/2021 | EPI_ISL_3716705 | 2021-02-15 | Virology Unit, Institut Pasteur de Madagascar | Institut Pateur de Dakar | Norosoa Razanajatovo, Cara E. Brook, Diagne Moussa Mo√Øse, Diallo Amadou, Dia Ndongo, Christian Ranaivoson, Soa Fy Andriamandimby, Vololoniaina Raharinosy, Sankhe Safietou, Ndiaye Ndack, Tsiry Randriambolamanantsoa, Helisoa Razafimanjato, Jean-Michel Hera |
| hCoV-19/India/GJ-INSACOG-GBRC2178/2020 | EPI_ISL_3717689 | 2020-12-12 | SMIMER Hospital & Medical College, Surat | Gujarat Biotechnology Research Centre | Nitin Shukla,Ramesh Pandit,Janvi Raval,Zarna Patel,Dinesh Kumar,Arpit Shukla,Bhadreshsinh Gohil,Nimesh Patel,Manish Patel,Chaitanya Joshi,Madhvi Joshi |
| hCoV-19/India/GJ-INSACOG-GBRC1057/2021 | EPI_ISL_3717807 | 2021-05-17 | Government Medical College (GMC), Surat | Gujarat Biotechnology Research Centre | Bhadreshsinh Gohil,Nimesh Patel,Nitin Shukla,Ramesh Pandit,Janvi Raval,Zarna Patel,Dinesh Kumar,Arpit Shukla,Neeta Khandelwal,Chaitanya Joshi,Madhvi Joshi |
| hCoV-19/SouthAfrica/NICD-N10031/2021 | EPI_ISL_3717913 | 2021-06-10 | PORT ELIZABETH LABORATORY | Diseases of the National Health Laboratory Service | Amoako DG, Everatt J, Scheepers C, Mohale T, Ntuli N, Mahlangu B, Mnguni A, Ismail A, Bhiman JN |
| hCoV-19/SouthAfrica/NICD-R12076/2021 | EPI_ISL_3717919 | 2021-08-12 | National Institute for Communicable Diseases of the National Health Laboratory Service | Diseases of the National Health Laboratory Service | Amoako DG, Everatt J, Scheepers C, Mohale T, Ntuli N, Mahlangu B, Mnguni A, Ismail A, Bhiman JN |
| hCoV-19/USA/WY-WYPHL-21059927/2021 | EPI_ISL_3718910 | 2021-08-13 | Wyoming Public Health Laboratory | Wyoming Public Health Laboratory | Jim Mildenberger, Taylor Fearing, Channing Weber, Ashley Norberg, Chayse Rowley, Marley Goetz, Brian Dominguez, Elliot Thomasson, Sam Britz, Cari Sloma, Robert Petit, and Rob Christensen |
| hCoV-19/Australia/NSW-RPAH-1083/2021 | EPI_ISL_3719108 | 2021-07-28 | New South Wales Health Pathology Royal Prince Alfred Hospital | Microbiology RPAH | Foster, C.; Au, J.; Ruiz Silva, M.; Deveson, I.; Bull, R.; Van Hal, S.; Rawlinson, W. |
| hCoV-19/Colombia/ANT-CWOHC-VG-SEC01556V/2021 | EPI_ISL_3721595 | 2021-02-25 | Hospital Pablo Tobon | Universidad Nacional de Colombia - Laboratorio Gen√≥mico One Health | Maria Stella L√≥pez, Andres F. Cardona-Rios, Daniel O. Maldonado-Perez, Sim√≥n Villegas Vel√°squez, Lina M. Hurtado, Laura Silvana Perez, Karl A Ciuoderis, Maria Ang√©lica Maya, Carolina Mu√±oz-Arango, Celeny Ortiz, Rita Almanza Payares, Idabely Betancur |
| hCoV-19/SouthAfrica/KRISP-K021232/2021 | EPI_ISL_3722217 | 2021-07-31 | NHLS Charlotte Maxeke Johannesburg Hospital | KRISP, KZn Research Innovation and Sequencing Platform | Kathleen Subramoney, Florette Treurnicht, Giandhari J, Pillay S, Yajna Ramphal, Naidoo Yeshnee, Tshabuila Derek, Emmanuel S, Tegally H, Wilkinson E, de Oliveira T |
| hCoV-19/Zimbabwe/CERI-KRISP-K021322/2021 | EPI_ISL_3722279 | 2021-07-26 | National Microbiology Reference Laboratory, Ministry of Health, Harare, Zimbabwe | CERI, Centre for Epidemic Response and Innvoation, Stellenbosch University and KRISP, KZN Research Innovation and Sequencing Platform, UKZN. | Air Comodor Dr J. Chimedza,¬†Dr Raiva Simbi,¬†Agnes Juru,¬†Tapfumanei Mashe,¬†Kenneth Maeka,¬†Hlanai Gumbo,¬†Tatenda Takawira,¬†Charles Nyagupe, Giandhari J, Pillay S, Naidoo Y, Emmanuel SJ, Tegally H, Wilkinson E, de Oliveira T |
| hCoV-19/SouthAfrica/KRISP-K021402/2021 | EPI_ISL_3722336 | 2021-08-03 | AHRI | KRISP, KZn Research Innovation and Sequencing Platform | Sigal Alex, Cele Sandile, Karim Farina, Khan Khadija, Bernstein Mallory, Giandhari J, Pillay S, Yajna Ramphal, Naidoo Yeshnee, Tshabuila Derek, Emmanuel S, Tegally H, Wilkinson E, de Oliveira T |
| hCoV-19/Slovenia/177001/2020 | EPI_ISL_3039100 | 2020-11-01 | Institute of Microbiology and Immunology, Faculty of Medicine, University of Ljubljana | Institute of Microbiology and Immunology, Faculty of Medicine, University of Ljubljana | Alen Suljiįç, Samo Zakotnik, Toma≈æ Mark Zorec, Matic Brvar, Doroteja Vlaj, Andra≈æ Celar, Dominika ≈†turm, Patricija Pozvek, ≈†pela Pleh, Mi≈°a Korva, Mario Poljak, Tatjana Av≈°iįç ,Äì ≈Ωupanc |
| hCoV-19/USA/CA-CDPH-500001014/2021 | EPI_ISL_3722998 | 2021-07-07 | TPMG Regional Laboratory | California Department of Public Health | Emily Smith on behalf of CDPH-COVIDNet and UCI Genome Sciences Center/GHTF |

| | | | | | |
|---|---|---|---|---|---|
| hCoV-19/Australia/QLD2224/2021 | EPI_ISL_3614817 | 2021-08-11 | PHV-FSS | PHV-FSS | Chenwei Wang on behalf of Q-PHIRE Genomics |
| hCoV-19/Bonaire/BQ-RIVM-50292/2021 | EPI_ISL_3731686 | 2021-07-31 | Dutch COVID-19 response team | National Institute for Public Health and the Environment (RIVM) | Adam Meijer, Harry Vennema, Dirk Eggink, Jeroen Cremer, Sharon van den Brink, Bas van der Veer, AnneMarie van den Brandt, Lisa Wijsman, Kim Freriks, Ryanne Jaarsma, Eunv≠ce Then, Lynn Aarts, Sanne Bos, Stijn van Rossum, Florian Zwagemaker, Dennis Schmitz, |
| hCoV-19/Netherlands/UT-RIVM-50526/2021 | EPI_ISL_3731897 | 2021-07-29 | Dutch COVID-19 response team | National Institute for Public Health and the Environment (RIVM) | Adam Meijer, Harry Vennema, Dirk Eggink, Jeroen Cremer, Sharon van den Brink, Bas van der Veer, AnneMarie van den Brandt, Lisa Wijsman, Kim Freriks, Ryanne Jaarsma, Eunv≠ce Then, Lynn Aarts, Sanne Bos, Stijn van Rossum, Florian Zwagemaker, Dennis Schmitz, |
| hCoV-19/Aruba/AW-RIVM-51730/2021 | EPI_ISL_3732903 | 2021-08-02 | Dutch COVID-19 response team | National Institute for Public Health and the Environment (RIVM) | Adam Meijer, Harry Vennema, Dirk Eggink, Jeroen Cremer, Sharon van den Brink, Bas van der Veer, AnneMarie van den Brandt, Lisa Wijsman, Kim Freriks, Ryanne Jaarsma, Eunv≠ce Then, Lynn Aarts, Sanne Bos, Stijn van Rossum, Florian Zwagemaker, Dennis Schmitz, |
| hCoV-19/Italy/ABR-IZSGC-301692/2021 | EPI_ISL_3734067 | 2021-08-16 | PO AVEZZANO - U.O.C. Laboratorio Analisi | Istituto Zooprofilattico Sperimentale dell‚ÄôAbruzzo e Molise ‚ÄúG. Caporale‚Äù | Lorusso A, Marcacci M, Di Domenico M, Ancora M, Curini V, Di Lollo Valeria, Mangone I, Rinaldi A, Delli Compagni E, Scialabba S, Caporale M, Di Pasquale A, Cammv† C, Puglia I, Calistri P, Savini G |
| hCoV-19/Switzerland/GE-HUG-35061439/2021 | EPI_ISL_3741100 | 2021-08-16 | University Hospitals of Geneva, Laboratory of Virology | HUG, Laboratory of Virology and the Health2030 Genome Center | Samuel Cordey, Aline Mamin, Francisco Perez, Ana Rita Goncalves, Laurent Kaiser, Lorenzo Cerutti, Cedric Howald, Henri Pegeot, Melyssa Elies, Deborah Penet, Keith Harshman, Ioannis Xenarios |
| hCoV-19/USA/UT-UPHL-210710486066-6-24/2021 | EPI_ISL_3756317 | 2021-06-24 | Utah Public Health Laboratory | Utah Public Health Laboratory | Erin L. Young, Kelly F. Oakeson, Tara Gallagher |
| hCoV-19/Sweden/SUS0003802/2021 | EPI_ISL_3761410 | 2021-08-19 | Clinical Microbiology, Infection Prevention and Control | Section for Molecular Diagnostics | Bjv∂rn Hallstrv∂m, Jonas Bjv∂rkman |
| hCoV-19/Brazil/SP-SJRP50810/2021 | EPI_ISL_3761572 | 2021-07-12 | Laboratorio de Pesquisa em Virologia, FAMERP, SJRP | Laboratorio de Pesquisa em Virologia, FAMERP, SJRP | Cecv≠lia Artico Banho; Beatriz de Carvalho Marques; Lv≠via Sacchetto; Marv≠lia Moraes; Guilherme Campos; Fv°bio Sossai Possebon; Leila Sabrina Ullmann; Cv≠ntia Bittar; Helena Lage Ferreira; Jorge A. Petroli Marchesi; Maisa C. Pereira Parra; Paula Rahal; |
| hCoV-19/Bangladesh/icddrb-GSB14-010/2021 | EPI_ISL_3762673 | 2021-07-18 | Shaheed Ziaur Rahman Medical College, Bogra | Virology Laboratory, International Centre for Diarrhoeal Disease Research, Bangladesh (ICDDR,B) | Mojnu Miah, Md. Mahfuzur Rahman, Mohsina Alam Shaheed, Manjur Hossain Khan, Senjuti Saha, Hassan Afrad, Mohammed Ziaur Rahman, Mohammad Jubair, Mohammad Enayet Hossain, Razib Mazumder, Mohammad Shahidul Islam, Ahmed Nawsher Alam, ASM Alamgir, Mahbubur Rah |
| hCoV-19/DominicanRepublic/Yale-5362/2021 | EPI_ISL_3105537 | 2021-01-22 | IMTSAG | Grubaugh Lab - Yale School of Public Health | Joseph Fauver, Mallery Breban, Isabell Ott, Tara Alpert, Mary Petrone, Anderson Brito, Chantal Vogels, Annie Watkins, Chaney Kalinich, Jessica Rothman, Robert Paulino-Ramirez, Alejandaro Vallejo Degaudenzi, Victor Virgilio Calderon, Elisa Contreras, Esper |

| | | | | | |
|---|---|---|---|---|---|
| hCoV-19/Mexico/TAB-InDRE_FB27536_S5864/2021 | EPI_ISL_3769238 | 2021-07-20 | LESP Tabasco | Instituto de Diagnostico y Referencia Epidemiologicos (INDRE) | Claudia Wong-Arambula, Abril Rodriguez-Maldonado, Vanessa Rivero-Arredondo, Ariadna Medina-Benitez, Joaquin Quiroz-Mercado, Sergio Rangel-Guerrero, Maribel Gonzalez-Villa, Natividad Cruz-Ortiz, Tatiana Nunez-Garcia, Gisela Barrera-Badillo, Lucia Hernandez |
| hCoV-19/Japan/IC-1789/2021 | EPI_ISL_3769589 | 2021-08-11 | SARS-CoV-2 testing team, National Institute of Infectious Diseases | Pathogen Genomics Center, National Institute of Infectious Diseases | Tsuyoshi Sekizuka, Kentaro Itokawa, Rina Tanaka, Masanori Hashino, Nozomu Hanaoka, Masumichi Saito, Naomi Nojiri, Hazuka Y Furihata, Tsuguto Fujimoto, Makoto Kuroda |
| hCoV-19/Malawi/MLW-00047/2021 | EPI_ISL_3770667 | 2021-07-12 | Malawi Liverpool Wellcome Trust Clinical Research Program | Malawi Liverpool Wellcome Trust CLinical Research Program | Catherine Anscombe, Philip Ashton, Ben Morton, Sam Lissauer, Kondwani Jambo |
| hCoV-19/USA/CA-CDC-FG-093509/2021 | EPI_ISL_4203209 | 2021-08-19 | Fulgent Genetics | Centers for Disease Control and Prevention Division of Viral Diseases, Pathogen Discovery | Dakota Howard,Dhwani Batra,Peter Cook,Jason Caravas,Benjamin Rambo-Martin,Scott Sammons,Yvette Unoarumhi,Matthew Schmerer,Kristine Lacek,Tymeckia Kendall,Victoria Caban Figueroa,Shatavia Morrison,Christopher Gulvick,Erisa Sula,Harry Gao,Mickey Li,John Gao |
| hCoV-19/USA/CA-CDC-FG-093459/2021 | EPI_ISL_4203267 | 2021-08-19 | Fulgent Genetics | Centers for Disease Control and Prevention Division of Viral Diseases, Pathogen Discovery | Dakota Howard,Dhwani Batra,Peter Cook,Jason Caravas,Benjamin Rambo-Martin,Scott Sammons,Yvette Unoarumhi,Matthew Schmerer,Kristine Lacek,Tymeckia Kendall,Victoria Caban Figueroa,Shatavia Morrison,Christopher Gulvick,Erisa Sula,Harry Gao,Mickey Li,John Gao |
| hCoV-19/England/NORT-1BF701D/2021 | EPI_ISL_3772084 | 2021-07-29 | Northumbria University / South Tees Hospitals NHS Foundation Trust / North Cumbria Integrated Care NHS Foundation Trust / North Tees and Hartlepool NHS Foundation Trust / Newcastle Hospitals NHS Foundation Trust | COVID-19 Genomics UK (COG-UK) Consortium | Darren L Smith,Andrew Nelson,Matthew Bashton,Greg R Young,Joshua Loh,John Allan,Mohammad A Tariq,Giles S Holt,Gary Black,Wen C Yew,Lynn Dover,Paul Baker,Steve Liggett,Sarah Essex,Jane Greenaway,Debra Padgett,Clive Graham,Garren Scott,Edward Barton,Emma Sw |
| hCoV-19/England/PHEC-U303UA0C/2021 | EPI_ISL_3776267 | 2021-03-21 | Respiratory Virus Unit, Microbiology Services Colindale, Public Health England | COVID-19 Genomics UK (COG-UK) Consortium | PHE Covid Sequencing Team |
| hCoV-19/England/PHEC-U303UC9D/2021 | EPI_ISL_3776302 | 2021-03-23 | Respiratory Virus Unit, Microbiology Services Colindale, Public Health England | COVID-19 Genomics UK (COG-UK) Consortium | PHE Covid Sequencing Team |
| hCoV-19/England/PHEC-U307U7F2/2021 | EPI_ISL_3776757 | 2021-03-25 | Respiratory Virus Unit, Microbiology Services Colindale, Public Health England | COVID-19 Genomics UK (COG-UK) Consortium | PHE Covid Sequencing Team |
| hCoV-19/England/PHEC-U30AU540/2021 | EPI_ISL_3777132 | 2021-04-20 | Respiratory Virus Unit, Microbiology Services Colindale, Public Health England | COVID-19 Genomics UK (COG-UK) Consortium | PHE Covid Sequencing Team |
| hCoV-19/England/PHEP-004089/2021 | EPI_ISL_3779148 | 2021-07-08 | Respiratory Virus Unit, Microbiology Services Colindale, Public Health England | COVID-19 Genomics UK (COG-UK) Consortium | PHE Covid Sequencing Team |
| hCoV-19/SouthAfrica/KRISP-K021777/2021 | EPI_ISL_3799042 | 2021-08-02 | National Health Laboratory Service, South Africa | KRISP, KZn Research Innovation and Sequencing Platform | Giandhari J, Pillay S, Yajna Ramphal, Naidoo Yeshnee, Tshabuila Derek, Emmanuel S, Tegally H, Wilkinson E, de Oliveira T |

| | | | | | |
|---|---|---|---|---|---|
| hCoV-19/USA/TN-CDC-4198495-001/2021 | EPI_ISL_3800424 | 2021-06-24 | TN Division of Laboratory Services | Centers for Disease Control and Prevention Division of Viral Diseases, Pathogen Discovery | Mili Sheth,Sarah Nobles,Jasmine Padilla,Alex Burgin,Meghan Bentz,Mark Burroughs,Peter Cook,Clinton Paden,Dhwani Batra,Krista Queen,Kristen Knipe,Dakota Howard,Yvette Unoarumhi,Matthew Schmerer,Ben Rambo-Martin,Kristine Lacek,Sam Shepard,Dave Wentworth,Viv |
| hCoV-19/USA/WI-WSLH-215750/2021 | EPI_ISL_3131937 | 2021-06-17 | Wisconsin State Laboratory of Hygiene Communicable Disease Division | Wisconsin State Laboratory of Hygiene Communicable Disease Division | Abigail C. Shockey, Alicia J. Mooney, Erika M. Hanson, Tonya Danz, Richard Griesser, Sara Wagner, Kelsey R. Florek |
| hCoV-19/Brazil/RJ-FIOCRUZ-38803/2021 | EPI_ISL_3801837 | 2021-07-14 | Laboratorio Central de Saude Publica do Estado do Rio de Janeiro (LACEN/RJ) | Laboratory of Respiratory Viruses and Measles, Oswaldo Cruz Institute, FIOCRUZ | Paola Resende, Luciana Appolinario, Fernando Motta, Anna Carolina Paixao, Ana Carolina Mendonca, Alice Sampaio Rocha, Taina Venas, Elisa Cavalcante Pereira, Renata Serrano Lopes, Agatha Cristinne Prudencio Soares, Ighor Leonardo Arantes Gomes, Andrea Cony |
| hCoV-19/Brazil/PR-FIOCRUZ-43888/2021 | EPI_ISL_3802979 | 2021-08-04 | Laboratorio Central de Saude Publica do Estado do Parana (LACEN/PR) | Laboratory of Respiratory Viruses and Measles, Oswaldo Cruz Institute, FIOCRUZ | Paola Resende, Luciana Appolinario, Fernando Motta, Anna Carolina Paixao, Ighor Leonardo Arantes, Agatha Soares, Ana Carolina Mendonca, Alice Sampaio Rocha, Taina Venas, Elisa Cavalcante Pereira, Renata Serrano Lopes, Irina Riediger, Marilda Siqueira on b |
| hCoV-19/Brazil/PR-FIOCRUZ-43802/2021 | EPI_ISL_3802986 | 2021-07-29 | Laboratorio Central de Saude Publica do Estado do Parana (LACEN/PR) | Laboratory of Respiratory Viruses and Measles, Oswaldo Cruz Institute, FIOCRUZ | Paola Resende, Luciana Appolinario, Fernando Motta, Anna Carolina Paixao, Ighor Leonardo Arantes, Agatha Soares, Ana Carolina Mendonca, Alice Sampaio Rocha, Taina Venas, Elisa Cavalcante Pereira, Renata Serrano Lopes, Irina Riediger, Marilda Siqueira on b |
| hCoV-19/Brazil/PR-FIOCRUZ-43881/2021 | EPI_ISL_3802991 | 2021-08-04 | Laboratorio Central de Saude Publica do Estado do Parana (LACEN/PR) | Laboratory of Respiratory Viruses and Measles, Oswaldo Cruz Institute, FIOCRUZ | Paola Resende, Luciana Appolinario, Fernando Motta, Anna Carolina Paixao, Ighor Leonardo Arantes, Agatha Soares, Ana Carolina Mendonca, Alice Sampaio Rocha, Taina Venas, Elisa Cavalcante Pereira, Renata Serrano Lopes, Irina Riediger, Marilda Siqueira on b |
| hCoV-19/NewZealand/21MV0731/2021 | EPI_ISL_3804548 | 2021-08-17 | LabPLUS | Institute of Environmental Science and Research (ESR) | Rachel Boyle, SallyAnn Harbison, Olivia Stroeven, Xiaoyun Ren, Matt Storey, Nikki Freed, Muhammad Faisal, Jing Wang, Hermes Perez, Anja Werno, Antje van der Linden, Arlo Upton, Chris Mansell, David Hammer, Dragana Drinkovic, Gary McAuliffe, Hana Sofia And |
| hCoV-19/USA/TN-SPHL-0261/2021 | EPI_ISL_3153315 | 2021-02-24 | TN DOH Lab Services | TN DOH Lab Services | Karen Beasley-Maynard, Brian Selinsky |
| hCoV-19/Australia/NSW-ICPMR-7007/2021 | EPI_ISL_4205293 | 2021-09-03 | Pathology West - NSW Health Pathology | Clinical Pathology and Medical Research; Westmead Hospital; University of Sydney | Arnott A., Draper J., Gall M., Martinez E., Rockett R., Sintchenko V., on behalf of ICPMR |
| hCoV-19/USA/FL-CDC-ASC210206322/2021 | EPI_ISL_3819707 | 2021-08-01 | Aegis Sciences Corporation | Centers for Disease Control and Prevention Division of Viral Diseases, Pathogen Discovery | Dakota Howard,Dhwani Batra,Peter Cook,Kara Moser,Adrian Paskey,Jason Caravas,Benjamin Rambo-Martin,Shatavia Morrison,Christopher Gulvick,Scott Sammons,Yvette Unoarumhi,Darlene Wagner,Matthew Schmerer,Cyndi Clark,Patrick Campbell,Rob Case,Vikramsinha Ghorp |

| | | | | | |
|---|---|---|---|---|---|
| hCoV-19/Brazil/SP-BT50788/2021 | EPI_ISL_3824310 | 2021-08-13 | Instituto de Biotecnologia - UNESP-Botucatu-SP | Instituto de Biotecnologia - UNESP-Botucatu-SP | F√°bio Sossai Possebon; Leila Sabrina Ullmann; Cec√≠lia Artico Banho; C√≠ntia Bittar; Guilherme Campos; Helena Lage Ferreira; Jorge A. Petroli Marchesi; L√≠via Sacchetto; Maisa C. Pereira Parra; Mar√≠lia Moraes; Maur√≠cio L. Nogueira; Paula Rahal; Paulo I |
| hCoV-19/Brazil/SP-BT2021081102/2021 | EPI_ISL_3824314 | 2021-08-10 | Instituto de Biotecnologia - UNESP-Botucatu-SP | Instituto de Biotecnologia - UNESP-Botucatu-SP | F√°bio Sossai Possebon; Leila Sabrina Ullmann; Cec√≠lia Artico Banho; C√≠ntia Bittar; Guilherme Campos; Helena Lage Ferreira; Jorge A. Petroli Marchesi; L√≠via Sacchetto; Maisa C. Pereira Parra; Mar√≠lia Moraes; Maur√≠cio L. Nogueira; Paula Rahal; Paulo I |
| hCoV-19/Ecuador/USFQ-2033/2021 | EPI_ISL_3824603 | 2021-07-30 | Institute of Microbiology, Universidad San Francisco de Quito | Institute of Microbiology, Universidad San Francisco de Quito | Bel√©n Prado-Vivar, Erika B. Mu√±oz, Fernanda Zurita, Mateo Carvajal, Monica Becerra-Wong, Juan Jos√© Guadalupe, Sully M√°rquez, Bernardo Guti√©rrez, Stephanie Esp√≠n-Arroba, Ronny Javier Pibaque, Liu Yuqian, Marco Viteri Yanez, Valeria Armijos, Luis Fuen |
| hCoV-19/USA/NV-NSPHL-392347/2021 | EPI_ISL_3825081 | 2021-02-03 | Nevada State Public Health Laboratory | Nevada State Public Health Laboratory | Andrew Gorzalski, Mark Pandori |
| hCoV-19/SouthAfrica/CERI-KRISP-Tyg1861/2021 | EPI_ISL_3827563 | 2021-07-29 | Division of Medical Virology, National Health Laboratory Service (NHLS), Tygerberg Hospital / Stellenbosch University | CERI, Centre for Epidemic Response and Innovation, Stellenbosch University and CERI-KRISP, KZN Research Innovation and Sequencing Platform | Susan Engelbrecht,Tongai Maponga,Wolfgang Preiser,Gert van Zyl,Shannon Wilson,Karabo Phadu,Bronwyn Kleinhans,Kamela Mahlakwane,Mathilda Claassen,Tania Stander,Carel J van Heerden,Alvera Vorster,Ren Veikondis, Giandhari Jennifer, Pillay Sureshnee, Yajna Ra |
| hCoV-19/SouthAfrica/CERI-KRISP-Tyg1745/2021 | EPI_ISL_3827620 | 2021-07-28 | Division of Medical Virology, National Health Laboratory Service (NHLS), Tygerberg Hospital / Stellenbosch University | CERI, Centre for Epidemic Response and Innovation, Stellenbosch University and CERI-KRISP, KZN Research Innovation and Sequencing Platform | Susan Engelbrecht,Tongai Maponga,Wolfgang Preiser,Gert van Zyl,Shannon Wilson,Karabo Phadu,Bronwyn Kleinhans,Kamela Mahlakwane,Mathilda Claassen,Tania Stander,Carel J van Heerden,Alvera Vorster,Ren Veikondis, Giandhari Jennifer, Pillay Sureshnee, Yajna Ra |
| hCoV-19/SouthAfrica/NICD-N9210/2021 | EPI_ISL_2827099 | 2021-06-06 | HELEN JOSEPH LABORATORY | Diseases of the National Health Laboratory Service | Amoako DG, Everatt J, Scheepers C, Mohale T, Ntuli N, Mahlangu B, Mnguni A, Ismail A, Bhiman JN |
| hCoV-19/USA/NY-PRL-2021_0701_51D19/2020 | EPI_ISL_2827578 | 2020-10-09 | Pandemic Response Lab - NYC | Pandemic Response Lab, R&D | Henry Lee, Michael Hammerling, Melissa Hopkins, Cybill del Castillo, Shinyoung Clair Kang, William Ward, Pradeep Bugga, Sol Rey, Dylan Law, Isabel Fernandez Escapa, Haiping Hao, Jon Laurent |
| hCoV-19/Guatemala/GUA-LNS-48495/2020 | EPI_ISL_2827836 | 2020-10-08 | Laboratorio Nacional de Salud | Laboratorio Nacional de Salud | Linda Mendoza, Gabriela Garc√≠a |
| hCoV-19/England/PLYM-180EA63/2021 | EPI_ISL_2828787 | 2021-06-30 | Lighthouse Laboratory Plymouth | Wellcome Sanger Institute for the COVID-19 Genomics UK (COG-UK) Consortium | Lighthouse Laboratory Plymouth and Alex Alderton, Roberto Amato, Jeffrey Barrett, Sonia Goncalves, Ewan Harrison, David K. Jackson, Ian Johnston, Dominic Kwiatkowski, Cordelia Langford, John Sillitoe on behalf of the Wellcome Sanger Institute COVID-19 Sur |

| | | | | | |
|---|---|---|---|---|---|
| hCoV-19/Spain/CT-HUGTiPM024WK4E1/2021 | EPI_ISL_2832182 | 2021-06-25 | Microbiology Department, Laboratori Cl√≠nic Metropolitana Nord. Hospital Universitari Germans Trias i Pujol | Can Ruti SARS-CoV-2 Sequencing Hub (HUGTiP/IrsiCaixa/IGTP) | Marc Noguera-Julian, Pilar Armengol, Ignacio Blanco, Antoni E Bordoy, Francesc Catala-Moll, Pere-Joan Cardona, Maria Casadell√†, Cristina Casa√±, Gemma Clara, Bonaventura Clotet, Montserrat Gim√©nez, Mercedes Guerrero, Anna Not, David Panisello, Roger Par |
| hCoV-19/Belgium/rega-7599/2021 | EPI_ISL_2832259 | 2021-04-22 | KU Leuven, Rega Institute, Clinical and Epidemiological Virology | KU Leuven, Rega Institute, Clinical and Epidemiological Virology | Tony Wawina-Bokalanga, Bert Vanmechelen, Joan Marti-Carerras, Piet Maes |
| hCoV-19/Belgium/rega-7861/2021 | EPI_ISL_2832606 | 2021-04-26 | KU Leuven, Rega Institute, Clinical and Epidemiological Virology | KU Leuven, Rega Institute, Clinical and Epidemiological Virology | Tony Wawina-Bokalanga, Bert Vanmechelen, Joan Marti-Carerras, Piet Maes |
| hCoV-19/Belgium/rega-7877/2021 | EPI_ISL_2832622 | 2021-04-27 | KU Leuven, Rega Institute, Clinical and Epidemiological Virology | KU Leuven, Rega Institute, Clinical and Epidemiological Virology | Tony Wawina-Bokalanga, Bert Vanmechelen, Joan Marti-Carerras, Piet Maes |
| hCoV-19/Israel/SMC-7003867/2021 | EPI_ISL_2834411 | 2021-06-20 | Shamir Medical Center (Asaf Harofe) | Shamir Medical Center (Asaf Harofe) | Nir Rainy,Chen Weiner,Reut Sorek Abramovich,Yevgeni Yegorov,Anna Vishnevsky,Patricia Benveniste-Lekovitz,Abu Hamad Ramzia,Adina Bar Chaim |
| hCoV-19/Colombia/CES-CGBM-S24082021-05/2021 | EPI_ISL_4207069 | 2021-08-04 | Laboratorio Cl√≠nico Nancy Florez Garc√≠a | Centro de Gen√©tica y Biolog√≠a Molecular - Universidad del Magdalena | Maria Teresa Mojica-Ortiz, Daniel Bautista, Andrea M. Ram√≠rez Hernandez, Edison Lea-Ch, Angel Oviedo Marquez, Lyda R. Castro |
| hCoV-19/Australia/NSW-ICPMR-5214/2021 | EPI_ISL_3833827 | 2021-08-21 | Pathology West - NSW Health Pathology | Clinical Pathology and Medical Research; Westmead Hospital; University of Sydney | Arnott A., Draper J., Gall M., Martinez E., Rockett R., Sintchenko V., on behalf of ICPMR |
| hCoV-19/FrenchGuiana/IPG2021564/2021 | EPI_ISL_2835991 | 2021-06-09 | Laboratoire Carage | Institut Pasteur de la Guyane | Anne Lavergne, Dominique Rousset, Antoine Enfissi, Arielle Salmier |
| hCoV-19/FrenchGuiana/IPG2021565/2021 | EPI_ISL_2835992 | 2021-06-18 | Eurofins Montjoly | Institut Pasteur de la Guyane | Anne Lavergne, Dominique Rousset, Antoine Enfissi, Arielle Salmier |
| hCoV-19/Australia/SA0782/2021 | EPI_ISL_2839992 | 2021-06-28 | SA Pathology | SA Pathology | Caitlin Selway, Lex Leong, Chuan Kok Lim, Mark Turra, Ivan Bastian, Geoff Higgins |
| hCoV-19/Australia/SA0798/2021 | EPI_ISL_2839997 | 2021-07-04 | SA Pathology | SA Pathology | Caitlin Selway, Lex Leong, Chuan Kok Lim, Mark Turra, Ivan Bastian, Geoff Higgins |
| hCoV-19/Israel/SMC-7012570/2021 | EPI_ISL_3834697 | 2021-08-06 | Shamir Medical Center (Asaf Harofe) | Shamir Medical Center (Asaf Harofe) | Nir Rainy,Chen Weiner,Reut Sorek Abramovich,Yevgeni Yegorov,Anna Vishnevsky,Patricia Benveniste-Lekovitz,Abu Hamad Ramzia,Adina Bar Chaim |
| hCoV-19/Israel/SMC-7012925/2021 | EPI_ISL_3834758 | 2021-08-07 | Shamir Medical Center (Asaf Harofe) | Shamir Medical Center (Asaf Harofe) | Nir Rainy,Chen Weiner,Reut Sorek Abramovich,Yevgeni Yegorov,Anna Vishnevsky,Patricia Benveniste-Lekovitz,Abu Hamad Ramzia,Adina Bar Chaim |
| hCoV-19/Israel/SMC-7012673/2021 | EPI_ISL_3834830 | 2021-08-05 | Shamir Medical Center (Asaf Harofe) | Shamir Medical Center (Asaf Harofe) | Nir Rainy,Chen Weiner,Reut Sorek Abramovich,Yevgeni Yegorov,Anna Vishnevsky,Patricia Benveniste-Lekovitz,Abu Hamad Ramzia,Adina Bar Chaim |
| hCoV-19/Israel/SMC-7012661/2021 | EPI_ISL_3834927 | 2021-08-05 | Shamir Medical Center (Asaf Harofe) | Shamir Medical Center (Asaf Harofe) | Nir Rainy,Chen Weiner,Reut Sorek Abramovich,Yevgeni Yegorov,Anna Vishnevsky,Patricia Benveniste-Lekovitz,Abu Hamad Ramzia,Adina Bar Chaim |
| hCoV-19/SouthAfrica/UFS-K018909-KRISP/2021 | EPI_ISL_2841638 | 2021-05-31 | UFS Virology | KRISP, KZn Research Innovation and Sequencing Platform | Giandhari Jennifer, Pillay Sureshnee, Yajna Ramphal, Naidoo Yeshnee, Tshabuila Derek,Tegally Houriiyah, San James, Wilkinson Eduan, de Oliveira Tulio |

| | | | | | |
|---|---|---|---|---|---|
| hCoV-19/India/KA-NIMH-SEQ-1844/2021 | EPI_ISL_2842358 | 2021-05-28 | BBMP Urban | INSACOG-KA, NIMHANS | Chitra Pattabiraman, Anson Kunjumon George,Darshan Sreenivas, Ellango Ramasamy, Sony Sharma, Mahesh Kumar.C.S, Ananthapadmanabha Kotambail, Chetan G K, Gautham Arunachal Udupi, Anita S Desai, V Ravi |
| hCoV-19/India/KA-NIMH-SEQ-1077/2021 | EPI_ISL_2842441 | 2021-05-25 | RAILWAY HOSPITAL | INSACOG-KA, NIMHANS | Chitra Pattabiraman, Anson Kunjumon George,Darshan Sreenivas, Ellango Ramasamy, Sony Sharma, Mahesh Kumar.C.S, Ananthapadmanabha Kotambail, Chetan G K, Gautham Arunachal Udupi, Anita S Desai, V Ravi |
| hCoV-19/India/KA-NIMH-SEQ-1130/2021 | EPI_ISL_2842481 | 2021-05-15 | DSU KALABURAGI | INSACOG-KA, NIMHANS | Chitra Pattabiraman, Anson Kunjumon George,Darshan Sreenivas, Ellango Ramasamy, Sony Sharma, Mahesh Kumar.C.S, Ananthapadmanabha Kotambail, Chetan G K, Gautham Arunachal Udupi, Anita S Desai, V Ravi |
| hCoV-19/India/KA-NIMH-SEQ-1153/2021 | EPI_ISL_2842498 | 2021-05-13 | DSU HASSAN | INSACOG-KA, NIMHANS | Chitra Pattabiraman, Anson Kunjumon George,Darshan Sreenivas, Ellango Ramasamy, Sony Sharma, Mahesh Kumar.C.S, Ananthapadmanabha Kotambail, Chetan G K, Gautham Arunachal Udupi, Anita S Desai, V Ravi |
| hCoV-19/Colombia/ANT-CIB-07-18/2021 | EPI_ISL_4207311 | 2021-08-26 | Laboratorio Las Am√©ricas | Corporaci√≥n para Investigaciones Biol√≥gicas-CIB | Uriel A. Hurtado Paez, Katterine Molina Hoyos, Jeanneth Mosquera Rendon, Marcela Mercado Reyes |
| hCoV-19/USA/CO-CDPHE-2101445393/2021 | EPI_ISL_4210453 | 2021-07-18 | Colorado Department of Public Health and Environment | Colorado Department of Public Health and Environment | Laura Bankers, Molly C. Hetherington-Rauth, Diana Ir, Alexandria Rossheim, Michael A. Martin, Mandy Waters, Shannon R. Matzinger, Sarah Elizabeth Totten, Emily A. Travanty |
| hCoV-19/Poland/8567962/2021 | EPI_ISL_3838167 | 2021-08-18 | Medyczne Laboratoria Diagnostyczne INVICTA | Wojew√≥dzka Stacja Sanitarno-Epidemiologiczna w Katowicach | Beata Rozwadowska |
| hCoV-19/SouthAfrica/NICD-N11767/2021 | EPI_ISL_3838323 | 2021-07-12 | EAST LONDON LABORATORY | Diseases of the National Health Laboratory Service | Amoako DG, Everatt J, Scheepers C, Mohale T, Ntuli N, Mahlangu B, Mnguni A, Ismail A, Bhiman JN |
| hCoV-19/Denmark/DCGC-123279/2021 | EPI_ISL_2854279 | 2021-06-30 | Department of Bacteria, Parasites and Fungi, Statens Serum Institut, Copenhagen, Denmark | Statens Serum Institut Bioinformatics and Microbial Genomics | Danish Covid-19 Genome Consortium |
| hCoV-19/USA/NV-CDC-FG-119907/2021 | EPI_ISL_4359286 | 2021-09-07 | Fulgent Genetics | Centers for Disease Control and Prevention Division of Viral Diseases, Pathogen Discovery | Dakota Howard,Dhwani Batra,Peter Cook,Jason Caravas,Benjamin Rambo-Martin,Scott Sammons,Yvette Unoarumhi,Matthew Schmerer,Kristine Lacek,Tymeckia Kendall,Victoria Caban Figueroa,Shatavia Morrison,Christopher Gulvick,Erisa Sula,Harry Gao,Mickey Li,John Gao |
| hCoV-19/USA/UT-UPHL-210617691764/2021 | EPI_ISL_2856332 | 2021-06-02 | Utah Public Health Laboratory | Utah Public Health Laboratory | Olinto Linares-Perdomo, Erin L. Young, Kelly F. Oakeson, Tara Gallagher |
| hCoV-19/Philippines/PH-PGC-45422/2021 | EPI_ISL_2859175 | 2021-04-28 | Zamboanga City Medical Center-Department of Agriculture Satellite Laboratory for COVID-19 Testing | Philippine Genome Center | Francis A. Tablizo, Kenneth M. Kim, Maria Sofia L. Yangzon, Elcid Aaron R. Pangilinan, Renato Jacinto Q. Mantaring, Benedict A. Maralit, Marc Edsel C. Ayes, Eva Maria Cutiongco-de la Paz, Alethea R. de Guzman, Jan Michael C. Yap, Jo-Hannah S. Llames, Shei |

| | | | | | |
|---|---|---|---|---|---|
| hCoV-19/Philippines/PH-PGC-34396/2021 | EPI_ISL_2860213 | 2021-04-20 | First Aide Diagnostic Center | Philippine Genome Center | Francis A. Tablizo, Kenneth M. Kim, Maria Sofia L. Yangzon, Elcid Aaron R. Pangilinan, Renato Jacinto Q. Mantaring, Benedict A. Maralit, Marc Edsel C. Ayes, Eva Maria Cutiongco-de la Paz, Alethea R. de Guzman, Jan Michael C. Yap, Jo-Hannah S. Llames, Shei |
| hCoV-19/USA/MD-MDH-4064/2021 | EPI_ISL_3841528 | 2021-07-24 | MD PHL | Maryland Department of Health Laboratories Administration | Eric N. Keller, Jillian Loomis, Kwang Low, Terence L. Moore, Ami Patel, and Robert Myers |
| hCoV-19/Netherlands/NB-RIVM-41920/2021 | EPI_ISL_2863244 | 2021-06-17 | Dutch COVID-19 response team | National Institute for Public Health and the Environment (RIVM) | Adam Meijer, Harry Vennema, Dirk Eggink, Jeroen Cremer, Sharon van den Brink, Bas van der Veer, AnneMarie van den Brandt, Lisa Wijsman, Kim Freriks, Ryanne Jaarsma, Eun√≠ce Then, Lynn Aarts, Sanne Bos, Melissa van Tuil, Florian Zwagemaker, Dennis Schmitz, |
| hCoV-19/Norway/Ahus-877/2021 | EPI_ISL_2868453 | 2021-06-27 | Dept. of Microbiology and Infection Control, Akershus University Hospital HF | Control, Akershus University Hospital HF | Hege Vangstein Aamot, Alexander Hesselberg L√∏vestad |
| hCoV-19/India/GJ-INSACOG-GBRC1971/2021 | EPI_ISL_3844046 | 2021-04-30 | Smimer Medical College, Surat | Gujarat Biotechnology Research Centre | Arpit Shukla,Bhadreshsinh Gohil,Nimesh Patel,Nitin Shukla,Ramesh Pandit,Janvi Raval,Zarna Patel,Dinesh Kumar,Manish Patel,Chaitanya Joshi,Madhvi Joshi |
| hCoV-19/India/GJ-INSACOG-GBRC1978/2021 | EPI_ISL_3844147 | 2021-04-20 | Smimer Medical College, Surat | Gujarat Biotechnology Research Centre | Dinesh Kumar,Arpit Shukla,Bhadreshsinh Gohil,Nimesh Patel,Nitin Shukla,Ramesh Pandit,Janvi Raval,Zarna Patel,Manish Patel,Chaitanya Joshi,Madhvi Joshi |
| hCoV-19/USA/CA-CDC-QDX26118061/2021 | EPI_ISL_2869602 | 2021-06-22 | Quest Diagnostics Incorporated | Centers for Disease Control and Prevention Division of Viral Diseases, Pathogen Discovery | Dakota Howard, Dhwani Batra, Peter W. Cook, Kara Moser, Adrian Paskey, Jason Caravas, Benjamin Rambo-Martin, Shatavia Morrison, Christopher Gulvick, Scott Sammons, Yvette Unoarumhi, Darlene Wagner, Matthew Schmerer, S. H. Rosenthal, A. Gerasimova, R. M. K |
| hCoV-19/India/GJ-INSACOG-GBRC2077/2021 | EPI_ISL_3844231 | 2021-04-15 | Smimer Medical College, Surat | Gujarat Biotechnology Research Centre | Nimesh Patel,Nitin Shukla,Ramesh Pandit,Janvi Raval,Zarna Patel,Dinesh Kumar,Arpit Shukla,Bhadreshsinh Gohil,Manish Patel,Chaitanya Joshi,Madhvi Joshi |
| hCoV-19/India/GJ-INSACOG-GBRC1940/2021 | EPI_ISL_3844351 | 2021-04-11 | Smimer Medical College, Surat | Gujarat Biotechnology Research Centre | Bhadreshsinh Gohil,Nimesh Patel,Nitin Shukla,Ramesh Pandit,Janvi Raval,Zarna Patel,Dinesh Kumar,Arpit Shukla,Manish Patel,Chaitanya Joshi,Madhvi Joshi |
| hCoV-19/USA/FL-CDC-QDX25848320/2021 | EPI_ISL_2871075 | 2021-06-12 | Quest Diagnostics Incorporated | Centers for Disease Control and Prevention Division of Viral Diseases, Pathogen Discovery | Dakota Howard, Dhwani Batra, Peter W. Cook, Kara Moser, Adrian Paskey, Jason Caravas, Benjamin Rambo-Martin, Shatavia Morrison, Christopher Gulvick, Scott Sammons, Yvette Unoarumhi, Darlene Wagner, Matthew Schmerer, S. H. Rosenthal, A. Gerasimova, R. M. K |
| hCoV-19/USA/TX-CDC-QDX25890811/2021 | EPI_ISL_2871249 | 2021-06-13 | Quest Diagnostics Incorporated | Centers for Disease Control and Prevention Division of Viral Diseases, Pathogen Discovery | Dakota Howard, Dhwani Batra, Peter W. Cook, Kara Moser, Adrian Paskey, Jason Caravas, Benjamin Rambo-Martin, Shatavia Morrison, Christopher Gulvick, Scott Sammons, Yvette Unoarumhi, Darlene Wagner, Matthew Schmerer, S. H. Rosenthal, A. Gerasimova, R. M. K |

| | | | | | |
|---|---|---|---|---|---|
| hCoV-19/USA/FL-CDC-ASC210231121/2021 | EPI_ISL_3845167 | 2021-07-27 | Aegis Sciences Corporation | Centers for Disease Control and Prevention Division of Viral Diseases, Pathogen Discovery | Dakota Howard,Dhwani Batra,Peter Cook,Kara Moser,Adrian Paskey,Jason Caravas,Benjamin Rambo-Martin,Shatavia Morrison,Christopher Gulvick,Scott Sammons,Yvette Unoarumhi,Darlene Wagner,Matthew Schmerer,Cyndi Clark,Patrick Campbell,Rob Case,Vikramsinha Ghorp |
| hCoV-19/USA/TX-CDC-LC0079983/2021 | EPI_ISL_2873234 | 2021-06-17 | Laboratory Corporation of America | Centers for Disease Control and Prevention Division of Viral Diseases, Pathogen Discovery | Dakota Howard, Dhwani Batra, Peter W. Cook, Kara Moser, Adrian Paskey, Jason Caravas, Benjamin Rambo-Martin, Shatavia Morrison, Christopher Gulvick, Scott Sammons, Yvette Unoarumhi, Darlene Wagner, Matthew Schmerer, Minoo Agarwal, Eyad Almasri, Debbie Bol |
| hCoV-19/Ghana/nCoV-TRA-656/2021 | EPI_ISL_2873845 | 2021-04-06 | National Influenza Centre | National Influenza Centre | William K. Ampofo, Michael Marks, Ivy A. Asante, Sharon Hsu, Benjamiin B. Lindsey, Benjamin H. Foulkes, Mildred Adusei-Poku, Linda Boatemaa, Lorreta Kwasah, Joseph Oliver-Commey, Ernest Asiedu, Franklin Asiedu-Bekoe, Gordon Awandare, Joyce Ngoi, Dennis La |
| hCoV-19/USA/AK-CDC-ASC210232249/2021 | EPI_ISL_3845509 | 2021-07-26 | Aegis Sciences Corporation | Centers for Disease Control and Prevention Division of Viral Diseases, Pathogen Discovery | Dakota Howard,Dhwani Batra,Peter Cook,Kara Moser,Adrian Paskey,Jason Caravas,Benjamin Rambo-Martin,Shatavia Morrison,Christopher Gulvick,Scott Sammons,Yvette Unoarumhi,Darlene Wagner,Matthew Schmerer,Cyndi Clark,Patrick Campbell,Rob Case,Vikramsinha Ghorp |
| hCoV-19/India/TG-CDFD-W65/2021 | EPI_ISL_3243212 | 2021-06-11 | Gandhi Hospital | CDFD | Murali Bashyam, Asmita Gupta, Reelina Basu, Vinay Donipadi, Nagamani Kammili, Divya Vashisht, Ashwin Dalal |
| hCoV-19/USA/FL-QDX-25847663/2021 | EPI_ISL_2876622 | 2021-06-13 | Quest Diagnostics Incorporated | Centers for Disease Control and Prevention Division of Viral Diseases, Pathogen Discovery | Dakota Howard, Dhwani Batra, Peter W. Cook, Kara Moser, Adrian Paskey, Jason Caravas, Benjamin Rambo-Martin, Shatavia Morrison, Christopher Gulvick, Scott Sammons, Yvette Unoarumhi, Darlene Wagner, Matthew Schmerer, S. H. Rosenthal, A. Gerasimova, R. M. K |
| hCoV-19/USA/GA-CDC-QDX26156973/2021 | EPI_ISL_2876857 | 2021-06-21 | Quest Diagnostics Incorporated | Centers for Disease Control and Prevention Division of Viral Diseases, Pathogen Discovery | Dakota Howard, Dhwani Batra, Peter W. Cook, Kara Moser, Adrian Paskey, Jason Caravas, Benjamin Rambo-Martin, Shatavia Morrison, Christopher Gulvick, Scott Sammons, Yvette Unoarumhi, Darlene Wagner, Matthew Schmerer, S. H. Rosenthal, A. Gerasimova, R. M. K |
| hCoV-19/Belgium/rega-10606/2021 | EPI_ISL_2877543 | 2021-06-10 | KU Leuven, Rega Institute, Clinical and Epidemiological Virology | KU Leuven, Rega Institute, Clinical and Epidemiological Virology | Tony Wawina-Bokalanga, Bert Vanmechelen, Joan Marti-Carerras, Piet Maes |
| hCoV-19/India/AS-1829500220127/2021 | EPI_ISL_2877789 | 2021-05-20 | INSACOG-Assam | National Institute of Biomedical Genomics ‚Äì INSACOG | Arindam Maitra, Ajanta Sharma, Nidhan Kumar Biswas, Lahari Saikia, Sreedhar Chinnaswamy,¬†Kailash Chamuah, Saumitra Das |
| hCoV-19/India/AS-1829500220220/2021 | EPI_ISL_2877790 | 2021-05-20 | INSACOG-Assam | National Institute of Biomedical Genomics ‚Äì INSACOG | Arindam Maitra, Ajanta Sharma, Nidhan Kumar Biswas, Lahari Saikia, Sreedhar Chinnaswamy,¬†Kailash Chamuah, Saumitra Das |
| hCoV-19/India/HR-IGIB-60128/2021 | EPI_ISL_2878570 | 2021-02-01 | INSACOG Surveillance | INSACOG at CSIR Institute of Genomics and Integrative Biology | INSACOG |

| | | | | | |
|---|---|---|---|---|---|
| hCoV-19/India/HR-IGIB-60230/2021 | EPI_ISL_2878575 | 2021-02-01 | INSACOG Surveillance | INSACOG at CSIR Institute of Genomics and Integrative Biology | INSACOG |
| hCoV-19/India/HR-IGIB-60260/2021 | EPI_ISL_2878578 | 2021-02-01 | INSACOG Surveillance | INSACOG at CSIR Institute of Genomics and Integrative Biology | INSACOG |
| hCoV-19/India/MH-IGIB-GSEQ_2146/2021 | EPI_ISL_2879089 | 2021-03-27 | INSACOG Surveillance | INSACOG at CSIR Institute of Genomics and Integrative Biology | INSACOG |
| hCoV-19/India/MH-IGIB-GSEQ_2155/2021 | EPI_ISL_2879141 | 2021-03-30 | INSACOG Surveillance | INSACOG at CSIR Institute of Genomics and Integrative Biology | INSACOG |
| hCoV-19/Australia/QLD2108/2021 | EPI_ISL_3247191 | 2021-08-01 | Queensland Health Forensic and Scientific Services | Queensland Health Forensic and Scientific Services | Q-PHIRE Genomics |
| hCoV-19/India/HR-IGIB-606201188966/2021 | EPI_ISL_2880022 | 2021-04-28 | INSACOG Surveillance | INSACOG at CSIR Institute of Genomics and Integrative Biology | INSACOG |
| hCoV-19/India/RJ-IGIB-135573/2021 | EPI_ISL_2880127 | 2021-04-29 | INSACOG Surveillance | INSACOG at CSIR Institute of Genomics and Integrative Biology | INSACOG |
| hCoV-19/India/HR-IGIB-607500293501/2021 | EPI_ISL_2880277 | 2021-04-30 | INSACOG Surveillance | INSACOG at CSIR Institute of Genomics and Integrative Biology | INSACOG |
| hCoV-19/India/HR-IGIB-607500319758/2021 | EPI_ISL_2880354 | 2021-04-30 | INSACOG Surveillance | INSACOG at CSIR Institute of Genomics and Integrative Biology | INSACOG |
| hCoV-19/India/PB-IGIB-360900225127/2021 | EPI_ISL_2881358 | 2021-04-25 | INSACOG Surveillance | INSACOG at CSIR Institute of Genomics and Integrative Biology | INSACOG |
| hCoV-19/India/UP-IGIB-MEEN0039598504/2021 | EPI_ISL_2881530 | 2021-06-04 | INSACOG Surveillance | INSACOG at CSIR Institute of Genomics and Integrative Biology | INSACOG |
| hCoV-19/India/UP-IGIB-AURN0035421630/2021 | EPI_ISL_2881674 | 2021-05-16 | INSACOG Surveillance | INSACOG at CSIR Institute of Genomics and Integrative Biology | INSACOG |
| hCoV-19/USA/MD-HP06584-PIDSAYMIYT/2021 | EPI_ISL_2884908 | 2021-06-17 | Johns Hopkins Hospital Department of Pathology | Johns Hopkins Hospital Department of Pathology | C. Paul Morris, Chun Huai Luo, Adannaya Amadi, Matthew Schwartz, Nicholas Gallagher, Heba H. Mostafa |
| hCoV-19/USA/IL-S21WGS2276/2021 | EPI_ISL_2885413 | 2021-06-24 | Illinois Department of Public Health - Springfield Lab | Illinois Department of Public Health - Springfield Lab | Bryan Sim, Gordon McCall |
| hCoV-19/Senegal/SN-IRFSFOU-32/2021 | EPI_ISL_2887857 | 2021-04-02 | Iressef Genomics lab | IRESSEF | Souleymane MBOUP, Abdou PADANE , Khadim GUEYE, Diabou Diagne , Samba Ndiour, Papa Alassane DIAW, Birahim Piere NDIAYE, Barada CISSE, Aminata MBOUP, Moustapha MBOW, Ndeye Coumba Toure KANE, Nafisatou LEYE, Gora LO, Ambroise AHOUIDI , Astou Gaye GAYE, Amina |
| hCoV-19/Lebanon/LAU-CVD-27/2021 | EPI_ISL_2893216 | 2021-07-01 | Laboratory of Molecular Biology and Cancer Immunology, Faculty of Sciences, Lebanese University | Microbial Pathogenomics Lab - LAU | Sima tokajian, Georgi Merhi, Jad Koweyes, Bassam Badran, Fadi Abdel Sater, Rawan Makki, Hamad Hassan |
| hCoV-19/SouthAfrica/KRISP-K016797/2021 | EPI_ISL_2893248 | 2021-05-30 | NHLS_VIRO Eastern Cape | KRISP, KZN Research Innovation and Sequencing Platform | Laguda-Akingba O, Giandhari J, Pillay S, Lessells R, Mdlalose K, York D, Khan S, Emmanuel SJ, Tegally H, Wilkinson E, de Oliveira T |
| hCoV-19/SouthAfrica/KRISP-K019020/2021 | EPI_ISL_2893311 | 2021-06-21 | BARC - Sisonke | KRISP, KZN Research Innovation and Sequencing Platform | Sisonke, Giandhari Jennifer, Pillay Sureshnee, Yajna Ramphal, Naidoo Yeshnee, Tshabuila Derek,Tegally Houriiyah, San James, Wilkinson Eduan, de Oliveira Tulio |
| hCoV-19/USA/AK-PHL9806/2021 | EPI_ISL_2893340 | 2021-06-13 | Alaska State Virology Laboratory | Alaska State Virology Laboratory | Stephanie DeRonde, Elva House, Jacob Zidek, Lisa Smith, Ph.D., Jack Chen, Ph.D. |

| | | | | | |
|---|---|---|---|---|---|
| hCoV-19/USA/AL-CDC-ASC210236739/2021 | EPI_ISL_3851067 | 2021-07-29 | Aegis Sciences Corporation | Centers for Disease Control and Prevention Division of Viral Diseases, Pathogen Discovery | Dakota Howard,Dhwani Batra,Peter Cook,Kara Moser,Adrian Paskey,Jason Caravas,Benjamin Rambo-Martin,Shatavia Morrison,Christopher Gulvick,Scott Sammons,Yvette Unoarumhi,Darlene Wagner,Matthew Schmerer,Cyndi Clark,Patrick Campbell,Rob Case,Vikramsinha Ghorp |
| hCoV-19/USA/SC-MCRDPI_20_0522-T56/2020 | EPI_ISL_2894683 | 2020-07-22 | Sealfon Laboratory, Department of Neurology, Icahn School of Medicine at Mount Sinai | van Bakel Laboratory, Genetics and Genomics Sciences, Icahn School of Medicine at Mount Sinai | Rhonda A. Lizewski, Rachel S. G. Sealfon, Sang Woo Park, Gregory R. Smith, Chad K. Porter, Ana S. Gonzalez-Reiche, Yongchao Ge, Carl W. Goforth, Hanna Pincas, Michael S. Termini, Irene Ramos, Venugopalan D. Nair, Stephen E. Lizewski, Hala Alshammary, Regi |
| hCoV-19/Chile/VS-117141/2021 | EPI_ISL_2894965 | 2021-06-26 | Genetica Molecular and Subdepartamento de Virologia ISP Chile | Instituto de Salud Publica de Chile | Karen Orostica, Constanza Campano, Barbara Parra, Loredana Arata, Gisselle Barra, Patricia Bustos, Rodrigo Fasce, Javier Tognarelli, Andres Castillo, Soledad Ulloa, Jorge Fernandez |
| hCoV-19/Ecuador/NIC-INSPI-334429/2021 | EPI_ISL_2895665 | 2021-06-24 | INSPI-CRN DE INFLUENZA Y OTROS VIRUS RESPIRATORIOS | NIC-INSPI | Alfredo Bruno , Maritza Olmedo, Michelle Pv°ez, Jimmy Garcv©s, Johanna Laines, Lizbeth Pativ±o, Manuel Gonzalez, Domenica de Mora. |
| hCoV-19/Ecuador/NIC-INSPI-334444/2021 | EPI_ISL_2895674 | 2021-06-24 | INSPI-CRN DE INFLUENZA Y OTROS VIRUS RESPIRATORIOS | NIC-INSPI | Alfredo Bruno , Maritza Olmedo, Michelle Pv°ez, Jimmy Garcv©s, Johanna Laines, Lizbeth Pativ±o, Manuel Gonzalez, Domenica de Mora. |
| hCoV-19/SouthAfrica/NICD-N9829/2021 | EPI_ISL_2895798 | 2021-06-04 | PORT ELIZABETH LABORATORY | Diseases of the National Health Laboratory Service | Amoako DG, Everatt J, Scheepers C, Mohale T, Ntuli N, Mahlangu B, Mnguni A, Ismail A, Bhiman JN |
| hCoV-19/Australia/QLD2014/2021 | EPI_ISL_2896246 | 2021-07-08 | Queensland Health Forensic and Scientific Services | Queensland Health Forensic and Scientific Services | Son Nguyen |
| hCoV-19/Wales/PHWC-PYCBYP/2021 | EPI_ISL_2904697 | 2021-06-27 | Originating lab: Wales Specialist Virology Centre Sequencing lab: Pathogen Genomics Unit | Public Health Wales Microbiology Cardiff Wales Specialist Virology Centre | Catherine Moore, Johnathan Evans, Laura Gifford, Malorie Perry, Simon Cottrell, Angela Marchbank, Alec Birchley, Alexander Adams, Amy Gaskin, Bree Gatica-Wilcox, Jason Coombes, Joel Southgate, Lauren Gilbert, Lee Graham, Nicole Pacchiarini, Sara Kumziene- |
| hCoV-19/Wales/PHWC-PYMJG5/2021 | EPI_ISL_2906364 | 2021-06-27 | Originating lab: Wales Specialist Virology Centre Sequencing lab: Pathogen Genomics Unit | Public Health Wales Microbiology Cardiff Wales Specialist Virology Centre | Catherine Moore, Johnathan Evans, Laura Gifford, Malorie Perry, Simon Cottrell, Angela Marchbank, Alec Birchley, Alexander Adams, Amy Gaskin, Bree Gatica-Wilcox, Jason Coombes, Joel Southgate, Lauren Gilbert, Lee Graham, Nicole Pacchiarini, Sara Kumziene- |
| hCoV-19/Italy/TAA-1900619758/2021 | EPI_ISL_2907541 | 2021-06-12 | Azienda Sanitaria dell'Alto Adige - Laboratorio Aziendale di Microbiologia e Virologia | Azienda Sanitaria dell'Alto Adige | Irene Bianconi |
| hCoV-19/Australia/VIC18904/2021 | EPI_ISL_2907549 | 2021-06-28 | Microbiological Diagnostic Unit - Public Health Laboratory (MDU-PHL) | MDU-PHL | Seemann T., Sait, M.L., Sherry, N.L. |
| hCoV-19/England/MILK-18656C0/2021 | EPI_ISL_2908563 | 2021-07-02 | Lighthouse Lab in Milton Keynes | Wellcome Sanger Institute for the COVID-19 Genomics UK (COG-UK) Consortium | The Lighthouse Lab in Milton Keynes and Alex Alderton, Roberto Amato, Jeffrey Barrett, Sonia Goncalves, Ewan Harrison, David K. Jackson, Ian Johnston, Dominic Kwiatkowski, Cordelia Langford, John Sillitoe on behalf of the Wellcome Sanger Institute COVID-1 |

| | | | | | |
|---|---|---|---|---|---|
| hCoV-19/Switzerland/TI-EOC-902_26016241/2021 | EPI_ISL_2918883 | 2021-07-02 | Laboratorio di Microbiologia | Laboratorio di Microbiologia | Martinetti Lucchini Gladys, Valeria Spina |
| hCoV-19/Switzerland/TI-EOC-LB2126914/2021 | EPI_ISL_2918897 | 2021-06-30 | Laboratorio di Microbiologia | Laboratorio di Microbiologia | Martinetti Lucchini Gladys, Valeria Spina |
| hCoV-19/USA/AZ-TG955621/2020 | EPI_ISL_2919685 | 2020-09-29 | TGen North | TGen North | Jolene Bowers, Heather Centner, Brett Van Tassel, Chris French, Hayley Yaglom, Darrin Lemmer, Dave Engelthaler |
| hCoV-19/Australia/VIC18596/2021 | EPI_ISL_2920954 | 2021-05-26 | Victorian Infectious Diseases Reference Laboratory (VIDRL) | VIDRL and MDU-PHL | Caly L., Seemann T., Sait, M.L., Druce J., Sherry, N.L. |
| hCoV-19/Australia/VIC18679/2021 | EPI_ISL_2920958 | 2021-06-04 | Victorian Infectious Diseases Reference Laboratory (VIDRL) | VIDRL and MDU-PHL | Caly L., Seemann T., Sait, M.L., Druce J., Sherry, N.L. |
| hCoV-19/Australia/VIC18784/2021 | EPI_ISL_2920973 | 2021-06-13 | Victorian Infectious Diseases Reference Laboratory (VIDRL) | VIDRL and MDU-PHL | Caly L., Seemann T., Sait, M.L., Druce J., Sherry, N.L. |
| hCoV-19/Australia/VIC18808/2021 | EPI_ISL_2920976 | 2021-06-16 | Victorian Infectious Diseases Reference Laboratory (VIDRL) | VIDRL and MDU-PHL | Caly L., Seemann T., Sait, M.L., Druce J., Sherry, N.L. |
| hCoV-19/Australia/VIC18821/2021 | EPI_ISL_2920984 | 2021-06-20 | Victorian Infectious Diseases Reference Laboratory (VIDRL) | VIDRL and MDU-PHL | Caly L., Seemann T., Sait, M.L., Druce J., Sherry, N.L. |
| hCoV-19/Monaco/IPP14905/2021 | EPI_ISL_2923154 | 2021-02-22 | Labo Analyses Med | National Reference Center for Viruses of Respiratory Infections, Institut Pasteur, Paris | Marion Barbet, Sylvie Behillil, M√©line Bizard, Angela Brisebarre, Camille Capel, Vincent Enouf, Louise Lefran√ßois, Fr√©d√©ric Lemoine, Christophe Malabat, Corinne Maufrais, Hub Bioinformatique Biostatistiques, Etienne Simon-Lori√®re, Maud Vanpeene, Sylv |
| hCoV-19/France/BRE-IPP15053/2021 | EPI_ISL_2923266 | 2021-06-29 | Labo Analyses Med | National Reference Center for Viruses of Respiratory Infections, Institut Pasteur, Paris | Marion Barbet, Sylvie Behillil, M√©line Bizard, Angela Brisebarre, Camille Capel, Vincent Enouf, Louise Lefran√ßois, Fr√©d√©ric Lemoine, Christophe Malabat, Corinne Maufrais, Hub Bioinformatique Biostatistiques, Etienne Simon-Lori√®re, Maud Vanpeene, Sylv |
| hCoV-19/Italy/SIC_CQRC_21057151/2021 | EPI_ISL_2925922 | 2021-07-05 | Laboratorio CQRC | CQRC_QUALITY CONTROL CHEMICAL BIOLOGICAL RISK_AOOR Villa Sofia Cervello Palermo | Di Gaudio F., Contino F., Brunacci G. |
| hCoV-19/USA/CA-CDC-LC0083825/2021 | EPI_ISL_2926828 | 2021-06-24 | Laboratory Corporation of America | Centers for Disease Control and Prevention Division of Viral Diseases, Pathogen Discovery | Dakota Howard, Dhwani Batra, Peter W. Cook, Kara Moser, Adrian Paskey, Jason Caravas, Benjamin Rambo-Martin, Shatavia Morrison, Christopher Gulvick, Scott Sammons, Yvette Unoarumhi, Darlene Wagner, Matthew Schmerer, Minoo Agarwal, Eyad Almasri, Debbie Bol |
| hCoV-19/Canada/BC-BCCDC-92029/2021 | EPI_ISL_3485999 | 2021-05-17 | British Columbia Centre For Disease Control | BCCDC Public Health Laboratory | Prystajecky Natalie, Linda Hoang, Dan Fornika, John Tyson, Shannon Russell, Kim Macdonald, Kimia Kamelian, Ana Pacagnella, Corrinne Ng, Loretta Janz, Robert Azana, Mel Krajden |
| hCoV-19/Burundi/UG522/2021 | EPI_ISL_2928006 | 2021-05-27 | National Institute of Public Health Burundi, WHO Burundi, ASLM Burundi, MRC/UVRI & LSHTM Uganda Research Unit | MRC/UVRI & LSHTM Uganda Research Unit, National Institute of Public Health Burundi, WHO Burundi, ASLM Burundi | Jerome Nkurunziza, Anatole Nkeshimana, Joseph Nyandwi, Jean Baptiste Niyibigira, Butoyi Pascal, Cassien Nduwimana, Francine Kabatesi, Eric Kezakarayagwa, Xavier Crespin, Alexis Niyomwungere, Kouadio Theodore Yao, Jerome Ndaruhutse, Blessing T. Marondera, |

| | | | | | |
|---|---|---|---|---|---|
| hCoV-19/USA/TX-CDC-QDX26300187/2021 | EPI_ISL_2929191 | 2021-06-27 | Quest Diagnostics Incorporated | Centers for Disease Control and Prevention Division of Viral Diseases, Pathogen Discovery | Dakota Howard, Dhwani Batra, Peter W. Cook, Kara Moser, Adrian Paskey, Jason Caravas, Benjamin Rambo-Martin, Shatavia Morrison, Christopher Gulvick, Scott Sammons, Yvette Unoarumhi, Darlene Wagner, Matthew Schmerer, S. H. Rosenthal, A. Gerasimova, R. M. K |
| hCoV-19/USA/GA-QDX-26343988/2021 | EPI_ISL_2929699 | 2021-06-27 | Quest Diagnostics Incorporated | Centers for Disease Control and Prevention Division of Viral Diseases, Pathogen Discovery | Dakota Howard, Dhwani Batra, Peter W. Cook, Kara Moser, Adrian Paskey, Jason Caravas, Benjamin Rambo-Martin, Shatavia Morrison, Christopher Gulvick, Scott Sammons, Yvette Unoarumhi, Darlene Wagner, Matthew Schmerer, S. H. Rosenthal, A. Gerasimova, R. M. K |
| hCoV-19/USA/HI-QDX-26343987/2021 | EPI_ISL_2929940 | 2021-06-27 | Quest Diagnostics Incorporated | Centers for Disease Control and Prevention Division of Viral Diseases, Pathogen Discovery | Dakota Howard, Dhwani Batra, Peter W. Cook, Kara Moser, Adrian Paskey, Jason Caravas, Benjamin Rambo-Martin, Shatavia Morrison, Christopher Gulvick, Scott Sammons, Yvette Unoarumhi, Darlene Wagner, Matthew Schmerer, S. H. Rosenthal, A. Gerasimova, R. M. K |
| hCoV-19/HongKong/CM21000251.2/2021 | EPI_ISL_2931330 | 2021-03-25 | Temporary Specimen Collection Centre at the AsiaWorld-Expo | Hong Kong Department of Health | Alan K.L. Tsang, Peter C.W. Yip, Ken H.L. Ng, Edman T.K. Lam, Rickjason C.W. Chan |
| hCoV-19/Italy/APU-IZSPB-1560PT/2021 | EPI_ISL_2931690 | 2021-06-30 | I.R.C.C.S. "S. De Bellis" - Ente Ospedaliero | Istituto Zooprofilattico Sperimentale della Puglia e della Basilicata | Parisi A., Bianco A., Capozzi L., Del Sambro L., Simone D., Difato L., Notarnicola M., Lippolis A. |
| hCoV-19/Mexico/CMX-INMEGEN-04-07-11/2020 | EPI_ISL_4219934 | 2020-06-25 | Instituto Nacional de Medicina Genomica | Instituto Nacional de Medicina Genomica | Cedro-Tanda A, Escobar-Arrazola MA, Ramirez-Vega O, Rangel-DeLeon D, Yair Alfaro-Mora, Hidalgo-Miranda A, Mendoza-Vargas A, Reyes-Grajeda JP, Herrera-Montalvo LA. |
| hCoV-19/Turkey/HSGM-B13838/2021 | EPI_ISL_2936604 | 2021-05-25 | Ministry of Health Turkey | Ministry of Health Turkey | Fatma Bayrakdar, Yasemin Cosgun, Suleyman Yalcin, Gulay Korukluoglu |
| hCoV-19/Brazil/ES-FIOCRUZ-49165/2021 | EPI_ISL_4220215 | 2021-08-14 | Laboratorio Central de Saude Publica do Estado do Espirito Santo (LACEN/ES) | Laboratory of Respiratory Viruses and Measles, Oswaldo Cruz Institute, FIOCRUZ | Paola Resende, Luciana Appolinario, Fernando Motta, Anna Carolina Paixao, Ana Carolina Mendonca, Alice Sampaio Rocha, Taina Venas, Elisa Cavalcante Pereira, Renata Serrano Lopes, Rodrigo Ribeiro Rodrigues, Marilda Siqueira on behalf of the Fiocruz COVID-1 |
| hCoV-19/USA/WA-UW-21061516940/2021 | EPI_ISL_2937934 | 2021-06-15 | UW Virology Lab | UW Virology Lab | Pavitra Roychoudhury, Hong Xie, Lasata Shrestha, Shah Mohamed Bakhash, Tien V. Nguyen, Noah R. Baker, Sean Ellis, Meei-Li Huang, Keith R Jerome, Alexander Greninger |
| hCoV-19/Canada/QC-L00369034001/2021 | EPI_ISL_4220692 | 2021-07-21 | Laboratoire de sant√© publique du Qu√©bec | Laboratoire de sant√© publique du Qu√©bec | Sandrine Moreira, Ioannis Ragoussis, Guillaume Bourque, Jesse Shapiro, Mark Lathrop and Michel Roger on behalf of the CoVSeQ research group |
| hCoV-19/Canada/QC-L00371834001/2021 | EPI_ISL_4220846 | 2021-07-30 | Laboratoire de sant√© publique du Qu√©bec | Laboratoire de sant√© publique du Qu√©bec | Sandrine Moreira, Ioannis Ragoussis, Guillaume Bourque, Jesse Shapiro, Mark Lathrop and Michel Roger on behalf of the CoVSeQ research group |
| hCoV-19/Canada/AB-ABPHL-19259/2021 | EPI_ISL_2940480 | 2021-05-13 | Edmonton Provincial Lab | Public Health Agency of Canada (PHAC) National Microbiology Laboratory | Buss, E, Croxen M, Deo A, Dieu P, Gill K, Ferrato C, Khan F, Koleva P, Li V, Lloyd C, Lynch T, Ma R, Murphy S, Pabbaraju K, Shokoples S, Tipples G, Thayer J, Whitehouse M, Wong A, Yu C, Zelyas N |

| | | | | | |
|---|---|---|---|---|---|
| hCoV-19/Canada/AB-ABPHL-19155/2021 | EPI_ISL_2940681 | 2021-05-15 | Edmonton Provincial Lab | Public Health Agency of Canada (PHAC) National Microbiology Laboratory | Buss, E, Croxen M, Deo A, Dieu P, Gill K, Ferrato C, Khan F, Koleva P, Li V, Lloyd C, Lynch T, Ma R, Murphy S, Pabbaraju K, Shokoples S, Tipples G, Thayer J, Whitehouse M, Wong A, Yu C, Zelyas N |
| hCoV-19/Canada/AB-ABPHL-19162/2021 | EPI_ISL_2940688 | 2021-05-21 | Edmonton Provincial Lab | Public Health Agency of Canada (PHAC) National Microbiology Laboratory | Buss, E, Croxen M, Deo A, Dieu P, Gill K, Ferrato C, Khan F, Koleva P, Li V, Lloyd C, Lynch T, Ma R, Murphy S, Pabbaraju K, Shokoples S, Tipples G, Thayer J, Whitehouse M, Wong A, Yu C, Zelyas N |
| hCoV-19/Canada/QC-L00371203001/2021 | EPI_ISL_4221422 | 2021-07-31 | Laboratoire de sant√© publique du Qu√©bec | Laboratoire de sant√© publique du Qu√©bec | Sandrine Moreira, Ioannis Ragoussis, Guillaume Bourque, Jesse Shapiro, Mark Lathrop and Michel Roger on behalf of the CoVSeQ research group |
| hCoV-19/USA/AL-Mobile-SL-21052860003/2021 | EPI_ISL_2941518 | 2021-05-28 | Mobile County Health Department | Synergy Laboratories | Megan Cornwell |
| hCoV-19/Canada/BC-BCCDC-92956/2021 | EPI_ISL_3485690 | 2021-05-16 | British Columbia Centre For Disease Control | BCCDC Public Health Laboratory | Prystajecky Natalie, Linda Hoang, Dan Fornika, John Tyson, Shannon Russell, Kim Macdonald, Kimia Kamelian, Ana Pacagnella, Corrinne Ng, Loretta Janz, Robert Azana, Mel Krajden |
| hCoV-19/Gambia/15061/2021 | EPI_ISL_2941575 | 2021-02-23 | MRCG at LSHTM Genomics lab | MRCG at LSHTM Genomics lab | Abdul Karim sesay, Abdoulie Kanteh, Jarra Manneh, Mariama Kujabi, Bakary Sanyang,Dabiri Damilari |
| hCoV-19/USA/TX-Curative-067628/2021 | EPI_ISL_2941944 | 2021-05-18 | Curative Labs | Curative Labs | Mikhail Hanewich-Hollatz, J.Cesar Ignacio-Espinoza, Elias L. Salfati, Nabjot Sandhu, George Way, Zhiyi Xie, Eugenia Khorosheva, Sophia Quasem, Janet Chen, Vladimir Slepnev |
| hCoV-19/Canada/QC-L00375237001/2021 | EPI_ISL_4221726 | 2021-08-10 | Laboratoire de sant√© publique du Qu√©bec | Laboratoire de sant√© publique du Qu√©bec | Sandrine Moreira, Ioannis Ragoussis, Guillaume Bourque, Jesse Shapiro, Mark Lathrop and Michel Roger on behalf of the CoVSeQ research group |
| hCoV-19/Turkey/HSGM-B17850/2021 | EPI_ISL_2958416 | 2021-06-17 | Ministry of Health Turkey | Ministry of Health Turkey | Fatma Bayrakdar, Yasemin Cosgun, Suleyman Yalcin, Gulay Korukluoglu |
| hCoV-19/Turkey/HSGM-B18014/2021 | EPI_ISL_2958470 | 2021-06-23 | Ministry of Health Turkey | Ministry of Health Turkey | Fatma Bayrakdar, Yasemin Cosgun, Suleyman Yalcin, Gulay Korukluoglu |
| hCoV-19/Turkey/HSGM-B19011/2021 | EPI_ISL_2958620 | 2021-06-29 | Ministry of Health Turkey | Ministry of Health Turkey | Fatma Bayrakdar, Yasemin Cosgun, Suleyman Yalcin, Gulay Korukluoglu |
| hCoV-19/Gambia/36747/2021 | EPI_ISL_2958645 | 2021-05-19 | MRCG at LSHTM Genomics lab | MRCG at LSHTM Genomics lab | Abdul Karim sesay, Abdoulie Kanteh, Jarra Manneh, Mariama Kujabi, Bakary Sanyang,Dabiri Damilari |
| hCoV-19/USA/MN-MDH-8491/2021 | EPI_ISL_2958968 | 2021-06-23 | Mayo Clinic & Mayo Clinic Laboratories | Minnesota Department of Health, Public Health Laboratory | Alexandra Lorentz, Jacob Garfin, Matt Plumb, and Xiong Wang |
| hCoV-19/Liberia/LIB-0197/2021 | EPI_ISL_4232134 | 2021-04-06 | National Public Health Institute of Liberia Reference Lab | Center for Infection and Immunity, Columbia University | Bode Shobayo, Mitali Mishra, Komal Jain, Jane MaCauley, Thomas Briese, W. Ian Lipkin, Nischay Mishra |
| hCoV-19/NewZealand/21MV0555/2021 | EPI_ISL_2964930 | 2021-06-27 | LabPLUS | Institute of Environmental Science and Research (ESR) | Rachel Boyle, SallyAnn Harbison, Olivia Stroeven, Xiaoyun Ren, Matt Storey, Nikki Freed, Muhammad Faisal, Jing Wang, Hermes Perez, Anja Werno, Antje van der Linden, Arlo Upton, Chris Mansell, David Hammer, Dragana Drinkovic, Gary McAuliffe, Hana Sofia And |

| | | | | | |
|---|---|---|---|---|---|
| hCoV-19/NewZealand/21MV0574/2021 | EPI_ISL_2964936 | 2021-07-02 | Middlemore Hospital | Institute of Environmental Science and Research (ESR) | Rachel Boyle, SallyAnn Harbison, Olivia Stroeven, Xiaoyun Ren, Matt Storey, Nikki Freed, Muhammad Faisal, Jing Wang, Hermes Perez, Anja Werno, Antje van der Linden, Arlo Upton, Chris Mansell, David Hammer, Dragana Drinkovic, Gary McAuliffe, Hana Sofia And |
| hCoV-19/India/MH-ICMR-NIV-INSACOG-GSEQ-4177/2021 | EPI_ISL_2965888 | 2021-06-07 | ICMR-National Institute of Virology - INSACOG | NIV Influenza | Potdar, Varsha on behalf of National Influenza Centre, NIV, Pune |
| hCoV-19/Bulgaria/21BG-NC_003581_R14/2021 | EPI_ISL_2967177 | 2021-06-23 | National Center of Infectious and Parasitic Diseases | National Center of Infectious and Parasitic Diseases | Alexiev et al |
| hCoV-19/Turkey/HSGM-B15134/2021 | EPI_ISL_2968531 | 2021-06-01 | Ministry of Health Turkey | Ministry of Health Turkey | Fatma Bayrakdar, Yasemin Cosgun, Suleyman Yalcin, Gulay Korukluoglu |
| hCoV-19/DRC/RDC-75/2021 | EPI_ISL_2968690 | 2021-05-11 | Viral Respiratory Lab, National Institute for Biomedical Research (INRB) | Pathogen Sequencing Lab, National Institute for Biomedical Research (INRB) | Placide Mbala-Kingebeni, Edith Nkwembe, Eddy Kinganda-Lusamaki, Amuri Aziza, Francisca Muyembe Mawete, Emmanuel Lokilo Lofiko, Jean Claude Makangara, Catherine Pratt, Matthias Pauthner, Josh Quick, Allison Black, James Hadfield, Trevor Bedford, Ian Goodfe |
| hCoV-19/Switzerland/UN-ETHZ-670143/2021 | EPI_ISL_4233747 | 2021-08-05 | labor team w AG | Department of Biosystems Science and Engineering, ETH Z√ºrich | Ivan Topolsky, Sarah Nadeau, Chaoran Chen, Louis du Plessis, Rebekka Pohl, Monika Bucher, Andreas Lindauer, Catharine Aquino, Daniel Ehrsam, Isabel St√ºrmer, Andreia Cabral de Gouvea, Maria Domenica Moccia, Simon Gr√üter, Timothy Sykes, Lennart Opitz, Gri |
| hCoV-19/Australia/SA0808/2021 | EPI_ISL_2978690 | 2021-07-07 | SA Pathology | SA Pathology | Caitlin Selway, Lex Leong, Chuan Kok Lim, Mark Turra, Ivan Bastian, Geoff Higgins |
| hCoV-19/Australia/SA0810/2021 | EPI_ISL_2978692 | 2021-07-07 | SA Pathology | SA Pathology | Caitlin Selway, Lex Leong, Chuan Kok Lim, Mark Turra, Ivan Bastian, Geoff Higgins |
| hCoV-19/Australia/SA0818/2021 | EPI_ISL_2978698 | 2021-07-10 | SA Pathology | SA Pathology | Caitlin Selway, Lex Leong, Chuan Kok Lim, Mark Turra, Ivan Bastian, Geoff Higgins |
| hCoV-19/Australia/SA0823/2021 | EPI_ISL_2978700 | 2021-07-12 | SA Pathology | SA Pathology | Caitlin Selway, Lex Leong, Chuan Kok Lim, Mark Turra, Ivan Bastian, Geoff Higgins |
| hCoV-19/Sweden/20460162P/2021 | EPI_ISL_2980514 | 2021-06-22 | Swedish national genomic surveillance program of SARS-CoV-2 | The Public Health Agency of Sweden | Maximilian Riess, Maria Lind Karlberg, Alma Brolund, Swedish national genomic surveillance program of SARS-CoV-2 |
| hCoV-19/Sweden/1260503240VK/2021 | EPI_ISL_2980588 | 2021-06-17 | Swedish national genomic surveillance program of SARS-CoV-2 | The Public Health Agency of Sweden | Maximilian Riess, Maria Lind Karlberg, Alma Brolund, Swedish national genomic surveillance program of SARS-CoV-2 |
| hCoV-19/Brazil/PR-FIOCRUZ-47437/2021 | EPI_ISL_4237184 | 2021-07-28 | Laboratorio Central de Saude Publica do Estado do Parana (LACEN/PR) | Laboratory of Respiratory Viruses and Measles, Oswaldo Cruz Institute, FIOCRUZ | Paola Resende, Luciana Appolinario, Fernando Motta, Anna Carolina Paixao, Ighor Leonardo Arantes, Agatha Soares, Ana Carolina Mendonca, Alice Sampaio Rocha, Taina Venas, Elisa Cavalcante Pereira, Renata Serrano Lopes, Irina Riediger, Marilda Siqueira on b |
| hCoV-19/Brazil/PR-FIOCRUZ-47441/2021 | EPI_ISL_4237185 | 2021-07-28 | Laboratorio Central de Saude Publica do Estado do Parana (LACEN/PR) | Laboratory of Respiratory Viruses and Measles, Oswaldo Cruz Institute, FIOCRUZ | Paola Resende, Luciana Appolinario, Fernando Motta, Anna Carolina Paixao, Ighor Leonardo Arantes, Agatha Soares, Ana Carolina Mendonca, Alice Sampaio Rocha, Taina Venas, Elisa Cavalcante Pereira, Renata Serrano Lopes, Irina Riediger, Marilda Siqueira on b |

| | | | | | |
|---|---|---|---|---|---|
| hCoV-19/Netherlands/OV-RIVM-43000/2021 | EPI_ISL_2981476 | 2021-06-28 | Dutch COVID-19 response team | National Institute for Public Health and the Environment (RIVM) | Adam Meijer, Harry Vennema, Dirk Eggink, Jeroen Cremer, Sharon van den Brink, Bas van der Veer, AnneMarie van den Brandt, Lisa Wijsman, Kim Freriks, Ryanne Jaarsma, Eun√≠ce Then, Lynn Aarts, Sanne Bos, Melissa van Tuil, Florian Zwagemaker, Dennis Schmitz, |
| hCoV-19/Aruba/AW-RIVM-43082/2021 | EPI_ISL_2981892 | 2021-06-30 | Dutch COVID-19 response team | National Institute for Public Health and the Environment (RIVM) | Adam Meijer, Harry Vennema, Dirk Eggink, Jeroen Cremer, Sharon van den Brink, Bas van der Veer, AnneMarie van den Brandt, Lisa Wijsman, Kim Freriks, Ryanne Jaarsma, Eun√≠ce Then, Lynn Aarts, Sanne Bos, Melissa van Tuil, Florian Zwagemaker, Dennis Schmitz, |
| hCoV-19/Bonaire/BQ-RIVM-42362/2021 | EPI_ISL_2981898 | 2021-06-29 | Dutch COVID-19 response team | National Institute for Public Health and the Environment (RIVM) | Adam Meijer, Harry Vennema, Dirk Eggink, Jeroen Cremer, Sharon van den Brink, Bas van der Veer, AnneMarie van den Brandt, Lisa Wijsman, Kim Freriks, Ryanne Jaarsma, Eun√≠ce Then, Lynn Aarts, Sanne Bos, Melissa van Tuil, Florian Zwagemaker, Dennis Schmitz, |
| hCoV-19/USA/MA-CDCBI-CRSP_SHW7NSZFLUXC74TW/2021 | EPI_ISL_2991837 | 2021-05-29 | Broad Institute Clinical Research Sequencing Platform | Infectious Disease Program, Broad Institute of Harvard and MIT | Lemieux,J.E., Siddle,K.J., Shaw,B., Adams,G., Pierce,V., Turbett,S., Anahtar,M., Branda,J., Slater,D., Harris,J., Lin,A.E., Gladden-Young,A., Lagerborg,K., Rudy,M., DeRuff,K., Carter,A., Normandin,E., Bauer,M., Reilly,S., Tomkins-Tinch,C., Loreth,C., Chal |
| hCoV-19/SouthAfrica/NICD-N10778/2021 | EPI_ISL_2987246 | 2021-06-29 | ERMELO LABORATORY | Diseases of the National Health Laboratory Service | Amoako DG, Everatt J, Scheepers C, Mohale T, Ntuli N, Mahlangu B, Mnguni A, Ismail A, Bhiman JN |
| hCoV-19/Turkey/HSGM-B17788/2021 | EPI_ISL_2987974 | 2021-06-21 | Ministry of Health Turkey | Ministry of Health Turkey | Fatma Bayrakdar, Yasemin Cosgun, Suleyman Yalcin, Gulay Korukluoglu |
| hCoV-19/Netherlands/GR-UMCG-MMB_2236/2021 | EPI_ISL_2988928 | 2021-07-09 | UMC Groningen, Clinical Virology, Department of Medical Microbiology and Infection Prevention | UMC Groningen, Clinical Virology, Department of Medical Microbiology and Infection Prevention | Hubert Niesters, Alexander Friedrich, Erley Lizarazo-Forero, Monika Fliss, Lilli Gard, Sigrid Rosema, Coretta Van Leer-Buter, Xuewei Zhou, Marjolein Knoester |
| hCoV-19/England/PLYM-18DC2A2/2021 | EPI_ISL_2998100 | 2021-07-10 | Lighthouse Laboratory Plymouth | Wellcome Sanger Institute for the COVID-19 Genomics UK (COG-UK) Consortium | Lighthouse Laboratory Plymouth and Alex Alderton, Roberto Amato, Jeffrey Barrett, Sonia Goncalves, Ewan Harrison, David K. Jackson, Ian Johnston, Dominic Kwiatkowski, Cordelia Langford, John Sillitoe on behalf of the Wellcome Sanger Institute COVID-19 Sur |
| hCoV-19/India/OR-ILSGS02629/2021 | EPI_ISL_3007047 | 2021-05-01 | District Head Quarters Hospital, Nayagarh | Institute of Life Sciences - INSACOG | Sunil K. Raghav, Safal Walia, Arup Ghosh, Atimukta Jha, Amol M. Kanampalliwar, Omprakash Shiriwas, Sana Fatma, Shifu Aggarwal, Rupesh Dash, Rajeeb Swain, Punit Prasad, INSACOG Consortium, Ajay Parida |
| hCoV-19/India/OR-ILSGS02738/2021 | EPI_ISL_3007096 | 2021-04-24 | Regional Medical Research Center, Bhubaneswar | Institute of Life Sciences - INSACOG | Sunil K. Raghav, Safal Walia, Arup Ghosh, Atimukta Jha, Amol M. Kanampalliwar, Omprakash Shiriwas, Sana Fatma, Shifu Aggarwal, Rupesh Dash, Rajeeb Swain, Punit Prasad, INSACOG Consortium, Ajay Parida |
| hCoV-19/Switzerland/ZH-ETHZ-33317997/2021 | EPI_ISL_3007624 | 2021-06-26 | Viollier AG | Department of Biosystems Science and Engineering, ETH Z√ºrich | Andrea Patrizia Salzmann, Henriette Kurth, Christiane Beckmann, Ivan Topolsky, Chaoran Chen, Sarah Nadeau, Kim Philipp Jablonski, Lara Fuhrmann, Niko Beerenwinkel, Christoph Noppen, Olivier Kobel, Maurice Redondo, Tanja Stadler |

| | | | | | |
|---|---|---|---|---|---|
| hCoV-19/Norway/3746/2020 | EPI_ISL_3008431 | 2020-09-01 | Ostfold Hospital Trust - Kalnes, Centre for Laboratory Medicine, Section for gene technology and infection serology | Norwegian Institute of Public Health, Department of Virology | Kathrine Stene-Johansen, Kamilla Heddeland Instefjord, Hilde Elshaug, Garcia Llorente Ignacio, Jon Br√•te, Engebretsen Serina Beate,Pedersen Benedikte Nevjen, Line Victoria Moen, Debech Nadia, Atiya R Ali,Marie Paulsen Madsen, Rasmus Riis Kopperud, Hilde |
| hCoV-19/England/MILK-1915EB7/2021 | EPI_ISL_3013730 | 2021-07-13 | Lighthouse Lab in Milton Keynes | Wellcome Sanger Institute for the COVID-19 Genomics UK (COG-UK) Consortium | The Lighthouse Lab in Milton Keynes and Alex Alderton, Roberto Amato, Jeffrey Barrett, Sonia Goncalves, Ewan Harrison, David K. Jackson, Ian Johnston, Dominic Kwiatkowski, Cordelia Langford, John Sillitoe on behalf of the Wellcome Sanger Institute COVID-1 |
| hCoV-19/Finland/8063/2021 | EPI_ISL_3015007 | 2021-05-29 | Department of Virology and Immunology, University of Helsinki and Helsinki University Hospital, Huslab Finland | Department of Virology, Faculty of Medicine, University of Helsinki, Helsinki, Finland | Teemu Smura, Ravi Kant, Phuoc Truong, Hussein Alburkat, Hannimari Kallio-Kokko, Jenni Virtanen, Maija Suvanto, Essi Korhonen, Sari Hannula, Harri Kangas, Hanna Liimatainen, Satu Kurkela, Hanna Jarva, Maija Lappalainen, Pekka Ellonen, Olli Vapalahti |
| hCoV-19/India/CT-ILSGS04064/2021 | EPI_ISL_3015518 | 2021-04-17 | Pt. Jawahar Lal Nehru Memorial Medical College, Raipur | Institute of Life Sciences - INSACOG | Sunil K. Raghav, Safal Walia, Arup Ghosh, Atimukta Jha, Amol M. Kanampalliwar, Omprakash Shiriwas, Sana Fatma, Shifu Aggarwal, Rupesh Dash, Rajeeb Swain, Punit Prasad, INSACOG Consortium, Ajay Parida |
| hCoV-19/USA/TX-CDC-LC0090133/2021 | EPI_ISL_3017027 | 2021-06-23 | Laboratory Corporation of America | Centers for Disease Control and Prevention Division of Viral Diseases, Pathogen Discovery | Dakota Howard, Dhwani Batra, Peter W. Cook, Kara Moser, Adrian Paskey, Jason Caravas, Benjamin Rambo-Martin, Shatavia Morrison, Christopher Gulvick, Scott Sammons, Yvette Unoarumhi, Darlene Wagner, Matthew Schmerer, Minoo Agarwal, Eyad Almasri, Debbie Bol |
| hCoV-19/India/WB-1930400538182/2021 | EPI_ISL_3019259 | 2021-06-08 | INSACOG-WB | National Institute of Biomedical Genomics ,Äì INSACOG | Arindam Maitra, Bhaswati Bandyopadhyay, Nidhan Kumar Biswas, Tamal Ghosh, Sreedhar Chinnaswamy, Ajay Chakraborti, Saumitra Das |
| hCoV-19/Israel/SMC-7004607/2021 | EPI_ISL_3019614 | 2021-06-25 | Shamir Medical Center (Asaf Harofe) | Shamir Medical Center (Asaf Harofe) | Nir Rainy,Chen Weiner,Reut Sorek Abramovich,Yevgeni Yegorov,Anna Vishnevsky,Patricia Benveniste-Lekovitz,Abu Hamad Ramzia,Adina Bar Chaim |
| hCoV-19/Finland/THL-202118915/2021 | EPI_ISL_3020544 | 2021-05-24 | NordLab Kokkola | Expert Microbiology, National Institute for Health and Welfare | Soile Blomqvist, Jani Halkilahti, Kirsi Liitsola, Haider al-Hello, P√§ivi Laurila, Erika Lindh, Teemu Smura, Ravi Kant, Phuoc Truong, Olli Vapalahti, Sari Hannula, Pekka Ellonen, Niina Ikonen, Carita Savolainen-Kopra |
| hCoV-19/USA/CA-CDC-2-4691929/2021 | EPI_ISL_4249127 | 2021-07-15 | CA-Los Angeles County Public Health Laboratory | Centers for Disease Control and Prevention Division of Viral Diseases, Pathogen Discovery | Mili Sheth,Sarah Nobles,Jasmine Padilla,Alex Burgin,Meghan Bentz,Mark Burroughs,Peter Cook,Clinton Paden,Dhwani Batra,Krista Queen,Kristen Knipe,Dakota Howard,Yvette Unoarumhi,Matthew Schmerer,Ben Rambo-Martin,Kristine Lacek,Sam Shepard,Dave Wentworth,Viv |
| hCoV-19/Israel/SMC-7004998/2021 | EPI_ISL_3021589 | 2021-06-28 | Shamir Medical Center (Asaf Harofe) | Shamir Medical Center (Asaf Harofe) | Nir Rainy,Chen Weiner,Reut Sorek Abramovich,Yevgeni Yegorov,Anna Vishnevsky,Patricia Benveniste-Lekovitz,Abu Hamad Ramzia,Adina Bar Chaim |
| hCoV-19/Israel/SMC-7005601/2021 | EPI_ISL_3021866 | 2021-06-30 | Shamir Medical Center (Asaf Harofe) | Shamir Medical Center (Asaf Harofe) | Nir Rainy,Chen Weiner,Reut Sorek Abramovich,Yevgeni Yegorov,Anna Vishnevsky,Patricia Benveniste-Lekovitz,Abu Hamad Ramzia,Adina Bar Chaim |

| | | | | | |
|---|---|---|---|---|---|
| hCoV-19/Israel/SMC-7005149/2021 | EPI_ISL_3022133 | 2021-06-28 | Shamir Medical Center (Asaf Harofe) | Shamir Medical Center (Asaf Harofe) | Nir Rainy,Chen Weiner,Reut Sorek Abramovich,Yevgeni Yegorov,Anna Vishnevsky,Patricia Benveniste-Lekovitz,Abu Hamad Ramzia,Adina Bar Chaim |
| hCoV-19/Israel/SMC-7006435/2021 | EPI_ISL_3024467 | 2021-07-07 | Shamir Medical Center (Asaf Harofe) | Shamir Medical Center (Asaf Harofe) | Nir Rainy,Chen Weiner,Reut Sorek Abramovich,Yevgeni Yegorov,Anna Vishnevsky,Patricia Benveniste-Lekovitz,Abu Hamad Ramzia,Adina Bar Chaim |
| hCoV-19/SouthAfrica/NICD-N10144/2021 | EPI_ISL_3024517 | 2021-06-25 | LEBOWAKGOMO LABORATORY | Diseases of the National Health Laboratory Service | Amoako DG, Everatt J, Scheepers C, Mohale T, Ntuli N, Mahlangu B, Mnguni A, Ismail A, Bhiman JN |
| hCoV-19/SouthAfrica/NICD-N10169/2021 | EPI_ISL_3024538 | 2021-06-27 | MANKWENG LABORATORY | Diseases of the National Health Laboratory Service | Amoako DG, Everatt J, Scheepers C, Mohale T, Ntuli N, Mahlangu B, Mnguni A, Ismail A, Bhiman JN |
| hCoV-19/SouthAfrica/NICD-N7653/2021 | EPI_ISL_3024680 | 2021-05-23 | CHRIS HANI BARAGWANATH LABORATORY | Diseases of the National Health Laboratory Service | Amoako DG, Everatt J, Scheepers C, Mohale T, Ntuli N, Mahlangu B, Mnguni A, Ismail A, Bhiman JN |
| hCoV-19/SouthKorea/KDCA6312/2021 | EPI_ISL_3026398 | 2021-06-28 | Division of Emerging Infectious Diseases, Bureau of Infectious Diseases Diagnosis Control, Korea Disease Control and Prevention Agency | Diseases, Bureau of Infectious Diseases Diagnosis Control, Korea Disease Control and Prevention Agency | Jeong-Ah Kim, Ae Kyung Park, Il-Hwan Kim, Heui Man Kim, Jeong-Min Kim, Chae Young Lee, Eun-Jin Kim |
| hCoV-19/England/ALDP-1DCF86F/2021 | EPI_ISL_4250816 | 2021-09-06 | Lighthouse Lab in Alderley Park | Wellcome Sanger Institute for the COVID-19 Genomics UK (COG-UK) Consortium | Jacquelyn Wynn, Mairead Hyland, The Lighthouse Lab in Alderley Park and Alex Alderton, Roberto Amato, Jeffrey Barrett, Sonia Goncalves, Ewan Harrison, David K. Jackson, Ian Johnston, Dominic Kwiatkowski, Cordelia Langford, John Sillitoe on behalf of the W |
| hCoV-19/Kenya/ILRI_COVC20167/2020 | EPI_ISL_4252059 | 2020-10-27 | International Livestock Research Institute | International Livestock Research Institute | Samuel O. Oyola, Gilbert Kibet, Daniel Ouso, Shebbar Osiany, Edward Kiritu, Paul Dobi, Collins Muli, Patrick Amoth, Vishvanath Nene, Sonal P. Henson, Edward O. Abworo |
| hCoV-19/Australia/VIC18952/2021 | EPI_ISL_3030413 | 2021-07-16 | Microbiological Diagnostic Unit - Public Health Laboratory (MDU-PHL) | MDU-PHL | Seemann T., Sait, M.L., Sherry, N.L. |
| hCoV-19/Kenya/ILRI_COVC18975/2020 | EPI_ISL_4252102 | 2020-11-05 | International Livestock Research Institute | International Livestock Research Institute | Samuel O. Oyola, Gilbert Kibet, Daniel Ouso, Shebbar Osiany, Edward Kiritu, Paul Dobi, Collins Muli, Patrick Amoth, Vishvanath Nene, Sonal P. Henson, Edward O. Abworo |
| hCoV-19/Ireland/TA-NVRL-S21IRL41977/2021 | EPI_ISL_3030770 | 2021-07-04 | National Virus Reference Laboratory | National Virus Reference Laboratory | Zoe Yandle, Charlene Bennett, Gabriel Gonzalez, Michael Carr, Jonathan Dean, Cillian F De Gascun |
| hCoV-19/Kenya/ILRI_COVC24195/2021 | EPI_ISL_4252259 | 2021-06-03 | International Livestock Research Institute | International Livestock Research Institute | Samuel O. Oyola, Gilbert Kibet, Daniel Ouso, Shebbar Osiany, Edward Kiritu, Paul Dobi, Collins Muli, Patrick Amoth, Vishvanath Nene, Sonal P. Henson, Edward O. Abworo |
| hCoV-19/Kenya/ILRI_COVC23915/2021 | EPI_ISL_4252302 | 2021-07-07 | International Livestock Research Institute | International Livestock Research Institute | Samuel O. Oyola, Gilbert Kibet, Daniel Ouso, Shebbar Osiany, Edward Kiritu, Paul Dobi, Collins Muli, Patrick Amoth, Vishvanath Nene, Sonal P. Henson, Edward O. Abworo |
| hCoV-19/France/IDF-IPP15497/2021 | EPI_ISL_3031887 | 2021-07-07 | Labo Analyses Med | National Reference Center for Viruses of Respiratory Infections, Institut Pasteur, Paris | Marion Barbet, Sylvie Behillil, M√©line Bizard, Angela Brisebarre, Camille Capel, Vincent Enouf, Louise Lefran√ßois, Fr√©d√©ric Lemoine, Christophe Malabat, Corinne Maufrais, Hub de Bioinformatique et Statistique, Etienne Simon-Lori√®re, Maud Vanpeene, Sy |

| | | | | | |
|---|---|---|---|---|---|
| hCoV-19/Italy/VEN-IZSVe-21RS1817-40_VR/2021 | EPI_ISL_3039631 | 2021-07-08 | AULSS 9 Scaligera - Villafranca | Istituto Zooprofilattico Sperimentale delle Venezie | Adelaide Milani, Alessia Schivo, Annalisa Salviato, Elisa Palumbo, Erika Giorgia Quaranta, Luca Tassoni, Ambra Pastori, Edoardo Giussani, Alice Fusaro, Isabella Monne, Calogero Terregino, Antonia Ricci |
| hCoV-19/Ghana/nCoV-TRA-1780/2021 | EPI_ISL_4253801 | 2021-08-18 | National Influenza Centre | National Influenza Centre | William K. Ampofo, Michael Marks, Ivy A. Asante, Sharon Hsu, Benjamiin B. Lindsey, Benjamin H. Foulkes, Mildred Adusei-Poku, Linda Boatemaa, Lorreta Kwasah, Joseph Oliver-Commey, Ernest Asiedu, Franklin Asiedu-Bekoe, Gordon Awandare, Joyce Ngoi, Dennis La |
| hCoV-19/Ghana/FS-21-2274/2021 | EPI_ISL_4253832 | 2021-08-13 | National Influenza Centre | National Influenza Centre | William K. Ampofo, Michael Marks, Ivy A. Asante, Sharon Hsu, Benjamiin B. Lindsey, Benjamin H. Foulkes, Mildred Adusei-Poku, Linda Boatemaa, Lorreta Kwasah, Joseph Oliver-Commey, Ernest Asiedu, Franklin Asiedu-Bekoe, Gordon Awandare, Joyce Ngoi, Dennis La |
| hCoV-19/Australia/NSW-SAVID-4737/2021 | EPI_ISL_4254513 | 2021-06-06 | Area of Virology, Serology and Virology Division (SAViD), New South Wales Health Pathology Randwick | Virology, Serology and Virology Division (SAViD), New South Wales Health Pathology Randwick | Foster, C.; Au, J.; Ruiz Silva, M.; Deveson, I.; Bull, R.; Van Hal, S.; Rawlinson, W. |
| hCoV-19/Australia/NSW-SAVID-5128/2021 | EPI_ISL_4254603 | 2021-08-18 | Area of Virology, Serology and Virology Division (SAViD), New South Wales Health Pathology Randwick | Virology, Serology and Virology Division (SAViD), New South Wales Health Pathology Randwick | Foster, C.; Au, J.; Ruiz Silva, M.; Deveson, I.; Bull, R.; Van Hal, S.; Rawlinson, W. |
| hCoV-19/USA/NC-SLPH-0226/2021 | EPI_ISL_3045292 | 2021-06-24 | NCSLPH | NCSLPH | Chase K, Miller MC, Greene S, Glover W |
| hCoV-19/Australia/QLD2041/2021 | EPI_ISL_3049193 | 2021-07-18 | Queensland Health Forensic and Scientific Services | Queensland Health Forensic and Scientific Services | Son Nguyen |
| hCoV-19/Kenya/C50996/2020 | EPI_ISL_3049392 | 2020-08-28 | KEMRI-Wellcome Trust Research Programme,Kilifi | KEMRI-Wellcome Trust Research Programme,Kilifi | Githinji G.,Matoke D.,Mohamed K.S.,de Laurent Z.,Mburu M.W.,Thiongo K. |
| hCoV-19/Kenya/C99548/2021 | EPI_ISL_3049435 | 2021-06-08 | KEMRI-Wellcome Trust Research Programme,Kilifi | KEMRI-Wellcome Trust Research Programme,Kilifi | Githinji G.,Matoke D.,Mohamed K.S.,de Laurent Z.,Mburu M.W.,Thiongo K. |
| hCoV-19/Kuwait/Jaber2120290383/2021 | EPI_ISL_2958688 | 2021-07-11 | Jaber Al Ahmad Hospital | Virology Lab, Jaber Al Ahmad Hospital | Dr. Zahrah Buhamad, Dr. Ebaa Al-Awadhi, Haroon Masih, Khubaid-ur-Rehman |
| hCoV-19/Ecuador/USFQ-2045/2021 | EPI_ISL_4256768 | 2021-08-15 | Institute of Microbiology, Universidad San Francisco de Quito | Institute of Microbiology, Universidad San Francisco de Quito | Monica Becerra-Wong, Juan Jos√© Guadalupe, Sully M√°rquez, Bel√©n Prado-Vivar, Fernanda Zurita, Erika B. Mu√±oz, Mateo Carvajal, Bernardo Guti√©rrez, Melva Morales, Ver√≥nica Barrag√°n, Patricio Rojas-Silva, Gabriel Trueba, Michelle Grunauer, Pa√∫l C√°rde |
| hCoV-19/Ecuador/USFQ-2065/2021 | EPI_ISL_4256794 | 2021-08-22 | Institute of Microbiology, Universidad San Francisco de Quito | Institute of Microbiology, Universidad San Francisco de Quito | Bel√©n Prado-Vivar, Erika B. Mu√±oz, Fernanda Zurita, Mateo Carvajal, Monica Becerra-Wong, Juan Jos√© Guadalupe, Sully M√°rquez, Bernardo Guti√©rrez, Melva morales, Ver√≥nica Barrag√°n, Patricio Rojas-Silva, Gabriel Trueba, Michelle Grunauer, Pa√∫l C√°rde |
| hCoV-19/Netherlands/UT-RIVM-43585/2021 | EPI_ISL_3057208 | 2021-07-06 | Dutch COVID-19 response team | National Institute for Public Health and the Environment (RIVM) | Adam Meijer, Harry Vennema, Dirk Eggink, Jeroen Cremer, Sharon van den Brink, Bas van der Veer, AnneMarie van den Brandt, Lisa Wijsman, Kim Freriks, Ryanne Jaarsma, Eun√≠ce Then, Lynn Aarts, Sanne Bos, Melissa van Tuil, Florian Zwagemaker, Dennis Schmitz, |

| | | | | | |
|---|---|---|---|---|---|
| hCoV-19/USA/NY-URMC-21070681-1/2021 | EPI_ISL_4258736 | 2021-07-29 | URMC LABS | URMC LABS | Ghinwa Dumyati, Dwight Hardy, Andrew Cameron, Stepan Voljehn, Kelly DeLary |
| hCoV-19/India/MH-NEERI-CIA5560/2021 | EPI_ISL_3060555 | 2021-06-10 | CSIR-National Environmental Engineering Research Institute | CSIR-Centre for Cellular and Molecular Biology-INSACOG | Payel Mukherjee, Lamuk Zaveri, Onkar Kulkarni,Tulasi Nagabandi, Sofia Banu, Ara Sreenivas,Shreekant Verma, Amareshwar Vodapalli ,B Himasri,Valli Nagalakshmi Undamatla, Sumedha Avadhanula, Vidhyadhari Methuku, Priya Nurkurthy, Archana Bharadwaj Siva, Karth |
| hCoV-19/India/TG-CCMB-CIA5806/2021 | EPI_ISL_3060639 | 2021-06-22 | Mapmygenome | CSIR-Centre for Cellular and Molecular Biology-INSACOG | Amareshwar Vodapalli,Ara Sreenivas,Shreekant Verma, B Himasri,Valli Nagalakshmi Undamatla,Payel Mukherjee, Lamuk Zaveri, Onkar Kulkarni,Tulasi Nagabandi, Sofia Banu, Jandhyala Sai Krishna,Priya Nurkurthy, Vidhyadhari Methuku, Sumedha Avadhanula, Archana B |
| hCoV-19/India/TG-CCMB-CIA6136/2021 | EPI_ISL_3060892 | 2021-06-28 | CSIR-Centre for Cellular and Molecular Biology | CSIR-Centre for Cellular and Molecular Biology-INSACOG | Vidhyadhari Methuku,Tulasi Nagabandi,Lamuk Zaveri, Ara Sreenivas,Shreekant Verma, Amareshwar Vodapalli ,B Himasri,Valli Nagalakshmi Undamatla, Priya Nurkurthy,Sumedha Avadhanula, Archana Bharadwaj Siva,Payel Mukherjee,Onkar Kulkarni, Jandhyala Sai Krishna |
| hCoV-19/USA/NC-UNC-1620/2021 | EPI_ISL_3061570 | 2021-06-13 | Clinical Molecular Microbiology Laboratory, UNC Hospitals | Jeremy Wang | Jeremy Wang, Shawn Hawken, Corbin Jones, Melissa Miller |
| hCoV-19/Ecuador/USFQ-1681-UTPL86/2021 | EPI_ISL_3063272 | 2021-07-05 | Health sciences Department, Universidad T√©cnica Particular de Loja | Institute of Microbiology, Universidad San Francisco de Quito | Monica Becerra-Wong, Erika Mu√±oz, Bel√©n Prado-Vivar, Sully M√°rquez, Juan Jos√© Guadalupe, Fernanda Zurita, Bernardo Guti√©rrez, Katty Ojeda, Fernando Serrano, Paola Dalgo, Ligia Brice√±o, Ver√≥nica Barrag√°n, Patricio Rojas-Silva, Gabriel Trueba, Miche |
| hCoV-19/Turkey/HSGM-B4146/2021 | EPI_ISL_3063470 | 2021-03-31 | Ministry of Health Turkey | Ministry of Health Turkey | Fatma Bayrakdar, Yasemin Cosgun, Suleyman Yalcin, Gulay Korukluoglu |
| hCoV-19/USA/WA-CDC-UW21071064559/2021 | EPI_ISL_3063528 | 2021-07-10 | UW Virology Lab | UW Virology Lab | Pavitra Roychoudhury, Shah Mohamed Bakhash, Ricardo Perez, Hong Xie, Tien V. Nguyen, Maria Lukes, Lasata Shrestha, Sean Ellis, Nathan Breit, Robert J. Livingston, Meei-Li Huang, Keith R Jerome, Patrick Mathias, Alexander Greninger |
| hCoV-19/USA/CO-CDC-MMB09241269/2021 | EPI_ISL_3065081 | 2021-06-24 | Mako Medical | Centers for Disease Control and Prevention Division of Viral Diseases, Pathogen Discovery | Dakota Howard, Dhwani Batra, Peter W. Cook, Kara Moser, Adrian Paskey, Jason Caravas, Benjamin Rambo-Martin, Shatavia Morrison, Christopher Gulvick, Scott Sammons, Yvette Unoarumhi, Darlene Wagner, Matthew Schmerer,Matthew Tugwell, Lauren Moon, Clinton R. |
| hCoV-19/Turkey/HSGM-B15483/2021 | EPI_ISL_3066689 | 2021-06-03 | Ministry of Health Turkey | Ministry of Health Turkey | Fatma Bayrakdar, Yasemin Cosgun, Suleyman Yalcin, Gulay Korukluoglu |
| hCoV-19/USA/NY-PRL-2021_0719_00B18/2021 | EPI_ISL_3071901 | 2021-07-16 | Pandemic Response Lab - NYC | Pandemic Response Lab, R&D | Henry Lee, Michael Hammerling, Melissa Hopkins, Cybill del Castillo, Shinyoung Clair Kang, William Ward, Pradeep Bugga, Sol Rey, Dylan Law, Isabel Fernandez Escapa, Haiping Hao, Jon Laurent |

| | | | | | |
|---|---|---|---|---|---|
| hCoV-19/Denmark/DCGC-133516/2021 | EPI_ISL_3072363 | 2021-07-19 | Department of Bacteria, Parasites and Fungi, Statens Serum Institut, Copenhagen, Denmark | Statens Serum Institut Bioinformatics and Microbial Genomics | Danish Covid-19 Genome Consortium |
| hCoV-19/SouthAfrica/NICD-N10917/2021 | EPI_ISL_3073961 | 2021-06-28 | MANKWENG LABORATORY | Diseases of the National Health Laboratory Service | Amoako DG, Everatt J, Scheepers C, Mohale T, Ntuli N, Mahlangu B, Mnguni A, Ismail A, Bhiman JN |
| hCoV-19/SouthAfrica/NICD-N11264/2021 | EPI_ISL_3074030 | 2021-07-02 | TAMBO MEMORIAL LABORATORY | Diseases of the National Health Laboratory Service | Amoako DG, Everatt J, Scheepers C, Mohale T, Ntuli N, Mahlangu B, Mnguni A, Ismail A, Bhiman JN |
| hCoV-19/India/AP-CCMB-CIA6058/2021 | EPI_ISL_3076726 | 2021-06-18 | AP SSO | CSIR-Centre for Cellular and Molecular Biology-INSACOG | Ara Sreenivas,Shreekant Verma, Amareshwar Vodapalli ,B Himasri,Valli Nagalakshmi Undamatla,Payel Mukherjee, Lamuk Zaveri, Onkar Kulkarni,Jandhyala Sai Krishna,Tulasi Nagabandi, Sofia Banu, ,Priya Nurkurthy, Vidhyadhari Methuku, Sumedha Avadhanula, Archana |
| hCoV-19/Scotland/CVR8617/2020 | EPI_ISL_3077206 | 2020-12-27 | West of Scotland Specialist Virology Centre, NHSGGC / MRC-University of Glasgow Centre for Virus Research | COVID-19 Genomics UK (COG-UK) Consortium | Ana da Silva Filipe, Natasha Johnson, Kathy Smollett, Daniel Mair, Stephen Carmichael, Alice Broos, Lily Tong, Jenna Nichols, Kyriaki Nomikou; Sarah McDonald; Ioulia Tsatsani, Richard Orton, Joseph Hughes, Sreenu Vattipally, David L Robertson; Alasdair Ma |
| hCoV-19/Wales/PHWC-PYOXC6/2021 | EPI_ISL_3082464 | 2021-07-11 | Originating lab: Wales Specialist Virology Centre Sequencing lab: Pathogen Genomics Unit | Public Health Wales Microbiology Cardiff Wales Specialist Virology Centre | Catherine Moore, Johnathan Evans, Laura Gifford, Malorie Perry, Simon Cottrell, Angela Marchbank, Alec Birchley, Alexander Adams, Amy Gaskin, Bree Gatica-Wilcox, Jason Coombes, Joel Southgate, Lauren Gilbert, Lee Graham, Nicole Pacchiarini, Sara Kumziene- |
| hCoV-19/USA/WI-UW-5381/2021 | EPI_ISL_3087569 | 2021-07-07 | University of Wisconsin-Madison AIDS Vaccine Research Laboratories | University of Wisconsin-Madison AIDS Vaccine Research Laboratories | Gage Moreno, Katarina Braun, et al. AIDS Vaccine Research Laboratories |
| hCoV-19/USA/TN-CDC-ASC210187083/2021 | EPI_ISL_3859781 | 2021-08-10 | Aegis Sciences Corporation | Centers for Disease Control and Prevention Division of Viral Diseases, Pathogen Discovery | Dakota Howard,Dhwani Batra,Peter Cook,Kara Moser,Adrian Paskey,Jason Caravas,Benjamin Rambo-Martin,Shatavia Morrison,Christopher Gulvick,Scott Sammons,Yvette Unoarumhi,Darlene Wagner,Matthew Schmerer,Cyndi Clark,Patrick Campbell,Rob Case,Vikramsinha Ghorp |
| hCoV-19/USA/IL-IDPH-WLS-SV-00367/2021 | EPI_ISL_2960191 | 2021-06-05 | Illinois Department of Public Health | Gagnon Lab, Southern Illinois University | Keith Gagnon |
| hCoV-19/Australia/QLD2045/2021 | EPI_ISL_3090751 | 2021-07-22 | Queensland Health Forensic and Scientific Services | Queensland Health Forensic and Scientific Services | Chenwei-Wang |
| hCoV-19/Australia/QLD2049/2021 | EPI_ISL_3090754 | 2021-07-23 | Queensland Health Forensic and Scientific Services | Queensland Health Forensic and Scientific Services | Chenwei-Wang |
| hCoV-19/Italy/CAM-TIGEM-IZSM-COLLI-17641/2021 | EPI_ISL_3092259 | 2021-07-12 | Cotugno | TIGEM | Antonio Grimaldi Patrizia Annunziata Francesco Panariello Biancamaria Pierri Claudia Tiberio Teresa Giuliano Valentina Bouche Chiara Colantuono Maria Concetta Cuomo Denise Di Concilio Lucio Di Filippo Anna Manfredi Marcello Salvi Antonio Limone Luigi Atri |
| hCoV-19/HongKong/HKPU-11792/2021 | EPI_ISL_3099408 | 2021-05-05 | Department of Health Technology and Informatics, The Hong Kong Polytechnic University | Department of Health Technology and Informatics, The Hong Kong Polytechnic University | Gilman Kit-Hang Siu, Lam-Kwong Lee, Kenneth Siu-Sing Leung, Jake Siu-Lun Leung, Wing-Hei Lo, Timothy Ting-Leung Ng, Chloe Toi-Mei Chan, Hoi-Ching Jim, Ivan Tak-Fai Wong, Kingsley King-Gee Tam, Hiu-Yin Lao, Denise Sze-Hang Wong, Alan Ka-Lun Wu, Miranda Cho |

| | | | | | |
|---|---|---|---|---|---|
| hCoV-19/USA/NV-NSPHL-373898/2021 | EPI_ISL_3102589 | 2021-07-19 | Nevada State Public Health Laboratory | Nevada State Public Health Laboratory | Andrew Gorzalski, Mark Pandori |
| hCoV-19/Brazil/SP-IB_130334/2021 | EPI_ISL_4275643 | 2021-08-30 | UPA TANGARA ESTORIL | Instituto Butantan | Dimas Tadeu Covas, Antonio Jorge Martins, Claudia Renata dos Santos Barros, David Schlesinger, Debora Botequio Moretti, Elaine Cristina Marqueze, Elaine Vieira Santos, Evandra Strazza Rodrigues, Heidge Fukumasu, Jayme Augusto de Souza-Neto, Jos√© Salvator |
| hCoV-19/USA/FL-CDC-LC0095092/2021 | EPI_ISL_3111953 | 2021-07-10 | Laboratory Corporation of America | Centers for Disease Control and Prevention Division of Viral Diseases, Pathogen Discovery | Dakota Howard, Dhwani Batra, Peter W. Cook, Kara Moser, Adrian Paskey, Jason Caravas, Benjamin Rambo-Martin, Shatavia Morrison, Christopher Gulvick, Scott Sammons, Yvette Unoarumhi, Darlene Wagner, Matthew Schmerer, Minoo Agarwal, Eyad Almasri, Debbie Bol |
| hCoV-19/USA/NC-CDC-LC0096114/2021 | EPI_ISL_3112769 | 2021-07-09 | Laboratory Corporation of America | Centers for Disease Control and Prevention Division of Viral Diseases, Pathogen Discovery | Dakota Howard, Dhwani Batra, Peter W. Cook, Kara Moser, Adrian Paskey, Jason Caravas, Benjamin Rambo-Martin, Shatavia Morrison, Christopher Gulvick, Scott Sammons, Yvette Unoarumhi, Darlene Wagner, Matthew Schmerer, Minoo Agarwal, Eyad Almasri, Debbie Bol |
| hCoV-19/USA/OH-CDC-QDX26532313/2021 | EPI_ISL_3114144 | 2021-06-28 | Quest Diagnostics Incorporated | Centers for Disease Control and Prevention Division of Viral Diseases, Pathogen Discovery | Dakota Howard, Dhwani Batra, Peter W. Cook, Kara Moser, Adrian Paskey, Jason Caravas, Benjamin Rambo-Martin, Shatavia Morrison, Christopher Gulvick, Scott Sammons, Yvette Unoarumhi, Darlene Wagner, Matthew Schmerer, S. H. Rosenthal, A. Gerasimova, R. M. K |
| hCoV-19/USA/OH-CDC-QDX26532312/2021 | EPI_ISL_3114580 | 2021-06-28 | Quest Diagnostics Incorporated | Centers for Disease Control and Prevention Division of Viral Diseases, Pathogen Discovery | Dakota Howard, Dhwani Batra, Peter W. Cook, Kara Moser, Adrian Paskey, Jason Caravas, Benjamin Rambo-Martin, Shatavia Morrison, Christopher Gulvick, Scott Sammons, Yvette Unoarumhi, Darlene Wagner, Matthew Schmerer, S. H. Rosenthal, A. Gerasimova, R. M. K |
| hCoV-19/Belgium/UZA-UA-CV2313730488/2021 | EPI_ISL_3115157 | 2021-07-20 | Platform BIS UZA/UAntwerpen | Labo Klinische Biologie, UZA | Marie Le Mercier, Jasmine Coppens, Kathleen Holemans, Ines Verbesselt, Basil Britto Xavier, Christine Lammens, Veerle Matheeussen, Herman Goossens |
| hCoV-19/USA/IN-INSPHL-04261/2021 | EPI_ISL_3115702 | 2021-07-20 | IN State Department of Health Laboratory Services | IN State Department of Health Laboratory Services | Cassandra Campion, Jamie Yeadon, Brian Pope, Lixia Liu, Kyle Brownlee, Melissa Hindenlang, Mark Glazier |
| hCoV-19/Canada/AB-ABPHL-23212/2021 | EPI_ISL_3117393 | 2021-06-19 | Edmonton Provincial Lab | Public Health Agency of Canada (PHAC) National Microbiology Laboratory | Buss, E, Croxen M, Deo A, Dieu P, Gill K, Ferrato C, Khan F, Koleva P, Li V, Lloyd C, Lynch T, Ma R, Murphy S, Pabbaraju K, Shokoples S, Tipples G, Thayer J, Whitehouse M, Wong A, Yu C, Zelyas N |
| hCoV-19/Ireland/LK-NVRL-S21IRL34538/2021 | EPI_ISL_3118150 | 2021-07-01 | National Virus Reference Laboratory | National Virus Reference Laboratory | Zoe Yandle, Charlene Bennett, Gabriel Gonzalez, Michael Carr, Jonathan Dean, Cillian F De Gascun |
| hCoV-19/Austria/KILM994/2021 | EPI_ISL_3119352 | 2021-07-17 | Lifebrain Covid Labor GmbH | Dept. of Laboratory Medicine | Harald Esterbauer, Anna Gschaider, Petra Jurkowitsch, Fabian Konig, Sabina Plumer, Robert Strassl, Claudia Weber, Oswald Wagner |
| hCoV-19/Russia/IVA-RII-MH28057S/2021 | EPI_ISL_3122979 | 2021-06-14 | City Clinical Hospital No 1 | WHO National Influenza Centre Russian Federation | Andrey Komissarov, Artem Fadeev, Kseniya Komissarova, Oula Mansour, Kirill Varchenko, Mikhail Bakaev, Tamila Musaeva, Veronika Eder, Maria Pisareva, Nikita Yolshin, Daria Danilenko, Maria Baturova, Alexey Masharsky, Ksenia Safina, Elena Nabieva, Georgii B |

| | | | | | |
|---|---|---|---|---|---|
| hCoV-19/Austria/CeMM12948/2021 | EPI_ISL_3123140 | 2021-06-28 | Austrian Agency for Health and Food Safety (AGES) | Bergthaler laboratory, CeMM Research Center for Molecular Medicine of the Austrian Academy of Sciences | Lukas Endler, Anna Schedl, Fabian Amman, Petr Triska, Matthew Thornton, Thomas Penz, Benedikt Agerer, Maelle Le Moing, Michael Schuster, Bekir Erguner, Jan Laine, Martin Senekowitsch, Christoph Bock, Andreas Bergthaler |
| hCoV-19/France/NAQ-HCL021126558801/2021 | EPI_ISL_3130446 | 2021-05-06 | LABORATOIRE FORTE BIO DAX | CNR Virus des Infections Respiratoires - France SUD | Antonin Bal, Gregory Destras, Gwendolyne Burfin, Hadrien Regue, Quentin Semanas, Martine Valette, Bruno Lina, Laurence Josset |
| hCoV-19/USA/WI-WSLH-215688/2021 | EPI_ISL_3131911 | 2021-06-10 | Wisconsin State Laboratory of Hygiene Communicable Disease Division | Wisconsin State Laboratory of Hygiene Communicable Disease Division | Abigail C. Shockey, Alicia J. Mooney, Erika M. Hanson, Tonya Danz, Richard Griesser, Sara Wagner, Kelsey R. Florek |
| hCoV-19/Gambia/49243/2021 | EPI_ISL_3132334 | 2021-07-10 | MRCG at LSHTM Genomics lab | MRCG at LSHTM Genomics lab | Abdul Karim sesay, Abdoulie Kanteh, Jarra Manneh, Mariama Kujabi, Bakary Sanyang,Dabiri Damilari |
| hCoV-19/USA/UT-UPHL-210624490751/2021 | EPI_ISL_3132910 | 2021-01-04 | Utah Public Health Laboratory | Utah Public Health Laboratory | Olinto Linares-Perdomo, Erin L. Young, Kelly F. Oakeson |
| hCoV-19/Ecuador/USFQ-1711/2021 | EPI_ISL_3133383 | 2021-07-16 | Institute of Microbiology, Universidad San Francisco de Quito | Institute of Microbiology, Universidad San Francisco de Quito | Sully M√°rquez, Juan Jos√© Guadalupe, Bel√©n Prado-Vivar, Monica Becerra-Wong, Fernanda Zurita, Erika Mu√±oz, Bernardo Guti√©rrez, Ver√≥nica Barrag√°n, Patricio Rojas-Silva, Gabriel Trueba, Michelle Grunauer, Pa√∫l C√°rdenas |
| hCoV-19/USA/IL-S21WGS2563/2021 | EPI_ISL_3133476 | 2021-07-14 | Illinois Department of Public Health - Springfield Lab | Illinois Department of Public Health - Springfield Lab | Bryan Sim, Gordon McCall |
| hCoV-19/Curacao/CW-RIVM-44546/2021 | EPI_ISL_3136618 | 2021-07-08 | Dutch COVID-19 response team | National Institute for Public Health and the Environment (RIVM) | Adam Meijer, Harry Vennema, Dirk Eggink, Jeroen Cremer, Sharon van den Brink, Bas van der Veer, AnneMarie van den Brandt, Lisa Wijsman, Kim Freriks, Ryanne Jaarsma, Eun√≠ce Then, Lynn Aarts, Sanne Bos, Melissa van Tuil, Florian Zwagemaker, Dennis Schmitz, |
| hCoV-19/Luxembourg/LNS9351900/2021 | EPI_ISL_3147643 | 2021-07-03 | Hospital Center Emile Mayrisch | Laboratoire national de sante, Microbiology, Microbial Genomics Platform | Anke Wienecke-Baldacchino, Catherine Ragimbeau,Jessica Tapp, Fatu Djabi, Lise Pignon, Raoul Salmon, Virginie Jover, Elodie Solarino, Cynthia Oxacelay, Tamir Abdelrahman |
| hCoV-19/Uganda/UG651/2021 | EPI_ISL_3149436 | 2021-06-14 | MRC/UVRI & LSHTM Uganda Research Unit, Uganda Virus Research Institute | Research Unit, Uganda Virus Research Institute | Matthew Cotten, Dan Lule Bugembe, My V.T Phan, Pontiano Kaleebu, John Kayiwa, |
| hCoV-19/Uganda/UG661/2021 | EPI_ISL_3149441 | 2021-06-14 | MRC/UVRI & LSHTM Uganda Research Unit, Uganda Virus Research Institute | Research Unit, Uganda Virus Research Institute | Matthew Cotten, Dan Lule Bugembe, My V.T Phan, Pontiano Kaleebu, John Kayiwa, |
| hCoV-19/Gambia/46728/2021 | EPI_ISL_3150942 | 2021-06-30 | MRCG at LSHTM Genomics lab | MRCG at LSHTM Genomics lab | Abdul Karim sesay, Abdoulie Kanteh, Jarra Manneh, Mariama Kujabi, Bakary Sanyang,Dabiri Damilari |
| hCoV-19/Gambia/46581/2021 | EPI_ISL_3150943 | 2021-06-29 | MRCG at LSHTM Genomics lab | MRCG at LSHTM Genomics lab | Abdul Karim sesay, Abdoulie Kanteh, Jarra Manneh, Mariama Kujabi, Bakary Sanyang,Dabiri Damilari |
| hCoV-19/USA/CT-PHL-1023443001/2021 | EPI_ISL_3151600 | 2021-07-19 | CT Department of Public Health | CT Department of Public Health | Claire_Pearson,Tu_N_Nguyen |
| hCoV-19/Senegal/SC20-504/2020 | EPI_ISL_3152079 | 2020-08-28 | MRCG | LBV Le Dantec | Halimatou Diop Ndiaye, Assane Dieng, Adjiratou Aissatou BA, Jarra Manneh, Abdul Karim Cesay, , Khadim Gueye, Abdoulie Kante, Mbengu√© Fall, Pauline Yacine Sene, Oumy DIOP, Sada Diallo, Anna julienne Selbe NDiaye, Pasacaline Manga, Dianke Samat√©, Aminata |

| | | | | | |
|---|---|---|---|---|---|
| hCoV-19/Senegal/SC20-507/2020 | EPI_ISL_3152080 | 2020-08-02 | MRCG | LBV Le Dantec | Halimatou Diop Ndiaye, Assane Dieng, Adjiratou Aissatou BA, Jarra Manneh, Abdul Karim Cesay, , Khadim Gueye, Abdoulie Kante, Mbenguv© Fall, Pauline Yacine Sene, Oumy DIOP, Sada Diallo, Anna julienne Selbe NDiaye, Pasacaline Manga, Dianke Samatv©, Aminata |
| hCoV-19/Reunion/PIMIT_15861/2021 | EPI_ISL_3153870 | 2021-07-12 | CERBALLIANCE | UMR PIMIT | Dr David A Wilkinson, Dr Patrick Mavingui, Dr Camille Lebarbenchon, Magali Turpin, Marie-Alice Simbi |
| hCoV-19/Senegal/SN-IR1-39599/2021 | EPI_ISL_3154886 | 2021-05-31 | Iressef Genomics lab | IRESSEF | Souleymane MBOUP, Abdou PADANE , Khadim GUEYE, Diabou Diagne , Samba Ndiour, Papa Alassane DIAW, Birahim Piere NDIAYE, Barada CISSE, Aminata MBOUP, Moustapha MBOW, Ndeye Coumba Toure KANE, Nafisatou LEYE, Gora LO, Ambroise AHOUIDI , Astou Gaye GAYE, Amina |
| hCoV-19/Senegal/SN-IRFSFOU-32_bis/2021 | EPI_ISL_3154893 | 2021-04-04 | Iressef Genomics lab | IRESSEF | Souleymane MBOUP, Abdou PADANE , Khadim GUEYE, Diabou Diagne , Samba Ndiour, Papa Alassane DIAW, Birahim Piere NDIAYE, Barada CISSE, Aminata MBOUP, Moustapha MBOW, Ndeye Coumba Toure KANE, Nafisatou LEYE, Gora LO, Ambroise AHOUIDI , Astou Gaye GAYE, Amina |
| hCoV-19/Senegal/SN-RUF-216276/2021 | EPI_ISL_3154898 | 2021-06-01 | Iressef Genomics lab | IRESSEF | Souleymane MBOUP, Abdou PADANE , Khadim GUEYE, Diabou Diagne , Samba Ndiour, Papa Alassane DIAW, Birahim Piere NDIAYE, Barada CISSE, Aminata MBOUP, Moustapha MBOW, Ndeye Coumba Toure KANE, Nafisatou LEYE, Gora LO, Ambroise AHOUIDI , Astou Gaye GAYE, Amina |
| hCoV-19/France/IDF-IPP16235/2021 | EPI_ISL_3154990 | 2021-07-13 | Labo Analyses Med | National Reference Center for Viruses of Respiratory Infections, Institut Pasteur, Paris | Marion Barbet, Sylvie Behillil, Mv©line Bizard, Angela Brisebarre, Camille Capel, Vincent Enouf, Louise Lefranv ßois, Frv©dv©ric Lemoine, Christophe Malabat, Corinne Maufrais, Hub Bioinformatique Biostatistiques, Etienne Simon-Loriv®re, Maud Vanpeene, Sylv |
| hCoV-19/France/IDF-IPP16455/2021 | EPI_ISL_3155069 | 2021-07-13 | Hospital | National Reference Center for Viruses of Respiratory Infections, Institut Pasteur, Paris | Marion Barbet, Sylvie Behillil, Mv©line Bizard, Angela Brisebarre, Camille Capel, Vincent Enouf, Louise Lefranv ßois, Frv©dv©ric Lemoine, Christophe Malabat, Corinne Maufrais, Hub Bioinformatique Biostatistiques, Etienne Simon-Loriv®re, Maud Vanpeene, Sylv |
| hCoV-19/Sweden/20502880S/2021 | EPI_ISL_3159146 | 2021-06-30 | Swedish national genomic surveillance program of SARS-CoV-2 | The Public Health Agency of Sweden | Maximilian Riess, Maria Lind Karlberg, Alma Brolund, Swedish national genomic surveillance program of SARS-CoV-2 |
| hCoV-19/SouthAfrica/KRISP-K019774/2021 | EPI_ISL_3161771 | 2021-07-02 | NHLS_VIRO | KRISP, KZN Research Innovation and Sequencing Platform | Giandhari Jennifer, Pillay Sureshnee, Yajna Ramphal, Naidoo Yeshnee, Tshabuila Derek,Tegally Houriiyah, San James, Wilkinson Eduan, de Oliveira Tulio |
| hCoV-19/Algeria/36420/2021 | EPI_ISL_3161807 | 2021-05-10 | NIC, Viral Respiratory Unit | Virology Departement | Alia Grad, Fayez Khardine, Amel Benyahia, Fetouma Doudou, Nardjes Hihi, Mohamed Amine Beloufa, Aissam Hachid, Aicha Bensalem, Fawzi Derrar |
| hCoV-19/Australia/QLD2057/2021 | EPI_ISL_3161814 | 2021-07-27 | Queensland Health Forensic and Scientific Services | Queensland Health Forensic and Scientific Services | Chenwei Wang on behalf of Q-PHIRE Genomics |

| | | | | | |
|---|---|---|---|---|---|
| hCoV-19/Australia/QLD2059/2021 | EPI_ISL_3161816 | 2021-07-27 | Queensland Health Forensic and Scientific Services | Queensland Health Forensic and Scientific Services | Chenwei Wang on behalf of Q-PHIRE Genomics |
| hCoV-19/USA/RI-RISHL-E01375/2021 | EPI_ISL_3161953 | 2021-07-13 | Rhode Island State Health Laboratory | Rhode Island State Health Laboratory | Ewa King, Richard C. Huard, Kristin Carpenter-Azevedo |
| hCoV-19/NewZealand/21MV0602/2021 | EPI_ISL_3164090 | 2021-07-15 | LabPLUS | Institute of Environmental Science and Research (ESR) | Rachel Boyle, SallyAnn Harbison, Olivia Stroeven, Xiaoyun Ren, Matt Storey, Nikki Freed, Muhammad Faisal, Jing Wang, Hermes Perez, Anja Werno, Antje van der Linden, Arlo Upton, Chris Mansell, David Hammer, Dragana Drinkovic, Gary McAuliffe, Hana Sofia And |
| hCoV-19/Australia/NSW-ICPMR-541/2021 | EPI_ISL_3185119 | 2021-07-18 | St Vincent's Pathology (SydPath) | Clinical Pathology and Medical Research; Westmead Hospital; University of Sydney | CIDM-PH et al. |
| hCoV-19/Australia/NSW-ICPMR-510/2021 | EPI_ISL_3185236 | 2021-07-17 | St Vincent's Pathology (SydPath) | Clinical Pathology and Medical Research; Westmead Hospital; University of Sydney | CIDM-PH et al. |
| hCoV-19/Chile/RM-125188/2021 | EPI_ISL_3185660 | 2021-07-09 | Genetica Molecular and Subdepartamento de Virologia ISP Chile | Instituto de Salud Publica de Chile | Karen Orostica, Constanza Campano, Barbara Parra, Loredana Arata, Gisselle Barra, Patricia Bustos, Rodrigo Fasce, Javier Tognarelli, Andres Castillo, Soledad Ulloa, Jorge Fernandez |
| hCoV-19/Sweden/1469449864/2021 | EPI_ISL_3186394 | 2021-07-13 | Sweden/Sodermanland/Unilabs | Unilabs/Eskilstuna/Sweden | Emma Arvidsson |
| hCoV-19/Australia/NSW-ICPMR-850/2021 | EPI_ISL_3186850 | 2021-07-22 | Medlab Pathology | Clinical Pathology and Medical Research; Westmead Hospital; University of Sydney | Arnott A., Draper J., Gall M., Martinez E., Rockett R., Sintchenko V., on behalf of ICPMR |
| hCoV-19/India/AS-1829000369826/2021 | EPI_ISL_3189070 | 2021-06-18 | INSACOG-Assam | National Institute of Biomedical Genomics ‚Äì INSACOG | Arindam Maitra, Ajanta Sharma, Nidhan Kumar Biswas, Lahari Saikia, Sreedhar Chinnaswamy, Kailash Chamuah, Saumitra Das |
| hCoV-19/India/WB-1931300402439/2021 | EPI_ISL_3189176 | 2021-07-03 | INSACOG-WB | National Institute of Biomedical Genomics ‚Äì INSACOG | Arindam Maitra, Bhaswati Bandyopadhyay, Nidhan Kumar Biswas, Tamal Ghosh, Sreedhar Chinnaswamy, Ajay Chakraborti, Saumitra Das |
| hCoV-19/India/WB-1931300404411/2021 | EPI_ISL_3189201 | 2021-07-05 | INSACOG-WB | National Institute of Biomedical Genomics ‚Äì INSACOG | Arindam Maitra, Bhaswati Bandyopadhyay, Nidhan Kumar Biswas, Tamal Ghosh, Sreedhar Chinnaswamy, Ajay Chakraborti, Saumitra Das |
| hCoV-19/Scotland/QEUH-1A213A9/2021 | EPI_ISL_3201446 | 2021-07-26 | Lighthouse Lab in Glasgow | Wellcome Sanger Institute for the COVID-19 Genomics UK (COG-UK) Consortium | Harper VanSteenhouse, Yumi Kasai, David Gray, Carol Clugston, Anna Dominiczak and Alex Alderton, Roberto Amato, Jeffrey Barrett, Sonia Goncalves, Ewan Harrison, David K. Jackson, Ian Johnston, Dominic Kwiatkowski, Cordelia Langford, John Sillitoe on behal |
| hCoV-19/England/MILK-1A2C3AE/2021 | EPI_ISL_3203410 | 2021-07-26 | Lighthouse Lab in Milton Keynes | Wellcome Sanger Institute for the COVID-19 Genomics UK (COG-UK) Consortium | The Lighthouse Lab in Milton Keynes and Alex Alderton, Roberto Amato, Jeffrey Barrett, Sonia Goncalves, Ewan Harrison, David K. Jackson, Ian Johnston, Dominic Kwiatkowski, Cordelia Langford, John Sillitoe on behalf of the Wellcome Sanger Institute COVID-1 |

| | | | | | |
|---|---|---|---|---|---|
| hCoV-19/USA/CA-CDC-FG-047707/2021 | EPI_ISL_3212634 | 2021-07-22 | Fulgent Genetics | Centers for Disease Control and Prevention Division of Viral Diseases, Pathogen Discovery | Dakota Howard, Dhwani Batra, Peter W. Cook, Kara Moser, Adrian Paskey, Jason Caravas, Benjamin Rambo-Martin, Shatavia Morrison, Christopher Gulvick, Scott Sammons, Yvette Unoarumhi, Darlene Wagner, Matthew Schmerer, Harry Gao, Mickey Li, John Gao, Joseph |
| hCoV-19/Italy/EMR-C036_21_25/2021 | EPI_ISL_3215182 | 2021-06-03 | U.O. Microbiologia Laboratorio Unico Centro Servizi - AUSL della Romagna | U.O. Microbiologia, Laboratorio Unico Centro Servizi - AUSL della Romagna | Giorgio Dirani |
| hCoV-19/USA/FL-CDC-ASC210112444/2021 | EPI_ISL_3218720 | 2021-07-08 | Aegis Sciences Corporation | Centers for Disease Control and Prevention Division of Viral Diseases, Pathogen Discovery | Dakota Howard, Dhwani Batra, Peter W. Cook, Kara Moser, Adrian Paskey, Jason Caravas, Benjamin Rambo-Martin, Shatavia Morrison, Christopher Gulvick, Scott Sammons, Yvette Unoarumhi, Darlene Wagner, Matthew Schmerer, Cyndi Clark, Patrick Campbell, Rob Case |
| hCoV-19/USA/CA-CDC-ASC210155168/2021 | EPI_ISL_3220549 | 2021-06-30 | Aegis Sciences Corporation | Centers for Disease Control and Prevention Division of Viral Diseases, Pathogen Discovery | Dakota Howard, Dhwani Batra, Peter W. Cook, Kara Moser, Adrian Paskey, Jason Caravas, Benjamin Rambo-Martin, Shatavia Morrison, Christopher Gulvick, Scott Sammons, Yvette Unoarumhi, Darlene Wagner, Matthew Schmerer, Cyndi Clark, Patrick Campbell, Rob Case |
| hCoV-19/Denmark/DCGC-140816/2021 | EPI_ISL_3228989 | 2021-07-28 | Department of Bacteria, Parasites and Fungi, Statens Serum Institut, Copenhagen, Denmark | Statens Serum Institut Bioinformatics and Microbial Genomics | Danish Covid-19 Genome Consortium |
| hCoV-19/Slovakia/UVZ_PL25_C1_13994/2021 | EPI_ISL_3229335 | 2021-04-01 | Public Health Authority of the Slovak Republic | Laboratory of Genomics and Bioinformatics, Comenius University Science Park | Tatiana Sedl√°ƒçkov√°, Diana Rus≈à√°kov√°, Miroslav B√∂hmer, Anna Gifçov√°, Jaroslav Budi≈°, Tom√°≈° Szemes, Jakub Styk |
| hCoV-19/Bulgaria/21BG-EU_003858_Pl33/2021 | EPI_ISL_3232676 | 2021-07-13 | National Center of Infectious and Parasitic Diseases | National Center of Infectious and Parasitic Diseases | Alexiev et al |
| hCoV-19/India/KA-NIMH-SEQ-2188/2021 | EPI_ISL_3232885 | 2021-07-13 | MMCRI | INSACOG-KA, NIMHANS | Chitra Pattabiraman, Anson Kunjumon George,Darshan Sreenivas, Ellango Ramasamy, Sony Sharma, Mahesh Kumar.C.S, Ananthapadmanabha Kotambail, Chetan G K, Gautham Arunachal Udupi, Anita S Desai, V Ravi |
| hCoV-19/SouthAfrica/NICD-N11609/2021 | EPI_ISL_3236978 | 2021-07-05 | TSHEPONG LABORATORY | Diseases of the National Health Laboratory Service | Amoako DG, Everatt J, Scheepers C, Mohale T, Ntuli N, Mahlangu B, Mnguni A, Ismail A, Bhiman JN |
| hCoV-19/SaudiArabia/DM-21-107803A/2021 | EPI_ISL_3237050 | 2021-04-21 | King Faisal Specialist Hospital & Research Centre | King Faisal Specialist Hospital & Research Centre | Abeer N Alshukairi, Waseem A Al Mousa, Rayan Bawayan, Mohamad K Al Hroub, Ashraf Dada |
| hCoV-19/Denmark/DCGC-141835/2021 | EPI_ISL_3240702 | 2021-07-31 | Department of Bacteria, Parasites and Fungi, Statens Serum Institut, Copenhagen, Denmark | Statens Serum Institut Bioinformatics and Microbial Genomics | Danish Covid-19 Genome Consortium |
| hCoV-19/Guam/GU-CDC-2-4651479/2021 | EPI_ISL_3241995 | 2021-07-07 | Guam Public Health Laboratory | Centers for Disease Control and Prevention Division of Viral Diseases, Pathogen Discovery | Mili Sheth, Sarah Nobles, Jasmine Padilla, Alex Burgin, Meghan Bentz, Mark Burroughs, Peter Cook, Clinton R. Paden, Dhwani Batra, Krista Queen, Kristen Knipe, Dakota Howard, Yvette Unoarumhi, Matthew Schmerer, Ben L. Rambo-Martin, Kristine Lacek, Sam Shep |
| hCoV-19/India/GA-CDFD-C53232/2021 | EPI_ISL_3242315 | 2021-06-08 | Goa Medical College | CDFD | Murali Bashyam, Asmita Gupta, Reelina Basu, Vinay Donipadi, Savio Rodrigues, Divya Vashisht, Ashwin Dalal |
| hCoV-19/India/GA-CDFD-C55264/2021 | EPI_ISL_3242383 | 2021-06-27 | Goa Medical College | CDFD | Murali Bashyam, Asmita Gupta, Reelina Basu, Vinay Donipadi, Savio Rodrigues, Divya Vashisht, Ashwin Dalal |

| | | | | | |
|---|---|---|---|---|---|
| hCoV-19/Chile/RM-UNAB-1237/2021 | EPI_ISL_3242492 | 2021-07-02 | Clinica INDISA | Facultad de Ciencias de la Vida, UNAB | Claudio Meneses, Eduardo Castro, Waldo Diaz, Macarena Bastias, Ricardo Yusta, Susana Saez, Dayan Sanhueza, Sebastian Wolter, Daniel Leon, Gonzalo Campa√±a, Paola Pidal, V√≠ctor Monreal, Claudio Olmos, Ariel Orellana |
| hCoV-19/Chile/RM-UNAB-1202/2021 | EPI_ISL_3242547 | 2021-06-29 | Clinica INDISA | Facultad de Ciencias de la Vida, UNAB | Claudio Meneses, Eduardo Castro, Waldo Diaz, Macarena Bastias, Ricardo Yusta, Susana Saez, Dayan Sanhueza, Sebastian Wolter, Daniel Leon, Gonzalo Campa√±a, Paola Pidal, V√≠ctor Monreal, Claudio Olmos, Ariel Orellana |
| hCoV-19/Chile/RM-UNAB-1154/2021 | EPI_ISL_3242560 | 2021-06-23 | Clinica INDISA | Facultad de Ciencias de la Vida, UNAB | Claudio Meneses, Eduardo Castro, Waldo Diaz, Macarena Bastias, Ricardo Yusta, Susana Saez, Dayan Sanhueza, Sebastian Wolter, Daniel Leon, Gonzalo Campa√±a, Paola Pidal, V√≠ctor Monreal, Claudio Olmos, Ariel Orellana |
| hCoV-19/Chile/RM-UNAB-0263-04987181/2021 | EPI_ISL_3242563 | 2021-05-07 | Clinica INDISA | Facultad de Ciencias de la Vida, UNAB | Claudio Meneses, Eduardo Castro, Waldo Diaz, Macarena Bastias, Ricardo Yusta, Susana Saez, Dayan Sanhueza, Sebastian Wolter, Daniel Leon, Gonzalo Campa√±a, Paola Pidal, V√≠ctor Monreal, Claudio Olmos, Ariel Orellana |
| hCoV-19/Chile/RM-UNAB-0263-05190500/2021 | EPI_ISL_3242599 | 2021-06-21 | Clinica INDISA | Facultad de Ciencias de la Vida, UNAB | Claudio Meneses, Eduardo Castro, Waldo Diaz, Macarena Bastias, Ricardo Yusta, Susana Saez, Dayan Sanhueza, Sebastian Wolter, Daniel Leon, Gonzalo Campa√±a, Paola Pidal, V√≠ctor Monreal, Claudio Olmos, Ariel Orellana |
| hCoV-19/USA/NY-APDN-S2374/2021 | EPI_ISL_3243023 | 2021-05-18 | Applied DNA Sciences Inc | Applied DNA Sciences Inc | Huan Dong; Spencer William; Lucas Liu; Yuhua Sun |
| hCoV-19/India/TG-CDFD-Xi577/2021 | EPI_ISL_3243223 | 2021-06-22 | CDFD | CDFD | Murali Bashyam, Asmita Gupta, Reelina Basu, Vinay Donipadi, Divya Vashisht, Ashwin Dalal |
| hCoV-19/India/TG-CDFD-W48/2021 | EPI_ISL_3243278 | 2021-06-04 | Gandhi Hospital | CDFD | Murali Bashyam, Asmita Gupta, Reelina Basu, Vinay Donipadi, Nagamani Kammili, Divya Vashisht, Ashwin Dalal |
| hCoV-19/USA/CA-CDPH-2000048639/2021 | EPI_ISL_3244422 | 2021-05-21 | CDPH VBL | California Department of Public Health | Emily Smith on behalf of CDPH-COVIDNet and UCSD EXCITE lab |
| hCoV-19/India/MH-ICMR-NIV-INSACOG-GSEQ-4728/2021 | EPI_ISL_3244655 | 2021-07-10 | ICMR-National Institute of Virology - INSACOG | NIV Influenza | Potdar, Varsha on behalf of National Influenza Centre, NIV, Pune |
| hCoV-19/Brazil/SP-LNN03648/2021 | EPI_ISL_3245342 | 2021-06-30 | Laboratorio Central Noel Nutels | Bioinformatics Laboratory / LNCC | Luiz G P de Almeida, Alessandra P Lamarca, Ronaldo da Silva F Jr, Liliane Cavalcante, Alexandra L Gerber, Ana Paula de C Guimaraes, Douglas Terra Machado, Erica Ramos dos Santos Nascimento, Fernanda Leitao dos Santos, Ricardo Jose Barbosa Salviano, Cintia |
| hCoV-19/USA/NY-NYCPHL-005020/2021 | EPI_ISL_3254239 | 2021-07-05 | Quest Diagnostics | New York City Public Health Laboratory | Jade Wang, et al. |
| hCoV-19/Curacao/CW-RIVM-46574/2021 | EPI_ISL_3257914 | 2021-07-14 | Dutch COVID-19 response team | National Institute for Public Health and the Environment (RIVM) | Adam Meijer, Harry Vennema, Dirk Eggink, Jeroen Cremer, Sharon van den Brink, Bas van der Veer, AnneMarie van den Brandt, Lisa Wijsman, Kim Freriks, Ryanne Jaarsma, Eun√≠ce Then, Lynn Aarts, Sanne Bos, Melissa van Tuil, Florian Zwagemaker, Dennis Schmitz, |

| | | | | | |
|---|---|---|---|---|---|
| hCoV-19/Netherlands/DR-RIVM-47648/2021 | EPI_ISL_3258702 | 2021-07-20 | Dutch COVID-19 response team | National Institute for Public Health and the Environment (RIVM) | Adam Meijer, Harry Vennema, Dirk Eggink, Jeroen Cremer, Sharon van den Brink, Bas van der Veer, AnneMarie van den Brandt, Lisa Wijsman, Kim Freriks, Ryanne Jaarsma, Eun√≠ce Then, Lynn Aarts, Sanne Bos, Melissa van Tuil, Florian Zwagemaker, Dennis Schmitz, |
| hCoV-19/USA/UT-UPHL-210729652440/2020 | EPI_ISL_3263174 | 2020-12-16 | Utah Public Health Laboratory | Utah Public Health Laboratory | Erin L. Young, Kelly F. Oakeson, Olinto Linares-Perdomo |
| hCoV-19/India/PY-34785_S351_R1_001/2021 | EPI_ISL_3265298 | 2021-05-17 | JIPMER Puducherry | inStem NCBS ,Äì INSACOG | Uma Ramakrishnan Dasaradhi Palakodeti Aswin SaiNarain |
| hCoV-19/USA/FL-CDC-QDX25891100/2021 | EPI_ISL_4372485 | 2021-06-14 | Quest Diagnostics Incorporated | Centers for Disease Control and Prevention Division of Viral Diseases, Pathogen Discovery | Dakota Howard,Dhwani Batra,Peter Cook,Jason Caravas,Benjamin Rambo-Martin,Scott Sammons,Yvette Unoarumhi,Matthew Schmerer,Kristine Lacek,Tymeckia Kendall,Victoria Caban Figueroa,Shatavia Morrison,Christopher Gulvick,Erisa Sula,S. Rosenthal,A. Gerasimova,R |
| hCoV-19/India/RJ-IGIB-B042519/2021 | EPI_ISL_3267272 | 2021-06-10 | INSACOG Surveillance | INSACOG at CSIR Institute of Genomics and Integrative Biology | INSACOG |
| hCoV-19/Ecuador/UEES-INTERLAB-6879811/2021 | EPI_ISL_3267867 | 2021-07-30 | INTERLAB | Omics Sciences Laboratory | Derly Andrade Molina, Gabriel Morey Le√≥n, Darlyn Amaya, Rub√©n Armas Gonz√°lez, Katheryn Sacheri, Daniela Bravo Prieto, Elena Romero Cevallos, Esteban Fern√°ndez, Karen Mu√±oz, Juan Carlos Fern√°ndez Cadena |
| hCoV-19/Ghana/WACCBIP-GS1017/2021 | EPI_ISL_3268043 | 2021-06-22 | WACCBIP, University of Ghana, Accra, Ghana | WACCBIP, University of Ghana, Volta Road, Legon-Accra, Ghana | Collins M. Morang'a, Joyce M. Ngoi, Dominic S. Y. Amuzu, Vincent Appiah, Evelyn B. Quansah, Samirah Saiid, Bright K. Yemi, Israel Osei-Wusu, Deborah N. A. Mettle, Philip M. Soglo, Violette V. M,Äôcormack, Paul Owusu-Oduro, Samuel Armoo, Oliver Commey, Syl |
| hCoV-19/Ghana/WACCBIP-GS1130/2021 | EPI_ISL_3268168 | 2021-07-06 | WACCBIP, University of Ghana, Accra, Ghana | WACCBIP, University of Ghana, Volta Road, Legon-Accra, Ghana | Collins M. Morang'a, Joyce M. Ngoi, Dominic S. Y. Amuzu, Vincent Appiah, Evelyn B. Quansah, Samirah Saiid, Bright K. Yemi, Israel Osei-Wusu, Deborah N. A. Mettle, Philip M. Soglo, Violette V. M,Äôcormack, Paul Owusu-Oduro, Samuel Armoo, Oliver Commey, Syl |
| hCoV-19/Ghana/WACCBIP-GS1199/2021 | EPI_ISL_3268172 | 2021-07-12 | WACCBIP, University of Ghana, Accra, Ghana | WACCBIP, University of Ghana, Volta Road, Legon-Accra, Ghana | Collins M. Morang'a, Joyce M. Ngoi, Dominic S. Y. Amuzu, Vincent Appiah, Evelyn B. Quansah, Samirah Saiid, Bright K. Yemi, Israel Osei-Wusu, Deborah N. A. Mettle, Philip M. Soglo, Violette V. M,Äôcormack, Paul Owusu-Oduro, Samuel Armoo, Oliver Commey, Syl |
| hCoV-19/FrenchGuiana/IPG2021800/2021 | EPI_ISL_3269614 | 2021-07-28 | CH de l'Ouest Guyanais Frank Joly | Institut Pasteur de la Guyane | Anne Lavergne, Dominique Rousset, Antoine Enfissi, Arielle Salmier |
| hCoV-19/Ecuador/NIC-INSPI-340883/2021 | EPI_ISL_3274106 | 2021-07-15 | INSPI-CRN DE INFLUENZA Y OTROS VIRUS RESPIRATORIOS | NIC-INSPI | Alfredo Bruno, Maritza Olmedo, Michelle P√°ez, Jimmy Garc√©s, Johanna Laines, Lizbeth Pati√±o, Manuel Gonzalez, Domenica de Mora. |
| hCoV-19/Ecuador/NIC-INSPI-342544/2021 | EPI_ISL_3274571 | 2021-07-21 | INSPI-CRN DE INFLUENZA Y OTROS VIRUS RESPIRATORIOS | NIC-INSPI | Alfredo Bruno , Maritza Olmedo, Michelle P√°ez, Jimmy Garc√©s, Johanna Laines, Lizbeth Pati√±o, Manuel Gonzalez, Domenica de Mora. |

| | | | | | |
|---|---|---|---|---|---|
| hCoV-19/USA/ID-UNM-IBL722038/2021 | EPI_ISL_3275107 | 2021-04-06 | Idaho Bureau of Laboratories | Center for Global Health, University of New Mexico Health Sciences Center | Daryl Domman, Kurt Schwalm, Valerie Morley, Matthew Burns, Robert Voermans, Christopher Ball, Darrell Dinwiddie |
| hCoV-19/USA/ID-UNM-IBL720559/2021 | EPI_ISL_3275133 | 2021-03-27 | Idaho Bureau of Laboratories | Center for Global Health, University of New Mexico Health Sciences Center | Daryl Domman, Kurt Schwalm, Valerie Morley, Matthew Burns, Robert Voermans, Christopher Ball, Darrell Dinwiddie |
| hCoV-19/Belgium/CHUNamur13529659/2021 | EPI_ISL_3275674 | 2021-08-03 | Plateforme de testing Namuroise | Plateforme de testing Namuroise | Lesly Nyinkeu Kemamen; Nicolas Gilliard; Demars Aurore ; Maschietto C√©line ; Nobis Chlo√© ; Otto Gaetan ; Denis Olivier ; Degosserie Jonathan ; Mullier Fran√ßois |
| hCoV-19/India/TN-SS-63_S258_R1_001/2021 | EPI_ISL_3275990 | 2021-05-23 | Dr S Raju, Director of Public Health and Preventive Medicine | inStem NCBS ,Äì INSACOG | Uma Ramakrishnan Dasaradhi Palakodeti Aswin SaiNarain |
| hCoV-19/India/TN-SS-253_S67_R1_001/2021 | EPI_ISL_3276069 | 2021-06-12 | Dr S raju, Director of public heath and preventive medicine | inStem NCBS ,Äì INSACOG | Uma Ramakrishnan Dasaradhi Palakodeti Aswin SaiNarain |
| hCoV-19/India/KA-144399_2_S289_R1_001/2021 | EPI_ISL_3276269 | 2021-05-19 | Kodigehalli PHC | inStem NCBS ,Äì INSACOG | Uma Ramakrishnan Dasaradhi Palakodeti Aswin SaiNarain |
| hCoV-19/Colombia/ANT-CWOHC-VG-PSEC01139U/2021 | EPI_ISL_3276577 | 2021-04-16 | Ayudas Diagnosticas Sura | Universidad Nacional de Colombia - Laboratorio Gen√≥mico One Health | Maria Stella L√≥pez, Andres F. Cardona-Rios, Daniel O. Maldonado-Perez, Sim√≥n Villegas Vel√°squez, Lina M. Hurtado, Laura Silvana Perez, Karl A Ciuoderis, Maria Ang√©lica Maya, Carolina Mu√±oz-Arango, Celeny Ortiz, Rita Almanza Payares, Idabely Betancur |
| hCoV-19/India/DL-ILBS-17824/2020 | EPI_ISL_3276880 | 2020-06-05 | Clinical Virology Laboratory, Institute of Liver and Biliary Sciences | ILBS | Ekta Gupta, Varun Suroliya, Diptanu Paul, Amit Pandey, Reshu Agarwal, Chhagan Bihari Sharma, Shiv Kumar Sarin |
| hCoV-19/Brazil/SP-IB_127594/2021 | EPI_ISL_4374592 | 2021-08-17 | AFIP | Instituto Butantan | Dimas Tadeu Covas, Antonio Jorge Martins, Claudia Renata dos Santos Barros, David Schlesinger, Debora Botequio Moretti, Elaine Cristina Marqueze, Elaine Vieira Santos, Evandra Strazza Rodrigues, Heidge Fukumasu, Jayme Augusto de Souza-Neto, Jos√© Salvator |
| hCoV-19/Brazil/SP-IB_127607/2021 | EPI_ISL_4374653 | 2021-08-16 | BIOFAST | Instituto Butantan | Dimas Tadeu Covas, Antonio Jorge Martins, Claudia Renata dos Santos Barros, David Schlesinger, Debora Botequio Moretti, Elaine Cristina Marqueze, Elaine Vieira Santos, Evandra Strazza Rodrigues, Heidge Fukumasu, Jayme Augusto de Souza-Neto, Jos√© Salvator |
| hCoV-19/India/DL-ILBS-64830/2020 | EPI_ISL_3276964 | 2020-11-02 | Clinical Virology Laboratory, Institute of Liver and Biliary Sciences | ILBS | Ekta Gupta, Varun Suroliya, Diptanu Paul, Amit Pandey, Reshu Agarwal, Chhagan Bihari Sharma, Shiv Kumar Sarin |
| hCoV-19/India/DL-ILBS-67122/2020 | EPI_ISL_3276988 | 2020-11-06 | Clinical Virology Laboratory, Institute of Liver and Biliary Sciences | ILBS | Ekta Gupta, Varun Suroliya, Diptanu Paul, Amit Pandey, Reshu Agarwal, Chhagan Bihari Sharma, Shiv Kumar Sarin |
| hCoV-19/India/DL-ILBS-70109/2020 | EPI_ISL_3276998 | 2020-11-14 | Clinical Virology Laboratory, Institute of Liver and Biliary Sciences | ILBS | Ekta Gupta, Varun Suroliya, Diptanu Paul, Amit Pandey, Reshu Agarwal, Chhagan Bihari Sharma, Shiv Kumar Sarin |
| hCoV-19/India/DL-ILBS-70935/2020 | EPI_ISL_3277001 | 2020-11-13 | Clinical Virology Laboratory, Institute of Liver and Biliary Sciences | ILBS | Ekta Gupta, Varun Suroliya, Diptanu Paul, Amit Pandey, Reshu Agarwal, Chhagan Bihari Sharma, Shiv Kumar Sarin |

| | | | | | |
|---|---|---|---|---|---|
| hCoV-19/FrenchGuiana/IPG2021833/2021 | EPI_ISL_3277503 | 2021-07-30 | Laboratoire Carage | Institut Pasteur de la Guyane | Anne Lavergne, Dominique Rousset, Antoine Enfissi, Arielle Salmier |
| hCoV-19/Israel/CVL-14635/2021 | EPI_ISL_3278492 | 2021-07-01 | Israel Central Virology laboratory | Israel National Consortium for SARS-CoV-2 sequencing | Neta Zuckerman, Efrat Dahan Bucris, Michal Mandelboim, Dana Bar-Ilan, Miranda Geva, Omer Asraf, Oran Erster, Efrat Glick-Saar, Omri Nayshool, Gideon Rechavi, Danit Sofer, Ella Mendelson, Orna Mor |
| hCoV-19/Israel/CVL-14649/2021 | EPI_ISL_3278504 | 2021-07-01 | Israel Central Virology laboratory | Israel National Consortium for SARS-CoV-2 sequencing | Neta Zuckerman, Efrat Dahan Bucris, Michal Mandelboim, Dana Bar-Ilan, Miranda Geva, Omer Asraf, Oran Erster, Efrat Glick-Saar, Omri Nayshool, Gideon Rechavi, Danit Sofer, Ella Mendelson, Orna Mor |
| hCoV-19/Israel/CVL-15078/2021 | EPI_ISL_3278822 | 2021-07-03 | Israel Central Virology laboratory | Israel National Consortium for SARS-CoV-2 sequencing | Neta Zuckerman, Efrat Dahan Bucris, Michal Mandelboim, Dana Bar-Ilan, Miranda Geva, Omer Asraf, Oran Erster, Efrat Glick-Saar, Omri Nayshool, Gideon Rechavi, Danit Sofer, Ella Mendelson, Orna Mor |
| hCoV-19/Israel/CVL-15571/2021 | EPI_ISL_3279135 | 2021-07-21 | Israel Central Virology laboratory | Israel National Consortium for SARS-CoV-2 sequencing | Neta Zuckerman, Efrat Dahan Bucris, Michal Mandelboim, Dana Bar-Ilan, Miranda Geva, Omer Asraf, Oran Erster, Efrat Glick-Saar, Omri Nayshool, Gideon Rechavi, Danit Sofer, Ella Mendelson, Orna Mor |
| hCoV-19/Israel/CVL-15656/2021 | EPI_ISL_3279181 | 2021-07-23 | Israel Central Virology laboratory | Israel National Consortium for SARS-CoV-2 sequencing | Neta Zuckerman, Efrat Dahan Bucris, Michal Mandelboim, Dana Bar-Ilan, Miranda Geva, Omer Asraf, Oran Erster, Efrat Glick-Saar, Omri Nayshool, Gideon Rechavi, Danit Sofer, Ella Mendelson, Orna Mor |
| hCoV-19/Israel/CVL-15671/2021 | EPI_ISL_3279190 | 2021-07-20 | Israel Central Virology laboratory | Israel National Consortium for SARS-CoV-2 sequencing | Neta Zuckerman, Efrat Dahan Bucris, Michal Mandelboim, Dana Bar-Ilan, Miranda Geva, Omer Asraf, Oran Erster, Efrat Glick-Saar, Omri Nayshool, Gideon Rechavi, Danit Sofer, Ella Mendelson, Orna Mor |
| hCoV-19/Australia/NSW-RPAH-0879/2021 | EPI_ISL_3280812 | 2021-07-26 | New South Wales Health Pathology Royal Prince Alfred Hospital | Microbiology RPAH | Foster, C.; Au, J.; Ruiz Silva, M.; Deveson, I.; Bull, R.; Van Hal, S.; Rawlinson, W. |
| hCoV-19/Australia/NSW-RPAH-0882/2021 | EPI_ISL_3280814 | 2021-08-03 | New South Wales Health Pathology Royal Prince Alfred Hospital | Microbiology RPAH | Foster, C.; Au, J.; Ruiz Silva, M.; Deveson, I.; Bull, R.; Van Hal, S.; Rawlinson, W. |
| hCoV-19/USA/CO-CDC-LC0044739/2021 | EPI_ISL_4379166 | 2021-04-13 | Laboratory Corporation of America | Centers for Disease Control and Prevention Division of Viral Diseases, Pathogen Discovery | Dakota Howard,Dhwani Batra,Peter Cook,Jason Caravas,Benjamin Rambo-Martin,Scott Sammons,Yvette Unoarumhi,Matthew Schmerer,Kristine Lacek,Tymeckia Kendall,Victoria Caban Figueroa,Shatavia Morrison,Christopher Gulvick,Minoo Agarwal,Eyad Almasri,Debbie Boles |
| hCoV-19/England/NORW-13F8560/2021 | EPI_ISL_3289487 | 2021-02-27 | Quadram Institute Bioscience | COVID-19 Genomics UK (COG-UK) Consortium | Dave J. Baker, Gemma L. Kay, Alp Aydin, Thanh Le-Viet, Steven Rudder, Ana P. Tedim, Anastasia Kolyva, Maria Diaz, Leonardo de Oliveira Martins, Nabil-Fareed Alikhan, Lizzie Meadows, Rachael Stanley, Rose K Davidson, Ngozi Elumogo, Muhammed Yasir, Sophie J |

| | | | | | |
|---|---|---|---|---|---|
| hCoV-19/USA/FL-CDC-LC0082034/2021 | EPI_ISL_4381376 | 2021-06-25 | Laboratory Corporation of America | Centers for Disease Control and Prevention Division of Viral Diseases, Pathogen Discovery | Dakota Howard,Dhwani Batra,Peter Cook,Jason Caravas,Benjamin Rambo-Martin,Scott Sammons,Yvette Unoarumhi,Matthew Schmerer,Kristine Lacek,Tymeckia Kendall,Victoria Caban Figueroa,Shatavia Morrison,Christopher Gulvick,Minoo Agarwal,Eyad Almasri,Debbie Boles |
| hCoV-19/USA/FL-CDC-ASC210126185/2021 | EPI_ISL_4381853 | 2021-09-01 | Aegis Sciences Corporation | Centers for Disease Control and Prevention Division of Viral Diseases, Pathogen Discovery | Dakota Howard,Dhwani Batra,Peter Cook,Jason Caravas,Benjamin Rambo-Martin,Scott Sammons,Yvette Unoarumhi,Matthew Schmerer,Kristine Lacek,Tymeckia Kendall,Victoria Caban Figueroa,Shatavia Morrison,Christopher Gulvick,Erisa Sula,Cyndi Clark,Patrick Campbell |
| hCoV-19/Switzerland/VD-ETHZ-33563505/2021 | EPI_ISL_3297100 | 2021-07-22 | Viollier AG | Department of Biosystems Science and Engineering, ETH Z√ºrich | Christian Beisel, Sarah Nadeau, Chaoran Chen, Ivan Topolsky, Kim Philipp Jablonski, Lara Fuhrmann, Rebecca Denes, Mirjam Feldkamp, Ina Nissen, Natascha Santacroce, Elodie Burcklen, Christiane Beckmann, Maurice Redondo, Olivier Kobel, Christoph Noppen, Nik |
| hCoV-19/Italy/VEN-IZSVe-21RS8032-12_VR/2021 | EPI_ISL_3308800 | 2021-07-21 | AULSS 9 Scaligera - San Bonifacio | Istituto Zooprofilattico Sperimentale delle Venezie | Adelaide Milani, Alessia Schivo, Annalisa Salviato, Elisa Palumbo, Erika Giorgia Quaranta, Luca Tassoni, Ambra Pastori, Edoardo Giussani, Alice Fusaro, Isabella Monne, Calogero Terregino, Antonia Ricci |
| hCoV-19/Turkey/HSGM-GF13466/2021 | EPI_ISL_3315670 | 2021-07-31 | Ministry of Health Turkey | Ministry of Health Turkey | Fatma Bayrakdar, Yasemin Co≈ügun, S√ºleyman Yalcin, G√ºlay Korukluoƒülu |
| hCoV-19/Slovenia/08-146777-MB/2021 | EPI_ISL_3316596 | 2021-07-22 | National Laboratory for Health, Environment and Food, OMM, Maribor | [illegible] Health, Environment and Food) / CISLD (Clinical Institute of Special Laboratory Diagnostics), University Children's Hospital, University Medical Center Ljubljana | Sandra Janezic, Aleksander Mahnic, Maja Rupnik, Tja≈°a ≈Ωohar ƒåretnik, Alenka ≈†torman, Nika Gobec, Aleksander Kocuvan, Kaja Tominc, Ma≈°a Jarƒçiƒç, Mojca Cimerman, Nika Volmajer, Andrej Golle / Jernej Kovaƒç, Barbara Jenko Bizjan, Tine Tesovnik, Robert |
| hCoV-19/Belgium/UGent-8088/2021 | EPI_ISL_3867774 | 2021-07-12 | Lab voor klinische biologie | Lab voor klinische biologie | Marija Janevska, Hannelore Hamerlinck, Bruno Verhasselt |
| hCoV-19/Kenya/SS2154_27_0/2021 | EPI_ISL_4196977 | 2021-08-27 | KEMRI-Wellcome Trust Research Programme,Kilifi | KEMRI-Wellcome Trust Research Programme,Kilifi | Githinji G.,Ochola I, Agoti C.,Mohamed K.S.,de Laurent Z.,Mburu M.W.,Ndwiga L., Ong'era M.Edidah,Morobe J.,Lambisia A. |
| hCoV-19/Australia/WA811/2021 | EPI_ISL_4392616 | 2021-09-16 | PathWest Laboratory Medicine WA Microbial Surveillance Unit | PathWest Laboratory Medicine WA Microbial Surveillance Unit | PathWest Laboratory Medicine WA Microbial Surveillance Unit |
| hCoV-19/Japan/PG-73920/2021 | EPI_ISL_3880686 | 2021-06-28 | Pathogen Genomics Center, National Institute of Infectious Diseases | Pathogen Genomics Center, National Institute of Infectious Diseases | Tsuyoshi Sekizuka, Kentaro Itokawa, Rina Tanaka, Masanori Hashino, Makoto Kuroda |
| hCoV-19/Japan/PG-73938/2021 | EPI_ISL_3880724 | 2021-06-27 | Pathogen Genomics Center, National Institute of Infectious Diseases | Pathogen Genomics Center, National Institute of Infectious Diseases | Tsuyoshi Sekizuka, Kentaro Itokawa, Rina Tanaka, Masanori Hashino, Makoto Kuroda |
| hCoV-19/Sweden/10097141/2020 | EPI_ISL_3791379 | 2020-11-19 | Swedish national genomic surveillance program of SARS-CoV-2 | The Public Health Agency of Sweden | Maximilian Riess, Maria Lind Karlberg, Alma Brolund, Swedish national genomic surveillance program of SARS-CoV-2 |
| hCoV-19/Japan/PG-79233/2021 | EPI_ISL_3891076 | 2021-07-24 | Pathogen Genomics Center, National Institute of Infectious Diseases | Pathogen Genomics Center, National Institute of Infectious Diseases | Tsuyoshi Sekizuka, Kentaro Itokawa, Rina Tanaka, Masanori Hashino, Makoto Kuroda |
| hCoV-19/USA/HI-H214088/2021 | EPI_ISL_4728342 | 2021-09-01 | State Laboratories Division, Hawaii State Department of Health | State Laboratories Division, Hawaii State Department of Health | Pamela O'Brien, Drew Kuwazaki, Ayana Garnet, Daniel Strange, Razvan Sultana, Edward Desmond |

| | | | | | |
|---|---|---|---|---|---|
| hCoV-19/Japan/PG-81690/2021 | EPI_ISL_3894054 | 2021-07-29 | Pathogen Genomics Center, National Institute of Infectious Diseases | Pathogen Genomics Center, National Institute of Infectious Diseases | Tsuyoshi Sekizuka, Kentaro Itokawa, Rina Tanaka, Masanori Hashino, Makoto Kuroda |
| hCoV-19/Japan/PG-82283/2021 | EPI_ISL_3894757 | 2021-06-29 | Pathogen Genomics Center, National Institute of Infectious Diseases | Pathogen Genomics Center, National Institute of Infectious Diseases | Tsuyoshi Sekizuka, Kentaro Itokawa, Rina Tanaka, Masanori Hashino, Makoto Kuroda |
| hCoV-19/Japan/PG-83761/2021 | EPI_ISL_3896414 | 2021-08-02 | SARS-CoV-2 testing team, National Institute of Infectious Diseases | Pathogen Genomics Center, National Institute of Infectious Diseases | Tsuyoshi Sekizuka, Kentaro Itokawa, Rina Tanaka, Masanori Hashino, Nozomu Hanaoka, Masumichi Saito, Naomi Nojiri, Hazuka Y Furihata, Tsuguto Fujimoto, Makoto Kuroda |
| hCoV-19/USA/KY-KSPHL-100850/2021 | EPI_ISL_4398735 | 2021-05-27 | Kentucky State Public Health Lab | Kentucky State Public Health Lab | Anjayya Tummala, Karim George, Joshua Tobias, William Grooms, Vaneet Arora, Matthew Johnson, Rachel Zinner, Rhonda Lucas, Chitralipi Kattunga |
| hCoV-19/Japan/PG-88537/2021 | EPI_ISL_3900259 | 2021-07-06 | Pathogen Genomics Center, National Institute of Infectious Diseases | Pathogen Genomics Center, National Institute of Infectious Diseases | Tsuyoshi Sekizuka, Kentaro Itokawa, Rina Tanaka, Masanori Hashino, Makoto Kuroda |
| hCoV-19/USA/FL-BPHL-5651/2021 | EPI_ISL_3903229 | 2021-07-13 | Florida Bureau of Public Health Laboratories | Florida Bureau of Public Health Laboratories | Sarah Schmedes, Jiaqi Li, Namratha Tarigopula, Jason Blanton |
| hCoV-19/Kenya/SS1201/2021 | EPI_ISL_3909850 | 2021-07-26 | KEMRI-Wellcome Trust Research Programme,Kilifi | KEMRI-Wellcome Trust Research Programme,Kilifi | Githinji G.,Ochola I, Agoti C.,Mohamed K.S.,de Laurent Z.,Mburu M.W.,Ndwiga L., Ong'era M.Edidah,Morobe J.,Lambisia A. |
| hCoV-19/Canada/AB-ABPHL-24218/2021 | EPI_ISL_3909873 | 2021-06-28 | Edmonton Provincial Lab | Public Health Agency of Canada (PHAC) National Microbiology Laboratory | Buss, E, Croxen M, Deo A, Dieu P, Gill K, Ferrato C, Khan F, Koleva P, Li V, Lloyd C, Lynch T, Ma R, Murphy S, Pabbaraju K, Shokoples S, Tipples G, Thayer J, Whitehouse M, Wong A, Yu C, Zelyas N |
| hCoV-19/Canada/AB-ABPHL-24245/2021 | EPI_ISL_3909910 | 2021-07-01 | Edmonton Provincial Lab | Public Health Agency of Canada (PHAC) National Microbiology Laboratory | Buss, E, Croxen M, Deo A, Dieu P, Gill K, Ferrato C, Khan F, Koleva P, Li V, Lloyd C, Lynch T, Ma R, Murphy S, Pabbaraju K, Shokoples S, Tipples G, Thayer J, Whitehouse M, Wong A, Yu C, Zelyas N |
| hCoV-19/Netherlands/OV-RIVM-57596/2021 | EPI_ISL_4400897 | 2021-08-30 | Dutch COVID-19 response team | National Institute for Public Health and the Environment (RIVM) | Adam Meijer, Harry Vennema, Dirk Eggink, Jeroen Cremer, Sharon van den Brink, Bas van der Veer, AnneMarie van den Brandt, Lisa Wijsman, Kim Freriks, Ryanne Jaarsma, Eun√≠ce Then, Lynn Aarts, Sanne Bos, Stijn van Rossum, Florian Zwagemaker, Dennis Schmitz, |
| hCoV-19/USA/TX-CDC-ASC210120121/2021 | EPI_ISL_4401197 | 2021-08-20 | Aegis Sciences Corporation | Centers for Disease Control and Prevention Division of Viral Diseases, Pathogen Discovery | Dakota Howard,Dhwani Batra,Peter Cook,Jason Caravas,Benjamin Rambo-Martin,Scott Sammons,Yvette Unoarumhi,Matthew Schmerer,Kristine Lacek,Tymeckia Kendall,Victoria Caban Figueroa,Shatavia Morrison,Christopher Gulvick,Erisa Sula,Cyndi Clark,Patrick Campbell |
| hCoV-19/Togo/NMIMR-21-TGS-433/2021 | EPI_ISL_3915116 | 2021-07-01 | Institut National D,Äôhygiene, Lome, Togo | Noguchi Memorial Institute for Medical Research, University of Ghana, Legon, Ghana | Afiwa W. Halatoko, Issaka Maman, Yawo A. Sadji, Komlan Kossi, Quaneeta Mohktar, Hilda Opoku Frempong, Peace O. Uche, Keren O. Attiku, Djimabi Salah, Yao Layibo, Lallepak Lamboni, Seyram B. Agbenyo, Hamadi Assane, Joyce Appiah-Kubi, Zoukaneirii Issa, Sena |

| | | | | | |
|---|---|---|---|---|---|
| hCoV-19/Aruba/AW-RIVM-53921/2021 | EPI_ISL_3918647 | 2021-08-15 | Dutch COVID-19 response team | National Institute for Public Health and the Environment (RIVM) | Adam Meijer, Harry Vennema, Dirk Eggink, Jeroen Cremer, Sharon van den Brink, Bas van der Veer, AnneMarie van den Brandt, Lisa Wijsman, Kim Freriks, Ryanne Jaarsma, Eun√≠ce Then, Lynn Aarts, Sanne Bos, Stijn van Rossum, Florian Zwagemaker, Dennis Schmitz, |
| hCoV-19/Peru/PIU-INS-1630/2021 | EPI_ISL_3023399 | 2021-02-17 | Laboratorio de Referencial Nacional de Virus Respiratorios | Laboratorio de Referencial Nacional de Virus Respiratorios | Carlos Padilla Rojas, Veronica Hurtado Vela, Orson Mestanza Millones, Iris Silva Molina, Luis Barcena Flores, Sandra Morales Ruiz, Steve Acedo Lazo, Omar Caceres Rey, Marco Galarza Perez, Henri Bailon Calderon, Nancy Rojas Serrano, Priscila Lope Pari, Lel |
| hCoV-19/Curacao/CW-RIVM-56706/2021 | EPI_ISL_4401882 | 2021-08-23 | Dutch COVID-19 response team | National Institute for Public Health and the Environment (RIVM) | Adam Meijer, Harry Vennema, Dirk Eggink, Jeroen Cremer, Sharon van den Brink, Bas van der Veer, AnneMarie van den Brandt, Lisa Wijsman, Kim Freriks, Ryanne Jaarsma, Eun√≠ce Then, Lynn Aarts, Sanne Bos, Stijn van Rossum, Florian Zwagemaker, Dennis Schmitz, |
| hCoV-19/France/ARA-HCL021142530801/2021 | EPI_ISL_3634187 | 2021-07-26 | LABORATOIRE UNILIANS DECINES | CNR Virus des Infections Respiratoires - France SUD | Antonin Bal, Gregory Destras, Gwendolyne Burfin, Hadrien Regue, Quentin Semanas, Martine Valette, Bruno Lina, Laurence Josset |
| hCoV-19/Chile/RM-TAAG-UNAB-594663/2021 | EPI_ISL_3922161 | 2021-07-29 | TAAG Genetics | Facultad de Ciencias de la Vida, UNAB | Claudio Meneses, Eduardo Castro, Macarena Bastias, Ricardo Yusta, Susana Saez, Dayan Sanhueza, Patricio Tapia, Ariel Orellana |
| hCoV-19/Iceland/9344/2021 | EPI_ISL_3928308 | 2021-08-09 | The National University Hospital of Iceland | deCODE genetics | Daniel F Gudbjartsson; Agnar Helgason; Hakon Jonsson; Olafur T Magnusson; Pall Melsted; Gudmundur L Norddahl; Jona Saemundsdottir; Asgeir Sigurdsson; Patrick Sulem; Arna B Agustsdottir; Hannes Eggertsson; Berglind Eiriksdottir; Run Fridriksdottir; Elisab |
| hCoV-19/USA/CA-CDC-FG-070459/2021 | EPI_ISL_3608965 | 2021-08-09 | Fulgent Genetics | Centers for Disease Control and Prevention Division of Viral Diseases, Pathogen Discovery | Dakota Howard,Dhwani Batra,Peter Cook,Kara Moser,Adrian Paskey,Jason Caravas,Benjamin Rambo-Martin,Shatavia Morrison,Christopher Gulvick,Scott Sammons,Yvette Unoarumhi,Darlene Wagner,Matthew Schmerer,Harry Gao,Mickey Li,John Gao,Joseph Fierro,Benafsh Sapr |
| hCoV-19/USA/NY-PRL-2021_0514_00I05/2021 | EPI_ISL_3933416 | 2021-05-11 | Pandemic Response Lab - NYC | Pandemic Response Lab, R&D | Isabel Fernandez Escapa, Henry Lee, Michael Hammerling, Melissa Hopkins, Cybill del Castillo, Shinyoung Clair Kang, William Ward, Pradeep Bugga, Sol Rey, Dylan Law, Katharine Nelson, Haiping Hao, Jon Laurent |
| hCoV-19/India/AP-CCMB-CIA7610/2021 | EPI_ISL_3935056 | 2021-07-13 | AP SSO | CSIR-Centre for Cellular and Molecular Biology - INSACOG | Sumedha Avadhanula, Lamuk Zaveri, Tulasi Nagabandi, Ara Sreenivas,Shreekant Verma, Amareshwar Vodapalli ,B Himasri,Valli Nagalakshmi Undamatla, Vidhyadhari Methuku, Priya Nurkurthy, Payel Mukherjee,Jandhyala Sai Krishna,Onkar Kulkarni, Sofia Banu, Archana |

| | | | | | |
|---|---|---|---|---|---|
| hCoV-19/India/TG-CCMB-CIA7650/2021 | EPI_ISL_3935061 | 2021-08-03 | CSIR-Centre for Cellular and Molecular Biology - INSACOG | CSIR-Centre for Cellular and Molecular Biology - INSACOG | Shreekant Verma, Amareshwar Vodapalli ,Ara Sreenivas,B Himasri,Valli Nagalakshmi Undamatla,Payel Mukherjee, Lamuk Zaveri, Onkar Kulkarni,Tulasi Nagabandi, Sofia Banu, Jandhyala Sai Krishna,Sumedha Avadhanula, Priya Nurkurthy, Vidhyadhari Methuku, Archana |
| hCoV-19/India/AP-CCMB-CIA7674/2021 | EPI_ISL_3935064 | 2021-07-18 | AP SSO | CSIR-Centre for Cellular and Molecular Biology - INSACOG | Tulasi Nagabandi,Lamuk Zaveri,Payel Mukherjee,Onkar Kulkarni,Jandhyala Sai Krishna, Sofia Banu, Ara Sreenivas,Shreekant Verma, Amareshwar Vodapalli ,B Himasri,Valli Nagalakshmi Undamatla, Priya Nurkurthy,Sumedha Avadhanula, Vidhyadhari Methuku, Archana Bh |
| hCoV-19/India/TG-CCMB-CIA7736/2021 | EPI_ISL_3935220 | 2021-08-05 | Mapmygenome | CSIR-Centre for Cellular and Molecular Biology - INSACOG | Lamuk Zaveri,Tulasi Nagabandi,Payel Mukherjee,Onkar Kulkarni, Sofia Banu, Jandhyala Sai Krishna,Ara Sreenivas,Shreekant Verma, Amareshwar Vodapalli ,B Himasri,Valli Nagalakshmi Undamatla, Sumedha Avadhanula, Priya Nurkurthy, Vidhyadhari Methuku, Archana B |
| hCoV-19/Argentina/PAIS-A0999/2021 | EPI_ISL_4405619 | 2021-08-02 | Laboratorio de Virolog√≠a del Hospital de Ni√±os Dr. Ricardo Gutierrez | Laboratorio de Virolog√≠a del Hospital de Ni√±os Dr. Ricardo Gutierrez on behalf of 'Proyecto Argentino Interinstitucional de genomica de SARS-CoV-2' (PAIS | Nabaes Jodar, MS; Goya, S; Acu√±a, D; Natale, MI; Lusso, S; Alexay, S; Gravis, E; Acevedo, ME; Alvarez Lopez, C; Jacques, O; Mistchenko, AS; Valinotto, LE; Viegas, M. |
| hCoV-19/Nigeria/NCDC-NR256/2021 | EPI_ISL_3939012 | 2021-07-28 | IBOM MOLECULAR LABORATORY | National Reference Laboratory, Nigeria Centre for Disease Control | Dr Ndodo Nnaemeka, Olusola Anuoluwapo Akanbi, Chimaobi Chukwu, Dr Omoare Adesuyi, Celestina Obiekea, Anthony Ahumibe, Catherine Okoi, Nwando Mba, Dr Chikwe Ihekweazu |
| hCoV-19/SouthAfrica/KRISP-K022367/2021 | EPI_ISL_3939051 | 2021-08-12 | National Health Laboratory Service, South Africa | KRISP, KZn Research Innovation and Sequencing Platform | Giandhari J, Pillay S, Yajna Ramphal, Naidoo Yeshnee, Tshabuila Derek, Emmanuel S, Tegally H, Wilkinson E, de Oliveira T |
| hCoV-19/Brazil/BA-LACEN-BA527/2021 | EPI_ISL_3944571 | 2021-08-12 | Central Public Health Laboratory - LACEN - Bahia, Salvador, Brazil | Central Public Health Laboratory - LACEN -Bahia, Salvador, Brazil | Stephane Tosta, Luciana Oliveira, Vanessa Nardy,Marcela G√≥mez, Jaqueline Gomes,Gabriela Menezes, Lenisa Dandara,Vagner Fonseca,Marta Giovanetti,Luiz Alcantara, Felicidade Pereira, Arabela Leal |
| hCoV-19/Bangladesh/CHRF-0554_Pr/2021 | EPI_ISL_3946173 | 2021-06-18 | Praava Health | Child Health Research Foundation | CHRF Bangladesh Genomics Team, Dr. Zaheed Husain, Shafiul Azam |
| hCoV-19/Nigeria/CV1984/2021 | EPI_ISL_3946301 | 2021-07-02 | Nigerian Centre for Disease Control (NCDC) | Africa Centre for Excellence for Genomics of Infectious Diseases (ACEGID), Redeemer,Äôs University | Olawoye, I.B., Oluniyi, P.E., Eromon, P.E., Oguzie, J.U., Kayode, A.T., Uwanibe, J.N., Ugwu, C.A., Akano, K.O., Ajogbasile, F.V., Abechi, P.S., Olumade, T.J., Nosamiefan, I., Folarin, O., Happi, C.T. |
| hCoV-19/NorthernMarianaIslands/MP-CDC-2-4813561/2021 | EPI_ISL_3947115 | 2021-07-17 | Commonwealth Healthcare Center | Centers for Disease Control and Prevention Division of Viral Diseases, Pathogen Discovery | Mili Sheth,Sarah Nobles,Jasmine Padilla,Alex Burgin,Meghan Bentz,Mark Burroughs,Peter Cook,Clinton Paden,Dhwani Batra,Krista Queen,Kristen Knipe,Dakota Howard,Yvette Unoarumhi,Matthew Schmerer,Ben Rambo-Martin,Kristine Lacek,Sam Shepard,Dave Wentworth,Viv |

| | | | | | |
|---|---|---|---|---|---|
| hCoV-19/Scotland/QEUH-1E73C03/2021 | EPI_ISL_4408075 | 2021-09-14 | Lighthouse Lab in Glasgow | Wellcome Sanger Institute for the COVID-19 Genomics UK (COG-UK) Consortium | Harper VanSteenhouse, Yumi Kasai, David Gray, Carol Clugston, Anna Dominiczak and Alex Alderton, Roberto Amato, Jeffrey Barrett, Sonia Goncalves, Ewan Harrison, David K. Jackson, Ian Johnston, Dominic Kwiatkowski, Cordelia Langford, John Sillitoe on behal |
| hCoV-19/Botswana/R29B9_BHP_000726046/2021 | EPI_ISL_3948382 | 2021-08-24 | National Health Laboratory | Botswana Harvard HIV Reference Laboratory | Sikhulile Moyo, Wonderful T. Choga, Dorcas Maruapula, Keoratile Ntshambiwa, Sefetogi Ramaologa, Thongbotho Mphoyakgosi, Boitumelo Zuze, Botshelo Radibe, Legodile Kooepile, Ontlametse T. Bareng, Patrick T. Mokgethi, Pamela Smith-Lawrence, Kgomotso Moruisi, |
| hCoV-19/England/MILK-1C6151E/2021 | EPI_ISL_3957088 | 2021-08-22 | Lighthouse Lab in Milton Keynes | Wellcome Sanger Institute for the COVID-19 Genomics UK (COG-UK) Consortium | The Lighthouse Lab in Milton Keynes and Alex Alderton, Roberto Amato, Jeffrey Barrett, Sonia Goncalves, Ewan Harrison, David K. Jackson, Ian Johnston, Dominic Kwiatkowski, Cordelia Langford, John Sillitoe on behalf of the Wellcome Sanger Institute COVID-1 |
| hCoV-19/SouthAfrica/NHLS-UCT-LA-Z621/2021 | EPI_ISL_3957787 | 2021-08-10 | Lancet | NHLS/UCT | Arash Iranzadeh, Deelan Doolabh, Lynn Tyers, Bruna Galvao, Innocent Mudau, Marvin Hsiao, Gert Marais, Diana Hardie, Stephen Korsman, Rageema Joseph, Carolyn Williamson |
| hCoV-19/England/PHEC-V307V7C5/2021 | EPI_ISL_3960221 | 2021-08-10 | Respiratory Virus Unit, Microbiology Services Colindale, Public Health England | COVID-19 Genomics UK (COG-UK) Consortium | PHE Covid Sequencing Team |
| hCoV-19/Australia/WA800/2021 | EPI_ISL_4414961 | 2021-08-26 | PathWest Laboratory Medicine WA | PathWest Laboratory Medicine WA Microbial Surveillance Unit | PathWest Laboratory Medicine WA Microbial Surveillance Unit |
| hCoV-19/India/MH-IISER-Pune-IP-00994/2021 | EPI_ISL_4415417 | 2021-02-09 | Department of Microbiology, Government Medical College, Baramati | IISER Pune-INSACOG | Krishanpal Karmodiya, Aurnab Ghose, Joy Merwin Monteiro |
| hCoV-19/India/MH-IISER-Pune-IP-01036/2021 | EPI_ISL_4415434 | 2021-02-09 | Department of Microbiology, Government Medical College, Baramati | IISER Pune-INSACOG | Krishanpal Karmodiya, Aurnab Ghose, Joy Merwin Monteiro |
| hCoV-19/India/MH-IISER-Pune-IP-01041/2021 | EPI_ISL_4415439 | 2021-02-09 | Department of Microbiology, Government Medical College, Baramati | IISER Pune-INSACOG | Krishanpal Karmodiya, Aurnab Ghose, Joy Merwin Monteiro |
| hCoV-19/India/MH-IISER-Pune-IP01482/2021 | EPI_ISL_4415507 | 2021-01-09 | BJMC, Pune | IISER Pune-INSACOG | Krishanpal Karmodiya, Aurnab Ghose, Joy Merwin Monteiro, Suvarna Joshi, Rajesh Karyakarte |
| hCoV-19/India/MH-IISER-Pune-IP01455/2021 | EPI_ISL_4415531 | 2021-01-09 | BJMC, Pune | IISER Pune-INSACOG | Krishanpal Karmodiya, Aurnab Ghose, Joy Merwin Monteiro, Suvarna Joshi, Rajesh Karyakarte |
| hCoV-19/India/MH-IISER-Pune-IP01471/2021 | EPI_ISL_4415599 | 2021-01-09 | BJMC, Pune | IISER Pune-INSACOG | Krishanpal Karmodiya, Aurnab Ghose, Joy Merwin Monteiro, Suvarna Joshi, Rajesh Karyakarte |
| hCoV-19/India/MH-IISER-Pune-IP-00999/2021 | EPI_ISL_4415634 | 2021-02-09 | Department of Microbiology, Government Medical College, Baramati | IISER Pune-INSACOG | Krishanpal Karmodiya, Aurnab Ghose, Joy Merwin Monteiro |
| hCoV-19/India/MH-IISER-Pune-IP01439/2021 | EPI_ISL_4415686 | 2021-01-09 | BJMC, Pune | IISER Pune-INSACOG | Krishanpal Karmodiya, Aurnab Ghose, Joy Merwin Monteiro, Suvarna Joshi, Rajesh Karyakarte |
| hCoV-19/India/MH-IISER-Pune-IP-01017/2021 | EPI_ISL_4415721 | 2021-02-09 | Department of Microbiology, Government Medical College, Baramati | IISER Pune-INSACOG | Krishanpal Karmodiya, Aurnab Ghose, Joy Merwin Monteiro |
| hCoV-19/India/MH-IISER-Pune-IP01450/2021 | EPI_ISL_4415751 | 2021-01-09 | BJMC, Pune | IISER Pune-INSACOG | Krishanpal Karmodiya, Aurnab Ghose, Joy Merwin Monteiro, Suvarna Joshi, Rajesh Karyakarte |
| hCoV-19/Portugal/PT15248/2021 | EPI_ISL_3981014 | 2021-08-24 | H Guimaraes | Instituto Nacional de Saude (INSA) | Borges et al |

| | | | | | |
|---|---|---|---|---|---|
| hCoV-19/Slovakia/16304/2021 | EPI_ISL_3982352 | 2021-08-04 | Public Health Authority of the Slovak Republic | Public Health Authority of the Slovak Republic | Barbora Kotvasovv°, Lucia ≈†evƒçv≠kovv°, Terv©zia Vrabƒæovv°, Anna Giƒçovv°, Elena Tichv°, Miroslav Bv∂hmer, Tomv°≈° Szemes, Pavol Mi≈°enko |
| hCoV-19/India/MH-ICMR-NIV-INSACOG-GSEQ-5245/2021 | EPI_ISL_3982475 | 2021-08-01 | ICMR-National Institute of Virology - INSACOG | NIV Influenza | Potdar, Varsha on behalf of National Influenza Centre, NIV, Pune |
| hCoV-19/USA/CA-SC-258416/2021 | EPI_ISL_4418137 | 2021-08-31 | Santa Clara Valley Medical Center | Santa Clara County Public Health Laboratory | Santa Clara County PHL |
| hCoV-19/India/AS-1861802605951/2021 | EPI_ISL_3982965 | 2021-07-26 | INSACOG-Assam | National Institute of Biomedical Genomics ‚Äì INSACOG | Arindam Maitra, Ajanta Sharma, Nidhan Kumar Biswas, Lahari Saikia, Sreedhar Chinnaswamy,¬†Kailash Chamuah, Saumitra Das |
| hCoV-19/India/AS-1828600416658/2021 | EPI_ISL_3983071 | 2021-07-13 | INSACOG-Assam | National Institute of Biomedical Genomics ‚Äì INSACOG | Arindam Maitra, Ajanta Sharma, Nidhan Kumar Biswas, Lahari Saikia, Sreedhar Chinnaswamy,¬†Kailash Chamuah, Saumitra Das |
| hCoV-19/India/AS-1828600407521/2021 | EPI_ISL_3983076 | 2021-07-11 | INSACOG-Assam | National Institute of Biomedical Genomics ‚Äì INSACOG | Arindam Maitra, Ajanta Sharma, Nidhan Kumar Biswas, Lahari Saikia, Sreedhar Chinnaswamy,¬†Kailash Chamuah, Saumitra Das |
| hCoV-19/Sweden/20600642P/2021 | EPI_ISL_3986667 | 2021-08-16 | Swedish national genomic surveillance program of SARS-CoV-2 | The Public Health Agency of Sweden | Maximilian Riess, Maria Lind Karlberg, Alma Brolund, Swedish national genomic surveillance program of SARS-CoV-2 |
| hCoV-19/Italy/VEN-IZSVe-21RS8091-2_RO/2021 | EPI_ISL_3987942 | 2021-08-12 | AULSS 5 Polesana | Istituto Zooprofilattico Sperimentale delle Venezie | Adelaide Milani, Alessia Schivo, Annalisa Salviato, Elisa Palumbo, Erika Giorgia Quaranta, Luca Tassoni, Ambra Pastori, Edoardo Giussani, Alice Fusaro, Isabella Monne, Calogero Terregino, Antonia Ricci |
| hCoV-19/Guinea/Conakry_20912/2021 | EPI_ISL_3988370 | 2021-04-27 | Institut Pasteur de Guinv©e | Institut Pateur de Dakar | Grayo Solene, Diagne Moussa Mov Øse, Diallo Amadou, Dia Ndongo, Mbengue Safietou Sankhe, Ndiaye Ndack, Diop Mamadou, Maimouna Mbanne, Mohamed Kane, Loucoubar Cheikh, Tordo Noel, Faye Ousmane, Sall Amadou Alpha |
| hCoV-19/USA/NC-WFSM-1086-41/2021 | EPI_ISL_4419731 | 2021-07-27 | Atrium Health Wake Forest Baptist Microbiology Lab | Wake Forest School of Medicine, Internal Medicine | Kimberly Reeves, Gregory Hawkins |
| hCoV-19/Canada/BC-BCCDC-168688/2021 | EPI_ISL_4422391 | 2021-02-26 | British Columbia Centre For Disease Control | BCCDC Public Health Laboratory | Prystajecky Natalie, Linda Hoang, Dan Fornika, John Tyson, Shannon Russell, Kim Macdonald, Kimia Kamelian, Ana Pacagnella, Corrinne Ng, Loretta Janz, Robert Azana, Mel Krajden |
| hCoV-19/Canada/BC-BCCDC-168976/2021 | EPI_ISL_4423106 | 2021-03-25 | British Columbia Centre For Disease Control | BCCDC Public Health Laboratory | Prystajecky Natalie, Linda Hoang, Dan Fornika, John Tyson, Shannon Russell, Kim Macdonald, Kimia Kamelian, Ana Pacagnella, Corrinne Ng, Loretta Janz, Robert Azana, Mel Krajden |
| hCoV-19/Canada/BC-BCCDC-169109/2021 | EPI_ISL_4423254 | 2021-03-31 | British Columbia Centre For Disease Control | BCCDC Public Health Laboratory | Prystajecky Natalie, Linda Hoang, Dan Fornika, John Tyson, Shannon Russell, Kim Macdonald, Kimia Kamelian, Ana Pacagnella, Corrinne Ng, Loretta Janz, Robert Azana, Mel Krajden |
| hCoV-19/Sweden/461847581967/2021 | EPI_ISL_3999071 | 2021-06-28 | Swedish national genomic surveillance program of SARS-CoV-2 | The Public Health Agency of Sweden | Maximilian Riess, Maria Lind Karlberg, Alma Brolund, Swedish national genomic surveillance program of SARS-CoV-2 |

| | | | | | |
|---|---|---|---|---|---|
| hCoV-19/Turkey/HSGM-C19560/2021 | EPI_ISL_4001155 | 2021-08-20 | Ministry of Health Turkey | Ministry of Health Turkey | Fatma Bayrakdar, Yasemin Co≈ügun, S√ºleyman Yalcin, G√ºlay Korukluoƒülu |
| hCoV-19/Sweden/21-54652/2021 | EPI_ISL_4003259 | 2021-08-04 | Swedish national genomic surveillance program of SARS-CoV-2 | The Public Health Agency of Sweden | Maximilian Riess, Maria Lind Karlberg, Alma Brolund, Swedish national genomic surveillance program of SARS-CoV-2 |
| hCoV-19/Australia/QLD2162/2021 | EPI_ISL_3333193 | 2021-08-05 | Queensland Health Forensic and Scientific Services | Queensland Health Forensic and Scientific Services | Chenwei Wang on behalf of Q-PHIRE Genomics |
| hCoV-19/Belgium/rega-11137/2021 | EPI_ISL_4007310 | 2021-06-15 | KU Leuven, Rega Institute, Clinical and Epidemiological Virology | KU Leuven, Rega Institute, Clinical and Epidemiological Virology | Tony Wawina-Bokalanga, Bert Vanmechelen, Joan Marti-Carerras, Piet Maes |
| hCoV-19/Belgium/rega-11196/2021 | EPI_ISL_4008009 | 2021-06-17 | KU Leuven, Rega Institute, Clinical and Epidemiological Virology | KU Leuven, Rega Institute, Clinical and Epidemiological Virology | Tony Wawina-Bokalanga, Bert Vanmechelen, Joan Marti-Carerras, Piet Maes |
| hCoV-19/USA/RI-RISHL-079216/2021 | EPI_ISL_4011339 | 2021-08-02 | Rhode Island State Health Laboratory | Rhode Island State Health Laboratory | Ewa King, Richard C. Huard, Kristin Carpenter-Azevedo |
| hCoV-19/USA/RI-RISHL-E01535/2021 | EPI_ISL_4011354 | 2021-08-06 | Rhode Island State Health Laboratory | Rhode Island State Health Laboratory | Ewa King, Richard C. Huard, Kristin Carpenter-Azevedo |
| hCoV-19/Turkey/HSGM-E5876/2021 | EPI_ISL_4428985 | 2021-09-09 | Ministry of Health Turkey | Ministry of Health Turkey | Fatma Bayrakdar, Yasemin Co≈ügun, S√ºleyman Yalcin, G√ºlay Korukluoƒülu |
| hCoV-19/England/ALDP-1D3FB70/2021 | EPI_ISL_4012106 | 2021-09-01 | Lighthouse Lab in Alderley Park | Wellcome Sanger Institute for the COVID-19 Genomics UK (COG-UK) Consortium | Jacquelyn Wynn, Mairead Hyland, The Lighthouse Lab in Alderley Park and Alex Alderton, Roberto Amato, Jeffrey Barrett, Sonia Goncalves, Ewan Harrison, David K. Jackson, Ian Johnston, Dominic Kwiatkowski, Cordelia Langford, John Sillitoe on behalf of the W |
| hCoV-19/Gambia/49011/2021 | EPI_ISL_3150961 | 2021-07-09 | MRCG at LSHTM Genomics lab | MRCG at LSHTM Genomics lab | Abdul Karim sesay, Abdoulie Kanteh, Jarra Manneh, Mariama Kujabi, Bakary Sanyang,Dabiri Damilari |
| hCoV-19/India/DL-NCDC-3009275/2021 | EPI_ISL_4435806 | 2021-04-22 | National Centre for Disease Control (NCDC) Biotechnology Division, Delhi - INSACOG | NCDC Delhi, Biotechnology Division - INSACOG | Radhakrishnan V. S, Robin Marwal, Mahesh S Dhar, Kalaiarasan Ponnusamy, Meena Datta, Priyanka Singh,¬†Uma Sharma,¬†Hemlata Lall, Manoj K Singh, Hema Gogia, Preeti Madan, Sandhya Kabra, Sujeet K Singh, Partha Rakshit |
| hCoV-19/India/DL-NCDC-3009298/2021 | EPI_ISL_4435821 | 2021-04-23 | National Centre for Disease Control (NCDC) Biotechnology Division, Delhi - INSACOG | NCDC Delhi, Biotechnology Division - INSACOG | Radhakrishnan V. S, Robin Marwal, Mahesh S Dhar, Kalaiarasan Ponnusamy, Meena Datta, Priyanka Singh,¬†Uma Sharma,¬†Hemlata Lall, Manoj K Singh, Hema Gogia, Preeti Madan, Sandhya Kabra, Sujeet K Singh, Partha Rakshit |
| hCoV-19/India/PB-NCDC-310935/2021 | EPI_ISL_4435930 | 2021-04-30 | National Centre for Disease Control (NCDC) Biotechnology Division, Delhi - INSACOG | NCDC Delhi, Biotechnology Division - INSACOG | Radhakrishnan V. S, Robin Marwal, Mahesh S Dhar, Kalaiarasan Ponnusamy, Meena Datta, Priyanka Singh,¬†Uma Sharma,¬†Hemlata Lall, Manoj K Singh, Hema Gogia, Preeti Madan, Sandhya Kabra, Sujeet K Singh, Partha Rakshit |
| hCoV-19/India/PB-NCDC-310990/2021 | EPI_ISL_4435969 | 2021-04-30 | National Centre for Disease Control (NCDC) Biotechnology Division, Delhi - INSACOG | NCDC Delhi, Biotechnology Division - INSACOG | Radhakrishnan V. S, Robin Marwal, Mahesh S Dhar, Kalaiarasan Ponnusamy, Meena Datta, Priyanka Singh,¬†Uma Sharma,¬†Hemlata Lall, Manoj K Singh, Hema Gogia, Preeti Madan, Sandhya Kabra, Sujeet K Singh, Partha Rakshit |

| | | | | | |
|---|---|---|---|---|---|
| hCoV-19/USA/CA-CDC-FG-099517/2021 | EPI_ISL_4023799 | 2021-08-23 | Fulgent Genetics | Centers for Disease Control and Prevention Division of Viral Diseases, Pathogen Discovery | Dakota Howard,Dhwani Batra,Peter Cook,Kara Moser,Adrian Paskey,Jason Caravas,Benjamin Rambo-Martin,Shatavia Morrison,Christopher Gulvick,Scott Sammons,Yvette Unoarumhi,Darlene Wagner,Matthew Schmerer,Harry Gao,Mickey Li,John Gao,Joseph Fierro,Benafsh Sapr |
| hCoV-19/USA/CA-CDC-FG-099815/2021 | EPI_ISL_4023926 | 2021-08-23 | Fulgent Genetics | Centers for Disease Control and Prevention Division of Viral Diseases, Pathogen Discovery | Dakota Howard,Dhwani Batra,Peter Cook,Kara Moser,Adrian Paskey,Jason Caravas,Benjamin Rambo-Martin,Shatavia Morrison,Christopher Gulvick,Scott Sammons,Yvette Unoarumhi,Darlene Wagner,Matthew Schmerer,Harry Gao,Mickey Li,John Gao,Joseph Fierro,Benafsh Sapr |
| hCoV-19/Nigeria/BCVL-3683/2020 | EPI_ISL_4026312 | 2020-07-17 | Biorepository and Clinical Virology Laboratory | Northwestern University - Center for Pathogen Genomics and Microbial Evolution | Ramon Lorenzo-Redondo, Adeola A. Fowotade, Ewean C. Omoruyi, Johnson A. Adeniji, Lacy M. Simons, Judd F. Hultquist, Babafemi O. Taiwo, Olubusuyi M. Adewumi, Egon A. Ozer |
| hCoV-19/Nigeria/BCVL-23078/2021 | EPI_ISL_4026421 | 2021-02-07 | Biorepository and Clinical Virology Laboratory | Northwestern University - Center for Pathogen Genomics and Microbial Evolution | Ramon Lorenzo-Redondo, Adeola A. Fowotade, Ewean C. Omoruyi, Johnson A. Adeniji, Lacy M. Simons, Judd F. Hultquist, Babafemi O. Taiwo, Olubusuyi M. Adewumi, Egon A. Ozer |
| hCoV-19/USA/CA-CDC-FG-103738/2021 | EPI_ISL_4027936 | 2021-08-26 | Fulgent Genetics | Centers for Disease Control and Prevention Division of Viral Diseases, Pathogen Discovery | Dakota Howard,Dhwani Batra,Peter Cook,Kara Moser,Adrian Paskey,Jason Caravas,Benjamin Rambo-Martin,Shatavia Morrison,Christopher Gulvick,Scott Sammons,Yvette Unoarumhi,Darlene Wagner,Matthew Schmerer,Harry Gao,Mickey Li,John Gao,Joseph Fierro,Benafsh Sapr |
| hCoV-19/SouthAfrica/NICD-R10194/2021 | EPI_ISL_4030086 | 2021-07-05 | National Institute for Communicable Diseases of the National Health Laboratory Service | Diseases of the National Health Laboratory Service | Amoako DG, Everatt J, Scheepers C, Mohale T, Ntuli N, Mahlangu B, Mnguni A, Ismail A, Bhiman JN |
| hCoV-19/Lithuania/MR-LUHS-16-10/2021 | EPI_ISL_4030576 | 2021-08-26 | UAB InMedica | Lithuanian University of Health Sciences | Arnoldas Pautienius, Emilija Gecyte, Gediminas Alzbutas, Dovydas Gecys, Lukas Zemaitis, Vaiva Lesauskaite |
| hCoV-19/Australia/VIC18908/2021 | EPI_ISL_2978348 | 2021-07-09 | Victorian Infectious Diseases Reference Laboratory (VIDRL) | VIDRL and MDU-PHL | Caly L., Seemann T., Sait, M.L., Druce J., Sherry, N.L. |
| hCoV-19/Australia/VIC18918/2021 | EPI_ISL_2978350 | 2021-07-12 | Victorian Infectious Diseases Reference Laboratory (VIDRL) | VIDRL and MDU-PHL | Caly L., Seemann T., Sait, M.L., Druce J., Sherry, N.L. |
| hCoV-19/USA/TX-CDC-FG-091573/2021 | EPI_ISL_4034236 | 2021-08-19 | Fulgent Genetics | Centers for Disease Control and Prevention Division of Viral Diseases, Pathogen Discovery | Dakota Howard,Dhwani Batra,Peter Cook,Kara Moser,Adrian Paskey,Jason Caravas,Benjamin Rambo-Martin,Shatavia Morrison,Christopher Gulvick,Scott Sammons,Yvette Unoarumhi,Darlene Wagner,Matthew Schmerer,Harry Gao,Mickey Li,John Gao,Joseph Fierro,Benafsh Sapr |
| hCoV-19/Peru/LIM-INS-3902/2021 | EPI_ISL_4036790 | 2021-07-19 | Laboratorio de Referencia Nacional de Virus Respiratorios. Centro Nacional de Salud Publica. Instituto Nacional de Salud Peru. | Laboratorio de Referencia Nacional de Virus Respiratorios. Centro Nacional de Salud Publica. Instituto Nacional de Salud Peru. | Carlos Padilla Rojas, Veronica Hurtado Vela, Orson Mestanza Millones, Iris Silva Molina, Luis Barcena Flores, Sandra Morales Ruiz, Steve Acedo Lazo, Wendy Lizarraga Olivares, Victor Jimenez Vasquez, Sara Gordillo Vilchez, Nieves Sevilla Casta√±eda, Prince |

| | | | | | |
|---|---|---|---|---|---|
| hCoV-19/Kenya/SS1507_1_0/2021 | EPI_ISL_4037003 | 2021-07-01 | KEMRI-Wellcome Trust Research Programme,Kilifi | KEMRI-Wellcome Trust Research Programme,Kilifi | Githinji G.,Ochola I, Agoti C.,Mohamed K.S.,de Laurent Z.,Mburu M.W.,Ndwiga L., Ong'era M.Edidah,Morobe J.,Lambisia A. |
| hCoV-19/USA/WY-WYPHL-21063402/2021 | EPI_ISL_4037053 | 2021-08-20 | Wyoming Public Health Laboratory | Wyoming Public Health Laboratory | Jim Mildenberger, Taylor Fearing, Channing Weber, Ashley Norberg, Chayse Rowley, Marley Goetz, Brian Dominguez, Elliot Thomasson, Sam Britz, Cari Sloma, Robert Petit, and Rob Christensen |
| hCoV-19/India/DL-NCDC-3409150/2021 | EPI_ISL_4444755 | 2021-04-18 | National Centre for Disease Control (NCDC) Biotechnology Division, Delhi - INSACOG | NCDC Delhi, Biotechnology Division - INSACOG | Robin Marwal, Mahesh S Dhar, Kalaiarasan Ponnusamy, Meena Datta, Priyanka Singh,¬†Uma Sharma,¬†Hemlata Lall, Manoj K Singh, Radhakrishnan V. S, Hema Gogia, Preeti Madan, Sandhya Kabra, Sujeet K Singh, Partha Rakshit |
| hCoV-19/India/UT-NCDC-3409244/2021 | EPI_ISL_4444858 | 2021-04-27 | National Centre for Disease Control (NCDC) Biotechnology Division, Delhi - INSACOG | NCDC Delhi, Biotechnology Division - INSACOG | Robin Marwal, Mahesh S Dhar, Kalaiarasan Ponnusamy, Meena Datta, Priyanka Singh,¬†Uma Sharma,¬†Hemlata Lall, Manoj K Singh, Radhakrishnan V. S, Hema Gogia, Preeti Madan, Sandhya Kabra, Sujeet K Singh, Partha Rakshit |
| hCoV-19/India/UT-NCDC-380911/2021 | EPI_ISL_4446596 | 2021-05-22 | National Centre for Disease Control (NCDC) Biotechnology Division, Delhi - INSACOG | NCDC Delhi, Biotechnology Division - INSACOG | Mahesh S Dhar, Kalaiarasan Ponnusamy, Meena Datta, Priyanka Singh,¬†Uma Sharma,¬†Hemlata Lall, Manoj K Singh, Radhakrishnan V. S, Robin Marwal, Hema Gogia, Preeti Madan, Sandhya Kabra, Sujeet K Singh, Partha Rakshit |
| hCoV-19/India/MP-NCDC-3809106/2021 | EPI_ISL_4446868 | 2021-05-02 | National Centre for Disease Control (NCDC) Biotechnology Division, Delhi - INSACOG | NCDC Delhi, Biotechnology Division - INSACOG | Mahesh S Dhar, Kalaiarasan Ponnusamy, Meena Datta, Priyanka Singh,¬†Uma Sharma,¬†Hemlata Lall, Manoj K Singh, Radhakrishnan V. S, Robin Marwal, Hema Gogia, Preeti Madan, Sandhya Kabra, Sujeet K Singh, Partha Rakshit |
| hCoV-19/Denmark/DCGC-173667/2021 | EPI_ISL_4448145 | 2021-09-15 | Department of Bacteria, Parasites and Fungi, Statens Serum Institut, Copenhagen, Denmark | Statens Serum Institut Bioinformatics and Microbial Genomics | Danish Covid-19 Genome Consortium |
| hCoV-19/Germany/NW-HHU-10990/2021 | EPI_ISL_4448517 | 2021-09-13 | ZOTZ KLIMAS MVZ D√ºsseldorf-Centrum GbR √úBAG f√ºr Labormedizin, Genetik, Zytologie, Pathologie | Center of Medical Microbiology, Virology, and Hospital Hygiene, University of Duesseldorf | Maximilian Damagnez; Alexander Dilthey; Patrick Finzer; Katrin Hoffmann; Torsten Houwaart; Lisanna H√ºlse; Malte Kohns Vasconcelos; Nadine L√ºbke; Jessica Nicolai; Klaus Pfeffer; Daniel Strelow; J√∂rg Timm; Andreas Walker; Tobias Wienemann; Rainer Zotz |
| hCoV-19/Germany/NW-HHU-10193/2021 | EPI_ISL_4456777 | 2021-09-01 | ZOTZ KLIMAS MVZ D√ºsseldorf-Centrum GbR √úBAG f√ºr Labormedizin, Genetik, Zytologie, Pathologie | Center of Medical Microbiology, Virology, and Hospital Hygiene, University of Duesseldorf | Maximilian Damagnez; Alexander Dilthey; Patrick Finzer; Katrin Hoffmann; Torsten Houwaart; Lisanna H√ºlse; Malte Kohns Vasconcelos; Nadine L√ºbke; Jessica Nicolai; Klaus Pfeffer; Daniel Strelow; J√∂rg Timm; Andreas Walker; Tobias Wienemann; Rainer Zotz |
| hCoV-19/India/MP-NCDC-360921/2021 | EPI_ISL_4459970 | 2021-05-01 | National Centre for Disease Control (NCDC) Biotechnology Division, Delhi - INSACOG | NCDC Delhi, Biotechnology Division - INSACOG | Robin Marwal, Mahesh S Dhar, Kalaiarasan Ponnusamy, Meena Datta, Priyanka Singh,¬†Uma Sharma,¬†Hemlata Lall, Manoj K Singh, Radhakrishnan V. S, Hema Gogia, Preeti Madan, Sandhya Kabra, Sujeet K Singh, Partha Rakshit |

| | | | | | |
|---|---|---|---|---|---|
| hCoV-19/India/PB-NCDC-4509307/2021 | EPI_ISL_4460812 | 2021-06-16 | National Centre for Disease Control (NCDC) Biotechnology Division, Delhi - INSACOG | NCDC Delhi, Biotechnology Division - INSACOG | Kalaiarasan Ponnusamy, Meena Datta, Priyanka Singh,¬†Uma Sharma,¬†Hemlata Lall, Manoj K Singh, Radhakrishnan V. S, Robin Marwal, Mahesh S Dhar, Hema Gogia, Preeti Madan, Sandhya Kabra, Sujeet K Singh, Partha Rakshit |
| hCoV-19/India/PB-NCDC-3109277/2021 | EPI_ISL_4462018 | 2021-05-09 | National Centre for Disease Control (NCDC) Biotechnology Division, Delhi - INSACOG | NCDC Delhi, Biotechnology Division - INSACOG | Radhakrishnan V. S, Robin Marwal, Mahesh S Dhar, Kalaiarasan Ponnusamy, Meena Datta, Priyanka Singh,¬†Uma Sharma,¬†Hemlata Lall, Manoj K Singh, Hema Gogia, Preeti Madan, Sandhya Kabra, Sujeet K Singh, Partha Rakshit |
| hCoV-19/India/MP-NCDC-3109341/2021 | EPI_ISL_4462051 | 2021-04-26 | National Centre for Disease Control (NCDC) Biotechnology Division, Delhi - INSACOG | NCDC Delhi, Biotechnology Division - INSACOG | Radhakrishnan V. S, Robin Marwal, Mahesh S Dhar, Kalaiarasan Ponnusamy, Meena Datta, Priyanka Singh,¬†Uma Sharma,¬†Hemlata Lall, Manoj K Singh, Hema Gogia, Preeti Madan, Sandhya Kabra, Sujeet K Singh, Partha Rakshit |
| hCoV-19/India/JK-NCDC-4209113/2021 | EPI_ISL_4462969 | 2021-06-08 | National Centre for Disease Control (NCDC) Biotechnology Division, Delhi - INSACOG | NCDC Delhi, Biotechnology Division - INSACOG | Kalaiarasan Ponnusamy, Meena Datta, Priyanka Singh,¬†Uma Sharma,¬†Hemlata Lall, Manoj K Singh, Radhakrishnan V. S, Robin Marwal, Mahesh S Dhar, Hema Gogia, Preeti Madan, Sandhya Kabra, Sujeet K Singh, Partha Rakshit |
| hCoV-19/India/MP-NCDC-3209165/2021 | EPI_ISL_4463537 | 2021-05-07 | National Centre for Disease Control (NCDC) Biotechnology Division, Delhi - INSACOG | NCDC Delhi, Biotechnology Division - INSACOG | Radhakrishnan V. S, Robin Marwal, Mahesh S Dhar, Kalaiarasan Ponnusamy, Meena Datta, Priyanka Singh,¬†Uma Sharma,¬†Hemlata Lall, Manoj K Singh, Hema Gogia, Preeti Madan, Sandhya Kabra, Sujeet K Singh, Partha Rakshit |
| hCoV-19/Brazil/SP-IB_130002/2021 | EPI_ISL_4469915 | 2021-08-29 | INSIDE DIAGN√ìSTICOS | Instituto Butantan | Dimas Tadeu Covas, Antonio Jorge Martins, Claudia Renata dos Santos Barros, David Schlesinger, Debora Botequio Moretti, Elaine Cristina Marqueze, Elaine Vieira Santos, Evandra Strazza Rodrigues, Heidge Fukumasu, Jayme Augusto de Souza-Neto, Jos√© Salvator |
| hCoV-19/USA/WY-WYPHL-21049304/2021 | EPI_ISL_4470572 | 2021-07-07 | Wyoming Public Health Laboratory | Wyoming Public Health Laboratory | Jim Mildenberger, Taylor Fearing, Channing Weber, Ashley Norberg, Chayse Rowley, Marley Goetz, Brian Dominguez, Elliot Thomasson, Sam Britz, Cari Sloma, Robert Petit, and Rob Christensen |
| hCoV-19/Malawi/CERI-KRISP-K022861/2021 | EPI_ISL_4473170 | 2021-07-26 | National HIV Reference Laboratory, Ministry of Health, Public Health Institute of Malawi | CERI, Centre for Epidemic Response and Innvoation, Stellenbosch University and KRISP, KZN Research Innovation and Sequencing Platform, UKZN. | Mvula B, Chilima B, Chiwaula M, Mwangomba W, Panja L, Kasambara W, Auld A, Kim L, Kampira E, Kaba M, Wadonda N, Maida A, Giandhari J, Pillay S, Naidoo Y, Lessells R, Emmanuel SJ, Tegally H, Wilkinson E, de Oliveira T |
| hCoV-19/SouthAfrica/CERI-KRISP-K022794/2021 | EPI_ISL_4473197 | 2021-08-18 | NHLS Charlotte Maxeke Johannesburg Hospital | [illegible] and Innvoation, Stellenbosch University and KRISP, KZN Research Innovation and Sequencing Platform, UKZN. | Kathleen Subramoney, Florette Treurnicht, Giandhari Jennifer, Pillay Sureshnee, Yajna Ramphal, Naidoo Yeshnee, Tshiabuila Derek,Tegally Houriiyah, San James, Wilkinson Eduan, de Oliveira Tulio |

| | | | | | |
|---|---|---|---|---|---|
| hCoV-19/India/HP-NCDC-3909337/2021 | EPI_ISL_4474105 | 2021-05-22 | National Centre for Disease Control (NCDC) Biotechnology Division, Delhi - INSACOG | NCDC Delhi, Biotechnology Division - INSACOG | Mahesh S Dhar, Kalaiarasan Ponnusamy, Meena Datta, Priyanka Singh,¬†Uma Sharma,¬†Hemlata Lall, Manoj K Singh, Radhakrishnan V. S, Robin Marwal, Hema Gogia, Preeti Madan, Sandhya Kabra, Sujeet K Singh, Partha Rakshit |
| hCoV-19/India/PB-NCDC-3909357/2021 | EPI_ISL_4474115 | 2021-05-28 | National Centre for Disease Control (NCDC) Biotechnology Division, Delhi - INSACOG | NCDC Delhi, Biotechnology Division - INSACOG | Mahesh S Dhar, Kalaiarasan Ponnusamy, Meena Datta, Priyanka Singh,¬†Uma Sharma,¬†Hemlata Lall, Manoj K Singh, Radhakrishnan V. S, Robin Marwal, Hema Gogia, Preeti Madan, Sandhya Kabra, Sujeet K Singh, Partha Rakshit |
| hCoV-19/India/LA-NCDC-3909118/2021 | EPI_ISL_4474209 | 2021-05-23 | National Centre for Disease Control (NCDC) Biotechnology Division, Delhi - INSACOG | NCDC Delhi, Biotechnology Division - INSACOG | Mahesh S Dhar, Kalaiarasan Ponnusamy, Meena Datta, Priyanka Singh,¬†Uma Sharma,¬†Hemlata Lall, Manoj K Singh, Radhakrishnan V. S, Robin Marwal, Hema Gogia, Preeti Madan, Sandhya Kabra, Sujeet K Singh, Partha Rakshit |
| hCoV-19/Malawi/CERI-KRISP-K024706/2021 | EPI_ISL_4474393 | 2021-07-24 | National HIV Reference Laboratory, Ministry of Health, Public Health Institute of Malawi | CERI, Centre for Epidemic Response and Innovation | Mvula B, Chilima B, Chiwaula M, Mwangomba W, Panja L, Kasambara W, Auld A, Kim L, Kampira E, Kaba M, Wadonda N, Maida A, Giandhari J, Pillay S, Yajna Ramphal, Naidoo Yeshnee, Tshabuila Derek, Emmanuel S, Tegally H, Wilkinson E, de Oliveira T |
| hCoV-19/USA/CA-CDPH1941/2021 | EPI_ISL_4474653 | 2021-09-17 | California Department of Public Health | California Department of Public Health | CDPH IDLB COVIDNet et al |
| hCoV-19/Ghana/nCoV-TRA-1775/2021 | EPI_ISL_4253804 | 2021-08-16 | National Influenza Centre | National Influenza Centre | William K. Ampofo, Michael Marks, Ivy A. Asante, Sharon Hsu, Benjamiin B. Lindsey, Benjamin H. Foulkes, Mildred Adusei-Poku, Linda Boatemaa, Lorreta Kwasah, Joseph Oliver-Commey, Ernest Asiedu, Franklin Asiedu-Bekoe, Gordon Awandare, Joyce Ngoi, Dennis La |
| hCoV-19/Slovakia/UVZ_PL36_H5_17811/2021 | EPI_ISL_4484009 | 2021-02-09 | Public Health Authority of the Slovak Republic | Laboratory of Genomics and Bioinformatics, Comenius University Science Park | Tatiana Sedl√°ƒçkov√°, Diana Rus≈à√°kov√°, Miroslav B√∂hmer, Anna Gifçov√°, Jaroslav Budi≈°, Tom√°≈° Szemes, Jakub Styk |
| hCoV-19/Slovakia/UVZ_PL37_G2_18013/2021 | EPI_ISL_4484081 | 2021-04-09 | Public Health Authority of the Slovak Republic | Laboratory of Genomics and Bioinformatics, Comenius University Science Park | Tatiana Sedl√°ƒçkov√°, Diana Rus≈à√°kov√°, Miroslav B√∂hmer, Anna Gifçov√°, Jaroslav Budi≈°, Tom√°≈° Szemes, Jakub Styk |
| hCoV-19/Slovakia/UVZ_PL36_C6_17814/2021 | EPI_ISL_4484111 | 2021-02-09 | Public Health Authority of the Slovak Republic | Laboratory of Genomics and Bioinformatics, Comenius University Science Park | Tatiana Sedl√°ƒçkov√°, Diana Rus≈à√°kov√°, Miroslav B√∂hmer, Anna Gifçov√°, Jaroslav Budi≈°, Tom√°≈° Szemes, Jakub Styk |
| hCoV-19/Slovakia/UVZ_PL37_C8_18063/2021 | EPI_ISL_4484346 | 2021-01-09 | Public Health Authority of the Slovak Republic | Laboratory of Genomics and Bioinformatics, Comenius University Science Park | Tatiana Sedl√°ƒçkov√°, Diana Rus≈à√°kov√°, Miroslav B√∂hmer, Anna Gifçov√°, Jaroslav Budi≈°, Tom√°≈° Szemes, Jakub Styk |
| hCoV-19/Slovakia/UVZ_PL36_D3_17791/2021 | EPI_ISL_4484480 | 2021-02-09 | Public Health Authority of the Slovak Republic | Laboratory of Genomics and Bioinformatics, Comenius University Science Park | Tatiana Sedl√°ƒçkov√°, Diana Rus≈à√°kov√°, Miroslav B√∂hmer, Anna Gifçov√°, Jaroslav Budi≈°, Tom√°≈° Szemes, Jakub Styk |
| hCoV-19/Slovakia/UVZ_PL37_B5_18038/2021 | EPI_ISL_4484548 | 2021-03-09 | Public Health Authority of the Slovak Republic | Laboratory of Genomics and Bioinformatics, Comenius University Science Park | Tatiana Sedl√°ƒçkov√°, Diana Rus≈à√°kov√°, Miroslav B√∂hmer, Anna Gifçov√°, Jaroslav Budi≈°, Tom√°≈° Szemes, Jakub Styk |

| | | | | | |
|---|---|---|---|---|---|
| hCoV-19/Belgium/ULG-19382/2021 | EPI_ISL_4489665 | 2021-09-02 | Clinique Notre Dame √† Hermalle | GIGA Medical Genomics | Keith Durkin, Maria Artesi, Bouchra Boujemla, Nathalie Renotte, Nadine Cambisano, S√©bastien Bontems, C√©cile Meex, Claire Gourzon√®s, Olivier Ek, Laurent Gillet, Marie-Pierre Hayette, Vincent Bours |
| hCoV-19/Cambodia/783802/2021 | EPI_ISL_4489790 | 2021-09-16 | Virology Unit, Institut Pasteur du Cambodge | Virology Unit, Institut Pasteur du Cambodge | Leakhena Pum, Sophoannadedh Rath, Jurre Y Siegers, Cecile Troupin, Ly Sovann, Kraing Sidonn, Yi Sengdoeurn, Chin Savuth, Chau Darapheak, Veasna Duong, Erik A Karlsson |
| hCoV-19/Nigeria/CV2717/2021 | EPI_ISL_4490294 | 2021-08-09 | Rivers State University Teaching Hospital | Africa Centre for Excellence for Genomics of Infectious Diseases (ACEGID), Redeemer‚Äôs University | Olawoye, I.B., Oluniyi, P.E., Eromon, P.E., Oguzie, J.U., Kayode, A.T., Uwanibe, J.N., Ugwu, C.A., Akano, K.O., Ajogbasile, F.V., Abechi, P.S., Olumade, T.J., Nosamiefan, I., Folarin, O., Happi, C.T. |
| hCoV-19/USA/CA-CDPH-3000128013/2021 | EPI_ISL_4491179 | 2021-06-25 | CDPH VBL | California Department of Public Health | Emily Smith on behalf of CDPH-COVIDNet |
| hCoV-19/India/HP-NCDC-4309334/2021 | EPI_ISL_4492971 | 2021-06-16 | National Centre for Disease Control (NCDC) Biotechnology Division, Delhi - INSACOG | NCDC Delhi, Biotechnology Division - INSACOG | Kalaiarasan Ponnusamy, Meena Datta, Priyanka Singh,¬†Uma Sharma,¬†Hemlata Lall, Manoj K Singh, Radhakrishnan V. S, Robin Marwal, Mahesh S Dhar, Hema Gogia, Preeti Madan, Sandhya Kabra, Sujeet K Singh, Partha Rakshit |
| hCoV-19/India/HP-NCDC-4309382/2021 | EPI_ISL_4493095 | 2021-06-20 | National Centre for Disease Control (NCDC) Biotechnology Division, Delhi - INSACOG | NCDC Delhi, Biotechnology Division - INSACOG | Kalaiarasan Ponnusamy, Meena Datta, Priyanka Singh,¬†Uma Sharma,¬†Hemlata Lall, Manoj K Singh, Radhakrishnan V. S, Robin Marwal, Mahesh S Dhar, Hema Gogia, Preeti Madan, Sandhya Kabra, Sujeet K Singh, Partha Rakshit |
| hCoV-19/Canada/MB-NML-157706/2021 | EPI_ISL_4495045 | 2021-07-12 | Dynacare Manitoba | Dynacare Manitoba | Hilary Racher, Melissa Desaulnier, Tintu Abraham, Hongbin Li, David Alexander |
| hCoV-19/India/GJ-INSACOG-GBRC1508/2021 | EPI_ISL_3717832 | 2021-05-19 | Government Medical College (GMC), Surat | Gujarat Biotechnology Research Centre | Janvi Raval,Zarna Patel,Dinesh Kumar,Arpit Shukla,Bhadreshsinh Gohil,Nimesh Patel,Nitin Shukla,Ramesh Pandit,Neeta Khandelwal,Chaitanya Joshi,Madhvi Joshi |
| hCoV-19/USA/CA-Stanford-24_S40/2021 | EPI_ISL_4496842 | 2021-04-23 | Stanford Health Care | Stanford University School of Medicine, Clinical Virology Laboratory | Malaya K. Sahoo, Becky Jiang, Selamawit Bihon, James Zehnder, and Benjamin A. Pinsky |
| hCoV-19/India/WB-INSACOG-1930400632588/2021 | EPI_ISL_4504301 | 2021-08-04 | INSACOG-WB | National Institute of Biomedical Genomics - INSACOG | Arindam Maitra, Bhaswati Bandyopadhyay, Nidhan Kumar Biswas, Tamal Ghosh, Sreedhar Chinnaswamy, Ajay Chakraborti, Saumitra Das |
| hCoV-19/India/WB-INSACOG-1966400167169/2021 | EPI_ISL_4504396 | 2021-08-03 | INSACOG-WB | National Institute of Biomedical Genomics - INSACOG | Arindam Maitra, Bhaswati Bandyopadhyay, Nidhan Kumar Biswas, Tamal Ghosh, Sreedhar Chinnaswamy, Ajay Chakraborti, Saumitra Das |
| hCoV-19/Kenya/KEM-CVR-100390/2021 | EPI_ISL_4505350 | 2021-04-19 | ARI lab, Center for Virus Research | ARI lab, Center for Virus Research | Silvanos Mukunzi Opanda, Wallace Bulimo, Samoel Ashimosi Khamadi, Samwel Lifumo Symekher, Samson Konongoi Limbaso, Janet Majanja, Solomon Langat, Meshack Wadegu,Rachel Achilla, Esther Chitechi, Milkah Mwangi, Nicholas Mwikwabe, Seth Okeyo, Edith Koskei, J |

| | | | | | |
|---|---|---|---|---|---|
| hCoV-19/Indonesia/JK-NIHRD-MI2112001/2021 | EPI_ISL_4508290 | 2021-08-06 | WISMA ATLET | National Institute of Health Research and Development | Hana Apsari Pawestri, Subangkit, Kartika Dewi Puspa, Arie Ardiansyah Nugraha, Hartanti Dian Ikawati, Krisna Pangesti, Triyani Soekarso, Fajar Nur Sulistiyohadi, Nelly Puspandari, Vivi Setiawaty |
| hCoV-19/Zambia/ZMB-107635/2021 | EPI_ISL_4513618 | 2021-06-30 | University of Zambia, School of Veterinary Medicine | UNZAVET and PATH | Mulenga Mwenda-Chimfwembe, Ngonda Saasa, Daniel Bridges, ZNPHI and ZGSC |
| hCoV-19/Zambia/ZMB-107636/2021 | EPI_ISL_4513619 | 2021-06-30 | University of Zambia, School of Veterinary Medicine | UNZAVET and PATH | Mulenga Mwenda-Chimfwembe, Ngonda Saasa, Daniel Bridges, ZNPHI and ZGSC |
| hCoV-19/Zambia/ZMB-108404/2021 | EPI_ISL_4513632 | 2021-06-30 | University of Zambia, School of Veterinary Medicine | UNZAVET and PATH | Mulenga Mwenda-Chimfwembe, Ngonda Saasa, Daniel Bridges, ZNPHI and ZGSC |
| hCoV-19/Zambia/ZMB-108731/2021 | EPI_ISL_4513654 | 2021-07-05 | University of Zambia, School of Veterinary Medicine | UNZAVET and PATH | Mulenga Mwenda-Chimfwembe, Ngonda Saasa, Daniel Bridges, ZNPHI and ZGSC |
| hCoV-19/England/PHEC-3G04BGF9/2021 | EPI_ISL_4529921 | 2021-09-23 | Respiratory Virus Unit, Microbiology Services Colindale, Public Health England | COVID-19 Genomics UK (COG-UK) Consortium | PHE Covid Sequencing Team |
| hCoV-19/England/PHEC-3G04DG99/2021 | EPI_ISL_4529942 | 2021-09-23 | Respiratory Virus Unit, Microbiology Services Colindale, Public Health England | COVID-19 Genomics UK (COG-UK) Consortium | PHE Covid Sequencing Team |
| hCoV-19/England/PHEC-3G04EG69/2021 | EPI_ISL_4529952 | 2021-09-23 | Respiratory Virus Unit, Microbiology Services Colindale, Public Health England | COVID-19 Genomics UK (COG-UK) Consortium | PHE Covid Sequencing Team |
| hCoV-19/England/PHEC-3G076G13/2021 | EPI_ISL_4530226 | 2021-09-14 | Respiratory Virus Unit, Microbiology Services Colindale, Public Health England | COVID-19 Genomics UK (COG-UK) Consortium | PHE Covid Sequencing Team |
| hCoV-19/Slovakia/UVZ_PL36_B3_17789/2021 | EPI_ISL_4533755 | 2021-01-09 | Public Health Authority of the Slovak Republic | Laboratory of Genomics and Bioinformatics, Comenius University Science Park | Tatiana Sedl√°ƒçkov√°, Diana Rus≈à√°kov√°, Miroslav B√∂hmer, Anna Giƒçov√°, Jaroslav Budi≈°, Tom√°≈° Szemes, Jakub Styk |
| hCoV-19/India/GJ-INSACOG-GBRC2210/2021 | EPI_ISL_4533952 | 2021-09-01 | Sterling Accuris, Bhavnagar | Gujarat Biotechnology Research Centre | Janvi Raval,Apurvasinh Puvar,Zarna Patel,Dinesh Kumar,Arpit Shukla,Bhadreshsinh Gohil,Nimesh Patel,Nitin Shukla,Ramesh Pandit,Nitin Savaliya,Priyank Chavda,Sonal Sharma ,Hemal Salot,Chaitanya Joshi,Madhvi Joshi |
| hCoV-19/USA/RI-CDC-ASC210181367/2021 | EPI_ISL_4535667 | 2021-09-01 | Aegis Sciences Corporation | Centers for Disease Control and Prevention Division of Viral Diseases, Pathogen Discovery | Dakota Howard,Dhwani Batra,Peter Cook,Jason Caravas,Benjamin Rambo-Martin,Scott Sammons,Yvette Unoarumhi,Matthew Schmerer,Kristine Lacek,Tymeckia Kendall,Victoria Caban Figueroa,Shatavia Morrison,Christopher Gulvick,Erisa Sula,Cyndi Clark,Patrick Campbell |
| hCoV-19/Sweden/01_SE100_21CS510269/2021 | EPI_ISL_4535989 | 2021-09-13 | Karolinska University Hospital Solna | Karolinska University Hospital | Jan Albert ,Tobias Allander ,Annelie Bjerkner ,Sandra Broddesson ,Robert Dyrdak ,Martin Ekman ,Lynda Eneh ,Lina Guerra Blomqvist ,Karolina Ininbergs ,Tanja Normark ,Isak Sylvin ,Zhibing Yun ,Martina Wahlund ,Valtteri Wirta |
| hCoV-19/Russia/MOS-CRIE-L106P3037u/2021 | EPI_ISL_4536728 | 2021-03-26 | Molecular diagnostic laboratory of Federal Budget Institution of Science "Central Research Institute of Epidemiology" of The Federal Service on Customers' Rights Protection and Human Well-being Surveillance | Group of Genomics and Postgenomic Technologies of Central Research Institute of Epidemiology | Samoilov A.E., Korneenko E.V., Smirnova Y.S., Nadtoka M.I., Sinitsyn S.O., Roev G.V., Buharina A.Y., Vyhodceva A.V., Shipulina O.Y., Svetlichnyj D.V., Speranskaya A.S., Kondrasheva L.Y., Akimkin V.G. |

| | | | | | |
|---|---|---|---|---|---|
| hCoV-19/USA/GU-CDC-2-4918946/2021 | EPI_ISL_4539464 | 2021-07-26 | Guam Public Health Laboratory | Centers for Disease Control and Prevention Division of Viral Diseases, Pathogen Discovery | Mili Sheth,Sarah Nobles,Jasmine Padilla,Alex Burgin,Meghan Bentz,Mark Burroughs,Peter Cook,Clinton Paden,Dhwani Batra,Krista Queen,Kristen Knipe,Dakota Howard,Yvette Unoarumhi,Matthew Schmerer,Ben Rambo-Martin,Kristine Lacek,Sam Shepard,Dave Wentworth,Viv |
| hCoV-19/USA/GU-CDC-2-4919006/2021 | EPI_ISL_4539491 | 2021-08-04 | Guam Public Health Laboratory | Centers for Disease Control and Prevention Division of Viral Diseases, Pathogen Discovery | Mili Sheth,Sarah Nobles,Jasmine Padilla,Alex Burgin,Meghan Bentz,Mark Burroughs,Peter Cook,Clinton Paden,Dhwani Batra,Krista Queen,Kristen Knipe,Dakota Howard,Yvette Unoarumhi,Matthew Schmerer,Ben Rambo-Martin,Kristine Lacek,Sam Shepard,Dave Wentworth,Viv |
| hCoV-19/USA/GU-CDC-2-4919045/2021 | EPI_ISL_4539523 | 2021-08-09 | Guam Public Health Laboratory | Centers for Disease Control and Prevention Division of Viral Diseases, Pathogen Discovery | Mili Sheth,Sarah Nobles,Jasmine Padilla,Alex Burgin,Meghan Bentz,Mark Burroughs,Peter Cook,Clinton Paden,Dhwani Batra,Krista Queen,Kristen Knipe,Dakota Howard,Yvette Unoarumhi,Matthew Schmerer,Ben Rambo-Martin,Kristine Lacek,Sam Shepard,Dave Wentworth,Viv |
| hCoV-19/USA/IL-RIPHL_10357_V/2021 | EPI_ISL_4542893 | 2021-08-11 | Rush University Medical Center | RIPHL at Rush University Medical Center | Stefan Green, Kevin Kunstman, Marieta Hyde, Laura Furtado, Felix Araujo Perez, Edith Perez, Sofiya Bobrovska, Mary Hayden, Joyce Houlihan, Diane Springer |
| hCoV-19/Bonaire/BQ-RIVM-59022/2021 | EPI_ISL_4543517 | 2021-09-05 | Dutch COVID-19 response team | National Institute for Public Health and the Environment (RIVM) | Adam Meijer, Harry Vennema, Dirk Eggink, Jeroen Cremer, Sharon van den Brink, Bas van der Veer, AnneMarie van den Brandt, Lisa Wijsman, Kim Freriks, Ryanne Jaarsma, Eun√≠ce Then, Lynn Aarts, Sanne Bos, Stijn van Rossum, Florian Zwagemaker, Dennis Schmitz, |
| hCoV-19/Netherlands/UT-RIVM-59668/2021 | EPI_ISL_4544363 | 2021-08-27 | Dutch COVID-19 response team | National Institute for Public Health and the Environment (RIVM) | Adam Meijer, Harry Vennema, Dirk Eggink, Jeroen Cremer, Sharon van den Brink, Bas van der Veer, AnneMarie van den Brandt, Lisa Wijsman, Kim Freriks, Ryanne Jaarsma, Eun√≠ce Then, Lynn Aarts, Sanne Bos, Stijn van Rossum, Florian Zwagemaker, Dennis Schmitz, |
| hCoV-19/Netherlands/NB-RIVM-58271/2021 | EPI_ISL_4544768 | 2021-09-05 | Dutch COVID-19 response team | National Institute for Public Health and the Environment (RIVM) | Adam Meijer, Harry Vennema, Dirk Eggink, Jeroen Cremer, Sharon van den Brink, Bas van der Veer, AnneMarie van den Brandt, Lisa Wijsman, Kim Freriks, Ryanne Jaarsma, Eun√≠ce Then, Lynn Aarts, Sanne Bos, Stijn van Rossum, Florian Zwagemaker, Dennis Schmitz, |
| hCoV-19/Kosovo/CO-00367-XXK000_9329/2021 | EPI_ISL_4545860 | 2021-07-28 | Department of Microbiology, National Institute of Public Health of Kosovo | Department of Microbiology, National Institute of Public Health of Kosovo | Xhevat Jakupi, Zana Deva, Pranvera Abazi, Donjeta Hajdari, Aferdita Hyseni, Nazmi Mehmeti, Blendi Jerliu, Robert Ramadani |
| hCoV-19/India/AP-CCMB-CIA7945/2021 | EPI_ISL_4545926 | 2021-08-08 | AP SSO | CSIR-Centre for Cellular and Molecular Biology - INSACOG | Sofia Banu,Payel Mukherjee, Lamuk Zaveri, Onkar Kulkarni,Tulasi Nagabandi, Jandhyala Sai Krishna, Ara Sreenivas,Shreekant Verma, Amareshwar Vodapalli ,B Himasri,Valli Nagalakshmi Undamatla, Sumedha Avadhanula, Vidhyadhari Methuku, Priya Nurkurthy, Archana |

| | | | | | |
|---|---|---|---|---|---|
| hCoV-19/India/AP-CCMB-CIA8125/2021 | EPI_ISL_4546085 | 2021-08-23 | AP SSO | CSIR-Centre for Cellular and Molecular Biology - INSACOG | Shreekant Verma, Amareshwar Vodapalli ,Ara Sreenivas,B Himasri,Valli Nagalakshmi Undamatla,Payel Mukherjee, Lamuk Zaveri, Onkar Kulkarni,Tulasi Nagabandi, Sofia Banu, Jandhyala Sai Krishna,Sumedha Avadhanula, Priya Nurkurthy, Vidhyadhari Methuku, Archana |
| hCoV-19/Colombia/ANT-CWOHC-VG-SEC01738V/2021 | EPI_ISL_4551031 | 2021-08-31 | AYUDAS DIAGNOSTICAS SURA | Universidad Nacional de Colombia - Laboratorio Gen√≥mico One Health | Maria Stella L√≥pez, Andres F. Cardona-Rios, Daniel O. Maldonado-Perez, Sim√≥n Villegas Vel√°squez, Lina M. Hurtado, Laura Silvana Perez, Karl A Ciuoderis, Maria Ang√©lica Maya, Carolina Mu√±oz-Arango, Celeny Ortiz, Rita Almanza Payares, Idabely Betancur |
| hCoV-19/Slovakia/UVZ_PL36_B11_17969/2021 | EPI_ISL_4551293 | 2021-02-09 | Public Health Authority of the Slovak Republic | Laboratory of Genomics and Bioinformatics, Comenius University Science Park | Tatiana Sedl√°ƒçkov√°, Diana Rus≈à√°kov√°, Miroslav B√∂hmer, Anna Gifçov√°, Jaroslav Budi≈°, Tom√°≈° Szemes, Jakub Styk |
| hCoV-19/Slovakia/UVZ_PL36_G2_17786/2021 | EPI_ISL_4551303 | 2021-01-09 | Public Health Authority of the Slovak Republic | Laboratory of Genomics and Bioinformatics, Comenius University Science Park | Tatiana Sedl√°ƒçkov√°, Diana Rus≈à√°kov√°, Miroslav B√∂hmer, Anna Gifçov√°, Jaroslav Budi≈°, Tom√°≈° Szemes, Jakub Styk |
| hCoV-19/Slovakia/UVZ_PL37_E1_18003/2021 | EPI_ISL_4551306 | 2021-02-09 | Public Health Authority of the Slovak Republic | Laboratory of Genomics and Bioinformatics, Comenius University Science Park | Tatiana Sedl√°ƒçkov√°, Diana Rus≈à√°kov√°, Miroslav B√∂hmer, Anna Gifçov√°, Jaroslav Budi≈°, Tom√°≈° Szemes, Jakub Styk |
| hCoV-19/USA/CA-CDC-ASC210181805/2021 | EPI_ISL_4554223 | 2021-09-01 | Aegis Sciences Corporation | Centers for Disease Control and Prevention Division of Viral Diseases, Pathogen Discovery | Dakota Howard,Dhwani Batra,Peter Cook,Jason Caravas,Benjamin Rambo-Martin,Scott Sammons,Yvette Unoarumhi,Matthew Schmerer,Kristine Lacek,Tymeckia Kendall,Victoria Caban Figueroa,Shatavia Morrison,Christopher Gulvick,Erisa Sula,Cyndi Clark,Patrick Campbell |
| hCoV-19/Portugal/PT17010/2021 | EPI_ISL_4555703 | 2021-09-13 | Avelab | Instituto Nacional de Saude (INSA) | Borges et al |
| hCoV-19/India/WB-IICB-S165-08/2021 | EPI_ISL_4558073 | 2021-08-08 | MEDICA Superspeciality Hospital | CSIR-Indian Institute of Chemical Biology | Siddik Sarkar, Saikat Chakrabarti, Partha Chakrabarti, Poulomi Sarkar, Sarthak Banerjee,Aviral Roy, Soumen Saha, Arpita Ghosh Mitra,Arun Bandyopadhyay |
| hCoV-19/USA/NC-CDC-QDX28740139/2021 | EPI_ISL_4566618 | 2021-09-13 | Quest Diagnostics Incorporated | Centers for Disease Control and Prevention Division of Viral Diseases, Pathogen Discovery | Dakota Howard,Dhwani Batra,Peter Cook,Jason Caravas,Benjamin Rambo-Martin,Scott Sammons,Yvette Unoarumhi,Matthew Schmerer,Kristine Lacek,Tymeckia Kendall,Victoria Caban Figueroa,Shatavia Morrison,Christopher Gulvick,Erisa Sula,S. Rosenthal,A. Gerasimova,R |
| hCoV-19/Benin/446709/2021 | EPI_ISL_4566898 | 2021-06-24 | Laboratoire des Fi√®vres H√©morragiques Virales du Benin | Institut f√ºr Virologie - Institute of Virology - Charite | Anges Yadouleton, Anna-Lena Sander, Praise Adewumi, Edmilson F. de Oliveira Filho, Carine Tchibozo, Gildas Hounkanrin, Keke K. Ren√©, Dossou Ange, Rodrigue K. Kohoun, Ramalia Chabi Nari, Sourakatou Salifou, Raoul Saizonou, Clement G. Kakai, Sonia V. Bedi√ |

| | | | | | |
|---|---|---|---|---|---|
| hCoV-19/Benin/453466/2021 | EPI_ISL_4566908 | 2021-06-30 | Laboratoire des Fi√®vres H√©morragiques Virales du Benin | Institut f√ºr Virologie - Institute of Virology - Charite | Anges Yadouleton, Anna-Lena Sander, Praise Adewumi, Edmilson F. de Oliveira Filho, Carine Tchibozo, Gildas Hounkanrin, Keke K. Ren√©, Dossou Ange, Rodrigue K. Kohoun, Ramalia Chabi Nari, Sourakatou Salifou, Raoul Saizonou, Clement G. Kakai, Sonia V. Bedi√ |
| hCoV-19/Benin/472982/2021 | EPI_ISL_4566938 | 2021-07-16 | Laboratoire des Fi√®vres H√©morragiques Virales du Benin | Institut f√ºr Virologie - Institute of Virology - Charite | Anges Yadouleton, Anna-Lena Sander, Praise Adewumi, Edmilson F. de Oliveira Filho, Carine Tchibozo, Gildas Hounkanrin, Keke K. Ren√©, Dossou Ange, Rodrigue K. Kohoun, Ramalia Chabi Nari, Sourakatou Salifou, Raoul Saizonou, Clement G. Kakai, Sonia V. Bedi√ |
| hCoV-19/Benin/FH1973/2021 | EPI_ISL_4566992 | 2021-07-09 | Laboratoire des Fi√®vres H√©morragiques Virales du Benin | Institut f√ºr Virologie - Institute of Virology - Charite | Anges Yadouleton, Anna-Lena Sander, Praise Adewumi, Edmilson F. de Oliveira Filho, Carine Tchibozo, Gildas Hounkanrin, Keke K. Ren√©, Dossou Ange, Rodrigue K. Kohoun, Ramalia Chabi Nari, Sourakatou Salifou, Raoul Saizonou, Clement G. Kakai, Sonia V. Bedi√ |
| hCoV-19/USA/CA-CDC-FG-129418/2021 | EPI_ISL_4567932 | 2021-09-17 | Fulgent Genetics | Centers for Disease Control and Prevention Division of Viral Diseases, Pathogen Discovery | Dakota Howard,Dhwani Batra,Peter Cook,Jason Caravas,Benjamin Rambo-Martin,Scott Sammons,Yvette Unoarumhi,Matthew Schmerer,Kristine Lacek,Tymeckia Kendall,Victoria Caban Figueroa,Shatavia Morrison,Christopher Gulvick,Erisa Sula,Harry Gao,Mickey Li,John Gao |
| hCoV-19/USA/CA-CDC-FG-129662/2021 | EPI_ISL_4568169 | 2021-09-18 | Fulgent Genetics | Centers for Disease Control and Prevention Division of Viral Diseases, Pathogen Discovery | Dakota Howard,Dhwani Batra,Peter Cook,Jason Caravas,Benjamin Rambo-Martin,Scott Sammons,Yvette Unoarumhi,Matthew Schmerer,Kristine Lacek,Tymeckia Kendall,Victoria Caban Figueroa,Shatavia Morrison,Christopher Gulvick,Erisa Sula,Harry Gao,Mickey Li,John Gao |
| hCoV-19/USA/CA-CDC-FG-130143/2021 | EPI_ISL_4568542 | 2021-09-20 | Fulgent Genetics | Centers for Disease Control and Prevention Division of Viral Diseases, Pathogen Discovery | Dakota Howard,Dhwani Batra,Peter Cook,Jason Caravas,Benjamin Rambo-Martin,Scott Sammons,Yvette Unoarumhi,Matthew Schmerer,Kristine Lacek,Tymeckia Kendall,Victoria Caban Figueroa,Shatavia Morrison,Christopher Gulvick,Erisa Sula,Harry Gao,Mickey Li,John Gao |
| hCoV-19/India/JK-NCDC-4009162/2021 | EPI_ISL_4569785 | 2021-06-01 | National Centre for Disease Control (NCDC) Biotechnology Division, Delhi - INSACOG | NCDC Delhi, Biotechnology Division - INSACOG | Mahesh S Dhar, Kalaiarasan Ponnusamy, Meena Datta, Priyanka Singh,¬†Uma Sharma,¬†Hemlata Lall, Manoj K Singh, Radhakrishnan V. S, Robin Marwal, Hema Gogia, Preeti Madan, Sandhya Kabra, Sujeet K Singh, Partha Rakshit |
| hCoV-19/Congo/FCRM-55-9674/2021 | EPI_ISL_4572266 | 2021-07-27 | Fondation Congolaise pour la recherche medicale (FCRM), Francine Ntoumi | Fondation Congolaise pour la Recherche M√©dicale | Mfoutou Mapanguy Claujens Chastel; Batchi-Bouyou Armel Landry; Dr. Jean Claude Djontu; Dr. Abel Lissom; Prof. Dr. Thirumalaisamy P. Velavan; Prof. Francine Ntoumi |
| hCoV-19/Morocco/FMP-338/2021 | EPI_ISL_4572736 | 2021-07-13 | Institut National d'Hygi√®ne / Consortium National de Veille G√©nomique - Maroc | Laboratoire de Biotechnologie | Mouna Ouadghiri, Safaa ElMazouri, Tarik Aanniz, Saa√Ød Amzazi, Lahcen Belyamani and Azeddine Ibrahimi |

| | | | | | |
|---|---|---|---|---|---|
| hCoV-19/Montenegro/CO-00421_MNE000_2746672102/2021 | EPI_ISL_4578355 | 2021-08-18 | Laboratory for Molecular Diagnostics, IPHMN | Eurofins Genomics Europe Sequencing GmbH | Danijela Vujo≈°evifá, Marija Govedarica |
| hCoV-19/India/KA-NIMH-SEQ-2753/2021 | EPI_ISL_4578361 | 2021-08-20 | AURIGA LABS | INSACOG-KA, NIMHANS | Anson Kunjumon George,Ellango Ramasamy, Sony Sharma, Mahesh Kumar.C.S, Ananthapadmanabha Kotambail, Chetan G K, Gautham Arunachal Udupi, Anita S Desai, V Ravi |
| hCoV-19/Ghana/nCoV-TRA-889/2021 | EPI_ISL_4602044 | 2021-04-26 | National Influenza Centre | National Influenza Centre | William K. Ampofo, Michael Marks, Ivy A. Asante, Sharon Hsu, Benjamiin B. Lindsey, Benjamin H. Foulkes, Mildred Adusei-Poku, Linda Boatemaa, Lorreta Kwasah, Joseph Oliver-Commey, Ernest Asiedu, Franklin Asiedu-Bekoe, Gordon Awandare, Joyce Ngoi, Dennis La |
| hCoV-19/Ghana/nCoV-TRA-713/2021 | EPI_ISL_4602069 | 2021-04-18 | National Influenza Centre | National Influenza Centre | William K. Ampofo, Michael Marks, Ivy A. Asante, Sharon Hsu, Benjamiin B. Lindsey, Benjamin H. Foulkes, Mildred Adusei-Poku, Linda Boatemaa, Lorreta Kwasah, Joseph Oliver-Commey, Ernest Asiedu, Franklin Asiedu-Bekoe, Gordon Awandare, Joyce Ngoi, Dennis La |
| hCoV-19/Australia/WA812/2021 | EPI_ISL_4602669 | 2021-09-25 | PathWest Laboratory Medicine WA | PathWest Laboratory Medicine WA | PathWest Laboratory Medicine WA Microbial Surveillance Unit |
| hCoV-19/France/ARA-HCL021160730601/2021 | EPI_ISL_4604604 | 2021-08-31 | LABORATOIRE UNILIANS DECINES | CNR Virus des Infections Respiratoires - France SUD | Antonin Bal, Gregory Destras, Gwendolyne Burfin, Hadrien Regue, Quentin Semanas, Martine Valette, Bruno Lina, Laurence Josset |
| hCoV-19/France/PAC-HCL021161519201/2021 | EPI_ISL_4605052 | 2021-08-30 | CERBALLIANCE COTE D'AZUR | CNR Virus des Infections Respiratoires - France SUD | Antonin Bal, Gregory Destras, Gwendolyne Burfin, Hadrien Regue, Quentin Semanas, Martine Valette, Bruno Lina, Laurence Josset |
| hCoV-19/Switzerland/SG-CLM-09216443/2021 | EPI_ISL_4624916 | 2021-09-21 | Center for Laboratory Medicine | Center for Laboratory Medicine | Yannick Gerth |
| hCoV-19/USA/CA-SDCPHL-092121-070/2021 | EPI_ISL_4629119 | 2021-08-11 | San Diego County Public Health Laboratory | San Diego County Public Health Laboratory | Ashleigh Murphy Schafer, Brett Austin |
| hCoV-19/Thailand/ChiangMai_SEQ16054/2021 | EPI_ISL_4634614 | 2021-09-08 | Regional Medical Sciences Center 1 Chiang Mai | National Institute of Health, Department of Medical Sciences, Ministry of Public Health, Thailand | Pilailuk Okada; Siripaporn Phuygun; Nuttida Thongpramul; Sittiporn Parnmen; Ratana Tacharoenmuang; Pakorn Piromtong; Natchaya¬† Khiadsang; Thanutsapa Thanadachakul; Warawan Wongboot; sirikanda wimol; Sunthareeya Waicharoen; Archawin Rojanawiwat |
| hCoV-19/Thailand/Songkhla_SEQ16080/2021 | EPI_ISL_4634615 | 2021-09-03 | Regional Medical Sciences Center 12 Songkhla | National Institute of Health, Department of Medical Sciences, Ministry of Public Health, Thailand | Pilailuk Okada; Siripaporn Phuygun; Nuttida Thongpramul; Sittiporn Parnmen; Ratana Tacharoenmuang; Pakorn Piromtong; Natchaya¬† Khiadsang; Thanutsapa Thanadachakul; Warawan Wongboot; sirikanda wimol; Sunthareeya Waicharoen; Archawin Rojanawiwat |
| hCoV-19/Norway/19669/2021 | EPI_ISL_4634775 | 2021-09-06 | Department Medical Microbiology, Baerum Hospital, Vestre Viken Health Trust | Norwegian Institute of Public Health, Department of Virology | Kathrine Stene-Johansen, Kamilla Heddeland Instefjord, Hilde Elshaug, Garcia Llorente Ignacio, Jon Br√•te, Engebretsen Serina Beate,Pedersen Benedikte Nevjen, Line Victoria Moen, Debech Nadia, Atiya R Ali,Marie Paulsen Madsen, Rasmus Riis Kopperud, Hilde |

| | | | | | |
|---|---|---|---|---|---|
| hCoV-19/Thailand/Narathiwat_SEQ16799/2021 | EPI_ISL_4635432 | 2021-09-14 | Regional Medical Sciences Center 12 Songkhla | National Institute of Health, Department of Medical Sciences, Ministry of Public Health, Thailand | Pilailuk Okada; Siripaporn Phuygun; Nuttida Thongpramul; Sittiporn Parnmen; Ratana Tacharoenmuang; Pakorn Piromtong; Natchaya¬† Khiadsang; Thanutsapa Thanadachakul; Warawan Wongboot; sirikanda wimol; Sunthareeya Waicharoen; Archawin Rojanawiwat |
| hCoV-19/Thailand/Narathiwat_SEQ16089/2021 | EPI_ISL_4636421 | 2021-09-02 | Regional Medical Sciences Center 12 Songkhla | National Institute of Health, Department of Medical Sciences, Ministry of Public Health, Thailand | Pilailuk Okada; Siripaporn Phuygun; Nuttida Thongpramul; Sittiporn Parnmen; Ratana Tacharoenmuang; Pakorn Piromtong; Natchaya¬† Khiadsang; Thanutsapa Thanadachakul; Warawan Wongboot; sirikanda wimol; Sunthareeya Waicharoen; Archawin Rojanawiwat |
| hCoV-19/SouthAfrica/NICD-N15560/2021 | EPI_ISL_4649729 | 2021-08-24 | KIMBERLEY LABORATORY | Diseases of the National Health Laboratory Service | Amoako DG, Everatt J, Scheepers C, Mohale T, Ntuli N, Mahlangu B, Mnguni A, Ismail A, Bhiman JN |
| hCoV-19/SouthAfrica/NICD-N14534/2021 | EPI_ISL_4650299 | 2021-07-23 | AMPATH | Diseases of the National Health Laboratory Service | Amoako DG, Everatt J, Scheepers C, Mohale T, Ntuli N, Mahlangu B, Mnguni A, Ismail A, Bhiman JN |
| hCoV-19/Japan/PG-102471/2021 | EPI_ISL_4651374 | 2021-08-06 | Nagasaki Prefectural Institute of Environment and Public Health | Nagasaki Prefectural Institute of Environment and Public Health | Fumiaki Matsumoto, Yumika Takaki, Fumika Nakamine, Motoki Ihara, Yuina Yamaguchi, Cai Guoxi, Yuji Migita, Akira Yoshikawa, Tsuyoshi Sekizuka, Kentaro Itokawa, Rina Tanaka, Masanori Hashino, Makoto Kuroda |
| hCoV-19/SouthAfrica/NICD-R13214/2021 | EPI_ISL_4651565 | 2021-08-30 | National Institute for Communicable Diseases of the National Health Laboratory Service | Diseases of the National Health Laboratory Service | Amoako DG, Everatt J, Scheepers C, Mohale T, Ntuli N, Mahlangu B, Mnguni A, Ismail A, Bhiman JN |
| hCoV-19/SouthAfrica/NICD-N15676/2021 | EPI_ISL_4651733 | 2021-08-24 | HELEN JOSEPH LABORATORY | Diseases of the National Health Laboratory Service | Amoako DG, Everatt J, Scheepers C, Mohale T, Ntuli N, Mahlangu B, Mnguni A, Ismail A, Bhiman JN |
| hCoV-19/SouthAfrica/NICD-N14378/2021 | EPI_ISL_4651786 | 2021-07-12 | AMPATH | Diseases of the National Health Laboratory Service | Amoako DG, Everatt J, Scheepers C, Mohale T, Ntuli N, Mahlangu B, Mnguni A, Ismail A, Bhiman JN |
| hCoV-19/SouthAfrica/NICD-R13310/2021 | EPI_ISL_4652437 | 2021-09-03 | National Institute for Communicable Diseases of the National Health Laboratory Service | Diseases of the National Health Laboratory Service | Amoako DG, Everatt J, Scheepers C, Mohale T, Ntuli N, Mahlangu B, Mnguni A, Ismail A, Bhiman JN |
| hCoV-19/Japan/PG-94145/2021 | EPI_ISL_4668611 | 2021-08-15 | SARS-CoV-2 testing team, National Institute of Infectious Diseases | Pathogen Genomics Center, National Institute of Infectious Diseases | Tsuyoshi Sekizuka, Kentaro Itokawa, Rina Tanaka, Masanori Hashino, Nozomu Hanaoka, Masumichi Saito, Naomi Nojiri, Hazuka Y Furihata, Tsuguto Fujimoto, Makoto Kuroda |
| hCoV-19/Japan/PG-108296/2021 | EPI_ISL_4709201 | 2021-07-26 | Pathogen Genomics Center, National Institute of Infectious Diseases | Pathogen Genomics Center, National Institute of Infectious Diseases | Tsuyoshi Sekizuka, Kentaro Itokawa, Rina Tanaka, Masanori Hashino, Makoto Kuroda |
| hCoV-19/Japan/PG-108870/2021 | EPI_ISL_4709945 | 2021-07-29 | Pathogen Genomics Center, National Institute of Infectious Diseases | Pathogen Genomics Center, National Institute of Infectious Diseases | Tsuyoshi Sekizuka, Kentaro Itokawa, Rina Tanaka, Masanori Hashino, Makoto Kuroda |
| hCoV-19/Japan/PG-113268/2021 | EPI_ISL_4720164 | 2021-08-03 | Pathogen Genomics Center, National Institute of Infectious Diseases | Pathogen Genomics Center, National Institute of Infectious Diseases | Tsuyoshi Sekizuka, Kentaro Itokawa, Rina Tanaka, Masanori Hashino, Makoto Kuroda |
| hCoV-19/India/AS-INSACOG-1829500321843/2021 | EPI_ISL_4739017 | 2021-07-09 | INSACOG-Assam | National Institute of Biomedical Genomics ,Äì¬†INSACOG | Arindam Maitra, Ajanta Sharma, Nidhan Kumar Biswas, Lahari Saikia, Sreedhar Chinnaswamy,¬†Kailash Chamuah, Saumitra Das |
| hCoV-19/India/AS-INSACOG-1829500335412/2021 | EPI_ISL_4739078 | 2021-07-16 | INSACOG-Assam | National Institute of Biomedical Genomics ,Äì¬†INSACOG | Arindam Maitra, Ajanta Sharma, Nidhan Kumar Biswas, Lahari Saikia, Sreedhar Chinnaswamy,¬†Kailash Chamuah, Saumitra Das |

| | | | | | |
|---|---|---|---|---|---|
| hCoV-19/India/WB-INSACOG-1931300457537/2021 | EPI_ISL_4739179 | 2021-09-06 | INSACOG-WB | National Institute of Biomedical Genomics - INSACOG | Arindam Maitra, Bhaswati Bandyopadhyay, Nidhan Kumar Biswas, Tamal Ghosh, Sreedhar Chinnaswamy, Ajay Chakraborti, Saumitra Das |
| hCoV-19/Italy/APU-IZSPB-1783PT/2021 | EPI_ISL_3485113 | 2021-07-16 | Presidio di Brindisi Di Summa - Perrino | Istituto Zooprofilattico Sperimentale della Puglia e della Basilicata | Parisi A., Bianco A., Capozzi L., Del Sambro L., Simone D., Difato L., Santoro A., Ridolfi D., Giannico A. |
| hCoV-19/SouthAfrica/NICD-N9291/2021 | EPI_ISL_2827155 | 2021-05-19 | TAMBO MEMORIAL LABORATORY | Diseases of the National Health Laboratory Service | Amoako DG, Everatt J, Scheepers C, Mohale T, Ntuli N, Mahlangu B, Mnguni A, Ismail A, Bhiman JN |
| hCoV-19/USA/IA-SHL-1780926/2021 | EPI_ISL_3939757 | 2021-02-08 | State Hygienic Laboratory at the University of Iowa | State Hygienic Laboratory at the University of Iowa | Valerie Reeb, Erik Twait, Anna Yakos, Mariah Knutson, Kris Eveland, Kristen Zanon, Jeff Benfer, Davis Rieckenberg, Cindy Toll, Wes Hottel, Alankar Kampoowale, Mohammed Allam |
| hCoV-19/Reunion/HCL0211532821/2021 | EPI_ISL_4282523 | 2021-08-17 | CNR Virus des Infections Respiratoires - France SUD | CNR Virus des Infections Respiratoires - France SUD | Antonin Bal, Gregory Destras, Gwendolyne Burfin, Hadrien Regue, Quentin Semanas, Martine Valette, Bruno Lina, Laurence Josset |
| hCoV-19/India/WB-INSACOG-1931300447032/2021 | EPI_ISL_4312283 | 2021-08-25 | INSACOG-WB | National Institute of Biomedical Genomics ‚Äì INSACOG | Arindam Maitra, Bhaswati Bandyopadhyay, Nidhan Kumar Biswas, Tamal Ghosh, Sreedhar Chinnaswamy, Ajay Chakraborti, Saumitra Das |
| hCoV-19/USA/TX-CDC-QDX26764086/2021 | EPI_ISL_3213789 | 2021-07-11 | Quest Diagnostics Incorporated | Centers for Disease Control and Prevention Division of Viral Diseases, Pathogen Discovery | Dakota Howard, Dhwani Batra, Peter W. Cook, Kara Moser, Adrian Paskey, Jason Caravas, Benjamin Rambo-Martin, Shatavia Morrison, Christopher Gulvick, Scott Sammons, Yvette Unoarumhi, Darlene Wagner, Matthew Schmerer, S. H. Rosenthal, A. Gerasimova, R. M. K |
| hCoV-19/Slovakia/UVZ_PL36_B7_17907/2021 | EPI_ISL_4483942 | 2021-02-09 | Public Health Authority of the Slovak Republic | Laboratory of Genomics and Bioinformatics, Comenius University Science Park | Tatiana Sedl√°ƒçkov√°, Diana Rus≈à√°kov√°, Miroslav B√∂hmer, Anna Gifçov√°, Jaroslav Budi≈°, Tom√°≈° Szemes, Jakub Styk |
| hCoV-19/Slovakia/UVZ_PL36_C7_17910/2021 | EPI_ISL_4483944 | 2021-02-09 | Public Health Authority of the Slovak Republic | Laboratory of Genomics and Bioinformatics, Comenius University Science Park | Tatiana Sedl√°ƒçkov√°, Diana Rus≈à√°kov√°, Miroslav B√∂hmer, Anna Gifçov√°, Jaroslav Budi≈°, Tom√°≈° Szemes, Jakub Styk |
| hCoV-19/Slovakia/UVZ_PL36_E2_17784/2021 | EPI_ISL_4483999 | 2021-01-09 | Public Health Authority of the Slovak Republic | Laboratory of Genomics and Bioinformatics, Comenius University Science Park | Tatiana Sedl√°ƒçkov√°, Diana Rus≈à√°kov√°, Miroslav B√∂hmer, Anna Gifçov√°, Jaroslav Budi≈°, Tom√°≈° Szemes, Jakub Styk |
| hCoV-19/Slovakia/UVZ_PL37_H5_18044/2021 | EPI_ISL_4484070 | 2021-03-09 | Public Health Authority of the Slovak Republic | Laboratory of Genomics and Bioinformatics, Comenius University Science Park | Tatiana Sedl√°ƒçkov√°, Diana Rus≈à√°kov√°, Miroslav B√∂hmer, Anna Gifçov√°, Jaroslav Budi≈°, Tom√°≈° Szemes, Jakub Styk |
| hCoV-19/Slovakia/UVZ_PL36_H11_17990/2021 | EPI_ISL_4484079 | 2021-03-09 | Public Health Authority of the Slovak Republic | Laboratory of Genomics and Bioinformatics, Comenius University Science Park | Tatiana Sedl√°ƒçkov√°, Diana Rus≈à√°kov√°, Miroslav B√∂hmer, Anna Gifçov√°, Jaroslav Budi≈°, Tom√°≈° Szemes, Jakub Styk |
| hCoV-19/India/MZ-INSACOG-1526500005999/2020 | EPI_ISL_4312739 | 2020-10-31 | INSACOG-Mizoram | National Institute of Biomedical Genomics ‚Äì INSACOG | Arindam Maitra, Swagnik Roy, Nidhan Kumar Biswas, Gracy Laldinmawii, Sreedhar Chinnaswamy, N Senthil Kumar, Saumitra Das |
| hCoV-19/France/IDF-HMN-21072190148/2021 | EPI_ISL_3391029 | 2021-07-13 | Biogroup Bio Lam-LCD Saint-Denis | Mondor University Hospital, Assistance Publique H√¥pitaux de Paris, Universit√© Paris-Est Cr√©teil, INSERM U955 | Christophe Rodriguez, Slim Fourati, Vanessa Demontant, Guillaume Gricourt, Melissa N'Debi, Alexandre Soulier, Elisabeth Trawinski, Jean-Michel Pawlotsky |

| | | | | | |
|---|---|---|---|---|---|
| hCoV-19/India/SK-INSACOG-1122500083560/2021 | EPI_ISL_4313081 | 2021-07-23 | INSACOG-Sikkim | National Institute of Biomedical Genomics ‚Äì†INSACOG | Arindam Maitra, Shrijana Gurung, Nidhan Kumar Biswas, Kaden Zangmu Bhutia, Sreedhar Chinnaswamy Tshering Ongmu Bhutia, Saumitra Das |
| hCoV-19/England/NEWC-1CD86B5/2021 | EPI_ISL_3994826 | 2021-08-27 | Integrated Covid Hub North East | Wellcome Sanger Institute for the COVID-19 Genomics UK (COG-UK) Consortium | Integrated Covid Hub North East and Alex Alderton, Roberto Amato, Jeffrey Barrett, Sonia Goncalves, Ewan Harrison, David K. Jackson, Ian Johnston, Dominic Kwiatkowski, Cordelia Langford, John Sillitoe on behalf of the Wellcome Sanger Institute COVID-19 Su |
| hCoV-19/Italy/FVG-UD-404586/2021 | EPI_ISL_4533460 | 2021-09-14 | Dipartimento di Medicina di Laboratorio, Azienda sanitaria universitaria Friuli Centrale (ASU FC) | Dipartimento di Medicina di Laboratorio, Azienda sanitaria universitaria Friuli Centrale (ASU FC) | Corrado Pipan, Catia Mio, Chiara Dal Secco, Federica Baldan, Stefania Marzinotto, Francesco Curcio |
| hCoV-19/Japan/PG-87015/2021 | EPI_ISL_3898730 | 2021-06-14 | Pathogen Genomics Center, National Institute of Infectious Diseases | Pathogen Genomics Center, National Institute of Infectious Diseases | Tsuyoshi Sekizuka, Kentaro Itokawa, Rina Tanaka, Masanori Hashino, Makoto Kuroda |
| hCoV-19/SouthAfrica/NICD-N9405/2021 | EPI_ISL_2984953 | 2021-05-28 | Pathcare Vaal Laboratory | Diseases of the National Health Laboratory Service | Amoako DG, Everatt J, Scheepers C, Mohale T, Ntuli N, Mahlangu B, Mnguni A, Ismail A, Bhiman JN |
| hCoV-19/Senegal/SN-IRFSFOU-34_bis/2021 | EPI_ISL_3154894 | 2021-04-05 | Iressef Genomics lab | IRESSEF | Souleymane MBOUP, Abdou PADANE , Khadim GUEYE, Diabou Diagne , Samba Ndiour, Papa Alassane DIAW, Birahim Piere NDIAYE, Barada CISSE, Aminata MBOUP, Moustapha MBOW, Ndeye Coumba Toure KANE, Nafisatou LEYE, Gora LO, Ambroise AHOUIDI , Astou Gaye GAYE, Amina |
| hCoV-19/Austria/CeMM12509/2021 | EPI_ISL_2985528 | 2021-06-25 | Austrian Agency for Health and Food Safety (AGES) | Bergthaler laboratory, CeMM Research Center for Molecular Medicine of the Austrian Academy of Sciences | Lukas Endler, Anna Schedl, Fabian Amman, Petr Triska, Matthew Thornton, Thomas Penz, Benedikt Agerer, Maelle Le Moing, Michael Schuster, Bekir Erguner, Jan Laine, Martin Senekowitsch, Christoph Bock, Andreas Bergthaler |
| hCoV-19/NewZealand/21MV1455/2021 | EPI_ISL_4063315 | 2021-09-07 | LabPLUS | Institute of Environmental Science and Research (ESR) | Rachel Boyle, SallyAnn Harbison, Olivia Stroeven, Xiaoyun Ren, Matt Storey, Nikki Freed, Muhammad Faisal, Jing Wang, Hermes Perez, Anja Werno, Antje van der Linden, Arlo Upton, Chris Mansell, David Hammer, Dragana Drinkovic, Gary McAuliffe, Hana Sofia And |
| hCoV-19/NewZealand/21MV1463/2021 | EPI_ISL_4063312 | 2021-09-07 | LabPLUS | Institute of Environmental Science and Research (ESR) | Rachel Boyle, SallyAnn Harbison, Olivia Stroeven, Xiaoyun Ren, Matt Storey, Nikki Freed, Muhammad Faisal, Jing Wang, Hermes Perez, Anja Werno, Antje van der Linden, Arlo Upton, Chris Mansell, David Hammer, Dragana Drinkovic, Gary McAuliffe, Hana Sofia And |
| hCoV-19/Turkey/HSGM-B17724/2021 | EPI_ISL_2987950 | 2021-06-21 | Ministry of Health Turkey | Ministry of Health Turkey | Fatma Bayrakdar, Yasemin Cosgun, Suleyman Yalcin, Gulay Korukluoglu |
| hCoV-19/Turkey/HSGM-B17730/2021 | EPI_ISL_2987954 | 2021-06-21 | Ministry of Health Turkey | Ministry of Health Turkey | Fatma Bayrakdar, Yasemin Cosgun, Suleyman Yalcin, Gulay Korukluoglu |
| hCoV-19/Turkey/HSGM-B17869/2021 | EPI_ISL_2987992 | 2021-06-22 | Ministry of Health Turkey | Ministry of Health Turkey | Fatma Bayrakdar, Yasemin Cosgun, Suleyman Yalcin, Gulay Korukluoglu |
| hCoV-19/USA/NV-NSPHL-390974/2021 | EPI_ISL_4491924 | 2021-08-16 | Nevada State Public Health Laboratory | Nevada State Public Health Laboratory | Andrew Gorzalski, Mark Pandori |

| | | | | | |
|---|---|---|---|---|---|
| hCoV-19/Peru/PIU-INS-2380/2021 | EPI_ISL_3401563 | 2021-02-03 | Laboratorio de Referencia Nacional de Virus Respiratorios. Centro Nacional de Salud Publica. Instituto Nacional de Salud Peru. | Laboratorio de Referencia Nacional de Virus Respiratorios. Centro Nacional de Salud Publica. Instituto Nacional de Salud Peru. | Carlos Padilla Rojas, Veronica Hurtado Vela, Orson Mestanza Millones, Iris Silva Molina, Luis Barcena Flores, Sandra Morales Ruiz, Steve Acedo Lazo, Wendy Lizarraga Olivares, Victor Jimenez Vasquez, Sara Gordillo Vilchez, Nieves Sevilla Casta√±eda, Prince |
| hCoV-19/Austria/CeMM14749/2021 | EPI_ISL_3797378 | 2021-07-31 | Austrian Agency for Health and Food Safety (AGES) | Bergthaler laboratory, CeMM Research Center for Molecular Medicine of the Austrian Academy of Sciences | Lukas Endler, Anna Schedl, Fabian Amman, Petr Triska, Matthew Thornton, Thomas Penz, Benedikt Agerer, Maelle Le Moing, Michael Schuster, Bekir Erguner, Jan Laine, Martin Senekowitsch, Christoph Bock, Andreas Bergthaler |
| hCoV-19/India/MP-NCDC-4309114/2021 | EPI_ISL_4493055 | 2021-06-02 | National Centre for Disease Control (NCDC) Biotechnology Division, Delhi - INSACOG | NCDC Delhi, Biotechnology Division - INSACOG | Kalaiarasan Ponnusamy, Meena Datta, Priyanka Singh,¬†Uma Sharma,¬†Hemlata Lall, Manoj K Singh, Radhakrishnan V. S, Robin Marwal, Mahesh S Dhar, Hema Gogia, Preeti Madan, Sandhya Kabra, Sujeet K Singh, Partha Rakshit |
| hCoV-19/England/NORT-YYNW7P/2020 | EPI_ISL_4306808 | 2020-10-27 | Northumbria University / South Tees Hospitals NHS Foundation Trust / North Cumbria Integrated Care NHS Foundation Trust / North Tees and Hartlepool NHS Foundation Trust / Newcastle Hospitals NHS Foundation Trust | COVID-19 Genomics UK (COG-UK) Consortium | Darren L Smith,Andrew Nelson,Matthew Bashton,Greg R Young,Joshua Loh,John Allan,Mohammad A Tariq,Giles S Holt,Gary Black,Wen C Yew,Lynn Dover,Paul Baker,Steve Liggett,Sarah Essex,Jane Greenaway,Debra Padgett,Clive Graham,Garren Scott,Edward Barton,Emma Sw |
| hCoV-19/India/TR-AGMC/COV/R25467/2021 | EPI_ISL_2877948 | 2021-04-26 | INSACOG-Tripura | National Institute of Biomedical Genomics ,Äì INSACOG | Arindam Maitra, Tapan Majumdar, Nidhan Kumar Biswas, Saikat Majumder, Sreedhar Chinnaswamy, Apurba Sarkar, Saumitra Das |
| hCoV-19/England/PHEP-005698/2021 | EPI_ISL_3779611 | 2021-07-09 | Respiratory Virus Unit, Microbiology Services Colindale, Public Health England | COVID-19 Genomics UK (COG-UK) Consortium | PHE Covid Sequencing Team |
| hCoV-19/USA/CO-CDPHE-2101334738/2021 | EPI_ISL_3404163 | 2021-05-26 | Colorado Department of Public Health and Environment | Colorado Department of Public Health and Environment | Laura Bankers, Molly C. Hetherington-Rauth, Diana Ir, Alexandria Rossheim, Michael Martin, Mandy Waters, Shannon R. Matzinger, Sarah Elizabeth Totten, Emily A. Travanty |
| hCoV-19/India/HR-IGIB-60252/2021 | EPI_ISL_2878577 | 2021-02-01 | INSACOG Surveillance | INSACOG at CSIR Institute of Genomics and Integrative Biology | INSACOG |
| hCoV-19/India/MH-IGIB-GSEQ_1945/2021 | EPI_ISL_2878988 | 2021-03-22 | INSACOG Surveillance | INSACOG at CSIR Institute of Genomics and Integrative Biology | INSACOG |
| hCoV-19/France/NAQ-Poitiers-2107_122333/2021 | EPI_ISL_3230774 | 2021-06-18 | FORTEBIO - UNILABS | CHU Poitiers | Caroline MICHAUD, Maxime PICHON, Manon PRAT, Valentin BON-BARET, Birama N'DIAYE, Magali GARCIA, Agnes BEBY-DEFAUX, Nicolas LEVEQUE |
| hCoV-19/India/MH-IGIB-GSEQ_2148/2021 | EPI_ISL_2879078 | 2021-03-26 | INSACOG Surveillance | INSACOG at CSIR Institute of Genomics and Integrative Biology | INSACOG |
| hCoV-19/Brazil/PE-FIOCRUZ-IAM2235/2020 | EPI_ISL_3046240 | 2020-09-09 | NUPIT/UFPE | WallauLab on behalf of Fiocruz COVID-19 Genomic Surveillance Network | Marcelo Henrique dos Santos Paiva, Cassia Docena, Matheus Filgueira Bezerra, Filipe Zimmer Dezordi, Lais Ceschini Machado, Lilian Caroliny Amorim Silva, Gustavo Barbosa de Lima, Alexandre Freitas da Silva, Sinval Pinto Brand√£o Filho, Const√¢ncia Fl√°via |
| hCoV-19/India/MH-IGIB-GSEQ_2737/2021 | EPI_ISL_2879537 | 2021-04-13 | INSACOG Surveillance | INSACOG at CSIR Institute of Genomics and Integrative Biology | INSACOG |

| | | | | | |
|---|---|---|---|---|---|
| hCoV-19/India/HR-IGIB-123500/2021 | EPI_ISL_2879594 | 2021-04-14 | INSACOG Surveillance | INSACOG at CSIR Institute of Genomics and Integrative Biology | INSACOG |
| hCoV-19/England/PHEC-U303U143/2021 | EPI_ISL_3776152 | 2021-08-17 | Respiratory Virus Unit, Microbiology Services Colindale, Public Health England | COVID-19 Genomics UK (COG-UK) Consortium | PHE Covid Sequencing Team |
| hCoV-19/Turkey/HSGM-FS2222/2021 | EPI_ISL_3405671 | 2021-08-06 | Ministry of Health Turkey | Ministry of Health Turkey | Fatma Bayrakdar, Yasemin Cosgun, Suleyman Yalcin, Gulay Korukluoglu |
| hCoV-19/Australia/NSW-1810/2021 | EPI_ISL_2828016 | 2021-06-28 | South Eastern Area Laboratory Services (SEALS) | Clinical Pathology and Medical Research; Westmead Hospital; University of Sydney | CIDM-PH et al. |
| hCoV-19/Chile/BI-142081/2021 | EPI_ISL_4631967 | 2021-08-20 | Genetica Molecular and Subdepartamento de Virologia ISP Chile | Instituto de Salud Publica de Chile | Karen Orostica, Constanza Campano, Barbara Parra, Loredana Arata, Gisselle Barra, Patricia Bustos, Rodrigo Fasce, Javier Tognarelli, Andres Castillo, Soledad Ulloa, Jorge Fernandez |
| hCoV-19/USA/VA-CDC-LC0074991/2021 | EPI_ISL_4374515 | 2021-06-09 | Laboratory Corporation of America | Centers for Disease Control and Prevention Division of Viral Diseases, Pathogen Discovery | Dakota Howard,Dhwani Batra,Peter Cook,Jason Caravas,Benjamin Rambo-Martin,Scott Sammons,Yvette Unoarumhi,Matthew Schmerer,Kristine Lacek,Tymeckia Kendall,Victoria Caban Figueroa,Shatavia Morrison,Christopher Gulvick,Minoo Agarwal,Eyad Almasri,Debbie Boles |
| hCoV-19/Chile/BI-142578/2021 | EPI_ISL_4632406 | 2021-08-23 | Genetica Molecular and Subdepartamento de Virologia ISP Chile | Instituto de Salud Publica de Chile | Karen Orostica, Constanza Campano, Barbara Parra, Loredana Arata, Gisselle Barra, Patricia Bustos, Rodrigo Fasce, Javier Tognarelli, Andres Castillo, Soledad Ulloa, Jorge Fernandez |
| hCoV-19/Australia/NSW-RPAH-1389/2021 | EPI_ISL_4254948 | 2021-08-28 | New South Wales Health Pathology Royal Prince Alfred Hospital | Microbiology RPAH | Foster, C.; Au, J.; Ruiz Silva, M.; Deveson, I.; Bull, R.; Van Hal, S.; Rawlinson, W. |
| hCoV-19/Chile/RM-147027/2021 | EPI_ISL_4632489 | 2021-09-01 | Genetica Molecular and Subdepartamento de Virologia ISP Chile | Instituto de Salud Publica de Chile | Karen Orostica, Constanza Campano, Barbara Parra, Loredana Arata, Gisselle Barra, Patricia Bustos, Rodrigo Fasce, Javier Tognarelli, Andres Castillo, Soledad Ulloa, Jorge Fernandez |
| hCoV-19/Bahrain/342422028/2021 | EPI_ISL_3914639 | 2021-07-31 | Communicable Disease Laboratory, Public Health Directorate | Communicable Disease Laboratory, Public Health Directorate | Alwasti,H., Altaif,Z., AlHujairi,Z., AlAbbas,Z., Marhoon,A., Touq,M. |
| hCoV-19/Turkey/HSGM-MK288/2021 | EPI_ISL_3540910 | 2021-08-13 | Ministry of Health Turkey | Ministry of Health Turkey | Fatma Bayrakdar, Yasemin Co≈ügun, S√ºleyman Yalcin, G√ºlay Korukluoƒülu |
| hCoV-19/Togo/NMIMR-21-TGS-267/2021 | EPI_ISL_3914992 | 2021-06-01 | Institut National D‚Äôhygiene, Lome, Togo | Noguchi Memorial Institute for Medical Research, University of Ghana, Legon, Ghana | Afiwa W. Halatoko, Issaka Maman, Yawo A. Sadji, Komlan Kossi, Quaneeta Mohktar, Hilda Opoku Frempong, Peace O. Uche, Keren O. Attiku, Djimabi Salah, Yao Layibo, Lallepak Lamboni, Seyram B. Agbenyo, Hamadi Assane, Joyce Appiah-Kubi, Zoukaneirii Issa, Sena |
| hCoV-19/India/MP-NCDC-360961/2021 | EPI_ISL_4460051 | 2021-05-17 | National Centre for Disease Control (NCDC) Biotechnology Division, Delhi - INSACOG | NCDC Delhi, Biotechnology Division - INSACOG | Robin Marwal, Mahesh S Dhar, Kalaiarasan Ponnusamy, Meena Datta, Priyanka Singh,¬†Uma Sharma,¬†Hemlata Lall, Manoj K Singh, Radhakrishnan V. S, Hema Gogia, Preeti Madan, Sandhya Kabra, Sujeet K Singh, Partha Rakshit |

| | | | | | |
|---|---|---|---|---|---|
| hCoV-19/USA/GA-CDC-QDX26400231/2021 | EPI_ISL_4369949 | 2021-06-30 | Quest Diagnostics Incorporated | Centers for Disease Control and Prevention Division of Viral Diseases, Pathogen Discovery | Dakota Howard,Dhwani Batra,Peter Cook,Jason Caravas,Benjamin Rambo-Martin,Scott Sammons,Yvette Unoarumhi,Matthew Schmerer,Kristine Lacek,Tymeckia Kendall,Victoria Caban Figueroa,Shatavia Morrison,Christopher Gulvick,Erisa Sula,S. Rosenthal,A. Gerasimova,R |
| hCoV-19/Sweden/01_SE100_21CS507296/2021 | EPI_ISL_3534402 | 2021-08-09 | Karolinska University Hospital Solna | Karolinska University Hospital | Jan Albert ,Tobias Allander ,Annelie Bjerkner ,Sandra Broddesson ,Robert Dyrdak ,Martin Ekman ,Lynda Eneh ,Lina Guerra Blomqvist ,Karolina Ininbergs ,Tanja Normark ,Isak Sylvin ,Zhibing Yun ,Martina Wahlund ,Valtteri Wirta |
| hCoV-19/India/UT-NCDC-4509193/2021 | EPI_ISL_4460860 | 2021-06-16 | National Centre for Disease Control (NCDC) Biotechnology Division, Delhi - INSACOG | NCDC Delhi, Biotechnology Division - INSACOG | Kalaiarasan Ponnusamy, Meena Datta, Priyanka Singh,¬†Uma Sharma,¬†Hemlata Lall, Manoj K Singh, Radhakrishnan V. S, Robin Marwal, Mahesh S Dhar, Hema Gogia, Preeti Madan, Sandhya Kabra, Sujeet K Singh, Partha Rakshit |
| hCoV-19/Brazil/SP-IB_130838/2021 | EPI_ISL_4275478 | 2021-08-30 | UNIDADE REFERENCIAL SUDOESTE | Instituto Butantan | Dimas Tadeu Covas, Antonio Jorge Martins, Claudia Renata dos Santos Barros, David Schlesinger, Debora Botequio Moretti, Elaine Cristina Marqueze, Elaine Vieira Santos, Evandra Strazza Rodrigues, Heidge Fukumasu, Jayme Augusto de Souza-Neto, Jos√© Salvator |
| hCoV-19/Brazil/PR-FIOCRUZ-HF462/2021 | EPI_ISL_3914313 | 2021-07-08 | Laboratorio Central de Saude Publica do Estado do Parana (Instituto de Biologia Molecular do Paran√° (LACEN-PR) | Instituto Carlos Chagas - Fiocruz | Mauro de Medeiros Oliveira, Michelle Orane Schemberger, Andreia Akemi Suzukawa, Irina Nastassja Riediger, Maria do Carmo Debur, Guilherme Becker, Paola Cristina Resende, Tiago Gr√§f, Eduardo Balsanelli, Valter Ant√¥nio de Baura, Emanuel Maltempi de Souza, |
| hCoV-19/India/DL-NCDC-4109305/2021 | EPI_ISL_4462327 | 2021-05-06 | National Centre for Disease Control (NCDC) Biotechnology Division, Delhi - INSACOG | NCDC Delhi, Biotechnology Division - INSACOG | Mahesh S Dhar, Kalaiarasan Ponnusamy, Meena Datta, Priyanka Singh,¬†Uma Sharma,¬†Hemlata Lall, Manoj K Singh, Radhakrishnan V. S, Robin Marwal, Hema Gogia, Preeti Madan, Sandhya Kabra, Sujeet K Singh, Partha Rakshit |
| hCoV-19/India/CH-NCDC-440984/2021 | EPI_ISL_4463364 | 2021-06-04 | National Centre for Disease Control (NCDC) Biotechnology Division, Delhi - INSACOG | NCDC Delhi, Biotechnology Division - INSACOG | Kalaiarasan Ponnusamy, Meena Datta, Priyanka Singh,¬†Uma Sharma,¬†Hemlata Lall, Manoj K Singh, Radhakrishnan V. S, Robin Marwal, Mahesh S Dhar, Hema Gogia, Preeti Madan, Sandhya Kabra, Sujeet K Singh, Partha Rakshit |
| hCoV-19/India/MP-NCDC-3209254/2021 | EPI_ISL_4463482 | 2021-05-10 | National Centre for Disease Control (NCDC) Biotechnology Division, Delhi - INSACOG | NCDC Delhi, Biotechnology Division - INSACOG | Radhakrishnan V. S, Robin Marwal, Mahesh S Dhar, Kalaiarasan Ponnusamy, Meena Datta, Priyanka Singh,¬†Uma Sharma,¬†Hemlata Lall, Manoj K Singh, Hema Gogia, Preeti Madan, Sandhya Kabra, Sujeet K Singh, Partha Rakshit |
| hCoV-19/Turkey/HSGM-E3104/2021 | EPI_ISL_4101759 | 2021-03-09 | Ministry of Health Turkey | Ministry of Health Turkey | Fatma Bayrakdar, Yasemin Cosgun, Suleyman Yalcin, Gulay Korukluoglu |
| hCoV-19/Turkey/HSGM-E3160/2021 | EPI_ISL_4101806 | 2021-03-09 | Ministry of Health Turkey | Ministry of Health Turkey | Fatma Bayrakdar, Yasemin Cosgun, Suleyman Yalcin, Gulay Korukluoglu |

| | | | | | |
|---|---|---|---|---|---|
| hCoV-19/Guam/GU-CDC-2-4611255/2021 | EPI_ISL_3355200 | 2021-06-01 | Guam Public Health Laboratory | Centers for Disease Control and Prevention Division of Viral Diseases, Pathogen Discovery | Mili Sheth,Sarah Nobles,Jasmine Padilla,Alex Burgin,Meghan Bentz,Mark Burroughs,Peter Cook,Clinton Paden,Dhwani Batra,Krista Queen,Kristen Knipe,Dakota Howard,Yvette Unoarumhi,Matthew Schmerer,Ben Rambo-Martin,Kristine Lacek,Sam Shepard,Dave Wentworth,Viv |
| hCoV-19/Chile/RM-152742/2021 | EPI_ISL_4658550 | 2021-09-12 | Genetica Molecular and Subdepartamento de Virologia ISP Chile | Instituto de Salud Publica de Chile | Karen Orostica, Constanza Campano, Barbara Parra, Loredana Arata, Gisselle Barra, Patricia Bustos, Rodrigo Fasce, Javier Tognarelli, Andres Castillo, Soledad Ulloa, Jorge Fernandez |
| hCoV-19/NorthernMarianaIslands/MP-CDC-2-3830939-/2020 | EPI_ISL_3353665 | 2020-12-21 | Commonwealth Healthcare Center | Centers for Disease Control and Prevention Division of Viral Diseases, Pathogen Discovery | Mili Sheth,Sarah Nobles,Jasmine Padilla,Alex Burgin,Meghan Bentz,Mark Burroughs,Peter Cook,Clinton Paden,Dhwani Batra,Krista Queen,Kristen Knipe,Dakota Howard,Yvette Unoarumhi,Matthew Schmerer,Ben Rambo-Martin,Kristine Lacek,Sam Shepard,Dave Wentworth,Viv |
| hCoV-19/NorthernMarianaIslands/MP-CDC-2-3831042-/2020 | EPI_ISL_3353695 | 2020-07-14 | Commonwealth Healthcare Center | Centers for Disease Control and Prevention Division of Viral Diseases, Pathogen Discovery | Mili Sheth,Sarah Nobles,Jasmine Padilla,Alex Burgin,Meghan Bentz,Mark Burroughs,Peter Cook,Clinton Paden,Dhwani Batra,Krista Queen,Kristen Knipe,Dakota Howard,Yvette Unoarumhi,Matthew Schmerer,Ben Rambo-Martin,Kristine Lacek,Sam Shepard,Dave Wentworth,Viv |
| hCoV-19/Bangladesh/IEDCR-ideSHi-GSB13-047/2021 | EPI_ISL_4467320 | 2021-08-23 | Mymensingh Medical College, Mymensingh | IEDCR-ideSHi Genomics Lab | Hamza O, Rahman SIA, Ahmed S, Hossain M, Khan MH, Saha S, Afrad MH, Rahman MZ, Jubair M, Hossain ME, Mazumder R, Islam MS, Alam AN, Alamgir ASM, Rahman M, Rahman M, Mondal D, Saha SK, Shirin T, Qadri F |
| hCoV-19/USA/FL-BPHL-4906/2021 | EPI_ISL_2836731 | 2021-06-09 | Florida Bureau of Public Health Laboratories | Florida Bureau of Public Health Laboratories | Sarah Schmedes, Jason Blanton |
| hCoV-19/Belize/CML-92/2021 | EPI_ISL_4296396 | 2021-02-07 | Central Medical Laboratory | Baylor College of Medicine | Alexander Kneubehl, Sarah Gunter, Shannon Ronca, Allison Lino, Adrianna Maliga, Sarah Strobel, Russell Manzanero, Gerhaldine Morazan, Kristy Murray |
| hCoV-19/Belize/CML-101/2021 | EPI_ISL_4296405 | 2021-01-07 | Central Medical Laboratory | Baylor College of Medicine | Alexander Kneubehl, Sarah Gunter, Shannon Ronca, Allison Lino, Adrianna Maliga, Sarah Strobel, Russell Manzanero, Gerhaldine Morazan, Kristy Murray |
| hCoV-19/Madagascar/LA2M-62987/2021 | EPI_ISL_4297556 | 2021-03-15 | Virology Unit, Institut Pasteur de Madagascar | Virology Unit, Institut Pasteur de Madagascar | Marion Barbet, Sylvie Behillil, Fr√©d√©ric Lemoine, Corinne Maufrais, Christophe Malabat, Emmanuelle Permal, M√©line Bizard, Angela Brisebarre, Camille Capel, Louise Lefran√ßois, Etienne Simon-Lori√®re, Vincent Enouf, Maud Vanpeene, Christian Ranaivoson, |
| hCoV-19/Guinea/IP-GNC_21228/2021 | EPI_ISL_3988377 | 2021-05-11 | Institut Pasteur de Guin√©e | Institut Pateur de Dakar | Grayo Solene, Diagne Moussa Mo√Øse, Diallo Amadou, Dia Ndongo, Mbengue Safietou Sankhe, Ndiaye Ndack, Diop Mamadou, Maimouna Mbanne, Mohamed Kane, Loucoubar Cheikh, Tordo Noel, Faye Ousmane, Sall Amadou Alpha |

| | | | | | |
|---|---|---|---|---|---|
| hCoV-19/India/OR-ILSGS06333/2021 | EPI_ISL_4102988 | 2021-06-14 | Srirama Chandra Bhanja Medical College and Hospital, Cuttack | Institute of Life Sciences-INSACOG | Sunil K. Raghav, Safal Walia, Arup Ghosh, Atimukta Jha, Amol M. Kanampalliwar, Sana Fatma, Shifu Aggarwal, Rupesh Dash, Rajeeb Swain, Punit Prasad, INSACOG Consortium, Ajay Parida |
| hCoV-19/Brazil/SP-IB_130648/2021 | EPI_ISL_4470051 | 2021-08-30 | INSIDE DIAGN√ìSTICOS | Instituto Butantan | Dimas Tadeu Covas, Antonio Jorge Martins, Claudia Renata dos Santos Barros, David Schlesinger, Debora Botequio Moretti, Elaine Cristina Marqueze, Elaine Vieira Santos, Evandra Strazza Rodrigues, Heidge Fukumasu, Jayme Augusto de Souza-Neto, Jos√© Salvator |
| hCoV-19/SierraLeone/CPHRL-004/2021 | EPI_ISL_4638335 | 2021-07-12 | Connaught Hospital Laboratory | Central Public Health Reference Laboratory | Allan Campbell, Samuel Sorie, Zein Souma, Santigie Kamara, Julian Campbell, Cyrus Yiaba, Anu Jegede-Williams, Doris Harding, Simon Ruhweza, Prof. Foday Sahr, Prof. Sahr Gevao, Prof. Isatta Wurie |
| hCoV-19/Peru/LAL-INS-1974/2021 | EPI_ISL_3376163 | 2021-02-12 | Laboratorio de Referencial Nacional de Virus Respiratorios | Laboratorio de Referencial Nacional de Virus Respiratorios | Carlos Padilla Rojas, Veronica Hurtado Vela, Orson Mestanza Millones, Iris Silva Molina, Luis Barcena Flores, Sandra Morales Ruiz, Steve Acedo Lazo, Omar Caceres Rey, Marco Galarza Perez, Henri Bailon Calderon, Nancy Rojas Serrano, Priscila Lope Pari, Le |
| hCoV-19/India/OR-ILSGS06656/2021 | EPI_ISL_4103576 | 2021-06-27 | District Public Health Laboratory, Angul | Institute of Life Sciences-INSACOG | Sunil K. Raghav, Safal Walia, Arup Ghosh, Atimukta Jha, Amol M. Kanampalliwar, Sana Fatma, Shifu Aggarwal, Rupesh Dash, Rajeeb Swain, Punit Prasad, INSACOG Consortium, Ajay Parida |
| hCoV-19/USA/TX-Curative-090958/2021 | EPI_ISL_2839933 | 2021-06-22 | Curative Labs | Curative Labs | Mikhail Hanewich-Hollatz, J.Cesar Ignacio-Espinoza, Elias L. Salfati, Nabjot Sandhu, George Way, Zhiyi Xie, Eugenia Khorosheva, Sophia Quasem, Janet Chen, Vladimir Slepnev |
| hCoV-19/Australia/SA0781/2021 | EPI_ISL_2839991 | 2021-06-28 | SA Pathology | SA Pathology | Caitlin Selway, Lex Leong, Chuan Kok Lim, Mark Turra, Ivan Bastian, Geoff Higgins |
| hCoV-19/Pakistan/NIH-B14-S31/2021 | EPI_ISL_3553528 | 2021-07-26 | Department of Virology | Department of Virology | Massab Umair, Aamer Ikram, Muhammad Salman, Nazish Badar, Zaira Rehman, Muhammad Ammar, Abdul Ahad, Syed Adnan Haider |
| hCoV-19/India/DL-NCDC-3309167/2021 | EPI_ISL_4470182 | 2021-05-07 | National Centre for Disease Control (NCDC) Biotechnology Division, Delhi - INSACOG | NCDC Delhi, Biotechnology Division - INSACOG | Radhakrishnan V. S, Robin Marwal, Mahesh S Dhar, Kalaiarasan Ponnusamy, Meena Datta, Priyanka Singh,¬†Uma Sharma,¬†Hemlata Lall, Manoj K Singh, Hema Gogia, Preeti Madan, Sandhya Kabra, Sujeet K Singh, Partha Rakshit |
| hCoV-19/India/PB-NCDC-3309370/2021 | EPI_ISL_4470259 | 2021-05-17 | National Centre for Disease Control (NCDC) Biotechnology Division, Delhi - INSACOG | NCDC Delhi, Biotechnology Division - INSACOG | Radhakrishnan V. S, Robin Marwal, Mahesh S Dhar, Kalaiarasan Ponnusamy, Meena Datta, Priyanka Singh,¬†Uma Sharma,¬†Hemlata Lall, Manoj K Singh, Hema Gogia, Preeti Madan, Sandhya Kabra, Sujeet K Singh, Partha Rakshit |
| hCoV-19/USA/UT-UPHL-210624031915/2021 | EPI_ISL_3132998 | 2021-06-10 | Utah Public Health Laboratory | Utah Public Health Laboratory | Olinto Linares-Perdomo, Erin L. Young, Kelly F. Oakeson |

| | | | | | |
|---|---|---|---|---|---|
| hCoV-19/Canada/BC-BCCDC-168985/2021 | EPI_ISL_4423121 | 2021-03-24 | British Columbia Centre For Disease Control | BCCDC Public Health Laboratory | Prystajecky Natalie, Linda Hoang, Dan Fornika, John Tyson, Shannon Russell, Kim Macdonald, Kimia Kamelian, Ana Pacagnella, Corrinne Ng, Loretta Janz, Robert Azana, Mel Krajden |
| hCoV-19/Canada/BC-BCCDC-158483/2021 | EPI_ISL_4423544 | 2021-04-21 | British Columbia Centre For Disease Control | BCCDC Public Health Laboratory | Prystajecky Natalie, Linda Hoang, Dan Fornika, John Tyson, Shannon Russell, Kim Macdonald, Kimia Kamelian, Ana Pacagnella, Corrinne Ng, Loretta Janz, Robert Azana, Mel Krajden |
| hCoV-19/Canada/BC-BCCDC-91563/2021 | EPI_ISL_3486773 | 2021-05-20 | British Columbia Centre For Disease Control | BCCDC Public Health Laboratory | Prystajecky Natalie, Linda Hoang, Dan Fornika, John Tyson, Shannon Russell, Kim Macdonald, Kimia Kamelian, Ana Pacagnella, Corrinne Ng, Loretta Janz, Robert Azana, Mel Krajden |
| hCoV-19/USA/WA-UW-21081704175/2021 | EPI_ISL_4434737 | 2021-08-17 | UW Virology Lab | UW Virology Lab | Pavitra Roychoudhury, Shah Mohamed Bakhash, Ricardo Perez, Maria Lukes, Hong Xie, Tien V. Nguyen, Savanna S. Carmack, Sean Ellis, Meei-Li Huang, Keith R Jerome, Alexander Greninger |
| hCoV-19/Madagascar/IPM-09269/2020 | EPI_ISL_3716408 | 2020-05-14 | Virology Unit, Institut Pasteur de Madagascar | Institut Pateur de Dakar | Norosoa Razanajatovo, Cara E. Brook, Diagne Moussa Mo√Øse, Diallo Amadou, Dia Ndongo, Christian Ranaivoson, Soa Fy Andriamandimby, Vololoniaina Raharinosy, Sankhe Safietou, Ndiaye Ndack, Tsiry Randriambolamanantsoa, Helisoa Razafimanjato, Jean-Michel Hera |
| hCoV-19/Madagascar/IPM-38381/2020 | EPI_ISL_3716665 | 2020-10-27 | Virology Unit, Institut Pasteur de Madagascar | Institut Pateur de Dakar | Norosoa Razanajatovo, Cara E. Brook, Diagne Moussa Mo√Øse, Diallo Amadou, Dia Ndongo, Christian Ranaivoson, Soa Fy Andriamandimby, Vololoniaina Raharinosy, Sankhe Safietou, Ndiaye Ndack, Tsiry Randriambolamanantsoa, Helisoa Razafimanjato, Jean-Michel Hera |
| hCoV-19/Madagascar/IPM-10139/2020 | EPI_ISL_3716413 | 2020-05-22 | Virology Unit, Institut Pasteur de Madagascar | Institut Pateur de Dakar | Norosoa Razanajatovo, Cara E. Brook, Diagne Moussa Mo√Øse, Diallo Amadou, Dia Ndongo, Christian Ranaivoson, Soa Fy Andriamandimby, Vololoniaina Raharinosy, Sankhe Safietou, Ndiaye Ndack, Tsiry Randriambolamanantsoa, Helisoa Razafimanjato, Jean-Michel Hera |
| hCoV-19/Madagascar/IPM-20000/2020 | EPI_ISL_3716491 | 2020-06-22 | Virology Unit, Institut Pasteur de Madagascar | Institut Pateur de Dakar | Norosoa Razanajatovo, Cara E. Brook, Diagne Moussa Mo√Øse, Diallo Amadou, Dia Ndongo, Christian Ranaivoson, Soa Fy Andriamandimby, Vololoniaina Raharinosy, Sankhe Safietou, Ndiaye Ndack, Tsiry Randriambolamanantsoa, Helisoa Razafimanjato, Jean-Michel Hera |
| hCoV-19/Madagascar/IPM-20523/2020 | EPI_ISL_3716498 | 2020-07-04 | Virology Unit, Institut Pasteur de Madagascar | Institut Pateur de Dakar | Norosoa Razanajatovo, Cara E. Brook, Diagne Moussa Mo√Øse, Diallo Amadou, Dia Ndongo, Christian Ranaivoson, Soa Fy Andriamandimby, Vololoniaina Raharinosy, Sankhe Safietou, Ndiaye Ndack, Tsiry Randriambolamanantsoa, Helisoa Razafimanjato, Jean-Michel Hera |

| | | | | | |
|---|---|---|---|---|---|
| hCoV-19/India/PB-NCDC-3109138/2021 | EPI_ISL_4436005 | 2021-04-30 | National Centre for Disease Control (NCDC) Biotechnology Division, Delhi - INSACOG | NCDC Delhi, Biotechnology Division - INSACOG | Radhakrishnan V. S, Robin Marwal, Mahesh S Dhar, Kalaiarasan Ponnusamy, Meena Datta, Priyanka Singh,¬†Uma Sharma,¬†Hemlata Lall, Manoj K Singh, Hema Gogia, Preeti Madan, Sandhya Kabra, Sujeet K Singh, Partha Rakshit |
| hCoV-19/India/GJ-ICMR-NIV-INSACOG-GSEQ-4122/2021 | EPI_ISL_2965852 | 2021-05-30 | ICMR-National Institute of Virology - INSACOG | NIV Influenza | Potdar, Varsha on behalf of National Influenza Centre, NIV, Pune |
| hCoV-19/Argentina/INEI106062/2021 | EPI_ISL_3867443 | 2021-06-04 | Servicio Virosis Respiratorias-Departamento Virolog√≠a-INEI | Instituto Nacional Enfermedades Infecciosas C.G.Malbran | Baumeister E., Avaro M., Benedetti E., Russo M., Dattero ME, Pontoriero A., Cisterna D., Molina V., Perandones C., Tuduri E., De Belder D., Sanchez Loria J., Haim MS., Lorenzo F., Poklepovich T., Campos J. |
| hCoV-19/Brazil/PR-FIOCRUZ-47436/2021 | EPI_ISL_4237183 | 2021-07-28 | Laboratorio Central de Saude Publica do Estado do Parana (LACEN/PR) | Laboratory of Respiratory Viruses and Measles, Oswaldo Cruz Institute, FIOCRUZ | Paola Resende, Luciana Appolinario, Fernando Motta, Anna Carolina Paixao, Ighor Leonardo Arantes, Agatha Soares, Ana Carolina Mendonca, Alice Sampaio Rocha, Taina Venas, Elisa Cavalcante Pereira, Renata Serrano Lopes, Irina Riediger, Marilda Siqueira on b |
| hCoV-19/India/DL-NCDC-3309186/2021 | EPI_ISL_4441264 | 2021-05-12 | National Centre for Disease Control (NCDC) Biotechnology Division, Delhi - INSACOG | NCDC Delhi, Biotechnology Division - INSACOG | Radhakrishnan V. S, Robin Marwal, Mahesh S Dhar, Kalaiarasan Ponnusamy, Meena Datta, Priyanka Singh,¬†Uma Sharma,¬†Hemlata Lall, Manoj K Singh, Hema Gogia, Preeti Madan, Sandhya Kabra, Sujeet K Singh, Partha Rakshit |
| hCoV-19/Turkey/HSGM-GF1727/2021 | EPI_ISL_3256451 | 2021-07-12 | Ministry of Health Turkey | Ministry of Health Turkey | Fatma Bayrakdar, Yasemin Co≈ügun, S√ºleyman Yalcin, G√ºlay Korukluofülu |
| hCoV-19/Turkey/HSGM-GF2007/2021 | EPI_ISL_3257000 | 2021-07-15 | Ministry of Health Turkey | Ministry of Health Turkey | Fatma Bayrakdar, Yasemin Co≈ügun, S√ºleyman Yalcin, G√ºlay Korukluofülu |
| hCoV-19/Turkey/HSGM-GF2323/2021 | EPI_ISL_3257211 | 2021-07-15 | Ministry of Health Turkey | Ministry of Health Turkey | Fatma Bayrakdar, Yasemin Co≈ügun, S√ºleyman Yalcin, G√ºlay Korukluofülu |
| hCoV-19/Turkey/HSGM-GF2249/2021 | EPI_ISL_3257139 | 2021-07-15 | Ministry of Health Turkey | Ministry of Health Turkey | Fatma Bayrakdar, Yasemin Co≈ügun, S√ºleyman Yalcin, G√ºlay Korukluofülu |
| hCoV-19/Turkey/HSGM-E3282/2021 | EPI_ISL_4101916 | 2021-03-09 | Ministry of Health Turkey | Ministry of Health Turkey | Fatma Bayrakdar, Yasemin Cosgun, Suleyman Yalcin, Gulay Korukluoglu |
| hCoV-19/Turkey/HSGM-E2979/2021 | EPI_ISL_4101660 | 2021-03-09 | Ministry of Health Turkey | Ministry of Health Turkey | Fatma Bayrakdar, Yasemin Cosgun, Suleyman Yalcin, Gulay Korukluoglu |
| hCoV-19/Turkey/HSGM-E2951/2021 | EPI_ISL_4101635 | 2021-03-09 | Ministry of Health Turkey | Ministry of Health Turkey | Fatma Bayrakdar, Yasemin Cosgun, Suleyman Yalcin, Gulay Korukluoglu |
| hCoV-19/Turkey/HSGM-E3019/2021 | EPI_ISL_4101691 | 2021-03-09 | Ministry of Health Turkey | Ministry of Health Turkey | Fatma Bayrakdar, Yasemin Cosgun, Suleyman Yalcin, Gulay Korukluoglu |
| hCoV-19/Turkey/HSGM-E3070/2021 | EPI_ISL_4101733 | 2021-03-09 | Ministry of Health Turkey | Ministry of Health Turkey | Fatma Bayrakdar, Yasemin Cosgun, Suleyman Yalcin, Gulay Korukluoglu |
| hCoV-19/Turkey/HSGM-E3243/2021 | EPI_ISL_4101882 | 2021-03-09 | Ministry of Health Turkey | Ministry of Health Turkey | Fatma Bayrakdar, Yasemin Cosgun, Suleyman Yalcin, Gulay Korukluoglu |
| hCoV-19/Turkey/HSGM-E3291/2021 | EPI_ISL_4101925 | 2021-03-09 | Ministry of Health Turkey | Ministry of Health Turkey | Fatma Bayrakdar, Yasemin Cosgun, Suleyman Yalcin, Gulay Korukluoglu |
| hCoV-19/Turkey/HSGM-E763/2021 | EPI_ISL_4102267 | 2021-08-20 | Ministry of Health Turkey | Ministry of Health Turkey | Fatma Bayrakdar, Yasemin Cosgun, Suleyman Yalcin, Gulay Korukluoglu |

| | | | | | |
|---|---|---|---|---|---|
| hCoV-19/Georgia/Tb-SNGS726/2021 | EPI_ISL_3996683 | 2021-08-06 | Department for Virology, Molecular Biology and Genome Research, R. G. Lugar Center for Public Health Research, National Center for Disease Control and Public Health (NCDC) of Georgia. | [illegible] Biology and Genome Research, R. G. Lugar Center for Public Health Research, National Center for Disease Control and Public Health (NCDC) of Georgia. | Giorgi Tomashvili, Giorgi Gogoladze, Meri Pantsulaia, Gvantsa Brachveli, Salome Javashvili, Nino Berishvili, Tata Imnadze, Ana Papkiauri, Gvantsa Chanturia, Ann Machablishvili, Nato Kotaria, Marine Murtskhvaladze, Lela Sabadze, Mari Gavashelidze, Tamar Ja |
| hCoV-19/USA/FL-CDC-ASC210243747/2021 | EPI_ISL_3817488 | 2021-07-24 | Aegis Sciences Corporation | Centers for Disease Control and Prevention Division of Viral Diseases, Pathogen Discovery | Dakota Howard,Dhwani Batra,Peter Cook,Kara Moser,Adrian Paskey,Jason Caravas,Benjamin Rambo-Martin,Shatavia Morrison,Christopher Gulvick,Scott Sammons,Yvette Unoarumhi,Darlene Wagner,Matthew Schmerer,Cyndi Clark,Patrick Campbell,Rob Case,Vikramsinha Ghorp |
| hCoV-19/Italy/LOM-Policlinico_Milano_18855887/2021 | EPI_ISL_3100596 | 2021-07-09 | Fondazione IRCCS Ca' Granda Ospedale Maggiore Policlinico | Fondazione IRCCS Ca' Granda Ospedale Maggiore Policlinico | Ferruccio Ceriotti, Sara Uceda Renteria |
| hCoV-19/Italy/SIC-CQRC-2450564-2036/2021 | EPI_ISL_4628171 | 2021-09-15 | Laboratorio CQRC | CQRC_QUALITY CONTROL CHEMICAL BIOLOGICAL RISK_AOOR Villa Sofia Cervello Palermo | Di Gaudio F., Contino F., Brunacci G. |
| hCoV-19/Italy/SIC-CQRC-2450593-2031/2021 | EPI_ISL_4628112 | 2021-09-15 | Laboratorio CQRC | CQRC_QUALITY CONTROL CHEMICAL BIOLOGICAL RISK_AOOR Villa Sofia Cervello Palermo | Di Gaudio F., Contino F., Brunacci G. |
| hCoV-19/Italy/SIC-CQRC-2443229-2023/2021 | EPI_ISL_4628142 | 2021-09-15 | Laboratorio CQRC | CQRC_QUALITY CONTROL CHEMICAL BIOLOGICAL RISK_AOOR Villa Sofia Cervello Palermo | Di Gaudio F., Contino F., Brunacci G. |
| hCoV-19/Australia/NSW-RPAH-1517/2021 | EPI_ISL_4520653 | 2021-09-02 | New South Wales Health Pathology Royal Prince Alfred Hospital | Microbiology RPAH | Foster, C.; Au, J.; Ruiz Silva, M.; Deveson, I.; Bull, R.; Van Hal, S.; Rawlinson, W. |
| hCoV-19/Lebanon/QIB-375/2021 | EPI_ISL_3343575 | 2021-02-24 | National Influenza Center | Quadram Institute Bioscience | Georgi Merhi, Alexander J. Trotter, Leonardo de Oliveira Martins, Jad Koweyes, Thanh Le-Viet, Hala Abou Naja, Mona Al Buaini, Sophie J. Prosolek, Nabil-Fareed Alikhan, Martin Lott, Orla J. Jupp, Sarah Gardener, Matthew W. Felgate, Kate A. Makin, Janine M. |
| hCoV-19/Japan/PG-82255/2021 | EPI_ISL_3894729 | 2021-06-29 | Pathogen Genomics Center, National Institute of Infectious Diseases | Pathogen Genomics Center, National Institute of Infectious Diseases | Tsuyoshi Sekizuka, Kentaro Itokawa, Rina Tanaka, Masanori Hashino, Makoto Kuroda |
| hCoV-19/Canada/QC-L00360044001/2021 | EPI_ISL_3458984 | 2021-06-13 | Laboratoire de sant√© publique du Qu√©bec | Laboratoire de sant√© publique du Qu√©bec | Sandrine Moreira, Ioannis Ragoussis, Guillaume Bourque, Jesse Shapiro, Mark Lathrop and Michel Roger on behalf of the CoVSeQ research group |
| hCoV-19/India/RJ-CCMB-CIA6208/2020 | EPI_ISL_3453615 | 2020-12-05 | RUHS College of Medical Sciences | CSIR-Centre for Cellular and Molecular Biology - INSACOG | Dr. Gaurav Dalela,Dr. Vaseem Naheed Baig,Dr. Sonali Sharma,Dr. Ramesh Sharma,Dr. Nilofer Khayyam,Dr. Jitendra Panda,Dr. Sudhanshu Kacker,Dr. Raja Babu Panwar,Dr. Rajeev Gupta, Ara Sreenivas,Shreekant Verma, Amareshwar Vodapalli ,B Himasri,Jandhyala Sai Kr |
| hCoV-19/USA/VI-Yale-10210/2020 | EPI_ISL_4577392 | 2020-08-21 | VIDOH (Virgin Islands Department of Health) | Grubaugh Lab - Yale School of Public Health | Joseph Fauver, Mallery Breban, Isabel Ott, Tara Alpert, Mary Petrone, Anderson Brito, Chantal Vogels, Annie Watkins, Chaney Kalinich, Jessica Rothman, Kendall Billig, Tobias Koch, Jendai Richards, TaLesa Aderohunmu, Esther Ellis, Marlon Lawrence, Brett El |

| | | | | | |
|---|---|---|---|---|---|
| hCoV-19/Kenya/ILRI_COVC15801/2020 | EPI_ISL_4252024 | 2020-10-23 | International Livestock Research Institute | International Livestock Research Institute | Samuel O. Oyola, Gilbert Kibet, Daniel Ouso, Shebbar Osiany, Edward Kiritu, Paul Dobi, Collins Muli, Patrick Amoth, Vishvanath Nene, Sonal P. Henson, Edward O. Abworo |
| hCoV-19/Kenya/ILRI_COVC24072/2021 | EPI_ISL_4252450 | 2021-08-06 | International Livestock Research Institute | International Livestock Research Institute | Samuel O. Oyola, Gilbert Kibet, Daniel Ouso, Shebbar Osiany, Edward Kiritu, Paul Dobi, Collins Muli, Patrick Amoth, Vishvanath Nene, Sonal P. Henson, Edward O. Abworo |
| hCoV-19/Luxembourg/LNS9383012/2021 | EPI_ISL_3148302 | 2021-07-05 | Laboratoires d'analyses medicales - Ketterthill | Laboratoire national de sante, Microbiology, Microbial Genomics Platform | Anke Wienecke-Baldacchino, Catherine Ragimbeau,Jessica Tapp, Fatu Djabi, Lise Pignon, Raoul Salmon, Virginie Jover, Elodie Solarino, Serge Vedy, Caroline Scheiber, Tamir Abdelrahman |
| hCoV-19/USA/CA-CDC-LC0101782/2021 | EPI_ISL_3326006 | 2021-07-10 | Laboratory Corporation of America | Centers for Disease Control and Prevention Division of Viral Diseases, Pathogen Discovery | Dakota Howard, Dhwani Batra, Peter W. Cook, Kara Moser, Adrian Paskey, Jason Caravas, Benjamin Rambo-Martin, Shatavia Morrison, Christopher Gulvick, Scott Sammons, Yvette Unoarumhi, Darlene Wagner, Matthew Schmerer, Minoo Agarwal, Eyad Almasri, Debbie Bol |
| hCoV-19/NewZealand/21MV0701/2021 | EPI_ISL_3506216 | 2021-08-10 | LabPLUS | Institute of Environmental Science and Research (ESR) | Rachel Boyle, SallyAnn Harbison, Olivia Stroeven, Xiaoyun Ren, Matt Storey, Nikki Freed, Muhammad Faisal, Jing Wang, Hermes Perez, Anja Werno, Antje van der Linden, Arlo Upton, Chris Mansell, David Hammer, Dragana Drinkovic, Gary McAuliffe, Hana Sofia And |
| hCoV-19/NewZealand/21MV0729/2021 | EPI_ISL_3543464 | 2021-08-17 | LabPLUS | Institute of Environmental Science and Research (ESR) | Rachel Boyle, SallyAnn Harbison, Olivia Stroeven, Xiaoyun Ren, Matt Storey, Nikki Freed, Muhammad Faisal, Jing Wang, Hermes Perez, Anja Werno, Antje van der Linden, Arlo Upton, Chris Mansell, David Hammer, Dragana Drinkovic, Gary McAuliffe, Hana Sofia And |
| hCoV-19/India/DL-NCDC-4109326/2021 | EPI_ISL_4462425 | 2021-05-16 | National Centre for Disease Control (NCDC) Biotechnology Division, Delhi - INSACOG | NCDC Delhi, Biotechnology Division - INSACOG | Mahesh S Dhar, Kalaiarasan Ponnusamy, Meena Datta, Priyanka Singh,¬†Uma Sharma,¬†Hemlata Lall, Manoj K Singh, Radhakrishnan V. S, Robin Marwal, Hema Gogia, Preeti Madan, Sandhya Kabra, Sujeet K Singh, Partha Rakshit |
| hCoV-19/Senegal/SN-RUF-216234/2021 | EPI_ISL_4276982 | 2021-05-19 | Iressef Genomics lab | IRESSEF | Souleymane MBOUP, Abdou PADANE , Khadim GUEYE, Diabou Diagne , Samba Ndiour, Papa Alassane DIAW, Birahim Piere NDIAYE, Barada CISSE, Aminata MBOUP, Moustapha MBOW, Ndeye Coumba Toure KANE, Nafisatou LEYE, Gora LO, Ambroise AHOUIDI , Astou Gaye GAYE, Amina |
| hCoV-19/Denmark/DCGC-172300/2021 | EPI_ISL_4256616 | 2021-09-12 | Department of Bacteria, Parasites and Fungi, Statens Serum Institut, Copenhagen, Denmark | Statens Serum Institut Bioinformatics and Microbial Genomics | Danish Covid-19 Genome Consortium |
| hCoV-19/Aruba/AW-RIVM-54051/2021 | EPI_ISL_3918772 | 2021-08-11 | Dutch COVID-19 response team | National Institute for Public Health and the Environment (RIVM) | Adam Meijer, Harry Vennema, Dirk Eggink, Jeroen Cremer, Sharon van den Brink, Bas van der Veer, AnneMarie van den Brandt, Lisa Wijsman, Kim Freriks, Ryanne Jaarsma, Eunv≠ce Then, Lynn Aarts, Sanne Bos, Stijn van Rossum, Florian Zwagemaker, Dennis Schmitz, |

| | | | | | |
|---|---|---|---|---|---|
| hCoV-19/India/PB-NCDC-4209312/2021 | EPI_ISL_4462808 | 2021-06-08 | National Centre for Disease Control (NCDC) Biotechnology Division, Delhi - INSACOG | NCDC Delhi, Biotechnology Division - INSACOG | Kalaiarasan Ponnusamy, Meena Datta, Priyanka Singh,¬†Uma Sharma,¬†Hemlata Lall, Manoj K Singh, Radhakrishnan V. S, Robin Marwal, Mahesh S Dhar, Hema Gogia, Preeti Madan, Sandhya Kabra, Sujeet K Singh, Partha Rakshit |
| hCoV-19/India/JK-NCDC-4209132/2021 | EPI_ISL_4462868 | 2021-06-08 | National Centre for Disease Control (NCDC) Biotechnology Division, Delhi - INSACOG | NCDC Delhi, Biotechnology Division - INSACOG | Kalaiarasan Ponnusamy, Meena Datta, Priyanka Singh,¬†Uma Sharma,¬†Hemlata Lall, Manoj K Singh, Radhakrishnan V. S, Robin Marwal, Mahesh S Dhar, Hema Gogia, Preeti Madan, Sandhya Kabra, Sujeet K Singh, Partha Rakshit |
| hCoV-19/India/JK-NCDC-4209257/2021 | EPI_ISL_4462883 | 2021-06-07 | National Centre for Disease Control (NCDC) Biotechnology Division, Delhi - INSACOG | NCDC Delhi, Biotechnology Division - INSACOG | Kalaiarasan Ponnusamy, Meena Datta, Priyanka Singh,¬†Uma Sharma,¬†Hemlata Lall, Manoj K Singh, Radhakrishnan V. S, Robin Marwal, Mahesh S Dhar, Hema Gogia, Preeti Madan, Sandhya Kabra, Sujeet K Singh, Partha Rakshit |
| hCoV-19/Chile/RM-132539/2021 | EPI_ISL_3546353 | 2021-07-31 | Genetica Molecular and Subdepartamento de Virologia ISP Chile | Instituto de Salud Publica de Chile | Karen Orostica, Constanza Campano, Barbara Parra, Loredana Arata, Gisselle Barra, Patricia Bustos, Rodrigo Fasce, Javier Tognarelli, Andres Castillo, Soledad Ulloa, Jorge Fernandez |
| hCoV-19/Turkey/HSGM-FS1229/2021 | EPI_ISL_3406440 | 2021-08-05 | Ministry of Health Turkey | Ministry of Health Turkey | Fatma Bayrakdar, Yasemin Cosgun, Suleyman Yalcin, Gulay Korukluoglu |
| hCoV-19/USA/VA-CDC-LC0260151/2021 | EPI_ISL_4488510 | 2021-09-09 | Laboratory Corporation of America | Centers for Disease Control and Prevention Division of Viral Diseases, Pathogen Discovery | Dakota Howard,Dhwani Batra,Peter Cook,Jason Caravas,Benjamin Rambo-Martin,Scott Sammons,Yvette Unoarumhi,Matthew Schmerer,Kristine Lacek,Tymeckia Kendall,Victoria Caban Figueroa,Shatavia Morrison,Christopher Gulvick,Minoo Agarwal,Eyad Almasri,Debbie Boles |
| hCoV-19/India/UP-IGIB-3346/2021 | EPI_ISL_2881190 | 2021-05-18 | INSACOG Surveillance | INSACOG at CSIR Institute of Genomics and Integrative Biology | INSACOG |
| hCoV-19/Denmark/DCGC-140251/2021 | EPI_ISL_3228487 | 2021-07-30 | Department of Bacteria, Parasites and Fungi, Statens Serum Institut, Copenhagen, Denmark | Statens Serum Institut Bioinformatics and Microbial Genomics | Danish Covid-19 Genome Consortium |
| hCoV-19/Kenya/C103184/2021 | EPI_ISL_3049825 | 2021-07-07 | KEMRI-Wellcome Trust Research Programme,Kilifi | KEMRI-Wellcome Trust Research Programme,Kilifi | Githinji G.,Matoke D.,Mohamed K.S.,de Laurent Z.,Mburu M.W.,Thiongo K. |
| hCoV-19/Italy/SIC-CQRC_2063617/2021 | EPI_ISL_3376098 | 2021-07-18 | Laboratorio CQRC | CQRC_QUALITY CONTROL CHEMICAL BIOLOGICAL RISK_AOOR Villa Sofia Cervello Palermo | Di Gaudio F., Contino F., Brunacci G. |
| hCoV-19/Canada/SK-NML-16581/2021 | EPI_ISL_2883125 | 2021-01-05 | SK-Roy Romanow Provincial Laboratory | National Microbiology Laboratory (NML) | Anna Majer, Shari Tyson, Grace Seo, Philip Mabon, Elsie Grudeski, Rhiannon Huzarewich, Russell Mandes, Anneliese Landgraff, Jennifer Tanner, Natalie Knox, Morag Graham, Gary Van Domselaar, Ryan McDonald, Keith MacKenzie, Meredith Faires, Kara Loos, Rachel |
| hCoV-19/Kenya/C99301/2021 | EPI_ISL_3049427 | 2021-06-06 | KEMRI-Wellcome Trust Research Programme,Kilifi | KEMRI-Wellcome Trust Research Programme,Kilifi | Githinji G.,Matoke D.,Mohamed K.S.,de Laurent Z.,Mburu M.W.,Thiongo K. |
| hCoV-19/Australia/QLD2283/2021 | EPI_ISL_4348498 | 2021-08-27 | unknown | PHV-FSS | Chenwei Wang on behalf of Q-PHIRE Genomics |

| | | | | | |
|---|---|---|---|---|---|
| hCoV-19/Australia/QLD2295/2021 | EPI_ISL_4348500 | 2021-09-01 | unknown | PHV-FSS | Chenwei Wang on behalf of Q-PHIRE Genomics |
| hCoV-19/Australia/VIC18930/2021 | EPI_ISL_3050783 | 2021-07-15 | Microbiological Diagnostic Unit - Public Health Laboratory (MDU-PHL) | MDU-PHL | Seemann T., Sait, M.L., Sherry, N.L. |
| hCoV-19/SaudiArabia/KAIMRC-S454/2021 | EPI_ISL_3799841 | 2021-02-12 | Clinical Microbiology Lab, King Abdulaziz Medical City, Ministry of National Guard ‚Äì Health Affairs, Riyadh, Saudi Arabia | Abdullah International Medical Research Center (KAIMRC), Riyadh, Saudi Arabia | Alghoribi,M.F., Alswaji,A.A., Okdah,L., Alhayli,S., Bu Ali,Z.,Aljohani,S.M. and Doumith,M. |
| hCoV-19/Switzerland/ZH-ETHZ-670917/2021 | EPI_ISL_4316799 | 2021-09-01 | labor team w AG | Department of Biosystems Science and Engineering, ETH Zürich | Ivan Topolsky, Sarah Nadeau, Chaoran Chen, Louis du Plessis, Rebekka Pohl, Monika Bucher, Andreas Lindauer, Catharine Aquino, Daniel Ehrsam, Isabel Stürmer, Andreia Cabral de Gouvea, Maria Domenica Moccia, Simon Grüter, Timothy Sykes, Lennart Opitz, Gri |
| hCoV-19/Turkey/HSGM-FS5254/2021 | EPI_ISL_3414682 | 2021-08-07 | Ministry of Health Turkey | Ministry of Health Turkey | Fatma Bayrakdar, Yasemin Cosgun, Suleyman Yalcin, Gulay Korukluoglu |
| hCoV-19/India/KA-NIMH-SEQ-2072/2021 | EPI_ISL_3232786 | 2021-07-05 | NIMHANS | INSACOG-KA, NIMHANS | Chitra Pattabiraman, Anson Kunjumon George,Darshan Sreenivas, Ellango Ramasamy, Sony Sharma, Mahesh Kumar.C.S, Ananthapadmanabha Kotambail, Chetan G K, Gautham Arunachal Udupi, Anita S Desai, V Ravi |
| hCoV-19/Bulgaria/21BG-NC_003976_R15/2021 | EPI_ISL_3233376 | 2021-07-20 | National Center of Infectious and Parasitic Diseases | National Center of Infectious and Parasitic Diseases | Alexiev et al |
| hCoV-19/Brazil/PR-FIOCRUZ-31708/2021 | EPI_ISL_3801875 | 2021-06-06 | Laboratorio Central de Saude Publica do Estado do Parana (LACEN/PR) | Laboratory of Respiratory Viruses and Measles, Oswaldo Cruz Institute, FIOCRUZ | Paola Resende, Luciana Appolinario, Fernando Motta, Anna Carolina Paixao, Ana Carolina Mendonca, Alice Sampaio Rocha, Taina Venas, Elisa Cavalcante Pereira, Renata Serrano Lopes, Irina Riediger, Marilda Siqueira on behalf of the Fiocruz COVID-19 Genomic S |
| hCoV-19/India/DL-ILBS-B_76/2021 | EPI_ISL_6025366 | 2021-01-27 | Max Super Speciality Hospital | ILBS | Ekta Gupta, Varun Suroliya, Reshu Agarwal, Diptanu Paul, Amit Pandey, Bansidhar Tarai, Chhagan Bihari Sharma, Shiv Kumar Sarin |
| hCoV-19/India/DL-ILBS-B_86/2021 | EPI_ISL_6025418 | 2021-02-14 | Max Super Speciality Hospital | ILBS | Ekta Gupta, Varun Suroliya, Reshu Agarwal, Diptanu Paul, Amit Pandey, Bansidhar Tarai, Chhagan Bihari Sharma, Shiv Kumar Sarin |
| hCoV-19/India/DL-ILBS-WGS_60/2021 | EPI_ISL_6025696 | 2021-09-04 | SANGHI MEDICAL CENTRE | ILBS | Ekta Gupta, Varun Suroliya, Reshu Agarwal, Diptanu Paul, Rahul Garg, Arjun Bhugra, urvinder kaur, santanu das, Pramod Gautam, Chhagan Bihari Sharma, Shiv Kumar Sarin |
| hCoV-19/Brazil/PA-FIOCRUZ-57981/2021 | EPI_ISL_6898704 | 2021-10-21 | Laboratorio Central de Saude Publica do Estado do Para (LACEN/PA) | Laboratory of Respiratory Viruses and Measles, Oswaldo Cruz Institute, FIOCRUZ | Paola Resende, Luciana Appolinario, Fernando Motta, Elisa Cavalcante Pereira, Ighor Arantes, Alice Sampaio Rocha, Bruna Mendonça da Silva, Jéssica Graça Macedo de Carvalho, Larissa Macedo Pinto, Victor Guimaraes, Valnete Andrade, Marilda Siqueira on be |
| hCoV-19/SouthAfrica/NICD-N20491/2021 | EPI_ISL_6900024 | 2021-10-20 | KIMBERLEY LABORATORY | Diseases of the National Health Laboratory Service | Amoako DG, Everatt J, Scheepers C, Mohale T, Ntuli N, Mahlangu B, Mnguni A, Ismail A, Bhiman JN |
| hCoV-19/SouthAfrica/NICD-N20511/2021 | EPI_ISL_6900031 | 2021-10-04 | AMPATH | Diseases of the National Health Laboratory Service | Amoako DG, Everatt J, Scheepers C, Mohale T, Ntuli N, Mahlangu B, Mnguni A, Ismail A, Bhiman JN |
| hCoV-19/SouthAfrica/NICD-N20522/2021 | EPI_ISL_6900039 | 2021-10-04 | AMPATH | Diseases of the National Health Laboratory Service | Amoako DG, Everatt J, Scheepers C, Mohale T, Ntuli N, Mahlangu B, Mnguni A, Ismail A, Bhiman JN |

| | | | | | |
|---|---|---|---|---|---|
| hCoV-19/SouthAfrica/NICD-N20551/2021 | EPI_ISL_6900057 | 2021-10-05 | AMPATH | Diseases of the National Health Laboratory Service | Amoako DG, Everatt J, Scheepers C, Mohale T, Ntuli N, Mahlangu B, Mnguni A, Ismail A, Bhiman JN |
| hCoV-19/SouthAfrica/NICD-N20564/2021 | EPI_ISL_6900068 | 2021-10-06 | AMPATH | Diseases of the National Health Laboratory Service | Amoako DG, Everatt J, Scheepers C, Mohale T, Ntuli N, Mahlangu B, Mnguni A, Ismail A, Bhiman JN |
| hCoV-19/SouthAfrica/NICD-N20740/2021 | EPI_ISL_6900095 | 2021-11-03 | HELEN JOSEPH LABORATORY | Diseases of the National Health Laboratory Service | Amoako DG, Everatt J, Scheepers C, Mohale T, Ntuli N, Mahlangu B, Mnguni A, Ismail A, Bhiman JN |
| hCoV-19/Israel/SMC-7022731/2021 | EPI_ISL_6900200 | 2021-11-20 | Shamir Medical Center (Asaf Harofe) | Shamir Medical Center (Asaf Harofe) | Nir Rainy,Chen Weiner,Reut Sorek Abramovich,Yevgeni Yegorov,Anna Vishnevsky,Patricia Benveniste-Lekovitz,Abu Hamad Ramzia,Adina Bar Chaim |
| hCoV-19/Spain/MD-HGUGM-5835945/2021 | EPI_ISL_6902675 | 2021-11-30 | Hospital General Universitario Gregorio Marav±V≥n | Hospital General Universitario Gregorio Marav±V≥n | Sergio Buenestado Serrano, Jorge RodrV≠guez-Grande, Pedro Sola Campoy, Laura PV©rez-Lago, Cristina Rodriguez-Grande, Marta Herranz Martin, Julia SuV°rez, Pilar CatalV°n, Patricia MuV±oz, DarV≠o GarcV≠a de Viedma |
| hCoV-19/India/TG-CDFD-K48/2021 | EPI_ISL_6903643 | 2021-11-10 | Gandhi Hospital | Centre for DNA Fingerprinting and Diagnostics | Murali Bashyam, Asmita Gupta, Arunkumar Karunanidhi, Vinay Donipadi, Nagamani Kammili, Divya Vashisht, Ashwin Dalal |
| hCoV-19/India/TG-CDFD-NMCVD48896/2021 | EPI_ISL_6903655 | 2021-11-05 | Nizam's Institute of Medical Sciences | Centre for DNA Fingerprinting and Diagnostics | Murali Bashyam, Asmita Gupta, Arunkumar Karunanidhi, Vinay Donipadi, Nagamani Kammili, Divya Vashisht, Ashwin Dalal |
| hCoV-19/USA/VT-CDCBI-CRSP_J72R7CK2GARCACGX/2021 | EPI_ISL_6035537 | 2021-10-30 | Broad Institute Clinical Research Sequencing Platform | Infectious Disease Program, Broad Institute of Harvard and MIT | Siddle,K.J., Adams,G., Pearlman,L., Gladden-Young,A., Vicente,G., Blumenstiel,B., DeFelice,M., Lee,M., McGovern,S., Lagerborg,K., Rudy,M., DeRuff,K., Carter,A., Normandin,E., Bauer,M., Reilly,S., Tomkins-Tinch,C., Loreth,C., Chaluvadi,S., Meldrim,J., Gran |
| hCoV-19/Netherlands/FR-UMCG-MMB_3239/2021 | EPI_ISL_6035907 | 2021-10-28 | UMC Groningen, Clinical Virology, Department of Medical Microbiology and Infection Prevention | UMC Groningen, Clinical Virology, Department of Medical Microbiology and Infection Prevention | Hubert Niesters, Alexander Friedrich, Erley Lizarazo-Forero, Monika Fliss, Lilli Gard, Sigrid Rosema, Coretta Van Leer-Buter, Xuewei Zhou, Marjolein Knoester |
| hCoV-19/Switzerland/TI-EOC-26328313/2021 | EPI_ISL_6036175 | 2021-10-29 | Laboratorio di Microbiologia | Laboratorio di Microbiologia | Martinetti Lucchini Gladys, Valeria Spina |
| hCoV-19/Sweden/01_SE100_21CS513207/2021 | EPI_ISL_6036964 | 2021-10-25 | Karolinska University Hospital Solna | Karolinska University Hospital | Jan Albert ,Tobias Allander ,Annelie Bjerkner ,Sandra Broddesson ,Robert Dyrdak ,Martin Ekman ,Lynda Eneh ,Lina Guerra Blomqvist ,Karolina Ininbergs ,Tanja Normark ,Isak Sylvin ,Zhibing Yun ,Martina Wahlund ,Valtteri Wirta |
| hCoV-19/SouthAfrica/NICD-N21190/2021 | EPI_ISL_6906383 | 2021-10-07 | TAMBO MEMORIAL LABORATORY | Diseases of the National Health Laboratory Service | Amoako DG, Everatt J, Scheepers C, Mohale T, Ntuli N, Mahlangu B, Mnguni A, Ismail A, Bhiman JN, Wolter N |
| hCoV-19/SouthAfrica/NICD-N19697/2020 | EPI_ISL_6906398 | 2020-12-31 | KIMBERLEY LABORATORY | Diseases of the National Health Laboratory Service | Amoako DG, Everatt J, Scheepers C, Mohale T, Ntuli N, Mahlangu B, Mnguni A, Ismail A, Bhiman JN, Wolter N |
| hCoV-19/England/ALDP-290B8A4/2021 | EPI_ISL_6038174 | 2021-11-01 | Lighthouse Lab in Alderley Park | Wellcome Sanger Institute for the COVID-19 Genomics UK (COG-UK) Consortium | Jacquelyn Wynn, Mairead Hyland, The Lighthouse Lab in Alderley Park and Alex Alderton, Roberto Amato, Jeffrey Barrett, Sonia Goncalves, Ewan Harrison, David K. Jackson, Ian Johnston, Dominic Kwiatkowski, Cordelia Langford, John Sillitoe on behalf of the W |
| hCoV-19/Zambia/ZMBID_6745/2020 | EPI_ISL_6838995 | 2020-08-14 | Right to care Zambia | Macha Research Trust | Muleya W, Mubemba B, Katowa B, Matoba J, Mupila Z, Kwenda G, Gill CJ, Simulundu E, Mwananyanda L |

| | | | | | |
|---|---|---|---|---|---|
| hCoV-19/Philippines/PH-PGC-64400/2021 | EPI_ISL_5544392 | 2021-07-28 | Philippine Red Cross - Port Area | Philippine Genome Center | Cynthia P. Saloma,Jan Michael C. Yap,Benedict A. Maralit,Marc Edsel C. Ayes,Eva Maria Cutiongco-de la Paz,Francis A. Tablizo,Carlo M. Lapid,El King D. Morado,Joshua Gregor A. Dizon,Kenneth M. Kim,Maria Sofia L. Yangzon,Elcid Aaron R. Pangilinan,Renato Jac |
| hCoV-19/USA/KS-CDC-QDX31046824/2021 | EPI_ISL_6907940 | 2021-11-13 | Quest Diagnostics Incorporated | Centers for Disease Control and Prevention Division of Viral Diseases, Pathogen Discovery | Dakota Howard,Dhwani Batra,Peter Cook,Jason Caravas,Benjamin Rambo-Martin,Scott Sammons,Yvette Unoarumhi,Matthew Schmerer,Kristine Lacek,Tymeckia Kendall,Victoria Caban Figueroa,Shatavia Morrison,Christopher Gulvick,Erisa Sula,S. Rosenthal,A. Gerasimova,R |
| hCoV-19/USA/CT-CDC-QDX31092359/2021 | EPI_ISL_6907950 | 2021-11-13 | Quest Diagnostics Incorporated | Centers for Disease Control and Prevention Division of Viral Diseases, Pathogen Discovery | Dakota Howard,Dhwani Batra,Peter Cook,Jason Caravas,Benjamin Rambo-Martin,Scott Sammons,Yvette Unoarumhi,Matthew Schmerer,Kristine Lacek,Tymeckia Kendall,Victoria Caban Figueroa,Shatavia Morrison,Christopher Gulvick,Erisa Sula,S. Rosenthal,A. Gerasimova,R |
| hCoV-19/Venezuela/Dtto1597/2021 | EPI_ISL_6989659 | 2021-08-10 | IVIC | Laboratorio de Virologí≠a Molecular | Rossana C Jaspe, Carmen L Loureiro, Yoneira Sulbaran, Pierina D¬¥Angelo, Zoila Moros, Lieska Rodr√≠guez, Jos√© Luis Zambrano, Mariana Hidalgo, V√≠ctor Alarc√≥n, Domingo J Garzaro, CoViVen Group, H√©ctor R Rangel, Flor H Pujol |
| hCoV-19/India/AP-CCMB-CIB0411/2021 | EPI_ISL_6047354 | 2021-09-02 | AP SSO | CSIR-Centre for Cellular and Molecular Biology-INSACOG | Lamuk Zaveri,Tulasi Nagabandi,Payel Mukherjee,Onkar Kulkarni, Sofia Banu, Jandhyala Sai Krishna,Ara Sreenivas,Shreekant Verma, Amareshwar Vodapalli ,B Himasri,Valli Nagalakshmi Undamatla, Sumedha Avadhanula, Priya Nurkurthy, Vidhyadhari Methuku, Archana B |
| hCoV-19/India/AP-CCMB-CIB0438/2021 | EPI_ISL_6047388 | 2021-09-07 | AP SSO | CSIR-Centre for Cellular and Molecular Biology-INSACOG | Tulasi Nagabandi,Lamuk Zaveri,Payel Mukherjee,Onkar Kulkarni,Jandhyala Sai Krishna, Sofia Banu, Ara Sreenivas,Shreekant Verma, Amareshwar Vodapalli ,B Himasri,Valli Nagalakshmi Undamatla, Priya Nurkurthy,Sumedha Avadhanula, Vidhyadhari Methuku, Archana Bh |
| hCoV-19/Brazil/SP-IAL-6479/2021 | EPI_ISL_6913917 | 2021-11-27 | Grupo CR Diagnosticos | Instituto Adolfo Lutz Strategic Laboratory | Claudio Tavares Sacchi, Karoline Rodrigues Campos |
| hCoV-19/SouthAfrica/NICD-N21437/2021 | EPI_ISL_6913991 | 2021-11-09 | Lancet Laboratories | Diseases of the National Health Laboratory Service | Amoako DG, Everatt J, Scheepers C, Mohale T, Ntuli N, Mahlangu B, Mnguni A, Ismail A, Bhiman JN, Wolter N |
| hCoV-19/SouthAfrica/NICD-N21624/2021 | EPI_ISL_6914015 | 2021-11-17 | Ampath Laboratories | Diseases of the National Health Laboratory Service | Amoako DG, Everatt J, Scheepers C, Mohale T, Ntuli N, Mahlangu B, Mnguni A, Ismail A, Bhiman JN, Wolter N |
| hCoV-19/SouthAfrica/NICD-N21656/2021 | EPI_ISL_6914032 | 2021-11-17 | Ampath Laboratories | Diseases of the National Health Laboratory Service | Amoako DG, Everatt J, Scheepers C, Mohale T, Ntuli N, Mahlangu B, Mnguni A, Ismail A, Bhiman JN, Wolter N |
| hCoV-19/Japan/IC-2279/2021 | EPI_ISL_6914908 | 2021-11-27 | SARS-CoV-2 testing team, National Institute of Infectious Diseases | Pathogen Genomics Center, National Institute of Infectious Diseases | Tsuyoshi Sekizuka, Kentaro Itokawa, Rina Tanaka, Masanori Hashino, Nozomu Hanaoka, Masumichi Saito, Naomi Nojiri, Hazuka Y Furihata, Tsuguto Fujimoto, Makoto Kuroda |

| | | | | | |
|---|---|---|---|---|---|
| hCoV-19/India/WB-INSACOG-1931300478870/2021 | EPI_ISL_6885834 | 2021-10-02 | INSACOG-WB | National Institute of Biomedical Genomics ,Äì INSACOG | Arindam Maitra, Bhaswati Bandyopadhyay, Nidhan Kumar Biswas, Tamal Ghosh, Sreedhar Chinnaswamy, Ajay Chakraborti, Saumitra Das |
| hCoV-19/France/PAC-CERBAHC-11489236/2021 | EPI_ISL_6916818 | 2021-11-10 | CERBALLIANCE PROVENCE | CERBA HealthCare | B√©n√©dicte Roquebert, Laura Verdurme, Sabine Trombert, St√©phanie Haim-Boukobza |
| hCoV-19/England/MILK-1F92C0D/2021 | EPI_ISL_4752610 | 2021-09-27 | Lighthouse Lab in Milton Keynes | Wellcome Sanger Institute for the COVID-19 Genomics UK (COG-UK) Consortium | The Lighthouse Lab in Milton Keynes and Alex Alderton, Roberto Amato, Jeffrey Barrett, Sonia Goncalves, Ewan Harrison, David K. Jackson, Ian Johnston, Dominic Kwiatkowski, Cordelia Langford, John Sillitoe on behalf of the Wellcome Sanger Institute COVID-1 |
| hCoV-19/UnitedArabEmirates/BTC-4951/2020 | EPI_ISL_5142661 | 2020-10-01 | BTC, Khalifa University | BTC, Khalifa University | Habiba Al Safar et al |
| hCoV-19/UnitedArabEmirates/BTC-5010/2020 | EPI_ISL_5142662 | 2020-10-06 | BTC, Khalifa University | BTC, Khalifa University | Habiba Al Safar et al |
| hCoV-19/Chile/AT-167340/2021 | EPI_ISL_6052399 | 2021-10-13 | Genetica Molecular and Subdepartamento de Virologia ISP Chile | Instituto de Salud Publica de Chile | Karen Orostica, Constanza Campano, Barbara Parra, Patricia Bustos, Rodrigo Fasce, Andres Castillo, Jorge Fernandez |
| hCoV-19/Israel/SMC-7014718/2021 | EPI_ISL_6055411 | 2021-08-26 | Shamir Medical Center (Asaf Harofe) | Shamir Medical Center (Asaf Harofe) | Nir Rainy,Chen Weiner,Reut Sorek Abramovich,Yevgeni Yegorov,Anna Vishnevsky,Patricia Benveniste-Lekovitz,Abu Hamad Ramzia,Adina Bar Chaim |
| hCoV-19/USA/NY-NYCPHL-007056/2021 | EPI_ISL_5145671 | 2021-10-04 | DOHMH Morrisania | New York City Public Health Laboratory | Jade Wang, et al. |
| hCoV-19/Colombia/QUI-UTP-VG-259/2021 | EPI_ISL_5146247 | 2021-08-18 | Universidad del Quind√≠o | Laboratorio de Biolog√≠a Molecular y Biotecnolog√≠a - Universidad Tecnol√≥gica de Pereira | Juan D. Anacona-Montilla, Fredy A. Tabares-Villa, Marcela Orjuela-Rodriguez, Augusto Zuluaga-Velez, Juan C. Sepulveda |
| hCoV-19/USA/VT-CDCBI-CRSP_7BXES7MCJR5IRMRA/2021 | EPI_ISL_5457057 | 2021-10-13 | Broad Institute Clinical Research Sequencing Platform | Infectious Disease Program, Broad Institute of Harvard and MIT | Siddle,K.J., Adams,G., Pearlman,L., Gladden-Young,A., Vicente,G., Blumenstiel,B., DeFelice,M., Lee,M., McGovern,S., Lagerborg,K., Rudy,M., DeRuff,K., Carter,A., Normandin,E., Bauer,M., Reilly,S., Tomkins-Tinch,C., Loreth,C., Chaluvadi,S., Meldrim,J., Gran |
| hCoV-19/Bolivia/SAN-UPCH-1022/2020 | EPI_ISL_5147922 | 2020-11-12 | Laboratorio de Diagn√≥stico e Investigaci√≥n BIOSCIENCE SRL | Laboratorio de Gen√≥mica Microbiana, Universidad Peruana Cayetano Heredia | Pablo Tsukayama, Alejandra D√°vila-Barclay, Guillermo Salvatierra, Luis Gonz√°lez, Pedro E. Romero, Diego Cuicapuza, Janet Huancachoque, Pool Marcos, Esdenka P√©rez, Mauricio Prado, Freddy Tinajeros, Paula Carballo |
| hCoV-19/USA/CA-CDPH-3000149029/2021 | EPI_ISL_5151273 | 2021-08-10 | CDPH VBL | California Department of Public Health | Emily Smith on behalf of CDPH-COVIDNet |
| hCoV-19/USA/PA-CDC-ASC210479015/2021 | EPI_ISL_6419225 | 2021-10-25 | Aegis Sciences Corporation | Centers for Disease Control and Prevention Division of Viral Diseases, Pathogen Discovery | Dakota Howard,Dhwani Batra,Peter Cook,Jason Caravas,Benjamin Rambo-Martin,Scott Sammons,Yvette Unoarumhi,Matthew Schmerer,Kristine Lacek,Tymeckia Kendall,Victoria Caban Figueroa,Shatavia Morrison,Christopher Gulvick,Erisa Sula,Cyndi Clark,Patrick Campbell |

| | | | | | |
|---|---|---|---|---|---|
| hCoV-19/USA/TX-CDC-ASC210089826/2021 | EPI_ISL_6061639 | 2021-10-05 | Aegis Sciences Corporation | Centers for Disease Control and Prevention Division of Viral Diseases, Pathogen Discovery | Dakota Howard,Dhwani Batra,Peter Cook,Jason Caravas,Benjamin Rambo-Martin,Scott Sammons,Yvette Unoarumhi,Matthew Schmerer,Kristine Lacek,Tymeckia Kendall,Victoria Caban Figueroa,Shatavia Morrison,Christopher Gulvick,Erisa Sula,Cyndi Clark,Patrick Campbell |
| hCoV-19/Argentina/INEI107884/2021 | EPI_ISL_5158413 | 2021-07-15 | Servicio Virosis Respiratorias-Departamento Virolog√≠a-INEI | Instituto Nacional Enfermedades Infecciosas C.G.Malbran | Baumeister E., Avaro M., Benedetti E., Russo M., Dattero ME, Pontoriero A., Cisterna D., Molina V., Perandones C., Tuduri E., De Belder D., Sanchez Loria J., Haim MS., Lorenzo F., Poklepovich T., Campos J. |
| hCoV-19/Argentina/INEI108962/2021 | EPI_ISL_5158684 | 2021-08-03 | Servicio Virosis Respiratorias-Departamento Virolog√≠a-INEI | Instituto Nacional Enfermedades Infecciosas C.G.Malbran | Baumeister E., Avaro M., Benedetti E., Russo M., Dattero ME, Pontoriero A., Cisterna D., Molina V., Perandones C., Tuduri E., De Belder D., Sanchez Loria J., Haim MS., Lorenzo F., Poklepovich T., Campos J. |
| hCoV-19/Argentina/INEI109186/2021 | EPI_ISL_5158707 | 2021-08-05 | Servicio Virosis Respiratorias-Departamento Virolog√≠a-INEI | Instituto Nacional Enfermedades Infecciosas C.G.Malbran | Baumeister E., Avaro M., Benedetti E., Russo M., Dattero ME, Pontoriero A., Cisterna D., Molina V., Perandones C., Tuduri E., De Belder D., Sanchez Loria J., Haim MS., Lorenzo F., Poklepovich T., Campos J. |
| hCoV-19/Argentina/INEI109772/2021 | EPI_ISL_5159054 | 2021-08-13 | Servicio Virosis Respiratorias-Departamento Virolog√≠a-INEI | Instituto Nacional Enfermedades Infecciosas C.G.Malbran | Baumeister E., Avaro M., Benedetti E., Russo M., Dattero ME, Pontoriero A., Cisterna D., Molina V., Perandones C., Tuduri E., De Belder D., Sanchez Loria J., Haim MS., Lorenzo F., Poklepovich T., Campos J. |
| hCoV-19/Argentina/INEI110028/2021 | EPI_ISL_5159140 | 2021-08-24 | Servicio Virosis Respiratorias-Departamento Virolog√≠a-INEI | Instituto Nacional Enfermedades Infecciosas C.G.Malbran | Baumeister E., Avaro M., Benedetti E., Russo M., Dattero ME, Pontoriero A., Cisterna D., Molina V., Perandones C., Tuduri E., De Belder D., Sanchez Loria J., Haim MS., Lorenzo F., Poklepovich T., Campos J. |
| hCoV-19/Argentina/INEI110221/2021 | EPI_ISL_5159152 | 2021-08-29 | Servicio Virosis Respiratorias-Departamento Virolog√≠a-INEI | Instituto Nacional Enfermedades Infecciosas C.G.Malbran | Baumeister E., Avaro M., Benedetti E., Russo M., Dattero ME, Pontoriero A., Cisterna D., Molina V., Perandones C., Tuduri E., De Belder D., Sanchez Loria J., Haim MS., Lorenzo F., Poklepovich T., Campos J. |
| hCoV-19/USA/FL-CDC-ASC210339937/2021 | EPI_ISL_6065720 | 2021-10-08 | Aegis Sciences Corporation | Centers for Disease Control and Prevention Division of Viral Diseases, Pathogen Discovery | Dakota Howard,Dhwani Batra,Peter Cook,Jason Caravas,Benjamin Rambo-Martin,Scott Sammons,Yvette Unoarumhi,Matthew Schmerer,Kristine Lacek,Tymeckia Kendall,Victoria Caban Figueroa,Shatavia Morrison,Christopher Gulvick,Erisa Sula,Cyndi Clark,Patrick Campbell |
| hCoV-19/Chile/VS-155708/2021 | EPI_ISL_5010725 | 2021-09-20 | Genetica Molecular and Subdepartamento de Virologia ISP Chile | Instituto de Salud Publica de Chile | Karen Orostica, Constanza Campano, Barbara Parra, Loredana Arata, Gisselle Barra, Patricia Bustos, Rodrigo Fasce, Javier Tognarelli, Andres Castillo, Soledad Ulloa, Jorge Fernandez |
| hCoV-19/India/GJ-INSACOG-GBRC2932/2021 | EPI_ISL_6931583 | 2021-11-07 | Sanjeevani Metropolis, Rajkot | Gujarat Biotechnology Research Centre | Nitin Shukla,Ramesh Pandit,Nitin Savaliya,Priyank Chavda,Sonal Sharma ,Janvi Raval,Apurvasinh Puvar,Zarna Patel,Dinesh Kumar,Bhadreshsinh Gohil,Nimesh Patel,Shafquat Jahan,Chaitanya Joshi,Madhvi Joshi |

| | | | | | |
|---|---|---|---|---|---|
| hCoV-19/India/GJ-INSACOG-GBRC2939/2021 | EPI_ISL_6931584 | 2021-10-16 | Mangalam Invitro Laboratory, Rajkot | Gujarat Biotechnology Research Centre | Zarna Patel,Dinesh Kumar,Bhadreshsinh Gohil,Nimesh Patel,Nitin Shukla,Ramesh Pandit,Nitin Savaliya,Priyank Chavda,Sonal Sharma ,Janvi Raval,Apurvasinh Puvar,Rahul Thumbar,Chaitanya Joshi,Madhvi Joshi |
| hCoV-19/TrinidadandTobago/103411/2021 | EPI_ISL_6932149 | 2021-11-01 | The Caribbean Public Health Agency | Carrington Lab, Department of Preclinical Sciences, Faculty of Medical Sciences, The University of the West Indies, St Augustine Campus | Nikita S. D. Sahadeo, Arianne Brown-Jordan, Sarah Hill, Vernie Ramkissoon, Anushka Ramjag, Soren Nicholls, Roshan Parasram, Avery Hinds, Risha Singh, SueMin Nathaniel, Nuno Faria, Oliver Pybus, Christopher Oura, Gabriel Escobar, Christine V. F. Carrington |
| hCoV-19/Rwanda/CV2197/2020 | EPI_ISL_4768101 | 2020-09-10 | Rwanda¬†National Joint Taskforce COVID-34 | Africa Centre for Excellence for Genomics of Infectious Diseases (ACEGID), Redeemer‚Äôs University | Enatha Mukantwari, Olawoye, I.B., Dieudonne Mutangana, Oluniyi, P.E., Umuringa Jeanne d'Arc, Kayode, A.T., Esperance Umumararungu, Uwanibe, J.N., Sindayiheba Reuben, Eromon, P.E., Arlene Uwituze, Oguzie, J.U., Murebwayire Clarisse, Ajogbasile, F.V., Izuwa |
| hCoV-19/India/MH-RFIP03983/2021 | EPI_ISL_6937564 | 2021-05-10 | Genepath, Pune | RF-IISER Pune | Krishanpal Karmodiya, Aurnab Ghose, Joy Merwin Monteiro, Shatakshi Ranade, Nikhil Phadke |
| hCoV-19/India/MH-RFIP03987/2021 | EPI_ISL_6937650 | 2021-05-10 | Genepath, Pune | RF-IISER Pune | Krishanpal Karmodiya, Aurnab Ghose, Joy Merwin Monteiro, Shatakshi Ranade, Nikhil Phadke |
| hCoV-19/India/MH-RFIP04073/2021 | EPI_ISL_6937660 | 2021-06-09 | BJMC,Pune | RF-IISER Pune | Krishanpal Karmodiya, Aurnab Ghose, Joy Merwin Monteiro, Suvarna Joshi, Rajesh Karyakarte |
| hCoV-19/India/MH-RFIP04164/2021 | EPI_ISL_6937737 | 2021-05-10 | BJMC,Pune | RF-IISER Pune | Krishanpal Karmodiya, Aurnab Ghose, Joy Merwin Monteiro, Suvarna Joshi, Rajesh Karyakarte |
| hCoV-19/Ghana/TRA-2232/2021 | EPI_ISL_6939024 | 2021-11-19 | National Influenza Centre | National Influenza Centre | William K. Ampofo, Michael Marks, Ivy A. Asante, Sharon Hsu, Benjamiin B. Lindsey, Benjamin H. Foulkes, Mildred Adusei-Poku, Bright Adu, Linda Boatemaa, Lorreta Kwasah, Quaneeta Mohktar, Joyce Appiah-Kubi, Hilda Opoku Frempong, Keren Okyerebea Attiku, Bl |
| hCoV-19/Ghana/TRA-2302/2021 | EPI_ISL_6939030 | 2021-11-25 | National Influenza Centre | National Influenza Centre | William K. Ampofo, Michael Marks, Ivy A. Asante, Sharon Hsu, Benjamiin B. Lindsey, Benjamin H. Foulkes, Mildred Adusei-Poku, Bright Adu, Linda Boatemaa, Lorreta Kwasah, Quaneeta Mohktar, Joyce Appiah-Kubi, Hilda Opoku Frempong, Keren Okyerebea Attiku, Bl |
| hCoV-19/Ghana/TRA-2294/2021 | EPI_ISL_6939039 | 2021-11-25 | National Influenza Centre | National Influenza Centre | William K. Ampofo, Michael Marks, Ivy A. Asante, Sharon Hsu, Benjamiin B. Lindsey, Benjamin H. Foulkes, Mildred Adusei-Poku, Bright Adu, Linda Boatemaa, Lorreta Kwasah, Quaneeta Mohktar, Joyce Appiah-Kubi, Hilda Opoku Frempong, Keren Okyerebea Attiku, Bl |
| hCoV-19/Ghana/TRA-2310/2021 | EPI_ISL_6939042 | 2021-11-25 | National Influenza Centre | National Influenza Centre | William K. Ampofo, Michael Marks, Ivy A. Asante, Sharon Hsu, Benjamiin B. Lindsey, Benjamin H. Foulkes, Mildred Adusei-Poku, Bright Adu, Linda Boatemaa, Lorreta Kwasah, Quaneeta Mohktar, Joyce Appiah-Kubi, Hilda Opoku Frempong, Keren Okyerebea Attiku, Bl |

| | | | | | |
|---|---|---|---|---|---|
| hCoV-19/Ghana/TRA-2280/2021 | EPI_ISL_6939053 | 2021-11-24 | National Influenza Centre | National Influenza Centre | William K. Ampofo, Michael Marks, Ivy A. Asante, Sharon Hsu, Benjamiin B. Lindsey, Benjamin H. Foulkes, Mildred Adusei-Poku, Bright Adu, Linda Boatemaa, Lorreta Kwasah, Quaneeta Mohktar, Joyce Appiah-Kubi, Hilda Opoku Frempong, Keren Okyerebea Attiku, Bl |
| hCoV-19/Ghana/TRA-2296/2021 | EPI_ISL_6939057 | 2021-11-25 | National Influenza Centre | National Influenza Centre | William K. Ampofo, Michael Marks, Ivy A. Asante, Sharon Hsu, Benjamiin B. Lindsey, Benjamin H. Foulkes, Mildred Adusei-Poku, Bright Adu, Linda Boatemaa, Lorreta Kwasah, Quaneeta Mohktar, Joyce Appiah-Kubi, Hilda Opoku Frempong, Keren Okyerebea Attiku, Bl |
| hCoV-19/Kenya/SS3875/2021 | EPI_ISL_6939093 | 2021-10-26 | The Nairobi Hospital | KEMRI-Wellcome Trust Research Programme,Kilifi | Githinji G.,Ochola I., Agoti C.,Mohamed K.S., de Laurent Z., Mburu M.W., Morobe J., Ndwiga L. Makori T., Ongera E., Lambisia A. Nokes D J., Kariuki P., Yala D.A., Muoki M. |
| hCoV-19/Kenya/SS4004/2021 | EPI_ISL_6940176 | 2021-10-07 | KEMRI-CMR | KEMRI-Wellcome Trust Research Programme,Kilifi | Githinji G.,Ochola I, Agoti C.,Mohamed K.S.,de Laurent Z.,Mburu M.W.,Morobe J.,Lambisia A.,Makori T.,Ongera E.,Tigoi C., Muema A., D.J.Nokes |
| hCoV-19/Kenya/SS4014/2021 | EPI_ISL_6940182 | 2021-09-15 | KEMRI-CMR | KEMRI-Wellcome Trust Research Programme,Kilifi | Githinji G.,Ochola I, Agoti C.,Mohamed K.S.,de Laurent Z.,Mburu M.W.,Morobe J.,Lambisia A.,Makori T.,Ongera E.,Tigoi C., Muema A., D.J.Nokes |
| hCoV-19/USA/NJ-CDC-FG-158837/2021 | EPI_ISL_6071634 | 2021-10-22 | Fulgent Genetics | Centers for Disease Control and Prevention Division of Viral Diseases, Pathogen Discovery | Dakota Howard,Dhwani Batra,Peter Cook,Jason Caravas,Benjamin Rambo-Martin,Scott Sammons,Yvette Unoarumhi,Matthew Schmerer,Kristine Lacek,Tymeckia Kendall,Victoria Caban Figueroa,Shatavia Morrison,Christopher Gulvick,Erisa Sula,Harry Gao,Mickey Li,John Gao |
| hCoV-19/Ghana/TRA-2181/2021 | EPI_ISL_6944046 | 2021-10-24 | National Influenza Centre | National Influenza Centre | William K. Ampofo, Michael Marks, Ivy A. Asante, Sharon Hsu, Benjamiin B. Lindsey, Benjamin H. Foulkes, Mildred Adusei-Poku, Bright Adu, Linda Boatemaa, Lorreta Kwasah, Quaneeta Mohktar, Joyce Appiah-Kubi, Hilda Opoku Frempong, Keren Okyerebea Attiku, Bl |
| hCoV-19/Ghana/TRA-2201/2021 | EPI_ISL_6944059 | 2021-11-01 | National Influenza Centre | National Influenza Centre | William K. Ampofo, Michael Marks, Ivy A. Asante, Sharon Hsu, Benjamiin B. Lindsey, Benjamin H. Foulkes, Mildred Adusei-Poku, Bright Adu, Linda Boatemaa, Lorreta Kwasah, Quaneeta Mohktar, Joyce Appiah-Kubi, Hilda Opoku Frempong, Keren Okyerebea Attiku, Bl |
| hCoV-19/Colombia/PUT-CGBM-S09112021-124/2021 | EPI_ISL_6945586 | 2021-10-11 | LDSP PUTUMAYO | Centro de Gen√©tica y Biolog√≠a Molecular - Universidad del Magdalena | Maria Teresa Mojica-Ortiz, Daniel Bautista, Andrea M. Ram√≠rez Hernandez, Edison Lea-Ch, Angel Oviedo Marquez, Lyda R. Castro |
| hCoV-19/Colombia/SAN-CGBM-S09112021-107/2021 | EPI_ISL_6945591 | 2021-09-07 | NEUROTRAUMA CENTER IPS | Centro de Gen√©tica y Biolog√≠a Molecular - Universidad del Magdalena | Maria Teresa Mojica-Ortiz, Daniel Bautista, Andrea M. Ram√≠rez Hernandez, Edison Lea-Ch, Angel Oviedo Marquez, Lyda R. Castro |
| hCoV-19/Colombia/NAR-CGBM-S09112021-108/2021 | EPI_ISL_6945609 | 2021-09-12 | HOSPITAL UNIVERSITARIO DEPARTAMENTAL DE NARI√ëO | Centro de Gen√©tica y Biolog√≠a Molecular - Universidad del Magdalena | Maria Teresa Mojica-Ortiz, Daniel Bautista, Andrea M. Ram√≠rez Hernandez, Edison Lea-Ch, Angel Oviedo Marquez, Lyda R. Castro |

| | | | | | |
|---|---|---|---|---|---|
| hCoV-19/Brazil/BA-LACEN-BA864-291931750/2021 | EPI_ISL_6945676 | 2021-10-25 | Central Public Health Laboratory - LACEN - Bahia, Salvador, Brazil | Central Public Health Laboratory - LACEN -Bahia, Salvador, Brazil | Stephane Tosta, Luciana Oliveira, Vanessa Nardy, Gabriela Menezes,Marcela G√≥mez, Lenisa Dandara, Jaqueline Gomes, Vagner Fonseca,Marta Giovanetti,Luiz Alcantara, Felicidade Pereira, Arabela Leal |
| hCoV-19/Peru/LIM-INS-9776/2021 | EPI_ISL_6946005 | 2021-11-08 | Laboratorio de Referencia Nacional de Virus Respiratorios. Centro Nacional de Salud Publica. Instituto Nacional de Salud Peru. | Laboratorio de Referencia Nacional de Virus Respiratorios. Centro Nacional de Salud Publica. Instituto Nacional de Salud Peru. | Carlos Padilla Rojas, Veronica Hurtado Vela, Orson Mestanza Millones, Iris Silva Molina, Luis Barcena Flores, Alicia Nu√±ez Llanos, Steve Acedo Lazo, Wendy Lizarraga Olivares, Victor Jimenez Vasquez, Sara Gordillo Vilchez, Kelly Vanessa Izarra Rojas, Edwa |
| hCoV-19/USA/CA-CDPH-2000054703/2021 | EPI_ISL_6946231 | 2021-10-26 | CDPH VBL | California Department of Public Health | Emily Smith on behalf of CDPH-COVIDNet and UCLA Technology Center for Genomics & Bioinformatics |
| hCoV-19/USA/CA-CDPH-2000054873/2021 | EPI_ISL_6946380 | 2021-10-26 | CDPH VBL | California Department of Public Health | Emily Smith on behalf of CDPH-COVIDNet and UCLA Technology Center for Genomics & Bioinformatics |
| hCoV-19/USA/CA-SC-260563/2021 | EPI_ISL_6074652 | 2021-10-20 | O'Connor Hospital | Santa Clara County Public Health Laboratory | Santa Clara County Public Health Department |
| hCoV-19/USA/CA-CDPH-3000269161/2021 | EPI_ISL_6947318 | 2021-11-07 | CDPH VBL | California Department of Public Health | Emily Smith on behalf of CDPH-COVIDNet |
| hCoV-19/Congo/FCRM-116-12738/2021 | EPI_ISL_6077269 | 2021-10-25 | Fondation Congolaise pour la recherche medicale (FCRM), Francine Ntoumi | Fondation Congolaise pour la Recherche M√©dicale | Mfoutou Mapanguy Claujens Chastel; Batchi-Bouyou Armel Landry; Dr. Jean Claude Djontu; Dr. Abel Lissom; Prof. Dr. Thirumalaisamy P. Velavan; Prof. Francine Ntoumi |
| hCoV-19/Ghana/TRA-2314/2021 | EPI_ISL_6939035 | 2021-11-25 | National Influenza Centre | National Influenza Centre | William K. Ampofo, Michael Marks, Ivy A. Asante, Sharon Hsu, Benjamiin B. Lindsey, Benjamin H. Foulkes, Mildred Adusei-Poku, Bright Adu, Linda Boatemaa, Lorreta Kwasah, Quaneeta Mohktar, Joyce Appiah-Kubi, Hilda Opoku Frempong, Keren Okyerebea Attiku, Bl |
| hCoV-19/Ghana/TRA-2292/2021 | EPI_ISL_6939038 | 2021-11-24 | National Influenza Centre | National Influenza Centre | William K. Ampofo, Michael Marks, Ivy A. Asante, Sharon Hsu, Benjamiin B. Lindsey, Benjamin H. Foulkes, Mildred Adusei-Poku, Bright Adu, Linda Boatemaa, Lorreta Kwasah, Quaneeta Mohktar, Joyce Appiah-Kubi, Hilda Opoku Frempong, Keren Okyerebea Attiku, Bl |
| hCoV-19/England/MILK-2C1C870/2021 | EPI_ISL_7023726 | 2021-11-27 | Lighthouse Lab in Milton Keynes | Wellcome Sanger Institute for the COVID-19 Genomics UK (COG-UK) Consortium | The Lighthouse Lab in Milton Keynes and Alex Alderton, Roberto Amato, Jeffrey Barrett, Sonia Goncalves, Ewan Harrison, David K. Jackson, Ian Johnston, Dominic Kwiatkowski, Cordelia Langford, John Sillitoe on behalf of the Wellcome Sanger Institute COVID-1 |
| hCoV-19/England/MILK-2BF595D/2021 | EPI_ISL_7029353 | 2021-11-25 | Lighthouse Lab in Milton Keynes | Wellcome Sanger Institute for the COVID-19 Genomics UK (COG-UK) Consortium | The Lighthouse Lab in Milton Keynes and Alex Alderton, Roberto Amato, Jeffrey Barrett, Sonia Goncalves, Ewan Harrison, David K. Jackson, Ian Johnston, Dominic Kwiatkowski, Cordelia Langford, John Sillitoe on behalf of the Wellcome Sanger Institute COVID-1 |
| hCoV-19/Norway/29450/2021 | EPI_ISL_7040235 | 2021-11-27 | Oslo University Hospital, Department of Medical Microbiology | Norwegian Institute of Public Health, Department of Virology | Kathrine Stene-Johansen, Kamilla Heddeland Instefjord, Hilde Elshaug, Garcia Llorente Ignacio, Jon Br√•te, Engebretsen Serina Beate,Pedersen Benedikte Nevjen, Line Victoria Moen, Debech Nadia, Atiya R Ali,Marie Paulsen Madsen, Rasmus Riis Kopperud, Hilde |

| | | | | | |
|---|---|---|---|---|---|
| hCoV-19/India/KA-RF-NCBS-Bengaluru-RFNB-1100/2021 | EPI_ISL_6963509 | 2021-11-22 | Sri Jayadeva Institute of Cardiovascular Sciences and Research / Strand Life Sciences | National Centre for Biological Sciences, TIFR - Rockefeller Foundation | Rohan Pais, Vamsi Veeramachaneni, Chitra Pattabiraman, Manoj Kumar Jha, Priyanka Ananta Mulay, Anson Kunjumon George, Chandrasekhar Vadlamudi, Madhusudhan J, Aarati Karaba, Aparnaa Ramanathan, Yasodha Kannan, Pradeep B P, Saumitra Mardikar, Mudasir Nazaar |
| hCoV-19/England/MILK-2BFA017/2021 | EPI_ISL_7026536 | 2021-11-26 | Lighthouse Lab in Milton Keynes | Wellcome Sanger Institute for the COVID-19 Genomics UK (COG-UK) Consortium | The Lighthouse Lab in Milton Keynes and Alex Alderton, Roberto Amato, Jeffrey Barrett, Sonia Goncalves, Ewan Harrison, David K. Jackson, Ian Johnston, Dominic Kwiatkowski, Cordelia Langford, John Sillitoe on behalf of the Wellcome Sanger Institute COVID-1 |
| hCoV-19/Austria/KILM2164/2021 | EPI_ISL_6972689 | 2021-11-27 | Dept. of Laboratory Medicine | Dept. of Laboratory Medicine | Harald Esterbauer, Fabian Konig, Sabina Plumer, Victoria Six, Robert Strassl, Claudia Weber, Oswald Wagner |
| hCoV-19/Portugal/PT21721/2021 | EPI_ISL_6892661 | 2021-11-27 | Germano de Sousa | Instituto Nacional de Saude (INSA) | Borges et al |
| hCoV-19/Brazil/SP-HIAE-ID989/2021 | EPI_ISL_6901960 | 2021-11-25 | LATE - Laboratv≥rio de Tv©cnicas Especiais - Hospital Israelita Albert Einstein | LATE - Laboratv≥rio de Tv©cnicas Especiais - Hospital Israelita Albert Einstein | Deyvid Amgarten, Fernanda de Mello Malta, Raquel Riyuzo, Ana Paula Moreira Salles, Luiz Vicente Rizzo, Pedro Henrique Sebe Rodrigues, Alexandre Hideaki Takara, Anelisie da Silva Santos, Erick Gustavo Dorlass, Marcio Anunciacao Menezes, Jov£o Renato Rebell |
| hCoV-19/HongKong/VM21044713_WHP5047-S5/2021 | EPI_ISL_6716902 | 2021-11-15 | Hong Kong Department of Health | School of Public Health, The University of Hong Kong | Dominic N.C. Tsang, Haogao Gu, Leo L.M. Poon, Malik Peiris |
| hCoV-19/Canada/ON-NML-249359/2021 | EPI_ISL_6826713 | 2021-11-23 | Dynacare | National Microbiology Laboratory (NML) | Anna Majer, Shari Tyson, Grace Seo, Philip Mabon, Elsie Grudeski, Rhiannon Huzarewich, Russell Mandes, Anneliese Landgraff, Jennifer Tanner, Natalie Knox, Morag Graham, Gary Van Domselaar, Gordon Jolly, Nathalie Bastien, Yan Li, Timothy Booth, Darian Hole |
| hCoV-19/Australia/NSW-RPAH-1933/2021 | EPI_ISL_6814922 | 2021-11-27 | New South Wales Health Pathology Royal Prince Alfred Hospital | Microbiology RPAH | Foster, C.; Au, J.; Ruiz Silva, M.; Deveson, I.; Bull, R.; Van Hal, S.; Rawlinson, W. |
| hCoV-19/Israel/SMC-7014472/2021 | EPI_ISL_6080820 | 2021-08-23 | Shamir Medical Center (Asaf Harofe) | Shamir Medical Center (Asaf Harofe) | Nir Rainy,Chen Weiner,Reut Sorek Abramovich,Yevgeni Yegorov,Anna Vishnevsky,Patricia Benveniste-Lekovitz,Abu Hamad Ramzia,Adina Bar Chaim |
| hCoV-19/Chile/RM-USS-UNAB-AEA392/2021 | EPI_ISL_5195628 | 2021-09-21 | Universidad San Sebastian | Facultad de Ciencias de la Vida, UNAB | Claudio Meneses, Eduardo Castro, Waldo Diaz, Macarena Bastias, Ricardo Yusta, Susana Saez, Dayan Sanhueza, Sebastian Wolter, Daniel Leon, Gonzalo Campav±a, Paola Pidal, Vv≠ctor Monreal, Claudio Olmos, Ariel Orellana |
| hCoV-19/Chile/RM-USS-UNAB-ALT132/2021 | EPI_ISL_5195641 | 2021-10-01 | Universidad San Sebastian | Facultad de Ciencias de la Vida, UNAB | Claudio Meneses, Eduardo Castro, Waldo Diaz, Macarena Bastias, Ricardo Yusta, Susana Saez, Dayan Sanhueza, Sebastian Wolter, Daniel Leon, Gonzalo Campav±a, Paola Pidal, Vv≠ctor Monreal, Claudio Olmos, Ariel Orellana |
| hCoV-19/Israel/SMC-7014431/2021 | EPI_ISL_6080984 | 2021-08-24 | Shamir Medical Center (Asaf Harofe) | Shamir Medical Center (Asaf Harofe) | Nir Rainy,Chen Weiner,Reut Sorek Abramovich,Yevgeni Yegorov,Anna Vishnevsky,Patricia Benveniste-Lekovitz,Abu Hamad Ramzia,Adina Bar Chaim |

| | | | | | |
|---|---|---|---|---|---|
| hCoV-19/Israel/SMC-7014496/2021 | EPI_ISL_6081123 | 2021-08-24 | Shamir Medical Center (Asaf Harofe) | Shamir Medical Center (Asaf Harofe) | Nir Rainy,Chen Weiner,Reut Sorek Abramovich,Yevgeni Yegorov,Anna Vishnevsky,Patricia Benveniste-Lekovitz,Abu Hamad Ramzia,Adina Bar Chaim |
| hCoV-19/Peru/JUN-INS-6938/2021 | EPI_ISL_5199104 | 2021-09-06 | Laboratorio de Referencia Nacional de Virus Respiratorios. Centro Nacional de Salud Publica. Instituto Nacional de Salud Peru. | Laboratorio de Referencia Nacional de Virus Respiratorios. Centro Nacional de Salud Publica. Instituto Nacional de Salud Peru. | Carlos Padilla Rojas, Veronica Hurtado Vela, Orson Mestanza Millones, Iris Silva Molina, Luis Barcena Flores, Sandra Morales Ruiz, Steve Acedo Lazo, Wendy Lizarraga Olivares, Victor Jimenez Vasquez, Sara Gordillo Vilchez, Nieves Sevilla Casta√±eda, Prince |
| hCoV-19/Israel/SMC-7015017/2021 | EPI_ISL_6081642 | 2021-08-29 | Shamir Medical Center (Asaf Harofe) | Shamir Medical Center (Asaf Harofe) | Nir Rainy,Chen Weiner,Reut Sorek Abramovich,Yevgeni Yegorov,Anna Vishnevsky,Patricia Benveniste-Lekovitz,Abu Hamad Ramzia,Adina Bar Chaim |
| hCoV-19/Australia/SA0998/2021 | EPI_ISL_5200352 | 2021-10-10 | SA Pathology | SA Pathology | Caitlin Selway, Lex Leong, Chuan Kok Lim, Mark Turra, Ivan Bastian |
| hCoV-19/Colombia/DC-LSPSDS-VG-1064510/2021 | EPI_ISL_5202325 | 2021-07-31 | SYNLAB | LSPSDS | Johana Hernandez Toloza, Marcela Castano Rodriguez,Gabriela Delgado Murcia, Alejandro Gomez Lopez, Marcela Mercado |
| hCoV-19/Canada/AB-ABPHL-42868/2021 | EPI_ISL_6949582 | 2021-11-23 | Edmonton Provincial Lab | Alberta Precision Labs (APL) | Buss, E, Croxen M, Deo A, Dieu P, Gill K, Ferrato C, Khan F, Koleva P, Li V, Lloyd C, Lynch T, Ma R, Murphy S, Pabbaraju K, Shokoples S, Tipples G, Thayer J, Whitehouse M, Wong A, Yu C, Zelyas N |
| hCoV-19/Bonaire/BQ-RIVM-63011/2021 | EPI_ISL_5209092 | 2021-09-26 | Dutch COVID-19 response team | National Institute for Public Health and the Environment (RIVM) | Adam Meijer, Harry Vennema, Dirk Eggink, Jeroen Cremer, Sharon van den Brink, Bas van der Veer, AnneMarie van den Brandt, Lisa Wijsman, Kim Freriks, Ryanne Jaarsma, Eun√≠ce Then, Lynn Aarts, Sanne Bos, Stijn van Rossum, Florian Zwagemaker, Dennis Schmitz, |
| hCoV-19/Netherlands/NB-RIVM-63651/2021 | EPI_ISL_5209385 | 2021-09-27 | Dutch COVID-19 response team | National Institute for Public Health and the Environment (RIVM) | Adam Meijer, Harry Vennema, Dirk Eggink, Jeroen Cremer, Sharon van den Brink, Bas van der Veer, AnneMarie van den Brandt, Lisa Wijsman, Kim Freriks, Ryanne Jaarsma, Eun√≠ce Then, Lynn Aarts, Sanne Bos, Stijn van Rossum, Florian Zwagemaker, Dennis Schmitz, |
| hCoV-19/Israel/SMC-7015773/2021 | EPI_ISL_6084260 | 2021-08-30 | Shamir Medical Center (Asaf Harofe) | Shamir Medical Center (Asaf Harofe) | Nir Rainy,Chen Weiner,Reut Sorek Abramovich,Yevgeni Yegorov,Anna Vishnevsky,Patricia Benveniste-Lekovitz,Abu Hamad Ramzia,Adina Bar Chaim |
| hCoV-19/USA/WI-CDC-LC0167440/2021 | EPI_ISL_5214045 | 2021-07-30 | Laboratory Corporation of America | Centers for Disease Control and Prevention Division of Viral Diseases, Pathogen Discovery | Dakota Howard,Dhwani Batra,Peter Cook,Jason Caravas,Benjamin Rambo-Martin,Scott Sammons,Yvette Unoarumhi,Matthew Schmerer,Kristine Lacek,Tymeckia Kendall,Victoria Caban Figueroa,Shatavia Morrison,Christopher Gulvick,Minoo Agarwal,Eyad Almasri,Debbie Boles |
| hCoV-19/Slovenia/08-134502-MB/2020 | EPI_ISL_6950152 | 2020-12-04 | National Laboratory for Health, Environment and Food, OMM, Maribor | [illegible] Health, Environment and Food) / CISLD (Clinical Institute of Special Laboratory Diagnostics), University Children's Hospital, University Medical Center Ljubljana | Sandra Janezic, Aleksander Mahnic, Maja Rupnik, Tja≈°a ≈Ωohar ƒåretnik, Alenka ≈†torman, Nika Gobec, Aleksander Kocuvan, Kaja Tominc, Ma≈°a Jarƒçiƒç, Mojca Cimerman, Nika Volmajer, Andrej Golle / Jernej Kovaƒç, Barbara Jenko Bizjan, Tine Tesovnik, Robert |

| | | | | | |
|---|---|---|---|---|---|
| hCoV-19/Israel/SMC-7016109/2021 | EPI_ISL_6086793 | 2021-09-04 | Shamir Medical Center (Asaf Harofe) | Shamir Medical Center (Asaf Harofe) | Nir Rainy,Chen Weiner,Reut Sorek Abramovich,Yevgeni Yegorov,Anna Vishnevsky,Patricia Benveniste-Lekovitz,Abu Hamad Ramzia,Adina Bar Chaim |
| hCoV-19/USA/KY-CDC-LC0347859/2021 | EPI_ISL_6086943 | 2021-10-21 | Laboratory Corporation of America | Centers for Disease Control and Prevention Division of Viral Diseases, Pathogen Discovery | Dakota Howard,Dhwani Batra,Peter Cook,Jason Caravas,Benjamin Rambo-Martin,Scott Sammons,Yvette Unoarumhi,Matthew Schmerer,Kristine Lacek,Tymeckia Kendall,Victoria Caban Figueroa,Shatavia Morrison,Christopher Gulvick,Minoo Agarwal,Eyad Almasri,Debbie Boles |
| hCoV-19/Canada/BC-BCCDC-208640/2021 | EPI_ISL_5227543 | 2021-09-19 | British Columbia Centre For Disease Control | BCCDC Public Health Laboratory | Prystajecky Natalie, Linda Hoang, Dan Fornika, John Tyson, Shannon Russell, Kim Macdonald, Kimia Kamelian, Ana Pacagnella, Corrinne Ng, Loretta Janz, Robert Azana, Mel Krajden |
| hCoV-19/Ghana/WACCBIP-GS1200/2021 | EPI_ISL_6088166 | 2021-07-12 | WACCBIP, University of Ghana | WACCBIP, University of Ghana | Collins M. Morang'a, Joyce M. Ngoi, Dominic S. Y. Amuzu, Vincent Appiah, Evelyn B. Quansah, Philip M. Soglo, Violette V. M'cormack, Samirah Saiid, Bright K. Yemi, Israel Osei-Wusu, Deborah N. A. Mettle, Aaron A. Manu, Paul Owusu-Oduro, Frederick Tei-Maya |
| hCoV-19/USA/MS-CDC-ASC210118190/2021 | EPI_ISL_5232196 | 2021-08-16 | Aegis Sciences Corporation | Centers for Disease Control and Prevention Division of Viral Diseases, Pathogen Discovery | Dakota Howard,Dhwani Batra,Peter Cook,Jason Caravas,Benjamin Rambo-Martin,Scott Sammons,Yvette Unoarumhi,Matthew Schmerer,Kristine Lacek,Tymeckia Kendall,Victoria Caban Figueroa,Shatavia Morrison,Christopher Gulvick,Erisa Sula,Cyndi Clark,Patrick Campbell |
| hCoV-19/USA/CA-CDC-FG-109833/2021 | EPI_ISL_5232450 | 2021-08-30 | Fulgent Genetics | Centers for Disease Control and Prevention Division of Viral Diseases, Pathogen Discovery | Dakota Howard,Dhwani Batra,Peter Cook,Jason Caravas,Benjamin Rambo-Martin,Scott Sammons,Yvette Unoarumhi,Matthew Schmerer,Kristine Lacek,Tymeckia Kendall,Victoria Caban Figueroa,Shatavia Morrison,Christopher Gulvick,Erisa Sula,Harry Gao,Mickey Li,John Gao |
| hCoV-19/USA/MN-CDC-IBX937058360097/2021 | EPI_ISL_5232788 | 2021-08-27 | Infinity Biologix | Centers for Disease Control and Prevention Division of Viral Diseases, Pathogen Discovery | Dakota Howard,Dhwani Batra,Peter Cook,Jason Caravas,Benjamin Rambo-Martin,Scott Sammons,Yvette Unoarumhi,Matthew Schmerer,Kristine Lacek,Tymeckia Kendall,Victoria Caban Figueroa,Shatavia Morrison,Christopher Gulvick,Erisa Sula,Christian Bixby,Yihe Wang,Jo |
| hCoV-19/Austria/CeMM18678/2021 | EPI_ISL_6092559 | 2021-10-11 | Austrian Agency for Health and Food Safety (AGES) | Bergthaler laboratory, CeMM Research Center for Molecular Medicine of the Austrian Academy of Sciences | Lukas Endler, Anna Schedl, Fabian Amman, Petr Triska, Matthew Thornton, Thomas Penz, Benedikt Agerer, Michelle Chan, Michael Schuster, Bekir Erguner, Jan Laine, Martin Senekowitsch, Christoph Bock, Andreas Bergthaler |
| hCoV-19/Kenya/ILRI_COVC24590/2021 | EPI_ISL_6093635 | 2021-09-24 | International Livestock Research Institute | International Livestock Research Institute | Samuel O. Oyola, Daniel Ouso, Gilbert Kibet, Shebbar Osiany, Edward Kiritu, Paul Dobi, Collins Muli, Patrick Amoth, Vishvanath Nene, Sonal P. Henson, Edward O. Abworo |

| | | | | | |
|---|---|---|---|---|---|
| hCoV-19/Kenya/ILRI_COVM00516/2021 | EPI_ISL_6093783 | 2021-09-08 | International Livestock Research Institute | International Livestock Research Institute | Samuel O. Oyola, Daniel Ouso, Gilbert Kibet, Shebbar Osiany, Edward Kiritu, Paul Dobi, Collins Muli, Patrick Amoth, Vishvanath Nene, Sonal P. Henson, Edward O. Abworo |
| hCoV-19/Kenya/ILRI_COVC24655/2021 | EPI_ISL_6094812 | 2021-10-01 | International Livestock Research Institute | International Livestock Research Institute | Samuel O. Oyola, Daniel Ouso, Gilbert Kibet, Shebbar Osiany, Edward Kiritu, Paul Dobi, Collins Muli, Patrick Amoth, Vishvanath Nene, Sonal P. Henson, Edward O. Abworo |
| hCoV-19/Kenya/ILRI_COVC18312/2020 | EPI_ISL_6095490 | 2020-11-04 | International Livestock Research Institute | International Livestock Research Institute | Samuel O. Oyola, Daniel Ouso, Gilbert Kibet, Shebbar Osiany, Edward Kiritu, Paul Dobi, Collins Muli, Patrick Amoth, Vishvanath Nene, Sonal P. Henson, Edward O. Abworo |
| hCoV-19/Kenya/ILRI_COVC24457/2021 | EPI_ISL_6095836 | 2021-09-02 | International Livestock Research Institute | International Livestock Research Institute | Samuel O. Oyola, Daniel Ouso, Gilbert Kibet, Shebbar Osiany, Edward Kiritu, Paul Dobi, Collins Muli, Patrick Amoth, Vishvanath Nene, Sonal P. Henson, Edward O. Abworo |
| hCoV-19/Kenya/ILRI_COVM01219/2021 | EPI_ISL_6095994 | 2021-04-08 | International Livestock Research Institute | International Livestock Research Institute | Samuel O. Oyola, Daniel Ouso, Gilbert Kibet, Shebbar Osiany, Edward Kiritu, Paul Dobi, Collins Muli, Patrick Amoth, Vishvanath Nene, Sonal P. Henson, Edward O. Abworo |
| hCoV-19/Kenya/ILRI_COVC24707/2021 | EPI_ISL_6096309 | 2021-10-06 | International Livestock Research Institute | International Livestock Research Institute | Samuel O. Oyola, Daniel Ouso, Gilbert Kibet, Shebbar Osiany, Edward Kiritu, Paul Dobi, Collins Muli, Patrick Amoth, Vishvanath Nene, Sonal P. Henson, Edward O. Abworo |
| hCoV-19/Kenya/ILRI_COVM01347/2021 | EPI_ISL_6096435 | 2021-01-09 | International Livestock Research Institute | International Livestock Research Institute | Samuel O. Oyola, Daniel Ouso, Gilbert Kibet, Shebbar Osiany, Edward Kiritu, Paul Dobi, Collins Muli, Patrick Amoth, Vishvanath Nene, Sonal P. Henson, Edward O. Abworo |
| hCoV-19/Kenya/ILRI_COVM01388/2021 | EPI_ISL_6096560 | 2021-08-18 | International Livestock Research Institute | International Livestock Research Institute | Samuel O. Oyola, Daniel Ouso, Gilbert Kibet, Shebbar Osiany, Edward Kiritu, Paul Dobi, Collins Muli, Patrick Amoth, Vishvanath Nene, Sonal P. Henson, Edward O. Abworo |
| hCoV-19/Denmark/DCGC-203661/2021 | EPI_ISL_6099459 | 2021-11-02 | Department of Bacteria, Parasites and Fungi, Statens Serum Institut, Copenhagen, Denmark | Statens Serum Institut Bioinformatics and Microbial Genomics | Danish Covid-19 Genome Consortium |
| hCoV-19/India/TN-SEQ_6000_S10_R1_001/2021 | EPI_ISL_6101066 | 2021-09-09 | Dr S Raju, Director of Public Health and Preventive Medicine | inStem NCBS ‚Äì INSACOG | Uma Ramakrishnan, Dasaradhi Palakodeti, Aswin SaiNarain |
| hCoV-19/Australia/NSW-ICPMR-14638/2021 | EPI_ISL_6956011 | 2021-11-30 | Pathology West - NSW Health Pathology | Clinical Pathology and Medical Research; Westmead Hospital; University of Sydney | Arnott A., Draper J., Gall M., Martinez E., Rockett R., Sintchenko V., on behalf of ICPMR |
| hCoV-19/HongKong/CM21000723/2021 | EPI_ISL_6958280 | 2021-11-27 | North Lantau Hospital | Hong Kong Department of Health | Alan K.L. Tsang, Peter C.W. Yip, Ken H.L. Ng, Edman T.K. Lam, Rickjason C.W. Chan |

| | | | | | |
|---|---|---|---|---|---|
| hCoV-19/SouthKorea/KDCA18124/2021 | EPI_ISL_6959926 | 2021-11-24 | Division of Emerging Infectious Diseases, Bureau of Infectious Diseases Diagnosis Control, Korea Disease Control and Prevention Agency | Diseases, Bureau of Infectious Diseases Diagnosis Control, Korea Disease Control and Prevention Agency | Ae Kyung Park, Il-Hwan Kim, Jeong-Ah Kim, Chae Young Lee, Hyuck Jin Lee, Eun-Jin Kim |
| hCoV-19/Nepal/CDBT-TU-QC5041/2021 | EPI_ISL_6963504 | 2021-09-05 | National Public Health Laboratory | Central Department Of Biotechnology, Tribhuvan University | Krishna Das Manandhar |
| hCoV-19/France/IDF-CERBAHC-12742321/2021 | EPI_ISL_6967758 | 2021-11-26 | CERBALLIANCE PARIS ET IDF EST | CERBA HealthCare | Judith Zerah, Johanna Roux, Laura Verdurme, Sabine Trombert, St√©phanie Haim-Boukobza, B√©n√©dicte Roquebert |
| hCoV-19/India/KA-NIMH-SEQ-3220/2021 | EPI_ISL_6112823 | 2021-09-05 | Mysuru Medical College and Research Institute | INSACOG-KA , NIMHANS | Ellango Ramasamy, Sony Sharma, Mahesh Kumar.C.S, Ananthapadmanabha Kotambail, Chetan G K, Gautham Arunachal Udupi, Anita S Desai, V Ravi |
| hCoV-19/SouthAfrica/UFS-VIRO-NGS-MSQ290921BC15/2021 | EPI_ISL_5249939 | 2021-09-23 | NHLS Universitas Academic | UFS Virology | PA Bester, MM Nyaga, P Nthiga, MT Mogotsi, Emmanuel Ogunbayo, D Goedhals, T de Oliveira |
| hCoV-19/USA/MA-CDCBI-CRSP_AL2DEVF6TPQBNIDM/2021 | EPI_ISL_5250997 | 2021-10-10 | Broad Institute Clinical Research Sequencing Platform | Infectious Disease Program, Broad Institute of Harvard and MIT | Siddle,K.J., Adams,G., Pearlman,L., Gladden-Young,A., Vicente,G., Blumenstiel,B., DeFelice,M., Lee,M., McGovern,S., Lagerborg,K., Rudy,M., DeRuff,K., Carter,A., Normandin,E., Bauer,M., Reilly,S., Tomkins-Tinch,C., Loreth,C., Chaluvadi,S., Meldrim,J., Gran |
| hCoV-19/Paraguay/266002/2021 | EPI_ISL_6971635 | 2021-10-19 | Instituto de Medicina Tropical | Laboratorio Central de Salud P√∫blica | Sandra Gonzalez, Andrea G√≥mez de la Fuente, Cynthia Vazquez, Julio Barrios, Cesar Cantero, Tania Alfonzo, Shirley Villalba, Juan Torales, Maria Liz Gamarra |
| hCoV-19/USA/CA-4014976721/2021 | EPI_ISL_5978050 | 2021-02-25 | UC Davis Genome Center | UC Davis Genome Center | UC Davis |
| hCoV-19/Scotland/EDB29553/2021 | EPI_ISL_6972088 | 2021-10-24 | Virology Department, Royal Infirmary of Edinburgh, NHS Lothian / School of Biological Sciences, University of Edinburgh | COVID-19 Genomics UK (COG-UK) Consortium | McHugh M, Dewar R, Cotton S, Gallagher A, Maloney D, Fernandez G, O,ÄôToole √Å, Scher E, Hill V, McCrone JT, Colquhoun R, Yu X, Jackson B, Rambaut A, Templeton K |
| hCoV-19/Cambodia/826598/2021 | EPI_ISL_5260557 | 2021-10-08 | Virology Unit, Institut Pasteur du Cambodge | Virology Unit, Institut Pasteur du Cambodge | Leakhena Pum, Sophoannadedh Rath, Jurre Y Siegers, Cecile Troupin, Ly Sovann, Kraing Sidonn, Yi Sengdoeurn, Chin Savuth, Chau Darapheak, Veasna Duong, Erik A Karlsson |
| hCoV-19/Reunion/PIMIT_23035/2021 | EPI_ISL_6980876 | 2021-11-29 | Cerballiance, Reunion | UMR PIMIT | David A Wilkinson; Patrick Mavingui |
| hCoV-19/England/PHEP-YYNYSXK/2021 | EPI_ISL_6988917 | 2021-11-11 | Respiratory Virus Unit, Microbiology Services Colindale, Public Health England | COVID-19 Genomics UK (COG-UK) Consortium | PHE Covid Sequencing Team |
| hCoV-19/CzechRepublic/UKDD-ZUUL-D540/2021 | EPI_ISL_6121492 | 2021-10-25 | Zdravotn√≠ √∫stav √öst√≠ nad Labem | Institute of Medical Microbiology and Virology, University Hospital Carl Gustav Carus, TU Dresden | Marlena Stadtm√ºller, Alexa Laubner, Fabian Rost, Sylvia Klemroth, Grit Mehnert, Montserrat Palau de Miquel, Susanne Reinhardt, Anett Zabzinski, Sylke Winkler, Alexander Dalpke, Romana Mike≈°ov√°, Jitka Poho≈ôsk√°, Ivana Stiborov√°, Eva Patrasov√°, Lenka |
| hCoV-19/USA/NC-ECU-CORVASEQ-109414238/2021 | EPI_ISL_5266147 | 2021-08-24 | Molecular Biology Laboratory, Vidant Medical Center | Brody Integrative Genomics Core, East Carolina University | Weihua Huang, Changhong Yin, Kimberly P. Briley, Heather Duncan, John T. Fallon |
| hCoV-19/Italy/SIC_CQRC_2728951/2021 | EPI_ISL_6122020 | 2021-01-11 | Laboratorio CQRC | CQRC_QUALITY CONTROL CHEMICAL BIOLOGICAL RISK_AOOR Villa Sofia Cervello Palermo | Di Gaudio F., Contino F., Brunacci G. |

| | | | | | |
|---|---|---|---|---|---|
| hCoV-19/France/BFC-HMN-21112020153/2021 | EPI_ISL_6124386 | 2021-10-22 | Laboratoire MAYMAT | [illegible] Mondor University Hospital, Assistance Publique H√¥pitaux de Paris, Universit√© Paris-Est Cr√©teil, INSERM U955 | Christophe Rodriguez, Slim Fourati, Vanessa Demontant, Guillaume Gricourt, Melissa N'Debi, Alexandre Soulier, Elisabeth Trawinski, Jean-Michel Pawlotsky |
| hCoV-19/Qatar/QA.QU..20.7.D7/2021 | EPI_ISL_6125608 | 2021-09-09 | Ministry of Public Health / Hamad Medical Corporation | Biomedical Research Center (BRC), Qatar University / Qatar Genome Project (QGP) | BRC:¬†Fatiha M. Benslimane, Heba A. Al-Khatib, Oal Al-Jamal, Dana Al-Batesh, Hadi M. Yassine, Asmaa A. Al-Thani. MOPH and HMC:¬†Abdullatif Al-Khal, Muna A. S. Al-Maslamani, Mashael A. Al-Bader, Hamda Alromaihi, Roberto Bertollini, Peter V. Coyle, Einas A. |
| hCoV-19/Qatar/QA.QU..20.7.C3/2021 | EPI_ISL_6125612 | 2021-09-15 | Ministry of Public Health / Hamad Medical Corporation | Biomedical Research Center (BRC), Qatar University / Qatar Genome Project (QGP) | BRC:¬†Fatiha M. Benslimane, Heba A. Al-Khatib, Oal Al-Jamal, Dana Al-Batesh, Hadi M. Yassine, Asmaa A. Al-Thani. MOPH and HMC:¬†Abdullatif Al-Khal, Muna A. S. Al-Maslamani, Mashael A. Al-Bader, Hamda Alromaihi, Roberto Bertollini, Peter V. Coyle, Einas A. |
| hCoV-19/Ecuador/NIC-INSPI-348343/2021 | EPI_ISL_6128461 | 2021-08-03 | INSPI-CRN DE INFLUENZA Y OTROS VIRUS RESPIRATORIOS | INSPI-CRN DE INFLUENZA Y OTROS VIRUS RESPIRATORIOS | Alfredo Bruno, Domenica de Mora, Maritza Olmedo, Michelle P√°ez, Jimmy Garc√©s, Maria Angelica Becerra, Marcela Mejia, Daniel Ramos, Fernando Llerena, Lizbeth Pati√±o, Ruben Armas |
| hCoV-19/Ecuador/NIC-INSPI-349638/2021 | EPI_ISL_6129897 | 2021-08-27 | INSPI-CRN DE INFLUENZA Y OTROS VIRUS RESPIRATORIOS | INSPI-CRN DE INFLUENZA Y OTROS VIRUS RESPIRATORIOS | Alfredo Bruno, Domenica de Mora, Maritza Olmedo, Michelle P√°ez, Jimmy Garc√©s, Maria Angelica Becerra, Marcela Mejia, Daniel Ramos, Fernando Llerena, Lizbeth Pati√±o, Ruben Armas |
| hCoV-19/Malaysia/A107/2021 | EPI_ISL_6132043 | 2021-10-04 | HRPB | Faculty of Medicine, University of Malaya; University of Malaya Medical Centre | Omar Khalilur Rahman, Jolene Yin Ling FU, I-Ching SAM, Yoke Fun Chan |
| hCoV-19/Qatar/QA.QU..18.25.C8/2021 | EPI_ISL_6132313 | 2021-08-14 | Ministry of Public Health / Hamad Medical Corporation | Biomedical Research Center (BRC), Qatar University / Qatar Genome Project (QGP) | BRC:¬†Fatiha M. Benslimane, Heba A. Al-Khatib, Oal Al-Jamal, Dana Al-Batesh, Hadi M. Yassine, Asmaa A. Al-Thani. MOPH and HMC:¬†Abdullatif Al-Khal, Muna A. S. Al-Maslamani, Mashael A. Al-Bader, Hamda Alromaihi, Roberto Bertollini, Peter V. Coyle, Einas A. |
| hCoV-19/Qatar/QA.QU..16.2.C5/2021 | EPI_ISL_6132625 | 2021-09-06 | Ministry of Public Health / Hamad Medical Corporation | Biomedical Research Center (BRC), Qatar University / Qatar Genome Project (QGP) | BRC:¬†Fatiha M. Benslimane, Heba A. Al-Khatib, Oal Al-Jamal, Dana Al-Batesh, Hadi M. Yassine, Asmaa A. Al-Thani. MOPH and HMC:¬†Abdullatif Al-Khal, Muna A. S. Al-Maslamani, Mashael A. Al-Bader, Hamda Alromaihi, Roberto Bertollini, Peter V. Coyle, Einas A. |
| hCoV-19/Wales/PHWC-PFKH4H/2021 | EPI_ISL_7009248 | 2021-11-10 | Originating lab: Wales Specialist Virology Centre Sequencing lab: Pathogen Genomics Unit | Public Health Wales Microbiology Cardiff Wales Specialist Virology Centre | Catherine Moore, Johnathan Evans, Laura Gifford, Malorie Perry, Simon Cottrell, Angela Marchbank, Alec Birchley, Alexander Adams, Amy Gaskin, Bree Gatica-Wilcox, Jason Coombes, Joel Southgate, Lauren Gilbert, Lee Graham, Nicole Pacchiarini, Sara Kumziene- |
| hCoV-19/India/MH-RFIP02800/2021 | EPI_ISL_6134373 | 2021-06-21 | BJMC,Pune | RF-IISER Pune | Krishanpal Karmodiya, Aurnab Ghose, Joy Merwin Monteiro, Suvarna Joshi, Rajesh Karyakarte |
| hCoV-19/England/PHEP-YYBIJUR/2021 | EPI_ISL_6137203 | 2021-10-22 | Respiratory Virus Unit, Microbiology Services Colindale, Public Health England | COVID-19 Genomics UK (COG-UK) Consortium | PHE Covid Sequencing Team |
| hCoV-19/England/PHEP-044260/2021 | EPI_ISL_5279101 | 2021-09-30 | Respiratory Virus Unit, Microbiology Services Colindale, Public Health England | COVID-19 Genomics UK (COG-UK) Consortium | PHE Covid Sequencing Team |

| | | | | | |
|---|---|---|---|---|---|
| hCoV-19/Malaysia/IMR_WC606483/2021 | EPI_ISL_7015235 | 2021-11-19 | Selangor State Health Department | Infectious Disease Research Centre, National Institutes of Health, Ministry of Health Malaysia | Suppiah J, Kamel K, Mohd Zawawi Z, Azizan MA, Ramly N, Robert F, Anasir MI, Ahmad Fazilah NA, Ahmad FA, Norhisham SN, Mohamad Sukri MZ, Rosli NR, Thayan R |
| hCoV-19/India/GJ-INSACOG-GBRC2789/2021 | EPI_ISL_7018777 | 2021-10-30 | Sanjeevani Metropolis, Rajkot | Gujarat Biotechnology Research Centre | Nimesh Patel,Nitin Shukla,Ramesh Pandit,Nitin Savaliya,Priyank Chavda,Sonal Sharma ,Janvi Raval,Apurvasinh Puvar,Zarna Patel,Dinesh Kumar,Bhadreshsinh Gohil,Shafquat Jahan,Chaitanya Joshi,Madhvi Joshi |
| hCoV-19/Australia/VIC19603/2021 | EPI_ISL_5288656 | 2021-06-11 | Microbiological Diagnostic Unit - Public Health Laboratory (MDU-PHL) | Microbiological Diagnostic Unit - Public Health Laboratory (MDU-PHL) | Seemann, T., Horan, K., Sherry, N.L. |
| hCoV-19/Australia/VIC19580/2021 | EPI_ISL_5288679 | 2021-05-26 | Microbiological Diagnostic Unit - Public Health Laboratory (MDU-PHL) | Microbiological Diagnostic Unit - Public Health Laboratory (MDU-PHL) | Seemann, T., Horan, K., Sherry, N.L. |
| hCoV-19/Australia/VIC19532/2021 | EPI_ISL_5288704 | 2021-02-18 | Microbiological Diagnostic Unit - Public Health Laboratory (MDU-PHL) | Microbiological Diagnostic Unit - Public Health Laboratory (MDU-PHL) | Seemann, T., Horan, K., Sherry, N.L. |
| hCoV-19/Australia/VIC19602/2021 | EPI_ISL_5288759 | 2021-05-27 | Microbiological Diagnostic Unit - Public Health Laboratory (MDU-PHL) | Microbiological Diagnostic Unit - Public Health Laboratory (MDU-PHL) | Seemann, T., Horan, K., Sherry, N.L. |
| hCoV-19/Australia/VIC18252/2021 | EPI_ISL_5288912 | 2021-02-11 | Microbiological Diagnostic Unit - Public Health Laboratory (MDU-PHL) | Microbiological Diagnostic Unit - Public Health Laboratory (MDU-PHL) | Seemann, T., Horan, K., Sherry, N.L. |
| hCoV-19/Australia/VIC18526/2021 | EPI_ISL_5289025 | 2021-05-14 | Microbiological Diagnostic Unit - Public Health Laboratory (MDU-PHL) | Microbiological Diagnostic Unit - Public Health Laboratory (MDU-PHL) | Seemann, T., Horan, K., Sherry, N.L. |
| hCoV-19/Australia/VIC18742/2021 | EPI_ISL_5289031 | 2021-06-11 | Microbiological Diagnostic Unit - Public Health Laboratory (MDU-PHL) | Microbiological Diagnostic Unit - Public Health Laboratory (MDU-PHL) | Seemann, T., Horan, K., Sherry, N.L. |
| hCoV-19/Australia/VIC14600/2020 | EPI_ISL_5297765 | 2020-08-26 | Microbiological Diagnostic Unit - Public Health Laboratory (MDU-PHL) | Microbiological Diagnostic Unit - Public Health Laboratory (MDU-PHL) | Seemann, T., Horan, K., Sherry, N.L. |
| hCoV-19/Australia/VIC10766/2020 | EPI_ISL_5300783 | 2020-09-02 | Microbiological Diagnostic Unit - Public Health Laboratory (MDU-PHL) | Microbiological Diagnostic Unit - Public Health Laboratory (MDU-PHL) | Seemann, T., Horan, K., Sherry, N.L. |
| hCoV-19/Portugal/PT18444/2021 | EPI_ISL_5304215 | 2021-10-02 | H Fernando Fonseca | Instituto Nacional de Saude (INSA) | Borges et al |
| hCoV-19/USA/CA-CDC-ASC210311130/2021 | EPI_ISL_6063077 | 2021-10-04 | Aegis Sciences Corporation | Centers for Disease Control and Prevention Division of Viral Diseases, Pathogen Discovery | Dakota Howard,Dhwani Batra,Peter Cook,Jason Caravas,Benjamin Rambo-Martin,Scott Sammons,Yvette Unoarumhi,Matthew Schmerer,Kristine Lacek,Tymeckia Kendall,Victoria Caban Figueroa,Shatavia Morrison,Christopher Gulvick,Erisa Sula,Cyndi Clark,Patrick Campbell |
| hCoV-19/England/MILK-2C1C6A3/2021 | EPI_ISL_7023675 | 2021-11-27 | Lighthouse Lab in Milton Keynes | Wellcome Sanger Institute for the COVID-19 Genomics UK (COG-UK) Consortium | The Lighthouse Lab in Milton Keynes and Alex Alderton, Roberto Amato, Jeffrey Barrett, Sonia Goncalves, Ewan Harrison, David K. Jackson, Ian Johnston, Dominic Kwiatkowski, Cordelia Langford, John Sillitoe on behalf of the Wellcome Sanger Institute COVID-1 |
| hCoV-19/Australia/NSW-ICPMR-10102/2021 | EPI_ISL_5305611 | 2021-10-03 | Australian Clinical Labs (formerly Healthscope Pathology) | Clinical Pathology and Medical Research; Westmead Hospital; University of Sydney | Arnott A., Draper J., Gall M., Martinez E., Rockett R., Sintchenko V., on behalf of ICPMR |
| hCoV-19/Australia/NSW-ICPMR-10817/2021 | EPI_ISL_5306008 | 2021-08-31 | St Vincent's Pathology (SydPath) | Clinical Pathology and Medical Research; Westmead Hospital; University of Sydney | Arnott A., Draper J., Gall M., Martinez E., Rockett R., Sintchenko V., on behalf of ICPMR |

| | | | | | |
|---|---|---|---|---|---|
| hCoV-19/FrenchGuiana/IPP29138/2021 | EPI_ISL_5306645 | 2021-09-13 | Outre mer | National Reference Center for Viruses of Respiratory Infections, Institut Pasteur, Paris | Marion Barbet, Sylvie Behillil, M√©line Bizard, Angela Brisebarre, Camille Capel, Vincent Enouf, Louise Lefran√ßois, Fr√©d√©ric Lemoine, Christophe Malabat, Corinne Maufrais, Slim El Khiari, Julien Fumey, Etienne Simon-Lori√®re, Maud Vanpeene, Sylvie Van |
| hCoV-19/USA/MI-CDC-FG-147465/2021 | EPI_ISL_5308065 | 2021-10-08 | Fulgent Genetics | Centers for Disease Control and Prevention Division of Viral Diseases, Pathogen Discovery | Dakota Howard,Dhwani Batra,Peter Cook,Jason Caravas,Benjamin Rambo-Martin,Scott Sammons,Yvette Unoarumhi,Matthew Schmerer,Kristine Lacek,Tymeckia Kendall,Victoria Caban Figueroa,Shatavia Morrison,Christopher Gulvick,Erisa Sula,Harry Gao,Mickey Li,John Gao |
| hCoV-19/Romania/SC24537/2021 | EPI_ISL_6157788 | 2021-10-04 | Medical Genetics Laboratory, Regional Centre of Medical Genetics, Emergency County Hospital Craiova | Medical Genetics Laboratory, Regional Centre of Medical Genetics, Emergency County Hospital Craiova | Anca-Lelia (Riza) Costache, Ioana Streata, Stefania Dorobantu, Adina Dragos, Mihai Cucu, Ana-Maria Buga, Razvan Plesea, Andrei Pirvu, Elena Plesea, Monica Cara, Mihai Ioana |
| hCoV-19/Canada/NB-CHUDGLD-219385/2021 | EPI_ISL_6157844 | 2021-10-15 | NB-H√¥pital Georges L. Dumont | New Brunswick - Vitalit√© Health Network | Chacko S., Shaw W., Lacroix J., Allain E., Lyons P., Garceau R., Desnoyers G., Crapoulet N. |
| hCoV-19/England/MILK-2C03E2F/2021 | EPI_ISL_7037264 | 2021-11-26 | Lighthouse Lab in Milton Keynes | Wellcome Sanger Institute for the COVID-19 Genomics UK (COG-UK) Consortium | The Lighthouse Lab in Milton Keynes and Alex Alderton, Roberto Amato, Jeffrey Barrett, Sonia Goncalves, Ewan Harrison, David K. Jackson, Ian Johnston, Dominic Kwiatkowski, Cordelia Langford, John Sillitoe on behalf of the Wellcome Sanger Institute COVID-1 |
| hCoV-19/Australia/VIC18865/2021 | EPI_ISL_5314293 | 2021-06-26 | Microbiological Diagnostic Unit - Public Health Laboratory (MDU-PHL) | Microbiological Diagnostic Unit - Public Health Laboratory (MDU-PHL) | Seemann, T., Horan, K., Sherry, N.L. |
| hCoV-19/Australia/VIC18862/2021 | EPI_ISL_5314390 | 2021-06-25 | Microbiological Diagnostic Unit - Public Health Laboratory (MDU-PHL) | Microbiological Diagnostic Unit - Public Health Laboratory (MDU-PHL) | Seemann, T., Horan, K., Sherry, N.L. |
| hCoV-19/Australia/VIC18812/2021 | EPI_ISL_5314444 | 2021-06-15 | Microbiological Diagnostic Unit - Public Health Laboratory (MDU-PHL) | Microbiological Diagnostic Unit - Public Health Laboratory (MDU-PHL) | Seemann, T., Horan, K., Sherry, N.L. |
| hCoV-19/USA/SC-CDC-ASC210317163/2021 | EPI_ISL_6160627 | 2021-09-24 | Aegis Sciences Corporation | Centers for Disease Control and Prevention Division of Viral Diseases, Pathogen Discovery | Dakota Howard,Dhwani Batra,Peter Cook,Jason Caravas,Benjamin Rambo-Martin,Scott Sammons,Yvette Unoarumhi,Matthew Schmerer,Kristine Lacek,Tymeckia Kendall,Victoria Caban Figueroa,Shatavia Morrison,Christopher Gulvick,Erisa Sula,Cyndi Clark,Patrick Campbell |
| hCoV-19/USA/CA-COLOR-UCSF-CC886/2021 | EPI_ISL_5323471 | 2021-05-09 | Color Genomics | Chiu Laboratory, University of California, San Francisco | Charles Chiu, Venice Servellita, Alicia Sotomayor-Gonzalez, Baolin Wang, Scott Topper, Erika Torres, Noah Brazer, Daniel Wong, Alicia Zhou |
| hCoV-19/SouthAfrica/CERI-KRISP-K025560/2021 | EPI_ISL_5327874 | 2021-09-15 | National Health Laboratory Service, Eastern Cape, South Africa | and Innvoation, Stellenbosch University and KRISP, KZN Research Innovation and Sequencing Platform, UKZN. | Oluwakemi Laguda-Akingba, Giandhari Jennifer, Pillay Sureshnee, Yajna Ramphal, Naidoo Yeshnee, Tshiabuila Derek,Tegally Houriiyah, San James, Wilkinson Eduan, de Oliveira Tulio |
| hCoV-19/SouthAfrica/CERI-KRISP-K025567/2021 | EPI_ISL_5327879 | 2021-09-15 | National Health Laboratory Service, Eastern Cape, South Africa | and Innvoation, Stellenbosch University and KRISP, KZN Research Innovation and Sequencing Platform, UKZN. | Oluwakemi Laguda-Akingba, Giandhari Jennifer, Pillay Sureshnee, Yajna Ramphal, Naidoo Yeshnee, Tshiabuila Derek,Tegally Houriiyah, San James, Wilkinson Eduan, de Oliveira Tulio |

| | | | | | |
|---|---|---|---|---|---|
| hCoV-19/USA/CT/2021/YPL-21-113931 | EPI_ISL_5330267 | 2021-09-24 | Yale Pathology Labs | Yale Pathology Labs | Sylvia White, Katherine Fajardo, Jinglan Wang, Guangxiao Yang, Richard Bouffard, Brian Daley, Peter Gershkovich, Stephanie Weirsman, Angelique Levi, Susan Bell, Monica Talmor, Kevin Schofield, Jianhui Wang, Minghao Zhong, Pei Hui, John Sinard, Chen Liu |
| hCoV-19/Turkey/HSGM-E11051/2021 | EPI_ISL_5331108 | 2021-09-28 | Ministry of Health Turkey | Ministry of Health Turkey | Fatma Bayrakdar, Yasemin Cosgun, Suleyman Yalcin, Gulay Korukluoglu |
| hCoV-19/Germany/NW-RKI-I-265476/2021 | EPI_ISL_4931139 | 2021-09-24 | SYNLAB MVZ Leverkusen | Robert Koch Institute | unknown |
| hCoV-19/Sweden/UUH-000200170848/2021 | EPI_ISL_5334072 | 2021-09-16 | Akademiska Sjukhuset, Clinical Microbiology and Hospital Hygiene | Uppsala University Hospital | Rene Kaden, Steinar Mannsverk, Julia Bergholm, Patrik Ellstr√∂m, Jonathan Haars |
| hCoV-19/Germany/NW-RKI-I-265794/2021 | EPI_ISL_4931872 | 2021-09-25 | SYNLAB MVZ Leverkusen | Robert Koch Institute | unknown |
| hCoV-19/Ghana/nCoV-TRA-971/2021 | EPI_ISL_5334593 | 2021-05-31 | National Influenza Centre | National Influenza Centre | William K. Ampofo, Michael Marks, Ivy A. Asante, Sharon Hsu, Benjamiin B. Lindsey, Benjamin H. Foulkes, Mildred Adusei-Poku, Linda Boatemaa, Lorreta Kwasah, Joseph Oliver-Commey, Ernest Asiedu, Franklin Asiedu-Bekoe, Gordon Awandare, Joyce Ngoi, Dennis La |
| hCoV-19/Ghana/nCoV-TRA-960/2021 | EPI_ISL_5334602 | 2021-05-24 | National Influenza Centre | National Influenza Centre | William K. Ampofo, Michael Marks, Ivy A. Asante, Sharon Hsu, Benjamiin B. Lindsey, Benjamin H. Foulkes, Mildred Adusei-Poku, Linda Boatemaa, Lorreta Kwasah, Joseph Oliver-Commey, Ernest Asiedu, Franklin Asiedu-Bekoe, Gordon Awandare, Joyce Ngoi, Dennis La |
| hCoV-19/Germany/BW-RKI-I-270959/2021 | EPI_ISL_4936618 | 2021-09-12 | Eurofins LifeCodexx GmbH | Robert Koch Institute | unknown |
| hCoV-19/Germany/NW-RKI-I-271382/2021 | EPI_ISL_4937186 | 2021-10-04 | Eurofins MVZ Labor Gelsenkirchen | Robert Koch Institute | unknown |
| hCoV-19/Brazil/SP-IB_134471/2021 | EPI_ISL_5336541 | 2021-09-20 | CENTRO DE SAUDE DR YOSHITO KANZAWA | Instituto Butantan | Dimas Tadeu Covas, Antonio Jorge Martins, Claudia Renata dos Santos Barros, David Schlesinger, Debora Botequio Moretti, Elaine Cristina Marqueze, Elaine Vieira Santos, Evandra Strazza Rodrigues, Heidge Fukumasu, Jayme Augusto de Souza-Neto, Jos√© Salvator |
| hCoV-19/Germany/BW-RKI-I-288563/2021 | EPI_ISL_5401270 | 2021-09-27 | Eurofins LifeCodexx GmbH | Robert Koch Institute | unknown |
| hCoV-19/Germany/BW-RKI-I-300720/2021 | EPI_ISL_5906887 | 2021-09-10 | Universit√§tsmedizin Mannheim | Robert Koch Institute | unknown |
| hCoV-19/Germany/NW-RKI-I-301441/2021 | EPI_ISL_5908630 | 2021-10-23 | SYNLAB MVZ Leverkusen | Robert Koch Institute | unknown |
| hCoV-19/Germany/BW-RKI-I-306790/2021 | EPI_ISL_6185328 | 2021-09-06 | Universit√§tsklinikum Heidelberg | Robert Koch Institute | unknown |
| hCoV-19/Germany/SH-RKI-I-307121/2021 | EPI_ISL_6186914 | 2021-10-20 | LADR Zentrallabor DR. Kramer & Kollegen Geesthacht | Robert Koch Institute | unknown |
| hCoV-19/Germany/BY-RKI-I-311539/2021 | EPI_ISL_6196770 | 2021-10-27 | LabKom - Labor Augsburg MVZ GmbH | Robert Koch Institute | unknown |
| hCoV-19/Germany/HE-RKI-I-319332/2021 | EPI_ISL_6449532 | 2021-11-05 | CENTOGENE Frankfurt Laboratory: Niederlassung Industriepark H√∂chst | Robert Koch Institute | unknown |

| | | | | | |
|---|---|---|---|---|---|
| hCoV-19/Germany/HE-RKI-I-319877/2021 | EPI_ISL_6452766 | 2021-11-04 | amedes MVZ f√ºr Labordiagnostik Rhein-Main | Robert Koch Institute | unknown |
| hCoV-19/Germany/BW-RKI-I-322903/2021 | EPI_ISL_6462413 | 2021-11-01 | Landesgesundheitsamt Baden-W√ºrttemberg | Robert Koch Institute | unknown |
| hCoV-19/Germany/SH-RKI-I-323536/2021 | EPI_ISL_6465180 | 2021-10-28 | LADR Zentrallabor DR. Kramer & Kollegen Geesthacht | Robert Koch Institute | unknown |
| hCoV-19/Germany/BY-RKI-I-324754/2021 | EPI_ISL_6467446 | 2021-11-05 | Synlab MVZ Augsburg | Robert Koch Institute | unknown |
| hCoV-19/Germany/ST-RKI-I-329382/2021 | EPI_ISL_6952596 | 2021-11-11 | amedes MVZ Halle/Leipzig | Robert Koch Institute | unknown |
| hCoV-19/Germany/NI-RKI-I-340653/2021 | EPI_ISL_7078176 | 2021-11-17 | amedes G√∂ttingen | Robert Koch Institute | unknown |
| hCoV-19/Slovakia/34343/2021 | EPI_ISL_7052004 | 2021-11-08 | Public Health Authority of the Slovak Republic | Faculty of Medicine in Martin, Comenius University | Dusan Loderer, Katarina Janikova, Ivana Kasubova, Maria Skerenova, Marian Grendar |
| hCoV-19/Germany/BY-RKI-I-345421/2021 | EPI_ISL_7091398 | 2021-11-11 | Labor Becker & Kollegen (Standort M√ºnchen) | Robert Koch Institute | unknown |
| hCoV-19/Germany/NW-RKI-I-351019/2021 | EPI_ISL_7111376 | 2021-11-26 | Labor Dr. Wisplinghoff - K√∂ln | Robert Koch Institute | unknown |
| hCoV-19/NewZealand/21MV2141/2021 | EPI_ISL_5348533 | 2021-10-12 | LabPLUS | Institute of Environmental Science and Research (ESR) | Sarah Cockerton, Clare Gebbie, Dhanisha Patel, Rachel Boyle, SallyAnn Harbison, Olivia Stroeven, Xiaoyun Ren, Matt Storey, Nikki Freed, Muhammad Faisal, Jing Wang, Hermes Perez, Anja Werno, Antje van der Linden, Arlo Upton, Chris Mansell, David Hammer, Dr |
| hCoV-19/NewZealand/21CV0939/2021 | EPI_ISL_5911346 | 2021-10-16 | LabPLUS | Institute of Environmental Science and Research (ESR) | Sarah Cockerton, Clare Gebbie, Dhanisha Patel, Rachel Boyle, SallyAnn Harbison, Olivia Stroeven, Xiaoyun Ren, Matt Storey, Nikki Freed, Muhammad Faisal, Jing Wang, Hermes Perez, Anja Werno, Antje van der Linden, Arlo Upton, Chris Mansell, David Hammer, Dr |
| hCoV-19/USA/TX-CDC-ASC210361235/2021 | EPI_ISL_6163851 | 2021-10-13 | Aegis Sciences Corporation | Centers for Disease Control and Prevention Division of Viral Diseases, Pathogen Discovery | Dakota Howard,Dhwani Batra,Peter Cook,Jason Caravas,Benjamin Rambo-Martin,Scott Sammons,Yvette Unoarumhi,Matthew Schmerer,Kristine Lacek,Tymeckia Kendall,Victoria Caban Figueroa,Shatavia Morrison,Christopher Gulvick,Erisa Sula,Cyndi Clark,Patrick Campbell |
| hCoV-19/India/TG-RFCH01242_CIB3564/2021 | EPI_ISL_7056437 | 2021-05-01 | MapMyGenome | CSIR-Centre for Cellular and Molecular Biology | Lamuk Zaveri,Tulasi Nagabandi,Jandhyala Sai Krishna,Malini Nemalikanti,Sreelekshmi MS, Ara Sreenivas,Shreekant Verma, Amareshwar Vodapalli,B Himasri,Valli Nagalakshmi Undamatla, Sumedha Avadhanula,Priya Nurkurthy,Vidhyadhari Methuku,M. Aravind Kumar, San |
| hCoV-19/India/TG-RFCH01323_CIB3647/2021 | EPI_ISL_7056717 | 2021-05-02 | MapMyGenome | CSIR-Centre for Cellular and Molecular Biology | Lamuk Zaveri,Tulasi Nagabandi,Jandhyala Sai Krishna,Malini Nemalikanti,Sreelekshmi MS, Ara Sreenivas,Shreekant Verma, Amareshwar Vodapalli,B Himasri,Valli Nagalakshmi Undamatla, Sumedha Avadhanula,Priya Nurkurthy,Vidhyadhari Methuku,M. Aravind Kumar, San |

| | | | | | |
|---|---|---|---|---|---|
| hCoV-19/India/TG-RFCH01394_CIB3718/2021 | EPI_ISL_7056967 | 2021-05-04 | MapMyGenome | CSIR-Centre for Cellular and Molecular Biology | Lamuk Zaveri,Tulasi Nagabandi,Jandhyala Sai Krishna,Malini Nemalikanti,Sreelekshmi MS, Ara Sreenivas,Shreekant Verma, Amareshwar Vodapalli,B Himasri,Valli Nagalakshmi Undamatla, Sumedha Avadhanula,Priya Nurkurthy,Vidhyadhari Methuku,M. Aravind Kumar, San |
| hCoV-19/India/TG-RFCH01416_CIB3742/2021 | EPI_ISL_7057021 | 2021-05-07 | MapMyGenome | CSIR-Centre for Cellular and Molecular Biology | Lamuk Zaveri,Tulasi Nagabandi,Jandhyala Sai Krishna,Malini Nemalikanti,Sreelekshmi MS, Ara Sreenivas,Shreekant Verma, Amareshwar Vodapalli,B Himasri,Valli Nagalakshmi Undamatla, Sumedha Avadhanula,Priya Nurkurthy,Vidhyadhari Methuku,M. Aravind Kumar, San |
| hCoV-19/USA/NV-NSPHL-433358/2021 | EPI_ISL_5362391 | 2021-09-24 | Nevada State Public Health Laboratory | Nevada State Public Health Laboratory | Andrew Gorzalski, Mark Pandori |
| hCoV-19/Slovakia/35953/2021 | EPI_ISL_7060995 | 2021-11-09 | Public Health Authority of the Slovak Republic | Faculty of Medicine in Martin, Comenius University | Dusan Loderer, Katarina Janikova, Ivana Kasubova, Maria Skerenova, Marian Grendar |
| hCoV-19/Denmark/DCGC-243048/2021 | EPI_ISL_7061585 | 2021-11-26 | Department of Bacteria, Parasites and Fungi, Statens Serum Institut, Copenhagen, Denmark | Statens Serum Institut Bioinformatics and Microbial Genomics | Danish Covid-19 Genome Consortium |
| hCoV-19/USA/NE-CDC-ASC210357374/2021 | EPI_ISL_6169008 | 2021-10-11 | Aegis Sciences Corporation | Centers for Disease Control and Prevention Division of Viral Diseases, Pathogen Discovery | Dakota Howard,Dhwani Batra,Peter Cook,Jason Caravas,Benjamin Rambo-Martin,Scott Sammons,Yvette Unoarumhi,Matthew Schmerer,Kristine Lacek,Tymeckia Kendall,Victoria Caban Figueroa,Shatavia Morrison,Christopher Gulvick,Erisa Sula,Cyndi Clark,Patrick Campbell |
| hCoV-19/USA/TX-HMH-MCoV-48887/2021 | EPI_ISL_5339223 | 2021-07-06 | Houston Methodist Hospital | Houston Methodist Hospital | Randall J. Olsen, Paul A. Christensen, S. Wesley Long, Sishir Subedi, Robert Olson, Marcus Nguyen, James J. Davis, Matthew Ojeda Saavedra, Prasanti Yerramilli, Layne Pruitt, Kristina Reppond, Madison N. Shyer, Jessica Cambric, Ryan Gadd, Ilya J. Finkels |
| hCoV-19/Reunion/CHU7211111300/2021 | EPI_ISL_6779428 | 2021-11-12 | LABM SAINT BENOIT | Laboratoire de virologie, CNR arbovirus Associ√©, Chu de la R√©union | Etienne Frumence, Nicolas M'nemosyme, Anne-Julie Gourd√©, Rubens Lhonneur, Nicolas Traversier, Marie-Christine Jaffar Bandjee |
| hCoV-19/Reunion/CHU798000081168/2021 | EPI_ISL_6779608 | 2021-08-09 | CHU - Saint-Denis | Laboratoire de virologie, CNR arbovirus Associ√©, Chu de la R√©union | Etienne Frumence, Anne-Julie Gourd√©, Rubens Lhonneur, Nicolas Traversier, Marie-Christine Jaffar Bandjee |
| hCoV-19/Reunion/CHU798000091333/2021 | EPI_ISL_6779793 | 2021-10-26 | CHU - Saint-Denis | Laboratoire de virologie, CNR arbovirus Associ√©, Chu de la R√©union | Etienne Frumence, Nicolas M'nemosyme, Anne-Julie Gourd√©, Rubens Lhonneur, Nicolas Traversier, Marie-Christine Jaffar Bandjee |
| hCoV-19/Peru/LIM-INS-7360/2021 | EPI_ISL_5595974 | 2021-09-25 | Laboratorio de Referencia Nacional de Virus Respiratorios. Centro Nacional de Salud Publica. Instituto Nacional de Salud Peru. | Laboratorio de Referencia Nacional de Virus Respiratorios. Centro Nacional de Salud Publica. Instituto Nacional de Salud Peru. | Carlos Padilla Rojas, Veronica Hurtado Vela, Orson Mestanza Millones, Iris Silva Molina, Luis Barcena Flores, Alicia Nu√±ez Llanos, Steve Acedo Lazo, Wendy Lizarraga Olivares, Victor Jimenez Vasquez, Sara Gordillo Vilchez, Nieves Sevilla Casta√±eda, Princ |

| | | | | | |
|---|---|---|---|---|---|
| hCoV-19/USA/WI-CDC-ASC210087434/2021 | EPI_ISL_5372003 | 2021-09-20 | Aegis Sciences Corporation | Centers for Disease Control and Prevention Division of Viral Diseases, Pathogen Discovery | Dakota Howard,Dhwani Batra,Peter Cook,Jason Caravas,Benjamin Rambo-Martin,Scott Sammons,Yvette Unoarumhi,Matthew Schmerer,Kristine Lacek,Tymeckia Kendall,Victoria Caban Figueroa,Shatavia Morrison,Christopher Gulvick,Erisa Sula,Cyndi Clark,Patrick Campbell |
| hCoV-19/USA/NC-ECU-CORVASEQ-108303219/2021 | EPI_ISL_6174224 | 2021-06-08 | Molecular Biology Laboratory, Vidant Medical Center | Brody Integrative Genomics Core, East Carolina University | Weihua Huang, Changhong Yin, Kimberly P. Briley, Heather Duncan, John T. Fallon |
| hCoV-19/USA/NC-ECU-CORVASEQ-108580629/2021 | EPI_ISL_6174310 | 2021-06-28 | Molecular Biology Laboratory, Vidant Medical Center | Brody Integrative Genomics Core, East Carolina University | Weihua Huang, Changhong Yin, Kimberly P. Briley, Heather Duncan, John T. Fallon |
| hCoV-19/USA/NC-ECU-CORVASEQ-108561047/2021 | EPI_ISL_6174317 | 2021-06-26 | Molecular Biology Laboratory, Vidant Medical Center | Brody Integrative Genomics Core, East Carolina University | Weihua Huang, Changhong Yin, Kimberly P. Briley, Heather Duncan, John T. Fallon |
| hCoV-19/USA/NC-ECU-CORVASEQ-108690260/2021 | EPI_ISL_6174356 | 2021-07-06 | Molecular Biology Laboratory, Vidant Medical Center | Brody Integrative Genomics Core, East Carolina University | Weihua Huang, Changhong Yin, Kimberly P. Briley, Heather Duncan, John T. Fallon |
| hCoV-19/USA/NC-ECU-CORVASEQ-108769628/2021 | EPI_ISL_6174600 | 2021-07-12 | Molecular Biology Laboratory, Vidant Medical Center | Brody Integrative Genomics Core, East Carolina University | Weihua Huang, Changhong Yin, Kimberly P. Briley, Heather Duncan, John T. Fallon |
| hCoV-19/Sweden/101664818/2021 | EPI_ISL_6179177 | 2021-10-18 | Swedish national genomic surveillance program of SARS-CoV-2 | The Public Health Agency of Sweden | Maximilian Riess, Maria Lind Karlberg, Alma Brolund, Swedish national genomic surveillance program of SARS-CoV-2 |
| hCoV-19/USA/TX-HMH-MCoV-48807/2020 | EPI_ISL_5383863 | 2020-07-15 | Houston Methodist Hospital | Houston Methodist Hospital | Randall J. Olsen, Paul A. Christensen, S. Wesley Long, Sishir Subedi, Robert Olson, Marcus Nguyen, James J. Davis, Matthew Ojeda Saavedra, Prasanti Yerramilli, Layne Pruitt, Kristina Reppond, Madison N. Shyer, Jessica Cambric, Ryan Gadd, Ilya J. Finkels |
| hCoV-19/USA/CA-CDC-FG-148043/2021 | EPI_ISL_5389886 | 2021-10-09 | Fulgent Genetics | Centers for Disease Control and Prevention Division of Viral Diseases, Pathogen Discovery | Dakota Howard,Dhwani Batra,Peter Cook,Jason Caravas,Benjamin Rambo-Martin,Scott Sammons,Yvette Unoarumhi,Matthew Schmerer,Kristine Lacek,Tymeckia Kendall,Victoria Caban Figueroa,Shatavia Morrison,Christopher Gulvick,Erisa Sula,Harry Gao,Mickey Li,John Gao |
| hCoV-19/Japan/IC-2100/2021 | EPI_ISL_5403381 | 2021-09-23 | SARS-CoV-2 testing team, National Institute of Infectious Diseases | Pathogen Genomics Center, National Institute of Infectious Diseases | Tsuyoshi Sekizuka, Kentaro Itokawa, Rina Tanaka, Masanori Hashino, Nozomu Hanaoka, Masumichi Saito, Naomi Nojiri, Hazuka Y Furihata, Tsuguto Fujimoto, Makoto Kuroda |
| hCoV-19/Spain/MD-H12O_1320/2021 | EPI_ISL_5410845 | 2021-09-08 | HOSPITAL UNIVERSITARIO 12 DE OCTUBRE | HOSPITAL UNIVERSITARIO 12 DE OCTUBRE | Carmen Mart√≠n-Higuera, Mar Aguilera, Sagrario Zurita,Noelia Moral, Irene M√∫√±oz-Gallego, Rafael Delgado, M.¬™ Dolores Folgueira, Esther Viedma |
| hCoV-19/USA/NE-NPHL-42616/2021 | EPI_ISL_7116461 | 2021-11-29 | Nebraska Public Health Laboratory | NPHL COVID-19 Response Team | NPHL COVID-19 Response Team |
| hCoV-19/USA/NE-NPHL-42617/2021 | EPI_ISL_7116439 | 2021-11-29 | Nebraska Public Health Laboratory | NPHL COVID-19 Response Team | NPHL COVID-19 Response Team |
| hCoV-19/USA/NE-NPHL-42615/2021 | EPI_ISL_7116738 | 2021-11-29 | Nebraska Public Health Laboratory | NPHL COVID-19 Response Team | NPHL COVID-19 Response Team |
| hCoV-19/Chile/VS-159467/2021 | EPI_ISL_5411178 | 2021-07-29 | Genetica Molecular and Subdepartamento de Virologia ISP Chile | Instituto de Salud Publica de Chile | Karen Orostica, Constanza Campano, Barbara Parra, Patricia Bustos, Rodrigo Fasce, Andres Castillo, Jorge Fernandez |

| | | | | | |
|---|---|---|---|---|---|
| hCoV-19/Chile/RM-157389/2021 | EPI_ISL_5411188 | 2021-09-26 | Genetica Molecular and Subdepartamento de Virologia ISP Chile | Instituto de Salud Publica de Chile | Karen Orostica, Constanza Campano, Barbara Parra, Patricia Bustos, Rodrigo Fasce, Andres Castillo, Jorge Fernandez |
| hCoV-19/Belgium/rega-12463/2021 | EPI_ISL_5414920 | 2021-05-17 | KU Leuven, Rega Institute, Clinical and Epidemiological Virology | KU Leuven, Rega Institute, Clinical and Epidemiological Virology | Tony Wawina-Bokalanga, Bert Vanmechelen, Joan Marti-Carerras, Piet Maes |
| hCoV-19/India/AP-CCMB-CIB0017/2021 | EPI_ISL_5416956 | 2021-09-06 | AP SSO | CSIR-Centre for Cellular and Molecular Biology-INSACOG | Lamuk Zaveri,Tulasi Nagabandi,Payel Mukherjee,Onkar Kulkarni, Sofia Banu, Jandhyala Sai Krishna,Ara Sreenivas,Shreekant Verma, Amareshwar Vodapalli ,B Himasri,Valli Nagalakshmi Undamatla, Sumedha Avadhanula, Priya Nurkurthy, Vidhyadhari Methuku, Archana B |
| hCoV-19/SouthAfrica/NICD-N16668/2021 | EPI_ISL_5416272 | 2021-08-05 | TAMBO MEMORIAL LABORATORY | Diseases of the National Health Laboratory Service | Amoako DG, Everatt J, Scheepers C, Mohale T, Ntuli N, Mahlangu B, Mnguni A, Ismail A, Bhiman JN |
| hCoV-19/SouthAfrica/NICD-N16631/2021 | EPI_ISL_5416384 | 2021-09-09 | KIMBERLEY LABORATORY | Diseases of the National Health Laboratory Service | Amoako DG, Everatt J, Scheepers C, Mohale T, Ntuli N, Mahlangu B, Mnguni A, Ismail A, Bhiman JN |
| hCoV-19/SouthAfrica/NICD-N16184/2021 | EPI_ISL_5416420 | 2021-08-25 | TSHEPONG LABORATORY | Diseases of the National Health Laboratory Service | Amoako DG, Everatt J, Scheepers C, Mohale T, Ntuli N, Mahlangu B, Mnguni A, Ismail A, Bhiman JN |
| hCoV-19/India/AP-CCMB-CIA9812/2021 | EPI_ISL_5416809 | 2021-09-02 | AP SSO | CSIR-Centre for Cellular and Molecular Biology-INSACOG | Vidhyadhari Methuku,Tulasi Nagabandi,Lamuk Zaveri, Ara Sreenivas,Shreekant Verma, Amareshwar Vodapalli ,B Himasri,Valli Nagalakshmi Undamatla, Priya Nurkurthy,Sumedha Avadhanula, Archana Bharadwaj Siva,Payel Mukherjee,Onkar Kulkarni, Jandhyala Sai Krishna |
| hCoV-19/India/AP-CCMB-CIA9785/2021 | EPI_ISL_5416872 | 2021-09-04 | AP SSO | CSIR-Centre for Cellular and Molecular Biology-INSACOG | Lamuk Zaveri,Tulasi Nagabandi,Payel Mukherjee,Onkar Kulkarni, Sofia Banu, Jandhyala Sai Krishna,Ara Sreenivas,Shreekant Verma, Amareshwar Vodapalli ,B Himasri,Valli Nagalakshmi Undamatla, Sumedha Avadhanula, Priya Nurkurthy, Vidhyadhari Methuku, Archana B |
| hCoV-19/Turkey/HSGM-E16040/2021 | EPI_ISL_5417846 | 2021-10-11 | Ministry of Health Turkey | Ministry of Health Turkey | Fatma Bayrakdar, Yasemin Co≈ügun, S√ºleyman Yalcin, G√ºlay Korukluoƒülu |
| hCoV-19/Luxembourg/LNS1481121/2021 | EPI_ISL_6187099 | 2021-10-19 | Laboratoires d'analyses medicales - Ketterthill | Laboratoire national de sante, Microbiology, Microbial Genomics Platform | Anke Wienecke-Baldacchino, Catherine Ragimbeau,Jessica Tapp, Fatu Djabi, Lise Pignon, Raoul Salmon,Virginie Jover, Elodie Solarino, Serge Vedy, Caroline Scheiber, Tamir Abdelrahman |
| hCoV-19/Japan/PG-158019/2021 | EPI_ISL_7091577 | 2021-09-08 | Department of Health Science, Gifu Prefectural Research Institute for Health and Environmental Sciences | Gifu Prefectural Research Institute for Health and Environmental Sciences | Yuta Uramoto, Masahiro Nishioka, Tomohiro Yamaguchi, Takuya Mizuno, Tsuyoshi Kuzuguchi, Yoshihiro Kameyama, Tsuyoshi Sekizuka, Kentaro Itokawa, Rina Tanaka, Masanori Hashino, Makoto Kuroda |
| hCoV-19/Switzerland/GE-HUG-36104667/2021 | EPI_ISL_7095010 | 2021-11-20 | University Hospitals of Geneva, Laboratory of Virology | HUG, Laboratory of Virology and the Health2030 Genome Center | Samuel Cordey, Aline Mamin, Francisco Perez, Ana Rita Goncalves, Laurent Kaiser, Lorenzo Cerutti, Cedric Howald, Henri Pegeot, Melyssa Elies, Deborah Penet, Keith Harshman, Ioannis Xenarios |

| | | | | | |
|---|---|---|---|---|---|
| hCoV-19/Sweden/01_SE100_21CS514903/2021 | EPI_ISL_7120688 | 2021-11-19 | Karolinska University Hospital Solna | Karolinska University Hospital | Jan Albert ,Tobias Allander ,Annelie Bjerkner ,Sandra Broddesson ,Robert Dyrdak ,Martin Ekman ,Lynda Eneh ,Lina Guerra Blomqvist ,Karolina Ininbergs ,Tanja Normark ,Isak Sylvin ,Zhibing Yun ,Martina Wahlund ,Valtteri Wirta |
| hCoV-19/Botswana/R42B81_BHP_000828260/2021 | EPI_ISL_7121202 | 2021-11-08 | National Health Laboratory | Botswana Harvard HIV Reference Laboratory | Sikhulile Moyo, Wonderful T. Choga, Dorcas Maruapula, Keoratile Ntshambiwa, Sefetogi Ramaologa, Thongbotho Mphoyakgosi, Boitumelo Zuze, Botshelo Radibe, Legodile Kooepile, Ontlametse T. Bareng, Pamela Smith-Lawrence, Kgomotso Moruisi, Roger Shapiro, Shahi |
| hCoV-19/Botswana/R44B92_BHP_000848712/2021 | EPI_ISL_7121217 | 2021-11-26 | National Health Laboratory | Botswana Harvard HIV Reference Laboratory | Sikhulile Moyo, Wonderful T. Choga, Dorcas Maruapula, Keoratile Ntshambiwa, Sefetogi Ramaologa, Thongbotho Mphoyakgosi, Boitumelo Zuze, Botshelo Radibe, Legodile Kooepile, Ontlametse T. Bareng, Pamela Smith-Lawrence, Kgomotso Moruisi, Roger Shapiro, Shahi |
| hCoV-19/Botswana/R44B37_BHP_2421009673/2021 | EPI_ISL_7121236 | 2021-11-25 | Diagnofirm Medical Laboratories | Botswana Harvard HIV Reference Laboratory | Sikhulile Moyo, Wonderful T. Choga, Dorcas Maruapula, Keoratile Ntshambiwa, Sefetogi Ramaologa, Thongbotho Mphoyakgosi, Boitumelo Zuze, Botshelo Radibe, Legodile Kooepile, Ontlametse T. Bareng, Pamela Smith-Lawrence, Kgomotso Moruisi, Roger Shapiro, Shahi |
| hCoV-19/USA/SC-CDC-MMB11359715/2021 | EPI_ISL_7121755 | 2021-10-30 | Mako Medical | Centers for Disease Control and Prevention Division of Viral Diseases, Pathogen Discovery | Dakota Howard,Dhwani Batra,Peter Cook,Jason Caravas,Benjamin Rambo-Martin,Scott Sammons,Yvette Unoarumhi,Matthew Schmerer,Kristine Lacek,Tymeckia Kendall,Victoria Caban Figueroa,Shatavia Morrison,Christopher Gulvick,Erisa Sula,Matthew Tugwell,Lauren Moon, |
| hCoV-19/USA/CO-CDC-MMB11365048/2021 | EPI_ISL_7122128 | 2021-10-31 | Mako Medical | Centers for Disease Control and Prevention Division of Viral Diseases, Pathogen Discovery | Dakota Howard,Dhwani Batra,Peter Cook,Jason Caravas,Benjamin Rambo-Martin,Scott Sammons,Yvette Unoarumhi,Matthew Schmerer,Kristine Lacek,Tymeckia Kendall,Victoria Caban Figueroa,Shatavia Morrison,Christopher Gulvick,Erisa Sula,Matthew Tugwell,Lauren Moon, |
| hCoV-19/Nigeria/CV3539/2021 | EPI_ISL_7122939 | 2021-01-30 | Genexpert Molecular Diagnostic Laboratory | Africa Centre for Excellence for Genomics of Infectious Diseases (ACEGID), Redeemer‚Äôs University | Olawoye, I.B., Oluniyi, P.E., Eromon, P.E., Oguzie, J.U., Kayode, A.T., Uwanibe, J.N., Ugwu, C.A., Akano, K.O., Ajogbasile, F.V., Abechi, P.S., Olumade, T.J., Nosamiefan, I., Folarin, O., Happi, C.T. |
| hCoV-19/Nigeria/CV3541/2021 | EPI_ISL_7122950 | 2021-02-02 | Genexpert Molecular Diagnostic Laboratory | Africa Centre for Excellence for Genomics of Infectious Diseases (ACEGID), Redeemer‚Äôs University | Olawoye, I.B., Oluniyi, P.E., Eromon, P.E., Oguzie, J.U., Kayode, A.T., Uwanibe, J.N., Ugwu, C.A., Akano, K.O., Ajogbasile, F.V., Abechi, P.S., Olumade, T.J., Nosamiefan, I., Folarin, O., Happi, C.T. |
| hCoV-19/Mozambique/CERI-KRISP-K025912/2021 | EPI_ISL_5425639 | 2021-06-25 | Instituto Nacional de Saude (INS), Mozambique | [illegible] and Innvoation, Stellenbosch University and KRISP, KZN Research Innovation and Sequencing Platform, UKZN. | Nalia Ismael, Nadia Sitoe, Paulo Arnaldo, Nedio Mabunda, Giandhari Jennifer, Pillay Sureshnee, Yajna Ramphal, Naidoo Yeshnee, Tshiabuila Derek,Tegally Houriiyah, San James, Wilkinson Eduan, de Oliveira Tulio |

| | | | | | |
|---|---|---|---|---|---|
| hCoV-19/Israel/SMC-7016827/2021 | EPI_ISL_6200240 | 2021-09-10 | Shamir Medical Center (Asaf Harofe) | Shamir Medical Center (Asaf Harofe) | Nir Rainy,Chen Weiner,Reut Sorek Abramovich,Yevgeni Yegorov,Anna Vishnevsky,Patricia Benveniste-Lekovitz,Abu Hamad Ramzia,Adina Bar Chaim |
| hCoV-19/SouthAfrica/NICD-N18962/2021 | EPI_ISL_6202232 | 2021-10-07 | KIMBERLEY LABORATORY | Diseases of the National Health Laboratory Service | Amoako DG, Everatt J, Scheepers C, Mohale T, Ntuli N, Mahlangu B, Mnguni A, Ismail A, Bhiman JN |
| hCoV-19/SouthAfrica/NICD-N18887/2021 | EPI_ISL_6203048 | 2021-10-12 | KIMBERLEY LABORATORY | Diseases of the National Health Laboratory Service | Amoako DG, Everatt J, Scheepers C, Mohale T, Ntuli N, Mahlangu B, Mnguni A, Ismail A, Bhiman JN |
| hCoV-19/Israel/SMC-7019073/2021 | EPI_ISL_6205367 | 2021-10-03 | Shamir Medical Center (Asaf Harofe) | Shamir Medical Center (Asaf Harofe) | Nir Rainy,Chen Weiner,Reut Sorek Abramovich,Yevgeni Yegorov,Anna Vishnevsky,Patricia Benveniste-Lekovitz,Abu Hamad Ramzia,Adina Bar Chaim |
| hCoV-19/Israel/SMC-7019076/2021 | EPI_ISL_6205419 | 2021-10-03 | Shamir Medical Center (Asaf Harofe) | Shamir Medical Center (Asaf Harofe) | Nir Rainy,Chen Weiner,Reut Sorek Abramovich,Yevgeni Yegorov,Anna Vishnevsky,Patricia Benveniste-Lekovitz,Abu Hamad Ramzia,Adina Bar Chaim |
| hCoV-19/Israel/SMC-7017460/2021 | EPI_ISL_6205851 | 2021-09-14 | Shamir Medical Center (Asaf Harofe) | Shamir Medical Center (Asaf Harofe) | Nir Rainy,Chen Weiner,Reut Sorek Abramovich,Yevgeni Yegorov,Anna Vishnevsky,Patricia Benveniste-Lekovitz,Abu Hamad Ramzia,Adina Bar Chaim |
| hCoV-19/Israel/SMC-7019414/2021 | EPI_ISL_6206353 | 2021-10-06 | Shamir Medical Center (Asaf Harofe) | Shamir Medical Center (Asaf Harofe) | Nir Rainy,Chen Weiner,Reut Sorek Abramovich,Yevgeni Yegorov,Anna Vishnevsky,Patricia Benveniste-Lekovitz,Abu Hamad Ramzia,Adina Bar Chaim |
| hCoV-19/Israel/SMC-7021069/2021 | EPI_ISL_6206457 | 2021-10-28 | Shamir Medical Center (Asaf Harofe) | Shamir Medical Center (Asaf Harofe) | Nir Rainy,Chen Weiner,Reut Sorek Abramovich,Yevgeni Yegorov,Anna Vishnevsky,Patricia Benveniste-Lekovitz,Abu Hamad Ramzia,Adina Bar Chaim |
| hCoV-19/Israel/SMC-7021323/2021 | EPI_ISL_6206624 | 2021-10-30 | Shamir Medical Center (Asaf Harofe) | Shamir Medical Center (Asaf Harofe) | Nir Rainy,Chen Weiner,Reut Sorek Abramovich,Yevgeni Yegorov,Anna Vishnevsky,Patricia Benveniste-Lekovitz,Abu Hamad Ramzia,Adina Bar Chaim |
| hCoV-19/Israel/SMC-7020363/2021 | EPI_ISL_6206752 | 2021-10-17 | Shamir Medical Center (Asaf Harofe) | Shamir Medical Center (Asaf Harofe) | Nir Rainy,Chen Weiner,Reut Sorek Abramovich,Yevgeni Yegorov,Anna Vishnevsky,Patricia Benveniste-Lekovitz,Abu Hamad Ramzia,Adina Bar Chaim |
| hCoV-19/Israel/SMC-7020173/2021 | EPI_ISL_6206968 | 2021-10-15 | Shamir Medical Center (Asaf Harofe) | Shamir Medical Center (Asaf Harofe) | Nir Rainy,Chen Weiner,Reut Sorek Abramovich,Yevgeni Yegorov,Anna Vishnevsky,Patricia Benveniste-Lekovitz,Abu Hamad Ramzia,Adina Bar Chaim |
| hCoV-19/Ecuador/NIC-INSPI-354245/2021 | EPI_ISL_6208209 | 2021-09-22 | INSPI-CRN DE INFLUENZA Y OTROS VIRUS RESPIRATORIOS | INSPI-CRN DE INFLUENZA Y OTROS VIRUS RESPIRATORIOS | Alfredo Bruno, Domenica de Mora, Maritza Olmedo, Michelle P√°ez, Jimmy Garc√©s, Maria Angelica Becerra, Marcela Mejia, Daniel Ramos, Fernando Llerena, Lizbeth Pati√±o, Ruben Armas |
| hCoV-19/Peru/CUS-INS-8741/2021 | EPI_ISL_6208633 | 2021-10-14 | Laboratorio de Referencia Nacional de Virus Respiratorios. Centro Nacional de Salud Publica. Instituto Nacional de Salud Peru. | Laboratorio de Referencia Nacional de Virus Respiratorios. Centro Nacional de Salud Publica. Instituto Nacional de Salud Peru. | Carlos Padilla Rojas, Veronica Hurtado Vela, Orson Mestanza Millones, Iris Silva Molina, Luis Barcena Flores, Alicia Nu√±ez Llanos, Steve Acedo Lazo, Wendy Lizarraga Olivares, Victor Jimenez Vasquez, Sara Gordillo Vilchez, Kelly Vanessa Izarra Rojas, Edwa |
| hCoV-19/Norway/24459/2021 | EPI_ISL_6211854 | 2021-10-15 | Unilabs Laboratory Medicine | Norwegian Institute of Public Health, Department of Virology | Kathrine Stene-Johansen, Kamilla Heddeland Instefjord, Hilde Elshaug, Garcia Llorente Ignacio, Jon Br√•te, Engebretsen Serina Beate, Pedersen Benedikte Nevjen, Line Victoria Moen, Debech Nadia, Atiya R Ali, Marie Paulsen Madsen, Rasmus Riis Kopperud, Hild |

| | | | | | |
|---|---|---|---|---|---|
| hCoV-19/England/MILK-295D7B5/2021 | EPI_ISL_6218261 | 2021-10-31 | Lighthouse Lab in Milton Keynes | Wellcome Sanger Institute for the COVID-19 Genomics UK (COG-UK) Consortium | The Lighthouse Lab in Milton Keynes and Alex Alderton, Roberto Amato, Jeffrey Barrett, Sonia Goncalves, Ewan Harrison, David K. Jackson, Ian Johnston, Dominic Kwiatkowski, Cordelia Langford, John Sillitoe on behalf of the Wellcome Sanger Institute COVID-1 |
| hCoV-19/Norway/OUS-14247/2021 | EPI_ISL_6226156 | 2021-09-11 | Oslo University Hospital, Department of Microbiology | Norwegian Institute of Public Health, Department of Virology | Mona Holberg-Petersen, Lise Andresen, Cathrine Fladeby, Mariann Nilsen, Teodora Plamenova Ribarska, Pv•l Marius Bjv∏rnstad, Gregor D. Gilfillan, Arvind Yegambaram Meenakshi Sundaram, Kathrine Stene-Johansen, Kamilla Heddeland Instefjord, Hilde Elshaug, Ga |
| hCoV-19/Kenya/SS3765/2021 | EPI_ISL_6227159 | 2021-09-23 | KEMRI-CMR | KEMRI-Wellcome Trust Research Programme,Kilifi | Githinji G.,Ochola I, Agoti C.,Mohamed K.S.,de Laurent Z.,Mburu M.W.,Morobe J.,Lambisia A.,Makori T.,Ongera E.,Tigoi C., Muema A., D.J.Nokes |
| hCoV-19/Netherlands/GL-RIVM-67848/2021 | EPI_ISL_6230229 | 2021-10-25 | Dutch COVID-19 response team | National Institute for Public Health and the Environment (RIVM) | Adam Meijer, Harry Vennema, Dirk Eggink, Jeroen Cremer, Sharon van den Brink, Bas van der Veer, AnneMarie van den Brandt, Lisa Wijsman, Kim Freriks, Ryanne Jaarsma, Eunv≠ce Then, Lynn Aarts, Sanne Bos, Stijn van Rossum, Florian Zwagemaker, Dennis Schmitz, |
| hCoV-19/Denmark/DCGC-207767/2021 | EPI_ISL_6231643 | 2021-11-05 | Department of Bacteria, Parasites and Fungi, Statens Serum Institut, Copenhagen, Denmark | Statens Serum Institut Bioinformatics and Microbial Genomics | Danish Covid-19 Genome Consortium |
| hCoV-19/Denmark/DCGC-189389/2021 | EPI_ISL_6232337 | 2021-10-21 | Department of Bacteria, Parasites and Fungi, Statens Serum Institut, Copenhagen, Denmark | Statens Serum Institut Bioinformatics and Microbial Genomics | Danish Covid-19 Genome Consortium |
| hCoV-19/USA/VA-CDC-MMB10875439/2021 | EPI_ISL_6255250 | 2021-10-04 | Mako Medical | Centers for Disease Control and Prevention Division of Viral Diseases, Pathogen Discovery | Dakota Howard,Dhwani Batra,Peter Cook,Jason Caravas,Benjamin Rambo-Martin,Scott Sammons,Yvette Unoarumhi,Matthew Schmerer,Kristine Lacek,Tymeckia Kendall,Victoria Caban Figueroa,Shatavia Morrison,Christopher Gulvick,Erisa Sula,Matthew Tugwell,Lauren Moon, |
| hCoV-19/USA/WY-WYPHL-20101855/2020 | EPI_ISL_6259572 | 2020-10-19 | Wyoming Public Health Laboratory | Wyoming Public Health Laboratory | Jim Mildenberger, Taylor Fearing, Channing Weber, Ashley Norberg, Chayse Rowley, Marley Goetz, Brian Dominguez, Elliot Thomasson, Sam Britz, Cari Sloma, Robert Petit, and Rob Christensen |
| hCoV-19/Kenya/C67070/2020 | EPI_ISL_6262694 | 2020-10-27 | KEMRI-Wellcome Trust Research Programme,Kilifi | KEMRI-Wellcome Trust Research Programme,Kilifi | Githinji G.,Ochola I, Agoti C.,Mohamed K.S.,de Laurent Z.,Mburu M.W.,Morobe J.,Lambisia A.,Makori T.,Ongera E, L. Ndwiga., D.J.Nokes. |
| hCoV-19/Uganda/CERI-UVRI-K022923/2021 | EPI_ISL_6262728 | 2021-06-14 | MRC/UVRI & LSHTM Uganda Research Unit, Uganda Virus Research Institute | CERI, Centre for Epidemic Response and Innvoation, Stellenbosch University and KRISP, KZN Research Innovation and Sequencing Platform, UKZN. | Deogratius Ssemwanga, Nicholas Bbosa, Hamidah Namagembe, Ronald Kiiza, Jocelyn Kiconco, John Kayiwa, Tom Lutalo, Julius Lutwama, Isaac Ssewanyana, Susan Nabadda, Henry Kyobe-Bbosa, Giandhari Jennifer, Pillay Sureshnee, Upasana Ramphal, Yajna Ramphal, Naid |

| | | | | | |
|---|---|---|---|---|---|
| hCoV-19/Uganda/CERI-UVRI-K022905/2021 | EPI_ISL_6262737 | 2021-06-14 | MRC/UVRI & LSHTM Uganda Research Unit, Uganda Virus Research Institute | CERI, Centre for Epidemic Response and Innvoation, Stellenbosch University and KRISP, KZN Research Innovation and Sequencing Platform, UKZN. | Deogratius Ssemwanga, Nicholas Bbosa, Hamidah Namagembe, Ronald Kiiza, Jocelyn Kiconco, John Kayiwa, Tom Lutalo, Julius Lutwama, Isaac Ssewanyana, Susan Nabadda, Henry Kyobe-Bbosa, Giandhari Jennifer, Pillay Sureshnee, Upasana Ramphal, Yajna Ramphal, Naid |
| hCoV-19/Uganda/CERI-UVRI-K022974/2021 | EPI_ISL_6262743 | 2021-06-15 | MRC/UVRI & LSHTM Uganda Research Unit, Uganda Virus Research Institute | CERI, Centre for Epidemic Response and Innvoation, Stellenbosch University and KRISP, KZN Research Innovation and Sequencing Platform, UKZN. | Deogratius Ssemwanga, Nicholas Bbosa, Hamidah Namagembe, Ronald Kiiza, Jocelyn Kiconco, John Kayiwa, Tom Lutalo, Julius Lutwama, Isaac Ssewanyana, Susan Nabadda, Henry Kyobe-Bbosa, Giandhari Jennifer, Pillay Sureshnee, Upasana Ramphal, Yajna Ramphal, Naid |
| hCoV-19/USA/WV-WVU-WV124649/2021 | EPI_ISL_6263378 | 2021-10-04 | Berkeley Medical Center | WVU and Marshall University Combined Genomics Core Facilities | James Denvir, Peter Stoilov, Peter Perrotta, Wesley Kimble, Ryan Percifield |
| hCoV-19/USA/WV-WVU-WV125251/2021 | EPI_ISL_6264000 | 2021-10-13 | Jefferson Medical Center | WVU and Marshall University Combined Genomics Core Facilities | James Denvir, Peter Stoilov, Peter Perrotta, Wesley Kimble, Ryan Percifield |
| hCoV-19/Australia/NSW-ICPMR-13298/2021 | EPI_ISL_6267930 | 2021-11-06 | Histopath | Clinical Pathology and Medical Research; Westmead Hospital; University of Sydney | Arnott A., Draper J., Gall M., Martinez E., Rockett R., Sintchenko V., on behalf of ICPMR |
| hCoV-19/senegal/SN-IR1-0071959/2021 | EPI_ISL_6274692 | 2021-10-13 | Iressef Genomics lab | IRESSEF | Souleymane MBOUP, Abdou PADANE , Khadim GUEYE, Diabou Diagne , Samba Ndiour, Papa Alassane DIAW, Birahim Piere NDIAYE, Barada CISSE, Aminata MBOUP, Moustapha MBOW, Ndeye Coumba Toure KANE, Nafisatou LEYE, Gora LO, Ambroise AHOUIDI , Astou Gaye GAYE, Amina |
| hCoV-19/senegal/SN-IR1-0074600/2021 | EPI_ISL_6274703 | 2021-10-28 | Iressef Genomics lab | IRESSEF | Souleymane MBOUP, Abdou PADANE , Khadim GUEYE, Diabou Diagne , Samba Ndiour, Papa Alassane DIAW, Birahim Piere NDIAYE, Barada CISSE, Aminata MBOUP, Moustapha MBOW, Ndeye Coumba Toure KANE, Nafisatou LEYE, Gora LO, Ambroise AHOUIDI , Astou Gaye GAYE, Amina |
| hCoV-19/senegal/SN-IR1-0074796/2021 | EPI_ISL_6274729 | 2021-10-28 | Iressef Genomics lab | IRESSEF | Souleymane MBOUP, Abdou PADANE , Khadim GUEYE, Diabou Diagne , Samba Ndiour, Papa Alassane DIAW, Birahim Piere NDIAYE, Barada CISSE, Aminata MBOUP, Moustapha MBOW, Ndeye Coumba Toure KANE, Nafisatou LEYE, Gora LO, Ambroise AHOUIDI , Astou Gaye GAYE, Amina |
| hCoV-19/senegal/SN-IR2-0040767/2021 | EPI_ISL_6274766 | 2021-11-02 | Iressef Genomics lab | IRESSEF | Souleymane MBOUP, Abdou PADANE , Khadim GUEYE, Diabou Diagne , Samba Ndiour, Papa Alassane DIAW, Birahim Piere NDIAYE, Barada CISSE, Aminata MBOUP, Moustapha MBOW, Ndeye Coumba Toure KANE, Nafisatou LEYE, Gora LO, Ambroise AHOUIDI , Astou Gaye GAYE, Amina |
| hCoV-19/England/PHEP-040529/2021 | EPI_ISL_6276768 | 2021-09-27 | Respiratory Virus Unit, Microbiology Services Colindale, Public Health England | COVID-19 Genomics UK (COG-UK) Consortium | PHE Covid Sequencing Team |
| hCoV-19/India/AS-INSACOG-1829500358569/2021 | EPI_ISL_5761113 | 2021-07-31 | INSACOG-Assam | National Institute of Biomedical Genomics ,Äì¬†INSACOG | Arindam Maitra, Ajanta Sharma, Nidhan Kumar Biswas, Lahari Saikia, Sreedhar Chinnaswamy,¬†Kailash Chamuah, Saumitra Das |

| | | | | | |
|---|---|---|---|---|---|
| hCoV-19/England/PHEP-YYB4BDD/2021 | EPI_ISL_6280020 | 2021-10-30 | Respiratory Virus Unit, Microbiology Services Colindale, Public Health England | COVID-19 Genomics UK (COG-UK) Consortium | PHE Covid Sequencing Team |
| hCoV-19/England/PHEP-YYBSI1A/2021 | EPI_ISL_6282942 | 2021-10-21 | Respiratory Virus Unit, Microbiology Services Colindale, Public Health England | COVID-19 Genomics UK (COG-UK) Consortium | PHE Covid Sequencing Team |
| hCoV-19/England/MILK-299BB0A/2021 | EPI_ISL_6300271 | 2021-11-06 | Lighthouse Lab in Milton Keynes | Wellcome Sanger Institute for the COVID-19 Genomics UK (COG-UK) Consortium | The Lighthouse Lab in Milton Keynes and Alex Alderton, Roberto Amato, Jeffrey Barrett, Sonia Goncalves, Ewan Harrison, David K. Jackson, Ian Johnston, Dominic Kwiatkowski, Cordelia Langford, John Sillitoe on behalf of the Wellcome Sanger Institute COVID-1 |
| hCoV-19/France/IDF-IPP31876/2021 | EPI_ISL_6315910 | 2021-10-25 | Labo Analyses Med | National Reference Center for Viruses of Respiratory Infections, Institut Pasteur, Paris | Marion Barbet, Sylvie Behillil, M√©line Bizard, Angela Brisebarre, Camille Capel, Vincent Enouf, Louise Lefran√ßois, Fr√©d√©ric Lemoine, Christophe Malabat, Corinne Maufrais, Slim El-Khiari, Julien Fumey, Etienne Simon-Lori√®re, Maud Vanpeene, Sylvie Van |
| hCoV-19/USA/MI-MDHHS-SC36172/2021 | EPI_ISL_6325724 | 2021-10-22 | Michigan Department of Health and Human Services, Bureau of Laboratories | Human Services, Bureau of Laboratories | Blankenship HM, Riner D, Soehnlen MK |
| hCoV-19/USA/NY-UB-CHS-05018/2021 | EPI_ISL_6327322 | 2021-08-29 | Catholic Health Laboratories | University at Buffalo Genomics and Bioinformatics Core | Jonathan Bard, Natalie Lamb, Alyssa Pohlman, Brandon Marzullo, Amanda Boccolucci, Norma Nowak, Donald Yergeau, Jennifer Surtees |
| hCoV-19/USA/NY-UB-KSL-00510/2021 | EPI_ISL_6327694 | 2021-08-31 | KSL Diagnostics | University at Buffalo Genomics and Bioinformatics Core | Jonathan Bard, Natalie Lamb, Alyssa Pohlman, Brandon Marzullo, Amanda Boccolucci, Norma Nowak, Donald Yergeau, Jennifer Surtees |
| hCoV-19/SouthAfrica/NICD-N14807/2021 | EPI_ISL_6327724 | 2021-08-06 | AMPATH | Diseases of the National Health Laboratory Service | Amoako DG, Everatt J, Scheepers C, Mohale T, Ntuli N, Mahlangu B, Mnguni A, Ismail A, Bhiman JN |
| hCoV-19/SouthAfrica/NICD-N20405/2021 | EPI_ISL_6327979 | 2021-10-11 | PORT ELIZABETH LABORATORY | Diseases of the National Health Laboratory Service | Amoako DG, Everatt J, Scheepers C, Mohale T, Ntuli N, Mahlangu B, Mnguni A, Ismail A, Bhiman JN |
| hCoV-19/Morocco/IPM20378282/2021 | EPI_ISL_6332507 | 2021-10-18 | Medical Virology and BSL3 Laboratory | Centre de S√©quen√ßage G√©nomique | EL Hamouchi Adil, Charoute Hicham , Bouzidi Aymane, Amalou Ghita, Harmak Houda, Anga Latifa, Faouzi Abdellah, Zemmouri Faouzia, El Oualid Abdelmjid, Nadifiyine Saloua, Zouheir Yassine, Omondi Francis Carey, Somda Soro Georgina Charlene, Abbad Anas, Chgour |
| hCoV-19/Uganda/UG727/2021 | EPI_ISL_6332920 | 2021-05-31 | MRC/UVRI & LSHTM Uganda¬†Research Unit , Rakai Health Sciences Program | MRC/UVRI & LSHTM Uganda¬†Research Unit , Rakai Health Sciences Program | Matthew Cotten Dan Lule Bugembe, My V.T. Phan, Charles Ssuuna, Ronald Moses Galiwango, Steven J Reynolds |
| hCoV-19/Uganda/UG969/2021 | EPI_ISL_6332965 | 2021-05-22 | MRC/UVRI & LSHTM Uganda¬†Research Unit , Rakai Health Sciences Program | MRC/UVRI & LSHTM Uganda¬†Research Unit , Rakai Health Sciences Program | Matthew Cotten Dan Lule Bugembe, My V.T. Phan, Charles Ssuuna, Ronald Moses Galiwango, Steven J Reynolds |
| hCoV-19/Guyana/GPR0359999/2021 | EPI_ISL_6342934 | 2021-09-07 | National Public Health Reference Laboratory, Ministry of Health, | Genomics and Proteomics Departament, Gorgas Memorial Institute For Health Studies | Claudia Gonzalez, Leyda Abrego, Marlene Castillo, Oris Chavarria, Jessica Gondola, Ambar Moreno, Alexander Martinez, Shemeeza Sonaram, Cyril Giddings, Horace Cox |
| hCoV-19/USA/CO-CDPHE-2101845927/2021 | EPI_ISL_5425937 | 2021-09-14 | Colorado Department of Public Health and Environment | Colorado Department of Public Health and Environment | Laura Bankers, Molly C. Hetherington-Rauth, Diana Ir, Alexandria Rossheim, Michael A. Martin, Mandy Waters, Shannon R. Matzinger, Sarah Elizabeth Totten, Emily A. Travanty |

| | | | | | |
|---|---|---|---|---|---|
| hCoV-19/Norway/24584/2021 | EPI_ISL_6347359 | 2021-01-21 | Haukeland University Hospital, Dept. of Microbiology | Norwegian Institute of Public Health, Department of Virology | Kathrine Stene-Johansen, Kamilla Heddeland Instefjord, Hilde Elshaug, Garcia Llorente Ignacio, Jon Br√•te, Engebretsen Serina Beate, Pedersen Benedikte Nevjen, Line Victoria Moen, Debech Nadia, Atiya R Ali, Marie Paulsen Madsen, Rasmus Riis Kopperud, Hild |
| hCoV-19/Ecuador/USFQ-2178/2021 | EPI_ISL_6367929 | 2021-10-04 | Institute of Microbiology, Universidad San Francisco de Quito | Institute of Microbiology, Universidad San Francisco de Quito | Erika B. Mu√±oz, Fernanda Zurita, Mateo Carvajal, Monica Becerra-Wong, Juan Jos√© Guadalupe, Sully M√°rquez, Bel√©n Prado-Vivar, Bernardo Guti√©rrez, Anaracelis Angarita, Ver√≥nica Barrag√°n, Patricio Rojas-Silva, Gabriel Trueba, Michelle Grunauer, Pa√∫l |
| hCoV-19/India/TG-CCMB-CIB0970/2021 | EPI_ISL_6368578 | 2021-09-06 | Osmania Medical College | CSIR-Centre for Cellular and Molecular Biology - INSACOG | Tulasi Nagabandi,Lamuk Zaveri,Payel Mukherjee,Onkar Kulkarni,Jandhyala Sai Krishna, Sofia Banu, Ara Sreenivas,Shreekant Verma, Amareshwar Vodapalli ,B Himasri,Valli Nagalakshmi Undamatla, Priya Nurkurthy,Sumedha Avadhanula, Vidhyadhari Methuku, Archana Bh |
| hCoV-19/Aruba/AW-RIVM-65005/2021 | EPI_ISL_5429780 | 2021-05-10 | Dutch COVID-19 response team | National Institute for Public Health and the Environment (RIVM) | Adam Meijer, Harry Vennema, Dirk Eggink, Jeroen Cremer, Sharon van den Brink, Bas van der Veer, AnneMarie van den Brandt, Lisa Wijsman, Kim Freriks, Ryanne Jaarsma, Eun√≠ce Then, Lynn Aarts, Sanne Bos, Stijn van Rossum, Florian Zwagemaker, Dennis Schmitz, |
| hCoV-19/Aruba/AW-RIVM-65028/2021 | EPI_ISL_5429787 | 2021-06-10 | Dutch COVID-19 response team | National Institute for Public Health and the Environment (RIVM) | Adam Meijer, Harry Vennema, Dirk Eggink, Jeroen Cremer, Sharon van den Brink, Bas van der Veer, AnneMarie van den Brandt, Lisa Wijsman, Kim Freriks, Ryanne Jaarsma, Eun√≠ce Then, Lynn Aarts, Sanne Bos, Stijn van Rossum, Florian Zwagemaker, Dennis Schmitz, |
| hCoV-19/Aruba/AW-RIVM-65070/2021 | EPI_ISL_5429802 | 2021-05-10 | Dutch COVID-19 response team | National Institute for Public Health and the Environment (RIVM) | Adam Meijer, Harry Vennema, Dirk Eggink, Jeroen Cremer, Sharon van den Brink, Bas van der Veer, AnneMarie van den Brandt, Lisa Wijsman, Kim Freriks, Ryanne Jaarsma, Eun√≠ce Then, Lynn Aarts, Sanne Bos, Stijn van Rossum, Florian Zwagemaker, Dennis Schmitz, |
| hCoV-19/Netherlands/UT-RIVM-64732/2021 | EPI_ISL_5430085 | 2021-10-05 | Dutch COVID-19 response team | National Institute for Public Health and the Environment (RIVM) | Adam Meijer, Harry Vennema, Dirk Eggink, Jeroen Cremer, Sharon van den Brink, Bas van der Veer, AnneMarie van den Brandt, Lisa Wijsman, Kim Freriks, Ryanne Jaarsma, Eun√≠ce Then, Lynn Aarts, Sanne Bos, Stijn van Rossum, Florian Zwagemaker, Dennis Schmitz, |
| hCoV-19/Netherlands/NH-RIVM-64503/2021 | EPI_ISL_5430223 | 2021-10-09 | Dutch COVID-19 response team | National Institute for Public Health and the Environment (RIVM) | Adam Meijer, Harry Vennema, Dirk Eggink, Jeroen Cremer, Sharon van den Brink, Bas van der Veer, AnneMarie van den Brandt, Lisa Wijsman, Kim Freriks, Ryanne Jaarsma, Eun√≠ce Then, Lynn Aarts, Sanne Bos, Stijn van Rossum, Florian Zwagemaker, Dennis Schmitz, |
| hCoV-19/Chile/RM-158957/2021 | EPI_ISL_5431489 | 2021-09-29 | Genetica Molecular and Subdepartamento de Virologia ISP Chile | Instituto de Salud Publica de Chile | Karen Orostica, Constanza Campano, Barbara Parra, Patricia Bustos, Rodrigo Fasce, Andres Castillo, Jorge Fernandez |

| | | | | | |
|---|---|---|---|---|---|
| hCoV-19/USA/NY-PRL-2021_0524_00D17/2021 | EPI_ISL_5438600 | 2021-05-22 | Pandemic Response Lab - NYC | Pandemic Response Lab, R&D | Henry Lee, Michael Hammerling, Melissa Hopkins, Cybill del Castillo, Shinyoung Clair Kang, William Ward, Pradeep Bugga, Sol Rey, Dylan Law, Alex Carpio, Isabel Fernandez Escapa, Haiping Hao, Jon Laurent |
| hCoV-19/Indonesia/JK-NIHRD-WGS11283/2021 | EPI_ISL_5441709 | 2021-08-13 | Lab Hamera Sarana Indonesia | National Institute of Health Research and Development | Vivi Setiawaty, Hana Apsari Pawestri, Subangkit, Kartika Dewi Puspa, Arie Ardiansyah Nugraha, Hartanti Dian Ikawati, Krisna Pangesti, Fajar Nur, Sulistiyohadi, Triyani Soekarso, Nelly Puspandari |
| hCoV-19/Cambodia/836160/2021 | EPI_ISL_5451644 | 2021-10-13 | Virology Unit, Institut Pasteur du Cambodge | Virology Unit, Institut Pasteur du Cambodge | Leakhena Pum, Sophoannadedh Rath, Jurre Y Siegers, Cecile Troupin, Ly Sovann, Kraing Sidonn, Yi Sengdoeurn, Chin Savuth, Chau Darapheak, Veasna Duong, Erik A Karlsson |
| hCoV-19/Denmark/DCGC-187504/2021 | EPI_ISL_5463476 | 2021-10-17 | Department of Bacteria, Parasites and Fungi, Statens Serum Institut, Copenhagen, Denmark | Statens Serum Institut Bioinformatics and Microbial Genomics | Danish Covid-19 Genome Consortium |
| hCoV-19/Reunion/HCL021182155901/2021 | EPI_ISL_5468576 | 2021-10-05 | CERBALLIANCE REUNION | CNR Virus des Infections Respiratoires - France SUD | Antonin Bal, Gregory Destras, Gwendolyne Burfin, Hadrien Regue, Quentin Semanas, Martine Valette, Bruno Lina, Laurence Josset |
| hCoV-19/India/WB-INSACOG-1931300468336/2021 | EPI_ISL_6391768 | 2021-09-19 | INSACOG-WB | National Institute of Biomedical Genomics ‚Äì INSACOG | Arindam Maitra, Bhaswati Bandyopadhyay, Nidhan Kumar Biswas, Tamal Ghosh, Sreedhar Chinnaswamy, Ajay Chakraborti, Saumitra Das |
| hCoV-19/India/WB-INSACOG-1931300468304/2021 | EPI_ISL_6391781 | 2021-09-19 | INSACOG-WB | National Institute of Biomedical Genomics ‚Äì INSACOG | Arindam Maitra, Bhaswati Bandyopadhyay, Nidhan Kumar Biswas, Tamal Ghosh, Sreedhar Chinnaswamy, Ajay Chakraborti, Saumitra Das |
| hCoV-19/USA/NC-CDC-LC0359749/2021 | EPI_ISL_6392161 | 2021-10-28 | Laboratory Corporation of America | Centers for Disease Control and Prevention Division of Viral Diseases, Pathogen Discovery | Dakota Howard,Dhwani Batra,Peter Cook,Jason Caravas,Benjamin Rambo-Martin,Scott Sammons,Yvette Unoarumhi,Matthew Schmerer,Kristine Lacek,Tymeckia Kendall,Victoria Caban Figueroa,Shatavia Morrison,Christopher Gulvick,Minoo Agarwal,Eyad Almasri,Debbie Boles |
| hCoV-19/USA/NY-CDC-LC0360098/2021 | EPI_ISL_6393033 | 2021-10-29 | Laboratory Corporation of America | Centers for Disease Control and Prevention Division of Viral Diseases, Pathogen Discovery | Dakota Howard,Dhwani Batra,Peter Cook,Jason Caravas,Benjamin Rambo-Martin,Scott Sammons,Yvette Unoarumhi,Matthew Schmerer,Kristine Lacek,Tymeckia Kendall,Victoria Caban Figueroa,Shatavia Morrison,Christopher Gulvick,Minoo Agarwal,Eyad Almasri,Debbie Boles |
| hCoV-19/India/SK-INSACOG-1122800013561/2021 | EPI_ISL_6393109 | 2021-09-11 | INSACOG-Sikkim | National Institute of Biomedical Genomics ‚Äì INSACOG | Arindam Maitra, Dr. Shrijana Gurung, Nidhan Kumar Biswas, Kaden Zangmu Bhutia, Sreedhar Chinnaswamy¬†Tshering Ongmu Bhutia, Saumitra Das |
| hCoV-19/India/MZ-INSACOG-1526100564008/2021 | EPI_ISL_6394162 | 2021-02-09 | INSACOG-Mizoram | National Institute of Biomedical Genomics ‚Äì INSACOG | Arindam Maitra, Swagnik Roy, Nidhan Kumar Biswas, Gracy Laldinmawii, Sreedhar Chinnaswamy, N Senthil Kumar, Saumitra Das |
| hCoV-19/India/WB-INSACOG-1931300460631/2021 | EPI_ISL_6395867 | 2021-09-09 | INSACOG-WB | National Institute of Biomedical Genomics ‚Äì¬†INSACOG | Arindam Maitra, Bhaswati Bandyopadhyay, Nidhan Kumar Biswas, Tamal Ghosh, Sreedhar Chinnaswamy, Ajay Chakraborti, Saumitra Das |

| | | | | | |
|---|---|---|---|---|---|
| hCoV-19/SouthAfrica/NICD-N17091/2021 | EPI_ISL_5477292 | 2021-06-10 | Glencore Western chrome mines | Diseases of the National Health Laboratory Service | Amoako DG, Everatt J, Scheepers C, Mohale T, Ntuli N, Mahlangu B, Mnguni A, Ismail A, Bhiman JN |
| hCoV-19/England/PHEC-3L04AL56/2021 | EPI_ISL_5479908 | 2021-10-13 | Respiratory Virus Unit, Microbiology Services Colindale, Public Health England | COVID-19 Genomics UK (COG-UK) Consortium | PHE Covid Sequencing Team |
| hCoV-19/Scotland/QEUH-27885D7/2021 | EPI_ISL_5482856 | 2021-10-16 | Lighthouse Lab in Glasgow | Wellcome Sanger Institute for the COVID-19 Genomics UK (COG-UK) Consortium | Harper VanSteenhouse, Yumi Kasai, David Gray, Carol Clugston, Anna Dominiczak and Alex Alderton, Roberto Amato, Jeffrey Barrett, Sonia Goncalves, Ewan Harrison, David K. Jackson, Ian Johnston, Dominic Kwiatkowski, Cordelia Langford, John Sillitoe on behal |
| hCoV-19/England/PHEP-YYBE15G/2021 | EPI_ISL_5485116 | 2021-10-10 | Respiratory Virus Unit, Microbiology Services Colindale, Public Health England | COVID-19 Genomics UK (COG-UK) Consortium | PHE Covid Sequencing Team |
| hCoV-19/England/PHEP-YYBFY74/2021 | EPI_ISL_5487183 | 2021-10-10 | Respiratory Virus Unit, Microbiology Services Colindale, Public Health England | COVID-19 Genomics UK (COG-UK) Consortium | PHE Covid Sequencing Team |
| hCoV-19/USA/AR-UMGC-17007/2021 | EPI_ISL_6404403 | 2021-09-14 | Arkansas Public Health Laboratory, Arkansas Department of Health | University of Minnesota Genomics Center | Daryl M. Gohl, John Garbe, Jaquelyn Kuriger-Laber, Corbin Dirkx |
| hCoV-19/USA/AR-UMGC-17682/2021 | EPI_ISL_6404732 | 2021-09-16 | Arkansas Public Health Laboratory, Arkansas Department of Health | University of Minnesota Genomics Center | Daryl M. Gohl, John Garbe, Jaquelyn Kuriger-Laber, Corbin Dirkx |
| hCoV-19/USA/AR-UMGC-17716/2021 | EPI_ISL_6404783 | 2021-09-20 | Arkansas Public Health Laboratory, Arkansas Department of Health | University of Minnesota Genomics Center | Daryl M. Gohl, John Garbe, Jaquelyn Kuriger-Laber, Corbin Dirkx |
| hCoV-19/Brazil/AM-FIOCRUZ-ILMD2100730/2021 | EPI_ISL_5490905 | 2021-09-23 | Laboratorio Central de Saude Publica do Amazonas - LACEN-AM | Laboratorio de Ecologia de Doencas Transmissiveis na Amazonia, Instituto Leonidas e Maria Deane - Fiocruz Amazonia | Arlesson Viana Da Silva, Valdinete Nascimento, Victor Souza, Andr√© Corado, Fernanda Nascimento, George Silva, Matilde Mej√≠a, Luciana Gon√ßalves, Maria J√∫lia Brand√£o, Felipe Naveca |
| hCoV-19/USA/MA-CDC-STM-CXX3TDTBW/2021 | EPI_ISL_6406616 | 2021-10-20 | Helix | Centers for Disease Control and Prevention Division of Viral Diseases, Pathogen Discovery | Dakota Howard,Dhwani Batra,Peter Cook,Jason Caravas,Benjamin Rambo-Martin,Scott Sammons,Yvette Unoarumhi,Matthew Schmerer,Kristine Lacek,Tymeckia Kendall,Victoria Caban Figueroa,Helix CA,Shatavia Morrison,Christopher Gulvick,Erisa Sula,Clinton Paden,Dunca |
| hCoV-19/USA/MN-UMGC-18081/2021 | EPI_ISL_6407148 | 2021-09-24 | Public Health Laboratory, Minnesota Department of Health | University of Minnesota Genomics Center | Daryl M. Gohl, John Garbe, Jaquelyn Kuriger-Laber, Corbin Dirkx |
| hCoV-19/Morocco/IPM20361572/2021 | EPI_ISL_5501117 | 2021-09-08 | Medical Virology and BSL3 Laboratory | Centre de S√©quen√ßage G√©nomique | EL Hamouchi Adil, Charoute Hicham , Bouzidi Aymane, Amalou Ghita, Harmak Houda, Anga Latifa, Faouzi Abdellah, Zemmouri Faouzia, El Oualid Abdelmjid, Nadifiyine Saloua, Zouheir Yassine, Omondi Francis Carey, Somda Soro Georgina Charlene, Abbad Anas, Chgour |
| hCoV-19/USA/MN-UMGC-16696/2021 | EPI_ISL_6410351 | 2021-09-09 | Public Health Laboratory, Minnesota Department of Health | University of Minnesota Genomics Center | Daryl M. Gohl, John Garbe, Jaquelyn Kuriger-Laber, Corbin Dirkx |
| hCoV-19/Morocco/IPM20361246/2021 | EPI_ISL_5501095 | 2021-09-07 | Medical Virology and BSL3 Laboratory | Centre de S√©quen√ßage G√©nomique | EL Hamouchi Adil, Charoute Hicham , Bouzidi Aymane, Amalou Ghita, Harmak Houda, Anga Latifa, Faouzi Abdellah, Zemmouri Faouzia, El Oualid Abdelmjid, Nadifiyine Saloua, Zouheir Yassine, Omondi Francis Carey, Somda Soro Georgina Charlene, Abbad Anas, Chgour |

| | | | | | |
|---|---|---|---|---|---|
| hCoV-19/USA/MN-UMGC-17475/2021 | EPI_ISL_6411026 | 2021-09-15 | Public Health Laboratory, Minnesota Department of Health | University of Minnesota Genomics Center | Daryl M. Gohl, John Garbe, Jaquelyn Kuriger-Laber, Corbin Dirkx |
| hCoV-19/USA/MN-UMGC-17550/2021 | EPI_ISL_6411166 | 2021-09-15 | Public Health Laboratory, Minnesota Department of Health | University of Minnesota Genomics Center | Daryl M. Gohl, John Garbe, Jaquelyn Kuriger-Laber, Corbin Dirkx |
| hCoV-19/USA/MI-CDC-STM-UA3VSYFZA/2021 | EPI_ISL_6415737 | 2021-10-28 | Helix | Centers for Disease Control and Prevention Division of Viral Diseases, Pathogen Discovery | Dakota Howard,Dhwani Batra,Peter Cook,Jason Caravas,Benjamin Rambo-Martin,Scott Sammons,Yvette Unoarumhi,Matthew Schmerer,Kristine Lacek,Tymeckia Kendall,Victoria Caban Figueroa,Helix CA,Shatavia Morrison,Christopher Gulvick,Erisa Sula,Clinton Paden,Dunca |
| hCoV-19/USA/PA-CDC-ASC210499139/2021 | EPI_ISL_6420034 | 2021-10-26 | Aegis Sciences Corporation | Centers for Disease Control and Prevention Division of Viral Diseases, Pathogen Discovery | Dakota Howard,Dhwani Batra,Peter Cook,Jason Caravas,Benjamin Rambo-Martin,Scott Sammons,Yvette Unoarumhi,Matthew Schmerer,Kristine Lacek,Tymeckia Kendall,Victoria Caban Figueroa,Shatavia Morrison,Christopher Gulvick,Erisa Sula,Cyndi Clark,Patrick Campbell |
| hCoV-19/Angola/CERI-KRISP-K026875/2021 | EPI_ISL_6422248 | 2021-09-14 | Instituto Nacional de Investiga√ß√£o em Sa√∫de | and Innvoation, Stellenbosch University and KRISP, KZN Research Innovation and Sequencing Platform, UKZN. | Morais J, Neto Z, Afonso P, Miranda J, David K, Ingl√™s L, Pereira A, Paulo A Carralero RR Paix√£o JP, Freitas RH, Mufinda M, Lutucuta S, Giandhari J, Pillay S, Naidoo Y, Emmanuel SJ, Tegally H, Wilkinson E, de Oliveira T |
| hCoV-19/Cameroon/CERI-KRISP-K026922/2021 | EPI_ISL_6422286 | 2021-08-31 | National Public Health Laboratory, Ministry of Public Health, | and Innvoation, Stellenbosch University and KRISP, KZN Research Innovation and Sequencing Platform, UKZN. | Dr Essomba Rene Ghislain, Pr Okomo Assoumou Marie Claire, Dr Alle Baba Dieng, Ibrahima Halilou and Dr Etoundi Mballa Alain, Giandhari J, Pillay S, Naidoo Y, Emmanuel SJ, Tegally H, Wilkinson E, de Oliveira T |
| hCoV-19/Cameroon/CERI-KRISP-K026996/2021 | EPI_ISL_6422325 | 2021-09-05 | National Public Health Laboratory, Ministry of Public Health, | and Innvoation, Stellenbosch University and KRISP, KZN Research Innovation and Sequencing Platform, UKZN. | Dr Essomba Rene Ghislain, Pr Okomo Assoumou Marie Claire, Dr Alle Baba Dieng, Ibrahima Halilou and Dr Etoundi Mballa Alain, Giandhari J, Pillay S, Naidoo Y, Emmanuel SJ, Tegally H, Wilkinson E, de Oliveira T |
| hCoV-19/Cameroon/CERI-KRISP-K027014/2021 | EPI_ISL_6422335 | 2021-09-24 | National Public Health Laboratory, Ministry of Public Health, | and Innvoation, Stellenbosch University and KRISP, KZN Research Innovation and Sequencing Platform, UKZN. | Dr Essomba Rene Ghislain, Pr Okomo Assoumou Marie Claire, Dr Alle Baba Dieng, Ibrahima Halilou and Dr Etoundi Mballa Alain, Giandhari J, Pillay S, Naidoo Y, Emmanuel SJ, Tegally H, Wilkinson E, de Oliveira T |
| hCoV-19/Cameroon/CERI-KRISP-K027015/2021 | EPI_ISL_6422336 | 2021-09-24 | National Public Health Laboratory, Ministry of Public Health, | and Innvoation, Stellenbosch University and KRISP, KZN Research Innovation and Sequencing Platform, UKZN. | Dr Essomba Rene Ghislain, Pr Okomo Assoumou Marie Claire, Dr Alle Baba Dieng, Ibrahima Halilou and Dr Etoundi Mballa Alain, Giandhari J, Pillay S, Naidoo Y, Emmanuel SJ, Tegally H, Wilkinson E, de Oliveira T |
| hCoV-19/Malawi/CERI-KRISP-K027018/2021 | EPI_ISL_6422342 | 2021-09-09 | National HIV Reference Laboratory, Ministry of Health, Public Health Institute of Malawi | and Innvoation, Stellenbosch University and KRISP, KZN Research Innovation and Sequencing Platform, UKZN. | Mvula B, Chilima B, Chiwaula M, Mwangomba W, Panja L, Kasambara W, Auld A, Kim L, Kampira E, Kaba M, Wadonda N, Maida A, Giandhari J, Pillay S, Naidoo Y, Emmanuel SJ, Tegally H, Wilkinson E, de Oliveira T |
| hCoV-19/England/ALDP-2A223C3/2021 | EPI_ISL_6431609 | 2021-11-10 | Lighthouse Lab in Alderley Park | Wellcome Sanger Institute for the COVID-19 Genomics UK (COG-UK) Consortium | Jacquelyn Wynn, Mairead Hyland, The Lighthouse Lab in Alderley Park and Alex Alderton, Roberto Amato, Jeffrey Barrett, Sonia Goncalves, Ewan Harrison, David K. Jackson, Ian Johnston, Dominic Kwiatkowski, Cordelia Langford, John Sillitoe on behalf of the W |

| | | | | | |
|---|---|---|---|---|---|
| hCoV-19/SouthAfrica/NHLS-UCT-LA-Z724/2021 | EPI_ISL_6436407 | 2021-10-12 | Lancet | NHLS/UCT | Arash Iranzadeh, Deelan Doolabh, Lynn Tyers, Bruna Galvao, Innocent Mudau, Marvin Hsiao, Gert Marais, Diana Hardie, Stephen Korsman, Rageema Joseph, Carolyn Williamson |
| hCoV-19/SouthAfrica/NHLS-UCT-GS-AE83/2021 | EPI_ISL_6436450 | 2021-10-21 | Oudtshoorn Sick Bay (SANDF) | NHLS/UCT | Arash Iranzadeh, Deelan Doolabh, Lynn Tyers, Bruna Galvao, Innocent Mudau, Marvin Hsiao, Gert Marais, Diana Hardie, Stephen Korsman, Rageema Joseph, Carolyn Williamson |
| hCoV-19/USA/MA-CDCBI-CRSP_JUFIIPYUOG2BUWZA/2021 | EPI_ISL_6447711 | 2021-11-06 | Broad Institute Clinical Research Sequencing Platform | Infectious Disease Program, Broad Institute of Harvard and MIT | Siddle,K.J., Adams,G., Pearlman,L., Gladden-Young,A., Vicente,G., Blumenstiel,B., DeFelice,M., Lee,M., McGovern,S., Lagerborg,K., Rudy,M., DeRuff,K., Carter,A., Normandin,E., Bauer,M., Reilly,S., Tomkins-Tinch,C., Loreth,C., Chaluvadi,S., Meldrim,J., Gran |
| hCoV-19/PapuaNewGuinea/PNG6076/2021 | EPI_ISL_6450123 | 2021-09-28 | Central Public Health Laboratory | Microbiological Diagnostic Unit - Public Health Laboratory (MDU-PHL) | Kumbu, J., Susapu, M., Palou, T., Seemann, T., Horan, K., Sherry, N.L. |
| hCoV-19/PapuaNewGuinea/PNG4034/2021 | EPI_ISL_6450588 | 2021-09-14 | Central Public Health Laboratory | Microbiological Diagnostic Unit - Public Health Laboratory (MDU-PHL) | Kumbu, J., Susapu, M., Palou, T., Seemann, T., Horan, K., Sherry, N.L. |
| hCoV-19/PapuaNewGuinea/PNG5137/2021 | EPI_ISL_6450609 | 2021-09-28 | Central Public Health Laboratory | Microbiological Diagnostic Unit - Public Health Laboratory (MDU-PHL) | Kumbu, J., Susapu, M., Palou, T., Seemann, T., Horan, K., Sherry, N.L. |
| hCoV-19/PapuaNewGuinea/PNG4074/2021 | EPI_ISL_6450754 | 2021-08-17 | Central Public Health Laboratory | Microbiological Diagnostic Unit - Public Health Laboratory (MDU-PHL) | Kumbu, J., Susapu, M., Palou, T., Seemann, T., Horan, K., Sherry, N.L. |
| hCoV-19/PapuaNewGuinea/PNG5782/2021 | EPI_ISL_6450909 | 2021-09-24 | Central Public Health Laboratory | Microbiological Diagnostic Unit - Public Health Laboratory (MDU-PHL) | Kumbu, J., Susapu, M., Palou, T., Seemann, T., Horan, K., Sherry, N.L. |
| hCoV-19/PapuaNewGuinea/PNG4483/2021 | EPI_ISL_6450934 | 2021-08-30 | Central Public Health Laboratory | Microbiological Diagnostic Unit - Public Health Laboratory (MDU-PHL) | Kumbu, J., Susapu, M., Palou, T., Seemann, T., Horan, K., Sherry, N.L. |
| hCoV-19/PapuaNewGuinea/PNG3817/2021 | EPI_ISL_6452123 | 2021-08-30 | Central Public Health Laboratory | Microbiological Diagnostic Unit - Public Health Laboratory (MDU-PHL) | Kumbu, J., Susapu, M., Palou, T., Seemann, T., Horan, K., Sherry, N.L. |
| hCoV-19/PapuaNewGuinea/PNG5843/2021 | EPI_ISL_6452345 | 2021-09-26 | Central Public Health Laboratory | Microbiological Diagnostic Unit - Public Health Laboratory (MDU-PHL) | Kumbu, J., Susapu, M., Palou, T., Seemann, T., Horan, K., Sherry, N.L. |
| hCoV-19/PapuaNewGuinea/PNG4447/2021 | EPI_ISL_6452491 | 2021-08-28 | Central Public Health Laboratory | Microbiological Diagnostic Unit - Public Health Laboratory (MDU-PHL) | Kumbu, J., Susapu, M., Palou, T., Seemann, T., Horan, K., Sherry, N.L. |
| hCoV-19/Chile/AN-168219/2021 | EPI_ISL_6453392 | 2021-10-14 | Genetica Molecular and Subdepartamento de Virologia ISP Chile | Instituto de Salud Publica de Chile | Karen Orostica, Constanza Campano, Barbara Parra, Patricia Bustos, Rodrigo Fasce, Andres Castillo, Jorge Fernandez |
| hCoV-19/Chile/AI-173598/2021 | EPI_ISL_6454035 | 2021-10-25 | Genetica Molecular and Subdepartamento de Virologia ISP Chile | Instituto de Salud Publica de Chile | Karen Orostica, Constanza Campano, Barbara Parra, Patricia Bustos, Rodrigo Fasce, Andres Castillo, Jorge Fernandez |

| | | | | | |
|---|---|---|---|---|---|
| hCoV-19/Chile/BI-157980/2021 | EPI_ISL_6455099 | 2021-09-24 | Genetica Molecular and Subdepartamento de Virologia ISP Chile | Instituto de Salud Publica de Chile | Karen Orostica, Constanza Campano, Barbara Parra, Patricia Bustos, Rodrigo Fasce, Andres Castillo, Jorge Fernandez |
| hCoV-19/Chile/LI-175684/2021 | EPI_ISL_6456433 | 2021-10-30 | Genetica Molecular and Subdepartamento de Virologia ISP Chile | Instituto de Salud Publica de Chile | Karen Orostica, Constanza Campano, Barbara Parra, Patricia Bustos, Rodrigo Fasce, Andres Castillo, Jorge Fernandez |
| hCoV-19/PapuaNewGuinea/PNG4040/2020 | EPI_ISL_6462933 | 2020-08-01 | Central Public Health Laboratory | Microbiological Diagnostic Unit - Public Health Laboratory (MDU-PHL) | Kumbu, J., Susapu, M., Palou, T., Seemann, T., Horan, K., Sherry, N.L. |
| hCoV-19/PapuaNewGuinea/PNG2080/2021 | EPI_ISL_6463323 | 2021-05-07 | Queensland Medical Laboratories | Microbiological Diagnostic Unit - Public Health Laboratory (MDU-PHL) | Susapu, M., Palou, T., Seemann, T., Horan, K., Sherry, N.L. |
| hCoV-19/PapuaNewGuinea/PNG1094/2021 | EPI_ISL_6463338 | 2021-03-31 | Queensland Medical Laboratories | Microbiological Diagnostic Unit - Public Health Laboratory (MDU-PHL) | Susapu, M., Palou, T., Seemann, T., Horan, K., Sherry, N.L. |
| hCoV-19/PapuaNewGuinea/PNG1103/2021 | EPI_ISL_6463589 | 2021-03-31 | Queensland Medical Laboratories | Microbiological Diagnostic Unit - Public Health Laboratory (MDU-PHL) | Susapu, M., Palou, T., Seemann, T., Horan, K., Sherry, N.L. |
| hCoV-19/Lithuania/MR-LUHS-25-08/2021 | EPI_ISL_6468529 | 2021-11-05 | UAB Medicina practica laboratorija | Lithuanian University of Health Sciences | Arnoldas Pautienius, Dovydas Gecys, Arnoldas Pautienius, Emilija Gecyte, Gediminas Alzbutas, Lukas Zemaitis, Vaiva Lesauskaite |
| hCoV-19/Gambia/13449/2021 | EPI_ISL_6471592 | 2021-03-16 | MRCG at LSHTM Genomics lab | MRCG at LSHTM Genomics lab | Abdul Karim sesay, Abdoulie Kanteh, Jarra Manneh, Mariama Kujabi, Bakary Sanyang,Dabiri Damilari,Yaya Bah |
| hCoV-19/Gambia/PF6266/2021 | EPI_ISL_6471601 | 2021-02-08 | MRCG at LSHTM Genomics lab | MRCG at LSHTM Genomics lab | Abdul Karim sesay, Abdoulie Kanteh, Jarra Manneh, Mariama Kujabi, Bakary Sanyang,Dabiri Damilari,Yaya Bah |
| hCoV-19/Gambia/PF0203/2021 | EPI_ISL_6471602 | 2021-02-08 | MRCG at LSHTM Genomics lab | MRCG at LSHTM Genomics lab | Abdul Karim sesay, Abdoulie Kanteh, Jarra Manneh, Mariama Kujabi, Bakary Sanyang,Dabiri Damilari,Yaya Bah |
| hCoV-19/Gambia/134114/2021 | EPI_ISL_6471609 | 2021-04-08 | MRCG at LSHTM Genomics lab | MRCG at LSHTM Genomics lab | Abdul Karim sesay, Abdoulie Kanteh, Jarra Manneh, Mariama Kujabi, Bakary Sanyang,Dabiri Damilari,Yaya Bah |
| hCoV-19/Brazil/CE-FIOCRUZ-26033/2021 | EPI_ISL_6474139 | 2021-09-28 | LACEN LABORATORIO DE SAUDE PUBLICA | ACME Lab, Oswaldo Cruz Foundation, FIOCRUZ/CE | Fabio Miyajima, Veridiana Pessoa Miyajima, Fernando Braga Stehling, Thais de Oliveira Costa, Thais Ferreira de Oliveira, Joaquim Cesar do Nascimento Sousa Junior, Jamille Maria Mendes Bezerra, Ticiane Cavalcante de Souza, Carlos Leonardo de Aragao Araujo, |
| hCoV-19/USA/LA-CDC-STM-000724781/2021 | EPI_ISL_6476578 | 2021-08-23 | Helix | Centers for Disease Control and Prevention Division of Viral Diseases, Pathogen Discovery | Dakota Howard,Dhwani Batra,Peter Cook,Jason Caravas,Benjamin Rambo-Martin,Scott Sammons,Yvette Unoarumhi,Matthew Schmerer,Kristine Lacek,Tymeckia Kendall,Victoria Caban Figueroa,Helix CA,Shatavia Morrison,Christopher Gulvick,Erisa Sula,Clinton Paden,Dunca |

| | | | | | |
|---|---|---|---|---|---|
| hCoV-19/USA/FL-CDC-STM-000612239/2021 | EPI_ISL_6484557 | 2021-07-06 | Helix | Centers for Disease Control and Prevention Division of Viral Diseases, Pathogen Discovery | Dakota Howard,Dhwani Batra,Peter Cook,Jason Caravas,Benjamin Rambo-Martin,Scott Sammons,Yvette Unoarumhi,Matthew Schmerer,Kristine Lacek,Tymeckia Kendall,Victoria Caban Figueroa,Helix CA,Shatavia Morrison,Christopher Gulvick,Erisa Sula,Clinton Paden,Dunca |
| hCoV-19/USA/AL-CDC-STM-000717563/2021 | EPI_ISL_6486223 | 2021-08-06 | Helix | Centers for Disease Control and Prevention Division of Viral Diseases, Pathogen Discovery | Dakota Howard,Dhwani Batra,Peter Cook,Jason Caravas,Benjamin Rambo-Martin,Scott Sammons,Yvette Unoarumhi,Matthew Schmerer,Kristine Lacek,Tymeckia Kendall,Victoria Caban Figueroa,Helix CA,Shatavia Morrison,Christopher Gulvick,Erisa Sula,Clinton Paden,Dunca |
| hCoV-19/USA/FL-CDC-STM-000710357/2021 | EPI_ISL_6486506 | 2021-07-22 | Helix | Centers for Disease Control and Prevention Division of Viral Diseases, Pathogen Discovery | Dakota Howard,Dhwani Batra,Peter Cook,Jason Caravas,Benjamin Rambo-Martin,Scott Sammons,Yvette Unoarumhi,Matthew Schmerer,Kristine Lacek,Tymeckia Kendall,Victoria Caban Figueroa,Helix CA,Shatavia Morrison,Christopher Gulvick,Erisa Sula,Clinton Paden,Dunca |
| hCoV-19/USA/OH-CDC-STM-000711522/2021 | EPI_ISL_6486873 | 2021-07-26 | Helix | Centers for Disease Control and Prevention Division of Viral Diseases, Pathogen Discovery | Dakota Howard,Dhwani Batra,Peter Cook,Jason Caravas,Benjamin Rambo-Martin,Scott Sammons,Yvette Unoarumhi,Matthew Schmerer,Kristine Lacek,Tymeckia Kendall,Victoria Caban Figueroa,Helix CA,Shatavia Morrison,Christopher Gulvick,Erisa Sula,Clinton Paden,Dunca |
| hCoV-19/USA/CA-COLOR-UCSF-CC890/2021 | EPI_ISL_5323475 | 2021-05-13 | Color Genomics | Chiu Laboratory, University of California, San Francisco | Charles Chiu, Venice Servellita, Alicia Sotomayor-Gonzalez, Baolin Wang, Scott Topper, Erika Torres, Noah Brazer, Daniel Wong, Alicia Zhou |
| hCoV-19/USA/FL-CDC-STM-000716044/2021 | EPI_ISL_6487242 | 2021-08-01 | Helix | Centers for Disease Control and Prevention Division of Viral Diseases, Pathogen Discovery | Dakota Howard,Dhwani Batra,Peter Cook,Jason Caravas,Benjamin Rambo-Martin,Scott Sammons,Yvette Unoarumhi,Matthew Schmerer,Kristine Lacek,Tymeckia Kendall,Victoria Caban Figueroa,Helix CA,Shatavia Morrison,Christopher Gulvick,Erisa Sula,Clinton Paden,Dunca |
| hCoV-19/CzechRepublic/UKDD-ZUUL-D889/2021 | EPI_ISL_6491326 | 2021-11-01 | Zdravotn√≠ √∫stav √öst√≠ nad Labem | Institute of Medical Microbiology and Virology, University Hospital Carl Gustav Carus, TU Dresden | Marlena Stadtm√ºller, Alexa Laubner, Fabian Rost, Sylvia Klemroth, Grit Mehnert, Montserrat Palau de Miquel, Susanne Reinhardt, Anett Zabzinski, Sylke Winkler, Alexander Dalpke, Romana Mike≈°ov√°, Jitka Poho≈ôsk√°, Ivana Stiborov√°, Eva Patrasov√°, Lenka |
| hCoV-19/USA/ND-NDDH-6337/2021 | EPI_ISL_6491481 | 2021-09-12 | North Dakota Department of Health, Public Health Laboratory | North Dakota Department of Health, Public Health Laboratory | Lisa Wingerter |
| hCoV-19/Brazil/MG-LBI812/2021 | EPI_ISL_6499901 | 2021-08-15 | Laborat√≥rio de Biologia Integrativa/ UFMG | Laborat√≥rio de Biologia Integrativa/ UFMG | Paula Luize Camargos Fonseca, Filipe Romero Rebello Moreira, Rafael Marques de Souza, Nat√°lia Rocha Guimar√£es, Nara Oliveira Carvalho, Talita Emile Ribeiro Adelino, Hugo Jos√© Alves, Luige Biciati Alvim, Alana Vitor Barbosa Costa, Walyson Coelho Costa, |

| | | | | | |
|---|---|---|---|---|---|
| hCoV-19/Brazil/MG-LBI810/2021 | EPI_ISL_6499995 | 2021-08-16 | Laboratv≥rio de Biologia Integrativa/ UFMG | Laboratv≥rio de Biologia Integrativa/ UFMG | Paula Luize Camargos Fonseca, Filipe Romero Rebello Moreira, Rafael Marques de Souza, Natv°lia Rocha Guimarv£es, Nara Oliveira Carvalho, Talita Emile Ribeiro Adelino, Hugo Josv© Alves, Luige Biciati Alvim, Alana Vitor Barbosa Costa, Walyson Coelho Costa, |
| hCoV-19/Mexico/CMX-INMEGEN-36-127/2021 | EPI_ISL_6507879 | 2021-11-01 | Salud Digna | Instituto Nacional de Medicina Genomica | Cedro-Tanda A, Escobar-Arrazola MA, Ramirez-Vega O, Cruz-Islas Jazmin, Garnica-Lopez Dora, Abraham Campos-Romero, Moreno-Camacho Josv© Luis, Rodriguez-Gallegos Jorge, Luna-Ruiz Marco, Rangel-DeLeon D, Yair Alfaro-Mora, Hidalgo-Miranda A, Mendoza-Vargas A |
| hCoV-19/Brazil/SP-IB_137798/2021 | EPI_ISL_6508964 | 2021-10-11 | SECRETARIA MUNICIPAL DA SAUDE DE ITAPEVA SP | Instituto Butantan | Dimas Tadeu Covas, Antonio Jorge Martins, Claudia Renata dos Santos Barros, David Schlesinger, Debora Botequio Moretti, Elaine Cristina Marqueze, Elaine Vieira Santos, Evandra Strazza Rodrigues, Heidge Fukumasu, Jayme Augusto de Souza-Neto, Josv© Salvator |
| hCoV-19/Brazil/SP-IB_138730/2021 | EPI_ISL_6509886 | 2021-10-17 | CENTRO DE ATENDIMENTO PARA ENFRENTAMENTO A COVID19 DE APARECIDA | Instituto Butantan | Dimas Tadeu Covas, Antonio Jorge Martins, Claudia Renata dos Santos Barros, David Schlesinger, Debora Botequio Moretti, Elaine Cristina Marqueze, Elaine Vieira Santos, Evandra Strazza Rodrigues, Heidge Fukumasu, Jayme Augusto de Souza-Neto, Josv© Salvator |
| hCoV-19/Brazil/SP-IB_138077/2021 | EPI_ISL_6510487 | 2021-10-18 | UNIDADE DE PRONTO ATENDIMENTO UPA SUL CARAGUATATUBA | Instituto Butantan | Dimas Tadeu Covas, Antonio Jorge Martins, Claudia Renata dos Santos Barros, David Schlesinger, Debora Botequio Moretti, Elaine Cristina Marqueze, Elaine Vieira Santos, Evandra Strazza Rodrigues, Heidge Fukumasu, Jayme Augusto de Souza-Neto, Josv© Salvator |
| hCoV-19/Kenya/SS2930/2021 | EPI_ISL_6511025 | 2021-01-18 | Aga Khan University Hospital | KEMRI-Wellcome Trust Research Programme,Kilifi | Githinji G.,Ochola I., Agoti C.,Mohamed K.S., de Laurent Z., Mburu M.W., Morobe J., Ndwiga L. Makori T., Ongera E., Lambisia A.Shah R.,Revathi G.,Sayed S., Adam R.Nokes D.J. |
| hCoV-19/Angola/CERI-KRISP-K027373/2021 | EPI_ISL_6511808 | 2021-09-07 | Instituto Nacional de Investigavßv£o em Sav∫de | CERI, Centre for Epidemic Response and Innvoation, Stellenbosch University and KRISP, KZN Research Innovation and Sequencing Platform, UKZN. | Morais J, Neto Z, Afonso P, Miranda J, David K, Inglv™s L, Pereira A, Paulo A Carralero RR Paixv£o JP, Freitas RH, Mufinda M, Lutucuta S,Giandhari J, Pillay S, Naidoo Y, Ramphal U, Ramphal Y, Moir M, San JE, Tshiabuila D, Tegally H, Wilkinson E, de Olivei |
| hCoV-19/Angola/CERI-KRISP-K027411/2021 | EPI_ISL_6511827 | 2021-09-18 | Instituto Nacional de Investigavßv£o em Sav∫de | CERI, Centre for Epidemic Response and Innvoation, Stellenbosch University and KRISP, KZN Research Innovation and Sequencing Platform, UKZN. | Morais J, Neto Z, Afonso P, Miranda J, David K, Inglv™s L, Pereira A, Paulo A Carralero RR Paixv£o JP, Freitas RH, Mufinda M, Lutucuta S,Giandhari J, Pillay S, Naidoo Y, Ramphal U, Ramphal Y, Moir M, San JE, Tshiabuila D, Tegally H, Wilkinson E, de Olivei |
| hCoV-19/USA/TN-CDC-ASC210463103/2021 | EPI_ISL_6515373 | 2021-10-15 | Aegis Sciences Corporation | Centers for Disease Control and Prevention Division of Viral Diseases, Pathogen Discovery | Dakota Howard,Dhwani Batra,Peter Cook,Jason Caravas,Benjamin Rambo-Martin,Scott Sammons,Yvette Unoarumhi,Matthew Schmerer,Kristine Lacek,Tymeckia Kendall,Victoria Caban Figueroa,Shatavia Morrison,Christopher Gulvick,Erisa Sula,Cyndi Clark,Patrick Campbell |

| | | | | | |
|---|---|---|---|---|---|
| hCoV-19/USA/CA-CDC-ASC210468235/2021 | EPI_ISL_6524937 | 2021-10-18 | Aegis Sciences Corporation | Centers for Disease Control and Prevention Division of Viral Diseases, Pathogen Discovery | Dakota Howard,Dhwani Batra,Peter Cook,Jason Caravas,Benjamin Rambo-Martin,Scott Sammons,Yvette Unoarumhi,Matthew Schmerer,Kristine Lacek,Tymeckia Kendall,Victoria Caban Figueroa,Shatavia Morrison,Christopher Gulvick,Erisa Sula,Cyndi Clark,Patrick Campbell |
| hCoV-19/Ecuador/NIC-INSPI-356170/2021 | EPI_ISL_6526332 | 2021-11-08 | INSPI-CRN DE INFLUENZA Y OTROS VIRUS RESPIRATORIOS | INSPI-CRN DE INFLUENZA Y OTROS VIRUS RESPIRATORIOS | Alfredo Bruno, Domenica de Mora, Maritza Olmedo, Michelle P√°ez, Jimmy Garc√©s, Marcela Mejia, Fernando Llerena, Lizbeth Pati√±o, Ruben Armas |
| hCoV-19/USA/AR-CDC-ASC210473352/2021 | EPI_ISL_6529331 | 2021-10-20 | Aegis Sciences Corporation | Centers for Disease Control and Prevention Division of Viral Diseases, Pathogen Discovery | Dakota Howard,Dhwani Batra,Peter Cook,Jason Caravas,Benjamin Rambo-Martin,Scott Sammons,Yvette Unoarumhi,Matthew Schmerer,Kristine Lacek,Tymeckia Kendall,Victoria Caban Figueroa,Shatavia Morrison,Christopher Gulvick,Erisa Sula,Cyndi Clark,Patrick Campbell |
| hCoV-19/Ecuador/NIC-INSPI-15466/2021 | EPI_ISL_6569606 | 2021-11-17 | INSPI-CRN DE INFLUENZA Y OTROS VIRUS RESPIRATORIOS | INSPI-CRN DE INFLUENZA Y OTROS VIRUS RESPIRATORIOS | Alfredo Bruno, Domenica de Mora, Maritza Olmedo, Michelle P√°ez, Jimmy Garc√©s, Maria Angelica Becerra, Marcela Mejia, Carlos Chiluiza, Fernando Llerena, Lizbeth Pati√±o, Ruben Armas |
| hCoV-19/Brazil/SP-IB_139332/2021 | EPI_ISL_6570315 | 2021-10-25 | UBS ESPIRITO SANTO DO TURVO | Instituto Butantan | Dimas Tadeu Covas, Antonio Jorge Martins, Claudia Renata dos Santos Barros, David Schlesinger, Debora Botequio Moretti, Elaine Cristina Marqueze, Elaine Vieira Santos, Evandra Strazza Rodrigues, Heidge Fukumasu, Jayme Augusto de Souza-Neto, Jos√© Salvator |
| hCoV-19/Brazil/AL-IB_138954/2021 | EPI_ISL_6570442 | 2021-09-24 | LACEN ALAGOAS | Instituto Butantan | Dimas Tadeu Covas, Antonio Jorge Martins, Claudia Renata dos Santos Barros, David Schlesinger, Debora Botequio Moretti, Elaine Cristina Marqueze, Elaine Vieira Santos, Evandra Strazza Rodrigues, Heidge Fukumasu, Jayme Augusto de Souza-Neto, Jos√© Salvator |
| hCoV-19/India/MH-RFIP03661/2021 | EPI_ISL_6575350 | 2021-05-03 | Genepath, Pune | RF-IISER Pune | Krishanpal Karmodiya, Aurnab Ghose, Joy Merwin Monteiro, Shatakshi Ranade, Nikhil Phadke |
| hCoV-19/India/MH-RFIP03802/2021 | EPI_ISL_6575486 | 2021-06-01 | BJMC,Pune | RF-IISER Pune | Krishanpal Karmodiya, Aurnab Ghose, Joy Merwin Monteiro, Suvarna Joshi, Rajesh Karyakarte |
| hCoV-19/SouthAfrica/CERI-KRISP-K027875/2021 | EPI_ISL_6585207 | 2021-11-04 | National Health Laboratory Service, Kwazulu-Natal, South Africa | [illegible] and Innvoation, Stellenbosch University and KRISP, KZN Research Innovation and Sequencing Platform, UKZN. | Nokukhanya Mdlalose, Giandhari J, Pillay S, Naidoo Y, Ramphal U, Ramphal Y, Arisha Maharaj, Moir M, San JE, Tshiabuila D, Tegally H, Wilkinson E, de Oliveira T |
| hCoV-19/USA/CO-CDPHE-2101975390/2021 | EPI_ISL_6585405 | 2021-10-14 | Colorado Department of Public Health and Environment | Colorado Department of Public Health and Environment | Laura Bankers, Molly C. Hetherington-Rauth, Diana Ir, Alexandria Rossheim, Michael A. Martin, Mandy Waters, Shannon R. Matzinger, Sarah Elizabeth Totten, Emily A. Travanty |
| hCoV-19/Australia/NSW-ICPMR-13801/2021 | EPI_ISL_6585934 | 2021-11-10 | Laverty Pathology | Clinical Pathology and Medical Research; Westmead Hospital; University of Sydney | Arnott A., Draper J., Gall M., Martinez E., Rockett R., Sintchenko V., on behalf of ICPMR |

| | | | | | |
|---|---|---|---|---|---|
| hCoV-19/Argentina/INEI110555/2021 | EPI_ISL_5159254 | 2021-09-06 | Servicio Virosis Respiratorias-Departamento Virologv≠a-INEI | Instituto Nacional Enfermedades Infecciosas C.G.Malbran | Baumeister E., Avaro M., Benedetti E., Russo M., Dattero ME, Pontoriero A., Cisterna D., Molina V., Perandones C., Tuduri E., De Belder D., Sanchez Loria J., Haim MS., Lorenzo F., Poklepovich T., Campos J. |
| hCoV-19/Australia/VIC25962/2021 | EPI_ISL_6588575 | 2021-10-18 | Microbiological Diagnostic Unit - Public Health Laboratory (MDU-PHL) | Microbiological Diagnostic Unit - Public Health Laboratory (MDU-PHL) | Seemann, T., Horan, K., Sherry, N.L. |
| hCoV-19/Australia/VIC26607/2021 | EPI_ISL_6588830 | 2021-10-25 | Microbiological Diagnostic Unit - Public Health Laboratory (MDU-PHL) | Microbiological Diagnostic Unit - Public Health Laboratory (MDU-PHL) | Seemann, T., Horan, K., Sherry, N.L. |
| hCoV-19/Australia/VIC26598/2021 | EPI_ISL_6589076 | 2021-10-16 | Microbiological Diagnostic Unit - Public Health Laboratory (MDU-PHL) | Microbiological Diagnostic Unit - Public Health Laboratory (MDU-PHL) | Seemann, T., Horan, K., Sherry, N.L. |
| hCoV-19/Australia/VIC26722/2021 | EPI_ISL_6589154 | 2021-11-04 | Microbiological Diagnostic Unit - Public Health Laboratory (MDU-PHL) | Microbiological Diagnostic Unit - Public Health Laboratory (MDU-PHL) | Seemann, T., Horan, K., Sherry, N.L. |
| hCoV-19/Australia/VIC21753/2021 | EPI_ISL_6589952 | 2021-08-26 | Microbiological Diagnostic Unit - Public Health Laboratory (MDU-PHL) | Microbiological Diagnostic Unit - Public Health Laboratory (MDU-PHL) | Seemann, T., Horan, K., Sherry, N.L. |
| hCoV-19/HongKong/VM21044713-1/2021 | EPI_ISL_6590782 | 2021-11-13 | Home Quarantine Taskforce | Hong Kong Department of Health | Alan K.L. Tsang, Peter C.W. Yip, Ken H.L. Ng, Edman T.K. Lam, Rickjason C.W. Chan |
| hCoV-19/USA/CO-CDPHE-2102025724/2021 | EPI_ISL_6599707 | 2021-10-14 | Colorado Department of Public Health and Environment | Colorado Department of Public Health and Environment | Laura Bankers, Molly C. Hetherington-Rauth, Diana Ir, Alexandria Rossheim, Michael Martin, Mandy Waters, Shannon R. Matzinger, Sarah Elizabeth Totten, Emily A. Travanty |
| hCoV-19/USA/CO-CDPHE-2006061187/2020 | EPI_ISL_6602338 | 2020-06-05 | Colorado Department of Public Health and Environment | Colorado Department of Public Health and Environment | Laura Bankers, Molly C. Hetherington-Rauth, Diana Ir, Alexandria Rossheim, Michael Martin, Mandy Waters, Shannon R. Matzinger, Sarah Elizabeth Totten, Emily A. Travanty |
| hCoV-19/SaudiArabia/KFSHRC_023A/2021 | EPI_ISL_6609689 | 2021-05-25 | Molecular Virology section, department of infection & immunity, KFSHRC | Molecular Virology section, department of infection & immunity, KFSHRC | Fatimah S. Alhamlan, Ahmed A. Al-Qahtani, Maysoon S. Mutabagani, Sahar I. Althawadi, Reem S. Almaghrabi Madain S. Alsanea, Basma M. Alahideb, Feda A. Alsuwairi, Maha Al-Abdulkarim, Mohamed S. Asiri, Sara A. Altamimi |
| hCoV-19/Denmark/DCGC-221110/2021 | EPI_ISL_6611967 | 2021-11-10 | Department of Bacteria, Parasites and Fungi, Statens Serum Institut, Copenhagen, Denmark | Statens Serum Institut Bioinformatics and Microbial Genomics | Danish Covid-19 Genome Consortium |
| hCoV-19/Scotland/QEUH-2AB7A37/2021 | EPI_ISL_6616893 | 2021-11-16 | Lighthouse Lab in Glasgow | Wellcome Sanger Institute for the COVID-19 Genomics UK (COG-UK) Consortium | Harper VanSteenhouse, Yumi Kasai, David Gray, Carol Clugston, Anna Dominiczak and Alex Alderton, Roberto Amato, Jeffrey Barrett, Sonia Goncalves, Ewan Harrison, David K. Jackson, Ian Johnston, Dominic Kwiatkowski, Cordelia Langford, John Sillitoe on behal |
| hCoV-19/Botswana/R34B19_BHP_AAC5803/2021 | EPI_ISL_5502135 | 2021-09-26 | Botswana Harvard HIV Reference Laboratory | Botswana Harvard HIV Reference Laboratory | Sikhulile Moyo, Wonderful T. Choga, Dorcas Maruapula, Keoratile Ntshambiwa, Sefetogi Ramaologa, Thongbotho Mphoyakgosi, Boitumelo J. L Zuze, Botshelo Radibe, Legodile Kooepile, Ontlametse T. Bareng, Letsibogo Gaoraelwe, Thela Tefelo, Pamela Smith-Lawrenc |

| | | | | | |
|---|---|---|---|---|---|
| hCoV-19/Botswana/R35B45_BHP_AAC12204/2021 | EPI_ISL_5502523 | 2021-04-24 | Nyanyangbwe HIV Reference Laboratory | Botswana Harvard HIV Reference Laboratory | Sikhulile Moyo, Wonderful T. Choga, Dorcas Maruapula, Keoratile Ntshambiwa, Sefetogi Ramaologa, Thongbotho Mphoyakgosi, Boitumelo J. L Zuze, Botshelo Radibe, Legodile Kooepile, Ontlametse T. Bareng, Letsibogo Gaoraelwe, Thela Tefelo, Pamela Smith-Lawrenc |
| hCoV-19/Peru/PIU-INS-7272/2021 | EPI_ISL_5505554 | 2021-09-14 | Laboratorio de Referencia Nacional de Virus Respiratorios. Centro Nacional de Salud Publica. Instituto Nacional de Salud Peru. | Laboratorio de Referencia Nacional de Virus Respiratorios. Centro Nacional de Salud Publica. Instituto Nacional de Salud Peru. | Carlos Padilla Rojas, Veronica Hurtado Vela, Orson Mestanza Millones, Iris Silva Molina, Luis Barcena Flores, Alicia Nu√±ez Llanos, Steve Acedo Lazo, Wendy Lizarraga Olivares, Victor Jimenez Vasquez, Sara Gordillo Vilchez, Nieves Sevilla Casta√±eda, Princ |
| hCoV-19/Philippines/PH-PGC-60395/2021 | EPI_ISL_5542843 | 2021-07-14 | Philippine General Hospital (PGH) | Philippine Genome Center | Cynthia P. Saloma,Jan Michael C. Yap,Benedict A. Maralit,Marc Edsel C. Ayes,Eva Maria Cutiongco-de la Paz,Francis A. Tablizo,Carlo M. Lapid,El King D. Morado,Joshua Gregor A. Dizon,Kenneth M. Kim,Maria Sofia L. Yangzon,Elcid Aaron R. Pangilinan,Renato Jac |
| hCoV-19/Israel/CVL-21657/2021 | EPI_ISL_5508269 | 2021-10-08 | Israel Central Virology laboratory | Israel Central Virology laboratory | Neta Zuckerman, Efrat Dahan Bucris, Michal Mandelboim, Miranda Geva, Omer Asraf, Efrat Glick-Saar, Gideon Rechavi, Oran Erster, Danit Sofer, Ella Mendelson, Orna Mor |
| hCoV-19/Israel/CVL-20823/2021 | EPI_ISL_5508496 | 2021-09-17 | Israel Central Virology laboratory | Israel Central Virology laboratory | Neta Zuckerman, Efrat Dahan Bucris, Michal Mandelboim, Miranda Geva, Omer Asraf, Efrat Glick-Saar, Gideon Rechavi, Oran Erster, Danit Sofer, Ella Mendelson, Orna Mor |
| hCoV-19/Israel/CVL-22319/2021 | EPI_ISL_5508827 | 2021-01-03 | Israel Central Virology laboratory | Israel Central Virology laboratory | Neta Zuckerman, Efrat Dahan Bucris, Michal Mandelboim, Miranda Geva, Omer Asraf, Efrat Glick-Saar, Gideon Rechavi, Oran Erster, Danit Sofer, Ella Mendelson, Orna Mor |
| hCoV-19/Israel/CVL-21501/2021 | EPI_ISL_5509300 | 2021-10-02 | Israel Central Virology laboratory | Israel Central Virology laboratory | Neta Zuckerman, Efrat Dahan Bucris, Michal Mandelboim, Miranda Geva, Omer Asraf, Efrat Glick-Saar, Gideon Rechavi, Oran Erster, Danit Sofer, Ella Mendelson, Orna Mor |
| hCoV-19/Aruba/AW-RIVM-65006/2021 | EPI_ISL_5429781 | 2021-04-10 | Dutch COVID-19 response team | National Institute for Public Health and the Environment (RIVM) | Adam Meijer, Harry Vennema, Dirk Eggink, Jeroen Cremer, Sharon van den Brink, Bas van der Veer, AnneMarie van den Brandt, Lisa Wijsman, Kim Freriks, Ryanne Jaarsma, Eun√≠ce Then, Lynn Aarts, Sanne Bos, Stijn van Rossum, Florian Zwagemaker, Dennis Schmitz, |
| hCoV-19/Aruba/AW-RIVM-65015/2021 | EPI_ISL_5429782 | 2021-04-10 | Dutch COVID-19 response team | National Institute for Public Health and the Environment (RIVM) | Adam Meijer, Harry Vennema, Dirk Eggink, Jeroen Cremer, Sharon van den Brink, Bas van der Veer, AnneMarie van den Brandt, Lisa Wijsman, Kim Freriks, Ryanne Jaarsma, Eun√≠ce Then, Lynn Aarts, Sanne Bos, Stijn van Rossum, Florian Zwagemaker, Dennis Schmitz, |
| hCoV-19/Aruba/AW-RIVM-65026/2021 | EPI_ISL_5429785 | 2021-04-10 | Dutch COVID-19 response team | National Institute for Public Health and the Environment (RIVM) | Adam Meijer, Harry Vennema, Dirk Eggink, Jeroen Cremer, Sharon van den Brink, Bas van der Veer, AnneMarie van den Brandt, Lisa Wijsman, Kim Freriks, Ryanne Jaarsma, Eun√≠ce Then, Lynn Aarts, Sanne Bos, Stijn van Rossum, Florian Zwagemaker, Dennis Schmitz, |

| | | | | | |
|---|---|---|---|---|---|
| hCoV-19/Israel/CVL-21429/2021 | EPI_ISL_5509881 | 2021-09-29 | Israel Central Virology laboratory | Israel Central Virology laboratory | Neta Zuckerman, Efrat Dahan Bucris, Michal Mandelboim, Miranda Geva, Omer Asraf, Efrat Glick-Saar, Gideon Rechavi, Oran Erster, Danit Sofer, Ella Mendelson, Orna Mor |
| hCoV-19/Israel/CVL-22344/2021 | EPI_ISL_5509966 | 2021-10-17 | Israel Central Virology laboratory | Israel Central Virology laboratory | Neta Zuckerman, Efrat Dahan Bucris, Michal Mandelboim, Miranda Geva, Omer Asraf, Efrat Glick-Saar, Gideon Rechavi, Oran Erster, Danit Sofer, Ella Mendelson, Orna Mor |
| hCoV-19/SouthAfrica/NICD-N13237/2021 | EPI_ISL_5510299 | 2021-07-20 | POLOKWANE | Diseases of the National Health Laboratory Service | Amoako DG, Everatt J, Scheepers C, Mohale T, Ntuli N, Mahlangu B, Mnguni A, Ismail A, Bhiman JN |
| hCoV-19/Aruba/AW-RIVM-61882/2021 | EPI_ISL_4992313 | 2021-09-15 | Dutch COVID-19 response team | National Institute for Public Health and the Environment (RIVM) | Adam Meijer, Harry Vennema, Dirk Eggink, Jeroen Cremer, Sharon van den Brink, Bas van der Veer, AnneMarie van den Brandt, Lisa Wijsman, Kim Freriks, Ryanne Jaarsma, Eun√≠ce Then, Lynn Aarts, Sanne Bos, Stijn van Rossum, Florian Zwagemaker, Dennis Schmitz, |
| hCoV-19/Israel/CVL-16005/2021 | EPI_ISL_5513073 | 2021-07-25 | Israel Central Virology laboratory | Israel Central Virology laboratory | Neta Zuckerman, Efrat Dahan Bucris, Michal Mandelboim, Miranda Geva, Omer Asraf, Efrat Glick-Saar, Gideon Rechavi, Oran Erster, Danit Sofer, Ella Mendelson, Orna Mor |
| hCoV-19/CzechRepublic/CSQ2808/2021 | EPI_ISL_6640384 | 2021-11-11 | Nemocnice Hodonin | University Hospital Brno, CMBG | Bezdicek Matej, Svaton Jan, Kristyna Dufkova, Lengerova Martina, Dolejska Monika |
| hCoV-19/Israel/ICH-760595093/2021 | EPI_ISL_6640467 | 2021-10-05 | Clinical Microbiology Laboratory, Tel Aviv Sourasky Medical Center | Clinical Microbiology Laboratory, Tel Aviv Sourasky Medical Center | Katya Levytskyi, Ora Halutz, Alon Ziv, Lior Handler, Amos Adler |
| hCoV-19/Israel/ICH-760595096/2021 | EPI_ISL_6640532 | 2021-10-07 | Clinical Microbiology Laboratory, Tel Aviv Sourasky Medical Center | Clinical Microbiology Laboratory, Tel Aviv Sourasky Medical Center | Katya Levytskyi, Ora Halutz, Alon Ziv, Lior Handler, Amos Adler |
| hCoV-19/Botswana/R41B2_BHP_000822289/2021 | EPI_ISL_6640823 | 2021-11-03 | National Health Laboratory | Botswana Harvard HIV Reference Laboratory | Sikhulile Moyo, Wonderful T. Choga, Dorcas Maruapula, Keoratile Ntshambiwa, Sefetogi Ramaologa, Thongbotho Mphoyakgosi, Boitumelo Zuze, Botshelo Radibe, Legodile Kooepile, Ontlametse T. Bareng, Pamela Smith-Lawrence, Kgomotso Moruisi, Roger Shapiro, Shahi |
| hCoV-19/Botswana/R41B19_BHP_000822902/2021 | EPI_ISL_6640840 | 2021-11-04 | National Health Laboratory | Botswana Harvard HIV Reference Laboratory | Sikhulile Moyo, Wonderful T. Choga, Dorcas Maruapula, Keoratile Ntshambiwa, Sefetogi Ramaologa, Thongbotho Mphoyakgosi, Boitumelo Zuze, Botshelo Radibe, Legodile Kooepile, Ontlametse T. Bareng, Pamela Smith-Lawrence, Kgomotso Moruisi, Roger Shapiro, Shahi |
| hCoV-19/Botswana/R41B87_BHP_AAC21748/2021 | EPI_ISL_6640873 | 2021-11-10 | National Health Laboratory | Botswana Harvard HIV Reference Laboratory | Sikhulile Moyo, Wonderful T. Choga, Dorcas Maruapula, Keoratile Ntshambiwa, Sefetogi Ramaologa, Thongbotho Mphoyakgosi, Boitumelo Zuze, Botshelo Radibe, Legodile Kooepile, Ontlametse T. Bareng, Pamela Smith-Lawrence, Kgomotso Moruisi, Roger Shapiro, Shahi |
| hCoV-19/Botswana/R40B62_BHP_2121020383/2021 | EPI_ISL_6640876 | 2021-11-11 | Botswana Harvard HIV Reference Laboratory | Botswana Harvard HIV Reference Laboratory | Sikhulile Moyo, Wonderful T. Choga, Dorcas Maruapula, Keoratile Ntshambiwa, Sefetogi Ramaologa, Thongbotho Mphoyakgosi, Boitumelo Zuze, Botshelo Radibe, Legodile Kooepile, Ontlametse T. Bareng, Pamela Smith-Lawrence, Kgomotso Moruisi, Roger Shapiro, Shahi |

| | | | | | |
|---|---|---|---|---|---|
| hCoV-19/USA/CA-CDC-ASC210094588/2021 | EPI_ISL_5514961 | 2021-09-23 | Aegis Sciences Corporation | Centers for Disease Control and Prevention Division of Viral Diseases, Pathogen Discovery | Dakota Howard,Dhwani Batra,Peter Cook,Jason Caravas,Benjamin Rambo-Martin,Scott Sammons,Yvette Unoarumhi,Matthew Schmerer,Kristine Lacek,Tymeckia Kendall,Victoria Caban Figueroa,Shatavia Morrison,Christopher Gulvick,Erisa Sula,Cyndi Clark,Patrick Campbell |
| hCoV-19/India/TG-CDFD-ncov-119461/2021 | EPI_ISL_6642632 | 2021-10-21 | Institute of Preventive Medicine | Centre for DNA Fingerprinting and Diagnostics | Murali Bashyam, Asmita Gupta, Arunkumar Karunanidhi, Vinay Donipadi, Nagamani Kammili, Divya Vashisht, Ashwin Dalal |
| hCoV-19/USA/OH-CDC-ASC210092279/2021 | EPI_ISL_5515234 | 2021-09-23 | Aegis Sciences Corporation | Centers for Disease Control and Prevention Division of Viral Diseases, Pathogen Discovery | Dakota Howard,Dhwani Batra,Peter Cook,Jason Caravas,Benjamin Rambo-Martin,Scott Sammons,Yvette Unoarumhi,Matthew Schmerer,Kristine Lacek,Tymeckia Kendall,Victoria Caban Figueroa,Shatavia Morrison,Christopher Gulvick,Erisa Sula,Cyndi Clark,Patrick Campbell |
| hCoV-19/USA/HI-H214802/2021 | EPI_ISL_5515861 | 2021-09-21 | State Laboratories Division, Hawaii State Department of Health | State Laboratories Division, Hawaii State Department of Health | Pamela O'Brien, Drew Kuwazaki, Ayana Garnet, Daniel Strange, Razvan Sultana, Edward Desmond |
| hCoV-19/Colombia/SAN-INS-VG-9405/2021 | EPI_ISL_6641972 | 2021-10-20 | IDIME COMUNEROS | Instituto Nacional de Salud-Direcci√≥n de Investigaci√≥n en Salud P√∫blica | Katherine Laiton-Donato, Carlos Franco-Mu√±oz, Diego A. √Ålvarez-D√≠az, H√©ctor Alejandro Ruiz-Moreno, Diego Andr√©s Prada, Jhonnatan Reales-Gonz√°lez, Sheryll Corchuelo, Mar√≠a T. Herrera-Sep√∫lveda, Juli√°n Naizaque, Gerardo Santamar√≠a, Jorge Rivera, P |
| hCoV-19/Croatia/13805/2021 | EPI_ISL_6647189 | 2021-10-27 | Zavod za javno zdravstvo Osjeƒçko-Baranjske ≈æupanije | Hrvatski zavod za javno zdravstvo | Irena Tabain, Ivana Ferenƒçak, Josipa Kuzle, Anita Jurić, Dragan Jurić |
| hCoV-19/USA/MI-CDC-ASC210332851/2021 | EPI_ISL_5517179 | 2021-09-21 | Aegis Sciences Corporation | Centers for Disease Control and Prevention Division of Viral Diseases, Pathogen Discovery | Dakota Howard,Dhwani Batra,Peter Cook,Jason Caravas,Benjamin Rambo-Martin,Scott Sammons,Yvette Unoarumhi,Matthew Schmerer,Kristine Lacek,Tymeckia Kendall,Victoria Caban Figueroa,Shatavia Morrison,Christopher Gulvick,Erisa Sula,Cyndi Clark,Patrick Campbell |
| hCoV-19/Italy/FVG-UD-465763/2021 | EPI_ISL_6648709 | 2021-10-27 | Dipartimento di Medicina di Laboratorio, Azienda sanitaria universitaria Friuli Centrale (ASU FC) | Dipartimento di Medicina di Laboratorio, Azienda sanitaria universitaria Friuli Centrale (ASU FC) | Corrado Pipan, Catia Mio, Chiara Dal Secco, Federica Baldan, Stefania Marzinotto, Francesco Curcio |
| hCoV-19/USA/MD-CDC-ASC210307429/2021 | EPI_ISL_6650006 | 2021-10-04 | Aegis Sciences Corporation | Centers for Disease Control and Prevention Division of Viral Diseases, Pathogen Discovery | Dakota Howard,Dhwani Batra,Peter Cook,Jason Caravas,Benjamin Rambo-Martin,Scott Sammons,Yvette Unoarumhi,Matthew Schmerer,Kristine Lacek,Tymeckia Kendall,Victoria Caban Figueroa,Shatavia Morrison,Christopher Gulvick,Erisa Sula,Cyndi Clark,Patrick Campbell |
| hCoV-19/USA/NY-CDC-FG-141220/2021 | EPI_ISL_5082588 | 2021-09-24 | Fulgent Genetics | Centers for Disease Control and Prevention Division of Viral Diseases, Pathogen Discovery | Dakota Howard,Dhwani Batra,Peter Cook,Jason Caravas,Benjamin Rambo-Martin,Scott Sammons,Yvette Unoarumhi,Matthew Schmerer,Kristine Lacek,Tymeckia Kendall,Victoria Caban Figueroa,Shatavia Morrison,Christopher Gulvick,Erisa Sula,Harry Gao,Mickey Li,John Gao |

| | | | | | |
|---|---|---|---|---|---|
| hCoV-19/England/HSLL-27C69A4/2021 | EPI_ISL_5527308 | 2021-10-19 | Health Services Laboratories | Wellcome Sanger Institute for the COVID-19 Genomics UK (COG-UK) Consortium | Health Services Laboratories and Alex Alderton, Roberto Amato, Jeffrey Barrett, Sonia Goncalves, Ewan Harrison, David K. Jackson, Ian Johnston, Dominic Kwiatkowski, Cordelia Langford, John Sillitoe on behalf of the Wellcome Sanger Institute COVID-19 Surve |
| hCoV-19/Australia/NSW-ICPMR-11170/2021 | EPI_ISL_5535192 | 2021-10-14 | Histopath | Clinical Pathology and Medical Research; Westmead Hospital; University of Sydney | Arnott A., Draper J., Gall M., Martinez E., Rockett R., Sintchenko V., on behalf of ICPMR |
| hCoV-19/Philippines/PH-PGC-63860/2021 | EPI_ISL_5543440 | 2021-07-22 | Research Institute for Tropical Medicine, Inc. (RITM) | Philippine Genome Center | Cynthia P. Saloma,Jan Michael C. Yap,Benedict A. Maralit,Marc Edsel C. Ayes,Eva Maria Cutiongco-de la Paz,Francis A. Tablizo,Carlo M. Lapid,El King D. Morado,Joshua Gregor A. Dizon,Kenneth M. Kim,Maria Sofia L. Yangzon,Elcid Aaron R. Pangilinan,Renato Jac |
| hCoV-19/Philippines/PH-PGC-64585/2021 | EPI_ISL_5544118 | 2021-07-30 | University of the Philippines National Institutes Of Health (UP NIH) | Philippine Genome Center | Cynthia P. Saloma,Jan Michael C. Yap,Benedict A. Maralit,Marc Edsel C. Ayes,Eva Maria Cutiongco-de la Paz,Francis A. Tablizo,Carlo M. Lapid,El King D. Morado,Joshua Gregor A. Dizon,Kenneth M. Kim,Maria Sofia L. Yangzon,Elcid Aaron R. Pangilinan,Renato Jac |
| hCoV-19/Philippines/PH-PGC-65648/2021 | EPI_ISL_5546950 | 2021-07-30 | Jose B. Lingad Memorial General Hospital | Philippine Genome Center | Cynthia P. Saloma,Jan Michael C. Yap,Benedict A. Maralit,Marc Edsel C. Ayes,Eva Maria Cutiongco-de la Paz,Francis A. Tablizo,Carlo M. Lapid,El King D. Morado,Joshua Gregor A. Dizon,Kenneth M. Kim,Maria Sofia L. Yangzon,Elcid Aaron R. Pangilinan,Renato Jac |
| hCoV-19/Philippines/PH-PGC-68997/2021 | EPI_ISL_5547802 | 2021-08-02 | Southern Isabela Medical Center Molecular Diagnostic & Research Laboratory (SIMC - MDRL) | Philippine Genome Center | Cynthia P. Saloma,Jan Michael C. Yap,Benedict A. Maralit,Marc Edsel C. Ayes,Eva Maria Cutiongco-de la Paz,Francis A. Tablizo,Carlo M. Lapid,El King D. Morado,Joshua Gregor A. Dizon,Kenneth M. Kim,Maria Sofia L. Yangzon,Elcid Aaron R. Pangilinan,Renato Jac |
| hCoV-19/Philippines/PH-PGC-65111/2021 | EPI_ISL_5550652 | 2021-08-01 | Qualimed Hospital Sta. Rosa | Philippine Genome Center | Cynthia P. Saloma,Jan Michael C. Yap,Benedict A. Maralit,Marc Edsel C. Ayes,Eva Maria Cutiongco-de la Paz,Francis A. Tablizo,Carlo M. Lapid,El King D. Morado,Joshua Gregor A. Dizon,Kenneth M. Kim,Maria Sofia L. Yangzon,Elcid Aaron R. Pangilinan,Renato Jac |
| hCoV-19/Philippines/PH-PGC-54656/2021 | EPI_ISL_5562356 | 2021-06-07 | Baguio General Hospital Medical Center (BGHMC) | Philippine Genome Center | Cynthia P. Saloma,Jan Michael C. Yap,Benedict A. Maralit,Marc Edsel C. Ayes,Eva Maria Cutiongco-de la Paz,Francis A. Tablizo,Carlo M. Lapid,El King D. Morado,Joshua Gregor A. Dizon,Maria Sofia L. Yangzon,Elcid Aaron R. Pangilinan,Renato Jacinto Q. Mantari |
| hCoV-19/Philippines/PH-PGC-78473/2021 | EPI_ISL_5565275 | 2021-08-23 | Philippine Red Cross - Port Area | Philippine Genome Center | Cynthia P. Saloma,Jan Michael C. Yap,Benedict A. Maralit,Marc Edsel C. Ayes,Eva Maria Cutiongco-de la Paz,Francis A. Tablizo,Carlo M. Lapid,El King D. Morado,Joshua Gregor A. Dizon,Maria Sofia L. Yangzon,Elcid Aaron R. Pangilinan,Renato Jacinto Q. Mantari |

| | | | | | |
|---|---|---|---|---|---|
| hCoV-19/USA/CA-CDC-FG-150468/2021 | EPI_ISL_5581734 | 2021-10-14 | Fulgent Genetics | Centers for Disease Control and Prevention Division of Viral Diseases, Pathogen Discovery | Dakota Howard,Dhwani Batra,Peter Cook,Jason Caravas,Benjamin Rambo-Martin,Scott Sammons,Yvette Unoarumhi,Matthew Schmerer,Kristine Lacek,Tymeckia Kendall,Victoria Caban Figueroa,Shatavia Morrison,Christopher Gulvick,Erisa Sula,Harry Gao,Mickey Li,John Gao |
| hCoV-19/Australia/NSW-RPAH-1903/2021 | EPI_ISL_5587718 | 2021-10-24 | New South Wales Health Pathology Royal Prince Alfred Hospital | Microbiology RPAH | Foster, C.; Au, J.; Ruiz Silva, M.; Deveson, I.; Bull, R.; Van Hal, S.; Rawlinson, W. |
| hCoV-19/France/NOR-HMN-21082100510/2021 | EPI_ISL_5594585 | 2021-07-07 | CHU Rouen Normandie | Mondor University Hospital, Assistance Publique H√¥pitaux de Paris, Universit√© Paris-Est Cr√©teil, INSERM U955 | Christophe Rodriguez, Slim Fourati, Vanessa Demontant, Guillaume Gricourt, Melissa N'Debi, Alexandre Soulier, Elisabeth Trawinski, Jean-Michel Pawlotsky |
| hCoV-19/Cameroon/CERI-NPHL-K026064/2021 | EPI_ISL_5596873 | 2021-09-23 | National Public Health Laboratory, Ministry of Public Health of Cameroon | CERI, Centre for Epidemic Response and Innvoation, Stellenbosch University and KRISP, KZN Research Innovation and Sequencing Platform, UKZN. | Dr Essomba Rene Ghislain, Pr Okomo Assoumou Marie Claire, Dr Alle Baba Dieng, Ibrahima Halilou and Dr Etoundi Mballa Alain, Giandhari Jennifer, Pillay Sureshnee, Yajna Ramphal, Naidoo Yeshnee, Tshiabuila Derek,Tegally Houriiyah, San James, Wilkinson Eduan |
| hCoV-19/USA/MD-IGS-52125103849A/2021 | EPI_ISL_5597093 | 2021-09-08 | Maryland Genomics, Institute for Genome Sciences, University of Maryland School of Medicine | Maryland Genomics, Institute for Genome Sciences, University of Maryland School of Medicine | Tallon, Luke J; Sadzewicz, Lisa D; Humphrys, Mike; Ott, Sandra; Roussey, Holly; Santana-Cruz, Ivette; Vavikolanu, Kranthi; Lim, Jonathan; Hazen, Tracy; Fraser, Claire M; Ravel, Jacques |
| hCoV-19/Belgium/rega-14614/2021 | EPI_ISL_5600584 | 2021-08-11 | KU Leuven, Rega Institute, Clinical and Epidemiological Virology | KU Leuven, Rega Institute, Clinical and Epidemiological Virology | Tony Wawina-Bokalanga, Bert Vanmechelen, Joan Marti-Carerras, Piet Maes |
| hCoV-19/Israel/CVL-16500/2021 | EPI_ISL_5602675 | 2021-07-28 | Israel Central Virology laboratory | Israel Central Virology laboratory | Neta Zuckerman, Efrat Dahan Bucris, Michal Mandelboim, Miranda Geva, Omer Asraf, Efrat Glick-Saar, Gideon Rechavi, Oran Erster, Danit Sofer, Ella Mendelson, Orna Mor |
| hCoV-19/SouthAfrica/NICD-N17700/2021 | EPI_ISL_5603075 | 2021-08-31 | AMPATH | Diseases of the National Health Laboratory Service | Amoako DG, Everatt J, Scheepers C, Mohale T, Ntuli N, Mahlangu B, Mnguni A, Ismail A, Bhiman JN |
| hCoV-19/USA/IL-S21WGS5409/2021 | EPI_ISL_5603889 | 2021-01-10 | Illinois Department of Public Health - Springfield Lab | Illinois Department of Public Health - Springfield Lab | Bryan Sim, Gordon McCall |
| hCoV-19/France/IDF-IPP29740/2021 | EPI_ISL_5604545 | 2021-10-05 | Labo Analyses Med | National Reference Center for Viruses of Respiratory Infections, Institut Pasteur, Paris | Marion Barbet, Sylvie Behillil, M√©line Bizard, Angela Brisebarre, Camille Capel, Vincent Enouf, Louise Lefran√ßois, Fr√©d√©ric Lemoine, Christophe Malabat, Corinne Maufrais, Slim El Khiari, Julien Fumey, Etienne Simon-Lori√®re, Maud Vanpeene, Sylvie Van |
| hCoV-19/Israel/CVL-17670/2021 | EPI_ISL_5620750 | 2021-08-03 | Israel Central Virology laboratory | Israel Central Virology laboratory | Neta Zuckerman, Efrat Dahan Bucris, Michal Mandelboim, Miranda Geva, Omer Asraf, Efrat Glick-Saar, Gideon Rechavi, Oran Erster, Danit Sofer, Ella Mendelson, Orna Mor |
| hCoV-19/Israel/CVL-17383/2021 | EPI_ISL_5620956 | 2021-08-04 | Israel Central Virology laboratory | Israel Central Virology laboratory | Neta Zuckerman, Efrat Dahan Bucris, Michal Mandelboim, Miranda Geva, Omer Asraf, Efrat Glick-Saar, Gideon Rechavi, Oran Erster, Danit Sofer, Ella Mendelson, Orna Mor |

| | | | | | |
|---|---|---|---|---|---|
| hCoV-19/Israel/CVL-17446/2021 | EPI_ISL_5620967 | 2021-08-05 | Israel Central Virology laboratory | Israel Central Virology laboratory | Neta Zuckerman, Efrat Dahan Bucris, Michal Mandelboim, Miranda Geva, Omer Asraf, Efrat Glick-Saar, Gideon Rechavi, Oran Erster, Danit Sofer, Ella Mendelson, Orna Mor |
| hCoV-19/USA/VA-DCLS-9878/2021 | EPI_ISL_5622227 | 2021-09-04 | Virginia Division of Consolidated Laboratory Services | Virginia Division of Consolidated Laboratory Services | Virginia Division of Consolidated Laboratory Services |
| hCoV-19/USA/AZ-ASU19232/2021 | EPI_ISL_5629697 | 2021-02-01 | Arizona State University | Arizona State University | Ajeet Bains, LaRinda A. Holland, Matthew F. Smith, Regan A. Sullins, Nicholas J. Mellor, Nathaniel Johnson, Joshua LaBaer, Vel Murugan, Efrem S. Lim |
| hCoV-19/Kenya/SS2407/2021 | EPI_ISL_4739395 | 2021-09-05 | Machakos Level 5 Hospital | KEMRI-Wellcome Trust Research Programme,Kilifi | Githinji G.,Ochola I., Agoti C.,Mohamed K.S., de Laurent Z., Mburu M.W., Morobe J., Ndwiga L. Makori T., Ongera E., Lambisia A. Nokes D J., Mutunga D., Kituku A. |
| hCoV-19/Kenya/SS2304/2021 | EPI_ISL_4739397 | 2021-08-31 | Machakos Level 5 Hospital | KEMRI-Wellcome Trust Research Programme,Kilifi | Githinji G.,Ochola I., Agoti C.,Mohamed K.S., de Laurent Z., Mburu M.W., Morobe J., Ndwiga L. Makori T., Ongera E., Lambisia A. Nokes D J., Mutunga D., Kituku A. |
| hCoV-19/Kenya/SS2340/2021 | EPI_ISL_4739399 | 2021-09-02 | Machakos Level 5 Hospital | KEMRI-Wellcome Trust Research Programme,Kilifi | Githinji G.,Ochola I., Agoti C.,Mohamed K.S., de Laurent Z., Mburu M.W., Morobe J., Ndwiga L. Makori T., Ongera E., Lambisia A. Nokes D J., Mutunga D., Kituku A. |
| hCoV-19/Kenya/SS2345/2021 | EPI_ISL_4739410 | 2021-08-31 | Machakos Level 5 Hospital | KEMRI-Wellcome Trust Research Programme,Kilifi | Githinji G.,Ochola I., Agoti C.,Mohamed K.S., de Laurent Z., Mburu M.W., Morobe J., Ndwiga L. Makori T., Ongera E., Lambisia A. Nokes D J., Mutunga D., Kituku A. |
| hCoV-19/Kenya/C108502/2021 | EPI_ISL_4739411 | 2021-09-06 | KEMRI-Wellcome Trust Research Programme,Kilifi | KEMRI-Wellcome Trust Research Programme,Kilifi | Githinji G.,Ochola I., Agoti C.,Mohamed K.S., de Laurent Z., Mburu M.W., Morobe J., Ndwiga L. Makori T., Ongera E., Lambisia A. D.J Nokes |
| hCoV-19/Kenya/SS2397/2021 | EPI_ISL_4739415 | 2021-09-03 | Machakos Level 5 Hospital | KEMRI-Wellcome Trust Research Programme,Kilifi | Githinji G.,Ochola I., Agoti C.,Mohamed K.S., de Laurent Z., Mburu M.W., Morobe J., Ndwiga L. Makori T., Ongera E., Lambisia A. Nokes D J., Mutunga D., Kituku A. |
| hCoV-19/Kenya/SS2487/2021 | EPI_ISL_4739430 | 2021-09-15 | Machakos Level 5 Hospital | KEMRI-Wellcome Trust Research Programme,Kilifi | Githinji G.,Ochola I., Agoti C.,Mohamed K.S., de Laurent Z., Mburu M.W., Morobe J., Ndwiga L. Makori T., Ongera E., Lambisia A. Nokes D J., Mutunga D., Kituku A. |
| hCoV-19/Kenya/SS2348/2021 | EPI_ISL_4739445 | 2021-08-31 | Machakos Level 5 Hospital | KEMRI-Wellcome Trust Research Programme,Kilifi | Githinji G.,Ochola I., Agoti C.,Mohamed K.S., de Laurent Z., Mburu M.W., Morobe J., Ndwiga L. Makori T., Ongera E., Lambisia A. Nokes D J., Mutunga D., Kituku A. |
| hCoV-19/Kenya/SS2391/2021 | EPI_ISL_4739447 | 2021-09-02 | Machakos Level 5 Hospital | KEMRI-Wellcome Trust Research Programme,Kilifi | Githinji G.,Ochola I., Agoti C.,Mohamed K.S., de Laurent Z., Mburu M.W., Morobe J., Ndwiga L. Makori T., Ongera E., Lambisia A. Nokes D J., Mutunga D., Kituku A. |
| hCoV-19/Sudan/N6782/2020 | EPI_ISL_4739584 | 2020-06-11 | National Public Health Laboratory (NPHL) | National Institute for Communicable Diseases of the National Health Laboratory Service | Elham Rizgalla, Elamin Abualas, Muntaser elTayeb Ibrahim, Adba Alfatih AlEmam, Mohamed Hassan Abdoelraheem Amoako DG, Everatt J, Mohale T, Ntuli N, Mahlangu B, Mnguni A, Ismail A, Bhiman JN, Hatem ahmed babeker Tamadur tag elser, Amira suliaman wadi, Yo |

| | | | | | |
|---|---|---|---|---|---|
| hCoV-19/Norway/20508/2021 | EPI_ISL_4739700 | 2021-09-14 | Akershus University Hospital, Department for Microbiology and Infectious Disease Control | Norwegian Institute of Public Health, Department of Virology | Kathrine Stene-Johansen, Kamilla Heddeland Instefjord, Hilde Elshaug, Garcia Llorente Ignacio, Jon Br√•te, Engebretsen Serina Beate,Pedersen Benedikte Nevjen, Line Victoria Moen, Debech Nadia, Atiya R Ali,Marie Paulsen Madsen, Rasmus Riis Kopperud, Hilde |
| hCoV-19/Colombia/DC-UNIANDES-G029239/2021 | EPI_ISL_4740156 | 2021-08-09 | Gencore - Universidad de los Andes | Gencore - Universidad de los Andes | Luisa Sacristan,Felipe B√°ez, David Gonz√°lez, Cristian Barrera, Gabriela Ariza, Marcela Guevara, Silvia Restrepo |
| hCoV-19/Morocco/20356163/2021 | EPI_ISL_4741161 | 2021-08-31 | Medical Virology and BSL3 Laboratory | Centre de S√©quen√ßage G√©nomique | EL Hamouchi Adil, Charoute Hicham , Bouzidi Aymane, Amalou Ghita, Harmak Houda, Anga Latifa, Faouzi Abdellah, Zemmouri Faouzia, El Oualid Abdelmjid, Nadifiyine Saloua, Zouheir Yassine, Omondi Francis Carey, Somda Soro Georgina Charlene, Abbad Anas, Chgour |
| hCoV-19/Philippines/PH-PGC-56733/2021 | EPI_ISL_4741519 | 2021-06-14 | Philippine Red Cross - Zamboanga Molecular Laboratory | Philippine Genome Center | Cynthia P. Saloma,Jan Michael C. Yap,Benedict A. Maralit,Marc Edsel C. Ayes,Eva Maria Cutiongco-de la Paz,Francis A. Tablizo,Carlo M. Lapid,El King D. Morado,Joshua Gregor A. Dizon,Kenneth M. Kim,Maria Sofia L. Yangzon,Elcid Aaron R. Pangilinan,Renato Jac |
| hCoV-19/Philippines/PH-PGC-58628/2021 | EPI_ISL_4742085 | 2021-07-01 | Philippine General Hospital (PGH) | Philippine Genome Center | Cynthia P. Saloma,Jan Michael C. Yap,Benedict A. Maralit,Marc Edsel C. Ayes,Eva Maria Cutiongco-de la Paz,Francis A. Tablizo,Carlo M. Lapid,El King D. Morado,Joshua Gregor A. Dizon,Kenneth M. Kim,Maria Sofia L. Yangzon,Elcid Aaron R. Pangilinan,Renato Jac |
| hCoV-19/Philippines/PH-PGC-56963/2021 | EPI_ISL_4742345 | 2021-06-21 | Philippine General Hospital (PGH) | Philippine Genome Center | Cynthia P. Saloma,Jan Michael C. Yap,Benedict A. Maralit,Marc Edsel C. Ayes,Eva Maria Cutiongco-de la Paz,Francis A. Tablizo,Carlo M. Lapid,El King D. Morado,Joshua Gregor A. Dizon,Kenneth M. Kim,Maria Sofia L. Yangzon,Elcid Aaron R. Pangilinan,Renato Jac |
| hCoV-19/India/MH-IISER-Pune-IP00926/2020 | EPI_ISL_4743494 | 2020-12-19 | BJMC,Pune | IISER Pune | Krishanpal Karmodiya, Aurnab Ghose, Joy Merwin Monteiro, Suvarna Joshi, Rajesh Karyakarte |
| hCoV-19/Brazil/SP-IB_131494/2021 | EPI_ISL_4746502 | 2021-09-06 | BIOFAST | Instituto Butantan | Dimas Tadeu Covas, Antonio Jorge Martins, Claudia Renata dos Santos Barros, David Schlesinger, Debora Botequio Moretti, Elaine Cristina Marqueze, Elaine Vieira Santos, Evandra Strazza Rodrigues, Heidge Fukumasu, Jayme Augusto de Souza-Neto, Jos√© Salvator |
| hCoV-19/Brazil/SP-IB_132283/2021 | EPI_ISL_4746712 | 2021-09-08 | CS DE PARANAPUA | Instituto Butantan | Dimas Tadeu Covas, Antonio Jorge Martins, Claudia Renata dos Santos Barros, David Schlesinger, Debora Botequio Moretti, Elaine Cristina Marqueze, Elaine Vieira Santos, Evandra Strazza Rodrigues, Heidge Fukumasu, Jayme Augusto de Souza-Neto, Jos√© Salvator |
| hCoV-19/Cambodia/788713/2021 | EPI_ISL_4747992 | 2021-09-19 | Virology Unit, Institut Pasteur du Cambodge | Virology Unit, Institut Pasteur du Cambodge | Leakhena Pum, Sophoannadedh Rath, Jurre Y Siegers, Cecile Troupin, Ly Sovann, Kraing Sidonn, Yi Sengdoeurn, Chin Savuth, Chau Darapheak, Veasna Duong, Erik A Karlsson |

| | | | | | |
|---|---|---|---|---|---|
| hCoV-19/Egypt/ARMY-EVA-Pharma-Wave4-018/2021 | EPI_ISL_4748277 | 2021-09-04 | Main Chemical Laboratories Egypt Army | Main Chemical Laboratories Egypt Army | Mohamed G Seadawy, Riad Mounir Armanious, Ahmed F Gad, Tarek Refaat Elnagdy,Raafat Zaher, Mostfa Elnakib, Mohamed Abdel-Salam Elgohary, Wael Ali, Ahmed Gad,Manal Hamdy Zahran, Ehab Abdelkader Abuelenein,Ahmed S Elsafty, Khaled Amer |
| hCoV-19/India/KL-GENESCOV06323/2021 | EPI_ISL_4748384 | 2021-08-05 | Kerala State SARS-COV-2 Genome Surveillance Programme (GENESCOV2) | GENESCOV2 at CSIR Institute of Genomics and Integrative Biology | Kerala State SARS-COV-2 Genome Surveillance Programme (GENESCOV2) |
| hCoV-19/India/KL-GENESCOV06531/2021 | EPI_ISL_4748588 | 2021-07-12 | Kerala State SARS-COV-2 Genome Surveillance Programme (GENESCOV2) | GENESCOV2 at CSIR Institute of Genomics and Integrative Biology | Kerala State SARS-COV-2 Genome Surveillance Programme (GENESCOV2) |
| hCoV-19/India/KL-GENESCOV06535/2021 | EPI_ISL_4748592 | 2021-07-22 | Kerala State SARS-COV-2 Genome Surveillance Programme (GENESCOV2) | GENESCOV2 at CSIR Institute of Genomics and Integrative Biology | Kerala State SARS-COV-2 Genome Surveillance Programme (GENESCOV2) |
| hCoV-19/India/KL-GENESCOV06635/2021 | EPI_ISL_4748686 | 2021-07-16 | Kerala State SARS-COV-2 Genome Surveillance Programme (GENESCOV2) | GENESCOV2 at CSIR Institute of Genomics and Integrative Biology | Kerala State SARS-COV-2 Genome Surveillance Programme (GENESCOV2) |
| hCoV-19/Australia/VIC19187/2021 | EPI_ISL_4761468 | 2021-07-30 | Microbiological Diagnostic Unit - Public Health Laboratory (MDU-PHL) | Microbiological Diagnostic Unit - Public Health Laboratory (MDU-PHL) | Seemann, T., Horan, K., Sherry, N.L. |
| hCoV-19/Australia/VIC24311/2021 | EPI_ISL_4762257 | 2021-09-13 | Microbiological Diagnostic Unit - Public Health Laboratory (MDU-PHL) | Microbiological Diagnostic Unit - Public Health Laboratory (MDU-PHL) | Seemann, T., Horan, K., Sherry, N.L. |
| hCoV-19/England/MILK-1FA6EF1/2021 | EPI_ISL_4764355 | 2021-09-24 | Lighthouse Lab in Milton Keynes | Wellcome Sanger Institute for the COVID-19 Genomics UK (COG-UK) Consortium | The Lighthouse Lab in Milton Keynes and Alex Alderton, Roberto Amato, Jeffrey Barrett, Sonia Goncalves, Ewan Harrison, David K. Jackson, Ian Johnston, Dominic Kwiatkowski, Cordelia Langford, John Sillitoe on behalf of the Wellcome Sanger Institute COVID-1 |
| hCoV-19/Rwanda/CV2226/2021 | EPI_ISL_4768056 | 2021-01-27 | Rwanda¬†National Joint Taskforce COVID-62 | Africa Centre for Excellence for Genomics of Infectious Diseases (ACEGID), Redeemer‚Äôs University | Enatha Mukantwari, Olawoye, I.B., Dieudonne Mutangana, Oluniyi, P.E., Umuringa Jeanne d'Arc, Kayode, A.T., Esperance Umumararungu, Uwanibe, J.N., Sindayiheba Reuben, Eromon, P.E., Arlene Uwituze, Oguzie, J.U., Murebwayire Clarisse, Ajogbasile, F.V., Izuwa |
| hCoV-19/Nigeria/CV3091/2021 | EPI_ISL_4768652 | 2021-08-30 | Ekiti State Emergency Operation Centre | Africa Centre for Excellence for Genomics of Infectious Diseases (ACEGID), Redeemer‚Äôs University | Olawoye, I.B., Oluniyi, P.E., Eromon, P.E., Oguzie, J.U., Kayode, A.T., Uwanibe, J.N., Ugwu, C.A., Akano, K.O., Ajogbasile, F.V., Abechi, P.S., Olumade, T.J., Nosamiefan, I., Folarin, O., Happi, C.T. |
| hCoV-19/Australia/VIC20627/2021 | EPI_ISL_4777915 | 2021-08-24 | Microbiological Diagnostic Unit - Public Health Laboratory (MDU-PHL) | Microbiological Diagnostic Unit - Public Health Laboratory (MDU-PHL) | Seemann, T., Horan, K., Sherry, N.L. |
| hCoV-19/Wales/PHWC-PD3PUH/2021 | EPI_ISL_4788845 | 2021-09-13 | Wales Specialist Vir Pathogen Genomics Unit | Public Health Wales Microbiology Cardiff Wales Specialist Virology Centre | Catherine Moore, Johnathan Evans, Laura Gifford, Malorie Perry, Simon Cottrell, Angela Marchbank, Alec Birchley, Alexander Adams, Amy Gaskin, Bree Gatica-Wilcox, Jason Coombes, Joel Southgate, Lauren Gilbert, Lee Graham, Nicole Pacchiarini, Sara Kumziene- |
| hCoV-19/Portugal/PT17380/2021 | EPI_ISL_4797898 | 2021-09-21 | Synlab | Instituto Nacional de Saude (INSA) | Borges et al |
| hCoV-19/Spain/MD-HRYC-1052050/2020 | EPI_ISL_4798741 | 2020-11-18 | Servicio de Microbiologia Hospital Ramon y Cajal | Servicio de Microbiologia Hospital Ramon y Cajal | Ponce M, Galan JC, Martinez L. Abreu M, y Gonzalez-Alba JM |
| hCoV-19/Ireland/RN-NVRL-M37IRL60202/2021 | EPI_ISL_4800586 | 2021-09-08 | National Virus Reference Laboratory | National Virus Reference Laboratory | Zoe Yandle, Charlene Bennett, Gabriel Gonzalez, Michael Carr, Jonathan Dean, Cillian F De Gascun |

| | | | | | |
|---|---|---|---|---|---|
| hCoV-19/Reunion/HCL021169793801/2021 | EPI_ISL_4805554 | 2021-09-14 | CERBAILLIANCE LA REUNION | CNR Virus des Infections Respiratoires - France SUD | Antonin Bal, Gregory Destras, Gwendolyne Burfin, Hadrien Regue, Quentin Semanas, Martine Valette, Bruno Lina, Laurence Josset |
| hCoV-19/France/OCC-HCL021170154901/2021 | EPI_ISL_4805691 | 2021-09-20 | CERBALLIANCE OCCITANIE | CNR Virus des Infections Respiratoires - France SUD | Antonin Bal, Gregory Destras, Gwendolyne Burfin, Hadrien Regue, Quentin Semanas, Martine Valette, Bruno Lina, Laurence Josset |
| hCoV-19/Sweden/01_SE100_21CS510975/2021 | EPI_ISL_4816673 | 2021-09-25 | Karolinska University Hospital Solna | Karolinska University Hospital | Jan Albert ,Tobias Allander ,Annelie Bjerkner ,Sandra Broddesson ,Robert Dyrdak ,Martin Ekman ,Lynda Eneh ,Lina Guerra Blomqvist ,Karolina Ininbergs ,Tanja Normark ,Isak Sylvin ,Zhibing Yun ,Martina Wahlund ,Valtteri Wirta |
| hCoV-19/Fiji/FJ1093/2021 | EPI_ISL_4819760 | 2021-06-30 | Fiji Centre for Communicable Disease Control | Microbiological Diagnostic Unit - Public Health Laboratory (MDU-PHL) | Sahukhan, A., Faktaufon, D., Singh, S., Cabemaiwai, T., Seemann, T., Horan, K., Sherry, N.L. |
| hCoV-19/Fiji/FJ031/2021 | EPI_ISL_4819781 | 2021-04-16 | Fiji Centre for Communicable Disease Control | Microbiological Diagnostic Unit - Public Health Laboratory (MDU-PHL) | Sahukhan, A., Faktaufon, D., Singh, S., Cabemaiwai, T., Seemann, T., Horan, K., Sherry, N.L. |
| hCoV-19/Fiji/FJ077/2021 | EPI_ISL_4819889 | 2021-04-25 | Fiji Centre for Communicable Disease Control | Microbiological Diagnostic Unit - Public Health Laboratory (MDU-PHL) | Sahukhan, A., Faktaufon, D., Singh, S., Cabemaiwai, T., Seemann, T., Horan, K., Sherry, N.L. |
| hCoV-19/Fiji/FJ116/2021 | EPI_ISL_4819919 | 2021-05-08 | Fiji Centre for Communicable Disease Control | Microbiological Diagnostic Unit - Public Health Laboratory (MDU-PHL) | Sahukhan, A., Faktaufon, D., Singh, S., Cabemaiwai, T., Seemann, T., Horan, K., Sherry, N.L. |
| hCoV-19/Colombia/ANT-PRO-LDSP365/2021 | EPI_ISL_4820602 | 2021-06-23 | HPTU | Laboratorio Departamental de Salud Publica de Antioquia | Idabely Betancur Ortiz, Maria Stella L√≥pez, Gloria Isabel Escobar, Juan P. Hernandez-Ortiz, Cristian Arbey Velarde Hoyos |
| hCoV-19/USA/ND-NDDH-4397/2020 | EPI_ISL_4821381 | 2020-09-09 | North Dakota Department of Health, Public Health Laboratory | North Dakota Department of Health, Public Health Laboratory | Lisa Wingerter |
| hCoV-19/USA/MN-CDC-IBX670181700029/2021 | EPI_ISL_4822187 | 2021-09-16 | Infinity Biologix | Centers for Disease Control and Prevention Division of Viral Diseases, Pathogen Discovery | Dakota Howard,Dhwani Batra,Peter Cook,Jason Caravas,Benjamin Rambo-Martin,Scott Sammons,Yvette Unoarumhi,Matthew Schmerer,Kristine Lacek,Tymeckia Kendall,Victoria Caban Figueroa,Shatavia Morrison,Christopher Gulvick,Erisa Sula,Christian Bixby,Yihe Wang,Jo |
| hCoV-19/USA/CA-CDC-FG-136082/2021 | EPI_ISL_4827353 | 2021-09-25 | Fulgent Genetics | Centers for Disease Control and Prevention Division of Viral Diseases, Pathogen Discovery | Dakota Howard,Dhwani Batra,Peter Cook,Jason Caravas,Benjamin Rambo-Martin,Scott Sammons,Yvette Unoarumhi,Matthew Schmerer,Kristine Lacek,Tymeckia Kendall,Victoria Caban Figueroa,Shatavia Morrison,Christopher Gulvick,Erisa Sula,Harry Gao,Mickey Li,John Gao |
| hCoV-19/Canada/BC-BCCDC-190115/2021 | EPI_ISL_4835562 | 2021-09-04 | British Columbia Centre For Disease Control | BCCDC Public Health Laboratory | Prystajecky Natalie, Linda Hoang, Dan Fornika, John Tyson, Shannon Russell, Kim Macdonald, Kimia Kamelian, Ana Pacagnella, Corrinne Ng, Loretta Janz, Robert Azana, Mel Krajden |
| hCoV-19/Curacao/CW-RIVM-60493/2021 | EPI_ISL_4839699 | 2021-09-11 | Dutch COVID-19 response team | National Institute for Public Health and the Environment (RIVM) | Adam Meijer, Harry Vennema, Dirk Eggink, Jeroen Cremer, Sharon van den Brink, Bas van der Veer, AnneMarie van den Brandt, Lisa Wijsman, Kim Freriks, Ryanne Jaarsma, Eunv≠ce Then, Lynn Aarts, Sanne Bos, Stijn van Rossum, Florian Zwagemaker, Dennis Schmitz, |

| | | | | | |
|---|---|---|---|---|---|
| hCoV-19/Malawi/NHRL-COVseq0062/2021 | EPI_ISL_4857952 | 2021-05-31 | ZCH | National HIV Reference Laboratory, Ministry of Health, Public Health Institute of Malawi | Chilima BZ, Mvula BD, Chiwaula MJ, Bighignoli B, Malolo I, Chitenje M, Chabuka LB, Panja L, Mzumara TE, Gwayi S, Wolh S |
| hCoV-19/Malawi/NHRL-COVseq0082/2021 | EPI_ISL_4858078 | 2021-05-31 | KCH | National HIV Reference Laboratory, Ministry of Health, Public Health Institute of Malawi | Chilima BZ, Mvula BD, Chiwaula MJ, Bighignoli B, Malolo I, Chitenje M, Chabuka LB, Panja L, Mzumara TE, Gwayi S, Wolh S |
| hCoV-19/Ethiopia/CERI-KRISP-K024746/2021 | EPI_ISL_4874944 | 2021-05-25 | Addis Ababa University | and Innvoation, Stellenbosch University and KRISP, KZN Research Innovation and Sequencing Platform, UKZN. | Abay Sisay, Abraham Tesfaye, Adey Feleke Desta, Giandhari Jennifer, Pillay Sureshnee, Yajna Ramphal, Naidoo Yeshnee, Tshiabuila Derek,Tegally Houriiyah, San James, Wilkinson Eduan, de Oliveira Tulio |
| hCoV-19/Senegal/SC20-1628/2021 | EPI_ISL_4877364 | 2021-08-09 | LBV Le DANTEC | LBV Le DANTEC | Halimatou Diop Ndiaye, Adjiratou Aissatou BA, Assane Dieng, Khadim GUEYE, Dianke Samat√©, Samba NDOUR, Mbengu√© Fall, Pauline Yacine Sene, Oumy DIOP, Sada Diallo, Anna julienne Selbe NDiaye, Pascaline Manga, Aminata Sileymane Thiam, Serigne Saliou Niane, |
| hCoV-19/Australia/VIC23000/2021 | EPI_ISL_4880964 | 2021-09-06 | Microbiological Diagnostic Unit - Public Health Laboratory (MDU-PHL) | Microbiological Diagnostic Unit - Public Health Laboratory (MDU-PHL) | Seemann, T., Horan, K., Sherry, N.L. |
| hCoV-19/SouthAfrica/NICD-N14980/2021 | EPI_ISL_4891683 | 2021-07-19 | Rob Ferreira Laboratory | Diseases of the National Health Laboratory Service | Amoako DG, Everatt J, Scheepers C, Mohale T, Ntuli N, Mahlangu B, Mnguni A, Ismail A, Bhiman JN |
| hCoV-19/SouthAfrica/NICD-N14997/2021 | EPI_ISL_4891728 | 2021-07-19 | Rob Ferreira Laboratory | Diseases of the National Health Laboratory Service | Amoako DG, Everatt J, Scheepers C, Mohale T, Ntuli N, Mahlangu B, Mnguni A, Ismail A, Bhiman JN |
| hCoV-19/USA/TX-CDC-FG-137348/2021 | EPI_ISL_4901510 | 2021-09-18 | Fulgent Genetics | Centers for Disease Control and Prevention Division of Viral Diseases, Pathogen Discovery | Dakota Howard,Dhwani Batra,Peter Cook,Jason Caravas,Benjamin Rambo-Martin,Scott Sammons,Yvette Unoarumhi,Matthew Schmerer,Kristine Lacek,Tymeckia Kendall,Victoria Caban Figueroa,Shatavia Morrison,Christopher Gulvick,Erisa Sula,Harry Gao,Mickey Li,John Gao |
| hCoV-19/USA/CA-CDC-FG-139087/2021 | EPI_ISL_4903028 | 2021-09-28 | Fulgent Genetics | Centers for Disease Control and Prevention Division of Viral Diseases, Pathogen Discovery | Dakota Howard,Dhwani Batra,Peter Cook,Jason Caravas,Benjamin Rambo-Martin,Scott Sammons,Yvette Unoarumhi,Matthew Schmerer,Kristine Lacek,Tymeckia Kendall,Victoria Caban Figueroa,Shatavia Morrison,Christopher Gulvick,Erisa Sula,Harry Gao,Mickey Li,John Gao |
| hCoV-19/USA/WI-CDC-ASC210129547/2021 | EPI_ISL_4913885 | 2021-09-04 | Aegis Sciences Corporation | Centers for Disease Control and Prevention Division of Viral Diseases, Pathogen Discovery | Dakota Howard,Dhwani Batra,Peter Cook,Jason Caravas,Benjamin Rambo-Martin,Scott Sammons,Yvette Unoarumhi,Matthew Schmerer,Kristine Lacek,Tymeckia Kendall,Victoria Caban Figueroa,Shatavia Morrison,Christopher Gulvick,Erisa Sula,Cyndi Clark,Patrick Campbell |
| hCoV-19/Germany/NW-HHU-11717/2021 | EPI_ISL_4917564 | 2021-09-15 | ZOTZ KLIMAS MVZ D√ºsseldorf-Centrum GbR √úBAG f√ºr Labormedizin, Genetik, Zytologie, Pathologie | Center of Medical Microbiology, Virology, and Hospital Hygiene, University of Duesseldorf | Maximilian Damagnez; Alexander Dilthey; Patrick Finzer; Katrin Hoffmann; Torsten Houwaart; Lisanna H√ºlse; Malte Kohns Vasconcelos; Nadine L√ºbke; Jessica Nicolai; Klaus Pfeffer; Daniel Strelow; J√∂rg Timm; Andreas Walker; Tobias Wienemann; Rainer Zotz |

| | | | | | |
|---|---|---|---|---|---|
| hCoV-19/Ghana/FS-21-2563/2021 | EPI_ISL_4919705 | 2021-09-08 | National Influenza Centre | National Influenza Centre | William K. Ampofo, Michael Marks, Ivy A. Asante, Sharon Hsu, Benjamiin B. Lindsey, Benjamin H. Foulkes, Mildred Adusei-Poku, Linda Boatemaa, Lorreta Kwasah, Joseph Oliver-Commey, Ernest Asiedu, Franklin Asiedu-Bekoe, Gordon Awandare, Joyce Ngoi, Dennis La |
| hCoV-19/USA/DC-CDC-ASC210278012/2021 | EPI_ISL_4920820 | 2021-08-14 | Aegis Sciences Corporation | Centers for Disease Control and Prevention Division of Viral Diseases, Pathogen Discovery | Dakota Howard,Dhwani Batra,Peter Cook,Jason Caravas,Benjamin Rambo-Martin,Scott Sammons,Yvette Unoarumhi,Matthew Schmerer,Kristine Lacek,Tymeckia Kendall,Victoria Caban Figueroa,Shatavia Morrison,Christopher Gulvick,Erisa Sula,Cyndi Clark,Patrick Campbell |
| hCoV-19/Germany/NW-RKI-I-267641/2021 | EPI_ISL_4933823 | 2021-09-01 | MVZ Labor Dr. Quade & Kollegen GmbH | Robert Koch Institute | unknown |
| hCoV-19/Canada/AB-ABPHL-35358/2021 | EPI_ISL_5631937 | 2021-06-07 | Edmonton Provincial Lab | Public Health Agency of Canada (PHAC) National Microbiology Laboratory | Buss, E, Croxen M, Deo A, Dieu P, Gill K, Ferrato C, Khan F, Koleva P, Li V, Lloyd C, Lynch T, Ma R, Murphy S, Pabbaraju K, Shokoples S, Tipples G, Thayer J, Whitehouse M, Wong A, Yu C, Zelyas N |
| hCoV-19/Morocco/FMP-346/2021 | EPI_ISL_4945675 | 2021-07-13 | Institut National d'Hygi√®ne / Consortium National de Veille G√©nomique - Maroc | Laboratoire de Biotechnologie | Mouna Ouadghiri, Tarik Aanniz, Safae El Mazouri, Saa√Ød Amzazi, Lahcen Belyamani and Azeddine Ibrahimi |
| hCoV-19/Colombia/VAC-INS-VG-5651/2021 | EPI_ISL_4946248 | 2021-06-03 | Laboratorio Angel ( Synlab) | Instituto Nacional de Salud-Direcci√≥n de Investigaci√≥n en Salud P√∫blica | Katherine Laiton-Donato, Diego A. √Ålvarez-D√≠az, Carlos Franco-Mu√±oz, Hector Alejandro Ruiz-Moreno, Paola Rojas, Maria T. Herrera-Sep√∫lveda, Diego Andr√©s Prada, Jhonnatan Reales-Gonz√°lez, Sheryll Corchuelo, Julian Naizaque, Jorge Rivera, Gerardo Sant |
| hCoV-19/India/MH-RFIP03512/2021 | EPI_ISL_6575200 | 2021-03-28 | Genepath, Pune | RF-IISER Pune | Krishanpal Karmodiya, Aurnab Ghose, Joy Merwin Monteiro, Shatakshi Ranade, Nikhil Phadke |
| hCoV-19/USA/WV-CDC-ASC210083978/2021 | EPI_ISL_4948690 | 2021-09-12 | Aegis Sciences Corporation | Centers for Disease Control and Prevention Division of Viral Diseases, Pathogen Discovery | Dakota Howard,Dhwani Batra,Peter Cook,Jason Caravas,Benjamin Rambo-Martin,Scott Sammons,Yvette Unoarumhi,Matthew Schmerer,Kristine Lacek,Tymeckia Kendall,Victoria Caban Figueroa,Shatavia Morrison,Christopher Gulvick,Erisa Sula,Cyndi Clark,Patrick Campbell |
| hCoV-19/PapuaNewGuinea/PNG3550/2021 | EPI_ISL_4948813 | 2021-08-30 | Central Public Health Laboratory | Microbiological Diagnostic Unit - Public Health Laboratory (MDU-PHL) | Kumbu, J., Susapu, M., Palou, T., Seemann, T., Horan, K., Sherry, N.L. |
| hCoV-19/PapuaNewGuinea/PNG3751/2021 | EPI_ISL_4948860 | 2021-08-27 | Central Public Health Laboratory | Microbiological Diagnostic Unit - Public Health Laboratory (MDU-PHL) | Kumbu, J., Susapu, M., Palou, T., Seemann, T., Horan, K., Sherry, N.L. |
| hCoV-19/PapuaNewGuinea/PNG2875/2021 | EPI_ISL_4948866 | 2021-07-14 | Central Public Health Laboratory | Microbiological Diagnostic Unit - Public Health Laboratory (MDU-PHL) | Kumbu, J., Susapu, M., Palou, T., Seemann, T., Horan, K., Sherry, N.L. |
| hCoV-19/PapuaNewGuinea/PNG3607/2021 | EPI_ISL_4948880 | 2021-08-18 | Central Public Health Laboratory | Microbiological Diagnostic Unit - Public Health Laboratory (MDU-PHL) | Kumbu, J., Susapu, M., Palou, T., Seemann, T., Horan, K., Sherry, N.L. |

| | | | | | |
|---|---|---|---|---|---|
| hCoV-19/PapuaNewGuinea/PNG2623/2021 | EPI_ISL_4948888 | 2021-04-11 | Central Public Health Laboratory | Microbiological Diagnostic Unit - Public Health Laboratory (MDU-PHL) | Kumbu, J., Susapu, M., Palou, T., Seemann, T., Horan, K., Sherry, N.L. |
| hCoV-19/PapuaNewGuinea/PNG3738/2021 | EPI_ISL_4948892 | 2021-08-26 | Central Public Health Laboratory | Microbiological Diagnostic Unit - Public Health Laboratory (MDU-PHL) | Kumbu, J., Susapu, M., Palou, T., Seemann, T., Horan, K., Sherry, N.L. |
| hCoV-19/Burundi/UG878/2021 | EPI_ISL_4949203 | 2021-07-28 | National Institute of Public Health Burundi, WHO Burundi, ASLM Burundi, MRC/UVRI & LSHTM Uganda Research Unit | MRC/UVRI & LSHTM Uganda Research Unit, National Institute of Public Health Burundi, WHO Burundi, ASLM Burundi | Jerome Nkurunziza, Anatole Nkeshimana, Joseph Nyandwi, Jean Baptiste Niyibigira, Butoyi Pascal, Cassien Nduwimana, Francine Kabatesi, Eric Kezakarayagwa, Xavier Crespin, Alexis Niyomwungere, Kouadio Theodore Yao, Jerome Ndaruhutse, Blessing T. Marondera, |
| hCoV-19/India/GJ-ICMR-NIV-INSACOG-GSEQ-5656/2021 | EPI_ISL_4950302 | 2021-09-10 | ICMR-National Institute of Virology - INSACOG | NIV Influenza | Potdar, Varsha on behalf of National Influenza Centre, NIV, Pune |
| hCoV-19/USA/MT-FYR-312/2021 | EPI_ISL_4950658 | 2021-08-17 | FYR Diagnostics | FYR Diagnostics | Blohm, Tre; Ghai, Vikas |
| hCoV-19/USA/CA-OC-FG-130229/2021 | EPI_ISL_4960043 | 2021-09-14 | Ambry Genetics Corporation | Fulgent Genetics | Harry Gao, Mickey Li, John Gao, Joseph Fierro, Benafsh Sapra, Becky Tsai, Yan Meng, Doreen Ng, James Xie |
| hCoV-19/USA/CT-CDC-LC0003748/2021 | EPI_ISL_4966775 | 2021-01-12 | Laboratory Corporation of America | Centers for Disease Control and Prevention Division of Viral Diseases, Pathogen Discovery | Dakota Howard,Dhwani Batra,Peter Cook,Jason Caravas,Benjamin Rambo-Martin,Scott Sammons,Yvette Unoarumhi,Matthew Schmerer,Kristine Lacek,Tymeckia Kendall,Victoria Caban Figueroa,Shatavia Morrison,Christopher Gulvick,Minoo Agarwal,Eyad Almasri,Debbie Boles |
| hCoV-19/Australia/SA0977/2021 | EPI_ISL_4968885 | 2021-09-25 | SA Pathology | SA Pathology | Caitlin Selway, Lex Leong, Chuan Kok Lim, Mark Turra, Ivan Bastian |
| hCoV-19/USA/TN-CDC-ASC210131038/2021 | EPI_ISL_4981305 | 2021-09-07 | Aegis Sciences Corporation | Centers for Disease Control and Prevention Division of Viral Diseases, Pathogen Discovery | Dakota Howard,Dhwani Batra,Peter Cook,Jason Caravas,Benjamin Rambo-Martin,Scott Sammons,Yvette Unoarumhi,Matthew Schmerer,Kristine Lacek,Tymeckia Kendall,Victoria Caban Figueroa,Shatavia Morrison,Christopher Gulvick,Erisa Sula,Cyndi Clark,Patrick Campbell |
| hCoV-19/Switzerland/BE-IFIK-3159-3043/2021 | EPI_ISL_4982337 | 2021-10-02 | Institute for Infectious Diseases | Institute for Infectious Diseases, University of Bern | Nicole Liechti, Christian Baumann, Miguel A Terrazos Miani, Sonja Gempeler, Lo√Øc Borcard, Stefan Neuenschwander, Cora S√§gesser, Pascal Bittel, Peter Keller, Franziska Suter-Riniker, Stephen L Leib, Alban Ramette |
| hCoV-19/Kuwait/Jaber2120359771/2021 | EPI_ISL_4989885 | 2021-08-14 | Jarallah Germna Specialized Clinic | Virology Lab, Jaber Al Ahmad Hospital | Dr. Zahrah Buhamad, Dr. Ebaa Al-Awadhi, Haroon Masih, Khubaid-ur-Rehman, Estabraq Kathim |
| hCoV-19/Aruba/AW-RIVM-62855/2021 | EPI_ISL_4993188 | 2021-09-25 | Dutch COVID-19 response team | National Institute for Public Health and the Environment (RIVM) | Adam Meijer, Harry Vennema, Dirk Eggink, Jeroen Cremer, Sharon van den Brink, Bas van der Veer, AnneMarie van den Brandt, Lisa Wijsman, Kim Freriks, Ryanne Jaarsma, Eun√≠ce Then, Lynn Aarts, Sanne Bos, Stijn van Rossum, Florian Zwagemaker, Dennis Schmitz, |

| | | | | | |
|---|---|---|---|---|---|
| hCoV-19/Netherlands/NH-RIVM-62168/2021 | EPI_ISL_4994997 | 2021-09-14 | Dutch COVID-19 response team | National Institute for Public Health and the Environment (RIVM) | Adam Meijer, Harry Vennema, Dirk Eggink, Jeroen Cremer, Sharon van den Brink, Bas van der Veer, AnneMarie van den Brandt, Lisa Wijsman, Kim Freriks, Ryanne Jaarsma, Eunv≠ce Then, Lynn Aarts, Sanne Bos, Stijn van Rossum, Florian Zwagemaker, Dennis Schmitz, |
| hCoV-19/USA/MT-CDC-ASC210085527/2021 | EPI_ISL_4997459 | 2021-09-13 | Aegis Sciences Corporation | Centers for Disease Control and Prevention Division of Viral Diseases, Pathogen Discovery | Dakota Howard,Dhwani Batra,Peter Cook,Jason Caravas,Benjamin Rambo-Martin,Scott Sammons,Yvette Unoarumhi,Matthew Schmerer,Kristine Lacek,Tymeckia Kendall,Victoria Caban Figueroa,Shatavia Morrison,Christopher Gulvick,Erisa Sula,Cyndi Clark,Patrick Campbell |
| hCoV-19/SouthKorea/KDCA12945/2021 | EPI_ISL_5007484 | 2021-09-04 | Division of Emerging Infectious Diseases, Bureau of Infectious Diseases Diagnosis Control, Korea Disease Control and Prevention Agency | Diseases, Bureau of Infectious Diseases Diagnosis Control, Korea Disease Control and Prevention Agency | Ae Kyung Park, Il-Hwan Kim, Heui Man Kim, Jeong-Ah Kim, Chae Young Lee, Eun-Jin Kim |
| hCoV-19/Chile/VS-158165/2021 | EPI_ISL_5011933 | 2021-09-28 | Genetica Molecular and Subdepartamento de Virologia ISP Chile | Instituto de Salud Publica de Chile | Karen Orostica, Constanza Campano, Barbara Parra, Loredana Arata, Gisselle Barra, Patricia Bustos, Rodrigo Fasce, Javier Tognarelli, Andres Castillo, Soledad Ulloa, Jorge Fernandez |
| hCoV-19/USA/MD-IGS-52123510280A/2021 | EPI_ISL_5014523 | 2021-08-23 | Maryland Genomics, Institute for Genome Sciences, University of Maryland School of Medicine | Maryland Genomics, Institute for Genome Sciences, University of Maryland School of Medicine | Tallon, Luke J; Sadzewicz, Lisa D; Humphrys, Mike; Ott, Sandra; Roussey, Holly; Santana-Cruz, Ivette; Vavikolanu, Kranthi; Lim, Jonathan; Hazen, Tracy; Fraser, Claire M; Ravel, Jacques |
| hCoV-19/USA/CA-CDPH-3000143814/2021 | EPI_ISL_5016613 | 2021-08-06 | CDPH VBL | California Department of Public Health | Emily Smith on behalf of CDPH-COVIDNet |
| hCoV-19/USA/CA-CDPH-3000145663/2021 | EPI_ISL_5017141 | 2021-08-07 | CDPH VBL | California Department of Public Health | Emily Smith on behalf of CDPH-COVIDNet |
| hCoV-19/USA/CO-CDPHE-2101785697/2021 | EPI_ISL_5018477 | 2021-09-09 | Colorado Department of Public Health and Environment | Colorado Department of Public Health and Environment | Laura Bankers, Molly C. Hetherington-Rauth, Diana Ir, Alexandria Rossheim, Michael Martin, Mandy Waters, Shannon R. Matzinger, Sarah Elizabeth Totten, Emily A. Travanty |
| hCoV-19/Brazil/SP-IB_133816/2021 | EPI_ISL_5020434 | 2021-09-13 | BIOFAST | Instituto Butantan | Dimas Tadeu Covas, Antonio Jorge Martins, Claudia Renata dos Santos Barros, David Schlesinger, Debora Botequio Moretti, Elaine Cristina Marqueze, Elaine Vieira Santos, Evandra Strazza Rodrigues, Heidge Fukumasu, Jayme Augusto de Souza-Neto, Jos√© Salvator |
| hCoV-19/Indonesia/JK-GS-NIHRD-WGS11489/2021 | EPI_ISL_5022833 | 2021-09-09 | Lab Smartco | National Institute of Health Research and Development | Kartika Dewi Puspa, Hana Apsari Pawestri, Subangkit, Arie Ardiansyah Nugraha, Hartanti Dian Ikawati, Krisna Pangesti, Triyani Soekarso, Fajar Nur Sulistiyohadi, Nelly Puspandari, Vivi Setiawaty |
| hCoV-19/Thailand/DMSc-02259/2021 | EPI_ISL_5030346 | 2021-06-18 | Charoenkrung pracharak hospital | Division of Genomic Medicine and Innovation support,Department of Medical Sciences, Ministry of Public Health, Thailand | Surakameth Mahasirimongkol,Nuanjun Wichukchinda,Archawin Rojanawiwat,Pilailuk Akkapaiboon Okada ,Waritta Sawaengdee,Penpitcha Thawong,Pundharika Piboonsiri,Jirapha Pakdee,Natthakul Bunneang |

| | | | | | |
|---|---|---|---|---|---|
| hCoV-19/Gabon/CERMEL-JJ0691/2021 | EPI_ISL_5031219 | 2021-09-30 | Centre de Recherches M√©dicales de Lambar√©n√© | Centre de Recherches M√©dicales de Lambar√©n√© | Moustapha Nzamba Maloum, Rodrigue Bikangui, G√©d√©on Prince Manouana, Samira Zoa Assoumou, Sam O'neilla Oye Bingono, Rom√©o Aim√© Laclong Lontchi, Ayong More, Jean Bernard Lekana-Douki, Steffen Borrmann, Bertrand Lell, Srinivas reddy Pallerla, Jo√´l-Fl |
| hCoV-19/Bahrain/21342639141/2021 | EPI_ISL_5032816 | 2021-09-30 | Communicable Disease Laboratory, Public Health Directorate | Communicable Disease Laboratory, Public Health Directorate | Alwasti,H., AlHujairi,Z., AlAbbas,Z., Marhoon,A., Almoamen,G. |
| hCoV-19/Australia/NSW-SAVID-5606/2021 | EPI_ISL_5033187 | 2021-09-03 | Area of Virology, Serology and Virology Division (SAViD), New South Wales Health Pathology Randwick | Virology, Serology and Virology Division (SAViD), New South Wales Health Pathology Randwick | Foster, C.; Au, J.; Ruiz Silva, M.; Deveson, I.; Bull, R.; Van Hal, S.; Rawlinson, W. |
| hCoV-19/Australia/QLD2408/2021 | EPI_ISL_5033343 | 2021-10-03 | PHV-FSS | PHV-FSS | Chenwei Wang on behalf of Q-PHIRE Genomics |
| hCoV-19/India/WB-INSACOG-1931300466610/2021 | EPI_ISL_6395658 | 2021-09-17 | INSACOG-WB | National Institute of Biomedical Genomics ‚Äì INSACOG | Arindam Maitra, Bhaswati Bandyopadhyay, Nidhan Kumar Biswas, Tamal Ghosh, Sreedhar Chinnaswamy, Ajay Chakraborti, Saumitra Das |
| hCoV-19/England/NORW-306189D/2021 | EPI_ISL_5041098 | 2021-02-09 | Quadram Institute Bioscience | COVID-19 Genomics UK (COG-UK) Consortium | Dave J. Baker, Gemma L. Kay, Alp Aydin, Thanh Le-Viet, Steven Rudder, Ana P. Tedim, Anastasia Kolyva, Maria Diaz, Leonardo de Oliveira Martins, Nabil-Fareed Alikhan, Lizzie Meadows, Rachael Stanley, Rose K Davidson, Ngozi Elumogo, Muhammed Yasir, Sophie J |
| hCoV-19/Brazil/SP-IB_133181/2021 | EPI_ISL_5049673 | 2021-09-17 | UBS DR WALDOMIRO FERREIRA NEVES SCRPARDO | Instituto Butantan | Dimas Tadeu Covas, Antonio Jorge Martins, Claudia Renata dos Santos Barros, David Schlesinger, Debora Botequio Moretti, Elaine Cristina Marqueze, Elaine Vieira Santos, Evandra Strazza Rodrigues, Heidge Fukumasu, Jayme Augusto de Souza-Neto, Jos√© Salvator |
| hCoV-19/Bangladesh/icddrb-GSB15-048/2021 | EPI_ISL_5050334 | 2021-09-13 | Rajshahi Medical College, Rajshahi | Virology Laboratory, International Centre for Diarrhoeal Disease Research, Bangladesh (ICDDR,B) | Mojnu Miah, Md. Mahfuzur Rahman, Sabera Gul Nahar, Manjur Hossain Khan, Senjuti Saha, Hassan Afrad, Mohammed Ziaur Rahman, Mohammad Jubair, Mohammad Enayet Hossain, Razib Mazumder, Mohammad Shahidul Islam, Ahmed Nawsher Alam, ASM Alamgir, Mahbubur Rahman, |
| hCoV-19/Germany/BY-MVP-000007800/2021 | EPI_ISL_5050487 | 2021-09-13 | Max von Pettenkofer Institute, Virology, National Reference Center for Retroviruses, LMU Munich | Laboratory for Functional Genome Analysis; Dept. Genomics; Gene Center of the LMU Munich | Max Muenchhoff; Stefan Krebs; Alexander Graf; Oliver Keppler; Helmut Blum |
| hCoV-19/Germany/BY-MVP-000007775/2021 | EPI_ISL_5050601 | 2021-09-01 | Max von Pettenkofer Institute, Virology, National Reference Center for Retroviruses, LMU Munich | Laboratory for Functional Genome Analysis; Dept. Genomics; Gene Center of the LMU Munich | Max Muenchhoff; Stefan Krebs; Alexander Graf; Oliver Keppler; Helmut Blum |
| hCoV-19/Germany/BY-MVP-000007773/2021 | EPI_ISL_5050623 | 2021-09-01 | Max von Pettenkofer Institute, Virology, National Reference Center for Retroviruses, LMU Munich | Laboratory for Functional Genome Analysis; Dept. Genomics; Gene Center of the LMU Munich | Max Muenchhoff; Stefan Krebs; Alexander Graf; Oliver Keppler; Helmut Blum |
| hCoV-19/Germany/BY-MVP-000007820/2021 | EPI_ISL_5050624 | 2021-09-01 | Max von Pettenkofer Institute, Virology, National Reference Center for Retroviruses, LMU Munich | Laboratory for Functional Genome Analysis; Dept. Genomics; Gene Center of the LMU Munich | Max Muenchhoff; Stefan Krebs; Alexander Graf; Oliver Keppler; Helmut Blum |

| | | | | | |
|---|---|---|---|---|---|
| hCoV-19/Germany/BY-MVP-000007268/2021 | EPI_ISL_5050658 | 2021-08-11 | Max von Pettenkofer Institute, Virology, National Reference Center for Retroviruses, LMU Munich | Laboratory for Functional Genome Analysis; Dept. Genomics; Gene Center of the LMU Munich | Max Muenchhoff; Stefan Krebs; Alexander Graf; Oliver Keppler; Helmut Blum |
| hCoV-19/Germany/BY-MVP-000007954/2021 | EPI_ISL_5050881 | 2021-09-26 | Max von Pettenkofer Institute, Virology, National Reference Center for Retroviruses, LMU Munich | Laboratory for Functional Genome Analysis; Dept. Genomics; Gene Center of the LMU Munich | Max Muenchhoff; Stefan Krebs; Alexander Graf; Oliver Keppler; Helmut Blum |
| hCoV-19/Norway/21106/2021 | EPI_ISL_5051118 | 2021-09-07 | Innlandet Hospital Trust, Division Lillehammer, Department for Medical Microbiology | Norwegian Institute of Public Health, Department of Virology | Kathrine Stene-Johansen, Kamilla Heddeland Instefjord, Hilde Elshaug, Garcia Llorente Ignacio, Jon Br√•te, Engebretsen Serina Beate,Pedersen Benedikte Nevjen, Line Victoria Moen, Debech Nadia, Atiya R Ali,Marie Paulsen Madsen, Rasmus Riis Kopperud, Hilde |
| hCoV-19/Thailand/Chiang_Rai_SEQ17099/2021 | EPI_ISL_5052320 | 2021-09-15 | Regional Medical Sciences Center 1/1 Chiang Rai | National Institute of Health, Department of Medical Sciences, Ministry of Public Health, Thailand | Pilailuk Okada; Siripaporn Phuygun; Nuttida Thongpramul; Sittiporn Parnmen; Ratana Tacharoenmuang; Pakorn Piromtong; Natchaya¬† Khiadsang; Thanutsapa Thanadachakul; Warawan Wongboot; sirikanda wimol; Sunthareeya Waicharoen; Archawin Rojanawiwat |
| hCoV-19/India/TG-CDFD-Xi182/2021 | EPI_ISL_5053264 | 2021-04-17 | Centre for DNA Fingerprinting and Diagnostics | CDFD-INSACOG | Murali Bashyam, Asmita Gupta, Pratyusha Bala, Vinay Donipadi, Divya Vashisht, Ashwin Dalal |
| hCoV-19/Colombia/CUN-INS-VG-7924/2021 | EPI_ISL_5055220 | 2021-08-27 | COLCAN | Instituto Nacional de Salud-Direcci√≥n de Investigaci√≥n en Salud P√∫blica | Katherine Laiton-Donato, Diego A. √Ålvarez-D√≠az, Carlos Franco-Mu√±oz, Hector Alejandro Ruiz-Moreno, Paola Rojas, Maria T. Herrera-Sep√∫lveda, Diego Andr√©s Prada, Jhonnatan Reales-Gonz√°lez, Tatiana Cobos, Beatriz de Arco, Sheryll Corchuelo, Julian Naiz |
| hCoV-19/Colombia/DC-INS-VG-8035/2021 | EPI_ISL_5055314 | 2021-07-16 | LABORATORIO LORENA VEJARANO S.A.S. | Instituto Nacional de Salud-Direcci√≥n de Investigaci√≥n en Salud P√∫blica | Katherine Laiton-Donato, Diego A. √Ålvarez-D√≠az, Carlos Franco-Mu√±oz, Hector Alejandro Ruiz-Moreno, Paola Rojas, Maria T. Herrera-Sep√∫lveda, Diego Andr√©s Prada, Jhonnatan Reales-Gonz√°lez, Tatiana Cobos, Beatriz de Arco, Sheryll Corchuelo, Julian Naiz |
| hCoV-19/Italy/SIC-Palermo-CQRC-2290705-2021/2021 | EPI_ISL_5056499 | 2021-08-21 | Laboratorio CQRC | CQRC_QUALITY CONTROL CHEMICAL BIOLOGICAL RISK_AOOR Villa Sofia Cervello Palermo | Di Gaudio F., Contino F., Brunacci G. |
| hCoV-19/Italy/SIC-Palermo-CQRC-21078073-2021/2021 | EPI_ISL_5056614 | 2021-08-21 | Laboratorio CQRC | CQRC_QUALITY CONTROL CHEMICAL BIOLOGICAL RISK_AOOR Villa Sofia Cervello Palermo | Di Gaudio F., Contino F., Brunacci G. |
| hCoV-19/France/NOR-13435627/2021 | EPI_ISL_5062406 | 2021-08-25 | Centre Hospitalier Universitaire de Rouen Laboratoire de Virologie | Centre Hospitalier Universitaire de Rouen Laboratoire de Virologie | Alice Moisan, Fabienne De Oliveira, Marie Leoz |
| hCoV-19/Austria/CeMM17381/2021 | EPI_ISL_5063341 | 2021-09-24 | Austrian Agency for Health and Food Safety (AGES) | Bergthaler laboratory, CeMM Research Center for Molecular Medicine of the Austrian Academy of Sciences | Lukas Endler, Anna Schedl, Fabian Amman, Petr Triska, Matthew Thornton, Thomas Penz, Benedikt Agerer, Maelle Le Moing, Michael Schuster, Bekir Erguner, Jan Laine, Martin Senekowitsch, Christoph Bock, Andreas Bergthaler |
| hCoV-19/India/AR-CSIR-NEIST-INSACOG-4-CID0064/2021 | EPI_ISL_5064563 | 2021-07-19 | Microbiology Department, BPGHTC | Centre for Infectious Diseases, CSIR-NEIST | Romi Wahengbam, Moirangthem G Singh, Munmi Bora, Pankaj Bharali, Prasenjit Manna, Tridip Phukan, Gayatri Gogoi, Yasmin B Tapaddar, Jatin Kalita, G Narahari Sastry |

| | | | | | |
|---|---|---|---|---|---|
| hCoV-19/India/GJ-INSACOG-GBRC2303/2021 | EPI_ISL_5066147 | 2021-09-14 | Green Cross Pathology & Molecular Laboratory, Ahmedabad | Gujarat Biotechnology Research Centre | Ramesh Pandit,Nitin Savaliya,Priyank Chavda,Sonal Sharma ,Janvi Raval,Apurvasinh Puvar,Zarna Patel,Dinesh Kumar,Arpit Shukla,Bhadreshsinh Gohil,Nimesh Patel,Nitin Shukla,Narendra M Patel,Chaitanya Joshi,Madhvi Joshi |
| hCoV-19/USA/TX-HMH-MCoV-47040/2021 | EPI_ISL_5070989 | 2021-05-29 | Houston Methodist Hospital | Houston Methodist Hospital | Randall J. Olsen, Paul A. Christensen, S. Wesley Long, Sishir Subedi, Robert Olson, Marcus Nguyen, James J. Davis, Matthew Ojeda Saavedra, Prasanti Yerramilli, Layne Pruitt, Kristina Reppond, Madison N. Shyer, Jessica Cambric, Ryan Gadd, Ilya J. Finkels |
| hCoV-19/USA/NV-NSPHL-421127/2021 | EPI_ISL_5071242 | 2021-09-13 | Nevada State Public Health Laboratory | Nevada State Public Health Laboratory | Andrew Gorzalski, Lynette Gumbleton, Mark Pandori |
| hCoV-19/USA/TX-HMH-MCoV-47651/2021 | EPI_ISL_5073196 | 2021-06-18 | Houston Methodist Hospital | Houston Methodist Hospital | Randall J. Olsen, Paul A. Christensen, S. Wesley Long, Sishir Subedi, Robert Olson, Marcus Nguyen, James J. Davis, Matthew Ojeda Saavedra, Prasanti Yerramilli, Layne Pruitt, Kristina Reppond, Madison N. Shyer, Jessica Cambric, Ryan Gadd, Ilya J. Finkels |
| hCoV-19/USA/TX-HMH-MCoV-47668/2021 | EPI_ISL_5073213 | 2021-06-21 | Houston Methodist Hospital | Houston Methodist Hospital | Randall J. Olsen, Paul A. Christensen, S. Wesley Long, Sishir Subedi, Robert Olson, Marcus Nguyen, James J. Davis, Matthew Ojeda Saavedra, Prasanti Yerramilli, Layne Pruitt, Kristina Reppond, Madison N. Shyer, Jessica Cambric, Ryan Gadd, Ilya J. Finkels |
| hCoV-19/USA/TX-HMH-MCoV-42711/2020 | EPI_ISL_5074369 | 2020-07-18 | Houston Methodist Hospital | Houston Methodist Hospital | Randall J. Olsen, Paul A. Christensen, S. Wesley Long, Sishir Subedi, Robert Olson, Marcus Nguyen, James J. Davis, Matthew Ojeda Saavedra, Prasanti Yerramilli, Layne Pruitt, Kristina Reppond, Madison N. Shyer, Jessica Cambric, Ryan Gadd, Ilya J. Finkels |
| hCoV-19/USA/CA-CDC-FG-140745/2021 | EPI_ISL_5083050 | 2021-09-30 | Fulgent Genetics | Centers for Disease Control and Prevention Division of Viral Diseases, Pathogen Discovery | Dakota Howard,Dhwani Batra,Peter Cook,Jason Caravas,Benjamin Rambo-Martin,Scott Sammons,Yvette Unoarumhi,Matthew Schmerer,Kristine Lacek,Tymeckia Kendall,Victoria Caban Figueroa,Shatavia Morrison,Christopher Gulvick,Erisa Sula,Harry Gao,Mickey Li,John Gao |
| hCoV-19/USA/TX-HMH-MCoV-47116/2020 | EPI_ISL_5085821 | 2020-07-31 | Houston Methodist Hospital | Houston Methodist Hospital | Randall J. Olsen, Paul A. Christensen, S. Wesley Long, Sishir Subedi, Robert Olson, Marcus Nguyen, James J. Davis, Matthew Ojeda Saavedra, Prasanti Yerramilli, Layne Pruitt, Kristina Reppond, Madison N. Shyer, Jessica Cambric, Ryan Gadd, Ilya J. Finkels |
| hCoV-19/USA/TX-HMH-MCoV-46074/2020 | EPI_ISL_5086238 | 2020-07-24 | Houston Methodist Hospital | Houston Methodist Hospital | Randall J. Olsen, Paul A. Christensen, S. Wesley Long, Sishir Subedi, Robert Olson, Marcus Nguyen, James J. Davis, Matthew Ojeda Saavedra, Prasanti Yerramilli, Layne Pruitt, Kristina Reppond, Madison N. Shyer, Jessica Cambric, Ryan Gadd, Ilya J. Finkels |

| | | | | | |
|---|---|---|---|---|---|
| hCoV-19/USA/SC-CDC-LC0293229/2021 | EPI_ISL_5091020 | 2021-09-24 | Laboratory Corporation of America | Centers for Disease Control and Prevention Division of Viral Diseases, Pathogen Discovery | Dakota Howard,Dhwani Batra,Peter Cook,Jason Caravas,Benjamin Rambo-Martin,Scott Sammons,Yvette Unoarumhi,Matthew Schmerer,Kristine Lacek,Tymeckia Kendall,Victoria Caban Figueroa,Shatavia Morrison,Christopher Gulvick,Minoo Agarwal,Eyad Almasri,Debbie Boles |
| hCoV-19/USA/CA-CDPH-500015686/2021 | EPI_ISL_5637151 | 2021-09-04 | TPMG Regional Laboratory | California Department of Public Health | Emily Smith on behalf of CDPH-COVIDNet and UCLA Technology Center for Genomics & Bioinformatics |
| hCoV-19/USA/CA-CDPH-7000000765/2021 | EPI_ISL_5637621 | 2021-09-05 | CDPH VBL | California Department of Public Health | Emily Smith on behalf of CDPH-COVIDNet and UCLA Technology Center for Genomics & Bioinformatics |
| hCoV-19/USA/NV-NSPHL-459241/2021 | EPI_ISL_5638547 | 2021-10-19 | Nevada State Public Health Laboratory | Nevada State Public Health Laboratory | Andrew Gorzalski, Mark Pandori |
| hCoV-19/USA/MD-IGS-112125600023A/2021 | EPI_ISL_5640301 | 2021-09-13 | Maryland Genomics, Institute for Genome Sciences, University of Maryland School of Medicine | Maryland Genomics, Institute for Genome Sciences, University of Maryland School of Medicine | Tallon, Luke J; Sadzewicz, Lisa D; Humphrys, Mike; Ott, Sandra; Roussey, Holly; Santana-Cruz, Ivette; Vavikolanu, Kranthi; Lim, Jonathan; Hazen, Tracy; Fraser, Claire M; Ravel, Jacques |
| hCoV-19/Gambia/51372/2021 | EPI_ISL_5641073 | 2021-07-16 | MRCG at LSHTM Genomics lab | MRCG at LSHTM Genomics lab | Abdul Karim sesay, Abdoulie Kanteh, Jarra Manneh, Mariama Kujabi, Bakary Sanyang,Dabiri Damilari |
| hCoV-19/Belgium/Jessa_11-2143-001076LKO/2021 | EPI_ISL_5644663 | 2021-10-25 | LKO | Jessa | Severine Berden et al. on behalf of the Jessa_cmdLab |
| hCoV-19/Brazil/SP-IB_113502/2021 | EPI_ISL_5646292 | 2021-05-30 | UPA MARY DOTA | Instituto Butantan | Dimas Tadeu Covas, Antonio Jorge Martins, Claudia Renata dos Santos Barros, David Schlesinger, Debora Botequio Moretti, Elaine Cristina Marqueze, Elaine Vieira Santos, Evandra Strazza Rodrigues, Heidge Fukumasu, Jayme Augusto de Souza-Neto, Josv© Salvator |
| hCoV-19/Brazil/SP-IB_114212/2021 | EPI_ISL_5646955 | 2021-06-07 | UNIDADE BASICA DE SAUDE JARDIM DO LAGO | Instituto Butantan | Dimas Tadeu Covas, Antonio Jorge Martins, Claudia Renata dos Santos Barros, David Schlesinger, Debora Botequio Moretti, Elaine Cristina Marqueze, Elaine Vieira Santos, Evandra Strazza Rodrigues, Heidge Fukumasu, Jayme Augusto de Souza-Neto, Josv© Salvator |
| hCoV-19/Australia/NT386/2021 | EPI_ISL_5656388 | 2021-09-18 | Royal Darwin Hospital Pathology | Microbiological Diagnostic Unit - Public Health Laboratory (MDU-PHL) | Meumann, E., Baird, R., Seemann T., Horan, K., Sherry, N.L. |
| hCoV-19/Australia/NT130/2021 | EPI_ISL_5656398 | 2021-02-05 | Royal Darwin Hospital Pathology | Microbiological Diagnostic Unit - Public Health Laboratory (MDU-PHL) | Meumann, E., Baird, R., Seemann T., Horan, K., Sherry, N.L. |
| hCoV-19/Australia/NT188/2021 | EPI_ISL_5656412 | 2021-04-20 | Royal Darwin Hospital Pathology | Microbiological Diagnostic Unit - Public Health Laboratory (MDU-PHL) | Meumann, E., Baird, R., Seemann T., Horan, K., Sherry, N.L. |
| hCoV-19/Germany/HH-RKI-I-291544/2021 | EPI_ISL_5665007 | 2021-10-15 | Sonic - Labor Dr. von Froreich GmbH | Robert Koch Institute | unknown |
| hCoV-19/Germany/SL-RKI-I-294162/2021 | EPI_ISL_5673341 | 2021-10-14 | Sonic - Bioscientia - MVZ Labor Saar GmbH | Robert Koch Institute | unknown |
| hCoV-19/Brazil/SP-IB_120724/2021 | EPI_ISL_5679943 | 2021-07-12 | UPA DR FRANCO DA ROCHA | Instituto Butantan | Dimas Tadeu Covas, Antonio Jorge Martins, Claudia Renata dos Santos Barros, David Schlesinger, Debora Botequio Moretti, Elaine Cristina Marqueze, Elaine Vieira Santos, Evandra Strazza Rodrigues, Heidge Fukumasu, Jayme Augusto de Souza-Neto, Josv© Salvator |

| | | | | | |
|---|---|---|---|---|---|
| hCoV-19/Brazil/SP-IB_134862/2021 | EPI_ISL_5688387 | 2021-09-28 | CENTRO DE ATENDIMENTO E ENFRENTAMENTO A COVID 19 | Instituto Butantan | Dimas Tadeu Covas, Antonio Jorge Martins, Claudia Renata dos Santos Barros, David Schlesinger, Debora Botequio Moretti, Elaine Cristina Marqueze, Elaine Vieira Santos, Evandra Strazza Rodrigues, Heidge Fukumasu, Jayme Augusto de Souza-Neto, Jos√© Salvator |
| hCoV-19/Brazil/SP-IB_134584/2021 | EPI_ISL_5689459 | 2021-09-26 | UPA PORTE II 24HS JD CEREJEIRAS ATIBAIA | Instituto Butantan | Dimas Tadeu Covas, Antonio Jorge Martins, Claudia Renata dos Santos Barros, David Schlesinger, Debora Botequio Moretti, Elaine Cristina Marqueze, Elaine Vieira Santos, Evandra Strazza Rodrigues, Heidge Fukumasu, Jayme Augusto de Souza-Neto, Jos√© Salvator |
| hCoV-19/Australia/NSW-RPAH-1735/2021 | EPI_ISL_5689738 | 2021-09-09 | New South Wales Health Pathology Royal Prince Alfred Hospital | Microbiology RPAH | Foster, C.; Au, J.; Ruiz Silva, M.; Deveson, I.; Bull, R.; Van Hal, S.; Rawlinson, W. |
| hCoV-19/USA/MN-CDC-IBX948887288560/2021 | EPI_ISL_5690319 | 2021-10-12 | Infinity Biologix | Centers for Disease Control and Prevention Division of Viral Diseases, Pathogen Discovery | Dakota Howard,Dhwani Batra,Peter Cook,Jason Caravas,Benjamin Rambo-Martin,Scott Sammons,Yvette Unoarumhi,Matthew Schmerer,Kristine Lacek,Tymeckia Kendall,Victoria Caban Figueroa,Shatavia Morrison,Christopher Gulvick,Erisa Sula,Christian Bixby,Yihe Wang,Jo |
| hCoV-19/Australia/NSW-RPAH-1823/2021 | EPI_ISL_5691105 | 2021-09-08 | New South Wales Health Pathology Royal Prince Alfred Hospital | Microbiology RPAH | Foster, C.; Au, J.; Ruiz Silva, M.; Deveson, I.; Bull, R.; Van Hal, S.; Rawlinson, W. |
| hCoV-19/Chile/RM-162835/2021 | EPI_ISL_5694112 | 2021-10-04 | Genetica Molecular and Subdepartamento de Virologia ISP Chile | Instituto de Salud Publica de Chile | Karen Orostica, Constanza Campano, Barbara Parra, Patricia Bustos, Rodrigo Fasce, Andres Castillo, Jorge Fernandez |
| hCoV-19/Chile/VS-161415/2021 | EPI_ISL_5694189 | 2021-09-29 | Genetica Molecular and Subdepartamento de Virologia ISP Chile | Instituto de Salud Publica de Chile | Karen Orostica, Constanza Campano, Barbara Parra, Patricia Bustos, Rodrigo Fasce, Andres Castillo, Jorge Fernandez |
| hCoV-19/USA/TN-SPHL-0610/2021 | EPI_ISL_5703247 | 2021-01-14 | TN DOH Lab Services | TN DOH Lab Services | Karen Beasley-Maynard, Brian Selinsky |
| hCoV-19/USA/TN-SPHL-0979/2021 | EPI_ISL_5703453 | 2021-01-14 | TN DOH Lab Services | TN DOH Lab Services | Karen Beasley-Maynard, Brian Selinsky |
| hCoV-19/USA/CO-CDPHE-2101885524/2021 | EPI_ISL_5712967 | 2021-10-01 | Colorado Department of Public Health and Environment | Colorado Department of Public Health and Environment | Laura Bankers, Molly C. Hetherington-Rauth, Diana Ir, Alexandria Rossheim, Michael A. Martin, Mandy Waters, Shannon R. Matzinger, Sarah Elizabeth Totten, Emily A. Travanty |
| hCoV-19/FrenchGuiana/IPG202101199/2021 | EPI_ISL_6662608 | 2021-09-10 | CNR Institut Pasteur de la Guyane | Institut Pasteur de la Guyane | Anne Lavergne, Dominique Rousset, Antoine Enfissi, Arielle Salmier |
| hCoV-19/Algeria/55577/2021 | EPI_ISL_5052201 | 2021-07-14 | NIC, Viral Respiratory Unit | Virology Departement | Fayez Khardine, Fetouma Doudou, Mohamed Amine Beloufa, Aissam Hachid, Aicha Bensalem, Fawzi Derrar |
| hCoV-19/USA/WA-CDC-UW21111380322/2021 | EPI_ISL_6667365 | 2021-11-13 | UW Virology Lab | UW Virology Lab | Pavitra Roychoudhury, Shah Mohamed Bakhash, Pooneh Hajian, Ricardo Perez, Hong Xie, Tien V. Nguyen, Sean Ellis, Nathan Breit, Robert J. Livingston, Meei-Li Huang, Keith R Jerome, Patrick Mathias, Alexander Greninger |

| | | | | | |
|---|---|---|---|---|---|
| hCoV-19/Botswana/R39B44_BHP_823065/2021 | EPI_ISL_6670105 | 2021-11-02 | National Health Laboratory | Botswana Harvard HIV Reference Laboratory | Sikhulile Moyo, Wonderful T. Choga, Dorcas Maruapula, Keoratile Ntshambiwa, Sefetogi Ramaologa, Thongbotho Mphoyakgosi, Boitumelo Zuze, Botshelo Radibe, Legodile Kooepile, Ontlametse T. Bareng, Pamela Smith-Lawrence, Kgomotso Moruisi, Roger Shapiro, Shahi |
| hCoV-19/Chile/AN-178940/2021 | EPI_ISL_6675423 | 2021-11-05 | Genetica Molecular and Subdepartamento de Virologia ISP Chile | Instituto de Salud Publica de Chile | Karen Orostica, Constanza Campano, Barbara Parra, Patricia Bustos, Rodrigo Fasce, Andres Castillo, Jorge Fernandez |
| hCoV-19/Chile/RM-181298/2021 | EPI_ISL_6675738 | 2021-11-12 | Genetica Molecular and Subdepartamento de Virologia ISP Chile | Instituto de Salud Publica de Chile | Karen Orostica, Constanza Campano, Barbara Parra, Patricia Bustos, Rodrigo Fasce, Andres Castillo, Jorge Fernandez |
| hCoV-19/USA/CO-CDPHE-2102053644/2021 | EPI_ISL_6676102 | 2021-10-23 | Colorado Department of Public Health and Environment | Colorado Department of Public Health and Environment | Laura Bankers, Molly C. Hetherington-Rauth, Diana Ir, Alexandria Rossheim, Michael Martin, Mandy Waters, Arianna Smith, Shannon R. Matzinger, Sarah Elizabeth Totten, Emily A. Travanty |
| hCoV-19/Romania/SV_SJU_17854/2020 | EPI_ISL_6679225 | 2020-08-16 | Molecular Biology Lab, Suceava County Emergency Hospital | Stefan cel Mare University Metagenomics Lab | Lobiuc Andrei, Gheorghita Puscaselu Roxana |
| hCoV-19/Romania/SV_SJU_19042/2020 | EPI_ISL_6679231 | 2020-08-28 | Molecular Biology Lab, Suceava County Emergency Hospital | Stefan cel Mare University Metagenomics Lab | Lobiuc Andrei, Gheorghita Puscaselu Roxana |
| hCoV-19/ElSalvador/02-37/2021 | EPI_ISL_6681865 | 2021-07-23 | Laboratorio de Vigilancia en Salud P√ʃblica el Salvador | Genomics and Proteomics Departament, Gorgas Memorial Institute For Health Studies | Hilda Chinchilla, Mitzi Castro, Sofia Carolina Alvarado, Claudia D√≠az, Sandra Paola Paz, Leyda Abrego, Marlene Castillo, Oris Chavarria, Jessica Gondola, Ambar Moreno, Claudia Gonzalez, Alexander Martinez |
| hCoV-19/India/MH-MCGM-ICMR-INSACOG-WG1224/2021 | EPI_ISL_6684934 | 2021-10-20 | Kasturba Hospital Molecular Lab - INSACOG | Kasturba hospital WGS | Jayanthi Shastri, Vidushi Chitalia |
| hCoV-19/India/MH-MCGM-ICMR-INSACOG-WG1220/2021 | EPI_ISL_6684990 | 2021-10-22 | Kasturba Hospital Molecular Lab - INSACOG | Kasturba hospital WGS | Jayanthi Shastri, Vidushi Chitalia |
| hCoV-19/Canada/ON-SHL-21-02821/2021 | EPI_ISL_6685544 | 2021-07-26 | Shared Hospital Laboratory | Shared Hospital Laboratory | Patryk Aftanas, Finlay Maguire, Kevin Katz, Samira Mubareka, Christie Vermeiren, Robert Kozak |
| hCoV-19/Israel/ICH-760595282/2021 | EPI_ISL_6688421 | 2021-10-22 | Clinical Microbiology Laboratory, Tel Aviv Sourasky Medical Center | Clinical Microbiology Laboratory, Tel Aviv Sourasky Medical Center | Katya Levytskyi, Ora Halutz, Alon Ziv, Lior Handler, Amos Adler |
| hCoV-19/Israel/ICH-760595248/2021 | EPI_ISL_6689081 | 2021-10-21 | Clinical Microbiology Laboratory, Tel Aviv Sourasky Medical Center | Clinical Microbiology Laboratory, Tel Aviv Sourasky Medical Center | Katya Levytskyi, Ora Halutz, Alon Ziv, Lior Handler, Amos Adler |
| hCoV-19/Slovakia/33941/2021 | EPI_ISL_6689924 | 2021-10-25 | Public Health Authority of the Slovak Republic | Public Health Authority of the Slovak Republic | Barbora Kotvasov√°, Lucia ≈†evƒç√≠kov√°, Ter√©zia Vrabƒæeov√°, Anna Giƒçov√°, Elena Tich√°, Miroslav B√∂hmer, Tom√°≈° Szemes, Pavol Mi≈°enko |
| hCoV-19/India/MH-KasturbaMCGM-ICMR-INSACOG-WG136/2021 | EPI_ISL_6689970 | 2021-08-20 | Kasturba Hospital Molecular Lab - INSACOG | Kasturba hospital WGS | Jayanthi Shastri, Vidushi Chitalia |
| hCoV-19/India/MH-KasturbaMCGM-ICMR-INSACOG-WG68/2021 | EPI_ISL_6690074 | 2021-04-20 | Kasturba Hospital Molecular Lab - INSACOG | Kasturba hospital WGS | Jayanthi Shastri, Vidushi Chitalia |

| | | | | | |
|---|---|---|---|---|---|
| hCoV-19/India/MH-KasturbaMCGM-ICMR-INSACOG-WG805/2021 | EPI_ISL_6690997 | 2021-09-20 | Kasturba Hospital Molecular Lab - INSACOG | Kasturba hospital WGS | Jayanthi Shastri, Vidushi Chitalia |
| hCoV-19/Malawi/NHRL-COVseq0129/2021 | EPI_ISL_6692105 | 2021-09-22 | Queen Elizabeth Central Hospital | Ministry of Health, Public Health Institute of Malawi | Chiwaula MJ, Mvula BD, Chitenje M, Dzinza N, Kusakala G, Makaula KP |
| hCoV-19/SouthAfrica/NICD-N19828/2020 | EPI_ISL_6693552 | 2020-06-26 | TSHEPONG LABORATORY | Diseases of the National Health Laboratory Service | Amoako DG, Everatt J, Scheepers C, Mohale T, Ntuli N, Mahlangu B, Mnguni A, Ismail A, Bhiman JN |
| hCoV-19/SouthAfrica/NICD-N19829/2020 | EPI_ISL_6693624 | 2020-06-26 | TSHEPONG LABORATORY | Diseases of the National Health Laboratory Service | Amoako DG, Everatt J, Scheepers C, Mohale T, Ntuli N, Mahlangu B, Mnguni A, Ismail A, Bhiman JN |
| hCoV-19/Djibouti/NAMRU3_E43/2021 | EPI_ISL_6693779 | 2021-09-22 | Naval Medical Research Unit No. 3 | Naval Medical Research Center Biological Defense Research Directorate | Catherine E. Arnold, Logan J. Voegtly, Emily K. Stefanov, Haven L. Miner, Andrea E. Luquette, Robin H. Miller, Lindsay Glang, Francisco Malagon, Regina Z. Cer, Stephen M. Eggan, and Kimberly A. Bishop-Lilly |
| hCoV-19/Israel/ICH-7605955534/2021 | EPI_ISL_6694203 | 2021-11-01 | Clinical Microbiology Laboratory, Tel Aviv Sourasky Medical Center | Clinical Microbiology Laboratory, Tel Aviv Sourasky Medical Center | Katya Levytskyi, Ora Halutz, Alon Ziv, Lior Handler, Amos Adler |
| hCoV-19/Israel/ICH-7605955771/2021 | EPI_ISL_6694317 | 2021-11-02 | Clinical Microbiology Laboratory, Tel Aviv Sourasky Medical Center | Clinical Microbiology Laboratory, Tel Aviv Sourasky Medical Center | Katya Levytskyi, Ora Halutz, Alon Ziv, Lior Handler, Amos Adler |
| hCoV-19/Argentina/INEI111345/2021 | EPI_ISL_6694970 | 2021-09-26 | Servicio Virosis Respiratorias-Departamento Virologv≠a-INEI | Instituto Nacional Enfermedades Infecciosas C.G.Malbran | Baumeister E., Avaro M., Benedetti E., Russo M., Dattero ME, Pontoriero A., Cisterna D., Molina V., Perandones C., Tuduri E., De Belder D., Sanchez Loria J., Haim MS., Lorenzo F., Poklepovich T., Campos J. |
| hCoV-19/Argentina/INEI111365/2021 | EPI_ISL_6694986 | 2021-09-25 | Servicio Virosis Respiratorias-Departamento Virologv≠a-INEI | Instituto Nacional Enfermedades Infecciosas C.G.Malbran | Baumeister E., Avaro M., Benedetti E., Russo M., Dattero ME, Pontoriero A., Cisterna D., Molina V., Perandones C., Tuduri E., De Belder D., Sanchez Loria J., Haim MS., Lorenzo F., Poklepovich T., Campos J. |
| hCoV-19/Argentina/INEI111414/2021 | EPI_ISL_6695017 | 2021-09-23 | Servicio Virosis Respiratorias-Departamento Virologv≠a-INEI | Instituto Nacional Enfermedades Infecciosas C.G.Malbran | Baumeister E., Avaro M., Benedetti E., Russo M., Dattero ME, Pontoriero A., Cisterna D., Molina V., Perandones C., Tuduri E., De Belder D., Sanchez Loria J., Haim MS., Lorenzo F., Poklepovich T., Campos J. |
| hCoV-19/Argentina/INEI111961/2021 | EPI_ISL_6695193 | 2021-10-14 | Servicio Virosis Respiratorias-Departamento Virologv≠a-INEI | Instituto Nacional Enfermedades Infecciosas C.G.Malbran | Baumeister E., Avaro M., Benedetti E., Russo M., Dattero ME, Pontoriero A., Cisterna D., Molina V., Perandones C., Tuduri E., De Belder D., Sanchez Loria J., Haim MS., Lorenzo F., Poklepovich T., Campos J. |
| hCoV-19/India/TG-RFCH00522_CIB0205/2021 | EPI_ISL_6696747 | 2021-09-20 | CSIR-Centre for Cellular and Molecular Biology | CSIR-Centre for Cellular and Molecular Biology | Jandhyala Sai Krishna,Tulasi Nagabandi, Lamuk Zaveri,Malini Nemalikanti,Sreelekshmi MS, Ara Sreenivas,Shreekant Verma, Amareshwar Vodapalli,B Himasri,Valli Nagalakshmi Undamatla, Sumedha Avadhanula,Priya Nurkurthy,Vidhyadhari Methuku, Archana Bharadwaj Si |
| hCoV-19/Australia/WA832/2021 | EPI_ISL_6698576 | 2021-11-19 | PathWest Laboratory Medicine WA | PathWest Laboratory Medicine WA | PathWest Laboratory Medicine WA Microbial Surveillance Unit |
| hCoV-19/HongKong/VM21045145/2021 | EPI_ISL_6698790 | 2021-11-18 | HOME QUARANTINE TASKFORCE | Hong Kong Department of Health | Alan K.L. Tsang, Peter C.W. Yip, Ken H.L. Ng, Edman T.K. Lam, Rickjason C.W. Chan |

| | | | | | |
|---|---|---|---|---|---|
| hCoV-19/Zimbabwe/CERI-KRISP-K027283/2021 | EPI_ISL_6699692 | 2021-10-04 | National Microbiology Reference Laboratory, Ministry of Health, Harare, Zimbabwe | CERI, Centre for Epidemic Response and Innvoation, Stellenbosch University and KRISP, KZN Research Innovation and Sequencing Platform, UKZN. | Air Comodor Dr J. Chimedza, Dr Raiva Simbi, Agnes Juru, Tapfumanei Mashe, Kenneth Maeka, Hlanai Gumbo, Tatenda Takawira, Charles Nyagupe, Giandhari J, Pillay S, Naidoo Y, Ramphal U, Ramphal Y, Arisha Maharaj, Moir M, San JE, Tshiabuila D, Tegally H, Wilki |
| hCoV-19/SouthAfrica/CERI-KRISP-K027291/2021 | EPI_ISL_6699702 | 2021-10-24 | National Health Laboratory Service, Kwazulu-Natal, South Africa | and Innvoation, Stellenbosch University and KRISP, KZN Research Innovation and Sequencing Platform, UKZN. | Nokukhanya Mdlalose, Giandhari J, Pillay S, Naidoo Y, Ramphal U, Ramphal Y, Arisha Maharaj, Moir M, San JE, Tshiabuila D, Tegally H, Wilkinson E, de Oliveira T |
| hCoV-19/SouthAfrica/CERI-KRISP-K032226/2021 | EPI_ISL_6699752 | 2021-11-19 | National Health Laboratory Services, Virology, Charlotte Maxeke Johannesburg hospital, Parktown, Johannesburg, Gauteng | CERI, Centre for Epidemic Response and Innvoation, Stellenbosch University and KRISP, KZN Research Innovation and Sequencing Platform, UKZN. | Kathleen Subramoney, Florette Treurnicht, Avani Bharuthram, Nkhensani Mtileni, NGS-SA (Scheepers C, Hsiao M, von Gottberg A, Bhiman J, Lessells R, Bester P, Engelbrecht S, Preiser W, Mdlalose N, Mlisana K, Goedhals D, Everatt J, Amoaka D, Makatini Z, Hard |
| hCoV-19/SouthAfrica/CERI-KRISP-K032254/2021 | EPI_ISL_6699771 | 2021-11-22 | National Health Laboratory Services, Virology, Charlotte Maxeke Johannesburg hospital, Parktown, Johannesburg, Gauteng | CERI, Centre for Epidemic Response and Innvoation, Stellenbosch University and KRISP, KZN Research Innovation and Sequencing Platform, UKZN. | Kathleen Subramoney, Florette Treurnicht, Avani Bharuthram, Nkhensani Mtileni, NGS-SA (Scheepers C, Hsiao M, von Gottberg A, Bhiman J, Lessells R, Bester P, Engelbrecht S, Preiser W, Mdlalose N, Mlisana K, Goedhals D, Everatt J, Amoaka D, Makatini Z, Hard |
| hCoV-19/France/IDF-IPP29798/2021 | EPI_ISL_5717242 | 2021-10-12 | Hospital | National Reference Center for Viruses of Respiratory Infections, Institut Pasteur, Paris | Marion Barbet, Sylvie Behillil, M√©line Bizard, Angela Brisebarre, Camille Capel, Vincent Enouf, Louise Lefran√ßois, Fr√©d√©ric Lemoine, Christophe Malabat, Corinne Maufrais, Slim El Khiari, Julien Fumey, Etienne Simon-Lori√®re, Maud Vanpeene, Sylvie Van |
| hCoV-19/India/KA-NIMH-SEQ-2877/2021 | EPI_ISL_5726232 | 2021-08-14 | VRDL HASSAN | INSACOG-KA , NIMHANS | Ellango Ramasamy, Sony Sharma, Mahesh Kumar.C.S, Ananthapadmanabha Kotambail, Chetan G K, Gautham Arunachal Udupi, Anita S Desai, V Ravi |
| hCoV-19/India/KA-NIMH-SEQ-2943/2021 | EPI_ISL_5727225 | 2021-09-07 | BENGALURU URBAN | INSACOG-KA , NIMHANS | Ellango Ramasamy, Sony Sharma, Mahesh Kumar.C.S, Ananthapadmanabha Kotambail, Chetan G K, Gautham Arunachal Udupi, Anita S Desai, V Ravi |
| hCoV-19/France/BRE-HMN-21112160793/2021 | EPI_ISL_6701266 | 2021-10-26 | LABORIZON BRETAGNE | Mondor University Hospital, Assistance Publique H√¥pitaux de Paris, Universit√© Paris-Est Cr√©teil, INSERM U955 | Christophe Rodriguez, Slim Fourati, Vanessa Demontant, Guillaume Gricourt, Melissa N'Debi, Alexandre Soulier, Elisabeth Trawinski, Jean-Michel Pawlotsky |
| hCoV-19/Brazil/MG-FIOCRUZ-AR-7/2021 | EPI_ISL_6701366 | 2021-08-23 | Funda√ß√£o Ezequiel Dias | Instituto Ren√© Rachou / Fiocruz Minas | Tha√≠s Silva, Caroline Penido, Fl√°vio Ara√∫jo, Anna Salim, Andr√© Bernardes, Felipe Iani, Rubens do Monte Neto, Gabriel Fernandes, Pedro Alves |
| hCoV-19/Canada/SK-RRPL-151909/2021 | EPI_ISL_5845239 | 2021-07-15 | SK-Roy Romanow Provincial Laboratory | Saskatchewan - Roy Romanow Provincial Laboratory (RRPL) | Ryan McDonald, Keith MacKenzie, Meredith Faires, Kara Loos, Rachel DePaulo, Alanna Senecal, Amanda Lang, Jessica Minion, Roy Romanow Provincial Laboratory - Molecular Diagnostics |
| hCoV-19/Colombia/ANT-CIB-10-24/2021 | EPI_ISL_5737677 | 2021-09-20 | Laboratorio Cl√≠nico BioReferencia | Corporaci√≥n para Investigaciones Biol√≥gicas-CIB | Uriel A. Hurtado Paez, Katterine Molina Hoyos, Jeanneth Mosquera Rendon, Marcela Mercado Reyes |
| hCoV-19/Denmark/DCGC-227452/2021 | EPI_ISL_6702923 | 2021-11-16 | Department of Bacteria, Parasites and Fungi, Statens Serum Institut, Copenhagen, Denmark | Statens Serum Institut Bioinformatics and Microbial Genomics | Danish Covid-19 Genome Consortium |

| | | | | | |
|---|---|---|---|---|---|
| hCoV-19/Senegal/SN-IR2-0040124/2021 | EPI_ISL_5748471 | 2021-10-25 | Iressef Genomics lab | IRESSEF | Souleymane MBOUP, Abdou PADANE , Khadim GUEYE, Diabou Diagne , Samba Ndiour, Papa Alassane DIAW, Birahim Piere NDIAYE, Barada CISSE, Aminata MBOUP, Moustapha MBOW, Ndeye Coumba Toure KANE, Nafisatou LEYE, Gora LO, Ambroise AHOUIDI , Astou Gaye GAYE, Amina |
| hCoV-19/Senegal/SN-IRVAC-139/2021 | EPI_ISL_5748492 | 2021-07-18 | Iressef Genomics lab | IRESSEF | Souleymane MBOUP, Abdou PADANE , Khadim GUEYE, Diabou Diagne , Samba Ndiour, Papa Alassane DIAW, Birahim Piere NDIAYE, Barada CISSE, Aminata MBOUP, Moustapha MBOW, Ndeye Coumba Toure KANE, Nafisatou LEYE, Gora LO, Ambroise AHOUIDI , Astou Gaye GAYE, Amina |
| hCoV-19/Bahrain/21921742973/2021 | EPI_ISL_5632882 | 2021-10-21 | Communicable Disease Laboratory, Public Health Directorate | Communicable Disease Laboratory, Public Health Directorate | Alwasti,H., Altaif,Z., Marhoon,A., AlHujairi,Z., AlTooq,M., Khamees,F., AlAnsari,W., AlAbbas,Z., Almoamen,G. |
| hCoV-19/Bahrain/21342705029/2021 | EPI_ISL_5632863 | 2021-10-23 | Communicable Disease Laboratory, Public Health Directorate | Communicable Disease Laboratory, Public Health Directorate | Alwasti,H., Altaif,Z., Marhoon,A., AlHujairi,Z., AlTooq,M., Khamees,F., AlAnsari,W., AlAbbas,Z., Almoamen,G. |
| hCoV-19/Sweden/8125969991/2021 | EPI_ISL_6705746 | 2021-11-12 | Swedish national genomic surveillance program of SARS-CoV-2 | The Public Health Agency of Sweden | Maximilian Riess, Maria Lind Karlberg, Alma Brolund, Swedish national genomic surveillance program of SARS-CoV-2 |
| hCoV-19/India/MZ-INSACOG-1526100650508/2021 | EPI_ISL_5758390 | 2021-09-20 | INSACOG-Mizoram | National Institute of Biomedical Genomics ‚Äì†INSACOG | Arindam Maitra, Swagnik Roy, Nidhan Kumar Biswas, Gracy Laldinmawii, Sreedhar Chinnaswamy, N Senthil Kumar, Saumitra Das |
| hCoV-19/India/AS-INSACOG-1828300501192/2021 | EPI_ISL_5759387 | 2021-09-01 | INSACOG-Assam | National Institute of Biomedical Genomics ‚Äì†INSACOG | Arindam Maitra, Ajanta Sharma, Nidhan Kumar Biswas, Lahari Saikia, Sreedhar Chinnaswamy,¬†Kailash Chamuah, Saumitra Das |
| hCoV-19/Israel/ICH-760596237/2021 | EPI_ISL_6708464 | 2021-11-12 | Clinical Microbiology Laboratory, Tel Aviv Sourasky Medical Center | Clinical Microbiology Laboratory, Tel Aviv Sourasky Medical Center | Katya Levytskyi, Ora Halutz, Alon Ziv, Lior Handler, Amos Adler |
| hCoV-19/Japan/PG-124074/2021 | EPI_ISL_5767199 | 2021-09-08 | Department of Infectious Diseases, Kobe Institute of Health | Department of Infectious Diseases, Kobe Institute of Health | Ryohei Nomoto, Noriko Nakanishi, Tomotada Iwamoto, Tsuyoshi Sekizuka, Kentaro Itokawa, Rina Tanaka, Masanori Hashino, Makoto Kuroda |
| hCoV-19/USA/CA-CDC-ASC210321021/2021 | EPI_ISL_5774385 | 2021-09-29 | Aegis Sciences Corporation | Centers for Disease Control and Prevention Division of Viral Diseases, Pathogen Discovery | Dakota Howard,Dhwani Batra,Peter Cook,Jason Caravas,Benjamin Rambo-Martin,Scott Sammons,Yvette Unoarumhi,Matthew Schmerer,Kristine Lacek,Tymeckia Kendall,Victoria Caban Figueroa,Shatavia Morrison,Christopher Gulvick,Erisa Sula,Cyndi Clark,Patrick Campbell |
| hCoV-19/USA/CO-CDPHE-2101904979/2021 | EPI_ISL_5776207 | 2021-09-30 | Colorado Department of Public Health and Environment | Colorado Department of Public Health and Environment | Laura Bankers, Molly C. Hetherington-Rauth, Diana Ir, Alexandria Rossheim, Michael A. Martin, Mandy Waters, Shannon R. Matzinger, Sarah Elizabeth Totten, Emily A. Travanty |
| hCoV-19/Canada/AB-ABPHL-35023/2021 | EPI_ISL_5631679 | 2021-09-20 | Edmonton Provincial Lab | Public Health Agency of Canada (PHAC) National Microbiology Laboratory | Buss, E, Croxen M, Deo A, Dieu P, Gill K, Ferrato C, Khan F, Koleva P, Li V, Lloyd C, Lynch T, Ma R, Murphy S, Pabbaraju K, Shokoples S, Tipples G, Thayer J, Whitehouse M, Wong A, Yu C, Zelyas N |

| | | | | | |
|---|---|---|---|---|---|
| hCoV-19/USA/VT-CDCBI-CRSP_RSSKJHG2OL6K3SHR/2021 | EPI_ISL_5777068 | 2021-10-19 | Broad Institute Clinical Research Sequencing Platform | Infectious Disease Program, Broad Institute of Harvard and MIT | Siddle,K.J., Adams,G., Pearlman,L., Gladden-Young,A., Vicente,G., Blumenstiel,B., DeFelice,M., Lee,M., McGovern,S., Lagerborg,K., Rudy,M., DeRuff,K., Carter,A., Normandin,E., Bauer,M., Reilly,S., Tomkins-Tinch,C., Loreth,C., Chaluvadi,S., Meldrim,J., Gran |
| hCoV-19/Rwanda/NRL-CV50789-10/2021 | EPI_ISL_5633456 | 2021-08-09 | Nucleic Acid Testing Laboratory-Rwanda Biomedical Centre | Nucleic Acid Testing Laboratory-Rwanda Biomedical Centre | Dieudonne Mutangana, Enatha Mukantwari, Umuringa Jeanne d'Arc, Sindayiheba Reuben, Arlene Uwituze, Murebwayire Clarisse, Izuwayo Gerard, Alice Kabanda, Esperance Umumararungu, Robert Rutayisire, Yvan Butera , Leon Mutesa |
| hCoV-19/Rwanda/NRL-CJ97834-10/2021 | EPI_ISL_5633462 | 2021-07-21 | Nucleic Acid Testing Laboratory-Rwanda Biomedical Centre | Nucleic Acid Testing Laboratory-Rwanda Biomedical Centre | Dieudonne Mutangana, Enatha Mukantwari, Umuringa Jeanne d'Arc, Sindayiheba Reuben, Arlene Uwituze, Murebwayire Clarisse, Izuwayo Gerard, Alice Kabanda, Esperance Umumararungu, Robert Rutayisire, Yvan Butera , Leon Mutesa |
| hCoV-19/USA/KY-LMDC-20170413/2021 | EPI_ISL_5778141 | 2021-03-10 | Louisville Metro Public Health and Wellness Department/Louisville Metro Department of Corrections | University of Louisville Sequencing Technology Center | Elizabeth Hudson,William Lauer, Leslie A. Wolf, Julia H. Chariker, Eric C. Rouchka, Melissa L. Smith |
| hCoV-19/USA/KY-LMDC-21071772/2021 | EPI_ISL_5778144 | 2021-04-26 | Louisville Metro Public Health and Wellness Department/Louisville Metro Department of Corrections | University of Louisville Sequencing Technology Center | Elizabeth Hudson,William Lauer, Leslie A. Wolf, Julia H. Chariker, Eric C. Rouchka, Melissa L. Smith |
| hCoV-19/USA/KY-LMDC-21072134/2021 | EPI_ISL_5778148 | 2021-03-30 | Louisville Metro Public Health and Wellness Department/Louisville Metro Department of Corrections | University of Louisville Sequencing Technology Center | Elizabeth Hudson,William Lauer, Leslie A. Wolf, Julia H. Chariker, Eric C. Rouchka, Melissa L. Smith |
| hCoV-19/Australia/NSW-ICPMR-11781/2021 | EPI_ISL_5778526 | 2021-10-21 | Histopath | Clinical Pathology and Medical Research; Westmead Hospital; University of Sydney | Arnott A., Draper J., Gall M., Martinez E., Rockett R., Sintchenko V., on behalf of ICPMR |
| hCoV-19/Rwanda/NRL-BS17614-11/2021 | EPI_ISL_6714839 | 2021-04-08 | Nucleic Acid Testing Laboratory-Rwanda Biomedical Centre | Nucleic Acid Testing Laboratory-Rwanda Biomedical Centre | Dieudonne Mutangana, Enatha Mukantwari, Umuringa Jeanne d'Arc, Sindayiheba Reuben, Arlene Uwituze, Murebwayire Clarisse, Izuwayo Gerard, Alice Kabanda, Esperance Umumararungu, Robert Rutayisire, Yvan Butera , Leon Mutesa |
| hCoV-19/USA/TN-VUMC-000735/2021 | EPI_ISL_5780206 | 2021-05-16 | VUMC Molecular Infectious Diseases Laboratory (MIDL) | Dr. Suman Das Lab - Vanderbilt University Medical Center (VUMC) | Grant Vestal, Hunter Brown, Meghan Shilts, Bookyung Park, Helen Boone, Suman B. Pakala, Seesandra Rajagopala, Jonathan E. Schmitz, Suman Das |
| hCoV-19/HongKong/VM21045145_WHP5043-S1/2021 | EPI_ISL_6716890 | 2021-11-20 | Hong Kong Department of Health | School of Public Health, The University of Hong Kong | Dominic N.C. Tsang, Haogao Gu, Leo L.M. Poon, Malik Peiris |
| hCoV-19/England/MILK-283223E/2021 | EPI_ISL_5785783 | 2021-10-23 | Lighthouse Lab in Milton Keynes | Wellcome Sanger Institute for the COVID-19 Genomics UK (COG-UK) Consortium | The Lighthouse Lab in Milton Keynes and Alex Alderton, Roberto Amato, Jeffrey Barrett, Sonia Goncalves, Ewan Harrison, David K. Jackson, Ian Johnston, Dominic Kwiatkowski, Cordelia Langford, John Sillitoe on behalf of the Wellcome Sanger Institute COVID-1 |

| | | | | | |
|---|---|---|---|---|---|
| hCoV-19/USA/WA-CDC-LC0340552/2021 | EPI_ISL_5878440 | 2021-10-16 | Laboratory Corporation of America | Centers for Disease Control and Prevention Division of Viral Diseases, Pathogen Discovery | Dakota Howard,Dhwani Batra,Peter Cook,Jason Caravas,Benjamin Rambo-Martin,Scott Sammons,Yvette Unoarumhi,Matthew Schmerer,Kristine Lacek,Tymeckia Kendall,Victoria Caban Figueroa,Shatavia Morrison,Christopher Gulvick,Minoo Agarwal,Eyad Almasri,Debbie Boles |
| hCoV-19/Brazil/SP-reBUT_barcode01/2021 | EPI_ISL_5802461 | 2021-03-07 | Secretaria Municipal De Saude Sorocaba | Instituto Butantan | Dimas Tadeu Covas, Antonio Jorge Martins, Claudia Renata dos Santos Barros, David Schlesinger, Debora Botequio Moretti, Elaine Cristina Marqueze, Elaine Vieira Santos, Evandra Strazza Rodrigues, Heidge Fukumasu, Jayme Augusto de Souza-Neto, Jos√© Salvator |
| hCoV-19/Brazil/SP-reBUT_barcode37/2021 | EPI_ISL_5802462 | 2021-03-07 | Secretaria Municipal De Saude Sorocaba | Instituto Butantan | Dimas Tadeu Covas, Antonio Jorge Martins, Claudia Renata dos Santos Barros, David Schlesinger, Debora Botequio Moretti, Elaine Cristina Marqueze, Elaine Vieira Santos, Evandra Strazza Rodrigues, Heidge Fukumasu, Jayme Augusto de Souza-Neto, Jos√© Salvator |
| hCoV-19/Switzerland/SH-ETHZ-34539811/2021 | EPI_ISL_6720293 | 2021-10-26 | Viollier AG | Department of Biosystems Science and Engineering, ETH Z√ºrich | Andrea Patrizia Salzmann, Henriette Kurth, Christiane Beckmann, Ivan Topolsky, Chaoran Chen, Sarah Nadeau, Louis du Plessis, Kim Philipp Jablonski, Lara Fuhrmann, Niko Beerenwinkel, Christoph Noppen, Olivier Kobel, Maurice Redondo, Tanja Stadler |
| hCoV-19/Switzerland/VD-ETHZ-34577680/2021 | EPI_ISL_6720382 | 2021-10-27 | Viollier AG | Department of Biosystems Science and Engineering, ETH Z√ºrich | Andrea Patrizia Salzmann, Henriette Kurth, Christiane Beckmann, Ivan Topolsky, Chaoran Chen, Sarah Nadeau, Louis du Plessis, Kim Philipp Jablonski, Lara Fuhrmann, Niko Beerenwinkel, Christoph Noppen, Olivier Kobel, Maurice Redondo, Tanja Stadler |
| hCoV-19/India/MH-RF-NCL-Pune-NCL-361/2021 | EPI_ISL_5798006 | 2021-03-26 | GENEPATH | NCL, Pune | Anu Raghunathan, Dhanasekaran Shanmugam, Ajinkya Khilari, Divya Niveditha, Bhagyashree Litikar, Sucheta, Shikha Takur, |
| hCoV-19/India/MH-RF-NCL-Pune-NCL-366/2021 | EPI_ISL_5798020 | 2021-03-29 | GENEPATH | NCL, Pune | Anu Raghunathan, Dhanasekaran Shanmugam, Ajinkya Khilari, Divya Niveditha, Bhagyashree Litikar, Sucheta, Shikha Takur, |
| hCoV-19/India/MH-RF-NCL-Pune-NCL-371/2021 | EPI_ISL_5798036 | 2021-03-10 | GENEPATH | NCL, Pune | Anu Raghunathan, Dhanasekaran Shanmugam, Ajinkya Khilari, Divya Niveditha, Bhagyashree Litikar, Sucheta, Shikha Takur, |
| hCoV-19/India/MH-RF-NCL-Pune-NCL-426/2021 | EPI_ISL_5798184 | 2021-03-29 | GENEPATH | NCL, Pune | Anu Raghunathan, Dhanasekaran Shanmugam, Ajinkya Khilari, Divya Niveditha, Bhagyashree Litikar, Sucheta, Shikha Takur, |
| hCoV-19/India/MH-RF-NCL-Pune-NCL-433/2021 | EPI_ISL_5798200 | 2021-03-23 | GENEPATH | NCL, Pune | Anu Raghunathan, Dhanasekaran Shanmugam, Ajinkya Khilari, Divya Niveditha, Bhagyashree Litikar, Sucheta, Shikha Takur, |
| hCoV-19/India/MH-RF-NCL-Pune-NCL-463/2021 | EPI_ISL_5798287 | 2021-08-08 | BJMC | NCL, Pune | Anu Raghunathan, Dhanasekaran Shanmugam, Ajinkya Khilari, Divya Niveditha, Bhagyashree Litikar, Sucheta, Shikha Takur, |

| | | | | | |
|---|---|---|---|---|---|
| hCoV-19/India/MH-RF-NCL-Pune-NCL-524/2021 | EPI_ISL_5798449 | 2021-09-05 | BJMC | NCL, Pune | Anu Raghunathan, Dhanasekaran Shanmugam, Ajinkya Khilari, Divya Niveditha, Bhagyashree Litikar, Sucheta, Shikha Takur, |
| hCoV-19/India/MH-RF-NCL-Pune-NCL-651/2021 | EPI_ISL_5798797 | 2021-08-26 | BJMC | NCL, Pune | Anu Raghunathan, Dhanasekaran Shanmugam, Ajinkya Khilari, Divya Niveditha, Bhagyashree Litikar, Sucheta, Shikha Takur, |
| hCoV-19/Japan/PG-146069/2021 | EPI_ISL_5799669 | 2021-09-10 | Pathogen Genomics Center, National Institute of Infectious Diseases | Pathogen Genomics Center, National Institute of Infectious Diseases | Tsuyoshi Sekizuka, Kentaro Itokawa, Rina Tanaka, Masanori Hashino, Makoto Kuroda |
| hCoV-19/Guyana/GPR0360017/2021 | EPI_ISL_6342936 | 2021-09-08 | National Public Health Reference Laboratory, Ministry of Health, | Genomics and Proteomics Departament, Gorgas Memorial Institute For Health Studies | Claudia Gonzalez, Leyda Abrego, Marlene Castillo, Oris Chavarria, Jessica Gondola, Ambar Moreno, Alexander Martinez, Shemeeza Sonaram, Cyril Giddings, Horace Cox |
| hCoV-19/Iraq/Duhok_5/2021 | EPI_ISL_5804795 | 2021-07-22 | Duhok Central public health laboratory | Duhok Central public health laboratory | Jivan Q. Ahmed, Sazan Q. Maulud |
| hCoV-19/India/PY-JsCOVb-56771_S21_R1_001/2021 | EPI_ISL_5806999 | 2021-07-03 | JIPMER Puducherry | inStem NCBS ‚Äì INSACOG | Uma Ramakrishnan Dasaradhi Palakodeti Aswin SaiNarain |
| hCoV-19/England/NORW-30902CC/2021 | EPI_ISL_5807215 | 2021-01-21 | Quadram Institute Bioscience | COVID-19 Genomics UK (COG-UK) Consortium | Dave J. Baker, Gemma L. Kay, Alp Aydin, Thanh Le-Viet, Steven Rudder, Ana P. Tedim, Anastasia Kolyva, Maria Diaz, Leonardo de Oliveira Martins, Nabil-Fareed Alikhan, Lizzie Meadows, Rachael Stanley, Rose K Davidson, Ngozi Elumogo, Muhammed Yasir, Sophie J |
| hCoV-19/India/KA-SEQ-4251_S79_R1_001/2021 | EPI_ISL_5807295 | 2021-07-03 | VRDL, Wenlock District Hospital | inStem NCBS ‚Äì INSACOG | Uma Ramakrishnan Dasaradhi Palakodeti Aswin SaiNarain |
| hCoV-19/England/PHEC-3N032N40/2021 | EPI_ISL_5810550 | 2021-10-20 | Respiratory Virus Unit, Microbiology Services Colindale, Public Health England | COVID-19 Genomics UK (COG-UK) Consortium | PHE Covid Sequencing Team |
| hCoV-19/India/TG-CDFD-C-228605/2021 | EPI_ISL_5815641 | 2021-09-19 | Gandhi Medical College | Centre for DNA Fingerprinting and Diagnostics | Murali Bashyam, Asmita Gupta, Arunkumar Karunanidhi, Vinay Donipadi, Nagamani Kammili, Divya Vashisht, Ashwin Dalal |
| hCoV-19/Canada/BC-BCCDC-221120/2021 | EPI_ISL_6726137 | 2021-10-07 | British Columbia Centre For Disease Control | BCCDC Public Health Laboratory | Prystajecky Natalie, Linda Hoang, Dan Fornika, John Tyson, Shannon Russell, Kim Macdonald, Kimia Kamelian, Ana Pacagnella, Corrinne Ng, Loretta Janz, Robert Azana, Mel Krajden |
| hCoV-19/Bonaire/BQ-RIVM-65124/2021 | EPI_ISL_5822126 | 2021-10-11 | Dutch COVID-19 response team | National Institute for Public Health and the Environment (RIVM) | Adam Meijer, Harry Vennema, Dirk Eggink, Jeroen Cremer, Sharon van den Brink, Bas van der Veer, AnneMarie van den Brandt, Lisa Wijsman, Kim Freriks, Ryanne Jaarsma, Eun√≠ce Then, Lynn Aarts, Sanne Bos, Stijn van Rossum, Florian Zwagemaker, Dennis Schmitz, |
| hCoV-19/Mozambique/COV-2302-A1950/2021 | EPI_ISL_5822654 | 2021-05-20 | Instiuto Nacional de Saude, Mozambique | Forschungszentrum Borstel | Christian Utpatel, Ivan Barilar, Stefan Niemann, Vanessa Mohr, Tanja Niemann, Jasmin Scharnberg, Sofia Viegas, Nalia Ismael, Carla Madeira, Nuro Abilio |
| hCoV-19/Mozambique/COV-2155-A1802/2021 | EPI_ISL_5822666 | 2021-05-04 | Instiuto Nacional de Saude, Mozambique | Forschungszentrum Borstel | Christian Utpatel, Ivan Barilar, Stefan Niemann, Vanessa Mohr, Tanja Niemann, Jasmin Scharnberg, Sofia Viegas, Nalia Ismael, Carla Madeira, Nuro Abilio |
| hCoV-19/Mozambique/COV-2177-A1824/2021 | EPI_ISL_5822669 | 2021-06-17 | Instiuto Nacional de Saude, Mozambique | Forschungszentrum Borstel | Christian Utpatel, Ivan Barilar, Stefan Niemann, Vanessa Mohr, Tanja Niemann, Jasmin Scharnberg, Sofia Viegas, Nalia Ismael, Carla Madeira, Nuro Abilio |

| | | | | | |
|---|---|---|---|---|---|
| hCoV-19/Mozambique/COV-2157-A1804/2021 | EPI_ISL_5822694 | 2021-05-05 | Instiuto Nacional de Saude, Mozambique | Forschungszentrum Borstel | Christian Utpatel, Ivan Barilar, Stefan Niemann, Vanessa Mohr, Tanja Niemann, Jasmin Scharnberg, Sofia Viegas, Nalia Ismael, Carla Madeira, Nuro Abilio |
| hCoV-19/Israel/CVL-19547/2021 | EPI_ISL_5826953 | 2021-08-31 | Israel Central Virology laboratory | Israel Central Virology laboratory | Neta Zuckerman, Efrat Dahan Bucris, Michal Mandelboim, Miranda Geva, Omer Asraf, Efrat Glick-Saar, Gideon Rechavi, Oran Erster, Danit Sofer, Ella Mendelson, Orna Mor |
| hCoV-19/Israel/CVL-20344/2021 | EPI_ISL_5829214 | 2021-09-12 | Israel Central Virology laboratory | Israel Central Virology laboratory | Neta Zuckerman, Efrat Dahan Bucris, Michal Mandelboim, Miranda Geva, Omer Asraf, Efrat Glick-Saar, Gideon Rechavi, Oran Erster, Danit Sofer, Ella Mendelson, Orna Mor |
| hCoV-19/Israel/CVL-19737/2021 | EPI_ISL_5829258 | 2021-09-03 | Israel Central Virology laboratory | Israel Central Virology laboratory | Neta Zuckerman, Efrat Dahan Bucris, Michal Mandelboim, Miranda Geva, Omer Asraf, Efrat Glick-Saar, Gideon Rechavi, Oran Erster, Danit Sofer, Ella Mendelson, Orna Mor |
| hCoV-19/England/PHEP-YYB4Z3Y/2021 | EPI_ISL_6280568 | 2021-10-24 | Respiratory Virus Unit, Microbiology Services Colindale, Public Health England | COVID-19 Genomics UK (COG-UK) Consortium | PHE Covid Sequencing Team |
| hCoV-19/Philippines/PH-PGC-72157/2021 | EPI_ISL_5553472 | 2021-08-17 | First Aide Diagnostic Center | Philippine Genome Center | Cynthia P. Saloma,Jan Michael C. Yap,Benedict A. Maralit,Marc Edsel C. Ayes,Eva Maria Cutiongco-de la Paz,Francis A. Tablizo,Carlo M. Lapid,El King D. Morado,Joshua Gregor A. Dizon,Maria Sofia L. Yangzon,Elcid Aaron R. Pangilinan,Renato Jacinto Q. Mantari |
| hCoV-19/Sweden/1468980664/2021 | EPI_ISL_5839143 | 2021-10-12 | Sweden/Vastragotaland/Unilabs | Unilabs/Eskilstuna/Sweden | Emma Arvidsson |
| hCoV-19/Philippines/PH-PGC-72436/2021 | EPI_ISL_5551831 | 2021-08-01 | Eastern Visayas Regional Covid-19 Testing Center | Philippine Genome Center | Cynthia P. Saloma,Jan Michael C. Yap,Benedict A. Maralit,Marc Edsel C. Ayes,Eva Maria Cutiongco-de la Paz,Francis A. Tablizo,Carlo M. Lapid,El King D. Morado,Joshua Gregor A. Dizon,Maria Sofia L. Yangzon,Elcid Aaron R. Pangilinan,Renato Jacinto Q. Mantari |
| hCoV-19/Qatar/QA.QU.20.7.B8/2021 | EPI_ISL_5852602 | 2021-09-03 | Ministry of Public Health / Hamad Medical Corporation | Biomedical Research Center (BRC), Qatar University / Qatar Genome Project (QGP) | BRC:¬†Fatiha M. Benslimane, Heba A. Al-Khatib, Oal Al-Jamal, Dana Al-Batesh, Hadi M. Yassine, Asmaa A. Al-Thani. MOPH and HMC:¬†Abdullatif Al-Khal, Muna A. S. Al-Maslamani, Mashael A. Al-Bader, Hamda Alromaihi, Roberto Bertollini, Peter V. Coyle, Einas A. |
| hCoV-19/Philippines/PH-PGC-68350/2021 | EPI_ISL_5547994 | 2021-08-06 | Dr. Jose N. Rodriguez Memorial Hospital and Sanitarium (Tala) | Philippine Genome Center | Cynthia P. Saloma,Jan Michael C. Yap,Benedict A. Maralit,Marc Edsel C. Ayes,Eva Maria Cutiongco-de la Paz,Francis A. Tablizo,Carlo M. Lapid,El King D. Morado,Joshua Gregor A. Dizon,Kenneth M. Kim,Maria Sofia L. Yangzon,Elcid Aaron R. Pangilinan,Renato Jac |
| hCoV-19/USA/VA-Noblis-S512B18/2020 | EPI_ISL_5863380 | 2020-10-29 | unknown | Noblis | Lauren Brinkac, Shiela Diepold, Shane Mitchell, Minh Tran, Lee F. Kolakowski, William M. Nelson, Katharine Jennings |

| | | | | | |
|---|---|---|---|---|---|
| hCoV-19/Brazil/SP-IAL-5309/2021 | EPI_ISL_5863679 | 2021-09-03 | Instituto Adolfo Lutz - Regional de Santos | Instituto Adolfo Lutz, Interdiciplinary Procedures Center, Strategic Laboratory | Claudio Tavares Sacchi, Claudia Regina Gon√ßalves, Karoline Rodrigues Campos, Caio Vinicius Dias Lopes, Leonardo Tadeu de Araujo, Marlon Benedito Nascimento Santos, Ariadne Ferreira Amarante |
| hCoV-19/USA/FL-BPHL-10981/2021 | EPI_ISL_5866588 | 2021-08-16 | Florida Bureau of Public Health Laboratories | Florida Bureau of Public Health Laboratories | Sarah Schmedes, Namratha Tarigopula, Tiffany Splatt, Jason Blanton |
| hCoV-19/Colombia/ANT-PRO-LDSP46/2020 | EPI_ISL_4965774 | 2020-07-19 | SURA | Laboratorio Departamental de Salud Publica de Antioquia | Idabely Betancur Ortiz, Andres F. Cardona-Rios, Maria Stella L√≥pez, Gloria Isabel Escobar, Juan P. Hernandez-Ortiz, Cristian Arbey Velarde Hoyos |
| hCoV-19/Canada/BC-BCCDC-231745/2021 | EPI_ISL_6743187 | 2021-10-14 | British Columbia Centre For Disease Control | BCCDC Public Health Laboratory | Prystajecky Natalie, Linda Hoang, Dan Fornika, John Tyson, Shannon Russell, Kim Macdonald, Kimia Kamelian, Ana Pacagnella, Corrinne Ng, Loretta Janz, Robert Azana, Mel Krajden |
| hCoV-19/USA/MN-CDC-LC0336760/2021 | EPI_ISL_5874302 | 2021-10-08 | Laboratory Corporation of America | Centers for Disease Control and Prevention Division of Viral Diseases, Pathogen Discovery | Dakota Howard,Dhwani Batra,Peter Cook,Jason Caravas,Benjamin Rambo-Martin,Scott Sammons,Yvette Unoarumhi,Matthew Schmerer,Kristine Lacek,Tymeckia Kendall,Victoria Caban Figueroa,Shatavia Morrison,Christopher Gulvick,Minoo Agarwal,Eyad Almasri,Debbie Boles |
| hCoV-19/USA/NJ-CDC-LC0331326/2021 | EPI_ISL_5877523 | 2021-10-14 | Laboratory Corporation of America | Centers for Disease Control and Prevention Division of Viral Diseases, Pathogen Discovery | Dakota Howard,Dhwani Batra,Peter Cook,Jason Caravas,Benjamin Rambo-Martin,Scott Sammons,Yvette Unoarumhi,Matthew Schmerer,Kristine Lacek,Tymeckia Kendall,Victoria Caban Figueroa,Shatavia Morrison,Christopher Gulvick,Minoo Agarwal,Eyad Almasri,Debbie Boles |
| hCoV-19/SriLanka/idea_uoc_00118/2021 | EPI_ISL_5883466 | 2021-10-04 | Department of Molecular Biology, Medical Research Institute, Colombo, Sri Lanka | IDEA Lab | Inoka C. Perera, Nadeeka Janage, Sahan Siriwardhana, Thejanee Perera, Erandika Harshani Hemamali, Rochelle Anthonies, Ramesha Dharmasiri |
| hCoV-19/Brazil/SP-IB_125523/2021 | EPI_ISL_5885344 | 2021-08-13 | CENTRO DE TRIAGEM COVID 19 | Instituto Butantan | Dimas Tadeu Covas, Antonio Jorge Martins, Claudia Renata dos Santos Barros, David Schlesinger, Debora Botequio Moretti, Elaine Cristina Marqueze, Elaine Vieira Santos, Evandra Strazza Rodrigues, Heidge Fukumasu, Jayme Augusto de Souza-Neto, Jos√© Salvator |
| hCoV-19/India/PY-COV245034_S748_R1_001/2021 | EPI_ISL_5886139 | 2021-06-28 | CBR, IISc | inStem NCBS ‚Äì INSACOG | Uma Ramakrishnan, Dasaradhi Palakodeti, Aswin SaiNarain |
| hCoV-19/USA/CA-CDC-FG-143604/2021 | EPI_ISL_5092943 | 2021-10-04 | Fulgent Genetics | Centers for Disease Control and Prevention Division of Viral Diseases, Pathogen Discovery | Dakota Howard,Dhwani Batra,Peter Cook,Jason Caravas,Benjamin Rambo-Martin,Scott Sammons,Yvette Unoarumhi,Matthew Schmerer,Kristine Lacek,Tymeckia Kendall,Victoria Caban Figueroa,Shatavia Morrison,Christopher Gulvick,Erisa Sula,Harry Gao,Mickey Li,John Gao |
| hCoV-19/India/TN-CL1720_S340_R1_001/2021 | EPI_ISL_5886340 | 2021-06-24 | CBR, IISc | inStem NCBS ‚Äì INSACOG | Uma Ramakrishnan, Dasaradhi Palakodeti, Aswin SaiNarain |
| hCoV-19/India/TN-CL1752_S372_R1_001/2021 | EPI_ISL_5886511 | 2021-06-30 | CBR, IISc | inStem NCBS ‚Äì INSACOG | Uma Ramakrishnan, Dasaradhi Palakodeti, Aswin SaiNarain |

| | | | | | |
|---|---|---|---|---|---|
| hCoV-19/India/TN-CL1126_S32_R1_001/2021 | EPI_ISL_5886597 | 2021-06-23 | CBR, IISc | inStem NCBS ‚Äì INSACOG | Uma Ramakrishnan, Dasaradhi Palakodeti, Aswin SaiNarain |
| hCoV-19/Chile/RM-HCUCH-UNAB-16905078152/2021 | EPI_ISL_5887420 | 2021-10-21 | Lab. Hospital clinico Universidad de Chile | Facultad de Ciencias de la Vida, UNAB | Claudio Meneses, Eduardo Castro, Waldo Diaz, Macarena Bastias, Ricardo Yusta, Susana Saez, Dayan Sanhueza, Sebastian Wolter, Daniel Leon, Gonzalo Campa√±a, Paola Pidal, V√≠ctor Monreal, Claudio Olmos, Ariel Orellana |
| hCoV-19/Chile/RM-USS-UNAB-ALZ683/2021 | EPI_ISL_5887438 | 2021-10-12 | Universidad San Sebastian | Facultad de Ciencias de la Vida, UNAB | Claudio Meneses, Eduardo Castro, Waldo Diaz, Macarena Bastias, Ricardo Yusta, Susana Saez, Dayan Sanhueza, Sebastian Wolter, Daniel Leon, Gonzalo Campa√±a, Paola Pidal, V√≠ctor Monreal, Claudio Olmos, Ariel Orellana |
| hCoV-19/Chile/RM-USS-UNAB-AMS431/2021 | EPI_ISL_5887455 | 2021-10-14 | Universidad San Sebastian | Facultad de Ciencias de la Vida, UNAB | Claudio Meneses, Eduardo Castro, Waldo Diaz, Macarena Bastias, Ricardo Yusta, Susana Saez, Dayan Sanhueza, Sebastian Wolter, Daniel Leon, Gonzalo Campa√±a, Paola Pidal, V√≠ctor Monreal, Claudio Olmos, Ariel Orellana |
| hCoV-19/Chile/RM-HCUCH-UNAB-16905079645/2021 | EPI_ISL_5887463 | 2021-10-22 | Lab. Hospital clinico Universidad de Chile | Facultad de Ciencias de la Vida, UNAB | Claudio Meneses, Eduardo Castro, Waldo Diaz, Macarena Bastias, Ricardo Yusta, Susana Saez, Dayan Sanhueza, Sebastian Wolter, Daniel Leon, Gonzalo Campa√±a, Paola Pidal, V√≠ctor Monreal, Claudio Olmos, Ariel Orellana |
| hCoV-19/Chile/RM-USS-UNAB-AMI058/2021 | EPI_ISL_5887467 | 2021-10-06 | Universidad San Sebastian | Facultad de Ciencias de la Vida, UNAB | Claudio Meneses, Eduardo Castro, Waldo Diaz, Macarena Bastias, Ricardo Yusta, Susana Saez, Dayan Sanhueza, Sebastian Wolter, Daniel Leon, Gonzalo Campa√±a, Paola Pidal, V√≠ctor Monreal, Claudio Olmos, Ariel Orellana |
| hCoV-19/Brazil/SP-IB_137318/2021 | EPI_ISL_5899081 | 2021-10-07 | UNIDADE RESPIRATORIA NOVA HORTOLANDIA | Instituto Butantan | Dimas Tadeu Covas, Antonio Jorge Martins, Claudia Renata dos Santos Barros, David Schlesinger, Debora Botequio Moretti, Elaine Cristina Marqueze, Elaine Vieira Santos, Evandra Strazza Rodrigues, Heidge Fukumasu, Jayme Augusto de Souza-Neto, Jos√© Salvator |
| hCoV-19/Canada/BC-BCCDC-238210/2021 | EPI_ISL_6751445 | 2021-10-27 | British Columbia Centre For Disease Control | BCCDC Public Health Laboratory | Prystajecky Natalie, Linda Hoang, Dan Fornika, John Tyson, Shannon Russell, Kim Macdonald, Kimia Kamelian, Ana Pacagnella, Corrinne Ng, Loretta Janz, Robert Azana, Mel Krajden |
| hCoV-19/USA/CA-CDPH-2000052063/2021 | EPI_ISL_5900184 | 2021-09-08 | CDPH VBL | California Department of Public Health | Emily Smith on behalf of CDPH-COVIDNet and UCLA Technology Center for Genomics & Bioinformatics |
| hCoV-19/Italy/CAM-TIGEM-IZSM-COLLI-20338/2021 | EPI_ISL_4875753 | 2021-09-24 | Lab. Microbiologia e Virologia Cotugno A.O. dei Colli | TIGEM | Antonio Grimaldi Patrizia Annunziata Francesco Panariello Claudia Tiberio Teresa Giuliano Valentina Bouche Chiara Colantuono Lucio Di Filippo Anna Manfredi Marcello Salvi Antonio Limone Luigi Atripaldi Andrea Ballabio Davide Cacchiarelli |
| hCoV-19/Botswana/R42B90_BHP_000842207/2021 | EPI_ISL_6752027 | 2021-11-21 | Botswana Harvard HIV Reference Laboratory | Botswana Harvard HIV Reference Laboratory | Sikhulile Moyo, Wonderful T. Choga, Dorcas Maruapula, Keoratile Ntshambiwa, Sefetogi Ramaologa, Thongbotho Mphoyakgosi, Boitumelo Zuze, Botshelo Radibe, Legodile Kooepile, Ontlametse T. Bareng, Pamela Smith-Lawrence, Kgomotso Moruisi, Roger Shapiro, Shahi |

| | | | | | |
|---|---|---|---|---|---|
| hCoV-19/NewZealand/21CH0379/2021 | EPI_ISL_5910719 | 2021-10-30 | Canterbury Health Laboratories | Institute of Environmental Science and Research (ESR) | Sarah Cockerton, Clare Gebbie, Dhanisha Patel, Rachel Boyle, SallyAnn Harbison, Olivia Stroeven, Xiaoyun Ren, Matt Storey, Nikki Freed, Muhammad Faisal, Jing Wang, Hermes Perez, Anja Werno, Antje van der Linden, Arlo Upton, Chris Mansell, David Hammer, Dr |
| hCoV-19/NewZealand/21CV0924/2021 | EPI_ISL_5910923 | 2021-10-15 | LabPLUS | Institute of Environmental Science and Research (ESR) | Sarah Cockerton, Clare Gebbie, Dhanisha Patel, Rachel Boyle, SallyAnn Harbison, Olivia Stroeven, Xiaoyun Ren, Matt Storey, Nikki Freed, Muhammad Faisal, Jing Wang, Hermes Perez, Anja Werno, Antje van der Linden, Arlo Upton, Chris Mansell, David Hammer, Dr |
| hCoV-19/Denmark/DCGC-197108/2021 | EPI_ISL_5915806 | 2021-10-26 | Department of Bacteria, Parasites and Fungi, Statens Serum Institut, Copenhagen, Denmark | Statens Serum Institut Bioinformatics and Microbial Genomics | Danish Covid-19 Genome Consortium |
| hCoV-19/Colombia/ANT-LDSP1003/2021 | EPI_ISL_5916023 | 2021-08-20 | LIME | Laboratorio Departamental de Salud Publica de Antioquia | Idabely Betancur Ortiz, Maria Stella Lv≥pez, Gloria Isabel Escobar, Juan P. Hernandez-Ortiz, Cristian Arbey Velarde Hoyos, Juan Pablo Isaza Agudelo, Ana Victoria Valencia Duarte |
| hCoV-19/Colombia/ANT-LDSP1052/2021 | EPI_ISL_5916125 | 2021-09-19 | SOMER | Laboratorio Departamental de Salud Publica de Antioquia | Idabely Betancur Ortiz, Maria Stella Lv≥pez, Gloria Isabel Escobar, Juan P. Hernandez-Ortiz, Cristian Arbey Velarde Hoyos, Juan Pablo Isaza Agudelo, Ana Victoria Valencia Duarte |
| hCoV-19/Romania/DJ14962/2020 | EPI_ISL_5917807 | 2020-07-21 | Medical Genetics Laboratory, Regional Centre of Medical Genetics, Emergency County Hospital Craiova | Medical Genetics Laboratory, Regional Centre of Medical Genetics, Emergency County Hospital Craiova | Anca-Lelia (Riza) Costache, Ioana Streata, Stefania Dorobantu, Adina Dragos, Mihai Cucu, Ana-Maria Buga, Razvan Plesea, Andrei Pirvu, Elena Plesea, Monica Cara, Mihai Ioana |
| hCoV-19/SouthAfrica/NICD-N18253/2021 | EPI_ISL_5918036 | 2021-09-07 | Kimberley Laboratory | Diseases of the National Health Laboratory Service | Amoako DG, Everatt J, Scheepers C, Mohale T, Ntuli N, Mahlangu B, Mnguni A, Ismail A, Bhiman JN |
| hCoV-19/Philippines/PH-PGC-64482/2021 | EPI_ISL_5544500 | 2021-07-29 | Philippine Red Cross - Port Area | Philippine Genome Center | Cynthia P. Saloma,Jan Michael C. Yap,Benedict A. Maralit,Marc Edsel C. Ayes,Eva Maria Cutiongco-de la Paz,Francis A. Tablizo,Carlo M. Lapid,El King D. Morado,Joshua Gregor A. Dizon,Kenneth M. Kim,Maria Sofia L. Yangzon,Elcid Aaron R. Pangilinan,Renato Jac |
| hCoV-19/SouthAfrica/NICD-N18051/2021 | EPI_ISL_5920314 | 2021-09-12 | Kimberley Laboratory | Diseases of the National Health Laboratory Service | Amoako DG, Everatt J, Scheepers C, Mohale T, Ntuli N, Mahlangu B, Mnguni A, Ismail A, Bhiman JN |
| hCoV-19/Italy/CAM-TIGEM-IZSM-COLLI-20650/2021 | EPI_ISL_5923091 | 2021-10-02 | Lab. Microbiologia e Virologia Cotugno A.O. dei Colli | TIGEM | Antonio Grimaldi Patrizia Annunziata Francesco Panariello Claudia Tiberio Teresa Giuliano Valentina Bouche Chiara Colantuono Lucio Di Filippo Anna Manfredi Marcello Salvi Antonio Limone Luigi Atripaldi Andrea Ballabio Davide Cacchiarelli |
| hCoV-19/Italy/CAM-TIGEM-IZSM-COLLI-20676/2021 | EPI_ISL_5923108 | 2021-10-01 | Lab. Microbiologia e Virologia Cotugno A.O. dei Colli | TIGEM | Antonio Grimaldi Patrizia Annunziata Francesco Panariello Claudia Tiberio Teresa Giuliano Valentina Bouche Chiara Colantuono Lucio Di Filippo Anna Manfredi Marcello Salvi Antonio Limone Luigi Atripaldi Andrea Ballabio Davide Cacchiarelli |

| | | | | | |
|---|---|---|---|---|---|
| hCoV-19/Italy/CAM-TIGEM-IZSM-COLLI-20805/2021 | EPI_ISL_5923160 | 2021-10-15 | Istituto Zooprofilattico Sperimentale del Mezzogiorno | TIGEM | Antonio Grimaldi Patrizia Annunziata Francesco Panariello Claudia Tiberio Teresa Giuliano Valentina Bouche Chiara Colantuono Lucio Di Filippo Anna Manfredi Marcello Salvi Antonio Limone Luigi Atripaldi Andrea Ballabio Davide Cacchiarelli |
| hCoV-19/Italy/CAM-TIGEM-IZSM-COLLI-20834/2021 | EPI_ISL_5923169 | 2021-10-16 | Istituto Zooprofilattico Sperimentale del Mezzogiorno | TIGEM | Antonio Grimaldi Patrizia Annunziata Francesco Panariello Claudia Tiberio Teresa Giuliano Valentina Bouche Chiara Colantuono Lucio Di Filippo Anna Manfredi Marcello Salvi Antonio Limone Luigi Atripaldi Andrea Ballabio Davide Cacchiarelli |
| hCoV-19/Colombia/DC-INS-VG-8481/2021 | EPI_ISL_5924530 | 2021-10-09 | COMPENSAR - LABORATORIO CLINICO LOS COBOS | Instituto Nacional de Salud-Direcci√≥n de Investigaci√≥n en Salud P√∫blica | Katherine Laiton-Donato, Carlos Franco-Mu√±oz, Diego A. √Ålvarez-D√≠az, H√©ctor Alejandro Ruiz-Moreno, Diego Andr√©s Prada, Jhonnatan Reales-Gonz√°lez, Sheryll Corchuelo, Mar√≠a T. Herrera-Sep√∫lveda, Juli√°n Naizaque, Gerardo Santamar√≠a, Jorge Rivera, P |
| hCoV-19/Morocco/IPM20376705/2021 | EPI_ISL_5924600 | 2021-10-12 | Medical Virology and BSL3 Laboratory | Centre de S√©quen√ßage G√©nomique | EL Hamouchi Adil, Charoute Hicham , Bouzidi Aymane, Amalou Ghita, Harmak Houda, Anga Latifa, Faouzi Abdellah, Zemmouri Faouzia, El Oualid Abdelmjid, Nadifiyine Saloua, Zouheir Yassine, Omondi Francis Carey, Somda Soro Georgina Charlene, Abbad Anas, Chgour |
| hCoV-19/Morocco/IPM20377157/2021 | EPI_ISL_5924607 | 2021-10-13 | Medical Virology and BSL3 Laboratory | Centre de S√©quen√ßage G√©nomique | EL Hamouchi Adil, Charoute Hicham , Bouzidi Aymane, Amalou Ghita, Harmak Houda, Anga Latifa, Faouzi Abdellah, Zemmouri Faouzia, El Oualid Abdelmjid, Nadifiyine Saloua, Zouheir Yassine, Omondi Francis Carey, Somda Soro Georgina Charlene, Abbad Anas, Chgour |
| hCoV-19/France/IDF-CERBAHC-10287496/2021 | EPI_ISL_5926666 | 2021-10-25 | LBM Bois de Verrieres | CERBA HealthCare | B√©n√©dicte Roquebert, Laura Verdurme, Sabine Trombert, St√©phanie Haim-Boukobza |
| hCoV-19/Bahrain/21921809543/2021 | EPI_ISL_5931402 | 2021-10-30 | Communicable Disease Laboratory, Public Health Directorate | Communicable Disease Laboratory, Public Health Directorate | Alwasti,H., Altaif,Z., Marhoon,A., AlHujairi,Z., AlTooq,M., Khamees,F., AlAnsari,W., AlAbbas,Z., Almoamen,G. |
| hCoV-19/USA/NJ-PHEL-V21024553/2021 | EPI_ISL_5934139 | 2021-10-26 | NJDOH, Public Health and Environmental Laboratories | NJ_PHEL | Ryan Pachucki, Shiv K. Verma, Lindsey Bodnar, Maria-Magdalene Pugliese, Chelsea San Filippo, Mohammad M. Ali, Dana Woell, Byeong Jeong |
| hCoV-19/USA/MA-CDCBI-CRSP_CCU7FPJFCOY5JVVN/2021 | EPI_ISL_5937901 | 2021-10-28 | Broad Institute Clinical Research Sequencing Platform | Infectious Disease Program, Broad Institute of Harvard and MIT | Siddle,K.J., Adams,G., Pearlman,L., Gladden-Young,A., Vicente,G., Blumenstiel,B., DeFelice,M., Lee,M., McGovern,S., Lagerborg,K., Rudy,M., DeRuff,K., Carter,A., Normandin,E., Bauer,M., Reilly,S., Tomkins-Tinch,C., Loreth,C., Chaluvadi,S., Meldrim,J., Gran |
| hCoV-19/USA/ND-UNDGC-G2021_103_4/2020 | EPI_ISL_5170767 | 2020-10-21 | Altru Health System | Genomics Core University of North Dakota | Sara Strandquist, Sara Faraji Jalal Apostal, Antariksh Tyagi, Marijo Roiko, Bony De Kumar, Sergei Nechaev |
| hCoV-19/Denmark/DCGC-198873/2021 | EPI_ISL_5944519 | 2021-10-28 | Department of Bacteria, Parasites and Fungi, Statens Serum Institut, Copenhagen, Denmark | Statens Serum Institut Bioinformatics and Microbial Genomics | Danish Covid-19 Genome Consortium |
| hCoV-19/India/LD-ICMR-NIV-INSACOG-GSEQ-5757/2021 | EPI_ISL_4950303 | 2021-09-13 | ICMR-National Institute of Virology - INSACOG | NIV Influenza | Potdar, Varsha on behalf of National Influenza Centre, NIV, Pune |

| | | | | | |
|---|---|---|---|---|---|
| hCoV-19/Australia/NSW-ICPMR-14264/2021 | EPI_ISL_6757255 | 2021-11-13 | 4Cyte Pathology | Clinical Pathology and Medical Research; Westmead Hospital; University of Sydney | Arnott A., Draper J., Gall M., Martinez E., Rockett R., Sintchenko V., on behalf of ICPMR |
| hCoV-19/Brazil/SP-IB_114930/2021 | EPI_ISL_5647657 | 2021-06-14 | UBS DE NOVA CANAA PAULISTA | Instituto Butantan | Dimas Tadeu Covas, Antonio Jorge Martins, Claudia Renata dos Santos Barros, David Schlesinger, Debora Botequio Moretti, Elaine Cristina Marqueze, Elaine Vieira Santos, Evandra Strazza Rodrigues, Heidge Fukumasu, Jayme Augusto de Souza-Neto, Jos√© Salvator |
| hCoV-19/India/MH-ICMR-NIV-INSACOG-GSEQ-5583/2021 | EPI_ISL_4950531 | 2021-08-18 | ICMR-National Institute of Virology - INSACOG | NIV Influenza | Potdar, Varsha on behalf of National Influenza Centre, NIV, Pune |
| hCoV-19/Aruba/AW-RIVM-66165/2021 | EPI_ISL_5967852 | 2021-10-14 | Dutch COVID-19 response team | National Institute for Public Health and the Environment (RIVM) | Adam Meijer, Harry Vennema, Dirk Eggink, Jeroen Cremer, Sharon van den Brink, Bas van der Veer, AnneMarie van den Brandt, Lisa Wijsman, Kim Freriks, Ryanne Jaarsma, Eun√≠ce Then, Lynn Aarts, Sanne Bos, Stijn van Rossum, Florian Zwagemaker, Dennis Schmitz, |
| hCoV-19/Curacao/CW-RIVM-66191/2021 | EPI_ISL_5968486 | 2021-10-15 | Dutch COVID-19 response team | National Institute for Public Health and the Environment (RIVM) | Adam Meijer, Harry Vennema, Dirk Eggink, Jeroen Cremer, Sharon van den Brink, Bas van der Veer, AnneMarie van den Brandt, Lisa Wijsman, Kim Freriks, Ryanne Jaarsma, Eun√≠ce Then, Lynn Aarts, Sanne Bos, Stijn van Rossum, Florian Zwagemaker, Dennis Schmitz, |
| hCoV-19/England/QEUH-288F5A2/2021 | EPI_ISL_5971465 | 2021-10-25 | Lighthouse Lab in Glasgow | Wellcome Sanger Institute for the COVID-19 Genomics UK (COG-UK) Consortium | Harper VanSteenhouse, Yumi Kasai, David Gray, Carol Clugston, Anna Dominiczak and Alex Alderton, Roberto Amato, Jeffrey Barrett, Sonia Goncalves, Ewan Harrison, David K. Jackson, Ian Johnston, Dominic Kwiatkowski, Cordelia Langford, John Sillitoe on behal |
| hCoV-19/USA/CA-CDC-FG-157438/2021 | EPI_ISL_5979187 | 2021-10-26 | Fulgent Genetics | Centers for Disease Control and Prevention Division of Viral Diseases, Pathogen Discovery | Dakota Howard,Dhwani Batra,Peter Cook,Jason Caravas,Benjamin Rambo-Martin,Scott Sammons,Yvette Unoarumhi,Matthew Schmerer,Kristine Lacek,Tymeckia Kendall,Victoria Caban Figueroa,Shatavia Morrison,Christopher Gulvick,Erisa Sula,Harry Gao,Mickey Li,John Gao |
| hCoV-19/Israel/ICH-760596351/2021 | EPI_ISL_6825615 | 2021-11-19 | Clinical Microbiology Laboratory, Tel Aviv Sourasky Medical Center | Clinical Microbiology Laboratory, Tel Aviv Sourasky Medical Center | Katya Levytskyi, Ora Halutz, Alon Ziv, Lior Handler, Amos Adler |
| hCoV-19/Australia/WA829/2021 | EPI_ISL_6586123 | 2021-11-17 | PathWest Laboratory Medicine WA | PathWest Laboratory Medicine WA | PathWest Laboratory Medicine WA Microbial Surveillance Unit |
| hCoV-19/USA/TX-CDC-QDX30320178/2021 | EPI_ISL_5982099 | 2021-10-23 | Quest Diagnostics Incorporated | Centers for Disease Control and Prevention Division of Viral Diseases, Pathogen Discovery | Dakota Howard,Dhwani Batra,Peter Cook,Jason Caravas,Benjamin Rambo-Martin,Scott Sammons,Yvette Unoarumhi,Matthew Schmerer,Kristine Lacek,Tymeckia Kendall,Victoria Caban Figueroa,Shatavia Morrison,Christopher Gulvick,Erisa Sula,S. Rosenthal,A. Gerasimova,R |
| hCoV-19/USA/CA-CDPH-1002005680/2021 | EPI_ISL_5985818 | 2021-08-21 | CDPH VBL | California Department of Public Health | CDPH-COVIDNet on behalf of Priva Path Labs Inc. |

| | | | | | |
|---|---|---|---|---|---|
| hCoV-19/USA/TX-CDC-ASC210308115/2021 | EPI_ISL_5986440 | 2021-10-03 | Aegis Sciences Corporation | Centers for Disease Control and Prevention Division of Viral Diseases, Pathogen Discovery | Dakota Howard,Dhwani Batra,Peter Cook,Jason Caravas,Benjamin Rambo-Martin,Scott Sammons,Yvette Unoarumhi,Matthew Schmerer,Kristine Lacek,Tymeckia Kendall,Victoria Caban Figueroa,Shatavia Morrison,Christopher Gulvick,Erisa Sula,Cyndi Clark,Patrick Campbell |
| hCoV-19/Iran/GRC-R9550-Tehran-LaB/2020 | EPI_ISL_5990010 | 2020-10-26 | Kariminejad-Najmabadi Pathology & Genetics Center, Tehran, Iran | of Social Welfare and Rehabilitation Sciences | Zohreh Fattahi, Marzieh Mohseni, Kimia Kahrizi, Maryam Azad, Mahdieh Koshki, Hossein Najmabadi. |
| hCoV-19/Chile/RM-PUC_MVL_1665/2021 | EPI_ISL_5994639 | 2021-09-29 | Laboratorio de Infectologia y Virologia Molecular | Laboratory of Molecular Virology, School of Medicine, Pontificia Universidad Catolica de Chile | Catalina Pardo-Roa,¬†Leonardo I. Almonacid,¬†Eugenia L. Fuentes Luppichini,¬†Estefany Poblete,¬†¬†Erick Salinas, Andres E. Munoz-Marcos,¬†Jennifer Angulo,¬†Tamara Garcia-Salum, Maria Jose Avendano, ¬†Ana Maria Contreras, Carlos Palma,¬† Jorge Levican, Eil |
| hCoV-19/Germany/BW-RKI-I-286641/2021 | EPI_ISL_5399947 | 2021-10-12 | Sonic - Labor Staber Heilbronn | Robert Koch Institute | unknown |
| hCoV-19/Nigeria/CV3494/2021 | EPI_ISL_6002895 | 2021-08-02 | Genexpert Molecular Diagnostic Laboratory | Africa Centre for Excellence for Genomics of Infectious Diseases (ACEGID), Redeemer‚Äôs University | Olawoye, I.B., Oluniyi, P.E., Eromon, P.E., Oguzie, J.U., Kayode, A.T., Uwanibe, J.N., Ugwu, C.A., Akano, K.O., Ajogbasile, F.V., Abechi, P.S., Olumade, T.J., Nosamiefan, I., Folarin, O., Happi, C.T. |
| hCoV-19/SouthAfrica/NHLS-UCT-GS-AC15/2021 | EPI_ISL_6010180 | 2021-09-27 | 2 Military Hospital wc MAA | NHLS/UCT | Arash Iranzadeh, Deelan Doolabh, Lynn Tyers, Bruna Galvao, Innocent Mudau, Marvin Hsiao, Gert Marais, Diana Hardie, Stephen Korsman, Rageema Joseph, Carolyn Williamson |
| hCoV-19/Egypt/CCHE57357_Wave_4_077/2021 | EPI_ISL_6011748 | 2021-10-23 | Genomics Program, Children Cancer Hospital | Genomics Program, Children Cancer Hospital | Hatem,A., ElHaddad,A., Abouelnaga,S., Amer,K., Elnaqeeb,M., Hassan,W., Salah,H., ElHalafawy,A., Farawela,H., Saaid,M., Hossam,M., Ismail,J., Almeldin,A., Said,D., Abdelaziz,H., Mansour,T., Shalaby,L., Soliman,S., Hassan,R., Hammad,M., Hussein,S., Abdo,I., |
| hCoV-19/Egypt/CCHE57357_Wave_4_108/2021 | EPI_ISL_6011779 | 2021-10-19 | Genomics Program, Children Cancer Hospital | Genomics Program, Children Cancer Hospital | Hatem,A., ElHaddad,A., Abouelnaga,S., Amer,K., Elnaqeeb,M., Hassan,W., Salah,H., ElHalafawy,A., Farawela,H., Saaid,M., Hossam,M., Ismail,J., Almeldin,A., Said,D., Abdelaziz,H., Mansour,T., Shalaby,L., Soliman,S., Hassan,R., Hammad,M., Hussein,S., Abdo,I., |
| hCoV-19/Egypt/CCHE57357_Wave_4_111/2021 | EPI_ISL_6011782 | 2021-10-19 | Genomics Program, Children Cancer Hospital | Genomics Program, Children Cancer Hospital | Hatem,A., ElHaddad,A., Abouelnaga,S., Amer,K., Elnaqeeb,M., Hassan,W., Salah,H., ElHalafawy,A., Farawela,H., Saaid,M., Hossam,M., Ismail,J., Almeldin,A., Said,D., Abdelaziz,H., Mansour,T., Shalaby,L., Soliman,S., Hassan,R., Hammad,M., Hussein,S., Abdo,I., |
| hCoV-19/Sweden/SUS0005359/2021 | EPI_ISL_6012032 | 2021-10-26 | Clinical Microbiology, Infection Prevention and Control | Section for Molecular Diagnostics | Bj√∂rn Hallstr√∂m, Jonas Bj√∂rkman |
| hCoV-19/Portugal/PT19779/2021 | EPI_ISL_6012549 | 2021-10-22 | SYNLAB | Instituto Nacional de Saude (INSA) | Borges et al |

| | | | | | |
|---|---|---|---|---|---|
| hCoV-19/England/NORW-309AF90/2021 | EPI_ISL_6019468 | 2021-10-20 | Quadram Institute Bioscience | COVID-19 Genomics UK (COG-UK) Consortium | Dave J. Baker, Gemma L. Kay, Alp Aydin, Thanh Le-Viet, Steven Rudder, Ana P. Tedim, Anastasia Kolyva, Maria Diaz, Leonardo de Oliveira Martins, Nabil-Fareed Alikhan, Lizzie Meadows, Rachael Stanley, Rose K Davidson, Ngozi Elumogo, Muhammed Yasir, Sophie J |
| hCoV-19/England/NORW-30A0375/2021 | EPI_ISL_6020893 | 2021-02-04 | Quadram Institute Bioscience | COVID-19 Genomics UK (COG-UK) Consortium | Dave J. Baker, Gemma L. Kay, Alp Aydin, Thanh Le-Viet, Steven Rudder, Ana P. Tedim, Anastasia Kolyva, Maria Diaz, Leonardo de Oliveira Martins, Nabil-Fareed Alikhan, Lizzie Meadows, Rachael Stanley, Rose K Davidson, Ngozi Elumogo, Muhammed Yasir, Sophie J |
| hCoV-19/India/DL-ILBS-106333/2021 | EPI_ISL_6023932 | 2021-04-14 | Clinical Virology Laboratory, ILBS | ILBS | Ekta Gupta, Varun Suroliya, Reshu Agarwal, Diptanu Paul, Amit Pandey, Chhagan Bihari Sharma, Shiv Kumar Sarin |
| hCoV-19/England/PHEC-3P069P84/2021 | EPI_ISL_6024251 | 2021-10-19 | Respiratory Virus Unit, Microbiology Services Colindale, Public Health England | COVID-19 Genomics UK (COG-UK) Consortium | PHE Covid Sequencing Team |
| hCoV-19/Belgium/CHUNamur13754160/2021 | EPI_ISL_6761095 | 2021-11-14 | Plateforme de testing Namuroise | Plateforme de testing Namuroise | Gilliard Nicolas, Drugmand Jonathan; Demars Aurore; Maschietto C√©line; Nobis Chlo√©; Otto Gaetan; Denis Olivier; Degosserie Jonathan; Mullier Fran√ßois |
| hCoV-19/Malaysia/IMR_WC578270/2021 | EPI_ISL_6774033 | 2021-11-05 | Selangor State Health Department | Infectious Disease Research Centre, National Institutes of Health, Ministry of Health Malaysia | Kamel K, Suppiah J, Mohd Zawawi Z, Azizan MA, Ramly N, Robert F, Anasir MI, Ahmad Fazilah NA, Ahmad FA, Norhisham SN, Mohamad Sukri MZ, Rosli NR, Thayan R |
| hCoV-19/Malaysia/IMR_CV02233/2021 | EPI_ISL_6774035 | 2021-10-28 | Seberang Perai Tengah District Health Office | Infectious Disease Research Centre, National Institutes of Health, Ministry of Health Malaysia | Kamel K, Suppiah J, Mohd Zawawi Z, Azizan MA, Ramly N, Robert F, Anasir MI, Ahmad Fazilah NA, Ahmad FA, Norhisham SN, Mohamad Sukri MZ, Rosli NR, Thayan R |
| hCoV-19/Botswana/R43B67_BHP_2021010142/2021 | EPI_ISL_6774087 | 2021-11-23 | Botswana Harvard HIV Reference Laboratory | Botswana Harvard HIV Reference Laboratory | Sikhulile Moyo, Wonderful T. Choga, Dorcas Maruapula, Keoratile Ntshambiwa, Sefetogi Ramaologa, Thongbotho Mphoyakgosi, Boitumelo Zuze, Botshelo Radibe, Legodile Kooepile, Ontlametse T. Bareng, Pamela Smith-Lawrence, Kgomotso Moruisi, Roger Shapiro, Shahi |
| hCoV-19/Georgia/Tb-sNGS1808/2021 | EPI_ISL_6777285 | 2021-11-04 | Department for Virology, Molecular Biology and Genome Research, R. G. Lugar Center for Public Health Research, National Center for Disease Control and Public Health (NCDC) of Georgia. | Department for Virology, Molecular Biology and Genome Research, R. G. Lugar Center for Public Health Research, National Center for Disease Control and Public Health (NCDC) of Georgia. | Giorgi Tomashvili, Giorgi Gogoladze, Meri Pantsulaia, Gvantsa Brachveli, Salome Javashvili, Nino Berishvili, Tata Imnadze, Ana Papkiauri, Gvantsa Chanturia, Ann Machablishvili, Nato Kotaria, Marine Murtskhvaladze, Lela Sabadze, Mari Gavashelidze, Tamar Ja |
| hCoV-19/Switzerland/GE-HUG-36051624/2021 | EPI_ISL_6778108 | 2021-11-15 | University Hospitals of Geneva, Laboratory of Virology | HUG, Laboratory of Virology and the Health2030 Genome Center | Samuel Cordey, Aline Mamin, Francisco Perez, Ana Rita Goncalves, Laurent Kaiser, Lorenzo Cerutti, Cedric Howald, Henri Pegeot, Melyssa Elies, Deborah Penet, Keith Harshman, Ioannis Xenarios |
| hCoV-19/Reunion/CHU7211111294/2021 | EPI_ISL_6779427 | 2021-11-12 | LABM SAINT BENOIT | Laboratoire de virologie, CNR arbovirus Associ√©, Chu de la R√©union | Etienne Frumence, Nicolas M'nemosyme, Anne-Julie Gourd√©, Rubens Lhonneur, Nicolas Traversier, Marie-Christine Jaffar Bandjee |
| hCoV-19/Reunion/CHU7210815121/2021 | EPI_ISL_6779599 | 2021-08-09 | CHOR Laboratoire | Laboratoire de virologie, CNR arbovirus Associ√©, Chu de la R√©union | Etienne Frumence, Anne-Julie Gourd√©, Rubens Lhonneur, Nicolas Traversier, Marie-Christine Jaffar Bandjee |
| hCoV-19/Reunion/CHU7210731541/2021 | EPI_ISL_6779613 | 2021-07-29 | CHU - Saint-Denis | Laboratoire de virologie, CNR arbovirus Associ√©, Chu de la R√©union | Etienne Frumence, Sabrina Petit Genet, Anne-Julie Gourd√©, Rubens Lhonneur, Nicolas Traversier, Marie-Christine Jaffar Bandjee |

| | | | | | |
|---|---|---|---|---|---|
| hCoV-19/Reunion/CHU7211001842/2021 | EPI_ISL_6779774 | 2021-10-03 | CHU - Saint-Denis | Laboratoire de virologie, CNR arbovirus Associ√©, Chu de la R√©union | Etienne Frumence, Nicolas M'nemosyme, Anne-Julie Gourd√©, Rubens Lhonneur, Nicolas Traversier, Marie-Christine Jaffar Bandjee |
| hCoV-19/Reunion/CHU7211003348/2021 | EPI_ISL_6779778 | 2021-10-04 | CHU - Saint-Denis | Laboratoire de virologie, CNR arbovirus Associ√©, Chu de la R√©union | Etienne Frumence, Nicolas M'nemosyme, Anne-Julie Gourd√©, Rubens Lhonneur, Nicolas Traversier, Marie-Christine Jaffar Bandjee |
| hCoV-19/Reunion/CHU5211019202/2021 | EPI_ISL_6779829 | 2021-10-21 | CHU - Saint-Pierre | Laboratoire de virologie, CNR arbovirus Associ√©, Chu de la R√©union | Etienne Frumence, Nicolas M'nemosyme, Anne-Julie Gourd√©, Rubens Lhonneur, Nicolas Traversier, Marie-Christine Jaffar Bandjee |
| hCoV-19/USA/OH-CDC-LC0378695/2021 | EPI_ISL_6791144 | 2021-11-09 | Laboratory Corporation of America | Centers for Disease Control and Prevention Division of Viral Diseases, Pathogen Discovery | Dakota Howard,Dhwani Batra,Peter Cook,Jason Caravas,Benjamin Rambo-Martin,Scott Sammons,Yvette Unoarumhi,Matthew Schmerer,Kristine Lacek,Tymeckia Kendall,Victoria Caban Figueroa,Shatavia Morrison,Christopher Gulvick,Minoo Agarwal,Eyad Almasri,Debbie Boles |
| hCoV-19/USA/ID-IBL-771950/2021 | EPI_ISL_6795134 | 2021-01-10 | Idaho Bureau of Laboratories | Idaho Bureau of Laboratories | R. Beukelman, Matthew Charles Burns, Aimee Ceniseros, Christian Loera, Robert L. Voermans, Christopher Ball |
| hCoV-19/Israel/SMC-7022800/2021 | EPI_ISL_6795212 | 2021-11-20 | Shamir Medical Center (Asaf Harofe) | Shamir Medical Center (Asaf Harofe) | Nir Rainy,Chen Weiner,Reut Sorek Abramovich,Yevgeni Yegorov, Alona Frenkel, Anna Vishnevsky,Patricia Benveniste-Lekovitz,Abu Hamad Ramzia,Adina Bar Chaim |
| hCoV-19/SouthAfrica/CERI-KRISP-K032307/2021 | EPI_ISL_6795834 | 2021-11-17 | ZARV/NHLS,¬†Department Medical Virology,¬†University of Pretoria | CERI, Centre for Epidemic Response and Innvoation, Stellenbosch University and KRISP, KZN Research Innovation and Sequencing Platform, UKZN. | Amy Strydom, Adriano Mendes, Micheala Davids, Sim Mayaphi and Marietjie Venter, NGS-SA (Scheepers C, Hsiao M, von Gottberg A, Bhiman J, Lessells R, Bester P, Engelbrecht S, Preiser W, Mdlalose N, Mlisana K, Goedhals D, Everatt J, Amoaka D, Makatini Z, Har |
| hCoV-19/Slovakia/UVZ_PL40_B10_18444/2021 | EPI_ISL_5113977 | 2021-03-09 | Public Health Authority of the Slovak Republic | Laboratory of Genomics and Bioinformatics, Comenius University Science Park | Tatiana Sedl√°ƒçkov√°, Diana Rus≈à√°kov√°, Miroslav B√∂hmer, Anna Giƒçov√°, Jaroslav Budi≈°, Tom√°≈° Szemes, Jakub Styk |
| hCoV-19/USA/NC-CDC-ASC210095948/2021 | EPI_ISL_5583841 | 2021-09-27 | Aegis Sciences Corporation | Centers for Disease Control and Prevention Division of Viral Diseases, Pathogen Discovery | Dakota Howard,Dhwani Batra,Peter Cook,Jason Caravas,Benjamin Rambo-Martin,Scott Sammons,Yvette Unoarumhi,Matthew Schmerer,Kristine Lacek,Tymeckia Kendall,Victoria Caban Figueroa,Shatavia Morrison,Christopher Gulvick,Erisa Sula,Cyndi Clark,Patrick Campbell |
| hCoV-19/SouthAfrica/CERI-KRISP-K032187/2021 | EPI_ISL_6810481 | 2021-11-17 | National Health Laboratory Services, Virology, Charlotte Maxeke Johannesburg hospital, Parktown, Johannesburg, Gauteng | CERI, Centre for Epidemic Response and Innvoation, Stellenbosch University and KRISP, KZN Research Innovation and Sequencing Platform, UKZN. | Kathleen Subramoney, Florette Treurnicht, Avani Bharuthram, Nkhensani Mtileni, NGS-SA (Scheepers C, Hsiao M, von Gottberg A, Bhiman J, Lessells R, Bester P, Engelbrecht S, Preiser W, Mdlalose N, Mlisana K, Goedhals D, Everatt J, Amoaka D, Makatini Z, Hard |
| hCoV-19/Brazil/SP-IB_139827/2021 | EPI_ISL_6813392 | 2021-11-01 | SANTA CASA DE MISERICORDIA DE BRAGANCA PAULISTA | Instituto Butantan | Dimas Tadeu Covas, Antonio Jorge Martins, Claudia Renata dos Santos Barros, David Schlesinger, Debora Botequio Moretti, Elaine Cristina Marqueze, Elaine Vieira Santos, Evandra Strazza Rodrigues, Heidge Fukumasu, Jayme Augusto de Souza-Neto, Jos√© Salvator |

| | | | | | |
|---|---|---|---|---|---|
| hCoV-19/Brazil/SP-IB_140001/2021 | EPI_ISL_6813494 | 2021-11-01 | COMPLEXO HOSPITALAR OURO VERDE DE CAMPINAS | Instituto Butantan | Dimas Tadeu Covas, Antonio Jorge Martins, Claudia Renata dos Santos Barros, David Schlesinger, Debora Botequio Moretti, Elaine Cristina Marqueze, Elaine Vieira Santos, Evandra Strazza Rodrigues, Heidge Fukumasu, Jayme Augusto de Souza-Neto, Jos√© Salvator |
| hCoV-19/Belgium/ULB-IBC_CV2212048523/2021 | EPI_ISL_6813838 | 2021-09-08 | National Platform bis COVID ULB-IBC | National Platform bis COVID ULB-IBC | Ricardo De Mendon√ßa, Marie-Luce Delforge, Arnaud Marchant, Coralie Henin, Benoit Haerlingen |
| hCoV-19/Australia/NSW-RPAH-1934/2021 | EPI_ISL_6814923 | 2021-11-27 | New South Wales Health Pathology Royal Prince Alfred Hospital | Microbiology RPAH | Foster, C.; Au, J.; Ruiz Silva, M.; Deveson, I.; Bull, R.; Van Hal, S.; Rawlinson, W. |
| hCoV-19/Congo/FCRM-141-038/2021 | EPI_ISL_6542741 | 2021-10-26 | Fondation Congolaise pour la recherche medicale (FCRM), Francine Ntoumi | Fondation Congolaise pour la Recherche M√©dicale | Dr Batchi-Bouyou Armel Landry; Mfoutou Mapanguy Claujens Chastel; Dr. Jean Claude Djontu; Dr. Abel Lissom; Prof. Dr. Thirumalaisamy P. Velavan; Prof. Francine Ntoumi |
| hCoV-19/Chile/AN-19196954/2021 | EPI_ISL_6823632 | 2021-11-21 | Laboratorio Cl√≠nico, Secci√≥n Biolog√≠a Molecular. Hospital Dr. Carlos Cisternas, Calama. SAR Alemania. Serem√≠a Salud Antofagasta. | Laboratorio de Gen√≥mica Microbiana, Unidad de Bioqu√≠mica, Departamento Biom√©dico, Facultad de Ciencias de la Salud, | Alexandra Galetovi√ć, Jorge Araya, Camila Salazar-Ardiles, Benito G√≥mez-Silva, Milton Urrutia, Alexander P√©rez, Gabriel Pe√±a, Mart√≠n Vargas, Carlos Cornejo. |
| hCoV-19/Chile/AN-19149974/2021 | EPI_ISL_6823635 | 2021-11-19 | Laboratorio Cl√≠nico, Secci√≥n Biolog√≠a Molecular. Hospital Dr. Carlos Cisternas, Calama. Serem√≠a Salud Antofagasta. | Microbiana, Unidad de Bioqu√≠mica, Departamento Biom√©dico, Facultad de Ciencias de la Salud, Universidad de Antofagasta, Chile. Avenida Angamos 601, Antofagasta, Chile. | Alexandra Galetovi√ć, Jorge Araya, Camila Salazar-Ardiles, Benito G√≥mez-Silva, Milton Urrutia, Alexander P√©rez, Gabriel Pe√±a, Mart√≠n Vargas, Carlos Cornejo. |
| hCoV-19/Israel/ICH-760596576/2021 | EPI_ISL_6825822 | 2021-11-20 | Clinical Microbiology Laboratory, Tel Aviv Sourasky Medical Center | Clinical Microbiology Laboratory, Tel Aviv Sourasky Medical Center | Katya Levytskyi, Ora Halutz, Alon Ziv, Lior Handler, Amos Adler |
| hCoV-19/Portugal/PT21267/2021 | EPI_ISL_6826846 | 2021-11-16 | ABC Algarve | Instituto Nacional de Saude (INSA) and Institute of Biomedicine (iBiMed), Universidade de Aveiro | Borges et al |
| hCoV-19/NewZealand/21CV2114/2021 | EPI_ISL_6827875 | 2021-11-02 | LabPLUS | Institute of Environmental Science and Research (ESR) | Sarah Cockerton, Clare Gebbie, Dhanisha Patel, Rachel Boyle, SallyAnn Harbison, Olivia Stroeven, Xiaoyun Ren, Matt Storey, Nikki Freed, Muhammad Faisal, Jing Wang, Hermes Perez, Anja Werno, Antje van der Linden, Arlo Upton, Chris Mansell, David Hammer, Dr |
| hCoV-19/Australia/NSW-RPAH-1937/2021 | EPI_ISL_6829577 | 2021-11-28 | New South Wales Health Pathology Royal Prince Alfred Hospital | Microbiology RPAH | Foster, C.; Au, J.; Ruiz Silva, M.; Deveson, I.; Bull, R.; Van Hal, S.; Rawlinson, W. |
| hCoV-19/Reunion/CHU799000005251/2021 | EPI_ISL_6829617 | 2021-08-18 | CHU - Saint-Denis | Laboratoire de virologie, CNR arbovirus Associ√©, Chu de la R√©union | Etienne Frumence, Nicolas M'nemosyme, Anne-Julie Gourd√©, Rubens Lhonneur, Nicolas Traversier, Marie-Christine Jaffar Bandjee |
| hCoV-19/HongKong/VM21045834/2021 | EPI_ISL_6832108 | 2021-11-26 | Home Quarantine Taskforce | Hong Kong Department of Health | Alan K.L. Tsang, Peter C.W. Yip, Ken H.L. Ng, Edman T.K. Lam, Rickjason C.W. Chan |
| hCoV-19/Zambia/ZMBID_6555/2021 | EPI_ISL_6837264 | 2021-05-23 | Right to care Zambia | Macha Research Trust | Muleya W, Mubemba B, Katowa B, Matoba J, Mupila Z, Kwenda G, Gill CJ, Simulundu E, Mwananyanda L |
| hCoV-19/CzechRepublic/FNP0554/2021 | EPI_ISL_6841562 | 2021-09-08 | Department of Microbiology, University Hospital in Pilsen | Department of Microbiology, University Hospital in Pilsen | Katerina Chudejova, Katerina Vlkova, Vendula Heringova, Jaroslav Hrabak |

| | | | | | |
|---|---|---|---|---|---|
| hCoV-19/HongKong/HKU-211129-002/2021 | EPI_ISL_6841981 | 2021-11-21 | Department of Microbiology, The University of Hong Kong | Department of Microbiology, The University of Hong Kong | Kelvin K.W. To, Kwok-Yung Yuen |
| hCoV-19/India/GJ-INSACOG-GBRC2678/2021 | EPI_ISL_6842296 | 2021-07-10 | GMERS Medical College, Junagadh | Gujarat Biotechnology Research Centre | Bhadreshsinh Gohil,Nimesh Patel,Nitin Shukla,Ramesh Pandit,Nitin Savaliya,Priyank Chavda,Sonal Sharma ,Janvi Raval,Apurvasinh Puvar,Zarna Patel,Dinesh Kumar,Neeraj Biswas,Chaitanya Joshi,Madhvi Joshi |
| hCoV-19/Turkey/HSGM-F2046/2021 | EPI_ISL_6842562 | 2021-11-17 | Ministry of Health Turkey | Ministry of Health Turkey | Fatma Bayrakdar, Yasemin Co≈ügun, S√ºleyman Yalcin, G√ºlay Korukluoƒülu |
| hCoV-19/India/KA-RF-NCBS-RFNB-136/2021 | EPI_ISL_6851927 | 2021-08-29 | K R PURAM UPHC | National Centre for Biological Sciences, TIFR - Rockefeller Foundation | Darshan Sreenivas,Lakshminarayanan CP,Manisha Bharadwaj,Awadhesh Pandit,Satyajit Mayor,Uma Ramakrishnan,Dimple Notani |
| hCoV-19/Aruba/AW-RIVM-69703/2021 | EPI_ISL_6852658 | 2021-11-07 | Dutch COVID-19 response team | National Institute for Public Health and the Environment (RIVM) | Adam Meijer, Harry Vennema, Dirk Eggink, Jeroen Cremer, Sharon van den Brink, Bas van der Veer, AnneMarie van den Brandt, Lisa Wijsman, Kim Freriks, Ryanne Jaarsma, Lynn Aarts, Sanne Bos, Stijn van Rossum, Linda van Someren, Florian Zwagemaker, Dennis Sch |
| hCoV-19/Curacao/CW-RIVM-69945/2021 | EPI_ISL_6852926 | 2021-11-05 | Dutch COVID-19 response team | National Institute for Public Health and the Environment (RIVM) | Adam Meijer, Harry Vennema, Dirk Eggink, Jeroen Cremer, Sharon van den Brink, Bas van der Veer, AnneMarie van den Brandt, Lisa Wijsman, Kim Freriks, Ryanne Jaarsma, Lynn Aarts, Sanne Bos, Stijn van Rossum, Linda van Someren, Florian Zwagemaker, Dennis Sch |
| hCoV-19/Curacao/CW-RIVM-69947/2021 | EPI_ISL_6852928 | 2021-11-06 | Dutch COVID-19 response team | National Institute for Public Health and the Environment (RIVM) | Adam Meijer, Harry Vennema, Dirk Eggink, Jeroen Cremer, Sharon van den Brink, Bas van der Veer, AnneMarie van den Brandt, Lisa Wijsman, Kim Freriks, Ryanne Jaarsma, Lynn Aarts, Sanne Bos, Stijn van Rossum, Linda van Someren, Florian Zwagemaker, Dennis Sch |
| hCoV-19/Curacao/CW-RIVM-69949/2021 | EPI_ISL_6852930 | 2021-11-06 | Dutch COVID-19 response team | National Institute for Public Health and the Environment (RIVM) | Adam Meijer, Harry Vennema, Dirk Eggink, Jeroen Cremer, Sharon van den Brink, Bas van der Veer, AnneMarie van den Brandt, Lisa Wijsman, Kim Freriks, Ryanne Jaarsma, Lynn Aarts, Sanne Bos, Stijn van Rossum, Linda van Someren, Florian Zwagemaker, Dennis Sch |
| hCoV-19/Israel/SMC-7016338/2021 | EPI_ISL_6089892 | 2021-09-09 | Shamir Medical Center (Asaf Harofe) | Shamir Medical Center (Asaf Harofe) | Nir Rainy,Chen Weiner,Reut Sorek Abramovich,Yevgeni Yegorov,Anna Vishnevsky,Patricia Benveniste-Lekovitz,Abu Hamad Ramzia,Adina Bar Chaim |
| hCoV-19/Israel/SMC-7021730/2021 | EPI_ISL_6860176 | 2021-11-03 | Shamir Medical Center (Asaf Harofe) | Shamir Medical Center (Asaf Harofe) | Nir Rainy,Chen Weiner,Reut Sorek Abramovich,Yevgeni Yegorov,Anna Vishnevsky,Patricia Benveniste-Lekovitz,Abu Hamad Ramzia,Adina Bar Chaim |
| hCoV-19/Israel/SMC-7022587/2021 | EPI_ISL_6860346 | 2021-11-17 | Shamir Medical Center (Asaf Harofe) | Shamir Medical Center (Asaf Harofe) | Nir Rainy,Chen Weiner,Reut Sorek Abramovich,Yevgeni Yegorov,Anna Vishnevsky,Patricia Benveniste-Lekovitz,Abu Hamad Ramzia,Adina Bar Chaim |

| | | | | | |
|---|---|---|---|---|---|
| hCoV-19/Israel/SMC-7022338/2021 | EPI_ISL_6860593 | 2021-11-15 | Shamir Medical Center (Asaf Harofe) | Shamir Medical Center (Asaf Harofe) | Nir Rainy,Chen Weiner,Reut Sorek Abramovich,Yevgeni Yegorov,Anna Vishnevsky,Patricia Benveniste-Lekovitz,Abu Hamad Ramzia,Adina Bar Chaim |
| hCoV-19/USA/TX-HMH-MCoV-45481/2020 | EPI_ISL_5074140 | 2020-07-31 | Houston Methodist Hospital | Houston Methodist Hospital | Randall J. Olsen, Paul A. Christensen, S. Wesley Long, Sishir Subedi, Robert Olson, Marcus Nguyen, James J. Davis, Matthew Ojeda Saavedra, Prasanti Yerramilli, Layne Pruitt, Kristina Reppond, Madison N. Shyer, Jessica Cambric, Ryan Gadd, Ilya J. Finkels |
| hCoV-19/Israel/SMC-7016404/2021 | EPI_ISL_6090479 | 2021-09-09 | Shamir Medical Center (Asaf Harofe) | Shamir Medical Center (Asaf Harofe) | Nir Rainy,Chen Weiner,Reut Sorek Abramovich,Yevgeni Yegorov,Anna Vishnevsky,Patricia Benveniste-Lekovitz,Abu Hamad Ramzia,Adina Bar Chaim |
| hCoV-19/USA/CA-CDPH-3000267595/2021 | EPI_ISL_6862715 | 2021-10-31 | CDPH VBL | California Department of Public Health | Emily Smith on behalf of CDPH-COVIDNet |
| hCoV-19/Australia/NSW-ICPMR-14637/2021 | EPI_ISL_6864915 | 2021-11-28 | Pathology West - NSW Health Pathology | Clinical Pathology and Medical Research; Westmead Hospital; University of Sydney | Arnott A., Draper J., Gall M., Martinez E., Rockett R., Sintchenko V., on behalf of ICPMR |
| hCoV-19/Norway/26496/2021 | EPI_ISL_6864956 | 2021-01-11 | Haugesund Hospital, laboratory for Medical Microbiology | Norwegian Institute of Public Health, Department of Virology | Kathrine Stene-Johansen, Kamilla Heddeland Instefjord, Hilde Elshaug, Garcia Llorente Ignacio, Jon BrV•te, Engebretsen Serina Beate, Pedersen Benedikte Nevjen, Line Victoria Moen, Debech Nadia, Atiya R Ali, Marie Paulsen Madsen, Rasmus Riis Kopperud, Hild |
| hCoV-19/India/AP-CCMB-CIB2502/2021 | EPI_ISL_6877402 | 2021-10-02 | AP SSO | CSIR-Centre for Cellular and Molecular Biology-INSACOG | Malini Nemalikanti,Lamuk Zaveri,Tulasi Nagabandi,Jandhyala Sai Krishna,Sreelekshmi MS, Ara Sreenivas,Shreekant Verma, Amareshwar Vodapalli,B Himasri,Valli Nagalakshmi Undamatla, Sumedha Avadhanula,Priya Nurkurthy,Vidhyadhari Methuku, Archana Bharadwaj Siv |
| hCoV-19/India/AP-CCMB-CIB2522/2021 | EPI_ISL_6877422 | 2021-10-14 | CSIR-Centre for Cellular and Molecular Biology | CSIR-Centre for Cellular and Molecular Biology-INSACOG | Priya Nurkurthy,Vidhyadhari Methuku,Sumedha Avadhanula,Valli Nagalakshmi Undamatla,Amareshwar Vodapalli, B Himasri,Shreekant Verma,Ara Sreenivas, Sreelekshmi MS,Malini Nemalikanti,Lamuk Zaveri,Tulasi Nagabandi,Jandhyala Sai Krishna, Archana Bharadwaj Siv |
| hCoV-19/Israel/SMC-7016565/2021 | EPI_ISL_6090813 | 2021-09-10 | Shamir Medical Center (Asaf Harofe) | Shamir Medical Center (Asaf Harofe) | Nir Rainy,Chen Weiner,Reut Sorek Abramovich,Yevgeni Yegorov,Anna Vishnevsky,Patricia Benveniste-Lekovitz,Abu Hamad Ramzia,Adina Bar Chaim |
| hCoV-19/Sweden/20977760P/2021 | EPI_ISL_6882671 | 2021-11-17 | Swedish national genomic surveillance program of SARS-CoV-2 | The Public Health Agency of Sweden | Maximilian Riess, Maria Lind Karlberg, Alma Brolund, Swedish national genomic surveillance program of SARS-CoV-2 |
| hCoV-19/India/WB-INSACOG-1931300475289/2021 | EPI_ISL_6885577 | 2021-09-28 | INSACOG-WB | National Institute of Biomedical Genomics ,Äì INSACOG | Arindam Maitra, Bhaswati Bandyopadhyay, Nidhan Kumar Biswas, Tamal Ghosh, Sreedhar Chinnaswamy, Ajay Chakraborti, Saumitra Das |
| hCoV-19/India/WB-INSACOG-1931502224299/2021 | EPI_ISL_6885970 | 2021-09-30 | INSACOG-WB | National Institute of Biomedical Genomics ,Äì INSACOG | Arindam Maitra, Bhaswati Bandyopadhyay, Nidhan Kumar Biswas, Tamal Ghosh, Sreedhar Chinnaswamy, Ajay Chakraborti, Saumitra Das |

| | | | | | |
|---|---|---|---|---|---|
| hCoV-19/India/WB-INSACOG-1931300488599/2021 | EPI_ISL_6886098 | 2021-10-16 | INSACOG-WB | National Institute of Biomedical Genomics ,Äì INSACOG | Arindam Maitra, Bhaswati Bandyopadhyay, Nidhan Kumar Biswas, Tamal Ghosh, Sreedhar Chinnaswamy, Ajay Chakraborti, Saumitra Das |
| hCoV-19/India/WB-INSACOG-1931300497880/2021 | EPI_ISL_6886106 | 2021-10-27 | INSACOG-WB | National Institute of Biomedical Genomics ,Äì INSACOG | Arindam Maitra, Bhaswati Bandyopadhyay, Nidhan Kumar Biswas, Tamal Ghosh, Sreedhar Chinnaswamy, Ajay Chakraborti, Saumitra Das |
| hCoV-19/India/WB-INSACOG-1931300474320/2021 | EPI_ISL_6886132 | 2021-09-27 | INSACOG-WB | National Institute of Biomedical Genomics ,Äì INSACOG | Arindam Maitra, Bhaswati Bandyopadhyay, Nidhan Kumar Biswas, Tamal Ghosh, Sreedhar Chinnaswamy, Ajay Chakraborti, Saumitra Das |
| hCoV-19/Italy/SIC_CQRC_3421070508/2021 | EPI_ISL_4884770 | 2021-03-16 | Laboratorio CQRC | CQRC_QUALITY CONTROL CHEMICAL BIOLOGICAL RISK_AOOR Villa Sofia Cervello Palermo | Di Gaudio F., Contino F., Brunacci G. |
| hCoV-19/Italy/SIC_CQRC_21067056/2021 | EPI_ISL_4884886 | 2021-03-16 | Laboratorio CQRC | CQRC_QUALITY CONTROL CHEMICAL BIOLOGICAL RISK_AOOR Villa Sofia Cervello Palermo | Di Gaudio F., Contino F., Brunacci G. |
| hCoV-19/USA/MI-CDC-ATM-VQK66M82G/2021 | EPI_ISL_6887737 | 2021-11-02 | Helix | Centers for Disease Control and Prevention Division of Viral Diseases, Pathogen Discovery | Dakota Howard,Dhwani Batra,Peter Cook,Jason Caravas,Benjamin Rambo-Martin,Scott Sammons,Yvette Unoarumhi,Matthew Schmerer,Kristine Lacek,Tymeckia Kendall,Victoria Caban Figueroa,Helix CA,Shatavia Morrison,Christopher Gulvick,Erisa Sula,Clinton Paden,Dunca |
| hCoV-19/USA/CO-CDC-MMB11324095/2021 | EPI_ISL_6888795 | 2021-10-28 | Mako Medical | Centers for Disease Control and Prevention Division of Viral Diseases, Pathogen Discovery | Dakota Howard,Dhwani Batra,Peter Cook,Jason Caravas,Benjamin Rambo-Martin,Scott Sammons,Yvette Unoarumhi,Matthew Schmerer,Kristine Lacek,Tymeckia Kendall,Victoria Caban Figueroa,Shatavia Morrison,Christopher Gulvick,Erisa Sula,Matthew Tugwell,Lauren Moon, |
| hCoV-19/USA/CO-CDC-MMB11340472/2021 | EPI_ISL_6888905 | 2021-10-29 | Mako Medical | Centers for Disease Control and Prevention Division of Viral Diseases, Pathogen Discovery | Dakota Howard,Dhwani Batra,Peter Cook,Jason Caravas,Benjamin Rambo-Martin,Scott Sammons,Yvette Unoarumhi,Matthew Schmerer,Kristine Lacek,Tymeckia Kendall,Victoria Caban Figueroa,Shatavia Morrison,Christopher Gulvick,Erisa Sula,Matthew Tugwell,Lauren Moon, |
| hCoV-19/Austria/AGES_676319/2021 | EPI_ISL_6891760 | 2021-11-26 | AGES-Institute for medical Microbiology and Hygiene Vienna | AGES-Institute for medical Microbiology and Hygiene Vienna | Kathrin Lippert, Vera Wallner,Elisabeth Polster,Stefanie Dobrovolny, Florian Heger,Julia Klikovits, Patrick Hyden, Alexander Indra, Peter Hufnagl |
| hCoV-19/Brazil/SP-IB_140308/2021 | EPI_ISL_6894546 | 2021-11-08 | BIOFAST | Instituto Butantan | Dimas Tadeu Covas, Antonio Jorge Martins, Claudia Renata dos Santos Barros, David Schlesinger, Debora Botequio Moretti, Elaine Cristina Marqueze, Elaine Vieira Santos, Evandra Strazza Rodrigues, Heidge Fukumasu, Jayme Augusto de Souza-Neto, Jos√© Salvator |

| | | | | | |
|---|---|---|---|---|---|
| hCoV-19/USA/MN-CDC-IBX452541060679/2021 | EPI_ISL_6897721 | 2021-11-15 | Infinity Biologix | Centers for Disease Control and Prevention Division of Viral Diseases, Pathogen Discovery | Dakota Howard,Dhwani Batra,Peter Cook,Jason Caravas,Benjamin Rambo-Martin,Scott Sammons,Yvette Unoarumhi,Matthew Schmerer,Kristine Lacek,Tymeckia Kendall,Victoria Caban Figueroa,Shatavia Morrison,Christopher Gulvick,Erisa Sula,Christian Bixby,Yihe Wang,Jo |
| hCoV-19/Kenya/ILRI_COVM01149/2021 | EPI_ISL_6095104 | 2021-04-05 | International Livestock Research Institute | International Livestock Research Institute | Samuel O. Oyola, Daniel Ouso, Gilbert Kibet, Shebbar Osiany, Edward Kiritu, Paul Dobi, Collins Muli, Patrick Amoth, Vishvanath Nene, Sonal P. Henson, Edward O. Abworo |
| hCoV-19/Kenya/ILRI_COVM01233/2021 | EPI_ISL_6096028 | 2021-01-21 | International Livestock Research Institute | International Livestock Research Institute | Samuel O. Oyola, Daniel Ouso, Gilbert Kibet, Shebbar Osiany, Edward Kiritu, Paul Dobi, Collins Muli, Patrick Amoth, Vishvanath Nene, Sonal P. Henson, Edward O. Abworo |
| hCoV-19/Kenya/ILRI_COVM01278/2021 | EPI_ISL_6096112 | 2021-08-21 | International Livestock Research Institute | International Livestock Research Institute | Samuel O. Oyola, Daniel Ouso, Gilbert Kibet, Shebbar Osiany, Edward Kiritu, Paul Dobi, Collins Muli, Patrick Amoth, Vishvanath Nene, Sonal P. Henson, Edward O. Abworo |
| hCoV-19/Kenya/ILRI_COVM01381/2021 | EPI_ISL_6096523 | 2021-08-25 | International Livestock Research Institute | International Livestock Research Institute | Samuel O. Oyola, Daniel Ouso, Gilbert Kibet, Shebbar Osiany, Edward Kiritu, Paul Dobi, Collins Muli, Patrick Amoth, Vishvanath Nene, Sonal P. Henson, Edward O. Abworo |
| hCoV-19/Israel/CVL-16513/2021 | EPI_ISL_5602490 | 2021-07-28 | Israel Central Virology laboratory | Israel Central Virology laboratory | Neta Zuckerman, Efrat Dahan Bucris, Michal Mandelboim, Miranda Geva, Omer Asraf, Efrat Glick-Saar, Gideon Rechavi, Oran Erster, Danit Sofer, Ella Mendelson, Orna Mor |
| hCoV-19/India/DL-ILBS-WGS_20/2021 | EPI_ISL_6098295 | 2021-08-24 | Mahajan Imaging Pvt. Ltd | ILBS | Ekta Gupta, Varun Suroliya, Reshu Agarwal, Diptanu Paul, Rahul Garg, Arjun Bhugra, urvinder kaur, santanu das, Pramod Gautam, Chhagan Bihari Sharma, Shiv Kumar Sarin |
| hCoV-19/Colombia/DC-LSPSDS-VG-996231/2021 | EPI_ISL_5603810 | 2021-08-20 | Compensar Calle 26 | LSPSDS | Johana Hernandez Toloza, Marcela Castano Rodriguez,Gabriela Delgado Murcia, Alejandro Gomez Lopez, Marcela Mercado |
| hCoV-19/UnitedArabEmirates/BTC-4485/2021 | EPI_ISL_5142496 | 2021-01-02 | BTC, Khalifa University | BTC, Khalifa University | Habiba Al Safar et al |
| hCoV-19/India/TN-SEQ_6070_S80_R1_001/2021 | EPI_ISL_6101009 | 2021-09-02 | Dr S Raju, Director of Public Health and Preventive Medicine | inStem NCBS ,Äì INSACOG | Uma Ramakrishnan, Dasaradhi Palakodeti, Aswin SaiNarain |
| hCoV-19/Ireland/WD-NVRL-M38IRL54880/2021 | EPI_ISL_4912186 | 2021-09-17 | National Virus Reference Laboratory | National Virus Reference Laboratory | Zoe Yandle, Charlene Bennett, Gabriel Gonzalez, Michael Carr, Jonathan Dean, Cillian F De Gascun |
| hCoV-19/UnitedArabEmirates/BTC-5011/2020 | EPI_ISL_5142896 | 2020-08-28 | BTC, Khalifa University | BTC, Khalifa University | Habiba Al Safar et al |
| hCoV-19/England/PHEP-YYNBNUU/2021 | EPI_ISL_6987767 | 2021-11-12 | Respiratory Virus Unit, Microbiology Services Colindale, Public Health England | COVID-19 Genomics UK (COG-UK) Consortium | PHE Covid Sequencing Team |

| | | | | | |
|---|---|---|---|---|---|
| hCoV-19/India/TN-SEQ_6038_S48_R1_001/2021 | EPI_ISL_6101047 | 2021-09-20 | Dr S Raju, Director of Public Health and Preventive Medicine | inStem NCBS ‚Äì INSACOG | Uma Ramakrishnan, Dasaradhi Palakodeti, Aswin SaiNarain |
| hCoV-19/USA/MI-CDC-ASC210469414/2021 | EPI_ISL_6313001 | 2021-10-18 | Aegis Sciences Corporation | Centers for Disease Control and Prevention Division of Viral Diseases, Pathogen Discovery | Dakota Howard,Dhwani Batra,Peter Cook,Jason Caravas,Benjamin Rambo-Martin,Scott Sammons,Yvette Unoarumhi,Matthew Schmerer,Kristine Lacek,Tymeckia Kendall,Victoria Caban Figueroa,Shatavia Morrison,Christopher Gulvick,Erisa Sula,Cyndi Clark,Patrick Campbell |
| hCoV-19/UnitedArabEmirates/BTC-4659/2020 | EPI_ISL_5142992 | 2020-12-20 | BTC, Khalifa University | BTC, Khalifa University | Habiba Al Safar et al |
| hCoV-19/USA/CA-CDC-IBX971745701617/2021 | EPI_ISL_4904636 | 2021-08-31 | Infinity Biologix | Centers for Disease Control and Prevention Division of Viral Diseases, Pathogen Discovery | Dakota Howard,Dhwani Batra,Peter Cook,Jason Caravas,Benjamin Rambo-Martin,Scott Sammons,Yvette Unoarumhi,Matthew Schmerer,Kristine Lacek,Tymeckia Kendall,Victoria Caban Figueroa,Shatavia Morrison,Christopher Gulvick,Erisa Sula,Christian Bixby,Yihe Wang,Jo |
| hCoV-19/Denmark/DCGC-189676/2021 | EPI_ISL_5593966 | 2021-10-19 | Department of Bacteria, Parasites and Fungi, Statens Serum Institut, Copenhagen, Denmark | Statens Serum Institut Bioinformatics and Microbial Genomics | Danish Covid-19 Genome Consortium |
| hCoV-19/USA/CA-CDPH-500019132/2021 | EPI_ISL_6568921 | 2021-09-15 | TPMG Regional Laboratory | California Department of Public Health | Emily Smith on behalf of CDPH-COVIDNet and UCR Institute for Integrative Genome Biology |
| hCoV-19/USA/TN-CDC-LC0350505/2021 | EPI_ISL_6079533 | 2021-10-18 | Laboratory Corporation of America | Centers for Disease Control and Prevention Division of Viral Diseases, Pathogen Discovery | Dakota Howard,Dhwani Batra,Peter Cook,Jason Caravas,Benjamin Rambo-Martin,Scott Sammons,Yvette Unoarumhi,Matthew Schmerer,Kristine Lacek,Tymeckia Kendall,Victoria Caban Figueroa,Shatavia Morrison,Christopher Gulvick,Minoo Agarwal,Eyad Almasri,Debbie Boles |
| hCoV-19/Bangladesh/IEDCR-icddrb-GSB17-050/2021 | EPI_ISL_5858344 | 2021-09-17 | Rangamati Medical College, Rangamati | Virology Laboratory, International Centre for Diarrhoeal Disease Research, Bangladesh (ICDDR,B) | Md. Awlad Hossain, Md. Syed Muktadir Al Sium, Shiny Talukder, Manjur Hossain Khan, Senjuti Saha, Hassan Afrad, Mohammed Ziaur Rahman, Mohammad Jubair, Mohammad Enayet Hossain, Razib Mazumder, Mohammad Shahidul Islam, Ahmed Nawsher Alam, ASM Alamgir, Mahbu |
| hCoV-19/Turkey/HSGM-E15370/2021 | EPI_ISL_6570000 | 2021-10-06 | Ministry of Health Turkey | Ministry of Health Turkey | Fatma Bayrakdar, Yasemin Cosgun, Suleyman Yalcin, Gulay Korukluoglu |
| hCoV-19/Slovakia/34246/2021 | EPI_ISL_7051296 | 2021-11-10 | Public Health Authority of the Slovak Republic | Faculty of Medicine in Martin, Comenius University | Dusan Loderer, Katarina Janikova, Ivana Kasubova, Maria Skerenova, Marian Grendar |
| hCoV-19/Chile/RM-PUC_MVL_1588/2021 | EPI_ISL_5144814 | 2021-07-01 | Laboratorio de Infectologia y Virologia Molecular | Laboratory of Molecular Virology, School of Medicine, Pontificia Universidad Catolica de Chile | Catalina Pardo-Roa, Eugenia L. Fuentes Luppichini, Constanza Martinez-Valdevenito, Jennifer Angulo, Tamara Garcia-Salum,¬† Estefany Poblete, Maria Jose Avendano, Leonardo I. Almonacid, Ana Maria Contreras, Carlos Palma,¬† Jorge Levican, Eileen Serrano, Co |
| hCoV-19/England/PHEC-3T055TEB/2021 | EPI_ISL_6978601 | 2021-10-25 | Respiratory Virus Unit, Microbiology Services Colindale, Public Health England | COVID-19 Genomics UK (COG-UK) Consortium | PHE Covid Sequencing Team |

| | | | | | |
|---|---|---|---|---|---|
| hCoV-19/Norway/23508/2021 | EPI_ISL_5623671 | 2021-10-13 | Furst Medical Laboratory | Norwegian Institute of Public Health, Department of Virology | Kathrine Stene-Johansen, Kamilla Heddeland Instefjord, Hilde Elshaug, Garcia Llorente Ignacio, Jon Br√•te, Engebretsen Serina Beate,Pedersen Benedikte Nevjen, Line Victoria Moen, Debech Nadia, Atiya R Ali,Marie Paulsen Madsen, Rasmus Riis Kopperud, Hilde |
| hCoV-19/USA/AZ-ASU22615/2021 | EPI_ISL_6573187 | 2021-11-07 | Arizona State University | Arizona State University | Steven C. Holland, Matthew F. Smith, Regan A. Sullins, LaRinda A. Holland, Nathaniel Johnson, Joshua LaBaer, Vel Murugan, Efrem S. Lim |
| hCoV-19/Brazil/BA-LACEN-BA718-291682285/2021 | EPI_ISL_5146198 | 2021-09-16 | Central Public Health Laboratory - LACEN - Bahia, Salvador, Brazil | Central Public Health Laboratory - LACEN -Bahia, Salvador, Brazil | Stephane Tosta, Luciana Oliveira, Vanessa Nardy, Gabriela Menezes,Marcela G√≥mez, Lenisa Dandara, Jaqueline Gomes, Vagner Fonseca,Marta Giovanetti,Luiz Alcantara, Felicidade Pereira, Arabela Leal |
| hCoV-19/SouthAfrica/NICD-N20471/2021 | EPI_ISL_6327870 | 2021-10-18 | KIMBERLEY LABORATORY | Diseases of the National Health Laboratory Service | Amoako DG, Everatt J, Scheepers C, Mohale T, Ntuli N, Mahlangu B, Mnguni A, Ismail A, Bhiman JN |
| hCoV-19/SouthAfrica/NICD-N14854/2021 | EPI_ISL_6327953 | 2021-08-11 | AMPATH | Diseases of the National Health Laboratory Service | Amoako DG, Everatt J, Scheepers C, Mohale T, Ntuli N, Mahlangu B, Mnguni A, Ismail A, Bhiman JN |
| hCoV-19/France/HDF-P225-21374M1466/2021 | EPI_ISL_5624543 | 2021-09-15 | CH VALENCIENNES | CHU Lille - Laboratoire de Virologie | AIT YAHYA Emilie, ALIDJINOU Enagnon Kazali, BOCKET Laurence, DEMAY Christophe, ENGELMANN Ilka, GEFFROY Sandrine, GUIGON Aur√©lie, LAMBERT Val√©rie, LAZREK Mouna, NOBILLIAUX Florian, PREVOST Brigitte, THUILLIER Caroline, TINEZ Claire |
| hCoV-19/USA/CA-CDPH-3000135318/2021 | EPI_ISL_4926087 | 2021-07-31 | CDPH VBL | California Department of Public Health | Emily Smith on behalf of CDPH-COVIDNet |
| hCoV-19/USA/CA-CDPH-3000140136/2021 | EPI_ISL_4926138 | 2021-08-04 | CDPH VBL | California Department of Public Health | Emily Smith on behalf of CDPH-COVIDNet |
| hCoV-19/Ethiopia/CERI-KRISP-K025398/2021 | EPI_ISL_5146890 | 2021-07-29 | Addis Ababa University | and Innvoation, Stellenbosch University and KRISP, KZN Research Innovation and Sequencing Platform, UKZN. | Abay Sisay, Abraham Tesfaye, Adey Feleke Desta, Giandhari Jennifer, Pillay Sureshnee, Yajna Ramphal, Naidoo Yeshnee, Tshiabuila Derek,Tegally Houriiyah, San James, Wilkinson Eduan, de Oliveira Tulio |
| hCoV-19/USA/MD-MDH-5456/2021 | EPI_ISL_5624992 | 2021-09-30 | MD PHL | Maryland Department of Health Laboratories Administration | Eric N. Keller, Jillian Loomis, Kwang Low, Terence L. Moore, Ami Patel, and Robert Myers |
| hCoV-19/Bolivia/SAN-UPCH-1002/2020 | EPI_ISL_5148501 | 2020-12-29 | Laboratorio de Diagn√≥stico e Investigaci√≥n BIOSCIENCE SRL | Laboratorio de Gen√≥mica Microbiana, Universidad Peruana Cayetano Heredia | Pablo Tsukayama, Alejandra D√°vila-Barclay, Guillermo Salvatierra, Luis Gonz√°lez, Pedro E. Romero, Diego Cuicapuza, Janet Huancachoque, Pool Marcos, Esdenka P√©rez, Mauricio Prado, Freddy Tinajeros, Paula Carballo |
| hCoV-19/France/IDF-HMN-21082270183/2021 | EPI_ISL_5626344 | 2021-08-24 | laboratoire Belle Epine | Mondor University Hospital, Assistance Publique H√¥pitaux de Paris, Universit√© Paris-Est Cr√©teil, INSERM U955 | Christophe Rodriguez, Slim Fourati, Vanessa Demontant, Guillaume Gricourt, Melissa N'Debi, Alexandre Soulier, Elisabeth Trawinski, Jean-Michel Pawlotsky |
| hCoV-19/TrinidadandTobago/98799/2021 | EPI_ISL_6250793 | 2021-10-11 | The Caribbean Public Health Agency | Carrington Lab, Department of Preclinical Sciences, Faculty of Medical Sciences, The University of the West Indies, St Augustine Campus | Nikita S. D. Sahadeo, Arianne Brown-Jordan, Sarah Hill, Vernie Ramkissoon, Anushka Ramjag, Roshan Parasram, Avery Hinds, Risha Singh, SueMin Nathaniel, Nuno Faria, Oliver Pybus, Christopher Oura, Gabriel Escobar, Christine V. F. Carrington |

| | | | | | |
|---|---|---|---|---|---|
| hCoV-19/India/TG-CDFD-50041/2021 | EPI_ISL_5058628 | 2021-04-16 | District Hospital | CDFD | Murali Bashyam, Asmita Gupta, Arunkumar Karunanidhi, Vinay Donipadi, Nagamani Kammili, Divya Vashisht, Ashwin Dalal |
| hCoV-19/Australia/NSW-ICPMR-11480/2021 | EPI_ISL_5533336 | 2021-10-19 | Histopath | Clinical Pathology and Medical Research; Westmead Hospital; University of Sydney | Arnott A., Draper J., Gall M., Martinez E., Rockett R., Sintchenko V., on behalf of ICPMR |
| hCoV-19/Egypt/CCHE57357_Wave_4_094/2021 | EPI_ISL_6011765 | 2021-10-21 | Genomics Program, Children Cancer Hospital | Genomics Program, Children Cancer Hospital | Hatem,A., ElHaddad,A., Abouelnaga,S., Amer,K., Elnaqeeb,M., Hassan,W., Salah,H., ElHalafawy,A., Farawela,H., Saaid,M., Hossam,M., Ismail,J., Almeldin,A., Said,D., Abdelaziz,H., Mansour,T., Shalaby,L., Soliman,S., Hassan,R., Hammad,M., Hussein,S., Abdo,I., |
| hCoV-19/India/MH-ICMR-NIV-INSACOG-GSEQ-5963/2021 | EPI_ISL_5536049 | 2021-08-28 | ICMR-National Institute of Virology - INSACOG | NIV Influenza | Potdar, Varsha on behalf of National Influenza Centre, NIV, Pune |
| hCoV-19/Australia/NSW-ICPMR-13200/2021 | EPI_ISL_6252013 | 2021-11-02 | Laverty Pathology | Clinical Pathology and Medical Research; Westmead Hospital; University of Sydney | Arnott A., Draper J., Gall M., Martinez E., Rockett R., Sintchenko V., on behalf of ICPMR |
| hCoV-19/SouthKorea/KDCA14680/2021 | EPI_ISL_6013327 | 2021-09-07 | Division of Emerging Infectious Diseases, Bureau of Infectious Diseases Diagnosis Control, Korea Disease Control and Prevention Agency | Diseases, Bureau of Infectious Diseases Diagnosis Control, Korea Disease Control and Prevention Agency | Ae Kyung Park, Il-Hwan Kim, Jeong-Ah Kim, Chae Young Lee, Eun-Jin Kim |
| hCoV-19/SouthKorea/KDCA14860/2021 | EPI_ISL_6013507 | 2021-08-31 | Division of Emerging Infectious Diseases, Bureau of Infectious Diseases Diagnosis Control, Korea Disease Control and Prevention Agency | Diseases, Bureau of Infectious Diseases Diagnosis Control, Korea Disease Control and Prevention Agency | Ae Kyung Park, Il-Hwan Kim, Jeong-Ah Kim, Chae Young Lee, Eun-Jin Kim |
| hCoV-19/India/MH-RF-NCL-Pune-NCL-385/2021 | EPI_ISL_5798064 | 2021-03-26 | GENEPATH | NCL, Pune | Anu Raghunathan, Dhanasekaran Shanmugam, Ajinkya Khilari, Divya Niveditha, Bhagyashree Litikar, Sucheta, Shikha Takur, |
| hCoV-19/India/MH-RF-NCL-Pune-NCL-430/2021 | EPI_ISL_5798192 | 2021-03-22 | GENEPATH | NCL, Pune | Anu Raghunathan, Dhanasekaran Shanmugam, Ajinkya Khilari, Divya Niveditha, Bhagyashree Litikar, Sucheta, Shikha Takur, |
| hCoV-19/India/MH-RF-NCL-Pune-NCL-473/2021 | EPI_ISL_5798307 | 2021-08-08 | BJMC | NCL, Pune | Anu Raghunathan, Dhanasekaran Shanmugam, Ajinkya Khilari, Divya Niveditha, Bhagyashree Litikar, Sucheta, Shikha Takur, |
| hCoV-19/India/MH-RF-NCL-Pune-NCL-490/2021 | EPI_ISL_5798349 | 2021-08-13 | BJMC | NCL, Pune | Anu Raghunathan, Dhanasekaran Shanmugam, Ajinkya Khilari, Divya Niveditha, Bhagyashree Litikar, Sucheta, Shikha Takur, |
| hCoV-19/India/AP-CCMB-CIA9294/2021 | EPI_ISL_5063915 | 2021-08-27 | AP SSO | CSIR-Centre for Cellular and Molecular Biology-INSACOG | Jandhyala Sai Krishna, Lamuk Zaveri, Tulasi Nagabandi,Sofia Banu,Payel Mukherjee, Ara Sreenivas,Shreekant Verma, Amareshwar Vodapalli ,B Himasri,Valli Nagalakshmi Undamatla, Sumedha Avadhanula, Vidhyadhari Methuku, Priya Nurkurthy, Archana Bharadwaj Siva, |
| hCoV-19/Kenya/SS2863/2020 | EPI_ISL_5797347 | 2020-12-20 | Epidemiology and Demography, KEMRI-Wellcome Trust Research Programme | Epidemiology and Demography, KEMRI-Wellcome Trust Research Programme | Githinji,G., Ochola-Oyier,I.L., Agoti,C.N., Mohammed,K.S., Mburu,M.W., Morobe,J.M., Lambisia,A.W., Makori,T., Ongera,E., Ndwiga,L., de Laurent,Z., Nokes,D.J. |

| | | | | | |
|---|---|---|---|---|---|
| hCoV-19/India/KA-SEQ_4717_S115_R1_001/2021 | EPI_ISL_6019829 | 2021-09-06 | District Surveillance Unit, Raichur | inStem NCBS ‚Äì INSACOG | Uma Ramakrishnan, Dasaradhi Palakodeti, Aswin SaiNarain |
| hCoV-19/Croatia/11044/2021 | EPI_ISL_6023169 | 2021-09-02 | Klinifçki bolnifçki centar Zagreb | Hrvatski zavod za javno zdravstvo | Irena Tabain, Ivana Ferenfçak, Josipa Kuzle, Anita Jurifá, Dragan Jurifá |
| hCoV-19/Ghana/TRA-2194/2021 | EPI_ISL_6944054 | 2021-10-28 | National Influenza Centre | National Influenza Centre | William K. Ampofo, Michael Marks, Ivy A. Asante, Sharon Hsu, Benjamiin B. Lindsey, Benjamin H. Foulkes, Mildred Adusei-Poku, Bright Adu, Linda Boatemaa, Lorreta Kwasah, Quaneeta Mohktar, Joyce Appiah-Kubi, Hilda Opoku Frempong, Keren Okyerebea Attiku, Bl |
| hCoV-19/India/DL-ILBS-B_67/2021 | EPI_ISL_6025332 | 2021-01-13 | Max Super Speciality Hospital | ILBS | Ekta Gupta, Varun Suroliya, Reshu Agarwal, Diptanu Paul, Amit Pandey, Bansidhar Tarai, Chhagan Bihari Sharma, Shiv Kumar Sarin |
| hCoV-19/India/DL-ILBS-B_84/2021 | EPI_ISL_6025404 | 2021-02-12 | Max Super Speciality Hospital | ILBS | Ekta Gupta, Varun Suroliya, Reshu Agarwal, Diptanu Paul, Amit Pandey, Bansidhar Tarai, Chhagan Bihari Sharma, Shiv Kumar Sarin |
| hCoV-19/India/DL-ILBS-B_85/2021 | EPI_ISL_6025411 | 2021-02-14 | Max Super Speciality Hospital | ILBS | Ekta Gupta, Varun Suroliya, Reshu Agarwal, Diptanu Paul, Amit Pandey, Bansidhar Tarai, Chhagan Bihari Sharma, Shiv Kumar Sarin |
| hCoV-19/USA/NY-MSHSPSP-PV39732/2021 | EPI_ISL_6493930 | 2021-09-09 | MSHS Clinical Microbiology Laboratories | MSHS Pathogen Surveillance Program | Ana S. Gonzalez-Reiche, Hala Alshammary, Mitchell J. Sullivan, Zain Khalil, Brianne Ciferri, Ajay Obla, Angela Amoako, Mahmoud Awawda, Daniel Floda, Julia Matthews, Ashley Salimbangon, Levy Sominsky, Katherine Beach, Kayla Russo, Charles Gleason, Shelcie |
| hCoV-19/Turkey/HSGM-E10703/2021 | EPI_ISL_5067198 | 2021-09-27 | Ministry of Health Turkey | Ministry of Health Turkey | Fatma Bayrakdar, Yasemin Co≈ügun, S√ºleyman Yalcin, G√ºlay Korukluoƒülu |
| hCoV-19/Israel/CVL-19371/2021 | EPI_ISL_5729412 | 2021-08-25 | Israel Central Virology laboratory | Israel Central Virology laboratory | Neta Zuckerman, Efrat Dahan Bucris, Michal Mandelboim, Miranda Geva, Omer Asraf, Efrat Glick-Saar, Gideon Rechavi, Oran Erster, Danit Sofer, Ella Mendelson, Orna Mor |
| hCoV-19/Israel/CVL-18125/2021 | EPI_ISL_5729472 | 2021-08-10 | Israel Central Virology laboratory | Israel Central Virology laboratory | Neta Zuckerman, Efrat Dahan Bucris, Michal Mandelboim, Miranda Geva, Omer Asraf, Efrat Glick-Saar, Gideon Rechavi, Oran Erster, Danit Sofer, Ella Mendelson, Orna Mor |
| hCoV-19/India/KL-GENESCOV06900/2021 | EPI_ISL_4748933 | 2021-07-14 | Kerala State SARS-COV-2 Genome Surveillance Programme (GENESCOV2) | GENESCOV2 at CSIR Institute of Genomics and Integrative Biology | Kerala State SARS-COV-2 Genome Surveillance Programme (GENESCOV2) |
| hCoV-19/India/KL-GENESCOV07262/2021 | EPI_ISL_4749291 | 2021-08-09 | Kerala State SARS-COV-2 Genome Surveillance Programme (GENESCOV2) | GENESCOV2 at CSIR Institute of Genomics and Integrative Biology | Kerala State SARS-COV-2 Genome Surveillance Programme (GENESCOV2) |
| hCoV-19/Gambia/17909/2021 | EPI_ISL_5926607 | 2021-08-09 | MRCG at LSHTM Genomics lab | MRCG at LSHTM Genomics lab | Abdul Karim sesay, Abdoulie Kanteh, Jarra Manneh, Mariama Kujabi, Bakary Sanyang,Dabiri Damilari |
| hCoV-19/Gambia/17582/2021 | EPI_ISL_5926602 | 2021-08-05 | MRCG at LSHTM Genomics lab | MRCG at LSHTM Genomics lab | Abdul Karim sesay, Abdoulie Kanteh, Jarra Manneh, Mariama Kujabi, Bakary Sanyang,Dabiri Damilari |

| | | | | | |
|---|---|---|---|---|---|
| hCoV-19/USA/TN-CDC-ASC210085583/2021 | EPI_ISL_4990924 | 2021-09-10 | Aegis Sciences Corporation | Centers for Disease Control and Prevention Division of Viral Diseases, Pathogen Discovery | Dakota Howard,Dhwani Batra,Peter Cook,Jason Caravas,Benjamin Rambo-Martin,Scott Sammons,Yvette Unoarumhi,Matthew Schmerer,Kristine Lacek,Tymeckia Kendall,Victoria Caban Figueroa,Shatavia Morrison,Christopher Gulvick,Erisa Sula,Cyndi Clark,Patrick Campbell |
| hCoV-19/Israel/CVL-18785/2021 | EPI_ISL_5731793 | 2021-08-18 | Israel Central Virology laboratory | Israel Central Virology laboratory | Neta Zuckerman, Efrat Dahan Bucris, Michal Mandelboim, Miranda Geva, Omer Asraf, Efrat Glick-Saar, Gideon Rechavi, Oran Erster, Danit Sofer, Ella Mendelson, Orna Mor |
| hCoV-19/Israel/CVL-18924/2021 | EPI_ISL_5731967 | 2021-08-19 | Israel Central Virology laboratory | Israel Central Virology laboratory | Neta Zuckerman, Efrat Dahan Bucris, Michal Mandelboim, Miranda Geva, Omer Asraf, Efrat Glick-Saar, Gideon Rechavi, Oran Erster, Danit Sofer, Ella Mendelson, Orna Mor |
| hCoV-19/Australia/NSW-ICPMR-12109/2021 | EPI_ISL_5936092 | 2021-10-18 | 4Cyte Pathology | Clinical Pathology and Medical Research; Westmead Hospital; University of Sydney | Arnott A., Draper J., Gall M., Martinez E., Rockett R., Sintchenko V., on behalf of ICPMR |
| hCoV-19/USA/MD-CDC-ASC210191994/2021 | EPI_ISL_5229904 | 2021-08-09 | Aegis Sciences Corporation | Centers for Disease Control and Prevention Division of Viral Diseases, Pathogen Discovery | Dakota Howard,Dhwani Batra,Peter Cook,Jason Caravas,Benjamin Rambo-Martin,Scott Sammons,Yvette Unoarumhi,Matthew Schmerer,Kristine Lacek,Tymeckia Kendall,Victoria Caban Figueroa,Shatavia Morrison,Christopher Gulvick,Erisa Sula,Cyndi Clark,Patrick Campbell |
| hCoV-19/Switzerland/BS-Risch-21C0305232/2021 | EPI_ISL_7062590 | 2021-11-26 | Dr. Risch Ostschweiz AG | Dr Risch Laboratory | Nadia Wohlwend, Faina Wehrli, Dominique Fabien Hilti, Sinem Kas, Martin Risch, Thomas Bodmer, Lorenz Risch |
| hCoV-19/India/TG-RFCH00570_CIB2483/2021 | EPI_ISL_6877384 | 2021-10-12 | MapMyGenome | CSIR-Centre for Cellular and Molecular Biology | Lamuk Zaveri,Tulasi Nagabandi,Jandhyala Sai Krishna,Malini Nemalikanti,Sreelekshmi MS, Ara Sreenivas,Shreekant Verma, Amareshwar Vodapalli,B Himasri,Valli Nagalakshmi Undamatla, Sumedha Avadhanula,Priya Nurkurthy,Vidhyadhari Methuku,M. Aravind Kumar, San |
| hCoV-19/India/AP-CCMB-CIB2496/2021 | EPI_ISL_6877396 | 2021-10-01 | AP SSO | CSIR-Centre for Cellular and Molecular Biology-INSACOG | Valli Nagalakshmi Undamatla,Amareshwar Vodapalli, Sumedha Avadhanula,B Himasri,Priya Nurkurthy,Vidhyadhari Methuku,Shreekant Verma,Ara Sreenivas, Sreelekshmi MS,Malini Nemalikanti,Lamuk Zaveri,Tulasi Nagabandi,Jandhyala Sai Krishna, Archana Bharadwaj Siv |
| hCoV-19/India/AP-CCMB-CIB2708/2021 | EPI_ISL_6877559 | 2021-10-11 | AP SSO | CSIR-Centre for Cellular and Molecular Biology-INSACOG | Sumedha Avadhanula,Valli Nagalakshmi Undamatla,Amareshwar Vodapalli, B Himasri,Priya Nurkurthy,Vidhyadhari Methuku,Shreekant Verma,Ara Sreenivas, Sreelekshmi MS,Malini Nemalikanti,Lamuk Zaveri,Tulasi Nagabandi,Jandhyala Sai Krishna, Archana Bharadwaj Siv |

| | | | | | |
|---|---|---|---|---|---|
| hCoV-19/USA/MN-CDC-IBX349324102571/2021 | EPI_ISL_6400594 | 2021-11-01 | Infinity Biologix | Centers for Disease Control and Prevention Division of Viral Diseases, Pathogen Discovery | Dakota Howard,Dhwani Batra,Peter Cook,Jason Caravas,Benjamin Rambo-Martin,Scott Sammons,Yvette Unoarumhi,Matthew Schmerer,Kristine Lacek,Tymeckia Kendall,Victoria Caban Figueroa,Shatavia Morrison,Christopher Gulvick,Erisa Sula,Christian Bixby,Yihe Wang,Jo |
| hCoV-19/India/AP-CCMB-CIB2758/2021 | EPI_ISL_6877599 | 2021-10-18 | AP SSO | CSIR-Centre for Cellular and Molecular Biology-INSACOG | Jandhyala Sai Krishna,Tulasi Nagabandi, Lamuk Zaveri,Malini Nemalikanti,Sreelekshmi MS, Ara Sreenivas,Shreekant Verma, Amareshwar Vodapalli,B Himasri,Valli Nagalakshmi Undamatla, Sumedha Avadhanula,Priya Nurkurthy,Vidhyadhari Methuku, Archana Bharadwaj Si |
| hCoV-19/Australia/WA827/2021 | EPI_ISL_6423034 | 2021-11-14 | PathWest Laboratory Medicine WA | PathWest Laboratory Medicine WA | PathWest Laboratory Medicine WA Microbial Surveillance Unit |
| hCoV-19/Canada/QC-L00396226001/2021 | EPI_ISL_6423194 | 2021-09-30 | Laboratoire de sant√© publique du Qu√©bec | Laboratoire de sant√© publique du Qu√©bec | Sandrine Moreira, Ioannis Ragoussis, Guillaume Bourque, Jesse Shapiro, Mark Lathrop and Michel Roger on behalf of the CoVSeQ research group |
| hCoV-19/Italy/SIC_CQRC_21065203/2021 | EPI_ISL_4884596 | 2021-03-16 | Laboratorio CQRC | CQRC_QUALITY CONTROL CHEMICAL BIOLOGICAL RISK_AOOR Villa Sofia Cervello Palermo | Di Gaudio F., Contino F., Brunacci G. |
| hCoV-19/Italy/SIC_CQRC_2191520/2021 | EPI_ISL_4884591 | 2021-03-16 | Laboratorio CQRC | CQRC_QUALITY CONTROL CHEMICAL BIOLOGICAL RISK_AOOR Villa Sofia Cervello Palermo | Di Gaudio F., Contino F., Brunacci G. |
| hCoV-19/Italy/SIC_CQRC_2200975/2021 | EPI_ISL_4884601 | 2021-03-16 | Laboratorio CQRC | CQRC_QUALITY CONTROL CHEMICAL BIOLOGICAL RISK_AOOR Villa Sofia Cervello Palermo | Di Gaudio F., Contino F., Brunacci G. |
| hCoV-19/Italy/SIC_CQRC_21071070/2021 | EPI_ISL_4884597 | 2021-03-16 | Laboratorio CQRC | CQRC_QUALITY CONTROL CHEMICAL BIOLOGICAL RISK_AOOR Villa Sofia Cervello Palermo | Di Gaudio F., Contino F., Brunacci G. |
| hCoV-19/Italy/SIC_CQRC_2202373/2021 | EPI_ISL_4884560 | 2021-03-16 | Laboratorio CQRC | CQRC_QUALITY CONTROL CHEMICAL BIOLOGICAL RISK_AOOR Villa Sofia Cervello Palermo | Di Gaudio F., Contino F., Brunacci G. |
| hCoV-19/Italy/SIC_CQRC_2206896/2021 | EPI_ISL_4884544 | 2021-03-16 | Laboratorio CQRC | CQRC_QUALITY CONTROL CHEMICAL BIOLOGICAL RISK_AOOR Villa Sofia Cervello Palermo | Di Gaudio F., Contino F., Brunacci G. |
| hCoV-19/Italy/SIC_CQRC_2188480/2021 | EPI_ISL_4884776 | 2021-03-16 | Laboratorio CQRC | CQRC_QUALITY CONTROL CHEMICAL BIOLOGICAL RISK_AOOR Villa Sofia Cervello Palermo | Di Gaudio F., Contino F., Brunacci G. |
| hCoV-19/Italy/SIC_CQRC_2193918/2021 | EPI_ISL_4884871 | 2021-03-16 | Laboratorio CQRC | CQRC_QUALITY CONTROL CHEMICAL BIOLOGICAL RISK_AOOR Villa Sofia Cervello Palermo | Di Gaudio F., Contino F., Brunacci G. |
| hCoV-19/Italy/SIC-CQRC-2434755/2021 | EPI_ISL_5034162 | 2021-10-09 | Laboratorio CQRC | CQRC_QUALITY CONTROL CHEMICAL BIOLOGICAL RISK_AOOR Villa Sofia Cervello Palermo | Di Gaudio F., Contino F., Brunacci G. |

| | | | | | |
|---|---|---|---|---|---|
| hCoV-19/Italy/SIC-CQRC-2385741/2021 | EPI_ISL_5034247 | 2021-10-09 | Laboratorio CQRC | CQRC_QUALITY CONTROL CHEMICAL BIOLOGICAL RISK_AOOR Villa Sofia Cervello Palermo | Di Gaudio F., Contino F., Brunacci G. |
| hCoV-19/Italy/SIC_CQRC_2204377/2021 | EPI_ISL_4884830 | 2021-03-16 | Laboratorio CQRC | CQRC_QUALITY CONTROL CHEMICAL BIOLOGICAL RISK_AOOR Villa Sofia Cervello Palermo | Di Gaudio F., Contino F., Brunacci G. |
| hCoV-19/Italy/SIC_CQRC_21065191/2021 | EPI_ISL_4884796 | 2021-03-16 | Laboratorio CQRC | CQRC_QUALITY CONTROL CHEMICAL BIOLOGICAL RISK_AOOR Villa Sofia Cervello Palermo | Di Gaudio F., Contino F., Brunacci G. |
| hCoV-19/Italy/SIC_CQRC_2764327/2021 | EPI_ISL_6444806 | 2021-11-06 | Laboratorio CQRC | CQRC_QUALITY CONTROL CHEMICAL BIOLOGICAL RISK_AOOR Villa Sofia Cervello Palermo | Di Gaudio F., Contino F., Brunacci G. |
| hCoV-19/Italy/SIC_CQRC_2764326/2021 | EPI_ISL_6444830 | 2021-11-06 | Laboratorio CQRC | CQRC_QUALITY CONTROL CHEMICAL BIOLOGICAL RISK_AOOR Villa Sofia Cervello Palermo | Di Gaudio F., Contino F., Brunacci G. |
| hCoV-19/Italy/LOM_Sacco_Var_T79882/2021 | EPI_ISL_5881166 | 2021-10-26 | Policlinico Ponte S. Pietro | Virology and Bioemergencies, ASST Fatebenefratelli Sacco - Sacco Hospital | Valeria Micheli |
| hCoV-19/Australia/NT353/2021 | EPI_ISL_4761381 | 2021-08-04 | Royal Darwin Hospital Pathology | Microbiological Diagnostic Unit - Public Health Laboratory (MDU-PHL) | Meumann, E., Baird, R., Seemann T., Horan, K., Sherry, N.L. |
| hCoV-19/Australia/VIC19629/2021 | EPI_ISL_4761443 | 2021-07-26 | Microbiological Diagnostic Unit - Public Health Laboratory (MDU-PHL) | Microbiological Diagnostic Unit - Public Health Laboratory (MDU-PHL) | Seemann, T., Horan, K., Sherry, N.L. |
| hCoV-19/USA/NC-CDC-ASC210323096/2021 | EPI_ISL_6644416 | 2021-09-30 | Aegis Sciences Corporation | Centers for Disease Control and Prevention Division of Viral Diseases, Pathogen Discovery | Dakota Howard,Dhwani Batra,Peter Cook,Jason Caravas,Benjamin Rambo-Martin,Scott Sammons,Yvette Unoarumhi,Matthew Schmerer,Kristine Lacek,Tymeckia Kendall,Victoria Caban Figueroa,Shatavia Morrison,Christopher Gulvick,Erisa Sula,Cyndi Clark,Patrick Campbell |
| hCoV-19/FrenchPolynesia/IPP35916/2021 | EPI_ISL_6885057 | 2021-10-13 | Outre Mer | National Reference Center for Viruses of Respiratory Infections, Institut Pasteur, Paris | Marion Barbet, Sylvie Behillil, M√©line Bizard, Angela Brisebarre, Camille Capel, Vincent Enouf, Louise Lefran√ßois, Fr√©d√©ric Lemoine, Christophe Malabat, Corinne Maufrais, Slim El Khiari, Julien Fumey, Etienne Simon-Lori√®re, Maud Vanpeene, Sylvie Van |
| hCoV-19/Canada/QC-L00383873001/2021 | EPI_ISL_6423791 | 2021-08-31 | Laboratoire de sant√© publique du Qu√©bec | Laboratoire de sant√© publique du Qu√©bec | Sandrine Moreira, Ioannis Ragoussis, Guillaume Bourque, Jesse Shapiro, Mark Lathrop and Michel Roger on behalf of the CoVSeQ research group |
| hCoV-19/Brazil/BA-FIOCRUZ-PVM86772/2021 | EPI_ISL_6908710 | 2021-01-29 | Hospital S√£o Rafael - H.S.R/BA | Plataforma de Vigilancia Molecular (PVM) - FIOCRUZ/BA | Laise de Moraes, Francys A Rangel, Eduardo Oyama, Laila A da Silva, Marina Cucco, Pedro Silveira, Camila Ara√∫jo de Lorenzo Barcia, Ana Verena Almeida Mendes, Marilia Miranda Franco, Carolina Kymie Vasques Nonaka, Bruno Solano de Freitas Souza, Tiago Graf |
| hCoV-19/Malawi/NHRL-COVseq0058/2021 | EPI_ISL_4857897 | 2021-05-31 | ZCH | National HIV Reference Laboratory, Ministry of Health, Public Health Institute of Malawi | Chilima BZ, Mvula BD, Chiwaula MJ, Bighignoli B, Malolo I, Chitenje M, Chabuka LB, Panja L, Mzumara TE, Gwayi S, Wolh S |

| | | | | | |
|---|---|---|---|---|---|
| hCoV-19/Philippines/PH-PGC-61991/2021 | EPI_ISL_5543071 | 2021-07-21 | San Antonio City of Ilagan Hospital | Philippine Genome Center | Cynthia P. Saloma,Jan Michael C. Yap,Benedict A. Maralit,Marc Edsel C. Ayes,Eva Maria Cutiongco-de la Paz,Francis A. Tablizo,Carlo M. Lapid,El King D. Morado,Joshua Gregor A. Dizon,Kenneth M. Kim,Maria Sofia L. Yangzon,Elcid Aaron R. Pangilinan,Renato Jac |
| hCoV-19/Philippines/PH-PGC-56585/2021 | EPI_ISL_5540606 | 2021-06-23 | Lung Center of the Philippines (LCP) | Philippine Genome Center | Cynthia P. Saloma,Jan Michael C. Yap,Benedict A. Maralit,Marc Edsel C. Ayes,Eva Maria Cutiongco-de la Paz,Francis A. Tablizo,Carlo M. Lapid,El King D. Morado,Joshua Gregor A. Dizon,Kenneth M. Kim,Maria Sofia L. Yangzon,Elcid Aaron R. Pangilinan,Renato Jac |
| hCoV-19/Australia/QLD2505/2021 | EPI_ISL_6258812 | 2021-11-04 | PHV-FSS | PHV-FSS | Chenwei Wang on behalf of Q-PHIRE Genomics |
| hCoV-19/India/TN-SEQ_5168_S268_R1_001/2021 | EPI_ISL_6033380 | 2021-07-30 | Dr S raju, Director of Public Health and Preventive Medicine | inStem NCBS ,Äì INSACOG | Uma Ramakrishnan, Dasaradhi Palakodeti, Aswin SaiNarain |
| hCoV-19/India/TN-SEQ_5037_S137_R1_001/2021 | EPI_ISL_6033396 | 2021-07-05 | Dr S raju, Director of Public Health and Preventive Medicine | inStem NCBS ,Äì INSACOG | Uma Ramakrishnan, Dasaradhi Palakodeti, Aswin SaiNarain |
| hCoV-19/Congo/RC-207/2021 | EPI_ISL_6951062 | 2021-10-28 | Molecular Lab, National Laboratory of Public Health | Pathogen Sequencing Lab, National Institute for Biomedical Research (INRB) | Fabien Rock Niama, Placide Mbala-Kingebeni, Edith Nkwembe, Missamou Mayengue Pembe, Brunel M Angounda, Eddy Kinganda-Lusamaki, Amuri Aziza, Francisca Muyembe Mawete, Emmanuel Lokilo Lofiko, Jean Claude Makangara, Vishnou AMPIRI, Rapha√´l Lumembe, Gabriel |
| hCoV-19/Congo/RC-201/2021 | EPI_ISL_6951086 | 2021-11-05 | Molecular Lab, National Laboratory of Public Health | Pathogen Sequencing Lab, National Institute for Biomedical Research (INRB) | Fabien Rock Niama, Placide Mbala-Kingebeni, Edith Nkwembe, Missamou Mayengue Pembe, Brunel M Angounda, Eddy Kinganda-Lusamaki, Amuri Aziza, Francisca Muyembe Mawete, Emmanuel Lokilo Lofiko, Jean Claude Makangara, Vishnou AMPIRI, Rapha√´l Lumembe, Gabriel |
| hCoV-19/Congo/RC-202/2021 | EPI_ISL_6951087 | 2021-10-29 | Molecular Lab, National Laboratory of Public Health | Pathogen Sequencing Lab, National Institute for Biomedical Research (INRB) | Fabien Rock Niama, Placide Mbala-Kingebeni, Edith Nkwembe, Missamou Mayengue Pembe, Brunel M Angounda, Eddy Kinganda-Lusamaki, Amuri Aziza, Francisca Muyembe Mawete, Emmanuel Lokilo Lofiko, Jean Claude Makangara, Vishnou AMPIRI, Rapha√´l Lumembe, Gabriel |
| hCoV-19/Philippines/PH-PGC-60177/2021 | EPI_ISL_5540128 | 2021-07-12 | Philippine Red Cross - Port Area | Philippine Genome Center | Cynthia P. Saloma,Jan Michael C. Yap,Benedict A. Maralit,Marc Edsel C. Ayes,Eva Maria Cutiongco-de la Paz,Francis A. Tablizo,Carlo M. Lapid,El King D. Morado,Joshua Gregor A. Dizon,Kenneth M. Kim,Maria Sofia L. Yangzon,Elcid Aaron R. Pangilinan,Renato Jac |
| hCoV-19/Zambia/SP37/2020 | EPI_ISL_6761027 | 2020-12-01 | Macha Research Trust | Macha Research Trust | Katowa B, Kalonda A, Matoba J, Muleya W, Mubemba B, Simulundu E |
| hCoV-19/England/PHEP-YYBYUTJ/2020 | EPI_ISL_5283939 | 2020-08-03 | Respiratory Virus Unit, Microbiology Services Colindale, Public Health England | COVID-19 Genomics UK (COG-UK) Consortium | PHE Covid Sequencing Team |

| | | | | | |
|---|---|---|---|---|---|
| hCoV-19/USA/VT-CDCBI-CRSP_6ZIIIMZIB4S4NTVW/2021 | EPI_ISL_6755713 | 2021-11-15 | Broad Institute Clinical Research Sequencing Platform | Infectious Disease Program, Broad Institute of Harvard and MIT | Siddle,K.J., Adams,G., Pearlman,L., Gladden-Young,A., Vicente,G., Blumenstiel,B., DeFelice,M., Lee,M., McGovern,S., Lagerborg,K., Rudy,M., DeRuff,K., Carter,A., Normandin,E., Bauer,M., Reilly,S., Tomkins-Tinch,C., Loreth,C., Chaluvadi,S., Meldrim,J., Gran |
| hCoV-19/England/PHEC-3K049KC2/2021 | EPI_ISL_5274500 | 2021-10-04 | Respiratory Virus Unit, Microbiology Services Colindale, Public Health England | COVID-19 Genomics UK (COG-UK) Consortium | PHE Covid Sequencing Team |
| hCoV-19/USA/RI-CDCBI-RIDOH_KQHAFHHJF2EIAV2Y/2021 | EPI_ISL_6491133 | 2021-11-06 | Rhode Island Department of Health | Infectious Disease Program, Broad Institute of Harvard and MIT | Siddle,K.J., Azevedo,K., Miller,A., Adams,G., Pearlman,L., Gladden-Young,A., Lagerborg,K., Rudy,M., DeRuff,K., Carter,A., Normandin,E., Bauer,M., Reilly,S., Tomkins-Tinch,C., Loreth,C., Chaluvadi,S., Lemieux,J.E., Birren,B.W., Sabeti,P.C., Huard,R., King, |
| hCoV-19/USA/OH-CDC-QDX28939267/2021 | EPI_ISL_5068070 | 2021-09-14 | Quest Diagnostics Incorporated | Centers for Disease Control and Prevention Division of Viral Diseases, Pathogen Discovery | Dakota Howard,Dhwani Batra,Peter Cook,Jason Caravas,Benjamin Rambo-Martin,Scott Sammons,Yvette Unoarumhi,Matthew Schmerer,Kristine Lacek,Tymeckia Kendall,Victoria Caban Figueroa,Shatavia Morrison,Christopher Gulvick,Erisa Sula,S. Rosenthal,A. Gerasimova,R |
| hCoV-19/England/PHEP-YYBYJ59/2021 | EPI_ISL_5283253 | 2021-09-27 | Respiratory Virus Unit, Microbiology Services Colindale, Public Health England | COVID-19 Genomics UK (COG-UK) Consortium | PHE Covid Sequencing Team |
| hCoV-19/NorthernMarianaIslands/CDC-2-4972261/2021 | EPI_ISL_5534629 | 2021-09-18 | Commonwealth Healthcare Center | Centers for Disease Control and Prevention Division of Viral Diseases, Pathogen Discovery | Mili Sheth,Sarah Nobles,Joseph Madden,Jasmine Padilla,Alex Burgin,Meghan Bentz,Mark Burroughs,Peter Cook,Clinton Paden,Dhwani Batra,Kristen Knipe,Dakota Howard,Yvette Unoarumhi,Matthew Schmerer,Ben Rambo-Martin,Kristine Lacek,Sam Shepard,Dave Wentworth,Vi |
| hCoV-19/Belgium/ULG-20423/2021 | EPI_ISL_4917356 | 2021-09-18 | SYNLAB | GIGA Medical Genomics | Keith Durkin, Maria Artesi, Bouchra Boujemla, Nathalie Renotte, Nadine Cambisano, S√©bastien Bontems, C√©cile Meex, Claire Gourzon√®s, Olivier Ek, Laurent Gillet, Marie-Pierre Hayette, Vincent Bours |
| hCoV-19/Belgium/rega-18455/2021 | EPI_ISL_6075852 | 2021-10-23 | KU Leuven, Rega Institute, Clinical and Epidemiological Virology | KU Leuven, Rega Institute, Clinical and Epidemiological Virology | Tony Wawina-Bokalanga, Bert Vanmechelen, Joan Marti-Carerras, Piet Maes |
| hCoV-19/USA/MD-CDC-IBX051090799770/2021 | EPI_ISL_6544214 | 2021-11-11 | Infinity Biologix | Centers for Disease Control and Prevention Division of Viral Diseases, Pathogen Discovery | Dakota Howard,Dhwani Batra,Peter Cook,Jason Caravas,Benjamin Rambo-Martin,Scott Sammons,Yvette Unoarumhi,Matthew Schmerer,Kristine Lacek,Tymeckia Kendall,Victoria Caban Figueroa,Shatavia Morrison,Christopher Gulvick,Erisa Sula,Christian Bixby,Yihe Wang,Jo |
| hCoV-19/Qatar/QA.QU.18.26.E12/2021 | EPI_ISL_5852519 | 2021-08-23 | Ministry of Public Health / Hamad Medical Corporation | Biomedical Research Center (BRC), Qatar University / Qatar Genome Project (QGP) | BRC:¬†Fatiha M. Benslimane, Heba A. Al-Khatib, Oal Al-Jamal, Dana Al-Batesh, Hadi M. Yassine, Asmaa A. Al-Thani. MOPH and HMC:¬†Abdullatif Al-Khal, Muna A. S. Al-Maslamani, Mashael A. Al-Bader, Hamda Alromaihi, Roberto Bertollini, Peter V. Coyle, Einas A. |

| | | | | | |
|---|---|---|---|---|---|
| hCoV-19/Qatar/QA.QU.18.27.H6/2021 | EPI_ISL_5852704 | 2021-09-21 | Ministry of Public Health / Hamad Medical Corporation | Biomedical Research Center (BRC), Qatar University / Qatar Genome Project (QGP) | BRC:¬†Fatiha M. Benslimane, Heba A. Al-Khatib, Oal Al-Jamal, Dana Al-Batesh, Hadi M. Yassine, Asmaa A. Al-Thani. MOPH and HMC:¬†Abdullatif Al-Khal, Muna A. S. Al-Maslamani, Mashael A. Al-Bader, Hamda Alromaihi, Roberto Bertollini, Peter V. Coyle, Einas A. |
| hCoV-19/USA/MI-MDHHS-SC34278/2021 | EPI_ISL_5367992 | 2021-09-05 | Michigan Department of Health and Human Services, Bureau of Laboratories | Human Services, Bureau of Laboratories | Blankenship HM, Riner D, Soehnlen MK |
| hCoV-19/SriLanka/aicbu2348/2021 | EPI_ISL_6810414 | 2021-11-21 | Allergy, Immunology and Cell Biology Unit (AICBU) | Allergy, Immunology and Cell Biology Unit (AICBU) | Chandima Jeewandara, Deshni Jayathilaka, Dumni Guasinghe, Tibutius Thanesh, Dinuka Ariyaratne, Ayesha Wijesinghe, Diyanath Ranasinghe, Gathsaurie Neelika Malavige |
| hCoV-19/Israel/CVL-17807/2021 | EPI_ISL_5620309 | 2021-07-18 | Israel Central Virology laboratory | Israel Central Virology laboratory | Neta Zuckerman, Efrat Dahan Bucris, Michal Mandelboim, Miranda Geva, Omer Asraf, Efrat Glick-Saar, Gideon Rechavi, Oran Erster, Danit Sofer, Ella Mendelson, Orna Mor |
| hCoV-19/Israel/CVL-17492/2021 | EPI_ISL_5620740 | 2021-08-01 | Israel Central Virology laboratory | Israel Central Virology laboratory | Neta Zuckerman, Efrat Dahan Bucris, Michal Mandelboim, Miranda Geva, Omer Asraf, Efrat Glick-Saar, Gideon Rechavi, Oran Erster, Danit Sofer, Ella Mendelson, Orna Mor |
| hCoV-19/USA/CO-CDPHE-2102227549/2021 | EPI_ISL_7048600 | 2021-11-07 | Colorado Department of Public Health and Environment | Colorado Department of Public Health and Environment | Laura Bankers, Molly C. Hetherington-Rauth, Diana Ir, Alexandria Rossheim, Michael Martin, Mandy Waters, Arianna Smith, Shannon R. Matzinger, Sarah Elizabeth Totten, Emily A. Travanty |
| hCoV-19/Israel/CVL-17648/2021 | EPI_ISL_5621066 | 2021-08-09 | Israel Central Virology laboratory | Israel Central Virology laboratory | Neta Zuckerman, Efrat Dahan Bucris, Michal Mandelboim, Miranda Geva, Omer Asraf, Efrat Glick-Saar, Gideon Rechavi, Oran Erster, Danit Sofer, Ella Mendelson, Orna Mor |
| hCoV-19/Australia/SA1019/2021 | EPI_ISL_6827510 | 2021-11-17 | SA Pathology | SA Pathology | Caitlin Selway, Lex Leong, Chuan Kok Lim, Mark Turra, Ivan Bastian |
| hCoV-19/SouthAfrica/UFS-VIRO-NGS-MSQ220921BC19/2021 | EPI_ISL_4955142 | 2021-09-03 | NHLS Universitas Academic | UFS Virology | PA Bester, MM Nyaga, P Nthiga, MT Mogotsi, Emmanuel Ogunbayo, D Goedhals, T de Oliveira |
| hCoV-19/USA/CA-CDPH-2000052089/2021 | EPI_ISL_5900210 | 2021-09-08 | CDPH VBL | California Department of Public Health | Emily Smith on behalf of CDPH-COVIDNet and UCLA Technology Center for Genomics & Bioinformatics |
| hCoV-19/Zambia/ZMBID_5579/2021 | EPI_ISL_6831757 | 2021-03-26 | Right to care Zambia | Macha Research Trust | Muleya W, Mubemba B, Katowa B, Matoba J, Mupila Z, Kwenda G, Gill CJ, Simulundu E, Mwananyanda L |
| hCoV-19/USA/CA-CDPH-500018348/2021 | EPI_ISL_5900608 | 2021-09-13 | TPMG Regional Laboratory | California Department of Public Health | Emily Smith on behalf of CDPH-COVIDNet and UCLA Technology Center for Genomics & Bioinformatics |
| hCoV-19/Denmark/DCGC-232091/2021 | EPI_ISL_6831328 | 2021-11-19 | Department of Bacteria, Parasites and Fungi, Statens Serum Institut, Copenhagen, Denmark | Statens Serum Institut Bioinformatics and Microbial Genomics | Danish Covid-19 Genome Consortium |
| hCoV-19/USA/NY-CDC-ASC210495436/2021 | EPI_ISL_6378348 | 2021-10-24 | Aegis Sciences Corporation | Centers for Disease Control and Prevention Division of Viral Diseases, Pathogen Discovery | Dakota Howard,Dhwani Batra,Peter Cook,Jason Caravas,Benjamin Rambo-Martin,Scott Sammons,Yvette Unoarumhi,Matthew Schmerer,Kristine Lacek,Tymeckia Kendall,Victoria Caban Figueroa,Shatavia Morrison,Christopher Gulvick,Erisa Sula,Cyndi Clark,Patrick Campbell |

| | | | | | |
|---|---|---|---|---|---|
| hCoV-19/SouthAfrica/NICD-N16445/2021 | EPI_ISL_5196548 | 2021-09-03 | KIMBERLEY LABORATORY | Diseases of the National Health Laboratory Service | Amoako DG, Everatt J, Scheepers C, Mohale T, Ntuli N, Mahlangu B, Mnguni A, Ismail A, Bhiman JN |
| hCoV-19/Australia/NT388/2021 | EPI_ISL_5656389 | 2021-09-20 | Royal Darwin Hospital Pathology | Microbiological Diagnostic Unit - Public Health Laboratory (MDU-PHL) | Meumann, E., Baird, R., Seemann T., Horan, K., Sherry, N.L. |
| hCoV-19/USA/VA-VCUVAS3-RRNR537502/2021 | EPI_ISL_6147641 | 2021-09-22 | Virginia Division of Consolidated Laboratory Services | Virginia Commonwealth University | Gregory A Buck, Myrna G Serrano, Laahirie Edupuganti |
| hCoV-19/India/MH-RFIP01523/2020 | EPI_ISL_6151339 | 2020-12-26 | Genepath, Pune | RF-IISER Pune | Krishanpal Karmodiya, Aurnab Ghose, Joy Merwin Monteiro, Shatakshi Ranade, Nikhil Phadke |
| hCoV-19/Argentina/PAIS-G0384/2021 | EPI_ISL_6155785 | 2021-09-06 | Laboratorio Central, Ministerio de Salud Cv≥rdoba | [illegible] INTA) on behalf of 'Proyecto Argentino Interinstitucional de genomica de SARS-CoV-2' (PAIS Consortium) | Fernv°ndez, FD; Marquez, N.; Debat, HJ.; Amadio, A; Irazoqui, M; Re, V.; Pisano, M.B.; Castro, G.; Barbas, G. |
| hCoV-19/Botswana/R44B19_BHP_000844008/2021 | EPI_ISL_7121205 | 2021-11-23 | Botswana Harvard HIV Reference Laboratory | Botswana Harvard HIV Reference Laboratory | Sikhulile Moyo, Wonderful T. Choga, Dorcas Maruapula, Keoratile Ntshambiwa, Sefetogi Ramaologa, Thongbotho Mphoyakgosi, Boitumelo Zuze, Botshelo Radibe, Legodile Kooepile, Ontlametse T. Bareng, Pamela Smith-Lawrence, Kgomotso Moruisi, Roger Shapiro, Shahi |
| hCoV-19/USA/TX-CDC-FG-176575/2021 | EPI_ISL_7118375 | 2021-11-16 | Fulgent Genetics | Centers for Disease Control and Prevention Division of Viral Diseases, Pathogen Discovery | Dakota Howard,Dhwani Batra,Peter Cook,Jason Caravas,Benjamin Rambo-Martin,Scott Sammons,Yvette Unoarumhi,Matthew Schmerer,Kristine Lacek,Tymeckia Kendall,Victoria Caban Figueroa,Shatavia Morrison,Christopher Gulvick,Erisa Sula,Harry Gao,Mickey Li,John Gao |
| hCoV-19/Australia/VIC27030/2021 | EPI_ISL_7825574 | 2021-11-14 | Microbiological Diagnostic Unit - Public Health Laboratory (MDU-PHL) | Microbiological Diagnostic Unit - Public Health Laboratory (MDU-PHL) | Seemann, T., Horan, K., Sherry, N.L. |
| hCoV-19/Australia/VIC27113/2021 | EPI_ISL_7825791 | 2021-11-12 | Microbiological Diagnostic Unit - Public Health Laboratory (MDU-PHL) | Microbiological Diagnostic Unit - Public Health Laboratory (MDU-PHL) | Seemann, T., Horan, K., Sherry, N.L. |
| hCoV-19/Argentina/PAIS-D0254/2021 | EPI_ISL_8181671 | 2021-04-05 | Laboratorio del Hospital Regional Ushuaia Gdor. Ernesto Campos | [illegible] Fuego - Hospital Regional Ushuaia - Centro Austral De Investigaciones Cientv≠ficas - Universidad Nacional De Tierra Del Fuego on behalf PAIS project and ANLIS-Malbran | Gramundi, ID; Gallego, F; Ceballos, SG; Nardi, CF; Rojas, AE; De Roccis, CA; Castro, G; Cv°ceres, SB; Yulan, CB; Boutureira, MF. |
| hCoV-19/Brazil/SP-NVBS8095GENOV828238948280/2021 | EPI_ISL_7366547 | 2021-10-08 | DASA | DASA | Rodrigo Guarischi, Paulo Pierry, Adriano Bonaldi, Cristina Oliveira, Bianca Cota, Luciane Sussuchi, Camila Romano, Angelica Hristov, Lidia Yamamoto, Rodrigo Salazar, Annelise Lopes, Jose Levi |
| hCoV-19/Singapore/9664/2021 | EPI_ISL_8435263 | 2021-12-28 | National Public Health Laboratory, National Centre for Infectious Diseases | National Centre for Infectious Diseases | Benny Yeo, Zhenyang Zhou, Royce Ang, Samuel Loo, Lin Cui, Raymond Tzer Pin Lin |
| hCoV-19/Botswana/R54B17_BHP_AAC37778/2021 | EPI_ISL_7834497 | 2021-12-12 | Botswana Harvard HIV Reference Laboratory | Botswana Harvard HIV Reference Laboratory | Sikhulile Moyo, Wonderful T. Choga, Dorcas Maruapula, Keoratile Ntshambiwa, Sefetogi Ramaologa, Thongbotho Mphoyakgosi, Boitumelo Zuze, Botshelo Radibe, Legodile Kooepile, Ontlametse T. Bareng, Pamela Smith-Lawrence, Kgomotso Moruisi, Roger Shapiro, Shahi |

| | | | | | |
|---|---|---|---|---|---|
| hCoV-19/Botswana/R54B81_BHP_362183260/2021 | EPI_ISL_7834549 | 2021-12-09 | Lancet Laboratories Gaborone | Botswana Harvard HIV Reference Laboratory | Sikhulile Moyo, Wonderful T. Choga, Dorcas Maruapula, Ellen Nkitse, Leyi Ncube, Mosana Moyo, Keoratile Ntshambiwa, Sefetogi Ramaologa, Thongbotho Mphoyakgosi, Boitumelo Zuze, Botshelo Radibe, Legodile Kooepile, Ontlametse T. Bareng, Pamela Smith-Lawrence, |
| hCoV-19/USA/PA-CDC-STM-EVNB7TCJY/2021 | EPI_ISL_8181942 | 2021-12-15 | Helix | Centers for Disease Control and Prevention Division of Viral Diseases, Pathogen Discovery | Dakota Howard,Dhwani Batra,Peter Cook,Jason Caravas,Benjamin Rambo-Martin,Scott Sammons,Yvette Unoarumhi,Matthew Schmerer,Kristine Lacek,Tymeckia Kendall,Victoria Caban Figueroa,Helix CA,Shatavia Morrison,Christopher Gulvick,Erisa Sula,Clinton Paden,Dunca |
| hCoV-19/Australia/NSW-ICPMR-18002/2021 | EPI_ISL_7987826 | 2021-12-15 | 4Cyte Pathology | Clinical Pathology and Medical Research; Westmead Hospital; University of Sydney | Arnott A., Draper J., Gall M., Martinez E., Rockett R., Sintchenko V., on behalf of ICPMR |
| hCoV-19/Germany/BY-RKI-I-400677/2021 | EPI_ISL_8229962 | 2021-12-20 | Medizinisch-Diagnostisches Labor Kempten allgV§ulab | Robert Koch Institute | unknown |
| hCoV-19/Ireland/LH-NVRL-S21IRL00559591/2021 | EPI_ISL_7845718 | 2021-12-11 | National Virus Reference Laboratory | National Virus Reference Laboratory | Zoe Yandle, Charlene Bennett, Gabriel Gonzalez, Michael Carr, Jonathan Dean, Cillian F De Gascun |
| hCoV-19/USA/CA-CDC-FG-191878/2021 | EPI_ISL_7848742 | 2021-12-11 | Fulgent Genetics | Centers for Disease Control and Prevention Division of Viral Diseases, Pathogen Discovery | Dakota Howard,Dhwani Batra,Peter Cook,Jason Caravas,Benjamin Rambo-Martin,Scott Sammons,Yvette Unoarumhi,Matthew Schmerer,Kristine Lacek,Tymeckia Kendall,Victoria Caban Figueroa,Shatavia Morrison,Christopher Gulvick,Erisa Sula,Harry Gao,Mickey Li,John Gao |
| hCoV-19/Kenya/SS2750/2021 | EPI_ISL_7850858 | 2021-09-13 | Nanyuki Teaching and Referral Hospital | KEMRI-Wellcome Trust Research Programme,Kilifi | Githinji G.,Ochola I., Agoti C.,Mohamed K.S., de Laurent Z., Mburu M.W., Morobe J., Ndwiga L. Makori T., Ongera E., Lambisia A. Nokes D.J. |
| hCoV-19/India/MH-RFIP04226/2021 | EPI_ISL_7850938 | 2021-10-12 | KEM Hospital, Pune | RF-IISER Pune | Krishanpal Karmodiya, Aurnab Ghose, Joy Merwin Monteiro, Shatakshi Ranade, Nikhil Phadke |
| hCoV-19/India/MH-RFIP04527/2021 | EPI_ISL_7851051 | 2021-05-06 | BJMC,Pune | RF-IISER Pune | Krishanpal Karmodiya, Aurnab Ghose, Joy Merwin Monteiro, Suvarna Joshi, Rajesh Karyakarte |
| hCoV-19/SouthAfrica/NCV261/2021 | EPI_ISL_7852860 | 2021-12-03 | National Health Laboratory Services, Virology | National Health Laboratory Services, Virology | Kathleen Subramoney, Florette Treurnicht, Nkhensani Mtileni, Avani Bharuthram |
| hCoV-19/SouthAfrica/NCV281/2021 | EPI_ISL_7852872 | 2021-12-06 | National Health Laboratory Services, Virology | National Health Laboratory Services, Virology | Kathleen Subramoney, Florette Treurnicht, Nkhensani Mtileni, Avani Bharuthram |
| hCoV-19/Australia/VIC27058/2021 | EPI_ISL_7853557 | 2021-10-20 | Microbiological Diagnostic Unit - Public Health Laboratory (MDU-PHL) | Microbiological Diagnostic Unit - Public Health Laboratory (MDU-PHL) | Seemann, T., Horan, K., Sherry, N.L. |
| hCoV-19/Germany/BY-RKI-I-399554/2021 | EPI_ISL_8228938 | 2021-12-13 | Sonic - Labor Staber Bayreuth | Robert Koch Institute | unknown |
| hCoV-19/England/MILK-2E598EE/2021 | EPI_ISL_7867321 | 2021-12-11 | Lighthouse Lab in Milton Keynes | Wellcome Sanger Institute for the COVID-19 Genomics UK (COG-UK) Consortium | The Lighthouse Lab in Milton Keynes and Alex Alderton, Roberto Amato, Jeffrey Barrett, Sonia Goncalves, Ewan Harrison, David K. Jackson, Ian Johnston, Dominic Kwiatkowski, Cordelia Langford, John Sillitoe on behalf of the Wellcome Sanger Institute COVID-1 |

| | | | | | |
|---|---|---|---|---|---|
| hCoV-19/Spain/IB-HUSE-03278/2021 | EPI_ISL_7867807 | 2021-12-10 | HOSPITAL CAN MISSES | HOSPITAL UNIVERSITARIO SON ESPASES | Dr. Antonio Oliver, Dr. Carla L√≥pez-Causap√©, Dr. Gabriel Cabot, on behalf of Servicio de Microbiolog√≠a, Hospital Universitario Son Espases |
| hCoV-19/England/NORT-YY73FN/2021 | EPI_ISL_7868715 | 2021-11-29 | Northumbria University / South Tees Hospitals NHS Foundation Trust / North Cumbria Integrated Care NHS Foundation Trust / North Tees and Hartlepool NHS Foundation Trust / Newcastle Hospitals NHS Foundation Trust | COVID-19 Genomics UK (COG-UK) Consortium | Darren L Smith,Andrew Nelson,Matthew Bashton,Greg R Young,Joshua Loh,John Allan,Mohammad A Tariq,Giles S Holt,Gary Black,Wen C Yew,Lynn Dover,Paul Baker,Steve Liggett,Sarah Essex,Jane Greenaway,Debra Padgett,Clive Graham,Garren Scott,Edward Barton,Emma Sw |
| hCoV-19/SouthAfrica/NICD-N22263/2021 | EPI_ISL_8182749 | 2021-11-23 | TSHEPONG LABORATORY | National Institute for Communicable Diseases of the National Health Laboratory Service | Amoako DG, Everatt J, Kekana D, Scheepers C, Mohale T, Ntuli N, Mahlangu B, Mnguni A, Ismail A, Bhiman JN, Wolter N |
| hCoV-19/Bangladesh/IEDCR-icddrb-GSB01-146/2021 | EPI_ISL_7870211 | 2021-11-06 | Institute of Epidemiology Disease Control And Research (IEDCR) | Virology Laboratory, International Centre for Diarrhoeal Disease Research, Bangladesh (ICDDR,B) | Mojnu Miah, Rashedul Hasan, Md. Mahfuzur Rahman, Manjur Hossain Khan, Senjuti Saha, Hassan Afrad, Mohammed Ziaur Rahman, Mohammad Jubair, Mohammad Enayet Hossain, Razib Mazumder, Mohammad Shahidul Islam, Ahmed Nawsher Alam, ASM Alamgir, Mahbubur Rahman, M |
| hCoV-19/SouthAfrica/NICD-N23137/2021 | EPI_ISL_8182789 | 2021-12-02 | KIMBERLEY LABORATORY | National Institute for Communicable Diseases of the National Health Laboratory Service | Amoako DG, Everatt J, Kekana D, Scheepers C, Mohale T, Ntuli N, Mahlangu B, Mnguni A, Ismail A, Bhiman JN, Wolter N |
| hCoV-19/SouthAfrica/NICD-N23148/2021 | EPI_ISL_8182799 | 2021-12-02 | KIMBERLEY LABORATORY | National Institute for Communicable Diseases of the National Health Laboratory Service | Amoako DG, Everatt J, Kekana D, Scheepers C, Mohale T, Ntuli N, Mahlangu B, Mnguni A, Ismail A, Bhiman JN, Wolter N |
| hCoV-19/SouthAfrica/NICD-N23210/2021 | EPI_ISL_8182809 | 2021-12-02 | KIMBERLEY LABORATORY | National Institute for Communicable Diseases of the National Health Laboratory Service | Amoako DG, Everatt J, Kekana D, Scheepers C, Mohale T, Ntuli N, Mahlangu B, Mnguni A, Ismail A, Bhiman JN, Wolter N |
| hCoV-19/SouthAfrica/NICD-N22913/2021 | EPI_ISL_8182859 | 2021-11-28 | CHRIS HANI BARAGWANATH LABORATORY | National Institute for Communicable Diseases of the National Health Laboratory Service | Amoako DG, Everatt J, Kekana D, Scheepers C, Mohale T, Ntuli N, Mahlangu B, Mnguni A, Ismail A, Bhiman JN, Wolter N |
| hCoV-19/SouthAfrica/NICD-N22916/2021 | EPI_ISL_8182862 | 2021-11-30 | CHRIS HANI BARAGWANATH LABORATORY | National Institute for Communicable Diseases of the National Health Laboratory Service | Amoako DG, Everatt J, Kekana D, Scheepers C, Mohale T, Ntuli N, Mahlangu B, Mnguni A, Ismail A, Bhiman JN, Wolter N |
| hCoV-19/Israel/ICH-741105961/2021 | EPI_ISL_7989020 | 2021-12-14 | Clinical Microbiology Laboratory, Tel Aviv Sourasky Medical Center | Clinical Microbiology Laboratory, Tel Aviv Sourasky Medical Center | Katya Levytskyi, Ora Halutz, Alon Ziv, Lior Handler, Amos Adler |
| hCoV-19/Canada/ON-PHL-21-44325/2021 | EPI_ISL_7872306 | 2021-12-02 | Public Health Ontario Laboratory | Public Health Ontario Laboratory | Vanessa G Allen, Philip Banh, Yao Chen, Richard de Borja, Alireza Eshaghi, Nahuel Fittipaldi, Christine Frantz, Jonathan B Gubbay, Jennifer L Guthrie, Lawrence Heisler, Esha Joshi, Michael Laszloffy, Aimin Li, Michael CY Li, Dean Maxwell, Sandeep Nagra, S |
| hCoV-19/Canada/ON-PHL-21-44326/2021 | EPI_ISL_7872307 | 2021-12-03 | Public Health Ontario Laboratory | Public Health Ontario Laboratory | Vanessa G Allen, Philip Banh, Yao Chen, Richard de Borja, Alireza Eshaghi, Nahuel Fittipaldi, Christine Frantz, Jonathan B Gubbay, Jennifer L Guthrie, Lawrence Heisler, Esha Joshi, Michael Laszloffy, Aimin Li, Michael CY Li, Dean Maxwell, Sandeep Nagra, S |

| | | | | | |
|---|---|---|---|---|---|
| hCoV-19/Canada/ON-PHL-21-44327/2021 | EPI_ISL_7872308 | 2021-12-03 | Public Health Ontario Laboratory | Public Health Ontario Laboratory | Vanessa G Allen, Philip Banh, Yao Chen, Richard de Borja, Alireza Eshaghi, Nahuel Fittipaldi, Christine Frantz, Jonathan B Gubbay, Jennifer L Guthrie, Lawrence Heisler, Esha Joshi, Michael Laszloffy, Aimin Li, Michael CY Li, Dean Maxwell, Sandeep Nagra, S |
| hCoV-19/Israel/ICH-741105927/2021 | EPI_ISL_7989062 | 2021-12-15 | Clinical Microbiology Laboratory, Tel Aviv Sourasky Medical Center | Clinical Microbiology Laboratory, Tel Aviv Sourasky Medical Center | Katya Levytskyi, Ora Halutz, Alon Ziv, Lior Handler, Amos Adler |
| hCoV-19/India/OR-ILSGS08185/2021 | EPI_ISL_7873897 | 2021-07-22 | Srirama Chandra Bhanja Medical College and Hospital, Cuttack | Institute of Life Sciences-INSACOG | Sunil K. Raghav, Safal Walia, Arup Ghosh, Atimukta Jha, Shifu Aggarwal, Amiyaranjan Das, Sana Fatma, Punyatoya Panda, Rupesh Dash, Rajeeb Swain, Punit Prasad, INSACOG Consortium, Ajay Parida |
| hCoV-19/Turkey/HSGM-F7590/2021 | EPI_ISL_7803109 | 2021-12-03 | Ministry of Health Turkey | Ministry of Health Turkey | Fatma Bayrakdar, Yasemin Co≈ügun, S√ºleyman Yalcin, G√ºlay Korukluoƒülu |
| hCoV-19/Brazil/SP-IB_SEQ21006/2021 | EPI_ISL_8149414 | 2021-12-14 | RIBEIRAO PRETO - UPA NORTE ,Äì DR NELSON MANDELA | Instituto Butantan | Dimas Tadeu Covas, Antonio Jorge Martins, Claudia Renata dos Santos Barros, David Schlesinger, Debora Botequio Moretti, Elaine Cristina Marqueze, Elaine Vieira Santos, Evandra Strazza Rodrigues, Heidge Fukumasu, Jayme Augusto de Souza-Neto, Jos√© Salvator |
| hCoV-19/India/OR-ILSGS08881/2021 | EPI_ISL_7877566 | 2021-06-27 | Angul, DHH | Institute of Life Sciences-INSACOG | Sunil K. Raghav, Safal Walia, Arup Ghosh, Atimukta Jha, Shifu Aggarwal, Amiyaranjan Das, Sana Fatma, Punyatoya Panda, Rupesh Dash, Rajeeb Swain, Punit Prasad, INSACOG Consortium, Ajay Parida |
| hCoV-19/Ecuador/USFQ-2330/2021 | EPI_ISL_8183455 | 2021-11-19 | Institute of Microbiology, Universidad San Francisco de Quito | Institute of Microbiology, Universidad San Francisco de Quito | Mateo Carvajal, Monica Becerra-Wong, Juan Jos√© Guadalupe, Sully M√°rquez, Bel√©n Prado-Vivar, Erika B. Mu√±oz, Fernanda Zurita, Bernardo Guti√©rrez, Andrea Pinos, Ver√≥nica Barrag√°n, Patricio Rojas-Silva, Gabriel Trueba, Michelle Grunauer, Pa√∫l C√°rden |
| hCoV-19/Ecuador/USFQ-2461/2021 | EPI_ISL_8183501 | 2021-12-17 | Institute of Microbiology, Universidad San Francisco de Quito | Institute of Microbiology, Universidad San Francisco de Quito | Bel√©n Prado-Vivar, Fernanda Zurita, Monica Becerra-Wong, Erika B. Mu√±oz, Mateo Carvajal, Juan Jos√© Guadalupe, Sully M√°rquez, Bernardo Guti√©rrez, Ver√≥nica Pacheco, Ver√≥nica Barrag√°n, Patricio Rojas-Silva, Gabriel Trueba, Michelle Grunauer, Pa√∫l C√ |
| hCoV-19/Canada/QC-1nDJXW-506876481/2021 | EPI_ISL_8184273 | 2021-11-19 | Laboratoire de sant√© publique du Qu√©bec | Laboratoire de sant√© publique du Qu√©bec | Sandrine Moreira, Ioannis Ragoussis, Guillaume Bourque, Jesse Shapiro, Mark Lathrop and Judith Fafard on behalf of the CoVSeQ research group |
| hCoV-19/Brazil/SP-IB_142957/2021 | EPI_ISL_7808759 | 2021-12-02 | CENTRO DE SAUDE CAMPOS ELISEOS TANCREDO NEVES | Instituto Butantan | Dimas Tadeu Covas, Antonio Jorge Martins, Claudia Renata dos Santos Barros, David Schlesinger, Debora Botequio Moretti, Elaine Cristina Marqueze, Elaine Vieira Santos, Evandra Strazza Rodrigues, Heidge Fukumasu, Jayme Augusto de Souza-Neto, Jos√© Salvator |
| hCoV-19/USA/MA-UMASSMED-202103859/2021 | EPI_ISL_8150861 | 2021-09-28 | UMass Memorial Medical Center | Center for Microbiome Research | Richard T. Ellison III, Karl J. Simon, Timothy F. Kowalik, Doyle V. Ward |
| hCoV-19/Sweden/192717922V/2021 | EPI_ISL_8063334 | 2021-11-29 | Vastragotaland, Unilabs | Unilabs, Eskilstuna | Emma Arvidsson |
| hCoV-19/Sweden/200618102V/2021 | EPI_ISL_8063583 | 2021-12-16 | Vastragotaland, Unilabs | Unilabs, Eskilstuna | Emma Arvidsson |

| | | | | | |
|---|---|---|---|---|---|
| hCoV-19/USA/MD-IGS-012133201018A/2021 | EPI_ISL_8151903 | 2021-11-28 | Maryland Genomics, Institute for Genome Sciences, University of Maryland School of Medicine | Maryland Genomics, Institute for Genome Sciences, University of Maryland School of Medicine | Tallon, Luke J; Sadzewicz, Lisa D; Humphrys, Mike; Ott, Sandra; Roussey, Holly; Vavikolanu, Kranthi; Regan, Kevin; George, Jain; Hazen, Tracy; Fraser, Claire M; Ravel, Jacques |
| hCoV-19/Spain/CT-HUGTiPM064WB3C9/2021 | EPI_ISL_8150054 | 2021-12-17 | Microbiology Department, Laboratori Cl√≠nic Metropolitana Nord. Hospital Universitari Germans Trias i Pujol | Can Ruti SARS-CoV-2 Sequencing Hub (HUGTiP/IrsiCaixa/IGTP) | Marc Noguera-Julian, Pilar Armengol, Ignacio Blanco, Ana Blanco, Antoni E Bordoy, Francesc Catala-Moll, Pere-Joan Cardona, Gemma Clara, Bonaventura Clotet, Montserrat Gim√©nez, Mercedes Guerrero, Anna Not, David Panisello, Roger Paredes, Alexia Par√≠s, Ve |
| hCoV-19/PuertoRico/PR-UPRRP-3592/2021 | EPI_ISL_7705562 | 2021-07-21 | Laboratorio Cinico Toledo | Sequencing and Genomics facility (SGF) | Riccardo Papa, Steven Van Belleghem, Silvia Planas, Yadira Ortiz, Camille Zenon, Limari Cora-Huertas, Israel Candelaria Velez, Marcia Cruz, Jose Rodriguez Orengo, Filipa Godoy, Jose Carlos Velez, Carlos Sariol |
| hCoV-19/Argentina/PAIS-D0301/2021 | EPI_ISL_8152086 | 2021-03-04 | Laboratorio del Hospital Regional Ushuaia Gdor. Ernesto Campos | Nodo de Secuenciaci√≥n Tierra del Fuego - Hospital Regional Ushuaia - Centro Austral De Investigaciones Cient√≠ficas - Universidad Nacional De Tierra Del Fuego on behalf Proyecto Argentino Interinstitucional de genomica de SARS-CoV-2' (PAIS Consortium) an | Gramundi, ID; Gallego, F; Ceballos, SG; Nardi, CF; Rojas, AE; De Roccis, CA; Castro, G; C√°ceres, SB; Yulan, CB; Boutureira, MF. |
| hCoV-19/Brazil/PR-FIOCRUZ-63865/2021 | EPI_ISL_8152428 | 2021-11-03 | Laboratorio Central de Saude Publica do Estado do Parana (LACEN/PR) | Laboratory of Respiratory Viruses and Measles, Oswaldo Cruz Institute, FIOCRUZ | Paola Resende, Luciana Appolinario, Fernando Motta, Elisa Cavalcante Pereira, Ighor Arantes, Alice Sampaio Rocha, Bruna Mendon√ßa da Silva, J√©ssica Gra√ßa Macedo de Carvalho, Larissa Macedo Pinto, Victor Guimaraes, Irina Riediger, Marilda Siqueira on beh |
| hCoV-19/Brazil/TO-FIOCRUZ-63560/2021 | EPI_ISL_8152486 | 2021-10-28 | Laboratorio Central de Saude Publica do Estado do Tocantins (LACEN/TO) | Laboratory of Respiratory Viruses and Measles, Oswaldo Cruz Institute, FIOCRUZ | Paola Resende, Luciana Appolinario, Fernando Motta, Elisa Cavalcante Pereira, Ighor Arantes, Alice Sampaio Rocha, Bruna Mendon√ßa da Silva, J√©ssica Gra√ßa Macedo de Carvalho, Larissa Macedo Pinto, Victor Guimaraes, Jucimaria Dantas Galvao, Marilda Siquei |
| hCoV-19/England/MILK-2E40783/2021 | EPI_ISL_7816757 | 2021-12-10 | Lighthouse Lab in Milton Keynes | Wellcome Sanger Institute for the COVID-19 Genomics UK (COG-UK) Consortium | The Lighthouse Lab in Milton Keynes and Alex Alderton, Roberto Amato, Jeffrey Barrett, Sonia Goncalves, Ewan Harrison, David K. Jackson, Ian Johnston, Dominic Kwiatkowski, Cordelia Langford, John Sillitoe on behalf of the Wellcome Sanger Institute COVID-1 |
| hCoV-19/England/MILK-2E257A2/2021 | EPI_ISL_7818612 | 2021-12-10 | Lighthouse Lab in Milton Keynes | Wellcome Sanger Institute for the COVID-19 Genomics UK (COG-UK) Consortium | The Lighthouse Lab in Milton Keynes and Alex Alderton, Roberto Amato, Jeffrey Barrett, Sonia Goncalves, Ewan Harrison, David K. Jackson, Ian Johnston, Dominic Kwiatkowski, Cordelia Langford, John Sillitoe on behalf of the Wellcome Sanger Institute COVID-1 |
| hCoV-19/USA/CO-CDPHE-2102373788/2021 | EPI_ISL_8064434 | 2021-12-03 | Colorado Department of Public Health and Environment | Colorado Department of Public Health and Environment | Laura Bankers, Molly C. Hetherington-Rauth, Diana Ir, Alexandria Rossheim, Michael Martin, Mandy Waters, Arianna Smith, Shannon R. Matzinger, Emily A. Travanty |
| hCoV-19/Australia/QLD4423/2021 | EPI_ISL_8437710 | 2021-12-24 | PHV-FSS | PHV-FSS | Chenwei Wang on behalf of Q-PHIRE Genomics |

| | | | | | |
|---|---|---|---|---|---|
| hCoV-19/Australia/QLD4724/2021 | EPI_ISL_8437738 | 2021-12-23 | PHV-FSS | PHV-FSS | Chenwei Wang on behalf of Q-PHIRE Genomics |
| hCoV-19/Australia/QLD4896/2021 | EPI_ISL_8437673 | 2021-12-24 | PHV-FSS | PHV-FSS | Chenwei Wang on behalf of Q-PHIRE Genomics |
| hCoV-19/England/MILK-2E180E3/2021 | EPI_ISL_7820103 | 2021-12-10 | Lighthouse Lab in Milton Keynes | Wellcome Sanger Institute for the COVID-19 Genomics UK (COG-UK) Consortium | The Lighthouse Lab in Milton Keynes and Alex Alderton, Roberto Amato, Jeffrey Barrett, Sonia Goncalves, Ewan Harrison, David K. Jackson, Ian Johnston, Dominic Kwiatkowski, Cordelia Langford, John Sillitoe on behalf of the Wellcome Sanger Institute COVID-1 |
| hCoV-19/England/MILK-2E18283/2021 | EPI_ISL_7820070 | 2021-12-10 | Lighthouse Lab in Milton Keynes | Wellcome Sanger Institute for the COVID-19 Genomics UK (COG-UK) Consortium | The Lighthouse Lab in Milton Keynes and Alex Alderton, Roberto Amato, Jeffrey Barrett, Sonia Goncalves, Ewan Harrison, David K. Jackson, Ian Johnston, Dominic Kwiatkowski, Cordelia Langford, John Sillitoe on behalf of the Wellcome Sanger Institute COVID-1 |
| hCoV-19/Australia/QLD4941/2021 | EPI_ISL_8438365 | 2021-12-25 | PHV-FSS | PHV-FSS | Chenwei Wang on behalf of Q-PHIRE Genomics |
| hCoV-19/Sweden/UUH-000200238157/2021 | EPI_ISL_7434645 | 2021-11-18 | Akademiska Sjukhuset, Clinical Microbiology and Hospital Hygiene | Uppsala University Hospital | Rene Kaden, Steinar Mannsverk, Julia Bergholm, Patrik Ellstr√∂m, Jonathan Haars |
| hCoV-19/USA/MO-CDC-LC0434859/2021 | EPI_ISL_8153681 | 2021-12-06 | Laboratory Corporation of America | Centers for Disease Control and Prevention Division of Viral Diseases, Pathogen Discovery | Dakota Howard,Dhwani Batra,Peter Cook,Jason Caravas,Benjamin Rambo-Martin,Scott Sammons,Yvette Unoarumhi,Matthew Schmerer,Kristine Lacek,Tymeckia Kendall,Victoria Caban Figueroa,Shatavia Morrison,Christopher Gulvick,Minoo Agarwal,Eyad Almasri,Debbie Boles |
| hCoV-19/Canada/MB-NML-260515/2021 | EPI_ISL_7413422 | 2021-11-30 | MB-Cadham Provincial laboratory | National Microbiology Laboratory (NML) | Anna Majer, Shari Tyson, Grace Seo, Philip Mabon, Elsie Grudeski, Rhiannon Huzarewich, Russell Mandes, Anneliese Landgraff, Jennifer Tanner, Natalie Knox, Morag Graham, Gary Van Domselaar, Paul Van Caeseele, Jared Bullard, David Alexander, Kerry Dust, Nat |
| hCoV-19/USA/MD-IGS-52132503862A/2021 | EPI_ISL_7404794 | 2021-11-21 | Maryland Genomics, Institute for Genome Sciences, University of Maryland School of Medicine | Maryland Genomics, Institute for Genome Sciences, University of Maryland School of Medicine | Tallon, Luke J; Sadzewicz, Lisa D; Humphrys, Mike; Ott, Sandra; Roussey, Holly; Vavikolanu, Kranthi; Regan, Kevin; George, Jain; Hazen, Tracy; Fraser, Claire M; Ravel, Jacques |
| hCoV-19/SouthAfrica/NICD-N21765/2021 | EPI_ISL_7337511 | 2021-11-17 | Charlotte Maxeke Johannesburg Academic Hospital | Diseases of the National Health Laboratory Service | Amoako DG, Everatt J, Scheepers C, Mohale T, Ntuli N, Mahlangu B, Mnguni A, Ismail A, Bhiman JN |
| hCoV-19/SouthAfrica/NICD-N21826/2021 | EPI_ISL_7337561 | 2021-11-24 | Chris Hani Baragwanath Laboratory | Diseases of the National Health Laboratory Service | Amoako DG, Everatt J, Scheepers C, Mohale T, Ntuli N, Mahlangu B, Mnguni A, Ismail A, Bhiman JN |
| hCoV-19/Canada/ON-NML-255882/2021 | EPI_ISL_7265456 | 2021-11-26 | Alpha Labs | National Microbiology Laboratory (NML) | Anna Majer, Shari Tyson, Grace Seo, Philip Mabon, Elsie Grudeski, Rhiannon Huzarewich, Russell Mandes, Anneliese Landgraff, Jennifer Tanner, Natalie Knox, Morag Graham, Gary Van Domselaar, Gordon Jolly, Nathalie Bastien, Yan Li, Timothy Booth, Darian Hole |

| | | | | | |
|---|---|---|---|---|---|
| hCoV-19/England/MILK-2C4356A/2021 | EPI_ISL_7185784 | 2021-11-28 | Lighthouse Lab in Milton Keynes | Wellcome Sanger Institute for the COVID-19 Genomics UK (COG-UK) Consortium | The Lighthouse Lab in Milton Keynes and Alex Alderton, Roberto Amato, Jeffrey Barrett, Sonia Goncalves, Ewan Harrison, David K. Jackson, Ian Johnston, Dominic Kwiatkowski, Cordelia Langford, John Sillitoe on behalf of the Wellcome Sanger Institute COVID-1 |
| hCoV-19/Australia/QLD5052/2021 | EPI_ISL_8438299 | 2021-12-25 | PHV-FSS | PHV-FSS | Chenwei Wang on behalf of Q-PHIRE Genomics |
| hCoV-19/Denmark/DCGC-245674/2021 | EPI_ISL_7272249 | 2021-11-22 | Department of Bacteria, Parasites and Fungi, Statens Serum Institut, Copenhagen, Denmark | Statens Serum Institut Bioinformatics and Microbial Genomics | Danish Covid-19 Genome Consortium |
| hCoV-19/England/ALDP-2C9F3ED/2021 | EPI_ISL_7294185 | 2021-11-27 | Lighthouse Lab in Alderley Park | Wellcome Sanger Institute for the COVID-19 Genomics UK (COG-UK) Consortium | Jacquelyn Wynn, Mairead Hyland, The Lighthouse Lab in Alderley Park and Alex Alderton, Roberto Amato, Jeffrey Barrett, Sonia Goncalves, Ewan Harrison, David K. Jackson, Ian Johnston, Dominic Kwiatkowski, Cordelia Langford, John Sillitoe on behalf of the W |
| hCoV-19/Canada/ON-NML-256530/2021 | EPI_ISL_7264139 | 2021-11-27 | Alpha Labs | National Microbiology Laboratory (NML) | Anna Majer, Shari Tyson, Grace Seo, Philip Mabon, Elsie Grudeski, Rhiannon Huzarewich, Russell Mandes, Anneliese Landgraff, Jennifer Tanner, Natalie Knox, Morag Graham, Gary Van Domselaar, Gordon Jolly, Nathalie Bastien, Yan Li, Timothy Booth, Darian Hole |
| hCoV-19/England/MILK-2C67481/2021 | EPI_ISL_7203016 | 2021-11-29 | Lighthouse Lab in Milton Keynes | Wellcome Sanger Institute for the COVID-19 Genomics UK (COG-UK) Consortium | The Lighthouse Lab in Milton Keynes and Alex Alderton, Roberto Amato, Jeffrey Barrett, Sonia Goncalves, Ewan Harrison, David K. Jackson, Ian Johnston, Dominic Kwiatkowski, Cordelia Langford, John Sillitoe on behalf of the Wellcome Sanger Institute COVID-1 |
| hCoV-19/Scotland/QEUH-2C30BE6/2021 | EPI_ISL_7158402 | 2021-11-27 | Lighthouse Lab in Glasgow | Wellcome Sanger Institute for the COVID-19 Genomics UK (COG-UK) Consortium | Harper VanSteenhouse, Yumi Kasai, David Gray, Carol Clugston, Anna Dominiczak and Alex Alderton, Roberto Amato, Jeffrey Barrett, Sonia Goncalves, Ewan Harrison, David K. Jackson, Ian Johnston, Dominic Kwiatkowski, Cordelia Langford, John Sillitoe on behal |
| hCoV-19/Brazil/DF-IAL-6820/2021 | EPI_ISL_7224971 | 2021-11-25 | LACEN do Distrito Federal | Instituto Adolfo Lutz Strategic Laboratory | Claudio Tavares Sacchi, Karoline Rodrigues Campos, Marlon Benedito Nascimento Santos |
| hCoV-19/England/PHEC-3U088UFE/2021 | EPI_ISL_7463979 | 2021-11-25 | Respiratory Virus Unit, Microbiology Services Colindale, Public Health England | COVID-19 Genomics UK (COG-UK) Consortium | PHE Covid Sequencing Team |
| hCoV-19/SouthAfrica/Tygerberg_3265/2021 | EPI_ISL_7452754 | 2021-11-29 | PathCare, Cape Town | Health Laboratory Service (NHLS), Tygerberg Hospital / Stellenbosch University | Jean Maritz, Nadine Cronje, Petra Raimond, Tongai Maponga, Shannon Wilson, Kamela Mahlakwane, Tania Stander, Gert van Zyl, Wolfgang Preiser |
| hCoV-19/SouthAfrica/Tygerberg_3206/2021 | EPI_ISL_7456511 | 2021-12-01 | PathCare, Cape Town | Health Laboratory Service (NHLS), Tygerberg Hospital / Stellenbosch University | Jean Maritz, Nadine Cronje, Petra Raimond, Tongai Maponga, Shannon Wilson, Kamela Mahlakwane, Tania Stander, Gert van Zyl, Wolfgang Preiser |
| hCoV-19/England/PHEC-3U087U4C/2021 | EPI_ISL_7463952 | 2021-11-24 | Respiratory Virus Unit, Microbiology Services Colindale, Public Health England | COVID-19 Genomics UK (COG-UK) Consortium | PHE Covid Sequencing Team |
| hCoV-19/Ireland/MH-NVRL-S21IRL00519096/2021 | EPI_ISL_7406048 | 2021-11-26 | National Virus Reference Laboratory | National Virus Reference Laboratory | Zoe Yandle, Charlene Bennett, Gabriel Gonzalez, Michael Carr, Jonathan Dean, Cillian F De Gascun |

| | | | | | |
|---|---|---|---|---|---|
| hCoV-19/England/MILK-2CAFFE6/2021 | EPI_ISL_7353981 | 2021-11-29 | Lighthouse Lab in Milton Keynes | Wellcome Sanger Institute for the COVID-19 Genomics UK (COG-UK) Consortium | The Lighthouse Lab in Milton Keynes and Alex Alderton, Roberto Amato, Jeffrey Barrett, Sonia Goncalves, Ewan Harrison, David K. Jackson, Ian Johnston, Dominic Kwiatkowski, Cordelia Langford, John Sillitoe on behalf of the Wellcome Sanger Institute COVID-1 |
| hCoV-19/Scotland/QEUH-2C7DDE7/2021 | EPI_ISL_7302122 | 2021-11-29 | Lighthouse Lab in Glasgow | Wellcome Sanger Institute for the COVID-19 Genomics UK (COG-UK) Consortium | Harper VanSteenhouse, Yumi Kasai, David Gray, Carol Clugston, Anna Dominiczak and Alex Alderton, Roberto Amato, Jeffrey Barrett, Sonia Goncalves, Ewan Harrison, David K. Jackson, Ian Johnston, Dominic Kwiatkowski, Cordelia Langford, John Sillitoe on behal |
| hCoV-19/India/DL-NCDC-6409265/2021 | EPI_ISL_7285023 | 2021-11-23 | National Centre for Disease Control (NCDC) Biotechnology Division, Delhi | NCDC Delhi, Biotechnology Division INSACOG | Robin Marwal, Mahesh S Dhar, Kalaiarasan Ponnusamy, Meena Datta, Priyanka Singh,¬†Uma Sharma, Hemlata Lall, Manoj K Singh,¬†Radhakrishnan V. S, Hema Gogia, Preeti Madan, Sandhya Kabra, Sujeet K Singh, Partha Rakshit |
| hCoV-19/USA/NY-Wadsworth-21103394-01/2021 | EPI_ISL_7334889 | 2021-11-30 | SUNRISE MEDICAL LABORATORIES | Wadsworth Center, New York State Department of Health | Kirsten St. George, Daryl M. Lamson, Alexis Russell, Matthew Shudt, Melissa A Leisner, Jonathan Plitnick, Catharine Prussing, Navjot Singh, John Kelly, Erasmus Schneider, Erica Lasek-Nesselquist |
| hCoV-19/England/MILK-2C8CD30/2021 | EPI_ISL_7300159 | 2021-11-30 | Lighthouse Lab in Milton Keynes | Wellcome Sanger Institute for the COVID-19 Genomics UK (COG-UK) Consortium | The Lighthouse Lab in Milton Keynes and Alex Alderton, Roberto Amato, Jeffrey Barrett, Sonia Goncalves, Ewan Harrison, David K. Jackson, Ian Johnston, Dominic Kwiatkowski, Cordelia Langford, John Sillitoe on behalf of the Wellcome Sanger Institute COVID-1 |
| hCoV-19/Israel/ICH-760596848/2021 | EPI_ISL_7265979 | 2021-11-30 | Clinical Microbiology Laboratory, Tel Aviv Sourasky Medical Center | Clinical Microbiology Laboratory, Tel Aviv Sourasky Medical Center | Katya Levytskyi, Ora Halutz, Alon Ziv, Lior Handler, Amos Adler |
| hCoV-19/SouthAfrica/NICD-N21740/2021 | EPI_ISL_7337483 | 2021-11-16 | Charlotte Maxeke Johannesburg Academic Hospital | Diseases of the National Health Laboratory Service | Amoako DG, Everatt J, Scheepers C, Mohale T, Ntuli N, Mahlangu B, Mnguni A, Ismail A, Bhiman JN |
| hCoV-19/SouthAfrica/NICD-N21759/2021 | EPI_ISL_7337505 | 2021-11-22 | Charlotte Maxeke Johannesburg Academic Hospital | Diseases of the National Health Laboratory Service | Amoako DG, Everatt J, Scheepers C, Mohale T, Ntuli N, Mahlangu B, Mnguni A, Ismail A, Bhiman JN |
| hCoV-19/England/MILK-2C90D93/2021 | EPI_ISL_7304476 | 2021-11-30 | Lighthouse Lab in Milton Keynes | Wellcome Sanger Institute for the COVID-19 Genomics UK (COG-UK) Consortium | The Lighthouse Lab in Milton Keynes and Alex Alderton, Roberto Amato, Jeffrey Barrett, Sonia Goncalves, Ewan Harrison, David K. Jackson, Ian Johnston, Dominic Kwiatkowski, Cordelia Langford, John Sillitoe on behalf of the Wellcome Sanger Institute COVID-1 |
| hCoV-19/HongKong/HK-HKPU-12388/2021 | EPI_ISL_7192723 | 2021-11-15 | Department of Health Technology and Informatics, The Hong Kong Polytechnic University | Department of Health Technology and Informatics, The Hong Kong Polytechnic University | Gilman Kit-Hang Siu, Lam-Kwong Lee, Kenneth Siu-Sing Leung, Jake Siu-Lun Leung, Wing-Hei Lo, Timothy Ting-Leung Ng, Chloe Toi-Mei Chan, Hoi-Ching Jim, Ivan Tak-Fai Wong, Kingsley King-Gee Tam, Hiu-Yin Lao, Denise Sze-Hang Wong, Alan Ka-Lun Wu, Miranda Cho |
| hCoV-19/Sweden/01_SE100_21CS106609/2021 | EPI_ISL_7219990 | 2021-11-27 | Karolinska University Hospital Huddinge | Karolinska University Hospital Huddinge | Jan Albert, Tobias Allander, Sandra Broddesson, Robert Dyrdak, Martin Ekman, Lynda Eneh, Shambhu Ganeshappa Aralaguppe, Natalija Gerasimcik, Karina Hentrich, Tanja Normark, Henning Onsbring, Annika Tiveljung Lindell, Zhibing Yun, Valtteri Wirta |

| | | | | | |
|---|---|---|---|---|---|
| hCoV-19/USA/MD-HP20874-PIDUYWZOWA/2021 | EPI_ISL_7160424 | 2021-11-27 | Johns Hopkins Hospital Department of Pathology | Johns Hopkins Hospital Department of Pathology | C. Paul Morris, David Gaston, Julie M. Norton, Amary Fall, Raghda Eldesouki, Michael Forman, Matthew Schwartz, Heba H. Mostafa |
| hCoV-19/England/BRBR-2C7ECBC/2021 | EPI_ISL_7197950 | 2021-11-27 | Berkshire and Surrey Pathology Services Lighthouse Laboratory | Wellcome Sanger Institute for the COVID-19 Genomics UK (COG-UK) Consortium | Berkshire and Surrey Pathology Services Lighthouse Laboratory and Alex Alderton, Roberto Amato, Jeffrey Barrett, Sonia Goncalves, Ewan Harrison, David K. Jackson, Ian Johnston, Dominic Kwiatkowski, Cordelia Langford, John Sillitoe on behalf of the Wellcom |
| hCoV-19/Norway/29741/2021 | EPI_ISL_7248772 | 2021-11-29 | Furst Medical Laboratory | Norwegian Institute of Public Health, Department of Virology | Kathrine Stene-Johansen, Kamilla Heddeland Instefjord, Hilde Elshaug, Garcia Llorente Ignacio, Jon BrV•te, Engebretsen Serina Beate, Pedersen Benedikte Nevjen, Line Victoria Moen, Debech Nadia, Atiya R Ali, Marie Paulsen Madsen, Rasmus Riis Kopperud, Hild |
| hCoV-19/Nepal/NPHL-S-263/2021 | EPI_ISL_7196120 | 2021-11-26 | Bidh Lab | National Public Health Laboratory | National Public Health Laboratory Team, Bidh Laboartory |
| hCoV-19/USA/WA-UW-21112997440/2021 | EPI_ISL_7160037 | 2021-11-29 | UW Virology Lab | UW Virology Lab | Pavitra Roychoudhury, Shah Mohamed Bakhash, Ricardo Perez, Pooneh Hajian, Hong Xie, Tien V. Nguyen, Sean Ellis, Saraswathi Sathees, Meei-Li Huang, Keith R Jerome, Alexander Greninger |
| hCoV-19/England/MILK-2C27A9A/2021 | EPI_ISL_7146517 | 2021-11-26 | Lighthouse Lab in Milton Keynes | Wellcome Sanger Institute for the COVID-19 Genomics UK (COG-UK) Consortium | The Lighthouse Lab in Milton Keynes and Alex Alderton, Roberto Amato, Jeffrey Barrett, Sonia Goncalves, Ewan Harrison, David K. Jackson, Ian Johnston, Dominic Kwiatkowski, Cordelia Langford, John Sillitoe on behalf of the Wellcome Sanger Institute COVID-1 |
| hCoV-19/Israel/SMC-7025961/2021 | EPI_ISL_7594083 | 2021-12-06 | Shamir Medical Center (Asaf Harofe) | Shamir Medical Center (Asaf Harofe) | Nir Rainy,Chen Weiner,Reut Sorek Abramovich,Yevgeni Yegorov,Anna Vishnevsky,Patricia Benveniste-Lekovitz,Abu Hamad Ramzia,Adina Bar Chaim |
| hCoV-19/Israel/SMC-7026072/2021 | EPI_ISL_7594030 | 2021-12-07 | Shamir Medical Center (Asaf Harofe) | Shamir Medical Center (Asaf Harofe) | Nir Rainy,Chen Weiner,Reut Sorek Abramovich,Yevgeni Yegorov,Anna Vishnevsky,Patricia Benveniste-Lekovitz,Abu Hamad Ramzia,Adina Bar Chaim |
| hCoV-19/Australia/QLD14191/2021 | EPI_ISL_8438461 | 2021-12-28 | PHV-FSS | PHV-FSS | Chenwei Wang on behalf of Q-PHIRE Genomics |
| hCoV-19/Australia/QLD7980/2021 | EPI_ISL_8437900 | 2021-12-28 | PHV-FSS | PHV-FSS | Chenwei Wang on behalf of Q-PHIRE Genomics |
| hCoV-19/USA/NJ-PHEL-V21031373-confirm/2021 | EPI_ISL_7605516 | 2021-11-29 | NJDOH, Public Health and Environmental Laboratories | NJ_PHEL | Lindsey Bodnar, Ryan Pachucki, Shiv K. Verma, Allison Roder, Maria-Magdalene Pugliese, Chelsea San Filippo, Jacquelyn Deverell, Mohammad M. Ali, Dana Woell, Byeong Jeong |
| hCoV-19/SouthAfrica/NICD-N22371/2021 | EPI_ISL_7605740 | 2021-11-26 | RUSTENBURG LABORATORY | Diseases of the National Health Laboratory Service | Amoako DG, Everatt J, Scheepers C, Mohale T, Ntuli N, Mahlangu B, Mnguni A, Ismail A, Bhiman JN, Wolter N |
| hCoV-19/SouthAfrica/NICD-N22253/2021 | EPI_ISL_7605746 | 2021-11-27 | TSHEPONG LABORATORY | Diseases of the National Health Laboratory Service | Amoako DG, Everatt J, Scheepers C, Mohale T, Ntuli N, Mahlangu B, Mnguni A, Ismail A, Bhiman JN, Wolter N |
| hCoV-19/USA/UT-UPHL-211118796588/2021 | EPI_ISL_8438730 | 2021-09-15 | Utah Public Health Laboratory | Utah Public Health Laboratory | Erin L. Young, Kelly F. Oakeson, Pooja Gupta, John Arnn, Tom Iverson, Olinto Linares-Perdomo |

| | | | | | |
|---|---|---|---|---|---|
| hCoV-19/Israel/SMC-7025431/2021 | EPI_ISL_7613573 | 2021-11-30 | Shamir Medical Center (Asaf Harofe) | Shamir Medical Center (Asaf Harofe) | Nir Rainy,Chen Weiner,Reut Sorek Abramovich,Yevgeni Yegorov,Anna Vishnevsky,Patricia Benveniste-Lekovitz,Abu Hamad Ramzia,Adina Bar Chaim |
| hCoV-19/Nigeria/NCDC-NR1463/2021 | EPI_ISL_8064550 | 2021-12-03 | EVERIGHT | National Reference Laboratory, Nigeria Centre for Disease Control | Dr Ndodo Nnaemeka, Olusola Anuoluwapo Akanbi, Chimaobi Chukwu, Oyeronke Ayansola, Dr Omoare Adesuyi, Anthony Ahumibe, Catherine Okoi, Nwando Mba, Dr Ifedayo Adetifa |
| hCoV-19/Nigeria/NCDC-NR1153/2021 | EPI_ISL_8064536 | 2021-11-26 | E-clinic & Diagnostics | National Reference Laboratory, Nigeria Centre for Disease Control | Dr Ndodo Nnaemeka, Olusola Anuoluwapo Akanbi, Chimaobi Chukwu, Oyeronke Ayansola, Dr Omoare Adesuyi, Anthony Ahumibe, Catherine Okoi, Nwando Mba, Dr Ifedayo Adetifa |
| hCoV-19/Nigeria/NCDC-NR1517/2021 | EPI_ISL_8064564 | 2021-12-07 | DNA LABS | National Reference Laboratory, Nigeria Centre for Disease Control | Dr Ndodo Nnaemeka, Olusola Anuoluwapo Akanbi, Chimaobi Chukwu, Oyeronke Ayansola, Dr Omoare Adesuyi, Anthony Ahumibe, Catherine Okoi, Nwando Mba, Dr Ifedayo Adetifa |
| hCoV-19/India/MH-IISER-Pune-IP-00991/2021 | EPI_ISL_7824855 | 2021-02-09 | Department of Microbiology, Government Medical College, Baramati | IISER Pune-INSACOG | Krishanpal Karmodiya, Aurnab Ghose, Joy Merwin Monteiro |
| hCoV-19/USA/NJ-CDC-LC0433030/2021 | EPI_ISL_8156641 | 2021-12-17 | Laboratory Corporation of America | Centers for Disease Control and Prevention Division of Viral Diseases, Pathogen Discovery | Dakota Howard,Dhwani Batra,Peter Cook,Jason Caravas,Benjamin Rambo-Martin,Scott Sammons,Yvette Unoarumhi,Matthew Schmerer,Kristine Lacek,Tymeckia Kendall,Victoria Caban Figueroa,Shatavia Morrison,Christopher Gulvick,Minoo Agarwal,Eyad Almasri,Debbie Boles |
| hCoV-19/USA/WI-MHDL-2021120268/2021 | EPI_ISL_7828340 | 2021-11-26 | NOAH Clinical Laboratory | City of Milwaukee Health Department Laboratory | Manjeet Khubbar, Amy Bauer, Samantha Scott, Joshua Weiner, Nandhakumar Balakrishnan, Sanjib Bhattacharyya |
| hCoV-19/USA/CO-CDC-MMB11856324/2021 | EPI_ISL_8157733 | 2021-11-28 | Mako Medical | Centers for Disease Control and Prevention Division of Viral Diseases, Pathogen Discovery | Dakota Howard,Dhwani Batra,Peter Cook,Jason Caravas,Benjamin Rambo-Martin,Scott Sammons,Yvette Unoarumhi,Matthew Schmerer,Kristine Lacek,Tymeckia Kendall,Victoria Caban Figueroa,Shatavia Morrison,Christopher Gulvick,Erisa Sula,Matthew Tugwell,Lauren Moon, |
| hCoV-19/USA/OR-OHSU-213570608/2021 | EPI_ISL_8130839 | 2021-12-16 | OHSU MM Lab | OHSU MM Lab | Yun Wu, Jeannine Lama, Amber Halse, Xuan Qin |
| hCoV-19/Argentina/PAIS-D0215/2021 | EPI_ISL_7620338 | 2021-11-01 | Laboratorio del Hospital Regional Ushuaia Gdor. Ernesto Campos | [illegible] Fuego - Hospital Regional Ushuaia - Centro Austral De Investigaciones Cientv≠ficas - Universidad Nacional De Tierra Del Fuego on behalf PAIS project and ANLIS-Malbran | Gramundi, ID; Gallego, F; Ceballos, SG; Nardi, CF; Rojas, AE; De Roccis, CA; Castro, G; Cv°ceres, SB; Yulan, CB; Boutureira, MF. |
| hCoV-19/Ghana/NMIMR-PUR-21-6929/2021 | EPI_ISL_8065577 | 2021-06-15 | Noguchi Memorial Institute for Medical Research, University of Ghana, Legon, Ghana | Noguchi Memorial Institute for Medical Research, University of Ghana, Legon, Ghana | Bright Adu, Joyce Appiah-Kubi, Quaneeta, Mohktar, Hilda Opoku Frempong, Seyram B. Avonyo, Peace Uche, Keren O. Attiku, Joseph K. H. Bonney, John K. Odoom |
| hCoV-19/Ghana/NMIMR-NCOV-21-55847/2021 | EPI_ISL_8065629 | 2021-07-09 | Noguchi Memorial Institute for Medical Research, University of Ghana, Legon, Ghana | Noguchi Memorial Institute for Medical Research, University of Ghana, Legon, Ghana | Bright Adu, Joyce Appiah-Kubi, Quaneeta, Mohktar, Hilda Opoku Frempong, Seyram B. Avonyo, Peace Uche, Keren O. Attiku, Joseph K. H. Bonney, John K. Odoom |

| | | | | | |
|---|---|---|---|---|---|
| hCoV-19/Ghana/NMIMR-CV-21-203/2021 | EPI_ISL_8065650 | 2021-07-16 | Noguchi Memorial Institute for Medical Research, University of Ghana, Legon, Ghana | Noguchi Memorial Institute for Medical Research, University of Ghana, Legon, Ghana | Bright Adu, Joyce Appiah-Kubi, Quaneeta, Mohktar, Hilda Opoku Frempong, Seyram B. Avonyo, Peace Uche, Keren O. Attiku, Joseph K. H. Bonney, John K. Odoom |
| hCoV-19/Ghana/NMIMR-RT-PUR-10425/2021 | EPI_ISL_8065675 | 2021-09-02 | Noguchi Memorial Institute for Medical Research, University of Ghana, Legon, Ghana | Noguchi Memorial Institute for Medical Research, University of Ghana, Legon, Ghana | Bright Adu, Joyce Appiah-Kubi, Quaneeta, Mohktar, Hilda Opoku Frempong, Seyram B. Avonyo, Peace Uche, Keren O. Attiku, Joseph K. H. Bonney, John K. Odoom |
| hCoV-19/USA/MN-CDC-ASC210523147/2021 | EPI_ISL_7920595 | 2021-11-05 | Aegis Sciences Corporation | Centers for Disease Control and Prevention Division of Viral Diseases, Pathogen Discovery | Dakota Howard,Dhwani Batra,Peter Cook,Jason Caravas,Benjamin Rambo-Martin,Scott Sammons,Yvette Unoarumhi,Matthew Schmerer,Kristine Lacek,Tymeckia Kendall,Victoria Caban Figueroa,Shatavia Morrison,Christopher Gulvick,Erisa Sula,Cyndi Clark,Patrick Campbell |
| hCoV-19/Ghana/NMIMR-NUP-21-9479/2021 | EPI_ISL_8065740 | 2021-09-13 | Noguchi Memorial Institute for Medical Research, University of Ghana, Legon, Ghana | Noguchi Memorial Institute for Medical Research, University of Ghana, Legon, Ghana | Bright Adu, Joyce Appiah-Kubi, Quaneeta, Mohktar, Hilda Opoku Frempong, Seyram B. Avonyo, Peace Uche, Keren O. Attiku, Joseph K. H. Bonney, John K. Odoom |
| hCoV-19/Ghana/NMIMR-NCOV-21-56407/2021 | EPI_ISL_8065727 | 2021-09-13 | Noguchi Memorial Institute for Medical Research, University of Ghana, Legon, Ghana | Noguchi Memorial Institute for Medical Research, University of Ghana, Legon, Ghana | Bright Adu, Joyce Appiah-Kubi, Quaneeta, Mohktar, Hilda Opoku Frempong, Seyram B. Avonyo, Peace Uche, Keren O. Attiku, Joseph K. H. Bonney, John K. Odoom |
| hCoV-19/Ghana/NMIMR-NCOV-21-60353/2021 | EPI_ISL_8065960 | 2021-11-22 | Noguchi Memorial Institute for Medical Research, University of Ghana, Legon, Ghana | Noguchi Memorial Institute for Medical Research, University of Ghana, Legon, Ghana | Bright Adu, Joyce Appiah-Kubi, Quaneeta, Mohktar, Hilda Opoku Frempong, Seyram B. Avonyo, Peace Uche, Keren O. Attiku, Joseph K. H. Bonney, John K. Odoom |
| hCoV-19/Ghana/NMIMR-NCOV-21-59339/2021 | EPI_ISL_8065939 | 2021-10-26 | Noguchi Memorial Institute for Medical Research, University of Ghana, Legon, Ghana | Noguchi Memorial Institute for Medical Research, University of Ghana, Legon, Ghana | Bright Adu, Joyce Appiah-Kubi, Quaneeta, Mohktar, Hilda Opoku Frempong, Seyram B. Avonyo, Peace Uche, Keren O. Attiku, Joseph K. H. Bonney, John K. Odoom |
| hCoV-19/Ghana/NMIMR-CT-21-18630/2021 | EPI_ISL_8065751 | 2021-09-14 | Noguchi Memorial Institute for Medical Research, University of Ghana, Legon, Ghana | Noguchi Memorial Institute for Medical Research, University of Ghana, Legon, Ghana | Bright Adu, Joyce Appiah-Kubi, Quaneeta, Mohktar, Hilda Opoku Frempong, Seyram B. Avonyo, Peace Uche, Keren O. Attiku, Joseph K. H. Bonney, John K. Odoom |
| hCoV-19/Ghana/NMIMR-NUR-21-33307/2021 | EPI_ISL_8065891 | 2021-10-09 | Noguchi Memorial Institute for Medical Research, University of Ghana, Legon, Ghana | Noguchi Memorial Institute for Medical Research, University of Ghana, Legon, Ghana | Bright Adu, Joyce Appiah-Kubi, Quaneeta, Mohktar, Hilda Opoku Frempong, Seyram B. Avonyo, Peace Uche, Keren O. Attiku, Joseph K. H. Bonney, John K. Odoom |
| hCoV-19/USA/MD-HP22227-PIDQHWRHTF/2021 | EPI_ISL_8160051 | 2021-12-20 | Johns Hopkins Hospital Department of Pathology | Johns Hopkins Hospital Department of Pathology | C. Paul Morris, David Gaston, Julie M. Norton, Amary Fall, Raghda Eldesouki, Michael Forman, Matthew Schwartz, Heba H. Mostafa |
| hCoV-19/Botswana/R55B01_BHP_AAC38061/2021 | EPI_ISL_7834393 | 2021-12-13 | Botswana Harvard HIV Reference Laboratory | Botswana Harvard HIV Reference Laboratory | Sikhulile Moyo, Wonderful T. Choga, Dorcas Maruapula, Keoratile Ntshambiwa, Sefetogi Ramaologa, Thongbotho Mphoyakgosi, Boitumelo Zuze, Botshelo Radibe, Legodile Kooepile, Ontlametse T. Bareng, Pamela Smith-Lawrence, Kgomotso Moruisi, Roger Shapiro, Shahi |
| hCoV-19/Kenya/SS4145/2021 | EPI_ISL_7834679 | 2021-11-12 | Pathologist Lancet Kenya | KEMRI-Wellcome Trust Research Programme,Kilifi | Githinji G.,Ochola I, Agoti C.,Mohamed K.S.,de Laurent Z.,Mburu M.W.,Morobe J.,Lambisia A.,Makori T.,Ongera E., Ndwiga L., Mukadam R, Ngari C., Munoko A., D.J.Nokes |

| | | | | | |
|---|---|---|---|---|---|
| hCoV-19/Switzerland/ZH-UZH-IMV-3ba578cc/2021 | EPI_ISL_7632035 | 2021-11-30 | Stadtspital Triemli | Institute of Medical Virology, University of Zurich | Daniel Ehrsam, Isabel St√ºrmer, Catharine Aquino, Joel Wirz, Weihong Qi, Hubert Rehrauer, Verena Kufner, Gabriela Ziltener, Maryam Zaheri, Stefan Schmutz, Annette Audig√©, Maria Gr√ºnberg, Kevin Steiner, Jon Huder, Cyril Shah, Riccarda Capaul, Guido Bloem |
| hCoV-19/USA/CO-CDPHE-2102420738/2021 | EPI_ISL_8162122 | 2021-12-01 | Colorado Department of Public Health and Environment | Colorado Department of Public Health and Environment | Laura Bankers, Molly C. Hetherington-Rauth, Diana Ir, Alexandria Rossheim, Michael Martin, Mandy Waters, Arianna Smith, Shannon R. Matzinger, Emily A. Travanty |
| hCoV-19/PapuaNewGuinea/PNG1863/2021 | EPI_ISL_7835312 | 2021-04-29 | Queensland Medical Laboratories | Microbiological Diagnostic Unit - Public Health Laboratory (MDU-PHL) | Susapu, M., Palou, T., Seemann, T., Horan, K., Sherry, N.L. |
| hCoV-19/TrinidadandTobago/109065/2021 | EPI_ISL_7835340 | 2021-11-26 | Trinidad Public Health Laboratory | CARPHA | SueMin Nathaniel on behalf of CARPHA |
| hCoV-19/USA/NE-CUMC_21347088/2021 | EPI_ISL_7934829 | 2021-12-06 | Creighton University Medical Center | Creighton COVID Consortium | Holly Stessman, Michael Belshan, Richard Goering, Cynthia Watson, Jake Siedlik |
| hCoV-19/SouthAfrica/NICD-N23600/2021 | EPI_ISL_8439593 | 2021-11-29 | AMPATH | National Institute for Communicable Diseases of the National Health Laboratory Service | Amoako DG, Everatt J, Kekana D, Scheepers C, Mohale T, Ntuli N, Mahlangu B, Mnguni A, Ismail A, Bhiman JN, Wolter N |
| hCoV-19/SouthAfrica/NICD-N23569/2021 | EPI_ISL_8439570 | 2021-11-29 | AMPATH | National Institute for Communicable Diseases of the National Health Laboratory Service | Amoako DG, Everatt J, Kekana D, Scheepers C, Mohale T, Ntuli N, Mahlangu B, Mnguni A, Ismail A, Bhiman JN, Wolter N |
| hCoV-19/Jamaica/109795/2021 | EPI_ISL_7837220 | 2021-08-16 | National Public Health Laboratory | CARPHA | SueMin Nathaniel on behalf of CARPHA |
| hCoV-19/SouthAfrica/NICD-N23626/2021 | EPI_ISL_8439611 | 2021-11-29 | AMPATH | National Institute for Communicable Diseases of the National Health Laboratory Service | Amoako DG, Everatt J, Kekana D, Scheepers C, Mohale T, Ntuli N, Mahlangu B, Mnguni A, Ismail A, Bhiman JN, Wolter N |
| hCoV-19/SouthAfrica/NICD-N23721/2021 | EPI_ISL_8439615 | 2021-11-29 | LANCET | National Institute for Communicable Diseases of the National Health Laboratory Service | Amoako DG, Everatt J, Kekana D, Scheepers C, Mohale T, Ntuli N, Mahlangu B, Mnguni A, Ismail A, Bhiman JN, Wolter N |
| hCoV-19/SouthAfrica/NICD-N23766/2021 | EPI_ISL_8439640 | 2021-11-30 | ROB FERREIRA LABORATORY | National Institute for Communicable Diseases of the National Health Laboratory Service | Amoako DG, Everatt J, Kekana D, Scheepers C, Mohale T, Ntuli N, Mahlangu B, Mnguni A, Ismail A, Bhiman JN, Wolter N |
| hCoV-19/SouthAfrica/NICD-N23611/2021 | EPI_ISL_8439600 | 2021-11-29 | AMPATH | National Institute for Communicable Diseases of the National Health Laboratory Service | Amoako DG, Everatt J, Kekana D, Scheepers C, Mohale T, Ntuli N, Mahlangu B, Mnguni A, Ismail A, Bhiman JN, Wolter N |
| hCoV-19/England/BRBR-2E52F95/2021 | EPI_ISL_7838073 | 2021-12-12 | Berkshire and Surrey Pathology Services Lighthouse Laboratory | Wellcome Sanger Institute for the COVID-19 Genomics UK (COG-UK) Consortium | Berkshire and Surrey Pathology Services Lighthouse Laboratory and Alex Alderton, Roberto Amato, Jeffrey Barrett, Sonia Goncalves, Ewan Harrison, David K. Jackson, Ian Johnston, Dominic Kwiatkowski, Cordelia Langford, John Sillitoe on behalf of the Wellcom |
| hCoV-19/Gambia/98998/2021 | EPI_ISL_8439743 | 2021-12-29 | MRCG at LSHTM Genomics lab | MRCG at LSHTM Genomics lab | Abdul Karim sesay, Abdoulie Kanteh, Jarra Manneh, Mariama Kujabi, Bakary Sanyang,Dabiri Damilari, Yaya bah |

| | | | | | |
|---|---|---|---|---|---|
| hCoV-19/England/MILK-2EF6F93/2021 | EPI_ISL_8073016 | 2021-12-15 | Lighthouse Lab in Milton Keynes | Wellcome Sanger Institute for the COVID-19 Genomics UK (COG-UK) Consortium | The Lighthouse Lab in Milton Keynes and Alex Alderton, Roberto Amato, Jeffrey Barrett, Sonia Goncalves, Ewan Harrison, David K. Jackson, Ian Johnston, Dominic Kwiatkowski, Cordelia Langford, John Sillitoe on behalf of the Wellcome Sanger Institute COVID-1 |
| hCoV-19/Zambia/ZMB-129103/2021 | EPI_ISL_7988178 | 2021-12-11 | University of Zambia, School of Veterinary Medicine | UNZAVET and PATH | Mulenga Mwenda-Chimfwembe, Ngonda Saasa, Daniel Bridges, ZNPHI and ZGSC |
| hCoV-19/USA/CA-SEARCH-58946/2021 | EPI_ISL_8186184 | 2021-11-19 | EXCITE Lab | Andersen lab at Scripps Research | David Pride, Sharon Reed, Chip Schooley, Angela Scioscia, Natasha Martin Cheryl Anderson, Sawyer Farmer, Abigail Schnapper, Helena Tubb, Tommy Valles + SEARCH |
| hCoV-19/England/MILK-30788B7/2021 | EPI_ISL_8440701 | 2021-12-30 | Lighthouse Lab in Milton Keynes | Wellcome Sanger Institute for the COVID-19 Genomics UK (COG-UK) Consortium | The Lighthouse Lab in Milton Keynes and Alex Alderton, Roberto Amato, Jeffrey Barrett, Sonia Goncalves, Ewan Harrison, David K. Jackson, Ian Johnston, Dominic Kwiatkowski, Cordelia Langford, John Sillitoe on behalf of the Wellcome Sanger Institute COVID-1 |
| hCoV-19/Morocco/IPM20383438/2021 | EPI_ISL_8186745 | 2021-11-15 | Medical Virology and BSL3 Laboratory | Centre de S√©quen√ßage G√©nomique | EL Hamouchi Adil, Charoute Hicham, Harmak Houda, Bouzidi Aymane, Amalou Ghita, Anga Latifa, Faouzi Abdellah, Zemmouri Faouzia, El Oualid Abdelmjid, Nadifiyine Saloua, Zouheir Yassine, Omondi Francis Carey, Somda Soro Georgina Charlene, Abbad Anas, Chgouri |
| hCoV-19/Morocco/IPM20382805/2021 | EPI_ISL_8186755 | 2021-11-10 | Medical Virology and BSL3 Laboratory | Centre de S√©quen√ßage G√©nomique | EL Hamouchi Adil, Charoute Hicham, Harmak Houda, Bouzidi Aymane, Amalou Ghita, Anga Latifa, Faouzi Abdellah, Zemmouri Faouzia, El Oualid Abdelmjid, Nadifiyine Saloua, Zouheir Yassine, Omondi Francis Carey, Somda Soro Georgina Charlene, Abbad Anas, Chgouri |
| hCoV-19/England/PHEP-YYNUCWN/2021 | EPI_ISL_8075909 | 2021-12-08 | Respiratory Virus Unit, Microbiology Services Colindale, Public Health England | COVID-19 Genomics UK (COG-UK) Consortium | PHE Covid Sequencing Team |
| hCoV-19/Colombia/VAC-LSB-112102988/2021 | EPI_ISL_8187203 | 2021-11-20 | Laboratorio Clinica Sanitas - Sebastian de Belalcazar | CIAT, Laboratorio de Virologia | Ana M. Leiva, Diana Lopez-Alvarez, WilmerJ. Cuellar, Programa Nacional de Caracterizaci√≥n Gen√≥mica de SARS-CoV-2 |
| hCoV-19/Colombia/VAC-LSB-112213980/2021 | EPI_ISL_8187208 | 2021-11-22 | Laboratorio Clinica Sanitas - Sebastian de Belalcazar | CIAT, Laboratorio de Virologia | Ana M. Leiva, Diana Lopez-Alvarez, WilmerJ. Cuellar, Programa Nacional de Caracterizaci√≥n Gen√≥mica de SARS-CoV-2 |
| hCoV-19/Colombia/VAC-LSB-112214422/2021 | EPI_ISL_8187209 | 2021-11-22 | Laboratorio Clinica Sanitas - Sebastian de Belalcazar | CIAT, Laboratorio de Virologia | Ana M. Leiva, Diana Lopez-Alvarez, WilmerJ. Cuellar, Programa Nacional de Caracterizaci√≥n Gen√≥mica de SARS-CoV-2 |
| hCoV-19/India/DL-ILBS-WGS970/2021 | EPI_ISL_8166590 | 2021-12-21 | NRL, DR LAL PATH LAB LTD | ILBS | Ekta Gupta, Varun Suroliya, Pramod Gautam, Reshu Agarwal, Rahul Garg, Urvinder Kaur, Arjun Bhugra, Chhagan Bihari Sharma, Shiv Kumar Sarin |
| hCoV-19/India/DL-ILBS-WGS995/2021 | EPI_ISL_8166613 | 2021-12-22 | CITY X RAY AND SCAN CLINIC PVT LTD | ILBS | Ekta Gupta, Varun Suroliya, Pramod Gautam, Reshu Agarwal, Rahul Garg, Urvinder Kaur, Arjun Bhugra, Chhagan Bihari Sharma, Shiv Kumar Sarin |

| | | | | | |
|---|---|---|---|---|---|
| hCoV-19/England/NORW-310489B/2021 | EPI_ISL_8239884 | 2021-12-13 | Quadram Institute Bioscience | COVID-19 Genomics UK (COG-UK) Consortium | Dave J. Baker, Gemma L. Kay, Alp Aydin, Thanh Le-Viet, Steven Rudder, Ana P. Tedim, Anastasia Kolyva, Maria Diaz, Leonardo de Oliveira Martins, Nabil-Fareed Alikhan, Lizzie Meadows, Rachael Stanley, Rose K Davidson, Ngozi Elumogo, Muhammed Yasir, Sophie J |
| hCoV-19/England/NORW-3104BC2/2021 | EPI_ISL_8239984 | 2021-12-13 | Quadram Institute Bioscience | COVID-19 Genomics UK (COG-UK) Consortium | Dave J. Baker, Gemma L. Kay, Alp Aydin, Thanh Le-Viet, Steven Rudder, Ana P. Tedim, Anastasia Kolyva, Maria Diaz, Leonardo de Oliveira Martins, Nabil-Fareed Alikhan, Lizzie Meadows, Rachael Stanley, Rose K Davidson, Ngozi Elumogo, Muhammed Yasir, Sophie J |
| hCoV-19/England/MILK-2FB2D98/2021 | EPI_ISL_8240843 | 2021-12-20 | Lighthouse Lab in Milton Keynes | Wellcome Sanger Institute for the COVID-19 Genomics UK (COG-UK) Consortium | The Lighthouse Lab in Milton Keynes and Alex Alderton, Roberto Amato, Jeffrey Barrett, Sonia Goncalves, Ewan Harrison, David K. Jackson, Ian Johnston, Dominic Kwiatkowski, Cordelia Langford, John Sillitoe on behalf of the Wellcome Sanger Institute COVID-1 |
| hCoV-19/USA/MA-CDCBI-CRSP_VSVN3PGHIESJSESE/2021 | EPI_ISL_7544427 | 2021-11-30 | Broad Institute Clinical Research Sequencing Platform | Infectious Disease Program, Broad Institute of Harvard and MIT | Siddle,K.J., Adams,G., Pearlman,L., Gladden-Young,A., Vicente,G., Blumenstiel,B., DeFelice,M., Lee,M., McGovern,S., Lagerborg,K., Rudy,M., DeRuff,K., Carter,A., Normandin,E., Bauer,M., Reilly,S., Tomkins-Tinch,C., Loreth,C., Chaluvadi,S., Meldrim,J., Gran |
| hCoV-19/England/MILK-2F9343E/2021 | EPI_ISL_8168662 | 2021-12-20 | Lighthouse Lab in Milton Keynes | Wellcome Sanger Institute for the COVID-19 Genomics UK (COG-UK) Consortium | The Lighthouse Lab in Milton Keynes and Alex Alderton, Roberto Amato, Jeffrey Barrett, Sonia Goncalves, Ewan Harrison, David K. Jackson, Ian Johnston, Dominic Kwiatkowski, Cordelia Langford, John Sillitoe on behalf of the Wellcome Sanger Institute COVID-1 |
| hCoV-19/Chile/RM-193647/2021 | EPI_ISL_8187566 | 2021-12-13 | Genetica Molecular and Subdepartamento de Virologia ISP Chile | Instituto de Salud Publica de Chile | Karen Orostica, Constanza Campano, Barbara Parra, Patricia Bustos, Rodrigo Fasce, Andres Castillo, Jorge Fernandez |
| hCoV-19/Chile/VS-194271/2021 | EPI_ISL_8187593 | 2021-12-14 | Genetica Molecular and Subdepartamento de Virologia ISP Chile | Instituto de Salud Publica de Chile | Karen Orostica, Constanza Campano, Barbara Parra, Patricia Bustos, Rodrigo Fasce, Andres Castillo, Jorge Fernandez |
| hCoV-19/Chile/RM-194483/2021 | EPI_ISL_8187628 | 2021-12-16 | Genetica Molecular and Subdepartamento de Virologia ISP Chile | Instituto de Salud Publica de Chile | Karen Orostica, Constanza Campano, Barbara Parra, Patricia Bustos, Rodrigo Fasce, Andres Castillo, Jorge Fernandez |
| hCoV-19/Chile/RM-194498/2021 | EPI_ISL_8187647 | 2021-12-15 | Genetica Molecular and Subdepartamento de Virologia ISP Chile | Instituto de Salud Publica de Chile | Karen Orostica, Constanza Campano, Barbara Parra, Patricia Bustos, Rodrigo Fasce, Andres Castillo, Jorge Fernandez |
| hCoV-19/Chile/RM-194500/2021 | EPI_ISL_8187649 | 2021-12-15 | Genetica Molecular and Subdepartamento de Virologia ISP Chile | Instituto de Salud Publica de Chile | Karen Orostica, Constanza Campano, Barbara Parra, Patricia Bustos, Rodrigo Fasce, Andres Castillo, Jorge Fernandez |
| hCoV-19/Chile/RM-195236/2021 | EPI_ISL_8187860 | 2021-12-18 | Genetica Molecular and Subdepartamento de Virologia ISP Chile | Instituto de Salud Publica de Chile | Karen Orostica, Constanza Campano, Barbara Parra, Patricia Bustos, Rodrigo Fasce, Andres Castillo, Jorge Fernandez |
| hCoV-19/Chile/VS-194985/2021 | EPI_ISL_8187850 | 2021-12-17 | Genetica Molecular and Subdepartamento de Virologia ISP Chile | Instituto de Salud Publica de Chile | Karen Orostica, Constanza Campano, Barbara Parra, Patricia Bustos, Rodrigo Fasce, Andres Castillo, Jorge Fernandez |

| | | | | | |
|---|---|---|---|---|---|
| hCoV-19/Chile/RM-195244/2021 | EPI_ISL_8187864 | 2021-12-17 | Genetica Molecular and Subdepartamento de Virologia ISP Chile | Instituto de Salud Publica de Chile | Karen Orostica, Constanza Campano, Barbara Parra, Patricia Bustos, Rodrigo Fasce, Andres Castillo, Jorge Fernandez |
| hCoV-19/USA/OH-CDC-ASC210535767/2021 | EPI_ISL_8078046 | 2021-12-14 | Aegis Sciences Corporation | Centers for Disease Control and Prevention Division of Viral Diseases, Pathogen Discovery | Dakota Howard,Dhwani Batra,Peter Cook,Jason Caravas,Benjamin Rambo-Martin,Scott Sammons,Yvette Unoarumhi,Matthew Schmerer,Kristine Lacek,Tymeckia Kendall,Victoria Caban Figueroa,Shatavia Morrison,Christopher Gulvick,Erisa Sula,Cyndi Clark,Patrick Campbell |
| hCoV-19/USA/PA-CDC-ASC210536055/2021 | EPI_ISL_8078111 | 2021-12-14 | Aegis Sciences Corporation | Centers for Disease Control and Prevention Division of Viral Diseases, Pathogen Discovery | Dakota Howard,Dhwani Batra,Peter Cook,Jason Caravas,Benjamin Rambo-Martin,Scott Sammons,Yvette Unoarumhi,Matthew Schmerer,Kristine Lacek,Tymeckia Kendall,Victoria Caban Figueroa,Shatavia Morrison,Christopher Gulvick,Erisa Sula,Cyndi Clark,Patrick Campbell |
| hCoV-19/Chile/VS-195986/2021 | EPI_ISL_8187991 | 2021-12-20 | Genetica Molecular and Subdepartamento de Virologia ISP Chile | Instituto de Salud Publica de Chile | Karen Orostica, Constanza Campano, Barbara Parra, Patricia Bustos, Rodrigo Fasce, Andres Castillo, Jorge Fernandez |
| hCoV-19/Chile/AT-194893/2021 | EPI_ISL_8188040 | 2021-12-15 | Genetica Molecular and Subdepartamento de Virologia ISP Chile | Instituto de Salud Publica de Chile | Karen Orostica, Constanza Campano, Barbara Parra, Patricia Bustos, Rodrigo Fasce, Andres Castillo, Jorge Fernandez |
| hCoV-19/Mexico/CMX-INMEGEN-43-93/2021 | EPI_ISL_7852284 | 2021-11-24 | ARION GENETICA | Instituto Nacional de Medicina Genomica | Cedro-Tanda A, Escobar-Arrazola MA, Ramirez-Vega O, Garnica-Lopez Dora,Uribe-Figueroa Laura, Roldan-Castillo Magaly, Verea Jazmin, Rangel-DeLeon D, Yair Alfaro-Mora, Hidalgo-Miranda A, Mendoza-Vargas A, Reyes-Grajeda JP, Herrera-Montalvo LA. |
| hCoV-19/USA/CO-CDC-MMB11841786/2021 | EPI_ISL_8078673 | 2021-11-26 | Mako Medical | Centers for Disease Control and Prevention Division of Viral Diseases, Pathogen Discovery | Dakota Howard,Dhwani Batra,Peter Cook,Jason Caravas,Benjamin Rambo-Martin,Scott Sammons,Yvette Unoarumhi,Matthew Schmerer,Kristine Lacek,Tymeckia Kendall,Victoria Caban Figueroa,Shatavia Morrison,Christopher Gulvick,Erisa Sula,Matthew Tugwell,Lauren Moon, |
| hCoV-19/USA/CO-CDC-MMB11842688/2021 | EPI_ISL_8078967 | 2021-11-26 | Mako Medical | Centers for Disease Control and Prevention Division of Viral Diseases, Pathogen Discovery | Dakota Howard,Dhwani Batra,Peter Cook,Jason Caravas,Benjamin Rambo-Martin,Scott Sammons,Yvette Unoarumhi,Matthew Schmerer,Kristine Lacek,Tymeckia Kendall,Victoria Caban Figueroa,Shatavia Morrison,Christopher Gulvick,Erisa Sula,Matthew Tugwell,Lauren Moon, |
| hCoV-19/USA/CO-CDC-MMB11841007/2021 | EPI_ISL_8078966 | 2021-11-26 | Mako Medical | Centers for Disease Control and Prevention Division of Viral Diseases, Pathogen Discovery | Dakota Howard,Dhwani Batra,Peter Cook,Jason Caravas,Benjamin Rambo-Martin,Scott Sammons,Yvette Unoarumhi,Matthew Schmerer,Kristine Lacek,Tymeckia Kendall,Victoria Caban Figueroa,Shatavia Morrison,Christopher Gulvick,Erisa Sula,Matthew Tugwell,Lauren Moon, |

| | | | | | |
|---|---|---|---|---|---|
| hCoV-19/USA/CO-CDC-MMB11839884/2021 | EPI_ISL_8078985 | 2021-11-26 | Mako Medical | Centers for Disease Control and Prevention Division of Viral Diseases, Pathogen Discovery | Dakota Howard,Dhwani Batra,Peter Cook,Jason Caravas,Benjamin Rambo-Martin,Scott Sammons,Yvette Unoarumhi,Matthew Schmerer,Kristine Lacek,Tymeckia Kendall,Victoria Caban Figueroa,Shatavia Morrison,Christopher Gulvick,Erisa Sula,Matthew Tugwell,Lauren Moon, |
| hCoV-19/USA/CO-CDC-MMB11844153/2021 | EPI_ISL_8079013 | 2021-11-26 | Mako Medical | Centers for Disease Control and Prevention Division of Viral Diseases, Pathogen Discovery | Dakota Howard,Dhwani Batra,Peter Cook,Jason Caravas,Benjamin Rambo-Martin,Scott Sammons,Yvette Unoarumhi,Matthew Schmerer,Kristine Lacek,Tymeckia Kendall,Victoria Caban Figueroa,Shatavia Morrison,Christopher Gulvick,Erisa Sula,Matthew Tugwell,Lauren Moon, |
| hCoV-19/USA/NY-PRL-2021_1208_03J14/2021 | EPI_ISL_7852168 | 2021-12-03 | Pandemic Response Lab - NYC | Pandemic Response Lab, R&D | Henry Lee, Michael Hammerling, Melissa Hopkins, Cybill del Castillo, Shinyoung Clair Kang, William Ward, Pradeep Bugga, Sol Rey, Dylan Law, Alex Carpio, Isabel Fernandez Escapa, Haiping Hao, Jon Laurent |
| hCoV-19/USA/CO-CDC-MMB11848258/2021 | EPI_ISL_8079122 | 2021-11-26 | Mako Medical | Centers for Disease Control and Prevention Division of Viral Diseases, Pathogen Discovery | Dakota Howard,Dhwani Batra,Peter Cook,Jason Caravas,Benjamin Rambo-Martin,Scott Sammons,Yvette Unoarumhi,Matthew Schmerer,Kristine Lacek,Tymeckia Kendall,Victoria Caban Figueroa,Shatavia Morrison,Christopher Gulvick,Erisa Sula,Matthew Tugwell,Lauren Moon, |
| hCoV-19/USA/CO-CDC-MMB11853884/2021 | EPI_ISL_8079263 | 2021-11-27 | Mako Medical | Centers for Disease Control and Prevention Division of Viral Diseases, Pathogen Discovery | Dakota Howard,Dhwani Batra,Peter Cook,Jason Caravas,Benjamin Rambo-Martin,Scott Sammons,Yvette Unoarumhi,Matthew Schmerer,Kristine Lacek,Tymeckia Kendall,Victoria Caban Figueroa,Shatavia Morrison,Christopher Gulvick,Erisa Sula,Matthew Tugwell,Lauren Moon, |
| hCoV-19/USA/NY-CDC-LC0408204/2021 | EPI_ISL_7915270 | 2021-12-02 | Laboratory Corporation of America | Centers for Disease Control and Prevention Division of Viral Diseases, Pathogen Discovery | Dakota Howard,Dhwani Batra,Peter Cook,Jason Caravas,Benjamin Rambo-Martin,Scott Sammons,Yvette Unoarumhi,Matthew Schmerer,Kristine Lacek,Tymeckia Kendall,Victoria Caban Figueroa,Shatavia Morrison,Christopher Gulvick,Minoo Agarwal,Eyad Almasri,Debbie Boles |
| hCoV-19/Netherlands/ZH-MZ-1035/2021 | EPI_ISL_8170696 | 2021-01-11 | Maasstad Ziekenhuis | Maasstad ziekenhuis | Pieter Smit |
| hCoV-19/Netherlands/ZH-MZ-1036/2021 | EPI_ISL_8170674 | 2021-01-11 | Maasstad Ziekenhuis | Maasstad ziekenhuis | Pieter Smit |
| hCoV-19/Netherlands/ZH-MZ-1038/2021 | EPI_ISL_8170675 | 2021-02-11 | Maasstad Ziekenhuis | Maasstad ziekenhuis | Pieter Smit |
| hCoV-19/Netherlands/ZH-MZ-1039/2021 | EPI_ISL_8170668 | 2021-02-11 | Maasstad Ziekenhuis | Maasstad ziekenhuis | Pieter Smit |
| hCoV-19/Netherlands/ZH-MZ-1027/2021 | EPI_ISL_8170662 | 2021-01-11 | Maasstad Ziekenhuis | Maasstad ziekenhuis | Pieter Smit |
| hCoV-19/Netherlands/ZH-MZ-1059/2021 | EPI_ISL_8170684 | 2021-04-11 | Maasstad Ziekenhuis | Maasstad ziekenhuis | Pieter Smit |

| | | | | | |
|---|---|---|---|---|---|
| hCoV-19/Netherlands/ZH-MZ-1054/2021 | EPI_ISL_8170658 | 2021-04-11 | Maasstad Ziekenhuis | Maasstad ziekenhuis | Pieter Smit |
| hCoV-19/Netherlands/ZH-MZ-1260/2021 | EPI_ISL_8170974 | 2021-02-12 | Maasstad Ziekenhuis | Maasstad ziekenhuis | Pieter Smit |
| hCoV-19/Netherlands/ZH-MZ-1264/2021 | EPI_ISL_8171004 | 2021-03-12 | Maasstad Ziekenhuis | Maasstad ziekenhuis | Pieter Smit |
| hCoV-19/Japan/IC-2377/2021 | EPI_ISL_7860220 | 2021-12-11 | SARS-CoV-2 testing team, National Institute of Infectious Diseases | Pathogen Genomics Center, National Institute of Infectious Diseases | Tsuyoshi Sekizuka, Kentaro Itokawa, Rina Tanaka, Masanori Hashino, Nozomu Hanaoka, Masumichi Saito, Naomi Nojiri, Hazuka Y Furihata, Tsuguto Fujimoto, Makoto Kuroda |
| hCoV-19/USA/MS-CDC-ASC210535451/2021 | EPI_ISL_8082027 | 2021-12-12 | Aegis Sciences Corporation | Centers for Disease Control and Prevention Division of Viral Diseases, Pathogen Discovery | Dakota Howard,Dhwani Batra,Peter Cook,Jason Caravas,Benjamin Rambo-Martin,Scott Sammons,Yvette Unoarumhi,Matthew Schmerer,Kristine Lacek,Tymeckia Kendall,Victoria Caban Figueroa,Shatavia Morrison,Christopher Gulvick,Erisa Sula,Cyndi Clark,Patrick Campbell |
| hCoV-19/Wales/PHWC-PGG7GM/2021 | EPI_ISL_8172275 | 2021-12-11 | Originating lab: Wales Specialist Virology Centre Sequencing lab: Pathogen Genomics Unit | Public Health Wales Microbiology Cardiff Wales Specialist Virology Centre | Catherine Moore, Johnathan Evans, Laura Gifford, Malorie Perry, Simon Cottrell, Angela Marchbank, Alec Birchley, Alexander Adams, Amy Gaskin, Bree Gatica-Wilcox, Jason Coombes, Joel Southgate, Lauren Gilbert, Lee Graham, Nicole Pacchiarini, Sara Kumziene- |
| hCoV-19/England/QEUH-3079C8D/2021 | EPI_ISL_8444750 | 2021-12-29 | Lighthouse Lab in Glasgow | Wellcome Sanger Institute for the COVID-19 Genomics UK (COG-UK) Consortium | Harper VanSteenhouse, Yumi Kasai, David Gray, Carol Clugston, Anna Dominiczak and Alex Alderton, Roberto Amato, Jeffrey Barrett, Sonia Goncalves, Ewan Harrison, David K. Jackson, Ian Johnston, Dominic Kwiatkowski, Cordelia Langford, John Sillitoe on behal |
| hCoV-19/USA/TN-CDC-ASC210391853/2021 | EPI_ISL_7922070 | 2021-11-06 | Aegis Sciences Corporation | Centers for Disease Control and Prevention Division of Viral Diseases, Pathogen Discovery | Dakota Howard,Dhwani Batra,Peter Cook,Jason Caravas,Benjamin Rambo-Martin,Scott Sammons,Yvette Unoarumhi,Matthew Schmerer,Kristine Lacek,Tymeckia Kendall,Victoria Caban Figueroa,Shatavia Morrison,Christopher Gulvick,Erisa Sula,Cyndi Clark,Patrick Campbell |
| hCoV-19/England/MILK-2E70F55/2021 | EPI_ISL_7863347 | 2021-12-13 | Lighthouse Lab in Milton Keynes | Wellcome Sanger Institute for the COVID-19 Genomics UK (COG-UK) Consortium | The Lighthouse Lab in Milton Keynes and Alex Alderton, Roberto Amato, Jeffrey Barrett, Sonia Goncalves, Ewan Harrison, David K. Jackson, Ian Johnston, Dominic Kwiatkowski, Cordelia Langford, John Sillitoe on behalf of the Wellcome Sanger Institute COVID-1 |
| hCoV-19/Georgia/Tb-SNGS2138/2021 | EPI_ISL_7924965 | 2021-12-11 | Department for Virology, Molecular Biology and Genome Research, R. G. Lugar Center for Public Health Research, National Center for Disease Control and Public Health (NCDC) of Georgia. | [illegible] Biology and Genome Research, R. G. Lugar Center for Public Health Research, National Center for Disease Control and Public Health (NCDC) of Georgia. | Giorgi Tomashvili, Giorgi Gogoladze, Meri Pantsulaia, Gvantsa Brachveli, Salome Javashvili, Nino Berishvili, Tata Imnadze, Ana Papkiauri, Gvantsa Chanturia, Ann Machablishvili, Nato Kotaria, Marine Murtskhvaladze, Lela Sabadze, Mari Gavashelidze, Tamar Ja |

| | | | | | |
|---|---|---|---|---|---|
| hCoV-19/Canada/ON-PHL-21-41505/2021 | EPI_ISL_7334880 | 2021-11-26 | Public Health Ontario Laboratory | Public Health Ontario Laboratory | Vanessa G Allen, Philip Banh, Yao Chen, Richard de Borja, Alireza Eshaghi, Nahuel Fittipaldi, Christine Frantz, Jonathan B Gubbay, Jennifer L Guthrie, Lawrence Heisler, Esha Joshi, Michael Laszloffy, Aimin Li, Michael CY Li, Dean Maxwell, Sandeep Nagra, S |
| hCoV-19/Netherlands/ZH-MZ-1271/2021 | EPI_ISL_8173126 | 2021-04-12 | Maasstad Ziekenhuis | Maasstad ziekenhuis | Pieter Smit |
| hCoV-19/England/LSPA-2FBE89B/2021 | EPI_ISL_8243298 | 2021-12-20 | Rosalind Franklin Laboratory | Wellcome Sanger Institute for the COVID-19 Genomics UK (COG-UK) Consortium | Donald Fraser, Suki Lee, Rob Howes, The Rosalind Franklin Laboratory and Alex Alderton, Roberto Amato, Jeffrey Barrett, Sonia Goncalves, Ewan Harrison, David K. Jackson, Ian Johnston, Dominic Kwiatkowski, Cordelia Langford, John Sillitoe on behalf of the |
| hCoV-19/USA/MI-CDC-ASC210457566/2021 | EPI_ISL_7931449 | 2021-11-12 | Aegis Sciences Corporation | Centers for Disease Control and Prevention Division of Viral Diseases, Pathogen Discovery | Dakota Howard,Dhwani Batra,Peter Cook,Jason Caravas,Benjamin Rambo-Martin,Scott Sammons,Yvette Unoarumhi,Matthew Schmerer,Kristine Lacek,Tymeckia Kendall,Victoria Caban Figueroa,Shatavia Morrison,Christopher Gulvick,Erisa Sula,Cyndi Clark,Patrick Campbell |
| hCoV-19/Slovenia/21-17271/2021 | EPI_ISL_7931807 | 2021-11-22 | Department of Public Health Microbiology Ljubljana, National Laboratory for Health, Environment and Food | Microbiology Ljubljana, National Laboratory for Health, Environment and Food | Tom Koritnik, Martin Bosilj, Katarina Prosenc, Natasa Berginc, Verica Mioc, Marija Trkov |
| hCoV-19/Poland/WSSEGorzow-21S2057/2021 | EPI_ISL_7869977 | 2021-12-05 | NZOZ MEDITEST Diagnostyka Medyczna Jacek Podolski | Epidemiologiczna w Gorzowie Wielkopolskim | Elzbieta Justynska, Renata Siegel, Klaudia Kobendza-Wlodarczak, Patrycja Faberska and Marek Margol |
| hCoV-19/England/PHEP-YYNTR64/2021 | EPI_ISL_7993935 | 2021-12-06 | Respiratory Virus Unit, Microbiology Services Colindale, Public Health England | COVID-19 Genomics UK (COG-UK) Consortium | PHE Covid Sequencing Team |
| hCoV-19/USA/MI-CDC-STM-2QK6YXPN8/2021 | EPI_ISL_7942355 | 2021-12-03 | Helix | Centers for Disease Control and Prevention Division of Viral Diseases, Pathogen Discovery | Dakota Howard,Dhwani Batra,Peter Cook,Jason Caravas,Benjamin Rambo-Martin,Scott Sammons,Yvette Unoarumhi,Matthew Schmerer,Kristine Lacek,Tymeckia Kendall,Victoria Caban Figueroa,Helix CA,Shatavia Morrison,Christopher Gulvick,Erisa Sula,Clinton Paden,Dunca |
| hCoV-19/Singapore/9012/2021 | EPI_ISL_7877331 | 2021-12-14 | National Public Health Laboratory, National Centre for Infectious Diseases | National Centre for Infectious Diseases | Katherine Ching, Benny Yeo Zhenyang Zhou, Royce Ang, Samuel Loo, Lin Cui, Raymond Tzer Pin Lin |
| hCoV-19/Singapore/9010/2021 | EPI_ISL_7877372 | 2021-12-14 | National Public Health Laboratory, National Centre for Infectious Diseases | National Centre for Infectious Diseases | Katherine Ching, Benny Yeo Zhenyang Zhou, Royce Ang, Samuel Loo, Lin Cui, Raymond Tzer Pin Lin |
| hCoV-19/USA/NM-CDC-ASC210718179/2021 | EPI_ISL_7946535 | 2021-11-27 | Aegis Sciences Corporation | Centers for Disease Control and Prevention Division of Viral Diseases, Pathogen Discovery | Dakota Howard,Dhwani Batra,Peter Cook,Jason Caravas,Benjamin Rambo-Martin,Scott Sammons,Yvette Unoarumhi,Matthew Schmerer,Kristine Lacek,Tymeckia Kendall,Victoria Caban Figueroa,Shatavia Morrison,Christopher Gulvick,Erisa Sula,Cyndi Clark,Patrick Campbell |
| hCoV-19/USA/CA-SEARCH-58480/2021 | EPI_ISL_7946576 | 2021-02-09 | EXCITE Lab | Andersen lab at Scripps Research | Sydney Morgan + Marni Jacobs + Celestine Magallanes + Holly Valentine + Louise C. Laurent + SEARCH |
| hCoV-19/Australia/QLD2507C/2021 | EPI_ISL_7947856 | 2021-11-05 | Public Health Virology, Forensic and Scientific Services 39 Kessels Rd, Coopers Plains QLD 4108 | Public Health Virology, Forensic and Scientific Services 39 Kessels Rd, Coopers Plains QLD 4108 | Alyssa T. Pyke |

| | | | | | |
|---|---|---|---|---|---|
| hCoV-19/USA/un-UW-21120954582/2021 | EPI_ISL_7951092 | 2021-12-09 | UW Virology Lab | UW Virology Lab | Pavitra Roychoudhury, Shah Mohamed Bakhash, Pooneh Hajian, Seffir T. Wendm, Isabel Arnould, Ricardo Perez, Hong Xie, Tien V. Nguyen, Sean Ellis, Saraswathi Sathees, Meei-Li Huang, Keith R Jerome, Alexander Greninger |
| hCoV-19/USA/un-UW-21120982000/2021 | EPI_ISL_7951093 | 2021-12-09 | UW Virology Lab | UW Virology Lab | Pavitra Roychoudhury, Shah Mohamed Bakhash, Pooneh Hajian, Seffir T. Wendm, Isabel Arnould, Ricardo Perez, Hong Xie, Tien V. Nguyen, Sean Ellis, Saraswathi Sathees, Meei-Li Huang, Keith R Jerome, Alexander Greninger |
| hCoV-19/USA/WA-UW-21120695101/2021 | EPI_ISL_7951097 | 2021-12-06 | UW Virology Lab | UW Virology Lab | Pavitra Roychoudhury, Shah Mohamed Bakhash, Pooneh Hajian, Seffir T. Wendm, Isabel Arnould, Ricardo Perez, Hong Xie, Tien V. Nguyen, Sean Ellis, Saraswathi Sathees, Meei-Li Huang, Keith R Jerome, Alexander Greninger |
| hCoV-19/Colombia/PUT-INS-VG-10607/2021 | EPI_ISL_7951634 | 2021-11-24 | LDSP PUTUMAYO | Instituto Nacional de Salud-Direcci√≥n de Investigaci√≥n en Salud P√∫blica | Katherine Laiton-Donato, Carlos Franco-Mu√±oz, Diego A. √Ålvarez-D√≠az, H√©ctor Alejandro Ruiz-Moreno, Diego Andr√©s Prada, Jhonnatan Reales-Gonz√°lez, Sheryll Corchuelo, Mar√≠a T. Herrera-Sep√∫lveda, Juli√°n Naizaque, Gerardo Santamar√≠a, Jorge Rivera, P |
| hCoV-19/Israel/ICH-741105426/2021 | EPI_ISL_7952909 | 2021-12-14 | Clinical Microbiology Laboratory, Tel Aviv Sourasky Medical Center | Clinical Microbiology Laboratory, Tel Aviv Sourasky Medical Center | Katya Levytskyi, Ora Halutz, Alon Ziv, Lior Handler, Amos Adler |
| hCoV-19/Israel/ICH-741105544/2021 | EPI_ISL_7953010 | 2021-12-15 | Clinical Microbiology Laboratory, Tel Aviv Sourasky Medical Center | Clinical Microbiology Laboratory, Tel Aviv Sourasky Medical Center | Katya Levytskyi, Ora Halutz, Alon Ziv, Lior Handler, Amos Adler |
| hCoV-19/Scotland/EDB31384/2021 | EPI_ISL_7953239 | 2021-11-24 | Virology Department, Royal Infirmary of Edinburgh, NHS Lothian / School of Biological Sciences, University of Edinburgh | COVID-19 Genomics UK (COG-UK) Consortium | McHugh M, Dewar R, Cotton S, Gallagher A, Maloney D, Fernandez G, O‚ÄôToole √Å, Scher E, Hill V, McCrone JT, Colquhoun R, Yu X, Jackson B, Rambaut A, Templeton K |
| hCoV-19/USA/MN-MDH-18853/2021 | EPI_ISL_8173839 | 2021-11-28 | Regions Hospital | Minnesota Department of Health, Public Health Laboratory | Jacob Garfin, Matt Plumb, Kelly Pung, Elizabeth Horn, Sarah Namugenyi, Alyssa Mondelli, and Xiong Wang |
| hCoV-19/NewZealand/21MV3736/2021 | EPI_ISL_8188884 | 2021-12-26 | PathLab Bay of Plenty | Institute of Environmental Science and Research (ESR) | Sarah Cockerton, Clare Gebbie, Dhanisha Patel, Rachel Boyle, SallyAnn Harbison, Olivia Stroeven, Xiaoyun Ren, Matt Storey, Nikki Freed, Muhammad Faisal, Jing Wang, Hermes Perez, Anja Werno, Antje van der Linden, Arlo Upton, Chris Mansell, David Hammer, Dr |
| hCoV-19/NewZealand/21MV3745/2021 | EPI_ISL_8189015 | 2021-12-22 | LabPLUS | Institute of Environmental Science and Research (ESR) | Sarah Cockerton, Clare Gebbie, Dhanisha Patel, Rachel Boyle, SallyAnn Harbison, Olivia Stroeven, Xiaoyun Ren, Matt Storey, Nikki Freed, Muhammad Faisal, Jing Wang, Hermes Perez, Anja Werno, Antje van der Linden, Arlo Upton, Chris Mansell, David Hammer, Dr |
| hCoV-19/USA/MN-UMGC-25315/2021 | EPI_ISL_7960083 | 2021-11-18 | Public Health Laboratory, Minnesota Department of Health | University of Minnesota Genomics Center | Daryl M. Gohl, John Garbe, Jaquelyn Kuriger-Laber, Corbin Dirkx |
| hCoV-19/NewZealand/21MV3777/2021 | EPI_ISL_8189027 | 2021-12-28 | LabPLUS | Institute of Environmental Science and Research (ESR) | Sarah Cockerton, Clare Gebbie, Dhanisha Patel, Rachel Boyle, SallyAnn Harbison, Olivia Stroeven, Xiaoyun Ren, Matt Storey, Nikki Freed, Muhammad Faisal, Jing Wang, Hermes Perez, Anja Werno, Antje van der Linden, Arlo Upton, Chris Mansell, David Hammer, Dr |

| | | | | | |
|---|---|---|---|---|---|
| hCoV-19/Slovenia/474344/2021 | EPI_ISL_7966421 | 2021-11-05 | Institute of Microbiology and Immunology, Faculty of Medicine, University of Ljubljana | Institute of Microbiology and Immunology, Faculty of Medicine, University of Ljubljana | Alen Suljiƒç, Samo Zakotnik, Toma≈æ Mark Zorec, Doroteja Vlaj, Andra≈æ Celar, Tina ≈Ωiviƒç, Domen Lazar, Tina Gabrov≈°ek, Patricija Pozvek, ≈†pela Pleh, Mi≈°a Korva, Mario Poljak, Tatjana Av≈°iƒç ,Äì ≈Ωupanc |
| hCoV-19/Croatia/15833/2021 | EPI_ISL_8174423 | 2021-12-08 | Zavod za javno zdravstvo Karlovaƒçke ≈æupanije | Hrvatski zavod za javno zdravstvo | Irena Tabain, Ivana Ferenƒçak, Josipa Kuzle, Anita Juriƒá, Dragan Juriƒá |
| hCoV-19/England/MILK-2E954A6/2021 | EPI_ISL_7969530 | 2021-12-14 | Lighthouse Lab in Milton Keynes | Wellcome Sanger Institute for the COVID-19 Genomics UK (COG-UK) Consortium | The Lighthouse Lab in Milton Keynes and Alex Alderton, Roberto Amato, Jeffrey Barrett, Sonia Goncalves, Ewan Harrison, David K. Jackson, Ian Johnston, Dominic Kwiatkowski, Cordelia Langford, John Sillitoe on behalf of the Wellcome Sanger Institute COVID-1 |
| hCoV-19/NewZealand/21MV3699/2021 | EPI_ISL_8188936 | 2021-12-22 | LabPLUS | Institute of Environmental Science and Research (ESR) | Sarah Cockerton, Clare Gebbie, Dhanisha Patel, Rachel Boyle, SallyAnn Harbison, Olivia Stroeven, Xiaoyun Ren, Matt Storey, Nikki Freed, Muhammad Faisal, Jing Wang, Hermes Perez, Anja Werno, Antje van der Linden, Arlo Upton, Chris Mansell, David Hammer, Dr |
| hCoV-19/Denmark/DCGC-266605/2021 | EPI_ISL_7731064 | 2021-11-30 | Department of Bacteria, Parasites and Fungi, Statens Serum Institut, Copenhagen, Denmark | Statens Serum Institut Bioinformatics and Microbial Genomics | Danish Covid-19 Genome Consortium |
| hCoV-19/SouthAfrica/NICD-N22652/2021 | EPI_ISL_7971539 | 2021-09-03 | VERMAAK LABORATORY | National Institute for Communicable Diseases of the National Health Laboratory Service | Amoako DG, Everatt J, Kekana D, Scheepers C, Mohale T, Ntuli N, Mahlangu B, Mnguni A, Ismail A, Bhiman JN, Wolter N |
| hCoV-19/SouthAfrica/NICD-N22682/2021 | EPI_ISL_7971548 | 2021-11-25 | VERMAAK LABORATORY | National Institute for Communicable Diseases of the National Health Laboratory Service | Amoako DG, Everatt J, Kekana D, Scheepers C, Mohale T, Ntuli N, Mahlangu B, Mnguni A, Ismail A, Bhiman JN, Wolter N |
| hCoV-19/SouthAfrica/NICD-N23021/2021 | EPI_ISL_7971569 | 2021-11-30 | TSHEPONG LABORATORY | National Institute for Communicable Diseases of the National Health Laboratory Service | Amoako DG, Everatt J, Kekana D, Scheepers C, Mohale T, Ntuli N, Mahlangu B, Mnguni A, Ismail A, Bhiman JN, Wolter N |
| hCoV-19/SouthAfrica/NICD-N23188/2021 | EPI_ISL_7971630 | 2021-11-30 | KIMBERLEY LABORATORY | National Institute for Communicable Diseases of the National Health Laboratory Service | Amoako DG, Everatt J, Kekana D, Scheepers C, Mohale T, Ntuli N, Mahlangu B, Mnguni A, Ismail A, Bhiman JN, Wolter N |
| hCoV-19/SouthAfrica/NICD-N23195/2021 | EPI_ISL_7971635 | 2021-12-03 | KIMBERLEY LABORATORY | National Institute for Communicable Diseases of the National Health Laboratory Service | Amoako DG, Everatt J, Kekana D, Scheepers C, Mohale T, Ntuli N, Mahlangu B, Mnguni A, Ismail A, Bhiman JN, Wolter N |
| hCoV-19/SouthAfrica/NICD-N22855/2021 | EPI_ISL_7971684 | 2021-11-30 | CHRIS HANI BARAGWANATH LABORATORY | National Institute for Communicable Diseases of the National Health Laboratory Service | Amoako DG, Everatt J, Kekana D, Scheepers C, Mohale T, Ntuli N, Mahlangu B, Mnguni A, Ismail A, Bhiman JN, Wolter N |
| hCoV-19/SouthAfrica/NICD-N22894/2021 | EPI_ISL_7971715 | 2021-10-12 | EAST LONDON LABORATORY | National Institute for Communicable Diseases of the National Health Laboratory Service | Amoako DG, Everatt J, Kekana D, Scheepers C, Mohale T, Ntuli N, Mahlangu B, Mnguni A, Ismail A, Bhiman JN, Wolter N |
| hCoV-19/SouthAfrica/NICD-N22895/2021 | EPI_ISL_7971716 | 2021-11-29 | EAST LONDON LABORATORY | National Institute for Communicable Diseases of the National Health Laboratory Service | Amoako DG, Everatt J, Kekana D, Scheepers C, Mohale T, Ntuli N, Mahlangu B, Mnguni A, Ismail A, Bhiman JN, Wolter N |

| | | | | | |
|---|---|---|---|---|---|
| hCoV-19/Malaysia/USM_Monash_JKNK_NCOV16073/2021 | EPI_ISL_7972318 | 2021-02-25 | Kota Bharu Public Health Laboratory | Molecular Research Laboratory | Chan Yean Yean, Nurfadhlina Musa, Muhammad Fazli bin Khalid,Engku Nur Syafirah bt Engku Abd Rahman, Muhammad Zarul Hanifah Bin Md Zoqratt, Lim Shu Yong, Lau Nyok Sean, Syahida binti Omar, Wan Mohd Zahiruddin Wan Mohammad, Wilhelm Eng Wei Han, Aswini Leela |
| hCoV-19/Montenegro/5782162102/2021 | EPI_ISL_7908402 | 2021-12-04 | Laboratory for Molecular Diagnostics,IPHMN | Eurofins Genomics Europe Sequencing GmbH | Danijela Vujo≈°evịá, Marija Govedarica |
| hCoV-19/England/PHWC-PF96W5/2021 | EPI_ISL_7995230 | 2021-12-06 | Originating lab: Wales Specialist Virology Centre Sequencing lab: Pathogen Genomics Unit | Public Health Wales Microbiology Cardiff Wales Specialist Virology Centre | Catherine Moore, Johnathan Evans, Laura Gifford, Malorie Perry, Simon Cottrell, Angela Marchbank, Alec Birchley, Alexander Adams, Amy Gaskin, Bree Gatica-Wilcox, Jason Coombes, Joel Southgate, Lauren Gilbert, Lee Graham, Nicole Pacchiarini, Sara Kumziene- |
| hCoV-19/PapuaNewGuinea/PNG1986/2021 | EPI_ISL_7737646 | 2021-05-07 | Queensland Medical Laboratories | Microbiological Diagnostic Unit - Public Health Laboratory (MDU-PHL) | Susapu, M., Palou, T., Seemann, T., Horan, K., Sherry, N.L. |
| hCoV-19/England/MILK-2EA3733/2021 | EPI_ISL_7974761 | 2021-12-14 | Lighthouse Lab in Milton Keynes | Wellcome Sanger Institute for the COVID-19 Genomics UK (COG-UK) Consortium | The Lighthouse Lab in Milton Keynes and Alex Alderton, Roberto Amato, Jeffrey Barrett, Sonia Goncalves, Ewan Harrison, David K. Jackson, Ian Johnston, Dominic Kwiatkowski, Cordelia Langford, John Sillitoe on behalf of the Wellcome Sanger Institute COVID-1 |
| hCoV-19/France/CVL-HMN-21122150386/2021 | EPI_ISL_7976495 | 2021-12-06 | BPR Analyses sp√©cialis√©es | Mondor University Hospital, Assistance Publique H√¥pitaux de Paris, Universit√© Paris-Est Cr√©teil, INSERM U955 | Christophe Rodriguez, Slim Fourati, Vanessa Demontant, Guillaume Gricourt, Melissa N'Debi, Alexandre Soulier, Elisabeth Trawinski, Jean-Michel Pawlotsky |
| hCoV-19/Jamaica/109733/2021 | EPI_ISL_7978306 | 2021-08-03 | National Public Health Laboratory | CARPHA | SueMin Nathaniel on behalf of CARPHA |
| hCoV-19/USA/WV-CDC-IBX447204893048/2021 | EPI_ISL_7979573 | 2021-12-03 | Infinity Biologix | Centers for Disease Control and Prevention Division of Viral Diseases, Pathogen Discovery | Dakota Howard,Dhwani Batra,Peter Cook,Jason Caravas,Benjamin Rambo-Martin,Scott Sammons,Yvette Unoarumhi,Matthew Schmerer,Kristine Lacek,Tymeckia Kendall,Victoria Caban Figueroa,Shatavia Morrison,Christopher Gulvick,Erisa Sula,Christian Bixby,Yihe Wang,Jo |
| hCoV-19/USA/MA-CDCBI-CRSP_U2GLDZBCKE5CEOZO/2021 | EPI_ISL_7980796 | 2021-12-07 | Broad Institute Clinical Research Sequencing Platform | Infectious Disease Program, Broad Institute of Harvard and MIT | Siddle,K.J., Adams,G., Pearlman,L., Gladden-Young,A., Vicente,G., Blumenstiel,B., DeFelice,M., Lee,M., McGovern,S., Lagerborg,K., Rudy,M., DeRuff,K., Carter,A., Normandin,E., Bauer,M., Reilly,S., Tomkins-Tinch,C., Loreth,C., Chaluvadi,S., Meldrim,J., Gran |
| hCoV-19/India/MH-KasturbaMCGM-ICMR-INSACOG-WG1630/2021 | EPI_ISL_7986400 | 2021-11-24 | Kasturba Hospital Molecular Lab - INSACOG | Kasturba hospital WGS | Jayanthi Shastri, Vidushi Chitalia |
| hCoV-19/India/MH-KasturbaMCGM-ICMR-INSACOG-WG1683/2021 | EPI_ISL_7986450 | 2021-12-01 | Kasturba Hospital Molecular Lab - INSACOG | Kasturba hospital WGS | Jayanthi Shastri, Vidushi Chitalia |

| | | | | | |
|---|---|---|---|---|---|
| hCoV-19/India/MH-KasturbaMCGM-ICMR-INSACOG-WG1685/2021 | EPI_ISL_7986452 | 2021-12-01 | Kasturba Hospital Molecular Lab - INSACOG | Kasturba hospital WGS | Jayanthi Shastri, Vidushi Chitalia |
| hCoV-19/India/MH-KasturbaMCGM-ICMR-INSACOG-WG1887/2021 | EPI_ISL_7986633 | 2021-11-26 | Kasturba Hospital Molecular Lab - INSACOG | Kasturba hospital WGS | Jayanthi Shastri, Vidushi Chitalia |
| hCoV-19/India/MH-KasturbaMCGM-ICMR-INSACOG-WG1888/2021 | EPI_ISL_7986634 | 2021-11-26 | Kasturba Hospital Molecular Lab - INSACOG | Kasturba hospital WGS | Jayanthi Shastri, Vidushi Chitalia |
| hCoV-19/India/MH-KasturbaMCGM-ICMR-INSACOG-WG1908/2021 | EPI_ISL_7986649 | 2021-11-22 | Kasturba Hospital Molecular Lab - INSACOG | Kasturba hospital WGS | Jayanthi Shastri, Vidushi Chitalia |
| hCoV-19/Australia/NSW-ICPMR-17660/2021 | EPI_ISL_7987484 | 2021-12-12 | Histopath | Clinical Pathology and Medical Research; Westmead Hospital; University of Sydney | Arnott A., Draper J., Gall M., Martinez E., Rockett R., Sintchenko V., on behalf of ICPMR |
| hCoV-19/Australia/NSW-ICPMR-17825/2021 | EPI_ISL_7987649 | 2021-12-07 | Histopath | Clinical Pathology and Medical Research; Westmead Hospital; University of Sydney | Arnott A., Draper J., Gall M., Martinez E., Rockett R., Sintchenko V., on behalf of ICPMR |
| hCoV-19/Australia/NSW-ICPMR-17830/2021 | EPI_ISL_7987654 | 2021-12-10 | Histopath | Clinical Pathology and Medical Research; Westmead Hospital; University of Sydney | Arnott A., Draper J., Gall M., Martinez E., Rockett R., Sintchenko V., on behalf of ICPMR |
| hCoV-19/Australia/NSW-ICPMR-18046/2021 | EPI_ISL_7987870 | 2021-12-15 | 4Cyte Pathology | Clinical Pathology and Medical Research; Westmead Hospital; University of Sydney | Arnott A., Draper J., Gall M., Martinez E., Rockett R., Sintchenko V., on behalf of ICPMR |
| hCoV-19/Australia/NSW-ICPMR-18135/2021 | EPI_ISL_7987959 | 2021-12-15 | Medlab Pathology | Clinical Pathology and Medical Research; Westmead Hospital; University of Sydney | Arnott A., Draper J., Gall M., Martinez E., Rockett R., Sintchenko V., on behalf of ICPMR |
| hCoV-19/Australia/NSW-ICPMR-18219/2021 | EPI_ISL_7988043 | 2021-12-14 | 4Cyte Pathology | Clinical Pathology and Medical Research; Westmead Hospital; University of Sydney | Arnott A., Draper J., Gall M., Martinez E., Rockett R., Sintchenko V., on behalf of ICPMR |
| hCoV-19/Zambia/ZMB-128868/2021 | EPI_ISL_7988169 | 2021-12-04 | University of Zambia, School of Veterinary Medicine | UNZAVET and PATH | Mulenga Mwenda-Chimfwembe, Ngonda Saasa, Daniel Bridges, ZNPHI and ZGSC |
| hCoV-19/Israel/ICH-741105997/2021 | EPI_ISL_7988857 | 2021-12-15 | Clinical Microbiology Laboratory, Tel Aviv Sourasky Medical Center | Clinical Microbiology Laboratory, Tel Aviv Sourasky Medical Center | Katya Levytskyi, Ora Halutz, Alon Ziv, Lior Handler, Amos Adler |
| hCoV-19/Israel/ICH-741106107/2021 | EPI_ISL_7988871 | 2021-12-13 | Clinical Microbiology Laboratory, Tel Aviv Sourasky Medical Center | Clinical Microbiology Laboratory, Tel Aviv Sourasky Medical Center | Katya Levytskyi, Ora Halutz, Alon Ziv, Lior Handler, Amos Adler |
| hCoV-19/Israel/ICH-741105778/2021 | EPI_ISL_7988990 | 2021-12-14 | Clinical Microbiology Laboratory, Tel Aviv Sourasky Medical Center | Clinical Microbiology Laboratory, Tel Aviv Sourasky Medical Center | Katya Levytskyi, Ora Halutz, Alon Ziv, Lior Handler, Amos Adler |
| hCoV-19/Israel/ICH-741105792/2021 | EPI_ISL_7988992 | 2021-12-15 | Clinical Microbiology Laboratory, Tel Aviv Sourasky Medical Center | Clinical Microbiology Laboratory, Tel Aviv Sourasky Medical Center | Katya Levytskyi, Ora Halutz, Alon Ziv, Lior Handler, Amos Adler |
| hCoV-19/Sweden/10208766/2021 | EPI_ISL_7873578 | 2021-11-30 | Swedish national genomic surveillance program of SARS-CoV-2 | The Public Health Agency of Sweden | Maximilian Riess, Maria Lind Karlberg, Alma Brolund, Swedish national genomic surveillance program of SARS-CoV-2 |
| hCoV-19/Israel/ICH-741107385/2021 | EPI_ISL_8177156 | 2021-12-20 | Clinical Microbiology Laboratory, Tel Aviv Sourasky Medical Center | Clinical Microbiology Laboratory, Tel Aviv Sourasky Medical Center | Katya Levytskyi, Ora Halutz, Alon Ziv, Lior Handler, Amos Adler |

| | | | | | |
|---|---|---|---|---|---|
| hCoV-19/India/OR-ILSGS08861/2021 | EPI_ISL_7877556 | 2021-07-28 | Angul, DHH | Institute of Life Sciences-INSACOG | Sunil K. Raghav, Safal Walia, Arup Ghosh, Atimukta Jha, Shifu Aggarwal, Amiyaranjan Das, Sana Fatma, Punyatoya Panda, Rupesh Dash, Rajeeb Swain, Punit Prasad, INSACOG Consortium, Ajay Parida |
| hCoV-19/CotedIvoire/COV-BKE8587-bc14/2021 | EPI_ISL_8178292 | 2021-10-21 | Molecular diagnostic unit for viral haemorrhagic fevers and emerging viruses, Bouak√© CHU Laboratory | Molecular diagnostic unit for viral haemorrhagic fevers and emerging viruses, Bouak√© CHU Laboratory | Chantal Akoua-Koffi, Dian√© Bamourou, Etil√© Anoh, Oby Wayoro, Safiatou Karidioula, Adjaratou Traor√©, Sound√©l√© Ma√Øt√©, Monemo Pacome, Coulibaly Mbegnan, Bamba Fatoumata Tour√©, Kra Ouffou√©, Jasmin Schlotterbeck, Grit Schubert, Essia Belarbi, Fabian L |
| hCoV-19/SouthAfrica/NHLS-UCT-GP-M449/2021 | EPI_ISL_7882975 | 2021-11-12 | Helderberg Hospital wc HHH | NHLS/UCT | Arash Iranzadeh, Deelan Doolabh, Lynn Tyers, Bruna Galvao, Innocent Mudau, Marvin Hsiao, Gert Marais, Diana Hardie, Stephen Korsman, Rageema Joseph, Luicer Olubayo, Nokuzola Mbhele, Carolyn Williamson |
| hCoV-19/Canada/AB-ABPHL-44775/2021 | EPI_ISL_7883388 | 2021-11-30 | Edmonton Provincial Lab | Alberta Precision Labs (APL) | Buss, E, Croxen M, Deo A, Dieu P, Gill K, Ferrato C, Khan F, Koleva P, Li V, Lloyd C, Lynch T, Ma R, Murphy S, Pabbaraju K, Shokoples S, Tipples G, Thayer J, Whitehouse M, Wong A, Yu C, Zelyas N |
| hCoV-19/SouthAfrica/NHLS-UCT-GS-AF55/2021 | EPI_ISL_7883621 | 2021-11-30 | 2 Military Hospital wc MAA | NHLS/UCT | Arash Iranzadeh, Deelan Doolabh, Lynn Tyers, Bruna Galvao, Innocent Mudau, Marvin Hsiao, Gert Marais, Diana Hardie, Stephen Korsman, Rageema Joseph, Luicer Olubayo, Nokuzola Mbhele, Carolyn Williamson |
| hCoV-19/CzechRepublic/IMG30S_7257/21 | EPI_ISL_7883753 | 2021-05-17 | National Institute of Public Health | Institute of Molecular Genetics CAS | Jan PacÃåes, MilusÃåe HradilovaÃÅ, Jana ≈†√°chov√°, Martin Zmuda, OndrÃåej MoravcÃåiÃÅk, Lucie PfeiferovaÃÅ, Michal KolaÃÅrÃå |
| hCoV-19/Japan/NC929-1N/2021 | EPI_ISL_7890636 | 2021-11-27 | Institute of Medical Science, University of Tokyo | Center for Influenza and Respiratory Virus Research, National Institute of Infectious Diseases (NIID) | Seiichiro Fujisaki, Hideka Miura, Emi Takashita, Seiya Yamayoshi, Masaki Imai, Yoshihiro Kawaoka |
| hCoV-19/India/GJ-INSACOG-GBRC3272/2021 | EPI_ISL_7897814 | 2021-12-08 | Invitro Speciality laboratory, Ahmedabad | Gujarat Biotechnology Research Centre | Roshani Mishra,Disha Vora,Janvi Raval,Apurvasinh Puvar,Zarna Patel,Bhadreshsinh Gohil,Nimesh Patel,Nitin Shukla,Ramesh Pandit,Nitin Savaliya,Priyank Chavda,Sonal Sharma ,Viral Pathak,Chaitanya Joshi,Madhvi Joshi |
| hCoV-19/England/MILK-2FBC51E/2021 | EPI_ISL_8244891 | 2021-12-21 | Lighthouse Lab in Milton Keynes | Wellcome Sanger Institute for the COVID-19 Genomics UK (COG-UK) Consortium | The Lighthouse Lab in Milton Keynes and Alex Alderton, Roberto Amato, Jeffrey Barrett, Sonia Goncalves, Ewan Harrison, David K. Jackson, Ian Johnston, Dominic Kwiatkowski, Cordelia Langford, John Sillitoe on behalf of the Wellcome Sanger Institute COVID-1 |
| hCoV-19/Switzerland/un-HUG-36378398/2021 | EPI_ISL_7995707 | 2021-12-13 | University Hospitals of Geneva, Laboratory of Virology | HUG, Laboratory of Virology and the Health2030 Genome Center | Samuel Cordey, Aline Mamin, Francisco Perez, Ana Rita Goncalves, Laurent Kaiser, Lorenzo Cerutti, Cedric Howald, Henri Pegeot, Melyssa Elies, Deborah Penet, Keith Harshman, Ioannis Xenarios |

| | | | | | |
|---|---|---|---|---|---|
| hCoV-19/USA/NY-MSHSPSP-PV41873/2021 | EPI_ISL_7908114 | 2021-11-11 | MSHS Clinical Microbiology Laboratories | MSHS Pathogen Surveillance Program | Ana S. Gonzalez-Reiche, Hala Alshammary, Mitchell J. Sullivan, Brianne Ciferri, Jose Polanco, Angela Amoako, Mahmoud Awawda, Aria Rooker, Christian Cognigni, Daniel Floda, Adriana van de Guchte, Zain Khalil, Keith Farrugia, Bremy Alburquerque, Julia Matth |
| hCoV-19/Denmark/DCGC-272583/2021 | EPI_ISL_7908618 | 2021-12-05 | Department of Bacteria, Parasites and Fungi, Statens Serum Institut, Copenhagen, Denmark | Statens Serum Institut Bioinformatics and Microbial Genomics | Danish Covid-19 Genome Consortium |
| hCoV-19/England/LOND-12916A3/2021 | EPI_ISL_8043523 | 2021-11-22 | Barts Health NHS Trust | COVID-19 Genomics UK (COG-UK) Consortium | CUTINO-MOGUEL, Maria-Teresa; HARRINGTON, David; HARRIS, Kathryn; SULTANA, Shahiba; KELE, Beatrix |
| hCoV-19/Slovenia/514005/2021 | EPI_ISL_7997055 | 2021-11-18 | Institute of Microbiology and Immunology, Faculty of Medicine, University of Ljubljana | Institute of Microbiology and Immunology, Faculty of Medicine, University of Ljubljana | Alen Suljiƒç, Samo Zakotnik, Toma≈æ Mark Zorec, Doroteja Vlaj, Andra≈æ Celar, Tina ≈Ωiviƒç, Domen Lazar, Tina Gabrov≈°ek, Patricija Pozvek, ≈†pela Pleh, Mi≈°a Korva, Mario Poljak, Tatjana Av≈°iƒç ,Äì ≈Ωupanc |
| hCoV-19/Denmark/DCGC-280675/2021 | EPI_ISL_8000566 | 2021-12-08 | Department of Bacteria, Parasites and Fungi, Statens Serum Institut, Copenhagen, Denmark | Statens Serum Institut Bioinformatics and Microbial Genomics | Danish Covid-19 Genome Consortium |
| hCoV-19/Nigeria/NCDC-NR1420/2021 | EPI_ISL_8189684 | 2021-11-30 | CENTO LAB | National Reference Laboratory, Nigeria Centre for Disease Control | Dr Ndodo Nnaemeka, Olusola Anuoluwapo Akanbi, Chimaobi Chukwu, Oyeronke Ayansola, Dr Omoare Adesuyi, Anthony Ahumibe, Catherine Okoi, Nwando Mba, Dr Ifedayo Adetifa |
| hCoV-19/Brazil/SC-FIOCRUZ-61479/2021 | EPI_ISL_8004423 | 2021-11-19 | Laboratorio Central de Saude Publica do Estado de Santa Catarina (LACEN/SC) | Laboratory of Respiratory Viruses and Measles, Oswaldo Cruz Institute, FIOCRUZ | Paola Resende, Luciana Appolinario, Fernando Motta, Elisa Cavalcante Pereira, Ighor Arantes, Alice Sampaio Rocha, Bruna Mendon√ßa da Silva, J√©ssica Gra√ßa Macedo de Carvalho, Larissa Macedo Pinto, Victor Guimaraes, Aline Moreira, Thiago Estevam Parente, |
| hCoV-19/Italy/SIC_CQRC_2983957/2021 | EPI_ISL_8002717 | 2021-12-14 | Laboratorio CQRC | CQRC_QUALITY CONTROL CHEMICAL BIOLOGICAL RISK_AOOR Villa Sofia Cervello Palermo | Di Gaudio F., Contino F., Brunacci G. |
| hCoV-19/Brazil/PR-FIOCRUZ-59616/2021 | EPI_ISL_8005513 | 2021-03-18 | Laboratorio Central de Saude Publica do Estado do Parana (LACEN/PR) | Laboratory of Respiratory Viruses and Measles, Oswaldo Cruz Institute, FIOCRUZ | Paola Resende, Luciana Appolinario, Fernando Motta, Elisa Cavalcante Pereira, Ighor Arantes, Alice Sampaio Rocha, Bruna Mendon√ßa da Silva, J√©ssica Gra√ßa Macedo de Carvalho, Larissa Macedo Pinto, Victor Guimaraes, Irina Riediger, Aline Moreira, Thiago E |
| hCoV-19/USA/TX-CDC-QDX31730208/2021 | EPI_ISL_8008406 | 2021-12-03 | Quest Diagnostics Incorporated | Centers for Disease Control and Prevention Division of Viral Diseases, Pathogen Discovery | Dakota Howard,Dhwani Batra,Peter Cook,Jason Caravas,Benjamin Rambo-Martin,Scott Sammons,Yvette Unoarumhi,Matthew Schmerer,Kristine Lacek,Tymeckia Kendall,Victoria Caban Figueroa,Shatavia Morrison,Christopher Gulvick,Erisa Sula,S. Rosenthal,A. Gerasimova,R |
| hCoV-19/USA/CA-CDPH-500034920/2021 | EPI_ISL_8246683 | 2021-11-22 | TPMG Regional Laboratory | California Department of Public Health | CDPH-COVIDNet and UCLA Technology Center for Genomics & Bioinformatics |

| | | | | | |
|---|---|---|---|---|---|
| hCoV-19/England/MILK-2FA683C/2021 | EPI_ISL_8195836 | 2021-12-20 | Lighthouse Lab in Milton Keynes | Wellcome Sanger Institute for the COVID-19 Genomics UK (COG-UK) Consortium | The Lighthouse Lab in Milton Keynes and Alex Alderton, Roberto Amato, Jeffrey Barrett, Sonia Goncalves, Ewan Harrison, David K. Jackson, Ian Johnston, Dominic Kwiatkowski, Cordelia Langford, John Sillitoe on behalf of the Wellcome Sanger Institute COVID-1 |
| hCoV-19/USA/MI-CDC-ASC210659531/2021 | EPI_ISL_8011358 | 2021-12-01 | Aegis Sciences Corporation | Centers for Disease Control and Prevention Division of Viral Diseases, Pathogen Discovery | Dakota Howard,Dhwani Batra,Peter Cook,Jason Caravas,Benjamin Rambo-Martin,Scott Sammons,Yvette Unoarumhi,Matthew Schmerer,Kristine Lacek,Tymeckia Kendall,Victoria Caban Figueroa,Shatavia Morrison,Christopher Gulvick,Erisa Sula,Cyndi Clark,Patrick Campbell |
| hCoV-19/Ecuador/NIC-INSPI-61237/2021 | EPI_ISL_8011680 | 2021-12-11 | INSPI-CRN DE INFLUENZA Y OTROS VIRUS RESPIRATORIOS | INSPI-CRN DE INFLUENZA Y OTROS VIRUS RESPIRATORIOS | Alfredo Bruno, Maritza Olmedo, Michelle P√°ez, Jimmy Garc√©s, Maria Angelica Becerra, Daniel Ramos, Fernando Llerena, Lizbeth Pati√±o, Diego Morales, Ruben Armas, Gulnara Borja |
| hCoV-19/USA/IL-CDC-ASC210606570/2021 | EPI_ISL_8012935 | 2021-12-04 | Aegis Sciences Corporation | Centers for Disease Control and Prevention Division of Viral Diseases, Pathogen Discovery | Dakota Howard,Dhwani Batra,Peter Cook,Jason Caravas,Benjamin Rambo-Martin,Scott Sammons,Yvette Unoarumhi,Matthew Schmerer,Kristine Lacek,Tymeckia Kendall,Victoria Caban Figueroa,Shatavia Morrison,Christopher Gulvick,Erisa Sula,Cyndi Clark,Patrick Campbell |
| hCoV-19/Chile/MA-CADIUMAG-01823/2021 | EPI_ISL_8014324 | 2021-12-03 | Laboratorio de Biolog√≠a Molecular, Universidad de Magallanes | Investigacion, Universidad de Magallanes | Constanza Ceroni, Diego Alvarez, Jorge Gonz√°lez, Jacqueline Aldridge, Marcelo Navarrete |
| hCoV-19/Denmark/DCGC-288640/2021 | EPI_ISL_8249540 | 2021-12-20 | Department of Bacteria, Parasites and Fungi, Statens Serum Institut, Copenhagen, Denmark | Statens Serum Institut Bioinformatics and Microbial Genomics | Danish Covid-19 Genome Consortium |
| hCoV-19/Senegal/IR1-0079927/2021 | EPI_ISL_7400617 | 2021-11-09 | Iressef Genomics lab | IRESSEF | Souleymane MBOUP, Abdou PADANE , Khadim GUEYE, Cyrille Diedhiou , Samba Ndiour, Diabou Diagne, Papa Alassane DIAW, Birahim Piere NDIAYE, Barada CISSE, Aminata MBOUP, Moustapha MBOW, Ndeye Coumba Toure KANE, Nafisatou LEYE, Gora LO, Ambroise AHOUIDI , Asto |
| hCoV-19/Curacao/CW-RIVM-75214/2021 | EPI_ISL_8015567 | 2021-11-29 | Dutch COVID-19 response team | National Institute for Public Health and the Environment (RIVM) | Adam Meijer, Harry Vennema, Dirk Eggink, Jeroen Cremer, Sharon van den Brink, Bas van der Veer, AnneMarie van den Brandt, Lisa Wijsman, Kim Freriks, Ryanne Jaarsma, Lynn Aarts, Sanne Bos, Jil Kocken, Jordy de Bakker, Stijn van Rossum, Linda van Someren, F |
| hCoV-19/Aruba/AW-RIVM-75240/2021 | EPI_ISL_8015547 | 2021-11-30 | Dutch COVID-19 response team | National Institute for Public Health and the Environment (RIVM) | Adam Meijer, Harry Vennema, Dirk Eggink, Jeroen Cremer, Sharon van den Brink, Bas van der Veer, AnneMarie van den Brandt, Lisa Wijsman, Kim Freriks, Ryanne Jaarsma, Lynn Aarts, Sanne Bos, Jil Kocken, Jordy de Bakker, Stijn van Rossum, Linda van Someren, F |

| | | | | | |
|---|---|---|---|---|---|
| hCoV-19/Curacao/CW-RIVM-75223/2021 | EPI_ISL_8015576 | 2021-12-03 | Dutch COVID-19 response team | National Institute for Public Health and the Environment (RIVM) | Adam Meijer, Harry Vennema, Dirk Eggink, Jeroen Cremer, Sharon van den Brink, Bas van der Veer, AnneMarie van den Brandt, Lisa Wijsman, Kim Freriks, Ryanne Jaarsma, Lynn Aarts, Sanne Bos, Jil Kocken, Jordy de Bakker, Stijn van Rossum, Linda van Someren, F |
| hCoV-19/Denmark/DCGC-288872/2021 | EPI_ISL_8249734 | 2021-12-22 | Department of Bacteria, Parasites and Fungi, Statens Serum Institut, Copenhagen, Denmark | Statens Serum Institut Bioinformatics and Microbial Genomics | Danish Covid-19 Genome Consortium |
| hCoV-19/USA/NY-CDC-FG-194357/2021 | EPI_ISL_8017348 | 2021-12-09 | Fulgent Genetics | Centers for Disease Control and Prevention Division of Viral Diseases, Pathogen Discovery | Dakota Howard,Dhwani Batra,Peter Cook,Jason Caravas,Benjamin Rambo-Martin,Scott Sammons,Yvette Unoarumhi,Matthew Schmerer,Kristine Lacek,Tymeckia Kendall,Victoria Caban Figueroa,Shatavia Morrison,Christopher Gulvick,Erisa Sula,Harry Gao,Mickey Li,John Gao |
| hCoV-19/Curacao/CW-RIVM-76481/2021 | EPI_ISL_8015601 | 2021-11-26 | Dutch COVID-19 response team | National Institute for Public Health and the Environment (RIVM) | Adam Meijer, Harry Vennema, Dirk Eggink, Jeroen Cremer, Sharon van den Brink, Bas van der Veer, AnneMarie van den Brandt, Lisa Wijsman, Kim Freriks, Ryanne Jaarsma, Lynn Aarts, Sanne Bos, Jil Kocken, Jordy de Bakker, Stijn van Rossum, Linda van Someren, F |
| hCoV-19/Curacao/CW-RIVM-76482/2021 | EPI_ISL_8015602 | 2021-11-26 | Dutch COVID-19 response team | National Institute for Public Health and the Environment (RIVM) | Adam Meijer, Harry Vennema, Dirk Eggink, Jeroen Cremer, Sharon van den Brink, Bas van der Veer, AnneMarie van den Brandt, Lisa Wijsman, Kim Freriks, Ryanne Jaarsma, Lynn Aarts, Sanne Bos, Jil Kocken, Jordy de Bakker, Stijn van Rossum, Linda van Someren, F |
| hCoV-19/Netherlands/GL-RIVM-75321/2021 | EPI_ISL_8015725 | 2021-11-28 | Dutch COVID-19 response team | National Institute for Public Health and the Environment (RIVM) | Adam Meijer, Harry Vennema, Dirk Eggink, Jeroen Cremer, Sharon van den Brink, Bas van der Veer, AnneMarie van den Brandt, Lisa Wijsman, Kim Freriks, Ryanne Jaarsma, Lynn Aarts, Sanne Bos, Jil Kocken, Jordy de Bakker, Stijn van Rossum, Linda van Someren, F |
| hCoV-19/England/ALDP-2F65268/2021 | EPI_ISL_8084721 | 2021-12-15 | Lighthouse Lab in Alderley Park | Wellcome Sanger Institute for the COVID-19 Genomics UK (COG-UK) Consortium | Jacquelyn Wynn, Mairead Hyland, The Lighthouse Lab in Alderley Park and Alex Alderton, Roberto Amato, Jeffrey Barrett, Sonia Goncalves, Ewan Harrison, David K. Jackson, Ian Johnston, Dominic Kwiatkowski, Cordelia Langford, John Sillitoe on behalf of the W |
| hCoV-19/Portugal/PT23553/2021 | EPI_ISL_8020761 | 2021-12-13 | ABC Algarve | Instituto Nacional de Saude (INSA) | Borges et al |
| hCoV-19/Netherlands/LB-RIVM-75402/2021 | EPI_ISL_8020985 | 2021-11-26 | Dutch COVID-19 response team | National Institute for Public Health and the Environment (RIVM) | Adam Meijer, Harry Vennema, Dirk Eggink, Jeroen Cremer, Sharon van den Brink, Bas van der Veer, AnneMarie van den Brandt, Lisa Wijsman, Kim Freriks, Ryanne Jaarsma, Lynn Aarts, Sanne Bos, Jil Kocken, Jordy de Bakker, Stijn van Rossum, Linda van Someren, F |
| hCoV-19/Denmark/DCGC-289815/2021 | EPI_ISL_8250626 | 2021-12-26 | Department of Bacteria, Parasites and Fungi, Statens Serum Institut, Copenhagen, Denmark | Statens Serum Institut Bioinformatics and Microbial Genomics | Danish Covid-19 Genome Consortium |

| | | | | | |
|---|---|---|---|---|---|
| hCoV-19/USA/NM-CDC-LC0422811/2021 | EPI_ISL_8024555 | 2021-12-03 | Laboratory Corporation of America | Centers for Disease Control and Prevention Division of Viral Diseases, Pathogen Discovery | Dakota Howard,Dhwani Batra,Peter Cook,Jason Caravas,Benjamin Rambo-Martin,Scott Sammons,Yvette Unoarumhi,Matthew Schmerer,Kristine Lacek,Tymeckia Kendall,Victoria Caban Figueroa,Shatavia Morrison,Christopher Gulvick,Minoo Agarwal,Eyad Almasri,Debbie Boles |
| hCoV-19/Denmark/DCGC-290226/2021 | EPI_ISL_8251029 | 2021-12-13 | Department of Bacteria, Parasites and Fungi, Statens Serum Institut, Copenhagen, Denmark | Statens Serum Institut Bioinformatics and Microbial Genomics | Danish Covid-19 Genome Consortium |
| hCoV-19/Denmark/DCGC-290213/2021 | EPI_ISL_8251018 | 2021-12-20 | Department of Bacteria, Parasites and Fungi, Statens Serum Institut, Copenhagen, Denmark | Statens Serum Institut Bioinformatics and Microbial Genomics | Danish Covid-19 Genome Consortium |
| hCoV-19/England/BRBR-2F41578/2021 | EPI_ISL_8086564 | 2021-12-18 | Berkshire and Surrey Pathology Services Lighthouse Laboratory | Wellcome Sanger Institute for the COVID-19 Genomics UK (COG-UK) Consortium | Berkshire and Surrey Pathology Services Lighthouse Laboratory and Alex Alderton, Roberto Amato, Jeffrey Barrett, Sonia Goncalves, Ewan Harrison, David K. Jackson, Ian Johnston, Dominic Kwiatkowski, Cordelia Langford, John Sillitoe on behalf of the Wellcom |
| hCoV-19/England/BRBR-2F42BC0/2021 | EPI_ISL_8086611 | 2021-12-16 | Berkshire and Surrey Pathology Services Lighthouse Laboratory | Wellcome Sanger Institute for the COVID-19 Genomics UK (COG-UK) Consortium | Berkshire and Surrey Pathology Services Lighthouse Laboratory and Alex Alderton, Roberto Amato, Jeffrey Barrett, Sonia Goncalves, Ewan Harrison, David K. Jackson, Ian Johnston, Dominic Kwiatkowski, Cordelia Langford, John Sillitoe on behalf of the Wellcom |
| hCoV-19/Brazil/BA-LACEN-BA948-291725521/2021 | EPI_ISL_8200859 | 2021-10-01 | Central Public Health Laboratory - LACEN - Bahia, Salvador, Brazil | Central Public Health Laboratory - LACEN -Bahia, Salvador, Brazil | Stephane Tosta, Luciana Oliveira, Vanessa Nardy, Gabriela Menezes,Marcela G√≥mez, Lenisa Dandara, Jaqueline Gomes, Vagner Fonseca,Marta Giovanetti,Luiz Alcantara, Felicidade Pereira, Arabela Leal |
| hCoV-19/England/MILK-2F52B27/2021 | EPI_ISL_8086983 | 2021-12-18 | Lighthouse Lab in Milton Keynes | Wellcome Sanger Institute for the COVID-19 Genomics UK (COG-UK) Consortium | The Lighthouse Lab in Milton Keynes and Alex Alderton, Roberto Amato, Jeffrey Barrett, Sonia Goncalves, Ewan Harrison, David K. Jackson, Ian Johnston, Dominic Kwiatkowski, Cordelia Langford, John Sillitoe on behalf of the Wellcome Sanger Institute COVID-1 |
| hCoV-19/USA/VA-CDC-LC0421707/2021 | EPI_ISL_8026992 | 2021-12-08 | Laboratory Corporation of America | Centers for Disease Control and Prevention Division of Viral Diseases, Pathogen Discovery | Dakota Howard,Dhwani Batra,Peter Cook,Jason Caravas,Benjamin Rambo-Martin,Scott Sammons,Yvette Unoarumhi,Matthew Schmerer,Kristine Lacek,Tymeckia Kendall,Victoria Caban Figueroa,Shatavia Morrison,Christopher Gulvick,Minoo Agarwal,Eyad Almasri,Debbie Boles |
| hCoV-19/Canada/ON-PHL-21-45810/2021 | EPI_ISL_8027342 | 2021-11-09 | Public Health Ontario Laboratory | Public Health Ontario Laboratory | Vanessa G Allen, Philip Banh, Yao Chen, Richard de Borja, Alireza Eshaghi, Nahuel Fittipaldi, Christine Frantz, Jonathan B Gubbay, Jennifer L Guthrie, Lawrence Heisler, Esha Joshi, Michael Laszloffy, Aimin Li, Michael CY Li, Dean Maxwell, Sandeep Nagra, S |

| | | | | | |
|---|---|---|---|---|---|
| hCoV-19/England/MILK-2F9F9BB/2021 | EPI_ISL_8193713 | 2021-12-20 | Lighthouse Lab in Milton Keynes | Wellcome Sanger Institute for the COVID-19 Genomics UK (COG-UK) Consortium | The Lighthouse Lab in Milton Keynes and Alex Alderton, Roberto Amato, Jeffrey Barrett, Sonia Goncalves, Ewan Harrison, David K. Jackson, Ian Johnston, Dominic Kwiatkowski, Cordelia Langford, John Sillitoe on behalf of the Wellcome Sanger Institute COVID-1 |
| hCoV-19/India/MH-RFIP06378/2021 | EPI_ISL_8252227 | 2021-12-15 | BJMC, Pune | RF-IISER Pune | Krishanpal Karmodiya, Aurnab Ghose, Joy Merwin Monteiro, Suvarna Joshi, Rajesh Karyakarte |
| hCoV-19/Argentina/PAIS-A1256/2021 | EPI_ISL_8203668 | 2021-12-16 | Laboratorio de Virologia del Hospital de Ni√±os Dr Ricardo Gutierrez | Laboratorio de Virolog√≠a del Hospital de Ni√±os Dr. Ricardo Gutierrez on behalf of 'Proyecto Argentino Interinstitucional de genomica de SARS-CoV-2' (PAIS | Alicia Mistchenko; Erica Grandis; Mar√≠a Cristina Alvarez L√≥pez; Mar√≠a Elina Acevedo; Oscar Jacquez; Sof√≠a Alexay; Mari√°ngeles Barreda Fink; Mar√≠a Emilia Villegas; Raquel Barquez; Estela Chasc√≥n; Jorgelina Caruso; Mariana Campal; Karina Zacar√≠as; C |
| hCoV-19/India/MH-RFIP06117/2021 | EPI_ISL_8252322 | 2021-01-11 | BJMC, Pune | RF-IISER Pune | Krishanpal Karmodiya, Aurnab Ghose, Joy Merwin Monteiro, Suvarna Joshi, Rajesh Karyakarte |
| hCoV-19/India/MH-RFIP06262/2021 | EPI_ISL_8252264 | 2021-11-07 | BJMC, Pune | RF-IISER Pune | Krishanpal Karmodiya, Aurnab Ghose, Joy Merwin Monteiro, Suvarna Joshi, Rajesh Karyakarte |
| hCoV-19/India/MH-RFIP06121/2021 | EPI_ISL_8252257 | 2021-01-11 | BJMC, Pune | RF-IISER Pune | Krishanpal Karmodiya, Aurnab Ghose, Joy Merwin Monteiro, Suvarna Joshi, Rajesh Karyakarte |
| hCoV-19/India/MH-RFIP06205/2021 | EPI_ISL_8252281 | 2021-11-02 | BJMC, Pune | RF-IISER Pune | Krishanpal Karmodiya, Aurnab Ghose, Joy Merwin Monteiro, Suvarna Joshi, Rajesh Karyakarte |
| hCoV-19/Israel/ICH-760598309/2021 | EPI_ISL_7552269 | 2021-12-03 | Clinical Microbiology Laboratory, Tel Aviv Sourasky Medical Center | Clinical Microbiology Laboratory, Tel Aviv Sourasky Medical Center | Katya Levytskyi, Ora Halutz, Alon Ziv, Lior Handler, Amos Adler |
| hCoV-19/Sweden/1928635664/2021 | EPI_ISL_8253181 | 2021-12-20 | Sweden, Vastragotaland, Unilabs | Unilabs, Eskilstuna, Sweden | Emma Arvidsson |
| hCoV-19/Sweden/2003958664/2021 | EPI_ISL_8253256 | 2021-12-20 | Sweden, Vastragotaland, Unilabs | Unilabs, Eskilstuna, Sweden | Emma Arvidsson |
| hCoV-19/USA/MD-IGS-012133103687A/2021 | EPI_ISL_8204846 | 2021-11-28 | Maryland Genomics, Institute for Genome Sciences, University of Maryland School of Medicine | Maryland Genomics, Institute for Genome Sciences, University of Maryland School of Medicine | Tallon, Luke J; Sadzewicz, Lisa D; Humphrys, Mike; Ott, Sandra; Roussey, Holly; Vavikolanu, Kranthi; Regan, Kevin; George, Jain; Hazen, Tracy; Fraser, Claire M; Ravel, Jacques |
| hCoV-19/Sweden/2931103964/2021 | EPI_ISL_8253295 | 2021-12-20 | Sweden, Vastragotaland, Unilabs | Unilabs, Eskilstuna, Sweden | Emma Arvidsson |
| hCoV-19/Colombia/VAC-SUR-O1V31398CFN/2021 | EPI_ISL_8254204 | 2021-10-18 | SURA | CIAT, Laboratorio de Virologia | Ana M. Leiva, Diana Lopez-Alvarez, WilmerJ. Cuellar, Programa Nacional de Caracterizaci√≥n Gen√≥mica de SARS-CoV-33 |
| hCoV-19/Egypt/NRC-1323/2021 | EPI_ISL_8205828 | 2021-11-17 | Center of Scientific Excellence for Influenza Viruses (CSEIV), National Research Centre | Influenza Viruses (CSEIV), National Research Centre | Rabeh El-Shesheny, Yassmin Moatasim, Mina Kamel, Mokhtar Gomaa, Ahmed Kandeil, Mohamed Ahmed Ali |
| hCoV-19/Malaysia/TIDREC-NSC186/2021 | EPI_ISL_8090611 | 2021-09-07 | PEJABAT KESIHATAN DAERAH KUALA KRAI | Tropical Infectious Diseases Research & Education Centre (TIDREC), Universiti Malaya | Kim-Kee Tan, Jia-Yi Tan, Jo-Ern Wong, Siti-Sarah Nor‚Äôe, Che-Norainon Yaacob, Mulya-Mustika-Sari Zulkifli, AsmaAnati CheMatSeri, Nur-Hidayana Mahfodz, Noor Syahida Azizan, Sazaly AbuBakar |
| hCoV-19/Israel/ICH-741108519/2021 | EPI_ISL_8206800 | 2021-12-22 | Clinical Microbiology Laboratory, Tel Aviv Sourasky Medical Center | Clinical Microbiology Laboratory, Tel Aviv Sourasky Medical Center | Katya Levytskyi, Ora Halutz, Alon Ziv, Lior Handler, Amos Adler |
| hCoV-19/Israel/ICH-741108210/2021 | EPI_ISL_8206726 | 2021-12-20 | Clinical Microbiology Laboratory, Tel Aviv Sourasky Medical Center | Clinical Microbiology Laboratory, Tel Aviv Sourasky Medical Center | Katya Levytskyi, Ora Halutz, Alon Ziv, Lior Handler, Amos Adler |

| | | | | | |
|---|---|---|---|---|---|
| hCoV-19/Switzerland/VD-CHUV-GEN9435/2021 | EPI_ISL_8253861 | 2021-12-21 | CHUV | metagenomics, Institute of Microbiology, University Hospital Centre and University of Lausanne | Trestan Pillonel, Damien Jacot, Sv©bastien Aeby, Gilbert Greub, Claire Bertelli |
| hCoV-19/Israel/ICH-741108221/2021 | EPI_ISL_8206796 | 2021-12-21 | Clinical Microbiology Laboratory, Tel Aviv Sourasky Medical Center | Clinical Microbiology Laboratory, Tel Aviv Sourasky Medical Center | Katya Levytskyi, Ora Halutz, Alon Ziv, Lior Handler, Amos Adler |
| hCoV-19/Israel/ICH-741108547/2021 | EPI_ISL_8206802 | 2021-12-23 | Clinical Microbiology Laboratory, Tel Aviv Sourasky Medical Center | Clinical Microbiology Laboratory, Tel Aviv Sourasky Medical Center | Katya Levytskyi, Ora Halutz, Alon Ziv, Lior Handler, Amos Adler |
| hCoV-19/Israel/ICH-741108630/2021 | EPI_ISL_8206841 | 2021-12-22 | Clinical Microbiology Laboratory, Tel Aviv Sourasky Medical Center | Clinical Microbiology Laboratory, Tel Aviv Sourasky Medical Center | Katya Levytskyi, Ora Halutz, Alon Ziv, Lior Handler, Amos Adler |
| hCoV-19/FrenchGuiana/IPG202101535/2021 | EPI_ISL_8207145 | 2021-12-23 | CNR Institut Pasteur de la Guyane | Institut Pasteur de la Guyane | Anne Lavergne, Dominique Rousset, Antoine Enfissi |
| hCoV-19/SouthKorea/KDCA19608/2021 | EPI_ISL_7792650 | 2021-11-03 | Division of Emerging Infectious Diseases, Bureau of Infectious Diseases Diagnosis Control, Korea Disease Control and Prevention Agency | Diseases, Bureau of Infectious Diseases Diagnosis Control, Korea Disease Control and Prevention Agency | Ae Kyung Park, Il-Hwan Kim, Jeong-Ah Kim, Chae Young Lee, Hyuck Jin Lee, Eun-Jin Kim |
| hCoV-19/USA/CA-CDC-FG-200515/2021 | EPI_ISL_8091405 | 2021-12-17 | Fulgent Genetics | Centers for Disease Control and Prevention Division of Viral Diseases, Pathogen Discovery | Dakota Howard,Dhwani Batra,Peter Cook,Jason Caravas,Benjamin Rambo-Martin,Scott Sammons,Yvette Unoarumhi,Matthew Schmerer,Kristine Lacek,Tymeckia Kendall,Victoria Caban Figueroa,Shatavia Morrison,Christopher Gulvick,Erisa Sula,Harry Gao,Mickey Li,John Gao |
| hCoV-19/Argentina/PAIS-G0670/2021 | EPI_ISL_8207615 | 2021-12-13 | Laboratorio Central, Ministerio de Salud Cv≥rdoba | Medicina and Instituto de Patologia Vegetal (IPAVE-CIAP-INTA) on behalf of 'Proyecto Argentino Interinstitucional de genomica de SARS-CoV-2' (PAIS Consortium) | Zeballos, M.; Lucca, A.; Fernv°ndez, F.D.; Marquez, N.; Debat, H., Goya, S.; Re, V.; Pisano, M.B.; Castro, G.; Sicilia, P.; Barbas, G. |
| hCoV-19/Argentina/PAIS-G0588/2021 | EPI_ISL_8207542 | 2021-12-11 | Laboratorio Central, Ministerio de Salud Cv≥rdoba | Medicina and Instituto de Patologia Vegetal (IPAVE-CIAP-INTA) on behalf of 'Proyecto Argentino Interinstitucional de genomica de SARS-CoV-2' (PAIS Consortium) | Zeballos, M.; Lucca, A.; Fernv°ndez, F.D.; Marquez, N.; Debat, H., Goya, S.; Re, V.; Pisano, M.B.; Castro, G.; Sicilia, P.; Barbas, G. |
| hCoV-19/Argentina/PAIS-G0624/2021 | EPI_ISL_8207574 | 2021-12-13 | Laboratorio Central, Ministerio de Salud Cv≥rdoba | Medicina and Instituto de Patologia Vegetal (IPAVE-CIAP-INTA) on behalf of 'Proyecto Argentino Interinstitucional de genomica de SARS-CoV-2' (PAIS Consortium) | Zeballos, M.; Lucca, A.; Fernv°ndez, F.D.; Marquez, N.; Debat, H., Goya, S.; Re, V.; Pisano, M.B.; Castro, G.; Sicilia, P.; Barbas, G. |
| hCoV-19/Argentina/PAIS-G0630/2021 | EPI_ISL_8207580 | 2021-12-13 | Laboratorio Central, Ministerio de Salud Cv≥rdoba | Medicina and Instituto de Patologia Vegetal (IPAVE-CIAP-INTA) on behalf of 'Proyecto Argentino Interinstitucional de genomica de SARS-CoV-2' (PAIS Consortium) | Zeballos, M.; Lucca, A.; Fernv°ndez, F.D.; Marquez, N.; Debat, H., Goya, S.; Re, V.; Pisano, M.B.; Castro, G.; Sicilia, P.; Barbas, G. |
| hCoV-19/Colombia/BOL-UNIMOL-VG-203/2021 | EPI_ISL_8035471 | 2021-07-11 | UNIMOL | UNIMOL | Steev Loyola, Mashiel Fernv°ndez-Ruiz, Jaison Torres-Pacheco, Doris Gv≥mez-Camargo |

| | | | | | |
|---|---|---|---|---|---|
| hCoV-19/France/BFC-HMN-21122240669/2021 | EPI_ISL_8257111 | 2021-12-21 | SELAS MEDILYS | [illegible] Mondor University Hospital, Assistance Publique H√¥pitaux de Paris, Universit√© Paris-Est Cr√©teil, INSERM U955 | Christophe Rodriguez, Slim Fourati, Vanessa Demontant, Guillaume Gricourt, Melissa N'Debi, Alexandre Soulier, Elisabeth Trawinski, Jean-Michel Pawlotsky |
| hCoV-19/USA/NJ-GBW-EWR000003/2021 | EPI_ISL_8035614 | 2021-12-14 | Northeastern LSTC | Ginkgo Bioworks Clinical Laboratory / XpressCheck | Keith Robison, Dongjuan Dai, Brintha P. Girinathan, Allison L. Hicks, Bryan Cosca, Alex M. Plocik, Birgitte B. Simen |
| hCoV-19/Brazil/MG-FUNED-210101003972/2021 | EPI_ISL_7795735 | 2021-12-06 | Funda√ß√£o Ezequiel Dias (FUNED) | Funda√ß√£o Ezequiel Dias (FUNED) | Talita Adelino, Natalia Guimaraes, Felipe Iani, Andre Leal, Vagner Fonseca, Marta Giovanetti, Joilson Xavier, Luiz Alcantara, Glauco Carvalho. |
| hCoV-19/USA/NY-CDC-FG-190079/2021 | EPI_ISL_7801013 | 2021-11-30 | Fulgent Genetics | Centers for Disease Control and Prevention Division of Viral Diseases, Pathogen Discovery | Dakota Howard,Dhwani Batra,Peter Cook,Jason Caravas,Benjamin Rambo-Martin,Scott Sammons,Yvette Unoarumhi,Matthew Schmerer,Kristine Lacek,Tymeckia Kendall,Victoria Caban Figueroa,Shatavia Morrison,Christopher Gulvick,Erisa Sula,Harry Gao,Mickey Li,John Gao |
| hCoV-19/USA/UT-CDC-ASC210512152/2021 | EPI_ISL_7801204 | 2021-11-29 | Aegis Sciences Corporation | Centers for Disease Control and Prevention Division of Viral Diseases, Pathogen Discovery | Dakota Howard,Dhwani Batra,Peter Cook,Jason Caravas,Benjamin Rambo-Martin,Scott Sammons,Yvette Unoarumhi,Matthew Schmerer,Kristine Lacek,Tymeckia Kendall,Victoria Caban Figueroa,Shatavia Morrison,Christopher Gulvick,Erisa Sula,Cyndi Clark,Patrick Campbell |
| hCoV-19/USA/GA-CDC-STM-F436C64CG/2021 | EPI_ISL_7801174 | 2021-11-30 | Helix | Centers for Disease Control and Prevention Division of Viral Diseases, Pathogen Discovery | Dakota Howard,Dhwani Batra,Peter Cook,Jason Caravas,Benjamin Rambo-Martin,Scott Sammons,Yvette Unoarumhi,Matthew Schmerer,Kristine Lacek,Tymeckia Kendall,Victoria Caban Figueroa,Helix CA,Shatavia Morrison,Christopher Gulvick,Erisa Sula,Clinton Paden,Dunca |
| hCoV-19/SouthAfrica/NICD-N22540/2021 | EPI_ISL_7740828 | 2021-11-26 | HELEN JOSEPH LABORATORY | Diseases of the National Health Laboratory Service | Amoako DG, Everatt J, Scheepers C, Mohale T, Ntuli N, Mahlangu B, Mnguni A, Ismail A, Bhiman JN, Wolter N |
| hCoV-19/Germany/HE-RKI-I-356248/2021 | EPI_ISL_7418464 | 2021-11-30 | Universit√§tsklinikum Frankfurt - Institut f√ºr Medizinische Virologie | Robert Koch Institute | unknown |
| hCoV-19/Germany/BW-RKI-I-356316/2021 | EPI_ISL_7418593 | 2021-11-29 | Limbach - MVZ Humangenetik Ulm | Robert Koch Institute | unknown |
| hCoV-19/Germany/BY-RKI-I-363298/2021 | EPI_ISL_7439547 | 2021-11-29 | IMD - MVZ Labor Martinsried | Robert Koch Institute | unknown |
| hCoV-19/Germany/BY-RKI-I-363324/2021 | EPI_ISL_7439650 | 2021-11-29 | IMD - MVZ Labor Martinsried | Robert Koch Institute | unknown |
| hCoV-19/Germany/BY-RKI-I-363325/2021 | EPI_ISL_7439657 | 2021-11-29 | IMD - MVZ Labor Martinsried | Robert Koch Institute | unknown |
| hCoV-19/Germany/BY-RKI-I-363340/2021 | EPI_ISL_7439730 | 2021-11-29 | IMD - MVZ Labor Martinsried | Robert Koch Institute | unknown |
| hCoV-19/Germany/NW-RKI-I-363836/2021 | EPI_ISL_7441981 | 2021-12-02 | SYNLAB MVZ Leverkusen | Robert Koch Institute | unknown |
| hCoV-19/Germany/BB-RKI-I-364364/2021 | EPI_ISL_7443742 | 2021-12-04 | Klinikum Ernst von Bergmann gemeinn√ºtzige GmbH - station√§rer Bereich | Robert Koch Institute | unknown |

| | | | | | |
|---|---|---|---|---|---|
| hCoV-19/Germany/NW-RKI-I-365404/2021 | EPI_ISL_7748675 | 2021-12-03 | SYNLAB MVZ Leverkusen | Robert Koch Institute | unknown |
| hCoV-19/Germany/NW-RKI-I-367094/2021 | EPI_ISL_7749828 | 2021-12-04 | Labor Dr. Wisplinghoff - K√∂ln | Robert Koch Institute | unknown |
| hCoV-19/Germany/HE-RKI-I-368792/2021 | EPI_ISL_7751329 | 2021-12-04 | CENTOGENE Frankfurt Laboratory: Niederlassung Industriepark H√∂chst | Robert Koch Institute | unknown |
| hCoV-19/Germany/MV-RKI-I-376956/2021 | EPI_ISL_7761036 | 2021-01-01 | IMD - Medizinisches Labor Rostock | Robert Koch Institute | unknown |
| hCoV-19/Germany/NW-RKI-I-378079/2021 | EPI_ISL_7763748 | 2021-12-08 | Labor Dr. Wisplinghoff - K√∂ln | Robert Koch Institute | unknown |
| hCoV-19/Germany/NW-RKI-I-382293/2021 | EPI_ISL_7770121 | 2021-12-16 | Eurofins MVZ Labor Gelsenkirchen | Robert Koch Institute | unknown |
| hCoV-19/Germany/SH-RKI-I-396948/2021 | EPI_ISL_8226499 | 2021-12-13 | LADR Zentrallabor DR. Kramer & Kollegen Geesthacht | Robert Koch Institute | unknown |
| hCoV-19/Germany/HE-RKI-I-397896/2021 | EPI_ISL_8227383 | 2021-12-15 | CENTOGENE Frankfurt Laboratory: Niederlassung Industriepark H√∂chst | Robert Koch Institute | unknown |
| hCoV-19/Germany/HE-RKI-I-397997/2021 | EPI_ISL_8227482 | 2021-12-17 | CENTOGENE Frankfurt Laboratory: Niederlassung Industriepark H√∂chst | Robert Koch Institute | unknown |
| hCoV-19/Germany/BW-RKI-I-401596/2021 | EPI_ISL_8232369 | 2021-12-20 | SYNLAB MVZ Leinfelden-Echterdingen | Robert Koch Institute | unknown |
| hCoV-19/Germany/BW-RKI-I-410179/2021 | EPI_ISL_8248539 | 2021-12-21 | MVZ Labor Dr. Limbach & Kollegen GbR | Robert Koch Institute | unknown |
| hCoV-19/Israel/ICH-760598358/2021 | EPI_ISL_7552311 | 2021-12-02 | Clinical Microbiology Laboratory, Tel Aviv Sourasky Medical Center | Clinical Microbiology Laboratory, Tel Aviv Sourasky Medical Center | Katya Levytskyi, Ora Halutz, Alon Ziv, Lior Handler, Amos Adler |
| hCoV-19/Botswana/R50B78_BHP_AAC33163/2021 | EPI_ISL_7552710 | 2021-12-07 | National Health Laboratory | Botswana Harvard AIDS Institute Partnership, Plot 1836 North Ring Road, Princess Marina Hospital, Gaborone | Sikhulile Moyo, Wonderful T. Choga, Ontlametse T. Bareng, Doreen Ditshwanelo, Dorcas Maruapula, Keoratile Ntshambiwa, Sefetogi Ramaologa, Thongbotho Mphoyakgosi, Boitumelo Zuze, Botshelo Radibe, Legodile Kooepile, Pamela Smith-Lawrence, Kgomotso Moruisi, |
| hCoV-19/USA/MA-CDCBI-CRSP_2E4FHYG2P3AUVWQ2/2021 | EPI_ISL_7981605 | 2021-12-10 | Broad Institute Clinical Research Sequencing Platform | Infectious Disease Program, Broad Institute of Harvard and MIT | Siddle,K.J., Adams,G., Pearlman,L., Gladden-Young,A., Vicente,G., Blumenstiel,B., DeFelice,M., Lee,M., McGovern,S., Lagerborg,K., Rudy,M., DeRuff,K., Carter,A., Normandin,E., Bauer,M., Reilly,S., Tomkins-Tinch,C., Loreth,C., Chaluvadi,S., Meldrim,J., Gran |
| hCoV-19/Indonesia/BA-EIJK-UNWAR-688/2021 | EPI_ISL_8257987 | 2021-08-21 | RS Ari Canti | [illegible] Molecular Biology, National Research and Innovation Agency; Molecular Biology Laboratory, Faculty Medicine and Health Sciences, Warmadewa University | Lidwina Priliani, Sukma Oktavianthi, Lydia V Panggalo, Willy Agustine, Iskandar Adnan, Jessica R Saragih, Fiqry Kleib, Nabilla Zeina Sudrajat, Shahnaz Mutia Dewi, Edison Johar, Muhammad Rezki Rasyak, Sri Masyeni, Erly Sintya, Frilasita A Yudhaputri, Hiday |
| hCoV-19/Colombia/ANT-CWOHC-VG-SEC02262Z/2021 | EPI_ISL_8259903 | 2021-10-09 | AYUDAS DIAGNOSTICAS SURA | Universidad Nacional de Colombia - Laboratorio Gen√≥mico One Health | Sim√≥n Villegas Vel√°squez, Andres F. Cardona-Rios, Maria Stella L√≥pez, Daniel O. Maldonado-Perez, Lina M. Hurtado, Laura Silvana Perez, Karl A Ciuoderis, Maria Ang√©lica Maya, Carolina Mu√±oz-Arango, Celeny Ortiz, Rita Almanza Payares, Idabely Betancur |

| | | | | | |
|---|---|---|---|---|---|
| hCoV-19/USA/DC-GWU-EPI-0001/2021 | EPI_ISL_7983319 | 2021-11-29 | Dept. of Epidemiology/George Washington University | Dept. of Epidemiology/George Washington University | Kamwing Jair, Karina Rivas, Amanda Castel, Jack Villani, Jorge Sepulveda, Megan Landry, Lynn Goldman, Jeanne Jordan |
| hCoV-19/USA/TX-HHD-2112233836/2021 | EPI_ISL_8208957 | 2021-12-17 | Houston Health Dept. | Houston Health Dept. | Ryker Penn, Pamela Brown, Adolfo Lara, Yanlai Lai |
| hCoV-19/USA/TX-HHD-2112234800/2021 | EPI_ISL_8208966 | 2021-12-17 | MAGNOLIA HEALTH CENTER - 7037 CAPITOL | Houston Health Dept. | Ryker Penn, Pamela Brown, Adolfo Lara, Yanlai Lai |
| hCoV-19/Belgium/LHUB-ULB_25160722/2021 | EPI_ISL_8439727 | 2021-11-29 | Department of Microbiology, Laboratoire Hospitalier Universitaire de Bruxelles, Universitï¿½ Libre de Bruxelles | Laboratoire Hospitalier Universitaire de Bruxelles, Universitï¿½ Libre de Bruxelles | Charlotte Michel, Sigi Van Den Wijngaert, Olivier Vandenberg |
| hCoV-19/England/MILK-2EE7A76/2021 | EPI_ISL_8042563 | 2021-12-15 | Lighthouse Lab in Milton Keynes | Wellcome Sanger Institute for the COVID-19 Genomics UK (COG-UK) Consortium | The Lighthouse Lab in Milton Keynes and Alex Alderton, Roberto Amato, Jeffrey Barrett, Sonia Goncalves, Ewan Harrison, David K. Jackson, Ian Johnston, Dominic Kwiatkowski, Cordelia Langford, John Sillitoe on behalf of the Wellcome Sanger Institute COVID-1 |
| hCoV-19/Italy/APU-IZSPB-PT5540/2021 | EPI_ISL_8444093 | 2021-12-23 | Istituto Zooprofilattico Sperimentale della Puglia e della Basilicata | Istituto Zooprofilattico Sperimentale della Puglia e della Basilicata | Parisi A., Bianco A., Capozzi L., Del Sambro L., Simone D., Barbieri G., Quarato C., Ridolfi D., Giannico A., Pace L., Galante D. |
| hCoV-19/USA/NY-PRL-2021_1219_01J18/2021 | EPI_ISL_8093009 | 2021-12-17 | Pandemic Response Lab - NYC | Pandemic Response Lab, R&D | Henry Lee, Michael Hammerling, Melissa Hopkins, Cybill del Castillo, Shinyoung Clair Kang, William Ward, Pradeep Bugga, Sol Rey, Dylan Law, Alex Carpio, Isabel Fernandez Escapa, Haiping Hao, Jon Laurent |
| hCoV-19/USA/CA-HLX-ATM-9NFUZ49QM/2021 | EPI_ISL_8036471 | 2021-11-30 | Kaiser Permanente Southern California | Helix | Kaiser Permanente Southern California, Helix |
| hCoV-19/NewZealand/21MV3581/2021 | EPI_ISL_7985263 | 2021-12-17 | Middlemore Hospital | Institute of Environmental Science and Research (ESR) | Sarah Cockerton, Clare Gebbie, Dhanisha Patel, Rachel Boyle, SallyAnn Harbison, Olivia Stroeven, Xiaoyun Ren, Matt Storey, Nikki Freed, Muhammad Faisal, Jing Wang, Hermes Perez, Anja Werno, Antje van der Linden, Arlo Upton, Chris Mansell, David Hammer, Dr |
| hCoV-19/England/MILK-2EE4736/2021 | EPI_ISL_8045772 | 2021-12-16 | Lighthouse Lab in Milton Keynes | Wellcome Sanger Institute for the COVID-19 Genomics UK (COG-UK) Consortium | The Lighthouse Lab in Milton Keynes and Alex Alderton, Roberto Amato, Jeffrey Barrett, Sonia Goncalves, Ewan Harrison, David K. Jackson, Ian Johnston, Dominic Kwiatkowski, Cordelia Langford, John Sillitoe on behalf of the Wellcome Sanger Institute COVID-1 |
| hCoV-19/USA/NY-PRL-2021_1216_00F18/2021 | EPI_ISL_8095002 | 2021-12-10 | Pandemic Response Lab - NYC | Pandemic Response Lab, R&D | Henry Lee, Michael Hammerling, Melissa Hopkins, Cybill del Castillo, Shinyoung Clair Kang, William Ward, Pradeep Bugga, Sol Rey, Dylan Law, Alex Carpio, Isabel Fernandez Escapa, Haiping Hao, Jon Laurent |
| hCoV-19/TrinidadandTobago/108649/2021 | EPI_ISL_8262769 | 2021-12-06 | Trinidad Public Health Laboratory | Carrington Lab, Department of Preclinical Sciences, Faculty of Medical Sciences, The University of the West Indies, St Augustine Campus | Nikita S. D. Sahadeo, Anushka Ramjag, Arianne Brown-Jordan, Vernie Ramkissoon, Soren Nicholls, Nicholas Mohammed, Sarah Hill, Roshan Parasram, Avery Hinds, Nuno Faria, Oliver Pybus, Christopher Oura, Gabriel Escobar, Christine V. F. Carrington |

| | | | | | |
|---|---|---|---|---|---|
| hCoV-19/England/MILK-2EFB4BD/2021 | EPI_ISL_8046610 | 2021-12-15 | Lighthouse Lab in Milton Keynes | Wellcome Sanger Institute for the COVID-19 Genomics UK (COG-UK) Consortium | The Lighthouse Lab in Milton Keynes and Alex Alderton, Roberto Amato, Jeffrey Barrett, Sonia Goncalves, Ewan Harrison, David K. Jackson, Ian Johnston, Dominic Kwiatkowski, Cordelia Langford, John Sillitoe on behalf of the Wellcome Sanger Institute COVID-1 |
| hCoV-19/NewZealand/21MV3554/2021 | EPI_ISL_7985237 | 2021-12-04 | LabPLUS | Institute of Environmental Science and Research (ESR) | Sarah Cockerton, Clare Gebbie, Dhanisha Patel, Rachel Boyle, SallyAnn Harbison, Olivia Stroeven, Xiaoyun Ren, Matt Storey, Nikki Freed, Muhammad Faisal, Jing Wang, Hermes Perez, Anja Werno, Antje van der Linden, Arlo Upton, Chris Mansell, David Hammer, Dr |
| hCoV-19/NewZealand/21CV3421/2021 | EPI_ISL_7984977 | 2021-11-29 | LabPLUS | Institute of Environmental Science and Research (ESR) | Sarah Cockerton, Clare Gebbie, Dhanisha Patel, Rachel Boyle, SallyAnn Harbison, Olivia Stroeven, Xiaoyun Ren, Matt Storey, Nikki Freed, Muhammad Faisal, Jing Wang, Hermes Perez, Anja Werno, Antje van der Linden, Arlo Upton, Chris Mansell, David Hammer, Dr |
| hCoV-19/England/QEUH-2EF92A6/2021 | EPI_ISL_8047263 | 2021-12-15 | Lighthouse Lab in Glasgow | Wellcome Sanger Institute for the COVID-19 Genomics UK (COG-UK) Consortium | Harper VanSteenhouse, Yumi Kasai, David Gray, Carol Clugston, Anna Dominiczak and Alex Alderton, Roberto Amato, Jeffrey Barrett, Sonia Goncalves, Ewan Harrison, David K. Jackson, Ian Johnston, Dominic Kwiatkowski, Cordelia Langford, John Sillitoe on behal |
| hCoV-19/England/ALDP-2EF162A/2021 | EPI_ISL_8044906 | 2021-12-15 | Lighthouse Lab in Alderley Park | Wellcome Sanger Institute for the COVID-19 Genomics UK (COG-UK) Consortium | Jacquelyn Wynn, Mairead Hyland, The Lighthouse Lab in Alderley Park and Alex Alderton, Roberto Amato, Jeffrey Barrett, Sonia Goncalves, Ewan Harrison, David K. Jackson, Ian Johnston, Dominic Kwiatkowski, Cordelia Langford, John Sillitoe on behalf of the W |
| hCoV-19/Japan/IC-2449/2021 | EPI_ISL_8097033 | 2021-12-18 | SARS-CoV-2 testing team, National Institute of Infectious Diseases | Pathogen Genomics Center, National Institute of Infectious Diseases | Tsuyoshi Sekizuka, Kentaro Itokawa, Rina Tanaka, Masanori Hashino, Nozomu Hanaoka, Masumichi Saito, Naomi Nojiri, Hazuka Y Furihata, Tsuguto Fujimoto, Makoto Kuroda |
| hCoV-19/USA/MA-CDCBI-CRSP_AJSZPA3XZ6CDKXX5/2021 | EPI_ISL_8211060 | 2021-12-21 | Broad Institute Clinical Research Sequencing Platform | Infectious Disease Program, Broad Institute of Harvard and MIT | Siddle,K.J., Adams,G., Pearlman,L., Gladden-Young,A., Vicente,G., Blumenstiel,B., DeFelice,M., Lee,M., McGovern,S., Lagerborg,K., Rudy,M., DeRuff,K., Carter,A., Normandin,E., Bauer,M., Reilly,S., Tomkins-Tinch,C., Loreth,C., Chaluvadi,S., Meldrim,J., Gran |
| hCoV-19/Japan/IC-2473/2021 | EPI_ISL_8097057 | 2021-12-19 | SARS-CoV-2 testing team, National Institute of Infectious Diseases | Pathogen Genomics Center, National Institute of Infectious Diseases | Tsuyoshi Sekizuka, Kentaro Itokawa, Rina Tanaka, Masanori Hashino, Nozomu Hanaoka, Masumichi Saito, Naomi Nojiri, Hazuka Y Furihata, Tsuguto Fujimoto, Makoto Kuroda |
| hCoV-19/Nigeria/CV3548/2021 | EPI_ISL_7123015 | 2021-06-02 | Genexpert Molecular Diagnostic Laboratory | Africa Centre for Excellence for Genomics of Infectious Diseases (ACEGID), Redeemer‚Äôs University | Olawoye, I.B., Oluniyi, P.E., Eromon, P.E., Oguzie, J.U., Kayode, A.T., Uwanibe, J.N., Ugwu, C.A., Akano, K.O., Ajogbasile, F.V., Abechi, P.S., Olumade, T.J., Nosamiefan, I., Folarin, O., Happi, C.T. |
| hCoV-19/Nigeria/CV3551/2021 | EPI_ISL_7123041 | 2021-03-02 | Genexpert Molecular Diagnostic Laboratory | Africa Centre for Excellence for Genomics of Infectious Diseases (ACEGID), Redeemer‚Äôs University | Olawoye, I.B., Oluniyi, P.E., Eromon, P.E., Oguzie, J.U., Kayode, A.T., Uwanibe, J.N., Ugwu, C.A., Akano, K.O., Ajogbasile, F.V., Abechi, P.S., Olumade, T.J., Nosamiefan, I., Folarin, O., Happi, C.T. |

| | | | | | |
|---|---|---|---|---|---|
| hCoV-19/Nigeria/CV3559/2021 | EPI_ISL_7123108 | 2021-06-02 | Genexpert Molecular Diagnostic Laboratory | Africa Centre for Excellence for Genomics of Infectious Diseases (ACEGID), Redeemer‚Äôs University | Olawoye, I.B., Oluniyi, P.E., Eromon, P.E., Oguzie, J.U., Kayode, A.T., Uwanibe, J.N., Ugwu, C.A., Akano, K.O., Ajogbasile, F.V., Abechi, P.S., Olumade, T.J., Nosamiefan, I., Folarin, O., Happi, C.T. |
| hCoV-19/Nigeria/CV3565/2021 | EPI_ISL_7123155 | 2021-04-02 | Genexpert Molecular Diagnostic Laboratory | Africa Centre for Excellence for Genomics of Infectious Diseases (ACEGID), Redeemer‚Äôs University | Olawoye, I.B., Oluniyi, P.E., Eromon, P.E., Oguzie, J.U., Kayode, A.T., Uwanibe, J.N., Ugwu, C.A., Akano, K.O., Ajogbasile, F.V., Abechi, P.S., Olumade, T.J., Nosamiefan, I., Folarin, O., Happi, C.T. |
| hCoV-19/Nigeria/CV3576/2021 | EPI_ISL_7123249 | 2021-02-24 | Genexpert Molecular Diagnostic Laboratory | Africa Centre for Excellence for Genomics of Infectious Diseases (ACEGID), Redeemer‚Äôs University | Olawoye, I.B., Oluniyi, P.E., Eromon, P.E., Oguzie, J.U., Kayode, A.T., Uwanibe, J.N., Ugwu, C.A., Akano, K.O., Ajogbasile, F.V., Abechi, P.S., Olumade, T.J., Nosamiefan, I., Folarin, O., Happi, C.T. |
| hCoV-19/Nigeria/CV3578/2021 | EPI_ISL_7123263 | 2021-02-24 | Genexpert Molecular Diagnostic Laboratory | Africa Centre for Excellence for Genomics of Infectious Diseases (ACEGID), Redeemer‚Äôs University | Olawoye, I.B., Oluniyi, P.E., Eromon, P.E., Oguzie, J.U., Kayode, A.T., Uwanibe, J.N., Ugwu, C.A., Akano, K.O., Ajogbasile, F.V., Abechi, P.S., Olumade, T.J., Nosamiefan, I., Folarin, O., Happi, C.T. |
| hCoV-19/Nigeria/CV3579/2021 | EPI_ISL_7123274 | 2021-02-24 | Genexpert Molecular Diagnostic Laboratory | Africa Centre for Excellence for Genomics of Infectious Diseases (ACEGID), Redeemer‚Äôs University | Olawoye, I.B., Oluniyi, P.E., Eromon, P.E., Oguzie, J.U., Kayode, A.T., Uwanibe, J.N., Ugwu, C.A., Akano, K.O., Ajogbasile, F.V., Abechi, P.S., Olumade, T.J., Nosamiefan, I., Folarin, O., Happi, C.T. |
| hCoV-19/Nigeria/CV3587/2021 | EPI_ISL_7123343 | 2021-01-27 | Genexpert Molecular Diagnostic Laboratory | Africa Centre for Excellence for Genomics of Infectious Diseases (ACEGID), Redeemer‚Äôs University | Olawoye, I.B., Oluniyi, P.E., Eromon, P.E., Oguzie, J.U., Kayode, A.T., Uwanibe, J.N., Ugwu, C.A., Akano, K.O., Ajogbasile, F.V., Abechi, P.S., Olumade, T.J., Nosamiefan, I., Folarin, O., Happi, C.T. |
| hCoV-19/Nigeria/CV3591/2021 | EPI_ISL_7123386 | 2021-03-02 | Genexpert Molecular Diagnostic Laboratory | Africa Centre for Excellence for Genomics of Infectious Diseases (ACEGID), Redeemer‚Äôs University | Olawoye, I.B., Oluniyi, P.E., Eromon, P.E., Oguzie, J.U., Kayode, A.T., Uwanibe, J.N., Ugwu, C.A., Akano, K.O., Ajogbasile, F.V., Abechi, P.S., Olumade, T.J., Nosamiefan, I., Folarin, O., Happi, C.T. |
| hCoV-19/Senegal/IR1-0077385/2021 | EPI_ISL_7123544 | 2021-10-11 | Iressef Genomics lab | IRESSEF | Souleymane MBOUP, Abdou PADANE , Khadim GUEYE, Cyrille Diedhiou , Samba Ndiour, Diabou Diagne, Papa Alassane DIAW, Birahim Piere NDIAYE, Barada CISSE, Aminata MBOUP, Moustapha MBOW, Ndeye Coumba Toure KANE, Nafisatou LEYE, Gora LO, Ambroise AHOUIDI , Asto |
| hCoV-19/Senegal/IR1-0077683/2021 | EPI_ISL_7123549 | 2021-11-01 | Iressef Genomics lab | IRESSEF | Souleymane MBOUP, Abdou PADANE , Khadim GUEYE, Cyrille Diedhiou , Samba Ndiour, Diabou Diagne, Papa Alassane DIAW, Birahim Piere NDIAYE, Barada CISSE, Aminata MBOUP, Moustapha MBOW, Ndeye Coumba Toure KANE, Nafisatou LEYE, Gora LO, Ambroise AHOUIDI , Asto |
| hCoV-19/USA/NY-PRL-2021_1126_01A06/2021 | EPI_ISL_7129868 | 2021-11-24 | Pandemic Response Lab - NYC | Pandemic Response Lab, R&D | Henry Lee, Michael Hammerling, Melissa Hopkins, Cybill del Castillo, Shinyoung Clair Kang, William Ward, Pradeep Bugga, Sol Rey, Dylan Law, Alex Carpio, Isabel Fernandez Escapa, Haiping Hao, Jon Laurent |

| | | | | | |
|---|---|---|---|---|---|
| hCoV-19/Ghana/FS-21-2655/2021 | EPI_ISL_7130080 | 2021-09-24 | National Influenza Centre | National Influenza Centre | William K. Ampofo, Michael Marks, Ivy A. Asante, Sharon Hsu, Benjamiin B. Lindsey, Benjamin H. Foulkes, Mildred Adusei-Poku, Bright Adu, Linda Boatemaa, Lorreta Kwasah, Quaneeta Mohktar, Joyce Appiah-Kubi, Hilda Opoku Frempong, Keren Okyerebea Attiku, Bl |
| hCoV-19/Brazil/SP-IB_141500/2021 | EPI_ISL_7132681 | 2021-11-18 | CENTRO DE SAUDE DIC I | Instituto Butantan | Dimas Tadeu Covas, Antonio Jorge Martins, Claudia Renata dos Santos Barros, David Schlesinger, Debora Botequio Moretti, Elaine Cristina Marqueze, Elaine Vieira Santos, Evandra Strazza Rodrigues, Heidge Fukumasu, Jayme Augusto de Souza-Neto, Jos√© Salvator |
| hCoV-19/Israel/CVL-22622/2021 | EPI_ISL_7133485 | 2021-01-24 | Israel Central Virology laboratory | Israel Central Virology laboratory | Neta Zuckerman, Efrat Dahan Bucris, Michal Mandelboim, Miranda Geva, Omer Asraf, Efrat Glick-Saar, Gideon Rechavi, Oran Erster, Danit Sofer, Ella Mendelson, Orna Mor |
| hCoV-19/Israel/CVL-8000540/2021 | EPI_ISL_7133942 | 2021-11-21 | Israel Central Virology laboratory | Israel Central Virology laboratory | Neta Zuckerman, Efrat Dahan Bucris, Michal Mandelboim, Miranda Geva, Omer Asraf, Efrat Glick-Saar, Gideon Rechavi, Oran Erster, Danit Sofer, Ella Mendelson, Orna Mor |
| hCoV-19/Israel/CVL-22623/2021 | EPI_ISL_7134152 | 2021-01-26 | Israel Central Virology laboratory | Israel Central Virology laboratory | Neta Zuckerman, Efrat Dahan Bucris, Michal Mandelboim, Miranda Geva, Omer Asraf, Efrat Glick-Saar, Gideon Rechavi, Oran Erster, Danit Sofer, Ella Mendelson, Orna Mor |
| hCoV-19/Israel/CVL-8000066/2021 | EPI_ISL_7134654 | 2021-11-12 | Israel Central Virology laboratory | Israel Central Virology laboratory | Neta Zuckerman, Efrat Dahan Bucris, Michal Mandelboim, Miranda Geva, Omer Asraf, Efrat Glick-Saar, Gideon Rechavi, Oran Erster, Danit Sofer, Ella Mendelson, Orna Mor |
| hCoV-19/Israel/CVL-22620/2021 | EPI_ISL_7134711 | 2021-01-26 | Israel Central Virology laboratory | Israel Central Virology laboratory | Neta Zuckerman, Efrat Dahan Bucris, Michal Mandelboim, Miranda Geva, Omer Asraf, Efrat Glick-Saar, Gideon Rechavi, Oran Erster, Danit Sofer, Ella Mendelson, Orna Mor |
| hCoV-19/France/IDF-HMN-21112220699/2021 | EPI_ISL_7134912 | 2021-11-16 | BIOPATH LABM | Mondor University Hospital, Assistance Publique H√¥pitaux de Paris, Universit√© Paris-Est Cr√©teil, INSERM U955 | Christophe Rodriguez, Slim Fourati, Vanessa Demontant, Guillaume Gricourt, Melissa N'Debi, Alexandre Soulier, Elisabeth Trawinski, Jean-Michel Pawlotsky |
| hCoV-19/Canada/ON-PHL-21-40785/2021 | EPI_ISL_7135499 | 2021-11-25 | Public Health Ontario Laboratory | Public Health Ontario Laboratory | Vanessa G Allen, Philip Banh, Yao Chen, Richard de Borja, Alireza Eshaghi, Nahuel Fittipaldi, Christine Frantz, Jonathan B Gubbay, Jennifer L Guthrie, Lawrence Heisler, Esha Joshi, Michael Laszloffy, Aimin Li, Michael CY Li, Dean Maxwell, Sandeep Nagra, S |
| hCoV-19/USA/CO-CDPHE-2102258147/2021 | EPI_ISL_7137016 | 2021-11-18 | Colorado Department of Public Health and Environment | Colorado Department of Public Health and Environment | Laura Bankers, Molly C. Hetherington-Rauth, Diana Ir, Alexandria Rossheim, Michael A. Martin, Mandy Waters, Shannon R. Matzinger, Sarah Elizabeth Totten, Emily A. Travanty |
| hCoV-19/USA/CA-CDPH-6000005435/2021 | EPI_ISL_7137326 | 2021-11-30 | California Department of Public Health | California Department of Public Health | CDPH IDLB COVIDNet |

| | | | | | |
|---|---|---|---|---|---|
| hCoV-19/HongKong/HKU-691/2021 | EPI_ISL_7138045 | 2021-11-14 | Department of Microbiology, The University of Hong Kong | Department of Microbiology, The University of Hong Kong | Kelvin K.W. To, Kwok-Yung Yuen |
| hCoV-19/Germany/RP-USAFSAM-S8816/2021 | EPI_ISL_7138443 | 2021-10-30 | US Air Force School of Aerospace Medicine | US Air Force School of Aerospace Medicine | Anthony Fries, Deanna Muehleman, Jennifer Meyer, William Gruner, Kelsey Lanter, Carol Garrett, Sarah Purves, Clarise Starr, Elizabeth Macias, Fritz Castillo, Jennifer Black, Cole Anderson |
| hCoV-19/Benin/NMIMR-316200/2021 | EPI_ISL_7139563 | 2021-03-04 | National Reference Laboratory, Laboratories Department, Cotonou, Benin | Noguchi Memorial Institute for Medical Research, University of Ghana, Legon, Ghana | Rene K. Keke, Anges W. Yadouleton, Quaneeta Mohktar, Hilda Opoku Frempong, Ange D. Dossou, Keren O. Attiku, Joyce Appiah-Kubi, Benjamin I.B. Hounkpatin, Bright Adu |
| hCoV-19/Benin/NMIMR-316947/2021 | EPI_ISL_7139583 | 2021-03-04 | National Reference Laboratory, Laboratories Department, Cotonou, Benin | Noguchi Memorial Institute for Medical Research, University of Ghana, Legon, Ghana | Rene K. Keke, Anges W. Yadouleton, Quaneeta Mohktar, Hilda Opoku Frempong, Ange D. Dossou, Keren O. Attiku, Joyce Appiah-Kubi, Benjamin I.B. Hounkpatin, Bright Adu |
| hCoV-19/Benin/NMIMR-LFHV520243/2021 | EPI_ISL_7139894 | 2021-08-17 | National Reference Laboratory, Laboratories Department, Cotonou, Benin | Noguchi Memorial Institute for Medical Research, University of Ghana, Legon, Ghana | Rene K. Keke, Anges W. Yadouleton, Quaneeta Mohktar, Hilda Opoku Frempong, Ange D. Dossou, Keren O. Attiku, Joyce Appiah-Kubi, Benjamin I.B. Hounkpatin, Bright Adu |
| hCoV-19/Benin/NMIMR-LFHV530999/2021 | EPI_ISL_7139930 | 2021-08-24 | National Reference Laboratory, Laboratories Department, Cotonou, Benin | Noguchi Memorial Institute for Medical Research, University of Ghana, Legon, Ghana | Rene K. Keke, Anges W. Yadouleton, Quaneeta Mohktar, Hilda Opoku Frempong, Ange D. Dossou, Keren O. Attiku, Joyce Appiah-Kubi, Benjamin I.B. Hounkpatin, Bright Adu |
| hCoV-19/Australia/NT-RDH-39/2021 | EPI_ISL_7141549 | 2021-11-14 | Territory Pathology | Territory Pathology | Ella Meumann, Dimitrios Menouhos, Stefanos Christofis, Farshid Dakh, Kevin Freeman, Robert Baird |
| hCoV-19/Australia/NT-RDH-07/2021 | EPI_ISL_7141551 | 2021-10-11 | Territory Pathology | Territory Pathology | Ella Meumann, Dimitrios Menouhos, Stefanos Christofis, Farshid Dakh, Kevin Freeman, Robert Baird |
| hCoV-19/Australia/NT-RDH-04/2021 | EPI_ISL_7141557 | 2021-09-20 | Territory Pathology | Territory Pathology | Ella Meumann, Dimitrios Menouhos, Stefanos Christofis, Farshid Dakh, Kevin Freeman, Robert Baird |
| hCoV-19/England/MILK-2BFB5F9/2021 | EPI_ISL_7146933 | 2021-11-26 | Lighthouse Lab in Milton Keynes | Wellcome Sanger Institute for the COVID-19 Genomics UK (COG-UK) Consortium | The Lighthouse Lab in Milton Keynes and Alex Alderton, Roberto Amato, Jeffrey Barrett, Sonia Goncalves, Ewan Harrison, David K. Jackson, Ian Johnston, Dominic Kwiatkowski, Cordelia Langford, John Sillitoe on behalf of the Wellcome Sanger Institute COVID-1 |
| hCoV-19/Scotland/QEUH-2C2C664/2021 | EPI_ISL_7148719 | 2021-11-27 | Lighthouse Lab in Glasgow | Wellcome Sanger Institute for the COVID-19 Genomics UK (COG-UK) Consortium | Harper VanSteenhouse, Yumi Kasai, David Gray, Carol Clugston, Anna Dominiczak and Alex Alderton, Roberto Amato, Jeffrey Barrett, Sonia Goncalves, Ewan Harrison, David K. Jackson, Ian Johnston, Dominic Kwiatkowski, Cordelia Langford, John Sillitoe on behal |
| hCoV-19/USA/CO-CDC-MMB11469309/2021 | EPI_ISL_7153140 | 2021-11-05 | Mako Medical | Centers for Disease Control and Prevention Division of Viral Diseases, Pathogen Discovery | Dakota Howard,Dhwani Batra,Peter Cook,Jason Caravas,Benjamin Rambo-Martin,Scott Sammons,Yvette Unoarumhi,Matthew Schmerer,Kristine Lacek,Tymeckia Kendall,Victoria Caban Figueroa,Shatavia Morrison,Christopher Gulvick,Erisa Sula,Matthew Tugwell,Lauren Moon, |
| hCoV-19/USA/CA-CDC-FG-177176/2021 | EPI_ISL_7154399 | 2021-11-26 | Fulgent Genetics | Centers for Disease Control and Prevention Division of Viral Diseases, Pathogen Discovery | Dakota Howard,Dhwani Batra,Peter Cook,Jason Caravas,Benjamin Rambo-Martin,Scott Sammons,Yvette Unoarumhi,Matthew Schmerer,Kristine Lacek,Tymeckia Kendall,Victoria Caban Figueroa,Shatavia Morrison,Christopher Gulvick,Erisa Sula,Harry Gao,Mickey Li,John Gao |

| | | | | | |
|---|---|---|---|---|---|
| hCoV-19/USA/CO-CDC-MMB11864811/2021 | EPI_ISL_7154401 | 2021-11-29 | Mako Medical | Centers for Disease Control and Prevention Division of Viral Diseases, Pathogen Discovery | Dakota Howard,Dhwani Batra,Peter Cook,Jason Caravas,Benjamin Rambo-Martin,Scott Sammons,Yvette Unoarumhi,Matthew Schmerer,Kristine Lacek,Tymeckia Kendall,Victoria Caban Figueroa,Shatavia Morrison,Christopher Gulvick,Erisa Sula,Matthew Tugwell,Lauren Moon, |
| hCoV-19/USA/MO-CDC-ASC210713936/2021 | EPI_ISL_7154405 | 2021-11-24 | Aegis Sciences Corporation | Centers for Disease Control and Prevention Division of Viral Diseases, Pathogen Discovery | Dakota Howard,Dhwani Batra,Peter Cook,Jason Caravas,Benjamin Rambo-Martin,Scott Sammons,Yvette Unoarumhi,Matthew Schmerer,Kristine Lacek,Tymeckia Kendall,Victoria Caban Figueroa,Shatavia Morrison,Christopher Gulvick,Erisa Sula,Cyndi Clark,Patrick Campbell |
| hCoV-19/Nigeria/NCDC-NR833/2021 | EPI_ISL_7154463 | 2021-11-25 | IFAIN Labs | National Reference Laboratory, Nigeria Centre for Disease Control | Dr Ndodo Nnaemeka, Olusola Anuoluwapo Akanbi, Chimaobi Chukwu, Oyeronke Ayansola, Dr Omoare Adesuyi, Catherine Okoi, Olajumoke Babatunde, Nwando Mba, Dr Ifedayo Adetifa |
| hCoV-19/Mexico/CMX-InDRE_FD122437_S9550/2021 | EPI_ISL_7156454 | 2021-11-29 | Hospital √Öngeles Lomas | Instituto de diagn√≥stico y Referencia Epidemiologicos (INDRE) | Claudia Wong-Arambula, Abril Rodriguez-Maldonado, Raymundo Rodr√≠guez Sandoval, Vanessa Rivero-Arredondo, Ariadna Medina-Benitez, Joaquin Quiroz-Mercado, Maribel Gonzalez-Villa, Natividad Cruz-Ortiz, Tatiana Nunez-Garcia, Gisela Barrera-Badillo, Fernando |
| hCoV-19/France/PAC-IHU-45003/2021 | EPI_ISL_7156955 | 2021-11-22 | IHU Mediterranee Infection | IHU Mediterranee Infection | Philippe Colson et al. |
| hCoV-19/Australia/NT-RDH-13/2021 | EPI_ISL_7161992 | 2021-09-26 | Territory Pathology | Territory Pathology | Ella Meumann, Dimitrios Menouhos, Stefanos Christofis, Farshid Dakh, Kevin Freeman, Robert Baird |
| hCoV-19/England/PHEC-3U06FU0A/2021 | EPI_ISL_7166216 | 2021-11-23 | Respiratory Virus Unit, Microbiology Services Colindale, Public Health England | COVID-19 Genomics UK (COG-UK) Consortium | PHE Covid Sequencing Team |
| hCoV-19/Qatar/QA.QU.18.28.B4/2021 | EPI_ISL_7166983 | 2021-09-25 | Ministry of Public Health / Hamad Medical Corporation | Biomedical Research Center (BRC), Qatar University / Qatar Genome Project (QGP) | BRC:¬†Heba A. Al-Khatib, Fatiha M. Benslimane,Maria K. Smatti, Hadeel T. Mohammed, Fatma Hassan Ali, Swapna Thomas, Ola Al-Jamal, Dana Al-Batsh, Hadi M. Yassine, Asmaa A. Al-Thani. MOPH and HMC:¬†Abdullatif Al-Khal, Muna A. S. Al-Maslamani, Mashael A. Al- |
| hCoV-19/Qatar/QA.QU.19.7.B10/2021 | EPI_ISL_7166998 | 2021-09-29 | Ministry of Public Health / Hamad Medical Corporation | Biomedical Research Center (BRC), Qatar University / Qatar Genome Project (QGP) | BRC:¬†Heba A. Al-Khatib, Fatiha M. Benslimane,Maria K. Smatti, Hadeel T. Mohammed, Fatma Hassan Ali, Swapna Thomas, Ola Al-Jamal, Dana Al-Batsh, Hadi M. Yassine, Asmaa A. Al-Thani. MOPH and HMC:¬†Abdullatif Al-Khal, Muna A. S. Al-Maslamani, Mashael A. Al- |
| hCoV-19/Qatar/QA.QU.18.28.C9/2021 | EPI_ISL_7167014 | 2021-10-05 | Ministry of Public Health / Hamad Medical Corporation | Biomedical Research Center (BRC), Qatar University / Qatar Genome Project (QGP) | BRC:¬†Heba A. Al-Khatib, Fatiha M. Benslimane,Maria K. Smatti, Hadeel T. Mohammed, Fatma Hassan Ali, Swapna Thomas, Ola Al-Jamal, Dana Al-Batsh, Hadi M. Yassine, Asmaa A. Al-Thani. MOPH and HMC:¬†Abdullatif Al-Khal, Muna A. S. Al-Maslamani, Mashael A. Al- |

| | | | | | |
|---|---|---|---|---|---|
| hCoV-19/Qatar/QA.QU.20.8.E4/2021 | EPI_ISL_7167052 | 2021-10-23 | Ministry of Public Health / Hamad Medical Corporation | Biomedical Research Center (BRC), Qatar University / Qatar Genome Project (QGP) | BRC:¬†Heba A. Al-Khatib, Fatiha M. Benslimane,Maria K. Smatti, Hadeel T. Mohammed, Fatma Hassan Ali, Swapna Thomas, Ola Al-Jamal, Dana Al-Batsh, Hadi M. Yassine, Asmaa A. Al-Thani. MOPH and HMC:¬†Abdullatif Al-Khal, Muna A. S. Al-Maslamani, Mashael A. Al- |
| hCoV-19/Qatar/QA.QU.19.7.G4/2021 | EPI_ISL_7167084 | 2021-11-02 | Ministry of Public Health / Hamad Medical Corporation | Biomedical Research Center (BRC), Qatar University / Qatar Genome Project (QGP) | BRC:¬†Heba A. Al-Khatib, Fatiha M. Benslimane,Maria K. Smatti, Hadeel T. Mohammed, Fatma Hassan Ali, Swapna Thomas, Ola Al-Jamal, Dana Al-Batsh, Hadi M. Yassine, Asmaa A. Al-Thani. MOPH and HMC:¬†Abdullatif Al-Khal, Muna A. S. Al-Maslamani, Mashael A. Al- |
| hCoV-19/Slovakia/UVZ_PL79_A1_36063/2021 | EPI_ISL_7167629 | 2021-11-12 | Public Health Authority of the Slovak Republic | Laboratory of Genomics and Bioinformatics, Comenius University Science Park | Tatiana Sedl√°ƒçkov√°, Diana Rus≈à√°kov√°, Miroslav B√∂hmer, Anna Gifçov√°, Jaroslav Budi≈°, Tom√°≈° Szemes, Jakub Styk |
| hCoV-19/Colombia/ANT-LDSP1261/2021 | EPI_ISL_7170017 | 2021-10-11 | SOMER | Laboratorio Departamental de Salud Publica de Antioquia | Idabely Betancur Ortiz, Maria Stella L√≥pez, Gloria Isabel Escobar, Juan P. Hernandez-Ortiz, Cristian Arbey Velarde Hoyos, Juan Pablo Isaza Agudelo, Ana Victoria Valencia Duarte |
| hCoV-19/Switzerland/ZH-UZH-IMV-3ba57505/2021 | EPI_ISL_7170972 | 2021-11-28 | Stadtspital Triemli | Institute of Medical Virology | Verena Kufner, Gabriela Ziltener, Maryam Zaheri, Stefan Schmutz, Annette Audig√©, Maria Gr√ºnberg, Kevin Steiner, Jon Huder, Cyril Shah, Riccarda Capaul, Guido Bloemberg, J√ºrg B√∂ni, Michael Huber, Alexandra Trkola |
| hCoV-19/India/MH-Pune-INSACOG_RF-NCL-1870/2021 | EPI_ISL_7173962 | 2021-11-29 | GENEPATH | NCL, Pune | Anu Raghunathan, Dhanasekaran Shanmugam, Ajinkya Khilari, Divya Niveditha, Bhagyashree Litikar, Jugal Kanekar, Shikha Takur |
| hCoV-19/Chile/AN_1108157/2021 | EPI_ISL_7174193 | 2021-11-26 | Hospital Regional Antofagasta (HRA) | Microbiana, Unidad de Bioqu√≠mica, Departamento Biom√©dico, Facultad de Ciencias de la Salud, Universidad de Antofagasta, Chile. | Alexandra Galetoviƒá, Jorge Araya, Camila Salazar-Ardiles, Benito G√≥mez-Silva, Milton Urrutia, Alexander P√©rez, Gabriel Pe√±a, Mart√≠n Vargas, Carlos Cornejo. |
| hCoV-19/Chile/AN_19353604/2021 | EPI_ISL_7174205 | 2021-11-25 | Laboratorio Cl√≠nico, Secci√≥n Biolog√≠a Molecular. Hospital Dr. Carlos Cisternas, Calama. Seremi√≠a Salud Antofagasta. | Microbiana, Unidad de Bioqu√≠mica, Departamento Biom√©dico, Facultad de Ciencias de la Salud, Universidad de Antofagasta, Chile. | Alexandra Galetoviƒá, Jorge Araya, Camila Salazar-Ardiles, Benito G√≥mez-Silva, Milton Urrutia, Alexander P√©rez, Gabriel Pe√±a, Mart√≠n Vargas, Carlos Cornejo. |
| hCoV-19/USA/OH-CDC-ASC210486405/2021 | EPI_ISL_7178993 | 2021-11-19 | Aegis Sciences Corporation | Centers for Disease Control and Prevention Division of Viral Diseases, Pathogen Discovery | Dakota Howard,Dhwani Batra,Peter Cook,Jason Caravas,Benjamin Rambo-Martin,Scott Sammons,Yvette Unoarumhi,Matthew Schmerer,Kristine Lacek,Tymeckia Kendall,Victoria Caban Figueroa,Shatavia Morrison,Christopher Gulvick,Erisa Sula,Cyndi Clark,Patrick Campbell |
| hCoV-19/England/ALDP-2C384D4/2021 | EPI_ISL_7188438 | 2021-11-25 | Lighthouse Lab in Alderley Park | Wellcome Sanger Institute for the COVID-19 Genomics UK (COG-UK) Consortium | Jacquelyn Wynn, Mairead Hyland, The Lighthouse Lab in Alderley Park and Alex Alderton, Roberto Amato, Jeffrey Barrett, Sonia Goncalves, Ewan Harrison, David K. Jackson, Ian Johnston, Dominic Kwiatkowski, Cordelia Langford, John Sillitoe on behalf of the W |
| hCoV-19/Australia/WA835/2021 | EPI_ISL_7189373 | 2021-12-01 | PathWest Laboratory Medicine WA | PathWest Laboratory Medicine WA Microbial Surveillance Unit | PathWest Laboratory Medicine WA Microbial Surveillance Unit |

| | | | | | |
|---|---|---|---|---|---|
| hCoV-19/Australia/QLD2562/2021 | EPI_ISL_7190352 | 2021-11-29 | PHV-FSS | PHV-FSS | Chenwei Wang on behalf of Q-PHIRE Genomics |
| hCoV-19/USA/CA-CDPH-500022757/2021 | EPI_ISL_7192241 | 2021-09-29 | TPMG Regional Laboratory | California Department of Public Health | Emily Smith on behalf of CDPH-COVIDNet and UCSF Center for Advanced Technology |
| hCoV-19/Japan/IC-2269/2021 | EPI_ISL_7192648 | 2021-11-21 | SARS-CoV-2 testing team, National Institute of Infectious Diseases | Pathogen Genomics Center, National Institute of Infectious Diseases | Tsuyoshi Sekizuka, Kentaro Itokawa, Rina Tanaka, Masanori Hashino, Nozomu Hanaoka, Masumichi Saito, Naomi Nojiri, Hazuka Y Furihata, Tsuguto Fujimoto, Makoto Kuroda |
| hCoV-19/Colombia/AMA-INS-VG-11087/2021 | EPI_ISL_8097188 | 2021-11-13 | LSPD Amazonas | Instituto Nacional de Salud-Direcci√≥n de Investigaci√≥n en Salud P√∫blica | Katherine Laiton-Donato, Carlos Franco-Mu√±oz, Diego A. √Ålvarez-D√≠az, H√©ctor Alejandro Ruiz-Moreno, Diego Andr√©s Prada, Jhonnatan Reales-Gonz√°lez, Sheryll Corchuelo, Mar√≠a T. Herrera-Sep√∫lveda, Juli√°n Naizaque, Gerardo Santamar√≠a, Jorge Rivera, P |
| hCoV-19/Botswana/R58B4_BHP_AAC42258/2021 | EPI_ISL_8097206 | 2021-12-09 | Nyanyangbwe HIV Reference Laboratory | Botswana Harvard HIV Reference Laboratory | Sikhulile Moyo, Wonderful T. Choga, Dorcas Maruapula,Lindani Bochena, David Matema, Keoratile Ntshambiwa, Thongbotho Mphoyakgosi, Boitumelo Zuze, Botshelo Radibe, Legodile Kooepile, Ontlametse T. Bareng, Thela Tefelo, Sefetogi Ramaologa, Mphapi B. Mbulawa |
| hCoV-19/Botswana/R58B51_BHP_AAC42307/2021 | EPI_ISL_8097211 | 2021-12-09 | Nyanyangbwe HIV Reference Laboratory | Botswana Harvard HIV Reference Laboratory | Sikhulile Moyo, Wonderful T. Choga, Dorcas Maruapula,Lindani Bochena, David Matema, Keoratile Ntshambiwa, Thongbotho Mphoyakgosi, Boitumelo Zuze, Botshelo Radibe, Legodile Kooepile, Ontlametse T. Bareng, Thela Tefelo, Sefetogi Ramaologa, Mphapi B. Mbulawa |
| hCoV-19/Botswana/R59B9_BHP_AAC44117/2021 | EPI_ISL_8097227 | 2021-12-08 | Kasane Primary Hospital Laboratory | Botswana Harvard HIV Reference Laboratory | Sikhulile Moyo, Wonderful T. Choga, Dorcas Maruapula, Agnes Karutwaeng, Rose Munyere, Keoratile Ntshambiwa, Thongbotho Mphoyakgosi, Boitumelo Zuze, Botshelo Radibe, Legodile Kooepile, Ontlametse T. Bareng, Thela Tefelo, Sefetogi Ramaologa, Mphapi B. Mbula |
| hCoV-19/India/MH-ICMR-NIV-INSACOG-GSEQ-6710/2021 | EPI_ISL_8049888 | 2021-11-03 | ICMR-National Institute of Virology - INSACOG | NIV Influenza | Dr. Varsha Potdar and NIC Team |
| hCoV-19/India/MH-ICMR-NIV-INSACOG-GSEQ-7055/2021 | EPI_ISL_8050104 | 2021-11-24 | ICMR-National Institute of Virology - INSACOG | NIV Influenza | Dr. Varsha Potdar and NIC Team |
| hCoV-19/USA/TX-DSHS-12631/2021 | EPI_ISL_8051841 | 2021-08-23 | TXDSHS | TXDSHS | Rashmi Tuladhar, Bonnie Oh, Maliha Rahman, Mayela Pedrueza, Anita Pokharel, Karen Bobier, Yan Sun, Rachel Lee, Grace Kubin |
| hCoV-19/Thailand/DMSc-05495/2021 | EPI_ISL_8098150 | 2021-11-27 | Regional Medical Sciences Center8 Udon Thani | Division of Genomic Medicine and Innovation support,Department of Medical Sciences, Ministry of Public Health, Thailand | Surakameth Mahasirimongkol,Nuanjun Wichukchinda,Archawin Rojanawiwat,Pilailuk Akkapaiboon Okada ,Waritta Sawaengdee,Penpitcha Thawong,Pundharika Piboonsiri,Jirapha Pakdee,Natthakul Bunneang |

| | | | | | |
|---|---|---|---|---|---|
| hCoV-19/USA/NY-MSHSPSP-PV44450/2021 | EPI_ISL_7908023 | 2021-11-28 | MSHS Clinical Microbiology Laboratories | MSHS Pathogen Surveillance Program | Ana S. Gonzalez-Reiche, Hala Alshammary, Mitchell J. Sullivan, Brianne Ciferri, Jose Polanco, Angela Amoako, Mahmoud Awawda, Aria Rooker, Christian Cognigni, Daniel Floda, Adriana van de Guchte, Zain Khalil, Keith Farrugia, Bremy Alburquerque, Julia Matth |
| hCoV-19/Portugal/PT23928/2021 | EPI_ISL_8098704 | 2021-12-14 | UNILABS | Instituto Nacional de Saude (INSA) | Borges et al |
| hCoV-19/England/MILK-2C69FBB/2021 | EPI_ISL_7203085 | 2021-11-29 | Lighthouse Lab in Milton Keynes | Wellcome Sanger Institute for the COVID-19 Genomics UK (COG-UK) Consortium | The Lighthouse Lab in Milton Keynes and Alex Alderton, Roberto Amato, Jeffrey Barrett, Sonia Goncalves, Ewan Harrison, David K. Jackson, Ian Johnston, Dominic Kwiatkowski, Cordelia Langford, John Sillitoe on behalf of the Wellcome Sanger Institute COVID-1 |
| hCoV-19/USA/NY-CDC-ASC210629010/2021 | EPI_ISL_8265217 | 2021-12-17 | Aegis Sciences Corporation | Centers for Disease Control and Prevention Division of Viral Diseases, Pathogen Discovery | Dakota Howard,Dhwani Batra,Peter Cook,Jason Caravas,Benjamin Rambo-Martin,Scott Sammons,Yvette Unoarumhi,Matthew Schmerer,Kristine Lacek,Tymeckia Kendall,Victoria Caban Figueroa,Shatavia Morrison,Christopher Gulvick,Erisa Sula,Cyndi Clark,Patrick Campbell |
| hCoV-19/England/MILK-2C69EDC/2021 | EPI_ISL_7203182 | 2021-11-29 | Lighthouse Lab in Milton Keynes | Wellcome Sanger Institute for the COVID-19 Genomics UK (COG-UK) Consortium | The Lighthouse Lab in Milton Keynes and Alex Alderton, Roberto Amato, Jeffrey Barrett, Sonia Goncalves, Ewan Harrison, David K. Jackson, Ian Johnston, Dominic Kwiatkowski, Cordelia Langford, John Sillitoe on behalf of the Wellcome Sanger Institute COVID-1 |
| hCoV-19/Chile/RM-USS-UNAB-BFU666/2021 | EPI_ISL_8053173 | 2021-09-04 | Universidad San Sebastian | Facultad de Ciencias de la Vida, UNAB | Claudio Meneses, Eduardo Castro, Waldo Diaz, Macarena Bastias, Ricardo Yusta, Susana Saez, Dayan Sanhueza, Sebastian Wolter, Daniel Leon, Gonzalo Campa√±a, Paola Pidal, V√≠ctor Monreal, Claudio Olmos, Ariel Orellana |
| hCoV-19/Chile/RM-USS-UNAB-BFU694/2021 | EPI_ISL_8053176 | 2021-09-04 | Universidad San Sebastian | Facultad de Ciencias de la Vida, UNAB | Claudio Meneses, Eduardo Castro, Waldo Diaz, Macarena Bastias, Ricardo Yusta, Susana Saez, Dayan Sanhueza, Sebastian Wolter, Daniel Leon, Gonzalo Campa√±a, Paola Pidal, V√≠ctor Monreal, Claudio Olmos, Ariel Orellana |
| hCoV-19/England/MILK-2C1908A/2021 | EPI_ISL_8053007 | 2021-11-26 | Lighthouse Lab in Milton Keynes | Wellcome Sanger Institute for the COVID-19 Genomics UK (COG-UK) Consortium | The Lighthouse Lab in Milton Keynes and Alex Alderton, Roberto Amato, Jeffrey Barrett, Sonia Goncalves, Ewan Harrison, David K. Jackson, Ian Johnston, Dominic Kwiatkowski, Cordelia Langford, John Sillitoe on behalf of the Wellcome Sanger Institute COVID-1 |
| hCoV-19/USA/MA-CDCBI-CRSP_7HV72OJ6TREMAKRZ/2021 | EPI_ISL_8215131 | 2021-12-18 | Broad Institute Clinical Research Sequencing Platform | Infectious Disease Program, Broad Institute of Harvard and MIT | Siddle,K.J., Adams,G., Pearlman,L., Gladden-Young,A., Vicente,G., Blumenstiel,B., DeFelice,M., Lee,M., McGovern,S., Lagerborg,K., Rudy,M., DeRuff,K., Carter,A., Normandin,E., Bauer,M., Reilly,S., Tomkins-Tinch,C., Loreth,C., Chaluvadi,S., Meldrim,J., Gran |
| hCoV-19/Denmark/DCGC-283852/2021 | EPI_ISL_8054357 | 2021-11-25 | Department of Bacteria, Parasites and Fungi, Statens Serum Institut, Copenhagen, Denmark | Statens Serum Institut Bioinformatics and Microbial Genomics | Danish Covid-19 Genome Consortium |
| hCoV-19/USA/CA-HLX-ATM-2SK2QZV5N/2021 | EPI_ISL_8035742 | 2021-11-30 | Kaiser Permanente Southern California | Helix | Kaiser Permanente Southern California, Helix |

| | | | | | |
|---|---|---|---|---|---|
| hCoV-19/Singapore/9813/2021 | EPI_ISL_8449974 | 2021-12-30 | National Public Health Laboratory, National Centre for Infectious Diseases | National Centre for Infectious Diseases | Grace Ngan, Benny Yeo, Zhenyang Zhou, Royce Ang, Samuel Loo, Lin Cui, Raymond Tzer Pin Lin |
| hCoV-19/France/PAC-CERBAHC-1225867/2021 | EPI_ISL_8057576 | 2021-12-06 | CERBALLIANCE ALPES DURANCE SITE TOKORO | CERBA HealthCare | B√©n√©dicte Roquebert, Laura Verdurme, Sabine Trombert, St√©phanie Haim-Boukobza |
| hCoV-19/India/TG-RFCH02576_CIB7429/2021 | EPI_ISL_8058382 | 2021-05-28 | MapMyGenome | CSIR-Centre for Cellular and Molecular Biology | Lamuk Zaveri,Tulasi Nagabandi,Jandhyala Sai Krishna,Malini Nemalikanti,Sreelekshmi MS, Ara Sreenivas,Shreekant Verma, Amareshwar Vodapalli,B Himasri,Valli Nagalakshmi Undamatla, Sumedha Avadhanula,Priya Nurkurthy,Vidhyadhari Methuku,M. Aravind Kumar, San |
| hCoV-19/India/AP-CCMB-CIB7606/2021 | EPI_ISL_8058485 | 2021-11-05 | AP SSO | CSIR-Centre for Cellular and Molecular Biology-INSACOG | Priya Nurkurthy,Vidhyadhari Methuku,Sumedha Avadhanula,Valli Nagalakshmi Undamatla,Amareshwar Vodapalli, B Himasri,Shreekant Verma,Ara Sreenivas, Sreelekshmi MS,Malini Nemalikanti,Lamuk Zaveri,Tulasi Nagabandi,Jandhyala Sai Krishna, Archana Bharadwaj Siv |
| hCoV-19/England/MILK-2F87725/2021 | EPI_ISL_8101564 | 2021-12-20 | Lighthouse Lab in Milton Keynes | Wellcome Sanger Institute for the COVID-19 Genomics UK (COG-UK) Consortium | The Lighthouse Lab in Milton Keynes and Alex Alderton, Roberto Amato, Jeffrey Barrett, Sonia Goncalves, Ewan Harrison, David K. Jackson, Ian Johnston, Dominic Kwiatkowski, Cordelia Langford, John Sillitoe on behalf of the Wellcome Sanger Institute COVID-1 |
| hCoV-19/England/MILK-3068708/2021 | EPI_ISL_8443009 | 2021-12-29 | Lighthouse Lab in Milton Keynes | Wellcome Sanger Institute for the COVID-19 Genomics UK (COG-UK) Consortium | The Lighthouse Lab in Milton Keynes and Alex Alderton, Roberto Amato, Jeffrey Barrett, Sonia Goncalves, Ewan Harrison, David K. Jackson, Ian Johnston, Dominic Kwiatkowski, Cordelia Langford, John Sillitoe on behalf of the Wellcome Sanger Institute COVID-1 |
| hCoV-19/Brazil/SC-FIOCRUZ-65149/2021 | EPI_ISL_8266392 | 2021-12-13 | Laboratorio Central de Saude Publica do Estado de Santa Catarina (LACEN/SC) | Laboratory of Respiratory Viruses and Measles, Oswaldo Cruz Institute, FIOCRUZ | Paola Resende, Luciana Appolinario, Fernando Motta, Elisa Cavalcante Pereira, Ighor Arantes, Alice Sampaio Rocha, Bruna Mendon√ßa da Silva, J√©ssica Gra√ßa Macedo de Carvalho, Larissa Macedo Pinto, Victor Guimaraes, Darcita Buerger Rovaris, Sandra Bianchi |
| hCoV-19/Reunion/CHU721120520501/2021 | EPI_ISL_8103879 | 2021-12-06 | CHOR Laboratoire | Laboratoire de virologie, CNR arbovirus Associ√©, Chu de la R√©union | Etienne Frumence, Nicolas M'nemosyme, Anne-Julie Gourd√©, Rubens Lhonneur, Nicolas Traversier, Marie-Christine Jaffar Bandjee |
| hCoV-19/Australia/QLD3488/2021 | EPI_ISL_8215876 | 2021-12-21 | unknown | PHV-FSS | Son Nguyen on behalf of Q-PHIRE Genomics |
| hCoV-19/Australia/QLD4602/2021 | EPI_ISL_8216058 | 2021-12-21 | unknown | PHV-FSS | Son Nguyen on behalf of Q-PHIRE Genomics |
| hCoV-19/Australia/QLD3548/2021 | EPI_ISL_8216087 | 2021-12-21 | unknown | PHV-FSS | Son Nguyen on behalf of Q-PHIRE Genomics |
| hCoV-19/Australia/QLD3565/2021 | EPI_ISL_8216043 | 2021-12-21 | unknown | PHV-FSS | Son Nguyen on behalf of Q-PHIRE Genomics |
| hCoV-19/Australia/QLD3818/2021 | EPI_ISL_8216071 | 2021-12-21 | unknown | PHV-FSS | Son Nguyen on behalf of Q-PHIRE Genomics |
| hCoV-19/Slovakia/36691/2021 | EPI_ISL_7211998 | 2021-11-20 | Public Health Authority of the Slovak Republic | Faculty of Medicine in Martin, Comenius University | Dusan Loderer, Katarina Janikova, Ivana Kasubova, Maria Skerenova, Marian Grendar |

| | | | | | |
|---|---|---|---|---|---|
| hCoV-19/Australia/QLD3790/2021 | EPI_ISL_8216132 | 2021-12-21 | unknown | PHV-FSS | Son Nguyen on behalf of Q-PHIRE Genomics |
| hCoV-19/Australia/QLD4007/2021 | EPI_ISL_8216363 | 2021-12-23 | unknown | PHV-FSS | Son Nguyen on behalf of Q-PHIRE Genomics |
| hCoV-19/Reunion/CHU79800009704302/2021 | EPI_ISL_8103874 | 2021-11-24 | CHU - Saint-Denis | Laboratoire de virologie, CNR arbovirus Associ√©, Chu de la R√©union | Etienne Frumence, Nicolas M'nemosyme, Anne-Julie Gourd√©, Rubens Lhonneur, Nicolas Traversier, Marie-Christine Jaffar Bandjee |
| hCoV-19/Colombia/CUN-LSPSDS-VG-1181561/2021 | EPI_ISL_8216585 | 2021-10-25 | Colsanitas Keralty | LSPSDS | Johana Hernandez Toloza, Marcela Castano Rodriguez,Gabriela Delgado Murcia, Alejandro Gomez Lopez, Marcela Mercado |
| hCoV-19/Colombia/DC-LSPSDS-VG-1176339/2021 | EPI_ISL_8216581 | 2021-05-22 | Compensar Calle 63 | LSPSDS | Johana Hernandez Toloza, Marcela Castano Rodriguez,Gabriela Delgado Murcia, Alejandro Gomez Lopez, Marcela Mercado |
| hCoV-19/USA/TX-CDC-ASC210523314/2021 | EPI_ISL_7920824 | 2021-11-05 | Aegis Sciences Corporation | Centers for Disease Control and Prevention Division of Viral Diseases, Pathogen Discovery | Dakota Howard,Dhwani Batra,Peter Cook,Jason Caravas,Benjamin Rambo-Martin,Scott Sammons,Yvette Unoarumhi,Matthew Schmerer,Kristine Lacek,Tymeckia Kendall,Victoria Caban Figueroa,Shatavia Morrison,Christopher Gulvick,Erisa Sula,Cyndi Clark,Patrick Campbell |
| hCoV-19/India/KL-GENESCOV01205/2021 | EPI_ISL_7217313 | 2021-01-20 | INSACOG-IGIB and Kerala State SARS-COV-2 Genome Surveillance Programme (GENESCOV2) | INSACOG and GENESCOV2 at CSIR Institute of Genomics and Integrative Biology | INSACOG-IGIB and Kerala State SARS-COV-2 Genome Surveillance Programme (GENESCOV2) |
| hCoV-19/India/KL-GENESCOV01236/2021 | EPI_ISL_7217596 | 2021-01-29 | INSACOG-IGIB and Kerala State SARS-COV-2 Genome Surveillance Programme (GENESCOV2) | INSACOG and GENESCOV2 at CSIR Institute of Genomics and Integrative Biology | INSACOG-IGIB and Kerala State SARS-COV-2 Genome Surveillance Programme (GENESCOV2) |
| hCoV-19/India/KL-GENESCOV04543/2021 | EPI_ISL_7220359 | 2021-03-09 | INSACOG-IGIB and Kerala State SARS-COV-2 Genome Surveillance Programme (GENESCOV2) | INSACOG and GENESCOV2 at CSIR Institute of Genomics and Integrative Biology | INSACOG-IGIB and Kerala State SARS-COV-2 Genome Surveillance Programme (GENESCOV2) |
| hCoV-19/France/GES-CERBAHC-12947147/2021 | EPI_ISL_7977462 | 2021-12-08 | LABORATOIRE B2A EPINAL SAUT-LE-CERF | CERBA HealthCare | B√©n√©dicte Roquebert, Laura Verdurme, Sabine Trombert, St√©phanie Haim-Boukobza |
| hCoV-19/England/MILK-2F72514/2021 | EPI_ISL_8107539 | 2021-12-17 | Lighthouse Lab in Milton Keynes | Wellcome Sanger Institute for the COVID-19 Genomics UK (COG-UK) Consortium | The Lighthouse Lab in Milton Keynes and Alex Alderton, Roberto Amato, Jeffrey Barrett, Sonia Goncalves, Ewan Harrison, David K. Jackson, Ian Johnston, Dominic Kwiatkowski, Cordelia Langford, John Sillitoe on behalf of the Wellcome Sanger Institute COVID-1 |
| hCoV-19/India/KL-GENESCOV03210/2021 | EPI_ISL_7221733 | 2021-04-16 | INSACOG-IGIB and Kerala State SARS-COV-2 Genome Surveillance Programme (GENESCOV2) | INSACOG and GENESCOV2 at CSIR Institute of Genomics and Integrative Biology | INSACOG-IGIB and Kerala State SARS-COV-2 Genome Surveillance Programme (GENESCOV2) |
| hCoV-19/Israel/ICH-741107152/2021 | EPI_ISL_8107638 | 2021-12-21 | Clinical Microbiology Laboratory, Tel Aviv Sourasky Medical Center | Clinical Microbiology Laboratory, Tel Aviv Sourasky Medical Center | Katya Levytskyi, Ora Halutz, Alon Ziv, Lior Handler, Amos Adler |
| hCoV-19/India/KL-GENESCOV04048/2021 | EPI_ISL_7223390 | 2021-05-01 | INSACOG-IGIB and Kerala State SARS-COV-2 Genome Surveillance Programme (GENESCOV2) | INSACOG and GENESCOV2 at CSIR Institute of Genomics and Integrative Biology | INSACOG-IGIB and Kerala State SARS-COV-2 Genome Surveillance Programme (GENESCOV2) |
| hCoV-19/Israel/ICH-741107075/2021 | EPI_ISL_8107666 | 2021-12-21 | Clinical Microbiology Laboratory, Tel Aviv Sourasky Medical Center | Clinical Microbiology Laboratory, Tel Aviv Sourasky Medical Center | Katya Levytskyi, Ora Halutz, Alon Ziv, Lior Handler, Amos Adler |

| | | | | | |
|---|---|---|---|---|---|
| hCoV-19/Brazil/SP-IAL-6829/2021 | EPI_ISL_7898709 | 2021-12-03 | Instituto Adolfo Lutz - Regional de Rio Claro | Instituto Adolfo Lutz, Interdiciplinary Procedures Center, Strategic Laboratory | Claudio Tavares Sacchi, Claudia Regina Gonvßalves, Karoline Rodrigues Campos, Caio Vinicius Dias Lopes, Leonardo Tadeu de Araujo, Marlon Benedito Nascimento Santos, Ariadne Ferreira Amarante |
| hCoV-19/India/KL-GENESCOV05755/2021 | EPI_ISL_7225700 | 2021-06-23 | INSACOG-IGIB and Kerala State SARS-COV-2 Genome Surveillance Programme (GENESCOV2) | INSACOG and GENESCOV2 at CSIR Institute of Genomics and Integrative Biology | INSACOG-IGIB and Kerala State SARS-COV-2 Genome Surveillance Programme (GENESCOV2) |
| hCoV-19/India/KL-GENESCOV05870/2021 | EPI_ISL_7225820 | 2021-06-11 | INSACOG-IGIB and Kerala State SARS-COV-2 Genome Surveillance Programme (GENESCOV2) | INSACOG and GENESCOV2 at CSIR Institute of Genomics and Integrative Biology | INSACOG-IGIB and Kerala State SARS-COV-2 Genome Surveillance Programme (GENESCOV2) |
| hCoV-19/India/KL-GENESCOV06223/2021 | EPI_ISL_7226537 | 2021-05-28 | INSACOG-IGIB and Kerala State SARS-COV-2 Genome Surveillance Programme (GENESCOV2) | INSACOG and GENESCOV2 at CSIR Institute of Genomics and Integrative Biology | INSACOG-IGIB and Kerala State SARS-COV-2 Genome Surveillance Programme (GENESCOV2) |
| hCoV-19/India/KL-GENESCOV07626/2021 | EPI_ISL_7226653 | 2021-08-28 | INSACOG-IGIB and Kerala State SARS-COV-2 Genome Surveillance Programme (GENESCOV2) | INSACOG and GENESCOV2 at CSIR Institute of Genomics and Integrative Biology | INSACOG-IGIB and Kerala State SARS-COV-2 Genome Surveillance Programme (GENESCOV2) |
| hCoV-19/India/AP-CCMB-CIB4380/2021 | EPI_ISL_7232621 | 2021-10-11 | AP SSO | CSIR-Centre for Cellular and Molecular Biology-INSACOG | Tulasi Nagabandi, Lamuk Zaveri,Jandhyala Sai Krishna,Malini Nemalikanti,Sreelekshmi MS, Ara Sreenivas,Shreekant Verma, Amareshwar Vodapalli,B Himasri,Valli Nagalakshmi Undamatla, Sumedha Avadhanula,Priya Nurkurthy,Vidhyadhari Methuku, Archana Bharadwaj Si |
| hCoV-19/USA/MI-CDC-ASC210525496/2021 | EPI_ISL_7233678 | 2021-11-07 | Aegis Sciences Corporation | Centers for Disease Control and Prevention Division of Viral Diseases, Pathogen Discovery | Dakota Howard,Dhwani Batra,Peter Cook,Jason Caravas,Benjamin Rambo-Martin,Scott Sammons,Yvette Unoarumhi,Matthew Schmerer,Kristine Lacek,Tymeckia Kendall,Victoria Caban Figueroa,Shatavia Morrison,Christopher Gulvick,Erisa Sula,Cyndi Clark,Patrick Campbell |
| hCoV-19/India/AP-CCMB-CIB4557/2021 | EPI_ISL_7233680 | 2021-10-23 | AP SSO | CSIR-Centre for Cellular and Molecular Biology-INSACOG | Amareshwar Vodapalli,B Himasri,Valli Nagalakshmi Undamatla, Sumedha Avadhanula,Priya Nurkurthy,Vidhyadhari Methuku,Shreekant Verma,Ara Sreenivas, Sreelekshmi MS,Malini Nemalikanti,Lamuk Zaveri,Tulasi Nagabandi,Jandhyala Sai Krishna, Archana Bharadwaj Siv |
| hCoV-19/Kenya/SS4127/2021 | EPI_ISL_7236446 | 2021-11-16 | Kenya Red Cross - IFO Camp | KEMRI-Wellcome Trust Research Programme,Kilifi | Githinji G.,Ochola I., Agoti C.,Mohamed K.S., de Laurent Z., Mburu M.W., Morobe J., Ndwiga L. Makori T., Ongera E., Lambisia A. Nokes D J., Hussein M. |
| hCoV-19/Kenya/C106832/2021 | EPI_ISL_7236700 | 2021-08-11 | KEMRI-Wellcome Trust Research Programme,Kilifi | KEMRI-Wellcome Trust Research Programme,Kilifi | Githinji G.,Ochola I, Agoti C.,Mohamed K.S.,de Laurent Z.,Mburu M.W.,Morobe J.,Lambisia A.,Makori T.,Ongera E., Ndwiga L., D.J.Nokes |
| hCoV-19/Kenya/C74521/2020 | EPI_ISL_7236794 | 2020-11-26 | KEMRI-Wellcome Trust Research Programme,Kilifi | KEMRI-Wellcome Trust Research Programme,Kilifi | Githinji G.,Ochola I, Agoti C.,Mohamed K.S.,de Laurent Z.,Mburu M.W.,Morobe J.,Lambisia A.,Makori T.,Ongera E., Ndwiga L., D.J.Nokes |
| hCoV-19/Israel/SMC-7027265/2021 | EPI_ISL_8111025 | 2021-12-12 | Shamir Medical Center (Asaf Harofe) | Shamir Medical Center (Asaf Harofe) | Nir Rainy,Chen Weiner,Reut Sorek Abramovich,Yevgeni Yegorov,Anna Vishnevsky,Patricia Benveniste-Lekovitz,Abu Hamad Ramzia,Adina Bar Chaim |

| | | | | | |
|---|---|---|---|---|---|
| hCoV-19/USA/MD-CDC-LC0388573/2021 | EPI_ISL_7246525 | 2021-11-13 | Laboratory Corporation of America | Centers for Disease Control and Prevention Division of Viral Diseases, Pathogen Discovery | Dakota Howard,Dhwani Batra,Peter Cook,Jason Caravas,Benjamin Rambo-Martin,Scott Sammons,Yvette Unoarumhi,Matthew Schmerer,Kristine Lacek,Tymeckia Kendall,Victoria Caban Figueroa,Shatavia Morrison,Christopher Gulvick,Minoo Agarwal,Eyad Almasri,Debbie Boles |
| hCoV-19/Israel/SMC-7026730/2021 | EPI_ISL_8110567 | 2021-12-10 | Shamir Medical Center (Asaf Harofe) | Shamir Medical Center (Asaf Harofe) | Nir Rainy,Chen Weiner,Reut Sorek Abramovich,Yevgeni Yegorov,Anna Vishnevsky,Patricia Benveniste-Lekovitz,Abu Hamad Ramzia,Adina Bar Chaim |
| hCoV-19/Israel/CVL-8000512/2021 | EPI_ISL_7133933 | 2021-11-21 | Israel Central Virology laboratory | Israel Central Virology laboratory | Neta Zuckerman, Efrat Dahan Bucris, Michal Mandelboim, Miranda Geva, Omer Asraf, Efrat Glick-Saar, Gideon Rechavi, Oran Erster, Danit Sofer, Ella Mendelson, Orna Mor |
| hCoV-19/USA/AZ-CDC-LC0390626/2021 | EPI_ISL_7247823 | 2021-11-15 | Laboratory Corporation of America | Centers for Disease Control and Prevention Division of Viral Diseases, Pathogen Discovery | Dakota Howard,Dhwani Batra,Peter Cook,Jason Caravas,Benjamin Rambo-Martin,Scott Sammons,Yvette Unoarumhi,Matthew Schmerer,Kristine Lacek,Tymeckia Kendall,Victoria Caban Figueroa,Shatavia Morrison,Christopher Gulvick,Minoo Agarwal,Eyad Almasri,Debbie Boles |
| hCoV-19/Norway/29746/2021 | EPI_ISL_7248821 | 2021-11-29 | Furst Medical Laboratory | Norwegian Institute of Public Health, Department of Virology | Kathrine Stene-Johansen, Kamilla Heddeland Instefjord, Hilde Elshaug, Garcia Llorente Ignacio, Jon BrV•te, Engebretsen Serina Beate, Pedersen Benedikte Nevjen, Line Victoria Moen, Debech Nadia, Atiya R Ali, Marie Paulsen Madsen, Rasmus Riis Kopperud, Hild |
| hCoV-19/Israel/SMC-7026722/2021 | EPI_ISL_8116137 | 2021-12-07 | Shamir Medical Center (Asaf Harofe) | Shamir Medical Center (Asaf Harofe) | Nir Rainy,Chen Weiner,Reut Sorek Abramovich,Yevgeni Yegorov,Anna Vishnevsky,Patricia Benveniste-Lekovitz,Abu Hamad Ramzia,Adina Bar Chaim |
| hCoV-19/India/GJ-INSACOG-GBRC3249/2021 | EPI_ISL_7922707 | 2021-12-04 | Gujarat Adani Institute of Medical Sciences, Bhuj | Gujarat Biotechnology Research Centre | Disha Vora,Janvi Raval,Apurvasinh Puvar,Zarna Patel,Bhadreshsinh Gohil,Nimesh Patel,Nitin Shukla,Ramesh Pandit,Nitin Savaliya,Priyank Chavda,Sonal Sharma ,Roshani Mishra,Hitesh Sudani,Chaitanya Joshi,Madhvi Joshi |
| hCoV-19/Israel/SMC-7026521/2021 | EPI_ISL_8116074 | 2021-12-12 | Shamir Medical Center (Asaf Harofe) | Shamir Medical Center (Asaf Harofe) | Nir Rainy,Chen Weiner,Reut Sorek Abramovich,Yevgeni Yegorov,Anna Vishnevsky,Patricia Benveniste-Lekovitz,Abu Hamad Ramzia,Adina Bar Chaim |
| hCoV-19/Canada/ON-PHL-21-46363/2021 | EPI_ISL_8117886 | 2021-12-07 | Public Health Ontario Laboratory | Public Health Ontario Laboratory | Vanessa G Allen, Philip Banh, Yao Chen, Richard de Borja, Alireza Eshaghi, Nahuel Fittipaldi, Christine Frantz, Jonathan B Gubbay, Jennifer L Guthrie, Lawrence Heisler, Esha Joshi, Michael Laszloffy, Aimin Li, Michael CY Li, Dean Maxwell, Sandeep Nagra, S |
| hCoV-19/Slovenia/21-13030/2021 | EPI_ISL_7931848 | 2021-10-11 | Department of Public Health Microbiology Ljubljana, National Laboratory for Health, Environment and Food | Microbiology Ljubljana, National Laboratory for Health, Environment and Food | Tom Koritnik, Martin Bosilj, Katarina Prosenc, Natasa Berginc, Verica Mioc, Marija Trkov |

| | | | | | |
|---|---|---|---|---|---|
| hCoV-19/USA/NJ-CDC-LC0389088/2021 | EPI_ISL_7252807 | 2021-11-17 | Laboratory Corporation of America | Centers for Disease Control and Prevention Division of Viral Diseases, Pathogen Discovery | Dakota Howard,Dhwani Batra,Peter Cook,Jason Caravas,Benjamin Rambo-Martin,Scott Sammons,Yvette Unoarumhi,Matthew Schmerer,Kristine Lacek,Tymeckia Kendall,Victoria Caban Figueroa,Shatavia Morrison,Christopher Gulvick,Minoo Agarwal,Eyad Almasri,Debbie Boles |
| hCoV-19/USA/OH-CDC-ASC210427505/2021 | EPI_ISL_7934922 | 2021-11-18 | Aegis Sciences Corporation | Centers for Disease Control and Prevention Division of Viral Diseases, Pathogen Discovery | Dakota Howard,Dhwani Batra,Peter Cook,Jason Caravas,Benjamin Rambo-Martin,Scott Sammons,Yvette Unoarumhi,Matthew Schmerer,Kristine Lacek,Tymeckia Kendall,Victoria Caban Figueroa,Shatavia Morrison,Christopher Gulvick,Erisa Sula,Cyndi Clark,Patrick Campbell |
| hCoV-19/USA/CO-CDC-MMB11858717/2021 | EPI_ISL_8121961 | 2021-11-28 | Mako Medical | Centers for Disease Control and Prevention Division of Viral Diseases, Pathogen Discovery | Dakota Howard,Dhwani Batra,Peter Cook,Jason Caravas,Benjamin Rambo-Martin,Scott Sammons,Yvette Unoarumhi,Matthew Schmerer,Kristine Lacek,Tymeckia Kendall,Victoria Caban Figueroa,Shatavia Morrison,Christopher Gulvick,Erisa Sula,Matthew Tugwell,Lauren Moon, |
| hCoV-19/Slovenia/21-12330/2020 | EPI_ISL_7932310 | 2020-10-23 | Department of Public Health Microbiology Ljubljana, National Laboratory for Health, Environment and Food | Microbiology Ljubljana, National Laboratory for Health, Environment and Food | Tom Koritnik, Martin Bosilj, Katarina Prosenc, Natasa Berginc, Verica Mioc, Marija Trkov |
| hCoV-19/USA/CA-CDC-FG-201736/2021 | EPI_ISL_8123114 | 2021-12-18 | Fulgent Genetics | Centers for Disease Control and Prevention Division of Viral Diseases, Pathogen Discovery | Dakota Howard,Dhwani Batra,Peter Cook,Jason Caravas,Benjamin Rambo-Martin,Scott Sammons,Yvette Unoarumhi,Matthew Schmerer,Kristine Lacek,Tymeckia Kendall,Victoria Caban Figueroa,Shatavia Morrison,Christopher Gulvick,Erisa Sula,Harry Gao,Mickey Li,John Gao |
| hCoV-19/Colombia/MAG-UCCSMR-VG520/2021 | EPI_ISL_7938413 | 2021-11-23 | Servicios Medicos Olimpus | Laboratorio de Biologia Molecular, Universidad Cooperativa de Colombia, Santa Marta | Andrew S. Mu√±oz-Gamba, Daniel B. Ram√≠rez-Osorio, Paula A. Quintero-Cort√©s, Pricelis Paulin Polanco Fontalvo, Mayerlin Vasquez Cardenas, Jos√© A. Usme-Ciro |
| hCoV-19/USA/CA-CDC-FG-201164/2021 | EPI_ISL_8123559 | 2021-12-18 | Fulgent Genetics | Centers for Disease Control and Prevention Division of Viral Diseases, Pathogen Discovery | Dakota Howard,Dhwani Batra,Peter Cook,Jason Caravas,Benjamin Rambo-Martin,Scott Sammons,Yvette Unoarumhi,Matthew Schmerer,Kristine Lacek,Tymeckia Kendall,Victoria Caban Figueroa,Shatavia Morrison,Christopher Gulvick,Erisa Sula,Harry Gao,Mickey Li,John Gao |
| hCoV-19/Chile/RM-PUC_MVL_2053/2021 | EPI_ISL_8124688 | 2021-11-22 | Laboratorio de Infectologia y Virologia Molecular | Laboratory of Molecular Virology, School of Medicine, Pontificia Universidad Catolica de Chile | Catalina Pardo-Roa,¬†Leonardo I. Almonacid,¬†Eugenia L. Fuentes Luppichini,¬†Estefany Poblete,¬†¬†Erick Salinas, Andres E. Munoz-Marcos,¬†Jennifer Angulo,¬†Tamara Garcia-Salum, Maria Jose Avendano, ¬†Ana Maria Contreras, Carlos Palma,¬† Jorge Levican, Eil |

| | | | | | |
|---|---|---|---|---|---|
| hCoV-19/SouthAfrica/CERI-KRISP-K033501/2021 | EPI_ISL_8128477 | 2021-11-30 | ZARV/NHLS,¬†Department Medical Virology,¬†University of Pretoria | and Innvoation, Stellenbosch University and KRISP, KZN Research Innovation and Sequencing Platform, UKZN. | Amy Strydom, Adriano Mendes, Micheala Davids, Sim Mayaphi and Marietjie Venter, Giandhari J, Pillay S, Naidoo Y, Ramphal U, Ramphal Y, Arisha Maharaj, San JE, Tshiabuila D, Tegally H, Wilkinson E, de Oliveira T |
| hCoV-19/SouthAfrica/CERI-KRISP-K033493/2021 | EPI_ISL_8128471 | 2021-11-29 | ZARV/NHLS,¬†Department Medical Virology,¬†University of Pretoria | and Innvoation, Stellenbosch University and KRISP, KZN Research Innovation and Sequencing Platform, UKZN. | Amy Strydom, Adriano Mendes, Micheala Davids, Sim Mayaphi and Marietjie Venter, Giandhari J, Pillay S, Naidoo Y, Ramphal U, Ramphal Y, Arisha Maharaj, San JE, Tshiabuila D, Tegally H, Wilkinson E, de Oliveira T |
| hCoV-19/Turkey/HSGM-F10646/2021 | EPI_ISL_8130381 | 2021-12-20 | Ministry of Health Turkey | Ministry of Health Turkey | Fatma Bayrakdar, Yasemin Cosgun, Suleyman Yalcin, Gulay Korukluoglu |
| hCoV-19/Kenya/ILRI_COVC25142/2021 | EPI_ISL_8130637 | 2021-11-04 | International Livestock Research Institute | International Livestock Research Institute | Samuel O. Oyola, Gilbert Kibet, Daniel Ouso, Shebbar Osiany, Edward Kiritu, Paul Dobi, Collins Muli, Patrick Amoth, Vishvanath Nene, Sonal P. Henson, Edward O. Abworo |
| hCoV-19/Kenya/ILRI_COVC25146/2021 | EPI_ISL_8130640 | 2021-10-31 | International Livestock Research Institute | International Livestock Research Institute | Samuel O. Oyola, Gilbert Kibet, Daniel Ouso, Shebbar Osiany, Edward Kiritu, Paul Dobi, Collins Muli, Patrick Amoth, Vishvanath Nene, Sonal P. Henson, Edward O. Abworo |
| hCoV-19/Kenya/ILRI_COVM01669/2021 | EPI_ISL_8130708 | 2021-12-06 | International Livestock Research Institute | International Livestock Research Institute | Samuel O. Oyola, Gilbert Kibet, Daniel Ouso, Shebbar Osiany, Edward Kiritu, Paul Dobi, Collins Muli, Patrick Amoth, Vishvanath Nene, Sonal P. Henson, Edward O. Abworo |
| hCoV-19/Thailand/NIC_KBI_SEQ22771/2021 | EPI_ISL_8132404 | 2021-12-01 | Regional Medical Sciences Center 11/1 Phuket | National Institute of Health, Department of Medical Sciences, Ministry of Public Health, Thailand | Pilailuk Okada; Siripaporn Phuygun; Nuttida Thongpramul; Pakorn Piromtong; Natchaya¬† Khiadsang; Thanutsapa Thanadachakul; Sirikanda Wimol; Suratchana Mitrat; Sunthareeya Waicharoen; Archawin Rojanawiwat |
| hCoV-19/Australia/QLD2691/2021 | EPI_ISL_8132538 | 2021-12-16 | PHV-FSS | PHV-FSS | Chenwei Wang on behalf of Q-PHIRE Genomics |
| hCoV-19/Australia/QLD2878/2021 | EPI_ISL_8132593 | 2021-12-19 | PHV-FSS | PHV-FSS | Chenwei Wang on behalf of Q-PHIRE Genomics |
| hCoV-19/India/MH-ICMR-NIV-INSACOG-GSEQ-7642/2021 | EPI_ISL_8267002 | 2021-12-06 | ICMR-National Institute of Virology - INSACOG | NIV Influenza | Dr. Varsha Potdar and NIC Team |
| hCoV-19/Denmark/DCGC-286556/2021 | EPI_ISL_8138852 | 2021-12-15 | Department of Bacteria, Parasites and Fungi, Statens Serum Institut, Copenhagen, Denmark | Statens Serum Institut Bioinformatics and Microbial Genomics | Danish Covid-19 Genome Consortium |
| hCoV-19/England/MILK-2F96073/2021 | EPI_ISL_8140991 | 2021-12-19 | Lighthouse Lab in Milton Keynes | Wellcome Sanger Institute for the COVID-19 Genomics UK (COG-UK) Consortium | The Lighthouse Lab in Milton Keynes and Alex Alderton, Roberto Amato, Jeffrey Barrett, Sonia Goncalves, Ewan Harrison, David K. Jackson, Ian Johnston, Dominic Kwiatkowski, Cordelia Langford, John Sillitoe on behalf of the Wellcome Sanger Institute COVID-1 |
| hCoV-19/USA/DC-DFS-PHL-03511/2021 | EPI_ISL_8141654 | 2021-11-19 | DC Public Health Lab/ Dept. of Forensic Sciences | DC Public Health Lab/ Dept. of Forensic Sciences | Janis Doss, Scott Nguyen, Elizabeth Zelaya, Eric Vaughn, Sarah Scott, Nathan Bruns, Wadih Bchara, Connie Maza, Monica Mann, Brittany Hamilton, Jocelyn Hauser |
| hCoV-19/Israel/SMC-7028547/2021 | EPI_ISL_8136503 | 2021-12-17 | Shamir Medical Center (Asaf Harofe) | Shamir Medical Center (Asaf Harofe) | Nir Rainy,Chen Weiner,Reut Sorek Abramovich,Yevgeni Yegorov,Anna Vishnevsky,Patricia Benveniste-Lekovitz,Abu Hamad Ramzia,Adina Bar Chaim |

| | | | | | |
|---|---|---|---|---|---|
| hCoV-19/USA/FL-BPHL-16969/2021 | EPI_ISL_8145129 | 2021-11-14 | Florida Bureau of Public Health Laboratories | Florida Bureau of Public Health Laboratories | Sarah Schmedes, Namratha Tarigopula, Tiffany Splatt, Jason Blanton |
| hCoV-19/CzechRepublic/FNHK-Ps-001550/2021 | EPI_ISL_8143494 | 2021-11-26 | University Hospital Hradec Kralove | University Hospital Hradec Kralove | Marketa Gancarcikova, Helena Kovarikova, Lenka Rysava, Monika Berankova |
| hCoV-19/India/MH-RFIP05423/2021 | EPI_ISL_8145430 | 2021-10-04 | BJMC, Pune | RF-IISER Pune | Krishanpal Karmodiya, Aurnab Ghose, Joy Merwin Monteiro, Suvarna Joshi, Rajesh Karyakarte |
| hCoV-19/India/MH-RFIP05470/2021 | EPI_ISL_8145473 | 2021-10-07 | BJMC, Pune | RF-IISER Pune | Krishanpal Karmodiya, Aurnab Ghose, Joy Merwin Monteiro, Suvarna Joshi, Rajesh Karyakarte |
| hCoV-19/India/MH-RFIP05612/2021 | EPI_ISL_8145448 | 2021-10-19 | BJMC, Pune | RF-IISER Pune | Krishanpal Karmodiya, Aurnab Ghose, Joy Merwin Monteiro, Suvarna Joshi, Rajesh Karyakarte |
| hCoV-19/England/ALDP-2FE8546/2021 | EPI_ISL_8275415 | 2021-12-24 | Lighthouse Lab in Alderley Park | Wellcome Sanger Institute for the COVID-19 Genomics UK (COG-UK) Consortium | Jacquelyn Wynn, Mairead Hyland, The Lighthouse Lab in Alderley Park and Alex Alderton, Roberto Amato, Jeffrey Barrett, Sonia Goncalves, Ewan Harrison, David K. Jackson, Ian Johnston, Dominic Kwiatkowski, Cordelia Langford, John Sillitoe on behalf of the W |
| hCoV-19/Israel/ICH-741105488/2021 | EPI_ISL_7952960 | 2021-12-14 | Clinical Microbiology Laboratory, Tel Aviv Sourasky Medical Center | Clinical Microbiology Laboratory, Tel Aviv Sourasky Medical Center | Katya Levytskyi, Ora Halutz, Alon Ziv, Lior Handler, Amos Adler |
| hCoV-19/USA/CA-CDC-FG-206186/2021 | EPI_ISL_8283102 | 2021-12-21 | Fulgent Genetics | Centers for Disease Control and Prevention Division of Viral Diseases, Pathogen Discovery | Dakota Howard,Dhwani Batra,Peter Cook,Jason Caravas,Benjamin Rambo-Martin,Scott Sammons,Yvette Unoarumhi,Matthew Schmerer,Kristine Lacek,Tymeckia Kendall,Victoria Caban Figueroa,Shatavia Morrison,Christopher Gulvick,Erisa Sula,Harry Gao,Mickey Li,John Gao |
| hCoV-19/USA/CA-CDC-FG-208935/2021 | EPI_ISL_8285118 | 2021-12-22 | Fulgent Genetics | Centers for Disease Control and Prevention Division of Viral Diseases, Pathogen Discovery | Dakota Howard,Dhwani Batra,Peter Cook,Jason Caravas,Benjamin Rambo-Martin,Scott Sammons,Yvette Unoarumhi,Matthew Schmerer,Kristine Lacek,Tymeckia Kendall,Victoria Caban Figueroa,Shatavia Morrison,Christopher Gulvick,Erisa Sula,Harry Gao,Mickey Li,John Gao |
| hCoV-19/India/KA-RFNB-1222/2021 | EPI_ISL_8285498 | 2021-11-22 | VASANTHANAGAR UPHC/MOLECULAR SOLUTIONS CARE HEALTH LLP BANGALORE | National Center for Biological Sciences, TIFR - Rockefeller Foundation | Darshan Sreenivas,Bhagyashree Madhav Shelar,Lakshminarayanan CP,Manisha Bharadwaj,Awadhesh Pandit,Satyajit Mayor,Uma Ramakrishnan,Dimple Notani |
| hCoV-19/Canada/ON-PHL-21-41648/2021 | EPI_ISL_7259710 | 2021-11-30 | Public Health Ontario Laboratory | Public Health Ontario Laboratory | Vanessa G Allen, Philip Banh, Yao Chen, Richard de Borja, Alireza Eshaghi, Nahuel Fittipaldi, Christine Frantz, Jonathan B Gubbay, Jennifer L Guthrie, Lawrence Heisler, Esha Joshi, Michael Laszloffy, Aimin Li, Michael CY Li, Dean Maxwell, Sandeep Nagra, S |
| hCoV-19/Canada/ON-PHL-21-41649/2021 | EPI_ISL_7259716 | 2021-11-30 | Public Health Ontario Laboratory | Public Health Ontario Laboratory | Vanessa G Allen, Philip Banh, Yao Chen, Richard de Borja, Alireza Eshaghi, Nahuel Fittipaldi, Christine Frantz, Jonathan B Gubbay, Jennifer L Guthrie, Lawrence Heisler, Esha Joshi, Michael Laszloffy, Aimin Li, Michael CY Li, Dean Maxwell, Sandeep Nagra, S |

| | | | | | |
|---|---|---|---|---|---|
| hCoV-19/Netherlands/NH-AUMC-015395/2021 | EPI_ISL_7263925 | 2021-11-26 | Public Health Laboratory, Public Health Service Amsterdam, The Netherlands | Department of Medical Microbiology & Infection prevention, Amsterdam University Medical Centers location AMC | Matthijs Welkers, Jelle Koopsen, Robin van Houdt, Marcel Jonges, Sebastien Matamoros, Sjoerd Rebers, Fokla Zorgdrager, Sylvia Bruisten, Judith den Uil, Akke Cornelissen, Janke Schinkel, Menno de Jong, Tjalling Leenstra and Mariken van der Lubben on behalf |
| hCoV-19/USA/HI-H216115/2021 | EPI_ISL_7264087 | 2021-11-30 | State Laboratories Division, Hawaii State Department of Health | State Laboratories Division, Hawaii State Department of Health | Pamela O'Brien, Drew Kuwazaki, Ayana Garnet, Daniel Strange, Razvan Sultana, Edward Desmond |
| hCoV-19/Australia/NSW-ICPMR-14787/2021 | EPI_ISL_7264297 | 2021-11-20 | St Vincent's Pathology (SydPath) | Clinical Pathology and Medical Research; Westmead Hospital; University of Sydney | Arnott A., Draper J., Gall M., Martinez E., Rockett R., Sintchenko V., on behalf of ICPMR |
| hCoV-19/Australia/NSW-ICPMR-15280/2021 | EPI_ISL_7264797 | 2021-11-27 | Pathology West - NSW Health Pathology | Clinical Pathology and Medical Research; Westmead Hospital; University of Sydney | Arnott A., Draper J., Gall M., Martinez E., Rockett R., Sintchenko V., on behalf of ICPMR |
| hCoV-19/Australia/NSW-ICPMR-15464/2021 | EPI_ISL_7264992 | 2021-11-29 | Pathology West - NSW Health Pathology | Clinical Pathology and Medical Research; Westmead Hospital; University of Sydney | Arnott A., Draper J., Gall M., Martinez E., Rockett R., Sintchenko V., on behalf of ICPMR |
| hCoV-19/Australia/NSW-ICPMR-15554/2021 | EPI_ISL_7265083 | 2021-11-30 | Pathology North - Royal North Shore Hospital - NSW Health Pathology | Clinical Pathology and Medical Research; Westmead Hospital; University of Sydney | Arnott A., Draper J., Gall M., Martinez E., Rockett R., Sintchenko V., on behalf of ICPMR |
| hCoV-19/Canada/ON-NML-255881/2021 | EPI_ISL_7265455 | 2021-11-26 | Alpha Labs | National Microbiology Laboratory (NML) | Anna Majer, Shari Tyson, Grace Seo, Philip Mabon, Elsie Grudeski, Rhiannon Huzarewich, Russell Mandes, Anneliese Landgraff, Jennifer Tanner, Natalie Knox, Morag Graham, Gary Van Domselaar, Gordon Jolly, Nathalie Bastien, Yan Li, Timothy Booth, Darian Hole |
| hCoV-19/Israel/ICH-760596828/2021 | EPI_ISL_7265960 | 2021-11-29 | Clinical Microbiology Laboratory, Tel Aviv Sourasky Medical Center | Clinical Microbiology Laboratory, Tel Aviv Sourasky Medical Center | Katya Levytskyi, Ora Halutz, Alon Ziv, Lior Handler, Amos Adler |
| hCoV-19/Israel/ICH-760596975/2021 | EPI_ISL_7266104 | 2021-11-29 | Clinical Microbiology Laboratory, Tel Aviv Sourasky Medical Center | Clinical Microbiology Laboratory, Tel Aviv Sourasky Medical Center | Katya Levytskyi, Ora Halutz, Alon Ziv, Lior Handler, Amos Adler |
| hCoV-19/Norway/29463/2021 | EPI_ISL_7267259 | 2021-11-29 | Medical Microbiology Unit, Department for Laboratory Medicine, Drammen Hospital, Vestre Viken Health Trust | Norwegian Institute of Public Health, Department of Virology | Kathrine Stene-Johansen, Kamilla Heddeland Instefjord, Hilde Elshaug, Garcia Llorente Ignacio, Jon Br√•te, Engebretsen Serina Beate, Pedersen Benedikte Nevjen, Line Victoria Moen, Debech Nadia, Atiya R Ali, Marie Paulsen Madsen, Rasmus Riis Kopperud, Hild |
| hCoV-19/USA/UT-UPHL-211125979401/2021 | EPI_ISL_7268926 | 2021-09-10 | Utah Public Health Laboratory | Utah Public Health Laboratory | Erin L. Young, Kelly F. Oakeson, Pooja Gupta, John Arnn, Tom Iverson, Olinto Linares-Perdomo |
| hCoV-19/USA/NV-NSPHL-480877/2021 | EPI_ISL_7271657 | 2021-11-30 | Nevada State Public Health Laboratory | Nevada State Public Health Laboratory | Andrew Gorzalski, Lynette Gumbleton, Mark Pandori |
| hCoV-19/Denmark/DCGC-247557/2021 | EPI_ISL_7280348 | 2021-11-25 | Department of Bacteria, Parasites and Fungi, Statens Serum Institut, Copenhagen, Denmark | Statens Serum Institut Bioinformatics and Microbial Genomics | Danish Covid-19 Genome Consortium |
| hCoV-19/France/CVL-HMN-21112040324/2021 | EPI_ISL_7290436 | 2021-10-26 | BPR Analyses sp√©cialis√©es | Mondor University Hospital, Assistance Publique H√¥pitaux de Paris, Universit√© Paris-Est Cr√©teil, INSERM U955 | Christophe Rodriguez, Slim Fourati, Vanessa Demontant, Guillaume Gricourt, Melissa N'Debi, Alexandre Soulier, Elisabeth Trawinski, Jean-Michel Pawlotsky |

| | | | | | |
|---|---|---|---|---|---|
| hCoV-19/England/QEUH-2CA9FDD/2021 | EPI_ISL_7296139 | 2021-11-28 | Lighthouse Lab in Glasgow | Wellcome Sanger Institute for the COVID-19 Genomics UK (COG-UK) Consortium | Harper VanSteenhouse, Yumi Kasai, David Gray, Carol Clugston, Anna Dominiczak and Alex Alderton, Roberto Amato, Jeffrey Barrett, Sonia Goncalves, Ewan Harrison, David K. Jackson, Ian Johnston, Dominic Kwiatkowski, Cordelia Langford, John Sillitoe on behal |
| hCoV-19/Scotland/QEUH-2C8EB34/2021 | EPI_ISL_7299536 | 2021-11-30 | Lighthouse Lab in Glasgow | Wellcome Sanger Institute for the COVID-19 Genomics UK (COG-UK) Consortium | Harper VanSteenhouse, Yumi Kasai, David Gray, Carol Clugston, Anna Dominiczak and Alex Alderton, Roberto Amato, Jeffrey Barrett, Sonia Goncalves, Ewan Harrison, David K. Jackson, Ian Johnston, Dominic Kwiatkowski, Cordelia Langford, John Sillitoe on behal |
| hCoV-19/Scotland/QEUH-2C8C3E3/2021 | EPI_ISL_7299709 | 2021-11-30 | Lighthouse Lab in Glasgow | Wellcome Sanger Institute for the COVID-19 Genomics UK (COG-UK) Consortium | Harper VanSteenhouse, Yumi Kasai, David Gray, Carol Clugston, Anna Dominiczak and Alex Alderton, Roberto Amato, Jeffrey Barrett, Sonia Goncalves, Ewan Harrison, David K. Jackson, Ian Johnston, Dominic Kwiatkowski, Cordelia Langford, John Sillitoe on behal |
| hCoV-19/Scotland/QEUH-2C8C29B/2021 | EPI_ISL_7299880 | 2021-11-30 | Lighthouse Lab in Glasgow | Wellcome Sanger Institute for the COVID-19 Genomics UK (COG-UK) Consortium | Harper VanSteenhouse, Yumi Kasai, David Gray, Carol Clugston, Anna Dominiczak and Alex Alderton, Roberto Amato, Jeffrey Barrett, Sonia Goncalves, Ewan Harrison, David K. Jackson, Ian Johnston, Dominic Kwiatkowski, Cordelia Langford, John Sillitoe on behal |
| hCoV-19/England/MILK-2C9742E/2021 | EPI_ISL_7301307 | 2021-11-29 | Lighthouse Lab in Milton Keynes | Wellcome Sanger Institute for the COVID-19 Genomics UK (COG-UK) Consortium | The Lighthouse Lab in Milton Keynes and Alex Alderton, Roberto Amato, Jeffrey Barrett, Sonia Goncalves, Ewan Harrison, David K. Jackson, Ian Johnston, Dominic Kwiatkowski, Cordelia Langford, John Sillitoe on behalf of the Wellcome Sanger Institute COVID-1 |
| hCoV-19/USA/DE-DHSS-B1135591/2021 | EPI_ISL_7307888 | 2021-09-29 | DPHL | Delaware Public Health Lab | Holly Miller, Moneeb Bajwa |
| hCoV-19/SouthAfrica/NHLS-UCT-GS-D074/2021 | EPI_ISL_7310240 | 2021-06-27 | Groote Schuur Hospital wc GSH | NHLS/UCT | Arash Iranzadeh, Deelan Doolabh, Lynn Tyers, Bruna Galvao, Innocent Mudau, Marvin Hsiao, Gert Marais, Diana Hardie, Stephen Korsman, Rageema Joseph, Luicer Olubayo, Carolyn Williamson |
| hCoV-19/USA/FL-BPHL-16914/2021 | EPI_ISL_7313687 | 2021-11-26 | Florida Bureau of Public Health Laboratories | Florida Bureau of Public Health Laboratories | Sarah Schmedes, Namratha Tarigopula, Tiffany Splatt, Jason Blanton |
| hCoV-19/Nigeria/CV3743/2021 | EPI_ISL_7324917 | 2021-02-03 | Genexpert Molecular Diagnostic Laboratory | Africa Centre for Excellence for Genomics of Infectious Diseases (ACEGID), Redeemer,Äôs University | Olawoye, I.B., Oluniyi, P.E., Eromon, P.E., Oguzie, J.U., Kayode, A.T., Uwanibe, J.N., Ugwu, C.A., Akano, K.O., Ajogbasile, F.V., Abechi, P.S., Olumade, T.J., Nosamiefan, I., Folarin, O., Happi, C.T. |
| hCoV-19/USA/IA-SHL-1946552/2021 | EPI_ISL_7325750 | 2021-11-24 | State Hygienic Laboratory at the University of Iowa | State Hygienic Laboratory at the University of Iowa | Valerie Reeb, Erik Twait, Anna Yakos, Mariah Knutson, Kris Eveland, Kristen Zanon, Jeff Benfer, Davis Rieckenberg, Cindy Toll, Wes Hottel, Alankar Kampoowale, Krishnaveni Sompallae, Mohammed Allam |
| hCoV-19/USA/NC-ECU-CORVASEQ-110475299/2021 | EPI_ISL_7329420 | 2021-10-27 | Molecular Biology Laboratory, Vidant Medical Center | Brody Integrative Genomics Core, East Carolina University | Weihua Huang, Changhong Yin, Kimberly P. Briley, Heather Duncan, John T. Fallon |

| | | | | | |
|---|---|---|---|---|---|
| hCoV-19/USA/CO-CDPHE-2101935858/2021 | EPI_ISL_7334349 | 2021-10-07 | Colorado Department of Public Health and Environment | Colorado Department of Public Health and Environment | Laura Bankers, Molly C. Hetherington-Rauth, Diana Ir, Alexandria Rossheim, Michael Martin, Mandy Waters, Arianna Smith, Shannon R. Matzinger, Sarah Elizabeth Totten, Emily A. Travanty |
| hCoV-19/USA/NY-Wadsworth-21103395-01/2021 | EPI_ISL_7334890 | 2021-11-30 | SUNRISE MEDICAL LABORATORIES | Wadsworth Center, New York State Department of Health | Kirsten St. George, Daryl M. Lamson, Alexis Russell, Matthew Shudt, Melissa A Leisner, Jonathan Plitnick, Catharine Prussing, Navjot Singh, John Kelly, Erasmus Schneider, Erica Lasek-Nesselquist |
| hCoV-19/USA/CA-CDPH-3000033569/2021 | EPI_ISL_7335711 | 2021-04-26 | CDPH VBL | California Department of Public Health | Emily Smith on behalf of CDPH-COVIDNet |
| hCoV-19/SouthAfrica/NICD-N21295/2021 | EPI_ISL_7337387 | 2021-09-24 | Kopanong Laboratory | Diseases of the National Health Laboratory Service | Amoako DG, Everatt J, Scheepers C, Mohale T, Ntuli N, Mahlangu B, Mnguni A, Ismail A, Bhiman JN |
| hCoV-19/SouthAfrica/NICD-N21696/2021 | EPI_ISL_7337442 | 2021-11-18 | Helen Joseph Laboratory | Diseases of the National Health Laboratory Service | Amoako DG, Everatt J, Scheepers C, Mohale T, Ntuli N, Mahlangu B, Mnguni A, Ismail A, Bhiman JN |
| hCoV-19/SouthAfrica/NICD-N21712/2021 | EPI_ISL_7337456 | 2021-11-19 | Helen Joseph Laboratory | Diseases of the National Health Laboratory Service | Amoako DG, Everatt J, Scheepers C, Mohale T, Ntuli N, Mahlangu B, Mnguni A, Ismail A, Bhiman JN |
| hCoV-19/SouthAfrica/NICD-N21783/2021 | EPI_ISL_7337526 | 2021-11-23 | Chris Hani Baragwanath Laboratory | Diseases of the National Health Laboratory Service | Amoako DG, Everatt J, Scheepers C, Mohale T, Ntuli N, Mahlangu B, Mnguni A, Ismail A, Bhiman JN |
| hCoV-19/SouthAfrica/NICD-N21787/2021 | EPI_ISL_7337530 | 2021-11-24 | Chris Hani Baragwanath Laboratory | Diseases of the National Health Laboratory Service | Amoako DG, Everatt J, Scheepers C, Mohale T, Ntuli N, Mahlangu B, Mnguni A, Ismail A, Bhiman JN |
| hCoV-19/SouthAfrica/NICD-N01329/2021 | EPI_ISL_7337568 | 2021-01-04 | Port Elizabeth Laboratory | Diseases of the National Health Laboratory Service | Amoako DG, Everatt J, Scheepers C, Mohale T, Ntuli N, Mahlangu B, Mnguni A, Ismail A, Bhiman JN |
| hCoV-19/Belgium/UGent-12012/2021 | EPI_ISL_7339434 | 2021-11-29 | Lab voor klinische biologie | Lab voor klinische biologie | May-Linh Truong, Marija Janevska, Hannelore Hamerlinck, Bruno Verhasselt |
| hCoV-19/England/MILK-2CC670F/2021 | EPI_ISL_7340046 | 2021-11-30 | Lighthouse Lab in Milton Keynes | Wellcome Sanger Institute for the COVID-19 Genomics UK (COG-UK) Consortium | The Lighthouse Lab in Milton Keynes and Alex Alderton, Roberto Amato, Jeffrey Barrett, Sonia Goncalves, Ewan Harrison, David K. Jackson, Ian Johnston, Dominic Kwiatkowski, Cordelia Langford, John Sillitoe on behalf of the Wellcome Sanger Institute COVID-1 |
| hCoV-19/England/ALDP-2CD1DC3/2021 | EPI_ISL_7344677 | 2021-11-30 | Lighthouse Lab in Alderley Park | Wellcome Sanger Institute for the COVID-19 Genomics UK (COG-UK) Consortium | Jacquelyn Wynn, Mairead Hyland, The Lighthouse Lab in Alderley Park and Alex Alderton, Roberto Amato, Jeffrey Barrett, Sonia Goncalves, Ewan Harrison, David K. Jackson, Ian Johnston, Dominic Kwiatkowski, Cordelia Langford, John Sillitoe on behalf of the W |
| hCoV-19/England/BRBR-2CBFCA0/2021 | EPI_ISL_7347522 | 2021-11-30 | Berkshire and Surrey Pathology Services Lighthouse Laboratory | Wellcome Sanger Institute for the COVID-19 Genomics UK (COG-UK) Consortium | Berkshire and Surrey Pathology Services Lighthouse Laboratory and Alex Alderton, Roberto Amato, Jeffrey Barrett, Sonia Goncalves, Ewan Harrison, David K. Jackson, Ian Johnston, Dominic Kwiatkowski, Cordelia Langford, John Sillitoe on behalf of the Wellcom |
| hCoV-19/Scotland/QEUH-2CBC3EA/2021 | EPI_ISL_7356256 | 2021-11-30 | Lighthouse Lab in Glasgow | Wellcome Sanger Institute for the COVID-19 Genomics UK (COG-UK) Consortium | Harper VanSteenhouse, Yumi Kasai, David Gray, Carol Clugston, Anna Dominiczak and Alex Alderton, Roberto Amato, Jeffrey Barrett, Sonia Goncalves, Ewan Harrison, David K. Jackson, Ian Johnston, Dominic Kwiatkowski, Cordelia Langford, John Sillitoe on behal |
| hCoV-19/HongKong/HKU-344/2021 | EPI_ISL_7357684 | 2021-11-27 | Department of Microbiology, The University of Hong Kong | Department of Microbiology, The University of Hong Kong | Kelvin K.W. To, Kwok-Yung Yuen |

| | | | | | |
|---|---|---|---|---|---|
| hCoV-19/SouthAfrica/CERI-KRISP-K032712/2021 | EPI_ISL_7358064 | 2021-11-30 | National Health Laboratory Service, Kwazulu-Natal, South Africa | and Innvoation, Stellenbosch University and KRISP, KZN Research Innovation and Sequencing Platform, UKZN. | Nokukhanya Mdlalose, Giandhari J, Pillay S, Naidoo Y, Ramphal U, Ramphal Y, Moir M, San JE, Tshiabuila D, Tegally H, Wilkinson E, de Oliveira T |
| hCoV-19/Denmark/DCGC-251958/2021 | EPI_ISL_7359618 | 2021-12-02 | Department of Bacteria, Parasites and Fungi, Statens Serum Institut, Copenhagen, Denmark | Statens Serum Institut Bioinformatics and Microbial Genomics | Danish Covid-19 Genome Consortium |
| hCoV-19/Canada/BC-BCCDC-263493/2021 | EPI_ISL_7370157 | 2021-11-29 | British Columbia Centre For Disease Control | BCCDC Public Health Laboratory | Prystajecky Natalie, Linda Hoang, Dan Fornika, John Tyson, Shannon Russell, Kim Macdonald, Kimia Kamelian, Ana Pacagnella, Corrinne Ng, Loretta Janz, Robert Azana, Mel Krajden |
| hCoV-19/India/DL-ILBS-WGS584/2021 | EPI_ISL_7379386 | 2021-11-15 | HOUSE OF DIAGNOSTICS | The Institute of Liver and Biliary Sciences (ILBS) | Ekta Gupta, Varun Suroliya, Pramod Gautam, Reshu Agarwal, Rahul Garg, Urvinder Kaur, Santanu Das, Arjun Bhugra, Chhagan Bihari Sharma, Shiv Kumar Sarin |
| hCoV-19/Georgia/Tb-SNGS1950/2021 | EPI_ISL_7380565 | 2021-11-27 | Department for Virology, Molecular Biology and Genome Research, R. G. Lugar Center for Public Health Research, National Center for Disease Control and Public Health (NCDC) of Georgia. | Biology and Genome Research, R. G. Lugar Center for Public Health Research, National Center for Disease Control and Public Health (NCDC) of Georgia. | Giorgi Tomashvili, Giorgi Gogoladze, Meri Pantsulaia, Gvantsa Brachveli, Salome Javashvili, Nino Berishvili, Tata Imnadze, Ana Papkiauri, Gvantsa Chanturia, Ann Machablishvili, Nato Kotaria, Marine Murtskhvaladze, Lela Sabadze, Mari Gavashelidze, Tamar Ja |
| hCoV-19/France/PAC-IHUvariant_49511/2021 | EPI_ISL_7381062 | 2021-11-22 | IHU Mediterranee Infection | IHU Mediterranee Infection | Philippe Colson et al. |
| hCoV-19/India/DL-ICMR-MCL_21_11514_7858/2021 | EPI_ISL_7381102 | 2021-11-23 | Indian Council of Medical Research-National Institute of Virology,¬†Microbial Containment Complex | National Institute of Virology,¬†Microbial Containment Complex | Pragya D. Yadav |
| hCoV-19/SouthAfrica/CERI-KRISP-K032772/2021 | EPI_ISL_7381208 | 2021-12-02 | NHLS Port Elizabeth Laboratory | and Innvoation, Stellenbosch University and KRISP, KZN Research Innovation and Sequencing Platform, UKZN. | Oluwakemi Laguda-Akingba and Nokukhanya Mdlalose, Giandhari J, Pillay S, Naidoo Y, Ramphal U, Ramphal Y, Arisha Maharaj, van Wyk S, Moir M, San JE, Tshiabuila D, Tegally H, Wilkinson E, de Oliveira T |
| hCoV-19/HongKong/HKU-211209-001/2021 | EPI_ISL_7385702 | 2021-11-27 | Department of Microbiology, The University of Hong Kong | Department of Microbiology, The University of Hong Kong | Kelvin K.W. To, Kwok-Yung Yuen |
| hCoV-19/USA/NH-CDCBI-CRSP_HWFPDCEGVL5ND6V6/2021 | EPI_ISL_7386466 | 2021-11-27 | Broad Institute Clinical Research Sequencing Platform | Infectious Disease Program, Broad Institute of Harvard and MIT | Siddle,K.J., Adams,G., Pearlman,L., Gladden-Young,A., Vicente,G., Blumenstiel,B., DeFelice,M., Lee,M., McGovern,S., Lagerborg,K., Rudy,M., DeRuff,K., Carter,A., Normandin,E., Bauer,M., Reilly,S., Tomkins-Tinch,C., Loreth,C., Chaluvadi,S., Meldrim,J., Gran |
| hCoV-19/England/QEUH-2CF52AC/2021 | EPI_ISL_7389585 | 2021-11-30 | Lighthouse Lab in Glasgow | Wellcome Sanger Institute for the COVID-19 Genomics UK (COG-UK) Consortium | Harper VanSteenhouse, Yumi Kasai, David Gray, Carol Clugston, Anna Dominiczak and Alex Alderton, Roberto Amato, Jeffrey Barrett, Sonia Goncalves, Ewan Harrison, David K. Jackson, Ian Johnston, Dominic Kwiatkowski, Cordelia Langford, John Sillitoe on behal |
| hCoV-19/England/MILK-2CD7FD6/2021 | EPI_ISL_7396628 | 2021-11-30 | Lighthouse Lab in Milton Keynes | Wellcome Sanger Institute for the COVID-19 Genomics UK (COG-UK) Consortium | The Lighthouse Lab in Milton Keynes and Alex Alderton, Roberto Amato, Jeffrey Barrett, Sonia Goncalves, Ewan Harrison, David K. Jackson, Ian Johnston, Dominic Kwiatkowski, Cordelia Langford, John Sillitoe on behalf of the Wellcome Sanger Institute COVID-1 |

| | | | | | |
|---|---|---|---|---|---|
| hCoV-19/Scotland/QEUH-2C95CCE/2021 | EPI_ISL_7397103 | 2021-11-30 | Lighthouse Lab in Glasgow | Wellcome Sanger Institute for the COVID-19 Genomics UK (COG-UK) Consortium | Harper VanSteenhouse, Yumi Kasai, David Gray, Carol Clugston, Anna Dominiczak and Alex Alderton, Roberto Amato, Jeffrey Barrett, Sonia Goncalves, Ewan Harrison, David K. Jackson, Ian Johnston, Dominic Kwiatkowski, Cordelia Langford, John Sillitoe on behal |
| hCoV-19/Thailand/NIC_BKK_SEQ22356/2021 | EPI_ISL_7398681 | 2021-11-24 | Vichaivej International Hospital Nongkhaem | National Institute of Health, Department of Medical Sciences, Ministry of Public Health, Thailand | Pilailuk Akkapaiboon Okada, Piroon Jenjaroenpun, Pongpun Sawatwong, Thidathip Wongsurawat, Siripaporn Phuygun, Suratchana Mitrat, Sirikanda Wimol, Nuttida Thongpramul, Kazuhisa Okada, Donlaya Maunplueng, Thanutsapa Thanadachakul, Pakorn Piromtong, Natchay |
| hCoV-19/England/PHEP-YYN8QFA/2021 | EPI_ISL_7399905 | 2021-11-16 | Respiratory Virus Unit, Microbiology Services Colindale, Public Health England | COVID-19 Genomics UK (COG-UK) Consortium | PHE Covid Sequencing Team |
| hCoV-19/Nepal/NPHL/S-253/2021 | EPI_ISL_7405756 | 2021-11-20 | Dhaulagiri Hospital PCR Lab | National Public Health Laboratory | National Public Health Laboratory Team, Dhaulagiri Hospital COVID PCR Lab Team |
| hCoV-19/USA/WV-CDC-ASC210398851/2021 | EPI_ISL_7410082 | 2021-11-12 | Aegis Sciences Corporation | Centers for Disease Control and Prevention Division of Viral Diseases, Pathogen Discovery | Dakota Howard,Dhwani Batra,Peter Cook,Jason Caravas,Benjamin Rambo-Martin,Scott Sammons,Yvette Unoarumhi,Matthew Schmerer,Kristine Lacek,Tymeckia Kendall,Victoria Caban Figueroa,Shatavia Morrison,Christopher Gulvick,Erisa Sula,Cyndi Clark,Patrick Campbell |
| hCoV-19/USA/TX-CDC-ASC210487088/2021 | EPI_ISL_7412262 | 2021-11-19 | Aegis Sciences Corporation | Centers for Disease Control and Prevention Division of Viral Diseases, Pathogen Discovery | Dakota Howard,Dhwani Batra,Peter Cook,Jason Caravas,Benjamin Rambo-Martin,Scott Sammons,Yvette Unoarumhi,Matthew Schmerer,Kristine Lacek,Tymeckia Kendall,Victoria Caban Figueroa,Shatavia Morrison,Christopher Gulvick,Erisa Sula,Cyndi Clark,Patrick Campbell |
| hCoV-19/USA/TX-CDC-ASC210487089/2021 | EPI_ISL_7412265 | 2021-11-19 | Aegis Sciences Corporation | Centers for Disease Control and Prevention Division of Viral Diseases, Pathogen Discovery | Dakota Howard,Dhwani Batra,Peter Cook,Jason Caravas,Benjamin Rambo-Martin,Scott Sammons,Yvette Unoarumhi,Matthew Schmerer,Kristine Lacek,Tymeckia Kendall,Victoria Caban Figueroa,Shatavia Morrison,Christopher Gulvick,Erisa Sula,Cyndi Clark,Patrick Campbell |
| hCoV-19/Gibraltar/EXP_01122021-1584256-80/2021 | EPI_ISL_7415145 | 2021-11-30 | Gibraltar Health Authority Lab | Gibraltar Health Authority Covid-19 Laboratory | Dr Nicholas Cortes, Dr Martyn Bell, Dr Zoe Vincent, Dr Daniel Cassaglia, Bruna Martins, Sofia Lavelle |
| hCoV-19/SouthAfrica/CERI-KRISP-K027906/2021 | EPI_ISL_7417255 | 2021-10-28 | National Health Laboratory Service, Kwazulu-Natal, South Africa | and Innvoation, Stellenbosch University and KRISP, KZN Research Innovation and Sequencing Platform, UKZN. | Nokukhanya Mdlalose, Giandhari J, Pillay S, Naidoo Y, Ramphal U, Ramphal Y, Moir M, San JE, Tshiabuila D, Tegally H, Wilkinson E, de Oliveira T |
| hCoV-19/SouthAfrica/CERI-KRISP-K027929/2021 | EPI_ISL_7417285 | 2021-11-03 | National Health Laboratory Services, Virology, Charlotte Maxeke Johannesburg hospital, Parktown, Johannesburg, Gauteng | and Innvoation, Stellenbosch University and KRISP, KZN Research Innovation and Sequencing Platform, UKZN. | Kathleen Subramoney, Florette Treurnicht, Giandhari J, Pillay S, Naidoo Y, Ramphal U, Ramphal Y, Moir M, San JE, Tshiabuila D, Tegally H, Wilkinson E, de Oliveira T |

| | | | | | |
|---|---|---|---|---|---|
| hCoV-19/Japan/TY38-873P0/2021 | EPI_ISL_7418017 | 2021-11-28 | National Institute of Infectious Diseases | National Institute of Infectious Diseases | Shun Iida, Harutaka Katano, Tadaki Suzuki, Yudai Kuroda, Ken Maeda, Shuetsu Fukushi, Tsuyoshi Sekizuka, Kentaro Itokawa, Makoto Kuroda |
| hCoV-19/Kenya/SS1408/2021 | EPI_ISL_7437759 | 2021-08-12 | Tigoni COVID-19 Centre | KEMRI-Wellcome Trust Research Programme,Kilifi | Githinji G.,Ochola I., Agoti C.,Mohamed K.S., de Laurent Z., Mburu M.W., Morobe J., Ndwiga L. Makori T., Ongera E., Lambisia A. Nokes D J., Njoki K. |
| hCoV-19/USA/MN-CDC-IBX614317678369/2021 | EPI_ISL_7444020 | 2021-11-23 | Infinity Biologix | Centers for Disease Control and Prevention Division of Viral Diseases, Pathogen Discovery | Dakota Howard,Dhwani Batra,Peter Cook,Jason Caravas,Benjamin Rambo-Martin,Scott Sammons,Yvette Unoarumhi,Matthew Schmerer,Kristine Lacek,Tymeckia Kendall,Victoria Caban Figueroa,Shatavia Morrison,Christopher Gulvick,Erisa Sula,Christian Bixby,Yihe Wang,Jo |
| hCoV-19/USA/CT-CDC-QDX31446953/2021 | EPI_ISL_7451070 | 2021-11-24 | Quest Diagnostics Incorporated | Centers for Disease Control and Prevention Division of Viral Diseases, Pathogen Discovery | Dakota Howard,Dhwani Batra,Peter Cook,Jason Caravas,Benjamin Rambo-Martin,Scott Sammons,Yvette Unoarumhi,Matthew Schmerer,Kristine Lacek,Tymeckia Kendall,Victoria Caban Figueroa,Shatavia Morrison,Christopher Gulvick,Erisa Sula,S. Rosenthal,A. Gerasimova,R |
| hCoV-19/Portugal/PT22545/2021 | EPI_ISL_7451261 | 2021-11-22 | UNILABS | Instituto Nacional de Saude (INSA) | Borges et al |
| hCoV-19/SouthAfrica/Tygerberg_3238/2021 | EPI_ISL_7452735 | 2021-11-29 | Division of Medical Virology, National Health Laboratory Service (NHLS), Tygerberg Hospital / Stellenbosch University | Health Laboratory Service (NHLS), Tygerberg Hospital / Stellenbosch University | Susan Engelbrecht, Tongai Maponga, Shannon Wilson, Kamela Mahlakwane, Tania Stander, Gert van Zyl, Wolfgang Preiser |
| hCoV-19/SouthAfrica/Tygerberg_3239/2021 | EPI_ISL_7452736 | 2021-11-29 | Division of Medical Virology, National Health Laboratory Service (NHLS), Tygerberg Hospital / Stellenbosch University | Health Laboratory Service (NHLS), Tygerberg Hospital / Stellenbosch University | Susan Engelbrecht, Tongai Maponga, Shannon Wilson, Kamela Mahlakwane, Tania Stander, Gert van Zyl, Wolfgang Preiser |
| hCoV-19/USA/MA-CDCBI-CRSP_ZYDXIR5ULCI2DM2R/2021 | EPI_ISL_7454277 | 2021-11-29 | Broad Institute Clinical Research Sequencing Platform | Infectious Disease Program, Broad Institute of Harvard and MIT | Siddle,K.J., Adams,G., Pearlman,L., Gladden-Young,A., Vicente,G., Blumenstiel,B., DeFelice,M., Lee,M., McGovern,S., Lagerborg,K., Rudy,M., DeRuff,K., Carter,A., Normandin,E., Bauer,M., Reilly,S., Tomkins-Tinch,C., Loreth,C., Chaluvadi,S., Meldrim,J., Gran |
| hCoV-19/SouthAfrica/Tygerberg_3182/2021 | EPI_ISL_7456496 | 2021-11-30 | PathCare, Cape Town | Health Laboratory Service (NHLS), Tygerberg Hospital / Stellenbosch University | Jean Maritz, Nadine Cronje, Petra Raimond, Tongai Maponga, Shannon Wilson, Kamela Mahlakwane, Tania Stander, Gert van Zyl, Wolfgang Preiser |
| hCoV-19/SouthAfrica/Tygerberg_3190/2021 | EPI_ISL_7456501 | 2021-11-29 | PathCare, Cape Town | Health Laboratory Service (NHLS), Tygerberg Hospital / Stellenbosch University | Jean Maritz, Nadine Cronje, Petra Raimond, Tongai Maponga, Shannon Wilson, Kamela Mahlakwane, Tania Stander, Gert van Zyl, Wolfgang Preiser |
| hCoV-19/SouthAfrica/Tygerberg_3204/2021 | EPI_ISL_7456509 | 2021-11-30 | PathCare, Cape Town | Health Laboratory Service (NHLS), Tygerberg Hospital / Stellenbosch University | Jean Maritz, Nadine Cronje, Petra Raimond, Tongai Maponga, Shannon Wilson, Kamela Mahlakwane, Tania Stander, Gert van Zyl, Wolfgang Preiser |
| hCoV-19/USA/CO-CDPHE-2102118269/2021 | EPI_ISL_7459053 | 2021-11-01 | Colorado Department of Public Health and Environment | Colorado Department of Public Health and Environment | Laura Bankers, Molly C. Hetherington-Rauth, Diana Ir, Alexandria Rossheim, Michael Martin, Mandy Waters, Arianna Smith, Shannon R. Matzinger, Sarah Elizabeth Totten, Emily A. Travanty |

| | | | | | |
|---|---|---|---|---|---|
| hCoV-19/Brazil/SP-IB_RP114924/2021 | EPI_ISL_7459683 | 2021-08-16 | Centro De Saude Ii Santa Rita Do Passa Quatro - Santa Rita Do Passa Quatro - 2774682 | Instituto Butantan | Dimas Tadeu Covas, Antonio Jorge Martins, Claudia Renata dos Santos Barros, David Schlesinger, Debora Botequio Moretti, Elaine Cristina Marqueze, Elaine Vieira Santos, Evandra Strazza Rodrigues, Heidge Fukumasu, Jayme Augusto de Souza-Neto, Jos√© Salvator |
| hCoV-19/Malta/MDxMDH720/2021 | EPI_ISL_7462375 | 2021-11-03 | Molecular Diagnostics Pathology Department Mater Dei Hospital Malta | Molecular Diagnostics Pathology Department Mater Dei Hospital Malta | G Zahra, R Borg, C Cilia, M Briffa, L Grech |
| hCoV-19/France/IDF-CERBAHC-11615853/2021 | EPI_ISL_7463934 | 2021-11-18 | LABM CLEMENT | CERBA HealthCare | B√©n√©dicte Roquebert, Laura Verdurme, Sabine Trombert, St√©phanie Haim-Boukobza |
| hCoV-19/England/PHEP-YYN891C/2021 | EPI_ISL_7464165 | 2021-11-26 | Respiratory Virus Unit, Microbiology Services Colindale, Public Health England | COVID-19 Genomics UK (COG-UK) Consortium | PHE Covid Sequencing Team |
| hCoV-19/England/PHEP-YYN89UK/2021 | EPI_ISL_7464177 | 2021-11-26 | Respiratory Virus Unit, Microbiology Services Colindale, Public Health England | COVID-19 Genomics UK (COG-UK) Consortium | PHE Covid Sequencing Team |
| hCoV-19/England/PHEP-YYN89WE/2021 | EPI_ISL_7464178 | 2021-11-26 | Respiratory Virus Unit, Microbiology Services Colindale, Public Health England | COVID-19 Genomics UK (COG-UK) Consortium | PHE Covid Sequencing Team |
| hCoV-19/Argentina/PAIS-E0571/2021 | EPI_ISL_7464424 | 2021-11-16 | Laboratorio Central de la Ciudad de Santa Fe | Bioinform√°tica del Instituto de Investigaci√≥n de la Cadena L√°ctea CONICET-INTA on behalf of 'Proyecto Argentino Interinstitucional de genomica de SARS-CoV-2' (PAIS | Eberhardt, MF; Irazoqui, JM; Ojeda, G; Rompato, G; Mugna, V; Pastor, C; Amadio, AF |
| hCoV-19/Argentina/PAIS-E0609/2021 | EPI_ISL_7464461 | 2021-11-24 | Laboratorio Central de la Ciudad de Santa Fe | Bioinform√°tica del Instituto de Investigaci√≥n de la Cadena L√°ctea CONICET-INTA on behalf of 'Proyecto Argentino Interinstitucional de genomica de SARS-CoV-2' (PAIS | Eberhardt, MF; Irazoqui, JM; Ojeda, G; Rompato, G; Mugna, V; Pastor, C; Amadio, AF |
| hCoV-19/Norway/Ahus-2435/2021 | EPI_ISL_7464516 | 2021-12-06 | Dept. of Microbiology and Infection Control, Akershus University Hospital HF | Control, Akershus University Hospital HF | Hege Vangstein Aamot, Alexander Hesselberg L√∏vestad |
| hCoV-19/India/MH-NEERI-CIB5131/2021 | EPI_ISL_7468124 | 2021-10-19 | CSIR-National Environmental Engineering Research Institute | CSIR-Centre for Cellular and Molecular Biology-INSACOG | Lamuk Zaveri,Tulasi Nagabandi, Sreelekshmi MS,Malini Nemalikanti,Ara Sreenivas,Shreekant Verma, Amareshwar Vodapalli ,B Himasri,Valli Nagalakshmi Undamatla, Sumedha Avadhanula, Vidhyadhari Methuku, Priya Nurkurthy, Archana Bharadwaj Siva, Karthik Bharadw |
| hCoV-19/India/GJ-INSACOG-GBRC3107/2021 | EPI_ISL_7469156 | 2021-11-11 | Pangenomics, Ahmedabad | Gujarat Biotechnology Research Centre | Janvi Raval,Apurvasinh Puvar,Zarna Patel,Dinesh Kumar,Bhadreshsinh Gohil,Nimesh Patel,Nitin Shukla,Ramesh Pandit,Nitin Savaliya,Priyank Chavda,Sonal Sharma ,KM Singh,Chaitanya Joshi,Madhvi Joshi |
| hCoV-19/Norway/29566/2021 | EPI_ISL_7470189 | 2021-11-30 | Ostfold Hospital Trust - Kalnes, Centre for Laboratory Medicine, Section for gene technology and infection serology | Norwegian Institute of Public Health, Department of Virology | Kathrine Stene-Johansen, Kamilla Heddeland Instefjord, Hilde Elshaug, Garcia Llorente Ignacio, Jon Br√•te, Engebretsen Serina Beate, Pedersen Benedikte Nevjen, Line Victoria Moen, Debech Nadia, Atiya R Ali, Marie Paulsen Madsen, Rasmus Riis Kopperud, Hild |

| | | | | | |
|---|---|---|---|---|---|
| hCoV-19/Norway/29647/2021 | EPI_ISL_7470216 | 2021-11-29 | Medical Microbiology Unit, Department for Laboratory Medicine, Drammen Hospital, Vestre Viken Health Trust | Norwegian Institute of Public Health, Department of Virology | Kathrine Stene-Johansen, Kamilla Heddeland Instefjord, Hilde Elshaug, Garcia Llorente Ignacio, Jon Br√•te, Engebretsen Serina Beate, Pedersen Benedikte Nevjen, Line Victoria Moen, Debech Nadia, Atiya R Ali, Marie Paulsen Madsen, Rasmus Riis Kopperud, Hild |
| hCoV-19/Norway/OUS-18315/2021 | EPI_ISL_7470728 | 2021-11-29 | Oslo University Hospital, Department of Microbiology | Norwegian Institute of Public Health, Department of Virology | Mona Holberg-Petersen, Lise Andresen, Cathrine Fladeby, Mariann Nilsen, Teodora Plamenova Ribarska, P√•l Marius Bj√∏rnstad, Gregor D. Gilfillan, Arvind Yegambaram Meenakshi Sundaram,Kathrine Stene-Johansen, Kamilla Heddeland Instefjord, Hilde Elshaug, Gar |
| hCoV-19/Netherlands/LB-RIVM-73013/2021 | EPI_ISL_7470823 | 2021-11-14 | Dutch COVID-19 response team | National Institute for Public Health and the Environment (RIVM) | Adam Meijer, Harry Vennema, Dirk Eggink, Jeroen Cremer, Sharon van den Brink, Bas van der Veer, AnneMarie van den Brandt, Lisa Wijsman, Kim Freriks, Ryanne Jaarsma, Lynn Aarts, Sanne Bos, Jil Kocken, Stijn van Rossum, Linda van Someren, Florian Zwagemaker |
| hCoV-19/Netherlands/NB-RIVM-71518/2021 | EPI_ISL_7470977 | 2021-11-21 | Dutch COVID-19 response team | National Institute for Public Health and the Environment (RIVM) | Adam Meijer, Harry Vennema, Dirk Eggink, Jeroen Cremer, Sharon van den Brink, Bas van der Veer, AnneMarie van den Brandt, Lisa Wijsman, Kim Freriks, Ryanne Jaarsma, Lynn Aarts, Sanne Bos, Jil Kocken, Stijn van Rossum, Linda van Someren, Florian Zwagemaker |
| hCoV-19/USA/CA-OC-1146/2021 | EPI_ISL_7479620 | 2021-11-14 | Orange County Public Health Laboratory | Laboratory | Angelica Torres, Julia Wolfe |
| hCoV-19/Brazil/SP-IB_RP126394/2021 | EPI_ISL_7494131 | 2021-09-30 | HOSPITAL E AMBULATORIO UNIDADE MISTA-SANTO ANTONIO ALEGRIA | Instituto Butantan | Dimas Tadeu Covas, Antonio Jorge Martins, Claudia Renata dos Santos Barros, David Schlesinger, Debora Botequio Moretti, Elaine Cristina Marqueze, Elaine Vieira Santos, Evandra Strazza Rodrigues, Heidge Fukumasu, Jayme Augusto de Souza-Neto, Jos√© Salvator |
| hCoV-19/Colombia/DC-LSPSDS-VG-1104981/2021 | EPI_ISL_7494437 | 2021-10-07 | Compensar Calle 63 | LSPSDS | Johana Hernandez Toloza, Marcela Castano Rodriguez,Gabriela Delgado Murcia, Alejandro Gomez Lopez, Marcela Mercado |
| hCoV-19/Colombia/DC-LSPSDS-VG-996204/2021 | EPI_ISL_7494451 | 2021-08-11 | Compensar Calle 26 | LSPSDS | Johana Hernandez Toloza, Marcela Castano Rodriguez,Gabriela Delgado Murcia, Alejandro Gomez Lopez, Marcela Mercado |
| hCoV-19/Peru/UCA-INS-10381/2021 | EPI_ISL_7494899 | 2021-11-11 | Laboratorio de Referencia Nacional de Virus Respiratorios. Centro Nacional de Salud Publica. Instituto Nacional de Salud Peru. | Laboratorio de Referencia Nacional de Virus Respiratorios. Centro Nacional de Salud Publica. Instituto Nacional de Salud Peru. | Carlos Padilla Rojas, Veronica Hurtado Vela, Orson Mestanza Millones, Iris Silva Molina, Luis Barcena Flores, Alicia Nu√±ez Llanos, Steve Acedo Lazo, Wendy Lizarraga Olivares, Victor Jimenez Vasquez, Sara Gordillo Vilchez, Kelly Vanessa Izarra Rojas, Edwa |
| hCoV-19/Peru/ICA-INS-10142/2021 | EPI_ISL_7495001 | 2021-11-16 | Laboratorio de Referencia Nacional de Virus Respiratorios. Centro Nacional de Salud Publica. Instituto Nacional de Salud Peru. | Laboratorio de Referencia Nacional de Virus Respiratorios. Centro Nacional de Salud Publica. Instituto Nacional de Salud Peru. | Carlos Padilla Rojas, Veronica Hurtado Vela, Orson Mestanza Millones, Iris Silva Molina, Luis Barcena Flores, Alicia Nu√±ez Llanos, Steve Acedo Lazo, Wendy Lizarraga Olivares, Victor Jimenez Vasquez, Sara Gordillo Vilchez, Kelly Vanessa Izarra Rojas, Edwa |
| hCoV-19/USA/UT-UPHL-211209180135/2021 | EPI_ISL_7503247 | 2021-11-29 | Utah Public Health Laboratory | Utah Public Health Laboratory | Erin L. Young, Kelly F. Oakeson, Pooja Gupta, John Arnn, Tom Iverson, Olinto Linares-Perdomo |

| | | | | | |
|---|---|---|---|---|---|
| hCoV-19/England/MILK-2CCFCF8/2021 | EPI_ISL_7512549 | 2021-11-29 | Lighthouse Lab in Milton Keynes | Wellcome Sanger Institute for the COVID-19 Genomics UK (COG-UK) Consortium | The Lighthouse Lab in Milton Keynes and Alex Alderton, Roberto Amato, Jeffrey Barrett, Sonia Goncalves, Ewan Harrison, David K. Jackson, Ian Johnston, Dominic Kwiatkowski, Cordelia Langford, John Sillitoe on behalf of the Wellcome Sanger Institute COVID-1 |
| hCoV-19/England/ALDP-2D52B47/2021 | EPI_ISL_7513735 | 2021-12-01 | Lighthouse Lab in Alderley Park | Wellcome Sanger Institute for the COVID-19 Genomics UK (COG-UK) Consortium | Jacquelyn Wynn, Mairead Hyland, The Lighthouse Lab in Alderley Park and Alex Alderton, Roberto Amato, Jeffrey Barrett, Sonia Goncalves, Ewan Harrison, David K. Jackson, Ian Johnston, Dominic Kwiatkowski, Cordelia Langford, John Sillitoe on behalf of the W |
| hCoV-19/England/ALDP-2D20368/2021 | EPI_ISL_7515719 | 2021-11-28 | Lighthouse Lab in Alderley Park | Wellcome Sanger Institute for the COVID-19 Genomics UK (COG-UK) Consortium | Jacquelyn Wynn, Mairead Hyland, The Lighthouse Lab in Alderley Park and Alex Alderton, Roberto Amato, Jeffrey Barrett, Sonia Goncalves, Ewan Harrison, David K. Jackson, Ian Johnston, Dominic Kwiatkowski, Cordelia Langford, John Sillitoe on behalf of the W |
| hCoV-19/England/MILK-2D2A2AD/2021 | EPI_ISL_7524405 | 2021-12-04 | Lighthouse Lab in Milton Keynes | Wellcome Sanger Institute for the COVID-19 Genomics UK (COG-UK) Consortium | The Lighthouse Lab in Milton Keynes and Alex Alderton, Roberto Amato, Jeffrey Barrett, Sonia Goncalves, Ewan Harrison, David K. Jackson, Ian Johnston, Dominic Kwiatkowski, Cordelia Langford, John Sillitoe on behalf of the Wellcome Sanger Institute COVID-1 |
| hCoV-19/USA/CA-SACPHL_21SAC0538/2021 | EPI_ISL_7542622 | 2021-12-01 | Sacramento County Public Health Laboratory | Sacramento County Public Health Laboratory | Nhi Duong |
| hCoV-19/USA/NC-CORVASEQ-1086-358/2021 | EPI_ISL_7542659 | 2021-11-20 | Wake Forest School of Medicine, Internal Medicine | Wake Forest School of Medicine, Internal Medicine | Kimberly Reeves, Gregory Hawkins |
| hCoV-19/USA/MA-CDCBI-CRSP_INFE47KV57ZUSYR6/2021 | EPI_ISL_7543579 | 2021-11-30 | Broad Institute Clinical Research Sequencing Platform | Infectious Disease Program, Broad Institute of Harvard and MIT | Siddle,K.J., Adams,G., Pearlman,L., Gladden-Young,A., Vicente,G., Blumenstiel,B., DeFelice,M., Lee,M., McGovern,S., Lagerborg,K., Rudy,M., DeRuff,K., Carter,A., Normandin,E., Bauer,M., Reilly,S., Tomkins-Tinch,C., Loreth,C., Chaluvadi,S., Meldrim,J., Gran |
| hCoV-19/SouthAfrica/NHLS-UCT-LA-Z744/2021 | EPI_ISL_7543960 | 2021-11-22 | Lancet | NHLS/UCT | Arash Iranzadeh, Deelan Doolabh, Lynn Tyers, Bruna Galvao, Innocent Mudau, Marvin Hsiao, Gert Marais, Diana Hardie, Stephen Korsman, Rageema Joseph, Luicer Olubayo, Nokuzola Mbhele, Carolyn Williamson |
| hCoV-19/SouthAfrica/NHLS-UCT-LA-Z750/2021 | EPI_ISL_7544030 | 2021-11-22 | Lancet | NHLS/UCT | Arash Iranzadeh, Deelan Doolabh, Lynn Tyers, Bruna Galvao, Innocent Mudau, Marvin Hsiao, Gert Marais, Diana Hardie, Stephen Korsman, Rageema Joseph, Luicer Olubayo, Nokuzola Mbhele, Carolyn Williamson |
| hCoV-19/SouthAfrica/NHLS-UCT-GS-AF35/2021 | EPI_ISL_7544319 | 2021-11-23 | 2 Military Hospital wc MAA | NHLS/UCT | Arash Iranzadeh, Deelan Doolabh, Lynn Tyers, Bruna Galvao, Innocent Mudau, Marvin Hsiao, Gert Marais, Diana Hardie, Stephen Korsman, Rageema Joseph, Luicer Olubayo, Nokuzola Mbhele, Carolyn Williamson |
| hCoV-19/Switzerland/ZH-UZH-IMV-3ba57f48/2021 | EPI_ISL_7544865 | 2021-11-28 | Zentrallabor Z√ºrich | Institute of Medical Virology, University of Zurich | Verena Kufner, Gabriela Ziltener, Maryam Zaheri, Stefan Schmutz, Annette Audig√©, Maria Gr√ºnberg, Kevin Steiner, Jon Huder, Cyril Shah, Riccarda Capaul, Guido Bloemberg, J√ºrg B√∂ni, Michael Huber, Alexandra Trkola |

| | | | | | |
|---|---|---|---|---|---|
| hCoV-19/USA/WA-CDC-UW21120375709/2021 | EPI_ISL_7545546 | 2021-12-03 | UW Virology Lab | UW Virology Lab | Pavitra Roychoudhury, Shah Mohamed Bakhash, Pooneh Hajian, Ricardo Perez, Seffir T. Wendm, Isabel Arnould, Hong Xie, Sean Ellis, Nathan Breit, Robert J. Livingston, Meei-Li Huang, Keith R Jerome, Patrick Mathias, Alexander Greninger |
| hCoV-19/SouthAfrica/CERI-KRISP-K032904/2021 | EPI_ISL_7545644 | 2021-11-30 | SAMRC | [illegible] and Innvoation, Stellenbosch University and KRISP, KZN Research Innovation and Sequencing Platform, UKZN. | MRC, Giandhari J, Pillay S, Naidoo Y, Ramphal U, Ramphal Y, Arisha Maharaj, San JE, Tshiabuila D, Tegally H, Wilkinson E, de Oliveira T |
| hCoV-19/SouthAfrica/CERI-KRISP-K032917/2021 | EPI_ISL_7545653 | 2021-12-03 | NHLS Livingstone Laboratory | [illegible] and Innvoation, Stellenbosch University and KRISP, KZN Research Innovation and Sequencing Platform, UKZN. | Oluwakemi Laguda-Akingba and Nokukhanya Mdlalose, Giandhari J, Pillay S, Naidoo Y, Ramphal U, Ramphal Y, Arisha Maharaj, San JE, Tshiabuila D, Tegally H, Wilkinson E, de Oliveira T |
| hCoV-19/SouthAfrica/CERI-KRISP-K032935/2021 | EPI_ISL_7545672 | 2021-12-02 | NHLS Livingstone Laboratory | [illegible] and Innvoation, Stellenbosch University and KRISP, KZN Research Innovation and Sequencing Platform, UKZN. | Oluwakemi Laguda-Akingba and Nokukhanya Mdlalose, Giandhari J, Pillay S, Naidoo Y, Ramphal U, Ramphal Y, Arisha Maharaj, San JE, Tshiabuila D, Tegally H, Wilkinson E, de Oliveira T |
| hCoV-19/Argentina/PAIS-G0461/2021 | EPI_ISL_7547024 | 2021-09-17 | Laboratorio Central, Ministerio de Salud C√≥rdoba | [illegible] Medicina and Instituto de Patologia Vegetal (IPAVE-CIAP-INTA) on behalf of 'Proyecto Argentino Interinstitucional de genomica de SARS-CoV-2' (PAIS Consortium) | Zeballos, M.; Lucca, A.; Fern√°ndez, F.D.; Marquez, N.; Debat, H., Goya, S.; Re, V.; Pisano, M.B.; Castro, G.; Sicilia, P.; Barbas, G. |
| hCoV-19/Nigeria/NCDC-NR1053/2021 | EPI_ISL_7547733 | 2021-11-29 | Everight diagnostics (Abuja) | National Reference Laboratory, Nigeria Centre for Disease Control | Dr Ndodo Nnaemeka, Olusola Anuoluwapo Akanbi, Chimaobi Chukwu, Oyeronke Ayansola, Dr Omoare Adesuyi, Catherine Okoi, Olajumoke Babatunde, Nwando Mba, Dr Ifedayo Adetifa |
| hCoV-19/Israel/ICH-760598069/2021 | EPI_ISL_7552046 | 2021-12-02 | Clinical Microbiology Laboratory, Tel Aviv Sourasky Medical Center | Clinical Microbiology Laboratory, Tel Aviv Sourasky Medical Center | Katya Levytskyi, Ora Halutz, Alon Ziv, Lior Handler, Amos Adler |
| hCoV-19/Scotland/EDB31454/2021 | EPI_ISL_7561464 | 2021-11-29 | Virology Department, Royal Infirmary of Edinburgh, NHS Lothian / School of Biological Sciences, University of Edinburgh | COVID-19 Genomics UK (COG-UK) Consortium | McHugh M, Dewar R, Cotton S, Gallagher A, Maloney D, Fernandez G, O‚ÄôToole √Å, Scher E, Hill V, McCrone JT, Colquhoun R, Yu X, Jackson B, Rambaut A, Templeton K |
| hCoV-19/Bangladesh/CHRF-GSB15-074/2021 | EPI_ISL_7566180 | 2021-11-01 | Rajshahi Medical College, Rajshahi | Child Health Research Foundation | CHRF Bangladesh Genomics Team, Sabera Gul Nahar, Manjur Hossain Khan, Senjuti Saha, Hassan Afrad, Mohammed Ziaur Rahman, Mohammad Jubair, Mohammad Enayet Hossain, Razib Mazumder, Mohammad Shahidul Islam, Ahmed Nawsher Alam, ASM Alamgir, Mahbubur Rahman, M |
| hCoV-19/Singapore/8709/2021 | EPI_ISL_7566286 | 2021-12-05 | National Public Health Laboratory, National Centre for Infectious Diseases | National Centre for Infectious Diseases | Katherine Ching, Zhenyang Zhou, Royce Ang, Samuel Loo, Lin Cui, Raymond Tzer Pin Lin |
| hCoV-19/Botswana/R50B95_BHP_000847976/2021 | EPI_ISL_7566353 | 2021-11-26 | National Health Laboratory | Botswana Harvard AIDS Institute Partnership, Plot 1836 North Ring Road, Princess Marina Hospital, Gaborone | Sikhulile Moyo, Wonderful T. Choga, Ontlametse T. Bareng, Doreen Ditshwanelo, Dorcas Maruapula, Keoratile Ntshambiwa, Sefetogi Ramaologa, Thongbotho Mphoyakgosi, Boitumelo Zuze, Botshelo Radibe, Legodile Kooepile, Pamela Smith-Lawrence, Kgomotso Moruisi, |

| | | | | | |
|---|---|---|---|---|---|
| hCoV-19/Botswana/R50B39_BHP_AAC29687/2021 | EPI_ISL_7566379 | 2021-12-01 | Botswana Harvard HIV Reference Laboratory | Botswana Harvard AIDS Institute Partnership, Plot 1836 North Ring Road, Princess Marina Hospital, Gaborone | Sikhulile Moyo, Wonderful T. Choga, Ontlametse T. Bareng, Doreen Ditshwanelo, Dorcas Maruapula, Keoratile Ntshambiwa, Sefetogi Ramaologa, Thongbotho Mphoyakgosi, Boitumelo Zuze, Botshelo Radibe, Legodile Kooepile, Pamela Smith-Lawrence, Kgomotso Moruisi, |
| hCoV-19/Switzerland/SO-ETHZ-35052074/2021 | EPI_ISL_7566947 | 2021-11-30 | Viollier AG | Department of Biosystems Science and Engineering, ETH Zv°rich | Chaoran Chen, Sarah Nadeau, Louis du Plessis, Catharine Aquino Fournier, Andrea Patrizia Salzmann, Henriette Kurth, Ivan Topolsky, Kim Philipp Jablonski, Lara Fuhrmann, David Dreifuss, Pelin Icer, Daniel Ehrsam, Isabel Stv°rmer, Andreia Cabral de Gouvea, |
| hCoV-19/Australia/ACT1769/2021 | EPI_ISL_7569737 | 2021-11-08 | ACT Pathology | Schwessinger Lab | Austin Bird, Carolina Correa Ospina, Emma Crean, Scott Ferguson, Robyn Hall, Evie Hodgson, Ashley Jones, Elise Kellett, Craig Kennedy, Karina Kennedy, Rachel Leonard, Carl McCombe, Catalina Barragv°n Quintero, Rene Riedelbauch, Benjamin Schwessinger, Gabr |
| hCoV-19/Senegal/SN-IR8-005670/2021 | EPI_ISL_7571613 | 2021-12-04 | Iressef Genomics lab | IRESSEF | Souleymane MBOUP, Abdou PADANE , Khadim GUEYE, Cyrille Diedhiou , Samba Ndiour, Diabou Diagne, Papa Alassane DIAW, Birahim Piere NDIAYE, Barada CISSE, Aminata MBOUP, Moustapha MBOW, Ndeye Coumba Toure KANE, Nafisatou LEYE, Gora LO, Ambroise AHOUIDI , Ast |
| hCoV-19/Bangladesh/TND-icddrb-02-0131/2021 | EPI_ISL_7571095 | 2021-10-03 | International Centre for Diarrhoeal Disease Research, Bangladesh (icddr,b) | Disease Research, Bangladesh (icddr,b) | Afrad MH, Khanam F, Islam T, Hamza O, Bhuyian TR, Firdausi Qadri |
| hCoV-19/England/PHEC-3U0A6U7A/2021 | EPI_ISL_7583286 | 2021-11-26 | Respiratory Virus Unit, Microbiology Services Colindale, Public Health England | COVID-19 Genomics UK (COG-UK) Consortium | PHE Covid Sequencing Team |
| hCoV-19/Germany/BY-MVP-000009532/2021 | EPI_ISL_7590785 | 2021-11-12 | Max von Pettenkofer Institute, Virology, National Reference Center for Retroviruses, LMU Munich | Laboratory for Functional Genome Analysis; Dept. Genomics; Gene Center of the LMU Munich | Max Muenchhoff; Stefan Krebs; Alexander Graf; Oliver Keppler; Helmut Blum |
| hCoV-19/Germany/BY-MVP-000009797/2021 | EPI_ISL_7591021 | 2021-11-17 | Max von Pettenkofer Institute, Virology, National Reference Center for Retroviruses, LMU Munich | Laboratory for Functional Genome Analysis; Dept. Genomics; Gene Center of the LMU Munich | Max Muenchhoff; Stefan Krebs; Alexander Graf; Oliver Keppler; Helmut Blum |
| hCoV-19/Bangladesh/TND-icddrb-09-0231/2021 | EPI_ISL_7592283 | 2021-11-24 | International Centre for Diarrhoeal Disease Research, Bangladesh (icddr,b) | Disease Research, Bangladesh (icddr,b) | Afrad MH, Khanam F, Islam T, Hamza O, Bhuyian TR, Firdausi Qadri |
| hCoV-19/Israel/SMC-7026144/2021 | EPI_ISL_7594672 | 2021-12-06 | Shamir Medical Center (Asaf Harofe) | Shamir Medical Center (Asaf Harofe) | Nir Rainy,Chen Weiner,Reut Sorek Abramovich,Yevgeni Yegorov,Anna Vishnevsky,Patricia Benveniste-Lekovitz,Abu Hamad Ramzia,Adina Bar Chaim |
| hCoV-19/Israel/SMC-7025490/2021 | EPI_ISL_7604219 | 2021-12-05 | Shamir Medical Center (Asaf Harofe) | Shamir Medical Center (Asaf Harofe) | Nir Rainy,Chen Weiner,Reut Sorek Abramovich,Yevgeni Yegorov,Anna Vishnevsky,Patricia Benveniste-Lekovitz,Abu Hamad Ramzia,Adina Bar Chaim |
| hCoV-19/USA/NJ-PHEL-V21032782-rep2/2021 | EPI_ISL_7605460 | 2021-11-26 | NJDOH, Public Health and Environmental Laboratories | NJ_PHEL | Lindsey Bodnar, Shiv K. Verma, Ryan Pachucki, Maria-Magdalene Pugliese, Chelsea San Filippo, Mohammad M. Ali, Dana Woell, Byeong Jeong |
| hCoV-19/SouthAfrica/NICD-N22061/2021 | EPI_ISL_7605601 | 2021-11-24 | KIMBERLEY LABORATORY | Diseases of the National Health Laboratory Service | Amoako DG, Everatt J, Scheepers C, Mohale T, Ntuli N, Mahlangu B, Mnguni A, Ismail A, Bhiman JN, Wolter N |
| hCoV-19/SouthAfrica/NICD-N22225/2021 | EPI_ISL_7605615 | 2021-11-22 | TSHEPONG LABORATORY | Diseases of the National Health Laboratory Service | Amoako DG, Everatt J, Scheepers C, Mohale T, Ntuli N, Mahlangu B, Mnguni A, Ismail A, Bhiman JN, Wolter N |

| | | | | | |
|---|---|---|---|---|---|
| hCoV-19/SouthAfrica/NICD-N22384/2021 | EPI_ISL_7605652 | 2021-11-30 | ROB FERREIRA LABORATORY | Diseases of the National Health Laboratory Service | Amoako DG, Everatt J, Scheepers C, Mohale T, Ntuli N, Mahlangu B, Mnguni A, Ismail A, Bhiman JN, Wolter N |
| hCoV-19/SouthAfrica/NICD-N22390/2021 | EPI_ISL_7605654 | 2021-11-30 | ROB FERREIRA LABORATORY | Diseases of the National Health Laboratory Service | Amoako DG, Everatt J, Scheepers C, Mohale T, Ntuli N, Mahlangu B, Mnguni A, Ismail A, Bhiman JN, Wolter N |
| hCoV-19/SouthAfrica/NICD-N22353/2021 | EPI_ISL_7605689 | 2021-11-30 | DE AAR LABORATORY | Diseases of the National Health Laboratory Service | Amoako DG, Everatt J, Scheepers C, Mohale T, Ntuli N, Mahlangu B, Mnguni A, Ismail A, Bhiman JN, Wolter N |
| hCoV-19/SouthAfrica/NICD-N22380/2021 | EPI_ISL_7605706 | 2021-11-30 | KIMBERLEY LABORATORY | Diseases of the National Health Laboratory Service | Amoako DG, Everatt J, Scheepers C, Mohale T, Ntuli N, Mahlangu B, Mnguni A, Ismail A, Bhiman JN, Wolter N |
| hCoV-19/SouthAfrica/NICD-N22337/2021 | EPI_ISL_7605720 | 2021-12-01 | National Institute for Communicable Diseases of the National Health Laboratory Service | Diseases of the National Health Laboratory Service | Amoako DG, Everatt J, Scheepers C, Mohale T, Ntuli N, Mahlangu B, Mnguni A, Ismail A, Bhiman JN, Wolter N |
| hCoV-19/SouthAfrica/NICD-N22086/2021 | EPI_ISL_7605758 | 2021-10-15 | PORT ELIZABETH LABORATORY | Diseases of the National Health Laboratory Service | Amoako DG, Everatt J, Scheepers C, Mohale T, Ntuli N, Mahlangu B, Mnguni A, Ismail A, Bhiman JN, Wolter N |
| hCoV-19/SouthAfrica/NICD-N22074/2021 | EPI_ISL_7605764 | 2021-11-22 | PORT ELIZABETH LABORATORY | Diseases of the National Health Laboratory Service | Amoako DG, Everatt J, Scheepers C, Mohale T, Ntuli N, Mahlangu B, Mnguni A, Ismail A, Bhiman JN, Wolter N |
| hCoV-19/Switzerland/ZH-UZH-IMV-3ba57443/2021 | EPI_ISL_7605921 | 2021-11-29 | Universit√§t Z√ºrich | Institute of Medical Virology, University of Zurich | Daniel Ehrsam, Isabel St√ºrmer, Catharine Aquino, Joel Wirz, Weihong Qi, Hubert Rehrauer, Verena Kufner, Gabriela Ziltener, Maryam Zaheri, Stefan Schmutz, Annette Audig√©, Maria Gr√ºnberg, Kevin Steiner, Jon Huder, Cyril Shah, Riccarda Capaul, Guido Bloem |
| hCoV-19/Israel/SMC-7025297/2021 | EPI_ISL_7606158 | 2021-11-29 | Shamir Medical Center (Asaf Harofe) | Shamir Medical Center (Asaf Harofe) | Nir Rainy,Chen Weiner,Reut Sorek Abramovich,Yevgeni Yegorov,Anna Vishnevsky,Patricia Benveniste-Lekovitz,Abu Hamad Ramzia,Adina Bar Chaim |
| hCoV-19/Israel/SMC-7025155/2021 | EPI_ISL_7606202 | 2021-11-29 | Shamir Medical Center (Asaf Harofe) | Shamir Medical Center (Asaf Harofe) | Nir Rainy,Chen Weiner,Reut Sorek Abramovich,Yevgeni Yegorov,Anna Vishnevsky,Patricia Benveniste-Lekovitz,Abu Hamad Ramzia,Adina Bar Chaim |
| hCoV-19/USA/NY-PRL-2021_1201_00K20/2021 | EPI_ISL_7614032 | 2021-11-28 | Pandemic Response Lab - NYC | Pandemic Response Lab, R&D | Henry Lee, Michael Hammerling, Melissa Hopkins, Cybill del Castillo, Shinyoung Clair Kang, William Ward, Pradeep Bugga, Sol Rey, Dylan Law, Alex Carpio, Isabel Fernandez Escapa, Haiping Hao, Jon Laurent |
| hCoV-19/USA/NY-PRL-2021_1201_01G08/2021 | EPI_ISL_7614035 | 2021-11-26 | Pandemic Response Lab - NYC | Pandemic Response Lab, R&D | Henry Lee, Michael Hammerling, Melissa Hopkins, Cybill del Castillo, Shinyoung Clair Kang, William Ward, Pradeep Bugga, Sol Rey, Dylan Law, Alex Carpio, Isabel Fernandez Escapa, Haiping Hao, Jon Laurent |
| hCoV-19/Israel/SMC-7024836/2021 | EPI_ISL_7614185 | 2021-11-27 | Shamir Medical Center (Asaf Harofe) | Shamir Medical Center (Asaf Harofe) | Nir Rainy,Chen Weiner,Reut Sorek Abramovich,Yevgeni Yegorov,Anna Vishnevsky,Patricia Benveniste-Lekovitz,Abu Hamad Ramzia,Adina Bar Chaim |
| hCoV-19/Israel/SMC-7024934/2021 | EPI_ISL_7614287 | 2021-11-28 | Shamir Medical Center (Asaf Harofe) | Shamir Medical Center (Asaf Harofe) | Nir Rainy,Chen Weiner,Reut Sorek Abramovich,Yevgeni Yegorov,Anna Vishnevsky,Patricia Benveniste-Lekovitz,Abu Hamad Ramzia,Adina Bar Chaim |
| hCoV-19/Israel/SMC-7023482/2021 | EPI_ISL_7615823 | 2021-11-23 | Shamir Medical Center (Asaf Harofe) | Shamir Medical Center (Asaf Harofe) | Nir Rainy,Chen Weiner,Reut Sorek Abramovich,Yevgeni Yegorov,Anna Vishnevsky,Patricia Benveniste-Lekovitz,Abu Hamad Ramzia,Adina Bar Chaim |
| hCoV-19/Canada/QC-1nIEOQ-15782162/2020 | EPI_ISL_7619799 | 2020-06-02 | Laboratoire de sant√© publique du Qu√©bec | Laboratoire de sant√© publique du Qu√©bec | Sandrine Moreira, Ioannis Ragoussis, Guillaume Bourque, Jesse Shapiro, Mark Lathrop and Judith Fafard on behalf of the CoVSeQ research group |

| | | | | | |
|---|---|---|---|---|---|
| hCoV-19/SierraLeone/CPHRL_SEQ_21_297/2021 | EPI_ISL_7620047 | 2021-11-24 | Makeni Molecular Laboratory | Central Public Health Reference Laboratory | Samuel Sorie, Zein Souma, Anu J. Williams, Allan K.O. Campbell, Warren C. Yiaba, Choe Miller, Dr. Raoul E.G. Wadoum, Doris Harding, Prof. Sahr Gevao, Prof. Foday Sahr, Prof. Isatta Wurie |
| hCoV-19/USA/AK-PHL15938/2021 | EPI_ISL_7620089 | 2021-11-29 | Alaska State Virology Laboratory | Alaska State Virology Laboratory | Elva House, Jeremy Roe, Jacob Zidek, Lisa Smith, Ph.D., Jack Chen, Ph.D. |
| hCoV-19/Argentina/PAIS-D0211/2021 | EPI_ISL_7620334 | 2021-10-22 | Laboratorio del Hospital Regional Ushuaia Gdor. Ernesto Campos | Fuego - Hospital Regional Ushuaia - Centro Austral De Investigaciones Cient√≠ficas - Universidad Nacional De Tierra Del Fuego on behalf PAIS project and ANLIS-Malbran | Gramundi, ID; Gallego, F; Ceballos, SG; Nardi, CF; Rojas, AE; De Roccis, CA; Castro, G; C√°ceres, SB; Yulan, CB; Boutureira, MF. |
| hCoV-19/Argentina/PAIS-D0213/2021 | EPI_ISL_7620336 | 2021-11-01 | Laboratorio del Hospital Regional Ushuaia Gdor. Ernesto Campos | Fuego - Hospital Regional Ushuaia - Centro Austral De Investigaciones Cient√≠ficas - Universidad Nacional De Tierra Del Fuego on behalf PAIS project and ANLIS-Malbran | Gramundi, ID; Gallego, F; Ceballos, SG; Nardi, CF; Rojas, AE; De Roccis, CA; Castro, G; C√°ceres, SB; Yulan, CB; Boutureira, MF. |
| hCoV-19/Argentina/PAIS-D0214/2021 | EPI_ISL_7620337 | 2021-11-01 | Laboratorio del Hospital Regional Ushuaia Gdor. Ernesto Campos | Fuego - Hospital Regional Ushuaia - Centro Austral De Investigaciones Cient√≠ficas - Universidad Nacional De Tierra Del Fuego on behalf PAIS project and ANLIS-Malbran | Gramundi, ID; Gallego, F; Ceballos, SG; Nardi, CF; Rojas, AE; De Roccis, CA; Castro, G; C√°ceres, SB; Yulan, CB; Boutureira, MF. |
| hCoV-19/Australia/NSW-ICPMR-15821/2021 | EPI_ISL_7620856 | 2021-11-30 | Histopath | Clinical Pathology and Medical Research; Westmead Hospital; University of Sydney | Arnott A., Draper J., Gall M., Martinez E., Rockett R., Sintchenko V., on behalf of ICPMR |
| hCoV-19/Australia/NSW-ICPMR-16160/2021 | EPI_ISL_7621211 | 2021-12-05 | Pathology West - NSW Health Pathology | Clinical Pathology and Medical Research; Westmead Hospital; University of Sydney | Arnott A., Draper J., Gall M., Martinez E., Rockett R., Sintchenko V., on behalf of ICPMR |
| hCoV-19/Australia/NSW-ICPMR-16363/2021 | EPI_ISL_7621423 | 2021-12-04 | St Vincent's Pathology (SydPath) | Clinical Pathology and Medical Research; Westmead Hospital; University of Sydney | Arnott A., Draper J., Gall M., Martinez E., Rockett R., Sintchenko V., on behalf of ICPMR |
| hCoV-19/Australia/NSW-ICPMR-16637/2021 | EPI_ISL_7621713 | 2021-12-06 | Histopath | Clinical Pathology and Medical Research; Westmead Hospital; University of Sydney | Arnott A., Draper J., Gall M., Martinez E., Rockett R., Sintchenko V., on behalf of ICPMR |
| hCoV-19/Australia/NSW-ICPMR-16781/2021 | EPI_ISL_7621867 | 2021-12-06 | St Vincent's Pathology (SydPath) | Clinical Pathology and Medical Research; Westmead Hospital; University of Sydney | Arnott A., Draper J., Gall M., Martinez E., Rockett R., Sintchenko V., on behalf of ICPMR |
| hCoV-19/Australia/NSW-ICPMR-16831/2021 | EPI_ISL_7621920 | 2021-12-06 | 4Cyte Pathology | Clinical Pathology and Medical Research; Westmead Hospital; University of Sydney | Arnott A., Draper J., Gall M., Martinez E., Rockett R., Sintchenko V., on behalf of ICPMR |
| hCoV-19/Australia/NSW-ICPMR-16862/2021 | EPI_ISL_7621951 | 2021-12-09 | Australian Clinical Labs (formerly Healthscope Pathology) | Clinical Pathology and Medical Research; Westmead Hospital; University of Sydney | Arnott A., Draper J., Gall M., Martinez E., Rockett R., Sintchenko V., on behalf of ICPMR |
| hCoV-19/Australia/NSW-ICPMR-16868/2021 | EPI_ISL_7621957 | 2021-12-09 | Australian Clinical Labs (formerly Healthscope Pathology) | Clinical Pathology and Medical Research; Westmead Hospital; University of Sydney | Arnott A., Draper J., Gall M., Martinez E., Rockett R., Sintchenko V., on behalf of ICPMR |

| | | | | | |
|---|---|---|---|---|---|
| hCoV-19/Denmark/DCGC-259865/2021 | EPI_ISL_7638902 | 2021-12-04 | Department of Bacteria, Parasites and Fungi, Statens Serum Institut, Copenhagen, Denmark | Statens Serum Institut Bioinformatics and Microbial Genomics | Danish Covid-19 Genome Consortium |
| hCoV-19/India/TG-CDFD-KRM41/2021 | EPI_ISL_7649560 | 2021-11-20 | Gandhi Medical College | Centre for DNA Fingerprinting and Diagnostics | Asmita Gupta, Rupin Shelake, Arunkumar Karunanidhi, Vinay Donipadi, Divya Vashisht, Ashwin Dalal, Nagamani Kammili, Murali Bashyam |
| hCoV-19/Canada/QC-1nDJXW-49951631/2021 | EPI_ISL_7651682 | 2021-09-09 | Laboratoire de sant√© publique du Qu√©bec | Laboratoire de sant√© publique du Qu√©bec | Sandrine Moreira, Ioannis Ragoussis, Guillaume Bourque, Jesse Shapiro, Mark Lathrop and Judith Fafard on behalf of the CoVSeQ research group |
| hCoV-19/Canada/QC-1nDWVQ-MX21176242/2021 | EPI_ISL_7651848 | 2021-05-03 | Laboratoire de sant√© publique du Qu√©bec | Laboratoire de sant√© publique du Qu√©bec | Sandrine Moreira, Ioannis Ragoussis, Guillaume Bourque, Jesse Shapiro, Mark Lathrop and Judith Fafard on behalf of the CoVSeQ research group |
| hCoV-19/England/BRBR-2EB3554/2021 | EPI_ISL_7963304 | 2021-12-14 | Berkshire and Surrey Pathology Services Lighthouse Laboratory | Wellcome Sanger Institute for the COVID-19 Genomics UK (COG-UK) Consortium | Berkshire and Surrey Pathology Services Lighthouse Laboratory and Alex Alderton, Roberto Amato, Jeffrey Barrett, Sonia Goncalves, Ewan Harrison, David K. Jackson, Ian Johnston, Dominic Kwiatkowski, Cordelia Langford, John Sillitoe on behalf of the Wellcom |
| hCoV-19/USA/VT-CDCBI-CRSP_7SF4QNNUQLXZ3CL4/2021 | EPI_ISL_7659112 | 2021-12-02 | Broad Institute Clinical Research Sequencing Platform | Infectious Disease Program, Broad Institute of Harvard and MIT | Siddle,K.J., Adams,G., Pearlman,L., Gladden-Young,A., Vicente,G., Blumenstiel,B., DeFelice,M., Lee,M., McGovern,S., Lagerborg,K., Rudy,M., DeRuff,K., Carter,A., Normandin,E., Bauer,M., Reilly,S., Tomkins-Tinch,C., Loreth,C., Chaluvadi,S., Meldrim,J., Gran |
| hCoV-19/Qatar/QA-QU-19-8-D3/2021 | EPI_ISL_7660083 | 2021-11-18 | Ministry of Public Health / Hamad Medical Corporation | Biomedical Research Center (BRC), Qatar University / Qatar Genome Project (QGP) | BRC:¬†Heba A. Al-Khatib, Fatiha M. Benslimane,Maria K. Smatti, Hadeel T. Mohammed, Fatma Hassan Ali, Swapna Thomas, Ola Al-Jamal, Dana Al-Batsh, Hadi M. Yassine, Asmaa A. Al-Thani. MOPH and HMC:¬†Abdullatif Al-Khal, Muna A. S. Al-Maslamani, Mashael A. Al- |
| hCoV-19/Canada/QC-1nMCY-S1130690/2020 | EPI_ISL_7664437 | 2020-05-13 | Laboratoire de sant√© publique du Qu√©bec | Laboratoire de sant√© publique du Qu√©bec | Sandrine Moreira, Ioannis Ragoussis, Guillaume Bourque, Jesse Shapiro, Mark Lathrop and Judith Fafard on behalf of the CoVSeQ research group |
| hCoV-19/India/KL-GENESCOV05007/2021 | EPI_ISL_7224909 | 2021-05-25 | INSACOG-IGIB and Kerala State SARS-COV-2 Genome Surveillance Programme (GENESCOV2) | INSACOG and GENESCOV2 at CSIR Institute of Genomics and Integrative Biology | INSACOG-IGIB and Kerala State SARS-COV-2 Genome Surveillance Programme (GENESCOV2) |
| hCoV-19/USA/KY-CDC-LC0396121/2021 | EPI_ISL_7668071 | 2021-11-23 | Laboratory Corporation of America | Centers for Disease Control and Prevention Division of Viral Diseases, Pathogen Discovery | Dakota Howard,Dhwani Batra,Peter Cook,Jason Caravas,Benjamin Rambo-Martin,Scott Sammons,Yvette Unoarumhi,Matthew Schmerer,Kristine Lacek,Tymeckia Kendall,Victoria Caban Figueroa,Shatavia Morrison,Christopher Gulvick,Minoo Agarwal,Eyad Almasri,Debbie Boles |
| hCoV-19/USA/MA-CDCBI-CRSP_TSH6YBTNZT6OB6PP/2021 | EPI_ISL_7673157 | 2021-12-02 | Broad Institute Clinical Research Sequencing Platform | Infectious Disease Program, Broad Institute of Harvard and MIT | Siddle,K.J., Adams,G., Pearlman,L., Gladden-Young,A., Vicente,G., Blumenstiel,B., DeFelice,M., Lee,M., McGovern,S., Lagerborg,K., Rudy,M., DeRuff,K., Carter,A., Normandin,E., Bauer,M., Reilly,S., Tomkins-Tinch,C., Loreth,C., Chaluvadi,S., Meldrim,J., Gran |
| hCoV-19/Australia/QLD2617/2021 | EPI_ISL_7674636 | 2021-12-11 | PHV-FSS | PHV-FSS | Chenwei Wang on behalf of Q-PHIRE Genomics |

| | | | | | |
|---|---|---|---|---|---|
| hCoV-19/USA/TX-HMH-MCoV-68909/2021 | EPI_ISL_7978244 | 2021-12-10 | Houston Methodist Hospital | Houston Methodist Hospital | Randall J. Olsen, Paul A. Christensen, S. Wesley Long, Richard Snehall, Robert Olson, Marcus Nguyen, James J. Davis, Matthew Ojeda Saavedra, Kristina Reppond, Madison N. Shyer, Jessica Cambric, Ryan Gadd, Akanksha Batajoo, Rashi Thakur, Sindy Pena, Regan |
| hCoV-19/England/PHEC-3U0ACU3C/2021 | EPI_ISL_7684916 | 2021-11-27 | Respiratory Virus Unit, Microbiology Services Colindale, Public Health England | COVID-19 Genomics UK (COG-UK) Consortium | PHE Covid Sequencing Team |
| hCoV-19/England/PHEC-3U0ACU4B/2021 | EPI_ISL_7684919 | 2021-11-26 | Respiratory Virus Unit, Microbiology Services Colindale, Public Health England | COVID-19 Genomics UK (COG-UK) Consortium | PHE Covid Sequencing Team |
| hCoV-19/England/PHEC-3U0AEU81/2021 | EPI_ISL_7684970 | 2021-11-29 | Respiratory Virus Unit, Microbiology Services Colindale, Public Health England | COVID-19 Genomics UK (COG-UK) Consortium | PHE Covid Sequencing Team |
| hCoV-19/Israel/ICH-741105127/2021 | EPI_ISL_7687295 | 2021-12-07 | Clinical Microbiology Laboratory, Tel Aviv Sourasky Medical Center | Clinical Microbiology Laboratory, Tel Aviv Sourasky Medical Center | Katya Levytskyi, Ora Halutz, Alon Ziv, Lior Handler, Amos Adler |
| hCoV-19/Israel/ICH-741105184/2021 | EPI_ISL_7687630 | 2021-12-07 | Clinical Microbiology Laboratory, Tel Aviv Sourasky Medical Center | Clinical Microbiology Laboratory, Tel Aviv Sourasky Medical Center | Katya Levytskyi, Ora Halutz, Alon Ziv, Lior Handler, Amos Adler |
| hCoV-19/Israel/ICH-741105038/2021 | EPI_ISL_7687720 | 2021-12-08 | Clinical Microbiology Laboratory, Tel Aviv Sourasky Medical Center | Clinical Microbiology Laboratory, Tel Aviv Sourasky Medical Center | Katya Levytskyi, Ora Halutz, Alon Ziv, Lior Handler, Amos Adler |
| hCoV-19/SouthAfrica/NICD-N22621/2021 | EPI_ISL_7971523 | 2021-09-30 | VERMAAK LABORATORY | National Institute for Communicable Diseases of the National Health Laboratory Service | Amoako DG, Everatt J, Kekana D, Scheepers C, Mohale T, Ntuli N, Mahlangu B, Mnguni A, Ismail A, Bhiman JN, Wolter N |
| hCoV-19/Colombia/MAG-UCCSMR-VG441/2021 | EPI_ISL_7694406 | 2021-10-27 | Servicios Medicos Olimpus | Laboratorio de Biologia Molecular, Universidad Cooperativa de Colombia, Santa Marta | Andrew S. Muv±oz-Gamba, Daniel B. Ramv≠rez-Osorio, Paula A. Quintero-Cortv©s, Pricelis Paulin Polanco Fontalvo, Mayerlin Vasquez Cardenas, Josv© A. Usme-Ciro |
| hCoV-19/USA/IN-CDC-STM-ZGTGFQFQZ/2021 | EPI_ISL_7695096 | 2021-12-01 | Helix | Centers for Disease Control and Prevention Division of Viral Diseases, Pathogen Discovery | Dakota Howard,Dhwani Batra,Peter Cook,Jason Caravas,Benjamin Rambo-Martin,Scott Sammons,Yvette Unoarumhi,Matthew Schmerer,Kristine Lacek,Tymeckia Kendall,Victoria Caban Figueroa,Helix CA,Shatavia Morrison,Christopher Gulvick,Erisa Sula,Clinton Paden,Dunca |
| hCoV-19/Thailand/DMSc-04892/2021 | EPI_ISL_7695246 | 2021-10-26 | So Phisai Hospital | Division of Genomic Medicine and Innovation support,Department of Medical Sciences, Ministry of Public Health, Thailand | Surakameth Mahasirimongkol,Nuanjun Wichukchinda,Archawin Rojanawiwat,Pilailuk Akkapaiboon Okada ,Waritta Sawaengdee,Penpitcha Thawong,Pundharika Piboonsiri,Jirapha Pakdee,Natthakul Bunneang |
| hCoV-19/USA/MD-HP21155-PIDFZAYXKG/2021 | EPI_ISL_7695690 | 2021-11-28 | Johns Hopkins Hospital Department of Pathology | Johns Hopkins Hospital Department of Pathology | C. Paul Morris, David Gaston, Julie M. Norton, Amary Fall, Raghda Eldesouki, Michael Forman, Matthew Schwartz, Heba H. Mostafa |
| hCoV-19/USA/NY-CDC-FG-188383/2021 | EPI_ISL_7698880 | 2021-12-03 | Fulgent Genetics | Centers for Disease Control and Prevention Division of Viral Diseases, Pathogen Discovery | Dakota Howard,Dhwani Batra,Peter Cook,Jason Caravas,Benjamin Rambo-Martin,Scott Sammons,Yvette Unoarumhi,Matthew Schmerer,Kristine Lacek,Tymeckia Kendall,Victoria Caban Figueroa,Shatavia Morrison,Christopher Gulvick,Erisa Sula,Harry Gao,Mickey Li,John Gao |
| hCoV-19/Germany/ST-MD4870/2021 | EPI_ISL_7698906 | 2021-11-12 | Institute of Medical Microbiology and Hospital Hygiene | Institute of Medical Microbiology and Hospital Hygiene | Prof. Dr. Achim Kaasch, Aljoscha Tersteegen |

| | | | | | |
|---|---|---|---|---|---|
| hCoV-19/India/KA-NIMH-SEQ-3563/2021 | EPI_ISL_7968672 | 2021-10-27 | NIMHANS | INSACOG-KA , NIMHANS | Sony Sharma, Mahesh Kumar.C.S, Ellango Ramasamy, Ananthapadmanabha Kotambail, Chetan G K, Gautham Arunachal Udupi, Anita S Desai, V Ravi |
| hCoV-19/SouthAfrica/NICD-N23732/2021 | EPI_ISL_7701120 | 2021-12-07 | LANCET LABORATORY | Diseases of the National Health Laboratory Service | Amoako DG, Everatt J, Scheepers C, Mohale T, Ntuli N, Mahlangu B, Mnguni A, Ismail A, Bhiman JN, Wolter N |
| hCoV-19/SouthAfrica/NICD-N23737/2021 | EPI_ISL_7701129 | 2021-12-09 | LANCET LABORATORY | Diseases of the National Health Laboratory Service | Amoako DG, Everatt J, Scheepers C, Mohale T, Ntuli N, Mahlangu B, Mnguni A, Ismail A, Bhiman JN, Wolter N |
| hCoV-19/USA/MN-CDC-IBX560743662207/2021 | EPI_ISL_7702322 | 2021-12-03 | Infinity Biologix | Centers for Disease Control and Prevention Division of Viral Diseases, Pathogen Discovery | Dakota Howard,Dhwani Batra,Peter Cook,Jason Caravas,Benjamin Rambo-Martin,Scott Sammons,Yvette Unoarumhi,Matthew Schmerer,Kristine Lacek,Tymeckia Kendall,Victoria Caban Figueroa,Shatavia Morrison,Christopher Gulvick,Erisa Sula,Christian Bixby,Yihe Wang,Jo |
| hCoV-19/Canada/QC-L00380078001/2021 | EPI_ISL_7704357 | 2021-08-22 | Laboratoire de santv© publique du Quv©bec | Laboratoire de santv© publique du Quv©bec | Sandrine Moreira, Ioannis Ragoussis, Guillaume Bourque, Jesse Shapiro, Mark Lathrop and Judith Fafard on behalf of the CoVSeQ research group |
| hCoV-19/Canada/QC-L00382534001/2021 | EPI_ISL_7705727 | 2021-08-26 | Laboratoire de santv© publique du Quv©bec | Laboratoire de santv© publique du Quv©bec | Sandrine Moreira, Ioannis Ragoussis, Guillaume Bourque, Jesse Shapiro, Mark Lathrop and Judith Fafard on behalf of the CoVSeQ research group |
| hCoV-19/NewZealand/21CH0797/2021 | EPI_ISL_7706806 | 2021-11-23 | Canterbury Health Laboratories | Institute of Environmental Science and Research (ESR) | Sarah Cockerton, Clare Gebbie, Dhanisha Patel, Rachel Boyle, SallyAnn Harbison, Olivia Stroeven, Xiaoyun Ren, Matt Storey, Nikki Freed, Muhammad Faisal, Jing Wang, Hermes Perez, Anja Werno, Antje van der Linden, Arlo Upton, Chris Mansell, David Hammer, Dr |
| hCoV-19/Canada/QC-1nIOU-B2053274/2021 | EPI_ISL_7707275 | 2021-08-04 | Laboratoire de santv© publique du Quv©bec | Laboratoire de santv© publique du Quv©bec | Sandrine Moreira, Ioannis Ragoussis, Guillaume Bourque, Jesse Shapiro, Mark Lathrop and Judith Fafard on behalf of the CoVSeQ research group |
| hCoV-19/Canada/QC-1nDJXQ-2122002000A/2021 | EPI_ISL_7707386 | 2021-08-08 | Laboratoire de santv© publique du Quv©bec | Laboratoire de santv© publique du Quv©bec | Sandrine Moreira, Ioannis Ragoussis, Guillaume Bourque, Jesse Shapiro, Mark Lathrop and Judith Fafard on behalf of the CoVSeQ research group |
| hCoV-19/Switzerland/BE-Risch-21C1304640/2021 | EPI_ISL_7710496 | 2021-11-30 | Dr. Risch Ostschweiz AG | Dr Risch Laboratory | Nadia Wohlwend, Faina Wehrli, Dominique Fabien Hilti, Sinem Kas, Martin Risch, Thomas Bodmer, Lorenz Risch |
| hCoV-19/Poland/WSSEGorzow-21S1959/2021 | EPI_ISL_7710836 | 2021-12-01 | Szpital Uniwersytecki imienia Karola Marcinkowskiego w Zielonej Gv≥rze spv≥≈Çka z o.o. | Wojewodzka Stacja Sanitarno-Epidemiologiczna w Gorzowie Wielkopolskim | Elzbieta Justynska, Renata Siegel, Klaudia Kobendza-Wlodarczak, Patrycja Faberska and Marek Margol |
| hCoV-19/Canada/QC-L00358133/2021 | EPI_ISL_7713683 | 2021-05-31 | Laboratoire de santv© publique du Quv©bec | Laboratoire de santv© publique du Quv©bec | Sandrine Moreira, Ioannis Ragoussis, Guillaume Bourque, Jesse Shapiro, Mark Lathrop and Judith Fafard on behalf of the CoVSeQ research group |
| hCoV-19/Canada/QC-L00361459001/2021 | EPI_ISL_7713710 | 2021-06-18 | Laboratoire de santv© publique du Quv©bec | Laboratoire de santv© publique du Quv©bec | Sandrine Moreira, Ioannis Ragoussis, Guillaume Bourque, Jesse Shapiro, Mark Lathrop and Judith Fafard on behalf of the CoVSeQ research group |
| hCoV-19/Canada/QC-L00375428001/2021 | EPI_ISL_7713817 | 2021-08-10 | Laboratoire de santv© publique du Quv©bec | Laboratoire de santv© publique du Quv©bec | Sandrine Moreira, Ioannis Ragoussis, Guillaume Bourque, Jesse Shapiro, Mark Lathrop and Judith Fafard on behalf of the CoVSeQ research group |

| | | | | | |
|---|---|---|---|---|---|
| hCoV-19/Canada/QC-L00375711001/2021 | EPI_ISL_7713957 | 2021-08-10 | Laboratoire de sant√© publique du Qu√©bec | Laboratoire de sant√© publique du Qu√©bec | Sandrine Moreira, Ioannis Ragoussis, Guillaume Bourque, Jesse Shapiro, Mark Lathrop and Judith Fafard on behalf of the CoVSeQ research group |
| hCoV-19/Canada/QC-L00375991001/2021 | EPI_ISL_7714022 | 2021-08-13 | Laboratoire de sant√© publique du Qu√©bec | Laboratoire de sant√© publique du Qu√©bec | Sandrine Moreira, Ioannis Ragoussis, Guillaume Bourque, Jesse Shapiro, Mark Lathrop and Judith Fafard on behalf of the CoVSeQ research group |
| hCoV-19/Canada/QC-L00377826001/2021 | EPI_ISL_7714339 | 2021-08-15 | Laboratoire de sant√© publique du Qu√©bec | Laboratoire de sant√© publique du Qu√©bec | Sandrine Moreira, Ioannis Ragoussis, Guillaume Bourque, Jesse Shapiro, Mark Lathrop and Judith Fafard on behalf of the CoVSeQ research group |
| hCoV-19/SouthAfrica/UFS-VIRO-NGS-MSQ101221BC82/2021 | EPI_ISL_7714413 | 2021-12-01 | NHLS Universitas Academic | UFS Virology | PA Bester, MM Nyaga, P Nthiga, MT Mogotsi, Emmanuel Ogunbayo, D Goedhals, T de Oliveira |
| hCoV-19/SouthAfrica/CERI-KRISP-K033221/2021 | EPI_ISL_7715374 | 2021-11-24 | National Health Laboratory Services, Virology | and Innvoation, Stellenbosch University and KRISP, KZN Research Innovation and Sequencing Platform, UKZN. | Kathleen Subramoney, Florette Treurnicht, Nkhensani Mtileni, Avani Bharuthram, Giandhari J, Pillay S, Naidoo Y, Ramphal U, Ramphal Y, Arisha Maharaj, San JE, Tshiabuila D, Tegally H, Wilkinson E, de Oliveira T |
| hCoV-19/Mozambique/CERI-KRISP-K032767/2021 | EPI_ISL_7715463 | 2021-11-30 | Instituto Nacional de Saude (INS), Mozambique | and Innvoation, Stellenbosch University and KRISP, KZN Research Innovation and Sequencing Platform, UKZN. | Nalia Ismael, Nadia Sitoe, Paulo Arnaldo, Nedio Mabunda, Giandhari J, Pillay S, Naidoo Y, Ramphal U, Ramphal Y, Arisha Maharaj, van Wyk S, Moir M, San JE, Tshiabuila D, Tegally H, Wilkinson E, de Oliveira T |
| hCoV-19/USA/IA-SHL-1959371/2021 | EPI_ISL_7716090 | 2021-12-05 | State Hygienic Laboratory at the University of Iowa | State Hygienic Laboratory at the University of Iowa | Valerie Reeb, Erik Twait, Anna Yakos, Mariah Knutson, Kris Eveland, Kristen Zanon, Jeff Benfer, Davis Rieckenberg, Cindy Toll, Wes Hottel, Alankar Kampoowale, Krishnaveni Sompallae, Mohammed Allam |
| hCoV-19/USA/NJ-PHEL-V21032052/2021 | EPI_ISL_7716282 | 2021-11-10 | NJDOH, Public Health and Environmental Laboratories | NJ_PHEL | Ryan Pachucki, Shiv K. Verma, Lindsey Bodnar, Maria-Magdalene Pugliese, Chelsea San Filippo, Mohammad M. Ali, Dana Woell, Byeong Jeong |
| hCoV-19/Estonia/17935831/2021 | EPI_ISL_7717818 | 2021-11-16 | Synlab Eesti O√ú | 1. Laboratory of Communicable Diseases (Estonia); 2. Eurofins Genomics Europe Sequencing GmbH | Abroi A., Lahesaare A., Shablinskaja A., Hoidmets D., Jaaniso E., Kallas E., Sepp H., Peterson H., Niglas H., Suija H., Lutsar I., Kuzmin I., Koppel I., Ep≈°tein J., Truusalu K., Ratnik K., Kaarna K., Huik K., Dotsenko L., Milani L., Metspalu M., H√§rma M |
| hCoV-19/Madagascar/LA2M-112743/2021 | EPI_ISL_7722899 | 2021-07-13 | Virology Unit, Institut Pasteur de Madagascar | Virology Unit, Institut Pasteur de Madagascar | Christian Ranaivoson, Cara E. Brook, Vida Ahyong, Soa Fy Andriamandimby, Vololoniaina Raharinosy, Tsiry Randriambolamanantsoa, Helisoa Razafimanjato, Norosoa Razanajatovo, Andry Andrianarivelo, Zely Arivelo Randriamanantany, Michelle Tan, Cristina M. Tato |
| hCoV-19/USA/MS-UMMC-M1645E6-518122/2021 | EPI_ISL_7724496 | 2021-12-01 | University of Mississippi Medical Center, Department of Pathology | University of Mississippi Medical Center, Molecular and Genomics Core Facility | Ashley C. Johnson, Wenjie Wu, Ithiel J. Frame, Krishna K. Ayyalasomayajula, Michael R. Garrett, D. Ashley Robinson |
| hCoV-19/England/PHEP-YYNQUJ7/2021 | EPI_ISL_7727570 | 2021-11-29 | Respiratory Virus Unit, Microbiology Services Colindale, Public Health England | COVID-19 Genomics UK (COG-UK) Consortium | PHE Covid Sequencing Team |
| hCoV-19/Suriname/SR-764/2021 | EPI_ISL_7730626 | 2021-10-10 | Central Laboratory, Bureau of Public Health (BOG) and Academic Hospital Paramaribo | Erasmus Medical Center | Bas B Oude Munnink, Cherise Beek, Consuella Partowidjojo, Dion Gajadin, Ed PF IJzerman, Emmanuelle Munger, Gary Gummels, Ingrid SK Krishnadath, Lycke Woittiez, Marion PG Koopmans, Mireille Van de Veer, Phyllis Pinas, Princes Wongsowidjojo, Radjesh Ori, Ra |

| | | | | | |
|---|---|---|---|---|---|
| hCoV-19/India/TG-RFCH01875_CIB5611/2020 | EPI_ISL_7734673 | 2020-11-21 | CSIR-Centre for Cellular and Molecular Biology | CSIR-Centre for Cellular and Molecular Biology | Valli Nagalakshmi Undamatla,Amareshwar Vodapalli, Sumedha Avadhanula,B Himasri,Priya Nurkurthy,Vidhyadhari Methuku,Shreekant Verma,Ara Sreenivas, Sreelekshmi MS,Malini Nemalikanti,Lamuk Zaveri,Tulasi Nagabandi,Jandhyala Sai Krishna, Archana Bharadwaj Siv |
| hCoV-19/India/TG-RFCH01944_CIB5682/2020 | EPI_ISL_7734730 | 2020-12-11 | CSIR-Centre for Cellular and Molecular Biology | CSIR-Centre for Cellular and Molecular Biology | Amareshwar Vodapalli,B Himasri,Valli Nagalakshmi Undamatla, Sumedha Avadhanula,Priya Nurkurthy,Vidhyadhari Methuku,Shreekant Verma,Ara Sreenivas, Sreelekshmi MS,Malini Nemalikanti,Lamuk Zaveri,Tulasi Nagabandi,Jandhyala Sai Krishna, Archana Bharadwaj Siv |
| hCoV-19/India/AP-CCMB-CIB5756/2021 | EPI_ISL_7734774 | 2021-11-12 | AP SSO | CSIR-Centre for Cellular and Molecular Biology-INSACOG | Amareshwar Vodapalli,B Himasri,Valli Nagalakshmi Undamatla, Sumedha Avadhanula,Priya Nurkurthy,Vidhyadhari Methuku,Shreekant Verma,Ara Sreenivas, Sreelekshmi MS,Malini Nemalikanti,Lamuk Zaveri,Tulasi Nagabandi,Jandhyala Sai Krishna, Archana Bharadwaj Siv |
| hCoV-19/India/TG-RFCH02044_CIB6091/2021 | EPI_ISL_7735101 | 2021-10-12 | MapMyGenome | CSIR-Centre for Cellular and Molecular Biology | Lamuk Zaveri,Tulasi Nagabandi,Jandhyala Sai Krishna,Malini Nemalikanti,Sreelekshmi MS, Ara Sreenivas,Shreekant Verma, Amareshwar Vodapalli,B Himasri,Valli Nagalakshmi Undamatla, Sumedha Avadhanula,Priya Nurkurthy,Vidhyadhari Methuku,M. Aravind Kumar, San |
| hCoV-19/India/TG-RFCH02076_CIB6305/2021 | EPI_ISL_7735115 | 2021-05-26 | MapMyGenome | CSIR-Centre for Cellular and Molecular Biology | Lamuk Zaveri,Tulasi Nagabandi,Jandhyala Sai Krishna,Malini Nemalikanti,Sreelekshmi MS, Ara Sreenivas,Shreekant Verma, Amareshwar Vodapalli,B Himasri,Valli Nagalakshmi Undamatla, Sumedha Avadhanula,Priya Nurkurthy,Vidhyadhari Methuku,M. Aravind Kumar, San |
| hCoV-19/France/IDF-HMN-21122080434/2021 | EPI_ISL_7735810 | 2021-11-12 | Laboratoire Ana-L | [illegible] Mondor University Hospital, Assistance Publique H√¥pitaux de Paris, Universit√© Paris-Est Cr√©teil, INSERM U955 | Christophe Rodriguez, Slim Fourati, Vanessa Demontant, Guillaume Gricourt, Melissa N'Debi, Alexandre Soulier, Elisabeth Trawinski, Jean-Michel Pawlotsky |
| hCoV-19/Chile/RM-186917/2021 | EPI_ISL_7736007 | 2021-11-25 | Genetica Molecular and Subdepartamento de Virologia ISP Chile | Instituto de Salud Publica de Chile | Karen Orostica, Constanza Campano, Barbara Parra, Patricia Bustos, Rodrigo Fasce, Andres Castillo, Jorge Fernandez |
| hCoV-19/PapuaNewGuinea/PNG853/2021 | EPI_ISL_7737616 | 2021-03-25 | Queensland Medical Laboratories | Microbiological Diagnostic Unit - Public Health Laboratory (MDU-PHL) | Susapu, M., Palou, T., Seemann, T., Horan, K., Sherry, N.L. |
| hCoV-19/PapuaNewGuinea/PNG4436/2021 | EPI_ISL_7737618 | 2021-08-28 | Central Public Health Laboratory | Microbiological Diagnostic Unit - Public Health Laboratory (MDU-PHL) | Kumbu, J., Susapu, M., Palou, T., Seemann, T., Horan, K., Sherry, N.L. |

| | | | | | |
|---|---|---|---|---|---|
| hCoV-19/USA/WA-PHL-007851/2021 | EPI_ISL_7738151 | 2021-11-30 | Washington State Department of Health Public Health Laboratories | Washington State Department of Health Public Health Laboratories | Drew MacKellar, Philip Dykema, Denny Russell, Lisa Jones, Holly Halstead, Kathryn Sickles, Kristin Roche, Ardizon Valdez, Claire Howell, Alex Lathum, JohnAric Peterson, Avi Singh, Rebecca Cao |
| hCoV-19/USA/WA-CDC-UW21112996251/2021 | EPI_ISL_7738943 | 2021-11-29 | UW Virology Lab | UW Virology Lab | Pavitra Roychoudhury, Shah Mohamed Bakhash, Seffir T. Wendm, Isabel Arnould, Pooneh Hajian, Hong Xie, Sean Ellis, Nathan Breit, Robert J. Livingston, Meei-Li Huang, Keith R Jerome, Patrick Mathias, Alexander Greninger |
| hCoV-19/SouthAfrica/NICD-N22522/2021 | EPI_ISL_7740821 | 2021-11-29 | HELEN JOSEPH LABORATORY | Diseases of the National Health Laboratory Service | Amoako DG, Everatt J, Scheepers C, Mohale T, Ntuli N, Mahlangu B, Mnguni A, Ismail A, Bhiman JN, Wolter N |
| hCoV-19/SouthAfrica/NICD-N22537/2021 | EPI_ISL_7740825 | 2021-11-26 | HELEN JOSEPH LABORATORY | Diseases of the National Health Laboratory Service | Amoako DG, Everatt J, Scheepers C, Mohale T, Ntuli N, Mahlangu B, Mnguni A, Ismail A, Bhiman JN, Wolter N |
| hCoV-19/SouthAfrica/NICD-N22544/2021 | EPI_ISL_7740831 | 2021-11-26 | HELEN JOSEPH LABORATORY | Diseases of the National Health Laboratory Service | Amoako DG, Everatt J, Scheepers C, Mohale T, Ntuli N, Mahlangu B, Mnguni A, Ismail A, Bhiman JN, Wolter N |
| hCoV-19/SouthAfrica/NICD-N22577/2021 | EPI_ISL_7740846 | 2021-11-28 | HELEN JOSEPH LABORATORY | Diseases of the National Health Laboratory Service | Amoako DG, Everatt J, Scheepers C, Mohale T, Ntuli N, Mahlangu B, Mnguni A, Ismail A, Bhiman JN, Wolter N |
| hCoV-19/SouthAfrica/NICD-N22582/2021 | EPI_ISL_7740849 | 2021-11-25 | HELEN JOSEPH LABORATORY | Diseases of the National Health Laboratory Service | Amoako DG, Everatt J, Scheepers C, Mohale T, Ntuli N, Mahlangu B, Mnguni A, Ismail A, Bhiman JN, Wolter N |
| hCoV-19/SouthAfrica/NICD-N22435/2021 | EPI_ISL_7740899 | 2021-11-29 | HELEN JOSEPH LABORATORY | Diseases of the National Health Laboratory Service | Amoako DG, Everatt J, Scheepers C, Mohale T, Ntuli N, Mahlangu B, Mnguni A, Ismail A, Bhiman JN, Wolter N |
| hCoV-19/USA/CA-CDC-FG-186214/2021 | EPI_ISL_7741531 | 2021-12-05 | Fulgent Genetics | Centers for Disease Control and Prevention Division of Viral Diseases, Pathogen Discovery | Dakota Howard,Dhwani Batra,Peter Cook,Jason Caravas,Benjamin Rambo-Martin,Scott Sammons,Yvette Unoarumhi,Matthew Schmerer,Kristine Lacek,Tymeckia Kendall,Victoria Caban Figueroa,Shatavia Morrison,Christopher Gulvick,Erisa Sula,Harry Gao,Mickey Li,John Gao |
| hCoV-19/USA/AZ-TG1088574/2021 | EPI_ISL_7743493 | 2021-07-21 | TGen North | TGen North | Jolene Bowers, Heather Centner, Brett Van Tassel, Chris French, Hayley Yaglom, Darrin Lemmer, Dave Engelthaler |
| hCoV-19/USA/MN-CDC-IBX581089872811/2021 | EPI_ISL_7744731 | 2021-11-24 | Infinity Biologix | Centers for Disease Control and Prevention Division of Viral Diseases, Pathogen Discovery | Dakota Howard,Dhwani Batra,Peter Cook,Jason Caravas,Benjamin Rambo-Martin,Scott Sammons,Yvette Unoarumhi,Matthew Schmerer,Kristine Lacek,Tymeckia Kendall,Victoria Caban Figueroa,Shatavia Morrison,Christopher Gulvick,Erisa Sula,Christian Bixby,Yihe Wang,Jo |
| hCoV-19/SouthAfrica/CERI-KRISP-K033360/2021 | EPI_ISL_7747537 | 2021-12-10 | National Health Laboratory Service, Kwazulu-Natal, South Africa | and Innvoation, Stellenbosch University and KRISP, KZN Research Innovation and Sequencing Platform, UKZN. | Nokukhanya Mdlalose,Giandhari J, Pillay S, Naidoo Y, Ramphal U, Ramphal Y, Arisha Maharaj, San JE, Tshiabuila D, Tegally H, Wilkinson E, de Oliveira T |
| hCoV-19/SouthAfrica/CERI-KRISP-K033361/2021 | EPI_ISL_7747538 | 2021-12-10 | National Health Laboratory Service, Kwazulu-Natal, South Africa | and Innvoation, Stellenbosch University and KRISP, KZN Research Innovation and Sequencing Platform, UKZN. | Nokukhanya Mdlalose,Giandhari J, Pillay S, Naidoo Y, Ramphal U, Ramphal Y, Arisha Maharaj, San JE, Tshiabuila D, Tegally H, Wilkinson E, de Oliveira T |
| hCoV-19/SouthAfrica/CERI-KRISP-K033441/2021 | EPI_ISL_7747604 | 2021-12-06 | National Health Laboratory Service, Kwazulu-Natal, South Africa | and Innvoation, Stellenbosch University and KRISP, KZN Research Innovation and Sequencing Platform, UKZN. | Nokukhanya Mdlalose,Giandhari J, Pillay S, Naidoo Y, Ramphal U, Ramphal Y, Arisha Maharaj, San JE, Tshiabuila D, Tegally H, Wilkinson E, de Oliveira T |

| | | | | | |
|---|---|---|---|---|---|
| hCoV-19/Australia/NSW-ICPMR-17200/2021 | EPI_ISL_7748798 | 2021-12-08 | Histopath | Clinical Pathology and Medical Research; Westmead Hospital; University of Sydney | Arnott A., Draper J., Gall M., Martinez E., Rockett R., Sintchenko V., on behalf of ICPMR |
| hCoV-19/Australia/NSW-ICPMR-17302/2021 | EPI_ISL_7749039 | 2021-12-12 | Pathology North - Royal North Shore Hospital - NSW Health Pathology | Clinical Pathology and Medical Research; Westmead Hospital; University of Sydney | Arnott A., Draper J., Gall M., Martinez E., Rockett R., Sintchenko V., on behalf of ICPMR |
| hCoV-19/Australia/NSW-ICPMR-17331/2021 | EPI_ISL_7749109 | 2021-12-12 | Histopath | Clinical Pathology and Medical Research; Westmead Hospital; University of Sydney | Arnott A., Draper J., Gall M., Martinez E., Rockett R., Sintchenko V., on behalf of ICPMR |
| hCoV-19/Argentina/INEI110155/2021 | EPI_ISL_7751232 | 2021-08-26 | Servicio Virosis Respiratorias-Departamento Virologia-INEI | Instituto Nacional Enfermedades Infecciosas C.G.Malbran | Baumeister E., Avaro M., Benedetti E., Russo M., Dattero ME, Pontoriero A., Cisterna D., Molina V., Perandones C., Tuduri E., De Belder D., Sanchez Loria J., Haim MS., Lorenzo F., Poklepovich T., Campos J. |
| hCoV-19/Argentina/INEI112386/2021 | EPI_ISL_7751972 | 2021-10-23 | Servicio Virosis Respiratorias-Departamento Virologia-INEI | Instituto Nacional Enfermedades Infecciosas C.G.Malbran | Baumeister E., Avaro M., Benedetti E., Russo M., Dattero ME, Pontoriero A., Cisterna D., Molina V., Perandones C., Tuduri E., De Belder D., Sanchez Loria J., Haim MS., Lorenzo F., Poklepovich T., Campos J. |
| hCoV-19/Argentina/INEI112898/2021 | EPI_ISL_7752883 | 2021-10-24 | Servicio Virosis Respiratorias-Departamento Virologia-INEI | Instituto Nacional Enfermedades Infecciosas C.G.Malbran | Baumeister E., Avaro M., Benedetti E., Russo M., Dattero ME, Pontoriero A., Cisterna D., Molina V., Perandones C., Tuduri E., De Belder D., Sanchez Loria J., Haim MS., Lorenzo F., Poklepovich T., Campos J. |
| hCoV-19/Argentina/INEI113050/2021 | EPI_ISL_7753148 | 2021-11-03 | Servicio Virosis Respiratorias-Departamento Virologia-INEI | Instituto Nacional Enfermedades Infecciosas C.G.Malbran | Baumeister E., Avaro M., Benedetti E., Russo M., Dattero ME, Pontoriero A., Cisterna D., Molina V., Perandones C., Tuduri E., De Belder D., Sanchez Loria J., Haim MS., Lorenzo F., Poklepovich T., Campos J. |
| hCoV-19/Argentina/INEI113249/2021 | EPI_ISL_7753627 | 2021-11-05 | Servicio Virosis Respiratorias-Departamento Virologia-INEI | Instituto Nacional Enfermedades Infecciosas C.G.Malbran | Baumeister E., Avaro M., Benedetti E., Russo M., Dattero ME, Pontoriero A., Cisterna D., Molina V., Perandones C., Tuduri E., De Belder D., Sanchez Loria J., Haim MS., Lorenzo F., Poklepovich T., Campos J. |
| hCoV-19/Canada/AB-ABPHL-42225/2021 | EPI_ISL_7298600 | 2021-11-17 | Edmonton Provincial Lab | Alberta Precision Labs (APL) | Buss, E, Croxen M, Deo A, Dieu P, Gill K, Ferrato C, Khan F, Koleva P, Li V, Lloyd C, Lynch T, Ma R, Murphy S, Pabbaraju K, Shokoples S, Tipples G, Thayer J, Whitehouse M, Wong A, Yu C, Zelyas N |
| hCoV-19/Australia/VIC27815/2021 | EPI_ISL_7766351 | 2021-11-24 | Microbiological Diagnostic Unit - Public Health Laboratory (MDU-PHL) | Microbiological Diagnostic Unit - Public Health Laboratory (MDU-PHL) | Seemann, T., Horan, K., Sherry, N.L. |
| hCoV-19/Australia/VIC28245/2021 | EPI_ISL_7768770 | 2021-11-26 | Microbiological Diagnostic Unit - Public Health Laboratory (MDU-PHL) | Microbiological Diagnostic Unit - Public Health Laboratory (MDU-PHL) | Seemann, T., Horan, K., Sherry, N.L. |
| hCoV-19/Australia/VIC28247/2021 | EPI_ISL_7768793 | 2021-11-25 | Microbiological Diagnostic Unit - Public Health Laboratory (MDU-PHL) | Microbiological Diagnostic Unit - Public Health Laboratory (MDU-PHL) | Seemann, T., Horan, K., Sherry, N.L. |
| hCoV-19/Australia/VIC28992/2021 | EPI_ISL_7773505 | 2021-12-10 | Microbiological Diagnostic Unit - Public Health Laboratory (MDU-PHL) | Microbiological Diagnostic Unit - Public Health Laboratory (MDU-PHL) | Seemann, T., Horan, K., Sherry, N.L. |
| hCoV-19/Reunion/CHU721111855901/2021 | EPI_ISL_7774149 | 2021-11-19 | LABM SAINT BENOIT | Laboratoire de virologie, CNR arbovirus Associ√©, Chu de la R√©union | Etienne Frumence, Nicolas M'nemosyme, Anne-Julie Gourd√©, Rubens Lhonneur, Nicolas Traversier, Marie-Christine Jaffar Bandjee |

| | | | | | |
|---|---|---|---|---|---|
| hCoV-19/Reunion/CHU721111615201/2021 | EPI_ISL_7774153 | 2021-11-17 | LABM SAINT BENOIT | Laboratoire de virologie, CNR arbovirus Associ√©, Chu de la R√©union | Etienne Frumence, Nicolas M'nemosyme, Anne-Julie Gourd√©, Rubens Lhonneur, Nicolas Traversier, Marie-Christine Jaffar Bandjee |
| hCoV-19/Reunion/CHU721120201006/2021 | EPI_ISL_7774155 | 2021-12-02 | CHU - Saint-Denis | Laboratoire de virologie, CNR arbovirus Associ√©, Chu de la R√©union | Etienne Frumence, Nicolas M'nemosyme, Anne-Julie Gourd√©, Rubens Lhonneur, Nicolas Traversier, Marie-Christine Jaffar Bandjee |
| hCoV-19/Reunion/CHU79800009699803/2021 | EPI_ISL_7774209 | 2021-11-29 | CHU - Saint-Denis | Laboratoire de virologie, CNR arbovirus Associ√©, Chu de la R√©union | Etienne Frumence, Nicolas M'nemosyme, Anne-Julie Gourd√©, Rubens Lhonneur, Nicolas Traversier, Marie-Christine Jaffar Bandjee |
| hCoV-19/Reunion/CHU721111736002/2021 | EPI_ISL_7774299 | 2021-11-17 | CHOR Laboratoire | Laboratoire de virologie, CNR arbovirus Associ√©, Chu de la R√©union | Etienne Frumence, Nicolas M'nemosyme, Anne-Julie Gourd√©, Rubens Lhonneur, Nicolas Traversier, Marie-Christine Jaffar Bandjee |
| hCoV-19/Reunion/CHU721111948903/2021 | EPI_ISL_7774344 | 2021-11-20 | CHU - Saint-Pierre | Laboratoire de virologie, CNR arbovirus Associ√©, Chu de la R√©union | Etienne Frumence, Nicolas M'nemosyme, Anne-Julie Gourd√©, Rubens Lhonneur, Nicolas Traversier, Marie-Christine Jaffar Bandjee |
| hCoV-19/Reunion/CHU721110606503/2021 | EPI_ISL_7774465 | 2021-11-06 | CHU - Saint-Pierre | Laboratoire de virologie, CNR arbovirus Associ√©, Chu de la R√©union | Etienne Frumence, Nicolas M'nemosyme, Anne-Julie Gourd√©, Rubens Lhonneur, Nicolas Traversier, Marie-Christine Jaffar Bandjee |
| hCoV-19/Reunion/CHU721120636801/2021 | EPI_ISL_7774478 | 2021-12-07 | LABM SAINT BENOIT | Laboratoire de virologie, CNR arbovirus Associ√©, Chu de la R√©union | Etienne Frumence, Nicolas M'nemosyme, Anne-Julie Gourd√©, Rubens Lhonneur, Nicolas Traversier, Marie-Christine Jaffar Bandjee |
| hCoV-19/Norway/29579/2021 | EPI_ISL_7775523 | 2021-11-26 | Haukeland University Hospital, Dept. of Microbiology | Norwegian Institute of Public Health, Department of Virology | Kathrine Stene-Johansen, Kamilla Heddeland Instefjord, Hilde Elshaug, Garcia Llorente Ignacio, Jon Br√•te, Engebretsen Serina Beate, Pedersen Benedikte Nevjen, Line Victoria Moen, Debech Nadia, Atiya R Ali, Marie Paulsen Madsen, Rasmus Riis Kopperud, Hild |
| hCoV-19/Chile/LL-182448/2021 | EPI_ISL_7778152 | 2021-11-11 | Genetica Molecular and Subdepartamento de Virologia ISP Chile | Instituto de Salud Publica de Chile | Karen Orostica, Constanza Campano, Barbara Parra, Patricia Bustos, Rodrigo Fasce, Andres Castillo, Jorge Fernandez |
| hCoV-19/USA/NY-Wadsworth-21104305-01/2021 | EPI_ISL_7782595 | 2021-11-30 | BIO-REFERENCE LABORATORIES | Wadsworth Center, New York State Department of Health | Kirsten St. George, Daryl M. Lamson, Alexis Russell, Matthew Shudt, Melissa A Leisner, Jonathan Plitnick, Catharine Prussing, Navjot Singh, John Kelly, Erasmus Schneider, Erica Lasek-Nesselquist |
| hCoV-19/France/NOR-HMN-21122080810/2021 | EPI_ISL_7783443 | 2021-12-06 | LBM du C√®dre | Mondor University Hospital, Assistance Publique H√¥pitaux de Paris, Universit√© Paris-Est Cr√©teil, INSERM U955 | Christophe Rodriguez, Slim Fourati, Vanessa Demontant, Guillaume Gricourt, Melissa N'Debi, Alexandre Soulier, Elisabeth Trawinski, Jean-Michel Pawlotsky |
| hCoV-19/USA/CT-CDC-ASC210488337/2021 | EPI_ISL_7783914 | 2021-11-19 | Aegis Sciences Corporation | Centers for Disease Control and Prevention Division of Viral Diseases, Pathogen Discovery | Dakota Howard,Dhwani Batra,Peter Cook,Jason Caravas,Benjamin Rambo-Martin,Scott Sammons,Yvette Unoarumhi,Matthew Schmerer,Kristine Lacek,Tymeckia Kendall,Victoria Caban Figueroa,Shatavia Morrison,Christopher Gulvick,Erisa Sula,Cyndi Clark,Patrick Campbell |

| | | | | | |
|---|---|---|---|---|---|
| hCoV-19/Aruba/AW-RIVM-74148/2021 | EPI_ISL_7785004 | 2021-12-01 | Dutch COVID-19 response team | National Institute for Public Health and the Environment (RIVM) | Adam Meijer, Harry Vennema, Dirk Eggink, Jeroen Cremer, Sharon van den Brink, Bas van der Veer, AnneMarie van den Brandt, Lisa Wijsman, Kim Freriks, Ryanne Jaarsma, Lynn Aarts, Sanne Bos, Jil Kocken, Stijn van Rossum, Linda van Someren, Florian Zwagemaker |
| hCoV-19/USA/IN-INSPHL-12931/2021 | EPI_ISL_7792354 | 2021-12-08 | IN State Department of Health Laboratory Services | IN State Department of Health Laboratory Services | Cassandra Campion, Jamie Yeadon, Brian Pope, Lixia Liu, Kyle Brownlee, Melissa Hindenlang, Mark Glazier |
| hCoV-19/SouthKorea/KDCA19840/2021 | EPI_ISL_7792855 | 2021-11-04 | Division of Emerging Infectious Diseases, Bureau of Infectious Diseases Diagnosis Control, Korea Disease Control and Prevention Agency | Diseases, Bureau of Infectious Diseases Diagnosis Control, Korea Disease Control and Prevention Agency | Ae Kyung Park, Il-Hwan Kim, Jeong-Ah Kim, Chae Young Lee, Hyuck Jin Lee, Eun-Jin Kim |
| hCoV-19/SouthKorea/KDCA21048/2021 | EPI_ISL_7793938 | 2021-11-03 | Division of Emerging Infectious Diseases, Bureau of Infectious Diseases Diagnosis Control, Korea Disease Control and Prevention Agency | Diseases, Bureau of Infectious Diseases Diagnosis Control, Korea Disease Control and Prevention Agency | Ae Kyung Park, Il-Hwan Kim, Jeong-Ah Kim, Chae Young Lee, Hyuck Jin Lee, Eun-Jin Kim |
| hCoV-19/SouthKorea/KDCA21142/2021 | EPI_ISL_7794007 | 2021-11-19 | Division of Emerging Infectious Diseases, Bureau of Infectious Diseases Diagnosis Control, Korea Disease Control and Prevention Agency | Diseases, Bureau of Infectious Diseases Diagnosis Control, Korea Disease Control and Prevention Agency | Ae Kyung Park, Il-Hwan Kim, Jeong-Ah Kim, Chae Young Lee, Hyuck Jin Lee, Eun-Jin Kim |
| hCoV-19/PapuaNewGuinea/PNG5947/2021 | EPI_ISL_7799136 | 2021-10-03 | Central Public Health Laboratory | Microbiological Diagnostic Unit - Public Health Laboratory (MDU-PHL) | Kumbu, J., Susapu, M., Palou, T., Seemann, T., Horan, K., Sherry, N.L. |
| hCoV-19/PapuaNewGuinea/PNG6832/2021 | EPI_ISL_7799471 | 2021-10-20 | Central Public Health Laboratory | Microbiological Diagnostic Unit - Public Health Laboratory (MDU-PHL) | Kumbu, J., Susapu, M., Palou, T., Seemann, T., Horan, K., Sherry, N.L. |
| hCoV-19/USA/WA-CDC-FG-189904/2021 | EPI_ISL_7799889 | 2021-12-05 | Fulgent Genetics | Centers for Disease Control and Prevention Division of Viral Diseases, Pathogen Discovery | Dakota Howard,Dhwani Batra,Peter Cook,Jason Caravas,Benjamin Rambo-Martin,Scott Sammons,Yvette Unoarumhi,Matthew Schmerer,Kristine Lacek,Tymeckia Kendall,Victoria Caban Figueroa,Shatavia Morrison,Christopher Gulvick,Erisa Sula,Harry Gao,Mickey Li,John Gao |
| hCoV-19/USA/WA-CDC-FG-189938/2021 | EPI_ISL_7799906 | 2021-12-06 | Fulgent Genetics | Centers for Disease Control and Prevention Division of Viral Diseases, Pathogen Discovery | Dakota Howard,Dhwani Batra,Peter Cook,Jason Caravas,Benjamin Rambo-Martin,Scott Sammons,Yvette Unoarumhi,Matthew Schmerer,Kristine Lacek,Tymeckia Kendall,Victoria Caban Figueroa,Shatavia Morrison,Christopher Gulvick,Erisa Sula,Harry Gao,Mickey Li,John Gao |
| hCoV-19/USA/WI-UW-6789/2021 | EPI_ISL_7800085 | 2021-12-09 | University of Wisconsin-Madison AIDS Vaccine Research Laboratories | University of Wisconsin-Madison AIDS Vaccine Research Laboratories | Gage Moreno, Katarina Braun, et al. AIDS Vaccine Research Laboratories |
| hCoV-19/USA/NY-Wadsworth-21104299-01/2021 | EPI_ISL_7800795 | 2021-11-29 | BIO-REFERENCE LABORATORIES | Wadsworth Center, New York State Department of Health | Kirsten St. George, Daryl M. Lamson, Alexis Russell, Matthew Shudt, Melissa A Leisner, Jonathan Plitnick, Catharine Prussing, Navjot Singh, John Kelly, Erasmus Schneider, Erica Lasek-Nesselquist |

| | | | | | |
|---|---|---|---|---|---|
| hCoV-19/USA/NJ-CDC-IBX667460567737/2021 | EPI_ISL_7801054 | 2021-11-30 | Infinity Biologix | Centers for Disease Control and Prevention Division of Viral Diseases, Pathogen Discovery | Dakota Howard,Dhwani Batra,Peter Cook,Jason Caravas,Benjamin Rambo-Martin,Scott Sammons,Yvette Unoarumhi,Matthew Schmerer,Kristine Lacek,Tymeckia Kendall,Victoria Caban Figueroa,Shatavia Morrison,Christopher Gulvick,Erisa Sula,Christian Bixby,Yihe Wang,Jo |
| hCoV-19/USA/NY-CDC-FG-188489/2021 | EPI_ISL_7801060 | 2021-11-30 | Fulgent Genetics | Centers for Disease Control and Prevention Division of Viral Diseases, Pathogen Discovery | Dakota Howard,Dhwani Batra,Peter Cook,Jason Caravas,Benjamin Rambo-Martin,Scott Sammons,Yvette Unoarumhi,Matthew Schmerer,Kristine Lacek,Tymeckia Kendall,Victoria Caban Figueroa,Shatavia Morrison,Christopher Gulvick,Erisa Sula,Harry Gao,Mickey Li,John Gao |
| hCoV-19/USA/NY-CDC-FG-184640/2021 | EPI_ISL_7801194 | 2021-11-29 | Fulgent Genetics | Centers for Disease Control and Prevention Division of Viral Diseases, Pathogen Discovery | Dakota Howard,Dhwani Batra,Peter Cook,Jason Caravas,Benjamin Rambo-Martin,Scott Sammons,Yvette Unoarumhi,Matthew Schmerer,Kristine Lacek,Tymeckia Kendall,Victoria Caban Figueroa,Shatavia Morrison,Christopher Gulvick,Erisa Sula,Harry Gao,Mickey Li,John Gao |
| hCoV-19/USA/MD-CDC-ASC210654250/2021 | EPI_ISL_7801203 | 2021-11-26 | Aegis Sciences Corporation | Centers for Disease Control and Prevention Division of Viral Diseases, Pathogen Discovery | Dakota Howard,Dhwani Batra,Peter Cook,Jason Caravas,Benjamin Rambo-Martin,Scott Sammons,Yvette Unoarumhi,Matthew Schmerer,Kristine Lacek,Tymeckia Kendall,Victoria Caban Figueroa,Shatavia Morrison,Christopher Gulvick,Erisa Sula,Cyndi Clark,Patrick Campbell |
| hCoV-19/USA/CA-CDC-ASC210657683/2021 | EPI_ISL_7801205 | 2021-11-29 | Aegis Sciences Corporation | Centers for Disease Control and Prevention Division of Viral Diseases, Pathogen Discovery | Dakota Howard,Dhwani Batra,Peter Cook,Jason Caravas,Benjamin Rambo-Martin,Scott Sammons,Yvette Unoarumhi,Matthew Schmerer,Kristine Lacek,Tymeckia Kendall,Victoria Caban Figueroa,Shatavia Morrison,Christopher Gulvick,Erisa Sula,Cyndi Clark,Patrick Campbell |
| hCoV-19/England/LSPA-2C8EF47/2021 | EPI_ISL_7302329 | 2021-11-22 | Rosalind Franklin Laboratory | Wellcome Sanger Institute for the COVID-19 Genomics UK (COG-UK) Consortium | Donald Fraser, Suki Lee, Rob Howes, The Rosalind Franklin Laboratory and Alex Alderton, Roberto Amato, Jeffrey Barrett, Sonia Goncalves, Ewan Harrison, David K. Jackson, Ian Johnston, Dominic Kwiatkowski, Cordelia Langford, John Sillitoe on behalf of the |
| hCoV-19/Brazil/SP-IB_142956/2021 | EPI_ISL_7808758 | 2021-11-29 | PRONTO ATENDIMENTO SAO JOSE | Instituto Butantan | Dimas Tadeu Covas, Antonio Jorge Martins, Claudia Renata dos Santos Barros, David Schlesinger, Debora Botequio Moretti, Elaine Cristina Marqueze, Elaine Vieira Santos, Evandra Strazza Rodrigues, Heidge Fukumasu, Jayme Augusto de Souza-Neto, Jos√© Salvator |
| hCoV-19/Brazil/SP-IB_143206/2021 | EPI_ISL_7809218 | 2021-11-26 | UBS NITRO OPER√ÅRIA (LESTE) | Instituto Butantan | Dimas Tadeu Covas, Antonio Jorge Martins, Claudia Renata dos Santos Barros, David Schlesinger, Debora Botequio Moretti, Elaine Cristina Marqueze, Elaine Vieira Santos, Evandra Strazza Rodrigues, Heidge Fukumasu, Jayme Augusto de Souza-Neto, Jos√© Salvator |

| | | | | | |
|---|---|---|---|---|---|
| hCoV-19/Brazil/SP-IB_143376/2021 | EPI_ISL_7809319 | 2021-12-02 | INSIDE | Instituto Butantan | Dimas Tadeu Covas, Antonio Jorge Martins, Claudia Renata dos Santos Barros, David Schlesinger, Debora Botequio Moretti, Elaine Cristina Marqueze, Elaine Vieira Santos, Evandra Strazza Rodrigues, Heidge Fukumasu, Jayme Augusto de Souza-Neto, Jos√© Salvator |
| hCoV-19/Brazil/SP-IB_143228/2021 | EPI_ISL_7809350 | 2021-12-02 | UNIDADE BASICA DE SAUDE II ESTRELA DO NORTE | Instituto Butantan | Dimas Tadeu Covas, Antonio Jorge Martins, Claudia Renata dos Santos Barros, David Schlesinger, Debora Botequio Moretti, Elaine Cristina Marqueze, Elaine Vieira Santos, Evandra Strazza Rodrigues, Heidge Fukumasu, Jayme Augusto de Souza-Neto, Jos√© Salvator |
| hCoV-19/USA/CA-CDC-FG-212270/2021 | EPI_ISL_8293668 | 2021-12-22 | Fulgent Genetics | Centers for Disease Control and Prevention Division of Viral Diseases, Pathogen Discovery | Dakota Howard,Dhwani Batra,Peter Cook,Jason Caravas,Benjamin Rambo-Martin,Scott Sammons,Yvette Unoarumhi,Matthew Schmerer,Kristine Lacek,Tymeckia Kendall,Victoria Caban Figueroa,Shatavia Morrison,Christopher Gulvick,Erisa Sula,Harry Gao,Mickey Li,John Gao |
| hCoV-19/USA/TX-CDC-FG-215309/2021 | EPI_ISL_8294102 | 2021-12-23 | Fulgent Genetics | Centers for Disease Control and Prevention Division of Viral Diseases, Pathogen Discovery | Dakota Howard,Dhwani Batra,Peter Cook,Jason Caravas,Benjamin Rambo-Martin,Scott Sammons,Yvette Unoarumhi,Matthew Schmerer,Kristine Lacek,Tymeckia Kendall,Victoria Caban Figueroa,Shatavia Morrison,Christopher Gulvick,Erisa Sula,Harry Gao,Mickey Li,John Gao |
| hCoV-19/Singapore/9434/2021 | EPI_ISL_8295092 | 2021-12-24 | National Public Health Laboratory, National Centre for Infectious Diseases | National Centre for Infectious Diseases | Grace Ngan, Zhenyang Zhou, Royce Ang, Samuel Loo, Lin Cui, Raymond Tzer Pin Lin |
| hCoV-19/Israel/ICH-741108464/2021 | EPI_ISL_8296012 | 2021-12-22 | Clinical Microbiology Laboratory, Tel Aviv Sourasky Medical Center | Clinical Microbiology Laboratory, Tel Aviv Sourasky Medical Center | Katya Levytskyi, Ora Halutz, Alon Ziv, Lior Handler, Amos Adler |
| hCoV-19/SouthAfrica/NHLS-UCT-GS-D145/2021 | EPI_ISL_7310245 | 2021-06-30 | Groote Schuur Hospital wc GSH | NHLS/UCT | Arash Iranzadeh, Deelan Doolabh, Lynn Tyers, Bruna Galvao, Innocent Mudau, Marvin Hsiao, Gert Marais, Diana Hardie, Stephen Korsman, Rageema Joseph, Luicer Olubayo, Carolyn Williamson |
| hCoV-19/SouthAfrica/NHLS-UCT-GS-AE94/2021 | EPI_ISL_7310321 | 2021-11-04 | Groote Schuur Hospital wc GSH | NHLS/UCT | Arash Iranzadeh, Deelan Doolabh, Lynn Tyers, Bruna Galvao, Innocent Mudau, Marvin Hsiao, Gert Marais, Diana Hardie, Stephen Korsman, Rageema Joseph, Luicer Olubayo, Carolyn Williamson |
| hCoV-19/Tunisia/F-2976/2021 | EPI_ISL_8298511 | 2021-09-07 | Laboratoire de Virologie Clinique, Institut Pasteur de Tunis | Laboratory of Clinical Virology, Institut Pasteur de Tunis | Fares Wasfi, Nissaf Ben Alaya, Triki Henda |
| hCoV-19/USA/MI-CDC-STM-XHKVWG4GB/2021 | EPI_ISL_7310789 | 2021-11-18 | Helix | Centers for Disease Control and Prevention Division of Viral Diseases, Pathogen Discovery | Dakota Howard,Dhwani Batra,Peter Cook,Jason Caravas,Benjamin Rambo-Martin,Scott Sammons,Yvette Unoarumhi,Matthew Schmerer,Kristine Lacek,Tymeckia Kendall,Victoria Caban Figueroa,Helix CA,Shatavia Morrison,Christopher Gulvick,Erisa Sula,Clinton Paden,Dunca |

| | | | | | |
|---|---|---|---|---|---|
| hCoV-19/England/MILK-30219BA/2021 | EPI_ISL_8300793 | 2021-12-28 | Lighthouse Lab in Milton Keynes | Wellcome Sanger Institute for the COVID-19 Genomics UK (COG-UK) Consortium | The Lighthouse Lab in Milton Keynes and Alex Alderton, Roberto Amato, Jeffrey Barrett, Sonia Goncalves, Ewan Harrison, David K. Jackson, Ian Johnston, Dominic Kwiatkowski, Cordelia Langford, John Sillitoe on behalf of the Wellcome Sanger Institute COVID-1 |
| hCoV-19/England/MILK-3026F77/2021 | EPI_ISL_8301355 | 2021-12-28 | Lighthouse Lab in Milton Keynes | Wellcome Sanger Institute for the COVID-19 Genomics UK (COG-UK) Consortium | The Lighthouse Lab in Milton Keynes and Alex Alderton, Roberto Amato, Jeffrey Barrett, Sonia Goncalves, Ewan Harrison, David K. Jackson, Ian Johnston, Dominic Kwiatkowski, Cordelia Langford, John Sillitoe on behalf of the Wellcome Sanger Institute COVID-1 |
| hCoV-19/England/LSPA-3019F3B/2021 | EPI_ISL_8301486 | 2021-12-24 | Rosalind Franklin Laboratory | Wellcome Sanger Institute for the COVID-19 Genomics UK (COG-UK) Consortium | Donald Fraser, Suki Lee, Rob Howes, The Rosalind Franklin Laboratory and Alex Alderton, Roberto Amato, Jeffrey Barrett, Sonia Goncalves, Ewan Harrison, David K. Jackson, Ian Johnston, Dominic Kwiatkowski, Cordelia Langford, John Sillitoe on behalf of the |
| hCoV-19/USA/NY-PRL-2021_1219_00L11/2021 | EPI_ISL_8304511 | 2021-12-15 | Pandemic Response Lab - NYC | Pandemic Response Lab, R&D | Henry Lee, Michael Hammerling, Melissa Hopkins, Cybill del Castillo, Shinyoung Clair Kang, William Ward, Pradeep Bugga, Sol Rey, Dylan Law, Alex Carpio, Isabel Fernandez Escapa, Haiping Hao, Jon Laurent |
| hCoV-19/USA/NY-PRL-2021_1219_06F01/2021 | EPI_ISL_8304880 | 2021-12-15 | Pandemic Response Lab - NYC | Pandemic Response Lab, R&D | Henry Lee, Michael Hammerling, Melissa Hopkins, Cybill del Castillo, Shinyoung Clair Kang, William Ward, Pradeep Bugga, Sol Rey, Dylan Law, Alex Carpio, Isabel Fernandez Escapa, Haiping Hao, Jon Laurent |
| hCoV-19/USA/CA-TCPHL-122921-25/2021 | EPI_ISL_8306858 | 2021-12-26 | Tulare County Public Health Laboratory | Tulare County Public Health Laboratory | Denise L Lopez, Joshua Kootstra |
| hCoV-19/USA/NY-PRL-2021_1222_00C16/2021 | EPI_ISL_8306144 | 2021-12-18 | Pandemic Response Lab - NYC | Pandemic Response Lab, R&D | Henry Lee, Michael Hammerling, Melissa Hopkins, Cybill del Castillo, Shinyoung Clair Kang, William Ward, Pradeep Bugga, Sol Rey, Dylan Law, Alex Carpio, Isabel Fernandez Escapa, Haiping Hao, Jon Laurent |
| hCoV-19/USA/NY-PRL-2021_1223_00C07/2021 | EPI_ISL_8305857 | 2021-12-20 | Pandemic Response Lab - NYC | Pandemic Response Lab, R&D | Henry Lee, Michael Hammerling, Melissa Hopkins, Cybill del Castillo, Shinyoung Clair Kang, William Ward, Pradeep Bugga, Sol Rey, Dylan Law, Alex Carpio, Isabel Fernandez Escapa, Haiping Hao, Jon Laurent |
| hCoV-19/Uganda/MRC-UVRI-CX23/2021 | EPI_ISL_8307323 | 2021-11-29 | MRC/UVRI & LSHTM Uganda Research Unit, Uganda Virus Research Institute | MRC/UVRI & LSHTM Uganda Research Unit, Uganda Virus Research Institute | Deogratius Ssemwanga, Nicholas Bbosa, Hamidah Namagembe, Ronald Kiiza, Jocelyn Kiconco, John Kayiwa, Tom Lutalo, Alfred Ssekagiri, Julius Lutwama, Isaac Ssewanyana, Susan Nabadda, Henry Kyobe-Bbosa, Pontiano Kaleebu |
| hCoV-19/Uganda/MRC-UVRI-EG70/2021 | EPI_ISL_8307367 | 2021-12-14 | MRC/UVRI & LSHTM Uganda Research Unit, Uganda Virus Research Institute | MRC/UVRI & LSHTM Uganda Research Unit, Uganda Virus Research Institute | Deogratius Ssemwanga, Nicholas Bbosa, Hamidah Namagembe, Ronald Kiiza, Jocelyn Kiconco, John Kayiwa, Tom Lutalo, Alfred Ssekagiri, Julius Lutwama, Isaac Ssewanyana, Susan Nabadda, Henry Kyobe-Bbosa, Pontiano Kaleebu |

| | | | | | |
|---|---|---|---|---|---|
| hCoV-19/Uganda/MRC-UVRI-GI48/2021 | EPI_ISL_8307376 | 2021-12-08 | MRC/UVRI & LSHTM Uganda Research Unit, Uganda Virus Research Institute | MRC/UVRI & LSHTM Uganda Research Unit, Uganda Virus Research Institute | Deogratius Ssemwanga, Nicholas Bbosa, Hamidah Namagembe, Ronald Kiiza, Jocelyn Kiconco, John Kayiwa, Tom Lutalo, Alfred Ssekagiri, Julius Lutwama, Isaac Ssewanyana, Susan Nabadda, Henry Kyobe-Bbosa, Pontiano Kaleebu |
| hCoV-19/Uganda/MRC-UVRI-NHLDSALG7778/2021 | EPI_ISL_8307408 | 2021-12-14 | MRC/UVRI & LSHTM Uganda Research Unit, Uganda Virus Research Institute | MRC/UVRI & LSHTM Uganda Research Unit, Uganda Virus Research Institute | Deogratius Ssemwanga, Nicholas Bbosa, Hamidah Namagembe, Ronald Kiiza, Jocelyn Kiconco, John Kayiwa, Tom Lutalo, Alfred Ssekagiri, Julius Lutwama, Isaac Ssewanyana, Susan Nabadda, Henry Kyobe-Bbosa, Pontiano Kaleebu |
| hCoV-19/USA/TX-CDC-ASC210635636/2021 | EPI_ISL_8308032 | 2021-12-18 | Aegis Sciences Corporation | Centers for Disease Control and Prevention Division of Viral Diseases, Pathogen Discovery | Dakota Howard,Dhwani Batra,Peter Cook,Jason Caravas,Benjamin Rambo-Martin,Scott Sammons,Yvette Unoarumhi,Matthew Schmerer,Kristine Lacek,Tymeckia Kendall,Victoria Caban Figueroa,Shatavia Morrison,Christopher Gulvick,Erisa Sula,Cyndi Clark,Patrick Campbell |
| hCoV-19/Turkey/HSGM-F12356/2021 | EPI_ISL_8308637 | 2021-12-28 | Ministry of Health Turkey | Ministry of Health Turkey | Fatma Bayrakdar, Yasemin Co≈ügun, S√ºleyman Yalcin, G√ºlay Korukluoƒülu |
| hCoV-19/Sweden/2002789064/2021 | EPI_ISL_8312088 | 2021-12-12 | Sweden, Vastragotaland, Unilabs | Unilabs, Eskilstuna, Sweden | Emma Arvidsson |
| hCoV-19/Lebanon/LAU-R248/2020 | EPI_ISL_8311724 | 2020-12-10 | LAUMC-Rizk Hospital | Multi-Omics Laboratory - School of Pharmacy - Lebanese American University | Nancy Fayad, Walid Abi Habib, Rabeh El-Shesheny, Ahmed Kandeil, Youmna Mourad, Jacques Mokhbat, Ghazi Kayali, Jimi Goldstein and Jad Abdallah |
| hCoV-19/Colombia/NSA-INS-VG-9803/2021 | EPI_ISL_7314433 | 2021-10-15 | IDIME | Instituto Nacional de Salud-Direcci√≥n de Investigaci√≥n en Salud P√∫blica | Katherine Laiton-Donato, Carlos Franco-Mu√±oz, Diego A. √Ålvarez-D√≠az, H√©ctor Alejandro Ruiz-Moreno, Diego Andr√©s Prada, Jhonnatan Reales-Gonz√°lez, Sheryll Corchuelo, Mar√≠a T. Herrera-Sep√∫lveda, Juli√°n Naizaque, Gerardo Santamar√≠a, Jorge Rivera, P |
| hCoV-19/USA/IL-CDC-STM-3X5BR8RY5/2021 | EPI_ISL_8316365 | 2021-12-14 | Helix | Centers for Disease Control and Prevention Division of Viral Diseases, Pathogen Discovery | Dakota Howard,Dhwani Batra,Peter Cook,Jason Caravas,Benjamin Rambo-Martin,Scott Sammons,Yvette Unoarumhi,Matthew Schmerer,Kristine Lacek,Tymeckia Kendall,Victoria Caban Figueroa,Helix CA,Shatavia Morrison,Christopher Gulvick,Erisa Sula,Clinton Paden,Dunca |
| hCoV-19/Sweden/Orebro-47829235/2021 | EPI_ISL_8316361 | 2021-12-22 | Orebro University Hospital, Dept Laboratory Medicine, Clinical Microbiology | Orebro University Hospital | Sundqvist M et al |
| hCoV-19/France/PAC-IHU-49241/2021 | EPI_ISL_7314302 | 2021-12-02 | IHU Mediterranee Infection | IHU Mediterranee Infection | Philippe Colson et al. |
| hCoV-19/Kenya/ILRI_COVC23193/2021 | EPI_ISL_8318558 | 2021-02-11 | International Livestock Research Institute | International Livestock Research Institute | Samuel O. Oyola, Gilbert Kibet, Daniel Ouso, Shebbar Osiany, Edward Kiritu, Paul Dobi, Collins Muli, Patrick Amoth, Vishvanath Nene, Sonal P. Henson, Edward O. Abworo |
| hCoV-19/Kenya/ILRI_KEMRI_01_90725/2021 | EPI_ISL_8318615 | 2021-01-26 | International Livestock Research Institute | International Livestock Research Institute | Samuel O. Oyola, Gilbert Kibet, Daniel Ouso, Shebbar Osiany, Edward Kiritu, Paul Dobi, Collins Muli, Patrick Amoth, Vishvanath Nene, Sonal P. Henson, Edward O. Abworo |

| | | | | | |
|---|---|---|---|---|---|
| hCoV-19/Colombia/ATL-INS-VG-11520/2021 | EPI_ISL_8318705 | 2021-12-21 | LABORATORIO CLINICO KHENEYZIR FAYAD | Instituto Nacional de Salud-Direcci√≥n de Investigaci√≥n en Salud P√∫blica | Katherine Laiton-Donato, Carlos Franco-Mu√±oz, Diego A. √Ålvarez-D√≠az, H√©ctor Alejandro Ruiz-Moreno, Diego Andr√©s Prada, Jhonnatan Reales-Gonz√°lez, Sheryll Corchuelo, Mar√≠a T. Herrera-Sep√∫lveda, Juli√°n Naizaque, Gerardo Santamar√≠a, Jorge Rivera, P |
| hCoV-19/India/MH-Pune-INSACOG-RF-NCL-2631/2021 | EPI_ISL_8319053 | 2021-12-13 | BJMC | NCL, Pune | Anu Raghunathan, Dhanasekaran Shanmugam, Ajinkya Khilari, Divya Niveditha, Bhagyashree Litikar, Jugal Kanekar, Shikha Takur, Kadir Phatan |
| hCoV-19/Canada/ON-PHL-21-46943/2021 | EPI_ISL_8323301 | 2021-12-08 | Public Health Ontario Laboratory | Public Health Ontario Laboratory | Vanessa G Allen, Philip Banh, Yao Chen, Richard de Borja, Alireza Eshaghi, Nahuel Fittipaldi, Christine Frantz, Jonathan B Gubbay, Jennifer L Guthrie, Lawrence Heisler, Esha Joshi, Michael Laszloffy, Aimin Li, Michael CY Li, Dean Maxwell, Sandeep Nagra, S |
| hCoV-19/Canada/ON-PHL-21-48537/2021 | EPI_ISL_8324636 | 2021-12-10 | Public Health Ontario Laboratory | Public Health Ontario Laboratory | Vanessa G Allen, Philip Banh, Yao Chen, Richard de Borja, Alireza Eshaghi, Nahuel Fittipaldi, Christine Frantz, Jonathan B Gubbay, Jennifer L Guthrie, Lawrence Heisler, Esha Joshi, Michael Laszloffy, Aimin Li, Michael CY Li, Dean Maxwell, Sandeep Nagra, S |
| hCoV-19/USA/NC-CLT-CORVASEQ-004130/2021 | EPI_ISL_8322566 | 2021-12-21 | StarMed Healthcare | UNC Charlotte Environmental Monitoring Laboratory | Jessica Schlueter, Jannatul Ferdous, Torri Weathers, Samuel Kunkleman, Juan Bolanos, Kevin Lambirth, Cynthia Gibas |
| hCoV-19/Brazil/GO-FIOCRUZ-65292/2021 | EPI_ISL_8325065 | 2021-12-07 | Laboratorio Central de Saude Publica do Estado de Goias (LACEN/GO) | Laboratory of Respiratory Viruses and Measles, Oswaldo Cruz Institute, FIOCRUZ | Paola Resende, Luciana Appolinario, Fernando Motta, Elisa Cavalcante Pereira, Ighor Arantes, Alice Sampaio Rocha, Bruna Mendon√ßa da Silva, J√©ssica Gra√ßa Macedo de Carvalho, Larissa Macedo Pinto, Victor Guimaraes, Rafael Souza Guedes, Carmen Helena Ramo |
| hCoV-19/Brazil/SC-FIOCRUZ-66119/2021 | EPI_ISL_8325370 | 2021-12-17 | Laboratorio Central de Saude Publica do Estado de Santa Catarina (LACEN/SC) | Laboratory of Respiratory Viruses and Measles, Oswaldo Cruz Institute, FIOCRUZ | Paola Resende, Luciana Appolinario, Fernando Motta, Elisa Cavalcante Pereira, Ighor Arantes, Alice Sampaio Rocha, Bruna Mendon√ßa da Silva, J√©ssica Gra√ßa Macedo de Carvalho, Larissa Macedo Pinto, Victor Guimaraes, Darcita Buerger Rovaris, Sandra Bianchi |
| hCoV-19/Brazil/SC-FIOCRUZ-66112/2021 | EPI_ISL_8325364 | 2021-12-17 | Laboratorio Central de Saude Publica do Estado de Santa Catarina (LACEN/SC) | Laboratory of Respiratory Viruses and Measles, Oswaldo Cruz Institute, FIOCRUZ | Paola Resende, Luciana Appolinario, Fernando Motta, Elisa Cavalcante Pereira, Ighor Arantes, Alice Sampaio Rocha, Bruna Mendon√ßa da Silva, J√©ssica Gra√ßa Macedo de Carvalho, Larissa Macedo Pinto, Victor Guimaraes, Darcita Buerger Rovaris, Sandra Bianchi |
| hCoV-19/Australia/NSW-ICPMR-18895/2021 | EPI_ISL_8334346 | 2021-12-27 | Pathology West - NSW Health Pathology | Clinical Pathology and Medical Research; Westmead Hospital; University of Sydney | Arnott A., Draper J., Gall M., Martinez E., Rockett R., Sintchenko V., on behalf of ICPMR |
| hCoV-19/Israel/ICH-741109968/2021 | EPI_ISL_8327689 | 2021-12-28 | Clinical Microbiology Laboratory, Tel Aviv Sourasky Medical Center | Clinical Microbiology Laboratory, Tel Aviv Sourasky Medical Center | Katya Levytskyi, Ora Halutz, Alon Ziv, Lior Handler, Goel Morad, Amos Adler |
| hCoV-19/Israel/ICH-741109918/2021 | EPI_ISL_8327647 | 2021-12-28 | Clinical Microbiology Laboratory, Tel Aviv Sourasky Medical Center | Clinical Microbiology Laboratory, Tel Aviv Sourasky Medical Center | Katya Levytskyi, Ora Halutz, Alon Ziv, Lior Handler, Goel Morad, Amos Adler |
| hCoV-19/India/MH-RFIP06538/2021 | EPI_ISL_8328922 | 2021-12-20 | BJMC Pune | RF-IISER Pune | Krishanpal Karmodiya, Aurnab Ghose, Joy Merwin Monteiro, Suvarna Joshi, Rajesh Karyakarte |

| | | | | | |
|---|---|---|---|---|---|
| hCoV-19/India/MH-RFIP06483/2021 | EPI_ISL_8328887 | 2021-12-20 | BJMC Pune | RF-IISER Pune | Krishanpal Karmodiya, Aurnab Ghose, Joy Merwin Monteiro, Suvarna Joshi, Rajesh Karyakarte |
| hCoV-19/India/MH-RFIP06640/2021 | EPI_ISL_8328974 | 2021-12-19 | BJMC Pune | RF-IISER Pune | Krishanpal Karmodiya, Aurnab Ghose, Joy Merwin Monteiro, Suvarna Joshi, Rajesh Karyakarte |
| hCoV-19/Australia/ACT-2096/2021 | EPI_ISL_8331484 | 2021-12-15 | Capital Pathology | Schwessinger Lab | Austin Bird, Bayantes Dagvadorj, Scott Ferguson, Robyn Hall, Evie Hodgson, Ashley Jones, Elise Kellett, Karina Kennedy, Rachel Leonard, Sandra Molloy, Carl McCombe, Catalina Barrag√°n Quintero, Rene Riedelbauch, Benjamin Schwessinger, Gabrielle Smith, Pau |
| hCoV-19/Australia/ACT-2420/2021 | EPI_ISL_8331766 | 2021-12-26 | ACT Pathology | Schwessinger Lab | Austin Bird, Bayantes Dagvadorj, Scott Ferguson, Robyn Hall, Evie Hodgson, Ashley Jones, Elise Kellett, Craig Kennedy, Karina Kennedy, Rachel Leonard, Carl McCombe, Catalina Barrag√°n Quintero, Rene Riedelbauch, Benjamin Schwessinger, Gabrielle Smith, Sal |
| hCoV-19/Australia/ACT-2481/2021 | EPI_ISL_8331826 | 2021-12-28 | ACT Pathology | Schwessinger Lab | Austin Bird, Bayantes Dagvadorj, Scott Ferguson, Robyn Hall, Evie Hodgson, Ashley Jones, Elise Kellett, Craig Kennedy, Karina Kennedy, Rachel Leonard, Carl McCombe, Catalina Barrag√°n Quintero, Rene Riedelbauch, Benjamin Schwessinger, Gabrielle Smith, Sal |
| hCoV-19/Australia/ACT-2097/2021 | EPI_ISL_8331485 | 2021-12-15 | Capital Pathology | Schwessinger Lab | Austin Bird, Bayantes Dagvadorj, Scott Ferguson, Robyn Hall, Evie Hodgson, Ashley Jones, Elise Kellett, Karina Kennedy, Rachel Leonard, Sandra Molloy, Carl McCombe, Catalina Barrag√°n Quintero, Rene Riedelbauch, Benjamin Schwessinger, Gabrielle Smith, Pau |
| hCoV-19/Brazil/RJ-FIOCRUZ-65318/2021 | EPI_ISL_8332308 | 2021-12-23 | Laboratorio Central de Saude Publica do Estado do Rio de Janeiro (LACEN/RJ) | Laboratory of Respiratory Viruses and Measles, Oswaldo Cruz Institute, FIOCRUZ | Paola Resende, Luciana Appolinario, Fernando Motta, Elisa Cavalcante Pereira, Ighor Arantes, Alice Sampaio Rocha, Bruna Mendon√ßa da Silva, J√©ssica Gra√ßa Macedo de Carvalho, Larissa Macedo Pinto, Victor Guimaraes, Andrea Cony Cavalcanti, Marilda Siqueir |
| hCoV-19/Australia/ACT-2526/2021 | EPI_ISL_8331865 | 2021-12-30 | ACT Pathology | Schwessinger Lab | Austin Bird, Bayantes Dagvadorj, Scott Ferguson, Robyn Hall, Evie Hodgson, Ashley Jones, Elise Kellett, Craig Kennedy, Karina Kennedy, Rachel Leonard, Carl McCombe, Catalina Barrag√°n Quintero, Rene Riedelbauch, Benjamin Schwessinger, Gabrielle Smith, Sal |
| hCoV-19/Australia/ACT-2531/2021 | EPI_ISL_8331870 | 2021-12-30 | ACT Pathology | Schwessinger Lab | Austin Bird, Bayantes Dagvadorj, Scott Ferguson, Robyn Hall, Evie Hodgson, Ashley Jones, Elise Kellett, Craig Kennedy, Karina Kennedy, Rachel Leonard, Carl McCombe, Catalina Barrag√°n Quintero, Rene Riedelbauch, Benjamin Schwessinger, Gabrielle Smith, Sal |

| | | | | | |
|---|---|---|---|---|---|
| hCoV-19/Australia/ACT-2520/2021 | EPI_ISL_8331860 | 2021-12-30 | ACT Pathology | Schwessinger Lab | Austin Bird, Bayantes Dagvadorj, Scott Ferguson, Robyn Hall, Evie Hodgson, Ashley Jones, Elise Kellett, Craig Kennedy, Karina Kennedy, Rachel Leonard, Carl McCombe, Catalina Barrag√°n Quintero, Rene Riedelbauch, Benjamin Schwessinger, Gabrielle Smith, Sal |
| hCoV-19/Australia/ACT-2274/2021 | EPI_ISL_8331646 | 2021-12-23 | Capital Pathology | Schwessinger Lab | Austin Bird, Bayantes Dagvadorj, Scott Ferguson, Robyn Hall, Evie Hodgson, Ashley Jones, Elise Kellett, Karina Kennedy, Rachel Leonard, Sandra Molloy, Carl McCombe, Catalina Barrag√°n Quintero, Rene Riedelbauch, Benjamin Schwessinger, Gabrielle Smith, Pau |
| hCoV-19/Australia/NSW-ICPMR-18309/2021 | EPI_ISL_8333760 | 2021-12-16 | St Vincent's Pathology (SydPath) | Clinical Pathology and Medical Research; Westmead Hospital; University of Sydney | Arnott A., Draper J., Gall M., Martinez E., Rockett R., Sintchenko V., on behalf of ICPMR |
| hCoV-19/Australia/NSW-ICPMR-18392/2021 | EPI_ISL_8333843 | 2021-12-18 | 4Cyte Pathology | Clinical Pathology and Medical Research; Westmead Hospital; University of Sydney | Arnott A., Draper J., Gall M., Martinez E., Rockett R., Sintchenko V., on behalf of ICPMR |
| hCoV-19/Australia/NSW-ICPMR-18446/2021 | EPI_ISL_8333897 | 2021-12-18 | 4Cyte Pathology | Clinical Pathology and Medical Research; Westmead Hospital; University of Sydney | Arnott A., Draper J., Gall M., Martinez E., Rockett R., Sintchenko V., on behalf of ICPMR |
| hCoV-19/Australia/NSW-ICPMR-18545/2021 | EPI_ISL_8333996 | 2021-12-21 | Pathology West - NSW Health Pathology | Clinical Pathology and Medical Research; Westmead Hospital; University of Sydney | Arnott A., Draper J., Gall M., Martinez E., Rockett R., Sintchenko V., on behalf of ICPMR |
| hCoV-19/Australia/NSW-ICPMR-18699/2021 | EPI_ISL_8334150 | 2021-12-26 | Pathology West - NSW Health Pathology | Clinical Pathology and Medical Research; Westmead Hospital; University of Sydney | Arnott A., Draper J., Gall M., Martinez E., Rockett R., Sintchenko V., on behalf of ICPMR |
| hCoV-19/Australia/NSW-ICPMR-18736/2021 | EPI_ISL_8334187 | 2021-12-21 | Pathology West - NSW Health Pathology | Clinical Pathology and Medical Research; Westmead Hospital; University of Sydney | Arnott A., Draper J., Gall M., Martinez E., Rockett R., Sintchenko V., on behalf of ICPMR |
| hCoV-19/India/GJ-INSACOG-GBRC3662/2021 | EPI_ISL_8334413 | 2021-11-23 | Sterling Accuris, Surat | Gujarat Biotechnology Research Centre | Jaykumar Rangani,Ramesh Pandit,Nitin Savaliya,Priyank Chavda,Sonal Sharma ,Roshani Mishra,Disha Vora,Janvi Raval,Apurvasinh Puvar,Zarna Patel,Bhadreshsinh Gohil,Nimesh Patel,Nitin Shukla,Akhilesh Modi,Tasnim Trivedi,Bharat Tandel,Chaitanya Joshi,Madhvi Jo |
| hCoV-19/Australia/NSW-ICPMR-18569/2021 | EPI_ISL_8334020 | 2021-12-21 | Pathology West - NSW Health Pathology | Clinical Pathology and Medical Research; Westmead Hospital; University of Sydney | Arnott A., Draper J., Gall M., Martinez E., Rockett R., Sintchenko V., on behalf of ICPMR |
| hCoV-19/Australia/QLD3688/2021 | EPI_ISL_8158398 | 2021-12-22 | PHV-FSS | PHV-FSS | Chenwei Wang on behalf of Q-PHIRE Genomics |
| hCoV-19/England/ALDP-302FF50/2021 | EPI_ISL_8339903 | 2021-12-24 | Lighthouse Lab in Alderley Park | Wellcome Sanger Institute for the COVID-19 Genomics UK (COG-UK) Consortium | Jacquelyn Wynn, Mairead Hyland, The Lighthouse Lab in Alderley Park and Alex Alderton, Roberto Amato, Jeffrey Barrett, Sonia Goncalves, Ewan Harrison, David K. Jackson, Ian Johnston, Dominic Kwiatkowski, Cordelia Langford, John Sillitoe on behalf of the W |

| | | | | | |
|---|---|---|---|---|---|
| hCoV-19/Denmark/DCGC-290621/2021 | EPI_ISL_8344207 | 2021-12-23 | Department of Bacteria, Parasites and Fungi, Statens Serum Institut, Copenhagen, Denmark | Statens Serum Institut Bioinformatics and Microbial Genomics | Danish Covid-19 Genome Consortium |
| hCoV-19/Denmark/DCGC-291550/2021 | EPI_ISL_8345185 | 2021-12-13 | Department of Bacteria, Parasites and Fungi, Statens Serum Institut, Copenhagen, Denmark | Statens Serum Institut Bioinformatics and Microbial Genomics | Danish Covid-19 Genome Consortium |
| hCoV-19/Denmark/DCGC-292815/2021 | EPI_ISL_8346381 | 2021-12-23 | Department of Bacteria, Parasites and Fungi, Statens Serum Institut, Copenhagen, Denmark | Statens Serum Institut Bioinformatics and Microbial Genomics | Danish Covid-19 Genome Consortium |
| hCoV-19/Denmark/DCGC-292325/2021 | EPI_ISL_8345954 | 2021-12-23 | Department of Clinical Microbiology, Odense University Hospital, Odense, Denmark | Bioinformatics and Microbial Genomics | Danish Covid-19 Genome Consortium |
| hCoV-19/USA/AZ-TG1114139/2021 | EPI_ISL_8319416 | 2021-11-27 | TGen North | TGen North | Jolene Bowers, Heather Centner, Brett Van Tassel, Chris French, Hayley Yaglom, Darrin Lemmer, Dave Engelthaler |
| hCoV-19/Denmark/DCGC-293379/2021 | EPI_ISL_8346937 | 2021-12-15 | Department of Bacteria, Parasites and Fungi, Statens Serum Institut, Copenhagen, Denmark | Statens Serum Institut Bioinformatics and Microbial Genomics | Danish Covid-19 Genome Consortium |
| hCoV-19/Denmark/DCGC-293661/2021 | EPI_ISL_8347187 | 2021-12-26 | Department of Bacteria, Parasites and Fungi, Statens Serum Institut, Copenhagen, Denmark | Statens Serum Institut Bioinformatics and Microbial Genomics | Danish Covid-19 Genome Consortium |
| hCoV-19/USA/CA-CDC-FG-180156/2021 | EPI_ISL_7497741 | 2021-11-30 | Fulgent Genetics | Centers for Disease Control and Prevention Division of Viral Diseases, Pathogen Discovery | Dakota Howard,Dhwani Batra,Peter Cook,Jason Caravas,Benjamin Rambo-Martin,Scott Sammons,Yvette Unoarumhi,Matthew Schmerer,Kristine Lacek,Tymeckia Kendall,Victoria Caban Figueroa,Shatavia Morrison,Christopher Gulvick,Erisa Sula,Harry Gao,Mickey Li,John Gao |
| hCoV-19/Denmark/DCGC-293747/2021 | EPI_ISL_8347265 | 2021-12-12 | Department of Bacteria, Parasites and Fungi, Statens Serum Institut, Copenhagen, Denmark | Statens Serum Institut Bioinformatics and Microbial Genomics | Danish Covid-19 Genome Consortium |
| hCoV-19/Denmark/DCGC-294055/2021 | EPI_ISL_8347538 | 2021-12-15 | Department of Bacteria, Parasites and Fungi, Statens Serum Institut, Copenhagen, Denmark | Statens Serum Institut Bioinformatics and Microbial Genomics | Danish Covid-19 Genome Consortium |
| hCoV-19/Ireland/LS-NVRL-S21IRL00563864/2021 | EPI_ISL_8350034 | 2021-12-13 | National Virus Reference Laboratory | National Virus Reference Laboratory | Zoe Yandle, Charlene Bennett, Gabriel Gonzalez, Michael Carr, Jonathan Dean, Cillian F De Gascun |
| hCoV-19/CaboVerde/Santiago-35855/2021 | EPI_ISL_8351173 | 2021-08-28 | Virology Laboratory of Praia | Institut Pasteur de Dakar | Sanders Kevin, Diagne Moussa Mo√Øse, Diallo Amadou, Dia Ndongo, Sankhe Safietou, Diop Mamadou, Ndiaye Ndack, Loucoubar Cheikh, Faye Ousmane, da Luz Lima Mendon√ßa Maria, Sall Amadou Alpha |
| hCoV-19/Belgium/Jessa_55-2151-001419/2021 | EPI_ISL_8351242 | 2021-12-26 | Jessa | Jessa | Severine Berden et al. on behalf of the Jessa_cmdLab |
| hCoV-19/USA/FL-BPHL-18426/2021 | EPI_ISL_8352666 | 2021-12-12 | Florida Bureau of Public Health Laboratories | Florida Bureau of Public Health Laboratories | Sarah Schmedes, Namratha Tarigopula, Tiffany Splatt, Jason Blanton |

| | | | | | |
|---|---|---|---|---|---|
| hCoV-19/USA/GA-CDC-ASC210631967/2021 | EPI_ISL_8353217 | 2021-12-20 | Aegis Sciences Corporation | Centers for Disease Control and Prevention Division of Viral Diseases, Pathogen Discovery | Dakota Howard,Dhwani Batra,Peter Cook,Jason Caravas,Benjamin Rambo-Martin,Scott Sammons,Yvette Unoarumhi,Matthew Schmerer,Kristine Lacek,Tymeckia Kendall,Victoria Caban Figueroa,Shatavia Morrison,Christopher Gulvick,Erisa Sula,Cyndi Clark,Patrick Campbell |
| hCoV-19/Brazil/SP-IB_145381/2021 | EPI_ISL_8353537 | 2021-12-20 | CENTRO DE SAUDE SAO QUIRINO | Instituto Butantan | Dimas Tadeu Covas, Antonio Jorge Martins, Claudia Renata dos Santos Barros, David Schlesinger, Debora Botequio Moretti, Elaine Cristina Marqueze, Elaine Vieira Santos, Evandra Strazza Rodrigues, Heidge Fukumasu, Jayme Augusto de Souza-Neto, Jos√© Salvator |
| hCoV-19/Brazil/SP-IB_145390/2021 | EPI_ISL_8353608 | 2021-12-19 | UNIDADE DE INFECTOLOGIA WILLIAN ROCHA GUARUJA | Instituto Butantan | Dimas Tadeu Covas, Antonio Jorge Martins, Claudia Renata dos Santos Barros, David Schlesinger, Debora Botequio Moretti, Elaine Cristina Marqueze, Elaine Vieira Santos, Evandra Strazza Rodrigues, Heidge Fukumasu, Jayme Augusto de Souza-Neto, Jos√© Salvator |
| hCoV-19/USA/NM-CDC-ASC210523401/2021 | EPI_ISL_7229579 | 2021-11-04 | Aegis Sciences Corporation | Centers for Disease Control and Prevention Division of Viral Diseases, Pathogen Discovery | Dakota Howard,Dhwani Batra,Peter Cook,Jason Caravas,Benjamin Rambo-Martin,Scott Sammons,Yvette Unoarumhi,Matthew Schmerer,Kristine Lacek,Tymeckia Kendall,Victoria Caban Figueroa,Shatavia Morrison,Christopher Gulvick,Erisa Sula,Cyndi Clark,Patrick Campbell |
| hCoV-19/Argentina/PAIS-E0681/2021 | EPI_ISL_8355442 | 2021-12-22 | Hospital Jaime Ferre - SAMCO Rafaela | [illegible] Bioinform√°tica del Instituto de Investigaci√≥n de la Cadena L√°ctea CONICET-INTA on behalf of 'Proyecto Argentino Interinstitucional de genomica de SARS-CoV-2' (PAIS | Eberhardt, MF; Irazoqui, JM; Aliprandi, D; Isaia C; Pandolfi V; Quaranta, JF; Soratti R; Amadio, AF |
| hCoV-19/Belgium/UMONS-P047988501/2021 | EPI_ISL_8357042 | 2021-11-30 | National Platform bis UMONS/Jolimont | National Platform bis UMONS/Jolimont | Guillaume Bayon-Vicente, Florian Juszczak, Laetitia Gheysen, Eric Tarantino, Gautier Detry, Ruddy Wattiez |
| hCoV-19/Colombia/DC-LSPSDS-VG-1018562/2021 | EPI_ISL_8357552 | 2021-08-27 | Colsanitas Keralty | LSPSDS | Johana Hernandez Toloza,Marcela Castano, Gabriela Delgado Murcia, Alejandro Gomez Lopez, Marcela Mercado |
| hCoV-19/USA/TX-CDC-ASC210613501/2021 | EPI_ISL_8358018 | 2021-12-16 | Aegis Sciences Corporation | Centers for Disease Control and Prevention Division of Viral Diseases, Pathogen Discovery | Dakota Howard,Dhwani Batra,Peter Cook,Jason Caravas,Benjamin Rambo-Martin,Scott Sammons,Yvette Unoarumhi,Matthew Schmerer,Kristine Lacek,Tymeckia Kendall,Victoria Caban Figueroa,Shatavia Morrison,Christopher Gulvick,Erisa Sula,Cyndi Clark,Patrick Campbell |
| hCoV-19/Brazil/RR-FIOCRUZ-ILMD2200016/2021 | EPI_ISL_8360701 | 2021-10-18 | LACEN/RR | Laborat√≥rio de Ecologia de Doen√ßas Transmiss√≠veis na Amaz√¥nia, Instituto Le√¥nidas e Maria Deane - Fiocruz Amaz√¥nia | C√°tia Alexandra Ribeiro Meneses, Rodrigo Melo Maito, Ilma de Aguiar Antony, Pricila Carlos Veloso, Janis Cassimiro Lopes, Arlesson Viana Da Silva, Dejanane Silva, Valdinete Nascimento, Victor Souza, Andr√© Corado, Fernanda Nascimento, George Silva, Matil |

| | | | | | |
|---|---|---|---|---|---|
| hCoV-19/USA/CA-CDC-FG-217644/2021 | EPI_ISL_8360894 | 2021-12-23 | Fulgent Genetics | Centers for Disease Control and Prevention Division of Viral Diseases, Pathogen Discovery | Dakota Howard,Dhwani Batra,Peter Cook,Jason Caravas,Benjamin Rambo-Martin,Scott Sammons,Yvette Unoarumhi,Matthew Schmerer,Kristine Lacek,Tymeckia Kendall,Victoria Caban Figueroa,Shatavia Morrison,Christopher Gulvick,Erisa Sula,Harry Gao,Mickey Li,John Gao |
| hCoV-19/Brazil/PR-FIOCRUZ-43883/2021 | EPI_ISL_8361512 | 2021-07-29 | Laboratorio Central de Saude Publica do Estado do Parana (LACEN/PR) | Laboratory of Respiratory Viruses and Measles, Oswaldo Cruz Institute, FIOCRUZ | Paola Resende, Luciana Appolinario, Fernando Motta, Anna Carolina Paixao, Ighor Leonardo Arantes, Agatha Soares, Ana Carolina Mendonca, Alice Sampaio Rocha, Taina Venas, Elisa Cavalcante Pereira, Renata Serrano Lopes, Irina Riediger, Marilda Siqueira on b |
| hCoV-19/USA/FL-CDC-LC0438388/2021 | EPI_ISL_8364696 | 2021-12-20 | Laboratory Corporation of America | Centers for Disease Control and Prevention Division of Viral Diseases, Pathogen Discovery | Dakota Howard,Dhwani Batra,Peter Cook,Jason Caravas,Benjamin Rambo-Martin,Scott Sammons,Yvette Unoarumhi,Matthew Schmerer,Kristine Lacek,Tymeckia Kendall,Victoria Caban Figueroa,Shatavia Morrison,Christopher Gulvick,Minoo Agarwal,Eyad Almasri,Debbie Boles |
| hCoV-19/USA/CT-CDC-LC0439386/2021 | EPI_ISL_8366831 | 2021-12-23 | Laboratory Corporation of America | Centers for Disease Control and Prevention Division of Viral Diseases, Pathogen Discovery | Dakota Howard,Dhwani Batra,Peter Cook,Jason Caravas,Benjamin Rambo-Martin,Scott Sammons,Yvette Unoarumhi,Matthew Schmerer,Kristine Lacek,Tymeckia Kendall,Victoria Caban Figueroa,Shatavia Morrison,Christopher Gulvick,Minoo Agarwal,Eyad Almasri,Debbie Boles |
| hCoV-19/USA/NY-NYCPHL-008485/2021 | EPI_ISL_8372557 | 2021-12-23 | DOHMH PHL | New York City Public Health Laboratory | Jade Wang, et al. |
| hCoV-19/Thailand/NIC_SNI_SEQ23246/2021 | EPI_ISL_8373277 | 2021-12-16 | Ko Samui Hospital | National Institute of Health, Department of Medical Sciences, Ministry of Public Health, Thailand | Pilailuk Okada; Siripaporn Phuygun; Nuttida Thongpramul; Pakorn Piromtong; Natchaya¬† Khiadsang; Thanutsapa Thanadachakul; Sirikanda Wimol; Suratchana Mitrat; Sunthareeya Waicharoen; Archawin Rojanawiwat |
| hCoV-19/USA/AZ-ASU30197/2021 | EPI_ISL_8374566 | 2021-12-28 | Arizona State University | Arizona State University | Steven C. Holland, Matthew F. Smith, Regan A. Sullins, James C. Hu, LaRinda A. Holland, Nathaniel Johnson, Joshua LaBaer, Vel Murugan, Efrem S. Lim |
| hCoV-19/Netherlands/ZH-EMC-4051/2021 | EPI_ISL_8377350 | 2021-01-21 | Dutch COVID-19 response team | Erasmus Medical Center | Bas Oude Munnink, Reina Sikkema, David Nieuwenhuijse, Irina Chestakova, Anne van der Linden, Marjan Boter, Emmanuelle Munger, Corine GeurtsvanKessel, Annemiek van der Eijk, Richard Molenkamp, Marion Koopmans, on behalf of the Dutch national COVID-19 respo |
| hCoV-19/Germany/BY-LGL-IMS-10272-CVDP-807f2e35-56d6-4000-a148-ef8aff4d6bfe/2021 | EPI_ISL_8379756 | 2021-11-29 | Bayerisches Landesamt fuer Gesundheit und Lebensmittelsicherheit (LGL) | Bayerisches Landesamt fuer Gesundheit und Lebensmittelsicherheit (LGL) | Alexandra Dangel, Carola Berger, Sabrina Hepner, Laura Weise, Annika Sprenger, Nikolaus Ackermann, Andreas Sing |
| hCoV-19/England/PHEP-YYN5E6N/2021 | EPI_ISL_8381169 | 2021-12-14 | Respiratory Virus Unit, Microbiology Services Colindale, Public Health England | COVID-19 Genomics UK (COG-UK) Consortium | PHE Covid Sequencing Team |
| hCoV-19/Denmark/DCGC-294624/2021 | EPI_ISL_8382244 | 2021-12-15 | Department of Bacteria, Parasites and Fungi, Statens Serum Institut, Copenhagen, Denmark | Statens Serum Institut Bioinformatics and Microbial Genomics | Danish Covid-19 Genome Consortium |

| | | | | | |
|---|---|---|---|---|---|
| hCoV-19/Thailand/DMSc-05885/2021 | EPI_ISL_8437307 | 2021-12-22 | Regional Medical Sciences Center 7 Khon Kaen | Division of Genomic Medicine and Innovation support,Department of Medical Sciences, Ministry of Public Health, Thailand | Surakameth Mahasirimongkol,Nuanjun Wichukchinda,Archawin Rojanawiwat,Pilailuk Akkapaiboon Okada ,Waritta Sawaengdee,Penpitcha Thawong,Pundharika Piboonsiri,Jirapha Pakdee,Natthakul Bunneang |
| hCoV-19/Thailand/DMSc-05955/2021 | EPI_ISL_8437373 | 2021-12-22 | Regional Medical Sciences Center 7 Khon Kaen | Division of Genomic Medicine and Innovation support,Department of Medical Sciences, Ministry of Public Health, Thailand | Surakameth Mahasirimongkol,Nuanjun Wichukchinda,Archawin Rojanawiwat,Pilailuk Akkapaiboon Okada ,Waritta Sawaengdee,Penpitcha Thawong,Pundharika Piboonsiri,Jirapha Pakdee,Natthakul Bunneang |
| hCoV-19/Italy/SIC_CQRC_3421124513/2021 | EPI_ISL_7398901 | 2021-11-23 | Laboratorio CQRC | CQRC_QUALITY CONTROL CHEMICAL BIOLOGICAL RISK_AOOR Villa Sofia Cervello Palermo | Di Gaudio F., Contino F., Brunacci G. |
| hCoV-19/Denmark/DCGC-294164/2021 | EPI_ISL_8381472 | 2021-12-21 | Molekylv¶r Medicinsk Afdeling, Aarhus University Hospital, Aarhus, Denmark | Bioinformatics and Microbial Genomics | Danish Covid-19 Genome Consortium |
| hCoV-19/Denmark/DCGC-294172/2021 | EPI_ISL_8381480 | 2021-12-14 | Department of Bacteria, Parasites and Fungi, Statens Serum Institut, Copenhagen, Denmark | Statens Serum Institut Bioinformatics and Microbial Genomics | Danish Covid-19 Genome Consortium |
| hCoV-19/Italy/SIC_CQRC_2837553/2021 | EPI_ISL_7399690 | 2021-11-23 | Laboratorio CQRC | CQRC_QUALITY CONTROL CHEMICAL BIOLOGICAL RISK_AOOR Villa Sofia Cervello Palermo | Di Gaudio F., Contino F., Brunacci G. |
| hCoV-19/Denmark/DCGC-294641/2021 | EPI_ISL_8382261 | 2021-12-27 | Department of Bacteria, Parasites and Fungi, Statens Serum Institut, Copenhagen, Denmark | Statens Serum Institut Bioinformatics and Microbial Genomics | Danish Covid-19 Genome Consortium |
| hCoV-19/Denmark/DCGC-294546/2021 | EPI_ISL_8382180 | 2021-12-22 | Department of Bacteria, Parasites and Fungi, Statens Serum Institut, Copenhagen, Denmark | Statens Serum Institut Bioinformatics and Microbial Genomics | Danish Covid-19 Genome Consortium |
| hCoV-19/Sweden/2931553263/2021 | EPI_ISL_8382072 | 2021-12-21 | Sweden, Vastragotaland, Unilabs | Unilabs, Eskilstuna, Sweden | Emma Arvidsson |
| hCoV-19/France/ARA-HCL121031708701/2021 | EPI_ISL_8376888 | 2021-12-14 | CNR Virus des Infections Respiratoires - France SUD | CNR Virus des Infections Respiratoires - France SUD | Antonin Bal, Bruno Simon, Gregory Destras, Gwendolyne Burfin, Hadrien Regue, Quentin Semanas, Martine Valette, Bruno Lina, Laurence Josset |
| hCoV-19/France/NAQ-HCL021231010201/2021 | EPI_ISL_8376676 | 2021-12-21 | LABO 17 | CNR Virus des Infections Respiratoires - France SUD | Antonin Bal, Bruno Simon, Gregory Destras, Gwendolyne Burfin, Hadrien Regue, Quentin Semanas, Martine Valette, Bruno Lina, Laurence Josset |
| hCoV-19/Germany/NW-HHU-16698/2021 | EPI_ISL_8377184 | 2021-12-20 | ZOTZ KLIMAS MVZ Dvºsseldorf-Centrum GbR √úBAG fvºr Labormedizin, Genetik, Zytologie, Pathologie | Center of Medical Microbiology, Virology, and Hospital Hygiene, University of Duesseldorf | Maximilian Damagnez; Alexander Dilthey; Patrick Finzer; Katrin Hoffmann; Torsten Houwaart; Lisanna Hvºlse; Malte Kohns Vasconcelos; Nadine Lvºbke; Jessica Nicolai; Klaus Pfeffer; Daniel Strelow; J√∂rg Timm; Andreas Walker; Tobias Wienemann; Rainer Zotz |
| hCoV-19/Guam/H216836/2021 | EPI_ISL_8383085 | 2021-12-06 | Guam Public Health Laboratory | State Laboratories Division, Hawaii State Department of Health | Pamela O'Brien, Drew Kuwazaki, Ayana Garnet, Samantha Cotter, Cheyenne Barela, Cheryl-lynn Daquip, Mark Nagata, Samantha Sruba, Jeffrey Au, Briana Ofilas, Remedios Gose, Daniel Strange, Edward Desmond |

| | | | | | |
|---|---|---|---|---|---|
| hCoV-19/England/ALDP-3014176/2021 | EPI_ISL_8386437 | 2021-12-25 | Lighthouse Lab in Alderley Park | Wellcome Sanger Institute for the COVID-19 Genomics UK (COG-UK) Consortium | Jacquelyn Wynn, Mairead Hyland, The Lighthouse Lab in Alderley Park and Alex Alderton, Roberto Amato, Jeffrey Barrett, Sonia Goncalves, Ewan Harrison, David K. Jackson, Ian Johnston, Dominic Kwiatkowski, Cordelia Langford, John Sillitoe on behalf of the W |
| hCoV-19/England/ALDP-3012F9C/2021 | EPI_ISL_8386480 | 2021-12-23 | Lighthouse Lab in Alderley Park | Wellcome Sanger Institute for the COVID-19 Genomics UK (COG-UK) Consortium | Jacquelyn Wynn, Mairead Hyland, The Lighthouse Lab in Alderley Park and Alex Alderton, Roberto Amato, Jeffrey Barrett, Sonia Goncalves, Ewan Harrison, David K. Jackson, Ian Johnston, Dominic Kwiatkowski, Cordelia Langford, John Sillitoe on behalf of the W |
| hCoV-19/England/QEUH-3047541/2021 | EPI_ISL_8386260 | 2021-12-28 | Lighthouse Lab in Glasgow | Wellcome Sanger Institute for the COVID-19 Genomics UK (COG-UK) Consortium | Harper VanSteenhouse, Yumi Kasai, David Gray, Carol Clugston, Anna Dominiczak and Alex Alderton, Roberto Amato, Jeffrey Barrett, Sonia Goncalves, Ewan Harrison, David K. Jackson, Ian Johnston, Dominic Kwiatkowski, Cordelia Langford, John Sillitoe on behal |
| hCoV-19/England/ALDP-3014264/2021 | EPI_ISL_8387542 | 2021-12-25 | Lighthouse Lab in Alderley Park | Wellcome Sanger Institute for the COVID-19 Genomics UK (COG-UK) Consortium | Jacquelyn Wynn, Mairead Hyland, The Lighthouse Lab in Alderley Park and Alex Alderton, Roberto Amato, Jeffrey Barrett, Sonia Goncalves, Ewan Harrison, David K. Jackson, Ian Johnston, Dominic Kwiatkowski, Cordelia Langford, John Sillitoe on behalf of the W |
| hCoV-19/TrinidadandTobago/99815/2021 | EPI_ISL_7647245 | 2021-11-18 | Trinidad Public Health Laboratory | Carrington Lab, Department of Preclinical Sciences, Faculty of Medical Sciences, The University of the West Indies, St Augustine Campus | Nikita S. D. Sahadeo, Arianne Brown-Jordan, Sarah Hill, Vernie Ramkissoon, Anushka Ramjag, Soren Nicholls, Roshan Parasram, Avery Hinds, Risha Singh, SueMin Nathaniel, Nuno Faria, Oliver Pybus, Christopher Oura, Gabriel Escobar, Christine V. F. Carrington |
| hCoV-19/Switzerland/AG-UZH-IMV-3ba56d97/2021 | EPI_ISL_7284494 | 2021-11-27 | Universit√§t Z√ºrich | Institute of Medical Virology, University of Zurich | Daniel Ehrsam, Isabel St√ºrmer, Catharine Aquino, Joel Wirz, Weihong Qi, Hubert Rehrauer, Verena Kufner, Gabriela Ziltener, Maryam Zaheri, Stefan Schmutz, Annette Audig√©, Maria Gr√ºnberg, Kevin Steiner, Jon Huder, Cyril Shah, Riccarda Capaul, Guido Bloem |
| hCoV-19/India/GJ-INSACOG-GBRC3748/2021 | EPI_ISL_8388886 | 2021-12-29 | Supratech Micropath Laboratory Research Institute Pvt Ltd, Ahmedabad | Gujarat Biotechnology Research Centre | Zarna Patel,Bhadreshsinh Gohil,Nimesh Patel,Nitin Shukla,Akhilesh Modi,Tasnim Trivedi,Jaykumar Rangani,Ramesh Pandit,Nitin Savaliya,Priyank Chavda,Sonal Sharma ,Roshani Mishra,Disha Vora,Janvi Raval,Apurvasinh Puvar,Shiva,Chaitanya Joshi,Madhvi Joshi |
| hCoV-19/Kenya/SS4390/2021 | EPI_ISL_8389245 | 2021-12-03 | Aga Khan University Hospital | KEMRI-Wellcome Trust Research Programme,Kilifi | Githinji G., Ochola I., Agoti C.,Mohamed K.S., de Laurent Z., Mburu M.W., Morobe J., Ndwiga L. Makori T., Ongera E., Lambisia A., Shah R.,Revathi G., Sayed S., Adam R.Nokes D.J. |
| hCoV-19/Kenya/SS4918/2021 | EPI_ISL_8390137 | 2021-12-10 | Machakos Level 5 Hospital | KEMRI-Wellcome Trust Research Programme,Kilifi | Githinji G.,Ochola I, Agoti C.,Mohamed K.S.,de Laurent Z.,Mburu M.W,Morobe J.,Lambisia A. Makori T.,Ongera E.,D.J Nokes, Ndwiga L., Mutunga D., Kituku A. |
| hCoV-19/Kenya/SS4920/2021 | EPI_ISL_8390150 | 2021-12-10 | Machakos Level 5 Hospital | KEMRI-Wellcome Trust Research Programme,Kilifi | Githinji G.,Ochola I, Agoti C.,Mohamed K.S.,de Laurent Z.,Mburu M.W,Morobe J.,Lambisia A. Makori T.,Ongera E.,D.J Nokes, Ndwiga L., Mutunga D., Kituku A. |

| | | | | | |
|---|---|---|---|---|---|
| hCoV-19/Kenya/SS4229/2021 | EPI_ISL_8390168 | 2021-11-26 | KEMRI CMR Nairobi | KEMRI-Wellcome Trust Research Programme,Kilifi | Githinji G.,Ochola I, Agoti C.,Mohamed K.S.,de Laurent Z.,Mburu M.W,Morobe J.,Lambisia A. Makori T.,Ongera E.,D.J Nokes, Ndwiga L.Muema A., Tigoi C |
| hCoV-19/USA/AZ-TG1088721/2021 | EPI_ISL_8365728 | 2021-10-12 | Arizona Department of Health Services | TGen North | Jolene Bowers, Heather Centner, Brett Van Tassel, Chris French, Hayley Yaglom, Darrin Lemmer, Dave Engelthaler |
| hCoV-19/USA/AZ-TG935185/2021 | EPI_ISL_8363671 | 2021-06-04 | Arizona Department of Health Services | TGen North | Jolene Bowers, Heather Centner, Brett Van Tassel, Chris French, Hayley Yaglom, Darrin Lemmer, Dave Engelthaler |
| hCoV-19/USA/CA-CDC-FG-218468/2021 | EPI_ISL_8393135 | 2021-12-23 | Fulgent Genetics | Centers for Disease Control and Prevention Division of Viral Diseases, Pathogen Discovery | Dakota Howard,Dhwani Batra,Peter Cook,Jason Caravas,Benjamin Rambo-Martin,Scott Sammons,Yvette Unoarumhi,Matthew Schmerer,Kristine Lacek,Tymeckia Kendall,Victoria Caban Figueroa,Shatavia Morrison,Christopher Gulvick,Erisa Sula,Harry Gao,Mickey Li,John Gao |
| hCoV-19/USA/AZ-TG1046967/2021 | EPI_ISL_8365213 | 2021-09-03 | Arizona Department of Health Services | TGen North | Jolene Bowers, Heather Centner, Brett Van Tassel, Chris French, Hayley Yaglom, Darrin Lemmer, Dave Engelthaler |
| hCoV-19/USA/AZ-TG989877/2021 | EPI_ISL_8364409 | 2021-07-13 | Arizona Department of Health Services | TGen North | Jolene Bowers, Heather Centner, Brett Van Tassel, Chris French, Hayley Yaglom, Darrin Lemmer, Dave Engelthaler |
| hCoV-19/USA/IL-RIPHL_80237_G/2021 | EPI_ISL_7149806 | 2021-10-05 | Alverno Labs | RIPHL at Rush University Medical Center | Stefan Green, Kevin Kunstman, Marieta Hyde, Laura Furtado, Felix Araujo Perez, Edith Perez, Sofiya Bobrovska, Alyse Kittner, Hannah Barbian, Melissa Mace |
| hCoV-19/USA/TX-CDC-ASC210632913/2021 | EPI_ISL_8394045 | 2021-12-21 | Aegis Sciences Corporation | Centers for Disease Control and Prevention Division of Viral Diseases, Pathogen Discovery | Dakota Howard,Dhwani Batra,Peter Cook,Jason Caravas,Benjamin Rambo-Martin,Scott Sammons,Yvette Unoarumhi,Matthew Schmerer,Kristine Lacek,Tymeckia Kendall,Victoria Caban Figueroa,Shatavia Morrison,Christopher Gulvick,Erisa Sula,Cyndi Clark,Patrick Campbell |
| hCoV-19/Gambia/76301/2021 | EPI_ISL_8401599 | 2021-09-23 | MRCG at LSHTM Genomics lab | MRCG at LSHTM Genomics lab | Abdul Karim sesay, Abdoulie Kanteh, Jarra Manneh, Mariama Kujabi, Bakary Sanyang,Dabiri Damilari, Yaya bah |
| hCoV-19/Canada/AB-ABPHL-46220/2021 | EPI_ISL_8391509 | 2021-12-13 | Edmonton Provincial Lab | Public Health Agency of Canada (PHAC) National Microbiology Laboratory | Buss, E, Croxen M, Deo A, Dieu P, Gill K, Ferrato C, Khan F, Koleva P, Li V, Lloyd C, Lynch T, Ma R, Murphy S, Pabbaraju K, Shokoples S, Tipples G, Thayer J, Whitehouse M, Wong A, Yu C, Zelyas N |
| hCoV-19/Italy/LOM_Policlinico_Milano_18967665/2021 | EPI_ISL_7548716 | 2021-12-06 | Fondazione IRCCS Ca' Granda Ospedale Maggiore Policlinico | Fondazione IRCCS Ca' Granda Ospedale Maggiore Policlinico | Ferruccio Ceriotti, Sara Uceda Renteria |
| hCoV-19/Italy/SIC_CQRC_2960174/2021 | EPI_ISL_7717659 | 2021-12-09 | Laboratorio CQRC | CQRC_QUALITY CONTROL CHEMICAL BIOLOGICAL RISK_AOOR Villa Sofia Cervello Palermo | Di Gaudio F., Contino F., Brunacci G. |
| hCoV-19/Italy/SIC_CQRC_2996322/2021 | EPI_ISL_8002139 | 2021-12-14 | Laboratorio CQRC | CQRC_QUALITY CONTROL CHEMICAL BIOLOGICAL RISK_AOOR Villa Sofia Cervello Palermo | Di Gaudio F., Contino F., Brunacci G. |
| hCoV-19/Italy/SIC-CQRC-3065534/2021 | EPI_ISL_8337828 | 2021-12-23 | Laboratorio CQRC | CQRC_QUALITY CONTROL CHEMICAL BIOLOGICAL RISK_AOOR Villa Sofia Cervello Palermo | Di Gaudio F., Contino F., Brunacci G. |

| | | | | | |
|---|---|---|---|---|---|
| hCoV-19/CzechRepublic/NRL_18531/2021 | EPI_ISL_8402344 | 2021-12-04 | National Institute of Public Health | National Institute of Public Health | Helena Jirincova, Jaromira Vecerova, Timotej Suri, Jan Moskalyk, Alexander Nagy |
| hCoV-19/SouthKorea/KDCA24699/2021 | EPI_ISL_8403656 | 2021-12-19 | Division of Emerging Infectious Diseases, Bureau of Infectious Diseases Diagnosis Control, Korea Disease Control and Prevention Agency | Diseases, Bureau of Infectious Diseases Diagnosis Control, Korea Disease Control and Prevention Agency | Jeong-Ah Kim, Ae Kyung Park, Il-Hwan Kim, Chae Young Lee, Hyuck Jin Lee, Eun-Jin Kim |
| hCoV-19/USA/NY-CDC-IBX0366522/2021 | EPI_ISL_8404589 | 2021-12-18 | Infinity Biologix | Centers for Disease Control and Prevention Division of Viral Diseases, Pathogen Discovery | Dakota Howard,Dhwani Batra,Peter Cook,Jason Caravas,Benjamin Rambo-Martin,Scott Sammons,Yvette Unoarumhi,Matthew Schmerer,Kristine Lacek,Tymeckia Kendall,Victoria Caban Figueroa,Shatavia Morrison,Christopher Gulvick,Erisa Sula,Christian Bixby,Yihe Wang,Jo |
| hCoV-19/Australia/WA873/2021 | EPI_ISL_8405491 | 2021-12-30 | PathWest Laboratory Medicine WA | PathWest Laboratory Medicine WA Microbial Surveillance Unit | PathWest Laboratory Medicine WA Microbial Surveillance Unit |
| hCoV-19/Ecuador/NIC-INSPI-202112211738/2021 | EPI_ISL_8406420 | 2021-12-22 | SYNLAB | INSPI-CRN DE INFLUENZA Y OTROS VIRUS RESPIRATORIOS | Alfredo Bruno, Maritza Olmedo, Michelle P√°ez, Jimmy Garc√©s, Fernando Llerena, Daniel Ramos, Maria Angelica Becerra, Lizbeth Pati√±o, Diego Morales, Ruben Armas, |
| hCoV-19/Ecuador/NIC-INSPI-211221442/2021 | EPI_ISL_8406424 | 2021-12-21 | INSPI-CRN DE INFLUENZA Y OTROS VIRUS RESPIRATORIOS | INSPI-CRN DE INFLUENZA Y OTROS VIRUS RESPIRATORIOS | Alfredo Bruno, Maritza Olmedo, Michelle P√°ez, Jimmy Garc√©s, Fernando Llerena, Lizbeth Pati√±o, Diego Morales, Ruben Armas, |
| hCoV-19/Nepal/NPHL/S-624/2021 | EPI_ISL_8408856 | 2021-08-06 | Bheri Hospital COVID PCR Labortaory | National Public Health Laboratory/Quadrum Institute Bioscience | National Public Health Laboratory Team /Quadrum Institute Bioscience UK Team (NVAP for funding /Administration) |
| hCoV-19/Australia/ACT2034/2021 | EPI_ISL_7569968 | 2021-12-07 | ACT Pathology | Schwessinger Lab | Austin Bird, Carolina Correa Ospina, Emma Crean, Bayantes Dagvadorj, Scott Ferguson, Robyn Hall, Evie Hodgson, Ashley Jones, Elise Kellett, Craig Kennedy, Karina Kennedy, Rachel Leonard, Carl McCombe, Catalina Barrag√°n Quintero, Rene Riedelbauch, Benjami |
| hCoV-19/Australia/ACT2010/2021 | EPI_ISL_7569944 | 2021-12-05 | ACT Pathology | Schwessinger Lab | Austin Bird, Carolina Correa Ospina, Emma Crean, Bayantes Dagvadorj, Scott Ferguson, Robyn Hall, Evie Hodgson, Ashley Jones, Elise Kellett, Craig Kennedy, Karina Kennedy, Rachel Leonard, Carl McCombe, Catalina Barrag√°n Quintero, Rene Riedelbauch, Benjami |
| hCoV-19/Australia/ACT2024/2021 | EPI_ISL_7569961 | 2021-12-05 | ACT Pathology | Schwessinger Lab | Austin Bird, Carolina Correa Ospina, Emma Crean, Bayantes Dagvadorj, Scott Ferguson, Robyn Hall, Evie Hodgson, Ashley Jones, Elise Kellett, Craig Kennedy, Karina Kennedy, Rachel Leonard, Carl McCombe, Catalina Barrag√°n Quintero, Rene Riedelbauch, Benjami |
| hCoV-19/South_Africa/SU-NHLS_3593/2021 | EPI_ISL_8409324 | 2021-12-28 | Division of Medical Virology, National Health Laboratory Service (NHLS), Tygerberg Hospital / Stellenbosch University | Health Laboratory Service (NHLS), Tygerberg Hospital / Stellenbosch University | Susan Engelbrecht, Tongai Maponga, Emilia Wilson, Mathilda Claassen, Tania Stander, Gert van Zyl, Wolfgang Preiser |

| | | | | | |
|---|---|---|---|---|---|
| hCoV-19/Scotland/QEUH-3035F66/2021 | EPI_ISL_8410469 | 2021-12-27 | Lighthouse Lab in Glasgow | Wellcome Sanger Institute for the COVID-19 Genomics UK (COG-UK) Consortium | Harper VanSteenhouse, Yumi Kasai, David Gray, Carol Clugston, Anna Dominiczak and Alex Alderton, Roberto Amato, Jeffrey Barrett, Sonia Goncalves, Ewan Harrison, David K. Jackson, Ian Johnston, Dominic Kwiatkowski, Cordelia Langford, John Sillitoe on behal |
| hCoV-19/SouthAfrica/SU-NHLS-3581/2021 | EPI_ISL_8409391 | 2021-12-26 | Division of Medical Virology, National Health Laboratory Service (NHLS), Tygerberg Hospital / Stellenbosch University | Health Laboratory Service (NHLS), Tygerberg Hospital / Stellenbosch University | Susan Engelbrecht, Tongai Maponga, Emilia Wilson, Mathilda Claassen, Tania Stander, Gert van Zyl, Wolfgang Preiser |
| hCoV-19/Slovenia/17-127938-CE/2021 | EPI_ISL_8417778 | 2021-12-17 | National Laboratory for Health, Environment and Food, OMM, Celje | Health, Environment and Food) / CISLD (Clinical Institute of Special Laboratory Diagnostics), University Children's Hospital, University Medical Center Ljubljana | Sandra Janezic, Aleksander Mahnic, Maja Rupnik, Tja≈°a ≈Ωohar ∫åretnik, Alenka ≈†torman, Nika Gobec, Aleksander Kocuvan, Kaja Tominc, Ma≈°a Jarƒçiƒç, Mateja Borinc, Da≈°a Kavka / Jernej Kovaƒç, Barbara Jenko Bizjan, Tine Tesovnik, Robert ≈†ket, Katarina K |
| hCoV-19/Denmark/DCGC-295490/2021 | EPI_ISL_8418172 | 2021-12-27 | Department of Bacteria, Parasites and Fungi, Statens Serum Institut, Copenhagen, Denmark | Statens Serum Institut Bioinformatics and Microbial Genomics | Danish Covid-19 Genome Consortium |
| hCoV-19/Denmark/DCGC-295903/2021 | EPI_ISL_8418746 | 2021-12-28 | Department of Bacteria, Parasites and Fungi, Statens Serum Institut, Copenhagen, Denmark | Statens Serum Institut Bioinformatics and Microbial Genomics | Danish Covid-19 Genome Consortium |
| hCoV-19/Denmark/DCGC-295999/2021 | EPI_ISL_8418838 | 2021-12-28 | Department of Bacteria, Parasites and Fungi, Statens Serum Institut, Copenhagen, Denmark | Statens Serum Institut Bioinformatics and Microbial Genomics | Danish Covid-19 Genome Consortium |
| hCoV-19/Denmark/DCGC-296018/2021 | EPI_ISL_8418856 | 2021-12-27 | Department of Bacteria, Parasites and Fungi, Statens Serum Institut, Copenhagen, Denmark | Statens Serum Institut Bioinformatics and Microbial Genomics | Danish Covid-19 Genome Consortium |
| hCoV-19/Denmark/DCGC-296015/2021 | EPI_ISL_8418853 | 2021-12-27 | Department of Bacteria, Parasites and Fungi, Statens Serum Institut, Copenhagen, Denmark | Statens Serum Institut Bioinformatics and Microbial Genomics | Danish Covid-19 Genome Consortium |
| hCoV-19/Denmark/DCGC-296086/2021 | EPI_ISL_8418922 | 2021-12-29 | Department of Bacteria, Parasites and Fungi, Statens Serum Institut, Copenhagen, Denmark | Statens Serum Institut Bioinformatics and Microbial Genomics | Danish Covid-19 Genome Consortium |
| hCoV-19/Denmark/DCGC-296466/2021 | EPI_ISL_8419269 | 2021-12-27 | Department of Bacteria, Parasites and Fungi, Statens Serum Institut, Copenhagen, Denmark | Statens Serum Institut Bioinformatics and Microbial Genomics | Danish Covid-19 Genome Consortium |
| hCoV-19/Denmark/DCGC-296525/2021 | EPI_ISL_8419324 | 2021-12-29 | Department of Bacteria, Parasites and Fungi, Statens Serum Institut, Copenhagen, Denmark | Statens Serum Institut Bioinformatics and Microbial Genomics | Danish Covid-19 Genome Consortium |
| hCoV-19/Denmark/DCGC-296659/2021 | EPI_ISL_8419442 | 2021-12-29 | Department of Bacteria, Parasites and Fungi, Statens Serum Institut, Copenhagen, Denmark | Statens Serum Institut Bioinformatics and Microbial Genomics | Danish Covid-19 Genome Consortium |
| hCoV-19/Denmark/DCGC-296841/2021 | EPI_ISL_8419703 | 2021-12-27 | Department of Bacteria, Parasites and Fungi, Statens Serum Institut, Copenhagen, Denmark | Statens Serum Institut Bioinformatics and Microbial Genomics | Danish Covid-19 Genome Consortium |
| hCoV-19/Denmark/DCGC-297044/2021 | EPI_ISL_8419884 | 2021-12-28 | Department of Bacteria, Parasites and Fungi, Statens Serum Institut, Copenhagen, Denmark | Statens Serum Institut Bioinformatics and Microbial Genomics | Danish Covid-19 Genome Consortium |

| | | | | | |
|---|---|---|---|---|---|
| hCoV-19/Denmark/DCGC-297048/2021 | EPI_ISL_8419888 | 2021-12-28 | Department of Bacteria, Parasites and Fungi, Statens Serum Institut, Copenhagen, Denmark | Statens Serum Institut Bioinformatics and Microbial Genomics | Danish Covid-19 Genome Consortium |
| hCoV-19/Denmark/DCGC-297122/2021 | EPI_ISL_8420649 | 2021-12-28 | Department of Bacteria, Parasites and Fungi, Statens Serum Institut, Copenhagen, Denmark | Statens Serum Institut Bioinformatics and Microbial Genomics | Danish Covid-19 Genome Consortium |
| hCoV-19/India/AP-CCMB-CIB7945/2021 | EPI_ISL_8421190 | 2021-11-29 | AP SSO | CSIR-Centre for Cellular and Molecular Biology-INSACOG | Jandhyala Sai Krishna,Tulasi Nagabandi, Lamuk Zaveri,Malini Nemalikanti,Sreelekshmi MS, Ara Sreenivas,Shreekant Verma, Amareshwar Vodapalli,B Himasri,Valli Nagalakshmi Undamatla, Sumedha Avadhanula,Priya Nurkurthy,Vidhyadhari Methuku, Archana Bharadwaj Si |
| hCoV-19/India/TG-RFCH02795_CIB8168/2021 | EPI_ISL_8421295 | 2021-02-05 | CSIR-Centre for Cellular and Molecular Biology | CSIR-Centre for Cellular and Molecular Biology | Shreekant Verma,Ara Sreenivas, Amareshwar Vodapalli,B Himasri,Valli Nagalakshmi Undamatla, Sumedha Avadhanula,Priya Nurkurthy,Vidhyadhari Methuku, Sreelekshmi MS,Malini Nemalikanti,Lamuk Zaveri,Tulasi Nagabandi,Jandhyala Sai Krishna, Archana Bharadwaj Siv |
| hCoV-19/India/AP-CCMB-CIB8285/2021 | EPI_ISL_8421336 | 2021-11-22 | AP SSO | CSIR-Centre for Cellular and Molecular Biology-INSACOG | Priya Nurkurthy,Vidhyadhari Methuku,Sumedha Avadhanula,Valli Nagalakshmi Undamatla,Amareshwar Vodapalli, B Himasri,Shreekant Verma,Ara Sreenivas, Sreelekshmi MS,Malini Nemalikanti,Lamuk Zaveri,Tulasi Nagabandi,Jandhyala Sai Krishna, Archana Bharadwaj Siv |
| hCoV-19/Bangladesh/icddrb-GAC2-01280/2021 | EPI_ISL_8422268 | 2021-08-25 | Virology Laboratory, International Centre for Diarrhoeal Disease Research, Bangladesh (ICDDR,B) | Virology Laboratory, International Centre for Diarrhoeal Disease Research, Bangladesh (ICDDR,B) | Mohammad Enayet Hossain, Mojnu Miah, Mst. Noorjahan Begum, Rashedul Hasan, Md. Mahfuzur Rahman, Mohammed Ziaur Rahman, Mustafa Mahfuz, Muntasir Alam, Kamrun Nahar, Shams El Arifeen, Mustafizur Rahman |
| hCoV-19/USA/CO-CDC-MMB12077039/2021 | EPI_ISL_8424795 | 2021-12-09 | Mako Medical | Centers for Disease Control and Prevention Division of Viral Diseases, Pathogen Discovery | Dakota Howard,Dhwani Batra,Peter Cook,Jason Caravas,Benjamin Rambo-Martin,Scott Sammons,Yvette Unoarumhi,Matthew Schmerer,Kristine Lacek,Tymeckia Kendall,Victoria Caban Figueroa,Shatavia Morrison,Christopher Gulvick,Erisa Sula,Matthew Tugwell,Lauren Moon, |
| hCoV-19/USA/PA-CDC-2-5321470/2021 | EPI_ISL_8426681 | 2021-12-06 | PA Department of Health, Bureau of Laboratories | Centers for Disease Control and Prevention Division of Viral Diseases, Pathogen Discovery | Mili Sheth,Sarah Nobles,Joseph Madden,Jasmine Padilla,Alex Burgin,Meghan Bentz,Mark Burroughs,Peter Cook,Clinton Paden,Dhwani Batra,Kristen Knipe,Dakota Howard,Yvette Unoarumhi,Matthew Schmerer,Ben Rambo-Martin,Kristine Lacek,Sam Shepard,Dave Wentworth,Vi |
| hCoV-19/USA/GA-CDC-STM-FQKVDPGX8/2021 | EPI_ISL_8427813 | 2021-12-21 | Helix | Centers for Disease Control and Prevention Division of Viral Diseases, Pathogen Discovery | Dakota Howard,Dhwani Batra,Peter Cook,Jason Caravas,Benjamin Rambo-Martin,Scott Sammons,Yvette Unoarumhi,Matthew Schmerer,Kristine Lacek,Tymeckia Kendall,Victoria Caban Figueroa,Helix CA,Shatavia Morrison,Christopher Gulvick,Erisa Sula,Clinton Paden,Dunca |

| | | | | | |
|---|---|---|---|---|---|
| hCoV-19/Brazil/RJ-FIOCRUZ-66290/2021 | EPI_ISL_8430500 | 2021-12-20 | Laboratorio Central de Saude Publica do Estado do Rio de Janeiro (LACEN/RJ) | Laboratory of Respiratory Viruses and Measles, Oswaldo Cruz Institute, FIOCRUZ | Paola Resende, Luciana Appolinario, Fernando Motta, Elisa Cavalcante Pereira, Ighor Arantes, Alice Sampaio Rocha, Bruna Mendon√ßa da Silva, J√©ssica Gra√ßa Macedo de Carvalho, Larissa Macedo Pinto, Victor Guimaraes, Andrea Cony Cavalcanti, Marilda Siqueir |
| hCoV-19/USA/MA-CDCBI-CRSP_72BOUR62RBQLFDOW/2021 | EPI_ISL_8431756 | 2021-12-27 | Broad Institute Clinical Research Sequencing Platform | Infectious Disease Program, Broad Institute of Harvard and MIT | Siddle,K.J., Adams,G., Pearlman,L., Gladden-Young,A., Vicente,G., Blumenstiel,B., DeFelice,M., Lee,M., McGovern,S., Lagerborg,K., Rudy,M., DeRuff,K., Carter,A., Normandin,E., Bauer,M., Reilly,S., Tomkins-Tinch,C., Loreth,C., Chaluvadi,S., Meldrim,J., Gran |
| hCoV-19/USA/MA-CDCBI-CRSP_SDKBDBCPTMSIKPHC/2021 | EPI_ISL_8433943 | 2021-12-26 | Broad Institute Clinical Research Sequencing Platform | Infectious Disease Program, Broad Institute of Harvard and MIT | Siddle,K.J., Adams,G., Pearlman,L., Gladden-Young,A., Vicente,G., Blumenstiel,B., DeFelice,M., Lee,M., McGovern,S., Lagerborg,K., Rudy,M., DeRuff,K., Carter,A., Normandin,E., Bauer,M., Reilly,S., Tomkins-Tinch,C., Loreth,C., Chaluvadi,S., Meldrim,J., Gran |
| hCoV-19/USA/NH-CDCBI-CRSP_ITUSILITB2R3OKJ6/2021 | EPI_ISL_8434746 | 2021-12-27 | Broad Institute Clinical Research Sequencing Platform | Infectious Disease Program, Broad Institute of Harvard and MIT | Siddle,K.J., Adams,G., Pearlman,L., Gladden-Young,A., Vicente,G., Blumenstiel,B., DeFelice,M., Lee,M., McGovern,S., Lagerborg,K., Rudy,M., DeRuff,K., Carter,A., Normandin,E., Bauer,M., Reilly,S., Tomkins-Tinch,C., Loreth,C., Chaluvadi,S., Meldrim,J., Gran |
| hCoV-19/Brazil/BA-FIOCRUZ-PVM88225/2021 | EPI_ISL_8127841 | 2021-10-25 | Laborat√≥rio Central de Sa√∫de P√∫blica - LACEN/BA | Plataforma de Vigilancia Molecular (PVM) - FIOCRUZ/BA | Laise de Moraes, Francys A Rangel, Eduardo Oyama, Laila A da Silva, Marina Cucco, Pedro Silveira, Stephane Tosta, Luciana Oliveira, Vanessa Nardy, Gabriela Menezes, Marcela Gomez, Lenisa Dandara, Jaqueline Gomes, Felicidade Pereira, Arabela Leal, Tiago Gr |
| hCoV-19/Brazil/BA-FIOCRUZ-PVM88257/2021 | EPI_ISL_8127903 | 2021-11-23 | Hospital S√£o Rafael - H.S.R/BA | Plataforma de Vigilancia Molecular (PVM) - FIOCRUZ/BA | Laise de Moraes, Francys A Rangel, Eduardo Oyama, Laila A da Silva, Marina Cucco, Pedro Silveira, Camila Araujo de Lorenzo Barcia, Ana Verena Almeida Mendes, Marilia Miranda Franco, Carolina Kymie Vasques Nonaka, Bruno Solano de Freitas Souza, Tiago Graf, |
| hCoV-19/India/TG-CDFD-C-272039/2021 | EPI_ISL_7649197 | 2021-11-17 | Gandhi Medical College | Centre for DNA Fingerprinting and Diagnostics | Asmita Gupta, Rupin Shelake, Arunkumar Karunanidhi, Vinay Donipadi, Divya Vashisht, Ashwin Dalal, Nagamani Kammili, Murali Bashyam |
| hCoV-19/Brazil/PR-NVBS7920GENOV828259240776/2021 | EPI_ISL_7366057 | 2021-10-14 | DASA | DASA | Rodrigo Guarischi, Paulo Pierry, Adriano Bonaldi, Cristina Oliveira, Bianca Cota, Luciane Sussuchi, Camila Romano, Angelica Hristov, Lidia Yamamoto, Rodrigo Salazar, Annelise Lopes, Jose Levi |
| hCoV-19/Cambodia/886723/2021 | EPI_ISL_7370866 | 2021-11-30 | Virology Unit, Institut Pasteur du Cambodge | Virology Unit, Institut Pasteur du Cambodge | Leakhena Pum, Sophoannadedh Rath, Cecile Troupin, Jurre Y Siegers, Ly Sovann, Kraing Sidonn, Yi Sengdoeurn, Chin Savuth, Chau Darapheak, Veasna Duong, Erik A Karlsson |
| hCoV-19/Switzerland/BL-ETHZ-34931518/2021 | EPI_ISL_7371675 | 2021-11-23 | Viollier AG | Department of Biosystems Science and Engineering, ETH Z√ºrich | Chaoran Chen, Sarah Nadeau, Louis du Plessis, Catharine Aquino, Ivan Topolsky, Kim Philipp Jablonski, Lara Fuhrmann, Daniel Ehrsam, Isabel St√ºrmer, Andreia Cabral de Gouvea, Maria Domenica Moccia, Simon Gr√ºter, Timothy Sykes, Lennart Opitz, Griffin Whit |

| | | | | | |
|---|---|---|---|---|---|
| hCoV-19/Thailand/NIC_BKK_SEQ23180/2021 | EPI_ISL_8373249 | 2021-12-14 | Samitivej Srinakarin Hospital | National Institute of Health, Department of Medical Sciences, Ministry of Public Health, Thailand | Pilailuk Okada; Siripaporn Phuygun; Nuttida Thongpramul; Pakorn Piromtong; Natchaya¬† Khiadsang; Thanutsapa Thanadachakul; Sirikanda Wimol; Suratchana Mitrat; Sunthareeya Waicharoen; Archawin Rojanawiwat |
| hCoV-19/Switzerland/BL-ETHZ-35010183/2021 | EPI_ISL_7620521 | 2021-11-29 | Viollier AG | Department of Biosystems Science and Engineering, ETH Zürich | Christian Beisel, Sarah Nadeau, Chaoran Chen, Louis du Plessis, Ivan Topolsky, Kim Philipp Jablonski, Lara Fuhrmann, David Dreifuss, Pelin Icer, Tobias Schv§r, Ina Nissen, Natascha Santacroce, Rebecca Denes, Mirjam Feldkamp, Elodie Burcklen, Christiane Be |
| hCoV-19/India/GJ-INSACOG-GBRC2991/2021 | EPI_ISL_7141056 | 2021-11-30 | Unipath Speciality Laboratory Limited, Ahmedabad | Gujarat Biotechnology Research Centre | Sonal Sharma ,Janvi Raval,Apurvasinh Puvar,Zarna Patel,Dinesh Kumar,Bhadreshsinh Gohil,Nimesh Patel,Nitin Shukla,Ramesh Pandit,Nitin Savaliya,Priyank Chavda,Jwalant Shah,Chaitanya Joshi,Madhvi Joshi |
| hCoV-19/USA/PA-VSP10158/2021 | EPI_ISL_7814073 | 2021-11-28 | Hospital of the University of Pennsylvania Molecular Pathology Lab | Bushman | Andrew D. Marques, John K. Everett, Kyle Rodino, Shantan Reddy, Aoife M. Roche, Young Hwang, Scott Sherrill-Mix, Samantha A. Whiteside, Jevon Graham-Wooten, Layla A. Khatib, Ayannah S. Fitzgerald, Arupa Ganguly, Mike Feldman, Brendan Kelly, Ronald G. Coll |
| hCoV-19/Kenya/ILRI_COVM01881/2021 | EPI_ISL_10011166 | 2021-12-21 | International Livestock Research Institute | International Livestock Research Institute | Samuel O. Oyola, Daniel Ouso, Gilbert Kibet, Shebbar Osiany, Edward Kiritu, Paul Dobi, Collins Muli, Patrick Amoth, Vishvanath Nene, Sonal P. Henson, Edward O. Abworo |
| hCoV-19/Kenya/ILRI_COVM01889/2021 | EPI_ISL_10011174 | 2021-12-21 | International Livestock Research Institute | International Livestock Research Institute | Samuel O. Oyola, Daniel Ouso, Gilbert Kibet, Shebbar Osiany, Edward Kiritu, Paul Dobi, Collins Muli, Patrick Amoth, Vishvanath Nene, Sonal P. Henson, Edward O. Abworo |
| hCoV-19/Kenya/ILRI_COVM01915/2021 | EPI_ISL_10011200 | 2021-12-22 | International Livestock Research Institute | International Livestock Research Institute | Samuel O. Oyola, Daniel Ouso, Gilbert Kibet, Shebbar Osiany, Edward Kiritu, Paul Dobi, Collins Muli, Patrick Amoth, Vishvanath Nene, Sonal P. Henson, Edward O. Abworo |
| hCoV-19/Kenya/ILRI_COVM01972/2021 | EPI_ISL_10011252 | 2021-12-23 | International Livestock Research Institute | International Livestock Research Institute | Samuel O. Oyola, Daniel Ouso, Gilbert Kibet, Shebbar Osiany, Edward Kiritu, Paul Dobi, Collins Muli, Patrick Amoth, Vishvanath Nene, Sonal P. Henson, Edward O. Abworo |
| hCoV-19/Kenya/ILRI_COVM01988/2021 | EPI_ISL_10011258 | 2021-12-28 | International Livestock Research Institute | International Livestock Research Institute | Samuel O. Oyola, Daniel Ouso, Gilbert Kibet, Shebbar Osiany, Edward Kiritu, Paul Dobi, Collins Muli, Patrick Amoth, Vishvanath Nene, Sonal P. Henson, Edward O. Abworo |
| hCoV-19/Kenya/ILRI_COVM02218/2021 | EPI_ISL_10011357 | 2021-06-18 | Molecular Biology laboratory, Universit√© Evang√©lique en Afrique | International Livestock Research Institute | Samuel O. Oyola, Daniel Ouso, Gilbert Kibet, Shebbar Osiany, Edward Kiritu, Paul Dobi, Collins Muli, Patrick Amoth, Vishvanath Nene, Sonal P. Henson, Edward O. Abworo |
| hCoV-19/Kenya/ILRI_COVM02266/2021 | EPI_ISL_10011394 | 2021-12-21 | International Livestock Research Institute | International Livestock Research Institute | Samuel O. Oyola, Daniel Ouso, Gilbert Kibet, Shebbar Osiany, Edward Kiritu, Paul Dobi, Collins Muli, Patrick Amoth, Vishvanath Nene, Sonal P. Henson, Edward O. Abworo |

| | | | | | |
|---|---|---|---|---|---|
| hCoV-19/Kenya/ILRI_COVM02295/2021 | EPI_ISL_10011411 | 2021-12-22 | International Livestock Research Institute | International Livestock Research Institute | Samuel O. Oyola, Daniel Ouso, Gilbert Kibet, Shebbar Osiany, Edward Kiritu, Paul Dobi, Collins Muli, Patrick Amoth, Vishvanath Nene, Sonal P. Henson, Edward O. Abworo |
| hCoV-19/Kenya/ILRI_COVM02341/2021 | EPI_ISL_10011440 | 2021-12-28 | International Livestock Research Institute | International Livestock Research Institute | Samuel O. Oyola, Daniel Ouso, Gilbert Kibet, Shebbar Osiany, Edward Kiritu, Paul Dobi, Collins Muli, Patrick Amoth, Vishvanath Nene, Sonal P. Henson, Edward O. Abworo |
| hCoV-19/Kenya/ILRI_COVM02423/2021 | EPI_ISL_10011516 | 2021-12-25 | International Livestock Research Institute | International Livestock Research Institute | Samuel O. Oyola, Daniel Ouso, Gilbert Kibet, Shebbar Osiany, Edward Kiritu, Paul Dobi, Collins Muli, Patrick Amoth, Vishvanath Nene, Sonal P. Henson, Edward O. Abworo |
| hCoV-19/Kenya/ILRI_COVM02487/2021 | EPI_ISL_10011576 | 2021-12-28 | International Livestock Research Institute | International Livestock Research Institute | Samuel O. Oyola, Daniel Ouso, Gilbert Kibet, Shebbar Osiany, Edward Kiritu, Paul Dobi, Collins Muli, Patrick Amoth, Vishvanath Nene, Sonal P. Henson, Edward O. Abworo |
| hCoV-19/Kenya/ILRI_COVM02505/2021 | EPI_ISL_10011594 | 2021-12-27 | International Livestock Research Institute | International Livestock Research Institute | Samuel O. Oyola, Daniel Ouso, Gilbert Kibet, Shebbar Osiany, Edward Kiritu, Paul Dobi, Collins Muli, Patrick Amoth, Vishvanath Nene, Sonal P. Henson, Edward O. Abworo |
| hCoV-19/Kenya/ILRI_COVM02509/2021 | EPI_ISL_10011598 | 2021-12-28 | International Livestock Research Institute | International Livestock Research Institute | Samuel O. Oyola, Daniel Ouso, Gilbert Kibet, Shebbar Osiany, Edward Kiritu, Paul Dobi, Collins Muli, Patrick Amoth, Vishvanath Nene, Sonal P. Henson, Edward O. Abworo |
| hCoV-19/Kenya/ILRI_COVM02527/2021 | EPI_ISL_10011606 | 2021-12-25 | International Livestock Research Institute | International Livestock Research Institute | Samuel O. Oyola, Daniel Ouso, Gilbert Kibet, Shebbar Osiany, Edward Kiritu, Paul Dobi, Collins Muli, Patrick Amoth, Vishvanath Nene, Sonal P. Henson, Edward O. Abworo |
| hCoV-19/England/PHEC-YYNCTGJ/2021 | EPI_ISL_10014470 | 2021-12-08 | Respiratory Virus Unit, Microbiology Services Colindale, Public Health England | COVID-19 Genomics UK (COG-UK) Consortium | PHE Covid Sequencing Team |
| hCoV-19/India/TN-CDFD-CO-1133/2021 | EPI_ISL_10019249 | 2021-12-30 | Directorate Of Public Health And Preventive Medicine | Centre for DNA Fingerprinting and Diagnostics | Asmita Gupta, Rupin Shelake, Arunkumar Karunanidhi, Vinay Donipadi, Divya Vashisht, Ashwin Dalal, Selvavinayagam Sivaprakasam, Darez Ahamed, Sampath Palani, Raju Sivadoss, Avudaiselvi Rathinasamy, Gurunathan Subramanian, Rajesh Kumar Manivannan, Devi Moni |
| hCoV-19/India/TN-CDFD-CO-1144/2021 | EPI_ISL_10019254 | 2021-12-30 | Directorate Of Public Health And Preventive Medicine | Centre for DNA Fingerprinting and Diagnostics | Asmita Gupta, Rupin Shelake, Arunkumar Karunanidhi, Vinay Donipadi, Divya Vashisht, Ashwin Dalal, Selvavinayagam Sivaprakasam, Darez Ahamed, Sampath Palani, Raju Sivadoss, Avudaiselvi Rathinasamy, Gurunathan Subramanian, Rajesh Kumar Manivannan, Devi Moni |
| hCoV-19/USA/CA-CDPH-500041685/2021 | EPI_ISL_10019912 | 2021-12-22 | TPMG Regional Laboratory | California Department of Public Health | CDPH COVIDNet and UCSC Genomics Institute |

| | | | | | |
|---|---|---|---|---|---|
| hCoV-19/Congo/RC-356/2021 | EPI_ISL_10023502 | 2021-08-12 | Molecular Lab, National Laboratory of Public Health | Pathogen Genomics Lab, National Institute for Biomedical Research (INRB) | Placide Mbala-Kingebeni, Frank Edgard ZONGO Ragomzingba, Edith Nkwembe, Eddy Kinganda-Lusamaki, Amuri Aziza, Francisca Muyembe Mawete, Emmanuel Lokilo Lofiko, Gradi Luakanda, Jean Claude Makangara, Raphael Lumembe, Gabriel Kabamba, Catherine Pratt, Matthi |
| hCoV-19/Congo/RC-383/2021 | EPI_ISL_10023512 | 2021-11-25 | Molecular Lab, National Laboratory of Public Health | Pathogen Genomics Lab, National Institute for Biomedical Research (INRB) | Placide Mbala-Kingebeni, Frank Edgard ZONGO Ragomzingba, Edith Nkwembe, Eddy Kinganda-Lusamaki, Amuri Aziza, Francisca Muyembe Mawete, Emmanuel Lokilo Lofiko, Gradi Luakanda, Jean Claude Makangara, Raphael Lumembe, Gabriel Kabamba, Catherine Pratt, Matthi |
| hCoV-19/Congo/RC-395/2021 | EPI_ISL_10023517 | 2021-03-12 | Molecular Lab, National Laboratory of Public Health | Pathogen Genomics Lab, National Institute for Biomedical Research (INRB) | Placide Mbala-Kingebeni, Frank Edgard ZONGO Ragomzingba, Edith Nkwembe, Eddy Kinganda-Lusamaki, Amuri Aziza, Francisca Muyembe Mawete, Emmanuel Lokilo Lofiko, Gradi Luakanda, Jean Claude Makangara, Raphael Lumembe, Gabriel Kabamba, Catherine Pratt, Matthi |
| hCoV-19/Congo/RC-396/2021 | EPI_ISL_10023518 | 2021-06-12 | Molecular Lab, National Laboratory of Public Health | Pathogen Genomics Lab, National Institute for Biomedical Research (INRB) | Placide Mbala-Kingebeni, Frank Edgard ZONGO Ragomzingba, Edith Nkwembe, Eddy Kinganda-Lusamaki, Amuri Aziza, Francisca Muyembe Mawete, Emmanuel Lokilo Lofiko, Gradi Luakanda, Jean Claude Makangara, Raphael Lumembe, Gabriel Kabamba, Catherine Pratt, Matthi |
| hCoV-19/Congo/RC-405/2021 | EPI_ISL_10023523 | 2021-08-12 | Molecular Lab, National Laboratory of Public Health | Pathogen Genomics Lab, National Institute for Biomedical Research (INRB) | Placide Mbala-Kingebeni, Frank Edgard ZONGO Ragomzingba, Edith Nkwembe, Eddy Kinganda-Lusamaki, Amuri Aziza, Francisca Muyembe Mawete, Emmanuel Lokilo Lofiko, Gradi Luakanda, Jean Claude Makangara, Raphael Lumembe, Gabriel Kabamba, Catherine Pratt, Matthi |
| hCoV-19/Congo/RC-409/2021 | EPI_ISL_10023526 | 2021-08-12 | Molecular Lab, National Laboratory of Public Health | Pathogen Genomics Lab, National Institute for Biomedical Research (INRB) | Placide Mbala-Kingebeni, Frank Edgard ZONGO Ragomzingba, Edith Nkwembe, Eddy Kinganda-Lusamaki, Amuri Aziza, Francisca Muyembe Mawete, Emmanuel Lokilo Lofiko, Gradi Luakanda, Jean Claude Makangara, Raphael Lumembe, Gabriel Kabamba, Catherine Pratt, Matthi |
| hCoV-19/Congo/RC-412/2021 | EPI_ISL_10023527 | 2021-10-12 | Molecular Lab, National Laboratory of Public Health | Pathogen Genomics Lab, National Institute for Biomedical Research (INRB) | Placide Mbala-Kingebeni, Frank Edgard ZONGO Ragomzingba, Edith Nkwembe, Eddy Kinganda-Lusamaki, Amuri Aziza, Francisca Muyembe Mawete, Emmanuel Lokilo Lofiko, Gradi Luakanda, Jean Claude Makangara, Raphael Lumembe, Gabriel Kabamba, Catherine Pratt, Matthi |

| | | | | | |
|---|---|---|---|---|---|
| hCoV-19/Congo/RC-413/2021 | EPI_ISL_10023528 | 2021-08-12 | Molecular Lab, National Laboratory of Public Health | Pathogen Genomics Lab, National Institute for Biomedical Research (INRB) | Placide Mbala-Kingebeni, Frank Edgard ZONGO Ragomzingba, Edith Nkwembe, Eddy Kinganda-Lusamaki, Amuri Aziza, Francisca Muyembe Mawete, Emmanuel Lokilo Lofiko, Gradi Luakanda, Jean Claude Makangara, Raphael Lumembe, Gabriel Kabamba, Catherine Pratt, Matthi |
| hCoV-19/Congo/RC-415/2021 | EPI_ISL_10023530 | 2021-09-12 | Molecular Lab, National Laboratory of Public Health | Pathogen Genomics Lab, National Institute for Biomedical Research (INRB) | Placide Mbala-Kingebeni, Frank Edgard ZONGO Ragomzingba, Edith Nkwembe, Eddy Kinganda-Lusamaki, Amuri Aziza, Francisca Muyembe Mawete, Emmanuel Lokilo Lofiko, Gradi Luakanda, Jean Claude Makangara, Raphael Lumembe, Gabriel Kabamba, Catherine Pratt, Matthi |
| hCoV-19/India/TG-CDFD-2684/2021 | EPI_ISL_10070476 | 2021-12-23 | Gandhi Hospital | Centre for DNA Fingerprinting and Diagnostics | Asmita Gupta, Rupin Shelake, Arunkumar Karunanidhi, Vinay Donipadi, Divya Vashisht, Ashwin Dalal, Nagamani Kammili, Murali Bashyam |
| hCoV-19/India/TG-CDFD-10662/2021 | EPI_ISL_10070510 | 2021-12-30 | Gandhi Hospital | Centre for DNA Fingerprinting and Diagnostics | Asmita Gupta, Rupin Shelake, Arunkumar Karunanidhi, Vinay Donipadi, Divya Vashisht, Ashwin Dalal, Nagamani Kammili, Murali Bashyam |
| hCoV-19/Australia/NT-RDH-57/2021 | EPI_ISL_10093182 | 2021-11-26 | Territory Pathology | Territory Pathology | Dimitrios Menouhos |
| hCoV-19/Zambia/CHAZ-CHS1452/2021 | EPI_ISL_10103896 | 2021-12-23 | CHAZ Laboratory | CHAZ Laboratory | Shempela. D, Sikalima. J, Sandala. D |
| hCoV-19/Rwanda/NRLNAT4339/2021 | EPI_ISL_10114891 | 2021-12-17 | Nucleic Acid Testing, National Reference Laboratory | GIGA Medical Genomics | Yvan Butera, Keith Durkin, Maria Artesi, Nadine Cambisano, Robert Rutayisire, Reuben Sindayiheba, Patrick Tuyisenge, Esperence Umumararungu, S√©bastien Bontems, Marie-Pierre Hayette, Nathalie Renotte, Jacob Souopgui, Claude Mao Muvunyi, Vincent Bours, L√© |
| hCoV-19/Coted'Ivoire/IPCI-cv482/2021 | EPI_ISL_10115878 | 2021-12-06 | Department of Epidemic Viruses, Institut Pasteur of C√¥te d'Ivoire, Abidjan, C√¥te d'Ivoire | Molecular Genetics Platform, Institut Pasteur of C√¥te d'Ivoire, Abidjan, C√¥te d'Ivoire | Aristide Berenger Ako AKO; Bernadette Tchepe Flore Diplo; Egomli Stanislas ASSOHOUN; Antoine Marie-David N'golo COULIBALY; Tata Gnire Safiatou COULIBALY; Jean-Ren√© Quassi ACQUAH; Djeda Franck Arnaud GNAHORE; Herv√© Alberic ADJE KADJO; Mireille DOSSO |
| hCoV-19/USA/CT-Yale-16105/2021 | EPI_ISL_10115914 | 2021-12-21 | Yale Clinical Virology Lab | Grubaugh Lab - Yale School of Public Health | Chantal Vogels, Mallery Breban, Kendall Billig, Tobias Koch, Kien Pham, Rebecca Earnest, Anne Hahn, Chaney Kalinich, Irina Tikhonova, Christopher Castaldi, Bony De Kumar, Nicholas Kerantzas, David Ferguson, Wade Schulz, David Peaper, Marie L. Landry, Nath |
| hCoV-19/India/TN-CDFD-CO-1196/2021 | EPI_ISL_10123101 | 2021-12-30 | Directorate Of Public Health And Preventive Medicine | CDFD | Asmita Gupta, Rupin Shelake, Arunkumar Karunanidhi, Vinay Donipadi, Divya Vashisht, Ashwin Dalal, Selvavinayagam Sivaprakasam, Darez Ahamed, Sampath Palani, Raju Sivadoss, Avudaiselvi Rathinasamy, Gurunathan Subramanian, Rajesh Kumar Manivannan, Devi Moni |

| | | | | | |
|---|---|---|---|---|---|
| hCoV-19/India/TN-CDFD-CO-1489/2021 | EPI_ISL_10123171 | 2021-12-30 | Directorate Of Public Health And Preventive Medicine | CDFD | Asmita Gupta, Rupin Shelake, Arunkumar Karunanidhi, Vinay Donipadi, Divya Vashisht, Ashwin Dalal, Selvavinayagam Sivaprakasam, Darez Ahamed, Sampath Palani, Raju Sivadoss, Avudaiselvi Rathinasamy, Gurunathan Subramanian, Rajesh Kumar Manivannan, Devi Moni |
| hCoV-19/NorthernIreland/NIRE-01847c/2021 | EPI_ISL_10175506 | 2021-12-26 | Regional Virus Laboratory, Belfast Health and Social Care Trust; and: Genomics Core Technology Unit, Queen's University Belfast. | COVID-19 Genomics UK (COG-UK) Consortium | [Regional Virus Laboratory, BHSCT]: Conall McCaughey, James McKenna, Tanya Curran, Susan Feeney, Alison Watt, Ciara Cox, Mairead Connor, Zoltan Molnar, David Simpson, Derek Fairley; [Genomics Core Technology Unit, QUB]: Marc Fuchs, Clara Radulescu, Miao T |
| hCoV-19/India/TN-CDFD-CO-1418/2021 | EPI_ISL_10199666 | 2021-12-30 | Directorate Of Public Health And Preventive Medicine | Centre for DNA Fingerprinting and Diagnostics | Asmita Gupta, Rupin Shelake, Arunkumar Karunanidhi, Vinay Donipadi, Divya Vashisht, Ashwin Dalal, Selvavinayagam Sivaprakasam, Darez Ahamed, Sampath Palani, Raju Sivadoss, Avudaiselvi Rathinasamy, Gurunathan Subramanian, Rajesh Kumar Manivannan, Devi Moni |
| hCoV-19/SouthAfrica/NICD-N30092/2021 | EPI_ISL_10210176 | 2021-12-22 | PORT ELIZABETH LABORATORY | National Institute for Communicable Diseases of the National Health Laboratory Service | Amoako DG, Everatt J, Kekana D, Scheepers C, Mohale T, Ntuli N, Mahlangu B, Mnguni A, Ismail A, Bhiman JN, Wolter N |
| hCoV-19/SouthAfrica/NICD-N32794/2021 | EPI_ISL_10210338 | 2021-12-02 | AMPATH | National Institute for Communicable Diseases of the National Health Laboratory Service | Amoako DG, Everatt J, Kekana D, Scheepers C, Mohale T, Ntuli N, Mahlangu B, Mnguni A, Ismail A, Bhiman JN, Wolter N |
| hCoV-19/SouthAfrica/NICD-N33131/2021 | EPI_ISL_10210357 | 2021-12-07 | AMPATH | National Institute for Communicable Diseases of the National Health Laboratory Service | Amoako DG, Everatt J, Kekana D, Scheepers C, Mohale T, Ntuli N, Mahlangu B, Mnguni A, Ismail A, Bhiman JN, Wolter N |
| hCoV-19/SouthAfrica/NICD-N33063/2021 | EPI_ISL_10210385 | 2021-12-13 | AMPATH | National Institute for Communicable Diseases of the National Health Laboratory Service | Amoako DG, Everatt J, Kekana D, Scheepers C, Mohale T, Ntuli N, Mahlangu B, Mnguni A, Ismail A, Bhiman JN, Wolter N |
| hCoV-19/SouthAfrica/NICD-N33088/2021 | EPI_ISL_10210393 | 2021-12-21 | AMPATH | National Institute for Communicable Diseases of the National Health Laboratory Service | Amoako DG, Everatt J, Kekana D, Scheepers C, Mohale T, Ntuli N, Mahlangu B, Mnguni A, Ismail A, Bhiman JN, Wolter N |
| hCoV-19/SouthAfrica/NICD-N32812/2021 | EPI_ISL_10210405 | 2021-12-30 | AMPATH | National Institute for Communicable Diseases of the National Health Laboratory Service | Amoako DG, Everatt J, Kekana D, Scheepers C, Mohale T, Ntuli N, Mahlangu B, Mnguni A, Ismail A, Bhiman JN, Wolter N |
| hCoV-19/SouthAfrica/NICD-N29404/2021 | EPI_ISL_10210463 | 2021-12-20 | ROB FERREIRA LABORATORY | National Institute for Communicable Diseases of the National Health Laboratory Service | Amoako DG, Everatt J, Kekana D, Scheepers C, Mohale T, Ntuli N, Mahlangu B, Mnguni A, Ismail A, Bhiman JN, Wolter N |
| hCoV-19/SouthAfrica/NICD-N29423/2021 | EPI_ISL_10210481 | 2021-12-24 | ROB FERREIRA LABORATORY | National Institute for Communicable Diseases of the National Health Laboratory Service | Amoako DG, Everatt J, Kekana D, Scheepers C, Mohale T, Ntuli N, Mahlangu B, Mnguni A, Ismail A, Bhiman JN, Wolter N |
| hCoV-19/USA/VA-DCLS-12944/2021 | EPI_ISL_10218981 | 2021-12-27 | Virginia Division of Consolidated Laboratory Services | Virginia Division of Consolidated Laboratory Services | Virginia Division of Consolidated Laboratory Services |
| hCoV-19/Zambia/CHAZ-CHS0834/2021 | EPI_ISL_10222451 | 2021-12-21 | CHAZ Laboratory | CHAZ Laboratory | Shempela. D, Sikalima. J, Sandala. D, Muyombo. A, Chipango. C, CHAZ Lab Staff |

| | | | | | |
|---|---|---|---|---|---|
| hCoV-19/Italy/LAZ-AMC-2112272558-DS/2021 | EPI_ISL_10228672 | 2021-12-27 | Virology Laboratory, Scientific Department, Army Medical Center | Virology Laboratory, Scientific Department, Army Medical Center | Silvia Fillo, Riccardo De Sanctis, Antonella Fortunato, Anella Monte, Rossella Brandi, Giulia Campoli, Marzia Cavalli, Anna Anselmo, Vanessa Vera Fain, Francesco Giordani, Giandomenico Cerreto, Filippo Molinari, Giancarlo Petralito, Florigio Lista |
| hCoV-19/India/DL-IGIB-0708401031025/2021 | EPI_ISL_10250693 | 2021-12-28 | INSACOG at CSIR Institute of Genomics and Integrative Biology | INSACOG at CSIR Institute of Genomics and Integrative Biology | INSACOG |
| hCoV-19/India/UN-IGIB-SRF_0708303345480/2021 | EPI_ISL_10250761 | 2021-12-29 | INSACOG at CSIR Institute of Genomics and Integrative Biology | INSACOG at CSIR Institute of Genomics and Integrative Biology | INSACOG |
| hCoV-19/India/UN-IGIB-SRF_0708303345491/2021 | EPI_ISL_10250792 | 2021-12-29 | INSACOG at CSIR Institute of Genomics and Integrative Biology | INSACOG at CSIR Institute of Genomics and Integrative Biology | INSACOG |
| hCoV-19/India/UN-IGIB-0708401034580/2021 | EPI_ISL_10250802 | 2021-12-29 | INSACOG at CSIR Institute of Genomics and Integrative Biology | INSACOG at CSIR Institute of Genomics and Integrative Biology | INSACOG |
| hCoV-19/India/DL-IGIB-SRF_0708303330288/2021 | EPI_ISL_10250851 | 2021-12-28 | INSACOG at CSIR Institute of Genomics and Integrative Biology | INSACOG at CSIR Institute of Genomics and Integrative Biology | INSACOG |
| hCoV-19/India/DL-IGIB-SRF_0708303331745/2021 | EPI_ISL_10250868 | 2021-12-28 | INSACOG at CSIR Institute of Genomics and Integrative Biology | INSACOG at CSIR Institute of Genomics and Integrative Biology | INSACOG |
| hCoV-19/India/DL-IGIB-SRF_0708303335565/2021 | EPI_ISL_10250896 | 2021-12-28 | INSACOG at CSIR Institute of Genomics and Integrative Biology | INSACOG at CSIR Institute of Genomics and Integrative Biology | INSACOG |
| hCoV-19/Australia/QLD5660/2021 | EPI_ISL_10262601 | 2021-12-25 | Pathology Queensland and Forensic Scientific Services | PHV-FSS | Chenwei Wang on behalf of Q-PHIRE Genomics |
| hCoV-19/USA/WY-WYPHL-22023456/2021 | EPI_ISL_10327857 | 2021-11-29 | Wyoming Public Health Laboratory | Wyoming Public Health Laboratory | Jim Mildenberger, Taylor Fearing, Channing Weber, Ashley Norberg, Chayse Rowley, Brian Dominguez, Elliot Thomasson, Emma Snow, Noah Cheshier, Cyrus Rautman, Carina Williams, Cameron Ward, Daniel Bolding, Robert Petit, Rob Christensen, and Joseph Reed |
| hCoV-19/Ghana/WACCBIP-GS2061/2021 | EPI_ISL_10334334 | 2021-12-09 | WACCBIP, University of Ghana | WACCBIP, University of Ghana | Collins M. Morang'a, Joyce M. Ngoi, Dominic S. Y. Amuzu, Vincent Appiah, Evelyn B. Quansah, Philip M. Soglo, Violette V. M'cormack, Samirah Saiid, Bright K. Yemi, Israel Osei-Wusu, Deborah N. A. Mettle, Aaron A. Manu, Paul Owusu-Oduro, Frederick Tei-Maya |
| hCoV-19/Ghana/WACCBIP-GS2386/2021 | EPI_ISL_10334462 | 2021-12-15 | WACCBIP, University of Ghana | WACCBIP, University of Ghana | Collins M. Morang'a, Peter K. Quashie, Joyce M. Ngoi, Dominic S. Y. Amuzu, Vincent Appiah, Evelyn B. Quansah, Philip M. Soglo, Violette Mcormack, Samirah Saiid, Frederick Tei-Maya, Edward Danso Fenteng, Patrick Tetteh Ababio, Theophilus Odoom, Emmanuel Ku |
| hCoV-19/France/GES-CERBAHC-12263014/2021 | EPI_ISL_10347112 | 2021-12-27 | B2A-LABORATOIRE DU BASSIN POTASSIQUE | CERBA HealthCare | B√©n√©dicte Roquebert, Laura Verdurme, Mathilde Roussel, Sabine Trombert, St√©phanie Haim-Boukobza |
| hCoV-19/England/DHSC-CYNX3MF/2021 | EPI_ISL_10385512 | 2021-11-30 | Oncologica UK Ltd | COVID-19 Genomics UK (COG-UK) Consortium | UKHSA; Oncologica UK Ltd |
| hCoV-19/England/DHSC-CYYSOMB/2021 | EPI_ISL_10393204 | 2021-12-20 | Oncologica UK Ltd | COVID-19 Genomics UK (COG-UK) Consortium | UKHSA; Oncologica UK Ltd |
| hCoV-19/SouthAfrica/1063959/2021 | EPI_ISL_10408472 | 2021-11-30 | NdlovuLaboratories | Ndlovu Laboratories | A. Mabaso, A. Mdunyelwa, K. Mlambo, M. Mathapelo, C. Umunnakwe |

| | | | | | |
|---|---|---|---|---|---|
| hCoV-19/Nigeria/NCDC-NR-GL-001274/2021 | EPI_ISL_10430162 | 2021-12-13 | MEDBURY MEDICAL SERVICES | National Reference Laboratory, Nigeria Centre for Disease Control | Dr Ndodo Nnaemeka, Olusola Anuoluwapo Akanbi, Oyeronke Ayansola, Chimaobi Chukwu, Richard Olulowo Ojedele, Victor Oripenaye, Khadijah Isa Imam, Bruno Nwokeocha, Kenneth Chukwuemeka Ikeata, Michael Mbata, Adelakun Sophiyah Sulaimon, Brownson Ekerete Akpan, |
| hCoV-19/Ghana/WACCBIP-GS2115/2021 | EPI_ISL_10438308 | 2021-12-20 | WACCBIP, University of Ghana | WACCBIP, University of Ghana | Collins M. Morang'a, Peter K. Quashie, Joyce M. Ngoi, Dominic S. Y. Amuzu, Vincent Appiah, Evelyn B. Quansah, Philip M. Soglo, Violette Mcormack, Samirah Saiid, Frederick Tei-Maya, Edward Danso Fenteng, Patrick Tetteh Ababio, Theophilus Odoom, Emmanuel Ku |
| hCoV-19/India/TN-CDFD-E130078/2021 | EPI_ISL_10504840 | 2021-11-25 | Christian Medical College | Centre for DNA Fingerprinting and Diagnostics | Asmita Gupta, Rupin Shelake, Arunkumar Karunanidhi, Vinay Donipadi, KPP Abhilash, Binila Chacko, Priya G, Ramya I, Mahesh Moorthy, Nitya Wadhwa, Shirali Raina Labroo, Bhabatosh Das, Divya Vashisht, Ashwin Dalal, Pramod Kumar Garg, Murali Bashyam |
| hCoV-19/India/TN-CDFD-E130084/2021 | EPI_ISL_10504847 | 2021-11-29 | Christian Medical College | Centre for DNA Fingerprinting and Diagnostics | Asmita Gupta, Rupin Shelake, Arunkumar Karunanidhi, Vinay Donipadi, KPP Abhilash, Binila Chacko, Priya G, Ramya I, Mahesh Moorthy, Nitya Wadhwa, Shirali Raina Labroo, Bhabatosh Das, Divya Vashisht, Ashwin Dalal, Pramod Kumar Garg, Murali Bashyam |
| hCoV-19/SouthSudan/UG1181/2021 | EPI_ISL_10512587 | 2021-12-14 | South Sudan Ministry of Health, WHO South Sudan, MRC/UVRI & LSHTM Uganda Research Unit | MRC/UVRI & LSHTM Uganda Research Unit, South Sudan Ministry of Health, WHO South Sudan | Abe G Abias, James Ayei, Lul Lojok Deng, Richard Lino Loro Lako, John Rumunu, Gregory Wani, Joseph Francis Wamala, Juma John HM, Sacha Bootsma, Dennis Kenyi Lodiongo, Sudhir Bunga, Dan Lule Bugembe, Matthew Cotten, My V.T. Phan |
| hCoV-19/Australia/VIC42276/2021 | EPI_ISL_10565618 | 2021-12-29 | Microbiological Diagnostic Unit - Public Health Laboratory (MDU-PHL), The Peter Doherty Institute for Infection and Immunity | Public Health Laboratory (MDU-PHL), The Peter Doherty Institute for Infection and Immunity | Seemann, T., Horan, K., Sherry, N.L. |
| hCoV-19/Australia/VIC43232/2021 | EPI_ISL_10565689 | 2021-12-29 | Microbiological Diagnostic Unit - Public Health Laboratory (MDU-PHL), The Peter Doherty Institute for Infection and Immunity | Public Health Laboratory (MDU-PHL), The Peter Doherty Institute for Infection and Immunity | Seemann, T., Horan, K., Sherry, N.L. |
| hCoV-19/Australia/VIC43248/2021 | EPI_ISL_10565703 | 2021-12-27 | Microbiological Diagnostic Unit - Public Health Laboratory (MDU-PHL), The Peter Doherty Institute for Infection and Immunity | Public Health Laboratory (MDU-PHL), The Peter Doherty Institute for Infection and Immunity | Seemann, T., Horan, K., Sherry, N.L. |
| hCoV-19/Australia/VIC43249/2021 | EPI_ISL_10565704 | 2021-12-27 | Microbiological Diagnostic Unit - Public Health Laboratory (MDU-PHL), The Peter Doherty Institute for Infection and Immunity | Public Health Laboratory (MDU-PHL), The Peter Doherty Institute for Infection and Immunity | Seemann, T., Horan, K., Sherry, N.L. |
| hCoV-19/Australia/VIC43253/2021 | EPI_ISL_10565707 | 2021-12-30 | Microbiological Diagnostic Unit - Public Health Laboratory (MDU-PHL), The Peter Doherty Institute for Infection and Immunity | Public Health Laboratory (MDU-PHL), The Peter Doherty Institute for Infection and Immunity | Seemann, T., Horan, K., Sherry, N.L. |
| hCoV-19/Australia/VIC42321/2021 | EPI_ISL_10565711 | 2021-12-27 | Microbiological Diagnostic Unit - Public Health Laboratory (MDU-PHL), The Peter Doherty Institute for Infection and Immunity | Public Health Laboratory (MDU-PHL), The Peter Doherty Institute for Infection and Immunity | Seemann, T., Horan, K., Sherry, N.L. |

| | | | | | |
|---|---|---|---|---|---|
| hCoV-19/Australia/VIC43465/2021 | EPI_ISL_10565917 | 2021-12-29 | Microbiological Diagnostic Unit - Public Health Laboratory (MDU-PHL), The Peter Doherty Institute for Infection and Immunity | Public Health Laboratory (MDU-PHL), The Peter Doherty Institute for Infection and Immunity | Seemann, T., Horan, K., Sherry, N.L. |
| hCoV-19/USA/VA-DCLS-13117/2021 | EPI_ISL_10592712 | 2021-12-28 | Virginia Division of Consolidated Laboratory Services | Virginia Division of Consolidated Laboratory Services | Virginia Division of Consolidated Laboratory Services |
| hCoV-19/Colombia/VAC-CSB-1153464349/2021 | EPI_ISL_10594761 | 2021-11-22 | Sebastian de Belalcazar | Universidad del Valle | Diana L√≥pez-Alvarez, Nelson Rivera Franco,Andres Castillo, David Valencia, Melissa Solarte, Erica M. Aristizabal, Flor Saa, Beatriz Parra & Programa Nacional de Caracterizaci√≥n Gen√≥mica de SARS-CoV-2 |
| hCoV-19/Singapore/9842/2021 | EPI_ISL_8450003 | 2021-12-30 | National Public Health Laboratory, National Centre for Infectious Diseases | National Centre for Infectious Diseases | Grace Ngan, Benny Yeo, Zhenyang Zhou, Royce Ang, Samuel Loo, Lin Cui, Raymond Tzer Pin Lin |
| hCoV-19/India/TG-GMCH-ICMR-INSACOG-K4/2021 | EPI_ISL_8452483 | 2021-12-22 | Gandhi Medical College and Hospital (GMCH), Secunderabad | NIV Influenza | Dr.K.Nagamani, Dr.Sunitha Pakalapaty, Dr.G.Sushma Rajya Lakshmi, D.R.Manisha Rani |
| hCoV-19/England/LSPA-3059272/2021 | EPI_ISL_8452942 | 2021-12-27 | Rosalind Franklin Laboratory | Wellcome Sanger Institute for the COVID-19 Genomics UK (COG-UK) Consortium | Donald Fraser, Suki Lee, Rob Howes, The Rosalind Franklin Laboratory and Alex Alderton, Roberto Amato, Jeffrey Barrett, Sonia Goncalves, Ewan Harrison, David K. Jackson, Ian Johnston, Dominic Kwiatkowski, Cordelia Langford, John Sillitoe on behalf of the |
| hCoV-19/England/MILK-3047F9E/2021 | EPI_ISL_8455236 | 2021-12-29 | Lighthouse Lab in Milton Keynes | Wellcome Sanger Institute for the COVID-19 Genomics UK (COG-UK) Consortium | The Lighthouse Lab in Milton Keynes and Alex Alderton, Roberto Amato, Jeffrey Barrett, Sonia Goncalves, Ewan Harrison, David K. Jackson, Ian Johnston, Dominic Kwiatkowski, Cordelia Langford, John Sillitoe on behalf of the Wellcome Sanger Institute COVID-1 |
| hCoV-19/England/MILK-303CCA4/2021 | EPI_ISL_8455997 | 2021-12-28 | Lighthouse Lab in Milton Keynes | Wellcome Sanger Institute for the COVID-19 Genomics UK (COG-UK) Consortium | The Lighthouse Lab in Milton Keynes and Alex Alderton, Roberto Amato, Jeffrey Barrett, Sonia Goncalves, Ewan Harrison, David K. Jackson, Ian Johnston, Dominic Kwiatkowski, Cordelia Langford, John Sillitoe on behalf of the Wellcome Sanger Institute COVID-1 |
| hCoV-19/England/MILK-2FF6008/2021 | EPI_ISL_8459188 | 2021-12-26 | Lighthouse Lab in Milton Keynes | Wellcome Sanger Institute for the COVID-19 Genomics UK (COG-UK) Consortium | The Lighthouse Lab in Milton Keynes and Alex Alderton, Roberto Amato, Jeffrey Barrett, Sonia Goncalves, Ewan Harrison, David K. Jackson, Ian Johnston, Dominic Kwiatkowski, Cordelia Langford, John Sillitoe on behalf of the Wellcome Sanger Institute COVID-1 |
| hCoV-19/Denmark/DCGC-298245/2021 | EPI_ISL_8459664 | 2021-12-30 | Department of Bacteria, Parasites and Fungi, Statens Serum Institut, Copenhagen, Denmark | Statens Serum Institut Bioinformatics and Microbial Genomics | Danish Covid-19 Genome Consortium |
| hCoV-19/England/ALDP-2FF0FD4/2021 | EPI_ISL_8459698 | 2021-12-18 | Lighthouse Lab in Alderley Park | Wellcome Sanger Institute for the COVID-19 Genomics UK (COG-UK) Consortium | Jacquelyn Wynn, Mairead Hyland, The Lighthouse Lab in Alderley Park and Alex Alderton, Roberto Amato, Jeffrey Barrett, Sonia Goncalves, Ewan Harrison, David K. Jackson, Ian Johnston, Dominic Kwiatkowski, Cordelia Langford, John Sillitoe on behalf of the W |
| hCoV-19/Denmark/DCGC-298280/2021 | EPI_ISL_8459737 | 2021-12-27 | Department of Bacteria, Parasites and Fungi, Statens Serum Institut, Copenhagen, Denmark | Statens Serum Institut Bioinformatics and Microbial Genomics | Danish Covid-19 Genome Consortium |

| | | | | | |
|---|---|---|---|---|---|
| hCoV-19/Netherlands/NH-RIVM-79723/2021 | EPI_ISL_8461235 | 2021-11-26 | Dutch COVID-19 response team | National Institute for Public Health and the Environment (RIVM) | Adam Meijer, Harry Vennema, Dirk Eggink, Jeroen Cremer, Sharon van den Brink, Bas van der Veer, AnneMarie van den Brandt, Lisa Wijsman, Kim Freriks, Ryanne Jaarsma, Lynn Aarts, Sanne Bos, Jil Kocken, Jordy de Bakker, Stijn van Rossum, Linda van Someren, F |
| hCoV-19/Netherlands/NH-RIVM-79728/2021 | EPI_ISL_8461295 | 2021-11-26 | Dutch COVID-19 response team | National Institute for Public Health and the Environment (RIVM) | Adam Meijer, Harry Vennema, Dirk Eggink, Jeroen Cremer, Sharon van den Brink, Bas van der Veer, AnneMarie van den Brandt, Lisa Wijsman, Kim Freriks, Ryanne Jaarsma, Lynn Aarts, Sanne Bos, Jil Kocken, Jordy de Bakker, Stijn van Rossum, Linda van Someren, F |
| hCoV-19/Netherlands/LB-RIVM-78789/2021 | EPI_ISL_8461609 | 2021-12-24 | Dutch COVID-19 response team | National Institute for Public Health and the Environment (RIVM) | Adam Meijer, Harry Vennema, Dirk Eggink, Jeroen Cremer, Sharon van den Brink, Bas van der Veer, AnneMarie van den Brandt, Lisa Wijsman, Kim Freriks, Ryanne Jaarsma, Lynn Aarts, Sanne Bos, Jil Kocken, Jordy de Bakker, Stijn van Rossum, Linda van Someren, F |
| hCoV-19/Netherlands/GL-RIVM-79159/2021 | EPI_ISL_8461807 | 2021-11-28 | Dutch COVID-19 response team | National Institute for Public Health and the Environment (RIVM) | Adam Meijer, Harry Vennema, Dirk Eggink, Jeroen Cremer, Sharon van den Brink, Bas van der Veer, AnneMarie van den Brandt, Lisa Wijsman, Kim Freriks, Ryanne Jaarsma, Lynn Aarts, Sanne Bos, Jil Kocken, Jordy de Bakker, Stijn van Rossum, Linda van Someren, F |
| hCoV-19/Denmark/DCGC-298689/2021 | EPI_ISL_8462279 | 2021-12-30 | Department of Bacteria, Parasites and Fungi, Statens Serum Institut, Copenhagen, Denmark | Statens Serum Institut Bioinformatics and Microbial Genomics | Danish Covid-19 Genome Consortium |
| hCoV-19/England/ALDP-2FE46A1/2021 | EPI_ISL_8462429 | 2021-12-19 | Lighthouse Lab in Alderley Park | Wellcome Sanger Institute for the COVID-19 Genomics UK (COG-UK) Consortium | Jacquelyn Wynn, Mairead Hyland, The Lighthouse Lab in Alderley Park and Alex Alderton, Roberto Amato, Jeffrey Barrett, Sonia Goncalves, Ewan Harrison, David K. Jackson, Ian Johnston, Dominic Kwiatkowski, Cordelia Langford, John Sillitoe on behalf of the W |
| hCoV-19/Denmark/DCGC-298971/2021 | EPI_ISL_8462653 | 2021-12-22 | Department of Clinical Microbiology and Center for Genomic Medicine, Rigshospitalet, Copenhagen, Denmark | Statens Serum Institut Bioinformatics and Microbial Genomics | Danish Covid-19 Genome Consortium |
| hCoV-19/Denmark/DCGC-299651/2021 | EPI_ISL_8463119 | 2021-12-29 | Department of Bacteria, Parasites and Fungi, Statens Serum Institut, Copenhagen, Denmark | Statens Serum Institut Bioinformatics and Microbial Genomics | Danish Covid-19 Genome Consortium |
| hCoV-19/Denmark/DCGC-299780/2021 | EPI_ISL_8463230 | 2021-12-30 | Department of Bacteria, Parasites and Fungi, Statens Serum Institut, Copenhagen, Denmark | Statens Serum Institut Bioinformatics and Microbial Genomics | Danish Covid-19 Genome Consortium |
| hCoV-19/Israel/ICH-741109440/2021 | EPI_ISL_8463571 | 2021-12-23 | Clinical Microbiology Laboratory, Tel Aviv Sourasky Medical Center | Clinical Microbiology Laboratory, Tel Aviv Sourasky Medical Center | Katya Levytskyi, Ora Halutz, Alon Ziv, Lior Handler, Goel Morad, Amos Adler |
| hCoV-19/Israel/ICH-741109469/2021 | EPI_ISL_8463640 | 2021-12-24 | Clinical Microbiology Laboratory, Tel Aviv Sourasky Medical Center | Clinical Microbiology Laboratory, Tel Aviv Sourasky Medical Center | Katya Levytskyi, Ora Halutz, Alon Ziv, Lior Handler, Goel Morad, Amos Adler |
| hCoV-19/SouthAfrica/NICD-R18375/2021 | EPI_ISL_8465009 | 2021-12-08 | Edendale Gateway Clinic | National Institute for Communicable Diseases of the National Health Laboratory Service | Amoako DG, Everatt J, Kekana D, Scheepers C, Mohale T, Ntuli N, Mahlangu B, Mnguni A, Ismail A, Bhiman JN, Wolter N |

| | | | | | |
|---|---|---|---|---|---|
| hCoV-19/SouthAfrica/NICD-R18399/2021 | EPI_ISL_8465013 | 2021-12-10 | Helen Joseph Hospital | National Institute for Communicable Diseases of the National Health Laboratory Service | Amoako DG, Everatt J, Kekana D, Scheepers C, Mohale T, Ntuli N, Mahlangu B, Mnguni A, Ismail A, Bhiman JN, Wolter N |
| hCoV-19/Switzerland/GE-HUG-36576659/2021 | EPI_ISL_8466230 | 2021-12-29 | University Hospitals of Geneva, Laboratory of Virology | HUG, Laboratory of Virology and the Health2030 Genome Center | Samuel Cordey, Aline Mamin, Francisco Perez, Ana Rita Goncalves, Laurent Kaiser, Lorenzo Cerutti, Cedric Howald, Henri Pegeot, Melyssa Elies, Deborah Penet, Keith Harshman, Ioannis Xenarios |
| hCoV-19/Brazil/PE-FIOCRUZ-IAM6354/2021 | EPI_ISL_8466874 | 2021-12-22 | LACEN/PE | WallauLab on behalf of Fiocruz COVID-19 Genomic Surveillance Network | Marcelo Henrique dos Santos Paiva, Cassia Docena, Matheus Filgueira Bezerra, Filipe Zimmer Dezordi, Antonio Marinho da Silva Neto, Lais Ceschini Machado, Lilian Caroliny Amorim Silva, Gustavo Barbosa de Lima, Alexandre Freitas da Silva, Sinval Pinto Brand |
| hCoV-19/USA/CA-CDC-FG-223413/2021 | EPI_ISL_8467061 | 2021-12-26 | Fulgent Genetics | Centers for Disease Control and Prevention Division of Viral Diseases, Pathogen Discovery | Dakota Howard,Dhwani Batra,Peter Cook,Jason Caravas,Benjamin Rambo-Martin,Scott Sammons,Yvette Unoarumhi,Matthew Schmerer,Kristine Lacek,Tymeckia Kendall,Victoria Caban Figueroa,Shatavia Morrison,Christopher Gulvick,Erisa Sula,Harry Gao,Mickey Li,John Gao |
| hCoV-19/USA/CA-CDC-FG-224578/2021 | EPI_ISL_8467678 | 2021-12-27 | Fulgent Genetics | Centers for Disease Control and Prevention Division of Viral Diseases, Pathogen Discovery | Dakota Howard,Dhwani Batra,Peter Cook,Jason Caravas,Benjamin Rambo-Martin,Scott Sammons,Yvette Unoarumhi,Matthew Schmerer,Kristine Lacek,Tymeckia Kendall,Victoria Caban Figueroa,Shatavia Morrison,Christopher Gulvick,Erisa Sula,Harry Gao,Mickey Li,John Gao |
| hCoV-19/USA/GA-CDC-ASC210568771/2021 | EPI_ISL_8470696 | 2021-12-20 | Aegis Sciences Corporation | Centers for Disease Control and Prevention Division of Viral Diseases, Pathogen Discovery | Dakota Howard,Dhwani Batra,Peter Cook,Jason Caravas,Benjamin Rambo-Martin,Scott Sammons,Yvette Unoarumhi,Matthew Schmerer,Kristine Lacek,Tymeckia Kendall,Victoria Caban Figueroa,Shatavia Morrison,Christopher Gulvick,Erisa Sula,Cyndi Clark,Patrick Campbell |
| hCoV-19/USA/AZ-TG1131535/2021 | EPI_ISL_8472821 | 2021-11-28 | TGen North | TGen North | Jolene Bowers, Heather Centner, Brett Van Tassel, Chris French, Hayley Yaglom, Darrin Lemmer, Dave Engelthaler |
| hCoV-19/USA/CA-CDC-FG-205819/2021 | EPI_ISL_8473407 | 2021-12-20 | Fulgent Genetics | Centers for Disease Control and Prevention Division of Viral Diseases, Pathogen Discovery | Dakota Howard,Dhwani Batra,Peter Cook,Jason Caravas,Benjamin Rambo-Martin,Scott Sammons,Yvette Unoarumhi,Matthew Schmerer,Kristine Lacek,Tymeckia Kendall,Victoria Caban Figueroa,Shatavia Morrison,Christopher Gulvick,Erisa Sula,Harry Gao,Mickey Li,John Gao |
| hCoV-19/USA/NY-CDC-FG-204092/2021 | EPI_ISL_8474619 | 2021-12-20 | Fulgent Genetics | Centers for Disease Control and Prevention Division of Viral Diseases, Pathogen Discovery | Dakota Howard,Dhwani Batra,Peter Cook,Jason Caravas,Benjamin Rambo-Martin,Scott Sammons,Yvette Unoarumhi,Matthew Schmerer,Kristine Lacek,Tymeckia Kendall,Victoria Caban Figueroa,Shatavia Morrison,Christopher Gulvick,Erisa Sula,Harry Gao,Mickey Li,John Gao |
| hCoV-19/Italy/EMR-AUSLRomagna-C001-22-39/2021 | EPI_ISL_8475468 | 2021-12-30 | U.O. Microbiologia Laboratorio Unico Centro Servizi - AUSL della Romagna | U.O. Microbiologia, Laboratorio Unico Centro Servizi - AUSL della Romagna | Giorgio Dirani |

| | | | | | |
|---|---|---|---|---|---|
| hCoV-19/USA/CA-HLX-ATM-8HFPZUC7M/2021 | EPI_ISL_8476965 | 2021-12-16 | Kaiser Permanente Southern California | Helix | Kaiser Permanente Southern California, Helix |
| hCoV-19/USA/MN-UMGC-26592/2021 | EPI_ISL_8477858 | 2021-12-06 | Public Health Laboratory, Minnesota Department of Health | University of Minnesota Genomics Center | Daryl M. Gohl, John Garbe, Jaquelyn Kuriger-Laber, Corbin Dirkx |
| hCoV-19/USA/MN-UMGC-26607/2021 | EPI_ISL_8477869 | 2021-12-08 | Public Health Laboratory, Minnesota Department of Health | University of Minnesota Genomics Center | Daryl M. Gohl, John Garbe, Jaquelyn Kuriger-Laber, Corbin Dirkx |
| hCoV-19/USA/MN-UMGC-27262/2021 | EPI_ISL_8478059 | 2021-12-01 | Public Health Laboratory, Minnesota Department of Health | University of Minnesota Genomics Center | Daryl M. Gohl, John Garbe, Jaquelyn Kuriger-Laber, Corbin Dirkx |
| hCoV-19/USA/CA-HLX-ATM-PACMJNTGR/2021 | EPI_ISL_8478561 | 2021-12-19 | Kaiser Permanente Southern California | Helix | Kaiser Permanente Southern California, Helix |
| hCoV-19/USA/LA-BIE-LSUH002491/2021 | EPI_ISL_8479557 | 2021-12-23 | Ochsner Health | BioInfoExperts | Amy Feehan, Judy Crabtree, Susanna L. Lamers, Rebecca Rose, Ben Lain, Chris Huston, Samual Moot, David J. Nolan, Tessa LaFleur, Julia-Garcia-Diaz, Lucio Miele |
| hCoV-19/USA/GA-GPHL-2882/2021 | EPI_ISL_8481150 | 2021-12-27 | GA Department of Public Health | GA Department of Public Health | Stacy Reeves, Sharmila Talekar, Jonathan Edwards, Tonia Parrott, Aliyah Fields, Taylor Smith |
| hCoV-19/USA/MA-CDCBI-CRSP_S5O27KZLGVW5LDSC/2021 | EPI_ISL_8484483 | 2021-12-29 | Broad Institute Clinical Research Sequencing Platform | Infectious Disease Program, Broad Institute of Harvard and MIT | Siddle,K.J., Adams,G., Pearlman,L., Gladden-Young,A., Vicente,G., Blumenstiel,B., DeFelice,M., Lee,M., McGovern,S., Lagerborg,K., Rudy,M., DeRuff,K., Carter,A., Normandin,E., Bauer,M., Reilly,S., Tomkins-Tinch,C., Loreth,C., Chaluvadi,S., Meldrim,J., Gran |
| hCoV-19/England/PHEC-YYBSI8X/2021 | EPI_ISL_8488167 | 2021-12-22 | Respiratory Virus Unit, Microbiology Services Colindale, Public Health England | COVID-19 Genomics UK (COG-UK) Consortium | PHE Covid Sequencing Team |
| hCoV-19/Mauritius/387764/2021 | EPI_ISL_8494988 | 2021-12-22 | Central Health Laboratory | Central Health Laboratory | Manraj SS, Sonoo J, Bahadoor BS, Mathur H, Sujeewon C, Jannoo N, Ramuth M,Ubheeram j,Bacorisen A |
| hCoV-19/Mauritius/388291/2021 | EPI_ISL_8494995 | 2021-12-23 | Central Health Laboratory | Central Health Laboratory | Manraj SS, Sonoo J, Bahadoor BS, Mathur H, Sujeewon C, Jannoo N, Ramuth M,Ubheeram j,Bacorisen A |
| hCoV-19/India/TG-RFCH02971_CIB5076/2021 | EPI_ISL_8499882 | 2021-10-29 | Apollo Hospital | CSIR-Centre for Cellular and Molecular Biology | Tulasi Nagabandi, Lamuk Zaveri,Jandhyala Sai Krishna,Malini Nemalikanti,Sreelekshmi MS, Ara Sreenivas,Shreekant Verma, Amareshwar Vodapalli,B Himasri,Valli Nagalakshmi Undamatla, Sumedha Avadhanula,Priya Nurkurthy,Vidhyadhari Methuku, M V Sasidhar, Rajes |
| hCoV-19/India/TG-RFCH03123_CIB8763/2021 | EPI_ISL_8500028 | 2021-04-09 | MapMyGenome | CSIR-Centre for Cellular and Molecular Biology | Lamuk Zaveri,Tulasi Nagabandi,Jandhyala Sai Krishna,Malini Nemalikanti,Sreelekshmi MS, Ara Sreenivas,Shreekant Verma, Amareshwar Vodapalli,B Himasri,Valli Nagalakshmi Undamatla, Sumedha Avadhanula,Priya Nurkurthy,Vidhyadhari Methuku,M. Aravind Kumar, San |
| hCoV-19/India/MH-NEERI-CIB9377/2021 | EPI_ISL_8500128 | 2021-12-08 | CSIR-National Environmental Engineering Research Institute | CSIR-Centre for Cellular and Molecular Biology-INSACOG | Lamuk Zaveri,Tulasi Nagabandi, Sreelekshmi MS,Malini Nemalikanti,Ara Sreenivas,Shreekant Verma, Amareshwar Vodapalli ,B Himasri,Valli Nagalakshmi Undamatla, Sumedha Avadhanula, Vidhyadhari Methuku, Priya Nurkurthy, Archana Bharadwaj Siva, Karthik Bharadw |

| | | | | | |
|---|---|---|---|---|---|
| hCoV-19/India/AP-CCMB-CIB9329/2021 | EPI_ISL_8500214 | 2021-12-12 | AP SSO | CSIR-Centre for Cellular and Molecular Biology-INSACOG | Sumedha Avadhanula,Valli Nagalakshmi Undamatla,Amareshwar Vodapalli, B Himasri,Priya Nurkurthy,Vidhyadhari Methuku,Shreekant Verma,Ara Sreenivas, Sreelekshmi MS,Malini Nemalikanti,Lamuk Zaveri,Tulasi Nagabandi,Jandhyala Sai Krishna, Archana Bharadwaj Siv |
| hCoV-19/India/AP-CCMB-CIB9330/2021 | EPI_ISL_8500215 | 2021-12-12 | AP SSO | CSIR-Centre for Cellular and Molecular Biology-INSACOG | Priya Nurkurthy,Vidhyadhari Methuku,Sumedha Avadhanula,Valli Nagalakshmi Undamatla,Amareshwar Vodapalli, B Himasri,Shreekant Verma,Ara Sreenivas, Sreelekshmi MS,Malini Nemalikanti,Lamuk Zaveri,Tulasi Nagabandi,Jandhyala Sai Krishna, Archana Bharadwaj Siv |
| hCoV-19/Wales/MILK-30815B4/2021 | EPI_ISL_8501577 | 2021-12-30 | Lighthouse Lab in Milton Keynes | Wellcome Sanger Institute for the COVID-19 Genomics UK (COG-UK) Consortium | The Lighthouse Lab in Milton Keynes and Alex Alderton, Roberto Amato, Jeffrey Barrett, Sonia Goncalves, Ewan Harrison, David K. Jackson, Ian Johnston, Dominic Kwiatkowski, Cordelia Langford, John Sillitoe on behalf of the Wellcome Sanger Institute COVID-1 |
| hCoV-19/Brazil/SP-IAL-8361/2021 | EPI_ISL_8501814 | 2021-12-21 | Instituto Adolfo Lutz - Regional de Rio Claro | Instituto Adolfo Lutz, Interdiciplinary Procedures Center, Strategic Laboratory | Claudio Tavares Sacchi, Claudia Regina Gon√ßalves, Karoline Rodrigues Campos, Caio Vinicius Dias Lopes, Marlon Benedito Nascimento Santos, Ariadne Ferreira Amarante |
| hCoV-19/Brazil/SP-IAL-8376/2021 | EPI_ISL_8501816 | 2021-12-17 | Instituto Adolfo Lutz - Regional de Sorocaba | Instituto Adolfo Lutz, Interdiciplinary Procedures Center, Strategic Laboratory | Claudio Tavares Sacchi, Claudia Regina Gon√ßalves, Karoline Rodrigues Campos, Caio Vinicius Dias Lopes, Marlon Benedito Nascimento Santos, Ariadne Ferreira Amarante |
| hCoV-19/USA/UT-UPHL-220107006738/2021 | EPI_ISL_8502811 | 2021-12-29 | Utah Public Health Laboratory | Utah Public Health Laboratory | Erin L. Young, Kelly F. Oakeson, Pooja Gupta, John Arnn, Tom Iverson, Olinto Linares-Perdomo |
| hCoV-19/USA/CA-CDPH-3000039512/2021 | EPI_ISL_8506005 | 2021-05-21 | CDPH VBL | California Department of Public Health | CDPH-COVIDNet |
| hCoV-19/Colombia/BOL-UNIMOL-VG-236/2021 | EPI_ISL_8508576 | 2021-12-28 | UNIMOL | UNIMOL | Steev Loyola, Mashiel Fern√°ndez-Ruiz, Jaison Torres-Pacheco, Doris G√≥mez-Camargo |
| hCoV-19/USA/NM-UNM-ED00872/2021 | EPI_ISL_8508931 | 2021-12-28 | University of New Mexico Hospital | Center for Global Health, University of New Mexico Health Sciences Center | Daryl Domman, Kurt Schwalm, Valerie Morley, Jesse Young, Justin Bacca, Jon Femling, Darrell Dinwiddie |
| hCoV-19/USA/MA-NEIDL-02783/2021 | EPI_ISL_8509575 | 2021-12-21 | Boston University CTL | Boston University/National Emerging Infectious Disease Laboratories | Jacquelyn Turcinovic, John H. Connor, Lena Landeverde, Catherine Klapperich, Lynn Doucette-Stamm |
| hCoV-19/Ireland/LH-NVRL-S21IRL00568953/2021 | EPI_ISL_8509931 | 2021-12-14 | National Virus Reference Laboratory | National Virus Reference Laboratory | Zoe Yandle, Charlene Bennett, Gabriel Gonzalez, Michael Carr, Jonathan Dean, Cillian F De Gascun |
| hCoV-19/Colombia/ANT-CWOHC-VG-SEC02337W/2021 | EPI_ISL_8514935 | 2021-10-05 | SYNLAB | Universidad Nacional de Colombia - Laboratorio Gen√≥mico One Health | Andres F. Cardona-Rios, Maria Stella L√≥pez, Daniel O. Maldonado-Perez, Sim√≥n Villegas Vel√°squez, Lina M. Hurtado, Laura Silvana Perez, Karl A Ciuoderis, Maria Ang√©lica Maya, Carolina Mu√±oz-Arango, Celeny Ortiz, Rita Almanza Payares, Idabely Betancur |
| hCoV-19/USA/CO-CDPHE-2102514119/2021 | EPI_ISL_8515536 | 2021-12-24 | Colorado Department of Public Health and Environment | Colorado Department of Public Health and Environment | Laura Bankers, Molly C. Hetherington-Rauth, Diana Ir, Alexandria Rossheim, Michael Martin, Mandy Waters, Arianna Smith, Shannon R. Matzinger, Emily A. Travanty |
| hCoV-19/England/PHEP-YYN6UBC/2021 | EPI_ISL_8517763 | 2021-12-21 | Respiratory Virus Unit, Microbiology Services Colindale, Public Health England | COVID-19 Genomics UK (COG-UK) Consortium | PHE Covid Sequencing Team |

| | | | | | |
|---|---|---|---|---|---|
| hCoV-19/England/MILK-30DBF08/2021 | EPI_ISL_8521882 | 2021-12-30 | Lighthouse Lab in Milton Keynes | Wellcome Sanger Institute for the COVID-19 Genomics UK (COG-UK) Consortium | The Lighthouse Lab in Milton Keynes and Alex Alderton, Roberto Amato, Jeffrey Barrett, Sonia Goncalves, Ewan Harrison, David K. Jackson, Ian Johnston, Dominic Kwiatkowski, Cordelia Langford, John Sillitoe on behalf of the Wellcome Sanger Institute COVID-1 |
| hCoV-19/CaboVerde/Santiago_32484_2/2021 | EPI_ISL_8523954 | 2021-07-19 | Virology Laboratory of Praia | Institut Pasteur de Dakar | Sanders Kevin, Diagne Moussa Mo√Øse, Diallo Amadou, Dia Ndongo, Sankhe Safietou, Ndiaye Ndack, Loucoubar Cheikh, Faye Ousmane, da Luz Lima Mendon√ßa Maria, Sall Amadou Alpha |
| hCoV-19/CaboVerde/Santiago-33168/2021 | EPI_ISL_8524323 | 2021-08-02 | Virology Laboratory of Praia | Institut Pasteur de Dakar | Sanders Kevin, Diagne Moussa Mo√Øse, Diallo Amadou, Dia Ndongo, Sankhe Safietou, Diop Mamadou, Ndiaye Ndack, Loucoubar Cheikh, Faye Ousmane, da Luz Lima Mendon√ßa Maria, Sall Amadou Alpha |
| hCoV-19/Maldives/MAV94016/2021 | EPI_ISL_8525200 | 2021-12-18 | Indira Gandhi Memorial Hospital | Indira Gandhi Memorial Hospital | D. Fathmath Nazla Rafeeq, Dr. Ibrahim Afzal, Ms. Aishath Shuhudha, Ms. Aminath Shazleena Abdul Rahman, Ms. Soafy Mohamed, Mr. Ibrahim Nishan Ahmed |
| hCoV-19/Senegal/24729/2020 | EPI_ISL_8525551 | 2020-05-09 | Institut Pasteur de Dakar | Institut Pasteur de Dakar | Diallo Amadou, Diagne Moussa Moise, Dia Ndongo, Sankhe Safietou, Ndiaye Ndack, Loucoubar Cheikh, Faye Ousmane, Sall Amadou Alpha |
| hCoV-19/Senegal/24961/2020 | EPI_ISL_8525574 | 2020-05-09 | Institut Pasteur de Dakar | Institut Pasteur de Dakar | Diallo Amadou, Diagne Moussa Moise, Dia Ndongo, Sankhe Safietou, Ndiaye Ndack, Loucoubar Cheikh, Faye Ousmane, Sall Amadou Alpha |
| hCoV-19/Senegal/25388/2020 | EPI_ISL_8525583 | 2020-05-09 | Institut Pasteur de Dakar | Institut Pasteur de Dakar | Diallo Amadou, Diagne Moussa Moise, Dia Ndongo, Sankhe Safietou, Ndiaye Ndack, Loucoubar Cheikh, Faye Ousmane, Sall Amadou Alpha |
| hCoV-19/Senegal/25616/2020 | EPI_ISL_8525591 | 2020-05-10 | Institut Pasteur de Dakar | Institut Pasteur de Dakar | Diallo Amadou, Diagne Moussa Moise, Dia Ndongo, Sankhe Safietou, Ndiaye Ndack, Loucoubar Cheikh, Faye Ousmane, Sall Amadou Alpha |
| hCoV-19/Senegal/25620/2020 | EPI_ISL_8525592 | 2020-05-10 | Institut Pasteur de Dakar | Institut Pasteur de Dakar | Diallo Amadou, Diagne Moussa Moise, Dia Ndongo, Sankhe Safietou, Ndiaye Ndack, Loucoubar Cheikh, Faye Ousmane, Sall Amadou Alpha |
| hCoV-19/Senegal/107888/2020 | EPI_ISL_8525870 | 2020-09-22 | Institut Pasteur de Dakar | Institut Pasteur de Dakar | Diallo Amadou, Diagne Moussa Moise, Dia Ndongo, Sankhe Safietou, Ndiaye Ndack, Loucoubar Cheikh, Faye Ousmane, Sall Amadou Alpha |
| hCoV-19/Senegal/1839194/2020 | EPI_ISL_8525944 | 2020-12-08 | Institut Pasteur de Dakar | Institut Pasteur de Dakar | Diallo Amadou, Diagne Moussa Moise, Dia Ndongo, Sankhe Safietou, Ndiaye Ndack, Loucoubar Cheikh, Faye Ousmane, Sall Amadou Alpha |
| hCoV-19/Senegal/196688/2021 | EPI_ISL_8526126 | 2021-06-25 | Institut Pasteur de Dakar | Institut Pasteur de Dakar | Diallo Amadou, Diagne Moussa Moise,¬† Dia Ndongo, Sankhe Safietou, Ndiaye Ndack, Loucoubar Cheikh, Faye Ousmane, Sall Amadou Alpha |
| hCoV-19/Senegal/10004662/2021 | EPI_ISL_8526133 | 2021-06-26 | Institut Pasteur de Dakar | Institut Pasteur de Dakar | Diallo Amadou, Diagne Moussa Moise,¬† Dia Ndongo, Sankhe Safietou, Ndiaye Ndack, Loucoubar Cheikh, Faye Ousmane, Sall Amadou Alpha |

| | | | | | |
|---|---|---|---|---|---|
| hCoV-19/Senegal/10009156/2021 | EPI_ISL_8526174 | 2021-06-28 | Institut Pasteur de Dakar | Institut Pasteur de Dakar | Diallo Amadou, Diagne Moussa Moise,¬† Dia Ndongo, Sankhe Safietou, Ndiaye Ndack, Loucoubar Cheikh, Faye Ousmane, Sall Amadou Alpha |
| hCoV-19/Senegal/196724/2021 | EPI_ISL_8526247 | 2021-06-28 | Institut Pasteur de Dakar | Institut Pasteur de Dakar | Diallo Amadou, Diagne Moussa Moise,¬† Dia Ndongo, Sankhe Safietou, Ndiaye Ndack, Loucoubar Cheikh, Faye Ousmane, Sall Amadou Alpha |
| hCoV-19/Senegal/11014681/2021 | EPI_ISL_8526303 | 2021-07-03 | Institut Pasteur de Dakar | Institut Pasteur de Dakar | Diallo Amadou, Diagne Moussa Moise,¬† Dia Ndongo, Sankhe Safietou, Ndiaye Ndack, Loucoubar Cheikh, Faye Ousmane, Sall Amadou Alpha |
| hCoV-19/Senegal/11046843/2021 | EPI_ISL_8526406 | 2021-07-05 | Institut Pasteur de Dakar | Institut Pasteur de Dakar | Diallo Amadou, Diagne Moussa Moise,¬† Dia Ndongo, Sankhe Safietou, Ndiaye Ndack, Loucoubar Cheikh, Faye Ousmane, Sall Amadou Alpha |
| hCoV-19/Senegal/196998/2021 | EPI_ISL_8526620 | 2021-07-16 | Institut Pasteur de Dakar | Institut Pasteur de Dakar | Diallo Amadou, Diagne Moussa Moise,¬† Dia Ndongo, Sankhe Safietou, Ndiaye Ndack, Loucoubar Cheikh, Faye Ousmane, Sall Amadou Alpha |
| hCoV-19/Senegal/244009/2021 | EPI_ISL_8526670 | 2021-07-23 | Institut Pasteur de Dakar | Institut Pasteur de Dakar | Diallo Amadou, Diagne Moussa Moise,¬† Dia Ndongo, Sankhe Safietou, Ndiaye Ndack, Loucoubar Cheikh, Faye Ousmane, Sall Amadou Alpha |
| hCoV-19/Senegal/246127/2021 | EPI_ISL_8526702 | 2021-07-28 | Institut Pasteur de Dakar | Institut Pasteur de Dakar | Diallo Amadou, Diagne Moussa Moise,¬† Dia Ndongo, Sankhe Safietou, Ndiaye Ndack, Loucoubar Cheikh, Faye Ousmane, Sall Amadou Alpha |
| hCoV-19/Senegal/246382/2021 | EPI_ISL_8526707 | 2021-08-28 | Institut Pasteur de Dakar | Institut Pasteur de Dakar | Diallo Amadou, Diagne Moussa Moise,¬† Dia Ndongo, Sankhe Safietou, Ndiaye Ndack, Loucoubar Cheikh, Faye Ousmane, Sall Amadou Alpha |
| hCoV-19/Senegal/246525/2021 | EPI_ISL_8526709 | 2021-07-27 | Institut Pasteur de Dakar | Institut Pasteur de Dakar | Diallo Amadou, Diagne Moussa Moise,¬† Dia Ndongo, Sankhe Safietou, Ndiaye Ndack, Loucoubar Cheikh, Faye Ousmane, Sall Amadou Alpha |
| hCoV-19/Senegal/246964/2021 | EPI_ISL_8526716 | 2021-07-29 | Institut Pasteur de Dakar | Institut Pasteur de Dakar | Diallo Amadou, Diagne Moussa Moise,¬† Dia Ndongo, Sankhe Safietou, Ndiaye Ndack, Loucoubar Cheikh, Faye Ousmane, Sall Amadou Alpha |
| hCoV-19/Senegal/284938/2021 | EPI_ISL_8526791 | 2021-08-19 | Institut Pasteur de Dakar | Institut Pasteur de Dakar | Diallo Amadou, Diagne Moussa Moise,¬† Dia Ndongo, Sankhe Safietou, Ndiaye Ndack, Loucoubar Cheikh, Faye Ousmane, Sall Amadou Alpha |
| hCoV-19/Senegal/285224/2021 | EPI_ISL_8526841 | 2021-08-23 | Institut Pasteur de Dakar | Institut Pasteur de Dakar | Diallo Amadou, Diagne Moussa Moise,¬† Dia Ndongo, Sankhe Safietou, Ndiaye Ndack, Loucoubar Cheikh, Faye Ousmane, Sall Amadou Alpha |
| hCoV-19/Senegal/285231/2021 | EPI_ISL_8526844 | 2021-08-25 | Institut Pasteur de Dakar | Institut Pasteur de Dakar | Diallo Amadou, Diagne Moussa Moise,¬† Dia Ndongo, Sankhe Safietou, Ndiaye Ndack, Loucoubar Cheikh, Faye Ousmane, Sall Amadou Alpha |
| hCoV-19/Senegal/285249/2021 | EPI_ISL_8526848 | 2021-08-27 | Institut Pasteur de Dakar | Institut Pasteur de Dakar | Diallo Amadou, Diagne Moussa Moise,¬† Dia Ndongo, Sankhe Safietou, Ndiaye Ndack, Loucoubar Cheikh, Faye Ousmane, Sall Amadou Alpha |
| hCoV-19/Senegal/285512/2021 | EPI_ISL_8526875 | 2021-09-01 | Institut Pasteur de Dakar | Institut Pasteur de Dakar | Diallo Amadou, Diagne Moussa Moise,¬† Dia Ndongo, Sankhe Safietou, Ndiaye Ndack, Loucoubar Cheikh, Faye Ousmane, Sall Amadou Alpha |

| | | | | | |
|---|---|---|---|---|---|
| hCoV-19/Senegal/285517/2021 | EPI_ISL_8526877 | 2021-09-02 | Institut Pasteur de Dakar | Institut Pasteur de Dakar | Diallo Amadou, Diagne Moussa Moise,¬† Dia Ndongo, Sankhe Safietou, Ndiaye Ndack, Loucoubar Cheikh, Faye Ousmane, Sall Amadou Alpha |
| hCoV-19/Senegal/285700/2021 | EPI_ISL_8526893 | 2021-09-02 | Institut Pasteur de Dakar | Institut Pasteur de Dakar | Diallo Amadou, Diagne Moussa Moise,¬† Dia Ndongo, Sankhe Safietou, Ndiaye Ndack, Loucoubar Cheikh, Faye Ousmane, Sall Amadou Alpha |
| hCoV-19/Senegal/285761/2021 | EPI_ISL_8526900 | 2021-09-04 | Institut Pasteur de Dakar | Institut Pasteur de Dakar | Diallo Amadou, Diagne Moussa Moise,¬† Dia Ndongo, Sankhe Safietou, Ndiaye Ndack, Loucoubar Cheikh, Faye Ousmane, Sall Amadou Alpha |
| hCoV-19/Senegal/260976/2021 | EPI_ISL_8526989 | 2021-08-09 | Institut Pasteur de Dakar | Institut Pasteur de Dakar | Diallo Amadou, Diagne Moussa Moise,¬† Dia Ndongo, Sankhe Safietou, Ndiaye Ndack, Loucoubar Cheikh, Faye Ousmane, Sall Amadou Alpha |
| hCoV-19/Senegal/259703/2021 | EPI_ISL_8527155 | 2021-07-25 | Institut Pasteur de Dakar | Institut Pasteur de Dakar | Diallo Amadou, Diagne Moussa Moise,¬† Dia Ndongo, Sankhe Safietou, Ndiaye Ndack, Loucoubar Cheikh, Faye Ousmane, Sall Amadou Alpha |
| hCoV-19/Senegal/135036/2021 | EPI_ISL_8527797 | 2021-11-12 | Institut Pasteur de Dakar | Institut Pasteur de Dakar | Diallo Amadou, Diagne Moussa Moise,¬† Dia Ndongo, Sankhe Safietou, Ndiaye Ndack, Loucoubar Cheikh, Faye Ousmane, Sall Amadou Alpha |
| hCoV-19/Senegal/135686/2021 | EPI_ISL_8527798 | 2021-01-09 | Institut Pasteur de Dakar | Institut Pasteur de Dakar | Diallo Amadou, Diagne Moussa Moise,¬† Dia Ndongo, Sankhe Safietou, Ndiaye Ndack, Loucoubar Cheikh, Faye Ousmane, Sall Amadou Alpha |
| hCoV-19/Senegal/4042305/2021 | EPI_ISL_8527807 | 2021-02-26 | Institut Pasteur de Dakar | Institut Pasteur de Dakar | Diallo Amadou, Diagne Moussa Moise,¬† Dia Ndongo, Sankhe Safietou, Ndiaye Ndack, Loucoubar Cheikh, Faye Ousmane, Sall Amadou Alpha |
| hCoV-19/Senegal/173072/2021 | EPI_ISL_8527853 | 2021-02-10 | Institut Pasteur de Dakar | Institut Pasteur de Dakar | Diallo Amadou, Diagne Moussa Moise,¬† Dia Ndongo, Sankhe Safietou, Ndiaye Ndack, Loucoubar Cheikh, Faye Ousmane, Sall Amadou Alpha |
| hCoV-19/Senegal/135911/2021 | EPI_ISL_8527885 | 2021-01-28 | Institut Pasteur de Dakar | Institut Pasteur de Dakar | Diallo Amadou, Diagne Moussa Moise,¬† Dia Ndongo, Sankhe Safietou, Ndiaye Ndack, Loucoubar Cheikh, Faye Ousmane, Sall Amadou Alpha |
| hCoV-19/Senegal/135928/2021 | EPI_ISL_8527910 | 2021-02-02 | Institut Pasteur de Dakar | Institut Pasteur de Dakar | Diallo Amadou, Diagne Moussa Moise,¬† Dia Ndongo, Sankhe Safietou, Ndiaye Ndack, Loucoubar Cheikh, Faye Ousmane, Sall Amadou Alpha |
| hCoV-19/Senegal/202083/2021 | EPI_ISL_8527946 | 2021-03-26 | Institut Pasteur de Dakar | Institut Pasteur de Dakar | Diallo Amadou, Diagne Moussa Moise,¬† Dia Ndongo, Sankhe Safietou, Ndiaye Ndack, Loucoubar Cheikh, Faye Ousmane, Sall Amadou Alpha |
| hCoV-19/Senegal/6998372/2021 | EPI_ISL_8528215 | 2021-05-01 | Institut Pasteur de Dakar | Institut Pasteur de Dakar | Diallo Amadou, Diagne Moussa Moise,¬† Dia Ndongo, Sankhe Safietou, Ndiaye Ndack, Loucoubar Cheikh, Faye Ousmane, Sall Amadou Alpha |
| hCoV-19/Senegal/6998658/2021 | EPI_ISL_8528219 | 2021-05-01 | Institut Pasteur de Dakar | Institut Pasteur de Dakar | Diallo Amadou, Diagne Moussa Moise,¬† Dia Ndongo, Sankhe Safietou, Ndiaye Ndack, Loucoubar Cheikh, Faye Ousmane, Sall Amadou Alpha |
| hCoV-19/Senegal/223855/2021 | EPI_ISL_8528221 | 2021-04-30 | Institut Pasteur de Dakar | Institut Pasteur de Dakar | Diallo Amadou, Diagne Moussa Moise,¬† Dia Ndongo, Sankhe Safietou, Ndiaye Ndack, Loucoubar Cheikh, Faye Ousmane, Sall Amadou Alpha |

| | | | | | |
|---|---|---|---|---|---|
| hCoV-19/Senegal/223915/2021 | EPI_ISL_8528225 | 2021-05-01 | Institut Pasteur de Dakar | Institut Pasteur de Dakar | Diallo Amadou, Diagne Moussa Moise,¬† Dia Ndongo, Sankhe Safietou, Ndiaye Ndack, Loucoubar Cheikh, Faye Ousmane, Sall Amadou Alpha |
| hCoV-19/Senegal/7924051/2021 | EPI_ISL_8528372 | 2021-05-25 | Institut Pasteur de Dakar | Institut Pasteur de Dakar | Diallo Amadou, Diagne Moussa Moise,¬† Dia Ndongo, Sankhe Safietou, Ndiaye Ndack, Loucoubar Cheikh, Faye Ousmane, Sall Amadou Alpha |
| hCoV-19/Senegal/7970308/2021 | EPI_ISL_8528431 | 2021-06-02 | Institut Pasteur de Dakar | Institut Pasteur de Dakar | Diallo Amadou, Diagne Moussa Moise,¬† Dia Ndongo, Sankhe Safietou, Ndiaye Ndack, Loucoubar Cheikh, Faye Ousmane, Sall Amadou Alpha |
| hCoV-19/Senegal/7970362/2021 | EPI_ISL_8528432 | 2021-06-02 | Institut Pasteur de Dakar | Institut Pasteur de Dakar | Diallo Amadou, Diagne Moussa Moise,¬† Dia Ndongo, Sankhe Safietou, Ndiaye Ndack, Loucoubar Cheikh, Faye Ousmane, Sall Amadou Alpha |
| hCoV-19/Senegal/9930726/2021 | EPI_ISL_8528589 | 2021-06-19 | Institut Pasteur de Dakar | Institut Pasteur de Dakar | Diallo Amadou, Diagne Moussa Moise,¬† Dia Ndongo, Sankhe Safietou, Ndiaye Ndack, Loucoubar Cheikh, Faye Ousmane, Sall Amadou Alpha |
| hCoV-19/Denmark/DCGC-300370/2021 | EPI_ISL_8529046 | 2021-12-28 | Department of Bacteria, Parasites and Fungi, Statens Serum Institut, Copenhagen, Denmark | Statens Serum Institut Bioinformatics and Microbial Genomics | Danish Covid-19 Genome Consortium |
| hCoV-19/Denmark/DCGC-300440/2021 | EPI_ISL_8529093 | 2021-12-28 | Department of Bacteria, Parasites and Fungi, Statens Serum Institut, Copenhagen, Denmark | Statens Serum Institut Bioinformatics and Microbial Genomics | Danish Covid-19 Genome Consortium |
| hCoV-19/Denmark/DCGC-301161/2021 | EPI_ISL_8529587 | 2021-12-23 | Department of Bacteria, Parasites and Fungi, Statens Serum Institut, Copenhagen, Denmark | Statens Serum Institut Bioinformatics and Microbial Genomics | Danish Covid-19 Genome Consortium |
| hCoV-19/Sweden/SUS0007685/2021 | EPI_ISL_8538870 | 2021-12-27 | Clinical Microbiology, Infection Prevention and Control | Section for Molecular Diagnostics | Bj√∂rn Hallstr√∂m, Jonas Bj√∂rkman |
| hCoV-19/Sweden/SUS0007718/2021 | EPI_ISL_8538903 | 2021-12-28 | Clinical Microbiology, Infection Prevention and Control | Section for Molecular Diagnostics | Bj√∂rn Hallstr√∂m, Jonas Bj√∂rkman |
| hCoV-19/Sweden/SUS0007804/2021 | EPI_ISL_8538989 | 2021-12-30 | Clinical Microbiology, Infection Prevention and Control | Section for Molecular Diagnostics | Bj√∂rn Hallstr√∂m, Jonas Bj√∂rkman |
| hCoV-19/England/PHEC-YYBAFSH/2021 | EPI_ISL_8539598 | 2021-12-20 | Respiratory Virus Unit, Microbiology Services Colindale, Public Health England | COVID-19 Genomics UK (COG-UK) Consortium | PHE Covid Sequencing Team |
| hCoV-19/Georgia/Tb-sNGS2514/2021 | EPI_ISL_8541184 | 2021-12-28 | JSC Megalab | [illegible] Biology and Genome Research, R. G. Lugar Center for Public Health Research, National Center for Disease Control and Public Health (NCDC) of Georgia. | Giorgi Tomashvili, Giorgi Gogoladze, Meri Pantsulaia, Gvantsa Brachveli, Salome Javashvili, Nino Berishvili, Tata Imnadze, Ana Papkiauri, Gvantsa Chanturia, Ann Machablishvili, Nato Kotaria, Marine Murtskhvaladze, Lela Sabadze, Mari Gavashelidze, Tamar Ja |
| hCoV-19/CzechRepublic/CSQ4569/2020 | EPI_ISL_8541491 | 2020-10-23 | SYNLAB | University Hospital Brno, CMBG | Bezdicek Matej, Svaton Jan, Kristyna Dufkova, Lengerova Martina, Dolejska Monika |
| hCoV-19/Indonesia/JB-NIHRD-MI_21_14762/2021 | EPI_ISL_8543103 | 2021-12-29 | Naraya Medical Center, Bekasi | National Institute of Health Research and Development | Kartika Dewi Puspa, Arie Ardiansyah Nugraha, Fajar Nur Sulistiyohadi, Hana Apsari Pawestri, Subangkit, Hartanti Dian Ikawati, Nelly Puspandari, Vivi Setiawaty |
| hCoV-19/USA/WI-UW-8277/2021 | EPI_ISL_8543583 | 2021-12-30 | University of Wisconsin-Madison AIDS Vaccine Research Laboratories | University of Wisconsin-Madison AIDS Vaccine Research Laboratories | Gage Moreno, Katarina Braun, et al. AIDS Vaccine Research Laboratories |

| | | | | | |
|---|---|---|---|---|---|
| hCoV-19/Italy/UMB-IZSGC-75.1.32/2021 | EPI_ISL_8543916 | 2021-12-28 | Istituto Zooprofilattico Sperimentale Umbria e Marche "Togo Rosati" | Istituto Zooprofilattico Sperimentale dell‚ÄôAbruzzo e Molise ‚ÄúG. Caporale‚Äù | Biagetti M, Giammarioli M, Mencacci A, Camilloni B, Lorusso A, Marcacci M, Di Domenico M, Ancora M, Curini V, Mangone I, Rinaldi A, Delli Compagni E, Scialabba S, Di Pasquale A, Camm√† C, Puglia I, Calistri P, Savini G |
| hCoV-19/USA/NY-PRL-2021_1226_02C11/2021 | EPI_ISL_8545395 | 2021-12-24 | Pandemic Response Lab - NYC | Pandemic Response Lab, R&D | Henry Lee, Michael Hammerling, Melissa Hopkins, Cybill del Castillo, Shinyoung Clair Kang, William Ward, Pradeep Bugga, Sol Rey, Dylan Law, Alex Carpio, Isabel Fernandez Escapa, Haiping Hao, Jon Laurent |
| hCoV-19/Italy/EMR_AUSLRomagna_C003-22-33/2020 | EPI_ISL_8547199 | 2020-12-30 | U.O. Microbiologia Laboratorio Unico Centro Servizi - AUSL della Romagna | U.O. Microbiologia, Laboratorio Unico Centro Servizi - AUSL della Romagna | Giorgio Dirani |
| hCoV-19/Chile/CO-USACH-00243/2021 | EPI_ISL_8547259 | 2021-12-28 | Hospital San Pablo | Unidad de Secuenciaci√≥n y Gen√≥mica, USACH | Carlos A. Villarroel, Mara Cuevas, Kamila Urbina, Yesseny V√°squez-Mart√≠nez, Marcelo Cortez-San Martin, Francisco A. Cubillos |
| hCoV-19/USA/FL-CDC-STM-FVF36RGN3/2021 | EPI_ISL_8548261 | 2021-12-26 | Helix | Centers for Disease Control and Prevention Division of Viral Diseases, Pathogen Discovery | Dakota Howard,Dhwani Batra,Peter Cook,Jason Caravas,Benjamin Rambo-Martin,Scott Sammons,Yvette Unoarumhi,Matthew Schmerer,Kristine Lacek,Tymeckia Kendall,Victoria Caban Figueroa,Helix CA,Shatavia Morrison,Christopher Gulvick,Erisa Sula,Clinton Paden,Dunca |
| hCoV-19/USA/MI-CDC-ASC210640267/2021 | EPI_ISL_8552015 | 2021-12-23 | Aegis Sciences Corporation | Centers for Disease Control and Prevention Division of Viral Diseases, Pathogen Discovery | Dakota Howard,Dhwani Batra,Peter Cook,Jason Caravas,Benjamin Rambo-Martin,Scott Sammons,Yvette Unoarumhi,Matthew Schmerer,Kristine Lacek,Tymeckia Kendall,Victoria Caban Figueroa,Shatavia Morrison,Christopher Gulvick,Erisa Sula,Cyndi Clark,Patrick Campbell |
| hCoV-19/USA/TX-CDC-ASC210640003/2021 | EPI_ISL_8552832 | 2021-12-23 | Aegis Sciences Corporation | Centers for Disease Control and Prevention Division of Viral Diseases, Pathogen Discovery | Dakota Howard,Dhwani Batra,Peter Cook,Jason Caravas,Benjamin Rambo-Martin,Scott Sammons,Yvette Unoarumhi,Matthew Schmerer,Kristine Lacek,Tymeckia Kendall,Victoria Caban Figueroa,Shatavia Morrison,Christopher Gulvick,Erisa Sula,Cyndi Clark,Patrick Campbell |
| hCoV-19/USA/NJ-CDC-ASC210568793/2021 | EPI_ISL_8555279 | 2021-12-27 | Aegis Sciences Corporation | Centers for Disease Control and Prevention Division of Viral Diseases, Pathogen Discovery | Dakota Howard,Dhwani Batra,Peter Cook,Jason Caravas,Benjamin Rambo-Martin,Scott Sammons,Yvette Unoarumhi,Matthew Schmerer,Kristine Lacek,Tymeckia Kendall,Victoria Caban Figueroa,Shatavia Morrison,Christopher Gulvick,Erisa Sula,Cyndi Clark,Patrick Campbell |
| hCoV-19/USA/NJ-CDC-IBX753898011089/2021 | EPI_ISL_8555636 | 2021-12-21 | Infinity Biologix | Centers for Disease Control and Prevention Division of Viral Diseases, Pathogen Discovery | Dakota Howard,Dhwani Batra,Peter Cook,Jason Caravas,Benjamin Rambo-Martin,Scott Sammons,Yvette Unoarumhi,Matthew Schmerer,Kristine Lacek,Tymeckia Kendall,Victoria Caban Figueroa,Shatavia Morrison,Christopher Gulvick,Erisa Sula,Christian Bixby,Yihe Wang,Jo |

| | | | | | |
|---|---|---|---|---|---|
| hCoV-19/Peru/MOQ-INS-11688/2021 | EPI_ISL_8558755 | 2021-12-01 | Laboratorio de Referencia Nacional de Virus Respiratorios. Centro Nacional de Salud Publica. Instituto Nacional de Salud Peru. | Laboratorio de Referencia Nacional de Virus Respiratorios. Centro Nacional de Salud Publica. Instituto Nacional de Salud Peru. | Carlos Padilla Rojas, Veronica Hurtado Vela, Orson Mestanza Millones, Iris Silva Molina, Luis Barcena Flores, Alicia Nu√±ez Llanos, Steve Acedo Lazo, Wendy Lizarraga Olivares, Victor Jimenez Vasquez, Sara Gordillo Vilchez, Kelly Vanessa Izarra Rojas, Edwa |
| hCoV-19/Peru/LIM-INS-11693/2021 | EPI_ISL_8558760 | 2021-12-11 | Laboratorio de Referencia Nacional de Virus Respiratorios. Centro Nacional de Salud Publica. Instituto Nacional de Salud Peru. | Laboratorio de Referencia Nacional de Virus Respiratorios. Centro Nacional de Salud Publica. Instituto Nacional de Salud Peru. | Carlos Padilla Rojas, Veronica Hurtado Vela, Orson Mestanza Millones, Iris Silva Molina, Luis Barcena Flores, Alicia Nu√±ez Llanos, Steve Acedo Lazo, Wendy Lizarraga Olivares, Victor Jimenez Vasquez, Sara Gordillo Vilchez, Kelly Vanessa Izarra Rojas, Edwa |
| hCoV-19/Zimbabwe/QIBZIM-70408/2021 | EPI_ISL_8559208 | 2021-11-27 | National Microbiology Reference Laboratory | Quadram Institute Bioscience | Tapfumanei Mashe, Faustinos T Takawira, Hlanai Gumbo, Kenneth K Maeka, Agnes Juru, Charles Nyagupe, Muchaneta Mugabe, Dave J. Baker, Thanh Le-Viet, Nabil-Fareed Alikhan, Lizzie Meadows, Martin Lott, Michaela Matthews, Alexander J Trotter, ¬†Andrew J. Page |
| hCoV-19/Japan/IC-2561/2021 | EPI_ISL_8559628 | 2021-12-22 | SARS-CoV-2 testing team, National Institute of Infectious Diseases | Pathogen Genomics Center, National Institute of Infectious Diseases | Tsuyoshi Sekizuka, Kentaro Itokawa, Rina Tanaka, Masanori Hashino, Nozomu Hanaoka, Masumichi Saito, Naomi Nojiri, Hazuka Y Furihata, Tsuguto Fujimoto, Makoto Kuroda |
| hCoV-19/Japan/IC-2598/2021 | EPI_ISL_8559666 | 2021-12-24 | SARS-CoV-2 testing team, National Institute of Infectious Diseases | Pathogen Genomics Center, National Institute of Infectious Diseases | Tsuyoshi Sekizuka, Kentaro Itokawa, Rina Tanaka, Masanori Hashino, Nozomu Hanaoka, Masumichi Saito, Naomi Nojiri, Hazuka Y Furihata, Tsuguto Fujimoto, Makoto Kuroda |
| hCoV-19/Japan/IC-2613/2021 | EPI_ISL_8559694 | 2021-12-25 | SARS-CoV-2 testing team, National Institute of Infectious Diseases | Pathogen Genomics Center, National Institute of Infectious Diseases | Tsuyoshi Sekizuka, Kentaro Itokawa, Rina Tanaka, Masanori Hashino, Nozomu Hanaoka, Masumichi Saito, Naomi Nojiri, Hazuka Y Furihata, Tsuguto Fujimoto, Makoto Kuroda |
| hCoV-19/Japan/IC-2616/2021 | EPI_ISL_8559702 | 2021-12-26 | SARS-CoV-2 testing team, National Institute of Infectious Diseases | Pathogen Genomics Center, National Institute of Infectious Diseases | Tsuyoshi Sekizuka, Kentaro Itokawa, Rina Tanaka, Masanori Hashino, Nozomu Hanaoka, Masumichi Saito, Naomi Nojiri, Hazuka Y Furihata, Tsuguto Fujimoto, Makoto Kuroda |
| hCoV-19/Colombia/MAG-CGBM-S30112021-27/2021 | EPI_ISL_8561302 | 2021-10-23 | Sociedad Cl√≠nica Iberoam√©rica | Centro de Gen√©tica y Biolog√≠a Molecular - Universidad del Magdalena | Maria Teresa Mojica-Ortiz, Daniel Bautista, Andrea M. Ram√≠rez Hernandez, Edison Lea-Ch, Angel Oviedo Marquez, Lyda R. Castro |
| hCoV-19/Colombia/TOL-INS-VG-11842/2021 | EPI_ISL_8562437 | 2021-12-27 | Clinica Tolima | Instituto Nacional de Salud-Direcci√≥n de Investigaci√≥n en Salud P√∫blica | Katherine Laiton-Donato, Carlos Franco-Mu√±oz, Diego A. √Ålvarez-D√≠az, H√©ctor Alejandro Ruiz-Moreno, Diego Andr√©s Prada, Jhonnatan Reales-Gonz√°lez, Sheryll Corchuelo, Mar√≠a T. Herrera-Sep√∫lveda, Juli√°n Naizaque, Gerardo Santamar√≠a, Jorge Rivera, P |
| hCoV-19/Colombia/DC-INS-VG-11937/2021 | EPI_ISL_8562465 | 2021-12-22 | LDSP - Cundinamarca | Instituto Nacional de Salud-Direcci√≥n de Investigaci√≥n en Salud P√∫blica | Katherine Laiton-Donato, Carlos Franco-Mu√±oz, Diego A. √Ålvarez-D√≠az, H√©ctor Alejandro Ruiz-Moreno, Diego Andr√©s Prada, Jhonnatan Reales-Gonz√°lez, Sheryll Corchuelo, Mar√≠a T. Herrera-Sep√∫lveda, Juli√°n Naizaque, Gerardo Santamar√≠a, Jorge Rivera, P |

| | | | | | |
|---|---|---|---|---|---|
| hCoV-19/Colombia/VAC-INS-VG-11780/2021 | EPI_ISL_8562484 | 2021-12-22 | Clinica Sebastian de Belarcazar | Instituto Nacional de Salud-Direcci√≥n de Investigaci√≥n en Salud P√∫blica | Katherine Laiton-Donato, Carlos Franco-Mu√±oz, Diego A. √Ålvarez-D√≠az, H√©ctor Alejandro Ruiz-Moreno, Diego Andr√©s Prada, Jhonnatan Reales-Gonz√°lez, Sheryll Corchuelo, Mar√≠a T. Herrera-Sep√∫lveda, Juli√°n Naizaque, Gerardo Santamar√≠a, Jorge Rivera, P |
| hCoV-19/USA/WA-PHL-009134/2021 | EPI_ISL_8564232 | 2021-12-28 | Washington State Department of Health Public Health Laboratories | Washington State Department of Health Public Health Laboratories | Drew MacKellar, Philip Dykema, Denny Russell, Lisa Jones, Holly Halstead, Kathryn Sickles, Kristin Roche, Ardizon Valdez, Claire Howell, Alex Lathum, JohnAric Peterson, Avi Singh, Rebecca Cao |
| hCoV-19/India/DL-ILBS-WGS1109/2021 | EPI_ISL_8565022 | 2021-12-24 | CITY X RAY AND SCAN CLINIC PVT LTD. | ILBS | Ekta Gupta, Varun Suroliya, Pramod Gautam, Reshu Agarwal, Rahul Garg, Urvinder Kaur, Arjun Bhugra, Chhagan Bihari Sharma, Shiv Kumar Sarin |
| hCoV-19/Suriname/SR-904/2021 | EPI_ISL_8572431 | 2021-12-13 | Central Laboratory, Bureau of Public Health (BOG) and Academic Hospital Paramaribo | Erasmus Medical Center | Bas B Oude Munnink, Cherise Beek, Consuella Partowidjojo, Dion Gajadin, Ed PF IJzerman, Emmanuelle Munger, Gary Gummels, Ingrid SK Krishnadath, Lycke Woittiez, Marion PG Koopmans, Mireille Van de Veer, Phyllis Pinas, Princes Wongsowidjojo, Radjesh Ori, Ra |
| hCoV-19/Scotland/QEUH-30FFAB2/2021 | EPI_ISL_8574998 | 2021-12-30 | Lighthouse Lab in Glasgow | Wellcome Sanger Institute for the COVID-19 Genomics UK (COG-UK) Consortium | Harper VanSteenhouse, Yumi Kasai, David Gray, Carol Clugston, Anna Dominiczak and Alex Alderton, Roberto Amato, Jeffrey Barrett, Sonia Goncalves, Ewan Harrison, David K. Jackson, Ian Johnston, Dominic Kwiatkowski, Cordelia Langford, John Sillitoe on behal |
| hCoV-19/Canada/ON-PHL-21-48836/2021 | EPI_ISL_8577725 | 2021-11-30 | Public Health Ontario Laboratory | Public Health Ontario Laboratory | Vanessa G Allen, Philip Banh, Yao Chen, Richard de Borja, Alireza Eshaghi, Nahuel Fittipaldi, Christine Frantz, Jonathan B Gubbay, Jennifer L Guthrie, Lawrence Heisler, Esha Joshi, Michael Laszloffy, Aimin Li, Michael CY Li, Dean Maxwell, Sandeep Nagra, S |
| hCoV-19/SouthAfrica/CERI-KRISP-K033966/2021 | EPI_ISL_8578306 | 2021-12-18 | CAPRISA | and Innvoation, Stellenbosch University and KRISP, KZN Research Innovation and Sequencing Platform, UKZN. | Giandhari J, Pillay S, Naidoo Y, Ramphal U, Ramphal Y, Arisha Maharaj, van Wyk S, Moir M, San JE, Tshiabuila D, Tegally H, Wilkinson E, de Oliveira T |
| hCoV-19/Denmark/DCGC-305410/2021 | EPI_ISL_8582551 | 2021-12-25 | Department of Bacteria, Parasites and Fungi, Statens Serum Institut, Copenhagen, Denmark | Statens Serum Institut Bioinformatics and Microbial Genomics | Danish Covid-19 Genome Consortium |
| hCoV-19/Denmark/DCGC-305397/2021 | EPI_ISL_8583756 | 2021-12-27 | Department of Clinical Microbiology and Center for Genomic Medicine, Rigshospitalet, Copenhagen, Denmark | Statens Serum Institut Bioinformatics and Microbial Genomics | Danish Covid-19 Genome Consortium |
| hCoV-19/Finland/THL-202134437/2021 | EPI_ISL_8585880 | 2021-12-06 | Fimlab Laboratoriot Oy Tampere | Expert Microbiology, National Institute for Health and Welfare | Soile Blomqvist, Jani Halkilahti, Kirsi Liitsola, Haider al-Hello, P√§ivi Laurila, Erika Lindh, Teemu Smura, Ravi Kant, Phuoc Truong, Olli Vapalahti, Sari Hannula, Pekka Ellonen, Niina Ikonen, Carita Savolainen-Kopra |
| hCoV-19/Aruba/AW-RIVM-77658/2021 | EPI_ISL_8586705 | 2021-12-15 | Dutch COVID-19 response team | National Institute for Public Health and the Environment (RIVM) | Adam Meijer, Harry Vennema, Dirk Eggink, Jeroen Cremer, Sharon van den Brink, Bas van der Veer, AnneMarie van den Brandt, Lisa Wijsman, Kim Freriks, Ryanne Jaarsma, Lynn Aarts, Sanne Bos, Jil Kocken, Jordy de Bakker, Stijn van Rossum, Linda van Someren, F |

| | | | | | |
|---|---|---|---|---|---|
| hCoV-19/Bonaire/BQ-RIVM-77958/2021 | EPI_ISL_8586778 | 2021-12-21 | Dutch COVID-19 response team | National Institute for Public Health and the Environment (RIVM) | Adam Meijer, Harry Vennema, Dirk Eggink, Jeroen Cremer, Sharon van den Brink, Bas van der Veer, AnneMarie van den Brandt, Lisa Wijsman, Kim Freriks, Ryanne Jaarsma, Lynn Aarts, Sanne Bos, Jil Kocken, Jordy de Bakker, Stijn van Rossum, Linda van Someren, F |
| hCoV-19/Curacao/CW-RIVM-76891/2021 | EPI_ISL_8586797 | 2021-12-08 | Dutch COVID-19 response team | National Institute for Public Health and the Environment (RIVM) | Adam Meijer, Harry Vennema, Dirk Eggink, Jeroen Cremer, Sharon van den Brink, Bas van der Veer, AnneMarie van den Brandt, Lisa Wijsman, Kim Freriks, Ryanne Jaarsma, Lynn Aarts, Sanne Bos, Jil Kocken, Jordy de Bakker, Stijn van Rossum, Linda van Someren, F |
| hCoV-19/Brazil/BA-LACEN-BA1015-291969891/2021 | EPI_ISL_8590659 | 2021-12-07 | Central Public Health Laboratory - LACEN - Bahia, Salvador, Brazil | Central Public Health Laboratory - LACEN -Bahia, Salvador, Brazil | Stephane Tosta, Luciana Oliveira, Vanessa Nardy, Gabriela Menezes,Marcela G√≥mez, Lenisa Dandara, Jaqueline Gomes, Vagner Fonseca,Marta Giovanetti,Luiz Alcantara, Felicidade Pereira, Arabela Leal |
| hCoV-19/USA/CA-CDC-FG-225898/2021 | EPI_ISL_8592292 | 2021-12-27 | Fulgent Genetics | Centers for Disease Control and Prevention Division of Viral Diseases, Pathogen Discovery | Dakota Howard,Dhwani Batra,Peter Cook,Jason Caravas,Benjamin Rambo-Martin,Scott Sammons,Yvette Unoarumhi,Matthew Schmerer,Kristine Lacek,Tymeckia Kendall,Victoria Caban Figueroa,Shatavia Morrison,Christopher Gulvick,Erisa Sula,Harry Gao,Mickey Li,John Gao |
| hCoV-19/USA/CA-CDC-FG-225169/2021 | EPI_ISL_8593026 | 2021-12-28 | Fulgent Genetics | Centers for Disease Control and Prevention Division of Viral Diseases, Pathogen Discovery | Dakota Howard,Dhwani Batra,Peter Cook,Jason Caravas,Benjamin Rambo-Martin,Scott Sammons,Yvette Unoarumhi,Matthew Schmerer,Kristine Lacek,Tymeckia Kendall,Victoria Caban Figueroa,Shatavia Morrison,Christopher Gulvick,Erisa Sula,Harry Gao,Mickey Li,John Gao |
| hCoV-19/USA/CO-CDPHE-2102508556/2021 | EPI_ISL_8594699 | 2021-12-22 | Colorado Department of Public Health and Environment | Colorado Department of Public Health and Environment | Laura Bankers, Molly C. Hetherington-Rauth, Diana Ir, Alexandria Rossheim, Michael Martin, Mandy Waters, Arianna Smith, Shannon R. Matzinger, Emily A. Travanty |
| hCoV-19/USA/CO-CDPHE-2102497611/2021 | EPI_ISL_8594773 | 2021-10-17 | Colorado Department of Public Health and Environment | Colorado Department of Public Health and Environment | Laura Bankers, Molly C. Hetherington-Rauth, Diana Ir, Alexandria Rossheim, Michael Martin, Mandy Waters, Arianna Smith, Shannon R. Matzinger, Emily A. Travanty |
| hCoV-19/Canada/AB-ABPHL-47814/2021 | EPI_ISL_8596019 | 2021-12-23 | Edmonton Provincial Lab | Public Health Agency of Canada (PHAC) National Microbiology Laboratory | Buss, E, Croxen M, Deo A, Dieu P, Gill K, Ferrato C, Khan F, Koleva P, Li V, Lloyd C, Lynch T, Ma R, Murphy S, Pabbaraju K, Shokoples S, Tipples G, Thayer J, Whitehouse M, Wong A, Yu C, Zelyas N |
| hCoV-19/Australia/QLD5243/2021 | EPI_ISL_8598287 | 2021-12-26 | PHV-FSS | PHV-FSS | Chenwei Wang on behalf of Q-PHIRE Genomics |
| hCoV-19/Australia/QLD4581/2021 | EPI_ISL_8598290 | 2021-12-24 | PHV-FSS | PHV-FSS | Chenwei Wang on behalf of Q-PHIRE Genomics |
| hCoV-19/USA/AZ-CDC-LC0452384/2021 | EPI_ISL_8599439 | 2021-12-22 | Laboratory Corporation of America | Centers for Disease Control and Prevention Division of Viral Diseases, Pathogen Discovery | Dakota Howard,Dhwani Batra,Peter Cook,Jason Caravas,Benjamin Rambo-Martin,Scott Sammons,Yvette Unoarumhi,Matthew Schmerer,Kristine Lacek,Tymeckia Kendall,Victoria Caban Figueroa,Shatavia Morrison,Christopher Gulvick,Minoo Agarwal,Eyad Almasri,Debbie Boles |

| | | | | | |
|---|---|---|---|---|---|
| hCoV-19/USA/MD-CDC-LC0453393/2021 | EPI_ISL_8601937 | 2021-12-29 | Laboratory Corporation of America | Centers for Disease Control and Prevention Division of Viral Diseases, Pathogen Discovery | Dakota Howard,Dhwani Batra,Peter Cook,Jason Caravas,Benjamin Rambo-Martin,Scott Sammons,Yvette Unoarumhi,Matthew Schmerer,Kristine Lacek,Tymeckia Kendall,Victoria Caban Figueroa,Shatavia Morrison,Christopher Gulvick,Minoo Agarwal,Eyad Almasri,Debbie Boles |
| hCoV-19/USA/NY-CDC-LC0454360/2021 | EPI_ISL_8603155 | 2021-12-30 | Laboratory Corporation of America | Centers for Disease Control and Prevention Division of Viral Diseases, Pathogen Discovery | Dakota Howard,Dhwani Batra,Peter Cook,Jason Caravas,Benjamin Rambo-Martin,Scott Sammons,Yvette Unoarumhi,Matthew Schmerer,Kristine Lacek,Tymeckia Kendall,Victoria Caban Figueroa,Shatavia Morrison,Christopher Gulvick,Minoo Agarwal,Eyad Almasri,Debbie Boles |
| hCoV-19/SouthAfrica/CERI-KRISP-K033618/2021 | EPI_ISL_8604929 | 2021-11-19 | AHRI | and Innvoation, Stellenbosch University and KRISP, KZN Research Innovation and Sequencing Platform, UKZN. | Cele S, Karim F, Khan K, Bernstein M, Giandhari J, Pillay S, Naidoo Y, Ramphal U, Ramphal Y, van Wyk S, Moir M, San JE, Tshiabuila D, Tegally H, Wilkinson E, Sigal A, de Oliveira T |
| hCoV-19/Italy/CAM-IZSM-RD058705D57-IZSM-COLLI-TIGEM/2021 | EPI_ISL_8605079 | 2021-01-03 | IZSM-Virologia | Istituto Zooprofilattico Sperimentale del Mezzogiorno - Unit√† Operativa Complessa di Virologia | Lorena Cardillo, Loredana Cozzolino, Maurizio Viscardi, Claudio de Martinis, Giovanna Fusco, Ester De Carlo, Antonio Limone |
| hCoV-19/Italy/CAM-IZSM-RD054078D56-IZSM-COLLI-TIGEM/2021 | EPI_ISL_8605090 | 2021-01-03 | IZSM-Virologia | Istituto Zooprofilattico Sperimentale del Mezzogiorno - Unit√† Operativa Complessa di Virologia | Lorena Cardillo, Loredana Cozzolino, Maurizio Viscardi, Claudio de Martinis, Giovanna Fusco, Ester De Carlo, Antonio Limone |
| hCoV-19/Botswana/R61B08_BHP_AAC44714/2021 | EPI_ISL_8607932 | 2021-12-17 | Botswana Harvard HIV Reference Laboratory | Botswana Harvard HIV Reference Laboratory | Sikhulile Moyo, Wonderful T. Choga, Dorcas Maruapula, Keoratile Ntshambiwa, Sefetogi Ramaologa, Thongbotho Mphoyakgosi, Boitumelo Zuze, Botshelo Radibe, Legodile Kooepile, Ontlametse T. Bareng, Pamela Smith-Lawrence, Kgomotso Moruisi, Roger Shapiro, Shahi |
| hCoV-19/Botswana/R61B72_BHP_925779/2021 | EPI_ISL_8607963 | 2021-12-29 | Botswana Harvard HIV Reference Laboratory | Botswana Harvard HIV Reference Laboratory | Sikhulile Moyo, Wonderful T. Choga, Dorcas Maruapula, Keoratile Ntshambiwa, Sefetogi Ramaologa, Thongbotho Mphoyakgosi, Boitumelo Zuze, Botshelo Radibe, Legodile Kooepile, Ontlametse T. Bareng, Pamela Smith-Lawrence, Kgomotso Moruisi, Roger Shapiro, Shahi |
| hCoV-19/Botswana/R61B80_BHP_923482/2021 | EPI_ISL_8607971 | 2021-12-28 | Botswana Harvard HIV Reference Laboratory | Botswana Harvard HIV Reference Laboratory | Sikhulile Moyo, Wonderful T. Choga, Dorcas Maruapula, Keoratile Ntshambiwa, Sefetogi Ramaologa, Thongbotho Mphoyakgosi, Boitumelo Zuze, Botshelo Radibe, Legodile Kooepile, Ontlametse T. Bareng, Pamela Smith-Lawrence, Kgomotso Moruisi, Roger Shapiro, Shahi |
| hCoV-19/Botswana/R63B05_BHP_AAC51505/2021 | EPI_ISL_8608043 | 2021-12-28 | Botswana Harvard HIV Reference Laboratory | Botswana Harvard HIV Reference Laboratory | Sikhulile Moyo, Wonderful T. Choga, Dorcas Maruapula, Keoratile Ntshambiwa, Sefetogi Ramaologa, Thongbotho Mphoyakgosi, Boitumelo Zuze, Botshelo Radibe, Legodile Kooepile, Ontlametse T. Bareng, Pamela Smith-Lawrence, Kgomotso Moruisi, Roger Shapiro, Shahi |

| | | | | | |
|---|---|---|---|---|---|
| hCoV-19/Botswana/R65B09_BHP_921181/2021 | EPI_ISL_8608131 | 2021-12-27 | Palapye Primary Hospital Laboratory | Botswana Harvard HIV Reference Laboratory | Sikhulile Moyo, Wonderful T. Choga, Dorcas Maruapula, Keoratile Ntshambiwa, Sefetogi Ramaologa, Thongbotho Mphoyakgosi, Boitumelo Zuze, Botshelo Radibe, Legodile Kooepile, Ontlametse T. Bareng, Pamela Smith-Lawrence, Kgomotso Moruisi, Roger Shapiro, Shahi |
| hCoV-19/Botswana/R65B24_BHP_921342/2021 | EPI_ISL_8608144 | 2021-12-26 | Palapye Primary Hospital Laboratory | Botswana Harvard HIV Reference Laboratory | Sikhulile Moyo, Wonderful T. Choga, Dorcas Maruapula, Keoratile Ntshambiwa, Sefetogi Ramaologa, Thongbotho Mphoyakgosi, Boitumelo Zuze, Botshelo Radibe, Legodile Kooepile, Ontlametse T. Bareng, Pamela Smith-Lawrence, Kgomotso Moruisi, Roger Shapiro, Shahi |
| hCoV-19/Botswana/R65B67_BHP_928796/2021 | EPI_ISL_8608186 | 2021-12-30 | Botswana Harvard HIV Reference Laboratory | Botswana Harvard HIV Reference Laboratory | Sikhulile Moyo, Wonderful T. Choga, Dorcas Maruapula, Keoratile Ntshambiwa, Sefetogi Ramaologa, Thongbotho Mphoyakgosi, Boitumelo Zuze, Botshelo Radibe, Legodile Kooepile, Ontlametse T. Bareng, Pamela Smith-Lawrence, Kgomotso Moruisi, Roger Shapiro, Shahi |
| hCoV-19/USA/CA-CDPH-3000009194/2021 | EPI_ISL_8613082 | 2021-04-19 | CDPH VBL | California Department of Public Health | CDPH-COVIDNet |
| hCoV-19/England/PHEP-YYN97BY/2021 | EPI_ISL_8614210 | 2021-12-16 | Respiratory Virus Unit, Microbiology Services Colindale, Public Health England | COVID-19 Genomics UK (COG-UK) Consortium | PHE Covid Sequencing Team |
| hCoV-19/SouthAfrica/NICD-N23321/2021 | EPI_ISL_8616590 | 2021-11-24 | AMPATH | National Institute for Communicable Diseases of the National Health Laboratory Service | Amoako DG, Everatt J, Kekana D, Scheepers C, Mohale T, Ntuli N, Mahlangu B, Mnguni A, Ismail A, Bhiman JN, Wolter N |
| hCoV-19/SouthAfrica/NICD-N23322/2021 | EPI_ISL_8616591 | 2021-11-24 | AMPATH | National Institute for Communicable Diseases of the National Health Laboratory Service | Amoako DG, Everatt J, Kekana D, Scheepers C, Mohale T, Ntuli N, Mahlangu B, Mnguni A, Ismail A, Bhiman JN, Wolter N |
| hCoV-19/SouthAfrica/NICD-N23342/2021 | EPI_ISL_8616610 | 2021-11-24 | AMPATH | National Institute for Communicable Diseases of the National Health Laboratory Service | Amoako DG, Everatt J, Kekana D, Scheepers C, Mohale T, Ntuli N, Mahlangu B, Mnguni A, Ismail A, Bhiman JN, Wolter N |
| hCoV-19/SouthAfrica/NICD-N23355/2021 | EPI_ISL_8616622 | 2021-11-24 | AMPATH | National Institute for Communicable Diseases of the National Health Laboratory Service | Amoako DG, Everatt J, Kekana D, Scheepers C, Mohale T, Ntuli N, Mahlangu B, Mnguni A, Ismail A, Bhiman JN, Wolter N |
| hCoV-19/SouthAfrica/NICD-N23411/2021 | EPI_ISL_8616662 | 2021-11-23 | AMPATH | National Institute for Communicable Diseases of the National Health Laboratory Service | Amoako DG, Everatt J, Kekana D, Scheepers C, Mohale T, Ntuli N, Mahlangu B, Mnguni A, Ismail A, Bhiman JN, Wolter N |
| hCoV-19/SouthAfrica/NICD-N23454/2021 | EPI_ISL_8616699 | 2021-11-22 | AMPATH | National Institute for Communicable Diseases of the National Health Laboratory Service | Amoako DG, Everatt J, Kekana D, Scheepers C, Mohale T, Ntuli N, Mahlangu B, Mnguni A, Ismail A, Bhiman JN, Wolter N |
| hCoV-19/SouthAfrica/NICD-N23473/2021 | EPI_ISL_8616705 | 2021-10-26 | AMPATH | National Institute for Communicable Diseases of the National Health Laboratory Service | Amoako DG, Everatt J, Kekana D, Scheepers C, Mohale T, Ntuli N, Mahlangu B, Mnguni A, Ismail A, Bhiman JN, Wolter N |
| hCoV-19/SouthAfrica/NICD-N23542/2021 | EPI_ISL_8616752 | 2021-11-28 | VERMAAK | National Institute for Communicable Diseases of the National Health Laboratory Service | Amoako DG, Everatt J, Kekana D, Scheepers C, Mohale T, Ntuli N, Mahlangu B, Mnguni A, Ismail A, Bhiman JN, Wolter N |
| hCoV-19/USA/NY-UB-CHS-05234/2021 | EPI_ISL_8617940 | 2021-11-28 | Catholic Health Laboratories | University at Buffalo Genomics and Bioinformatics Core | Jonathan Bard, Natalie Lamb, Alyssa Pohlman, Brandon Marzullo, Amanda Boccolucci, Norma Nowak, Donald Yergeau, Jennifer Surtees |

| | | | | | |
|---|---|---|---|---|---|
| hCoV-19/England/ALDP-30E33E3/2021 | EPI_ISL_8620261 | 2021-12-30 | Lighthouse Lab in Alderley Park | Wellcome Sanger Institute for the COVID-19 Genomics UK (COG-UK) Consortium | Jacquelyn Wynn, Mairead Hyland, The Lighthouse Lab in Alderley Park and Alex Alderton, Roberto Amato, Jeffrey Barrett, Sonia Goncalves, Ewan Harrison, David K. Jackson, Ian Johnston, Dominic Kwiatkowski, Cordelia Langford, John Sillitoe on behalf of the W |
| hCoV-19/Nigeria/NCDC-NR1597/2021 | EPI_ISL_8621077 | 2021-12-16 | UNILORIN MDRL | National Reference Laboratory, Nigeria Centre for Disease Control | Dr Ndodo Nnaemeka, Olusola Anuoluwapo Akanbi, Chimaobi Chukwu, Oyeronke Ayansola, Oluwapelumi Olufemi ADEYEMI, Mariam Kehinde SULAIMAN, Ireoluwa Yinka JOEL, Abayomi FADEYI, Kolawole Wasiu WAHAB, Mikhail Olayinka BUHARI, Dr Omoare Adesuyi, Anthony Ahumibe, |
| hCoV-19/Nigeria/NCDC-NR1724/2021 | EPI_ISL_8621130 | 2021-12-21 | 54Gene Ekiti | National Reference Laboratory, Nigeria Centre for Disease Control | Dr Ndodo Nnaemeka, Olusola Anuoluwapo Akanbi, Chimaobi Chukwu, Oyeronke Ayansola, Dr Omoare Adesuyi, Anthony Ahumibe, Catherine Okoi, Nwando Mba, Dr Ifedayo Adetifa |
| hCoV-19/USA/CA-CDC-FG-227858/2021 | EPI_ISL_8624994 | 2021-12-27 | Fulgent Genetics | Centers for Disease Control and Prevention Division of Viral Diseases, Pathogen Discovery | Dakota Howard,Dhwani Batra,Peter Cook,Jason Caravas,Benjamin Rambo-Martin,Scott Sammons,Yvette Unoarumhi,Matthew Schmerer,Kristine Lacek,Tymeckia Kendall,Victoria Caban Figueroa,Shatavia Morrison,Christopher Gulvick,Erisa Sula,Harry Gao,Mickey Li,John Gao |
| hCoV-19/USA/CA-CDC-FG-230086/2021 | EPI_ISL_8625553 | 2021-12-30 | Fulgent Genetics | Centers for Disease Control and Prevention Division of Viral Diseases, Pathogen Discovery | Dakota Howard,Dhwani Batra,Peter Cook,Jason Caravas,Benjamin Rambo-Martin,Scott Sammons,Yvette Unoarumhi,Matthew Schmerer,Kristine Lacek,Tymeckia Kendall,Victoria Caban Figueroa,Shatavia Morrison,Christopher Gulvick,Erisa Sula,Harry Gao,Mickey Li,John Gao |
| hCoV-19/Spain/CL-COV17806/2021 | EPI_ISL_8627959 | 2021-11-17 | SARS-CoV-2 Sequencing Castilla y Leon-Spain Consortium | SARS-CoV-2 Sequencing Castilla y Leon-Spain Consortium | Antonio Ordu√±a-Domingo, Marta Hernandez, David Abad, Marta Dominguez-Gil, Silvia Rojo, Gabriel March Rosello, Sonsoles Garcinu√±o P√©rez, Carmen Aldea-Mansilla, M¬™ Fe Brezmes-Valdivieso, Gregoria Meg√≠as Lob√≥n, Mar√≠a Antonia Garc√≠a Castro, Carmen Gim |
| hCoV-19/Morocco/CNRST_CHU01/2021 | EPI_ISL_8629695 | 2021-10-15 | LABORATOIRE CENTRAL DE VIROLOGIE | Functional Genomic Platform UATRS-biology, CNRST | Myrian SEFFAR, Hakima KABBAJ, Ghizlane EL AMIN, Amal ZOUAKI, Mouhssine HEMLALI, Taha Chouati, Hamza Gharmaz, Marouane Melloul, Sanaa Alaoui-Amine, Safae Ghoulame, Elmostafa EL FAHIME. |
| hCoV-19/Morocco/CNRST_CHU14/2021 | EPI_ISL_8629700 | 2021-10-23 | LABORATOIRE CENTRAL DE VIROLOGIE | Functional Genomic Platform UATRS-biology, CNRST | Myrian SEFFAR, Hakima KABBAJ, Ghizlane EL AMIN, Amal ZOUAKI, Mouhssine HEMLALI, Taha Chouati, Hamza Gharmaz, Marouane Melloul, Sanaa Alaoui-Amine, Safae Ghoulame, Elmostafa EL FAHIME. |
| hCoV-19/USA/MD-IGS-012135502174A/2021 | EPI_ISL_8629804 | 2021-12-21 | Maryland Genomics, Institute for Genome Sciences, University of Maryland School of Medicine | Maryland Genomics, Institute for Genome Sciences, University of Maryland School of Medicine | Tallon, Luke J; Sadzewicz, Lisa D; Humphrys, Mike; Ott, Sandra; Roussey, Holly; Vavikolanu, Kranthi; Regan, Kevin; George, Jain; Hazen, Tracy; Fraser, Claire M; Ravel, Jacques |

| | | | | | |
|---|---|---|---|---|---|
| hCoV-19/Brazil/MG-FIOCRUZ-299/2021 | EPI_ISL_8630253 | 2021-10-05 | Laboratorio Municipal de Biologia Molecular | Instituto Rene Rachou / Fiocruz Minas | Thais Silva, Eneida Oliveira, Anna Salim, Andre Menezes, Rubens do Monte Neto, Gabriel Fernandes, Pedro Alves |
| hCoV-19/USA/NY-Wadsworth-21108999-01/2021 | EPI_ISL_8632312 | 2021-12-27 | SUNY UPSTATE MEDICAL UNIVERSITY | Wadsworth Center, New York State Department of Health | Kirsten St. George, Daryl M. Lamson, Alexis Russell, Matthew Shudt, Melissa A Leisner, Jonathan Plitnick, Catharine Prussing, Navjot Singh, John Kelly, Erasmus Schneider, Erica Lasek-Nesselquist |
| hCoV-19/Brazil/RJ-FIOCRUZ-66267/2021 | EPI_ISL_8633889 | 2021-12-30 | Laboratory of Respiratory Viruses and Measles, Oswaldo Cruz Institute, FIOCRUZ | Laboratory of Respiratory Viruses and Measles, Oswaldo Cruz Institute, FIOCRUZ | Paola Resende, Luciana Appolinario, Fernando Motta, Elisa Cavalcante Pereira, Ighor Arantes, Alice Sampaio Rocha, Bruna Mendon√ßa da Silva, J√©ssica Gra√ßa Macedo de Carvalho, Larissa Macedo Pinto, Victor Guimaraes, Marilda Siqueira on behalf of the Fiocr |
| hCoV-19/USA/NY-Wadsworth-21108634-01/2021 | EPI_ISL_8634259 | 2021-12-17 | BIO-REFERENCE LABORATORIES | Wadsworth Center, New York State Department of Health | Kirsten St. George, Daryl M. Lamson, Alexis Russell, Matthew Shudt, Melissa A Leisner, Jonathan Plitnick, Catharine Prussing, Navjot Singh, John Kelly, Erasmus Schneider, Erica Lasek-Nesselquist |
| hCoV-19/USA/KS-KHEL-8650/2021 | EPI_ISL_8636370 | 2021-11-30 | Kansas Health and Environmental Lab | Kansas Health and Environmental Lab | Mike Grose, Jonathan Barnell, Carrie Welch, Gary Buruss, Victor Anderson, Amanda Bradley, and Phil Adam |
| hCoV-19/SouthAfrica/CERI-KRISP-K033643/2021 | EPI_ISL_8636460 | 2021-12-09 | ZARV/NHLS,¬†Department Medical Virology,¬†University of Pretoria | [illegible] and Innvoation, Stellenbosch University and KRISP, KZN Research Innovation and Sequencing Platform, UKZN. | Amy Strydom, Adriano Mendes, Micheala Davids, Sim Mayaphi and Marietjie Venter, Giandhari J, Pillay S, Naidoo Y, Ramphal U, Ramphal Y, Moir M, Van Wyk S, San JE, Tshiabuila D, Tegally H, Wilkinson E, de Oliveira T |
| hCoV-19/SouthAfrica/CERI-KRISP-K033708/2021 | EPI_ISL_8636514 | 2021-11-19 | ZARV/NHLS,¬†Department Medical Virology,¬†University of Pretoria | [illegible] and Innvoation, Stellenbosch University and KRISP, KZN Research Innovation and Sequencing Platform, UKZN. | Amy Strydom, Adriano Mendes, Micheala Davids, Sim Mayaphi and Marietjie Venter, Giandhari J, Pillay S, Naidoo Y, Ramphal U, Ramphal Y, Moir M, Van Wyk S, San JE, Tshiabuila D, Tegally H, Wilkinson E, de Oliveira T |
| hCoV-19/India/AP-CCMB-CIB9453/2021 | EPI_ISL_8637660 | 2021-12-14 | AP SSO | CSIR-Centre for Cellular and Molecular Biology-INSACOG | Malini Nemalikanti,Lamuk Zaveri,Tulasi Nagabandi,Jandhyala Sai Krishna,Sreelekshmi MS, Ara Sreenivas,Shreekant Verma, Amareshwar Vodapalli,B Himasri,Valli Nagalakshmi Undamatla, Sumedha Avadhanula,Priya Nurkurthy,Vidhyadhari Methuku, Archana Bharadwaj Siv |
| hCoV-19/Peru/LIM-INS-11176/2021 | EPI_ISL_8638608 | 2021-12-16 | Laboratorio de Referencia Nacional de Virus Respiratorios. Centro Nacional de Salud Publica. Instituto Nacional de Salud Peru. | Laboratorio de Referencia Nacional de Virus Respiratorios. Centro Nacional de Salud Publica. Instituto Nacional de Salud Peru. | Carlos Padilla Rojas, Veronica Hurtado Vela, Orson Mestanza Millones, Iris Silva Molina, Luis Barcena Flores, Alicia Nu√±ez Llanos, Steve Acedo Lazo, Wendy Lizarraga Olivares, Victor Jimenez Vasquez, Sara Gordillo Vilchez, Kelly Vanessa Izarra Rojas, Edwa |
| hCoV-19/Australia/VIC29887/2021 | EPI_ISL_8640310 | 2021-12-13 | Microbiological Diagnostic Unit - Public Health Laboratory (MDU-PHL) | Microbiological Diagnostic Unit - Public Health Laboratory (MDU-PHL) | Seemann, T., Horan, K., Sherry, N.L. |
| hCoV-19/Australia/VIC29901/2021 | EPI_ISL_8640389 | 2021-12-14 | Microbiological Diagnostic Unit - Public Health Laboratory (MDU-PHL) | Microbiological Diagnostic Unit - Public Health Laboratory (MDU-PHL) | Seemann, T., Horan, K., Sherry, N.L. |
| hCoV-19/Australia/VIC29904/2021 | EPI_ISL_8640391 | 2021-12-15 | Microbiological Diagnostic Unit - Public Health Laboratory (MDU-PHL) | Microbiological Diagnostic Unit - Public Health Laboratory (MDU-PHL) | Seemann, T., Horan, K., Sherry, N.L. |
| hCoV-19/Australia/VIC29923/2021 | EPI_ISL_8640396 | 2021-12-15 | Microbiological Diagnostic Unit - Public Health Laboratory (MDU-PHL) | Microbiological Diagnostic Unit - Public Health Laboratory (MDU-PHL) | Seemann, T., Horan, K., Sherry, N.L. |
| hCoV-19/Australia/VIC30677/2021 | EPI_ISL_8640501 | 2021-12-14 | Microbiological Diagnostic Unit - Public Health Laboratory (MDU-PHL) | Microbiological Diagnostic Unit - Public Health Laboratory (MDU-PHL) | Seemann, T., Horan, K., Sherry, N.L. |

| | | | | | |
|---|---|---|---|---|---|
| hCoV-19/Australia/VIC32728/2021 | EPI_ISL_8640666 | 2021-12-13 | Microbiological Diagnostic Unit - Public Health Laboratory (MDU-PHL) | Microbiological Diagnostic Unit - Public Health Laboratory (MDU-PHL) | Seemann, T., Horan, K., Sherry, N.L. |
| hCoV-19/Australia/NSW-RPAH-2214/2021 | EPI_ISL_8640778 | 2021-12-29 | New South Wales Health Pathology Royal Prince Alfred Hospital | Microbiology RPAH | Foster, C.; Au, J.; Ruiz Silva, M.; Deveson, I.; Bull, R.; Van Hal, S.; Rawlinson, W. |
| hCoV-19/Australia/VIC30231/2021 | EPI_ISL_8640885 | 2021-12-17 | Microbiological Diagnostic Unit - Public Health Laboratory (MDU-PHL) | Microbiological Diagnostic Unit - Public Health Laboratory (MDU-PHL) | Seemann, T., Horan, K., Sherry, N.L. |
| hCoV-19/Australia/VIC30276/2021 | EPI_ISL_8640918 | 2021-12-17 | Microbiological Diagnostic Unit - Public Health Laboratory (MDU-PHL) | Microbiological Diagnostic Unit - Public Health Laboratory (MDU-PHL) | Seemann, T., Horan, K., Sherry, N.L. |
| hCoV-19/Australia/VIC30429/2021 | EPI_ISL_8640962 | 2021-12-16 | Microbiological Diagnostic Unit - Public Health Laboratory (MDU-PHL) | Microbiological Diagnostic Unit - Public Health Laboratory (MDU-PHL) | Seemann, T., Horan, K., Sherry, N.L. |
| hCoV-19/Australia/VIC30398/2021 | EPI_ISL_8640991 | 2021-12-20 | Microbiological Diagnostic Unit - Public Health Laboratory (MDU-PHL) | Microbiological Diagnostic Unit - Public Health Laboratory (MDU-PHL) | Seemann, T., Horan, K., Sherry, N.L. |
| hCoV-19/Australia/VIC30470/2021 | EPI_ISL_8641003 | 2021-12-17 | Microbiological Diagnostic Unit - Public Health Laboratory (MDU-PHL) | Microbiological Diagnostic Unit - Public Health Laboratory (MDU-PHL) | Seemann, T., Horan, K., Sherry, N.L. |
| hCoV-19/Australia/VIC31766/2021 | EPI_ISL_8641264 | 2021-12-20 | Microbiological Diagnostic Unit - Public Health Laboratory (MDU-PHL) | Microbiological Diagnostic Unit - Public Health Laboratory (MDU-PHL) | Seemann, T., Horan, K., Sherry, N.L. |
| hCoV-19/Australia/VIC32116/2021 | EPI_ISL_8641275 | 2021-12-19 | Microbiological Diagnostic Unit - Public Health Laboratory (MDU-PHL) | Microbiological Diagnostic Unit - Public Health Laboratory (MDU-PHL) | Seemann, T., Horan, K., Sherry, N.L. |
| hCoV-19/Australia/VIC32143/2021 | EPI_ISL_8641368 | 2021-12-18 | Microbiological Diagnostic Unit - Public Health Laboratory (MDU-PHL) | Microbiological Diagnostic Unit - Public Health Laboratory (MDU-PHL) | Seemann, T., Horan, K., Sherry, N.L. |
| hCoV-19/Australia/VIC29660/2021 | EPI_ISL_8641645 | 2021-12-08 | Microbiological Diagnostic Unit - Public Health Laboratory (MDU-PHL) | Microbiological Diagnostic Unit - Public Health Laboratory (MDU-PHL) | Seemann, T., Horan, K., Sherry, N.L. |
| hCoV-19/Australia/VIC30667/2021 | EPI_ISL_8641850 | 2021-12-10 | Microbiological Diagnostic Unit - Public Health Laboratory (MDU-PHL) | Microbiological Diagnostic Unit - Public Health Laboratory (MDU-PHL) | Seemann, T., Horan, K., Sherry, N.L. |
| hCoV-19/Australia/VIC30710/2021 | EPI_ISL_8641955 | 2021-12-22 | Microbiological Diagnostic Unit - Public Health Laboratory (MDU-PHL) | Microbiological Diagnostic Unit - Public Health Laboratory (MDU-PHL) | Seemann, T., Horan, K., Sherry, N.L. |
| hCoV-19/Australia/VIC32080/2021 | EPI_ISL_8641994 | 2021-12-23 | Microbiological Diagnostic Unit - Public Health Laboratory (MDU-PHL) | Microbiological Diagnostic Unit - Public Health Laboratory (MDU-PHL) | Seemann, T., Horan, K., Sherry, N.L. |
| hCoV-19/Australia/VIC31263/2021 | EPI_ISL_8642046 | 2021-12-21 | Microbiological Diagnostic Unit - Public Health Laboratory (MDU-PHL) | Microbiological Diagnostic Unit - Public Health Laboratory (MDU-PHL) | Seemann, T., Horan, K., Sherry, N.L. |
| hCoV-19/Australia/VIC31091/2021 | EPI_ISL_8642051 | 2021-12-21 | Microbiological Diagnostic Unit - Public Health Laboratory (MDU-PHL) | Microbiological Diagnostic Unit - Public Health Laboratory (MDU-PHL) | Seemann, T., Horan, K., Sherry, N.L. |
| hCoV-19/Australia/VIC30998/2021 | EPI_ISL_8642182 | 2021-12-24 | Microbiological Diagnostic Unit - Public Health Laboratory (MDU-PHL) | Microbiological Diagnostic Unit - Public Health Laboratory (MDU-PHL) | Seemann, T., Horan, K., Sherry, N.L. |
| hCoV-19/Australia/VIC32180/2021 | EPI_ISL_8642412 | 2021-12-23 | Microbiological Diagnostic Unit - Public Health Laboratory (MDU-PHL) | Microbiological Diagnostic Unit - Public Health Laboratory (MDU-PHL) | Seemann, T., Horan, K., Sherry, N.L. |
| hCoV-19/Australia/VIC32182/2021 | EPI_ISL_8642413 | 2021-12-22 | Microbiological Diagnostic Unit - Public Health Laboratory (MDU-PHL) | Microbiological Diagnostic Unit - Public Health Laboratory (MDU-PHL) | Seemann, T., Horan, K., Sherry, N.L. |
| hCoV-19/Australia/VIC32528/2021 | EPI_ISL_8642433 | 2021-12-23 | Microbiological Diagnostic Unit - Public Health Laboratory (MDU-PHL) | Microbiological Diagnostic Unit - Public Health Laboratory (MDU-PHL) | Seemann, T., Horan, K., Sherry, N.L. |
| hCoV-19/Australia/VIC31837/2021 | EPI_ISL_8642509 | 2021-12-24 | Microbiological Diagnostic Unit - Public Health Laboratory (MDU-PHL) | Microbiological Diagnostic Unit - Public Health Laboratory (MDU-PHL) | Seemann, T., Horan, K., Sherry, N.L. |
| hCoV-19/Australia/VIC31525/2021 | EPI_ISL_8642864 | 2021-12-27 | Microbiological Diagnostic Unit - Public Health Laboratory (MDU-PHL) | Microbiological Diagnostic Unit - Public Health Laboratory (MDU-PHL) | Seemann, T., Horan, K., Sherry, N.L. |

| | | | | | |
|---|---|---|---|---|---|
| hCoV-19/Australia/VIC31367/2021 | EPI_ISL_8642921 | 2021-12-25 | Microbiological Diagnostic Unit - Public Health Laboratory (MDU-PHL) | Microbiological Diagnostic Unit - Public Health Laboratory (MDU-PHL) | Seemann, T., Horan, K., Sherry, N.L. |
| hCoV-19/Australia/VIC31964/2021 | EPI_ISL_8643039 | 2021-12-26 | Microbiological Diagnostic Unit - Public Health Laboratory (MDU-PHL) | Microbiological Diagnostic Unit - Public Health Laboratory (MDU-PHL) | Seemann, T., Horan, K., Sherry, N.L. |
| hCoV-19/Australia/VIC31697/2021 | EPI_ISL_8643061 | 2021-12-26 | Microbiological Diagnostic Unit - Public Health Laboratory (MDU-PHL) | Microbiological Diagnostic Unit - Public Health Laboratory (MDU-PHL) | Seemann, T., Horan, K., Sherry, N.L. |
| hCoV-19/Australia/VIC33425/2021 | EPI_ISL_8643130 | 2021-12-26 | Microbiological Diagnostic Unit - Public Health Laboratory (MDU-PHL) | Microbiological Diagnostic Unit - Public Health Laboratory (MDU-PHL) | Seemann, T., Horan, K., Sherry, N.L. |
| hCoV-19/Australia/VIC32604/2021 | EPI_ISL_8643156 | 2021-12-25 | Microbiological Diagnostic Unit - Public Health Laboratory (MDU-PHL) | Microbiological Diagnostic Unit - Public Health Laboratory (MDU-PHL) | Seemann, T., Horan, K., Sherry, N.L. |
| hCoV-19/Australia/VIC31830/2021 | EPI_ISL_8643180 | 2021-12-25 | Microbiological Diagnostic Unit - Public Health Laboratory (MDU-PHL) | Microbiological Diagnostic Unit - Public Health Laboratory (MDU-PHL) | Seemann, T., Horan, K., Sherry, N.L. |
| hCoV-19/Australia/VIC32186/2021 | EPI_ISL_8643256 | 2021-12-26 | Microbiological Diagnostic Unit - Public Health Laboratory (MDU-PHL) | Microbiological Diagnostic Unit - Public Health Laboratory (MDU-PHL) | Seemann, T., Horan, K., Sherry, N.L. |
| hCoV-19/Australia/VIC33935/2021 | EPI_ISL_8643298 | 2021-12-27 | Microbiological Diagnostic Unit - Public Health Laboratory (MDU-PHL) | Microbiological Diagnostic Unit - Public Health Laboratory (MDU-PHL) | Seemann, T., Horan, K., Sherry, N.L. |
| hCoV-19/Australia/VIC32548/2021 | EPI_ISL_8643344 | 2021-12-28 | Microbiological Diagnostic Unit - Public Health Laboratory (MDU-PHL) | Microbiological Diagnostic Unit - Public Health Laboratory (MDU-PHL) | Seemann, T., Horan, K., Sherry, N.L. |
| hCoV-19/Australia/VIC32557/2021 | EPI_ISL_8643351 | 2021-12-27 | Microbiological Diagnostic Unit - Public Health Laboratory (MDU-PHL) | Microbiological Diagnostic Unit - Public Health Laboratory (MDU-PHL) | Seemann, T., Horan, K., Sherry, N.L. |
| hCoV-19/Australia/VIC32879/2021 | EPI_ISL_8643433 | 2021-12-26 | Microbiological Diagnostic Unit - Public Health Laboratory (MDU-PHL) | Microbiological Diagnostic Unit - Public Health Laboratory (MDU-PHL) | Seemann, T., Horan, K., Sherry, N.L. |
| hCoV-19/Australia/VIC32893/2021 | EPI_ISL_8643447 | 2021-12-26 | Microbiological Diagnostic Unit - Public Health Laboratory (MDU-PHL) | Microbiological Diagnostic Unit - Public Health Laboratory (MDU-PHL) | Seemann, T., Horan, K., Sherry, N.L. |
| hCoV-19/Australia/VIC33944/2021 | EPI_ISL_8643503 | 2021-12-28 | Microbiological Diagnostic Unit - Public Health Laboratory (MDU-PHL) | Microbiological Diagnostic Unit - Public Health Laboratory (MDU-PHL) | Seemann, T., Horan, K., Sherry, N.L. |
| hCoV-19/Australia/VIC33196/2021 | EPI_ISL_8643515 | 2021-12-28 | Microbiological Diagnostic Unit - Public Health Laboratory (MDU-PHL) | Microbiological Diagnostic Unit - Public Health Laboratory (MDU-PHL) | Seemann, T., Horan, K., Sherry, N.L. |
| hCoV-19/Australia/VIC33226/2021 | EPI_ISL_8643544 | 2021-12-25 | Microbiological Diagnostic Unit - Public Health Laboratory (MDU-PHL) | Microbiological Diagnostic Unit - Public Health Laboratory (MDU-PHL) | Seemann, T., Horan, K., Sherry, N.L. |
| hCoV-19/Australia/VIC33250/2021 | EPI_ISL_8643568 | 2021-12-25 | Microbiological Diagnostic Unit - Public Health Laboratory (MDU-PHL) | Microbiological Diagnostic Unit - Public Health Laboratory (MDU-PHL) | Seemann, T., Horan, K., Sherry, N.L. |
| hCoV-19/USA/CO-CDPHE-2102544747/2021 | EPI_ISL_8643930 | 2021-12-29 | Colorado Department of Public Health and Environment | Colorado Department of Public Health and Environment | Laura Bankers, Molly C. Hetherington-Rauth, Diana Ir, Alexandria Rossheim, Michael Martin, Mandy Waters, Arianna Smith, Shannon R. Matzinger, Emily A. Travanty |
| hCoV-19/USA/CO-CDPHE-2102540992/2021 | EPI_ISL_8644245 | 2021-12-29 | Colorado Department of Public Health and Environment | Colorado Department of Public Health and Environment | Laura Bankers, Molly C. Hetherington-Rauth, Diana Ir, Alexandria Rossheim, Michael Martin, Mandy Waters, Arianna Smith, Shannon R. Matzinger, Emily A. Travanty |
| hCoV-19/Australia/VIC29695/2021 | EPI_ISL_8645833 | 2021-11-28 | Microbiological Diagnostic Unit - Public Health Laboratory (MDU-PHL) | Microbiological Diagnostic Unit - Public Health Laboratory (MDU-PHL) | Seemann, T., Horan, K., Sherry, N.L. |
| hCoV-19/Australia/VIC33122/2021 | EPI_ISL_8645868 | 2021-12-29 | Microbiological Diagnostic Unit - Public Health Laboratory (MDU-PHL) | Microbiological Diagnostic Unit - Public Health Laboratory (MDU-PHL) | Seemann, T., Horan, K., Sherry, N.L. |
| hCoV-19/Australia/VIC33689/2021 | EPI_ISL_8645880 | 2021-12-29 | Microbiological Diagnostic Unit - Public Health Laboratory (MDU-PHL) | Microbiological Diagnostic Unit - Public Health Laboratory (MDU-PHL) | Seemann, T., Horan, K., Sherry, N.L. |

| | | | | | |
|---|---|---|---|---|---|
| hCoV-19/Australia/VIC33695/2021 | EPI_ISL_8645885 | 2021-12-29 | Microbiological Diagnostic Unit - Public Health Laboratory (MDU-PHL) | Microbiological Diagnostic Unit - Public Health Laboratory (MDU-PHL) | Seemann, T., Horan, K., Sherry, N.L. |
| hCoV-19/Australia/VIC33700/2021 | EPI_ISL_8645889 | 2021-12-29 | Microbiological Diagnostic Unit - Public Health Laboratory (MDU-PHL) | Microbiological Diagnostic Unit - Public Health Laboratory (MDU-PHL) | Seemann, T., Horan, K., Sherry, N.L. |
| hCoV-19/Australia/VIC34087/2021 | EPI_ISL_8645936 | 2021-12-29 | Microbiological Diagnostic Unit - Public Health Laboratory (MDU-PHL) | Microbiological Diagnostic Unit - Public Health Laboratory (MDU-PHL) | Seemann, T., Horan, K., Sherry, N.L. |
| hCoV-19/Australia/VIC33744/2021 | EPI_ISL_8645969 | 2021-12-29 | Microbiological Diagnostic Unit - Public Health Laboratory (MDU-PHL) | Microbiological Diagnostic Unit - Public Health Laboratory (MDU-PHL) | Seemann, T., Horan, K., Sherry, N.L. |
| hCoV-19/Australia/VIC34233/2021 | EPI_ISL_8646048 | 2021-12-30 | Microbiological Diagnostic Unit - Public Health Laboratory (MDU-PHL) | Microbiological Diagnostic Unit - Public Health Laboratory (MDU-PHL) | Seemann, T., Horan, K., Sherry, N.L. |
| hCoV-19/Israel/CVL-8000831/2021 | EPI_ISL_8647322 | 2021-12-30 | Israel Central Virology laboratory | Israel Central Virology laboratory | Neta Zuckerman, Efrat Dahan Bucris, Michal Mandelboim, Miranda Geva, Omer Asraf, Efrat Glick-Saar, Gideon Rechavi, Oran Erster, Danit Sofer, Ella Mendelson, Orna Mor |
| hCoV-19/Israel/CVL-8000847/2021 | EPI_ISL_8647338 | 2021-12-30 | Israel Central Virology laboratory | Israel Central Virology laboratory | Neta Zuckerman, Efrat Dahan Bucris, Michal Mandelboim, Miranda Geva, Omer Asraf, Efrat Glick-Saar, Gideon Rechavi, Oran Erster, Danit Sofer, Ella Mendelson, Orna Mor |
| hCoV-19/Israel/CVL-8000849/2021 | EPI_ISL_8647340 | 2021-12-30 | Israel Central Virology laboratory | Israel Central Virology laboratory | Neta Zuckerman, Efrat Dahan Bucris, Michal Mandelboim, Miranda Geva, Omer Asraf, Efrat Glick-Saar, Gideon Rechavi, Oran Erster, Danit Sofer, Ella Mendelson, Orna Mor |
| hCoV-19/Israel/CVL-8000886/2021 | EPI_ISL_8647377 | 2021-12-30 | Israel Central Virology laboratory | Israel Central Virology laboratory | Neta Zuckerman, Efrat Dahan Bucris, Michal Mandelboim, Miranda Geva, Omer Asraf, Efrat Glick-Saar, Gideon Rechavi, Oran Erster, Danit Sofer, Ella Mendelson, Orna Mor |
| hCoV-19/Israel/CVL-8000917/2021 | EPI_ISL_8647407 | 2021-12-29 | Israel Central Virology laboratory | Israel Central Virology laboratory | Neta Zuckerman, Efrat Dahan Bucris, Michal Mandelboim, Miranda Geva, Omer Asraf, Efrat Glick-Saar, Gideon Rechavi, Oran Erster, Danit Sofer, Ella Mendelson, Orna Mor |
| hCoV-19/Italy/SIC_CQRC_3209140/2021 | EPI_ISL_8647619 | 2021-12-30 | Laboratorio CQRC | CQRC_QUALITY CONTROL CHEMICAL BIOLOGICAL RISK_AOOR Villa Sofia Cervello Palermo | Di Gaudio F., Contino F., Brunacci G. |
| hCoV-19/NewZealand/21MV3795/2021 | EPI_ISL_8648517 | 2021-12-28 | LabPLUS | Institute of Environmental Science and Research (ESR) | Sarah Cockerton, Clare Gebbie, Dhanisha Patel, Rachel Boyle, SallyAnn Harbison, Olivia Stroeven, Xiaoyun Ren, Matt Storey, Nikki Freed, Muhammad Faisal, Jing Wang, Hermes Perez, Anja Werno, Antje van der Linden, Arlo Upton, Chris Mansell, David Hammer, Dr |
| hCoV-19/NewZealand/21MV3793/2021 | EPI_ISL_8648607 | 2021-12-28 | LabPLUS | Institute of Environmental Science and Research (ESR) | Sarah Cockerton, Clare Gebbie, Dhanisha Patel, Rachel Boyle, SallyAnn Harbison, Olivia Stroeven, Xiaoyun Ren, Matt Storey, Nikki Freed, Muhammad Faisal, Jing Wang, Hermes Perez, Anja Werno, Antje van der Linden, Arlo Upton, Chris Mansell, David Hammer, Dr |

| | | | | | |
|---|---|---|---|---|---|
| hCoV-19/NewZealand/22CV0039/2021 | EPI_ISL_8648609 | 2021-12-29 | Middlemore Hospital | Institute of Environmental Science and Research (ESR) | Sarah Cockerton, Clare Gebbie, Dhanisha Patel, Rachel Boyle, SallyAnn Harbison, Olivia Stroeven, Xiaoyun Ren, Matt Storey, Nikki Freed, Muhammad Faisal, Jing Wang, Hermes Perez, Anja Werno, Antje van der Linden, Arlo Upton, Chris Mansell, David Hammer, Dr |
| hCoV-19/SouthAfrica/SU-NHLS_3637/2021 | EPI_ISL_8649305 | 2021-12-22 | Division of Medical Virology, National Health Laboratory Service (NHLS), Tygerberg Hospital / Stellenbosch University | Health Laboratory Service (NHLS), Tygerberg Hospital / Stellenbosch University | Susan Engelbrecht, Tongai Maponga, Shannon Wilson, Mathilda Claassen, Tania Stander, Gert van Zyl, Wolfgang Preiser |
| hCoV-19/SouthAfrica/SU-NHLS_3692/2021 | EPI_ISL_8649345 | 2021-12-26 | Division of Medical Virology, National Health Laboratory Service (NHLS), Tygerberg Hospital / Stellenbosch University | Health Laboratory Service (NHLS), Tygerberg Hospital / Stellenbosch University | Susan Engelbrecht, Tongai Maponga, Shannon Wilson, Mathilda Claassen, Tania Stander, Gert van Zyl, Wolfgang Preiser |
| hCoV-19/SouthAfrica/SU-NHLS_3707/2021 | EPI_ISL_8649355 | 2021-12-14 | Division of Medical Virology, National Health Laboratory Service (NHLS), Tygerberg Hospital / Stellenbosch University | Health Laboratory Service (NHLS), Tygerberg Hospital / Stellenbosch University | Susan Engelbrecht, Tongai Maponga, Shannon Wilson, Mathilda Claassen, Tania Stander, Gert van Zyl, Wolfgang Preiser |
| hCoV-19/Brazil/SP-IB_115632/2021 | EPI_ISL_8649481 | 2021-06-01 | HOSPITAL MUNICIPAL DE ILHABELA GOV MARIO COVAS JR | Instituto Butantan | Dimas Tadeu Covas, Antonio Jorge Martins, Claudia Renata dos Santos Barros, David Schlesinger, Debora Botequio Moretti, Elaine Cristina Marqueze, Elaine Vieira Santos, Evandra Strazza Rodrigues, Heidge Fukumasu, Jayme Augusto de Souza-Neto, Jos√© Salvator |
| hCoV-19/Brazil/SP-IB_115663/2021 | EPI_ISL_8649512 | 2021-05-27 | HOSPITAL MUNICIPAL DE ILHABELA GOV MARIO COVAS JR | Instituto Butantan | Dimas Tadeu Covas, Antonio Jorge Martins, Claudia Renata dos Santos Barros, David Schlesinger, Debora Botequio Moretti, Elaine Cristina Marqueze, Elaine Vieira Santos, Evandra Strazza Rodrigues, Heidge Fukumasu, Jayme Augusto de Souza-Neto, Jos√© Salvator |
| hCoV-19/India/MH-RFIP07502/2021 | EPI_ISL_8652462 | 2021-12-28 | BJMC, Pune | RF-IISER Pune | Krishanpal Karmodiya, Aurnab Ghose, Joy Merwin Monteiro, Suvarna Joshi, Rajesh Karyakarte |
| hCoV-19/Germany/BY-RKI-I-418739/2021 | EPI_ISL_8665161 | 2021-12-09 | Medizinisch-Diagnostisches Labor Kempten allg√§ulab | Robert Koch Institute | unknown |
| hCoV-19/Germany/SH-RKI-I-420105/2021 | EPI_ISL_8666814 | 2021-12-27 | LADR Zentrallabor DR. Kramer & Kollegen Geesthacht | Robert Koch Institute | unknown |
| hCoV-19/Germany/BY-RKI-I-420244/2021 | EPI_ISL_8666933 | 2021-12-22 | LabKom - Labor Augsburg MVZ GmbH | Robert Koch Institute | unknown |
| hCoV-19/Germany/RP-RKI-I-421249/2021 | EPI_ISL_8667777 | 2021-12-29 | SYNLAB MVZ Trier | Robert Koch Institute | unknown |
| hCoV-19/France/NAQ-HCL021232130401/2021 | EPI_ISL_8669978 | 2021-12-22 | LABORATOIRE d'ANALYSES de BIOLOGIE MEDICALES | CNR Virus des Infections Respiratoires - France SUD | Antonin Bal, Bruno Simon, Th√©ophile Boyer, Gregory Destras, Gwendolyne Burfin, Hadrien Regue, Quentin Semanas, Martine Valette, Bruno Lina, Laurence Josset |
| hCoV-19/Germany/BY-RKI-I-426234/2021 | EPI_ISL_8674546 | 2021-12-29 | SYNLAB MVZ Weiden | Robert Koch Institute | unknown |
| hCoV-19/Germany/BW-RKI-I-432870/2021 | EPI_ISL_8681191 | 2021-12-28 | MVZ Labor Dr. Limbach & Kollegen GbR | Robert Koch Institute | unknown |

| | | | | | |
|---|---|---|---|---|---|
| hCoV-19/AmericanSamoa/AS-CDC-2-5343531/2021 | EPI_ISL_8681208 | 2021-09-30 | Tafuna Health Center | Centers for Disease Control and Prevention Division of Viral Diseases, Pathogen Discovery | Mili Sheth,Sarah Nobles,Joseph Madden,Jasmine Padilla,Alex Burgin,Meghan Bentz,Mark Burroughs,Peter Cook,Clinton Paden,Dhwani Batra,Kristen Knipe,Dakota Howard,Yvette Unoarumhi,Matthew Schmerer,Ben Rambo-Martin,Kristine Lacek,Sam Shepard,Dave Wentworth,Vi |
| hCoV-19/Switzerland/ZH-UZH-IMV-3ba59af8/2021 | EPI_ISL_8682473 | 2021-12-20 | Kantonsspital Winterthur | Institute of Medical Virology, University of Zurich | Daniel Ehrsam, Isabel St√ºrmer, Catharine Aquino, Joel Wirz, Weihong Qi, Hubert Rehrauer, Verena Kufner, Gabriela Ziltener, Maryam Zaheri, Stefan Schmutz, Annette Audig√©, Maria Gr√ºnberg, Kevin Steiner, Jon Huder, Cyril Shah, Riccarda Capaul, Guido Bloem |
| hCoV-19/USA/GA-CDC-ASC210571857/2021 | EPI_ISL_8686683 | 2021-12-27 | Aegis Sciences Corporation | Centers for Disease Control and Prevention Division of Viral Diseases, Pathogen Discovery | Dakota Howard,Dhwani Batra,Peter Cook,Jason Caravas,Benjamin Rambo-Martin,Scott Sammons,Yvette Unoarumhi,Matthew Schmerer,Kristine Lacek,Tymeckia Kendall,Victoria Caban Figueroa,Shatavia Morrison,Christopher Gulvick,Erisa Sula,Cyndi Clark,Patrick Campbell |
| hCoV-19/Brazil/SP-IB_145230/2021 | EPI_ISL_8687529 | 2021-12-21 | CENTRO DE SAUDE DR NELSON DA CUNHA BASTOS | Instituto Butantan | Dimas Tadeu Covas, Antonio Jorge Martins, Claudia Renata dos Santos Barros, David Schlesinger, Debora Botequio Moretti, Elaine Cristina Marqueze, Elaine Vieira Santos, Evandra Strazza Rodrigues, Heidge Fukumasu, Jayme Augusto de Souza-Neto, Jos√© Salvator |
| hCoV-19/Brazil/SP-IB_145713/2021 | EPI_ISL_8687828 | 2021-12-20 | INSIDE | Instituto Butantan | Dimas Tadeu Covas, Antonio Jorge Martins, Claudia Renata dos Santos Barros, David Schlesinger, Debora Botequio Moretti, Elaine Cristina Marqueze, Elaine Vieira Santos, Evandra Strazza Rodrigues, Heidge Fukumasu, Jayme Augusto de Souza-Neto, Jos√© Salvator |
| hCoV-19/Brazil/SP-IB_145742/2021 | EPI_ISL_8687858 | 2021-12-21 | INSIDE | Instituto Butantan | Dimas Tadeu Covas, Antonio Jorge Martins, Claudia Renata dos Santos Barros, David Schlesinger, Debora Botequio Moretti, Elaine Cristina Marqueze, Elaine Vieira Santos, Evandra Strazza Rodrigues, Heidge Fukumasu, Jayme Augusto de Souza-Neto, Jos√© Salvator |
| hCoV-19/SouthAfrica/CERI-KRISP-K034489/2021 | EPI_ISL_8688119 | 2021-12-15 | National Health Laboratory Services, Virology | CERI, Centre for Epidemic Response and Innvoation, Stellenbosch University and KRISP, KZN Research Innovation and Sequencing Platform, UKZN. | Kathleen Subramoney, Florette Treurnicht, Nkhensani Mtileni, Avani Bharuthram, Giandhari J, Pillay S, Naidoo Y, Ramphal U, Ramphal Y, Arisha Maharaj, van Wyk S, Moir M, San JE, Tshiabuila D, Tegally H, Wilkinson E, de Oliveira T |
| hCoV-19/SouthAfrica/CERI-KRISP-K034516/2021 | EPI_ISL_8688140 | 2021-12-30 | National Health Laboratory Service, Kwazulu-Natal, South Africa | and Innvoation, Stellenbosch University and KRISP, KZN Research Innovation and Sequencing Platform, UKZN. | Nokukhanya Mdlalose, Giandhari J, Pillay S, Naidoo Y, Ramphal U, Ramphal Y, Arisha Maharaj, van Wyk S, Moir M, San JE, Tshiabuila D, Tegally H, Wilkinson E, de Oliveira T |
| hCoV-19/SouthAfrica/CERI-KRISP-K034520/2021 | EPI_ISL_8688142 | 2021-12-30 | National Health Laboratory Service, Kwazulu-Natal, South Africa | and Innvoation, Stellenbosch University and KRISP, KZN Research Innovation and Sequencing Platform, UKZN. | Nokukhanya Mdlalose, Giandhari J, Pillay S, Naidoo Y, Ramphal U, Ramphal Y, Arisha Maharaj, van Wyk S, Moir M, San JE, Tshiabuila D, Tegally H, Wilkinson E, de Oliveira T |

| | | | | | |
|---|---|---|---|---|---|
| hCoV-19/Argentina/INEI113562/2021 | EPI_ISL_8688596 | 2021-10-19 | Servicio Virosis Respiratorias-Departamento Virologv≠a-INEI | Instituto Nacional Enfermedades Infecciosas C.G.Malbran | Baumeister E., Avaro M., Benedetti E., Russo M., Dattero ME, Pontoriero A., Cisterna D., Molina V., Perandones C., Tuduri E., De Belder D., Sanchez Loria J., Haim MS., Lorenzo F., Poklepovich T., Campos J. |
| hCoV-19/Argentina/INEI113854/2021 | EPI_ISL_8689252 | 2021-11-10 | Servicio Virosis Respiratorias-Departamento Virologv≠a-INEI | Instituto Nacional Enfermedades Infecciosas C.G.Malbran | Baumeister E., Avaro M., Benedetti E., Russo M., Dattero ME, Pontoriero A., Cisterna D., Molina V., Perandones C., Tuduri E., De Belder D., Sanchez Loria J., Haim MS., Lorenzo F., Poklepovich T., Campos J. |
| hCoV-19/Chile/MA-CADIUMAG-01962/2021 | EPI_ISL_8692712 | 2021-12-14 | Laboratorio de Biologv≠a Molecular, Universidad de Magallanes | Investigacion, Universidad de Magallanes | Constanza Ceroni, Diego Alvarez, Jorge Gonzv°lez, Jacqueline Aldridge, Marcelo Navarrete |
| hCoV-19/Peru/CAJ-INS-12174/2021 | EPI_ISL_8693149 | 2021-12-13 | Laboratorio de Referencia Nacional de Virus Respiratorios. Centro Nacional de Salud Publica. Instituto Nacional de Salud Peru. | Laboratorio de Referencia Nacional de Virus Respiratorios. Centro Nacional de Salud Publica. Instituto Nacional de Salud Peru. | Carlos Padilla Rojas, Veronica Hurtado Vela, Orson Mestanza Millones, Iris Silva Molina, Luis Barcena Flores, Alicia Nuv±ez Llanos, Steve Acedo Lazo, Wendy Lizarraga Olivares, Victor Jimenez Vasquez, Sara Gordillo Vilchez, Kelly Vanessa Izarra Rojas, Edwa |
| hCoV-19/India/MH-KasturbaMCGM-ICMR-INSACOG-WG1343/2021 | EPI_ISL_8693403 | 2021-11-07 | Kasturba Hospital Molecular Lab - INSACOG | Kasturba hospital WGS | Jayanthi Shastri, Vidushi Chitalia |
| hCoV-19/India/MH-KasturbaMCGM-ICMR-INSACOG-WG1364/2021 | EPI_ISL_8693418 | 2021-11-24 | Kasturba Hospital Molecular Lab - INSACOG | Kasturba hospital WGS | Jayanthi Shastri, Vidushi Chitalia |
| hCoV-19/USA/CA-SCPHL-22WGS00055/2021 | EPI_ISL_8693648 | 2021-01-06 | Sonoma County Public Health Lab | Sonoma County Public Health Laboratory | Rachel Rees, Lisa Critchett, Carlos Gonzalez, Iryna Goraichuk |
| hCoV-19/India/MZ-INSACOG-1526100764708/2021 | EPI_ISL_8700670 | 2021-10-17 | INSACOG-Mizoram | National¬†Institute¬†of¬†Biomedical ¬†Genomics ‚Äì¬†INSACOG | Arindam Maitra, Swagnik Roy, Nidhan Kumar Biswas, Gracy Laldinmawii, Sreedhar Chinnaswamy, N Senthil Kumar, Saumitra Das |
| hCoV-19/India/MZ-INSACOG-1526100533085/2021 | EPI_ISL_8701003 | 2021-08-27 | INSACOG-Mizoram | National¬†Institute¬†of¬†Biomedical ¬†Genomics ‚Äì¬†INSACOG | Arindam Maitra, Swagnik Roy, Nidhan Kumar Biswas, Gracy Laldinmawii, Sreedhar Chinnaswamy, N Senthil Kumar, Saumitra Das |
| hCoV-19/India/WB-INSACOG-1931502516067/2021 | EPI_ISL_8701203 | 2021-12-04 | INSACOG-WB | National¬†Institute¬†of¬†Biomedical ¬†Genomics ‚Äì¬†INSACOG | Arindam Maitra, Bhaswati Bandyopadhyay, Nidhan Kumar Biswas, Tamal Ghosh, Sreedhar Chinnaswamy, Ajay Chakraborti, Saumitra Das |
| hCoV-19/England/PLYM-2FD8084/2021 | EPI_ISL_8706980 | 2021-12-23 | Lighthouse Laboratory Plymouth | Wellcome Sanger Institute for the COVID-19 Genomics UK (COG-UK) Consortium | Lighthouse Laboratory Plymouth and Alex Alderton, Roberto Amato, Jeffrey Barrett, Sonia Goncalves, Ewan Harrison, David K. Jackson, Ian Johnston, Dominic Kwiatkowski, Cordelia Langford, John Sillitoe on behalf of the Wellcome Sanger Institute COVID-19 Sur |
| hCoV-19/Brazil/PE-FIOCRUZ-IAM6612/2021 | EPI_ISL_8708717 | 2021-12-16 | LACEN/PE | WallauLab on behalf of Fiocruz COVID-19 Genomic Surveillance Network | Marcelo Henrique dos Santos Paiva, Cassia Docena, Matheus Filgueira Bezerra, Filipe Zimmer Dezordi, Antonio Marinho da Silva Neto, Lais Ceschini Machado, Lilian Caroliny Amorim Silva, Gustavo Barbosa de Lima, Alexandre Freitas da Silva, Sinval Pinto Brand |

| | | | | | |
|---|---|---|---|---|---|
| hCoV-19/Norway/34381/2021 | EPI_ISL_8712562 | 2021-12-30 | Furst Medical Laboratory | Norwegian Institute of Public Health, Department of Virology | Kathrine Stene-Johansen, Kamilla Heddeland Instefjord, Hilde Elshaug, Garcia Llorente Ignacio, Jon Br√•te, Engebretsen Serina Beate, Pedersen Benedikte Nevjen, Line Victoria Moen, Debech Nadia, Atiya R Ali, Marie Paulsen Madsen, Rasmus Riis Kopperud, Hild |
| hCoV-19/France/IDF-HMN-21122290226/2021 | EPI_ISL_8712678 | 2021-12-21 | BIO VSM LAB | Mondor University Hospital, Assistance Publique H√¥pitaux de Paris, Universit√© Paris-Est Cr√©teil, INSERM U955 | Christophe Rodriguez, Slim Fourati, Vanessa Demontant, Guillaume Gricourt, Melissa N'Debi, Alexandre Soulier, Elisabeth Trawinski, Jean-Michel Pawlotsky |
| hCoV-19/France/BFC-HMN-21122300847/2021 | EPI_ISL_8712774 | 2021-04-16 | LPA BESANCON | Mondor University Hospital, Assistance Publique H√¥pitaux de Paris, Universit√© Paris-Est Cr√©teil, INSERM U955 | Christophe Rodriguez, Slim Fourati, Vanessa Demontant, Guillaume Gricourt, Melissa N'Debi, Alexandre Soulier, Elisabeth Trawinski, Jean-Michel Pawlotsky |
| hCoV-19/France/BFC-HMN-21122300055/2021 | EPI_ISL_8712784 | 2021-12-27 | EVORIAL Laboratoire | Mondor University Hospital, Assistance Publique H√¥pitaux de Paris, Universit√© Paris-Est Cr√©teil, INSERM U955 | Christophe Rodriguez, Slim Fourati, Vanessa Demontant, Guillaume Gricourt, Melissa N'Debi, Alexandre Soulier, Elisabeth Trawinski, Jean-Michel Pawlotsky |
| hCoV-19/India/MH-KasturbaMCGM-ICMR-INSACOG-WG2329/2021 | EPI_ISL_8713073 | 2021-12-20 | Kasturba Hospital Molecular Lab - INSACOG | Kasturba hospital WGS | Jayanthi Shastri, Vidushi Chitalia |
| hCoV-19/India/MH-KasturbaMCGM-ICMR-INSACOG-WG2205/2021 | EPI_ISL_8713135 | 2021-12-21 | Kasturba Hospital Molecular Lab - INSACOG | Kasturba hospital WGS | Jayanthi Shastri, Vidushi Chitalia |
| hCoV-19/Croatia/19008/2021 | EPI_ISL_8713273 | 2021-12-17 | Klinika za infektivne bolesti "Dr. Fran Mihaljevifá" | Hrvatski zavod za javno zdravstvo | Irena Tabain, Ivana Ferenfçak, Josipa Kuzle, Anita Jurifá, Dragan Jurifá |
| hCoV-19/Peru/LIM-INS-11267/2021 | EPI_ISL_8716068 | 2021-12-20 | Laboratorio de Referencia Nacional de Virus Respiratorios. Centro Nacional de Salud Publica. Instituto Nacional de Salud Peru. | Laboratorio de Referencia Nacional de Virus Respiratorios. Centro Nacional de Salud Publica. Instituto Nacional de Salud Peru. | Carlos Padilla Rojas, Veronica Hurtado Vela, Orson Mestanza Millones, Iris Silva Molina, Luis Barcena Flores, Alicia Nu√±ez Llanos, Steve Acedo Lazo, Wendy Lizarraga Olivares, Victor Jimenez Vasquez, Sara Gordillo Vilchez, Kelly Vanessa Izarra Rojas, Edwa |
| hCoV-19/Peru/LIM-INS-11294/2021 | EPI_ISL_8716095 | 2021-12-19 | Laboratorio de Referencia Nacional de Virus Respiratorios. Centro Nacional de Salud Publica. Instituto Nacional de Salud Peru. | Laboratorio de Referencia Nacional de Virus Respiratorios. Centro Nacional de Salud Publica. Instituto Nacional de Salud Peru. | Carlos Padilla Rojas, Veronica Hurtado Vela, Orson Mestanza Millones, Iris Silva Molina, Luis Barcena Flores, Alicia Nu√±ez Llanos, Steve Acedo Lazo, Wendy Lizarraga Olivares, Victor Jimenez Vasquez, Sara Gordillo Vilchez, Kelly Vanessa Izarra Rojas, Edwa |
| hCoV-19/Peru/LIM-INS-11302/2021 | EPI_ISL_8716104 | 2021-12-19 | Laboratorio de Referencia Nacional de Virus Respiratorios. Centro Nacional de Salud Publica. Instituto Nacional de Salud Peru. | Laboratorio de Referencia Nacional de Virus Respiratorios. Centro Nacional de Salud Publica. Instituto Nacional de Salud Peru. | Carlos Padilla Rojas, Veronica Hurtado Vela, Orson Mestanza Millones, Iris Silva Molina, Luis Barcena Flores, Alicia Nu√±ez Llanos, Steve Acedo Lazo, Wendy Lizarraga Olivares, Victor Jimenez Vasquez, Sara Gordillo Vilchez, Kelly Vanessa Izarra Rojas, Edwa |
| hCoV-19/Poland/PZH-GUM-5511/2021 | EPI_ISL_8716834 | 2021-12-29 | Warsaw Genomics Laboratorium Analiz Genetycznych | [illegible] consortium: University of Gdansk, Medical University of Gdansk, Vaxican Ltd., Invicta Ltd. 2. National Institute of Public Health - National Institute of Hygiene, Warsaw, Poland | Maciej Kosinski, Celina Cybulska, Krystyna Bienkowska Szewczyk, Maciej Grzybek, Karolina Gackowska, Marcin Lubocki, Katarzyna Groth, Lukasz Rabalski, Katarzyna Zacharczuk, Magdalena Nowakowska, Ma≈Çgorzata Sadkowska-Todys, Tomasz Wo≈Çkowicz |

| | | | | | |
|---|---|---|---|---|---|
| hCoV-19/USA/CA-CDC-FG-230669/2021 | EPI_ISL_8717406 | 2021-12-30 | Fulgent Genetics | Centers for Disease Control and Prevention Division of Viral Diseases, Pathogen Discovery | Dakota Howard,Dhwani Batra,Peter Cook,Jason Caravas,Benjamin Rambo-Martin,Scott Sammons,Yvette Unoarumhi,Matthew Schmerer,Kristine Lacek,Tymeckia Kendall,Victoria Caban Figueroa,Shatavia Morrison,Christopher Gulvick,Erisa Sula,Harry Gao,Mickey Li,John Gao |
| hCoV-19/USA/PA-CDC-LC0455179/2021 | EPI_ISL_8718510 | 2021-12-29 | Laboratory Corporation of America | Centers for Disease Control and Prevention Division of Viral Diseases, Pathogen Discovery | Dakota Howard,Dhwani Batra,Peter Cook,Jason Caravas,Benjamin Rambo-Martin,Scott Sammons,Yvette Unoarumhi,Matthew Schmerer,Kristine Lacek,Tymeckia Kendall,Victoria Caban Figueroa,Shatavia Morrison,Christopher Gulvick,Minoo Agarwal,Eyad Almasri,Debbie Boles |
| hCoV-19/USA/TN-CDC-ASC210571237/2021 | EPI_ISL_8720121 | 2021-12-29 | Aegis Sciences Corporation | Centers for Disease Control and Prevention Division of Viral Diseases, Pathogen Discovery | Dakota Howard,Dhwani Batra,Peter Cook,Jason Caravas,Benjamin Rambo-Martin,Scott Sammons,Yvette Unoarumhi,Matthew Schmerer,Kristine Lacek,Tymeckia Kendall,Victoria Caban Figueroa,Shatavia Morrison,Christopher Gulvick,Erisa Sula,Cyndi Clark,Patrick Campbell |
| hCoV-19/Brazil/SP-IB_146500/2021 | EPI_ISL_8721239 | 2021-12-27 | BIOFAST | Instituto Butantan | Dimas Tadeu Covas, Antonio Jorge Martins, Claudia Renata dos Santos Barros, David Schlesinger, Debora Botequio Moretti, Elaine Cristina Marqueze, Elaine Vieira Santos, Evandra Strazza Rodrigues, Heidge Fukumasu, Jayme Augusto de Souza-Neto, Jos√© Salvator |
| hCoV-19/Brazil/SP-IB_146374/2021 | EPI_ISL_8721308 | 2021-12-29 | BIOFAST | Instituto Butantan | Dimas Tadeu Covas, Antonio Jorge Martins, Claudia Renata dos Santos Barros, David Schlesinger, Debora Botequio Moretti, Elaine Cristina Marqueze, Elaine Vieira Santos, Evandra Strazza Rodrigues, Heidge Fukumasu, Jayme Augusto de Souza-Neto, Jos√© Salvator |
| hCoV-19/Brazil/SP-IB_146362/2021 | EPI_ISL_8721466 | 2021-12-29 | BIOFAST | Instituto Butantan | Dimas Tadeu Covas, Antonio Jorge Martins, Claudia Renata dos Santos Barros, David Schlesinger, Debora Botequio Moretti, Elaine Cristina Marqueze, Elaine Vieira Santos, Evandra Strazza Rodrigues, Heidge Fukumasu, Jayme Augusto de Souza-Neto, Jos√© Salvator |
| hCoV-19/Brazil/SP-IB_146049/2021 | EPI_ISL_8721475 | 2021-12-27 | SECRETARIA MUNICIPAL DE SAUDE SOROCABA | Instituto Butantan | Dimas Tadeu Covas, Antonio Jorge Martins, Claudia Renata dos Santos Barros, David Schlesinger, Debora Botequio Moretti, Elaine Cristina Marqueze, Elaine Vieira Santos, Evandra Strazza Rodrigues, Heidge Fukumasu, Jayme Augusto de Souza-Neto, Jos√© Salvator |
| hCoV-19/Brazil/SP-IB_146553/2021 | EPI_ISL_8721547 | 2021-12-29 | FACULDADE DE MEDICINA ABC | Instituto Butantan | Dimas Tadeu Covas, Antonio Jorge Martins, Claudia Renata dos Santos Barros, David Schlesinger, Debora Botequio Moretti, Elaine Cristina Marqueze, Elaine Vieira Santos, Evandra Strazza Rodrigues, Heidge Fukumasu, Jayme Augusto de Souza-Neto, Jos√© Salvator |

| | | | | | |
|---|---|---|---|---|---|
| hCoV-19/Brazil/SP-IB_146508/2021 | EPI_ISL_8721805 | 2021-12-28 | BIOFAST | Instituto Butantan | Dimas Tadeu Covas, Antonio Jorge Martins, Claudia Renata dos Santos Barros, David Schlesinger, Debora Botequio Moretti, Elaine Cristina Marqueze, Elaine Vieira Santos, Evandra Strazza Rodrigues, Heidge Fukumasu, Jayme Augusto de Souza-Neto, Josv© Salvator |
| hCoV-19/Brazil/SP-IB_145994/2021 | EPI_ISL_8721816 | 2021-12-28 | CS II DR JOSE CARQUEIJO SCRPARDO | Instituto Butantan | Dimas Tadeu Covas, Antonio Jorge Martins, Claudia Renata dos Santos Barros, David Schlesinger, Debora Botequio Moretti, Elaine Cristina Marqueze, Elaine Vieira Santos, Evandra Strazza Rodrigues, Heidge Fukumasu, Jayme Augusto de Souza-Neto, Josv© Salvator |
| hCoV-19/Brazil/SP-IB_146597/2021 | EPI_ISL_8721854 | 2021-12-27 | INSIDE DIAGNvìSTICOS | Instituto Butantan | Dimas Tadeu Covas, Antonio Jorge Martins, Claudia Renata dos Santos Barros, David Schlesinger, Debora Botequio Moretti, Elaine Cristina Marqueze, Elaine Vieira Santos, Evandra Strazza Rodrigues, Heidge Fukumasu, Jayme Augusto de Souza-Neto, Josv© Salvator |
| hCoV-19/Brazil/SP-IB_146530/2021 | EPI_ISL_8721904 | 2021-12-29 | FACULDADE DE MEDICINA ABC | Instituto Butantan | Dimas Tadeu Covas, Antonio Jorge Martins, Claudia Renata dos Santos Barros, David Schlesinger, Debora Botequio Moretti, Elaine Cristina Marqueze, Elaine Vieira Santos, Evandra Strazza Rodrigues, Heidge Fukumasu, Jayme Augusto de Souza-Neto, Josv© Salvator |
| hCoV-19/Brazil/SP-IB_146481/2021 | EPI_ISL_8721927 | 2021-12-29 | BIOFAST | Instituto Butantan | Dimas Tadeu Covas, Antonio Jorge Martins, Claudia Renata dos Santos Barros, David Schlesinger, Debora Botequio Moretti, Elaine Cristina Marqueze, Elaine Vieira Santos, Evandra Strazza Rodrigues, Heidge Fukumasu, Jayme Augusto de Souza-Neto, Josv© Salvator |
| hCoV-19/Brazil/SP-IB_146728/2021 | EPI_ISL_8721932 | 2021-12-28 | CENTRO DE SAUDE DE RIBEIRAO DO SUL | Instituto Butantan | Dimas Tadeu Covas, Antonio Jorge Martins, Claudia Renata dos Santos Barros, David Schlesinger, Debora Botequio Moretti, Elaine Cristina Marqueze, Elaine Vieira Santos, Evandra Strazza Rodrigues, Heidge Fukumasu, Jayme Augusto de Souza-Neto, Josv© Salvator |
| hCoV-19/Ireland/KK-NVRL-S21IRL00580426/2021 | EPI_ISL_8722839 | 2021-12-18 | National Virus Reference Laboratory | National Virus Reference Laboratory | Zoe Yandle, Charlene Bennett, Gabriel Gonzalez, Michael Carr, Jonathan Dean, Cillian F De Gascun |
| hCoV-19/CzechRepublic/NRL_16769/2021 | EPI_ISL_8725203 | 2021-03-19 | National Institute of Public Health | National Institute of Public Health | Helena Jirincova, Jaromira Vecerova, Timotej Suri, Alexander Nagy |
| hCoV-19/USA/CA-CDPH-3000052990/2021 | EPI_ISL_8729848 | 2021-06-13 | CDPH VBL | California Department of Public Health | CDPH-COVIDNet |
| hCoV-19/Spain/CL-COV18828/2021 | EPI_ISL_8731452 | 2021-11-26 | SARS-CoV-2 Sequencing Castilla y Leon-Spain Consortium | SARS-CoV-2 Sequencing Castilla y Leon-Spain Consortium | Antonio Orduv±a-Domingo, Marta Hernandez, David Abad, Marta Dominguez-Gil, Silvia Rojo, Gabriel March Rosello, Sonsoles Garcinuv±o Pv©rez, Carmen Aldea-Mansilla, M¬™ Fe Brezmes-Valdivieso, Gregoria Megv≠as Lobv≥n, Marv≠a Antonia Garcv≠a Castro, Carmen Gim |
| hCoV-19/England/PHEP-YYDBAEU/2021 | EPI_ISL_8739344 | 2021-03-20 | Respiratory Virus Unit, Microbiology Services Colindale, Public Health England | COVID-19 Genomics UK (COG-UK) Consortium | PHE Covid Sequencing Team |

| | | | | | |
|---|---|---|---|---|---|
| hCoV-19/England/PHEP-YYDNX1F/2021 | EPI_ISL_8742630 | 2021-07-10 | Respiratory Virus Unit, Microbiology Services Colindale, Public Health England | COVID-19 Genomics UK (COG-UK) Consortium | PHE Covid Sequencing Team |
| hCoV-19/England/LSPA-30EEBC2/2021 | EPI_ISL_8743601 | 2021-12-29 | Rosalind Franklin Laboratory | Wellcome Sanger Institute for the COVID-19 Genomics UK (COG-UK) Consortium | Donald Fraser, Suki Lee, Rob Howes, The Rosalind Franklin Laboratory and Alex Alderton, Roberto Amato, Jeffrey Barrett, Sonia Goncalves, Ewan Harrison, David K. Jackson, Ian Johnston, Dominic Kwiatkowski, Cordelia Langford, John Sillitoe on behalf of the |
| hCoV-19/India/GJ-INSACOG-GBRC4008/2021 | EPI_ISL_8744139 | 2021-12-24 | Neuberg Supratech Reference Laboratory, Ahmedabad | Gujarat Biotechnology Research Centre | Priyank Chavda,Sonal Sharma,Roshani Mishra,Disha Vora,Janvi Raval,Apurvasinh Puvar,Zarna Patel,Bhadreshsinh Gohil,Nimesh Patel,Nitin Shukla,Akhilesh Modi,Tasnim Trivedi,Jaykumar Rangani,Ramesh Pandit,Nitin Savaliya,Jignesh Panchal,Chaitanya Joshi,Madhvi J |
| hCoV-19/Mexico/CMX_InDRE_FB54358_E09312665724_S9987/2021 | EPI_ISL_8747609 | 2021-11-19 | LESP Mexico City | Instituto de Diagnostico y Referencia Epidemiologicos (INDRE) | Claudia Wong-Arambula, Abril Rodriguez-Maldonado, Vanessa Rivero-Arredondo, Ariadna Medina-Benitez, Joaquin Quiroz-Mercado, Sergio Rangel-Guerrero, Maribel Gonzalez-Villa, Natividad Cruz-Ortiz, Tatiana Nunez-Garcia, Gisela Barrera-Badillo, Fernando Gonzal |
| hCoV-19/USA/CA-LACPHL-AF05859/2021 | EPI_ISL_8749341 | 2021-02-28 | Flow Diagnostics Lab | Los Angeles County Public Health Laboratories | A.R. Gener et al. |
| hCoV-19/USA/CA-CDPH-3000053352/2021 | EPI_ISL_8750559 | 2021-06-16 | CDPH VBL | California Department of Public Health | CDPH-COVIDNet |
| hCoV-19/USA/NJ-PHEL-V22001459/2021 | EPI_ISL_8751733 | 2021-01-05 | NJDOH, Public Health and Environmental Laboratories | NJ_PHEL | Lindsey Bodnar, Shiv K. Verma, Maria-Magdalene Pugliese, Chelsea San Filippo, Mohammad M. Ali, Dana Woell, Byeong Jeong |
| hCoV-19/Japan/BRL21-0392/2021 | EPI_ISL_8753746 | 2021-12-09 | National Center for Global Health and Medicine | [illegible] Laboratory, Department of Academic-Industrial Partnerships Promotion, Center for Clinical Sciences, National Center for Global Health and Medicine | Junko S. TAKEUCHI, Nobumasa OKUMURA, Shinya TSUZUKI, Sho SAITO, Yumiko KITO, Azusa KAMIKAWA, Moto KIMURA, Wataru SUGIURA, Norio OHMAGARI |
| hCoV-19/CzechRepublic/FNHK-Ps-002387/2021 | EPI_ISL_8754108 | 2021-01-30 | Oblastni nemocnice Nachod a.s. | University Hospital Hradec Kralove | Marketa Gancarcikova, Helena Kovarikova, Lenka Rysava, Monika Berankova |
| hCoV-19/England/PHEP-YYDGS7Y/2021 | EPI_ISL_8761524 | 2021-12-26 | Respiratory Virus Unit, Microbiology Services Colindale, Public Health England | COVID-19 Genomics UK (COG-UK) Consortium | PHE Covid Sequencing Team |
| hCoV-19/Wales/PHWC-PGURZG/2021 | EPI_ISL_8763668 | 2021-12-20 | Originating lab: Wales Specialist Virology Centre Sequencing lab: Pathogen Genomics Unit | Public Health Wales Microbiology Cardiff Wales Specialist Virology Centre | Catherine Moore, Johnathan Evans, Laura Gifford, Malorie Perry, Simon Cottrell, Angela Marchbank, Alec Birchley, Alexander Adams, Amy Gaskin, Bree Gatica-Wilcox, Jason Coombes, Joel Southgate, Lauren Gilbert, Lee Graham, Nicole Pacchiarini, Sara Kumziene- |
| hCoV-19/Brazil/SP-NVBS10589GENOV828368438217/2021 | EPI_ISL_8765828 | 2021-11-06 | DASA | DASA | Rodrigo Guarischi, Paulo Pierry, Adriano Bonaldi, Cristina Oliveira, Bianca Cota, Luciane Sussuchi, Camila Romano, Angelica Hristov, Lidia Yamamoto, Rodrigo Salazar, Annelise Lopes, Jose Levi |

| | | | | | |
|---|---|---|---|---|---|
| hCoV-19/Brazil/SP-NVBS10813GENOV828347431062/2021 | EPI_ISL_8765988 | 2021-11-02 | DASA | DASA | Rodrigo Guarischi, Paulo Pierry, Adriano Bonaldi, Cristina Oliveira, Bianca Cota, Luciane Sussuchi, Camila Romano, Angelica Hristov, Lidia Yamamoto, Rodrigo Salazar, Annelise Lopes, Jose Levi |
| hCoV-19/Brazil/SP-NVBS10886GENOV828366391583/2021 | EPI_ISL_8766035 | 2021-11-06 | DASA | DASA | Rodrigo Guarischi, Paulo Pierry, Adriano Bonaldi, Cristina Oliveira, Bianca Cota, Luciane Sussuchi, Camila Romano, Angelica Hristov, Lidia Yamamoto, Rodrigo Salazar, Annelise Lopes, Jose Levi |
| hCoV-19/Malaysia/A287/2021 | EPI_ISL_8767280 | 2021-12-27 | Kuala Lumpur Internation Airport (KLIA) Health Office | Faculty of Medicine, University of Malaya; University of Malaya Medical Centre | Omar Khalilur Rahman, Jolene Yin Ling FU, I-Ching SAM, Yoke Fun Chan |
| hCoV-19/env/Austria/CeMM21559/2021 | EPI_ISL_8769283 | 2021-12-26 | Institute for Water Quality and Resource Management, Technical University Vienna | Bergthaler laboratory, CeMM Research Center for Molecular Medicine of the Austrian Academy of Sciences | Lukas Endler, Anna Schedl, Fabian Amman, Petr Triska, Matthew Thornton, Thomas Penz, Benedikt Agerer, Michelle Chan, Michael Schuster, Bekir Erguner, Jan Laine, Martin Senekowitsch, Christoph Bock, Andreas Bergthaler |
| hCoV-19/India/DL-ILBS-WGS1320/2021 | EPI_ISL_8769709 | 2021-12-26 | CARINGdx-Mahajan Imaging | ILBS | Ekta Gupta, Varun Suroliya, Pramod Gautam, Reshu Agarwal, Rahul Garg, Urvinder Kaur, Arjun Bhugra, Chhagan Bihari Sharma, Shiv Kumar Sarin |
| hCoV-19/India/DL-ILBS-WGS1328/2021 | EPI_ISL_8769717 | 2021-12-23 | NRL, Dr Lal Path Lab Ltds | ILBS | Ekta Gupta, Varun Suroliya, Pramod Gautam, Reshu Agarwal, Rahul Garg, Urvinder Kaur, Arjun Bhugra, Chhagan Bihari Sharma, Shiv Kumar Sarin |
| hCoV-19/India/DL-ILBS-WGS1440/2021 | EPI_ISL_8769829 | 2021-12-24 | Lifeline Laboratory | ILBS | Ekta Gupta, Varun Suroliya, Pramod Gautam, Reshu Agarwal, Rahul Garg, Urvinder Kaur, Arjun Bhugra, Chhagan Bihari Sharma, Shiv Kumar Sarin |
| hCoV-19/India/DL-ILBS-WGS1561/2021 | EPI_ISL_8769851 | 2021-12-25 | GENE STORE | ILBS | Ekta Gupta, Varun Suroliya, Pramod Gautam, Reshu Agarwal, Rahul Garg, Urvinder Kaur, Arjun Bhugra, Chhagan Bihari Sharma, Shiv Kumar Sarin |
| hCoV-19/India/DL-ILBS-WGS1584/2021 | EPI_ISL_8769871 | 2021-12-28 | MAHAJAN IMAGING | ILBS | Ekta Gupta, Varun Suroliya, Pramod Gautam, Reshu Agarwal, Rahul Garg, Urvinder Kaur, Arjun Bhugra, Chhagan Bihari Sharma, Shiv Kumar Sarin |
| hCoV-19/India/DL-ILBS-WGS1585/2021 | EPI_ISL_8769872 | 2021-12-28 | MAHAJAN IMAGING | ILBS | Ekta Gupta, Varun Suroliya, Pramod Gautam, Reshu Agarwal, Rahul Garg, Urvinder Kaur, Arjun Bhugra, Chhagan Bihari Sharma, Shiv Kumar Sarin |
| hCoV-19/England/PHEP-YYD8EY8/2021 | EPI_ISL_8782083 | 2021-12-22 | Respiratory Virus Unit, Microbiology Services Colindale, Public Health England | COVID-19 Genomics UK (COG-UK) Consortium | PHE Covid Sequencing Team |
| hCoV-19/Wales/PHWC-PGWTHP/2021 | EPI_ISL_8785818 | 2021-02-21 | Originating lab: Wales Specialist Virology Centre Sequencing lab: Pathogen Genomics Unit | Public Health Wales Microbiology Cardiff Wales Specialist Virology Centre | Catherine Moore, Johnathan Evans, Laura Gifford, Malorie Perry, Simon Cottrell, Angela Marchbank, Alec Birchley, Alexander Adams, Amy Gaskin, Bree Gatica-Wilcox, Jason Coombes, Joel Southgate, Lauren Gilbert, Lee Graham, Nicole Pacchiarini, Sara Kumziene- |
| hCoV-19/France/ARA-HCL022001382101/2022 | EPI_ISL_8787158 | 2022-01-03 | CNR Virus des Infections Respiratoires - France SUD | CNR Virus des Infections Respiratoires - France SUD | Antonin Bal, Bruno Simon, Th√©ophile Boyer, Gregory Destras, Gwendolyne Burfin, Hadrien Regue, Quentin Semanas, Martine Valette, Bruno Lina, Laurence Josset |

| | | | | | |
|---|---|---|---|---|---|
| hCoV-19/France/IDF-IPP52314/2021 | EPI_ISL_8789586 | 2021-12-27 | Hopital | National Reference Center for Viruses of Respiratory Infections, Institut Pasteur, Paris | Marion Barbet, Sylvie Behillil, M√©line Bizard, Angela Brisebarre, Camille Capel, Vincent Enouf, Louise Lefran√ßois, Fr√©d√©ric Lemoine, Christophe Malabat, Corinne Maufrais, Slim El Khiari, Julien Fumey, Etienne Simon-Lori√®re, Maud Vanpeene, Sylvie Van |
| hCoV-19/France/IDF-IPP52965/2021 | EPI_ISL_8789837 | 2021-12-27 | Labo Analyses Med | National Reference Center for Viruses of Respiratory Infections, Institut Pasteur, Paris | Marion Barbet, Sylvie Behillil, M√©line Bizard, Angela Brisebarre, Camille Capel, Vincent Enouf, Louise Lefran√ßois, Fr√©d√©ric Lemoine, Christophe Malabat, Corinne Maufrais, Slim El Khiari, Julien Fumey, Etienne Simon-Lori√®re, Maud Vanpeene, Sylvie Van |
| hCoV-19/Luxembourg/LNS2757724/2021 | EPI_ISL_8791864 | 2021-12-21 | Centre Hospitalier du Nord | Laboratoire national de sante, Microbiology, Microbial Genomics Platform | Anke Wienecke-Baldacchino, Sibel Berger, Catherine Ragimbeau,Jessica Tapp, Fatu Djabi, Lise Pignon, Raoul Salmon,Virginie Jover, Elodie Solarino, Fatiha Boulmerka, Jacqueline Parmentier, Tamir Abdelrahman |
| hCoV-19/Brazil/SC-FIOCRUZ-66554/2021 | EPI_ISL_8797127 | 2021-12-20 | Laboratorio Central de Saude Publica do Estado de Santa Catarina (LACEN/SC) | Laboratory of Respiratory Viruses and Measles, Oswaldo Cruz Institute, FIOCRUZ | Paola Resende, Luciana Appolinario, Fernando Motta, Elisa Cavalcante Pereira, Ighor Arantes, Alice Sampaio Rocha, Bruna Mendon√ßa da Silva, J√©ssica Gra√ßa Macedo de Carvalho, Larissa Macedo Pinto, Victor Guimaraes, Darcita Buerger Rovaris, Sandra Bianchi |
| hCoV-19/SouthAfrica/NICD-N23983/2021 | EPI_ISL_8799131 | 2021-12-01 | ROB FERREIRA LABORATORY | National Institute for Communicable Diseases of the National Health Laboratory Service | Amoako DG, Everatt J, Kekana D, Scheepers C, Mohale T, Ntuli N, Mahlangu B, Mnguni A, Ismail A, Bhiman JN, Wolter N |
| hCoV-19/SouthAfrica/NICD-N24001/2021 | EPI_ISL_8799148 | 2021-11-30 | HELEN JOSEPH LABORATORY | National Institute for Communicable Diseases of the National Health Laboratory Service | Amoako DG, Everatt J, Kekana D, Scheepers C, Mohale T, Ntuli N, Mahlangu B, Mnguni A, Ismail A, Bhiman JN, Wolter N |
| hCoV-19/SouthAfrica/NICD-N24004/2021 | EPI_ISL_8799151 | 2021-11-30 | HELEN JOSEPH LABORATORY | National Institute for Communicable Diseases of the National Health Laboratory Service | Amoako DG, Everatt J, Kekana D, Scheepers C, Mohale T, Ntuli N, Mahlangu B, Mnguni A, Ismail A, Bhiman JN, Wolter N |
| hCoV-19/SouthAfrica/NICD-N24007/2021 | EPI_ISL_8799153 | 2021-11-30 | HELEN JOSEPH LABORATORY | National Institute for Communicable Diseases of the National Health Laboratory Service | Amoako DG, Everatt J, Kekana D, Scheepers C, Mohale T, Ntuli N, Mahlangu B, Mnguni A, Ismail A, Bhiman JN, Wolter N |
| hCoV-19/SouthAfrica/NICD-N24017/2021 | EPI_ISL_8799161 | 2021-11-30 | HELEN JOSEPH LABORATORY | National Institute for Communicable Diseases of the National Health Laboratory Service | Amoako DG, Everatt J, Kekana D, Scheepers C, Mohale T, Ntuli N, Mahlangu B, Mnguni A, Ismail A, Bhiman JN, Wolter N |
| hCoV-19/SouthAfrica/NICD-N24021/2021 | EPI_ISL_8799163 | 2021-11-30 | HELEN JOSEPH LABORATORY | National Institute for Communicable Diseases of the National Health Laboratory Service | Amoako DG, Everatt J, Kekana D, Scheepers C, Mohale T, Ntuli N, Mahlangu B, Mnguni A, Ismail A, Bhiman JN, Wolter N |
| hCoV-19/Mozambique/CERI-KRISP-K027205/2021 | EPI_ISL_8799680 | 2021-07-14 | Instituto Nacional de Saude (INS) | [illegible] and Innvoation, Stellenbosch University and KRISP, KZN Research Innovation and Sequencing Platform, UKZN. | Nalia Ismael, Nadia Sitoe, Paulo Arnaldo, Nedio Mabunda, Giandhari J, Pillay S, Naidoo Y, Ramphal U, Ramphal Y, Moir M, Van Wyk S, San JE, Tshiabuila D, Tegally H, Wilkinson E, de Oliveira T |
| hCoV-19/Bolivia/SAN-CABANA-PE-UPCH-2229/2021 | EPI_ISL_8800557 | 2021-06-14 | Laboratorio de Diagn√≥stico e Investigaci√≥n BIOSCIENCE SRL | Laboratorio de Gen√≥mica Microbiana, Universidad Peruana Cayetano Heredia | Esdenka P√©rez, Mauricio Prado, Freddy Tinajeros, Paula Carballo, Alejandra D√°vila-Barclay, Guillermo Salvatierra, Diego Cuicapuza, Janet Huancachoque, Pablo Tsukayama |

| | | | | | |
|---|---|---|---|---|---|
| hCoV-19/Bolivia/SAN-CABANA-PE-UPCH-2275/2021 | EPI_ISL_8800562 | 2021-09-16 | Laboratorio de Diagn√≥stico e Investigaci√≥n BIOSCIENCE SRL | Laboratorio de Gen√≥mica Microbiana, Universidad Peruana Cayetano Heredia | Esdenka P√©rez, Mauricio Prado, Freddy Tinajeros, Paula Carballo, Alejandra D√°vila-Barclay, Guillermo Salvatierra, Diego Cuicapuza, Janet Huancachoque, Pablo Tsukayama |
| hCoV-19/Bolivia/SAN-CABANA-PE-UPCH-2345/2021 | EPI_ISL_8800584 | 2021-11-05 | Laboratorio de Diagn√≥stico e Investigaci√≥n BIOSCIENCE SRL | Laboratorio de Gen√≥mica Microbiana, Universidad Peruana Cayetano Heredia | Esdenka P√©rez, Mauricio Prado, Freddy Tinajeros, Paula Carballo, Alejandra D√°vila-Barclay, Guillermo Salvatierra, Diego Cuicapuza, Janet Huancachoque, Pablo Tsukayama |
| hCoV-19/CzechRepublic/CSQ4732/2021 | EPI_ISL_8800893 | 2021-07-20 | Cytogeneticka laborator Brno | University Hospital Brno, CMBG | Bezdicek Matej, Svaton Jan, Kristyna Dufkova, Lengerova Martina, Dolejska Monika |
| hCoV-19/SouthAfrica/NICD-N25599/2021 | EPI_ISL_8801197 | 2021-12-04 | TSHEPONG LABORATORY | National Institute for Communicable Diseases of the National Health Laboratory Service | Amoako DG, Everatt J, Kekana D, Scheepers C, Mohale T, Ntuli N, Mahlangu B, Mnguni A, Ismail A, Bhiman JN, Wolter N |
| hCoV-19/SouthAfrica/NICD-N23929/2021 | EPI_ISL_8801248 | 2021-11-30 | ROB FERREIRA LABORATORY | National Institute for Communicable Diseases of the National Health Laboratory Service | Amoako DG, Everatt J, Kekana D, Scheepers C, Mohale T, Ntuli N, Mahlangu B, Mnguni A, Ismail A, Bhiman JN, Wolter N |
| hCoV-19/India/MH-NCCS-2748700556319/2021 | EPI_ISL_8803707 | 2021-08-02 | National Centre For Cell Science | National Centre For Cell Science ‚Äì INSACOG | Dhiraj Dhotre, Girish R Nair, Harshida S Ghode, Kartik J Deopujari, Avinash K Kshirsagar, Manoj Kumar Bhat, Ajay Pillai, INSACOG Consortium team, Yogesh Shouche |
| hCoV-19/India/GA-NCCS-3055200157065/2021 | EPI_ISL_8803715 | 2021-08-02 | National Centre For Cell Science | National Centre For Cell Science ‚Äì INSACOG | Dhiraj Dhotre, Girish R Nair, Harshida S Ghode, Kartik J Deopujari, Avinash K Kshirsagar, Manoj Kumar Bhat, Ajay Pillai, INSACOG Consortium team, Yogesh Shouche |
| hCoV-19/India/MH-NCCS-2748700557025/2021 | EPI_ISL_8803721 | 2021-08-02 | National Centre For Cell Science | National Centre For Cell Science ‚Äì INSACOG | Dhiraj Dhotre, Girish R Nair, Harshida S Ghode, Kartik J Deopujari, Avinash K Kshirsagar, Manoj Kumar Bhat, Ajay Pillai, INSACOG Consortium team, Yogesh Shouche |
| hCoV-19/India/GA-NCCS-C53804/2021 | EPI_ISL_8803968 | 2021-06-14 | National Centre For Cell Science | National Centre For Cell Science ‚Äì INSACOG | Dhiraj Dhotre, Girish R Nair, Harshida S Ghode, Kartik J Deopujari, Manoj Kumar Bhat, Ajay Pillai, INSACOG Consortium team, Yogesh Shouche |
| hCoV-19/Denmark/DCGC-314581/2021 | EPI_ISL_8805804 | 2021-12-30 | Department of Clinical Microbiology and Center for Genomic Medicine, Rigshospitalet, Copenhagen, Denmark | Statens Serum Institut Bioinformatics and Microbial Genomics | Danish Covid-19 Genome Consortium |
| hCoV-19/India/WB-IICB-S630-12/2021 | EPI_ISL_8806008 | 2021-12-01 | School of Tropical Medicine, Kolkata | CSIR-Indian Institute of Chemical Biology | Siddik Sarkar, Saikat Chakrabarti, Partha Chakrabarti, Poulomi Sarkar, Sarthak Banerjee, Arun Bandyopadhyay, INSACOG |
| hCoV-19/India/MH-NCCS-495904/2021 | EPI_ISL_8806949 | 2021-10-05 | National Centre For Cell Science | National Centre For Cell Science ‚Äì INSACOG | Dhiraj Dhotre, Girish R Nair, Harshida S Ghode, Kartik J Deopujari, Avinash K Kshirsagar, Manoj Kumar Bhat, Ajay Pillai, INSACOG Consortium team, Yogesh Shouche |
| hCoV-19/India/GA-NCCS-212690139/2021 | EPI_ISL_8806961 | 2021-09-26 | National Centre For Cell Science | National Centre For Cell Science ‚Äì INSACOG | Dhiraj Dhotre, Girish R Nair, Harshida S Ghode, Kartik J Deopujari, Avinash K Kshirsagar, Manoj Kumar Bhat, Ajay Pillai, INSACOG Consortium team, Yogesh Shouche |
| hCoV-19/India/GA-NCCS-3055200190658/2021 | EPI_ISL_8806994 | 2021-10-09 | National Centre For Cell Science | National Centre For Cell Science ‚Äì INSACOG | Dhiraj Dhotre, Girish R Nair, Harshida S Ghode, Kartik J Deopujari, Avinash K Kshirsagar, Manoj Kumar Bhat, Ajay Pillai, INSACOG Consortium team, Yogesh Shouche |

| | | | | | |
|---|---|---|---|---|---|
| hCoV-19/India/GA-NCCS-3055100258181/2021 | EPI_ISL_8807017 | 2021-10-09 | National Centre For Cell Science | National Centre For Cell Science ,Äì INSACOG | Dhiraj Dhotre, Girish R Nair, Harshida S Ghode, Kartik J Deopujari, Avinash K Kshirsagar, Manoj Kumar Bhat, Ajay Pillai, INSACOG Consortium team, Yogesh Shouche |
| hCoV-19/Finland/THL-202134790/2021 | EPI_ISL_8807657 | 2021-12-16 | SYNLAB Suomi | Expert Microbiology, National Institute for Health and Welfare | Soile Blomqvist, Jani Halkilahti, Kirsi Liitsola, Haider al-Hello, Pv§ivi Laurila, Erika Lindh, Teemu Smura, Ravi Kant, Phuoc Truong, Olli Vapalahti, Sari Hannula, Pekka Ellonen, Niina Ikonen, Carita Savolainen-Kopra |
| hCoV-19/USA/CA-CDC-ASC210562601/2021 | EPI_ISL_8815209 | 2021-12-29 | Aegis Sciences Corporation | Centers for Disease Control and Prevention Division of Viral Diseases, Pathogen Discovery | Dakota Howard,Dhwani Batra,Peter Cook,Jason Caravas,Benjamin Rambo-Martin,Scott Sammons,Yvette Unoarumhi,Matthew Schmerer,Kristine Lacek,Tymeckia Kendall,Victoria Caban Figueroa,Shatavia Morrison,Christopher Gulvick,Erisa Sula,Cyndi Clark,Patrick Campbell |
| hCoV-19/Indonesia/JB-GS-WJHL-ITB-WGS136/2020 | EPI_ISL_8825262 | 2020-11-12 | Dinas Kesehatan Kabupaten Purwakarta | West Java Health Laboratory; School of Life Sciences and Technology, Institut Teknologi Bandung | Azzania Fibriani, Miftahul Faridl, Karimatu Khoirunnisa, Ema Rahmawati, Ryan Bayusantika Ristandi, Rifky Waluyajati Rachman, Cut Nur Cinthia Alamanda, Rini Robiani, Karlina Mellyani |
| hCoV-19/Botswana/R66B35_BHP_362199714/2021 | EPI_ISL_8826406 | 2021-12-22 | Lancet Laboratories Gaborone | Botswana Harvard HIV Reference Laboratory | Sikhulile Moyo, Wonderful T. Choga, Dorcas Maruapula, Ellen Nkitse, Leyi Ncube, Mosana Moyo, Keoratile Ntshambiwa, Sefetogi Ramaologa, Thongbotho Mphoyakgosi, Boitumelo Zuze, Botshelo Radibe, Legodile Kooepile, Ontlametse T. Bareng, Pamela Smith-Lawrence, |
| hCoV-19/Botswana/R66B49_BHP_928643/2021 | EPI_ISL_8826414 | 2021-12-30 | National Health Laboratory | Botswana Harvard HIV Reference Laboratory | Sikhulile Moyo, Wonderful T. Choga, Dorcas Maruapula, Keoratile Ntshambiwa, Sefetogi Ramaologa, Thongbotho Mphoyakgosi, Boitumelo J. L Zuze, Botshelo Radibe, Legodile Kooepile, Ontlametse T. Bareng, Letsibogo Gaoraelwe, Thela Tefelo, Pamela Smith-Lawrenc |
| hCoV-19/Botswana/R66B92_BHP_925648/2021 | EPI_ISL_8826447 | 2021-12-29 | Botswana Harvard HIV Reference Laboratory | Botswana Harvard HIV Reference Laboratory | Sikhulile Moyo, Wonderful T. Choga, Dorcas Maruapula, Keoratile Ntshambiwa, Sefetogi Ramaologa, Thongbotho Mphoyakgosi, Boitumelo J. L Zuze, Botshelo Radibe, Legodile Kooepile, Ontlametse T. Bareng, Letsibogo Gaoraelwe, Thela Tefelo, Pamela Smith-Lawrenc |
| hCoV-19/USA/TX-CDC-ASC210572639/2021 | EPI_ISL_8827375 | 2021-12-28 | Aegis Sciences Corporation | Centers for Disease Control and Prevention Division of Viral Diseases, Pathogen Discovery | Dakota Howard,Dhwani Batra,Peter Cook,Jason Caravas,Benjamin Rambo-Martin,Scott Sammons,Yvette Unoarumhi,Matthew Schmerer,Kristine Lacek,Tymeckia Kendall,Victoria Caban Figueroa,Shatavia Morrison,Christopher Gulvick,Erisa Sula,Cyndi Clark,Patrick Campbell |
| hCoV-19/USA/CO-CDPHE-2102573767/2021 | EPI_ISL_8829286 | 2021-11-24 | Colorado Department of Public Health and Environment | Colorado Department of Public Health and Environment | Laura Bankers, Molly C. Hetherington-Rauth, Diana Ir, Alexandria Rossheim, Michael Martin, Mandy Waters, Arianna Smith, Shannon R. Matzinger, Emily A. Travanty |
| hCoV-19/USA/CO-CDPHE-2102573774/2021 | EPI_ISL_8829348 | 2021-11-23 | Colorado Department of Public Health and Environment | Colorado Department of Public Health and Environment | Laura Bankers, Molly C. Hetherington-Rauth, Diana Ir, Alexandria Rossheim, Michael Martin, Mandy Waters, Arianna Smith, Shannon R. Matzinger, Emily A. Travanty |

| | | | | | |
|---|---|---|---|---|---|
| hCoV-19/Thailand/DMSc-06059/2021 | EPI_ISL_8840714 | 2021-12-20 | Regional Medical Sciences Center 11/1 Phuket | Division of Genomic Medicine and Innovation support,Department of Medical Sciences, Ministry of Public Health, Thailand | Surakameth Mahasirimongkol,Nuanjun Wichukchinda,Archawin Rojanawiwat,Pilailuk Akkapaiboon Okada ,Waritta Sawaengdee,Penpitcha Thawong,Pundharika Piboonsiri,Jirapha Pakdee,Natthakul Bunneang |
| hCoV-19/Thailand/DMSc-06300/2021 | EPI_ISL_8840763 | 2021-12-26 | Regional Medical Sciences Center 7 Khon Kaen | Division of Genomic Medicine and Innovation support,Department of Medical Sciences, Ministry of Public Health, Thailand | Surakameth Mahasirimongkol,Nuanjun Wichukchinda,Archawin Rojanawiwat,Pilailuk Akkapaiboon Okada ,Waritta Sawaengdee,Penpitcha Thawong,Pundharika Piboonsiri,Jirapha Pakdee,Natthakul Bunneang |
| hCoV-19/Colombia/DC-SDS-1041250/2021 | EPI_ISL_8844711 | 2021-09-18 | Laboratorio de salud publica de Bogota | Gencore - Universidad de los Andes | Luisa Sacristan, David Gonzv°lez, Cristian Barrera, Gabriela Ariza, Mariana Restrepo, Carolina Valencia, Marcela Guevara, Silvia Restrepo, Johana Hernandez, Gabriela Delgado, Alejandro Gomez |
| hCoV-19/Gambia/87098/2021 | EPI_ISL_8845801 | 2021-11-10 | MRCG at LSHTM Genomics lab | MRCG at LSHTM Genomics lab | Abdul Karim sesay, Abdoulie Kanteh, Jarra Manneh, Mariama Kujabi, Bakary Sanyang,Dabiri Damilari, Yaya bah |
| hCoV-19/Canada/AB-ABPHL-48588/2021 | EPI_ISL_8846679 | 2021-12-26 | Edmonton Provincial Lab | Public Health Agency of Canada (PHAC) National Microbiology Laboratory | Buss, E, Croxen M, Deo A, Dieu P, Gill K, Ferrato C, Khan F, Koleva P, Li V, Lloyd C, Lynch T, Ma R, Murphy S, Pabbaraju K, Shokoples S, Tipples G, Thayer J, Whitehouse M, Wong A, Yu C, Zelyas N |
| hCoV-19/Wales/PHWC-PGA6C3/2021 | EPI_ISL_8851631 | 2021-12-28 | Originating lab: Wales Specialist Virology Centre Sequencing lab: Pathogen Genomics Unit | Public Health Wales Microbiology Cardiff Wales Specialist Virology Centre | Catherine Moore, Johnathan Evans, Laura Gifford, Malorie Perry, Simon Cottrell, Angela Marchbank, Alec Birchley, Alexander Adams, Amy Gaskin, Bree Gatica-Wilcox, Jason Coombes, Joel Southgate, Lauren Gilbert, Lee Graham, Nicole Pacchiarini, Sara Kumziene- |
| hCoV-19/Slovenia/17-034120-CE/2020 | EPI_ISL_8855479 | 2020-10-21 | National Laboratory for Health, Environment and Food, OMM, Celje | [illegible] Health, Environment and Food) / CISLD (Clinical Institute of Special Laboratory Diagnostics), University Children's Hospital, University Medical Center Ljubljana | Sandra Janezic, Aleksander Mahnic, Maja Rupnik, Tja≈°a ≈Ωohar ƒåretnik, Alenka ≈†torman, Nika Gobec, Aleksander Kocuvan, Kaja Tominc, Ma≈°a Jarƒçiƒç, Mateja Borinc, Da≈°a Kavka / Jernej Kovaƒç, Barbara Jenko Bizjan, Tine Tesovnik, Robert ≈†ket, Katarina K |
| hCoV-19/Slovenia/17-036839-CE/2020 | EPI_ISL_8855484 | 2020-10-26 | National Laboratory for Health, Environment and Food, OMM, Celje | [illegible] Health, Environment and Food) / CISLD (Clinical Institute of Special Laboratory Diagnostics), University Children's Hospital, University Medical Center Ljubljana | Sandra Janezic, Aleksander Mahnic, Maja Rupnik, Tja≈°a ≈Ωohar ƒåretnik, Alenka ≈†torman, Nika Gobec, Aleksander Kocuvan, Kaja Tominc, Ma≈°a Jarƒçiƒç, Mateja Borinc, Da≈°a Kavka / Jernej Kovaƒç, Barbara Jenko Bizjan, Tine Tesovnik, Robert ≈†ket, Katarina K |
| hCoV-19/Slovenia/17-037130-CE/2020 | EPI_ISL_8855485 | 2020-10-23 | National Laboratory for Health, Environment and Food, OMM, Celje | [illegible] Health, Environment and Food) / CISLD (Clinical Institute of Special Laboratory Diagnostics), University Children's Hospital, University Medical Center Ljubljana | Sandra Janezic, Aleksander Mahnic, Maja Rupnik, Tja≈°a ≈Ωohar ƒåretnik, Alenka ≈†torman, Nika Gobec, Aleksander Kocuvan, Kaja Tominc, Ma≈°a Jarƒçiƒç, Mateja Borinc, Da≈°a Kavka / Jernej Kovaƒç, Barbara Jenko Bizjan, Tine Tesovnik, Robert ≈†ket, Katarina K |
| hCoV-19/Slovenia/17-039558-CE/2020 | EPI_ISL_8855486 | 2020-10-29 | National Laboratory for Health, Environment and Food, OMM, Celje | [illegible] Health, Environment and Food) / CISLD (Clinical Institute of Special Laboratory Diagnostics), University Children's Hospital, University Medical Center Ljubljana | Sandra Janezic, Aleksander Mahnic, Maja Rupnik, Tja≈°a ≈Ωohar ƒåretnik, Alenka ≈†torman, Nika Gobec, Aleksander Kocuvan, Kaja Tominc, Ma≈°a Jarƒçiƒç, Mateja Borinc, Da≈°a Kavka / Jernej Kovaƒç, Barbara Jenko Bizjan, Tine Tesovnik, Robert ≈†ket, Katarina K |

| | | | | | |
|---|---|---|---|---|---|
| hCoV-19/Slovenia/17-042039-CE/2020 | EPI_ISL_8855488 | 2020-11-03 | National Laboratory for Health, Environment and Food, OMM, Celje | [illegible] Health, Environment and Food) / CISLD (Clinical Institute of Special Laboratory Diagnostics), University Children's Hospital, University Medical Center Ljubljana | Sandra Janezic, Aleksander Mahnic, Maja Rupnik, Tja≈°a ≈Ωohar ƒåretnik, Alenka ≈†torman, Nika Gobec, Aleksander Kocuvan, Kaja Tominc, Ma≈°a Jarƒçiƒç, Mateja Borinc, Da≈°a Kavka / Jernej Kovaƒç, Barbara Jenko Bizjan, Tine Tesovnik, Robert ≈†ket, Katarina K |
| hCoV-19/Slovenia/17-043868-CE/2020 | EPI_ISL_8855491 | 2020-11-06 | National Laboratory for Health, Environment and Food, OMM, Celje | [illegible] Health, Environment and Food) / CISLD (Clinical Institute of Special Laboratory Diagnostics), University Children's Hospital, University Medical Center Ljubljana | Sandra Janezic, Aleksander Mahnic, Maja Rupnik, Tja≈°a ≈Ωohar ƒåretnik, Alenka ≈†torman, Nika Gobec, Aleksander Kocuvan, Kaja Tominc, Ma≈°a Jarƒçiƒç, Mateja Borinc, Da≈°a Kavka / Jernej Kovaƒç, Barbara Jenko Bizjan, Tine Tesovnik, Robert ≈†ket, Katarina K |
| hCoV-19/PuertoRico/PR-CDC-FG-233175/2021 | EPI_ISL_8866444 | 2021-12-27 | Fulgent Genetics | Centers for Disease Control and Prevention Division of Viral Diseases, Pathogen Discovery | Dakota Howard,Dhwani Batra,Peter Cook,Jason Caravas,Benjamin Rambo-Martin,Scott Sammons,Yvette Unoarumhi,Matthew Schmerer,Kristine Lacek,Tymeckia Kendall,Victoria Caban Figueroa,Shatavia Morrison,Christopher Gulvick,Erisa Sula,Harry Gao,Mickey Li,John Gao |
| hCoV-19/USA/NY-PRL-2022_0104_05A09/2021 | EPI_ISL_8866786 | 2021-12-30 | Pandemic Response Lab - NYC | Pandemic Response Lab, R&D | Henry Lee, Michael Hammerling, Melissa Hopkins, Cybill del Castillo, Shinyoung Clair Kang, William Ward, Pradeep Bugga, Sol Rey, Dylan Law, Alex Carpio, Isabel Fernandez Escapa, Haiping Hao, Jon Laurent |
| hCoV-19/USA/FL-CDC-ASC210557907/2021 | EPI_ISL_8870629 | 2021-12-27 | Aegis Sciences Corporation | Centers for Disease Control and Prevention Division of Viral Diseases, Pathogen Discovery | Dakota Howard,Dhwani Batra,Peter Cook,Jason Caravas,Benjamin Rambo-Martin,Scott Sammons,Yvette Unoarumhi,Matthew Schmerer,Kristine Lacek,Tymeckia Kendall,Victoria Caban Figueroa,Shatavia Morrison,Christopher Gulvick,Erisa Sula,Cyndi Clark,Patrick Campbell |
| hCoV-19/USA/NY-CDC-ASC210559540/2021 | EPI_ISL_8870766 | 2021-12-27 | Aegis Sciences Corporation | Centers for Disease Control and Prevention Division of Viral Diseases, Pathogen Discovery | Dakota Howard,Dhwani Batra,Peter Cook,Jason Caravas,Benjamin Rambo-Martin,Scott Sammons,Yvette Unoarumhi,Matthew Schmerer,Kristine Lacek,Tymeckia Kendall,Victoria Caban Figueroa,Shatavia Morrison,Christopher Gulvick,Erisa Sula,Cyndi Clark,Patrick Campbell |
| hCoV-19/USA/CA-CDC-ASC210560068/2021 | EPI_ISL_8871840 | 2021-12-28 | Aegis Sciences Corporation | Centers for Disease Control and Prevention Division of Viral Diseases, Pathogen Discovery | Dakota Howard,Dhwani Batra,Peter Cook,Jason Caravas,Benjamin Rambo-Martin,Scott Sammons,Yvette Unoarumhi,Matthew Schmerer,Kristine Lacek,Tymeckia Kendall,Victoria Caban Figueroa,Shatavia Morrison,Christopher Gulvick,Erisa Sula,Cyndi Clark,Patrick Campbell |
| hCoV-19/USA/CO-CDC-ASC210560238/2021 | EPI_ISL_8872002 | 2021-12-28 | Aegis Sciences Corporation | Centers for Disease Control and Prevention Division of Viral Diseases, Pathogen Discovery | Dakota Howard,Dhwani Batra,Peter Cook,Jason Caravas,Benjamin Rambo-Martin,Scott Sammons,Yvette Unoarumhi,Matthew Schmerer,Kristine Lacek,Tymeckia Kendall,Victoria Caban Figueroa,Shatavia Morrison,Christopher Gulvick,Erisa Sula,Cyndi Clark,Patrick Campbell |

| | | | | | |
|---|---|---|---|---|---|
| hCoV-19/Colombia/CUN-UNIANDES-G033278/2021 | EPI_ISL_8877027 | 2021-12-21 | Gencore - Universidad de los Andes | Gencore - Universidad de los Andes | Luisa Sacristan, David Gonzv°lez, Cristian Barrera, Gabriela Ariza, Mariana Restrepo, Carolina Valencia, Marcela Guevara, Silvia Restrepo, Marcela Mercado |
| hCoV-19/Colombia/DC-UNIANDES-G033478/2021 | EPI_ISL_8877073 | 2021-12-24 | Gencore - Universidad de los Andes | Gencore - Universidad de los Andes | Luisa Sacristan, David Gonzv°lez, Cristian Barrera, Gabriela Ariza, Mariana Restrepo, Carolina Valencia, Marcela Guevara, Silvia Restrepo, Marcela Mercado |
| hCoV-19/Colombia/DC-LSPSDS-VG-1251625/2021 | EPI_ISL_8879529 | 2021-12-07 | Clinica los nogales | LSPSDS | Johana Hernandez Toloza, Marcela Castano Rodriguez,Gabriela Delgado Murcia, Alejandro Gomez Lopez, H√©ctor Alejandro Ruiz-Moreno, Marcela Mercado |
| hCoV-19/Brazil/SC-UFSC-48-83/2021 | EPI_ISL_8880801 | 2021-12-02 | LABORATORIO MUNICIPAL DE FLORIANOPOLIS - LAMUF | Laborat√≥rio de Bioinform√°tica - Universidade Federal de Santa Catarina | Glauber Wagner, Juliana Righetto Moser, Eric Kazuo Kawagoe, Milene Moehr de Moraes, Vilmar Benetti Filho, Dayane Azevedo Padilha, Fernando Hartmann Barazzetti, Marcos Andr√© Sch√∂rner, Doris Sobral Marques Souza, Ariane Nicaretta Amorim, Darcita Buerger R |
| hCoV-19/Peru/LIM-INS-12761/2021 | EPI_ISL_8882676 | 2021-12-24 | Laboratorio de Referencia Nacional de Virus Respiratorios. Centro Nacional de Salud Publica. Instituto Nacional de Salud Peru. | Laboratorio de Referencia Nacional de Virus Respiratorios. Centro Nacional de Salud Publica. Instituto Nacional de Salud Peru. | Carlos Padilla Rojas, Veronica Hurtado Vela, Orson Mestanza Millones, Iris Silva Molina, Luis Barcena Flores, Steve Acedo Lazo, Wendy Lizarraga Olivares, Victor Jimenez Vasquez, Princesa Medrano Alhuay, Alicia Nu√±ez Llanos, Kelly Izarra Rojas, Francisco |
| hCoV-19/India/GA-NCCS-VH78799/2021 | EPI_ISL_8882814 | 2021-06-29 | National Centre For Cell Science | National Centre For Cell Science ,Äì INSACOG | Dhiraj Dhotre, Girish R Nair, Harshida S Ghode, Kartik J Deopujari, Manoj Kumar Bhat, Ajay Pillai, INSACOG Consortium team, Yogesh Shouche |
| hCoV-19/USA/ID-IVREF-697930/2020 | EPI_ISL_8883176 | 2020-12-29 | Idaho Bureau of Laboratories | BVAMC, RD | R. Beukelman, Matthew Charles Burns, Aimee Ceniseros, Christian Loera, Robert L. Voermans, Christopher Ball, Mary Cloud Ammons, Jay Radke, Sarah Hobdey, Devin Bolz, Eric McIndoo, Hailey Burgoyne, Emily Price, Dennis Stevens |
| hCoV-19/USA/ID-IVREF-697988/2020 | EPI_ISL_8883179 | 2020-12-30 | Idaho Bureau of Laboratories | BVAMC, RD | R. Beukelman, Matthew Charles Burns, Aimee Ceniseros, Christian Loera, Robert L. Voermans, Christopher Ball, Mary Cloud Ammons, Jay Radke, Sarah Hobdey, Devin Bolz, Eric McIndoo, Hailey Burgoyne, Emily Price, Dennis Stevens |
| hCoV-19/India/PB-ICMR-NIV-INSACOG-560975098/2021 | EPI_ISL_8884408 | 2021-01-11 | Viral Research and Diagnostic Laboratories | NIV Influenza | Dr.Rupinder Bakshi |
| hCoV-19/India/PB-ICMR-NIV-INSACOG-600144988/2021 | EPI_ISL_8884441 | 2021-11-12 | Viral Research and Diagnostic Laboratories | NIV Influenza | Dr.Rupinder Bakshi |
| hCoV-19/England/PHEP-YYDKEHE/2021 | EPI_ISL_8884877 | 2021-03-21 | Respiratory Virus Unit, Microbiology Services Colindale, Public Health England | COVID-19 Genomics UK (COG-UK) Consortium | PHE Covid Sequencing Team |
| hCoV-19/SouthKorea/KDCA26106/2021 | EPI_ISL_8885871 | 2021-12-12 | Division of Emerging Infectious Diseases, Bureau of Infectious Diseases Diagnosis Control, Korea Disease Control and Prevention Agency | Diseases, Bureau of Infectious Diseases Diagnosis Control, Korea Disease Control and Prevention Agency | Jeong-Ah Kim, Ae Kyung Park, Il-Hwan Kim, Chae Young Lee, Hyuck Jin Lee, Eun-Jin Kim |

| | | | | | |
|---|---|---|---|---|---|
| hCoV-19/SouthKorea/KDCA26112/2021 | EPI_ISL_8885877 | 2021-12-14 | Division of Emerging Infectious Diseases, Bureau of Infectious Diseases Diagnosis Control, Korea Disease Control and Prevention Agency | Diseases, Bureau of Infectious Diseases Diagnosis Control, Korea Disease Control and Prevention Agency | Jeong-Ah Kim, Ae Kyung Park, Il-Hwan Kim, Chae Young Lee, Hyuck Jin Lee, Eun-Jin Kim |
| hCoV-19/SouthKorea/KDCA26126/2021 | EPI_ISL_8885891 | 2021-12-20 | Division of Emerging Infectious Diseases, Bureau of Infectious Diseases Diagnosis Control, Korea Disease Control and Prevention Agency | Diseases, Bureau of Infectious Diseases Diagnosis Control, Korea Disease Control and Prevention Agency | Jeong-Ah Kim, Ae Kyung Park, Il-Hwan Kim, Chae Young Lee, Hyuck Jin Lee, Eun-Jin Kim |
| hCoV-19/Gabon/7268L/2021 | EPI_ISL_8886396 | 2021-09-20 | Centre Interdisciplinaire de Recherches M√©dicales de Franceville (CIRMF) | Institut Hospitalo-Universitaire - M√©diterran√©e Infection | Sonia Etenna Lekana-Douki, Nadine N,Äôdilimabaka, Anthony Levasseur, Philippe Colson, Julia Cyrielle Andeko, Ornella Zong Minko, Octavie Banga Mve-Ella, Pierre-Edouard Fournier, Christian Devaux, Bertrand Mve Ondo, Falone Larissa Akombi, Laurianne Yacka M |
| hCoV-19/Indonesia/JT-Labkes-33A/2021 | EPI_ISL_8887863 | 2021-11-29 | Laboratorium Kesehatan Kota Semarang | Balai Labkes PAK Prov Jateng | Yekti, Purnomo, Dyah, Wiwik, Ismi, Sekti, Endah |
| hCoV-19/Indonesia/JT-Labkes-34A/2021 | EPI_ISL_8887864 | 2021-11-29 | Laboratorium Kesehatan Kota Semarang | Balai Labkes PAK Prov Jateng | Yekti, Purnomo, Dyah, Wiwik, Ismi, Sekti, Endah |
| hCoV-19/Indonesia/KB-NIHRD-WGS_22_00005/2021 | EPI_ISL_8889312 | 2021-12-23 | RS Universitas Tanjung Pura | National Institute of Health Research and Development | Vivi Setiawaty, Hana Apsari Pawestri, Subangkit, Kartika Dewi Puspa, Arie Ardiansyah Nugraha, Hartanti Dian Ikawati, Fajar Nur Sulistiyohadi, Nur Ika Hariastuti, Putri Widia, Nelly Puspandari |
| hCoV-19/Malawi/COM-SEQ6145/2020 | EPI_ISL_8889515 | 2020-06-22 | Kamuzu University of Health Sciences, College of Medicine, Department of Pathology | Kamuzu University of Health Sciences, College of Medicine, Department of Pathology | Tonney Nyirenda, Arox W. Kamng'ona, Chisomo Msefula, David Chaima, Chimwemwe Mhango, Jonathan Mandolo, Wanangwa Ndovie, Samuel Gwayi, Chrispin Chaguza, Benjamin Kumwenda, Alistair Darby, Kondwani C. Jambo, Victor Mwapasa, Khuzwayo C. Jere |
| hCoV-19/Peru/LIM-UPCH-1608/2020 | EPI_ISL_8889659 | 2020-09-14 | Instituto de Medicina Tropical Alexander von Humboldt, Universidad Peruana Cayetano Heredia | Laboratorio de Gen√≥mica Microbiana, Universidad Peruana Cayetano Heredia | Ericka Meza, Anthony Campos, Diego Cuicapuza, Alejandra D√°vila-Barclay, Guillermo Salvatierra, C√©sar Ugarte-Gil, Pablo Tsukayama |
| hCoV-19/Peru/LIM-UPCH-1984/2021 | EPI_ISL_8889969 | 2021-01-13 | Instituto de Medicina Tropical Alexander von Humboldt, Universidad Peruana Cayetano Heredia | Laboratorio de Gen√≥mica Microbiana, Universidad Peruana Cayetano Heredia | Ericka Meza, Anthony Campos, Diego Cuicapuza, Alejandra D√°vila-Barclay, Guillermo Salvatierra, C√©sar Ugarte-Gil, Pablo Tsukayama |
| hCoV-19/Brazil/SP-IB_SEQ01081/2021 | EPI_ISL_8890148 | 2021-09-16 | Unidade de Pronto Atendimento Zona Leste - Labmovel | Instituto Butantan | Dimas Tadeu Covas, Antonio Jorge Martins, Claudia Renata dos Santos Barros, David Schlesinger, Debora Botequio Moretti, Elaine Cristina Marqueze, Elaine Vieira Santos, Evandra Strazza Rodrigues, Heidge Fukumasu, Jayme Augusto de Souza-Neto, Jos√© Salvator |
| hCoV-19/Brazil/SP-IB_SEQ05490/2021 | EPI_ISL_8890330 | 2021-09-01 | UPA CENTRAL SANTOS - Labmovel | Instituto Butantan | Dimas Tadeu Covas, Antonio Jorge Martins, Claudia Renata dos Santos Barros, David Schlesinger, Debora Botequio Moretti, Elaine Cristina Marqueze, Elaine Vieira Santos, Evandra Strazza Rodrigues, Heidge Fukumasu, Jayme Augusto de Souza-Neto, Jos√© Salvator |

| | | | | | |
|---|---|---|---|---|---|
| hCoV-19/Brazil/SP-IB_SEQ05518/2021 | EPI_ISL_8890331 | 2021-09-01 | UPA CENTRAL SANTOS - Labmovel | Instituto Butantan | Dimas Tadeu Covas, Antonio Jorge Martins, Claudia Renata dos Santos Barros, David Schlesinger, Debora Botequio Moretti, Elaine Cristina Marqueze, Elaine Vieira Santos, Evandra Strazza Rodrigues, Heidge Fukumasu, Jayme Augusto de Souza-Neto, Josv© Salvator |
| hCoV-19/Brazil/SP-IB_SEQ07612/2021 | EPI_ISL_8890390 | 2021-09-14 | LABORATORIO MUNICIPAL DE ITANHAEM - Labmovel | Instituto Butantan | Dimas Tadeu Covas, Antonio Jorge Martins, Claudia Renata dos Santos Barros, David Schlesinger, Debora Botequio Moretti, Elaine Cristina Marqueze, Elaine Vieira Santos, Evandra Strazza Rodrigues, Heidge Fukumasu, Jayme Augusto de Souza-Neto, Josv© Salvator |
| hCoV-19/SouthAfrica/UFS-VIRO-NGS-ONT180122BC76/2021 | EPI_ISL_8890544 | 2021-12-25 | NHLS Universitas Academic | UFS Virology | PA Bester, MM Nyaga, P Nthiga, MT Mogotsi, Emmanuel Ogunbayo, D Goedhals, T de Oliveira |
| hCoV-19/SouthAfrica/UFS-VIRO-NGS-ONT180122BC12/2021 | EPI_ISL_8890599 | 2021-12-09 | NHLS Universitas Academic | UFS Virology | PA Bester, MM Nyaga, P Nthiga, MT Mogotsi, Emmanuel Ogunbayo, D Goedhals, T de Oliveira |
| hCoV-19/England/PHWC-PGAAIC/2021 | EPI_ISL_8892103 | 2021-12-28 | Originating lab: Wales Specialist Virology Centre Sequencing lab: Pathogen Genomics Unit | Public Health Wales Microbiology Cardiff Wales Specialist Virology Centre | Catherine Moore, Johnathan Evans, Laura Gifford, Malorie Perry, Simon Cottrell, Angela Marchbank, Alec Birchley, Alexander Adams, Amy Gaskin, Bree Gatica-Wilcox, Jason Coombes, Joel Southgate, Lauren Gilbert, Lee Graham, Nicole Pacchiarini, Sara Kumziene- |
| hCoV-19/Mauritius/NICD-N21990/2021 | EPI_ISL_8895410 | 2021-10-01 | Central Health Laboratory, Min of Health | Virology Department,Central Health Laboratory, Ministry of Health and Wellness | Ramuth M, Baboo SB,Ubheeram J, Jannoo N, Amoako DG, Everatt J, Kekana D, Scheepers C, Mohale T, Ntuli N, Mahlangu B, Mnguni A, Ismail A, Bhiman JN, Wolter N, Sonoo J |
| hCoV-19/Mauritius/NICD-N22009/2021 | EPI_ISL_8895417 | 2021-10-10 | NOVALAB | Virology Department,Central Health Laboratory, Ministry of Health and Wellness | Ramuth M,Leigh-Anne MC Intyre, Baboo SB,Ubheeram J, Jannoo N, Amoako DG, Everatt J, Kekana D, Scheepers C, Mohale T, Ntuli N, Mahlangu B, Mnguni A, Ismail A, Bhiman JN, Wolter N, Sonoo J |
| hCoV-19/Sweden/SE1410858855/2021 | EPI_ISL_8895729 | 2021-10-27 | Swedish national genomic surveillance program of SARS-CoV-2 | The Public Health Agency of Sweden | Maximilian Riess, Maria Lind Karlberg, Alma Brolund, Swedish national genomic surveillance program of SARS-CoV-2 |
| hCoV-19/USA/ID-IVREF-658492/2020 | EPI_ISL_8900632 | 2020-09-16 | Idaho Bureau of Laboratories | BVAMC, RD | R. Beukelman, Matthew Charles Burns, Aimee Ceniseros, Christian Loera, Robert L. Voermans, Christopher Ball, Mary Cloud Ammons, Jay Radke, Sarah Hobdey, Devin Bolz, Eric McIndoo, Hailey Burgoyne, Emily Price, Dennis Stevens |
| hCoV-19/Australia/NSW-ICPMR-19057/2021 | EPI_ISL_8900972 | 2021-12-30 | Pathology West - NSW Health Pathology | Clinical Pathology and Medical Research; Westmead Hospital; University of Sydney | Arnott A., Draper J., Gall M., Martinez E., Rockett R., Sintchenko V., on behalf of ICPMR |
| hCoV-19/Canada/BC-BCCDC-312101/2021 | EPI_ISL_8903728 | 2021-12-26 | British Columbia Centre For Disease Control | BCCDC Public Health Laboratory | Prystajecky Natalie, Linda Hoang, Dan Fornika, John Tyson, Shannon Russell, Kim Macdonald, Kimia Kamelian, James Zlosnik, Ana Pacagnella, Corrinne Ng, Loretta Janz, Robert Azana, Mel Krajden |

| | | | | | |
|---|---|---|---|---|---|
| hCoV-19/Russia/MOW-CSP-CK-275_S44/2020 | EPI_ISL_8904475 | 2020-10-04 | Chief Federal State Budgetary Healthcare Institution ,ÄúCentre of Hygiene and Epidemiology,Äù of the Federal Medical Biological Agency, Central laboratory of microbiological and clinical diagnostic tests | Federal State Budgetary Institution ,ÄúCentre for Strategic Planning and Management of Biomedical Health Risks,Äù of the Federal Medical Biological Agency, Laboratory for advanced molecular diagnostics of human diseases. | Mikhaylov, Ivan M.; Matsvay, Alina; Ayginin, Andrey A.; Say, Anna; Zakamornaya, Anastasiya I.; Khakhina, Anastasiya O.; Lisitsa, Tatiana S.; Abramov, Ivan S.; Kolbutova, Kseniya; Oleynikova, Daria; Avdeenko, Tatiana; Bogdan, Sergey; Shipulin, German A.; Y |
| hCoV-19/Russia/MOW-CSP-KOM-0135_S21/2020 | EPI_ISL_8904735 | 2020-09-08 | Chief Federal State Budgetary Healthcare Institution ,ÄúCentre of Hygiene and Epidemiology,Äù of the Federal Medical Biological Agency, Central laboratory of microbiological and clinical diagnostic tests | Federal State Budgetary Institution ,ÄúCentre for Strategic Planning and Management of Biomedical Health Risks,Äù of the Federal Medical Biological Agency, Laboratory for advanced molecular diagnostics of human diseases. | Mikhaylov, Ivan M.; Matsvay, Alina; Ayginin, Andrey A.; Say, Anna; Zakamornaya, Anastasiya I.; Khakhina, Anastasiya O.; Lisitsa, Tatiana S.; Abramov, Ivan S.; Kolbutova, Kseniya; Oleynikova, Daria; Avdeenko, Tatiana; Bogdan, Sergey; Shipulin, German A.; Y |
| hCoV-19/Canada/BC-BCCDC-309505/2021 | EPI_ISL_8911634 | 2021-12-23 | British Columbia Centre For Disease Control | BCCDC Public Health Laboratory | Prystajecky Natalie, Linda Hoang, Dan Fornika, John Tyson, Shannon Russell, Kim Macdonald, Kimia Kamelian, James Zlosnik, Ana Pacagnella, Corrinne Ng, Loretta Janz, Robert Azana, Mel Krajden |
| hCoV-19/Canada/BC-BCCDC-316421/2021 | EPI_ISL_8914749 | 2021-12-27 | British Columbia Centre For Disease Control | BCCDC Public Health Laboratory | Prystajecky Natalie, Linda Hoang, Dan Fornika, John Tyson, Shannon Russell, Kim Macdonald, Kimia Kamelian, James Zlosnik, Ana Pacagnella, Corrinne Ng, Loretta Janz, Robert Azana, Mel Krajden |
| hCoV-19/Canada/BC-BCCDC-316497/2021 | EPI_ISL_8914814 | 2021-12-29 | British Columbia Centre For Disease Control | BCCDC Public Health Laboratory | Prystajecky Natalie, Linda Hoang, Dan Fornika, John Tyson, Shannon Russell, Kim Macdonald, Kimia Kamelian, James Zlosnik, Ana Pacagnella, Corrinne Ng, Loretta Janz, Robert Azana, Mel Krajden |
| hCoV-19/Canada/BC-BCCDC-316700/2021 | EPI_ISL_8914930 | 2021-12-28 | British Columbia Centre For Disease Control | BCCDC Public Health Laboratory | Prystajecky Natalie, Linda Hoang, Dan Fornika, John Tyson, Shannon Russell, Kim Macdonald, Kimia Kamelian, James Zlosnik, Ana Pacagnella, Corrinne Ng, Loretta Janz, Robert Azana, Mel Krajden |
| hCoV-19/Canada/BC-BCCDC-316765/2021 | EPI_ISL_8914965 | 2021-12-27 | British Columbia Centre For Disease Control | BCCDC Public Health Laboratory | Prystajecky Natalie, Linda Hoang, Dan Fornika, John Tyson, Shannon Russell, Kim Macdonald, Kimia Kamelian, James Zlosnik, Ana Pacagnella, Corrinne Ng, Loretta Janz, Robert Azana, Mel Krajden |
| hCoV-19/Canada/BC-BCCDC-316810/2021 | EPI_ISL_8914993 | 2021-12-29 | British Columbia Centre For Disease Control | BCCDC Public Health Laboratory | Prystajecky Natalie, Linda Hoang, Dan Fornika, John Tyson, Shannon Russell, Kim Macdonald, Kimia Kamelian, James Zlosnik, Ana Pacagnella, Corrinne Ng, Loretta Janz, Robert Azana, Mel Krajden |
| hCoV-19/Morocco/40CC/2021 | EPI_ISL_8920489 | 2021-07-13 | Laboratoire de Recherche et d'Analyses M√©dicales de la Gendarmerie Royale | Laboratoire de Recherche et d'Analyses M√©dicales de la Gendarmerie Royale | Amal SOUIRI, Sana√¢ LEMRISS, Mohamed Mouatakid,Hajar LEMRISS, Mohamed LABIOUI, Nabil LEMZAOUI, Sa√¢d EL KABBAJ |
| hCoV-19/Myanmar/DSMRC-063/2021 | EPI_ISL_8920492 | 2021-11-30 | DSMRC | Defence Services Medical Research Centre (DSMRC) | Khine Zaw Oo, Zaw Win Htun, Kyaw Zawl Lin, Aung Sitt Hmue, Zaw Myo Lwin, Ko Ko Win, Kyee Myint, Ko Ko Lwin |
| hCoV-19/Myanmar/DSMRC-064/2021 | EPI_ISL_8920493 | 2021-11-30 | DSMRC | Defence Services Medical Research Centre (DSMRC) | Khine Zaw Oo, Zaw Win Htun, Kyaw Zawl Lin, Aung Sitt Hmue, Zaw Myo Lwin, Ko Ko Win, Kyee Myint, Ko Ko Lwin |
| hCoV-19/Italy/APU-IZSPB-PT5669/2021 | EPI_ISL_8920739 | 2021-10-07 | Ospedale Vito Fazzi | Istituto Zooprofilattico Sperimentale della Puglia e della Basilicata | Parisi A., Bianco A., Capozzi L., Del Sambro L., Simone D., Barbieri G., Quarato C., Ridolfi D., Giannico A., Lobreglio G. |

| | | | | | |
|---|---|---|---|---|---|
| hCoV-19/Ecuador/NIC-INSPI-0813-824/2021 | EPI_ISL_8920916 | 2021-12-29 | INSPI-CRN DE INFLUENZA Y OTROS VIRUS RESPIRATORIOS | INSPI-CRN DE INFLUENZA Y OTROS VIRUS RESPIRATORIOS | Alfredo Bruno, Maritza Olmedo, Michelle P√°ez, Jimmy Garc√©s, Fernando Llerena, Diego Morales, Ruben Armas |
| hCoV-19/England/PHEC-YYB5OEE/2021 | EPI_ISL_8923894 | 2021-12-28 | Respiratory Virus Unit, Microbiology Services Colindale, Public Health England | COVID-19 Genomics UK (COG-UK) Consortium | PHE Covid Sequencing Team |
| hCoV-19/USA/NY-UB-CHS-02167/2021 | EPI_ISL_8924319 | 2021-12-07 | Catholic Health Laboratories | University at Buffalo Genomics and Bioinformatics Core | Jonathan Bard, Natalie Lamb, Alyssa Pohlman, Brandon Marzullo, Amanda Boccolucci, Norma Nowak, Donald Yergeau, Jennifer Surtees |
| hCoV-19/Canada/QC-1nFWU-5278110/2021 | EPI_ISL_8924777 | 2021-12-03 | Laboratoire de sant√© publique du Qu√©bec | Laboratoire de sant√© publique du Qu√©bec | Sandrine Moreira, Ioannis Ragoussis, Guillaume Bourque, Jesse Shapiro, Mark Lathrop and Judith Fafard on behalf of the CoVSeQ research group |
| hCoV-19/USA/OH-IVY-ZZX9KOXB/2021 | EPI_ISL_8937072 | 2021-12-20 | IVY3 Central Lab, Vanderbilt University Medical Center | Department of Microbiology and Immunology | Gilbert |
| hCoV-19/Argentina/PAIS-G0755/2021 | EPI_ISL_8938334 | 2021-12-18 | Laboratorio Central, Ministerio de Salud C√≥rdoba | [illegible] Medicina and Instituto de Patologia Vegetal (IPAVE-CIAP-INTA) on behalf of 'Proyecto Argentino Interinstitucional de genomica de SARS-CoV-2' (PAIS Consortium) | Zeballos, M.; Lucca, A.; Fern√°ndez, F.D.; Marquez, N.; Debat, H., Goya, S.; Re, V.; Pisano, M.B.; Castro, G.; Sicilia, P.; Barbas, G. |
| hCoV-19/USA/MN-CDC-IBX393467659555/2021 | EPI_ISL_8946780 | 2021-12-28 | Infinity Biologix | Centers for Disease Control and Prevention Division of Viral Diseases, Pathogen Discovery | Dakota Howard,Dhwani Batra,Peter Cook,Jason Caravas,Benjamin Rambo-Martin,Scott Sammons,Yvette Unoarumhi,Matthew Schmerer,Kristine Lacek,Tymeckia Kendall,Victoria Caban Figueroa,Shatavia Morrison,Christopher Gulvick,Erisa Sula,Christian Bixby,Yihe Wang,Jo |
| hCoV-19/USA/FL-CDC-ASC210500751/2021 | EPI_ISL_8950351 | 2021-12-30 | Aegis Sciences Corporation | Centers for Disease Control and Prevention Division of Viral Diseases, Pathogen Discovery | Dakota Howard,Dhwani Batra,Peter Cook,Jason Caravas,Benjamin Rambo-Martin,Scott Sammons,Yvette Unoarumhi,Matthew Schmerer,Kristine Lacek,Tymeckia Kendall,Victoria Caban Figueroa,Shatavia Morrison,Christopher Gulvick,Erisa Sula,Cyndi Clark,Patrick Campbell |
| hCoV-19/USA/UT-UPHL-220119698550/2021 | EPI_ISL_8950921 | 2021-12-22 | Utah Public Health Laboratory | Utah Public Health Laboratory | Erin L. Young, Kelly F. Oakeson, Pooja Gupta, John Arnn, Tom Iverson, Olinto Linares-Perdomo |
| hCoV-19/USA/FL-CDC-ASC210501126/2021 | EPI_ISL_8951364 | 2021-12-30 | Aegis Sciences Corporation | Centers for Disease Control and Prevention Division of Viral Diseases, Pathogen Discovery | Dakota Howard,Dhwani Batra,Peter Cook,Jason Caravas,Benjamin Rambo-Martin,Scott Sammons,Yvette Unoarumhi,Matthew Schmerer,Kristine Lacek,Tymeckia Kendall,Victoria Caban Figueroa,Shatavia Morrison,Christopher Gulvick,Erisa Sula,Cyndi Clark,Patrick Campbell |
| hCoV-19/USA/CO-CDC-MMB12427410/2021 | EPI_ISL_8952874 | 2021-12-24 | Mako Medical | Centers for Disease Control and Prevention Division of Viral Diseases, Pathogen Discovery | Dakota Howard,Dhwani Batra,Peter Cook,Jason Caravas,Benjamin Rambo-Martin,Scott Sammons,Yvette Unoarumhi,Matthew Schmerer,Kristine Lacek,Tymeckia Kendall,Victoria Caban Figueroa,Shatavia Morrison,Christopher Gulvick,Erisa Sula,Matthew Tugwell,Lauren Moon, |

| | | | | | |
|---|---|---|---|---|---|
| hCoV-19/USA/MI-CDC-QDX32535277/2021 | EPI_ISL_8953151 | 2021-12-27 | Quest Diagnostics Incorporated | Centers for Disease Control and Prevention Division of Viral Diseases, Pathogen Discovery | Dakota Howard,Dhwani Batra,Peter Cook,Jason Caravas,Benjamin Rambo-Martin,Scott Sammons,Yvette Unoarumhi,Matthew Schmerer,Kristine Lacek,Tymeckia Kendall,Victoria Caban Figueroa,Shatavia Morrison,Christopher Gulvick,Erisa Sula,S. Rosenthal,A. Gerasimova,R |
| hCoV-19/Guam/GU-CDC-2-5400803/2021 | EPI_ISL_8954061 | 2021-12-03 | Guam Public Health Laboratory | Centers for Disease Control and Prevention Division of Viral Diseases, Pathogen Discovery | Mili Sheth,Sarah Nobles,Joseph Madden,Jasmine Padilla,Alex Burgin,Meghan Bentz,Mark Burroughs,Peter Cook,Clinton Paden,Dhwani Batra,Kristen Knipe,Dakota Howard,Yvette Unoarumhi,Matthew Schmerer,Ben Rambo-Martin,Kristine Lacek,Sam Shepard,Dave Wentworth,Vi |
| hCoV-19/SouthAfrica/CERI-KRISP-K033162/2021 | EPI_ISL_8957606 | 2021-11-29 | National Health Laboratory Service, Kwazulu-Natal, South Africa | and Innvoation, Stellenbosch University and KRISP, KZN Research Innovation and Sequencing Platform, UKZN. | Nokukhanya Mdlalose, Giandhari J, Pillay S, Naidoo Y, Ramphal U, Ramphal Y, Arisha Maharaj, San JE, Tshiabuila D, Tegally H, Wilkinson E, de Oliveira T |
| hCoV-19/Scotland/SCOT-2675/2021 | EPI_ISL_8963910 | 2021-12-28 | West of Scotland Specialist Virology Centre, NHSGGC / MRC-University of Glasgow Centre for Virus Research | COVID-19 Genomics UK (COG-UK) Consortium | Ana da Silva Filipe, Kathy Smollett, Daniel Mair, Lily Tong, Jessica Benkaroun, Nicolas Suarez, Jenna Nichols, Kyriaki Nomikou; Sarah McDonald; Alasdair MacLean, Emily Goldstein, Rachael Tomb, Rory Gunson; Richard Orton, Joseph Hughes, Sreenu Vattipally, |
| hCoV-19/SouthAfrica/NHLS-UCT-GP-N329/2021 | EPI_ISL_8975005 | 2021-12-17 | New Somerset Hospital wc NSH | NHLS/UCT | Arash Iranzadeh, Deelan Doolabh, Lynn Tyers, Bruna Galvao, Innocent Mudau, Marvin Hsiao, Gert Marais, Diana Hardie, Stephen Korsman, Rageema Joseph, Luicer Olubayo, Nokuzola Mbhele, Carolyn Williamson |
| hCoV-19/Brazil/MS-GD93/2020 | EPI_ISL_8975208 | 2020-11-20 | Health Science Research Laboratory | Fundaâ√ß√£o Ezequiel Dias | La√≠s Albuquerque de Oliveira, Vinicius Navarini, Silvana Beutinger Marchioro, Alex Jos√© Leite Torres, Julio Croda, Simone Simionatto, Joilson Xavier, Emerson de Castro, Vagner Fonseca, Flavia Aburjaile, Hegger Fritsch, Felipe Iani, Talita Adelino, Marta |
| hCoV-19/Vietnam/QNa2021-123/2021 | EPI_ISL_8979247 | 2021-12-23 | CDC Quang Nam, Vietnam | Department of Microbiology and Immunology | Trinh Hoang Long |
| hCoV-19/India/KL-GENESCOV08985/2021 | EPI_ISL_8985442 | 2021-11-16 | INSACOG-IGIB and Kerala State SARS-COV-2 Genome Surveillance Programme (GENESCOV2) | INSACOG-IGIB and Kerala State SARS-COV-2 Genome Surveillance Programme (GENESCOV2) | INSACOG-IGIB and Kerala State SARS-COV-2 Genome Surveillance Programme (GENESCOV2) |
| hCoV-19/England/LOND-YYBY9H8/2021 | EPI_ISL_8989562 | 2021-12-28 | University College London, Great Ormond Street Hospital for Children NHS Foundation Trust, Imperial College Healthcare NHS Trust | COVID-19 Genomics UK (COG-UK) Consortium | Sergi Castellano, Rachel Williams, Mark Kristiansen, Sunando Roy, Charlotte Williams, Nadua Bayzid, Marius Cotic, Helena Tutill, Patricia Dyal, Judith Breuer |
| hCoV-19/Botswana/R69B79_BHP_916378/2021 | EPI_ISL_9002845 | 2021-12-23 | National Health Laboratory | Botswana Harvard HIV Reference Laboratory | Sikhulile Moyo, Wonderful T. Choga, Dorcas Maruapula, Keoratile Ntshambiwa, Sefetogi Ramaologa, Thongbotho Mphoyakgosi, Boitumelo Zuze, Botshelo Radibe, Legodile Kooepile, Ontlametse T. Bareng, Pamela Smith-Lawrence, Kgomotso Moruisi, Roger Shapiro, Shahi |
| hCoV-19/Vietnam/DN2021-149/2021 | EPI_ISL_9003863 | 2021-12-08 | CDC Da Nang, Vietnam | Department of Microbiology and Immunology, Pasteur Institute in Nha Trang | Trinh Hoang Long, Nguyen Duc Duy, Nguyen Bao Trieu, Huynh Kim Mai, Vu Thi Ngoc, Hoang Tien Thanh, Nguyen Dinh LΔ∞Δ°ng, Do Thai Hung |

| | | | | | |
|---|---|---|---|---|---|
| hCoV-19/Mauritius/NICD-N18815/2021 | EPI_ISL_9005323 | 2021-10-03 | Airport Health Laboratory | Central Health Laboratory | Ramuth M, Patoo M, Ubheeram J Baboo SB,Mathur H, Sujeewon C, Jannoo N, Amoako DG, Everatt J, Kekana D, Scheepers C, Mohale T, Ntuli N, Mahlangu B, Mnguni A, Ismail A, Bhiman JN, Wolter N, Sonoo J |
| hCoV-19/Mauritius/NICD-N18816/2021 | EPI_ISL_9005324 | 2021-10-03 | Airport Health Laboratory | Central Health Laboratory | Ramuth M, Patoo M, Ubheeram J Baboo SB,Mathur H, Sujeewon C, Jannoo N, Amoako DG, Everatt J, Kekana D, Scheepers C, Mohale T, Ntuli N, Mahlangu B, Mnguni A, Ismail A, Bhiman JN, Wolter N, Sonoo J |
| hCoV-19/India/AS-INSACOG-1828000607577/2021 | EPI_ISL_9017605 | 2021-11-02 | INSACOG-Assam | National¬†Institute¬†of¬†Biomedical ¬†Genomics ,Äì¬†INSACOG | Arindam Maitra, Ajanta Sharma, Nidhan Kumar Biswas, Lahari Saikia, Sreedhar Chinnaswamy,¬†Kailash Chamuah, Saumitra Das |
| hCoV-19/Morocco/14CC/2021 | EPI_ISL_9019543 | 2021-07-12 | Laboratoire de Recherche et d'Analyses M√©dicales de la Gendarmerie Royale | Laboratoire de Recherche et d'Analyses M√©dicales de la Gendarmerie Royale | Sana√¢ LEMRISS, Amal SOUIRI, Mohamed Mouatakid, Hajar LEMRISS, Mohamed LABIOUI, Nabil LEMZAOUI, Sa√¢d EL KABBAJ |
| hCoV-19/Morocco/RA6/2021 | EPI_ISL_9020397 | 2021-12-03 | Laboratoire de Recherche et d'Analyses M√©dicales de la Gendarmerie Royale | d'Analyses M√©dicales de la Gendarmerie Royale | Sana√¢ LEMRISS, Amal SOUIRI, Hajar LEMRISS, Mohamed LABIOUI, Nabil LEMZAOUI, Sa√¢d EL KABBAJ |
| hCoV-19/Thailand/NIC_BKK_SEQ23886/2021 | EPI_ISL_9020979 | 2021-12-25 | Paolo Hospital Phahonyothin | National Institute of Health, Department of Medical Sciences, Ministry of Public Health, Thailand | Pilailuk Okada; Siripaporn Phuygun; Nuttida Thongpramul; Pakorn Piromtong; Natchaya¬† Khiadsang; Thanutsapa Thanadachakul; Sirikanda Wimol; Suratchana Mitrat; Sunthareeya Waicharoen; Archawin Rojanawiwat |
| hCoV-19/Thailand/NIC_RET_SEQ24275/2021 | EPI_ISL_9021066 | 2021-12-23 | Regional Medical Sciences Center 7 Khon Kaen | National Institute of Health, Department of Medical Sciences, Ministry of Public Health, Thailand | Pilailuk Okada; Siripaporn Phuygun; Nuttida Thongpramul; Pakorn Piromtong; Natchaya¬† Khiadsang; Thanutsapa Thanadachakul; Sirikanda Wimol; Suratchana Mitrat; Sunthareeya Waicharoen; Archawin Rojanawiwat |
| hCoV-19/Thailand/NIC_RET_SEQ24282/2021 | EPI_ISL_9021073 | 2021-12-23 | Regional Medical Sciences Center 7 Khon Kaen | National Institute of Health, Department of Medical Sciences, Ministry of Public Health, Thailand | Pilailuk Okada; Siripaporn Phuygun; Nuttida Thongpramul; Pakorn Piromtong; Natchaya¬† Khiadsang; Thanutsapa Thanadachakul; Sirikanda Wimol; Suratchana Mitrat; Sunthareeya Waicharoen; Archawin Rojanawiwat |
| hCoV-19/Thailand/NIC_RET_SEQ24288/2021 | EPI_ISL_9021079 | 2021-12-24 | Regional Medical Sciences Center 7 Khon Kaen | National Institute of Health, Department of Medical Sciences, Ministry of Public Health, Thailand | Pilailuk Okada; Siripaporn Phuygun; Nuttida Thongpramul; Pakorn Piromtong; Natchaya¬† Khiadsang; Thanutsapa Thanadachakul; Sirikanda Wimol; Suratchana Mitrat; Sunthareeya Waicharoen; Archawin Rojanawiwat |
| hCoV-19/Qatar/QA-QU-18-34-C3/2021 | EPI_ISL_9028471 | 2021-12-29 | Ministry of Public Health / Hamad Medical Corporation | Biomedical Research Center (BRC), Qatar University / Qatar Genome Project (QGP) | BRC:¬†Heba A. Al-Khatib, Fatiha M. Benslimane,Maria K. Smatti, Hadeel T. Mohammed, Fatma Hassan Ali, Swapna Thomas, Ola Al-Jamal, Dana Al-Batsh, Hadi M. Yassine, Asmaa A. Al-Thani. MOPH and HMC:¬†Abdullatif Al-Khal, Muna A. S. Al-Maslamani, Mashael A. Al- |
| hCoV-19/Qatar/QA-QU-18-34-H1/2021 | EPI_ISL_9028486 | 2021-12-26 | Ministry of Public Health / Hamad Medical Corporation | Biomedical Research Center (BRC), Qatar University / Qatar Genome Project (QGP) | BRC:¬†Heba A. Al-Khatib, Fatiha M. Benslimane,Maria K. Smatti, Hadeel T. Mohammed, Fatma Hassan Ali, Swapna Thomas, Ola Al-Jamal, Dana Al-Batsh, Hadi M. Yassine, Asmaa A. Al-Thani. MOPH and HMC:¬†Abdullatif Al-Khal, Muna A. S. Al-Maslamani, Mashael A. Al- |

| | | | | | |
|---|---|---|---|---|---|
| hCoV-19/Wales/PHWC-PG9AF9/2021 | EPI_ISL_9032473 | 2021-12-30 | Originating lab: Wales Specialist Virology Centre Sequencing lab: Pathogen Genomics Unit | Public Health Wales Microbiology Cardiff Wales Specialist Virology Centre | Catherine Moore, Johnathan Evans, Laura Gifford, Malorie Perry, Simon Cottrell, Angela Marchbank, Alec Birchley, Alexander Adams, Amy Gaskin, Bree Gatica-Wilcox, Jason Coombes, Joel Southgate, Lauren Gilbert, Lee Graham, Nicole Pacchiarini, Sara Kumziene- |
| hCoV-19/USA/UT-CDC-LC0459491/2021 | EPI_ISL_9036997 | 2021-12-30 | Laboratory Corporation of America | Centers for Disease Control and Prevention Division of Viral Diseases, Pathogen Discovery | Dakota Howard,Dhwani Batra,Peter Cook,Jason Caravas,Benjamin Rambo-Martin,Scott Sammons,Yvette Unoarumhi,Matthew Schmerer,Kristine Lacek,Tymeckia Kendall,Victoria Caban Figueroa,Shatavia Morrison,Christopher Gulvick,Minoo Agarwal,Eyad Almasri,Debbie Boles |
| hCoV-19/USA/AZ-TG1168954/2021 | EPI_ISL_9040190 | 2021-11-07 | TGen North | TGen North | Jolene Bowers, Heather Centner, Brett Van Tassel, Chris French, Hayley Yaglom, Darrin Lemmer, Dave Engelthaler |
| hCoV-19/USA/AZ-TG1169465/2021 | EPI_ISL_9040656 | 2021-12-22 | TGen North | TGen North | Jolene Bowers, Heather Centner, Brett Van Tassel, Chris French, Hayley Yaglom, Darrin Lemmer, Dave Engelthaler |
| hCoV-19/Peru/LIM-INS-12919/2021 | EPI_ISL_9046210 | 2021-11-28 | Laboratorio de Referencia Nacional de Virus Respiratorios. Centro Nacional de Salud Publica. Instituto Nacional de Salud Peru. | Laboratorio de Referencia Nacional de Virus Respiratorios. Centro Nacional de Salud Publica. Instituto Nacional de Salud Peru. | Carlos Padilla Rojas, Veronica Hurtado Vela, Orson Mestanza Millones, Iris Silva Molina, Luis Barcena Flores, Steve Acedo Lazo, Wendy Lizarraga Olivares, Victor Jimenez Vasquez, Princesa Medrano Alhuay, Alicia Nu√±ez Llanos, Kelly Izarra Rojas, Francisco |
| hCoV-19/Peru/ANC-INS-12916/2021 | EPI_ISL_9046219 | 2021-12-29 | Laboratorio de Referencia Nacional de Virus Respiratorios. Centro Nacional de Salud Publica. Instituto Nacional de Salud Peru. | Laboratorio de Referencia Nacional de Virus Respiratorios. Centro Nacional de Salud Publica. Instituto Nacional de Salud Peru. | Carlos Padilla Rojas, Veronica Hurtado Vela, Orson Mestanza Millones, Iris Silva Molina, Luis Barcena Flores, Steve Acedo Lazo, Wendy Lizarraga Olivares, Victor Jimenez Vasquez, Princesa Medrano Alhuay, Alicia Nu√±ez Llanos, Kelly Izarra Rojas, Francisco |
| hCoV-19/Peru/MOQ-INS-12797/2021 | EPI_ISL_9046299 | 2021-12-20 | Laboratorio de Referencia Nacional de Virus Respiratorios. Centro Nacional de Salud Publica. Instituto Nacional de Salud Peru. | Laboratorio de Referencia Nacional de Virus Respiratorios. Centro Nacional de Salud Publica. Instituto Nacional de Salud Peru. | Carlos Padilla Rojas, Veronica Hurtado Vela, Orson Mestanza Millones, Iris Silva Molina, Luis Barcena Flores, Steve Acedo Lazo, Wendy Lizarraga Olivares, Victor Jimenez Vasquez, Princesa Medrano Alhuay, Alicia Nu√±ez Llanos, Kelly Izarra Rojas, Francisco |
| hCoV-19/Egypt/CPHL-EGY21020/2021 | EPI_ISL_9047372 | 2021-12-01 | CPHL,MOH,EGYPT | CPHL | Wael H. Roshdy, Mohamed Kamal , Shymaa s. Ahmed, Ramy Galal,Nancy el guindy, Amel nagiub,yasser el hady,salma sayed, Abd Monaem Adel,Galal Mahmoud,Dalia Ramadan,Rabeh ..R. El-Shesheny, Mohamed A Ali,Mohamed Hassany, Amr Kandil. |
| hCoV-19/Egypt/CPHL-EGY21034/2021 | EPI_ISL_9047386 | 2021-12-12 | CPHL,MOH,EGYPT | CPHL | Wael H. Roshdy, Mohamed Kamal , Shymaa s. Ahmed, Ramy Galal,Nancy el guindy, Amel nagiub,yasser el hady,salma sayed, Abd Monaem Adel,Galal Mahmoud,Dalia Ramadan,Rabeh ..R. El-Shesheny, Mohamed A Ali,Mohamed Hassany, Amr Kandil. |
| hCoV-19/Egypt/CPHL-EGY21045/2021 | EPI_ISL_9047397 | 2021-12-12 | CPHL,MOH,EGYPT | CPHL | Wael H. Roshdy, Mohamed Kamal , Shymaa s. Ahmed, Ramy Galal,Nancy el guindy, Amel nagiub,yasser el hady,salma sayed, Abd Monaem Adel,Galal Mahmoud,Dalia Ramadan,Rabeh ..R. El-Shesheny, Mohamed A Ali,Mohamed Hassany, Amr Kandil. |

| | | | | | |
|---|---|---|---|---|---|
| hCoV-19/Egypt/CPHL-EGY21054/2021 | EPI_ISL_9047406 | 2021-12-13 | CPHL,MOH,EGYPT | CPHL | Wael H. Roshdy, Mohamed Kamal , Shymaa s. Ahmed, Ramy Galal,Nancy el guindy, Amel nagiub,yasser el hady,salma sayed, Abd Monaem Adel,Galal Mahmoud,Dalia Ramadan,Rabeh ..R. El-Shesheny, Mohamed A Ali,Mohamed Hassany, Amr Kandil. |
| hCoV-19/Egypt/CPHL-EGY21074/2021 | EPI_ISL_9047426 | 2021-12-18 | CPHL,MOH,EGYPT | CPHL | Wael H. Roshdy, Mohamed Kamal , Shymaa s. Ahmed, Ramy Galal,Nancy el guindy, Amel nagiub,yasser el hady,salma sayed, Abd Monaem Adel,Galal Mahmoud,Dalia Ramadan,Rabeh ..R. El-Shesheny, Mohamed A Ali,Mohamed Hassany, Amr Kandil. |
| hCoV-19/Egypt/CPHL-EGY21124/2021 | EPI_ISL_9047476 | 2021-12-15 | CPHL,MOH,EGYPT | CPHL | Wael H. Roshdy, Mohamed Kamal , Shymaa s. Ahmed, Ramy Galal,Nancy el guindy, Amel nagiub,yasser el hady,salma sayed, Abd Monaem Adel,Galal Mahmoud,Dalia Ramadan,Rabeh ..R. El-Shesheny, Mohamed A Ali,Mohamed Hassany, Amr Kandil. |
| hCoV-19/Egypt/CPHL-EGY21135/2021 | EPI_ISL_9047485 | 2021-11-28 | CPHL,MOH,EGYPT | CPHL | Wael H. Roshdy, Mohamed Kamal , Shymaa s. Ahmed, Ramy Galal,Nancy el guindy, Amel nagiub,yasser el hady,salma sayed, Abd Monaem Adel,Galal Mahmoud,Dalia Ramadan,Rabeh ..R. El-Shesheny, Mohamed A Ali,Mohamed Hassany, Amr Kandil. |
| hCoV-19/Egypt/CPHL-EGY21144/2020 | EPI_ISL_9047494 | 2020-12-13 | CPHL,MOH,EGYPT | CPHL | Wael H. Roshdy, Mohamed Kamal , Shymaa s. Ahmed, Ramy Galal,Nancy el guindy, Amel nagiub,yasser el hady,salma sayed, Abd Monaem Adel,Galal Mahmoud,Dalia Ramadan,Rabeh ..R. El-Shesheny, Mohamed A Ali,Mohamed Hassany, Amr Kandil. |
| hCoV-19/Egypt/CPHL-EGY21153/2021 | EPI_ISL_9047503 | 2021-11-07 | CPHL,MOH,EGYPT | CPHL | Wael H. Roshdy, Mohamed Kamal , Shymaa s. Ahmed, Ramy Galal,Nancy el guindy, Amel nagiub,yasser el hady,salma sayed, Abd Monaem Adel,Galal Mahmoud,Dalia Ramadan,Rabeh ..R. El-Shesheny, Mohamed A Ali,Mohamed Hassany, Amr Kandil. |
| hCoV-19/Egypt/CPHL-EGY21165/2021 | EPI_ISL_9047515 | 2021-12-06 | CPHL,MOH,EGYPT | CPHL | Wael H. Roshdy, Mohamed Kamal , Shymaa s. Ahmed, Ramy Galal,Nancy el guindy, Amel nagiub,yasser el hady,salma sayed, Abd Monaem Adel,Galal Mahmoud,Dalia Ramadan,Rabeh ..R. El-Shesheny, Mohamed A Ali,Mohamed Hassany, Amr Kandil. |
| hCoV-19/Egypt/CPHL-EGY21220/2021 | EPI_ISL_9047569 | 2021-02-12 | CPHL,MOH,EGYPT | CPHL | Wael H. Roshdy, Mohamed Kamal , Shymaa s. Ahmed, Ramy Galal,Nancy el guindy, Amel nagiub,yasser el hady,salma sayed, Abd Monaem Adel,Galal Mahmoud,Dalia Ramadan,Rabeh ..R. El-Shesheny, Mohamed A Ali,Mohamed Hassany, Amr Kandil. |
| hCoV-19/Egypt/CPHL-EGY21295/2021 | EPI_ISL_9047637 | 2021-12-01 | CPHL,MOH,EGYPT | CPHL | Wael H. Roshdy, Mohamed Kamal , Shymaa s. Ahmed, Ramy Galal,Nancy el guindy, Amel nagiub,yasser el hady,salma sayed, Abd Monaem Adel,Galal Mahmoud,Dalia Ramadan,Rabeh ..R. El-Shesheny, Mohamed A Ali,Mohamed Hassany, Amr Kandil. |

| | | | | | |
|---|---|---|---|---|---|
| hCoV-19/Egypt/CPHL-EGY21304/2021 | EPI_ISL_9047644 | 2021-12-15 | CPHL,MOH,EGYPT | CPHL | Wael H. Roshdy, Mohamed Kamal , Shymaa s. Ahmed, Ramy Galal,Nancy el guindy, Amel nagiub,yasser el hady,salma sayed, Abd Monaem Adel,Galal Mahmoud,Dalia Ramadan,Rabeh ..R. El-Shesheny, Mohamed A Ali,Mohamed Hassany, Amr Kandil. |
| hCoV-19/Egypt/CPHL-EGY21339/2021 | EPI_ISL_9047676 | 2021-12-01 | CPHL,MOH,EGYPT | CPHL | Wael H. Roshdy, Mohamed Kamal , Shymaa s. Ahmed, Ramy Galal,Nancy el guindy, Amel nagiub,yasser el hady,salma sayed, Abd Monaem Adel,Galal Mahmoud,Dalia Ramadan,Rabeh ..R. El-Shesheny, Mohamed A Ali,Mohamed Hassany, Amr Kandil. |
| hCoV-19/Egypt/CPHL-EGY21375/2021 | EPI_ISL_9047710 | 2021-12-01 | CPHL,MOH,EGYPT | CPHL | Wael H. Roshdy, Mohamed Kamal , Shymaa s. Ahmed, Ramy Galal,Nancy el guindy, Amel nagiub,yasser el hady,salma sayed, Abd Monaem Adel,Galal Mahmoud,Dalia Ramadan,Rabeh ..R. El-Shesheny, Mohamed A Ali,Mohamed Hassany, Amr Kandil. |
| hCoV-19/Germany/RP-RKI-I-437785/2021 | EPI_ISL_9054654 | 2021-12-30 | MVZ f√ºr Laboratoriumsmedizin und Mikrobiologie Koblenz-Mittelrhein (Labor Koblenz) | Robert Koch Institute | unknown |
| hCoV-19/India/AS-INSACOG-1828000563490/2021 | EPI_ISL_9056148 | 2021-09-09 | INSACOG-Assam | National Institute of Biomedical Genomics ,Äì INSACOG | Arindam Maitra, Ajanta Sharma, Nidhan Kumar Biswas, Lahari Saikia, Sreedhar Chinnaswamy,¬†Kailash Chamuah, Saumitra Das |
| hCoV-19/India/AS-INSACOG-1828000576487/2021 | EPI_ISL_9056334 | 2021-10-20 | INSACOG-Assam | National Institute of Biomedical Genomics ,Äì INSACOG | Arindam Maitra, Ajanta Sharma, Nidhan Kumar Biswas, Lahari Saikia, Sreedhar Chinnaswamy,¬†Kailash Chamuah, Saumitra Das |
| hCoV-19/India/AS-INSACOG-1828200331821/2021 | EPI_ISL_9056407 | 2021-09-14 | INSACOG-Assam | National Institute of Biomedical Genomics ,Äì INSACOG | Arindam Maitra, Ajanta Sharma, Nidhan Kumar Biswas, Lahari Saikia, Sreedhar Chinnaswamy,¬†Kailash Chamuah, Saumitra Das |
| hCoV-19/India/MZ-INSACOG-1526500063110/2021 | EPI_ISL_9056453 | 2021-10-04 | INSACOG-Mizoram | National Institute of Biomedical Genomics ,Äì INSACOG | Arindam Maitra, Swagnik Roy, Nidhan Kumar Biswas, Gracy Laldinmawii, Sreedhar Chinnaswamy, N Senthil Kumar, Saumitra Das |
| hCoV-19/USA/NJ-PHEL-V22002856/2020 | EPI_ISL_9060723 | 2020-12-31 | NJDOH, Public Health and Environmental Laboratories | NJ Public Health Environmental and Agricultural Laboratories | Lindsey Bodnar, Shiv K. Verma, Allison Roder, Maria-Magdalene Pugliese, Chelsea San Filippo, Jacquelyn Deverell, Mohammad M. Ali, Dana Woell, Byeong Jeong |
| hCoV-19/Italy/VEN-IRCCS-SCDC-134/2021 | EPI_ISL_9066389 | 2021-10-20 | IRCCS Sacro Cuore Don Calabria Hospital, Department of Infectious, Tropical Diseases & Microbiology | Department of Infectious, Tropical Diseases & Microbiology,IRCCS Sacro Cuore Don Calabria Hospital | Michela Deiana, Antonio Mori, Elena Pomari, Chiara Piubelli; |
| hCoV-19/Iraq/CPHL-051221-06/2021 | EPI_ISL_9068180 | 2021-12-01 | Central Public Health Laboratory | Central Public Health Laboratory | Dr. Hani Mousa Al-Akabi, Dr. Riyadh Al-Hilfi, Dr. Duraid Qasim Al-Sharif, Dr. Mohammad Issa Mohsin, Asaad Mahdi Asaad, Mohammaed Mohsin Gaeed, Manal Ahmed, Dr. Wissam Jasim, Iman Mutashar |
| hCoV-19/Iraq/CPHL-051221-08/2021 | EPI_ISL_9068183 | 2021-12-01 | Central Public Health Laboratory | Central Public Health Laboratory | Dr. Hani Mousa Al-Akabi, Dr. Riyadh Al-Hilfi, Dr. Duraid Qasim Al-Sharif, Dr. Mohammad Issa Mohsin, Asaad Mahdi Asaad, Mohammaed Mohsin Gaeed, Manal Ahmed, Dr. Wissam Jasim, Iman Mutashar |

| | | | | | |
|---|---|---|---|---|---|
| hCoV-19/Iraq/CPHL-051221-02/2021 | EPI_ISL_9068192 | 2021-12-01 | Central Public Health Laboratory | Central Public Health Laboratory | Dr. Hani Mousa Al-Akabi, Dr. Riyadh Al-Hilfi, Dr. Duraid Qasim Al-Sharif, Dr. Mohammad Issa Mohsin, Asaad Mahdi Asaad, Mohammaed Mohsin Gaeed, Manal Ahmed, Dr. Wissam Jasim, Iman Mutashar |
| hCoV-19/Iraq/CPHL-051221-40/2021 | EPI_ISL_9068198 | 2021-12-01 | Central Public Health Laboratory | Central Public Health Laboratory | Dr. Hani Mousa Al-Akabi, Dr. Riyadh Al-Hilfi, Dr. Duraid Qasim Al-Sharif, Dr. Mohammad Issa Mohsin, Asaad Mahdi Asaad, Mohammaed Mohsin Gaeed, Manal Ahmed, Dr. Wissam Jasim, Iman Mutashar |
| hCoV-19/Iraq/CPHL-051221-12/2021 | EPI_ISL_9068202 | 2021-12-01 | Central Public Health Laboratory | Central Public Health Laboratory | Dr. Hani Mousa Al-Akabi, Dr. Riyadh Al-Hilfi, Dr. Duraid Qasim Al-Sharif, Dr. Mohammad Issa Mohsin, Asaad Mahdi Asaad, Mohammaed Mohsin Gaeed, Manal Ahmed, Dr. Wissam Jasim, Iman Mutashar |
| hCoV-19/Iraq/CPHL-051221-05/2021 | EPI_ISL_9068205 | 2021-12-01 | Central Public Health Laboratory | Central Public Health Laboratory | Dr. Hani Mousa Al-Akabi, Dr. Riyadh Al-Hilfi, Dr. Duraid Qasim Al-Sharif, Dr. Mohammad Issa Mohsin, Asaad Mahdi Asaad, Mohammaed Mohsin Gaeed, Manal Ahmed, Dr. Wissam Jasim, Iman Mutashar |
| hCoV-19/Iraq/CPHL-061021-36/2021 | EPI_ISL_9069645 | 2021-10-01 | Central Public Health Laboratory | Central Public Health Laboratory | Dr. Hani Mousa Al-Akabi, Dr. Riyadh Al-Hilfi, Dr. Duraid Qasim Al-Sharif, Dr. Mohammad Issa Mohsin, Asaad Mahdi Asaad, Mohammaed Mohsin Gaeed, Manal Ahmed, Dr. Wissam Jasim, Iman Mutashar |
| hCoV-19/SouthAfrica/CERI-KRISP-K034752/2021 | EPI_ISL_9072346 | 2021-12-21 | CAPRISA Hillcrest | and Innvoation, Stellenbosch University and KRISP, KZN Research Innovation and Sequencing Platform, UKZN. | Natasha Samsunder, Aida Sivro, Giandhari J, Pillay S, Naidoo Y, Ramphal U, Ramphal Y, van Wyk S, Moir M, San JE, Tshiabuila D, Tegally H, Wilkinson E, de Oliveira T |
| hCoV-19/Germany/NW-RKI-I-443099/2021 | EPI_ISL_9075190 | 2021-12-14 | Labor ZOTZ\|KLIMAS; MVZ Dvºsseldorf-Centrum | Robert Koch Institute | unknown |
| hCoV-19/Netherlands/NH-RIVM-80798/2021 | EPI_ISL_9085496 | 2021-12-29 | Dutch COVID-19 response team | National Institute for Public Health and the Environment (RIVM) | Adam Meijer, Harry Vennema, Dirk Eggink, Jeroen Cremer, Sharon van den Brink, Bas van der Veer, AnneMarie van den Brandt, Lisa Wijsman, Kim Freriks, Ryanne Jaarsma, Lynn Aarts, Sanne Bos, Jil Kocken, Jordy de Bakker, Afke Vogelzang, Linda van Someren, Flo |
| hCoV-19/SintMaarten/SX-RIVM-81068/2021 | EPI_ISL_9086202 | 2021-12-28 | Dutch COVID-19 response team | National Institute for Public Health and the Environment (RIVM) | Adam Meijer, Harry Vennema, Dirk Eggink, Jeroen Cremer, Sharon van den Brink, Bas van der Veer, AnneMarie van den Brandt, Lisa Wijsman, Kim Freriks, Ryanne Jaarsma, Lynn Aarts, Sanne Bos, Jil Kocken, Jordy de Bakker, Afke Vogelzang, Linda van Someren, Flo |
| hCoV-19/SintMaarten/SX-RIVM-81082/2021 | EPI_ISL_9086214 | 2021-12-28 | Dutch COVID-19 response team | National Institute for Public Health and the Environment (RIVM) | Adam Meijer, Harry Vennema, Dirk Eggink, Jeroen Cremer, Sharon van den Brink, Bas van der Veer, AnneMarie van den Brandt, Lisa Wijsman, Kim Freriks, Ryanne Jaarsma, Lynn Aarts, Sanne Bos, Jil Kocken, Jordy de Bakker, Afke Vogelzang, Linda van Someren, Flo |
| hCoV-19/France/BRE-IPP53884/2021 | EPI_ISL_9087215 | 2021-12-27 | Labo Analyses Med | National Reference Center for Viruses of Respiratory Infections, Institut Pasteur, Paris | Marion Barbet, Sylvie Behillil, M√©line Bizard, Angela Brisebarre, Camille Capel, Vincent Enouf, Louise Lefran√ßois, Fr√©d√©ric Lemoine, Christophe Malabat, Corinne Maufrais, Slim El Khiari, Julien Fumey, Etienne Simon-Lori√®re, Maud Vanpeene, Sylvie Van |

| | | | | | |
|---|---|---|---|---|---|
| hCoV-19/India/KA-RFNB-3473/2021 | EPI_ISL_9091072 | 2021-12-25 | BANGALORE MEDICAL COLLEGE AND RESEARCH INSTITUTE VRDL (BMCRI), VICTORIA HOSPITAL CAMPUS | National Center for Biological Sciences, TIFR - Rockefeller Foundation | Darshan Sreenivas,Bhagyashree Madhav Shelar,Kathirvel Kandasamy,Lakshminarayanan CP,Awadhesh Pandit,Satyajit Mayor,Uma Ramakrishnan,Dimple Notani |
| hCoV-19/India/KA-RFNB-2225/2021 | EPI_ISL_9092081 | 2021-12-29 | Auriga Research Pvt Ltd, Laboratory, Bengaluru | National Center for Biological Sciences, TIFR - Rockefeller Foundation | Darshan Sreenivas,Bhagyashree Madhav Shelar,Kathirvel Kandasamy,Lakshminarayanan CP,Awadhesh Pandit,Satyajit Mayor,Uma Ramakrishnan,Dimple Notani |
| hCoV-19/Kenya/ILRI_COVM01621/2021 | EPI_ISL_9093416 | 2021-11-27 | International Livestock Research Institute | International Livestock Research Institute | Samuel O. Oyola, Daniel Ouso, Gilbert Kibet, Shebbar Osiany, Edward Kiritu, Paul Dobi, Collins Muli, Patrick Amoth, Vishvanath Nene, Sonal P. Henson, Edward O. Abworo |
| hCoV-19/Kenya/ILRI_COVM01623/2021 | EPI_ISL_9093418 | 2021-11-28 | International Livestock Research Institute | International Livestock Research Institute | Samuel O. Oyola, Daniel Ouso, Gilbert Kibet, Shebbar Osiany, Edward Kiritu, Paul Dobi, Collins Muli, Patrick Amoth, Vishvanath Nene, Sonal P. Henson, Edward O. Abworo |
| hCoV-19/Kenya/ILRI_COVM01686/2021 | EPI_ISL_9093447 | 2021-12-06 | International Livestock Research Institute | International Livestock Research Institute | Samuel O. Oyola, Daniel Ouso, Gilbert Kibet, Shebbar Osiany, Edward Kiritu, Paul Dobi, Collins Muli, Patrick Amoth, Vishvanath Nene, Sonal P. Henson, Edward O. Abworo |
| hCoV-19/Kenya/ILRI_COVM01692/2021 | EPI_ISL_9093452 | 2021-12-06 | International Livestock Research Institute | International Livestock Research Institute | Samuel O. Oyola, Daniel Ouso, Gilbert Kibet, Shebbar Osiany, Edward Kiritu, Paul Dobi, Collins Muli, Patrick Amoth, Vishvanath Nene, Sonal P. Henson, Edward O. Abworo |
| hCoV-19/Kenya/ILRI_COVM01787/2021 | EPI_ISL_9093527 | 2021-12-14 | International Livestock Research Institute | International Livestock Research Institute | Samuel O. Oyola, Daniel Ouso, Gilbert Kibet, Shebbar Osiany, Edward Kiritu, Paul Dobi, Collins Muli, Patrick Amoth, Vishvanath Nene, Sonal P. Henson, Edward O. Abworo |
| hCoV-19/Kenya/ILRI_COVM01866/2021 | EPI_ISL_9093603 | 2021-12-16 | International Livestock Research Institute | International Livestock Research Institute | Samuel O. Oyola, Daniel Ouso, Gilbert Kibet, Shebbar Osiany, Edward Kiritu, Paul Dobi, Collins Muli, Patrick Amoth, Vishvanath Nene, Sonal P. Henson, Edward O. Abworo |
| hCoV-19/DRC/ILRI_COVM01446/2021 | EPI_ISL_9093633 | 2021-04-02 | International Livestock Research Institute | International Livestock Research Institute | Samuel O. Oyola, Daniel Ouso, Gilbert Kibet, Shebbar Osiany, Edward Kiritu, Paul Dobi, Collins Muli, Patrick Amoth, Vishvanath Nene, Sonal P. Henson, Edward O. Abworo |
| hCoV-19/DRC/ILRI_COVM01516/2021 | EPI_ISL_9093689 | 2021-07-21 | International Livestock Research Institute | International Livestock Research Institute | Samuel O. Oyola, Daniel Ouso, Gilbert Kibet, Shebbar Osiany, Edward Kiritu, Paul Dobi, Collins Muli, Patrick Amoth, Vishvanath Nene, Sonal P. Henson, Edward O. Abworo |
| hCoV-19/USA/OH-2021063955/2021 | EPI_ISL_9094232 | 2021-08-12 | Ohio Department of Health Laboratory | Ohio Department of Health Laboratory | Jennifer Holmes, Eric Brandt, Keoni Omura, Glen McGillivary, Caitlin McDonnell, Jade Mowery, Stephanie Mccracken, Tyler Payne, Kirtana Ramadugu, Erica Leasure, Brent Lee, Kelsey Florek, Heather Blankenship, Allison Black, Quanta Brown, Tammy Bannerman, Sa |

| | | | | | |
|---|---|---|---|---|---|
| hCoV-19/USA/NJ-NYGC-V1265-AAYY607/2021 | EPI_ISL_9096406 | 2021-11-29 | Hackensack Medical Center | New York Genome Center | Michael Zody, Andre Corvelo, Dayna M. Oschwald, Amy Baldwin, Samantha Fennessey, Tom Maniatis, Liang Chen, Jose Mediavilla, Marcus Cunningham, Kaelea Composto, Kar Chow, David Perlin, Barry Kreiswirth |
| hCoV-19/USA/CA-HLX-STM-KZ9NDTS3Q/2021 | EPI_ISL_9098990 | 2021-12-29 | Kaiser Permanente Southern California | Helix | Kaiser Permanente Southern California, Helix |
| hCoV-19/Germany/SN-RKI-I-449602/2021 | EPI_ISL_9105652 | 2021-12-30 | Bioscientia Labor Wermsdorf | Robert Koch Institute | unknown |
| hCoV-19/USA/TX-HHD-2201057541/2021 | EPI_ISL_9112999 | 2021-12-22 | GALE, LETOSHA | Houston Health Dept. | Ryker Penn, Pamela Brown, Adolfo Lara, Yanlai Lai |
| hCoV-19/India/WB-INSACOG-1930900316278/2021 | EPI_ISL_9116543 | 2021-09-20 | INSACOG-WB | National Institute of Biomedical Genomics ‚Äì INSACOG | Arindam Maitra, Bhaswati Bandyopadhyay, Nidhan Kumar Biswas, Tamal Ghosh, Sreedhar Chinnaswamy, Ajay Chakraborti, Saumitra Das |
| hCoV-19/India/WB-INSACOG-1931300506701/2021 | EPI_ISL_9116743 | 2021-11-06 | INSACOG-WB | National Institute of Biomedical Genomics ‚Äì INSACOG | Arindam Maitra, Bhaswati Bandyopadhyay, Nidhan Kumar Biswas, Tamal Ghosh, Sreedhar Chinnaswamy, Ajay Chakraborti, Saumitra Das |
| hCoV-19/Bermuda/WR-NYGC-DEC-6122-FFRF784/2021 | EPI_ISL_9121153 | 2021-09-12 | Bermuda Molecular Diagnostics and Research Laboratory | New York Genome Center | Michael Zody, Andre Corvelo, Dayna M. Oschwald, Amy Baldwin, Samantha Fennessey, Tom Maniatis, Carika Weldon, Dy-Juan DeRoza, Ayoola Oyinloye |
| hCoV-19/Bermuda/WR-NYGC-DEC-6132-MZFB693/2021 | EPI_ISL_9121156 | 2021-11-12 | Bermuda Molecular Diagnostics and Research Laboratory | New York Genome Center | Michael Zody, Andre Corvelo, Dayna M. Oschwald, Amy Baldwin, Samantha Fennessey, Tom Maniatis, Carika Weldon, Dy-Juan DeRoza, Ayoola Oyinloye |
| hCoV-19/Bermuda/WR-NYGC-DEC-6126-GNOP644/2021 | EPI_ISL_9121170 | 2021-10-12 | Bermuda Molecular Diagnostics and Research Laboratory | New York Genome Center | Michael Zody, Andre Corvelo, Dayna M. Oschwald, Amy Baldwin, Samantha Fennessey, Tom Maniatis, Carika Weldon, Dy-Juan DeRoza, Ayoola Oyinloye |
| hCoV-19/USA/NY-SUNYQBR-61210501016904/2021 | EPI_ISL_9133926 | 2021-12-01 | Quadrant Laboratories, LLC | SUNY Upstate Medical University | Frank Middleton, Brian Pavlovitz |
| hCoV-19/Canada/ON-PHL-22-01561/2021 | EPI_ISL_9136991 | 2021-12-28 | Public Health Ontario Laboratory | Public Health Ontario Laboratory | Vanessa G Allen, Philip Banh, Yao Chen, Richard de Borja, Alireza Eshaghi, Nahuel Fittipaldi, Christine Frantz, Jonathan B Gubbay, Jennifer L Guthrie, Lawrence Heisler, Esha Joshi, Michael Laszloffy, Aimin Li, Michael CY Li, Dean Maxwell, Sandeep Nagra, S |
| hCoV-19/USA/AZ-TG1170254/2021 | EPI_ISL_9152653 | 2021-10-27 | TGen North | TGen North | Jolene Bowers, Heather Centner, Brett Van Tassel, Chris French, Hayley Yaglom, Darrin Lemmer, Dave Engelthaler |
| hCoV-19/France/ARA-HCL021233073501/2021 | EPI_ISL_9155607 | 2021-12-30 | CNR Virus des Infections Respiratoires - France SUD | CNR Virus des Infections Respiratoires - France SUD | Antonin Bal, Bruno Simon, Th√©ophile Boyer, Gregory Destras, Gwendolyne Burfin, Hadrien Regue, Quentin Semanas, Martine Valette, Bruno Lina, Laurence Josset |
| hCoV-19/France/ARA-HCL022006010901/2021 | EPI_ISL_9155687 | 2021-12-27 | LABORATOIRE NOVELAB | CNR Virus des Infections Respiratoires - France SUD | Antonin Bal, Bruno Simon, Th√©ophile Boyer, Gregory Destras, Gwendolyne Burfin, Hadrien Regue, Quentin Semanas, Martine Valette, Bruno Lina, Laurence Josset |
| hCoV-19/Chile/CO-197270/2021 | EPI_ISL_9156374 | 2021-12-21 | Genetica Molecular and Subdepartamento de Virologia ISP Chile | Instituto de Salud Publica de Chile | Karen Orostica, Constanza Campano, Barbara Parra, Patricia Bustos, Rodrigo Fasce, Andres Castillo, Jorge Fernandez |
| hCoV-19/Chile/VS-197305/2021 | EPI_ISL_9156409 | 2021-12-23 | Genetica Molecular and Subdepartamento de Virologia ISP Chile | Instituto de Salud Publica de Chile | Karen Orostica, Constanza Campano, Barbara Parra, Patricia Bustos, Rodrigo Fasce, Andres Castillo, Jorge Fernandez |

| | | | | | |
|---|---|---|---|---|---|
| hCoV-19/Chile/VS-197317/2021 | EPI_ISL_9156421 | 2021-12-23 | Genetica Molecular and Subdepartamento de Virologia ISP Chile | Instituto de Salud Publica de Chile | Karen Orostica, Constanza Campano, Barbara Parra, Patricia Bustos, Rodrigo Fasce, Andres Castillo, Jorge Fernandez |
| hCoV-19/Chile/RM-197435/2021 | EPI_ISL_9156513 | 2021-12-24 | Genetica Molecular and Subdepartamento de Virologia ISP Chile | Instituto de Salud Publica de Chile | Karen Orostica, Constanza Campano, Barbara Parra, Patricia Bustos, Rodrigo Fasce, Andres Castillo, Jorge Fernandez |
| hCoV-19/Chile/RM-197439/2021 | EPI_ISL_9156516 | 2021-12-23 | Genetica Molecular and Subdepartamento de Virologia ISP Chile | Instituto de Salud Publica de Chile | Karen Orostica, Constanza Campano, Barbara Parra, Patricia Bustos, Rodrigo Fasce, Andres Castillo, Jorge Fernandez |
| hCoV-19/Chile/VS-197517/2021 | EPI_ISL_9156573 | 2021-12-24 | Genetica Molecular and Subdepartamento de Virologia ISP Chile | Instituto de Salud Publica de Chile | Karen Orostica, Constanza Campano, Barbara Parra, Patricia Bustos, Rodrigo Fasce, Andres Castillo, Jorge Fernandez |
| hCoV-19/Chile/RM-197569/2021 | EPI_ISL_9156607 | 2021-12-25 | Genetica Molecular and Subdepartamento de Virologia ISP Chile | Instituto de Salud Publica de Chile | Karen Orostica, Constanza Campano, Barbara Parra, Patricia Bustos, Rodrigo Fasce, Andres Castillo, Jorge Fernandez |
| hCoV-19/Chile/RM-197579/2021 | EPI_ISL_9156617 | 2021-12-25 | Genetica Molecular and Subdepartamento de Virologia ISP Chile | Instituto de Salud Publica de Chile | Karen Orostica, Constanza Campano, Barbara Parra, Patricia Bustos, Rodrigo Fasce, Andres Castillo, Jorge Fernandez |
| hCoV-19/Chile/RM-197591/2021 | EPI_ISL_9156629 | 2021-12-25 | Genetica Molecular and Subdepartamento de Virologia ISP Chile | Instituto de Salud Publica de Chile | Karen Orostica, Constanza Campano, Barbara Parra, Patricia Bustos, Rodrigo Fasce, Andres Castillo, Jorge Fernandez |
| hCoV-19/USA/CA-CDPH-6000006956/2021 | EPI_ISL_9169187 | 2021-12-26 | Color | California Department of Public Health | VRDL COVID Sequencing Hub |
| hCoV-19/Chile/TA-199483/2021 | EPI_ISL_9172233 | 2021-12-28 | Genetica Molecular and Subdepartamento de Virologia ISP Chile | Instituto de Salud Publica de Chile | Karen Orostica, Constanza Campano, Barbara Parra, Patricia Bustos, Rodrigo Fasce, Andres Castillo, Jorge Fernandez |
| hCoV-19/Chile/TA-198263/2021 | EPI_ISL_9172254 | 2021-12-26 | Genetica Molecular and Subdepartamento de Virologia ISP Chile | Instituto de Salud Publica de Chile | Karen Orostica, Constanza Campano, Barbara Parra, Patricia Bustos, Rodrigo Fasce, Andres Castillo, Jorge Fernandez |
| hCoV-19/Chile/AN-197029/2021 | EPI_ISL_9172509 | 2021-12-22 | Genetica Molecular and Subdepartamento de Virologia ISP Chile | Instituto de Salud Publica de Chile | Karen Orostica, Constanza Campano, Barbara Parra, Patricia Bustos, Rodrigo Fasce, Andres Castillo, Jorge Fernandez |
| hCoV-19/Chile/TA-200255/2021 | EPI_ISL_9180081 | 2021-12-29 | Genetica Molecular and Subdepartamento de Virologia ISP Chile | Instituto de Salud Publica de Chile | Karen Orostica, Constanza Campano, Barbara Parra, Patricia Bustos, Rodrigo Fasce, Andres Castillo, Jorge Fernandez |
| hCoV-19/Chile/RM-197193/2021 | EPI_ISL_9181775 | 2021-12-23 | Genetica Molecular and Subdepartamento de Virologia ISP Chile | Instituto de Salud Publica de Chile | Karen Orostica, Constanza Campano, Barbara Parra, Patricia Bustos, Rodrigo Fasce, Andres Castillo, Jorge Fernandez |
| hCoV-19/Chile/RM-197105/2021 | EPI_ISL_9181816 | 2021-12-20 | Genetica Molecular and Subdepartamento de Virologia ISP Chile | Instituto de Salud Publica de Chile | Karen Orostica, Constanza Campano, Barbara Parra, Patricia Bustos, Rodrigo Fasce, Andres Castillo, Jorge Fernandez |
| hCoV-19/USA/MA-MASPHL-08943/2021 | EPI_ISL_9182171 | 2021-12-14 | Massachusetts State Public Health Lab | Massachusetts State Public Health Lab | Andrew Lang, Timelia Fink, Glen Gallagher, Sandra Smole |
| hCoV-19/USA/CO-CDPHE-2102615952/2021 | EPI_ISL_9183951 | 2021-12-27 | Colorado Department of Public Health and Environment | Colorado Department of Public Health and Environment | Laura Bankers, Molly C. Hetherington-Rauth, Diana Ir, Alexandria Rossheim, Michael Martin, Mandy Waters, Arianna Smith, Shannon R. Matzinger, Emily A. Travanty |

| | | | | | |
|---|---|---|---|---|---|
| hCoV-19/Chile/RM-197646/2021 | EPI_ISL_9184413 | 2021-12-24 | Genetica Molecular and Subdepartamento de Virologia ISP Chile | Instituto de Salud Publica de Chile | Karen Orostica, Constanza Campano, Barbara Parra, Patricia Bustos, Rodrigo Fasce, Andres Castillo, Jorge Fernandez |
| hCoV-19/Chile/ML-198016/2021 | EPI_ISL_9184489 | 2021-10-26 | Genetica Molecular and Subdepartamento de Virologia ISP Chile | Instituto de Salud Publica de Chile | Karen Orostica, Constanza Campano, Barbara Parra, Patricia Bustos, Rodrigo Fasce, Andres Castillo, Jorge Fernandez |
| hCoV-19/Chile/RM-198098/2021 | EPI_ISL_9184562 | 2021-12-26 | Genetica Molecular and Subdepartamento de Virologia ISP Chile | Instituto de Salud Publica de Chile | Karen Orostica, Constanza Campano, Barbara Parra, Patricia Bustos, Rodrigo Fasce, Andres Castillo, Jorge Fernandez |
| hCoV-19/Chile/RM-198663/2021 | EPI_ISL_9184578 | 2021-12-27 | Genetica Molecular and Subdepartamento de Virologia ISP Chile | Instituto de Salud Publica de Chile | Karen Orostica, Constanza Campano, Barbara Parra, Patricia Bustos, Rodrigo Fasce, Andres Castillo, Jorge Fernandez |
| hCoV-19/Chile/RM-199141/2021 | EPI_ISL_9184641 | 2021-12-27 | Genetica Molecular and Subdepartamento de Virologia ISP Chile | Instituto de Salud Publica de Chile | Karen Orostica, Constanza Campano, Barbara Parra, Patricia Bustos, Rodrigo Fasce, Andres Castillo, Jorge Fernandez |
| hCoV-19/Chile/RM-199151/2021 | EPI_ISL_9184650 | 2021-12-27 | Genetica Molecular and Subdepartamento de Virologia ISP Chile | Instituto de Salud Publica de Chile | Karen Orostica, Constanza Campano, Barbara Parra, Patricia Bustos, Rodrigo Fasce, Andres Castillo, Jorge Fernandez |
| hCoV-19/Argentina/INEI115523/2021 | EPI_ISL_9189258 | 2021-12-07 | Servicio Virosis Respiratorias-Departamento Virologv≠a-INEI | Instituto Nacional Enfermedades Infecciosas C.G.Malbran | Baumeister E., Avaro M., Benedetti E., Russo M., Dattero ME, Pontoriero A., Cisterna D., Molina V., Perandones C., Tuduri E., De Belder D., Sanchez Loria J., Haim MS., Lorenzo F., Poklepovich T., Campos J. |
| hCoV-19/Argentina/INEI116063/2021 | EPI_ISL_9189321 | 2021-12-07 | Servicio Virosis Respiratorias-Departamento Virologv≠a-INEI | Instituto Nacional Enfermedades Infecciosas C.G.Malbran | Baumeister E., Avaro M., Benedetti E., Russo M., Dattero ME, Pontoriero A., Cisterna D., Molina V., Perandones C., Tuduri E., De Belder D., Sanchez Loria J., Haim MS., Lorenzo F., Poklepovich T., Campos J. |
| hCoV-19/Argentina/INEI116103/2021 | EPI_ISL_9189323 | 2021-12-16 | Servicio Virosis Respiratorias-Departamento Virologv≠a-INEI | Instituto Nacional Enfermedades Infecciosas C.G.Malbran | Baumeister E., Avaro M., Benedetti E., Russo M., Dattero ME, Pontoriero A., Cisterna D., Molina V., Perandones C., Tuduri E., De Belder D., Sanchez Loria J., Haim MS., Lorenzo F., Poklepovich T., Campos J. |
| hCoV-19/Argentina/INEI115837/2021 | EPI_ISL_9189706 | 2021-12-16 | Servicio Virosis Respiratorias-Departamento Virologv≠a-INEI | Instituto Nacional Enfermedades Infecciosas C.G.Malbran | Baumeister E., Avaro M., Benedetti E., Russo M., Dattero ME, Pontoriero A., Cisterna D., Molina V., Perandones C., Tuduri E., De Belder D., Sanchez Loria J., Haim MS., Lorenzo F., Poklepovich T., Campos J. |
| hCoV-19/Argentina/INEI116125/2021 | EPI_ISL_9189819 | 2021-12-17 | Servicio Virosis Respiratorias-Departamento Virologv≠a-INEI | Instituto Nacional Enfermedades Infecciosas C.G.Malbran | Baumeister E., Avaro M., Benedetti E., Russo M., Dattero ME, Pontoriero A., Cisterna D., Molina V., Perandones C., Tuduri E., De Belder D., Sanchez Loria J., Haim MS., Lorenzo F., Poklepovich T., Campos J. |
| hCoV-19/Argentina/INEI117296/2021 | EPI_ISL_9191708 | 2021-12-29 | Servicio Virosis Respiratorias-Departamento Virologv≠a-INEI | Instituto Nacional Enfermedades Infecciosas C.G.Malbran | Baumeister E., Avaro M., Benedetti E., Russo M., Dattero ME, Pontoriero A., Cisterna D., Molina V., Perandones C., Tuduri E., De Belder D., Sanchez Loria J., Haim MS., Lorenzo F., Poklepovich T., Campos J. |
| hCoV-19/Argentina/INEI117094/2021 | EPI_ISL_9191736 | 2021-12-22 | Servicio Virosis Respiratorias-Departamento Virologv≠a-INEI | Instituto Nacional Enfermedades Infecciosas C.G.Malbran | Baumeister E., Avaro M., Benedetti E., Russo M., Dattero ME, Pontoriero A., Cisterna D., Molina V., Perandones C., Tuduri E., De Belder D., Sanchez Loria J., Haim MS., Lorenzo F., Poklepovich T., Campos J. |

| | | | | | |
|---|---|---|---|---|---|
| hCoV-19/Argentina/INEI116491/2021 | EPI_ISL_9191784 | 2021-12-23 | Servicio Virosis Respiratorias-Departamento Virologv≠a-INEI | Instituto Nacional Enfermedades Infecciosas C.G.Malbran | Baumeister E., Avaro M., Benedetti E., Russo M., Dattero ME, Pontoriero A., Cisterna D., Molina V., Perandones C., Tuduri E., De Belder D., Sanchez Loria J., Haim MS., Lorenzo F., Poklepovich T., Campos J. |
| hCoV-19/Argentina/INEI117239/2021 | EPI_ISL_9191854 | 2021-12-26 | Servicio Virosis Respiratorias-Departamento Virologv≠a-INEI | Instituto Nacional Enfermedades Infecciosas C.G.Malbran | Baumeister E., Avaro M., Benedetti E., Russo M., Dattero ME, Pontoriero A., Cisterna D., Molina V., Perandones C., Tuduri E., De Belder D., Sanchez Loria J., Haim MS., Lorenzo F., Poklepovich T., Campos J. |
| hCoV-19/Argentina/INEI117063/2021 | EPI_ISL_9191942 | 2021-12-25 | Servicio Virosis Respiratorias-Departamento Virologv≠a-INEI | Instituto Nacional Enfermedades Infecciosas C.G.Malbran | Baumeister E., Avaro M., Benedetti E., Russo M., Dattero ME, Pontoriero A., Cisterna D., Molina V., Perandones C., Tuduri E., De Belder D., Sanchez Loria J., Haim MS., Lorenzo F., Poklepovich T., Campos J. |
| hCoV-19/Argentina/INEI116420/2021 | EPI_ISL_9192113 | 2021-12-29 | Servicio Virosis Respiratorias-Departamento Virologv≠a-INEI | Instituto Nacional Enfermedades Infecciosas C.G.Malbran | Baumeister E., Avaro M., Benedetti E., Russo M., Dattero ME, Pontoriero A., Cisterna D., Molina V., Perandones C., Tuduri E., De Belder D., Sanchez Loria J., Haim MS., Lorenzo F., Poklepovich T., Campos J. |
| hCoV-19/Argentina/INEI116452/2021 | EPI_ISL_9192125 | 2021-12-27 | Servicio Virosis Respiratorias-Departamento Virologv≠a-INEI | Instituto Nacional Enfermedades Infecciosas C.G.Malbran | Baumeister E., Avaro M., Benedetti E., Russo M., Dattero ME, Pontoriero A., Cisterna D., Molina V., Perandones C., Tuduri E., De Belder D., Sanchez Loria J., Haim MS., Lorenzo F., Poklepovich T., Campos J. |
| hCoV-19/Argentina/INEI116463/2021 | EPI_ISL_9192129 | 2021-12-27 | Servicio Virosis Respiratorias-Departamento Virologv≠a-INEI | Instituto Nacional Enfermedades Infecciosas C.G.Malbran | Baumeister E., Avaro M., Benedetti E., Russo M., Dattero ME, Pontoriero A., Cisterna D., Molina V., Perandones C., Tuduri E., De Belder D., Sanchez Loria J., Haim MS., Lorenzo F., Poklepovich T., Campos J. |
| hCoV-19/Argentina/INEI116756/2021 | EPI_ISL_9192132 | 2021-12-22 | Servicio Virosis Respiratorias-Departamento Virologv≠a-INEI | Instituto Nacional Enfermedades Infecciosas C.G.Malbran | Baumeister E., Avaro M., Benedetti E., Russo M., Dattero ME, Pontoriero A., Cisterna D., Molina V., Perandones C., Tuduri E., De Belder D., Sanchez Loria J., Haim MS., Lorenzo F., Poklepovich T., Campos J. |
| hCoV-19/Argentina/INEI116939/2021 | EPI_ISL_9192145 | 2021-12-28 | Servicio Virosis Respiratorias-Departamento Virologv≠a-INEI | Instituto Nacional Enfermedades Infecciosas C.G.Malbran | Baumeister E., Avaro M., Benedetti E., Russo M., Dattero ME, Pontoriero A., Cisterna D., Molina V., Perandones C., Tuduri E., De Belder D., Sanchez Loria J., Haim MS., Lorenzo F., Poklepovich T., Campos J. |
| hCoV-19/Argentina/INEI117276/2021 | EPI_ISL_9192159 | 2021-12-28 | Servicio Virosis Respiratorias-Departamento Virologv≠a-INEI | Instituto Nacional Enfermedades Infecciosas C.G.Malbran | Baumeister E., Avaro M., Benedetti E., Russo M., Dattero ME, Pontoriero A., Cisterna D., Molina V., Perandones C., Tuduri E., De Belder D., Sanchez Loria J., Haim MS., Lorenzo F., Poklepovich T., Campos J. |
| hCoV-19/Argentina/INEI116504/2021 | EPI_ISL_9192219 | 2021-12-20 | Servicio Virosis Respiratorias-Departamento Virologv≠a-INEI | Instituto Nacional Enfermedades Infecciosas C.G.Malbran | Baumeister E., Avaro M., Benedetti E., Russo M., Dattero ME, Pontoriero A., Cisterna D., Molina V., Perandones C., Tuduri E., De Belder D., Sanchez Loria J., Haim MS., Lorenzo F., Poklepovich T., Campos J. |
| hCoV-19/Argentina/INEI116495/2021 | EPI_ISL_9192220 | 2021-12-22 | Servicio Virosis Respiratorias-Departamento Virologv≠a-INEI | Instituto Nacional Enfermedades Infecciosas C.G.Malbran | Baumeister E., Avaro M., Benedetti E., Russo M., Dattero ME, Pontoriero A., Cisterna D., Molina V., Perandones C., Tuduri E., De Belder D., Sanchez Loria J., Haim MS., Lorenzo F., Poklepovich T., Campos J. |

| | | | | | |
|---|---|---|---|---|---|
| hCoV-19/Argentina/INEI117202/2021 | EPI_ISL_9192250 | 2021-12-30 | Servicio Virosis Respiratorias-Departamento Virologv≠a-INEI | Instituto Nacional Enfermedades Infecciosas C.G.Malbran | Baumeister E., Avaro M., Benedetti E., Russo M., Dattero ME, Pontoriero A., Cisterna D., Molina V., Perandones C., Tuduri E., De Belder D., Sanchez Loria J., Haim MS., Lorenzo F., Poklepovich T., Campos J. |
| hCoV-19/Argentina/INEI116972/2021 | EPI_ISL_9192267 | 2021-12-27 | Servicio Virosis Respiratorias-Departamento Virologv≠a-INEI | Instituto Nacional Enfermedades Infecciosas C.G.Malbran | Baumeister E., Avaro M., Benedetti E., Russo M., Dattero ME, Pontoriero A., Cisterna D., Molina V., Perandones C., Tuduri E., De Belder D., Sanchez Loria J., Haim MS., Lorenzo F., Poklepovich T., Campos J. |
| hCoV-19/USA/TX-DSHS-14289/2021 | EPI_ISL_9196633 | 2021-09-30 | TXDSHS | TXDSHS | Rashmi Tuladhar, Bonnie Oh, Breanna Scorza, Maliha Rahman, Mayela Pedrueza, Anita Pokharel, Jie Lu, Karen Bobier, Yan Sun, Rachel Lee, Grace Kubin |
| hCoV-19/USA/TX-DSHS-14291/2021 | EPI_ISL_9196636 | 2021-09-30 | TXDSHS | TXDSHS | Rashmi Tuladhar, Bonnie Oh, Breanna Scorza, Maliha Rahman, Mayela Pedrueza, Anita Pokharel, Jie Lu, Karen Bobier, Yan Sun, Rachel Lee, Grace Kubin |
| hCoV-19/USA/TX-DSHS-14292/2021 | EPI_ISL_9196639 | 2021-10-01 | TXDSHS | TXDSHS | Rashmi Tuladhar, Bonnie Oh, Breanna Scorza, Maliha Rahman, Mayela Pedrueza, Anita Pokharel, Jie Lu, Karen Bobier, Yan Sun, Rachel Lee, Grace Kubin |
| hCoV-19/USA/TX-HMH-MCoV-71964/2021 | EPI_ISL_9199447 | 2021-12-22 | Houston Methodist Hospital | Houston Methodist Hospital | Randall J. Olsen, Paul A. Christensen, S. Wesley Long, Richard Snehall, Robert Olson, Marcus Nguyen, James J. Davis, Matthew Ojeda Saavedra, Kristina Reppond, Madison N. Shyer, Jessica Cambric, Ryan Gadd, Akanksha Batajoo, Rashi Thakur, Sindy Pena, Regan |
| hCoV-19/SouthAfrica/NICD-N23660/2021 | EPI_ISL_9201366 | 2021-12-05 | VERMAAK | National Institute for Communicable Diseases of the National Health Laboratory Service | Amoako DG, Everatt J, Kekana D, Scheepers C, Mohale T, Ntuli N, Mahlangu B, Mnguni A, Ismail A, Bhiman JN, Wolter N |
| hCoV-19/SouthAfrica/NICD-N24029/2021 | EPI_ISL_9201388 | 2021-11-30 | HELEN JOSEPH LABORATORY | National Institute for Communicable Diseases of the National Health Laboratory Service | Amoako DG, Everatt J, Kekana D, Scheepers C, Mohale T, Ntuli N, Mahlangu B, Mnguni A, Ismail A, Bhiman JN, Wolter N |
| hCoV-19/SouthAfrica/NICD-N25049/2021 | EPI_ISL_9201460 | 2021-11-29 | INDALOBIO (PTY) LTD | National Institute for Communicable Diseases of the National Health Laboratory Service | Amoako DG, Everatt J, Kekana D, Scheepers C, Mohale T, Ntuli N, Mahlangu B, Mnguni A, Ismail A, Bhiman JN, Wolter N |
| hCoV-19/SouthAfrica/NICD-N25376/2021 | EPI_ISL_9201500 | 2021-12-14 | KIMBERLEY LABORATORY | National Institute for Communicable Diseases of the National Health Laboratory Service | Amoako DG, Everatt J, Kekana D, Scheepers C, Mohale T, Ntuli N, Mahlangu B, Mnguni A, Ismail A, Bhiman JN, Wolter N |
| hCoV-19/SouthAfrica/NICD-N28566/2021 | EPI_ISL_9201591 | 2021-12-24 | LANCET | National Institute for Communicable Diseases of the National Health Laboratory Service | Amoako DG, Everatt J, Kekana D, Scheepers C, Mohale T, Ntuli N, Mahlangu B, Mnguni A, Ismail A, Bhiman JN, Wolter N |
| hCoV-19/SouthAfrica/NICD-N28587/2021 | EPI_ISL_9201612 | 2021-12-30 | LANCET | National Institute for Communicable Diseases of the National Health Laboratory Service | Amoako DG, Everatt J, Kekana D, Scheepers C, Mohale T, Ntuli N, Mahlangu B, Mnguni A, Ismail A, Bhiman JN, Wolter N |
| hCoV-19/Japan/PG-173542/2021 | EPI_ISL_9207480 | 2021-12-09 | Pathogen Genomics Center, National Institute of Infectious Diseases | Pathogen Genomics Center, National Institute of Infectious Diseases | Tsuyoshi Sekizuka, Kentaro Itokawa, Rina Tanaka, Masanori Hashino, Makoto Kuroda |
| hCoV-19/Japan/PG-175503/2021 | EPI_ISL_9207843 | 2021-12-23 | Pathogen Genomics Center, National Institute of Infectious Diseases | Pathogen Genomics Center, National Institute of Infectious Diseases | Tsuyoshi Sekizuka, Kentaro Itokawa, Rina Tanaka, Masanori Hashino, Makoto Kuroda |
| hCoV-19/Japan/PG-179772/2021 | EPI_ISL_9209138 | 2021-01-07 | Pathogen Genomics Center, National Institute of Infectious Diseases | Pathogen Genomics Center, National Institute of Infectious Diseases | Tsuyoshi Sekizuka, Kentaro Itokawa, Rina Tanaka, Masanori Hashino, Makoto Kuroda |

| | | | | | |
|---|---|---|---|---|---|
| hCoV-19/Japan/PG-179964/2021 | EPI_ISL_9209750 | 2021-01-06 | Pathogen Genomics Center, National Institute of Infectious Diseases | Pathogen Genomics Center, National Institute of Infectious Diseases | Tsuyoshi Sekizuka, Kentaro Itokawa, Rina Tanaka, Masanori Hashino, Makoto Kuroda |
| hCoV-19/Japan/PG-180032/2021 | EPI_ISL_9209943 | 2021-01-07 | Pathogen Genomics Center, National Institute of Infectious Diseases | Pathogen Genomics Center, National Institute of Infectious Diseases | Tsuyoshi Sekizuka, Kentaro Itokawa, Rina Tanaka, Masanori Hashino, Makoto Kuroda |
| hCoV-19/Japan/PG-174699/2021 | EPI_ISL_9215000 | 2021-12-28 | Kyoto City Institute of Public Health and Environmental Sciences | Kyoto City Institute of Public Health and Environmental Sciences | Yuta Tsukamoto, Akane Furuichi, Michiko Hayashi, Toshiko Fukushima, Masayoshi Watanabe, Manami Nakai, Tsuyoshi Sekizuka, Kentaro Itokawa, Rina Tanaka, Masanori Hashino, Makoto Kuroda |
| hCoV-19/Brazil/CE-FIOCRUZ-51014CE/2021 | EPI_ISL_9215919 | 2021-12-29 | UBASF AUTODROMO | Analytical Competence Molecular Epidemiology Lab/ACME, Oswaldo Cruz Foundation, Ceara (FIOCRUZ CE) | Fabio Miyajima, Veridiana Pessoa Miyajima, Fernando Braga Stehling, Thais de Oliveira Costa, Thais Ferreira de Oliveira, Joaquim Cesar do Nascimento Sousa Junior, Jamille Maria Mendes Bezerra, Ticiane Cavalcante de Souza, Carlos Leonardo de Aragao Araujo, |
| hCoV-19/Japan/PG-174362/2021 | EPI_ISL_9216986 | 2021-09-04 | Hiroshima Prefectural Technology Research Institute, Public Health and Environment Center | Hiroshima Prefectural Technology Research Institute, Public Health and Environment Center | Naoki Shigemoto, Akie Nakashima, Yukie Shimazu, Hiroko Akita, Syuhei Ikeda, Kanako Masuda, Takahiro Hiratsuka, Mana Suei, Ayano Ito, Shinichi Takao, Tsuyoshi Sekizuka, Kentaro Itokawa, Rina Tanaka, Masanori Hashino, Makoto Kuroda |
| hCoV-19/Japan/PG-174365/2021 | EPI_ISL_9216996 | 2021-12-24 | Hiroshima Prefectural Technology Research Institute, Public Health and Environment Center | Hiroshima Prefectural Technology Research Institute, Public Health and Environment Center | Naoki Shigemoto, Akie Nakashima, Yukie Shimazu, Hiroko Akita, Syuhei Ikeda, Kanako Masuda, Takahiro Hiratsuka, Mana Suei, Ayano Ito, Shinichi Takao, Tsuyoshi Sekizuka, Kentaro Itokawa, Rina Tanaka, Masanori Hashino, Makoto Kuroda |
| hCoV-19/Japan/PG-173705/2021 | EPI_ISL_9218404 | 2021-12-19 | Saitama Institute of Public Health | Saitama Institute of Public Health | Satoshi Daisaki, Eri Aonuma, Hayato Ehara, Tsuyoshi Sekizuka, Kentaro Itokawa, Rina Tanaka, Masanori Hashino, Makoto Kuroda |
| hCoV-19/Japan/PG-175855/2021 | EPI_ISL_9221540 | 2021-12-30 | Kanagawa Prefectural Institute of Public Health | Kanagawa Prefectural Institute of Public Health | Takayuki Hishiki, Rieko Suzuki, Takako Sano, Makiko Kondo, Sumi Watanabe, Itsumi Toyokura, Junichi Sakuragi, Tomohiko Takasaki, Tsuyoshi Sekizuka, Kentaro Itokawa, Rina Tanaka, Masanori Hashino, Makoto Kuroda |
| hCoV-19/Japan/PG-175858/2021 | EPI_ISL_9221549 | 2021-12-26 | Kanagawa Prefectural Institute of Public Health | Kanagawa Prefectural Institute of Public Health | Takayuki Hishiki, Rieko Suzuki, Takako Sano, Makiko Kondo, Sumi Watanabe, Itsumi Toyokura, Junichi Sakuragi, Tomohiko Takasaki, Tsuyoshi Sekizuka, Kentaro Itokawa, Rina Tanaka, Masanori Hashino, Makoto Kuroda |
| hCoV-19/Japan/PG-174340/2021 | EPI_ISL_9224815 | 2021-12-02 | Kawasaki City Institute for Public Health | Kawasaki City Institute for Public Health | Kuniharu Sasaki, Kouhei Natsui, Chihiro Arai, Azumi Wakana, Ayako Komane, Hideaki Shimizu, Tsuyoshi Sekizuka, Kentaro Itokawa, Rina Tanaka, Masanori Hashino, Makoto Kuroda |
| hCoV-19/Japan/PG-173699/2021 | EPI_ISL_9225501 | 2021-12-15 | Division of Microbiology, Osaka Institute of Public Health | Division of Microbiology, Osaka Institute of Public Health | Ryuji Kawahara, Satoshi Hiroi, Saeko Morikawa, Haruyo Mori, Kazushi Motomura, Jun Ogasawara, Tsuyoshi Sekizuka, Kentaro Itokawa, Rina Tanaka, Masanori Hashino, Makoto Kuroda |
| hCoV-19/Germany/NW-RKI-I-470864/2021 | EPI_ISL_9226004 | 2021-12-16 | Labor ZOTZ\|KLIMAS; MVZ Dvºsseldorf-Centrum | Robert Koch Institute | unknown |
| hCoV-19/Germany/NW-RKI-I-470937/2021 | EPI_ISL_9226140 | 2021-12-22 | Labor ZOTZ\|KLIMAS; MVZ Dvºsseldorf-Centrum | Robert Koch Institute | unknown |

| | | | | | |
|---|---|---|---|---|---|
| hCoV-19/England/NORT-YBUU9G/2021 | EPI_ISL_9227671 | 2021-12-29 | Northumbria University / South Tees Hospitals NHS Foundation Trust / North Cumbria Integrated Care NHS Foundation Trust / North Tees and Hartlepool NHS Foundation Trust / Newcastle Hospitals NHS Foundation Trust | COVID-19 Genomics UK (COG-UK) Consortium | Darren L Smith,Andrew Nelson,Matthew Bashton,Greg R Young,Joshua Loh,John Allan,Mohammad A Tariq,Giles S Holt,Gary Black,Wen C Yew,Lynn Dover,Paul Baker,Steve Liggett,Sarah Essex,Jane Greenaway,Debra Padgett,Clive Graham,Garren Scott,Edward Barton,Emma Sw |
| hCoV-19/England/NORT-YBUZR3/2021 | EPI_ISL_9227884 | 2021-02-05 | Northumbria University / South Tees Hospitals NHS Foundation Trust / North Cumbria Integrated Care NHS Foundation Trust / North Tees and Hartlepool NHS Foundation Trust / Newcastle Hospitals NHS Foundation Trust | COVID-19 Genomics UK (COG-UK) Consortium | Darren L Smith,Andrew Nelson,Matthew Bashton,Greg R Young,Joshua Loh,John Allan,Mohammad A Tariq,Giles S Holt,Gary Black,Wen C Yew,Lynn Dover,Paul Baker,Steve Liggett,Sarah Essex,Jane Greenaway,Debra Padgett,Clive Graham,Garren Scott,Edward Barton,Emma Sw |
| hCoV-19/India/AP-CCMB-CIC0459/2021 | EPI_ISL_9232179 | 2021-12-30 | AP SSO | CSIR-Centre for Cellular and Molecular Biology-INSACOG | Jandhyala Sai Krishna,Tulasi Nagabandi, Lamuk Zaveri,Malini Nemalikanti,Sreelekshmi MS, Ara Sreenivas,Shreekant Verma, Amareshwar Vodapalli,B Himasri,Valli Nagalakshmi Undamatla, Sumedha Avadhanula,Priya Nurkurthy,Vidhyadhari Methuku, Archana Bharadwaj Si |
| hCoV-19/India/TN-CLRI-CIC0568/2020 | EPI_ISL_9232324 | 2020-09-05 | CSIR-Central Leather Research Institute | CSIR-Centre for Cellular and Molecular Biology-INSACOG | Lamuk Zaveri,Tulasi Nagabandi,Jandhyala Sai Krishna,Malini Nemalikanti,Sreelekshmi MS, Ara Sreenivas,Shreekant Verma, Amareshwar Vodapalli,B Himasri,Valli Nagalakshmi Undamatla, Sumedha Avadhanula,Priya Nurkurthy,Vidhyadhari Methuku,A Gnanamani,Preethi V, |
| hCoV-19/India/TN-CLRI-CIC0594/2020 | EPI_ISL_9232349 | 2020-09-12 | CSIR-Central Leather Research Institute | CSIR-Centre for Cellular and Molecular Biology-INSACOG | Lamuk Zaveri,Tulasi Nagabandi,Jandhyala Sai Krishna,Malini Nemalikanti,Sreelekshmi MS, Ara Sreenivas,Shreekant Verma, Amareshwar Vodapalli,B Himasri,Valli Nagalakshmi Undamatla, Sumedha Avadhanula,Priya Nurkurthy,Vidhyadhari Methuku,A Gnanamani,Preethi V, |
| hCoV-19/India/TG-CCMB-CIC0181/2021 | EPI_ISL_9232477 | 2021-12-30 | CSIR-Centre for Cellular and Molecular Biology | CSIR-Centre for Cellular and Molecular Biology-INSACOG | Malini Nemalikanti,Lamuk Zaveri,Tulasi Nagabandi,Jandhyala Sai Krishna,Sreelekshmi MS, Ara Sreenivas,Shreekant Verma, Amareshwar Vodapalli,B Himasri,Valli Nagalakshmi Undamatla, Sumedha Avadhanula,Priya Nurkurthy,Vidhyadhari Methuku, Archana Bharadwaj Siv |
| hCoV-19/Japan/PG-176034/2021 | EPI_ISL_9235867 | 2021-11-30 | Wakayama Prefectural Research Center of Environment and Public Health | Wakayama Prefectural Research Center of Environment and Public Health | Fumio Terasoma, Akari Kajino, Yosuke Hamajima, Yasuyuki Hujimoto, Hideyuki Yanai, Tsuyoshi Sekizuka, Kentaro Itokawa, Rina Tanaka, Masanori Hashino, Makoto Kuroda |
| hCoV-19/India/UP-NBRI-3-1-22-83-CWS-980/2021 | EPI_ISL_9244100 | 2021-12-30 | King George's Medical University | CSIR-National Botanical Research Institute | Priti Prasad, Shantanu Prakash, Babita, Tripti Rungta, Suruchi Shukla, Sumit K Bag, Amita Jain, Samir V Sawant |
| hCoV-19/India/UP-NBRI-3-1-22-81-CWS-978/2021 | EPI_ISL_9244117 | 2021-12-29 | King George's Medical University | CSIR-National Botanical Research Institute | Priti Prasad, Shantanu Prakash, Babita, Tripti Rungta, Suruchi Shukla, Sumit K Bag, Amita Jain, Samir V Sawant |

| | | | | | |
|---|---|---|---|---|---|
| hCoV-19/Brazil/SC-UFSC-52-84/2021 | EPI_ISL_9248465 | 2021-12-30 | SMS - S√ÉO JOS√â | Laborat√≥rio de Bioinform√°tica - Universidade Federal de Santa Catarina | Glauber Wagner, Juliana Righetto Moser, Eric Kazuo Kawagoe, Milene Moehr de Moraes, Vilmar Benetti Filho, Dayane Azevedo Padilha, Fernando Hartmann Barazzetti, Marcos Andr√© Sch√∂rner, Doris Sobral Marques Souza, Ariane Nicaretta Amorim, Darcita Buerger R |
| hCoV-19/USA/AZ-TG846866/2021 | EPI_ISL_9260484 | 2021-04-11 | TGen North | TGen North | Jolene Bowers, Heather Centner, Brett Van Tassel, Chris French, Hayley Yaglom, Darrin Lemmer, Dave Engelthaler |
| hCoV-19/USA/AZ-TG1145865/2021 | EPI_ISL_9261192 | 2021-12-09 | TGen North | TGen North | Jolene Bowers, Heather Centner, Brett Van Tassel, Chris French, Hayley Yaglom, Darrin Lemmer, Dave Engelthaler |
| hCoV-19/USA/VA-Curative-139227/2021 | EPI_ISL_9261465 | 2021-12-18 | Curative Labs | Curative Labs | Mikhail Hanewich-Hollatz, J.Cesar Ignacio-Espinoza, Elias L. Salfati, Nabjot Sandhu, George Way, Zhiyi Xie, Eugenia Khorosheva, Sophia Quasem, Janet Chen, Vladimir Slepnev |
| hCoV-19/USA/ME-HETL-J11433/2021 | EPI_ISL_9263962 | 2021-01-27 | Maine Health and Environmental Testing Laboratory | Tewhey Lab, The Jackson Laboratory | Matluk,N., Dewey,H., Iosue,F., Barter,M., Lynch,R., Munger,H. and Tewhey,R. |
| hCoV-19/Brazil/SP-HIAE-ID1099/2021 | EPI_ISL_9265915 | 2021-12-29 | LATE - Laborat√≥rio de T√©cnicas Especiais - Hospital Israelita Albert Einstein | LATE - Laborat√≥rio de T√©cnicas Especiais - Hospital Israelita Albert Einstein | Deyvid Amgarten, Fernanda de Mello Malta, Raquel Riyuzo, Ana Paula Moreira Salles, Luiz Vicente Rizzo, Pedro Henrique Sebe Rodrigues, Alexandre Hideaki Takara, Anelisie da Silva Santos, Erick Gustavo Dorlass, Marcio Anunciacao Menezes, Jo√£o Renato Rebell |
| hCoV-19/India/TG-CDFD-71284/2021 | EPI_ISL_9274459 | 2021-12-08 | District Hospital, Warangal | Centre for DNA Fingerprinting and Diagnostics | Asmita Gupta, Rupin Shelake, Arunkumar Karunanidhi, Vinay Donipadi, Divya Vashisht, Ashwin Dalal, Nagmani, Murali Bashyam |
| hCoV-19/Chad/TCH-34/2021 | EPI_ISL_9274870 | 2021-08-09 | Minist√®re de Sant√© Publique et de la Solidarit√© Nationale | Pathogen Genomics Lab, National Institute for Biomedical Research (INRB) | Placide Mbala-Kingebeni, Frank Edgard ZONGO Ragomzingba, Edith Nkwembe, Eddy Kinganda-Lusamaki, Amuri Aziza, Francisca Muyembe Mawete, Emmanuel Lokilo Lofiko, Gradi Luakanda, Jean Claude Makangara, Raphael Lumembe, Gabriel Kabamba, Catherine Pratt, Matthi |
| hCoV-19/Chad/TCH-43/2021 | EPI_ISL_9274877 | 2021-11-05 | Minist√®re de Sant√© Publique et de la Solidarit√© Nationale | Pathogen Genomics Lab, National Institute for Biomedical Research (INRB) | Placide Mbala-Kingebeni, Frank Edgard ZONGO Ragomzingba, Edith Nkwembe, Eddy Kinganda-Lusamaki, Amuri Aziza, Francisca Muyembe Mawete, Emmanuel Lokilo Lofiko, Gradi Luakanda, Jean Claude Makangara, Raphael Lumembe, Gabriel Kabamba, Catherine Pratt, Matthi |
| hCoV-19/Chad/TCH-45/2021 | EPI_ISL_9274879 | 2021-11-05 | Minist√®re de Sant√© Publique et de la Solidarit√© Nationale | Pathogen Genomics Lab, National Institute for Biomedical Research (INRB) | Placide Mbala-Kingebeni, Frank Edgard ZONGO Ragomzingba, Edith Nkwembe, Eddy Kinganda-Lusamaki, Amuri Aziza, Francisca Muyembe Mawete, Emmanuel Lokilo Lofiko, Gradi Luakanda, Jean Claude Makangara, Raphael Lumembe, Gabriel Kabamba, Catherine Pratt, Matthi |

| | | | | | |
|---|---|---|---|---|---|
| hCoV-19/Chad/TCH-48/2021 | EPI_ISL_9274881 | 2021-11-08 | Ministv®re de Santv© Publique et de la Solidaritv© Nationale | Pathogen Genomics Lab, National Institute for Biomedical Research (INRB) | Placide Mbala-Kingebeni, Frank Edgard ZONGO Ragomzingba, Edith Nkwembe, Eddy Kinganda-Lusamaki, Amuri Aziza, Francisca Muyembe Mawete, Emmanuel Lokilo Lofiko, Gradi Luakanda, Jean Claude Makangara, Raphael Lumembe, Gabriel Kabamba, Catherine Pratt, Matthi |
| hCoV-19/Chad/TCH-49/2021 | EPI_ISL_9274882 | 2021-11-09 | Ministv®re de Santv© Publique et de la Solidaritv© Nationale | Pathogen Genomics Lab, National Institute for Biomedical Research (INRB) | Placide Mbala-Kingebeni, Frank Edgard ZONGO Ragomzingba, Edith Nkwembe, Eddy Kinganda-Lusamaki, Amuri Aziza, Francisca Muyembe Mawete, Emmanuel Lokilo Lofiko, Gradi Luakanda, Jean Claude Makangara, Raphael Lumembe, Gabriel Kabamba, Catherine Pratt, Matthi |
| hCoV-19/Chad/TCH-54/2021 | EPI_ISL_9274887 | 2021-11-05 | Ministv®re de Santv© Publique et de la Solidaritv© Nationale | Pathogen Genomics Lab, National Institute for Biomedical Research (INRB) | Placide Mbala-Kingebeni, Frank Edgard ZONGO Ragomzingba, Edith Nkwembe, Eddy Kinganda-Lusamaki, Amuri Aziza, Francisca Muyembe Mawete, Emmanuel Lokilo Lofiko, Gradi Luakanda, Jean Claude Makangara, Raphael Lumembe, Gabriel Kabamba, Catherine Pratt, Matthi |
| hCoV-19/Kyrgyzstan/Kyrg361310-090721/2021 | EPI_ISL_9288140 | 2021-07-09 | AQUA Lab | SRC VB "Vector", "Collection of microorganisms" Department. | Sergey A. Bodnev, Anastasiya M. Smirnova, Tatyana V. Tregubchak, Alexander N. Shvalov, Damira O.Ashyralieva, Kalila Zhumalieva, Irina A.Copova |
| hCoV-19/Kyrgyzstan/Kyrg512120-170721/2021 | EPI_ISL_9288153 | 2021-07-17 | National Virology Lab. | SRC VB "Vector", "Collection of microorganisms" Department. | Sergey A. Bodnev, Anastasiya M. Smirnova, Tatyana V. Tregubchak, Alexander N. Shvalov, Damira O.Ashyralieva, Kalila Zhumalieva. |
| hCoV-19/Mayotte/HCL022005374401/2021 | EPI_ISL_9314965 | 2021-12-26 | CENTRE HOSPITALIER DE MAYOTTE | CNR Virus des Infections Respiratoires - France SUD | Antonin Bal, Bruno Simon, Thv©ophile Boyer, Gregory Destras, Gwendolyne Burfin, Hadrien Regue, Quentin Semanas, Martine Valette, Bruno Lina, Laurence Josset |
| hCoV-19/Philippines/PH-PGC-102568/2021 | EPI_ISL_9320760 | 2021-12-26 | Philippine Red Cross Clark Molecular Laboratory | Philippine Genome Center | Cynthia P. Saloma,Jan Michael C. Yap,Benedict A. Maralit,Marc Edsel C. Ayes,Eva Maria Cutiongco-de la Paz,Francis A. Tablizo,Carlo M. Lapid,El King D. Morado,Joshua Gregor A. Dizon,Maria Sofia L. Yangzon,Elcid Aaron R. Pangilinan,Renato Jacinto Q. Mantari |
| hCoV-19/Philippines/PH-PGC-102577/2021 | EPI_ISL_9320816 | 2021-12-22 | Philippine Red Cross Clark Molecular Laboratory | Philippine Genome Center | Cynthia P. Saloma,Jan Michael C. Yap,Benedict A. Maralit,Marc Edsel C. Ayes,Eva Maria Cutiongco-de la Paz,Francis A. Tablizo,Carlo M. Lapid,El King D. Morado,Joshua Gregor A. Dizon,Maria Sofia L. Yangzon,Elcid Aaron R. Pangilinan,Renato Jacinto Q. Mantari |
| hCoV-19/Philippines/PH-PGC-102582/2021 | EPI_ISL_9320829 | 2021-12-27 | PRC Subic Molecular Laboratory | Philippine Genome Center | Cynthia P. Saloma,Jan Michael C. Yap,Benedict A. Maralit,Marc Edsel C. Ayes,Eva Maria Cutiongco-de la Paz,Francis A. Tablizo,Carlo M. Lapid,El King D. Morado,Joshua Gregor A. Dizon,Maria Sofia L. Yangzon,Elcid Aaron R. Pangilinan,Renato Jacinto Q. Mantari |

| | | | | | |
|---|---|---|---|---|---|
| hCoV-19/Philippines/PH-PGC-102719/2021 | EPI_ISL_9320961 | 2021-12-29 | Philippine Airport Diagnostic Laboratory | Philippine Genome Center | Cynthia P. Saloma,Jan Michael C. Yap,Benedict A. Maralit,Marc Edsel C. Ayes,Eva Maria Cutiongco-de la Paz,Francis A. Tablizo,Carlo M. Lapid,El King D. Morado,Joshua Gregor A. Dizon,Maria Sofia L. Yangzon,Elcid Aaron R. Pangilinan,Renato Jacinto Q. Mantari |
| hCoV-19/Brazil/RJ-LNN07834/2021 | EPI_ISL_9321218 | 2021-12-29 | Laboratorio Central Noel Nutels | Bioinformatics Laboratory / LNCC | Alessandra P Lamarca, Luiz G P de Almeida, Ronaldo da Silva Francisco Junior, Liliane Cavalcante, Otavio Brustolini, Alexandra L Gerber, Ana Paula de C Guimaraes, Thiago Henrique de Oliveira, Erica Ramos dos Santos Nascimento, Cintia Policarpo, Mario Serg |
| hCoV-19/Brazil/RJ-LNN07940/2021 | EPI_ISL_9321313 | 2021-12-22 | Unidade de apoio ao diagnostico da COVID - UNADIG | Bioinformatics Laboratory / LNCC | Alessandra P Lamarca, Luiz G P de Almeida, Ronaldo da Silva Francisco Junior, Liliane Cavalcante, Otavio Brustolini, Alexandra L Gerber, Ana Paula de C Guimaraes, Thiago Henrique de Oliveira, Erica Ramos dos Santos Nascimento, Cintia Policarpo, Mario Serg |
| hCoV-19/Brazil/RJ-LNN07997/2021 | EPI_ISL_9321370 | 2021-12-29 | Unidade de apoio ao diagnostico da COVID - UNADIG | Bioinformatics Laboratory / LNCC | Alessandra P Lamarca, Luiz G P de Almeida, Ronaldo da Silva Francisco Junior, Liliane Cavalcante, Otavio Brustolini, Alexandra L Gerber, Ana Paula de C Guimaraes, Thiago Henrique de Oliveira, Erica Ramos dos Santos Nascimento, Cintia Policarpo, Mario Serg |
| hCoV-19/Philippines/PH-PGC-102848/2021 | EPI_ISL_9321465 | 2021-12-28 | Valenzuela Hope Molecular Laboratory | Philippine Genome Center | Cynthia P. Saloma,Jan Michael C. Yap,Benedict A. Maralit,Marc Edsel C. Ayes,Eva Maria Cutiongco-de la Paz,Francis A. Tablizo,Carlo M. Lapid,El King D. Morado,Joshua Gregor A. Dizon,Maria Sofia L. Yangzon,Elcid Aaron R. Pangilinan,Renato Jacinto Q. Mantari |
| hCoV-19/Philippines/PH-PGC-102849/2021 | EPI_ISL_9321468 | 2021-12-28 | Valenzuela Hope Molecular Laboratory | Philippine Genome Center | Cynthia P. Saloma,Jan Michael C. Yap,Benedict A. Maralit,Marc Edsel C. Ayes,Eva Maria Cutiongco-de la Paz,Francis A. Tablizo,Carlo M. Lapid,El King D. Morado,Joshua Gregor A. Dizon,Maria Sofia L. Yangzon,Elcid Aaron R. Pangilinan,Renato Jacinto Q. Mantari |
| hCoV-19/Philippines/PH-PGC-103010/2021 | EPI_ISL_9321607 | 2021-12-30 | Lung Center Of The Philippines (LCP) | Philippine Genome Center | Cynthia P. Saloma,Jan Michael C. Yap,Benedict A. Maralit,Marc Edsel C. Ayes,Eva Maria Cutiongco-de la Paz,Francis A. Tablizo,Carlo M. Lapid,El King D. Morado,Joshua Gregor A. Dizon,Maria Sofia L. Yangzon,Elcid Aaron R. Pangilinan,Renato Jacinto Q. Mantari |
| hCoV-19/Philippines/PH-PGC-103027/2021 | EPI_ISL_9321733 | 2021-12-29 | Lung Center Of The Philippines (LCP) | Philippine Genome Center | Cynthia P. Saloma,Jan Michael C. Yap,Benedict A. Maralit,Marc Edsel C. Ayes,Eva Maria Cutiongco-de la Paz,Francis A. Tablizo,Carlo M. Lapid,El King D. Morado,Joshua Gregor A. Dizon,Maria Sofia L. Yangzon,Elcid Aaron R. Pangilinan,Renato Jacinto Q. Mantari |

| | | | | | |
|---|---|---|---|---|---|
| hCoV-19/Philippines/PH-PGC-101512/2021 | EPI_ISL_9322735 | 2021-11-24 | Baguio General Hospital And Medical Center | Philippine Genome Center | Cynthia P. Saloma,Jan Michael C. Yap,Benedict A. Maralit,Marc Edsel C. Ayes,Eva Maria Cutiongco-de la Paz,Francis A. Tablizo,Carlo M. Lapid,El King D. Morado,Joshua Gregor A. Dizon,Maria Sofia L. Yangzon,Elcid Aaron R. Pangilinan,Renato Jacinto Q. Mantari |
| hCoV-19/Philippines/PH-PGC-101528/2021 | EPI_ISL_9322738 | 2021-11-22 | Baguio General Hospital And Medical Center | Philippine Genome Center | Cynthia P. Saloma,Jan Michael C. Yap,Benedict A. Maralit,Marc Edsel C. Ayes,Eva Maria Cutiongco-de la Paz,Francis A. Tablizo,Carlo M. Lapid,El King D. Morado,Joshua Gregor A. Dizon,Maria Sofia L. Yangzon,Elcid Aaron R. Pangilinan,Renato Jacinto Q. Mantari |
| hCoV-19/Philippines/PH-PGC-102877/2021 | EPI_ISL_9322784 | 2021-12-29 | Laguna Provincial Hospital - San Pablo City District Hospital | Philippine Genome Center | Cynthia P. Saloma,Jan Michael C. Yap,Benedict A. Maralit,Marc Edsel C. Ayes,Eva Maria Cutiongco-de la Paz,Francis A. Tablizo,Carlo M. Lapid,El King D. Morado,Joshua Gregor A. Dizon,Maria Sofia L. Yangzon,Elcid Aaron R. Pangilinan,Renato Jacinto Q. Mantari |
| hCoV-19/Philippines/PH-PGC-103128/2021 | EPI_ISL_9322838 | 2021-12-27 | Baguio General Hospital And Medical Center | Philippine Genome Center | Cynthia P. Saloma,Jan Michael C. Yap,Benedict A. Maralit,Marc Edsel C. Ayes,Eva Maria Cutiongco-de la Paz,Francis A. Tablizo,Carlo M. Lapid,El King D. Morado,Joshua Gregor A. Dizon,Maria Sofia L. Yangzon,Elcid Aaron R. Pangilinan,Renato Jacinto Q. Mantari |
| hCoV-19/Philippines/PH-PGC-103317/2021 | EPI_ISL_9322889 | 2021-12-30 | Army General Hospital Molecular Laboratory (AGHML) | Philippine Genome Center | Cynthia P. Saloma,Jan Michael C. Yap,Benedict A. Maralit,Marc Edsel C. Ayes,Eva Maria Cutiongco-de la Paz,Francis A. Tablizo,Carlo M. Lapid,El King D. Morado,Joshua Gregor A. Dizon,Maria Sofia L. Yangzon,Elcid Aaron R. Pangilinan,Renato Jacinto Q. Mantari |
| hCoV-19/Philippines/PH-PGC-103743/2021 | EPI_ISL_9323211 | 2021-12-29 | University Of The East Ramon Magsaysay Memorial Medical Center (UERM) | Philippine Genome Center | Cynthia P. Saloma,Jan Michael C. Yap,Benedict A. Maralit,Marc Edsel C. Ayes,Eva Maria Cutiongco-de la Paz,Francis A. Tablizo,Carlo M. Lapid,El King D. Morado,Joshua Gregor A. Dizon,Maria Sofia L. Yangzon,Elcid Aaron R. Pangilinan,Renato Jacinto Q. Mantari |
| hCoV-19/Philippines/PH-PGC-103749/2021 | EPI_ISL_9323216 | 2021-12-30 | University Of The East Ramon Magsaysay Memorial Medical Center (UERM) | Philippine Genome Center | Cynthia P. Saloma,Jan Michael C. Yap,Benedict A. Maralit,Marc Edsel C. Ayes,Eva Maria Cutiongco-de la Paz,Francis A. Tablizo,Carlo M. Lapid,El King D. Morado,Joshua Gregor A. Dizon,Maria Sofia L. Yangzon,Elcid Aaron R. Pangilinan,Renato Jacinto Q. Mantari |
| hCoV-19/India/MZ-INSACOG-1526100736692/2021 | EPI_ISL_9323714 | 2021-10-11 | INSACOG-Mizoram | National Institute of Biomedical Genomics ‚Äì INSACOG | Arindam Maitra, Swagnik Roy, Nidhan Kumar Biswas, Gracy Laldinmawii, Sreedhar Chinnaswamy, N Senthil Kumar, Saumitra Das |

| | | | | | |
|---|---|---|---|---|---|
| hCoV-19/India/KA-SEQ_9092/2021 | EPI_ISL_9324293 | 2021-12-17 | Kims Hubli | inStem, NCBS-INSACOG | Uma Ramakrishnan, Dasaradhi Palakodeti, Aswin SaiNarain |
| hCoV-19/Thailand/NIC_CMI_SEQ25037/2021 | EPI_ISL_9324388 | 2021-12-27 | Regional Medical Sciences Center 1 Chiang Mai | National Institute of Health, Department of Medical Sciences, Ministry of Public Health, Thailand | Pilailuk Okada; Siripaporn Phuygun; Nuttida Thongpramul; Pakorn Piromtong; Natchaya¬† Khiadsang; Thanutsapa Thanadachakul; Sirikanda Wimol; Suratchana Mitrat; Sunthareeya Waicharoen; Archawin Rojanawiwat |
| hCoV-19/SouthAfrica/CERI-KRISP-K034314/2021 | EPI_ISL_9325960 | 2021-12-24 | National Health Laboratory Services, Virology | and Innovation, Stellenbosch University and KRISP, KZN Research Innovation and Sequencing Platform, UKZN. | Subramoney K, Treurnicht F, Mtileni N, Bharuthram A, Giandhari J, Pillay S, Naidoo Y, Ramphal U, Ramphal Y, Moir M, van Wyk S, San JE, Tshiabuila D, Tegally H, Wilkinson E, de Oliveira T |
| hCoV-19/SouthAfrica/CERI-KRISP-K034345/2021 | EPI_ISL_9325988 | 2021-12-28 | National Health Laboratory Services, Virology | and Innovation, Stellenbosch University and KRISP, KZN Research Innovation and Sequencing Platform, UKZN. | Subramoney K, Treurnicht F, Mtileni N, Bharuthram A, Giandhari J, Pillay S, Naidoo Y, Ramphal U, Ramphal Y, Moir M, van Wyk S, San JE, Tshiabuila D, Tegally H, Wilkinson E, de Oliveira T |
| hCoV-19/SouthAfrica/CERI-KRISP-K034364/2021 | EPI_ISL_9326005 | 2021-12-29 | National Health Laboratory Services, Virology | and Innovation, Stellenbosch University and KRISP, KZN Research Innovation and Sequencing Platform, UKZN. | Subramoney K, Treurnicht F, Mtileni N, Bharuthram A, Giandhari J, Pillay S, Naidoo Y, Ramphal U, Ramphal Y, Moir M, van Wyk S, San JE, Tshiabuila D, Tegally H, Wilkinson E, de Oliveira T |
| hCoV-19/SouthAfrica/CERI-KRISP-K033749/2021 | EPI_ISL_9328110 | 2021-12-17 | National Health Laboratory Service, Kwazulu-Natal, South Africa | and Innovation, Stellenbosch University and KRISP, KZN Research Innovation and Sequencing Platform, UKZN. | Nokukhanya Mdlalose, Giandhari J, Pillay S, Naidoo Y, Ramphal U, Ramphal Y, Maharaj A, San JE, Moir M, Tshiabuila D, Tegally H, Wilkinson E, de Oliveira T |
| hCoV-19/Mauritius/CERI-KRISP-K034987/2021 | EPI_ISL_9328447 | 2021-12-27 | Victoria Hospital | CERI, Centre for Epidemic Response and Innovation, Stellenbosch University and KRISP, KZN Research Innovation and Sequencing Platform, UKZN. | Manraj SS, Sonoo J, Bahadoor BS,Ubheeram A, Francois A, Jannoo N,Bacorisen A, Ramuth M, Giandhari J, Pillay S, Naidoo Y, Naicker D, Maharaj A, Anyaneji UJ, Moir M, Van Wyk S, San JE, Tshiabuila D, Tegally H, Wilkinson E, de Oliveira T |
| hCoV-19/PuertoRico/PR-CDC-2-5441370/2021 | EPI_ISL_9329077 | 2021-01-03 | PR Public Health Lab | Centers for Disease Control and Prevention Division of Viral Diseases, Pathogen Discovery | Mili Sheth,Sarah Nobles,Joseph Madden,Jasmine Padilla,Alex Burgin,Meghan Bentz,Mark Burroughs,Peter Cook,Clinton Paden,Dhwani Batra,Kristen Knipe,Dakota Howard,Yvette Unoarumhi,Matthew Schmerer,Ben Rambo-Martin,Kristine Lacek,Sam Shepard,Dave Wentworth,Vi |
| hCoV-19/PuertoRico/PR-CDC-2-5441299/2021 | EPI_ISL_9329079 | 2021-01-03 | PR Public Health Lab | Centers for Disease Control and Prevention Division of Viral Diseases, Pathogen Discovery | Mili Sheth,Sarah Nobles,Joseph Madden,Jasmine Padilla,Alex Burgin,Meghan Bentz,Mark Burroughs,Peter Cook,Clinton Paden,Dhwani Batra,Kristen Knipe,Dakota Howard,Yvette Unoarumhi,Matthew Schmerer,Ben Rambo-Martin,Kristine Lacek,Sam Shepard,Dave Wentworth,Vi |
| hCoV-19/USA/GA-CDC-MMB12565473/2021 | EPI_ISL_9330943 | 2021-12-29 | Mako Medical | Centers for Disease Control and Prevention Division of Viral Diseases, Pathogen Discovery | Dakota Howard,Dhwani Batra,Peter Cook,Jason Caravas,Benjamin Rambo-Martin,Scott Sammons,Yvette Unoarumhi,Matthew Schmerer,Kristine Lacek,Tymeckia Kendall,Victoria Caban Figueroa,Shatavia Morrison,Christopher Gulvick,Erisa Sula,Matthew Tugwell,Lauren Moon, |

| | | | | | |
|---|---|---|---|---|---|
| hCoV-19/USA/GA-CDC-MMB12617753/2021 | EPI_ISL_9331479 | 2021-12-30 | Mako Medical | Centers for Disease Control and Prevention Division of Viral Diseases, Pathogen Discovery | Dakota Howard,Dhwani Batra,Peter Cook,Jason Caravas,Benjamin Rambo-Martin,Scott Sammons,Yvette Unoarumhi,Matthew Schmerer,Kristine Lacek,Tymeckia Kendall,Victoria Caban Figueroa,Shatavia Morrison,Christopher Gulvick,Erisa Sula,Matthew Tugwell,Lauren Moon, |
| hCoV-19/Colombia/BOL-UNIMOL-VG-302/2021 | EPI_ISL_9338478 | 2021-12-26 | SERVCIOS INTREGALES DE SALUD | UNIMOL | Steev Loyola, Mashiel Fern√°ndez-Ruiz, Jaison Torres-Pacheco, Doris G√≥mez-Camargo |
| hCoV-19/Brazil/MG-FIOCRUZ-645/2021 | EPI_ISL_9340194 | 2021-12-23 | Laboratorio Municipal de Biologia Molecular | Instituto Rene Rachou / Fiocruz Minas | Thais Silva, Eneida Oliveira, Anna Salim, Andre Menezes, Rubens do Monte Neto, Gabriel Fernandes, Pedro Alves |
| hCoV-19/Colombia/PUT-INS-VG-10637/2021 | EPI_ISL_9340349 | 2021-11-16 | HOSPITAL UNIVERSITARIO DEPARTAMENTAL DE NARI√É‚ÄòO | Instituto Nacional de Salud-Direcci√≥n de Investigaci√≥n en Salud P√∫blica | Katherine Laiton-Donato, Carlos Franco-Mu√±oz, Diego A. √Ålvarez-D√≠az, H√©ctor Alejandro Ruiz-Moreno, Diego Andr√©s Prada, Jhonnatan Reales-Gonz√°lez, Sheryll Corchuelo, Mar√≠a T. Herrera-Sep√∫lveda, Juli√°n Naizaque, Gerardo Santamar√≠a, Jorge Rivera, P |
| hCoV-19/CzechRepublic/CSQ5299/2021 | EPI_ISL_9342977 | 2021-07-08 | Nemocnice Havlickuv Brod | University Hospital Brno, CMBG | Bezdicek Matej, Svaton Jan, Kristyna Dufkova, Lengerova Martina, Dolejska Monika |
| hCoV-19/CzechRepublic/CSQ5424/2021 | EPI_ISL_9343075 | 2021-01-23 | OKMI | University Hospital Brno, CMBG | Bezdicek Matej, Svaton Jan, Kristyna Dufkova, Lengerova Martina, Dolejska Monika |
| hCoV-19/USA/LA-EVTL11440/2021 | EPI_ISL_9347463 | 2021-12-18 | Willis-Knighton Medical Center Hospital Laboratory | LSUHS Emerging Viral Threat Laboratory | Lorie M. Atkins, April N. Johnson, Gregory L. Ware, Alexander Mijalis, Jeremy P. Kamil, Jennifer L. Carroll, Maarten Van Diest, Rona S. Scott, Andrew D. Yurochko, Christopher G. Kevil, John A. Vanchiere, Krista Queen, Joseph A. Bocchini |
| hCoV-19/USA/ND-NDDH-10407/2021 | EPI_ISL_9349208 | 2021-08-23 | North Dakota Department of Health, Public Health Laboratory | North Dakota Department of Health, Public Health Laboratory | Lisa Wingerter |
| hCoV-19/USA/TN-VUMC-002940/2021 | EPI_ISL_9357084 | 2021-07-26 | VUMC Molecular Infectious Diseases Laboratory (MIDL) | Dr. Suman Das Lab - Vanderbilt University Medical Center (VUMC) | Grant Vestal, Hunter Brown, Meghan Shilts, Bookyung Park, Helen Boone, Suman B. Pakala, Seesandra Rajagopala, Jonathan E. Schmitz, Suman Das |
| hCoV-19/India/TN-CBR-SEQ-7746/2021 | EPI_ISL_9361491 | 2021-10-18 | CBR,IISc | CBR,IISc-INSACOG | Bratati Kahali |
| hCoV-19/Colombia/DC-LSPSDS-46004/2021 | EPI_ISL_9362076 | 2021-12-27 | Ayudas Diagnosticas Sura | LSPSDS | Johana Hernandez Toloza, Gabriela Delgado Murcia, Alejandro Gomez Lopez, H√©ctor Alejandro Ruiz-Moreno, Marcela Mercado |
| hCoV-19/India/TN-CBR-SEQ-10499/2021 | EPI_ISL_9366426 | 2021-01-26 | CBR,IISc | CBR,IISc | Bratati Kahali |
| hCoV-19/India/TN-CBR-SEQ-10516/2021 | EPI_ISL_9366438 | 2021-11-13 | CBR,IISc | CBR,IISc | Bratati Kahali |
| hCoV-19/India/TN-CBR-SEQ-10604/2021 | EPI_ISL_9366509 | 2021-10-16 | CBR,IISc | CBR,IISc | Bratati Kahali |
| hCoV-19/Reunion/PIMIT_20028/2021 | EPI_ISL_9370420 | 2021-08-26 | CERBALLIANCE | UMR PIMIT | Dr David A Wilkinson, Dr Patrick Mavingui, Dr Camille Lebarbenchon, Magali Turpin, Marie-Alice Simbi, Dr Jonathan Turpin |

| | | | | | |
|---|---|---|---|---|---|
| hCoV-19/Reunion/PIMIT_23665/2021 | EPI_ISL_9370444 | 2021-12-03 | REUNILAB | UMR PIMIT | Dr David A Wilkinson, Dr Patrick Mavingui, Dr Camille Lebarbenchon, Magali Turpin, Marie-Alice Simbi, Dr Jonathan Turpin |
| hCoV-19/Reunion/PIMIT_18329/2021 | EPI_ISL_9370518 | 2021-08-07 | CERBALLIANCE | UMR PIMIT | Dr David A Wilkinson, Dr Patrick Mavingui, Dr Camille Lebarbenchon, Magali Turpin, Marie-Alice Simbi, Dr Jonathan Turpin |
| hCoV-19/Reunion/PIMIT_20090/2021 | EPI_ISL_9370617 | 2021-08-25 | REUNILAB | UMR PIMIT | Dr David A Wilkinson, Dr Patrick Mavingui, Dr Camille Lebarbenchon, Magali Turpin, Marie-Alice Simbi, Dr Jonathan Turpin |
| hCoV-19/Reunion/PIMIT_16978/2021 | EPI_ISL_9370859 | 2021-07-22 | REUNILAB | UMR PIMIT | Dr David A Wilkinson, Dr Patrick Mavingui, Dr Camille Lebarbenchon, Magali Turpin, Marie-Alice Simbi, Dr Jonathan Turpin |
| hCoV-19/Reunion/PIMIT_22672/2021 | EPI_ISL_9371137 | 2021-11-10 | CERBALLIANCE | UMR PIMIT | Dr David A Wilkinson, Dr Patrick Mavingui, Dr Camille Lebarbenchon, Magali Turpin, Marie-Alice Simbi, Dr Jonathan Turpin |
| hCoV-19/Reunion/PIMIT_24481/2021 | EPI_ISL_9371190 | 2021-12-13 | REUNILAB | UMR PIMIT | Dr David A Wilkinson, Dr Patrick Mavingui, Dr Camille Lebarbenchon, Magali Turpin, Marie-Alice Simbi, Dr Jonathan Turpin |
| hCoV-19/Reunion/PIMIT_23746/2021 | EPI_ISL_9371257 | 2021-12-06 | CERBALLIANCE | UMR PIMIT | Dr David A Wilkinson, Dr Patrick Mavingui, Dr Camille Lebarbenchon, Magali Turpin, Marie-Alice Simbi, Dr Jonathan Turpin |
| hCoV-19/Reunion/PIMIT_22526/2021 | EPI_ISL_9371580 | 2021-11-12 | REUNILAB | UMR PIMIT | Dr David A Wilkinson, Dr Patrick Mavingui, Dr Camille Lebarbenchon, Magali Turpin, Marie-Alice Simbi, Dr Jonathan Turpin |
| hCoV-19/Reunion/PIMIT_25274/2021 | EPI_ISL_9371663 | 2021-12-27 | REUNILAB | UMR PIMIT | Dr David A Wilkinson, Dr Patrick Mavingui, Dr Camille Lebarbenchon, Magali Turpin, Marie-Alice Simbi, Dr Jonathan Turpin |
| hCoV-19/Reunion/PIMIT_21022/2021 | EPI_ISL_9371761 | 2021-09-10 | REUNILAB | UMR PIMIT | Dr David A Wilkinson, Dr Patrick Mavingui, Dr Camille Lebarbenchon, Magali Turpin, Marie-Alice Simbi, Dr Jonathan Turpin |
| hCoV-19/Reunion/PIMIT_20011/2021 | EPI_ISL_9371788 | 2021-08-26 | CERBALLIANCE | UMR PIMIT | Dr David A Wilkinson, Dr Patrick Mavingui, Dr Camille Lebarbenchon, Magali Turpin, Marie-Alice Simbi, Dr Jonathan Turpin |
| hCoV-19/Reunion/PIMIT_23745/2021 | EPI_ISL_9371836 | 2021-12-06 | CERBALLIANCE | UMR PIMIT | Dr David A Wilkinson, Dr Patrick Mavingui, Dr Camille Lebarbenchon, Magali Turpin, Marie-Alice Simbi, Dr Jonathan Turpin |
| hCoV-19/Reunion/PIMIT_19724/2021 | EPI_ISL_9371853 | 2021-08-19 | REUNILAB | UMR PIMIT | Dr David A Wilkinson, Dr Patrick Mavingui, Dr Camille Lebarbenchon, Magali Turpin, Marie-Alice Simbi, Dr Jonathan Turpin |
| hCoV-19/Reunion/PIMIT_25188/2021 | EPI_ISL_9371888 | 2021-12-27 | REUNILAB | UMR PIMIT | Dr David A Wilkinson, Dr Patrick Mavingui, Dr Camille Lebarbenchon, Magali Turpin, Marie-Alice Simbi, Dr Jonathan Turpin |
| hCoV-19/Reunion/PIMIT_26515/2021 | EPI_ISL_9371889 | 2021-12-27 | REUNILAB | UMR PIMIT | Dr David A Wilkinson, Dr Patrick Mavingui, Dr Camille Lebarbenchon, Magali Turpin, Marie-Alice Simbi, Dr Jonathan Turpin |

| | | | | | |
|---|---|---|---|---|---|
| hCoV-19/Reunion/PIMIT_25235/2021 | EPI_ISL_9372005 | 2021-12-27 | REUNILAB | UMR PIMIT | Dr David A Wilkinson, Dr Patrick Mavingui, Dr Camille Lebarbenchon, Magali Turpin, Marie-Alice Simbi, Dr Jonathan Turpin |
| hCoV-19/Reunion/PIMIT_23585/2021 | EPI_ISL_9372132 | 2021-11-30 | REUNILAB | UMR PIMIT | Dr David A Wilkinson, Dr Patrick Mavingui, Dr Camille Lebarbenchon, Magali Turpin, Marie-Alice Simbi, Dr Jonathan Turpin |
| hCoV-19/Reunion/PIMIT_26719/2021 | EPI_ISL_9372178 | 2021-12-27 | REUNILAB | UMR PIMIT | Dr David A Wilkinson, Dr Patrick Mavingui, Dr Camille Lebarbenchon, Magali Turpin, Marie-Alice Simbi, Dr Jonathan Turpin |
| hCoV-19/Reunion/PIMIT_21989/2021 | EPI_ISL_9372294 | 2021-10-25 | REUNILAB | UMR PIMIT | Dr David A Wilkinson, Dr Patrick Mavingui, Dr Camille Lebarbenchon, Magali Turpin, Marie-Alice Simbi, Dr Jonathan Turpin |
| hCoV-19/Reunion/PIMIT_21968/2021 | EPI_ISL_9372489 | 2021-10-30 | CERBALLIANCE | UMR PIMIT | Dr David A Wilkinson, Dr Patrick Mavingui, Dr Camille Lebarbenchon, Magali Turpin, Marie-Alice Simbi, Dr Jonathan Turpin |
| hCoV-19/Reunion/PIMIT_22159/2021 | EPI_ISL_9372645 | 2021-11-03 | REUNILAB | UMR PIMIT | Dr David A Wilkinson, Dr Patrick Mavingui, Dr Camille Lebarbenchon, Magali Turpin, Marie-Alice Simbi, Dr Jonathan Turpin |
| hCoV-19/Reunion/PIMIT_20879/2021 | EPI_ISL_9372657 | 2021-09-06 | REUNILAB | UMR PIMIT | Dr David A Wilkinson, Dr Patrick Mavingui, Dr Camille Lebarbenchon, Magali Turpin, Marie-Alice Simbi, Dr Jonathan Turpin |
| hCoV-19/Reunion/PIMIT_21908/2021 | EPI_ISL_9372702 | 2021-10-29 | CERBALLIANCE | UMR PIMIT | Dr David A Wilkinson, Dr Patrick Mavingui, Dr Camille Lebarbenchon, Magali Turpin, Marie-Alice Simbi, Dr Jonathan Turpin |
| hCoV-19/Reunion/PIMIT_19184/2021 | EPI_ISL_9372718 | 2021-08-13 | REUNILAB | UMR PIMIT | Dr David A Wilkinson, Dr Patrick Mavingui, Dr Camille Lebarbenchon, Magali Turpin, Marie-Alice Simbi, Dr Jonathan Turpin |
| hCoV-19/Reunion/PIMIT_23654/2021 | EPI_ISL_9372769 | 2021-12-03 | REUNILAB | UMR PIMIT | Dr David A Wilkinson, Dr Patrick Mavingui, Dr Camille Lebarbenchon, Magali Turpin, Marie-Alice Simbi, Dr Jonathan Turpin |
| hCoV-19/Reunion/PIMIT_20395/2021 | EPI_ISL_9372843 | 2021-09-04 | CERBALLIANCE | UMR PIMIT | Dr David A Wilkinson, Dr Patrick Mavingui, Dr Camille Lebarbenchon, Magali Turpin, Marie-Alice Simbi, Dr Jonathan Turpin |
| hCoV-19/Reunion/PIMIT_21115/2021 | EPI_ISL_9373109 | 2021-09-13 | REUNILAB | UMR PIMIT | Dr David A Wilkinson, Dr Patrick Mavingui, Dr Camille Lebarbenchon, Magali Turpin, Marie-Alice Simbi, Dr Jonathan Turpin |
| hCoV-19/Reunion/PIMIT_20634/2021 | EPI_ISL_9373123 | 2021-09-03 | REUNILAB | UMR PIMIT | Dr David A Wilkinson, Dr Patrick Mavingui, Dr Camille Lebarbenchon, Magali Turpin, Marie-Alice Simbi, Dr Jonathan Turpin |
| hCoV-19/Reunion/PIMIT_26677/2021 | EPI_ISL_9373137 | 2021-12-27 | REUNILAB | UMR PIMIT | Dr David A Wilkinson, Dr Patrick Mavingui, Dr Camille Lebarbenchon, Magali Turpin, Marie-Alice Simbi, Dr Jonathan Turpin |
| hCoV-19/Reunion/PIMIT_20916/2021 | EPI_ISL_9373142 | 2021-09-08 | REUNILAB | UMR PIMIT | Dr David A Wilkinson, Dr Patrick Mavingui, Dr Camille Lebarbenchon, Magali Turpin, Marie-Alice Simbi, Dr Jonathan Turpin |

| | | | | | |
|---|---|---|---|---|---|
| hCoV-19/Reunion/PIMIT_26723/2021 | EPI_ISL_9373157 | 2021-12-27 | REUNILAB | UMR PIMIT | Dr David A Wilkinson, Dr Patrick Mavingui, Dr Camille Lebarbenchon, Magali Turpin, Marie-Alice Simbi, Dr Jonathan Turpin |
| hCoV-19/Reunion/PIMIT_21073/2021 | EPI_ISL_9373181 | 2021-09-17 | CERBALLIANCE | UMR PIMIT | Dr David A Wilkinson, Dr Patrick Mavingui, Dr Camille Lebarbenchon, Magali Turpin, Marie-Alice Simbi, Dr Jonathan Turpin |
| hCoV-19/Reunion/PIMIT_21636/2021 | EPI_ISL_9373185 | 2021-10-11 | CERBALLIANCE | UMR PIMIT | Dr David A Wilkinson, Dr Patrick Mavingui, Dr Camille Lebarbenchon, Magali Turpin, Marie-Alice Simbi, Dr Jonathan Turpin |
| hCoV-19/Reunion/PIMIT_20676/2021 | EPI_ISL_9373236 | 2021-09-01 | REUNILAB | UMR PIMIT | Dr David A Wilkinson, Dr Patrick Mavingui, Dr Camille Lebarbenchon, Magali Turpin, Marie-Alice Simbi, Dr Jonathan Turpin |
| hCoV-19/Reunion/PIMIT_19299/2021 | EPI_ISL_9373268 | 2021-08-14 | REUNILAB | UMR PIMIT | Dr David A Wilkinson, Dr Patrick Mavingui, Dr Camille Lebarbenchon, Magali Turpin, Marie-Alice Simbi, Dr Jonathan Turpin |
| hCoV-19/Reunion/PIMIT_26708/2021 | EPI_ISL_9373410 | 2021-12-27 | REUNILAB | UMR PIMIT | Dr David A Wilkinson, Dr Patrick Mavingui, Dr Camille Lebarbenchon, Magali Turpin, Marie-Alice Simbi, Dr Jonathan Turpin |
| hCoV-19/Reunion/PIMIT_21528/2021 | EPI_ISL_9373445 | 2021-10-04 | REUNILAB | UMR PIMIT | Dr David A Wilkinson, Dr Patrick Mavingui, Dr Camille Lebarbenchon, Magali Turpin, Marie-Alice Simbi, Dr Jonathan Turpin |
| hCoV-19/Reunion/PIMIT_21120/2021 | EPI_ISL_9373456 | 2021-09-15 | REUNILAB | UMR PIMIT | Dr David A Wilkinson, Dr Patrick Mavingui, Dr Camille Lebarbenchon, Magali Turpin, Marie-Alice Simbi, Dr Jonathan Turpin |
| hCoV-19/Reunion/PIMIT_20648/2021 | EPI_ISL_9373602 | 2021-09-04 | REUNILAB | UMR PIMIT | Dr David A Wilkinson, Dr Patrick Mavingui, Dr Camille Lebarbenchon, Magali Turpin, Marie-Alice Simbi, Dr Jonathan Turpin |
| hCoV-19/Reunion/PIMIT_21800/2021 | EPI_ISL_9373784 | 2021-10-20 | REUNILAB | UMR PIMIT | Dr David A Wilkinson, Dr Patrick Mavingui, Dr Camille Lebarbenchon, Magali Turpin, Marie-Alice Simbi, Dr Jonathan Turpin |
| hCoV-19/Venezuela/Tru460/2021 | EPI_ISL_9377166 | 2021-11-06 | IVIC | Laboratorio de Virologia Molecular | Rossana C Jaspe, Carmen L Loureiro, Yoneira Sulbaran, Zoila Moros, Pierina D¬¥Angelo, Lieska Rodr√≠guez, Jos√© Luis Zambrano, Mariana Hidalgo, V√≠ctor Alarc√≥n, Domingo J Garzaro, CoViMol Group, H√©ctor R Rangel, Flor H Pujol |
| hCoV-19/India/MZ-INSACOG-1526100705275/2021 | EPI_ISL_9389516 | 2021-10-02 | INSACOG-Mizoram | National Institute of Biomedical Genomics ‚Äì INSACOG | Arindam Maitra, Swagnik Roy, Nidhan Kumar Biswas, Gracy Laldinmawii, Sreedhar Chinnaswamy, N Senthil Kumar, Saumitra Das |
| hCoV-19/India/MZ-INSACOG-1526100706208/2021 | EPI_ISL_9389572 | 2021-10-02 | INSACOG-Mizoram | National Institute of Biomedical Genomics ‚Äì INSACOG | Arindam Maitra, Swagnik Roy, Nidhan Kumar Biswas, Gracy Laldinmawii, Sreedhar Chinnaswamy, N Senthil Kumar, Saumitra Das |
| hCoV-19/India/MZ-INSACOG-1526100744226/2021 | EPI_ISL_9389648 | 2021-10-12 | INSACOG-Mizoram | National Institute of Biomedical Genomics ‚Äì INSACOG | Arindam Maitra, Swagnik Roy, Nidhan Kumar Biswas, Gracy Laldinmawii, Sreedhar Chinnaswamy, N Senthil Kumar, Saumitra Das |

| | | | | | |
|---|---|---|---|---|---|
| hCoV-19/India/MZ-INSACOG-1526100706987/2021 | EPI_ISL_9389919 | 2021-10-02 | INSACOG-Mizoram | National Institute of Biomedical Genomics ‚Äì INSACOG | Arindam Maitra, Swagnik Roy, Nidhan Kumar Biswas, Gracy Laldinmawii, Sreedhar Chinnaswamy, N Senthil Kumar, Saumitra Das |
| hCoV-19/India/WB-INSACOG-1931300521737/2021 | EPI_ISL_9389928 | 2021-11-24 | INSACOG-WB | National Institute of Biomedical Genomics ‚Äì INSACOG | Arindam Maitra, Bhaswati Bandyopadhyay, Nidhan Kumar Biswas, Tamal Ghosh, Sreedhar Chinnaswamy, Ajay Chakraborti, Saumitra Das |
| hCoV-19/India/MZ-INSACOG-1526100744322/2021 | EPI_ISL_9389996 | 2021-10-12 | INSACOG-Mizoram | National Institute of Biomedical Genomics ‚Äì INSACOG | Arindam Maitra, Swagnik Roy, Nidhan Kumar Biswas, Gracy Laldinmawii, Sreedhar Chinnaswamy, N Senthil Kumar, Saumitra Das |
| hCoV-19/India/MZ-INSACOG-1526100708084/2021 | EPI_ISL_9390043 | 2021-10-03 | INSACOG-Mizoram | National Institute of Biomedical Genomics ‚Äì INSACOG | Arindam Maitra, Swagnik Roy, Nidhan Kumar Biswas, Gracy Laldinmawii, Sreedhar Chinnaswamy, N Senthil Kumar, Saumitra Das |
| hCoV-19/USA/CO-CDPHE-2102608411/2021 | EPI_ISL_9391357 | 2021-12-28 | Colorado Department of Public Health and Environment | Colorado Department of Public Health and Environment | Laura Bankers, Molly C. Hetherington-Rauth, Diana Ir, Alexandria Rossheim, Michael Martin, Mandy Waters, Arianna Smith, Shannon R. Matzinger, Emily A. Travanty |
| hCoV-19/USA/CA-CDPH-2000057531/2021 | EPI_ISL_9393323 | 2021-12-21 | CDPH VBL | California Department of Public Health | CDPH-COVIDNet and UCLA Technology Center for Genomics & Bioinformatics |
| hCoV-19/Thailand/64130096/2021 | EPI_ISL_9396148 | 2021-09-12 | Molecular HIV and EID, BIDI, DDC,Thai MOPH | Molecular HIV and EID, BIDI, DDC,Thai MOPH | Sumonmal Uttayamakul |
| hCoV-19/Australia/NSW-RPAH-2427/2021 | EPI_ISL_9396384 | 2021-01-01 | New South Wales Health Pathology Royal Prince Alfred Hospital | Microbiology RPAH | Foster, C.; Au, J.; Ruiz Silva, M.; Deveson, I.; Bull, R.; Van Hal, S.; Rawlinson, W. |
| hCoV-19/Australia/NSW-RPAH-2428/2021 | EPI_ISL_9396385 | 2021-01-02 | New South Wales Health Pathology Royal Prince Alfred Hospital | Microbiology RPAH | Foster, C.; Au, J.; Ruiz Silva, M.; Deveson, I.; Bull, R.; Van Hal, S.; Rawlinson, W. |
| hCoV-19/USA/AZ-TG1170123/2021 | EPI_ISL_9397113 | 2021-10-04 | TGen North | TGen North | Jolene Bowers, Heather Centner, Brett Van Tassel, Chris French, Hayley Yaglom, Darrin Lemmer, Dave Engelthaler |
| hCoV-19/SouthAfrica/NICD-N28160/2021 | EPI_ISL_9401660 | 2021-12-30 | VERMAAK | National Institute for Communicable Diseases of the National Health Laboratory Service | Amoako DG, Everatt J, Kekana D, Scheepers C, Mohale T, Ntuli N, Mahlangu B, Mnguni A, Ismail A, Bhiman JN, Wolter N |
| hCoV-19/USA/KY-GD_01669/2021 | EPI_ISL_9405167 | 2021-12-09 | Gravity Diagnostics, LLC | University of Louisville Sequencing Technology Center | Elizabeth Hudson, William Lauer, James P. Canner, Ryan Walker, Julia H. Chariker, Aryan Neupane, Eric C. Rouchka, Melissa L. Smith |
| hCoV-19/India/KA-SEQ_9489/2021 | EPI_ISL_9409007 | 2021-12-21 | BBMP- West | inStem, NCBS-INSACOG | Uma Ramakrishnan, Dasaradhi Palakodeti, Aswin SaiNarain |
| hCoV-19/India/KA-SEQ_9411/2021 | EPI_ISL_9409074 | 2021-12-30 | Bangalore Airport | inStem, NCBS-INSACOG | Uma Ramakrishnan, Dasaradhi Palakodeti, Aswin SaiNarain |
| hCoV-19/USA/TN-VUMC-O-0308/2021 | EPI_ISL_9409946 | 2021-11-29 | VUMC Molecular Infectious Diseases Laboratory (MIDL) | Dr. Suman Das Lab - Vanderbilt University Medical Center (VUMC) | Grant Vestal, Hunter Brown, Meghan Shilts, Bookyung Park, Helen Boone, Suman B. Pakala, Seesandra Rajagopala, Jonathan E. Schmitz, Suman Das |
| hCoV-19/DominicanRepublic/1420572/2021 | EPI_ISL_9413613 | 2021-01-11 | Laboratorio Nacional de Salud Pv∫blica Dr. Defillo (LNSPDD) | Genomics and Proteomics Departament, Gorgas Memorial Institute For Health Studies | Isaac Miguel , Yvonne Imbert, Lucia de la Cruz, Ronald Skewes, Grey Benoit, Leyda Abrego, Marlene Castillo, Melissa Gaitan ,Oris Chavarria, Jessica Gondola, Ambar Moreno, Claudia Gonzalez V, Alexander A Martinez |
| hCoV-19/DominicanRepublic/1423368/2021 | EPI_ISL_9413614 | 2021-02-11 | Laboratorio Nacional de Salud Pv∫blica Dr. Defillo (LNSPDD) | Genomics and Proteomics Departament, Gorgas Memorial Institute For Health Studies | Isaac Miguel , Yvonne Imbert, Lucia de la Cruz, Ronald Skewes, Grey Benoit, Leyda Abrego, Marlene Castillo, Melissa Gaitan ,Oris Chavarria, Jessica Gondola, Ambar Moreno, Claudia Gonzalez V, Alexander A Martinez |

| | | | | | |
|---|---|---|---|---|---|
| hCoV-19/DominicanRepublic/1437151/2021 | EPI_ISL_9413618 | 2021-02-11 | Laboratorio Nacional de Salud PV∫blica Dr. Defillo (LNSPDD) | Genomics and Proteomics Departament, Gorgas Memorial Institute For Health Studies | Isaac Miguel , Yvonne Imbert, Lucia de la Cruz, Ronald Skewes, Grey Benoit, Leyda Abrego, Marlene Castillo, Melissa Gaitan ,Oris Chavarria, Jessica Gondola, Ambar Moreno, Claudia Gonzalez V, Alexander A Martinez |
| hCoV-19/DominicanRepublic/1455157/2021 | EPI_ISL_9413623 | 2021-03-11 | Laboratorio Nacional de Salud PV∫blica Dr. Defillo (LNSPDD) | Genomics and Proteomics Departament, Gorgas Memorial Institute For Health Studies | Isaac Miguel , Yvonne Imbert, Lucia de la Cruz, Ronald Skewes, Grey Benoit, Leyda Abrego, Marlene Castillo, Melissa Gaitan ,Oris Chavarria, Jessica Gondola, Ambar Moreno, Claudia Gonzalez V, Alexander A Martinez |
| hCoV-19/DominicanRepublic/1423948/2021 | EPI_ISL_9413637 | 2021-01-11 | Laboratorio Nacional de Salud PV∫blica Dr. Defillo (LNSPDD) | Genomics and Proteomics Departament, Gorgas Memorial Institute For Health Studies | Isaac Miguel , Yvonne Imbert, Lucia de la Cruz, Ronald Skewes, Grey Benoit, Leyda Abrego, Marlene Castillo, Melissa Gaitan ,Oris Chavarria, Jessica Gondola, Ambar Moreno, Claudia Gonzalez V, Alexander A Martinez |
| hCoV-19/DominicanRepublic/1450964/2021 | EPI_ISL_9413653 | 2021-02-11 | Laboratorio Nacional de Salud PV∫blica Dr. Defillo (LNSPDD) | Genomics and Proteomics Departament, Gorgas Memorial Institute For Health Studies | Isaac Miguel , Yvonne Imbert, Lucia de la Cruz, Ronald Skewes, Grey Benoit, Leyda Abrego, Marlene Castillo, Melissa Gaitan ,Oris Chavarria, Jessica Gondola, Ambar Moreno, Claudia Gonzalez V, Alexander A Martinez |
| hCoV-19/DominicanRepublic/1423990/2021 | EPI_ISL_9413669 | 2021-01-11 | Laboratorio Nacional de Salud PV∫blica Dr. Defillo (LNSPDD) | Genomics and Proteomics Departament, Gorgas Memorial Institute For Health Studies | Isaac Miguel , Yvonne Imbert, Lucia de la Cruz, Ronald Skewes, Grey Benoit, Leyda Abrego, Marlene Castillo, Melissa Gaitan ,Oris Chavarria, Jessica Gondola, Ambar Moreno, Claudia Gonzalez V, Alexander A Martinez |
| hCoV-19/SouthAfrica/NCV476/2021 | EPI_ISL_9413697 | 2021-11-29 | National Health Laboratory Services, Virology | National Health Laboratory Services, Virology | Kathleen Subramoney, Nkhensani Mtileni, Avani Bharuthram, Florette K. Treurnicht |
| hCoV-19/SouthAfrica/NCV494/2021 | EPI_ISL_9413706 | 2021-11-04 | National Health Laboratory Services, Virology | National Health Laboratory Services, Virology | Kathleen Subramoney, Nkhensani Mtileni, Avani Bharuthram, Florette K. Treurnicht |
| hCoV-19/Mozambique/CERI-KRISP-K033862/2021 | EPI_ISL_9418715 | 2021-12-07 | Instituto Nacional de Saude (INS), Mozambique | and Innovation, Stellenbosch University and KRISP, KZN Research Innovation and Sequencing Platform, UKZN. | Nalia Ismael, Nadia Sitoe, Paulo Arnaldo, Nedio Mabunda, Giandhari J, Pillay S, Naidoo Y, Naicker D, Maharaj A, Anyaneji UJ, Moir M, Van Wyk S, San JE, Tshiabuila D, Tegally H, Wilkinson E, de Oliveira T |
| hCoV-19/India/MH-ICMR-NIV-INSACOG-GSEQ-8096/2021 | EPI_ISL_9419053 | 2021-12-30 | ICMR-National Institute of Virology - INSACOG | NIV Influenza | Dr. Varsha Potdar and NIC Team |
| hCoV-19/SouthAfrica/CERI-KRISP-K034206/2021 | EPI_ISL_9423050 | 2021-12-23 | ZARV/NHLS,¬†Department Medical Virology,¬†University of Pretoria | CERI, Centre for Epidemic Response and Innvoation, Stellenbosch University and KRISP, KZN Research Innovation and Sequencing Platform, UKZN. | Amy Strydom, Adriano Mendes, Micheala Davids, Sim Mayaphi and Marietjie Venter, Giandhari J, Pillay S, Naidoo Y, Naicker D, Maharaj A, Anyaneji UJ, Moir M, Van Wyk S, San JE, Tshiabuila D, Tegally H, Wilkinson E, de Oliveira T |
| hCoV-19/SouthAfrica/CERI-KRISP-K034241/2021 | EPI_ISL_9423052 | 2021-12-15 | ZARV/NHLS,¬†Department Medical Virology,¬†University of Pretoria | CERI, Centre for Epidemic Response and Innvoation, Stellenbosch University and KRISP, KZN Research Innovation and Sequencing Platform, UKZN. | Amy Strydom, Adriano Mendes, Micheala Davids, Sim Mayaphi and Marietjie Venter, Giandhari J, Pillay S, Naidoo Y, Naicker D, Maharaj A, Anyaneji UJ, Moir M, Van Wyk S, San JE, Tshiabuila D, Tegally H, Wilkinson E, de Oliveira T |
| hCoV-19/SouthAfrica/CERI-KRISP-K034258/2021 | EPI_ISL_9423053 | 2021-12-24 | National Health Laboratory Services, Virology | and Innvoation, Stellenbosch University and KRISP, KZN Research Innovation and Sequencing Platform, UKZN. | Kathleen Subramoney, Florette Treurnicht, Nkhensani Mtileni, Avani Bharuthram, Giandhari J, Pillay S, Naidoo Y, Naicker D, Maharaj A, Anyaneji UJ, Moir M, Van Wyk S, San JE, Tshiabuila D, Tegally H, Wilkinson E, de Oliveira T |

| | | | | | |
|---|---|---|---|---|---|
| hCoV-19/SouthAfrica/CERI-KRISP-K034272/2021 | EPI_ISL_9423085 | 2021-12-30 | Lancet Laboratoy | and Innvoation, Stellenbosch University and KRISP, KZN Research Innovation and Sequencing Platform, UKZN. | Giandhari J, Pillay S, Naidoo Y, Naicker D, Maharaj A, Anyaneji UJ, Moir M, Van Wyk S, San JE, Tshiabuila D, Tegally H, Wilkinson E, de Oliveira T |
| hCoV-19/SouthAfrica/CERI-KRISP-K034215/2021 | EPI_ISL_9423096 | 2021-12-21 | ZARV/NHLS,¬†Department Medical Virology,¬†University of Pretoria | CERI, Centre for Epidemic Response and Innvoation, Stellenbosch University and KRISP, KZN Research Innovation and Sequencing Platform, UKZN. | Amy Strydom, Adriano Mendes, Micheala Davids, Sim Mayaphi and Marietjie Venter, Giandhari J, Pillay S, Naidoo Y, Naicker D, Maharaj A, Anyaneji UJ, Moir M, Van Wyk S, San JE, Tshiabuila D, Tegally H, Wilkinson E, de Oliveira T |
| hCoV-19/Botswana/CERI-KRISP-K027669/2021 | EPI_ISL_9423143 | 2021-10-15 | National Health Laboratory | CERI, Centre for Epidemic Response and Innvoation, Stellenbosch University and KRISP, KZN Research Innovation and Sequencing Platform, UKZN. | Thongbotho Mphoyakgosi, Sefetogi Ramaologa, Keoratile Ntshambiwa, Thela Tefelo, Mphaphi B. Mbulawa,Pamela Smith-Lawrence, Dorcas Maruapula, Sikhulile Moyo, Wonderful T. Choga, Joseph Makhema, Simani Gaseitsiwe , Giandhari J, Pillay S, Naidoo Y, Naicker D |
| hCoV-19/France/CVL-HMN-22012180757/2021 | EPI_ISL_9428287 | 2021-12-27 | BPR Analyses sp√©cialis√©es | Mondor University Hospital, Assistance Publique H√¥pitaux de Paris, Universit√© Paris-Est Cr√©teil, INSERM U955 | Christophe Rodriguez, Slim Fourati, Vanessa Demontant, Guillaume Gricourt, Melissa N'Debi, Alexandre Soulier, Elisabeth Trawinski, Jean-Michel Pawlotsky |
| hCoV-19/France/IDF-IPP03138/2021 | EPI_ISL_9429418 | 2021-01-09 | Hopital | National Reference Center for Viruses of Respiratory Infections, Institut Pasteur, Paris | Marion Barbet, Sylvie Behillil, M√©line Bizard, Angela Brisebarre, Camille Capel, Vincent Enouf, Louise Lefran√ßois, Fr√©d√©ric Lemoine, Christophe Malabat, Corinne Maufrais, Slim El Khiari, Julien Fumey, Etienne Simon-Lori√®re, Maud Vanpeene, Sylvie Van |
| hCoV-19/India/KA-CBR-SEQ-10874/2021 | EPI_ISL_9430079 | 2021-12-29 | CBR,IISc | CBR,IISc | Bratati Kahali |
| hCoV-19/Canada/BC-BCCDC-318850/2021 | EPI_ISL_9434118 | 2021-12-27 | British Columbia Centre For Disease Control | BCCDC Public Health Laboratory | Prystajecky Natalie, Linda Hoang, Dan Fornika, John Tyson, Shannon Russell, Kim Macdonald, Kimia Kamelian, James Zlosnik, Ana Pacagnella, Corrinne Ng, Loretta Janz, Robert Azana, Mel Krajden |
| hCoV-19/Paraguay/306491/2021 | EPI_ISL_9438230 | 2021-12-06 | Laboratorio Central de Salud P√∫blica | Laboratorio Central de Salud P√∫blica | Sandra Gonzalez, Andrea G√≥mez de la Fuente, Cynthia Vazquez, Julio Barrios, Cesar Cantero, Tania Alfonzo, Shirley Villalba, Juan Torales, Maria Liz Gamarra |
| hCoV-19/Eswatini/N21907/2021 | EPI_ISL_9439133 | 2021-10-09 | Raleigh Fitkin Memorial | National Institute for Communicable Diseases of the National Health Laboratory Service | Maphalala G, Amoako DG, Everatt J, Kekana D, Scheepers C, Mohale T, Ntuli N, Mahlangu B, Mnguni A, Ismail A, Bhiman JN, Wolter N |
| hCoV-19/Eswatini/N22763/2021 | EPI_ISL_9439186 | 2021-11-28 | Good Shepard Hospital | National Institute for Communicable Diseases of the National Health Laboratory Service | Maphalala G, Amoako DG, Everatt J, Kekana D, Scheepers C, Mohale T, Ntuli N, Mahlangu B, Mnguni A, Ismail A, Bhiman JN, Wolter N |
| hCoV-19/Eswatini/N22764/2021 | EPI_ISL_9439187 | 2021-11-28 | Good Shepard Hospital | National Institute for Communicable Diseases of the National Health Laboratory Service | Maphalala G, Amoako DG, Everatt J, Kekana D, Scheepers C, Mohale T, Ntuli N, Mahlangu B, Mnguni A, Ismail A, Bhiman JN, Wolter N |
| hCoV-19/Eswatini/N22775/2021 | EPI_ISL_9439198 | 2021-11-27 | Good Shepard Hospital | National Institute for Communicable Diseases of the National Health Laboratory Service | Maphalala G, Amoako DG, Everatt J, Kekana D, Scheepers C, Mohale T, Ntuli N, Mahlangu B, Mnguni A, Ismail A, Bhiman JN, Wolter N |

| | | | | | |
|---|---|---|---|---|---|
| hCoV-19/Eswatini/N22777/2021 | EPI_ISL_9439200 | 2021-11-23 | Good Shepard Hospital | National Institute for Communicable Diseases of the National Health Laboratory Service | Maphalala G, Amoako DG, Everatt J, Kekana D, Scheepers C, Mohale T, Ntuli N, Mahlangu B, Mnguni A, Ismail A, Bhiman JN, Wolter N |
| hCoV-19/Eswatini/N22786/2021 | EPI_ISL_9439209 | 2021-11-28 | Good Shepard Hospital | National Institute for Communicable Diseases of the National Health Laboratory Service | Maphalala G, Amoako DG, Everatt J, Kekana D, Scheepers C, Mohale T, Ntuli N, Mahlangu B, Mnguni A, Ismail A, Bhiman JN, Wolter N |
| hCoV-19/Eswatini/N22789/2021 | EPI_ISL_9439212 | 2021-11-28 | Good Shepard Hospital | National Institute for Communicable Diseases of the National Health Laboratory Service | Maphalala G, Amoako DG, Everatt J, Kekana D, Scheepers C, Mohale T, Ntuli N, Mahlangu B, Mnguni A, Ismail A, Bhiman JN, Wolter N |
| hCoV-19/Eswatini/N22793/2021 | EPI_ISL_9439216 | 2021-11-29 | Hhohho RRT | National Institute for Communicable Diseases of the National Health Laboratory Service | Maphalala G, Amoako DG, Everatt J, Kekana D, Scheepers C, Mohale T, Ntuli N, Mahlangu B, Mnguni A, Ismail A, Bhiman JN, Wolter N |
| hCoV-19/Namibia/N25838/2021 | EPI_ISL_9454959 | 2021-12-06 | Namibia Instuition of Pathology | National Institute for Communicable Diseases of the National Health Laboratory Service | Konstantinus I, Amoako DG, Everatt J, Kekana D, Scheepers C, Mohale T, Ntuli N, Mahlangu B, Mnguni A, Ismail A, Bhiman JN, Wolter N |
| hCoV-19/Namibia/N25869/2021 | EPI_ISL_9454974 | 2021-12-07 | Namibia Instuition of Pathology | National Institute for Communicable Diseases of the National Health Laboratory Service | Konstantinus I, Amoako DG, Everatt J, Kekana D, Scheepers C, Mohale T, Ntuli N, Mahlangu B, Mnguni A, Ismail A, Bhiman JN, Wolter N |
| hCoV-19/India/PB-VRDLGMCPTA-ICMR-INSACOG-587666318/2021 | EPI_ISL_9457929 | 2021-11-29 | Virus Research and Diagnostic Laboratory | NIV Influenza | Dr.Rupinder Bakshi |
| hCoV-19/Norway/35184/2021 | EPI_ISL_9461045 | 2021-12-27 | Haugesund Hospital, laboratory for Medical Microbiology | Norwegian Institute of Public Health, Department of Virology | Kathrine Stene-Johansen, Kamilla Heddeland Instefjord, Hilde Elshaug, Garcia Llorente Ignacio, Jon BrV•te, Engebretsen Serina Beate, Pedersen Benedikte Nevjen, Line Victoria Moen, Hilde Nordby Falkenhaug, Debech Nadia, Atiya R Ali, Marie Paulsen Madsen, R |
| hCoV-19/India/WB-INSACOG-1931502627383/2021 | EPI_ISL_9466144 | 2021-12-28 | INSACOG-WB | National¬†Institute¬†of¬†Biomedical ¬†Genomics ‚Äì¬†INSACOG | Arindam Maitra, Bhaswati Bandyopadhyay, Nidhan Kumar Biswas, Tamal Ghosh, Sreedhar Chinnaswamy, Ajay Chakraborti, Saumitra Das |
| hCoV-19/India/WB-INSACOG-1931502628136/2021 | EPI_ISL_9466420 | 2021-12-28 | INSACOG-WB | National¬†Institute¬†of¬†Biomedical ¬†Genomics ‚Äì¬†INSACOG | Arindam Maitra, Bhaswati Bandyopadhyay, Nidhan Kumar Biswas, Tamal Ghosh, Sreedhar Chinnaswamy, Ajay Chakraborti, Saumitra Das |
| hCoV-19/India/WB-INSACOG-1931502640420/2021 | EPI_ISL_9466466 | 2021-12-30 | INSACOG-WB | National¬†Institute¬†of¬†Biomedical ¬†Genomics ‚Äì¬†INSACOG | Arindam Maitra, Bhaswati Bandyopadhyay, Nidhan Kumar Biswas, Tamal Ghosh, Sreedhar Chinnaswamy, Ajay Chakraborti, Saumitra Das |
| hCoV-19/India/WB-INSACOG-1931502631576/2021 | EPI_ISL_9466521 | 2021-12-29 | INSACOG-WB | National¬†Institute¬†of¬†Biomedical ¬†Genomics ‚Äì¬†INSACOG | Arindam Maitra, Bhaswati Bandyopadhyay, Nidhan Kumar Biswas, Tamal Ghosh, Sreedhar Chinnaswamy, Ajay Chakraborti, Saumitra Das |
| hCoV-19/India/WB-INSACOG-1931502634753/2021 | EPI_ISL_9466590 | 2021-12-29 | INSACOG-WB | National¬†Institute¬†of¬†Biomedical ¬†Genomics ‚Äì¬†INSACOG | Arindam Maitra, Bhaswati Bandyopadhyay, Nidhan Kumar Biswas, Tamal Ghosh, Sreedhar Chinnaswamy, Ajay Chakraborti, Saumitra Das |
| hCoV-19/India/WB-INSACOG-1931502636212/2021 | EPI_ISL_9466631 | 2021-12-29 | INSACOG-WB | National¬†Institute¬†of¬†Biomedical ¬†Genomics ‚Äì¬†INSACOG | Arindam Maitra, Bhaswati Bandyopadhyay, Nidhan Kumar Biswas, Tamal Ghosh, Sreedhar Chinnaswamy, Ajay Chakraborti, Saumitra Das |

| | | | | | |
|---|---|---|---|---|---|
| hCoV-19/India/WB-INSACOG-1931502630642/2021 | EPI_ISL_9466678 | 2021-12-29 | INSACOG-WB | National¬†Institute¬†of¬†Biomedical ¬†Genomics ,Äì¬†INSACOG | Arindam Maitra, Bhaswati Bandyopadhyay, Nidhan Kumar Biswas, Tamal Ghosh, Sreedhar Chinnaswamy, Ajay Chakraborti, Saumitra Das |
| hCoV-19/Paraguay/325191/2021 | EPI_ISL_9469690 | 2021-12-22 | Laboratorio Central de Salud P√∫blica | Laboratorio Central de Salud P√∫blica | Sandra Gonzalez, Andrea G√≥mez de la Fuente, Cynthia Vazquez, Julio Barrios,Tania Alfonzo, Shirley Villalba, Juan Torales, Maria Liz Gamarra |
| hCoV-19/Paraguay/326371/2021 | EPI_ISL_9469707 | 2021-12-22 | Laboratorio Central de Salud P√∫blica | Laboratorio Central de Salud P√∫blica | Sandra Gonzalez, Andrea G√≥mez de la Fuente, Cynthia Vazquez, Julio Barrios,Tania Alfonzo, Shirley Villalba, Juan Torales, Maria Liz Gamarra |
| hCoV-19/Paraguay/329277/2021 | EPI_ISL_9469720 | 2021-12-24 | Laboratorio Central de Salud P√∫blica | Laboratorio Central de Salud P√∫blica | Sandra Gonzalez, Andrea G√≥mez de la Fuente, Cynthia Vazquez, Julio Barrios,Tania Alfonzo, Shirley Villalba, Juan Torales, Maria Liz Gamarra |
| hCoV-19/India/TN-CDFD-CO-459/2021 | EPI_ISL_9472286 | 2021-12-29 | Directorate Of Public Health And Preventive Medicine | Centre for DNA Fingerprinting and Diagnostics | Asmita Gupta, Rupin Shelake, Arunkumar Karunanidhi, Vinay Donipadi, Divya Vashisht, Ashwin Dalal, Selvavinayagam Sivaprakasam, Darez Ahamed, Sampath Palani, Raju Sivadoss, Avudaiselvi Rathinasamy, Gurunathan Subramanian, Rajesh Kumar Manivannan, Devi Moni |
| hCoV-19/India/TN-CDFD-CO-470/2021 | EPI_ISL_9472294 | 2021-12-24 | Directorate Of Public Health And Preventive Medicine | Centre for DNA Fingerprinting and Diagnostics | Asmita Gupta, Rupin Shelake, Arunkumar Karunanidhi, Vinay Donipadi, Divya Vashisht, Ashwin Dalal, Selvavinayagam Sivaprakasam, Darez Ahamed, Sampath Palani, Raju Sivadoss, Avudaiselvi Rathinasamy, Gurunathan Subramanian, Rajesh Kumar Manivannan, Devi Moni |
| hCoV-19/India/TN-CDFD-CO-567/2021 | EPI_ISL_9472367 | 2021-12-27 | Directorate Of Public Health And Preventive Medicine | Centre for DNA Fingerprinting and Diagnostics | Asmita Gupta, Rupin Shelake, Arunkumar Karunanidhi, Vinay Donipadi, Divya Vashisht, Ashwin Dalal, Selvavinayagam Sivaprakasam, Darez Ahamed, Sampath Palani, Raju Sivadoss, Avudaiselvi Rathinasamy, Gurunathan Subramanian, Rajesh Kumar Manivannan, Devi Moni |
| hCoV-19/India/TN-CDFD-CO-719/2021 | EPI_ISL_9472464 | 2021-12-22 | Directorate Of Public Health And Preventive Medicine | Centre for DNA Fingerprinting and Diagnostics | Asmita Gupta, Rupin Shelake, Arunkumar Karunanidhi, Vinay Donipadi, Divya Vashisht, Ashwin Dalal, Selvavinayagam Sivaprakasam, Darez Ahamed, Sampath Palani, Raju Sivadoss, Avudaiselvi Rathinasamy, Gurunathan Subramanian, Rajesh Kumar Manivannan, Devi Moni |
| hCoV-19/India/TN-CDFD-CO-781/2021 | EPI_ISL_9472496 | 2021-12-29 | Directorate Of Public Health And Preventive Medicine | Centre for DNA Fingerprinting and Diagnostics | Asmita Gupta, Rupin Shelake, Arunkumar Karunanidhi, Vinay Donipadi, Divya Vashisht, Ashwin Dalal, Selvavinayagam Sivaprakasam, Darez Ahamed, Sampath Palani, Raju Sivadoss, Avudaiselvi Rathinasamy, Gurunathan Subramanian, Rajesh Kumar Manivannan, Devi Moni |
| hCoV-19/India/WB-INSACOG-1931502639242/2021 | EPI_ISL_9472738 | 2021-12-30 | INSACOG-WB | National¬†Institute¬†of¬†Biomedical ¬†Genomics ,Äì¬†INSACOG | Arindam Maitra, Bhaswati Bandyopadhyay, Nidhan Kumar Biswas, Tamal Ghosh, Sreedhar Chinnaswamy, Ajay Chakraborti, Saumitra Das |

| | | | | | |
|---|---|---|---|---|---|
| hCoV-19/India/WB-INSACOG-1931502624738/2021 | EPI_ISL_9472766 | 2021-12-28 | INSACOG-WB | National¬†Institute¬†of¬†Biomedical ¬†Genomics ‚Äì¬†INSACOG | Arindam Maitra, Bhaswati Bandyopadhyay, Nidhan Kumar Biswas, Tamal Ghosh, Sreedhar Chinnaswamy, Ajay Chakraborti, Saumitra Das |
| hCoV-19/India/WB-INSACOG-618283010/2021 | EPI_ISL_9472843 | 2021-12-28 | INSACOG-WB | National¬†Institute¬†of¬†Biomedical ¬†Genomics ‚Äì¬†INSACOG | Arindam Maitra, Bhaswati Bandyopadhyay, Nidhan Kumar Biswas, Tamal Ghosh, Sreedhar Chinnaswamy, Ajay Chakraborti, Saumitra Das |
| hCoV-19/India/WB-INSACOG-1931502625270/2021 | EPI_ISL_9472912 | 2021-12-28 | INSACOG-WB | National¬†Institute¬†of¬†Biomedical ¬†Genomics ‚Äì¬†INSACOG | Arindam Maitra, Bhaswati Bandyopadhyay, Nidhan Kumar Biswas, Tamal Ghosh, Sreedhar Chinnaswamy, Ajay Chakraborti, Saumitra Das |
| hCoV-19/France/HDF-IPP3837/2021 | EPI_ISL_9474890 | 2021-01-17 | Hopital | National Reference Center for Viruses of Respiratory Infections, Institut Pasteur, Paris | Marion Barbet, Sylvie Behillil, M√©line Bizard, Angela Brisebarre, Camille Capel, Vincent Enouf, Louise Lefran√ßois, Fr√©d√©ric Lemoine, Christophe Malabat, Corinne Maufrais, Slim El Khiari, Julien Fumey, Etienne Simon-Lori√®re, Maud Vanpeene, Sylvie Van |
| hCoV-19/France/HDF-IPP03845/2021 | EPI_ISL_9474898 | 2021-01-17 | Hopital | National Reference Center for Viruses of Respiratory Infections, Institut Pasteur, Paris | Marion Barbet, Sylvie Behillil, M√©line Bizard, Angela Brisebarre, Camille Capel, Vincent Enouf, Louise Lefran√ßois, Fr√©d√©ric Lemoine, Christophe Malabat, Corinne Maufrais, Slim El Khiari, Julien Fumey, Etienne Simon-Lori√®re, Maud Vanpeene, Sylvie Van |
| hCoV-19/Slovenia/590410/2021 | EPI_ISL_9477772 | 2021-12-30 | Institute of Microbiology and Immunology, Faculty of Medicine, University of Ljubljana | Institute of Microbiology and Immunology, Faculty of Medicine, University of Ljubljana | Alen Suljiƒç, Samo Zakotnik, Toma≈æ Mark Zorec, Doroteja Vlaj, Andra≈æ Celar, Tina ≈Ωiviƒç, Domen Lazar, Tina Gabrov≈°ek, Patricija Pozvek, ≈†pela Pleh, Mi≈°a Korva, Mario Poljak, Tatjana Av≈°iƒç ‚Äì ≈Ωupanc |
| hCoV-19/Canada/QC-L00413203001/2021 | EPI_ISL_9487343 | 2021-11-24 | Laboratoire de sant√© publique du Qu√©bec | Laboratoire de sant√© publique du Qu√©bec | Sandrine Moreira, Ioannis Ragoussis, Guillaume Bourque, Jesse Shapiro, Mark Lathrop and Judith Fafard on behalf of the CoVSeQ research group |
| hCoV-19/Canada/QC-L00413189001/2021 | EPI_ISL_9487345 | 2021-11-24 | Laboratoire de sant√© publique du Qu√©bec | Laboratoire de sant√© publique du Qu√©bec | Sandrine Moreira, Ioannis Ragoussis, Guillaume Bourque, Jesse Shapiro, Mark Lathrop and Judith Fafard on behalf of the CoVSeQ research group |
| hCoV-19/Canada/QC-L00413175001/2021 | EPI_ISL_9487347 | 2021-11-24 | Laboratoire de sant√© publique du Qu√©bec | Laboratoire de sant√© publique du Qu√©bec | Sandrine Moreira, Ioannis Ragoussis, Guillaume Bourque, Jesse Shapiro, Mark Lathrop and Judith Fafard on behalf of the CoVSeQ research group |
| hCoV-19/Canada/QC-L00413264001/2021 | EPI_ISL_9487358 | 2021-11-24 | Laboratoire de sant√© publique du Qu√©bec | Laboratoire de sant√© publique du Qu√©bec | Sandrine Moreira, Ioannis Ragoussis, Guillaume Bourque, Jesse Shapiro, Mark Lathrop and Judith Fafard on behalf of the CoVSeQ research group |
| hCoV-19/Ecuador/USFQ-2512/2021 | EPI_ISL_9489246 | 2021-12-02 | Institute of Microbiology, Universidad San Francisco de Quito | Institute of Microbiology, Universidad San Francisco de Quito | Sully M√°rquez, Erika B. Mu√±oz, Mateo Carvajal, Bel√©n Prado-Vivar, Fernanda Zurita, Rommel Guevara, Bernardo Guti√©rrez,Guzm√°n Bernab√©u LorenzoGuzm√°n Bernab√©u Lorenzo, Ver√≥nica Barrag√°n, Patricio Rojas-Silva, Gabriel Trueba, Michelle Grunauer, Pa√ |
| hCoV-19/Ecuador/USFQ-2513/2021 | EPI_ISL_9489247 | 2021-12-05 | Institute of Microbiology, Universidad San Francisco de Quito | Institute of Microbiology, Universidad San Francisco de Quito | Mateo Carvajal, Sully M√°rquez, Bel√©n Prado-Vivar, Erika B. Mu√±oz, Fernanda Zurita, Bernardo Guti√©rrez, Guzm√°n Bernab√©u Lorenzo, Ver√≥nica Barrag√°n, Patricio Rojas-Silva, Gabriel Trueba, Michelle Grunauer, Pa√∫l C√°rdenas |

| | | | | | |
|---|---|---|---|---|---|
| hCoV-19/Ecuador/USFQ-2568/2021 | EPI_ISL_9489291 | 2021-01-10 | Institute of Microbiology, Universidad San Francisco de Quito | Institute of Microbiology, Universidad San Francisco de Quito | Mateo Carvajal, Sully M√°rquez, Bel√©n Prado-Vivar, Erika B. Mu√±oz, Fernanda Zurita, Bernardo Guti√©rrez, Ver√≥nica Barrag√°n, Patricio Rojas-Silva, Gabriel Trueba, Michelle Grunauer, Pa√∫l C√°rdenas |
| hCoV-19/India/MH-ICMR-NIV-INSACOG-382936/2021 | EPI_ISL_9494481 | 2021-12-21 | ICMR-National Institute of Virology - INSACOG | NIV Influenza | Dr. Varsha Potdar and NIC Team |
| hCoV-19/India/MH-ICMR-NIV-INSACOG-383864/2021 | EPI_ISL_9494525 | 2021-12-24 | ICMR-National Institute of Virology - INSACOG | NIV Influenza | Dr. Varsha Potdar and NIC Team |
| hCoV-19/India/MH-ICMR-NIV-INSACOG-383865/2021 | EPI_ISL_9494526 | 2021-12-25 | ICMR-National Institute of Virology - INSACOG | NIV Influenza | Dr. Varsha Potdar and NIC Team |
| hCoV-19/India/MH-ICMR-NIV-INSACOG-384188/2021 | EPI_ISL_9494575 | 2021-12-23 | ICMR-National Institute of Virology - INSACOG | NIV Influenza | Dr. Varsha Potdar and NIC Team |
| hCoV-19/India/MH-ICMR-NIV-INSACOG-384981/2021 | EPI_ISL_9494716 | 2021-11-28 | ICMR-National Institute of Virology - INSACOG | NIV Influenza | Dr. Varsha Potdar and NIC Team |
| hCoV-19/India/MH-ICMR-NIV-INSACOG-385614/2021 | EPI_ISL_9494789 | 2021-12-28 | ICMR-National Institute of Virology - INSACOG | NIV Influenza | Dr. Varsha Potdar and NIC Team |
| hCoV-19/India/MH-ICMR-NIV-INSACOG-382015/2021 | EPI_ISL_9503938 | 2021-12-20 | ICMR-National Institute of Virology - INSACOG | NIV Influenza | Dr. Varsha Potdar and NIC Team |
| hCoV-19/India/MH-ICMR-NIV-INSACOG-382022/2021 | EPI_ISL_9503941 | 2021-12-20 | ICMR-National Institute of Virology - INSACOG | NIV Influenza | Dr. Varsha Potdar and NIC Team |
| hCoV-19/Canada/QC-L00426390001/2021 | EPI_ISL_9508708 | 2021-12-25 | Laboratoire de sant√© publique du Qu√©bec | Laboratoire de sant√© publique du Qu√©bec | Sandrine Moreira, Ioannis Ragoussis, Guillaume Bourque, Jesse Shapiro, Mark Lathrop and Judith Fafard on behalf of the CoVSeQ research group |
| hCoV-19/India/MH-KasturbaMCGM-ICMR-INSACOG-WG2505/2021 | EPI_ISL_9512293 | 2021-12-30 | Kasturba Hospital Molecular Lab - INSACOG | Kasturba hospital WGS | Jayanthi Shastri, Vidushi Chitalia |
| hCoV-19/Brazil/GO-LACEN-728/2021 | EPI_ISL_9534369 | 2021-12-07 | LACEN | LGBio (Laboratorio de Genetica & Biodiversidade) | Mariana Pires de Campos Telles, Daniela de Melo e Silva, Elisangela de Paula Silveira Lacerda, Renata de Oliveira Dias, Rhewter Nunes, Cintia Pelegrineti Targueta de Azevedo Brito, Ramilla dos Santos Braga, Thais Guimar√£es Castro, Thays Millena Alves Ped |
| hCoV-19/USA/CA-HLX-STM-3M8DBCYEJ/2021 | EPI_ISL_9534862 | 2021-12-25 | Kaiser Permanente Southern California | Helix | Kaiser Permanente Southern California, Helix |
| hCoV-19/India/MH-ICMR-NIV-INSACOG-383865/2021 | EPI_ISL_9494526 | 2021-12-25 | ICMR-National Institute of Virology - INSACOG | NIV Influenza | Dr. Varsha Potdar and NIC Team |
| hCoV-19/India/MH-ICMR-NIV-INSACOG-384188/2021 | EPI_ISL_9494575 | 2021-12-23 | ICMR-National Institute of Virology - INSACOG | NIV Influenza | Dr. Varsha Potdar and NIC Team |
| hCoV-19/India/MH-ICMR-NIV-INSACOG-384981/2021 | EPI_ISL_9494716 | 2021-11-28 | ICMR-National Institute of Virology - INSACOG | NIV Influenza | Dr. Varsha Potdar and NIC Team |
| hCoV-19/India/MH-ICMR-NIV-INSACOG-385614/2021 | EPI_ISL_9494789 | 2021-12-28 | ICMR-National Institute of Virology - INSACOG | NIV Influenza | Dr. Varsha Potdar and NIC Team |
| hCoV-19/India/MH-ICMR-NIV-INSACOG-382015/2021 | EPI_ISL_9503938 | 2021-12-20 | ICMR-National Institute of Virology - INSACOG | NIV Influenza | Dr. Varsha Potdar and NIC Team |
| hCoV-19/India/MH-ICMR-NIV-INSACOG-382022/2021 | EPI_ISL_9503941 | 2021-12-20 | ICMR-National Institute of Virology - INSACOG | NIV Influenza | Dr. Varsha Potdar and NIC Team |

| | | | | | |
|---|---|---|---|---|---|
| hCoV-19/Canada/QC-L00426390001/2021 | EPI_ISL_9508708 | 2021-12-25 | Laboratoire de sant√© publique du Qu√©bec | Laboratoire de sant√© publique du Qu√©bec | Sandrine Moreira, Ioannis Ragoussis, Guillaume Bourque, Jesse Shapiro, Mark Lathrop and Judith Fafard on behalf of the CoVSeQ research group |
| hCoV-19/India/MH-KasturbaMCGM-ICMR-INSACOG-WG2505/2021 | EPI_ISL_9512293 | 2021-12-30 | Kasturba Hospital Molecular Lab - INSACOG | Kasturba hospital WGS | Jayanthi Shastri, Vidushi Chitalia |
| hCoV-19/Brazil/GO-LACEN-728/2021 | EPI_ISL_9534369 | 2021-12-07 | LACEN | LGBio (Laboratorio de Genetica & Biodiversidade) | Mariana Pires de Campos Telles, Daniela de Melo e Silva, Elisangela de Paula Silveira Lacerda, Renata de Oliveira Dias, Rhewter Nunes, Cintia Pelegrineti Targueta de Azevedo Brito, Ramilla dos Santos Braga, Thais Guimar√£es Castro, Thays Millena Alves Ped |
| hCoV-19/USA/CA-HLX-STM-3M8DBCYEJ/2021 | EPI_ISL_9534862 | 2021-12-25 | Kaiser Permanente Southern California | Helix | Kaiser Permanente Southern California, Helix |
| hCoV-19/USA/CA-HLX-STM-0006331-H04/2021 | EPI_ISL_9536076 | 2021-07-23 | Kaiser Permanente Southern California | Helix | Kaiser Permanente Southern California, Helix |
| hCoV-19/USA/CA-HLX-STM-0006319-C07/2021 | EPI_ISL_9537048 | 2021-08-23 | Kaiser Permanente Southern California | Helix | Kaiser Permanente Southern California, Helix |
| hCoV-19/USA/CA-HLX-STM-0006298-H06/2021 | EPI_ISL_9537653 | 2021-06-03 | Kaiser Permanente Southern California | Helix | Kaiser Permanente Southern California, Helix |
| hCoV-19/USA/CA-HLX-STM-0006298-G08/2021 | EPI_ISL_9537677 | 2021-05-29 | Kaiser Permanente Southern California | Helix | Kaiser Permanente Southern California, Helix |
| hCoV-19/USA/CA-HLX-STM-0006255-A12/2021 | EPI_ISL_9540530 | 2021-07-28 | Kaiser Permanente Southern California | Helix | Kaiser Permanente Southern California, Helix |
| hCoV-19/USA/CA-HLX-STM-0006236-B12/2021 | EPI_ISL_9541365 | 2021-07-25 | Kaiser Permanente Southern California | Helix | Kaiser Permanente Southern California, Helix |
| hCoV-19/USA/CA-HLX-STM-0006176-E12/2021 | EPI_ISL_9544907 | 2021-07-20 | Kaiser Permanente Southern California | Helix | Kaiser Permanente Southern California, Helix |
| hCoV-19/USA/CA-HLX-STM-0006116-E08/2021 | EPI_ISL_9547706 | 2021-06-28 | Kaiser Permanente Southern California | Helix | Kaiser Permanente Southern California, Helix |
| hCoV-19/USA/CA-HLX-STM-0006077-E12/2021 | EPI_ISL_9548889 | 2021-06-14 | Kaiser Permanente Southern California | Helix | Kaiser Permanente Southern California, Helix |
| hCoV-19/USA/CA-HLX-STM-0006077-D09/2021 | EPI_ISL_9548956 | 2021-06-15 | Kaiser Permanente Southern California | Helix | Kaiser Permanente Southern California, Helix |
| hCoV-19/India/WB-INSACOG-1931502620882/2021 | EPI_ISL_9550982 | 2021-12-27 | INSACOG-WB | National¬†Institute¬†of¬†Biomedical ¬†Genomics ‚Äì¬†INSACOG | Arindam Maitra, Bhaswati Bandyopadhyay, Nidhan Kumar Biswas, Tamal Ghosh, Sreedhar Chinnaswamy, Ajay Chakraborti, Saumitra Das |
| hCoV-19/India/WB-INSACOG-1930301681781/2021 | EPI_ISL_9551016 | 2021-12-30 | INSACOG-WB | National¬†Institute¬†of¬†Biomedical ¬†Genomics ‚Äì¬†INSACOG | Arindam Maitra, Bhaswati Bandyopadhyay, Nidhan Kumar Biswas, Tamal Ghosh, Sreedhar Chinnaswamy, Ajay Chakraborti, Saumitra Das |
| hCoV-19/India/WB-INSACOG-1930301673523/2021 | EPI_ISL_9551063 | 2021-12-28 | INSACOG-WB | National¬†Institute¬†of¬†Biomedical ¬†Genomics ‚Äì¬†INSACOG | Arindam Maitra, Bhaswati Bandyopadhyay, Nidhan Kumar Biswas, Tamal Ghosh, Sreedhar Chinnaswamy, Ajay Chakraborti, Saumitra Das |
| hCoV-19/India/WB-INSACOG-1931502632731/2021 | EPI_ISL_9551188 | 2021-12-29 | INSACOG-WB | National¬†Institute¬†of¬†Biomedical ¬†Genomics ‚Äì¬†INSACOG | Arindam Maitra, Bhaswati Bandyopadhyay, Nidhan Kumar Biswas, Tamal Ghosh, Sreedhar Chinnaswamy, Ajay Chakraborti, Saumitra Das |

| | | | | | |
|---|---|---|---|---|---|
| hCoV-19/India/WB-INSACOG-1930301678657/2021 | EPI_ISL_9551212 | 2021-12-29 | INSACOG-WB | National¬†Institute¬†of¬†Biomedical ¬†Genomics ‚Äì¬†INSACOG | Arindam Maitra, Bhaswati Bandyopadhyay, Nidhan Kumar Biswas, Tamal Ghosh, Sreedhar Chinnaswamy, Ajay Chakraborti, Saumitra Das |
| hCoV-19/India/WB-INSACOG-1930301677626/2021 | EPI_ISL_9551342 | 2021-12-29 | INSACOG-WB | National¬†Institute¬†of¬†Biomedical ¬†Genomics ‚Äì¬†INSACOG | Arindam Maitra, Bhaswati Bandyopadhyay, Nidhan Kumar Biswas, Tamal Ghosh, Sreedhar Chinnaswamy, Ajay Chakraborti, Saumitra Das |
| hCoV-19/Ireland/G-Enfer-COV261221009_G1/2021 | EPI_ISL_9554263 | 2021-12-26 | Enfer | Enfer | Elaine M. Kenny, Suzie Coughlan |
| hCoV-19/DRC/INRB-RDC-706/2021 | EPI_ISL_9570964 | 2021-11-29 | Pathogen Genomics Lab, National Institute for Biomedical Research (INRB) | Pathogen Genomics Lab, National Institute for Biomedical Research (INRB) | Placide Mbala-Kingebeni, Edith Nkwembe, Eddy Kinganda-Lusamaki, Amuri Aziza, Francisca Muyembe Mawete, Emmanuel Lokilo Lofiko, Gradi Luakanda Ndelemo, Jean Claude Makangara, Raphaël Lumembe, Gabriel Kabamba, Catherine Pratt, Matthias Pauthner, Josh Quick |
| hCoV-19/DRC/INRB-RDC-791/2021 | EPI_ISL_9570996 | 2021-10-26 | Pathogen Genomics Lab, National Institute for Biomedical Research (INRB) | Pathogen Genomics Lab, National Institute for Biomedical Research (INRB) | Placide Mbala-Kingebeni, Edith Nkwembe, Eddy Kinganda-Lusamaki, Amuri Aziza, Francisca Muyembe Mawete, Emmanuel Lokilo Lofiko, Gradi Luakanda Ndelemo, Jean Claude Makangara, Raphaël Lumembe, Gabriel Kabamba, Catherine Pratt, Matthias Pauthner, Josh Quick |
| hCoV-19/DRC/INRB-RDC-981/2021 | EPI_ISL_9571096 | 2021-12-15 | Pathogen Genomics Lab, National Institute for Biomedical Research (INRB) | Pathogen Genomics Lab, National Institute for Biomedical Research (INRB) | Placide Mbala-Kingebeni, Edith Nkwembe, Eddy Kinganda-Lusamaki, Amuri Aziza, Francisca Muyembe Mawete, Emmanuel Lokilo Lofiko, Gradi Luakanda Ndelemo, Jean Claude Makangara, Raphaël Lumembe, Gabriel Kabamba, Catherine Pratt, Matthias Pauthner, Josh Quick |
| hCoV-19/DRC/INRB-RDC-1006/2021 | EPI_ISL_9571103 | 2021-12-17 | Pathogen Genomics Lab, National Institute for Biomedical Research (INRB) | Pathogen Genomics Lab, National Institute for Biomedical Research (INRB) | Placide Mbala-Kingebeni, Edith Nkwembe, Eddy Kinganda-Lusamaki, Amuri Aziza, Francisca Muyembe Mawete, Emmanuel Lokilo Lofiko, Gradi Luakanda Ndelemo, Jean Claude Makangara, Raphaël Lumembe, Gabriel Kabamba, Catherine Pratt, Matthias Pauthner, Josh Quick |
| hCoV-19/Brazil/SC-FIOCRUZ-66853/2021 | EPI_ISL_9571572 | 2021-12-30 | Laboratorio Central de Saude Publica do Estado de Santa Catarina (LACEN/SC) | Laboratory of Respiratory Viruses and Measles, Oswaldo Cruz Institute, FIOCRUZ | Paola Resende, Luciana Appolinario, Fernando Motta, Elisa Cavalcante Pereira, Ighor Arantes, Alice Sampaio Rocha, Bruna Mendonça da Silva, Jéssica Graça Macedo de Carvalho, Larissa Macedo Pinto, Victor Guimaraes, Darcita Buerger Rovaris, Sandra Bianchi |
| hCoV-19/Brazil/RS-FIOCRUZ-66744/2021 | EPI_ISL_9571611 | 2021-12-24 | Laboratorio Central de Saude Publica do Estado do Rio Grande do Sul (LACEN/RS) | Laboratory of Respiratory Viruses and Measles, Oswaldo Cruz Institute, FIOCRUZ | Paola Resende, Luciana Appolinario, Fernando Motta, Alice Sampaio Rocha, Elisa Cavalcante Pereira, Bruna Mendonça da Silva, Jéssica Graça Macedo de Carvalho, Larissa Macedo Pinto, Victor Guimaraes, Ighor Leonardo Arantes, Anderson Brandao Leite, Marild |
| hCoV-19/USA/GA-EHC-3410D/2021 | EPI_ISL_9571985 | 2021-12-21 | Emory Molecular Diagnostics Laboratory, Emory Healthcare | Piantadosi Lab, Emory Department of Pathology | Ludy Registre Carmola, Anne Piantadosi |
| hCoV-19/Malaysia/UiTM1051/2021 | EPI_ISL_9579120 | 2021-12-20 | PKD SPT | iPROMISE, UiTM | Mohd Zaki Salleh, Teh Lay Kek, Ariza Adnan, Sazzli Shahlan Kassim, Fadzilah Mohd Nor, Wang Seok Mui, Siti Hamimah Sheikh Abdul Kadir, Mohd Nur Fakhruzzaman Noorizhab, Siti Farah Alwani Mohd Nawi, Lim Wai Feng, Mohd Asif Mohd Sukri |

| | | | | | |
|---|---|---|---|---|---|
| hCoV-19/Malaysia/UiTM1152/2021 | EPI_ISL_9579184 | 2021-11-26 | Pejabat Kesihatan Daerah Timur Laut | iPROMISE, UiTM | Mohd Zaki Salleh, Teh Lay Kek, Ariza Adnan, Sazzli Shahlan Kassim, Fadzilah Mohd Nor, Wang Seok Mui, Siti Hamimah Sheikh Abdul Kadir, Mohd Nur Fakhruzzaman Noorizhab, Siti Farah Alwani Mohd Nawi, Lim Wai Feng, Mohd Asif Mohd Sukri |
| hCoV-19/Cameroon/DO10144/2021 | EPI_ISL_9585253 | 2021-11-13 | CREMER (Centre de Recherches sur les Maladies Emergentes et Re-emergentes) | CREMER (Centre de Recherches sur les Maladies Emergentes et Re-emergentes) | Celestin Godwe, Martin Maidadi Foudi, Dowbiss Meta Djomsi,Ginette Edoul Mbesse, Marie Amougou,Nicolas Fernandez Nunez, Eitel Mpoudi Ngole, Ahidjo Ayouba, Marcel Tongo, Martine Peeters, Eric Delaporte |
| hCoV-19/England/PHEP-YYD4GXR/2021 | EPI_ISL_9589414 | 2021-05-10 | Respiratory Virus Unit, Microbiology Services Colindale, Public Health England | COVID-19 Genomics UK (COG-UK) Consortium | PHE Covid Sequencing Team |
| hCoV-19/Reunion/CHU721122643603/2021 | EPI_ISL_9608023 | 2021-12-26 | CHU - Saint-Pierre | Laboratoire de virologie, CNR arbovirus Associ√©, Chu de la R√©union | Etienne Frumence, Nicolas M'nemosyme, Anne-Julie Gourd√©, Rubens Lhonneur, Nicolas Traversier, Marie-Christine Jaffar Bandjee |
| hCoV-19/Reunion/CHU721122346601/2021 | EPI_ISL_9608116 | 2021-12-23 | LABM SAINT BENOIT | Laboratoire de virologie, CNR arbovirus Associ√©, Chu de la R√©union | Etienne Frumence, Nicolas M'nemosyme, Anne-Julie Gourd√©, Rubens Lhonneur, Nicolas Traversier, Marie-Christine Jaffar Bandjee |
| hCoV-19/Reunion/CHU721122072101/2021 | EPI_ISL_9608195 | 2021-12-21 | LABM SAINT BENOIT | Laboratoire de virologie, CNR arbovirus Associ√©, Chu de la R√©union | Etienne Frumence, Nicolas M'nemosyme, Anne-Julie Gourd√©, Rubens Lhonneur, Nicolas Traversier, Marie-Christine Jaffar Bandjee |
| hCoV-19/Reunion/CHU721122070201/2021 | EPI_ISL_9608206 | 2021-12-21 | LABM SAINT BENOIT | Laboratoire de virologie, CNR arbovirus Associ√©, Chu de la R√©union | Etienne Frumence, Nicolas M'nemosyme, Anne-Julie Gourd√©, Rubens Lhonneur, Nicolas Traversier, Marie-Christine Jaffar Bandjee |
| hCoV-19/Reunion/CHU79800009540202/2021 | EPI_ISL_9608365 | 2021-12-10 | CHU - Saint-Denis | Laboratoire de virologie, CNR arbovirus Associ√©, Chu de la R√©union | Etienne Frumence, Nicolas M'nemosyme, Anne-Julie Gourd√©, Rubens Lhonneur, Nicolas Traversier, Marie-Christine Jaffar Bandjee |
| hCoV-19/India/WB-INSACOG-707903380074/2021 | EPI_ISL_9608751 | 2021-12-25 | INSACOG-WB | National¬†Institute¬†of¬†Biomedical ¬†Genomics ‚Äì¬†INSACOG | Arindam Maitra, Bhaswati Bandyopadhyay, Nidhan Kumar Biswas, Tamal Ghosh, Sreedhar Chinnaswamy, Ajay Chakraborti, Saumitra Das |
| hCoV-19/India/WB-INSACOG-1931502626643/2021 | EPI_ISL_9608755 | 2021-12-28 | INSACOG-WB | National¬†Institute¬†of¬†Biomedical ¬†Genomics ‚Äì¬†INSACOG | Arindam Maitra, Bhaswati Bandyopadhyay, Nidhan Kumar Biswas, Tamal Ghosh, Sreedhar Chinnaswamy, Ajay Chakraborti, Saumitra Das |
| hCoV-19/India/WB-INSACOG-1931502628163/2021 | EPI_ISL_9608870 | 2021-12-28 | INSACOG-WB | National¬†Institute¬†of¬†Biomedical ¬†Genomics ‚Äì¬†INSACOG | Arindam Maitra, Bhaswati Bandyopadhyay, Nidhan Kumar Biswas, Tamal Ghosh, Sreedhar Chinnaswamy, Ajay Chakraborti, Saumitra Das |
| hCoV-19/India/WB-INSACOG-1931502617383/2021 | EPI_ISL_9608900 | 2021-12-26 | INSACOG-WB | National¬†Institute¬†of¬†Biomedical ¬†Genomics ‚Äì¬†INSACOG | Arindam Maitra, Bhaswati Bandyopadhyay, Nidhan Kumar Biswas, Tamal Ghosh, Sreedhar Chinnaswamy, Ajay Chakraborti, Saumitra Das |
| hCoV-19/India/WB-INSACOG-1931502591073/2021 | EPI_ISL_9609035 | 2021-12-20 | INSACOG-WB | National¬†Institute¬†of¬†Biomedical ¬†Genomics ‚Äì¬†INSACOG | Arindam Maitra, Bhaswati Bandyopadhyay, Nidhan Kumar Biswas, Tamal Ghosh, Sreedhar Chinnaswamy, Ajay Chakraborti, Saumitra Das |
| hCoV-19/India/WB-INSACOG-1931502622699/2021 | EPI_ISL_9609118 | 2021-12-27 | INSACOG-WB | National¬†Institute¬†of¬†Biomedical ¬†Genomics ‚Äì¬†INSACOG | Arindam Maitra, Bhaswati Bandyopadhyay, Nidhan Kumar Biswas, Tamal Ghosh, Sreedhar Chinnaswamy, Ajay Chakraborti, Saumitra Das |

| | | | | | |
|---|---|---|---|---|---|
| hCoV-19/India/WB-INSACOG-707903365568/2021 | EPI_ISL_9609143 | 2021-12-24 | INSACOG-WB | National¬†Institute¬†of¬†Biomedical ¬†Genomics ‚Äì¬†INSACOG | Arindam Maitra, Bhaswati Bandyopadhyay, Nidhan Kumar Biswas, Tamal Ghosh, Sreedhar Chinnaswamy, Ajay Chakraborti, Saumitra Das |
| hCoV-19/India/WB-INSACOG-1931502615780/2021 | EPI_ISL_9609158 | 2021-12-26 | INSACOG-WB | National¬†Institute¬†of¬†Biomedical ¬†Genomics ‚Äì¬†INSACOG | Arindam Maitra, Bhaswati Bandyopadhyay, Nidhan Kumar Biswas, Tamal Ghosh, Sreedhar Chinnaswamy, Ajay Chakraborti, Saumitra Das |
| hCoV-19/Nigeria/NCDC-NRL-GL-000199/2021 | EPI_ISL_9609756 | 2021-12-24 | 54 GENE OGUN | National Reference Laboratory, Nigeria Centre for Disease Control | Dr Ndodo Nnaemeka, Olusola Anuoluwapo Akanbi, Chimaobi Chukwu, Oyeronke Ayansola, Dr Omoare Adesuyi, Anthony Ahumibe, Catherine Okoi, Nwando Mba, Dr Ifedayo Adetifa |
| hCoV-19/Nigeria/NCDC-NR2402/2021 | EPI_ISL_9610379 | 2021-12-21 | E-CLINIC | National Reference Laboratory, Nigeria Centre for Disease Control | Dr Ndodo Nnaemeka, Olusola Anuoluwapo Akanbi, Chimaobi Chukwu, Oyeronke Ayansola, Dr Omoare Adesuyi, Anthony Ahumibe, Catherine Okoi, Nwando Mba, Dr Ifedayo Adetifa |
| hCoV-19/France/ARA-CERBAHC-12646949/2021 | EPI_ISL_9617758 | 2021-12-27 | CERBALLIANCE RHONE ALPES | CERBA HealthCare | B√©n√©dicte Roquebert, Laura Verdurme, Mathilde Roussel, Sabine Trombert, St√©phanie Haim-Boukobza |
| hCoV-19/Norway/35622/2021 | EPI_ISL_9626450 | 2021-01-06 | Department of Medical Microbiology - section Molde, Molde Hospital | Norwegian Institute of Public Health, Department of Virology | Kathrine Stene-Johansen, Kamilla Heddeland Instefjord, Hilde Elshaug, Garcia Llorente Ignacio, Jon Br√•te, Engebretsen Serina Beate, Pedersen Benedikte Nevjen, Line Victoria Moen, Hilde Nordby Falkenhaug, Debech Nadia, Atiya R Ali, Marie Paulsen Madsen, R |
| hCoV-19/Suriname/SR-913/2021 | EPI_ISL_9628807 | 2021-01-01 | Central Laboratory, Bureau of Public Health (BOG) and Academic Hospital Paramaribo | Erasmus Medical Center | Bas B Oude Munnink, Cherise Beek, Consuella Partowidjojo, Dion Gajadin, Ed PF IJzerman, Emmanuelle Munger, Gary Gummels, Ingrid SK Krishnadath, Lycke Woittiez, Marion PG Koopmans, Mireille Van de Veer, Phyllis Pinas, Princes Wongsowidjojo, Radjesh Ori, Ra |
| hCoV-19/Suriname/SR-918/2021 | EPI_ISL_9628812 | 2021-01-04 | Central Laboratory, Bureau of Public Health (BOG) and Academic Hospital Paramaribo | Erasmus Medical Center | Bas B Oude Munnink, Cherise Beek, Consuella Partowidjojo, Dion Gajadin, Ed PF IJzerman, Emmanuelle Munger, Gary Gummels, Ingrid SK Krishnadath, Lycke Woittiez, Marion PG Koopmans, Mireille Van de Veer, Phyllis Pinas, Princes Wongsowidjojo, Radjesh Ori, Ra |
| hCoV-19/Suriname/SR-921/2021 | EPI_ISL_9628815 | 2021-01-06 | Central Laboratory, Bureau of Public Health (BOG) and Academic Hospital Paramaribo | Erasmus Medical Center | Bas B Oude Munnink, Cherise Beek, Consuella Partowidjojo, Dion Gajadin, Ed PF IJzerman, Emmanuelle Munger, Gary Gummels, Ingrid SK Krishnadath, Lycke Woittiez, Marion PG Koopmans, Mireille Van de Veer, Phyllis Pinas, Princes Wongsowidjojo, Radjesh Ori, Ra |
| hCoV-19/Suriname/SR-932/2021 | EPI_ISL_9628826 | 2021-01-16 | Central Laboratory, Bureau of Public Health (BOG) and Academic Hospital Paramaribo | Erasmus Medical Center | Bas B Oude Munnink, Cherise Beek, Consuella Partowidjojo, Dion Gajadin, Ed PF IJzerman, Emmanuelle Munger, Gary Gummels, Ingrid SK Krishnadath, Lycke Woittiez, Marion PG Koopmans, Mireille Van de Veer, Phyllis Pinas, Princes Wongsowidjojo, Radjesh Ori, Ra |
| hCoV-19/Suriname/SR-948/2021 | EPI_ISL_9628842 | 2021-01-19 | Central Laboratory, Bureau of Public Health (BOG) and Academic Hospital Paramaribo | Erasmus Medical Center | Bas B Oude Munnink, Cherise Beek, Consuella Partowidjojo, Dion Gajadin, Ed PF IJzerman, Emmanuelle Munger, Gary Gummels, Ingrid SK Krishnadath, Lycke Woittiez, Marion PG Koopmans, Mireille Van de Veer, Phyllis Pinas, Princes Wongsowidjojo, Radjesh Ori, Ra |

| | | | | | |
|---|---|---|---|---|---|
| hCoV-19/Suriname/SR-956/2021 | EPI_ISL_9628850 | 2021-01-08 | Central Laboratory, Bureau of Public Health (BOG) and Academic Hospital Paramaribo | Erasmus Medical Center | Bas B Oude Munnink, Cherise Beek, Consuella Partowidjojo, Dion Gajadin, Ed PF IJzerman, Emmanuelle Munger, Gary Gummels, Ingrid SK Krishnadath, Lycke Woittiez, Marion PG Koopmans, Mireille Van de Veer, Phyllis Pinas, Princes Wongsowidjojo, Radjesh Ori, Ra |
| hCoV-19/CotedIvoire/ECOV347459-cv012/2021 | EPI_ISL_9631805 | 2021-07-28 | Environment and Health Department, Institut Pasteur of C√¥te d'Ivoire, Abidjan, C√¥te d'Ivoire | Molecular Genetics Platform, Institut Pasteur of C√¥te d'Ivoire, Abidjan, C√¥te d'Ivoire | Aristide Berenger Ako AKO; Bernadette Tchepe Flore Diplo; Egomli Stanislas ASSOHOUN; Antoine Marie-David N'golo COULIBALY; Tata Gnire Safiatou COULIBALY; Jean-Ren√© Quassi ACQUAH; Djeda Franck Arnaud GNAHORE; Herv√© Alberic ADJE KADJO, Mireille DOSSO |
| hCoV-19/CotedIvoire/ECOV347350-cv017/2021 | EPI_ISL_9631922 | 2021-07-28 | Environment and Health Department, Institut Pasteur of C√¥te d'Ivoire, Abidjan, C√¥te d'Ivoire | Molecular Genetics Platform, Institut Pasteur of C√¥te d'Ivoire, Abidjan, C√¥te d'Ivoire | Aristide Berenger Ako AKO; Bernadette Tchepe Flore Diplo; Egomli Stanislas ASSOHOUN; Antoine Marie-David N'golo COULIBALY; Tata Gnire Safiatou COULIBALY; Jean-Ren√© Quassi ACQUAH; Djeda Franck Arnaud GNAHORE; Herv√© Alberic ADJE KADJO, Mireille DOSSO |
| hCoV-19/Thailand/NIC_BKK_SEQ25361/2021 | EPI_ISL_9636881 | 2021-11-29 | Bio Molecular Laboratory | National Institute of Health, Department of Medical Sciences, Ministry of Public Health, Thailand | Pilailuk Okada; Siripaporn Phuygun; Nuttida Thongpramul; Pakorn Piromtong; Natchaya¬† Khiadsang; Thanutsapa Thanadachakul; Sirikanda Wimol; Suratchana Mitrat; Sunthareeya Waicharoen; Archawin Rojanawiwat |
| hCoV-19/Thailand/NIC_CMI_SEQ25434/2021 | EPI_ISL_9636898 | 2021-11-29 | Regional Medical Sciences Center 1 Chiang Mai | National Institute of Health, Department of Medical Sciences, Ministry of Public Health, Thailand | Pilailuk Okada; Siripaporn Phuygun; Nuttida Thongpramul; Pakorn Piromtong; Natchaya¬† Khiadsang; Thanutsapa Thanadachakul; Sirikanda Wimol; Suratchana Mitrat; Sunthareeya Waicharoen; Archawin Rojanawiwat |
| hCoV-19/Thailand/NIC_CMI_SEQ25437/2021 | EPI_ISL_9636899 | 2021-11-29 | Regional Medical Sciences Center 1 Chiang Mai | National Institute of Health, Department of Medical Sciences, Ministry of Public Health, Thailand | Pilailuk Okada; Siripaporn Phuygun; Nuttida Thongpramul; Pakorn Piromtong; Natchaya¬† Khiadsang; Thanutsapa Thanadachakul; Sirikanda Wimol; Suratchana Mitrat; Sunthareeya Waicharoen; Archawin Rojanawiwat |
| hCoV-19/Thailand/NIC_BKK_SEQ00009/2021 | EPI_ISL_9636927 | 2021-11-29 | World Medical Hospital | National Institute of Health, Department of Medical Sciences, Ministry of Public Health, Thailand | Pilailuk Okada; Siripaporn Phuygun; Nuttida Thongpramul; Pakorn Piromtong; Natchaya¬† Khiadsang; Thanutsapa Thanadachakul; Sirikanda Wimol; Suratchana Mitrat; Sunthareeya Waicharoen; Archawin Rojanawiwat |
| hCoV-19/Thailand/NIC_BKK_SEQ00010/2021 | EPI_ISL_9637149 | 2021-11-29 | World Medical Hospital | National Institute of Health, Department of Medical Sciences, Ministry of Public Health, Thailand | Pilailuk Okada; Siripaporn Phuygun; Nuttida Thongpramul; Pakorn Piromtong; Natchaya¬† Khiadsang; Thanutsapa Thanadachakul; Sirikanda Wimol; Suratchana Mitrat; Sunthareeya Waicharoen; Archawin Rojanawiwat |
| hCoV-19/Thailand/NIC_BKK_SEQ00014/2021 | EPI_ISL_9637150 | 2021-11-29 | World Medical Hospital | National Institute of Health, Department of Medical Sciences, Ministry of Public Health, Thailand | Pilailuk Okada; Siripaporn Phuygun; Nuttida Thongpramul; Pakorn Piromtong; Natchaya¬† Khiadsang; Thanutsapa Thanadachakul; Sirikanda Wimol; Suratchana Mitrat; Sunthareeya Waicharoen; Archawin Rojanawiwat |
| hCoV-19/Thailand/NIC_BKK_SEQ00015/2021 | EPI_ISL_9637151 | 2021-11-29 | World Medical Hospital | National Institute of Health, Department of Medical Sciences, Ministry of Public Health, Thailand | Pilailuk Okada; Siripaporn Phuygun; Nuttida Thongpramul; Pakorn Piromtong; Natchaya¬† Khiadsang; Thanutsapa Thanadachakul; Sirikanda Wimol; Suratchana Mitrat; Sunthareeya Waicharoen; Archawin Rojanawiwat |

| | | | | | |
|---|---|---|---|---|---|
| hCoV-19/CotedIvoire/ECOV391701-cv545/2021 | EPI_ISL_9638536 | 2021-08-24 | Environment and Health Department, Institut Pasteur of C√¥te d'Ivoire, Abidjan, C√¥te d'Ivoire | Molecular Genetics Platform, Institut Pasteur of C√¥te d'Ivoire, Abidjan, C√¥te d'Ivoire | Aristide Berenger Ako AKO; Bernadette Tchepe Flore Diplo; Egomli Stanislas ASSOHOUN; Antoine Marie-David N'golo COULIBALY; Tata Gnire Safiatou COULIBALY; Jean-Ren√© Quassi ACQUAH; Djeda Franck Arnaud GNAHORE; Herv√© Alberic ADJE KADJO, Mireille DOSSO |
| hCoV-19/Australia/QLD8205/2021 | EPI_ISL_9638675 | 2021-12-29 | PHV-FSS | PHV-FSS | Chenwei Wang on behalf of Q-PHIRE Genomics |
| hCoV-19/Australia/QLD14415/2021 | EPI_ISL_9638908 | 2021-12-27 | PHV-FSS | PHV-FSS | Chenwei Wang on behalf of Q-PHIRE Genomics |
| hCoV-19/Australia/QLD7363/2021 | EPI_ISL_9639207 | 2021-12-28 | PHV-FSS | PHV-FSS | Chenwei Wang on behalf of Q-PHIRE Genomics |
| hCoV-19/Australia/QLD8436/2021 | EPI_ISL_9639221 | 2021-12-29 | PHV-FSS | PHV-FSS | Chenwei Wang on behalf of Q-PHIRE Genomics |
| hCoV-19/CotedIvoire/ECOV391589-cv560/2021 | EPI_ISL_9640013 | 2021-08-24 | Environment and Health Department, Institut Pasteur of C√¥te d'Ivoire, Abidjan, C√¥te d'Ivoire | Molecular Genetics Platform, Institut Pasteur of C√¥te d'Ivoire, Abidjan, C√¥te d'Ivoire | Aristide Berenger Ako AKO; Bernadette Tchepe Flore Diplo; Egomli Stanislas ASSOHOUN; Antoine Marie-David N'golo COULIBALY; Tata Gnire Safiatou COULIBALY; Jean-Ren√© Quassi ACQUAH; Djeda Franck Arnaud GNAHORE; Herv√© Alberic ADJE KADJO, Mireille DOSSO |
| hCoV-19/mink/Poland/53970/2021 | EPI_ISL_9640055 | 2021-12-15 | Department of Virology, National Veterinary Research Institute | Department of Omics Analysis, National Veterinary Research Institute | Orlowska Anna, Smreczak Marcin, Trebas Pawel, Lisowska Anna, Opolska Justyna, Bomba Arkadiusz, Katarzyna Tluscik, Iwan Ewelina, Niemczuk Krzysztof, Domanska-Blicharz Katarzyna, Rola Jerzy |
| hCoV-19/USA/IL-RIPHL_10916_G/2021 | EPI_ISL_9648573 | 2021-12-15 | Rush University Medical Center | RIPHL at Rush University Medical Center | Stefan Green, Kevin Kunstman, Laura Furtado, Felix Araujo Perez, Edith Perez, Sofiya Bobrovska, Alyse Kittner, Hannah Barbian, Cecilia Chau, Mary Hayden, Joyce Houlihan, Diane Springer |
| hCoV-19/SouthAfrica/UFS-VIRO-NGS-ONT030222NB12/2021 | EPI_ISL_9662361 | 2021-12-02 | NHLS Universitas Academic | UFS Virology | PA Bester, MM Nyaga, P Nthiga, MT Mogotsi, Emmanuel Ogunbayo, D Goedhals, T de Oliveira |
| hCoV-19/Nigeria/NCDC-NR1188/2021 | EPI_ISL_9673555 | 2021-10-12 | ZANKLI MEDICAL CENTRE | National Reference Laboratory, Nigeria Centre for Disease Control | Dr Ndodo Nnaemeka, Olusola Anuoluwapo Akanbi, Oyeronke Ayansola, Chimaobi Chukwu, Dr Omoare Adesuyi, Anthony Ahumibe, Catherine Okoi, Nwando Mba, Dr Ifedayo Adetifa |
| hCoV-19/Nigeria/NCDC-NR1239/2021 | EPI_ISL_9673570 | 2021-11-27 | EPICONSULT | National Reference Laboratory, Nigeria Centre for Disease Control | Dr Ndodo Nnaemeka, Olusola Anuoluwapo Akanbi, Oyeronke Ayansola, Chimaobi Chukwu, Dr Omoare Adesuyi, Anthony Ahumibe, Catherine Okoi, Nwando Mba, Dr Ifedayo Adetifa |
| hCoV-19/Nigeria/NCDC-NR-GL-000464/2021 | EPI_ISL_9673583 | 2021-12-28 | NIZAMIYE | National Reference Laboratory, Nigeria Centre for Disease Control | Dr Ndodo Nnaemeka, Olusola Anuoluwapo Akanbi, Oyeronke Ayansola, Chimaobi Chukwu, Dr Omoare Adesuyi, Anthony Ahumibe, Catherine Okoi, Nwando Mba, Dr Ifedayo Adetifa |
| hCoV-19/Nigeria/NCDC-NR-GL-000472/2021 | EPI_ISL_9673591 | 2021-12-29 | NIZAMIYE | National Reference Laboratory, Nigeria Centre for Disease Control | Dr Ndodo Nnaemeka, Olusola Anuoluwapo Akanbi, Oyeronke Ayansola, Chimaobi Chukwu, Dr Omoare Adesuyi, Anthony Ahumibe, Catherine Okoi, Nwando Mba, Dr Ifedayo Adetifa |

| | | | | | |
|---|---|---|---|---|---|
| hCoV-19/SouthAfrica/NICD-N28936/2021 | EPI_ISL_9679267 | 2021-12-11 | AMPATH | National Institute for Communicable Diseases of the National Health Laboratory Service | Amoako DG, Everatt J, Kekana D, Scheepers C, Mohale T, Ntuli N, Mahlangu B, Mnguni A, Ismail A, Bhiman JN, Wolter N |
| hCoV-19/SouthAfrica/NICD-N28994/2021 | EPI_ISL_9679276 | 2021-11-27 | AMPATH | National Institute for Communicable Diseases of the National Health Laboratory Service | Amoako DG, Everatt J, Kekana D, Scheepers C, Mohale T, Ntuli N, Mahlangu B, Mnguni A, Ismail A, Bhiman JN, Wolter N |
| hCoV-19/SouthAfrica/NICD-N29109/2021 | EPI_ISL_9679338 | 2021-12-02 | AMPATH | National Institute for Communicable Diseases of the National Health Laboratory Service | Amoako DG, Everatt J, Kekana D, Scheepers C, Mohale T, Ntuli N, Mahlangu B, Mnguni A, Ismail A, Bhiman JN, Wolter N |
| hCoV-19/SouthAfrica/NICD-N29265/2021 | EPI_ISL_9679450 | 2021-12-19 | AMPATH | National Institute for Communicable Diseases of the National Health Laboratory Service | Amoako DG, Everatt J, Kekana D, Scheepers C, Mohale T, Ntuli N, Mahlangu B, Mnguni A, Ismail A, Bhiman JN, Wolter N |
| hCoV-19/SouthAfrica/NICD-N29275/2021 | EPI_ISL_9679457 | 2021-12-19 | AMPATH | National Institute for Communicable Diseases of the National Health Laboratory Service | Amoako DG, Everatt J, Kekana D, Scheepers C, Mohale T, Ntuli N, Mahlangu B, Mnguni A, Ismail A, Bhiman JN, Wolter N |
| hCoV-19/SouthAfrica/NICD-N29940/2021 | EPI_ISL_9679482 | 2021-12-20 | PORT ELIZABETH LABORATORY | National Institute for Communicable Diseases of the National Health Laboratory Service | Amoako DG, Everatt J, Kekana D, Scheepers C, Mohale T, Ntuli N, Mahlangu B, Mnguni A, Ismail A, Bhiman JN, Wolter N |
| hCoV-19/SouthAfrica/NICD-N30013/2021 | EPI_ISL_9679542 | 2021-12-19 | PORT ELIZABETH LABORATORY | National Institute for Communicable Diseases of the National Health Laboratory Service | Amoako DG, Everatt J, Kekana D, Scheepers C, Mohale T, Ntuli N, Mahlangu B, Mnguni A, Ismail A, Bhiman JN, Wolter N |
| hCoV-19/SouthAfrica/NICD-N32969/2021 | EPI_ISL_9679693 | 2021-12-08 | AMPATH | National Institute for Communicable Diseases of the National Health Laboratory Service | Amoako DG, Everatt J, Kekana D, Scheepers C, Mohale T, Ntuli N, Mahlangu B, Mnguni A, Ismail A, Bhiman JN, Wolter N |
| hCoV-19/India/MH-RFIP06118/2021 | EPI_ISL_9682825 | 2021-01-11 | BJMC, Pune | RF-IISER Pune | Krishanpal Karmodiya, Aurnab Ghose, Joy Merwin Monteiro, Suvarna Joshi, Rajesh Karyakarte |
| hCoV-19/India/MH-RFIP06126/2021 | EPI_ISL_9682849 | 2021-02-11 | BJMC, Pune | RF-IISER Pune | Krishanpal Karmodiya, Aurnab Ghose, Joy Merwin Monteiro, Suvarna Joshi, Rajesh Karyakarte |
| hCoV-19/India/MH-RFIP06237/2021 | EPI_ISL_9682927 | 2021-11-04 | BJMC, Pune | RF-IISER Pune | Krishanpal Karmodiya, Aurnab Ghose, Joy Merwin Monteiro, Suvarna Joshi, Rajesh Karyakarte |
| hCoV-19/India/MH-RFIP06130/2021 | EPI_ISL_9682934 | 2021-02-11 | BJMC, Pune | RF-IISER Pune | Krishanpal Karmodiya, Aurnab Ghose, Joy Merwin Monteiro, Suvarna Joshi, Rajesh Karyakarte |
| hCoV-19/India/MH-RFIP06222/2021 | EPI_ISL_9682953 | 2021-11-03 | BJMC, Pune | RF-IISER Pune | Krishanpal Karmodiya, Aurnab Ghose, Joy Merwin Monteiro, Suvarna Joshi, Rajesh Karyakarte |
| hCoV-19/India/MH-RFIP06311/2021 | EPI_ISL_9682959 | 2021-12-06 | BJMC, Pune | RF-IISER Pune | Krishanpal Karmodiya, Aurnab Ghose, Joy Merwin Monteiro, Suvarna Joshi, Rajesh Karyakarte |
| hCoV-19/India/TN-CDFD-CO-203/2021 | EPI_ISL_9683518 | 2021-12-26 | Directorate Of Public Health And Preventive Medicine | Centre for DNA Fingerprinting and Diagnostics | Asmita Gupta, Rupin Shelake, Arunkumar Karunanidhi, Vinay Donipadi, Divya Vashisht, Ashwin Dalal, Selvavinayagam Sivaprakasam, Darez Ahamed, Sampath Palani, Raju Sivadoss, Avudaiselvi Rathinasamy, Gurunathan Subramanian, Rajesh Kumar Manivannan, Devi Moni |

| | | | | | |
|---|---|---|---|---|---|
| hCoV-19/India/TN-CDFD-CO-219/2021 | EPI_ISL_9683527 | 2021-12-25 | Directorate Of Public Health And Preventive Medicine | Centre for DNA Fingerprinting and Diagnostics | Asmita Gupta, Rupin Shelake, Arunkumar Karunanidhi, Vinay Donipadi, Divya Vashisht, Ashwin Dalal, Selvavinayagam Sivaprakasam, Darez Ahamed, Sampath Palani, Raju Sivadoss, Avudaiselvi Rathinasamy, Gurunathan Subramanian, Rajesh Kumar Manivannan, Devi Moni |
| hCoV-19/India/TN-CDFD-CO-229/2021 | EPI_ISL_9683535 | 2021-12-27 | Directorate Of Public Health And Preventive Medicine | Centre for DNA Fingerprinting and Diagnostics | Asmita Gupta, Rupin Shelake, Arunkumar Karunanidhi, Vinay Donipadi, Divya Vashisht, Ashwin Dalal, Selvavinayagam Sivaprakasam, Darez Ahamed, Sampath Palani, Raju Sivadoss, Avudaiselvi Rathinasamy, Gurunathan Subramanian, Rajesh Kumar Manivannan, Devi Moni |
| hCoV-19/Canada/ON-PHL-22-05160/2021 | EPI_ISL_9683893 | 2021-11-30 | Public Health Ontario Laboratory | Public Health Ontario Laboratory | Vanessa G Allen, Philip Banh, Yao Chen, Richard de Borja, Alireza Eshaghi, Nahuel Fittipaldi, Christine Frantz, Jonathan B Gubbay, Jennifer L Guthrie, Lawrence Heisler, Esha Joshi, Michael Laszloffy, Aimin Li, Michael CY Li, Dean Maxwell, Sandeep Nagra, S |
| hCoV-19/Canada/ON-PHL-22-05163/2021 | EPI_ISL_9683896 | 2021-11-04 | Public Health Ontario Laboratory | Public Health Ontario Laboratory | Vanessa G Allen, Philip Banh, Yao Chen, Richard de Borja, Alireza Eshaghi, Nahuel Fittipaldi, Christine Frantz, Jonathan B Gubbay, Jennifer L Guthrie, Lawrence Heisler, Esha Joshi, Michael Laszloffy, Aimin Li, Michael CY Li, Dean Maxwell, Sandeep Nagra, S |
| hCoV-19/Namibia/N32118/2021 | EPI_ISL_9694253 | 2021-12-07 | Namibia Institute of Pathology | National Institute for Communicable Diseases of the National Health Laboratory Service | Maphalala G, Amoako DG, Everatt J, Kekana D, Scheepers C, Mohale T, Ntuli N, Mahlangu B, Mnguni A, Ismail A, Bhiman JN, Wolter N |
| hCoV-19/Namibia/N32125/2021 | EPI_ISL_9694256 | 2021-12-08 | Namibia Institute of Pathology | National Institute for Communicable Diseases of the National Health Laboratory Service | Maphalala G, Amoako DG, Everatt J, Kekana D, Scheepers C, Mohale T, Ntuli N, Mahlangu B, Mnguni A, Ismail A, Bhiman JN, Wolter N |
| hCoV-19/Namibia/N32177/2021 | EPI_ISL_9694291 | 2021-12-20 | Namibia Institute of Pathology | National Institute for Communicable Diseases of the National Health Laboratory Service | Maphalala G, Amoako DG, Everatt J, Kekana D, Scheepers C, Mohale T, Ntuli N, Mahlangu B, Mnguni A, Ismail A, Bhiman JN, Wolter N |
| hCoV-19/Nigeria/CV4813/2021 | EPI_ISL_9695253 | 2021-12-08 | Irrua Specialist Teaching Hospital (ISTH) | African Centre of Excellence for Genomics of Infectious Diseases (ACEGID), Redeemer‚Äôs University | Oluniyi, P.E., Olawoye, I.B., Ajogbasile, F.V., Kayode, A.T., Oguzie, J.U., Uwanibe, J.N., Olumade, T.J., Eromon, P.E., Ugwu, C.A., Akano, K.O., Abechi, P.S., Ahmed, M., Chukwu, G., Ope-Ewe, O., Nosamiefan, I., Folarin, O., Happi, C.T. |
| hCoV-19/Nigeria/CV4830/2021 | EPI_ISL_9695271 | 2021-12-12 | Irrua Specialist Teaching Hospital (ISTH) | African Centre of Excellence for Genomics of Infectious Diseases (ACEGID), Redeemer‚Äôs University | Oluniyi, P.E., Olawoye, I.B., Ajogbasile, F.V., Kayode, A.T., Oguzie, J.U., Uwanibe, J.N., Olumade, T.J., Eromon, P.E., Ugwu, C.A., Akano, K.O., Abechi, P.S., Ahmed, M., Chukwu, G., Ope-Ewe, O., Nosamiefan, I., Folarin, O., Happi, C.T. |
| hCoV-19/Nigeria/CV4850/2021 | EPI_ISL_9695289 | 2021-12-13 | Irrua Specialist Teaching Hospital (ISTH) | African Centre of Excellence for Genomics of Infectious Diseases (ACEGID), Redeemer‚Äôs University | Oluniyi, P.E., Olawoye, I.B., Ajogbasile, F.V., Kayode, A.T., Oguzie, J.U., Uwanibe, J.N., Olumade, T.J., Eromon, P.E., Ugwu, C.A., Akano, K.O., Abechi, P.S., Ahmed, M., Chukwu, G., Ope-Ewe, O., Nosamiefan, I., Folarin, O., Happi, C.T. |

| | | | | | |
|---|---|---|---|---|---|
| hCoV-19/Nigeria/CV4940/2021 | EPI_ISL_9695378 | 2021-12-19 | Irrua Specialist Teaching Hospital (ISTH) | African Centre of Excellence for Genomics of Infectious Diseases (ACEGID), Redeemer‚Äôs University | Oluniyi, P.E., Olawoye, I.B., Ajogbasile, F.V., Kayode, A.T., Oguzie, J.U., Uwanibe, J.N., Olumade, T.J., Eromon, P.E., Ugwu, C.A., Akano, K.O., Abechi, P.S., Ahmed, M., Chukwu, G., Ope-Ewe, O., Nosamiefan, I., Folarin, O., Happi, C.T. |
| hCoV-19/Nigeria/ON189/2021 | EPI_ISL_9695421 | 2021-10-18 | Nigeria Centre for Disease Control (NCDC) | African Centre of Excellence for Genomics of Infectious Diseases (ACEGID), Redeemer‚Äôs University | Oluniyi, P.E., Olawoye, I.B., Ajogbasile, F.V., Kayode, A.T., Oguzie, J.U., Uwanibe, J.N., Olumade, T.J., Eromon, P.E., Ugwu, C.A., Akano, K.O., Abechi, P.S., Ahmed, M., Chukwu, G., Ope-Ewe, O., Nosamiefan, I., Folarin, O., Happi, C.T. |
| hCoV-19/India/MH-RFIP05743/2021 | EPI_ISL_9702175 | 2021-11-30 | NIV | RF-IISER Pune | Krishanpal Karmodiya, Aurnab Ghose, Joy Merwin Monteiro |
| hCoV-19/USA/CA-CCPHL-1530/2021 | EPI_ISL_9710442 | 2021-11-06 | Contra Costa Public Health Laboratory | Contra Costa Public Health Laboratory | Contra Costa Public Health Laboratory |
| hCoV-19/Gambia/99215/2021 | EPI_ISL_9712010 | 2021-12-29 | MRCG at LSHTM Genomics lab | MRCG at LSHTM Genomics lab | Abdul Karim sesay, Abdoulie Kanteh, Jarra Manneh, Mariama Kujabi, Bakary Sanyang,Dabiri Damilari, Yaya bah |
| hCoV-19/India/SK-INSACOG-1122500116007/2021 | EPI_ISL_9721021 | 2021-12-29 | INSACOG-Sikkim | National Institute of Biomedical Genomics | Arindam Maitra, Dr. Shrijana Gurung, Nidhan Kumar Biswas, Kaden Zangmu Bhutia, Sreedhar Chinnaswamy¬†Tshering Ongmu Bhutia, Saumitra Das |
| hCoV-19/India/SK-INSACOG-1122500116438/2021 | EPI_ISL_9721786 | 2021-12-29 | INSACOG-Sikkim | National Institute of Biomedical Genomics | Arindam Maitra, Dr. Shrijana Gurung, Nidhan Kumar Biswas, Kaden Zangmu Bhutia, Sreedhar Chinnaswamy¬†Tshering Ongmu Bhutia, Saumitra Das |
| hCoV-19/India/SK-INSACOG-589711611/2021 | EPI_ISL_9721788 | 2021-12-29 | INSACOG-Sikkim | National Institute of Biomedical Genomics | Arindam Maitra, Dr. Shrijana Gurung, Nidhan Kumar Biswas, Kaden Zangmu Bhutia, Sreedhar Chinnaswamy¬†Tshering Ongmu Bhutia, Saumitra Das |
| hCoV-19/India/MH-RFIP08858/2021 | EPI_ISL_9727096 | 2021-11-18 | BJMC | RF-IISER Pune | Krishanpal Karmodiya, Aurnab Ghose, Joy Merwin Monteiro, Suvarna Joshi, Rajesh Karyakarte |
| hCoV-19/Iran/AN0526/2021 | EPI_ISL_9735583 | 2021-12-21 | Shiraz University of Medical Sciences | COVID-19 National Reference Laboratory, Pasteur Institute of Iran | Zahra Ahmadi, Leila badiei-Maryam mahdavi, Mahsa Tavakoli, Zahra Fereydouni, Ali Maleki, Amir Hesam Nemati, Tahmineh Jalali, Mohammad Hassan Pouriayevali, Parastoo Yektay Sanati, Setareh Kashanian, Zahra Hosseini, Ahmad Ghasemi, Tahereh Mohammadi, Sanam A |
| hCoV-19/Iran/AN1121/2021 | EPI_ISL_9735610 | 2021-12-25 | Central Laboratory of East Azerbaijan | COVID-19 National Reference Laboratory, Pasteur Institute of Iran | Zahra Fereydouni, Vahdat Poortahmasebi, Mahsa Tavakoli, Zahra Ahmadi, ¬†Ali Maleki, Amir Hesam Nemati, Tahmineh Jalali, Mohammad Hassan Pouriayevali, Parastoo Yektay Sanati, Setareh Kashanian, Zahra Hosseini, Ahmad Ghasemi, Tahereh Mohammadi, Sanam Azad-M |
| hCoV-19/Belgium/UGent-12505/2021 | EPI_ISL_9753277 | 2021-05-23 | Lab voor klinische biologie | Lab voor klinische biologie | May-Linh Truong, Hannelore Hamerlinck, Bruno Verhasselt |
| hCoV-19/Brazil/PR-NVBS12112GENOV701807696167/2021 | EPI_ISL_9754623 | 2021-12-28 | DASA | DASA | Rodrigo Guarischi, Paulo Pierry, Adriano Bonaldi, Cristina Oliveira, Bianca Cota, Luciane Sussuchi, Camila Romano, Angelica Hristov, Lidia Yamamoto, Rodrigo Salazar, Annelise Lopes, Jose Levi |

| | | | | | |
|---|---|---|---|---|---|
| hCoV-19/USA/WI-WSLH-2203617/2021 | EPI_ISL_9755701 | 2021-01-07 | Wisconsin State Laboratory of Hygiene Communicable Disease Division | Wisconsin State Laboratory of Hygiene Communicable Disease Division | Christopher R. Jossart, Abigail C. Shockey, Alicia J. Mooney, Erika M. Hanson, Tonya Danz, Richard Griesser, Kelsey R. Florek |
| hCoV-19/Canada/BC-BCCDC-326797/2021 | EPI_ISL_9759797 | 2021-12-30 | British Columbia Centre For Disease Control | BCCDC Public Health Laboratory | Prystajecky Natalie, Linda Hoang, Dan Fornika, John Tyson, Shannon Russell, Kim Macdonald, Kimia Kamelian, James Zlosnik, Ana Pacagnella, Corrinne Ng, Loretta Janz, Robert Azana, Mel Krajden |
| hCoV-19/Canada/BC-BCCDC-327374/2021 | EPI_ISL_9760152 | 2021-12-30 | British Columbia Centre For Disease Control | BCCDC Public Health Laboratory | Prystajecky Natalie, Linda Hoang, Dan Fornika, John Tyson, Shannon Russell, Kim Macdonald, Kimia Kamelian, James Zlosnik, Ana Pacagnella, Corrinne Ng, Loretta Janz, Robert Azana, Mel Krajden |
| hCoV-19/USA/OR-UW-21123002431/2021 | EPI_ISL_9760615 | 2021-12-30 | UW Virology Lab | UW Virology Lab | Pavitra Roychoudhury, Shah Mohamed Bakhash, Pooneh Hajian, Seffir T. Wendm, Isabel Arnould, Ricardo Perez, Hong Xie, Sean Ellis, Saraswathi Sathees, Meei-Li Huang, Keith R Jerome, Alexander Greninger |
| hCoV-19/USA/AZ-TG1216064/2021 | EPI_ISL_9769955 | 2021-10-28 | TGen North | TGen North | Jolene Bowers, Heather Centner, Brett Van Tassel, Chris French, Hayley Yaglom, Darrin Lemmer, Dave Engelthaler |
| hCoV-19/Norway/OUS-12779/2021 | EPI_ISL_9793505 | 2021-08-19 | Oslo University Hospital, Department of Microbiology | Norwegian Institute of Public Health, Department of Virology | Mona Holberg-Petersen, Lise Andresen, Cathrine Fladeby, Mariann Nilsen, Teodora Plamenova Ribarska, P√•l Marius Bj√∏rnstad, Gregor D. Gilfillan, Arvind Yegambaram Meenakshi Sundaram, Kathrine Stene-Johansen, Kamilla Heddeland Instefjord, Hilde Elshaug, Ga |
| hCoV-19/Brazil/CE-FIOCRUZ-54090CE/2021 | EPI_ISL_9794298 | 2021-12-30 | UPA I UNIDADE DE PRONTO ATENDIMENTO | Analytical Competence Molecular Epidemiology Lab/ACME, Oswaldo Cruz Foundation, Ceara (FIOCRUZ CE) | Fabio Miyajima, Veridiana Pessoa Miyajima, Fernando Braga Stehling, Thais de Oliveira Costa, Thais Ferreira de Oliveira, Joaquim Cesar do Nascimento Sousa Junior, Jamille Maria Mendes Bezerra, Ticiane Cavalcante de Souza, Carlos Leonardo de Aragao Araujo, |
| hCoV-19/Brazil/SP-SEQ07638/2021 | EPI_ISL_9795130 | 2021-10-18 | DiviSao de Vigilancia Joao Luiz Perez Vigilancia Epidemiologica - Buritima - Labmovel | Instituto Butantan | Dimas Tadeu Covas, Antonio Jorge Martins, Claudia Renata dos Santos Barros, David Schlesinger, Debora Botequio Moretti, Elaine Cristina Marqueze, Elaine Vieira Santos, Evandra Strazza Rodrigues, Heidge Fukumasu, Jayme Augusto de Souza-Neto, Luiz Alcantara |
| hCoV-19/USA/MN-MDH-22317/2021 | EPI_ISL_9803031 | 2021-12-21 | LAKEWOOD HEALTH SYSTEM - STAPLES CLINIC | Minnesota Department of Health, Public Health Laboratory | Jacob Garfin, Matt Plumb, Kelly Pung, Elizabeth Horn, Sarah Namugenyi, Alyssa Mondelli, Haejin Hwang, and Xiong Wang |
| hCoV-19/USA/CA-SEARCH-66508/2021 | EPI_ISL_9806799 | 2021-11-29 | EXCITE Lab | Andersen lab at Scripps Research | Charles Chiu, Eric Delwart, Alicia Sotomayor-Gonzalez, Venice Servellita, San Francisco Southeast Wastewater Treatment Plant, Mel Leong Treatment Plant at San Francisco Airport (SFO), Smruthi Karthikeyan, Greg Humphrey, Sawyer Farmer, Abigail Schnapper, |
| hCoV-19/Nigeria/NCDC-NR1202/2021 | EPI_ISL_9829269 | 2021-12-15 | IRUAH SPECIALIST HOSPITAL | National Reference Laboratory, Nigeria Centre for Disease Control | Dr Ndodo Nnaemeka, Olusola Anuoluwapo Akanbi, Oyeronke Ayansola, Chimaobi Chukwu, Dr Omoare Adesuyi, Anthony Ahumibe, Catherine Okoi, Nwando Mba, Dr Ifedayo Adetifa |

| | | | | | |
|---|---|---|---|---|---|
| hCoV-19/Malaysia/TIDREC-NSC661/2021 | EPI_ISL_9829829 | 2021-12-23 | IMR | Tropical Infectious Diseases Research & Education Centre (TIDREC), Universiti Malaya | Kim-Kee Tan, Jia-Yi Tan, Jo-Ern Wong, Siti-Sarah Nor,Äôe, Che-Norainon Yaacob, Mulya-Mustika-Sari Zulkifli, AsmaAnati CheMatSeri, Nur-Hidayana Mahfodz, Noor Syahida Azizan, Sazaly AbuBakar |
| hCoV-19/England/LOND-YYBDAET/2021 | EPI_ISL_9832672 | 2021-09-28 | University College London, Great Ormond Street Hospital for Children NHS Foundation Trust, Imperial College Healthcare NHS Trust | COVID-19 Genomics UK (COG-UK) Consortium | Sergi Castellano, Rachel Williams, Mark Kristiansen, Sunando Roy, Charlotte Williams, Nadua Bayzid, Marius Cotic, Helena Tutill, Patricia Dyal, Judith Breuer |
| hCoV-19/HongKong/HKU-220213-154/2021 | EPI_ISL_9845567 | 2021-11-17 | Department of Microbiology, The University of Hong Kong | Department of Microbiology, The University of Hong Kong | Kelvin K.W. To, Kwok-Yung Yuen |
| hCoV-19/HongKong/HKU-220213-269/2021 | EPI_ISL_9845679 | 2021-12-24 | Department of Microbiology, The University of Hong Kong | Department of Microbiology, The University of Hong Kong | Kelvin K.W. To, Kwok-Yung Yuen |
| hCoV-19/HongKong/HKU-220213-294/2021 | EPI_ISL_9845703 | 2021-12-26 | Department of Microbiology, The University of Hong Kong | Department of Microbiology, The University of Hong Kong | Kelvin K.W. To, Kwok-Yung Yuen |
| hCoV-19/HongKong/HKU-220213-304/2021 | EPI_ISL_9845713 | 2021-12-27 | Department of Microbiology, The University of Hong Kong | Department of Microbiology, The University of Hong Kong | Kelvin K.W. To, Kwok-Yung Yuen |
| hCoV-19/Ghana/TRV/30014/2021 | EPI_ISL_9848204 | 2021-12-13 | Kumasi Centre for Collaborative Research in Tropical Medicine | Kumasi Centre for Collaborative Research in Tropical Medicine | Augustina Sylverken, Michael Owusu, Sherihane Aryeetey, Jesse Asamoah,Rita Ziem, Ernest Adankwa,James Osei-Mensah, Franklin Asiedu Bekoe, Norman Nausch, Sascha Knauf, Thorsten Thye, Simone Luert, Michele Kolasa, Victoria Crede,Richard Phillips |
| hCoV-19/Thailand/NIC_SRI_SEQ01603/2021 | EPI_ISL_9872444 | 2021-01-24 | Regional Medical Sciences Center 4 Saraburi | National Institute of Health, Department of Medical Sciences, Ministry of Public Health, Thailand | Pilailuk Okada; Siripaporn Phuygun; Nuttida Thongpramul; Pakorn Piromtong; Natchaya¬† Khiadsang; Thanutsapa Thanadachakul; Sirikanda Wimol; Suratchana Mitrat; Sunthareeya Waicharoen; Archawin Rojanawiwat |
| hCoV-19/SouthAfrica/NCV577/2020 | EPI_ISL_9877973 | 2020-12-18 | National Health Laboratory Services, Virology | National Health Laboratory Services, Virology | Kathleen Subramoney, Nkhensani Mtileni, Avani Bharuthram, Ashlyn S. C. Davis, Florette K. Treurnicht |
| hCoV-19/SouthAfrica/NCV575/2020 | EPI_ISL_9877975 | 2020-12-19 | National Health Laboratory Services, Virology | National Health Laboratory Services, Virology | Kathleen Subramoney, Nkhensani Mtileni, Avani Bharuthram, Ashlyn S. C. Davis, Florette K. Treurnicht |
| hCoV-19/Coted'Ivoire/347325-cv025/2021 | EPI_ISL_9888101 | 2021-07-28 | Environment and Health Department, Institut Pasteur of C√¥te d'Ivoire, Abidjan, C√¥te d'Ivoire | Molecular Genetics Platform, Institut Pasteur of C√¥te d'Ivoire, Abidjan, C√¥te d'Ivoire | Aristide Berenger Ako AKO; Bernadette Tchepe Flore Diplo; Egomli Stanislas ASSOHOUN; Antoine Marie-David N'golo COULIBALY; Tata Gnire Safiatou COULIBALY; Jean-Ren√© Quassi ACQUAH; Djeda Franck Arnaud GNAHORE; Herv√© Alberic ADJE KADJO; Haris ONYWERA; Adey |
| hCoV-19/Coted'Ivoire/225306-cv236/2021 | EPI_ISL_9894409 | 2021-03-01 | Department of Epidemic Viruses, Institut Pasteur of C√¥te d'Ivoire, Abidjan, C√¥te d'Ivoire | Molecular Genetics Platform, Institut Pasteur of C√¥te d'Ivoire, Abidjan, C√¥te d'Ivoire | Aristide Berenger Ako AKO; Bernadette Tchepe Flore Diplo; Egomli Stanislas ASSOHOUN; Antoine Marie-David N'golo COULIBALY; Tata Gnire Safiatou COULIBALY; Jean-Ren√© Quassi ACQUAH; Djeda Franck Arnaud GNAHORE; Herv√© Alberic ADJE KADJO; Mireille DOSSO |
| hCoV-19/Coted'Ivoire/225263-cv242/2021 | EPI_ISL_9894414 | 2021-03-01 | Department of Epidemic Viruses, Institut Pasteur of C√¥te d'Ivoire, Abidjan, C√¥te d'Ivoire | Molecular Genetics Platform, Institut Pasteur of C√¥te d'Ivoire, Abidjan, C√¥te d'Ivoire | Aristide Berenger Ako AKO; Bernadette Tchepe Flore Diplo; Egomli Stanislas ASSOHOUN; Antoine Marie-David N'golo COULIBALY; Tata Gnire Safiatou COULIBALY; Jean-Ren√© Quassi ACQUAH; Djeda Franck Arnaud GNAHORE; Herv√© Alberic ADJE KADJO; Mireille DOSSO |

| | | | | | |
|---|---|---|---|---|---|
| hCoV-19/Coted'Ivoire/225327-cv244/2021 | EPI_ISL_9894416 | 2021-03-01 | Department of Epidemic Viruses, Institut Pasteur of C√¥te d'Ivoire, Abidjan, C√¥te d'Ivoire | Molecular Genetics Platform, Institut Pasteur of C√¥te d'Ivoire, Abidjan, C√¥te d'Ivoire | Aristide Berenger Ako AKO; Bernadette Tchepe Flore Diplo; Egomli Stanislas ASSOHOUN; Antoine Marie-David N'golo COULIBALY; Tata Gnire Safiatou COULIBALY; Jean-Ren√© Quassi ACQUAH; Djeda Franck Arnaud GNAHORE; Herv√© Alberic ADJE KADJO; Mireille DOSSO |
| hCoV-19/Coted'Ivoire/225265-cv246/2021 | EPI_ISL_9894418 | 2021-03-01 | Department of Epidemic Viruses, Institut Pasteur of C√¥te d'Ivoire, Abidjan, C√¥te d'Ivoire | Molecular Genetics Platform, Institut Pasteur of C√¥te d'Ivoire, Abidjan, C√¥te d'Ivoire | Aristide Berenger Ako AKO; Bernadette Tchepe Flore Diplo; Egomli Stanislas ASSOHOUN; Antoine Marie-David N'golo COULIBALY; Tata Gnire Safiatou COULIBALY; Jean-Ren√© Quassi ACQUAH; Djeda Franck Arnaud GNAHORE; Herv√© Alberic ADJE KADJO; Mireille DOSSO |
| hCoV-19/Coted'Ivoire/225268-cv248/2021 | EPI_ISL_9894420 | 2021-03-01 | Department of Epidemic Viruses, Institut Pasteur of C√¥te d'Ivoire, Abidjan, C√¥te d'Ivoire | Molecular Genetics Platform, Institut Pasteur of C√¥te d'Ivoire, Abidjan, C√¥te d'Ivoire | Aristide Berenger Ako AKO; Bernadette Tchepe Flore Diplo; Egomli Stanislas ASSOHOUN; Antoine Marie-David N'golo COULIBALY; Tata Gnire Safiatou COULIBALY; Jean-Ren√© Quassi ACQUAH; Djeda Franck Arnaud GNAHORE; Herv√© Alberic ADJE KADJO; Mireille DOSSO |
| hCoV-19/India/RJ-SMS-ICMR-INSACOG-TS-1150/2021 | EPI_ISL_9905686 | 2021-12-24 | SMS MEDICAL COLLEGE,JAIPUR | NIV Influenza | Bharti Malhotra, Swati Gautam, Pratibha Sharma, Himanshu sharma,Dinesh parsoya, Farah Deeba, Neha Bhomia, Nita pal, Varsha Potdar, Pragya D Yadav, Nivedita Gupta, Sudhir Bhandari |
| hCoV-19/India/RJ-SMS-ICMR-INSACOG-TS-1333/2021 | EPI_ISL_9905759 | 2021-12-26 | SMS MEDICAL COLLEGE,JAIPUR | NIV Influenza | Bharti Malhotra, Swati Gautam, Pratibha Sharma, Himanshu sharma,Dinesh parsoya, Farah Deeba, Neha Bhomia, Nita pal, Varsha Potdar, Pragya D Yadav, Nivedita Gupta, Sudhir Bhandari |
| hCoV-19/India/RJ-SMS-ICMR-INSACOG-TS-1338/2021 | EPI_ISL_9905763 | 2021-12-26 | SMS MEDICAL COLLEGE,JAIPUR | NIV Influenza | Bharti Malhotra, Swati Gautam, Pratibha Sharma, Himanshu sharma,Dinesh parsoya, Farah Deeba, Neha Bhomia, Nita pal, Varsha Potdar, Pragya D Yadav, Nivedita Gupta, Sudhir Bhandari |
| hCoV-19/India/RJ-SMS-ICMR-INSACOG-TS-1697/2021 | EPI_ISL_9905869 | 2021-12-30 | SMS MEDICAL COLLEGE,JAIPUR | NIV Influenza | Bharti Malhotra, Swati Gautam, Pratibha Sharma, Himanshu sharma,Dinesh parsoya, Farah Deeba, Neha Bhomia, Nita pal, Varsha Potdar, Pragya D Yadav, Nivedita Gupta, Sudhir Bhandari |
| hCoV-19/India/RJ-SMS-ICMR-INSACOG-TS-1787/2021 | EPI_ISL_9905916 | 2021-12-30 | SMS MEDICAL COLLEGE,JAIPUR | NIV Influenza | Bharti Malhotra, Swati Gautam, Pratibha Sharma, Himanshu sharma,Dinesh parsoya, Farah Deeba, Neha Bhomia, Nita pal, Varsha Potdar, Pragya D Yadav, Nivedita Gupta, Sudhir Bhandari |
| hCoV-19/Coted'Ivoire/211157-cv255/2021 | EPI_ISL_9906189 | 2021-02-18 | Department of Epidemic Viruses, Institut Pasteur of C√¥te d'Ivoire, Abidjan, C√¥te d'Ivoire | Molecular Genetics Platform, Institut Pasteur of C√¥te d'Ivoire, Abidjan, C√¥te d'Ivoire | Aristide Berenger Ako AKO; Bernadette Tchepe Flore Diplo; Egomli Stanislas ASSOHOUN; Antoine Marie-David N'golo COULIBALY; Tata Gnire Safiatou COULIBALY; Jean-Ren√© Quassi ACQUAH; Djeda Franck Arnaud GNAHORE; Herv√© Alberic ADJE KADJO; Mireille DOSSO |
| hCoV-19/Coted'Ivoire/211162-cv256/2021 | EPI_ISL_9906190 | 2021-02-18 | Department of Epidemic Viruses, Institut Pasteur of C√¥te d'Ivoire, Abidjan, C√¥te d'Ivoire | Molecular Genetics Platform, Institut Pasteur of C√¥te d'Ivoire, Abidjan, C√¥te d'Ivoire | Aristide Berenger Ako AKO; Bernadette Tchepe Flore Diplo; Egomli Stanislas ASSOHOUN; Antoine Marie-David N'golo COULIBALY; Tata Gnire Safiatou COULIBALY; Jean-Ren√© Quassi ACQUAH; Djeda Franck Arnaud GNAHORE; Herv√© Alberic ADJE KADJO; Mireille DOSSO |

| | | | | | |
|---|---|---|---|---|---|
| hCoV-19/Coted'Ivoire/211213-cv260/2021 | EPI_ISL_9906194 | 2021-02-18 | Department of Epidemic Viruses, Institut Pasteur of CV¥te d'Ivoire, Abidjan, CV¥te d'Ivoire | Molecular Genetics Platform, Institut Pasteur of CV¥te d'Ivoire, Abidjan, CV¥te d'Ivoire | Aristide Berenger Ako AKO; Bernadette Tchepe Flore Diplo; Egomli Stanislas ASSOHOUN; Antoine Marie-David N'golo COULIBALY; Tata Gnire Safiatou COULIBALY; Jean-RenV© Quassi ACQUAH; Djeda Franck Arnaud GNAHORE; HervV© Alberic ADJE KADJO; Mireille DOSSO |
| hCoV-19/Coted'Ivoire/210786-cv265/2021 | EPI_ISL_9906198 | 2021-02-18 | Department of Epidemic Viruses, Institut Pasteur of CV¥te d'Ivoire, Abidjan, CV¥te d'Ivoire | Molecular Genetics Platform, Institut Pasteur of CV¥te d'Ivoire, Abidjan, CV¥te d'Ivoire | Aristide Berenger Ako AKO; Bernadette Tchepe Flore Diplo; Egomli Stanislas ASSOHOUN; Antoine Marie-David N'golo COULIBALY; Tata Gnire Safiatou COULIBALY; Jean-RenV© Quassi ACQUAH; Djeda Franck Arnaud GNAHORE; HervV© Alberic ADJE KADJO; Mireille DOSSO |
| hCoV-19/Coted'Ivoire/225188-cv277/2021 | EPI_ISL_9906208 | 2021-03-01 | Department of Epidemic Viruses, Institut Pasteur of CV¥te d'Ivoire, Abidjan, CV¥te d'Ivoire | Molecular Genetics Platform, Institut Pasteur of CV¥te d'Ivoire, Abidjan, CV¥te d'Ivoire | Aristide Berenger Ako AKO; Bernadette Tchepe Flore Diplo; Egomli Stanislas ASSOHOUN; Antoine Marie-David N'golo COULIBALY; Tata Gnire Safiatou COULIBALY; Jean-RenV© Quassi ACQUAH; Djeda Franck Arnaud GNAHORE; HervV© Alberic ADJE KADJO; Mireille DOSSO |
| hCoV-19/USA/UT-UPHL-220213537366/2021 | EPI_ISL_9919284 | 2021-12-27 | Utah Public Health Laboratory | Utah Public Health Laboratory | Erin L. Young, Kelly F. Oakeson, Pooja Gupta, John Arnn, Tom Iverson, Olinto Linares-Perdomo |
| hCoV-19/mink/USA/UT-CDC-4455676-001/2021 | EPI_ISL_9936929 | 2021-10-09 | One Health Office, Centers for Disease Control and Prevention | Genomics and Discovery, Respiratory Viruses Branch, Division of Viral Diseases, Centers for Disease Control and Prevention | Yan Li, Anna Kelleher, Jing Zhang, Brian Lynch, Ying Tao, Ausaf Ahmad, Paige Bunkley, Lauren Claire Godino, Owen Herzegh, Anna Uehara, Peter Cook, Han Jia Justin Ng, Caitlin M. Cossaboom, Natalie M. Wendling, Jonathan Towner, Brian Amman, Reid Harvey, Han |
| hCoV-19/mink/USA/UT-CDC-4455677-001/2021 | EPI_ISL_9936930 | 2021-10-09 | One Health Office, Centers for Disease Control and Prevention | Genomics and Discovery, Respiratory Viruses Branch, Division of Viral Diseases, Centers for Disease Control and Prevention | Yan Li, Anna Kelleher, Jing Zhang, Brian Lynch, Ying Tao, Ausaf Ahmad, Paige Bunkley, Lauren Claire Godino, Owen Herzegh, Anna Uehara, Peter Cook, Han Jia Justin Ng, Caitlin M. Cossaboom, Natalie M. Wendling, Jonathan Towner, Brian Amman, Reid Harvey, Han |
| hCoV-19/Australia/NSW-ICPMR-9654/2021 | EPI_ISL_9951667 | 2021-09-28 | Medlab Pathology | Clinical Pathology and Medical Research; Westmead Hospital; University of Sydney | Arnott A., Draper J., Gall M., Martinez E., Rockett R., Sintchenko V., on behalf of ICPMR |
| hCoV-19/SouthAfrica/NICD-N28962/2021 | EPI_ISL_9962672 | 2021-12-09 | AMPATH | National Institute for Communicable Diseases of the National Health Laboratory Service | Amoako DG, Everatt J, Kekana D, Scheepers C, Mohale T, Ntuli N, Mahlangu B, Mnguni A, Ismail A, Bhiman JN, Wolter N |
| hCoV-19/SouthAfrica/NICD-N32471/2021 | EPI_ISL_9962791 | 2021-12-30 | PORT ELIZABETH LABORATORY | National Institute for Communicable Diseases of the National Health Laboratory Service | Amoako DG, Everatt J, Kekana D, Scheepers C, Mohale T, Ntuli N, Mahlangu B, Mnguni A, Ismail A, Bhiman JN, Wolter N |
| hCoV-19/SouthAfrica/NICD-N32489/2021 | EPI_ISL_9962800 | 2021-12-29 | PORT ELIZABETH LABORATORY | National Institute for Communicable Diseases of the National Health Laboratory Service | Amoako DG, Everatt J, Kekana D, Scheepers C, Mohale T, Ntuli N, Mahlangu B, Mnguni A, Ismail A, Bhiman JN, Wolter N |
| hCoV-19/SouthAfrica/NICD-N32495/2021 | EPI_ISL_9962806 | 2021-12-08 | KIMBERLEY LABORATORY | National Institute for Communicable Diseases of the National Health Laboratory Service | Amoako DG, Everatt J, Kekana D, Scheepers C, Mohale T, Ntuli N, Mahlangu B, Mnguni A, Ismail A, Bhiman JN, Wolter N |
| hCoV-19/SouthAfrica/NICD-N32561/2021 | EPI_ISL_9962823 | 2021-12-18 | PORT ELIZABETH LABORATORY | National Institute for Communicable Diseases of the National Health Laboratory Service | Amoako DG, Everatt J, Kekana D, Scheepers C, Mohale T, Ntuli N, Mahlangu B, Mnguni A, Ismail A, Bhiman JN, Wolter N |

| | | | | | |
|---|---|---|---|---|---|
| hCoV-19/SouthAfrica/NICD-N32568/2021 | EPI_ISL_9962830 | 2021-12-20 | PORT ELIZABETH LABORATORY | National Institute for Communicable Diseases of the National Health Laboratory Service | Amoako DG, Everatt J, Kekana D, Scheepers C, Mohale T, Ntuli N, Mahlangu B, Mnguni A, Ismail A, Bhiman JN, Wolter N |
| hCoV-19/SouthAfrica/NICD-N32699/2021 | EPI_ISL_9962912 | 2021-12-30 | AMPATH | National Institute for Communicable Diseases of the National Health Laboratory Service | Amoako DG, Everatt J, Kekana D, Scheepers C, Mohale T, Ntuli N, Mahlangu B, Mnguni A, Ismail A, Bhiman JN, Wolter N |
| hCoV-19/SouthAfrica/NICD-N32722/2021 | EPI_ISL_9962922 | 2021-12-26 | AMPATH | National Institute for Communicable Diseases of the National Health Laboratory Service | Amoako DG, Everatt J, Kekana D, Scheepers C, Mohale T, Ntuli N, Mahlangu B, Mnguni A, Ismail A, Bhiman JN, Wolter N |
| hCoV-19/SouthAfrica/NICD-N32723/2021 | EPI_ISL_9962923 | 2021-12-26 | AMPATH | National Institute for Communicable Diseases of the National Health Laboratory Service | Amoako DG, Everatt J, Kekana D, Scheepers C, Mohale T, Ntuli N, Mahlangu B, Mnguni A, Ismail A, Bhiman JN, Wolter N |
| hCoV-19/SouthAfrica/NICD-N32910/2021 | EPI_ISL_9962936 | 2021-12-13 | AMPATH | National Institute for Communicable Diseases of the National Health Laboratory Service | Amoako DG, Everatt J, Kekana D, Scheepers C, Mohale T, Ntuli N, Mahlangu B, Mnguni A, Ismail A, Bhiman JN, Wolter N |
| hCoV-19/SouthAfrica/NICD-N33250/2021 | EPI_ISL_9963045 | 2021-12-20 | AMPATH | National Institute for Communicable Diseases of the National Health Laboratory Service | Amoako DG, Everatt J, Kekana D, Scheepers C, Mohale T, Ntuli N, Mahlangu B, Mnguni A, Ismail A, Bhiman JN, Wolter N |
| hCoV-19/SouthAfrica/NICD-N33376/2021 | EPI_ISL_9963107 | 2021-12-15 | AMPATH | National Institute for Communicable Diseases of the National Health Laboratory Service | Amoako DG, Everatt J, Kekana D, Scheepers C, Mohale T, Ntuli N, Mahlangu B, Mnguni A, Ismail A, Bhiman JN, Wolter N |
| hCoV-19/SouthAfrica/NICD-N33402/2021 | EPI_ISL_9963122 | 2021-12-26 | AMPATH | National Institute for Communicable Diseases of the National Health Laboratory Service | Amoako DG, Everatt J, Kekana D, Scheepers C, Mohale T, Ntuli N, Mahlangu B, Mnguni A, Ismail A, Bhiman JN, Wolter N |
| hCoV-19/SouthAfrica/NICD-N33496/2021 | EPI_ISL_9963142 | 2021-12-19 | AMPATH | National Institute for Communicable Diseases of the National Health Laboratory Service | Amoako DG, Everatt J, Kekana D, Scheepers C, Mohale T, Ntuli N, Mahlangu B, Mnguni A, Ismail A, Bhiman JN, Wolter N |
| hCoV-19/SouthAfrica/NICD-N33524/2021 | EPI_ISL_9963159 | 2021-12-19 | AMPATH | National Institute for Communicable Diseases of the National Health Laboratory Service | Amoako DG, Everatt J, Kekana D, Scheepers C, Mohale T, Ntuli N, Mahlangu B, Mnguni A, Ismail A, Bhiman JN, Wolter N |
| hCoV-19/SouthAfrica/NICD-N33555/2021 | EPI_ISL_9963175 | 2021-12-23 | AMPATH | National Institute for Communicable Diseases of the National Health Laboratory Service | Amoako DG, Everatt J, Kekana D, Scheepers C, Mohale T, Ntuli N, Mahlangu B, Mnguni A, Ismail A, Bhiman JN, Wolter N |
| hCoV-19/SouthAfrica/1063102/2021 | EPI_ISL_9977188 | 2021-11-30 | Ndlovu Laboratories | Ndlovu Laboratories | Anna Mabaso, Anele Mdunyelwa, Khamusi Mlambo, Mathapelo Maphanga, Chijioke Umunnakwe |
| hCoV-19/Montserrat/120966/2021 | EPI_ISL_9982743 | 2021-01-31 | Glendon Hospital Laboratory | CARPHA | Risha Singh on behalf of CARPHA |
| hCoV-19/Norway/35740/2021 | EPI_ISL_9984371 | 2021-12-26 | Nordland Hospital - Bodo, Laboratory Department, Molecular Biology Unit | Norwegian Institute of Public Health, Department of Virology | Kathrine Stene-Johansen, Kamilla Heddeland Instefjord, Hilde Elshaug, Garcia Llorente Ignacio, Jon Br√•te, Engebretsen Serina Beate, Pedersen Benedikte Nevjen, Line Victoria Moen, Hilde Nordby Falkenhaug, Debech Nadia, Atiya R Ali, Marie Paulsen Madsen, R |
